# Cross-Ancestry Investigation of Venous Thromboembolism Genomic Predictors

**DOI:** 10.1101/2022.03.04.22271003

**Authors:** Florian Thibord, Derek Klarin, Jennifer A. Brody, Ming-Huei Chen, Michael G. Levin, Daniel I. Chasman, Ellen L. Goode, Kristian Hveem, Maris Teder-Laving, Angel Martinez-Perez, Dylan Aïssi, Delphine Daian-Bacq, Kaoru Ito, Pradeep Natarajan, Pamela L. Lutsey, Girish N. Nadkarni, Gabriel Cuellar-Partida, Brooke N. Wolford, Jack W. Pattee, Charles Kooperberg, Sigrid K. Braekkan, Ruifang Li-Gao, Noemie Saut, Corriene Sept, Marine Germain, Renae L. Judy, Kerri L. Wiggins, Darae Ko, Christopher O’Donnell, Kent D. Taylor, Franco Giulianini, Mariza De Andrade, Therese H. Nøst, Anne Boland, Jean-Philippe Empana, Satoshi Koyama, Thomas Gilliland, Ron Do, Xin Wang, Wei Zhou, Jose Manuel Soria, Juan Carlos Souto, Nathan Pankratz, Jeffery Haessler, Kristian Hindberg, Frits R. Rosendaal, Constance Turman, Robert Olaso, Rachel L. Kember, Traci M. Bartz, Julie A. Lynch, Susan R. Heckbert, Sebastian M. Armasu, Ben Brumpton, David M. Smadja, Xavier Jouven, Issei Komuro, Katharine Clapham, Ruth J.F. Loos, Cristen Willer, Maria Sabater-Lleal, James S. Pankow, Alexander P. Reiner, Vania M. Morelli, Paul M. Ridker, Astrid van Hylckama Vlieg, Jean-François Deleuze, Peter Kraft, Daniel J. Rader, Barbara McKnight, Global Biobank Meta-Analysis Initiative, Estonian Biobank Research Team, 23andMe Research Team, Kyung Min Lee, Bruce M. Psaty, Anne Heidi Skogholt, Joseph Emmerich, Pierre Suchon, Biobank Japan, Stephen S. Rich, Ha My T. Vy, Weihong Tang, Rebecca D. Jackson, John-Bjarne Hansen, Pierre-Emmanuel Morange, Christopher Kabrhel, David-Alexandre Trégouët, Scott Damrauer, Andrew D. Johnson, Nicholas L. Smith

## Abstract

Venous thromboembolism (VTE) is a complex disease with environmental and genetic determinants. We present new cross-ancestry meta-analyzed genome-wide association study (GWAS) results from 30 studies, with replication of novel loci and their characterization through *in silico* genomic interrogations. In our initial genetic discovery effort that included 55,330 participants with VTE (47,822 European, 6,320 African, and 1,188 Hispanic ancestry), we identified 48 novel associations of which 34 replicated after correction for multiple testing. In our combined discovery-replication analysis (81,669 VTE participants) and ancestry-stratified meta-analyses (European, African and Hispanic), we identified another 44 novel associations, which are new candidate VTE-associated loci requiring replication. In total, across all GWAS meta-analyses, we identified 135 independent genomic loci significantly associated with VTE risk. We also identified 31 novel transcript associations in transcriptome-wide association studies and 8 novel candidate genes with protein QTL Mendelian randomization analyses. *In silico* interrogations of hemostasis and hematology traits and a large phenome-wide association analysis of the 135 novel GWAS loci provided insights to biological pathways contributing to VTE, indicating that some loci may contribute to VTE through well-characterized coagulation pathways while others provide new data on the role of hematology traits, particularly platelet function. Many of the replicated loci are outside of known or currently hypothesized pathways to thrombosis. In summary, these findings highlight new pathways to thrombosis and provide novel molecules that may be useful in the development of antithrombosis treatments with reduced risk of bleeds.

## INTRODUCTION

Venous thrombosis is a vascular event resulting from an imbalance in the regulation of hemostasis, with subsequent pathologic coagulation and vascular thrombosis formation. Clinically, venous thrombosis can manifest as deep vein thrombosis (DVT), when occurring in the deep veins primarily of the legs and trunk, or as a pulmonary embolism (PE), when the thrombus embolizes and obstructs the pulmonary arteries. Collectively, these events are known as venous thromboembolism (VTE), a life-threatening condition with an incidence of 1-2 events per 1,000 person-years.^1–3^ VTE is a complex disease with both environmental and genetic determinants. Family studies, candidate-gene approaches, and early genome-wide association studies (GWAS) primarily identified genetic risk factors in loci with well characterized effects on coagulation (*F2, F5, F11, FGG, ABO, SERPINC1, PROCR, PROC, PROS1*), supporting current therapeutic strategies that mainly target the coagulation cascade.^4–8^ In recent years, larger GWAS meta-analyses revealed unanticipated loci, such as *SLC44A2*,^9^ which was later characterized as a choline transporter involved in platelet activation,^10^ and in the adhesion and activation of neutrophils.^11,12^ Thus, genetic associations with VTE in larger and more diverse populations may uncover new biological pathways and molecular events contributing to the disease and potentially help identify novel targets for treatment. Most recently, 2 large efforts involving up to 30,000 VTE cases, led by the International Network Against Venous Thrombosis (INVENT) consortium ^13^ and the Million Veteran Program^14^ (MVP), identified up to 43 genetic loci associated with VTE. To expand discovery of novel VTE risk loci, we conducted a large, cross-ancestry GWAS meta-analysis involving more than 80,000 VTE cases, along with a replication of novel loci and their characterization through downstream analyses.

## METHODS

### Design and Study Participants

The current cross-ancestry GWAS meta-analysis is comprised of new analyses of data from 13 studies, including the Department of Veterans Affairs Million Veteran Program (MVP, version 4),^14^ UK Biobank (UKB), ^15,16^ FinnGen (freeze 5), Estonian Biobank (EGP),^17^ Biobank Japan (BBJ),^18^ Mass General Brigham biobank (MGB),^19^ BioMe, Penn Medicine BioBank (UPenn), FARIVE,^20^ MARTHA12,^21^ RETROVE,^22^ Multi-Ethnic Study of Atherosclerosis (MESA),^23^ and GAIT2,^24,25^ as well as previously published data from the INVENT consortium, a 17 study analysis of prospective cohorts and case-control data (designated INVENT-2019).^13^ A detailed description of participating studies is provided in supplemental **Table S1**.

The study design (see **Figure 1**) included a discovery meta-analysis of GWAS summary data from 4 consortium/studies (INVENT-2019, MVP, FinnGen, EGP) followed by a replication of discovery loci that exceed the genome-wide significance threshold (P < 5.00 × 10^−8^). Included in the replication were the remaining 10 participating studies plus 2 external collaborations that provided association results for the queried variants: 23andMe^26^ and Global Biobank Meta-Analysis Initiative (GBMI), limiting data to non-overlapping studies with our discovery.^27^ The combined discovery and replication data (with the exception of 23andMe and GBMI) were then meta-analyzed, and ancestry-stratified meta-analyses were performed for African (AFR)-ancestry, European (EUR)-ancestry, and Hispanic (HIS) participants to enable further downstream ancestry-specific analyses, such as fine mapping. Participants from studies provided written informed consent for use of their genetic and health information for analysis, and the studies were individually approved by the appropriate Institutional Review Boards (see **Supplemental Methods**).

**Figure 1:**
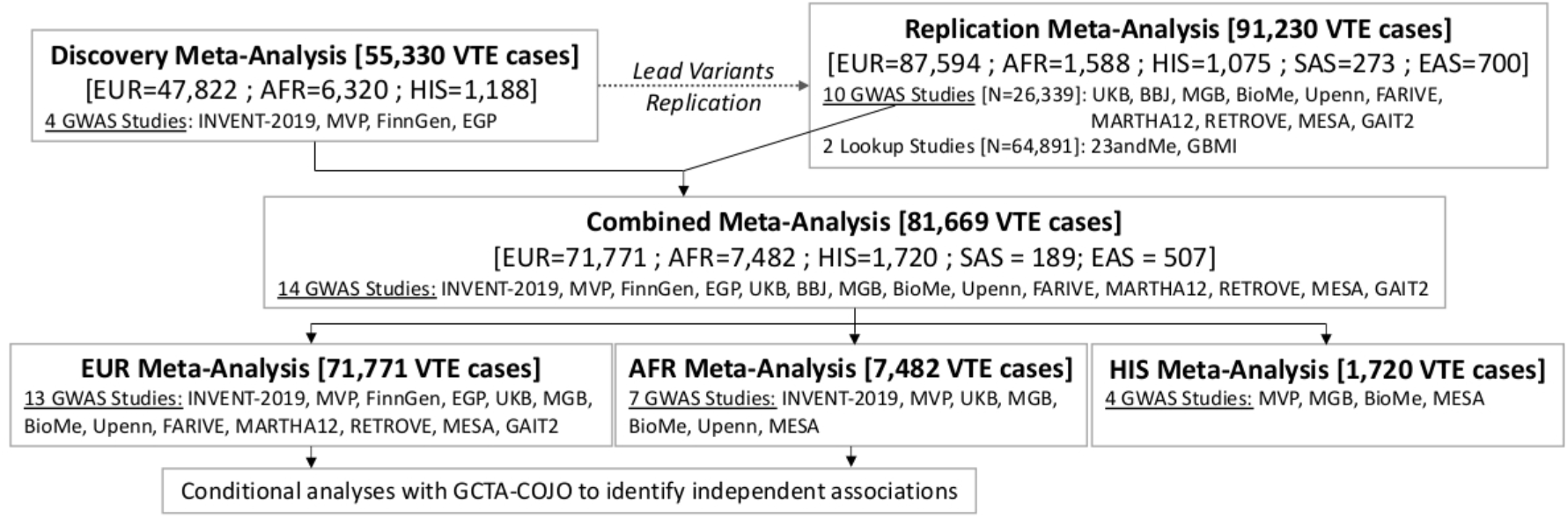
Analyses Workflow Workflow of genetic analyses conducted for this study.

### Study-specific GWAS

Genotyping arrays, imputation panels, and analyses performed by each participating study are detailed in supplemental **Table S1**. Briefly, studies performed association analyses (logistic regression analyses or generalized mixed models for case-control studies and Cox regression for cohort studies) using age and sex as covariates and further adjusting for participant relatedness, genetic principal components, and study site or other study-specific factors when applicable.

For each dataset, quality control was performed using EasyQC^28^ to remove variants with missing information (effect and/or standard error), low imputation quality (< 0.3), and rare variants (allele count < 5). For studies missing either imputation quality or variant frequency (BioMe, UPenn), a filter was added to remove variants with extreme effects (|Effect| > 10). Indels and marker names were then harmonized across all studies.

For the X chromosome, all studies performed sex-stratified GWAS, excluding variants from pseudo-autosomal regions. Results from males and females were then meta-analyzed.

### Discovery, Replication, and Combined GWAS Meta-analyses

All GWAS meta-analyses were conducted with METAL,^29^ using a fixed-effects inverse-variance weighted model. All variants were included and there was no lower minor allele frequency (MAF) limit beyond study-specific minor allele count. Genome-wide significant variants (P < 5.00 × 10^−8^) were kept if a concordant effect direction was observed in 2 or more studies and grouped into the same locus if they were within 1Mb. We used the closest gene to the lead variant to refer to each locus, except at known loci where the causal gene has been previously identified and is different from the closest gene (such as *PROCR* or *PROS1*). We defined a locus as novel if a genetic association with VTE has not been previously observed in the region according to our review of peer-reviewed published reports. We used PhenoGram^30^ to visually represent the genomic position of the loci, their significance in EUR- or AFR-ancestry analyses, and their discovery status of novel or known.

#### Discovery Meta-Analysis

For the discovery cross-ancestry GWAS meta-analysis, we meta-analyzed data from 4 consortium/studies: INVENT-2019, MVP, FinnGen and EGCUT. Participants were adult men and women and included 55,330 VTE cases (either DVT and/or PE cases) and 1,081,973 controls of EUR, AFR, or HIS ancestries. At each locus with a genome-wide significant signal, the lead variant was extracted and tested in an independent replication meta-analysis.

#### Replication

The replication GWAS meta-analysis consisted of the remaining 10 participating studies, as well as 2 external collaborators (GBMI ^27^ and 23andMe^26^), for a total of 91,230 VTE cases in replication. Replicating variants from the discovery were defined as those that had concordant effect direction in the discovery and the replication, and reached a Bonferroni-corrected p-value threshold in the replication population corresponding to the number of variants tested for replication with a 1-sided hypothesis: p-value threshold = [(0.05*2)/number of variants tested for replication] in the replication analysis.

#### Combined GWAS Meta-Analysis and Stratification by Ancestry

We performed a combined, cross-ancestry GWAS meta-analysis of discovery and replication data using participating studies with genome-wide summary data. We included variants with MAF ≥ 0.01 to maintain adequate statistical power by reducing the number of low-powered tests since replication was not available. Genome-wide data from GBMI and 23andMe data were not available and therefore excluded from combined analyses. We estimated the heterogeneity associated with each variant using Cochran’s Q test and the corresponding I^2^ statistic. We assessed the genomic inflation with the lambda genomic control.^31^ We report on variants exceeding the genome-wide threshold (P < 5.00 × 10^−8^) and view these as candidate novel loci associated with VTE and needing future replication.

We then stratified the analyses by ancestry and limited strata to EUR, AFR, and HIS ancestries as the remaining ancestries had too few VTE events to be informative: East Asian (EAS) in BBJ, n=507 VTE events; South Asian (SAS) in UKB, n=189 VTE events. As above, we estimated heterogeneity and assessed inflation with lambda genomic control; the LD-score intercept was computed for EUR-ancestry analysis, using the recommended Hapmap3 variants.^32^ We report all additional ancestry-specific variants exceeding the genome-wide threshold (P < 5.00 × 10^−8^) and view these as ancestry-specific candidate loci associated with VTE and needing future replication.

### Ancestry-Stratified Analyses: Conditional Analyses and Fine-mapping

To estimate the presence of multiple independent signals, we performed conditional analyses with GCTA-COJO^33^ at each locus with significant signals in the EUR- and AFR-ancestry GWAS meta-analyses. The Trans-Omics for Precision Medicine (TOPMed) trans-ancestry sequence data (freeze 8) was used as reference panel, selecting only EUR-ancestry participants from TOPMed (N=34,890) for the EUR conditional analyses and AFR-ancestry participants (N=17,322) for the AFR analyses.^34^ GCTA-COJO analyses were not performed for the HIS-ancestry stratum since a HIS reference panel was not available in TOPMed. Conditional analyses were performed at each locus, using a window that encompassed at least the genome-wide significant variants present in the locus with an additional buffer of ±100 Kb. A stepwise joint regression model was used to identify secondary signals with joint p-values < 5.00 × 10^−8^ and a linkage disequilibrium (LD) r^2^ < 0.2 with selected variants.

In addition, for each locus and for each ancestry-specific GWAS meta-analysis, we produced forest plots with the *forestplot* R library, and regional association plots with the *RACER*^35^ and *LDlinkR*^36^ R libraries, to visually inspect the local genetic architecture (available as supplemental **Figures 1-8**). We used the 1000 Genomes project EUR-ancestry dataset as reference panel to infer LD patterns for the EUR-ancestry participants and overall meta-analyses and the 1000 Genomes project AFR-ancestry reference panel for the AFR-ancestry meta-analysis. Furthermore, at each locus where distinct lead variants were identified in the different meta-analyses, we also extracted the lead variant from each analysis, as well as additional independent variants identified by the conditional analyses, and computed the LD between each variant (using both EUR- and AFR-ancestry reference panels) to verify the independence of the signals.

### Transcriptome-wide Association Studies (TWAS)

We performed TWAS with the FUSION pipeline to accomplish 2 tasks: (1) prioritize genes for those genome-significant signals with ambiguous gene associations; and (2) identify new candidate loci by linking gene expression with VTE risk using GWAS results not reaching genome-wide significance.^37^ This analysis was performed using the EUR-ancestry autosomal GWAS meta-analysis results, since FUSION depends on a EUR-ancestry LD reference panel (from 1000 genomes^38^) and does not include data for chromosome X. We first performed a series of single-tissue TWAS using gene expression from eQTL datasets relevant to blood and thrombosis disorders: whole blood, liver, lung, and spleen from GTEx v8,^39^ whole blood from the Young Finn Study,^40^ and peripheral blood from the Netherlands Twins Register.^41^ We then also employed an analysis using cross-tissue weights computed from GTEx v8 tissues, available as 3 canonical vectors (sCCA1-3) that capture most of the gene expression.^42^ All associations reaching a Bonferroni corrected significance threshold corresponding to the number of gene tested (N=14,219, P < 3.52 × 10^−6^) were deemed statistically significant. As several genes can be associated at the same locus, the TWAS results were subjected to a conditional analysis implemented in FUSION to select genes that remained conditionally independent. For each tissue, we further performed a colocalization test with COLOC^43^ for all significant associations, to identify and select genetic signals shared by both VTE risk and gene expression with high posterior probability (PP4 > 0.75). Selected genes located farther than 200kb from genetic loci identified in the meta-analyses were considered novel candidate VTE genes.

### Protein QTL Mendelian Randomization

Using the combined, cross-ancestry VTE GWAS meta-analysis results, we performed a proteome Mendelian randomization (MR) analysis with high-confidence genomic instruments corresponding to protein QTL (pQTL) for 1,216 circulating plasma proteins that passed consistency and pleiotropy filters, as previously described.^44^ When 2 or more genetic instruments were available for the exposure-outcome pairs, we performed inverse variance-weighted MR. If a single genetic variant was present, Wald-ratio MR was used instead. In addition, when multiple SOMAmers for a protein were available, which was the case for 40 proteins, tests were conducted separately for each SOMAmer. To account for multiple testing, associations passing the Bonferroni corrected threshold corresponding to the number of SOMAmers tested (N=1,256, P < 3.98 × 10^−5^) were considered statistically significant.

### Association of VTE Loci with Hemostasis and Hematology Traits

We conducted a series of *in silico* investigations involving hemostasis and hematology traits to better characterize the VTE-associated variants from the GWAS meta-analyses. To better understand if novel VTE-associated variants might operate through hemostasis pathways, we extracted associations from published GWAS of 10 hemostatic traits: fibrinogen;^45^ fibrin D-dimer;^46^ coagulation factors VII (FVII),^47^ VIII (FVIII),^48^ and XI (FXI);^49^ von Willebrand factor (vWF);^48^ tissue plasminogen activator (tPA);^50^ plasminogen-activator inhibitor 1 (PAI-1);^51^ activated partial thromboplastin time (aPTT); and prothrombin time (PT).^52^ Since each variant-association was investigated in 10 hemostasis traits, we set a p-value threshold of 0.005 (0.05/10 traits tested for each lead variant of a locus) to separate associations of interest from other associations.

Similarly, we extracted associations with complete blood count (CBC) measures using summary data from nearly 750,000 individuals on 15 leukocyte, erythrocyte, and platelet traits.^53^ Given the large sample size and high statistical power of these analyses, we used a more stringent threshold of interest that was a Bonferroni correction corresponding to the number of look-ups performed (P < 1.92 × 10^−5^).

### Phenome-wide Association Testing

To explore associations between VTE-associated variants and other traits agnostically, we performed a phenome-wide association study (PheWAS) using the MRC IEU infrastructure,^54^ which included the datasets from the 1,500 UKB analyses performed by the Neale lab on 337,000 individuals of British ancestry (pheWAS source codes: ukb-a and ukb-d). Only variants of interest reaching genome-wide significance (P < 5.00 × 10^−8^) were extracted for each trait and presented.

## RESULTS

### Discovery Cross-Ancestry Meta-analysis and Replication

The primary cross-ancestry discovery analysis was based on 4 contributing consortium/studies (INVENT-2019, MVP, FinnGen, EGP) and included 55,330 participants among 3 ancestry groups with VTE (47,822 EUR, 6,320 AFR, and 1,188 HIS) and 1,081,973 participants without VTE (918,195 EUR, 118,144 AFR, and 45,634 HIS). Over the 22 autosomal and X chromosomes, 35.5 million variants were analyzed, and the observed genomic inflation factor was 1.06. We identified 10,493 variants reaching genome-wide significance, corresponding to 85 loci, of which 48 have not been identified in previous genetic studies of VTE (see supplemental **Table S2**).

We tested these 85 variants for replication in 91,230 cases and 3,322,939 controls from the independent replication data. After meta-analyzing the results of these 85 tests in the replication population, we identified 83 variants with a concordant effect direction between the discovery and the replication, of which 68 replicated at the 1-sided Bonferroni corrected significance threshold (p < 0.1/83 = 0.0012) (**Table 1, Figure 2**, supplemental **Table S2**). The successfully replicated signals corresponded to 34 known and 34 novel loci. Among the 34 novel loci that replicated, heterogeneity was minimal (heterogeneity P > 0.05), odds ratios (ORs) ranged between 0.84-0.98 and 1.03-1.18, and MAFs were all ≥ 0.021. The majority of variants were gene-centric (4 exonic, 16 intronic, and 3 in 3’ or 5’ UTR regions or immediately downstream), 3 were linked to intronic non-coding RNA, and 8 were considered intergenic. The novel replicated variants were at the following loci (in chromosomal order): *H6PD*/*SPSB1, TENT5C, TRIM58, CALCRL* (near *TFPI*), *CPS1, SERPINE2, PIK3CB*/*LINC01391, MECOM, LINC01968*/*XXYLT1, SEC31A, ARHGAP24, LNPEP, ILRUN, AGPAT5*/*XKR5, ZNF367*/*HABP4, MIR1265*/*FAM107B, ZMIZ1, PLCE1, ST3GAL4, A2ML1*/*PHC1, COPZ1, SH2B3, RCOR1, MAP1A, RORA-AS1, HSD3B7, ZFPM1, ALOX12-AS1, MAPT-AS1, CEP112*/*APOH, RFX2, GIPR, FUT2*, and *SYN3*. Among the 17 variants and their associated loci that failed replication, 14 were novel and remain candidate loci that merit additional replication while 3 were known loci: *PROS1, JAK2*, and *STAB2*.

**Table 1:**
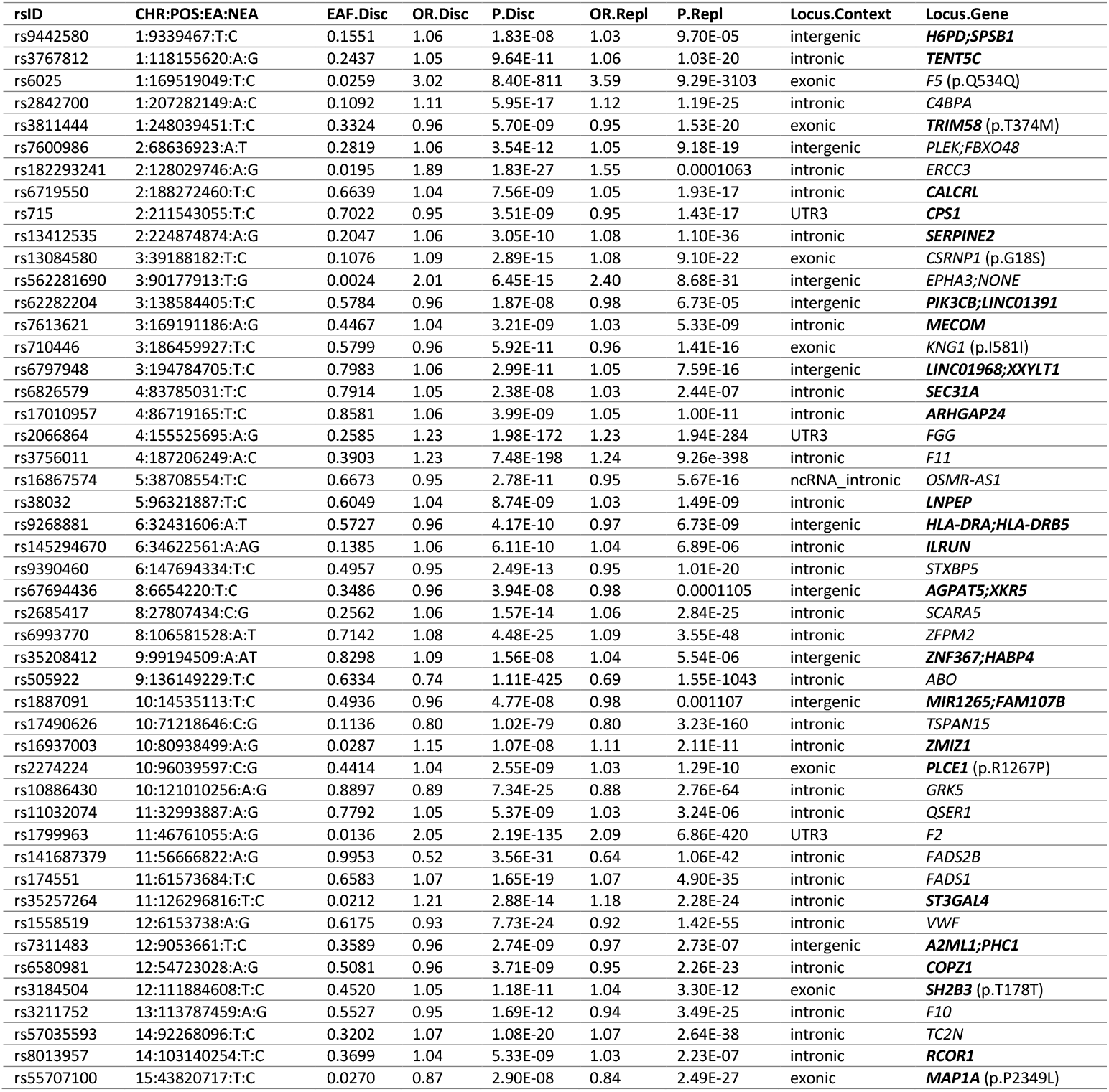
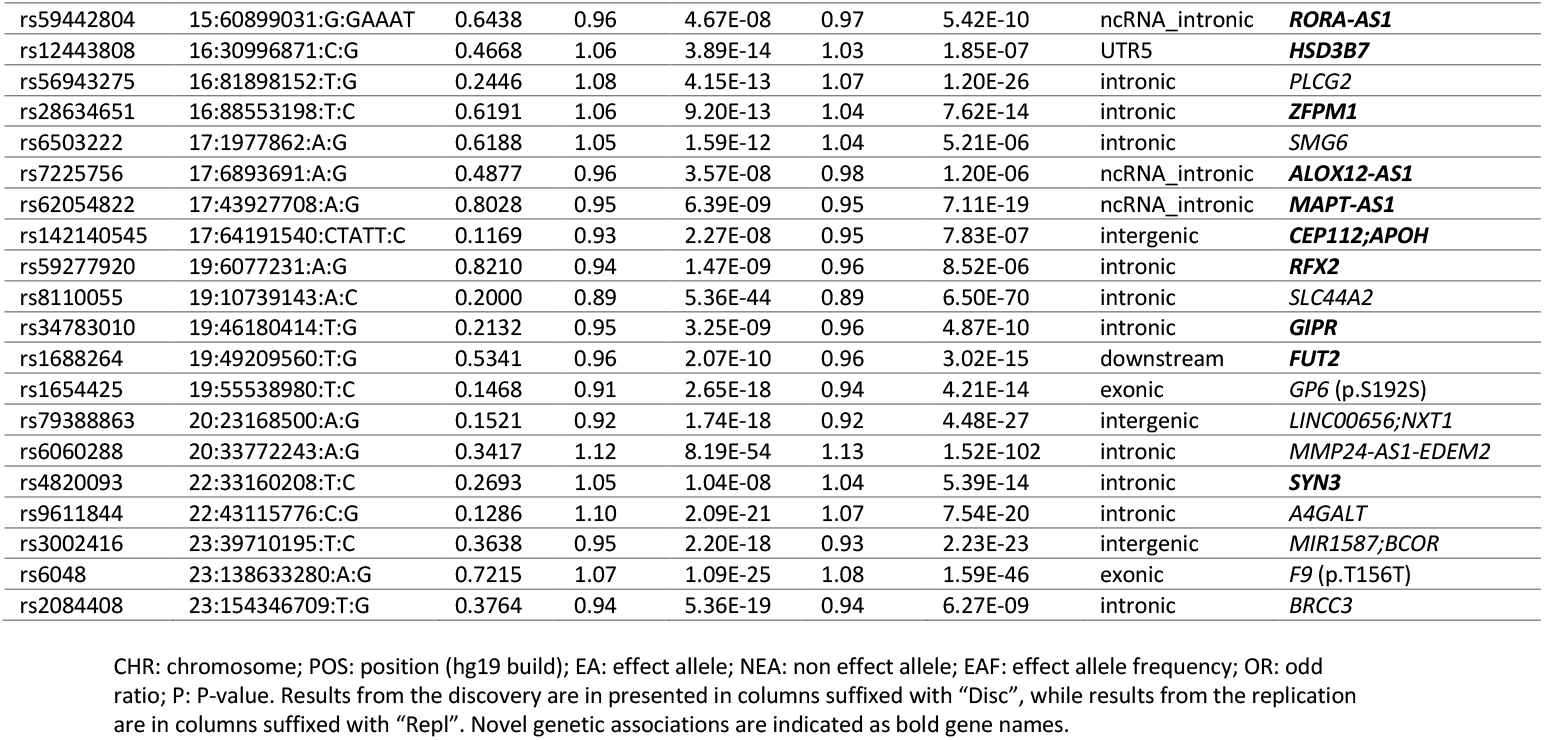
68 Lead variants from the Discovery meta-analysis successfully replicated

**Figure 2:**
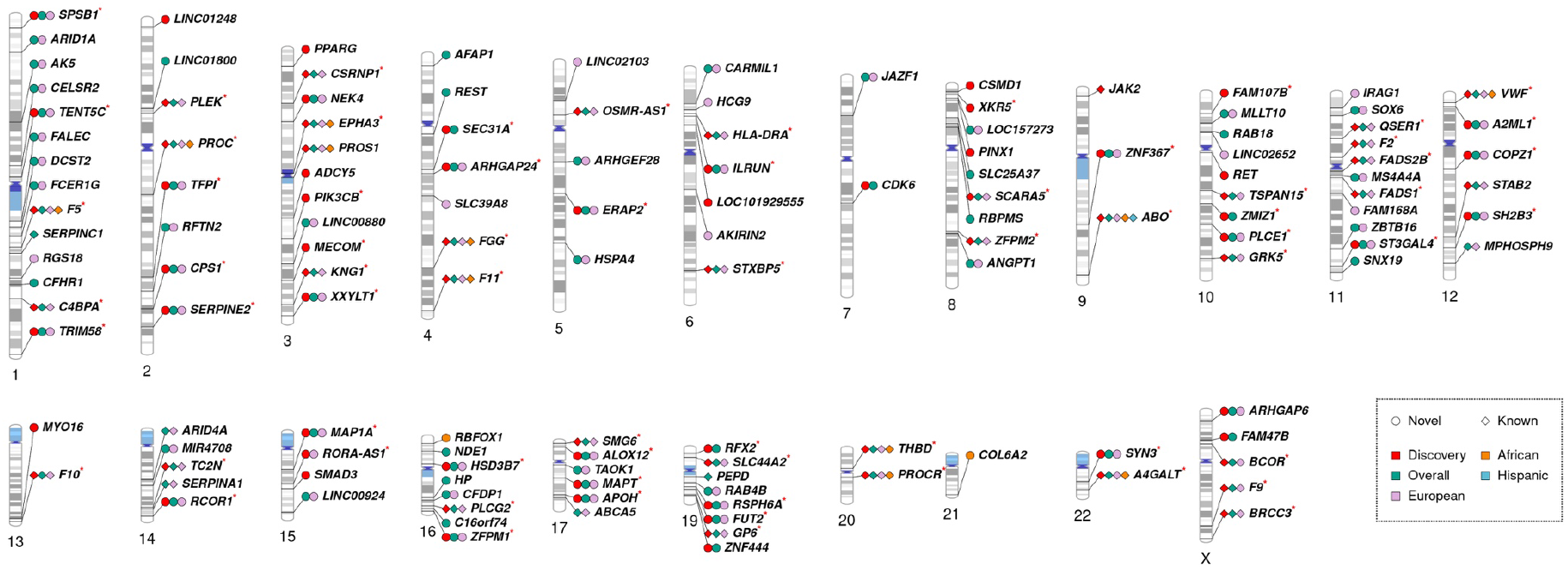
Genetic loci associated with VTE This figure presents the 130 loci significantly associated with VTE identified across all 4 meta-analyses: the Discovery (in blue), the overall meta-analysis (in green), the analysis restricted to individuals of European ancestry (in red) and the analysis restricted to individuals of African ancestry (in black). Novel loci are represented with circles and known loci with diamonds.

### Combined Cross-Ancestry GWAS Meta-analysis and Ancestry-Stratified Results

#### Combined

The combined, cross-ancestry meta-analysis of the studies with genome-wide markers included 81,669 individuals with VTE and 1,426,717 individuals without VTE. We analyzed 19.1 million common variants (MAF ≥ 0.01) and observed a genomic factor of 1.16 which is slightly elevated but expected for large scale meta-analyses of polygenic traits.^55^ We identified 16,550 variants reaching genome-wide significance located in 110 loci, of which 41 were not observed in the discovery analysis (supplemental **Table S3, Figure 2**). Of these 41 additional loci, 1 corresponded to a common variant at the known *SERPINC1* locus (rs6695940) which encodes antithrombin, 4 were previously identified in the INVENT-2019^13^ or MVP^14^ meta-analyses at the *PEPD, ABCA5, MPHOSPH9*, and *ARID4A* loci, and 1 was a known pathogenic missense variant located in *SERPINA1* (rs28929474, p.Glu366Lys).^56^ The remaining 35 loci were novel associations and are presented in **Table 2**. Among the 35 candidate loci, all had ORs within the range of 0.93-0.97 and 1.03-1.15 and had a minimum MAF of 0.021. The majority of the variants were gene-centric (18 intronic and 3 in 3’ UTR regions), 3 were intronic in non-coding RNA and 11 were considered intergenic. These candidate loci included (in chromosomal order): *ARID1A, AK5, CELSR2, FALEC*/*ADAMTSL4, DCST2, FCER1G, CFHR1*/*CFHR4, LINC01800, RFTN2*/*MARS2, LINC02029*/*LINC00880, AFAP1, REST, ARHGEF28, HSPA4, CARMIL1, HCG9, JAZF1-AS1, LOC101929128*/*LOC157273, ENTPD4*/*SLC25A37, RBPMS, ANGPT1, MLLT10, RAB18*/*MKX, SOX6, MS4A4A*/*MS4A6E, ZBTB16, SNX19, LINC02324*/*MIR4708, LINC00924*/*LOC105369212, NDE1, DHODH*/*HP, CFDP1, C16orf74, TAOK1*, and *RAB4B*.

**Table 2:**
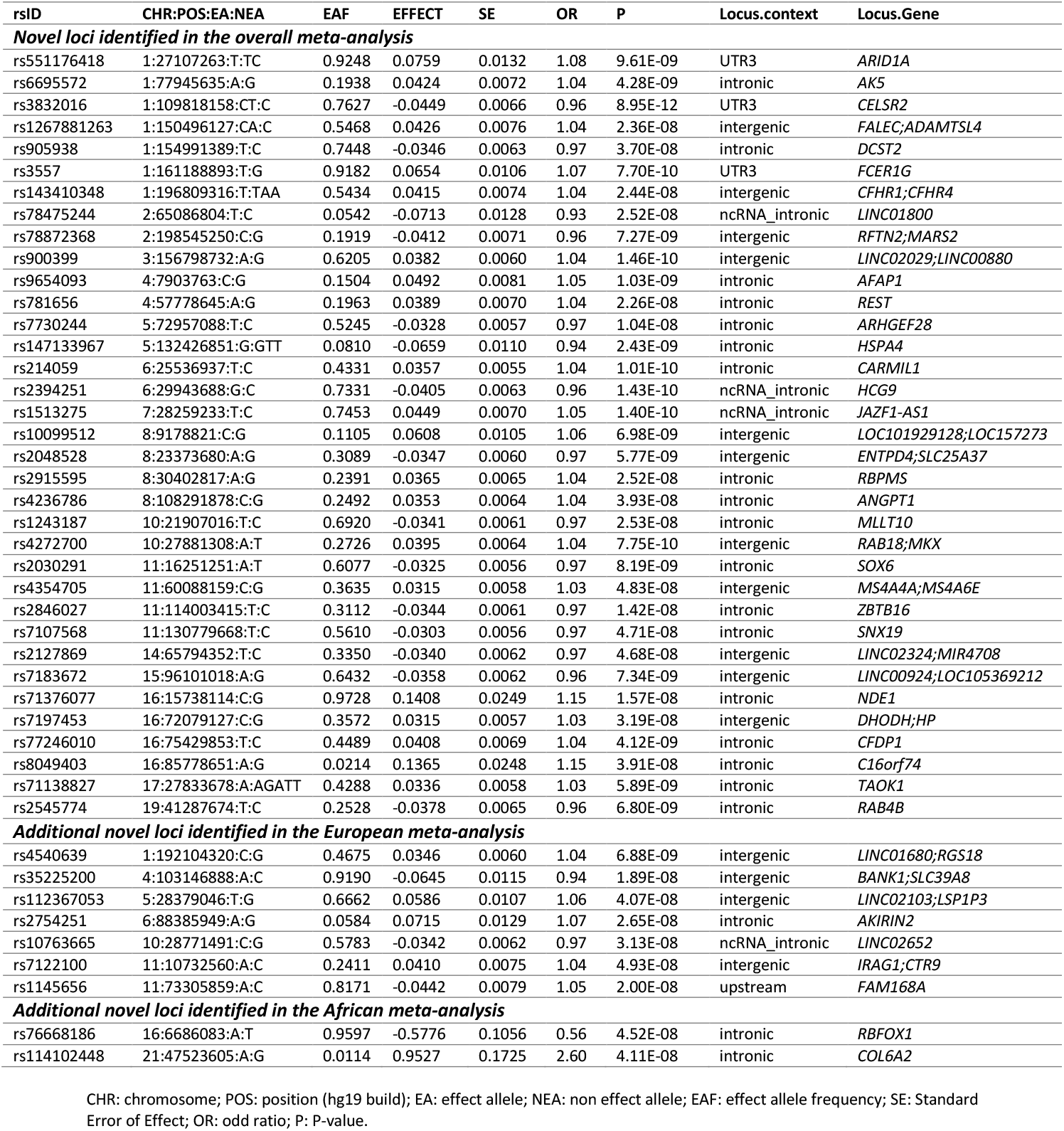
Additional 44 candidate novel loci identified in the Overall, European and African meta-analyses

#### European Ancestry

The EUR-ancestry meta-analysis, which included 71,771 participants with VTE and 1,059,740 participants without VTE, had a lambda genomic factor of 1.22. As population stratification might be introduced by founder effects in Finnish participants from FinnGen, we did a sensitivity analysis by removing this cohort, and observed a similar genomic factor of 1.19. We also observed an LD-score intercept of 1.07, indicating an inflation mainly due to polygenic architecture. Out of 11.1 million variants analyzed, 16,867 were genome-wide significant and clustered into 100 regions, of which 7 did not overlap with loci identified in the discovery or overall meta-analysis: *FAM168A, AKIRIN2, LINC02103, LINC02652, SLC39A8, IRAG1*, and *RGS18* (**Table 2, Figure 2**, supplemental **Table S4**). For these 7 additional candidate loci, the ORs ranged from 0.94-0.97 to 1.04-1.07 and the minimum MAF was 0.058.

Conditional analyses were performed with GCTA COJO at each of the 100 significant loci and revealed a subset of 21 loci with multiple independent signals (supplemental **Table S5**). These included 3 within novel loci: rs39840 (intronic, in *ERAP1*), rs28712647 (upstream of *NEURL4*, near *ALOX12*) and rs7412 (nonsynonymous, in *APOE*). At the remaining 18 known VTE loci, secondary signals at 5 loci had been previously observed, or were in LD with previously identified signals, at *F5, F11, ABO, F2* and *VWF*,^13^ while the rest were located at *EPHA3, PROS1, FGG*, the *HLA* region, *SCARA5, TSPAN15, FADS2B, FADS1, STAB2, F10, PLCG2, THBD*, and *A4GALT*.

#### African Ancestry

The AFR-ancestry meta-analysis included 7,482 participants with VTE and 129,975 participants without VTE from 7 cohorts and had a genomic inflation factor of 1.05. Here, 17.1 million variants were analyzed, of which 752 were genome-wide significant and located within 13 loci, of which 2 corresponded to novel ancestry-specific signals at *RBFOX1* (OR = 0.56; MAF = 0.04) and *COL6A2* (OR = 2.16; MAF = 0.011) (**Table 2, Figure 2**, supplemental **Table S6**).

Conditional analyses were performed with GCTA COJO at each of the 13 significant loci revealed 3 loci with additional independent signals (supplemental **Table S7**) at *EPHA3* (with LD r^2^ < 0.02 with the primary and secondary EUR signals, using the 1000 Genomes AFR reference LD panel), *PROS1* (rs6795524, LD r^2^ = 0.94 with rs28479320, the lead variant from the combined meta-analysis located in *ARL13B*), and *ABO* (several signals).

#### Hispanic Ancestry

The HIS-ancestry meta-analysis included 1,720 participants with VTE and 57,367 participants without VTE from 4 cohorts and had a genomic inflation factor of 1.02. We analyzed 11.1 million variants, of which 58 were genome-wide significant, all located at the *ABO* locus with rs2519093 as lead variant (MAF = 0.15, OR = 1.49, P = 3.08 × 10^−15^).

#### Comparison of Ancestry-Specific and Cross-Ancestry Meta-Analysis Results

We then investigated the lead variants from the AFR- and EUR-ancestry meta-analyses at the 11 loci (all known) identified in both analyses. At 5 loci (*PROC, EPHA3, PROS1, VWF* and *THBD*), none of the AFR lead variants were available in the EUR analyses, due to their low frequency in EUR (MAF < 0.0006 for all 5 lead variants in non-Finnish Europeans according to gnomAD^57^). At the remaining 6 loci, the lead variants from the AFR analysis were also genome wide significant in the EUR analysis, and shared similar effect sizes.

Across the discovery, combined, EUR, AFR and HIS meta-analyses, we identified 135 independent loci (**Figure 2**). A summary of each locus, including LD patterns between lead variants from each meta-analysis as well as independent signals and association test results across all meta-analyses, is available in supplemental **Table S8**.

### Gene Prioritization with TWAS and Protein QTL MR

#### Transcriptome Wide Association Study

Across the 6 single-tissue and 3 cross-tissues datasets analyzed, we identified 166 significant (P < 3.52 × 10^−6^) and conditionally independent associations with a high posterior probability of colocalization (> 0.75) between gene expression and VTE risk (see supplemental **Table S9**). These associations involved 108 genes, of which 77 were mapped to 46 genome-wide significant GWAS loci, leaving an additional 31 novel candidate genes that mapped outside of genome-wide significant GWAS loci (supplemental **Table S10**). The candidate genes included (in chromosomal order): *PRDX6, LMOD1, FEZ2, PNKD, BAP1, PRKCD, CHST13, CHST13, HGFAC, NIPAL1, ARHGAP10, LHFPL2, DND1, THBS2, MEST, COPG2, SYK, AAMDC, STAC3, KIF5A, NFKBIA, CINP, COMMD4, DCTPP1, NFAT5, MPDU1, KDM6B, C18orf8, NPC1, RAB3D, ISOC2*, and *SDCBP2*. At 33 GWAS loci, an associated gene matched the gene closest to the lead variant, supporting a role as a causal gene, while associated genes at the remaining 13 GWAS loci may point to genes of interest for further investigations.

#### Protein QTL Mendelian Randomization

We performed agnostic MR of 1,216 plasma circulating pQTL using the combined VTE meta-analysis results and identified 23 proteins with a significant causal association (P < 3.98 × 10^−5^, **Figure 3**, supplemental **Table S11**). For 13 proteins, the gene coordinates matched a genome-wide significant GWAS locus and included 5 of the novel GWAS loci: *TFPI, ERAP2, TIMP3* (at the *SYN3* locus), *TIMP4* (at the *SYN2/PPARG* locus) and *ECM1* (at the *FALEC* locus). Among the 10 candidate genes mapping outside of GWAS loci, *HGFAC* and *PRDX6* were also candidate genes identified in the TWAS. The remaining 8 candidate genes were: *EFEMP1, LCT, CLPS, MSR1, LGALS3, CD97, LILRB5, APOL3*.

**Figure 3:**
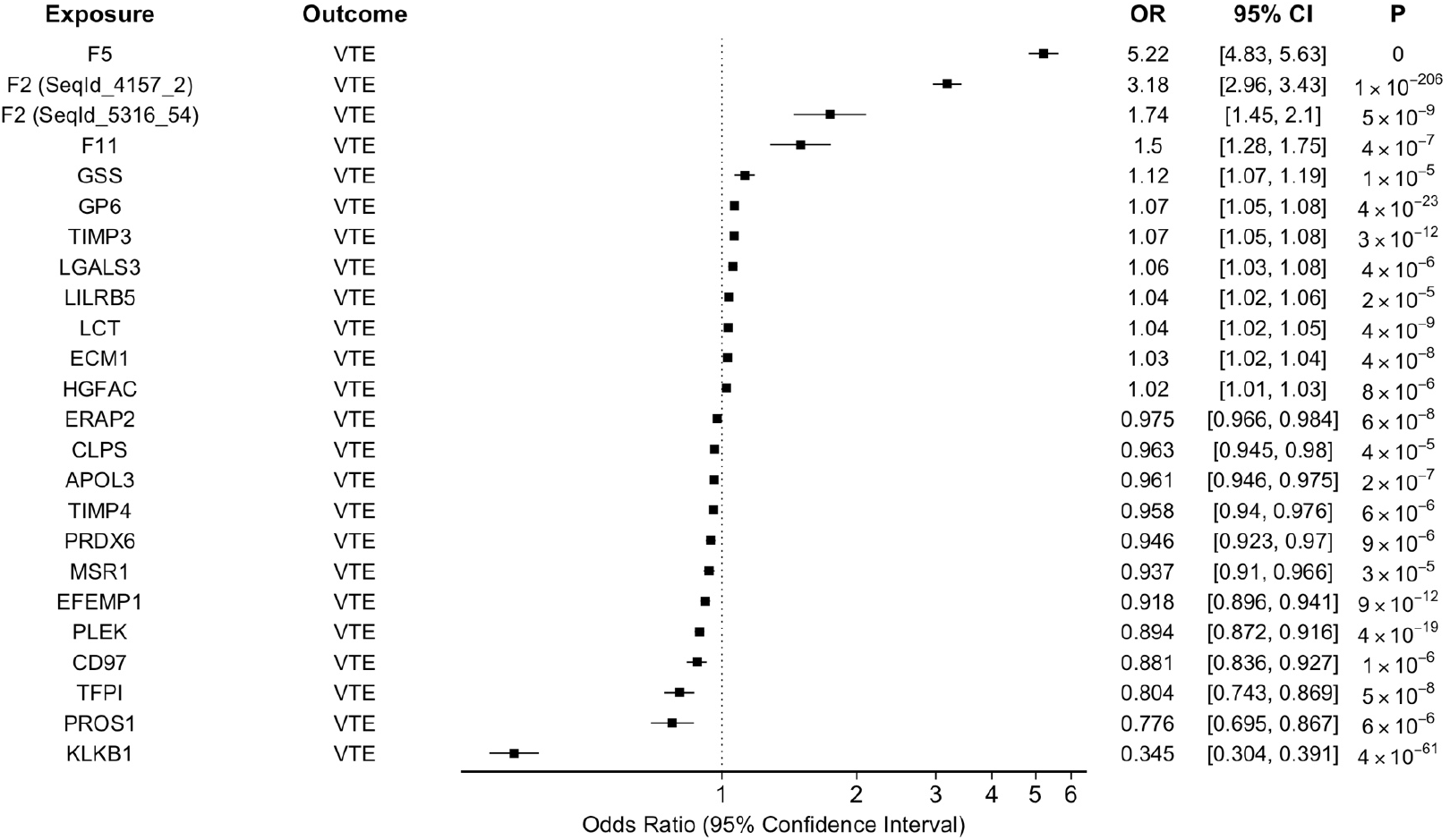
Significant associations of protein QTL Mendelian Randomization 23 genes with significant causal associations with VTE, out of 1,216 plasma protein analyzed, using the combined VTE summary statistics.

### Association of VTE-associated Variants with Hemostasis and Hematology Traits

The association of any lead or conditionally independent variant at the 135 GWAS loci with hemostasis traits is presented in **Figure 4.A** and supplemental **Table S12**. Among the 92 novel (replicated and candidate) loci reported above, 18 (19%) had a variant associated with 1 or more of the 10 hemostasis traits: fibrinogen (*CPS1, SLC39A8, ARHGEF28, LNPEP, PLCE1, MS4A4A, SH2B3, MIR4708, MAP1A, HP*); vWF (*SLC39A8, LNPEP, ST3GAL4, SH2B3, MAP1A, HP, APOE, FUT2*); FVII (*XXYLT1, JAZF1*-*AS1, MS4A6E, RCOR1, MAP1A*); FVIII (*ST3GAL4, COPZ1, HP, APOE*); PAI-1 (*COPZ1, GIPR*); or PT (*APOH*).

**Figure 4:**
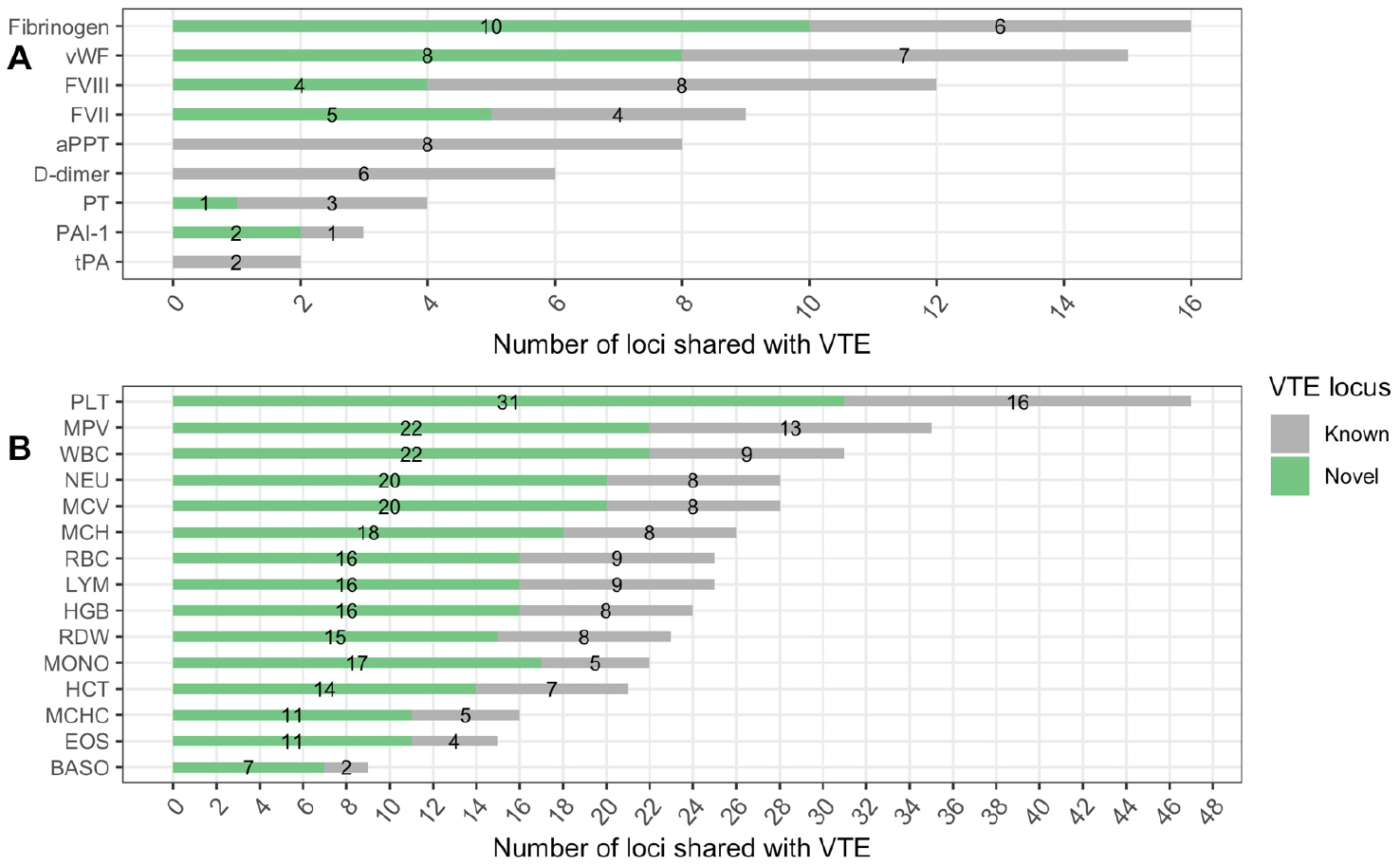
VTE genetic loci shared with hemostatic factors and blood traits (A) Number of known and novel VTE loci shared with each of the 10 hemostatic factors investigated. (B) Same analysis with complete blood count traits: PLT (platelet count), MPV (mean platelet volume), RBC (red blood cell count), MCV (mean corpuscular volume), HCT (hematocrit), MCH (mean corpuscular hemoglobin), MCHC (MCH concentration), HGB (hemoglobin concentration), RDW (red cell distribution width), WBC (white blood cell count), MONO (monocyte count), NEU (neutrophil count), EOS (eosinophil count), BASO (basophil count), LYM (lymphocyte count).

Next, we investigated associations of the 135 GWAS loci with hematology traits, presented in **Figure 4.B** and supplemental **Table S13**. Across all 15 CBC measures and among the 92 novel loci, we observed at least 1 association at 55 (59%) novel (replicated and candidate) loci.

### Phenome-wide Association Studies

We performed a pheWAS of lead and conditionally independent variants at the 135 significantly associated loci across 1,500 publicly available phenotypes involving European UKB participants (supplemental **Table S14**). For each trait, only genome-wide significant variants were retrieved, and we focused on traits sharing at least 10 loci with our VTE analyses (**Figure 5**, supplemental **Table S15**), which might indicate common biological pathways. Hematology traits, in particular platelet traits, shared the most loci with VTE (e.g. 33 for platelet count), consistent with our observations from the larger CBC GWAS (n∼750,000) sample (**Figure 4.B**). Several traits correspond to height and weight measurements, as well as enzymes mainly produced by the liver (e.g., albumin, sex-hormone binding globulin, or insulin growth factor-1), and plasma lipid-related traits (Apolipoprotein-A and B, HDL cholesterol, or triglycerides). Blood pressure (systolic and diastolic), glycated hemoglobin, calcium, cystatin C, and C-reactive protein levels were among additional traits sharing at least 10 loci with VTE. Few traits had a consistent direction of effect with respect to VTE risk across shared loci (**Figure 5**). For example, out of 10 loci shared between bilirubin levels and VTE, 9 (90%) were associated with an increase of both bilirubin levels and VTE risk. For albumin levels, glycated hemoglobin, and systolic blood pressure, an opposite direction of effect between these traits and VTE risk was observed at more than 75% of shared loci.

**Figure 5:**
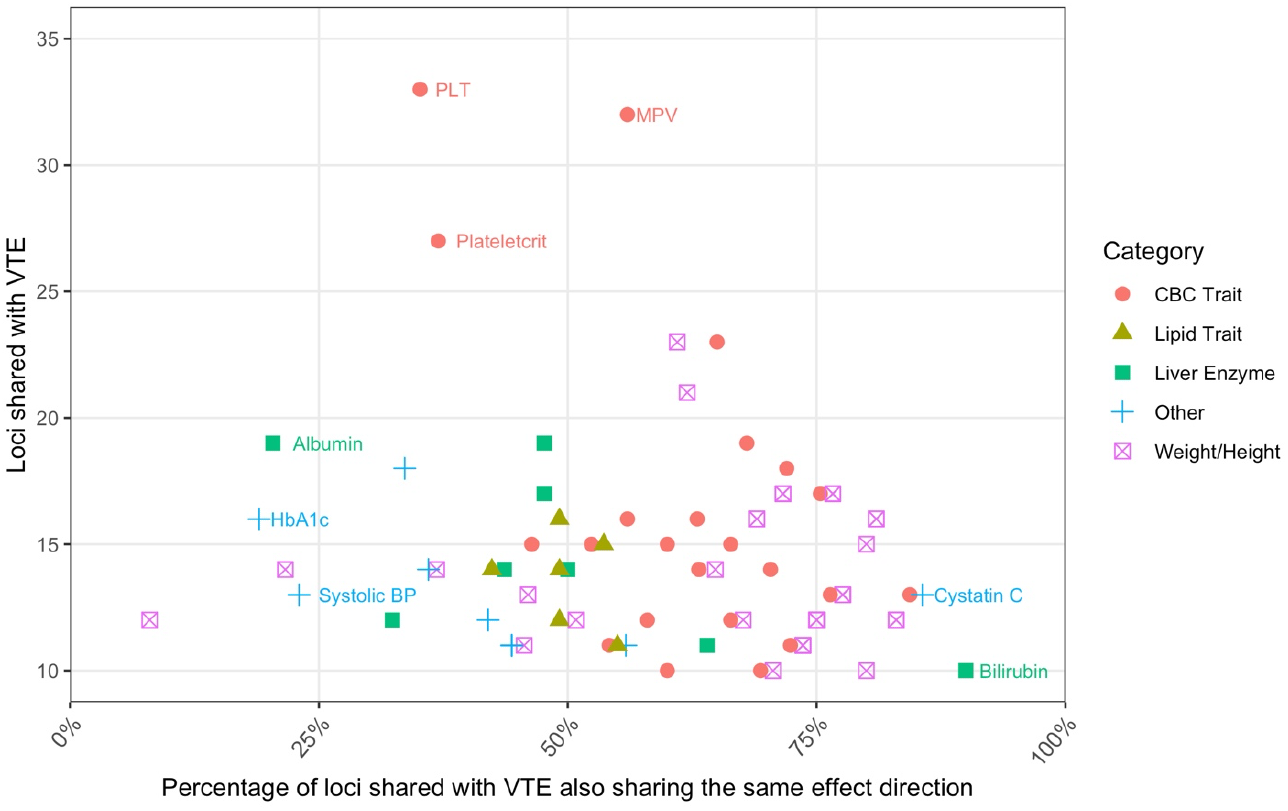
PheWAS traits sharing at least 10 loci with VTE This figure presents the pheWAS traits sharing at least 10 loci with VTE. Shape and color represent one of 5 categories: Complete Blood Count (CBC) traits, lipid traits, liver enzyme, height and weight traits, or other (if the trait did not fit in one of the aforementioned categories). The x-axis indicates the number of loci shared between VTE and the pheWAS trait, while the y-axis indicates the proportion of loci where the direction of effect was identical between the pheWAS trait and VTE. As a result, traits close to 100% have the same direction of effect than VTE at most shared loci, while traits close to 0% have an opposite direction than VTE at most shared loci

## DISCUSSION

We identified 135 independent genomic loci and 39 additional genes from TWAS and pQTL associated with an increased or decreased risk of VTE. This reflects a substantial increase in the number of validated and candidate loci for VTE risk beyond past genetic mapping efforts.^13,14^ Our results highlight genetic variation across the rare-to-common allele frequency spectrum in multiple ancestry groups and add new evidence of biologic predictors of VTE pathogenesis for further investigation. The *in silico* interrogations provide valuable clues regarding the putative causal gene at each locus and additional insights to biological pathways shared with VTE.

### Biological Insights

#### Novel Replicated Loci

Our strongest evidence supports 34 loci with novel VTE associations. Except for *TFPI* and *SERPINE2*, the novel genetic loci were not in established VTE pathophysiology pathways. A subset of these loci (12 loci, 35%) was associated with plasma levels of the 10 hemostasis traits interrogated and most (26 loci, 76%) were associated with a hematology trait. This contrast should be interpreted with caution as statistical power for the hemostasis traits was much smaller than for the hematology traits.

While most of the novel associations reported had an OR in the range of [0.90-0.98; 1.03-1.10], we were able to identify and replicate 3 uncommon variants with larger effects: an intronic variant (MAF = 0.021) in the glycosyltransferase *ST3GAL4* (discovery OR = 1.21, replication OR = 1.18), which was also associated with increased vWF and FVIII levels, an intronic variant (MAF = 0.029) in the transcriptional co-activator *ZMIZ1* (discovery OR = 1.15, replication OR = 1.11), and an exonic variant (MAF = 0.027) in *MAP1A* (p.Pro2349Leu, discovery OR = 0.87, replication OR = 0.84), which was also associated with decreased levels of vWF and fibrinogen, and had a protective effect against VTE.

Variants associated with hemostasis traits provide clues that the causal gene at these loci might directly or indirectly perturb the coagulation cascade. For instance, *XXYLT1* encodes a xylosyltransferase known to interact with coagulation factors^58^ and had a nearby variant (OR discovery = 1.06, OR replication = 1.06) also associated with decreased FVII levels. Another example is *FUT2*, a fucosyltransferase gene involved in the synthesis of the H antigen, a building block for the production of antigens within the ABO blood group. *FUT2* had a downstream variant (OR discovery= 0.96, OR replication = 0.96) that was also associated with decreased vWF levels, mirroring results observed with vWF at the *ABO* locus. In addition, some variants were associated with several hematology traits, suggesting common genetic regulatory pathways affecting hematopoiesis, such as the replicated *RCOR1* signal on chromosome 14, and the candidate gene *REST* on chromosome 4 identified in the combined meta-analysis, 2 genes that form the transcriptional repressor CoREST, known to mediate hematopoiesis.^59^

Among the 34 loci, 17 had TWAS evidence linking transcript expression with a gene in the locus and 3 were linked to protein measures. These results may help to prioritize biologically relevant genes for further investigations. Notably, at the *COPZ1* locus, the lead variant was associated with several CBC measures, including platelet count and red blood cell count, and the TWAS revealed an association with *NFE2*, known to regulate erythroid and megakaryocyte maturation.

#### Other Replicated and Non-Replicated Loci

Replicated variants included 2 rare variants at the known *EPHA3* (intergenic, MAF = 0.0024, OR = 2.40) and *FADS2B* (intronic, MAF = 0.0047, OR = 0.64) loci. Among variants that failed replication, only 1 rare variant displayed significant heterogeneity (P = 0.0001, *MYO16* locus), and 3 variants were located in known loci: *STAB2* was previously identified as associated with VTE in an independent gene-based study using exome sequencing,^60^ *ARL13B* (near *PROS1*) was identified in the previous VTE GWAS from MVP,^14^ and the *JAK2* V617F variant, which is known to increase the risk of myeloproliferative neoplasm, was recently identified as associated with VTE in an exome study of nearly 450,000 UKB participants.^61^ According to gnomAD,^57^ the *ARL13B* variant identified is mostly observed in AFR-ancestry individuals (rs79324379, AFR MAF = 0.026 against MAF < 0.0003 in other ancestries) and was not in LD with the lead variant identified in the previous MVP GWAS (rs6795524, LD r^2^= 0.01 in AFR); nonetheless, we would need additional information to validate this locus as a truly independent signal—and not just a marker—from any strong, uncharacterized signal in *PROS1*. Similarly, the *STAB2* variant identified is mostly observed in Finns (rs142351376, Finns MAF = 0.020 against MAF < 0.0003 in other ancestries); the lack of Finns in the replication likely impaired our ability to replicate the association. Out of the other 12 failed replications, 5 involved rare variants (MAF < 0.01) that did not reach nominal significance (P < 0.05), while 6 of the remaining 7 common variants reached nominal significance, suggesting that these common variants might need a larger replication sample to be validated. One of these signals, located between *SYN2* and *PPARG*, was associated with the protein levels of *TIMP4* in a previous study.^62^ This protein, known to inhibit matrix metalloproteinases and involved in platelet aggregation and recruitment,^63^ was confirmed by the pQTL MR analysis as a gene associated with VTE risk.

#### Novel Candidate Loci

Across the multiple interrogation approaches, we identified several score of candidate loci with evidence to support their association with VTE, though not yet replicated. This included 35 candidates from the combined GWAS, 7 candidates from the EUR-ancestry GWAS, and 2 candidates from the AFR-ancestry GWAS. Interestingly, the 2 variants (MAF 0.04 and 0.011) in the AFR-ancestry population were not present in EUR-ancestry participants and were associated with nearly 2-fold changes in risk of VTE. However, these 2 variants were only detected in a subset of studies, which included only 882 AFR-ancestry VTE cases out of 7,482, warranting additional investigations to confirm these 2 signals in *RBFOX1* (an RNA-binding protein) and *COL6A2* (a collagen-generating gene that contains several domains similar to *VWF* type A domains). For the remaining candidate GWAS loci, we saw similar attributes and associations as we did with the replicated loci. With additional replication resources in the future, these candidates may become fully replicated genetic associations. In addition, the conditional analyses revealed independently associated variants mapping to distinct genes that may be of interest for further investigations, such as *BRD3* at the *ABO* locus, a chromatin reader known to associate with the hematopoietic transcription factor *GATA1*.^64^ At the *EPHA3* locus, we also noted that the lead GWAS variant and the conditionally independent variant mapped upstream and downstream of *PROS2P*, a protein S pseudogene that might be of interest.

At these candidate loci, gene prioritized by the TWAS may also provide putative genes at these loci. For example *ZBTB7B*, a zinc-finger protein that represses the expression of extracellular matrix genes such as fibronectin and collagen^65^ was identified by TWAS at the candidate locus *DCST2*. The 31 candidate genes identified in the TWAS as well as the additional 8 from the pQTL MR analyses, although lacking a significant genetic association at these loci, might indicate relevant genes for future investigations. For instance, SYK is a critical platelet-activation protein and tyrosine kinase inhibitors of SYK have been explored for platelet inhibition.^66,67^

### Clinical Implications

Current anticoagulation therapy to prevent or treat VTE operate through the modulation of proteins produced in the liver (coumarin-based therapies) or through direct inhibition of coagulation factors IIa (thrombin) and Xa. Although the safety profile of anticoagulation treatments has evolved, bleeding remains a life-threatening off-target outcome. New approaches to preventing thrombosis while minimizing bleeds are in development, including a focus on contact (intrinsic) pathway proteins factor XI, factor XII, prekallikrein, and high-molecular-weight kininogen.^68^ Agnostic interrogations such as these may lead to discovery of novel proteins that “break the inexorable link between antithrombotic therapy and bleeding risk.”^69^

Remarkably, the hematology traits investigations and the pheWAS established that CBC measures share a large number of loci with VTE, and platelet phenotypes in particular are the most frequent traits shared with VTE variants: 51 loci were associated with either platelet count, mean platelet volume, plateletcrit or platelet distribution width in the pheWAS, and 35 of these loci are novel, which represents more than a third of all novel genetic associations. Several loci associated with VTE harbor genes with known roles in hematopoiesis and megakaryocyte development, or platelet turnover: *ARID1A, REST* and its co-repressor *RCOR1, CDK6, MECOM, RBPMS, ANGPT1, RET, NFE2, ST3GAL4, SH2B3, ZFPM2* and *ZFPM1*,^59,71–79^ or platelet aggregation: *SLC44A2, VWF, FGG, GP6, RGS18, GRK5, PIK3CB, PLCE1, PLCG2, IRAG1, TIMP4, FCER1G*, and *ALOX12*. ^10,63,80–89^ Altered platelet generation, turnover or reactivity may be a feature of VTE pathogenesis. For one, past prospective studies^90^ and case-control studies^91,92^ suggest that enlarged platelets, as measured by MPV, are associated with VTE and VTE outcomes. Studies of platelet function measures with VTE have been less conclusive which may relate to the limitations of these studies in assessing comprehensive and standardized platelet reactivity mechanisms.^93–95^ Collectively, these results suggest that treatments inhibiting platelet activation such as aspirin might be beneficial in the prevention of VTE, although previous studies and trials on aspirin and combinations with anticoagulants offered mixed results.^96^ Different antiplatelets, such as more targeted thrombin, PAR1 or PAR4 inhibitors, or intracellular PDE platelet signaling inhibitors like cilostazol, could be worthwhile for further study in VTE prevention.

### Strengths and Limitations

The major strength of this genetic discovery effort is the large sample size of the genetic variation interrogations. As the largest genetic association study of VTE to date, we were able to increase statistical power compared with previous VTE GWAS meta-analysis efforts and increase our ability to detect new associations, many of which were replicated. We were also better powered to detect less common genetic variation. The cross-ancestry meta-analyses also increased our potential to discover novel genetic associations where the allele frequency was more common in some populations compared with others.

Several limitations deserve mention. Case ascertainment varied by study and some studies provided validated VTE events while others relied on information from electronic health records. Further, some studies only included hospitalized VTE events and did not capture events in the outpatient setting. These differences may have introduced some bias if case ascertainment and hospitalization status have genetic determinants. We included all VTE cases and did not stratify by provoked status in order to increase statistical power. Furthermore, many of the studies had not classified the VTE events as provoked and unprovoked. In addition, although the cross-ancestry approach provided benefits as described above, the numbers of VTE cases were not evenly distributed by ancestry, thus reducing our ability to detect ancestry-specific VTE variants in the under-represented ancestry groups with more modest case counts. Due to the diversity of imputation panels used by the participating studies, genetic variants had variable coverage across studies which weakened our power to detect associations. Another limitation of our approach that used summary GWAS statistics from meta-analyses is the absence of participant-specific genotype-level information. This required us to rely on LD information extracted from external datasets, which can result in variants being missed and LD patterns not accurately captured. This may have introduced some bias in analyses that relied on LD, such as the conditional analyses and the TWAS. Further, *in silico* work was performed using external data sets such as the hemostatic factors and hematology traits summary statistics, where the size (and statistical power) of the datasets varied greatly. Although different significance thresholds were employed for significance, this may have biased the detection of significant associations to those traits that had large sample sizes.

### Conclusions

These cross-ancestry GWAS meta-analyzes have provided a list of 34 loci that replicated discovery findings. Some of the novel loci may contribute to VTE through well-characterized coagulation pathways while others provide new data on the role of hematology traits, particularly platelet function. Many of the replicated loci are outside of known or currently hypothesized pathways to thrombosis. We also provided a list of 44 new candidate loci including candidates from the combined cross-ancestry GWAS, from the EUR-ancestry GWAS, from the AFR-ancestry GWAS, and also 39 candidate genes from the TWAS and pQTL MR. These findings highlight new pathways to thrombosis and provide novel molecules that may be useful in the development of antithrombosis treatment that reduce bleeding adverse occurrences.

## Data Availability

All data produced in the present study are available upon reasonable request to the authors

## Acknowledgments

The INVENT Consortium would like to acknowledge all the participants across studies that provided their health information to support these analyses.

The INVENT Consortium is supported in part by HL134894 and HL154385. The Analysis Commons was funded by R01HL131136. Infrastructure for the CHARGE Consortium is supported in part by the National Heart, Lung, and Blood Institute grant R01HL105756.

Study acknowledges can be found in the **Supplemental Methods**.

The views expressed in this manuscript are those of the authors and do not necessarily represent the views of the National Heart, Lung and Blood Institute, the National Institute of Health, Department of Veterans Affairs, or the U.S. Department of Health and Human Services.

