## Supplemental Figure 1 for "Cross-Ancestry Investigation of Venous Thromboembolism Genomic Predictors"

### Supplementary Figure 1.
### Forest plots of Discovery analysis lead variants.

1.1 rs9442580 (chr1:9339467) in locus 1 (H6PD ;SPSB1) . . . 3
1.2 rs3767812 (chr1:118155620) in locus 5 (TENT5C) . . . 3
1.3 rs6025 (chr1:169519049) in locus 9 (F5) . . . 4
1.4 rs2842700 (chr1:207282149) in locus 13 (C4BPA) . . . 4
1.5 rs3811444 (chr1:248039451) in locus 14 (TRIM58) . . . 5
1.6 rs545891600 (chr2:5378301) in locus 15 (LINC01249 ;LINC01248) . . . 5
1.7 rs7600986 (chr2:68636923) in locus 17 (PLEK ;FBXO48) . . . 6
1.8 rs182293241 (chr2:128029746) in locus 18 (ERCC3) . . . 6
1.9 rs6719550 (chr2:188272460) in locus 19 (CALCRL) . . . 7
1.10 rs715 (chr2:211543055) in locus 21 (CPS1) . . . 7
1.11 rs13412535 (chr2:224874874) in locus 22 (SERPINE2) . . . 8
1.12 rs2960420 (chr3:12314512) in locus 23 (SYN2 ;PPARG) . . . 8
1.13 rs13084580 (chr3:39188182) in locus 24 (CSRNP1) . . . 9
1.14 rs11130326 (chr3:52771920) in locus 25 (NEK4) . . . 9
1.15 rs562281690 (chr3:90177913) in locus 26 (EPHA3 ;NONE) . . . 10
1.16 rs79324379 (chr3:93712671) in locus 27 (ARL13B) . . . 10
1.17 rs9872572 (chr3:123108613) in locus 28 (ADCY5) . . . 11
1.18 rs62282204 (chr3:138584405) in locus 29 (PIK3CB ;LINC01391) . . . 11
1.19 rs7613621 (chr3:169191186) in locus 31 (MECOM) . . . 12
1.20 rs710446 (chr3:186459927) in locus 32 (KNG1) . . . 12
1.21 rs6797948 (chr3:194784705) in locus 33 (LINC01968 ;XXYLT1) . . . 13
1.22 rs6826579 (chr4:83785031) in locus 36 (SEC31A) . . . 13
1.23 rs17010957 (chr4:86719165) in locus 37 (ARHGAP24) . . . 14
1.24 rs2066864 (chr4:155525695) in locus 39 (FGG) . . . 14
1.25 rs3756011 (chr4:187206249) in locus 40 (F11) . . . 15
1.26 rs16867574 (chr5:38708554) in locus 42 (OSMR-AS1) . . . 15
1.27 rs38032 (chr5:96321887) in locus 44 (LNPEP) . . . 16
1.28 rs9268881 (chr6:32431606) in locus 48 (HLA-DRA ;HLA-DRB5) . . . 16
1.29 rs145294670 (chr6:34622561) in locus 49 (ILRUN) . . . 17
1.30 rs903646797 (chr6:40985346) in locus 50 (LOC101929555) . . . 17
1.31 rs9390460 (chr6:147694334) in locus 52 (STXBP5) . . . 18
1.32 rs42036 (chr7:92241451) in locus 54 (CDK6) . . . 18
1.33 rs538161377 (chr8:4206326) in locus 55 (CSMD1) . . . 19
1.34 rs67694436 (chr8:6654220) in locus 56 (AGPAT5 ;XKR5) . . . 19
1.35 rs7010926 (chr8:10626791) in locus 58 (PINX1) . . . 20
1.36 rs2685417 (chr8:27807434) in locus 60 (SCARA5) . . . 20
1.37 rs6993770 (chr8:106581528) in locus 62 (ZFPM2) . . . 21
1.38 rs77375493 (chr9:5073770) in locus 64 (JAK2) . . . 21
1.39 rs35208412 (chr9:99194509) in locus 65 (ZNF367 ;HABP4) . . . 22
1.40 rs505922 (chr9:136149229) in locus 66 (ABO) . . . 22
1.41 rs1887091 (chr10:14535113) in locus 67 (MIR1265 ;FAM107B) . . . 23
1.42 rs529892067 (chr10:43568540) in locus 71 (MIR5100 ;RET) . . . 23
1.43 rs17490626 (chr10:71218646) in locus 72 (TSPAN15) . . . 24
1.44 rs16937003 (chr10:80938499) in locus 73 (ZMIZ1) . . . 24
1.45 rs2274224 (chr10:96039597) in locus 74 (PLCE1) . . . 25
1.46 rs10886430 (chr10:121010256) in locus 75 (GRK5) . . . 25
1.47 rs11032074 (chr11:32993887) in locus 78 (QSER1) . . . 26
1.48 rs1799963 (chr11:46761055) in locus 79 (F2) . . . 26
1.49 rs141687379 (chr11:56666822) in locus 80 (FADS2B) . . . 27
1.50 rs174551 (chr11:61573684) in locus 82 (FADS1) . . . 27

1.51 rs35257264 (chr11:126296816) in locus 85 (ST3GAL4) . . . . . 28
1.52 rs1558519 (chr12:6153738) in locus 87 (VWF) . . . . . 28
1.53 rs7311483 (chr12:9053661) in locus 88 (A2ML1;PHC1) . . . . . 29
1.54 rs6580981 (chr12:54723028) in locus 89 (COPZ1) . . . . . 29
1.55 rs142351376 (chr12:104136288) in locus 90 (STAB2) . . . . . 30
1.56 rs3184504 (chr12:111884608) in locus 91 (SH2B3) . . . . . 30
1.57 rs75940120 (chr13:109686619) in locus 93 (MYO16) . . . . . 31
1.58 rs3211752 (chr13:113787459) in locus 94 (F10) . . . . . 31
1.59 rs57035593 (chr14:92268096) in locus 97 (TC2N) . . . . . 32
1.60 rs8013957 (chr14:103140254) in locus 99 (RCOR1) . . . . . 32
1.61 rs55707100 (chr15:43820717) in locus 100 (MAP1A) . . . . . 33
1.62 rs59442804 (chr15:60899031) in locus 101 (RORA-AS1) . . . . . 33
1.63 rs182906510 (chr15:67372922) in locus 102 (SMAD3) . . . . . 34
1.64 rs12443808 (chr16:30996871) in locus 106 (HSD3B7) . . . . . 34
1.65 rs56943275 (chr16:81898152) in locus 109 (PLCG2) . . . . . 35
1.66 rs28634651 (chr16:88553198) in locus 111 (ZFPM1) . . . . . 35
1.67 rs6503222 (chr17:1977862) in locus 112 (SMG6) . . . . . 36
1.68 rs7225756 (chr17:6893691) in locus 113 (ALOX12-AS1) . . . . . 36
1.69 rs62054822 (chr17:43927708) in locus 115 (MAPT-AS1) . . . . . 37
1.70 rs142140545 (chr17:64191540) in locus 116 (CEP112;APOH) . . . . . 37
1.71 rs59277920 (chr19:6077231) in locus 118 (RFX2) . . . . . 38
1.72 rs8110055 (chr19:10739143) in locus 119 (SLC44A2) . . . . . 38
1.73 rs34783010 (chr19:46180414) in locus 122 (GIPR) . . . . . 39
1.74 rs1688264 (chr19:49209560) in locus 123 (FUT2) . . . . . 39
1.75 rs1654425 (chr19:55538980) in locus 124 (GP6) . . . . . 40
1.76 rs2631628 (chr19:56645250) in locus 125 (ZNF787;ZNF444) . . . . . 40
1.77 rs79388863 (chr20:23168500) in locus 126 (LINC00656;NXT1) . . . . . 41
1.78 rs6060288 (chr20:33772243) in locus 127 (MMP24-AS1-EDEM2) . . . . . 41
1.79 rs4820093 (chr22:33160208) in locus 129 (SYN3) . . . . . 42
1.80 rs9611844 (chr22:43115776) in locus 130 (A4GALT) . . . . . 42
1.81 rs34609587 (chr23:11657363) in locus 131 (ARHGAP6) . . . . . 43
1.82 rs6632109 (chr23:34894638) in locus 132 (TMEM47;FAM47B) . . . . . 43
1.83 rs3002416 (chr23:39710195) in locus 133 (MIR1587;BCOR) . . . . . 44
1.84 rs6048 (chr23:138633280) in locus 134 (F9) . . . . . 44
1.85 rs2084408 (chr23:154346709) in locus 135 (BRCC3) . . . . . 45

**Supplementary Figure 1.1** – rs9442580 (chr1:9339467) in locus 1 (H6PD ;SPSB1)

| Study | Cases | Controls | OR |
|---|---|---|---|
| INVENT-2019_EA | 15572 | 115280 | 1.1 |
| INVENT-2019_AA | 799 | 14353 | 0.99 |
| MVP_EA | 18952 | 411746 | 1.05 |
| MVP_AA | 5521 | 103791 | 1.01 |
| MVP_HIS | 1188 | 45634 | 1.06 |
| FinnGen_EA | 9176 | 209616 | 1.05 |
| EGP_EA | 4122 | 181553 | 1.02 |
| **Discovery** | **55330** | **1081973** | **1.06** |

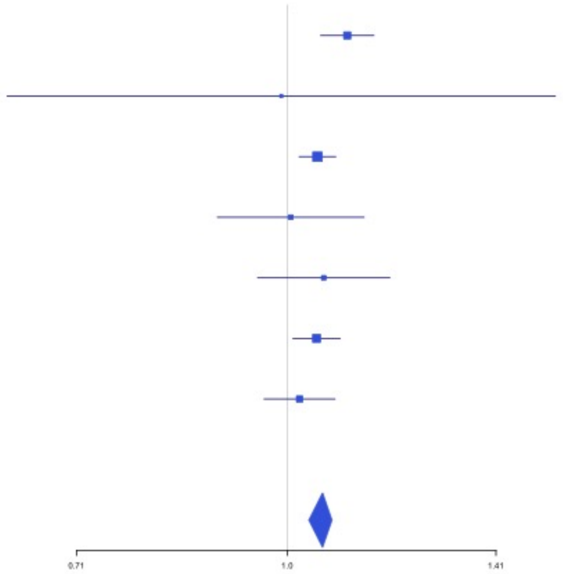


**Supplementary Figure 1.2** – rs3767812 (chr1:118155620) in locus 5 (TENT5C)

| Study | Cases | Controls | OR |
|---|---|---|---|
| INVENT-2019_EA | 15572 | 115280 | 1.04 |
| INVENT-2019_AA | 799 | 14353 | 1.14 |
| MVP_EA | 18952 | 411746 | 1.06 |
| MVP_AA | 5521 | 103791 | 1.08 |
| MVP_HIS | 1188 | 45634 | 1.05 |
| FinnGen_EA | 9176 | 209616 | 1.04 |
| EGP_EA | 4122 | 181553 | 1 |
| **Discovery** | **55330** | **1081973** | **1.05** |

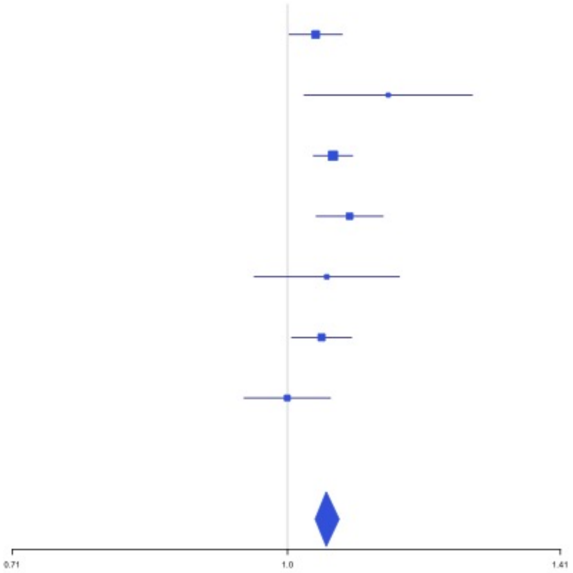

**Supplementary Figure 1.3** – rs6025 (chr1:169519049) in locus 9 (F5)

| Study | Cases | Controls | OR |
|---|---|---|---|
| INVENT-2019_EA | 15572 | 115280 | 2.39 |
| MVP_EA | 18952 | 411746 | 3.12 |
| MVP_AA | 5521 | 103791 | 2.61 |
| FinnGen_EA | 9176 | 209616 | 3.24 |
| EGP_EA | 4122 | 181553 | 3.07 |
| **Discovery** | **53343** | **1021986** | **3.02** |

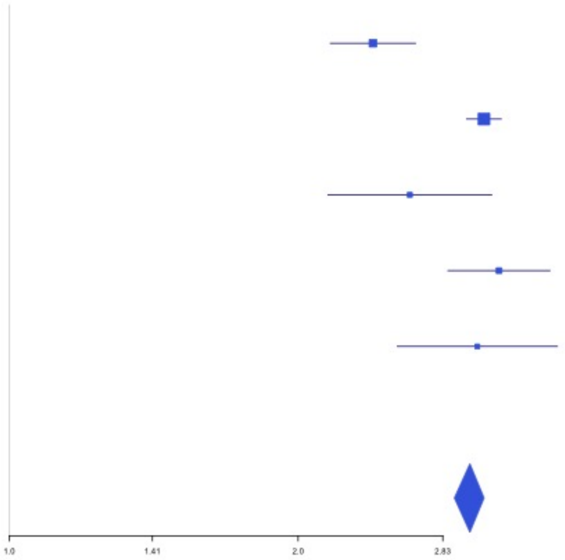


**Supplementary Figure 1.4** – rs2842700 (chr1:207282149) in locus 13 (C4BPA)

| Study | Cases | Controls | OR |
|---|---|---|---|
| INVENT-2019_EA | 15572 | 115280 | 1.18 |
| INVENT-2019_AA | 799 | 14353 | 1.04 |
| MVP_EA | 18952 | 411746 | 1.16 |
| MVP_AA | 5521 | 103791 | 1.07 |
| MVP_HIS | 1188 | 45634 | 1.13 |
| FinnGen_EA | 9176 | 209616 | 1.13 |
| EGP_EA | 4122 | 181553 | 1.02 |
| **Discovery** | **55330** | **1081973** | **1.11** |

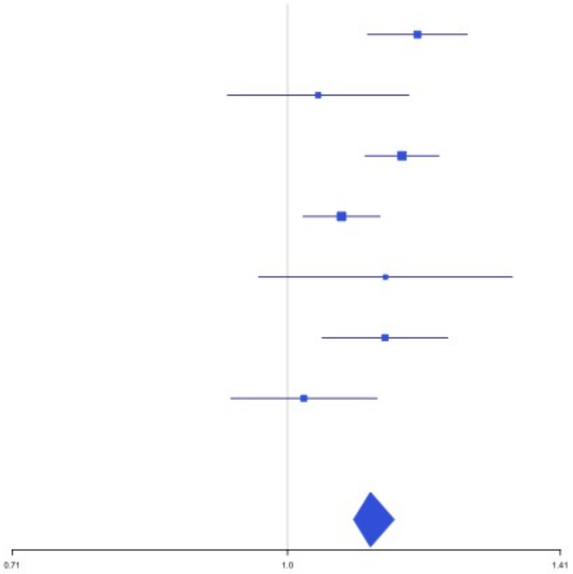

**Supplementary Figure 1.5** – rs3811444 (chr1:248039451) in locus 14 (TRIM58)

| Study | Cases | Controls | OR |
|---|---|---|---|
| INVENT-2019_EA | 15572 | 115280 | 0.99 |
| INVENT-2019_AA | 799 | 14353 | 1.02 |
| MVP_EA | 18952 | 411746 | 0.94 |
| MVP_AA | 5521 | 103791 | 1 |
| FinnGen_EA | 9176 | 209616 | 0.97 |
| EGP_EA | 4122 | 181553 | 0.95 |
| **Discovery** | **54142** | **1036339** | **0.96** |

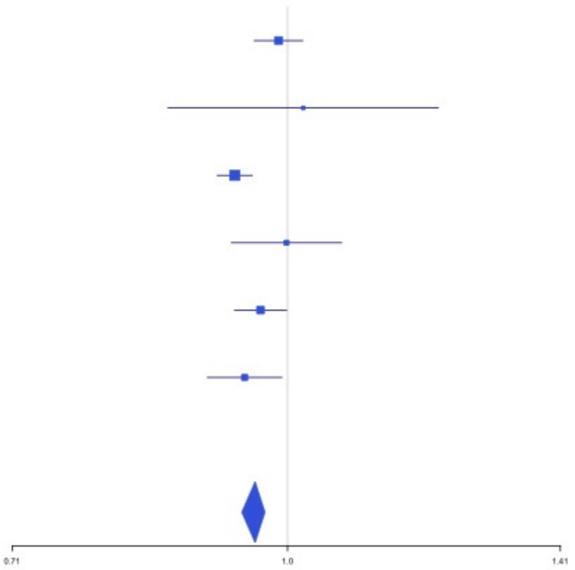


**Supplementary Figure 1.6** – rs545891600 (chr2:5378301) in locus 15 (LINC01249;LINC01248)

| Study | Cases | Controls | OR |
|---|---|---|---|
| MVP_EA | 18952 | 411746 | 0.61 |
| MVP_AA | 5521 | 103791 | 0.5 |
| MVP_HIS | 1188 | 45634 | 3.33 |
| **Discovery** | **25661** | **561171** | **0.5** |

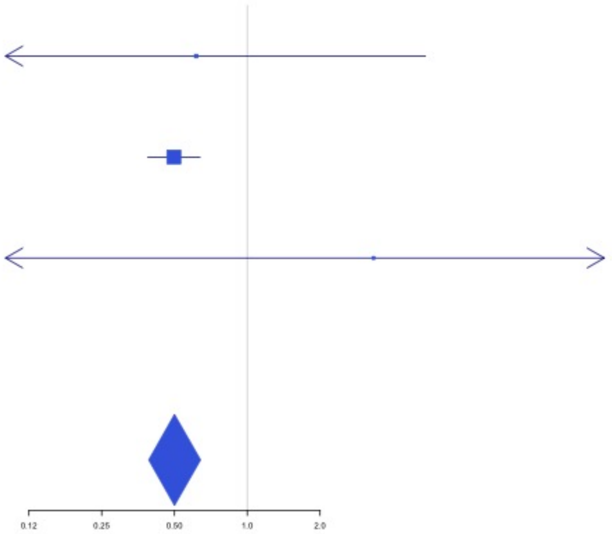

**Supplementary Figure 1.7** – rs7600986 (chr2:68636923) in locus 17 (PLEK ;FBXO48)

| Study | Cases | Controls | OR |
|---|---|---|---|
| INVENT-2019_EA | 15572 | 115280 | 1.05 |
| INVENT-2019_AA | 799 | 14353 | 0.86 |
| MVP_EA | 18952 | 411746 | 1.05 |
| MVP_AA | 5521 | 103791 | 1.1 |
| MVP_HIS | 1188 | 45634 | 1.08 |
| FinnGen_EA | 9176 | 209616 | 1.07 |
| EGP_EA | 4122 | 181553 | 1.05 |
| **Discovery** | **55330** | **1081973** | **1.06** |

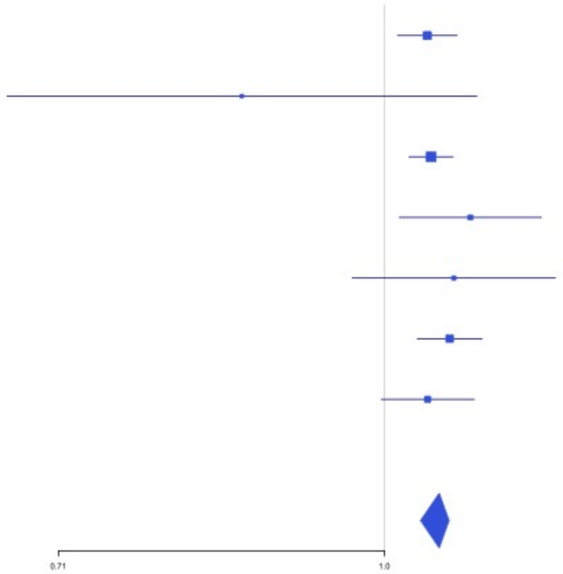


**Supplementary Figure 1.8** – rs182293241 (chr2:128029746) in locus 18 (ERCC3)

| Study | Cases | Controls | OR |
|---|---|---|---|
| MVP_AA | 5521 | 103791 | 1.89 |
| MVP_HIS | 1188 | 45634 | 1.83 |
| **Discovery** | **6709** | **149425** | **1.89** |

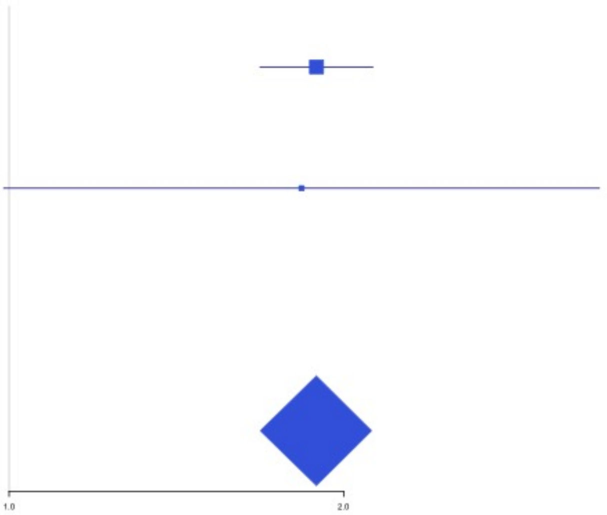

**Supplementary Figure 1.9** – rs6719550 (chr2:188272460) in locus 19 (CALCRL)

| Study | Cases | Controls | OR |
|---|---|---|---|
| INVENT-2019_EA | 15572 | 115280 | 1.04 |
| INVENT-2019_AA | 799 | 14353 | 1.13 |
| MVP_EA | 18952 | 411746 | 1.05 |
| MVP_AA | 5521 | 103791 | 1.05 |
| MVP_HIS | 1188 | 45634 | 1.08 |
| FinnGen_EA | 9176 | 209616 | 1.02 |
| EGP_EA | 4122 | 181553 | 1.03 |
| **Discovery** | **55330** | **1081973** | **1.04** |

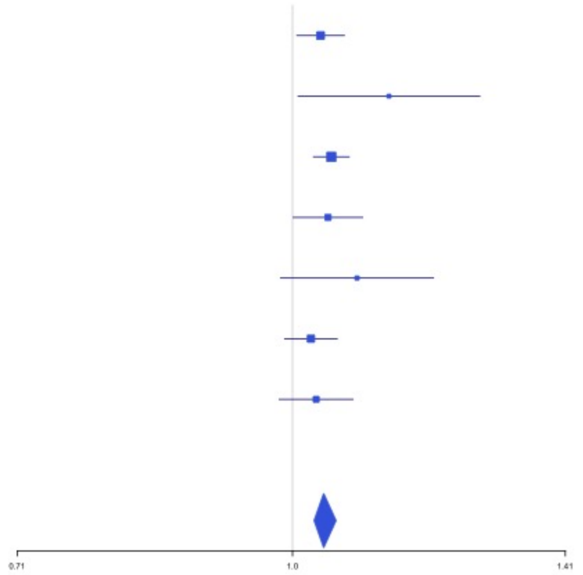


**Supplementary Figure 1.10** – rs715 (chr2:211543055) in locus 21 (CPS1)

| Study | Cases | Controls | OR |
|---|---|---|---|
| INVENT-2019_EA | 15572 | 115280 | 0.96 |
| INVENT-2019_AA | 799 | 14353 | 1 |
| MVP_EA | 18952 | 411746 | 0.95 |
| MVP_AA | 5521 | 103791 | 0.98 |
| MVP_HIS | 1188 | 45634 | 1.05 |
| FinnGen_EA | 9176 | 209616 | 0.93 |
| EGP_EA | 4122 | 181553 | 0.96 |
| **Discovery** | **55330** | **1081973** | **0.95** |

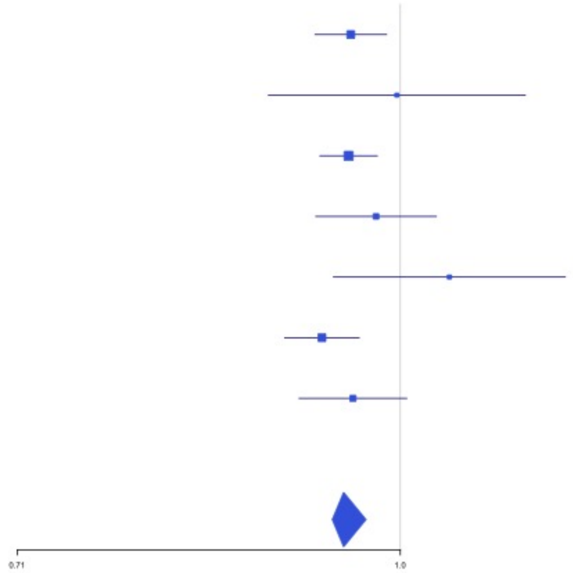

**Supplementary Figure 1.11** – rs13412535 (chr2:224874874) in locus 22 (SERPINE2)

| Study | Cases | Controls | OR |
|---|---|---|---|
| INVENT-2019_EA | 15572 | 115280 | 1.07 |
| INVENT-2019_AA | 799 | 14353 | 1.15 |
| MVP_EA | 18952 | 411746 | 1.07 |
| MVP_AA | 5521 | 103791 | 1.04 |
| MVP_HIS | 1188 | 45634 | 1.09 |
| FinnGen_EA | 9176 | 209616 | 1.05 |
| EGP_EA | 4122 | 181553 | 1.02 |
| **Discovery** | **55330** | **1081973** | **1.06** |

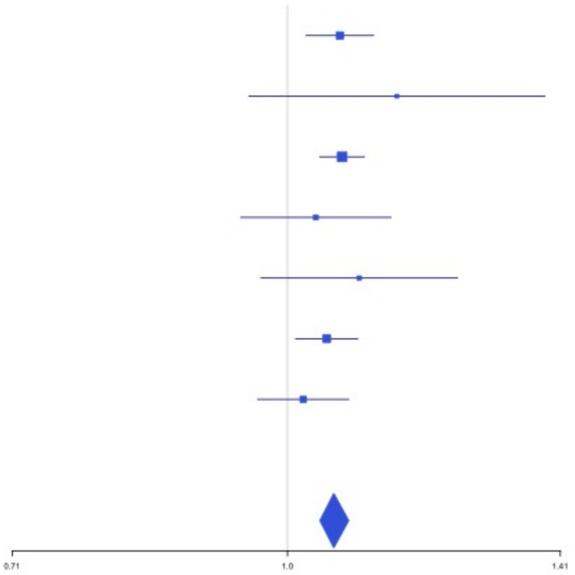


**Supplementary Figure 1.12** – rs2960420 (chr3:12314512) in locus 23 (SYN2;PPARG)

| Study | Cases | Controls | OR |
|---|---|---|---|
| INVENT-2019_EA | 15572 | 115280 | 1.03 |
| INVENT-2019_AA | 799 | 14353 | 1.02 |
| MVP_EA | 18952 | 411746 | 1.05 |
| MVP_AA | 5521 | 103791 | 1.03 |
| MVP_HIS | 1188 | 45634 | 1.09 |
| FinnGen_EA | 9176 | 209616 | 1.04 |
| EGP_EA | 4122 | 181553 | 1.03 |
| **Discovery** | **55330** | **1081973** | **1.04** |

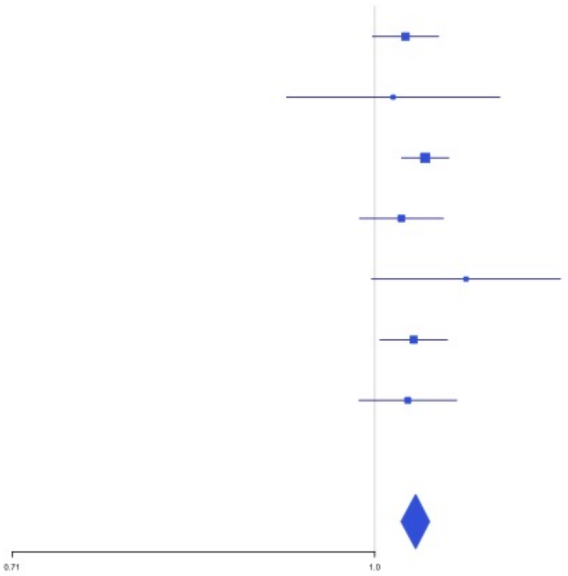

**Supplementary Figure 1.13** – rs13084580 (chr3:39188182) in locus 24 (CSRNP1)

| Study | Cases | Controls | OR |
|---|---|---|---|
| INVENT-2019_EA | 15572 | 115280 | 1.08 |
| INVENT-2019_AA | 799 | 14353 | 1.07 |
| MVP_EA | 18952 | 411746 | 1.1 |
| MVP_AA | 5521 | 103791 | 1.08 |
| MVP_HIS | 1188 | 45634 | 1.01 |
| FinnGen_EA | 9176 | 209616 | 1.12 |
| EGP_EA | 4122 | 181553 | 1.05 |
| **Discovery** | **55330** | **1081973** | **1.09** |

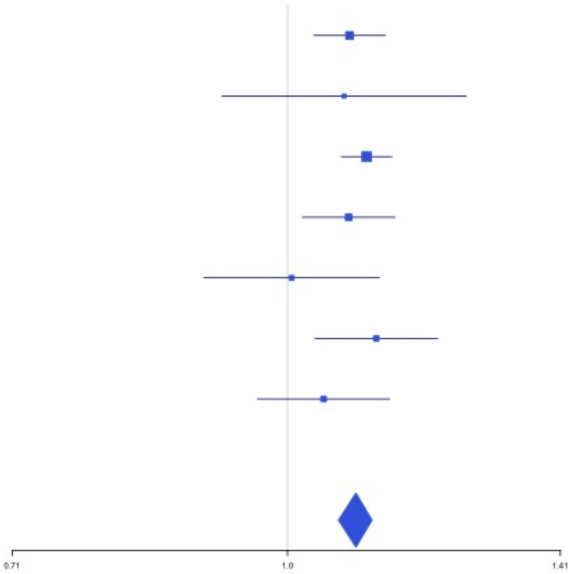


**Supplementary Figure 1.14** – rs11130326 (chr3:52771920) in locus 25 (NEK4)

| Study | Cases | Controls | OR |
|---|---|---|---|
| INVENT-2019_EA | 15572 | 115280 | 1.05 |
| INVENT-2019_AA | 799 | 14353 | 0.93 |
| MVP_EA | 18952 | 411746 | 1.04 |
| MVP_AA | 5521 | 103791 | 1.05 |
| MVP_HIS | 1188 | 45634 | 0.96 |
| FinnGen_EA | 9176 | 209616 | 1.05 |
| **Discovery** | **51208** | **900420** | **1.04** |

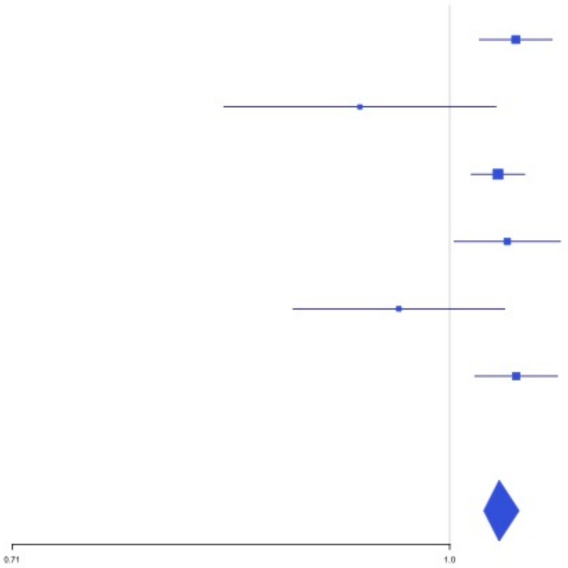

**Supplementary Figure 1.15** – rs562281690 (chr3:90177913) in locus 26 (EPHA3;NONE)

| Study | Cases | Controls | OR |
|---|---|---|---|
| MVP_EA | 18952 | 411746 | 2.08 |
| MVP_AA | 5521 | 103791 | 0.63 |
| MVP_HIS | 1188 | 45634 | 1.07 |
| FinnGen_EA | 9176 | 209616 | 1.86 |
| EGP_EA | 4122 | 181553 | 2.17 |
| **Discovery** | **38959** | **952340** | **2.01** |

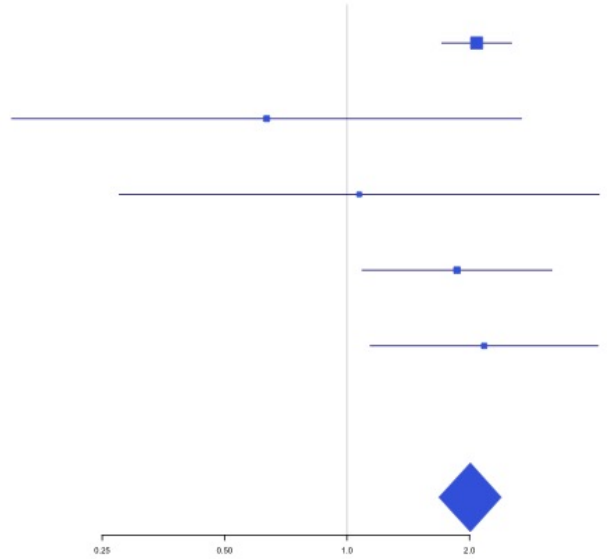


**Supplementary Figure 1.16** – rs79324379 (chr3:93712671) in locus 27 (ARL13B)

| Study | Cases | Controls | OR |
|---|---|---|---|
| MVP_EA | 18952 | 411746 | 2.07 |
| MVP_AA | 5521 | 103791 | 1.55 |
| MVP_HIS | 1188 | 45634 | 1.94 |
| **Discovery** | **25661** | **561171** | **1.57** |

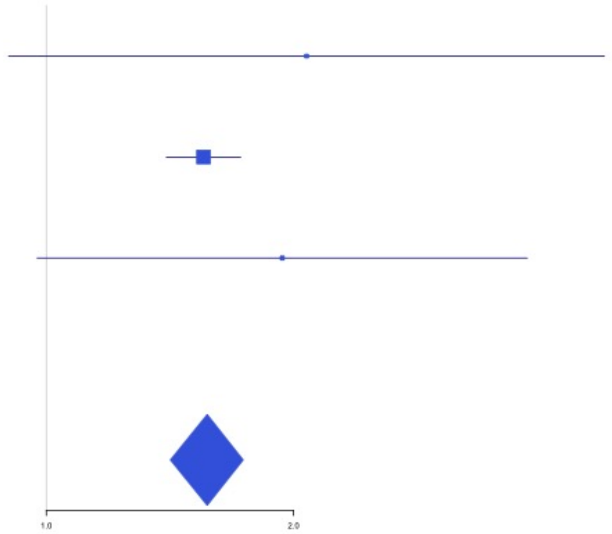

**Supplementary Figure 1.17** – rs9872572 (chr3:123108613) in locus 28 (ADCY5)

| Study | Cases | Controls | OR |
|---|---|---|---|
| INVENT-2019_EA | 15572 | 115280 | 1.04 |
| INVENT-2019_AA | 799 | 14353 | 1.03 |
| MVP_EA | 18952 | 411746 | 1.04 |
| MVP_AA | 5521 | 103791 | 1.02 |
| MVP_HIS | 1188 | 45634 | 1.01 |
| FinnGen_EA | 9176 | 209616 | 1.05 |
| EGP_EA | 4122 | 181553 | 1.04 |
| **Discovery** | **55330** | **1081973** | **1.04** |

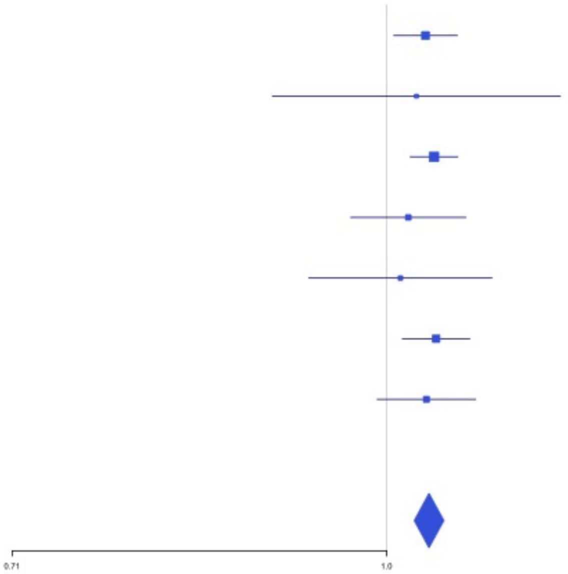


**Supplementary Figure 1.18** – rs62282204 (chr3:138584405) in locus 29 (PIK3CB ;LINC01391)

| Study | Cases | Controls | OR |
|---|---|---|---|
| INVENT-2019_EA | 15572 | 115280 | 0.96 |
| INVENT-2019_AA | 799 | 14353 | 1.01 |
| MVP_EA | 18952 | 411746 | 0.95 |
| MVP_AA | 5521 | 103791 | 0.96 |
| MVP_HIS | 1188 | 45634 | 1.01 |
| EGP_EA | 4122 | 181553 | 0.97 |
| **Discovery** | **46154** | **872357** | **0.96** |

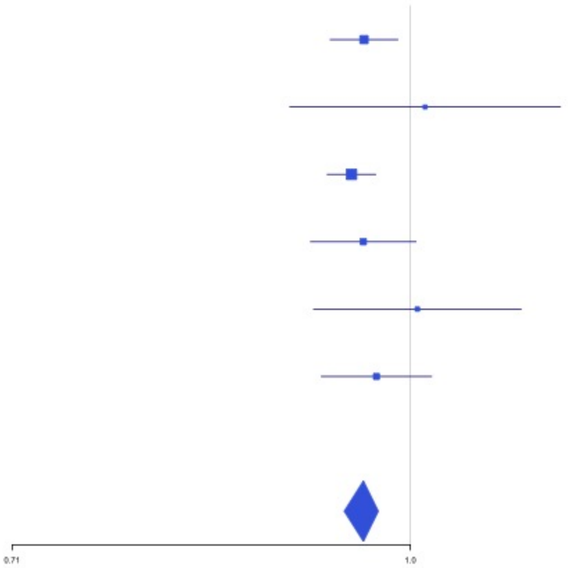

**Supplementary Figure 1.19** – rs7613621 (chr3:169191186) in locus 31 (MECOM)

| Study | Cases | Controls | OR |
|---|---|---|---|
| INVENT-2019_EA | 15572 | 115280 | 1.07 |
| INVENT-2019_AA | 799 | 14353 | 1.04 |
| MVP_EA | 18952 | 411746 | 1.04 |
| MVP_AA | 5521 | 103791 | 1.02 |
| MVP_HIS | 1188 | 45634 | 1.03 |
| FinnGen_EA | 9176 | 209616 | 1.02 |
| EGP_EA | 4122 | 181553 | 1.04 |
| **Discovery** | **55330** | **1081973** | **1.04** |

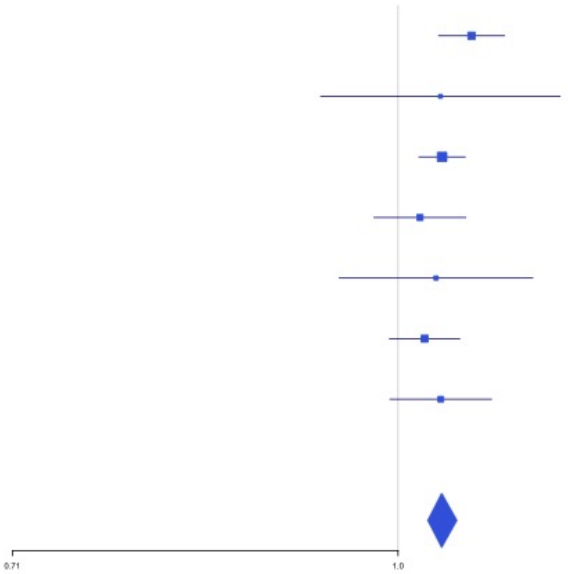


**Supplementary Figure 1.20** – rs710446 (chr3:186459927) in locus 32 (KNG1)

| Study | Cases | Controls | OR |
|---|---|---|---|
| INVENT-2019_EA | 15572 | 115280 | 0.94 |
| INVENT-2019_AA | 799 | 14353 | 0.95 |
| MVP_EA | 18952 | 411746 | 0.97 |
| MVP_AA | 5521 | 103791 | 0.98 |
| MVP_HIS | 1188 | 45634 | 0.95 |
| FinnGen_EA | 9176 | 209616 | 0.94 |
| EGP_EA | 4122 | 181553 | 0.96 |
| **Discovery** | **55330** | **1081973** | **0.96** |

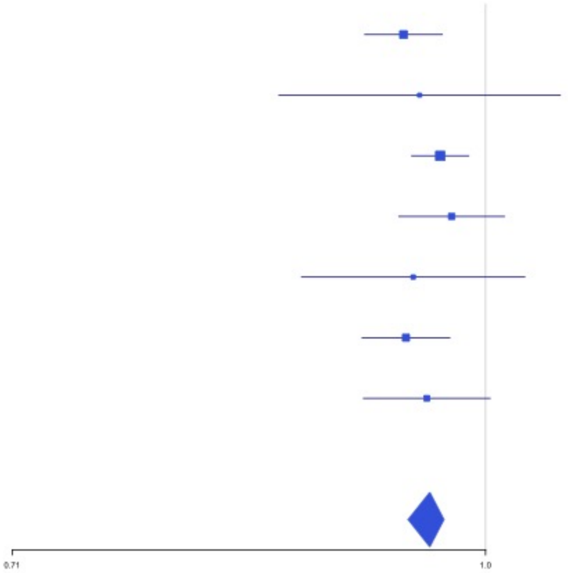

**Supplementary Figure 1.21** – rs6797948 (chr3:194784705) in locus 33 (LINC01968;XXYLT1)

| Study | Cases | Controls | OR |
|---|---|---|---|
| INVENT-2019_EA | 15572 | 115280 | 1.07 |
| INVENT-2019_AA | 799 | 14353 | 1.04 |
| MVP_EA | 18952 | 411746 | 1.07 |
| MVP_AA | 5521 | 103791 | 1.04 |
| FinnGen_EA | 9176 | 209616 | 1.04 |
| EGP_EA | 4122 | 181553 | 1.02 |
| **Discovery** | **54142** | **1036339** | **1.06** |

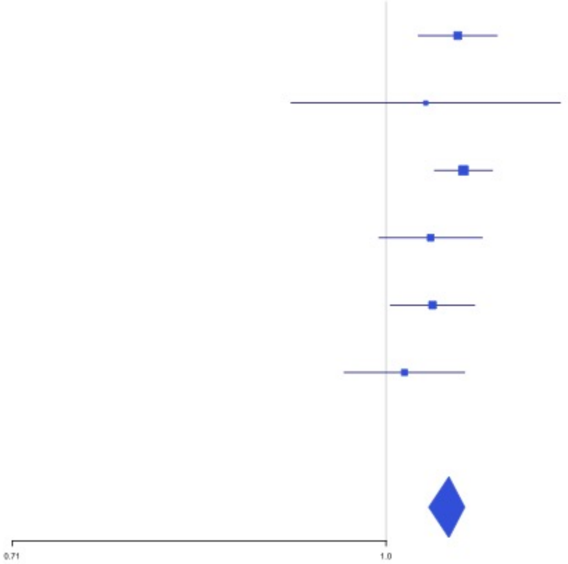


**Supplementary Figure 1.22** – rs6826579 (chr4:83785031) in locus 36 (SEC31A)

| Study | Cases | Controls | OR |
|---|---|---|---|
| INVENT-2019_EA | 15572 | 115280 | 1.05 |
| INVENT-2019_AA | 799 | 14353 | 1.07 |
| MVP_EA | 18952 | 411746 | 1.05 |
| MVP_AA | 5521 | 103791 | 1.03 |
| MVP_HIS | 1188 | 45634 | 0.99 |
| FinnGen_EA | 9176 | 209616 | 1.06 |
| EGP_EA | 4122 | 181553 | 1.05 |
| **Discovery** | **55330** | **1081973** | **1.05** |

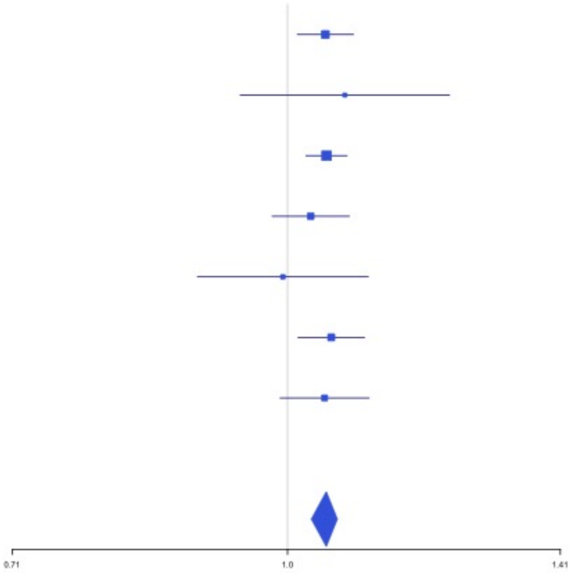

**Supplementary Figure 1.23** – rs17010957 (chr4:86719165) in locus 37 (ARHGAP24)

| Study | Cases | Controls | OR |
|---|---|---|---|
| INVENT-2019_EA | 15572 | 115280 | 1.04 |
| INVENT-2019_AA | 799 | 14353 | 1.17 |
| MVP_EA | 18952 | 411746 | 1.05 |
| MVP_AA | 5521 | 103791 | 1.05 |
| MVP_HIS | 1188 | 45634 | 1.18 |
| FinnGen_EA | 9176 | 209616 | 1.07 |
| EGP_EA | 4122 | 181553 | 1.09 |
| **Discovery** | **55330** | **1081973** | **1.06** |

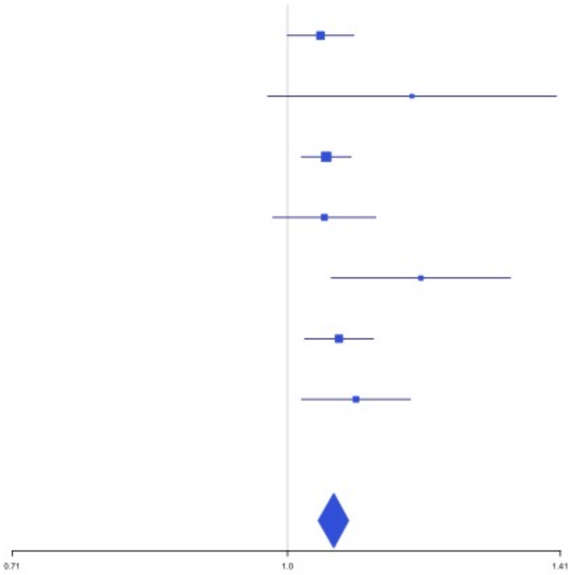


**Supplementary Figure 1.24** – rs2066864 (chr4:155525695) in locus 39 (FGG)

| Study | Cases | Controls | OR |
|---|---|---|---|
| INVENT-2019_EA | 15572 | 115280 | 1.2 |
| INVENT-2019_AA | 799 | 14353 | 1.17 |
| MVP_EA | 18952 | 411746 | 1.26 |
| MVP_AA | 5521 | 103791 | 1.19 |
| MVP_HIS | 1188 | 45634 | 1.26 |
| FinnGen_EA | 9176 | 209616 | 1.24 |
| EGP_EA | 4122 | 181553 | 1.18 |
| **Discovery** | **55330** | **1081973** | **1.23** |

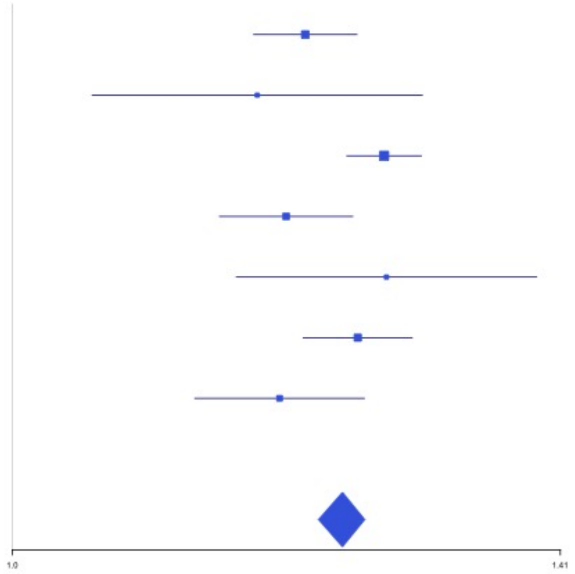

**Supplementary Figure 1.25** – rs3756011 (chr4:187206249) in locus 40 (F11)

| Study | Cases | Controls | OR |
|---|---|---|---|
| INVENT-2019_EA | 15572 | 115280 | 1.2 |
| INVENT-2019_AA | 799 | 14353 | 1.15 |
| MVP_EA | 18952 | 411746 | 1.26 |
| MVP_AA | 5521 | 103791 | 1.18 |
| MVP_HIS | 1188 | 45634 | 1.2 |
| FinnGen_EA | 9176 | 209616 | 1.23 |
| EGP_EA | 4122 | 181553 | 1.25 |
| **Discovery** | **55330** | **1081973** | **1.23** |

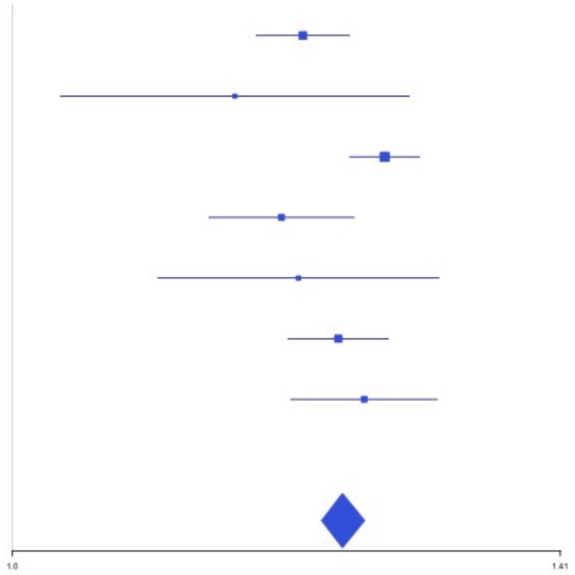


**Supplementary Figure 1.26** – rs16867574 (chr5:38708554) in locus 42 (OSMR-AS1)

| Study | Cases | Controls | OR |
|---|---|---|---|
| INVENT-2019_EA | 15572 | 115280 | 0.93 |
| INVENT-2019_AA | 799 | 14353 | 1.14 |
| MVP_EA | 18952 | 411746 | 0.95 |
| MVP_AA | 5521 | 103791 | 0.94 |
| MVP_HIS | 1188 | 45634 | 0.95 |
| FinnGen_EA | 9176 | 209616 | 0.99 |
| EGP_EA | 4122 | 181553 | 0.93 |
| **Discovery** | **55330** | **1081973** | **0.95** |

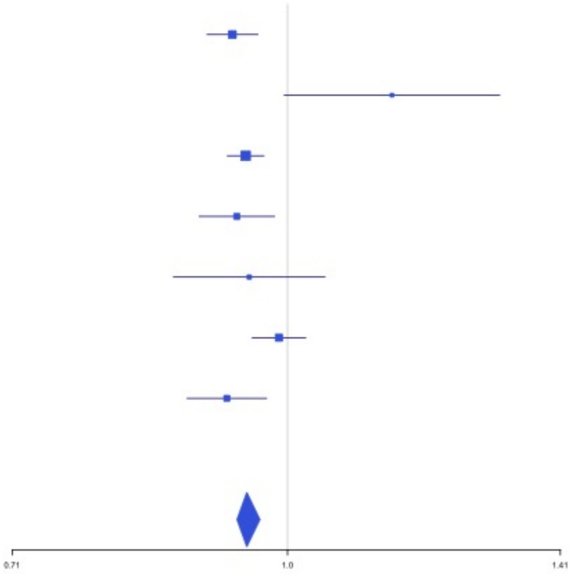

**Supplementary Figure 1.27** – rs38032 (chr5:96321887) in locus 44 (LNPEP)

| Study | Cases | Controls | OR |
|---|---|---|---|
| INVENT-2019_EA | 15572 | 115280 | 1.03 |
| INVENT-2019_AA | 799 | 14353 | 1.01 |
| MVP_EA | 18952 | 411746 | 1.05 |
| MVP_AA | 5521 | 103791 | 1.03 |
| MVP_HIS | 1188 | 45634 | 1.04 |
| FinnGen_EA | 9176 | 209616 | 1.05 |
| EGP_EA | 4122 | 181553 | 1.03 |
| **Discovery** | **55330** | **1081973** | **1.04** |

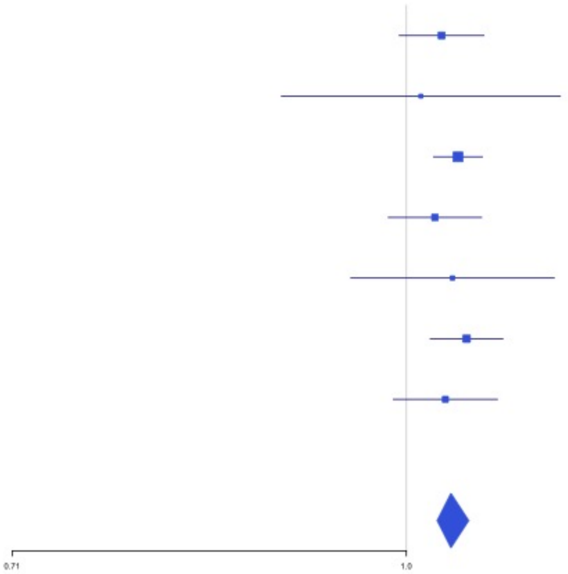


**Supplementary Figure 1.28** – rs9268881 (chr6:32431606) in locus 48 (HLA-DRA ;HLA-DRB5)

| Study | Cases | Controls | OR |
|---|---|---|---|
| INVENT-2019_EA | 15572 | 115280 | 0.96 |
| INVENT-2019_AA | 799 | 14353 | 0.97 |
| MVP_EA | 18952 | 411746 | 0.94 |
| MVP_AA | 5521 | 103791 | 1 |
| MVP_HIS | 1188 | 45634 | 0.99 |
| FinnGen_EA | 9176 | 209616 | 0.94 |
| EGP_EA | 4122 | 181553 | 0.98 |
| **Discovery** | **55330** | **1081973** | **0.96** |

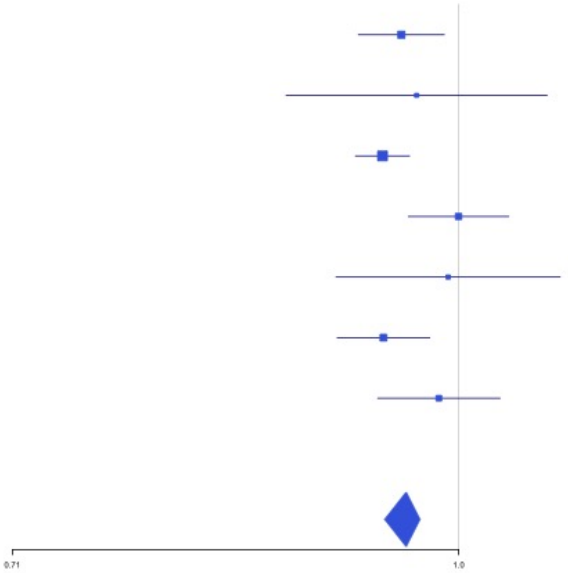

**Supplementary Figure 1.29** – rs145294670 (chr6:34622561) in locus 49 (ILRUN)

| Study | Cases | Controls | OR |
|---|---|---|---|
| INVENT-2019_EA | 15572 | 115280 | 1.03 |
| MVP_EA | 18952 | 411746 | 1.09 |
| MVP_AA | 5521 | 103791 | 1.17 |
| MVP_HIS | 1188 | 45634 | 1.02 |
| FinnGen_EA | 9176 | 209616 | 1.05 |
| EGP_EA | 4122 | 181553 | 1.05 |
| **Discovery** | **54531** | **1067620** | **1.06** |

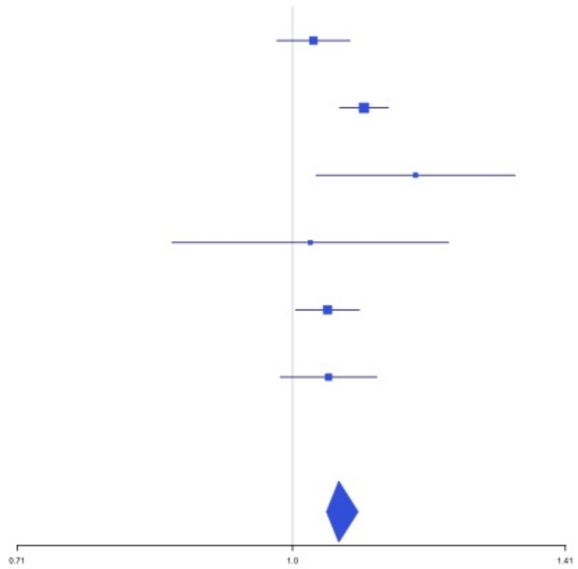


**Supplementary Figure 1.30** – rs903646797 (chr6:40985346) in locus 50 (LOC101929555)

| Study | Cases | Controls | OR |
|---|---|---|---|
| MVP_EA | 18952 | 411746 | 3.66 |
| MVP_AA | 5521 | 103791 | 3.05 |
| MVP_HIS | 1188 | 45634 | 0 |
| **Discovery** | **25661** | **561171** | **3.06** |

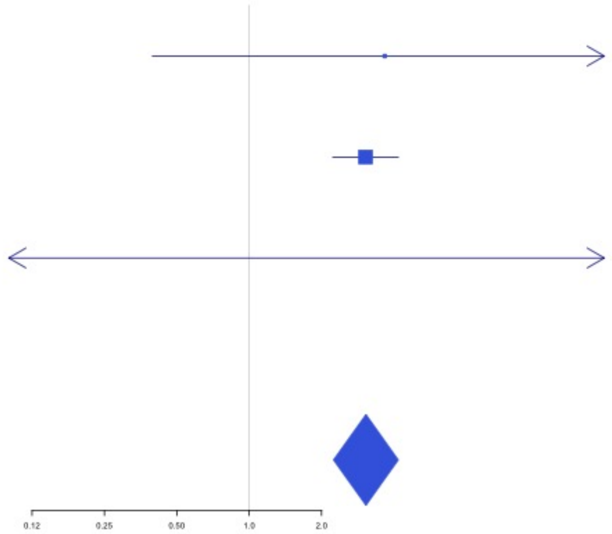

**Supplementary Figure 1.31** – rs9390460 (chr6:147694334) in locus 52 (STXBP5)

| Study | Cases | Controls | OR |
|---|---|---|---|
| INVENT-2019_EA | 15572 | 115280 | 0.94 |
| INVENT-2019_AA | 799 | 14353 | 1.11 |
| MVP_EA | 18952 | 411746 | 0.94 |
| MVP_AA | 5521 | 103791 | 0.93 |
| MVP_HIS | 1188 | 45634 | 0.93 |
| FinnGen_EA | 9176 | 209616 | 0.98 |
| EGP_EA | 4122 | 181553 | 0.98 |
| **Discovery** | **55330** | **1081973** | **0.95** |

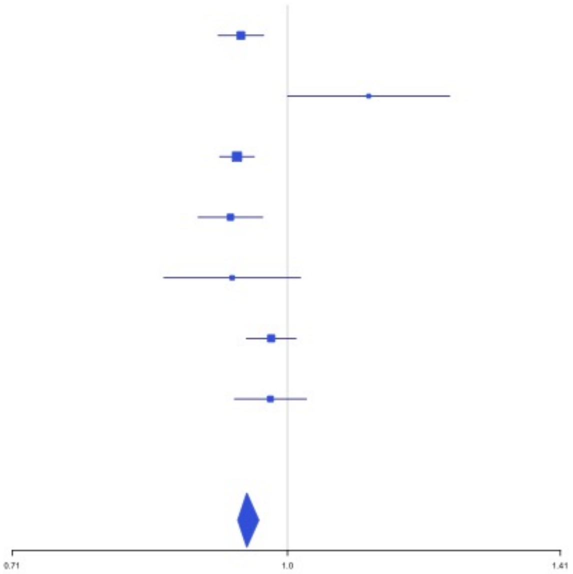


**Supplementary Figure 1.32** – rs42036 (chr7:92241451) in locus 54 (CDK6)

| Study | Cases | Controls | OR |
|---|---|---|---|
| INVENT-2019_EA | 15572 | 115280 | 0.95 |
| INVENT-2019_AA | 799 | 14353 | 1.01 |
| MVP_EA | 18952 | 411746 | 0.96 |
| MVP_AA | 5521 | 103791 | 0.93 |
| MVP_HIS | 1188 | 45634 | 0.93 |
| FinnGen_EA | 9176 | 209616 | 0.95 |
| EGP_EA | 4122 | 181553 | 0.95 |
| **Discovery** | **55330** | **1081973** | **0.96** |

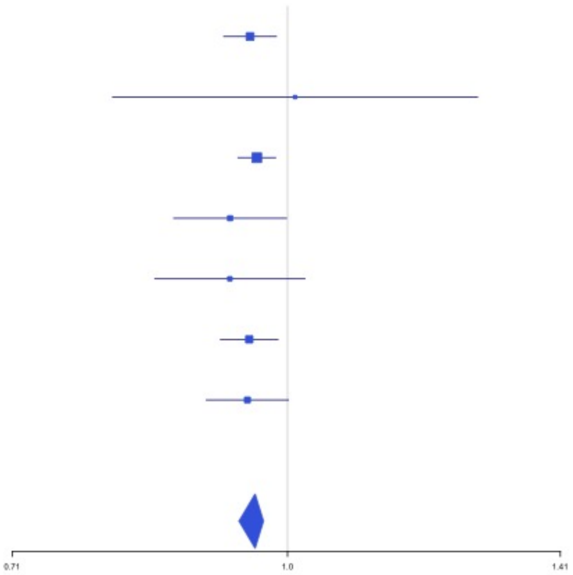

**Supplementary Figure 1.33** – rs538161377 (chr8:4206326) in locus 55 (CSMD1)

| Study | Cases | Controls | OR |
|---|---|---|---|
| MVP_AA | 5521 | 103791 | 0.35 |
| MVP_HIS | 1188 | 45634 | 0.46 |
| **Discovery** | **6709** | **149425** | **0.35** |

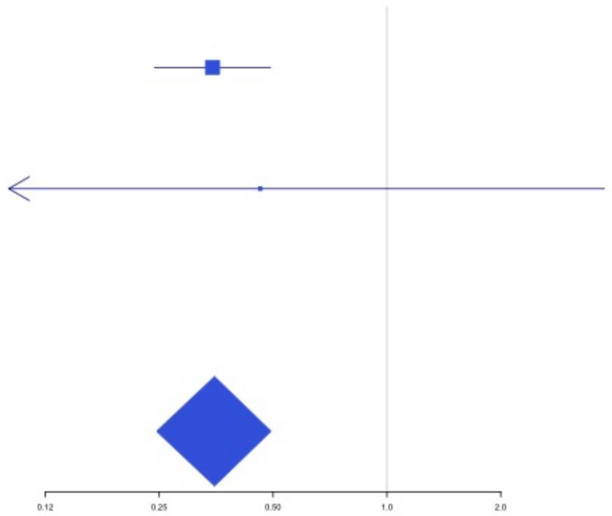


**Supplementary Figure 1.34** – rs67694436 (chr8:6654220) in locus 56 (AGPAT5;XKR5)

| Study | Cases | Controls | OR |
|---|---|---|---|
| INVENT-2019_EA | 15572 | 115280 | 0.97 |
| INVENT-2019_AA | 799 | 14353 | 0.98 |
| MVP_EA | 18952 | 411746 | 0.96 |
| MVP_AA | 5521 | 103791 | 0.98 |
| MVP_HIS | 1188 | 45634 | 1.06 |
| FinnGen_EA | 9176 | 209616 | 0.93 |
| EGP_EA | 4122 | 181553 | 0.96 |
| **Discovery** | **55330** | **1081973** | **0.96** |

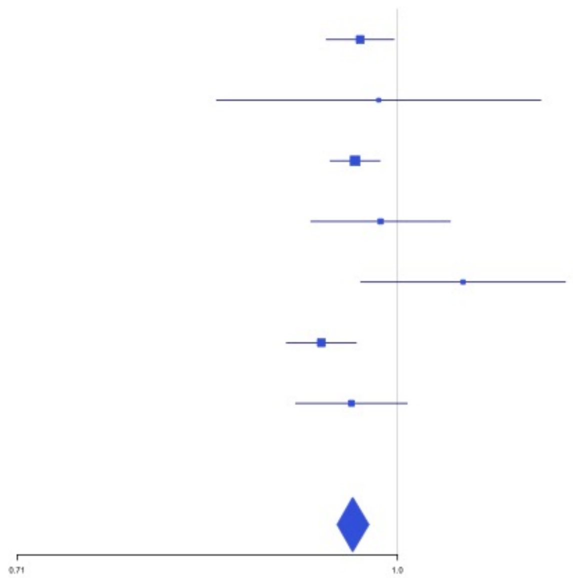

**Supplementary Figure 1.35** – rs7010926 (chr8:10626791) in locus 58 (PINX1)

| Study | Cases | Controls | OR |
|---|---|---|---|
| INVENT-2019_EA | 15572 | 115280 | 0.98 |
| INVENT-2019_AA | 799 | 14353 | 0.94 |
| MVP_EA | 18952 | 411746 | 0.95 |
| MVP_AA | 5521 | 103791 | 0.91 |
| MVP_HIS | 1188 | 45634 | 0.98 |
| FinnGen_EA | 9176 | 209616 | 0.96 |
| EGP_EA | 4122 | 181553 | 1.01 |
| **Discovery** | **55330** | **1081973** | **0.96** |

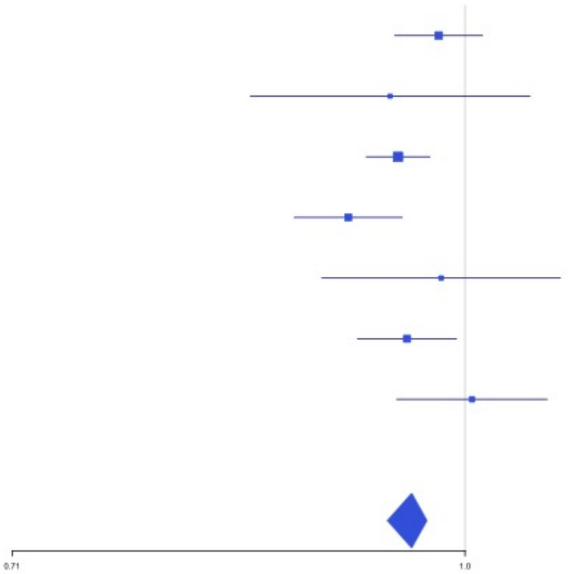


**Supplementary Figure 1.36** – rs2685417 (chr8:27807434) in locus 60 (SCARA5)

| Study | Cases | Controls | OR |
|---|---|---|---|
| INVENT-2019_EA | 15572 | 115280 | 1.04 |
| INVENT-2019_AA | 799 | 14353 | 1.09 |
| MVP_EA | 18952 | 411746 | 1.08 |
| MVP_AA | 5521 | 103791 | 1.01 |
| MVP_HIS | 1188 | 45634 | 1.08 |
| FinnGen_EA | 9176 | 209616 | 1.03 |
| EGP_EA | 4122 | 181553 | 1.11 |
| **Discovery** | **55330** | **1081973** | **1.06** |

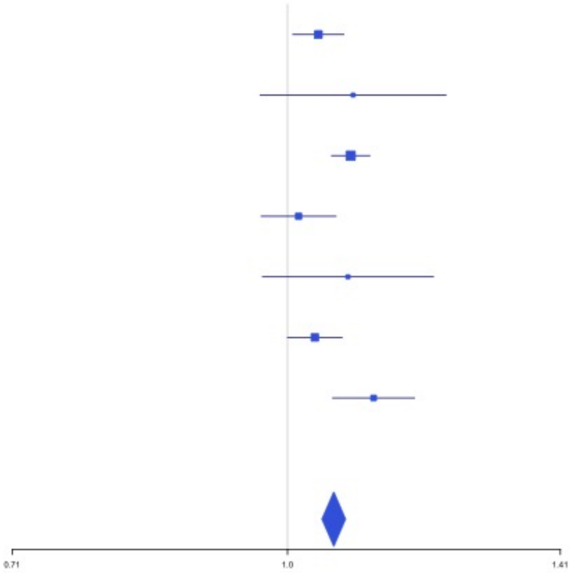

**Supplementary Figure 1.37** – rs6993770 (chr8:106581528) in locus 62 (ZFPM2)

| Study | Cases | Controls | OR |
|---|---|---|---|
| INVENT-2019_EA | 15572 | 115280 | 1.07 |
| INVENT-2019_AA | 799 | 14353 | 1.08 |
| MVP_EA | 18952 | 411746 | 1.1 |
| MVP_AA | 5521 | 103791 | 1.1 |
| MVP_HIS | 1188 | 45634 | 1.06 |
| FinnGen_EA | 9176 | 209616 | 1.05 |
| EGP_EA | 4122 | 181553 | 1.05 |
| **Discovery** | **55330** | **1081973** | **1.08** |

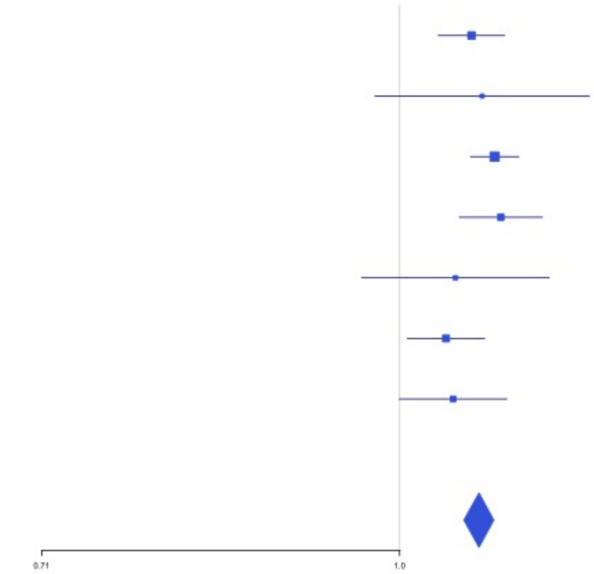


**Supplementary Figure 1.38** – rs77375493 (chr9:5073770) in locus 64 (JAK2)

| Study | Cases | Controls | OR |
|---|---|---|---|
| MVP_EA | 18952 | 411746 | 2.41 |
| MVP_AA | 5521 | 103791 | 2.47 |
| MVP_HIS | 1188 | 45634 | 1.43 |
| FinnGen_EA | 9176 | 209616 | 1.42 |
| **Discovery** | **34837** | **770787** | **2.14** |

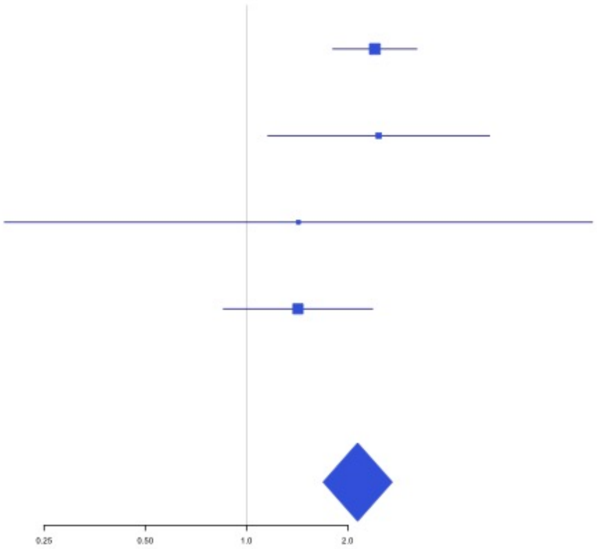

**Supplementary Figure 1.39** – rs35208412 (chr9:99194509) in locus 65 (ZNF367;HABP4)

| Study | Cases | Controls | OR |
|---|---|---|---|
| MVP_EA | 18952 | 411746 | 1.1 |
| MVP_AA | 5521 | 103791 | 1.07 |
| MVP_HIS | 1188 | 45634 | 1.08 |
| **Discovery** | **25661** | **561171** | **1.09** |

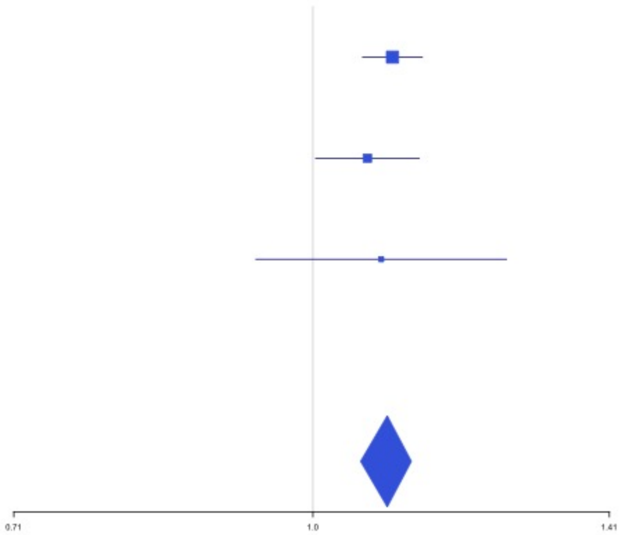


**Supplementary Figure 1.40** – rs505922 (chr9:136149229) in locus 66 (ABO)

| Study | Cases | Controls | OR |
|---|---|---|---|
| INVENT-2019_EA | 15572 | 115280 | 0.74 |
| INVENT-2019_AA | 799 | 14353 | 0.82 |
| MVP_EA | 18952 | 411746 | 0.71 |
| MVP_AA | 5521 | 103791 | 0.75 |
| MVP_HIS | 1188 | 45634 | 0.7 |
| FinnGen_EA | 9176 | 209616 | 0.78 |
| EGP_EA | 4122 | 181553 | 0.82 |
| **Discovery** | **55330** | **1081973** | **0.74** |

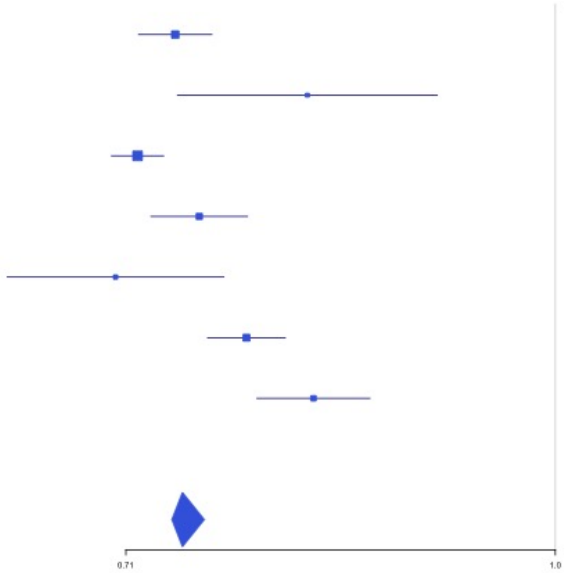

**Supplementary Figure 1.41** – rs1887091 (chr10:14535113) in locus 67 (MIR1265;FAM107B)

| Study | Cases | Controls | OR |
|---|---|---|---|
| INVENT-2019_EA | 15572 | 115280 | 0.95 |
| INVENT-2019_AA | 799 | 14353 | 0.97 |
| MVP_EA | 18952 | 411746 | 0.97 |
| MVP_AA | 5521 | 103791 | 0.96 |
| MVP_HIS | 1188 | 45634 | 0.94 |
| FinnGen_EA | 9176 | 209616 | 0.97 |
| EGP_EA | 4122 | 181553 | 0.96 |
| **Discovery** | **55330** | **1081973** | **0.96** |

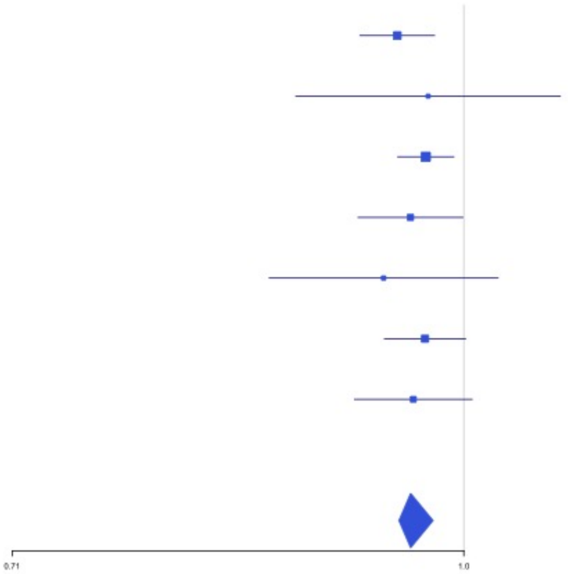


**Supplementary Figure 1.42** – rs529892067 (chr10:43568540) in locus 71 (MIR5100;RET)

| Study | Cases | Controls | OR |
|---|---|---|---|
| MVP_EA | 18952 | 411746 | 0.23 |
| MVP_HIS | 1188 | 45634 | 0.24 |
| **Discovery** | **20140** | **457380** | **0.24** |

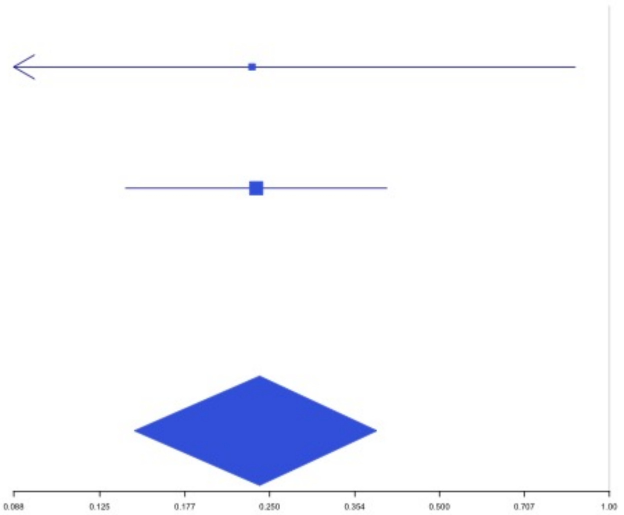

**Supplementary Figure 1.43** – rs17490626 (chr10:71218646) in locus 72 (TSPAN15)

| Study | Cases | Controls | OR |
|---|---|---|---|
| INVENT-2019_EA | 15572 | 115280 | 0.85 |
| INVENT-2019_AA | 799 | 14353 | 0.84 |
| MVP_EA | 18952 | 411746 | 0.75 |
| MVP_AA | 5521 | 103791 | 0.82 |
| MVP_HIS | 1188 | 45634 | 0.86 |
| FinnGen_EA | 9176 | 209616 | 0.83 |
| EGP_EA | 4122 | 181553 | 0.83 |
| **Discovery** | **55330** | **1081973** | **0.8** |

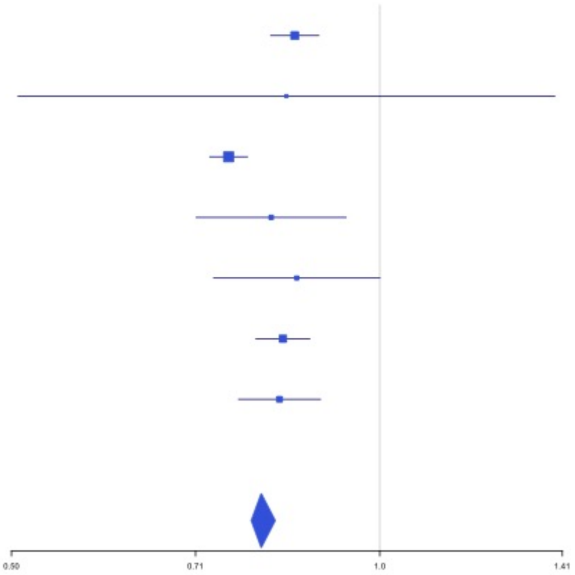


**Supplementary Figure 1.44** – rs16937003 (chr10:80938499) in locus 73 (ZMIZ1)

| Study | Cases | Controls | OR |
|---|---|---|---|
| INVENT-2019_EA | 15572 | 115280 | 1.15 |
| INVENT-2019_AA | 799 | 14353 | 1.03 |
| MVP_EA | 18952 | 411746 | 1.13 |
| MVP_AA | 5521 | 103791 | 1.24 |
| MVP_HIS | 1188 | 45634 | 1.04 |
| FinnGen_EA | 9176 | 209616 | 1.18 |
| EGP_EA | 4122 | 181553 | 1.11 |
| **Discovery** | **55330** | **1081973** | **1.15** |

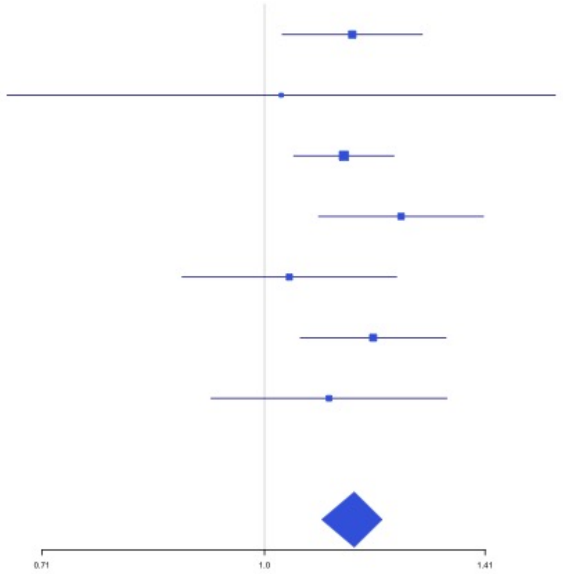

**Supplementary Figure 1.45** – rs2274224 (chr10:96039597) in locus 74 (PLCE1)

| Study | Cases | Controls | OR |
|---|---|---|---|
| INVENT-2019_EA | 15572 | 115280 | 1.03 |
| INVENT-2019_AA | 799 | 14353 | 1.17 |
| MVP_EA | 18952 | 411746 | 1.04 |
| MVP_AA | 5521 | 103791 | 1.05 |
| MVP_HIS | 1188 | 45634 | 0.98 |
| FinnGen_EA | 9176 | 209616 | 1.04 |
| EGP_EA | 4122 | 181553 | 1.04 |
| **Discovery** | **55330** | **1081973** | **1.04** |

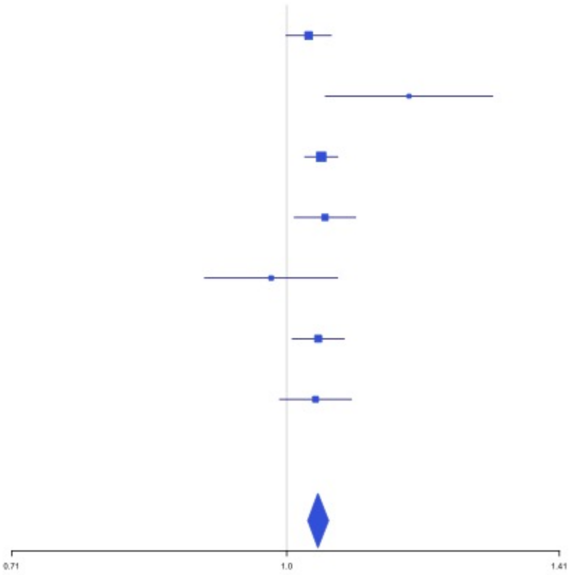


**Supplementary Figure 1.46** – rs10886430 (chr10:121010256) in locus 75 (GRK5)

| Study | Cases | Controls | OR |
|---|---|---|---|
| INVENT-2019_EA | 15572 | 115280 | 0.9 |
| INVENT-2019_AA | 799 | 14353 | 1.43 |
| MVP_EA | 18952 | 411746 | 0.86 |
| MVP_AA | 5521 | 103791 | 0.89 |
| MVP_HIS | 1188 | 45634 | 0.83 |
| FinnGen_EA | 9176 | 209616 | 0.91 |
| EGP_EA | 4122 | 181553 | 0.92 |
| **Discovery** | **55330** | **1081973** | **0.89** |

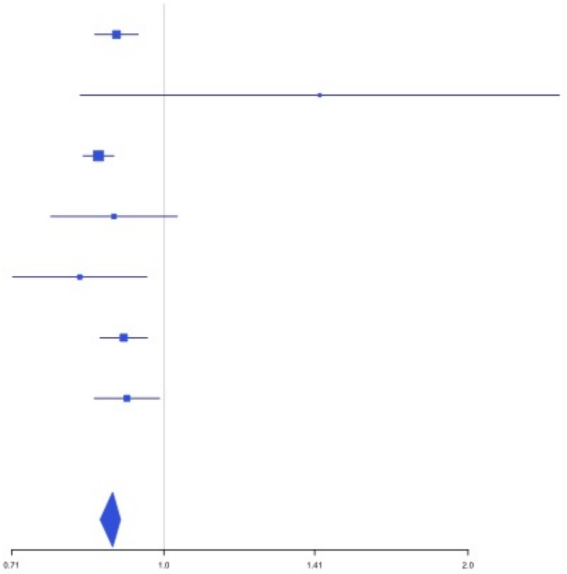

**Supplementary Figure 1.47** – rs11032074 (chr11:32993887) in locus 78 (QSER1)

| Study | Cases | Controls | OR |
|---|---|---|---|
| INVENT-2019_EA | 15572 | 115280 | 1.06 |
| INVENT-2019_AA | 799 | 14353 | 0.95 |
| MVP_EA | 18952 | 411746 | 1.05 |
| MVP_AA | 5521 | 103791 | 1.04 |
| MVP_HIS | 1188 | 45634 | 1.07 |
| FinnGen_EA | 9176 | 209616 | 1.06 |
| EGP_EA | 4122 | 181553 | 1.04 |
| **Discovery** | **55330** | **1081973** | **1.05** |

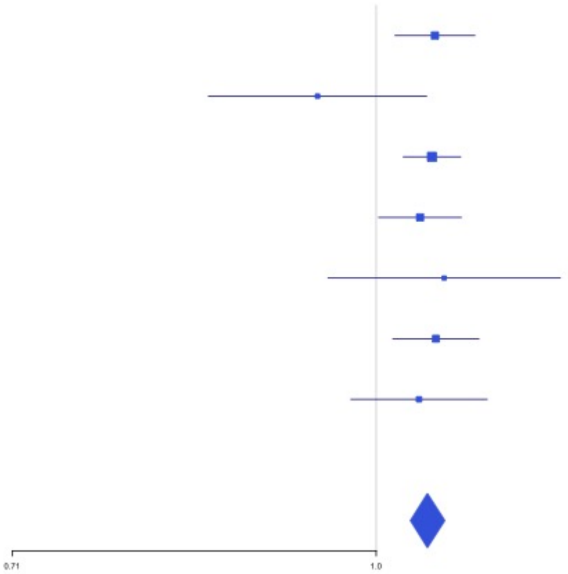


**Supplementary Figure 1.48** – rs1799963 (chr11:46761055) in locus 79 (F2)

| Study | Cases | Controls | OR |
|---|---|---|---|
| INVENT-2019_EA | 15572 | 115280 | 1.6 |
| MVP_EA | 18952 | 411746 | 2.08 |
| MVP_AA | 5521 | 103791 | 1.73 |
| MVP_HIS | 1188 | 45634 | 1.58 |
| FinnGen_EA | 9176 | 209616 | 3.42 |
| EGP_EA | 4122 | 181553 | 1.94 |
| **Discovery** | **54531** | **1067620** | **2.05** |

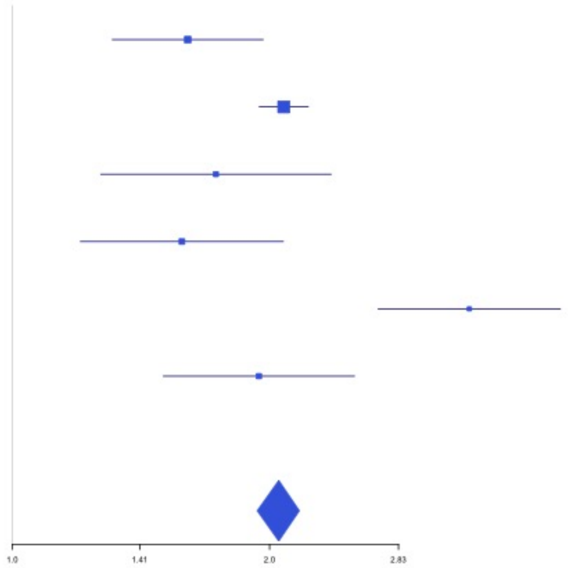

**Supplementary Figure 1.49** – rs141687379 (chr11:56666822) in locus 80 (FADS2B)

| **Study** | **Cases** | **Controls** | **OR** |
|---|---|---|---|
| MVP_EA | 18952 | 411746 | 0.55 |
| MVP_AA | 5521 | 103791 | 0.83 |
| MVP_HIS | 1188 | 45634 | 1.37 |
| FinnGen_EA | 9176 | 209616 | 0.3 |
| EGP_EA | 4122 | 181553 | 0.47 |
| **Discovery** | **38959** | **952340** | **0.52** |

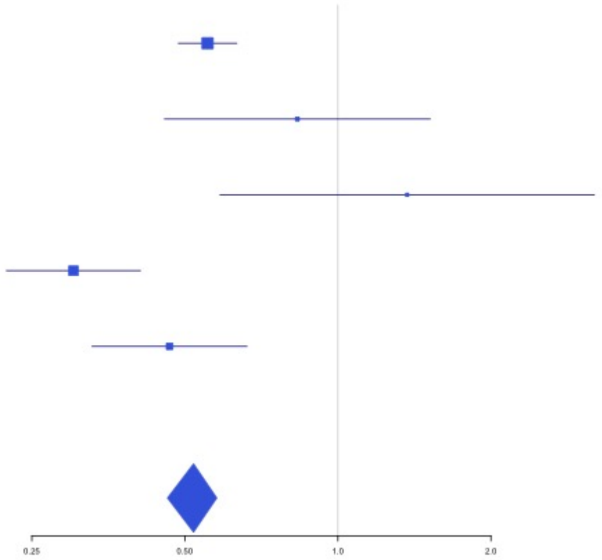


**Supplementary Figure 1.50** – rs174551 (chr11:61573684) in locus 82 (FADS1)

| **Study** | **Cases** | **Controls** | **OR** |
|---|---|---|---|
| INVENT-2019_EA | 15572 | 115280 | 1.05 |
| INVENT-2019_AA | 799 | 14353 | 0.97 |
| MVP_EA | 18952 | 411746 | 1.09 |
| MVP_AA | 5521 | 103791 | 1.05 |
| MVP_HIS | 1188 | 45634 | 1.08 |
| FinnGen_EA | 9176 | 209616 | 1.06 |
| **Discovery** | **51208** | **900420** | **1.07** |

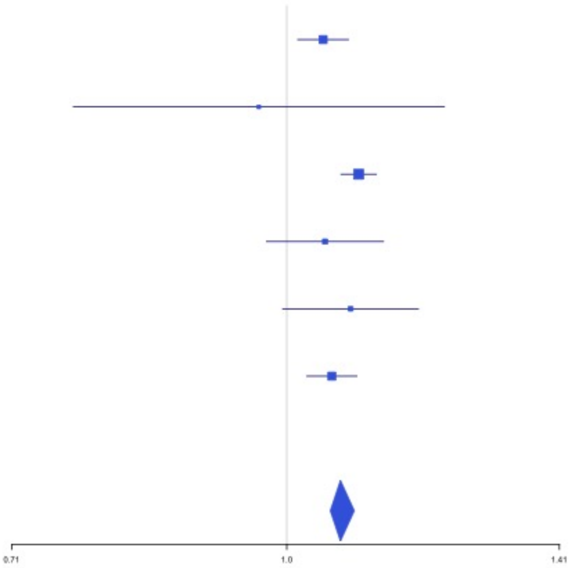

**Supplementary Figure 1.51** – rs35257264 (chr11:126296816) in locus 85 (ST3GAL4)

| Study | Cases | Controls | OR |
|---|---|---|---|
| INVENT-2019_EA | 15572 | 115280 | 1.2 |
| MVP_EA | 18952 | 411746 | 1.22 |
| MVP_AA | 5521 | 103791 | 1.42 |
| MVP_HIS | 1188 | 45634 | 1.53 |
| FinnGen_EA | 9176 | 209616 | 1.2 |
| EGP_EA | 4122 | 181553 | 1.1 |
| **Discovery** | **54531** | **1067620** | **1.21** |

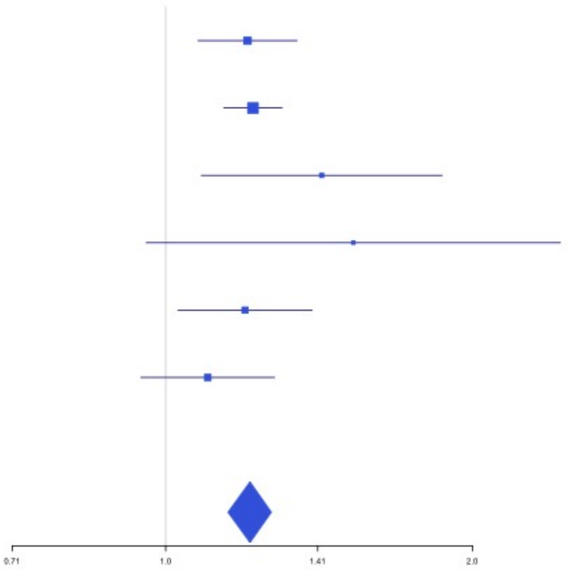


**Supplementary Figure 1.52** – rs1558519 (chr12:6153738) in locus 87 (VWF)

| Study | Cases | Controls | OR |
|---|---|---|---|
| INVENT-2019_EA | 15572 | 115280 | 0.92 |
| INVENT-2019_AA | 799 | 14353 | 0.99 |
| MVP_EA | 18952 | 411746 | 0.92 |
| MVP_AA | 5521 | 103791 | 0.96 |
| MVP_HIS | 1188 | 45634 | 1.01 |
| FinnGen_EA | 9176 | 209616 | 0.94 |
| EGP_EA | 4122 | 181553 | 0.96 |
| **Discovery** | **55330** | **1081973** | **0.93** |

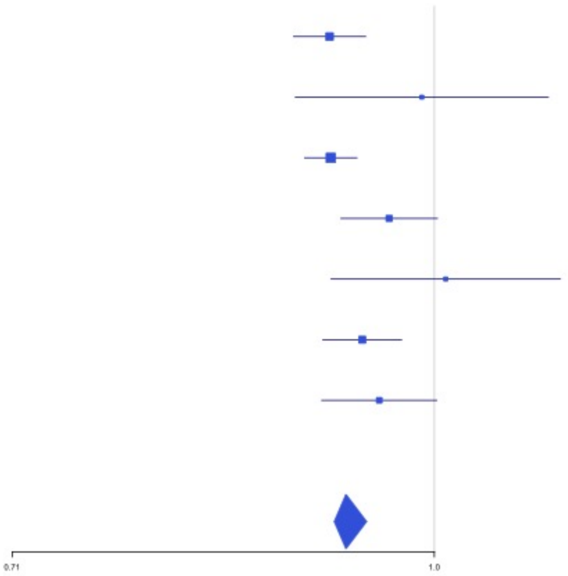

**Supplementary Figure 1.53** – rs7311483 (chr12:9053661) in locus 88 (A2ML1 ;PHC1)

| Study | Cases | Controls | OR |
|---|---|---|---|
| INVENT-2019_EA | 15572 | 115280 | 0.98 |
| INVENT-2019_AA | 799 | 14353 | 0.99 |
| MVP_EA | 18952 | 411746 | 0.95 |
| MVP_AA | 5521 | 103791 | 0.92 |
| MVP_HIS | 1188 | 45634 | 0.91 |
| FinnGen_EA | 9176 | 209616 | 0.95 |
| EGP_EA | 4122 | 181553 | 0.95 |
| **Discovery** | **55330** | **1081973** | **0.96** |

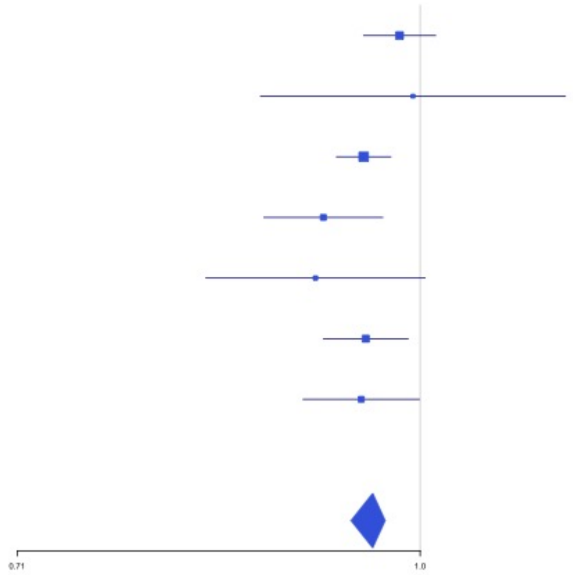


**Supplementary Figure 1.54** – rs6580981 (chr12:54723028) in locus 89 (COPZ1)

| Study | Cases | Controls | OR |
|---|---|---|---|
| INVENT-2019_EA | 15572 | 115280 | 0.96 |
| INVENT-2019_AA | 799 | 14353 | 1.03 |
| MVP_EA | 18952 | 411746 | 0.94 |
| MVP_AA | 5521 | 103791 | 0.97 |
| MVP_HIS | 1188 | 45634 | 0.95 |
| FinnGen_EA | 9176 | 209616 | 0.97 |
| EGP_EA | 4122 | 181553 | 0.98 |
| **Discovery** | **55330** | **1081973** | **0.96** |

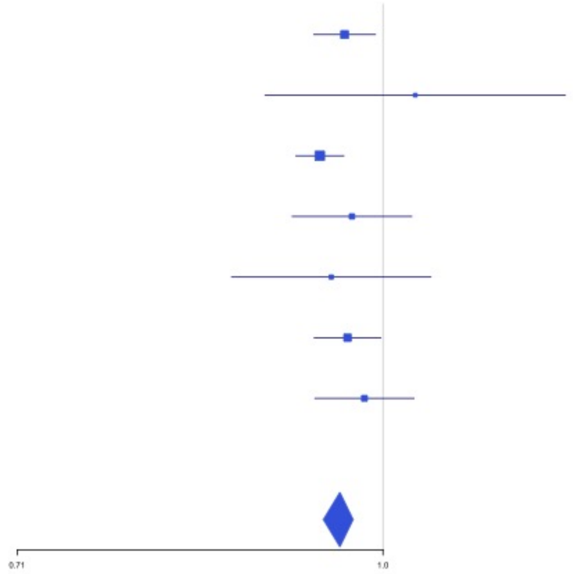

**Supplementary Figure 1.55** – rs142351376 (chr12:104136288) in locus 90 (STAB2)

| Study | Cases | Controls | OR |
|---|---|---|---|
| MVP_EA | 18952 | 411746 | 1.78 |
| FinnGen_EA | 9176 | 209616 | 1.67 |
| EGP_EA | 4122 | 181553 | 1.91 |
| **Discovery** | **32250** | **802915** | **1.68** |

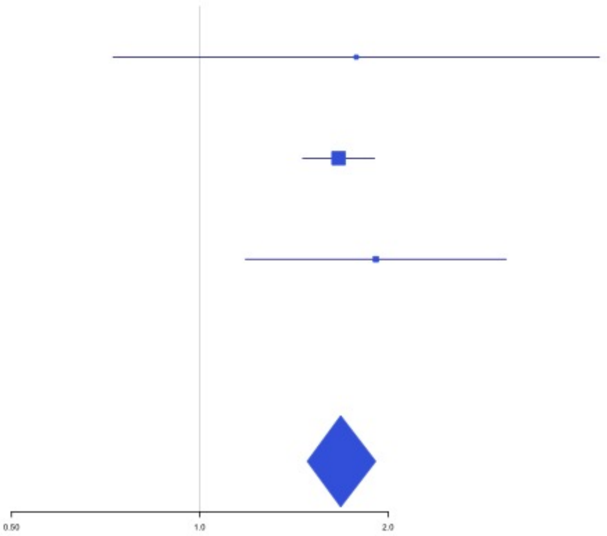


**Supplementary Figure 1.56** – rs3184504 (chr12:111884608) in locus 91 (SH2B3)

| Study | Cases | Controls | OR |
|---|---|---|---|
| INVENT-2019_EA | 15572 | 115280 | 1.04 |
| INVENT-2019_AA | 799 | 14353 | 0.99 |
| MVP_EA | 18952 | 411746 | 1.05 |
| MVP_AA | 5521 | 103791 | 1.02 |
| MVP_HIS | 1188 | 45634 | 1.05 |
| FinnGen_EA | 9176 | 209616 | 1.04 |
| EGP_EA | 4122 | 181553 | 1.05 |
| **Discovery** | **55330** | **1081973** | **1.05** |

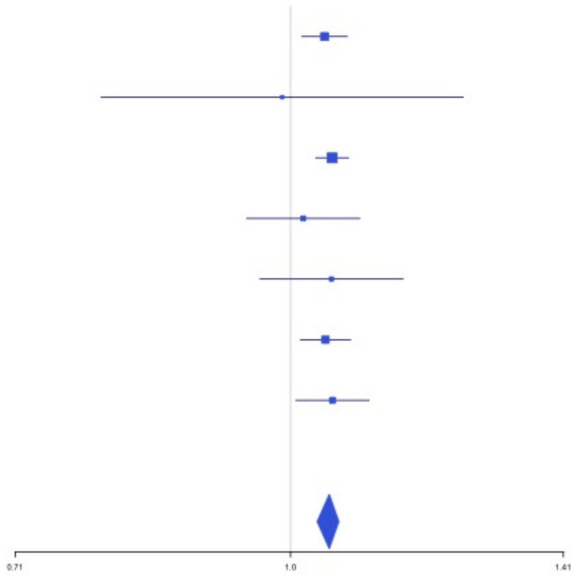

**Supplementary Figure 1.57** – rs75940120 (chr13:109686619) in locus 93 (MYO16)

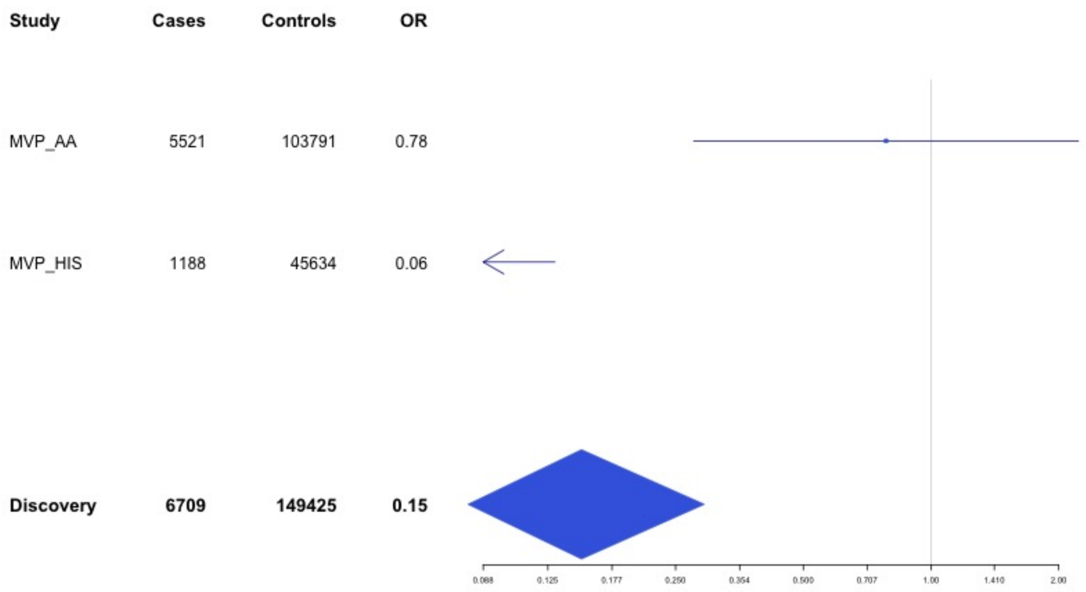


**Supplementary Figure 1.58** – rs3211752 (chr13:113787459) in locus 94 (F10)

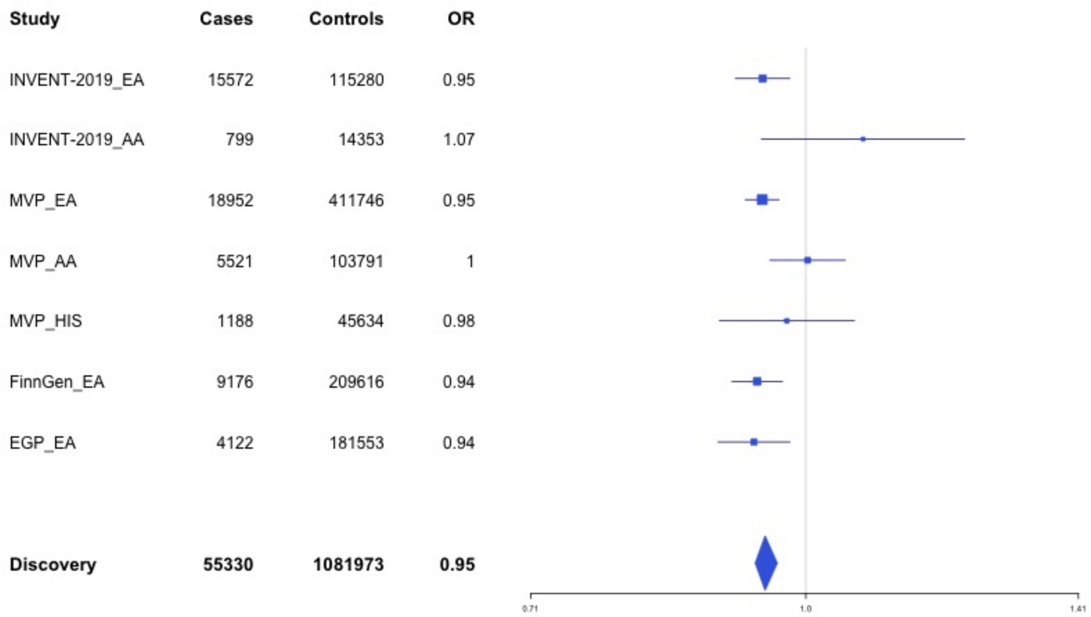

**Supplementary Figure 1.59** – rs57035593 (chr14:92268096) in locus 97 (TC2N)

| Study | Cases | Controls | OR |
|---|---|---|---|
| INVENT-2019_EA | 15572 | 115280 | 1.07 |
| INVENT-2019_AA | 799 | 14353 | 1 |
| MVP_EA | 18952 | 411746 | 1.07 |
| MVP_AA | 5521 | 103791 | 1.08 |
| MVP_HIS | 1188 | 45634 | 1.01 |
| FinnGen_EA | 9176 | 209616 | 1.06 |
| EGP_EA | 4122 | 181553 | 1.09 |
| **Discovery** | **55330** | **1081973** | **1.07** |

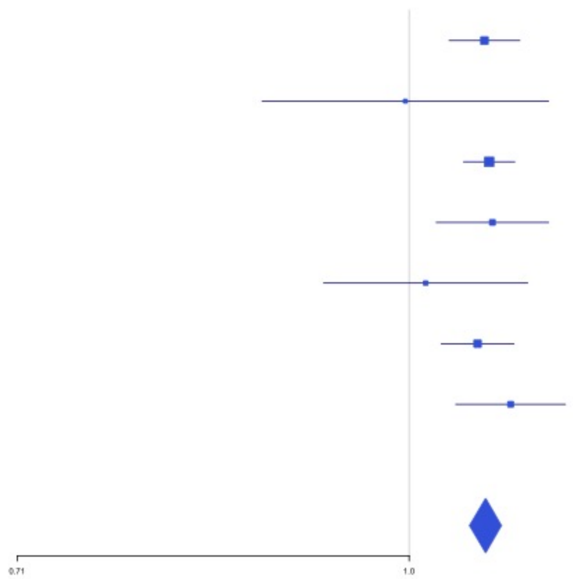


**Supplementary Figure 1.60** – rs8013957 (chr14:103140254) in locus 99 (RCOR1)

| Study | Cases | Controls | OR |
|---|---|---|---|
| INVENT-2019_EA | 15572 | 115280 | 1.03 |
| INVENT-2019_AA | 799 | 14353 | 1.11 |
| MVP_EA | 18952 | 411746 | 1.03 |
| MVP_AA | 5521 | 103791 | 1.05 |
| MVP_HIS | 1188 | 45634 | 1.18 |
| FinnGen_EA | 9176 | 209616 | 1.04 |
| EGP_EA | 4122 | 181553 | 1.06 |
| **Discovery** | **55330** | **1081973** | **1.04** |

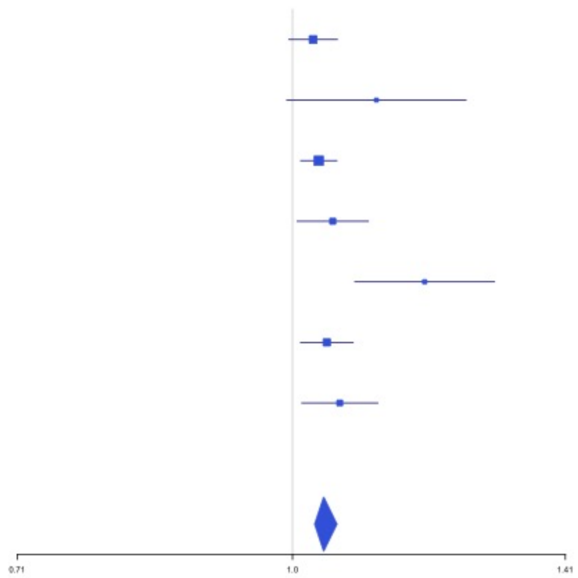

**Supplementary Figure 1.61** – rs55707100 (chr15:43820717) in locus 100 (MAP1A)

| Study | Cases | Controls | OR |
|---|---|---|---|
| INVENT-2019_EA | 15572 | 115280 | 0.92 |
| MVP_EA | 18952 | 411746 | 0.84 |
| MVP_AA | 5521 | 103791 | 1.09 |
| MVP_HIS | 1188 | 45634 | 0.66 |
| FinnGen_EA | 9176 | 209616 | 0.82 |
| EGP_EA | 4122 | 181553 | 0.9 |
| **Discovery** | **54531** | **1067620** | **0.87** |

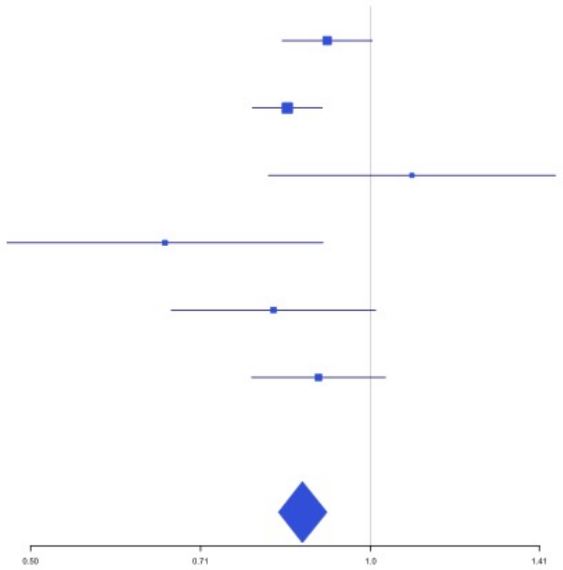


**Supplementary Figure 1.62** – rs59442804 (chr15:60899031) in locus 101 (RORA-AS1)

| Study | Cases | Controls | OR |
|---|---|---|---|
| INVENT-2019_EA | 15572 | 115280 | 0.96 |
| INVENT-2019_AA | 799 | 14353 | 1.01 |
| MVP_EA | 18952 | 411746 | 0.96 |
| MVP_AA | 5521 | 103791 | 0.96 |
| MVP_HIS | 1188 | 45634 | 0.92 |
| FinnGen_EA | 9176 | 209616 | 0.97 |
| EGP_EA | 4122 | 181553 | 0.95 |
| **Discovery** | **55330** | **1081973** | **0.96** |

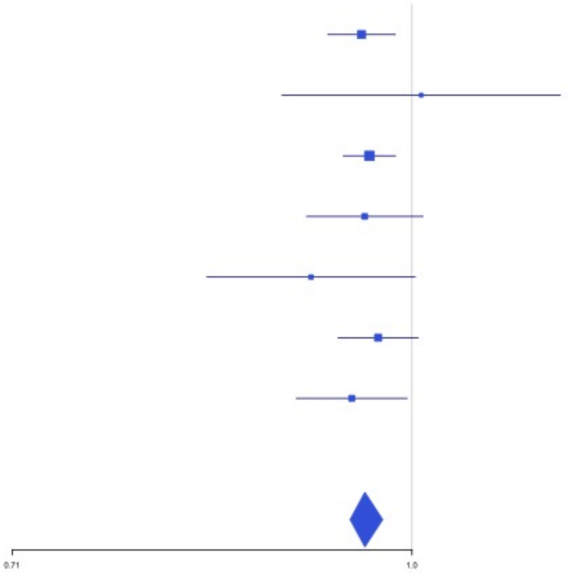

**Supplementary Figure 1.63** – rs182906510 (chr15:67372922) in locus 102 (SMAD3)

| Study | Cases | Controls | OR |
|---|---|---|---|
| MVP_EA | 18952 | 411746 | 6.99 |
| MVP_AA | 5521 | 103791 | 4.41 |
| MVP_HIS | 1188 | 45634 | 7.51 |
| **Discovery** | **25661** | **561171** | **5.95** |

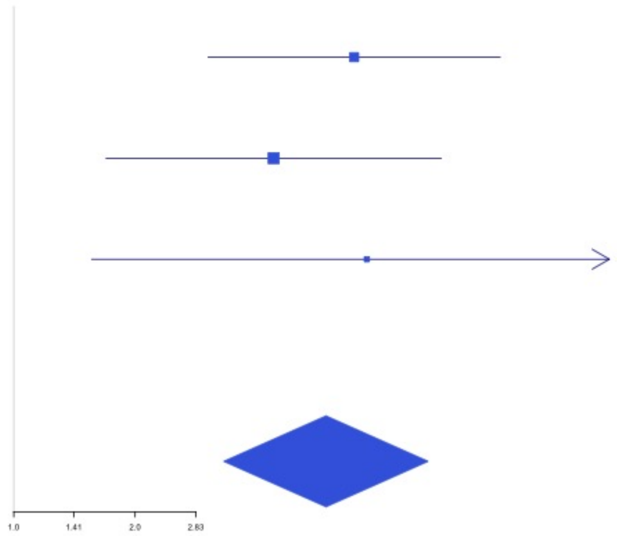


**Supplementary Figure 1.64** – rs12443808 (chr16:30996871) in locus 106 (HSD3B7)

| Study | Cases | Controls | OR |
|---|---|---|---|
| INVENT-2019_EA | 15572 | 115280 | 1.06 |
| INVENT-2019_AA | 799 | 14353 | 1.09 |
| MVP_EA | 18952 | 411746 | 1.06 |
| MVP_AA | 5521 | 103791 | 1.05 |
| MVP_HIS | 1188 | 45634 | 1.09 |
| FinnGen_EA | 9176 | 209616 | 1.05 |
| EGP_EA | 4122 | 181553 | 1.02 |
| **Discovery** | **55330** | **1081973** | **1.06** |

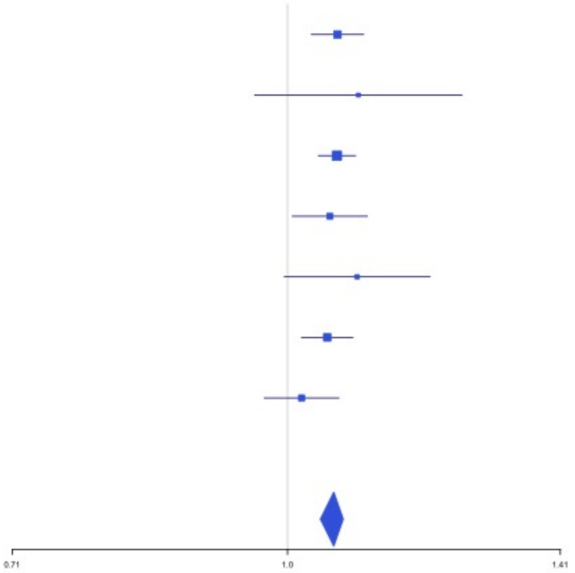

**Supplementary Figure 1.65** – rs56943275 (chr16:81898152) in locus 109 (PLCG2)

| Study | Cases | Controls | OR |
|---|---|---|---|
| INVENT-2019_EA | 15572 | 115280 | 1.1 |
| INVENT-2019_AA | 799 | 14353 | 1.22 |
| MVP_HIS | 1188 | 45634 | 1.09 |
| FinnGen_EA | 9176 | 209616 | 1.06 |
| EGP_EA | 4122 | 181553 | 1.08 |
| **Discovery** | **30857** | **566436** | **1.08** |

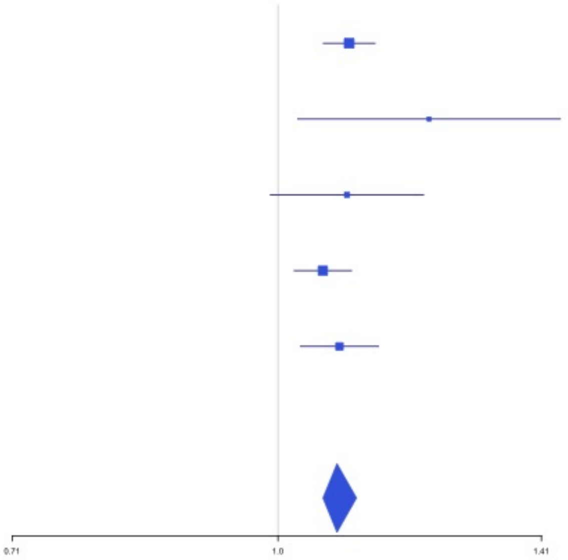


**Supplementary Figure 1.66** – rs28634651 (chr16:88553198) in locus 111 (ZFPM1)

| Study | Cases | Controls | OR |
|---|---|---|---|
| INVENT-2019_EA | 15572 | 115280 | 1.04 |
| INVENT-2019_AA | 799 | 14353 | 1.12 |
| MVP_EA | 18952 | 411746 | 1.06 |
| MVP_HIS | 1188 | 45634 | 1.05 |
| FinnGen_EA | 9176 | 209616 | 1.07 |
| **Discovery** | **45687** | **796629** | **1.06** |

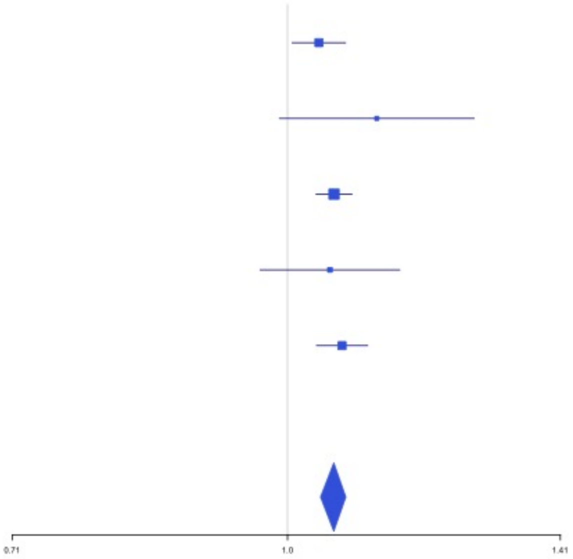

**Supplementary Figure 1.67** – rs6503222 (chr17:1977862) in locus 112 (SMG6)

| Study | Cases | Controls | OR |
|---|---|---|---|
| INVENT-2019_EA | 15572 | 115280 | 1.06 |
| INVENT-2019_AA | 799 | 14353 | 0.95 |
| MVP_EA | 18952 | 411746 | 1.06 |
| MVP_AA | 5521 | 103791 | 1.03 |
| MVP_HIS | 1188 | 45634 | 1.09 |
| FinnGen_EA | 9176 | 209616 | 1.03 |
| EGP_EA | 4122 | 181553 | 1.06 |
| **Discovery** | **55330** | **1081973** | **1.05** |

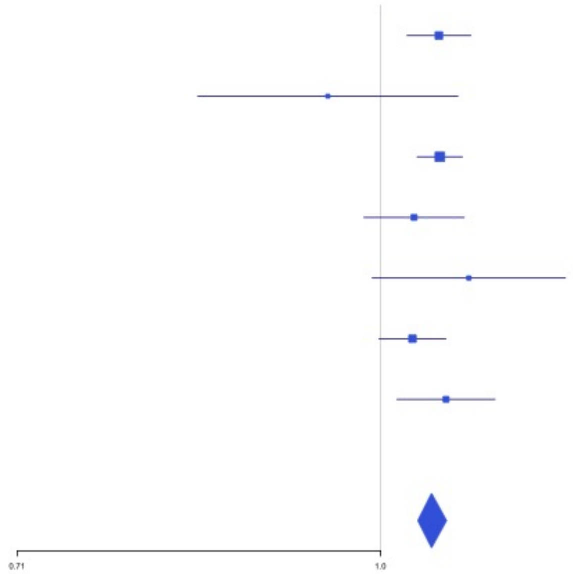


**Supplementary Figure 1.68** – rs7225756 (chr17:6893691) in locus 113 (ALOX12-AS1)

| Study | Cases | Controls | OR |
|---|---|---|---|
| INVENT-2019_EA | 15572 | 115280 | 0.95 |
| INVENT-2019_AA | 799 | 14353 | 1 |
| MVP_EA | 18952 | 411746 | 0.95 |
| MVP_AA | 5521 | 103791 | 0.96 |
| MVP_HIS | 1188 | 45634 | 0.94 |
| FinnGen_EA | 9176 | 209616 | 0.97 |
| EGP_EA | 4122 | 181553 | 1 |
| **Discovery** | **55330** | **1081973** | **0.96** |

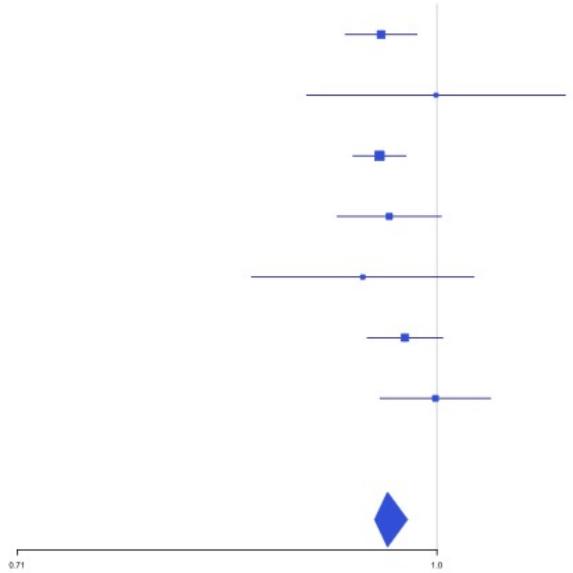

**Supplementary Figure 1.69** – rs62054822 (chr17:43927708) in locus 115 (MAPT-AS1)

| Study | Cases | Controls | OR |
|---|---|---|---|
| INVENT-2019_EA | 15572 | 115280 | 0.96 |
| INVENT-2019_AA | 799 | 14353 | 0.94 |
| MVP_EA | 18952 | 411746 | 0.95 |
| MVP_AA | 5521 | 103791 | 0.9 |
| MVP_HIS | 1188 | 45634 | 0.89 |
| FinnGen_EA | 9176 | 209616 | 0.97 |
| EGP_EA | 4122 | 181553 | 0.97 |
| **Discovery** | **55330** | **1081973** | **0.95** |

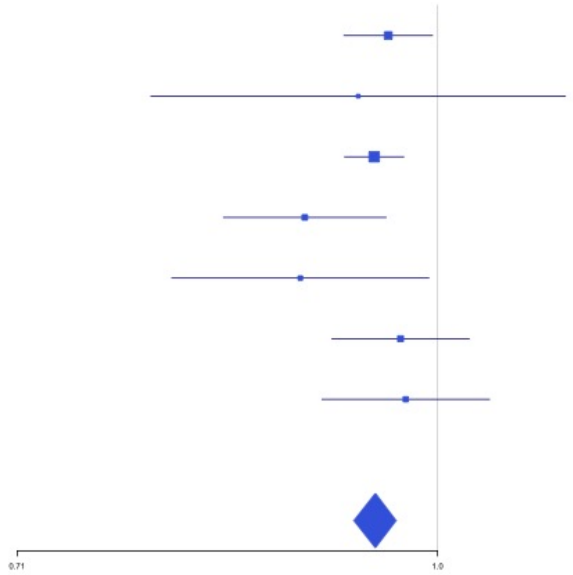


**Supplementary Figure 1.70** – rs142140545 (chr17:64191540) in locus 116 (CEP112;APOH)

| Study | Cases | Controls | OR |
|---|---|---|---|
| MVP_EA | 18952 | 411746 | 0.92 |
| MVP_AA | 5521 | 103791 | 0.94 |
| MVP_HIS | 1188 | 45634 | 0.89 |
| FinnGen_EA | 9176 | 209616 | 0.92 |
| EGP_EA | 4122 | 181553 | 0.98 |
| **Discovery** | **38959** | **952340** | **0.93** |

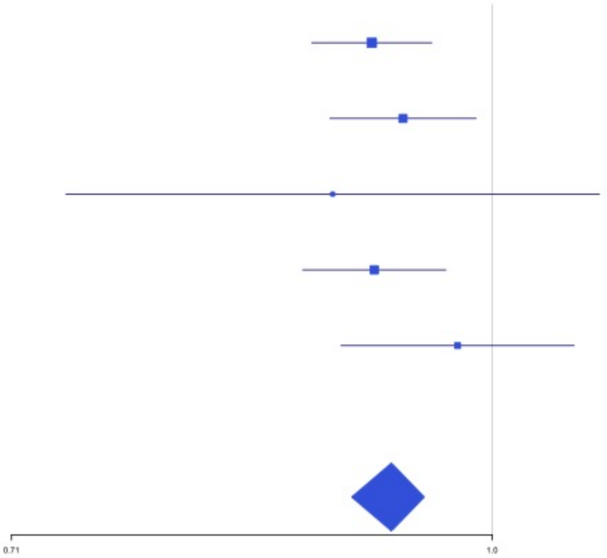

**Supplementary Figure 1.71** – rs59277920 (chr19:6077231) in locus 118 (RFX2)

| Study | Cases | Controls | OR |
|---|---|---|---|
| INVENT-2019_EA | 15572 | 115280 | 0.97 |
| INVENT-2019_AA | 799 | 14353 | 0.92 |
| MVP_EA | 18952 | 411746 | 0.92 |
| MVP_AA | 5521 | 103791 | 0.93 |
| MVP_HIS | 1188 | 45634 | 0.97 |
| FinnGen_EA | 9176 | 209616 | 0.98 |
| EGP_EA | 4122 | 181553 | 0.94 |
| **Discovery** | **55330** | **1081973** | **0.94** |

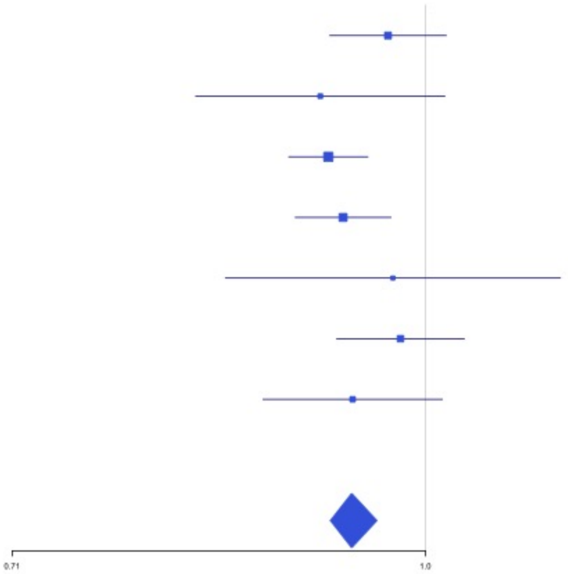


**Supplementary Figure 1.72** – rs8110055 (chr19:10739143) in locus 119 (SLC44A2)

| Study | Cases | Controls | OR |
|---|---|---|---|
| INVENT-2019_EA | 15572 | 115280 | 0.9 |
| INVENT-2019_AA | 799 | 14353 | 0.96 |
| MVP_EA | 18952 | 411746 | 0.88 |
| MVP_AA | 5521 | 103791 | 0.86 |
| MVP_HIS | 1188 | 45634 | 0.78 |
| FinnGen_EA | 9176 | 209616 | 0.87 |
| EGP_EA | 4122 | 181553 | 0.93 |
| **Discovery** | **55330** | **1081973** | **0.89** |

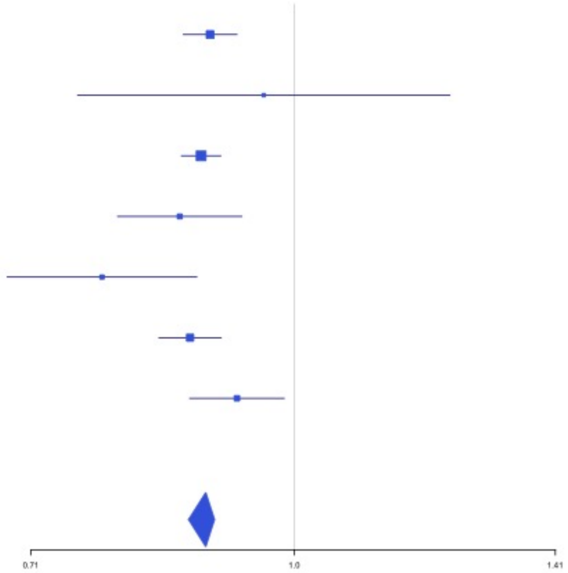

**Supplementary Figure 1.73** – rs34783010 (chr19:46180414) in locus 122 (GIPR)

| Study | Cases | Controls | OR |
|---|---|---|---|
| INVENT-2019_EA | 15572 | 115280 | 0.94 |
| INVENT-2019_AA | 799 | 14353 | 1.08 |
| MVP_EA | 18952 | 411746 | 0.94 |
| MVP_AA | 5521 | 103791 | 0.97 |
| MVP_HIS | 1188 | 45634 | 0.97 |
| FinnGen_EA | 9176 | 209616 | 0.96 |
| EGP_EA | 4122 | 181553 | 0.99 |
| **Discovery** | **55330** | **1081973** | **0.95** |

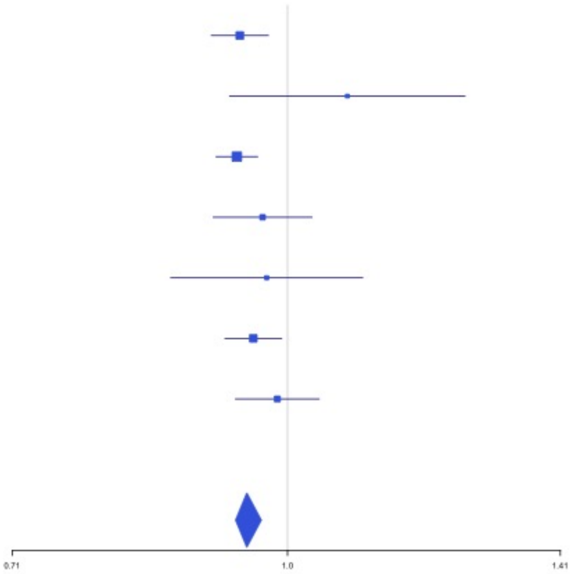


**Supplementary Figure 1.74** – rs1688264 (chr19:49209560) in locus 123 (FUT2)

| Study | Cases | Controls | OR |
|---|---|---|---|
| INVENT-2019_EA | 15572 | 115280 | 0.96 |
| INVENT-2019_AA | 799 | 14353 | 0.96 |
| MVP_EA | 18952 | 411746 | 0.96 |
| MVP_AA | 5521 | 103791 | 0.94 |
| MVP_HIS | 1188 | 45634 | 1 |
| FinnGen_EA | 9176 | 209616 | 0.97 |
| EGP_EA | 4122 | 181553 | 0.95 |
| **Discovery** | **55330** | **1081973** | **0.96** |

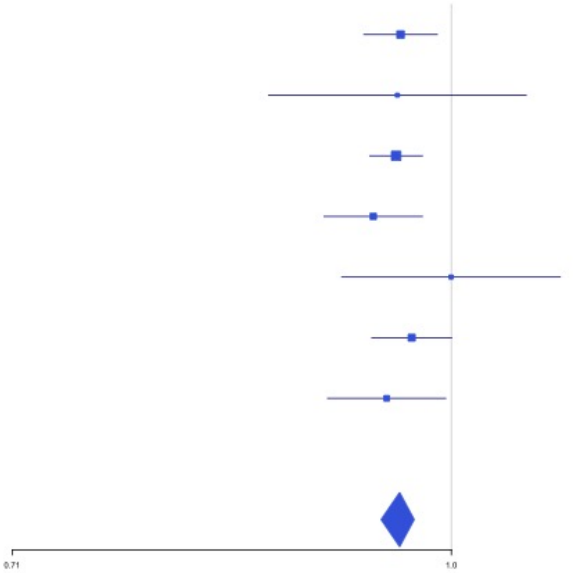

**Supplementary Figure 1.75** – rs1654425 (chr19:55538980) in locus 124 (GP6)

| Study | Cases | Controls | OR |
|---|---|---|---|
| INVENT-2019_EA | 15572 | 115280 | 0.93 |
| INVENT-2019_AA | 799 | 14353 | 0.83 |
| MVP_EA | 18952 | 411746 | 0.9 |
| MVP_AA | 5521 | 103791 | 0.95 |
| MVP_HIS | 1188 | 45634 | 0.86 |
| FinnGen_EA | 9176 | 209616 | 0.92 |
| EGP_EA | 4122 | 181553 | 0.93 |
| **Discovery** | **55330** | **1081973** | **0.91** |

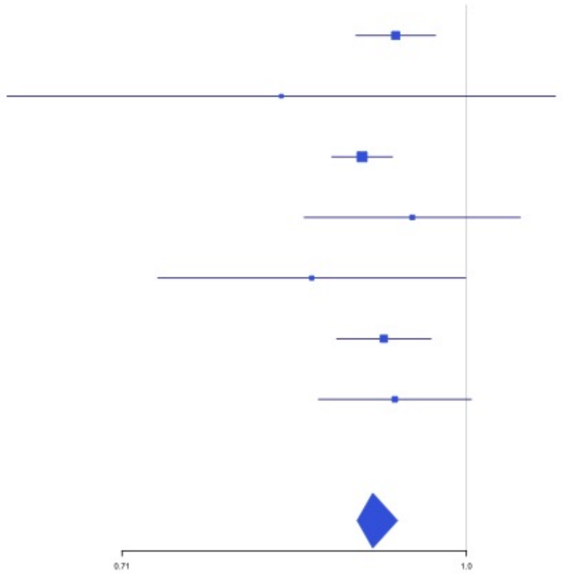


**Supplementary Figure 1.76** – rs2631628 (chr19:56645250) in locus 125 (ZNF787;ZNF444)

| Study | Cases | Controls | OR |
|---|---|---|---|
| INVENT-2019_AA | 799 | 14353 | 0.92 |
| MVP_EA | 18952 | 411746 | 0.9 |
| MVP_AA | 5521 | 103791 | 0.89 |
| MVP_HIS | 1188 | 45634 | 0.91 |
| FinnGen_EA | 9176 | 209616 | 0.9 |
| EGP_EA | 4122 | 181553 | 0.83 |
| **Discovery** | **39758** | **966693** | **0.89** |

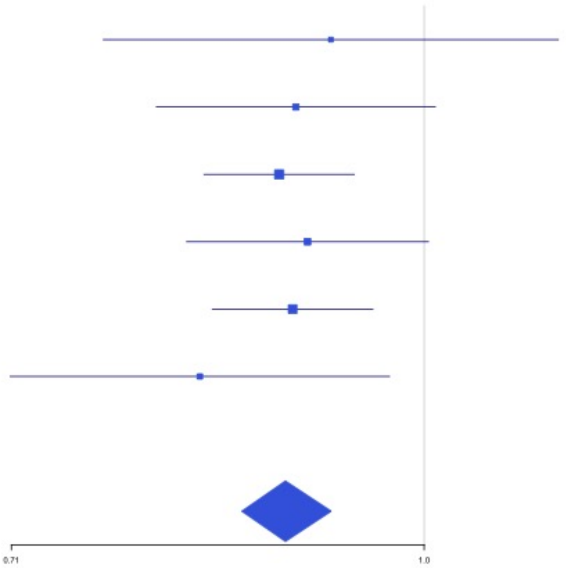

**Supplementary Figure 1.77** – rs79388863 (chr20:23168500) in locus 126 (LINC00656 ;NXT1)

| Study | Cases | Controls | OR |
|---|---|---|---|
| INVENT-2019_EA | 15572 | 115280 | 0.93 |
| INVENT-2019_AA | 799 | 14353 | 1.11 |
| MVP_EA | 18952 | 411746 | 0.9 |
| MVP_AA | 5521 | 103791 | 0.92 |
| MVP_HIS | 1188 | 45634 | 0.84 |
| FinnGen_EA | 9176 | 209616 | 0.91 |
| EGP_EA | 4122 | 181553 | 0.94 |
| **Discovery** | **55330** | **1081973** | **0.92** |

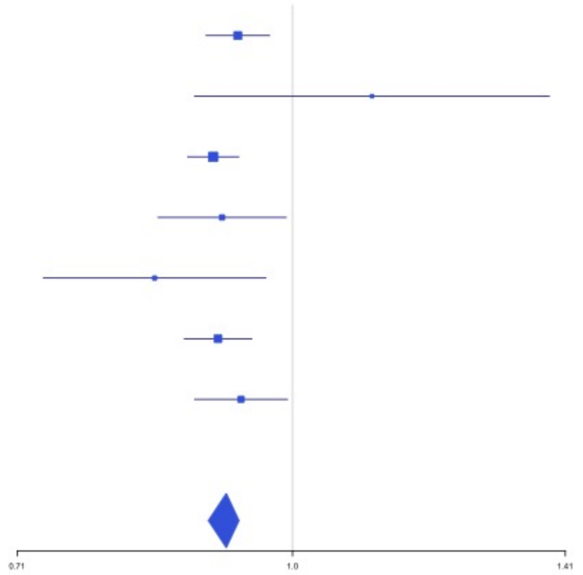


**Supplementary Figure 1.78** – rs6060288 (chr20:33772243) in locus 127 (MMP24-AS1-EDEM2)

| Study | Cases | Controls | OR |
|---|---|---|---|
| INVENT-2019_EA | 15572 | 115280 | 1.1 |
| INVENT-2019_AA | 799 | 14353 | 1.02 |
| MVP_EA | 18952 | 411746 | 1.15 |
| MVP_AA | 5521 | 103791 | 1.1 |
| MVP_HIS | 1188 | 45634 | 1.06 |
| FinnGen_EA | 9176 | 209616 | 1.12 |
| EGP_EA | 4122 | 181553 | 1.09 |
| **Discovery** | **55330** | **1081973** | **1.12** |

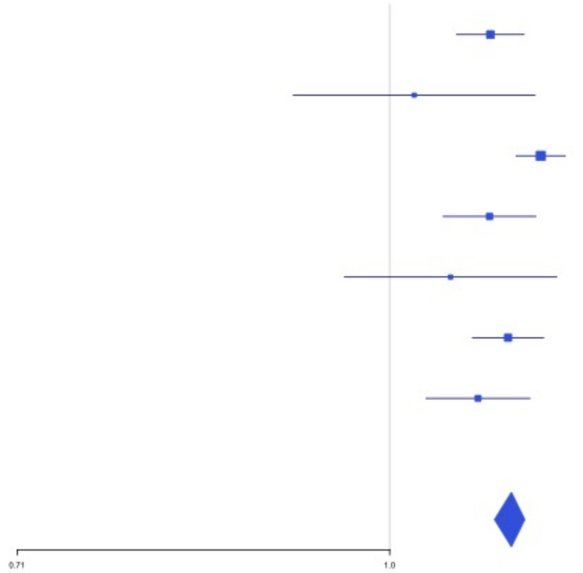

**Supplementary Figure 1.79** – rs4820093 (chr22:33160208) in locus 129 (SYN3)

| Study | Cases | Controls | OR |
|---|---|---|---|
| INVENT-2019_EA | 15572 | 115280 | 1.04 |
| INVENT-2019_AA | 799 | 14353 | 1.2 |
| MVP_EA | 18952 | 411746 | 1.05 |
| MVP_HIS | 1188 | 45634 | 1.05 |
| FinnGen_EA | 9176 | 209616 | 1.04 |
| EGP_EA | 4122 | 181553 | 1.08 |
| **Discovery** | **49809** | **978182** | **1.05** |

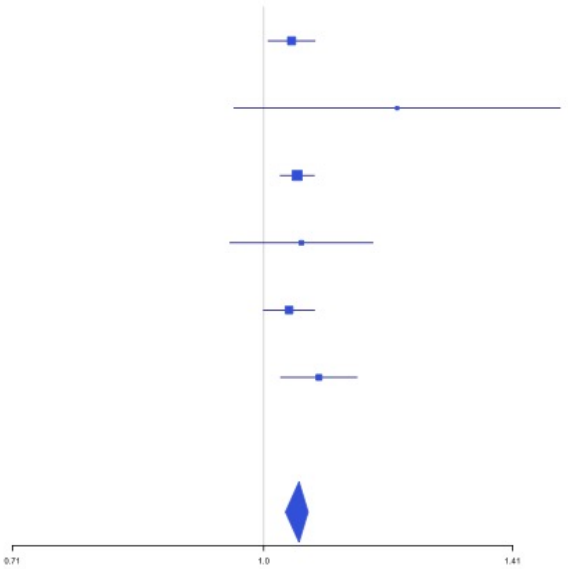


**Supplementary Figure 1.80** – rs9611844 (chr22:43115776) in locus 130 (A4GALT)

| Study | Cases | Controls | OR |
|---|---|---|---|
| INVENT-2019_EA | 15572 | 115280 | 1.04 |
| INVENT-2019_AA | 799 | 14353 | 1.08 |
| MVP_EA | 18952 | 411746 | 1.11 |
| MVP_AA | 5521 | 103791 | 1.15 |
| MVP_HIS | 1188 | 45634 | 1.11 |
| FinnGen_EA | 9176 | 209616 | 1.08 |
| EGP_EA | 4122 | 181553 | 1.17 |
| **Discovery** | **55330** | **1081973** | **1.1** |

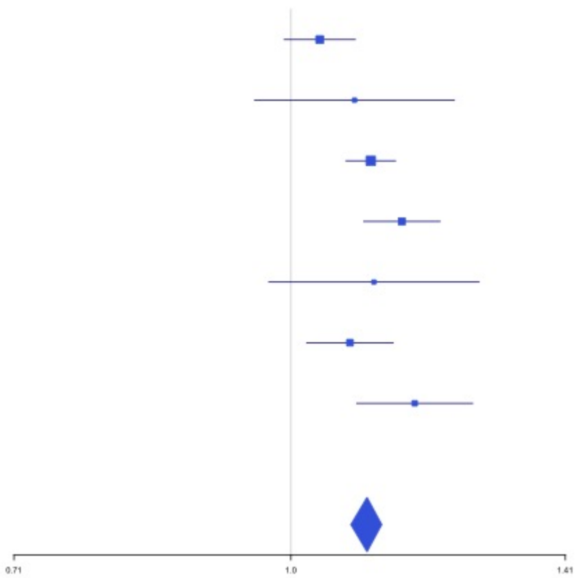

**Supplementary Figure 1.81** – rs34609587 (chr23:11657363) in locus 131 (ARHGAP6)

| Study | Cases | Controls | OR |
|---|---|---|---|
| INVENT-2019_EA | 8705 | 52578 | 0.97 |
| MVP_EA | 18952 | 411746 | 0.93 |
| MVP_AA | 5521 | 103791 | 0.97 |
| MVP_HIS | 1188 | 45634 | 0.98 |
| FinnGen_EA | 9176 | 209616 | 0.95 |
| **Discovery** | **43542** | **823365** | **0.95** |

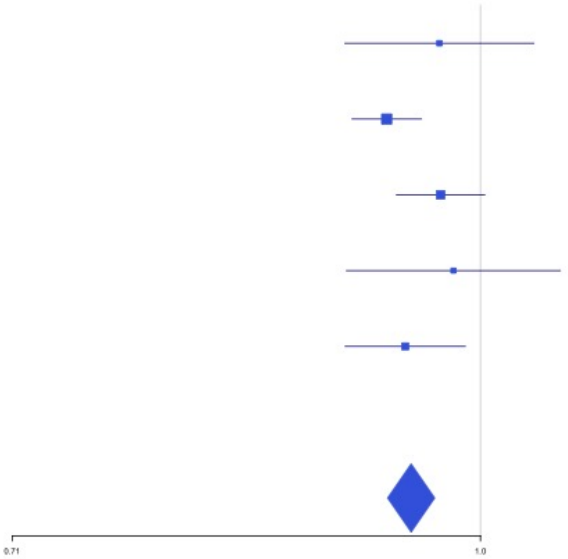


**Supplementary Figure 1.82** – rs6632109 (chr23:34894638) in locus 132 (TMEM47;FAM47B)

| Study | Cases | Controls | OR |
|---|---|---|---|
| INVENT-2019_EA | 8705 | 52578 | 1.04 |
| MVP_EA | 18952 | 411746 | 1.04 |
| MVP_AA | 5521 | 103791 | 1.09 |
| MVP_HIS | 1188 | 45634 | 1.01 |
| FinnGen_EA | 9176 | 209616 | 1.04 |
| EGP_EA | 4122 | 181553 | 1.05 |
| **Discovery** | **47664** | **1004918** | **1.04** |

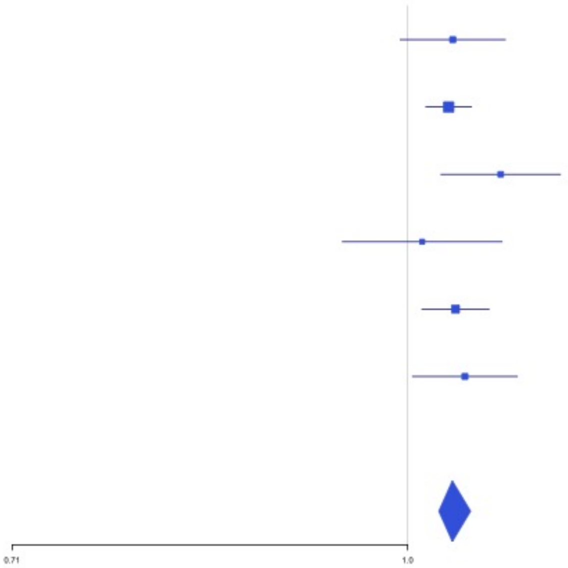

**Supplementary Figure 1.83** – rs3002416 (chr23:39710195) in locus 133 (MIR1587;BCOR)

| Study | Cases | Controls | OR |
|---|---|---|---|
| INVENT-2019_EA | 8705 | 52578 | 0.91 |
| MVP_EA | 18952 | 411746 | 0.94 |
| MVP_AA | 5521 | 103791 | 0.91 |
| MVP_HIS | 1188 | 45634 | 1 |
| FinnGen_EA | 9176 | 209616 | 0.96 |
| EGP_EA | 4122 | 181553 | 0.98 |
| **Discovery** | **47664** | **1004918** | **0.95** |

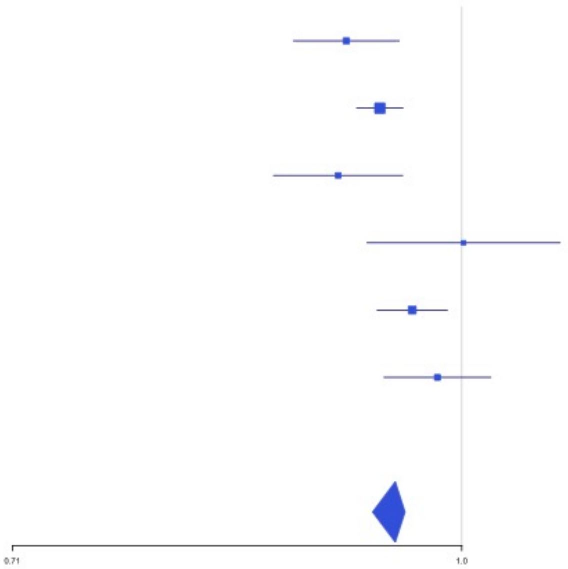


**Supplementary Figure 1.84** – rs6048 (chr23:138633280) in locus 134 (F9)

| Study | Cases | Controls | OR |
|---|---|---|---|
| INVENT-2019_EA | 8705 | 52578 | 1.09 |
| MVP_EA | 18952 | 411746 | 1.07 |
| MVP_AA | 5521 | 103791 | 1.1 |
| MVP_HIS | 1188 | 45634 | 1.04 |
| FinnGen_EA | 9176 | 209616 | 1.05 |
| EGP_EA | 4122 | 181553 | 1.07 |
| **Discovery** | **47664** | **1004918** | **1.07** |

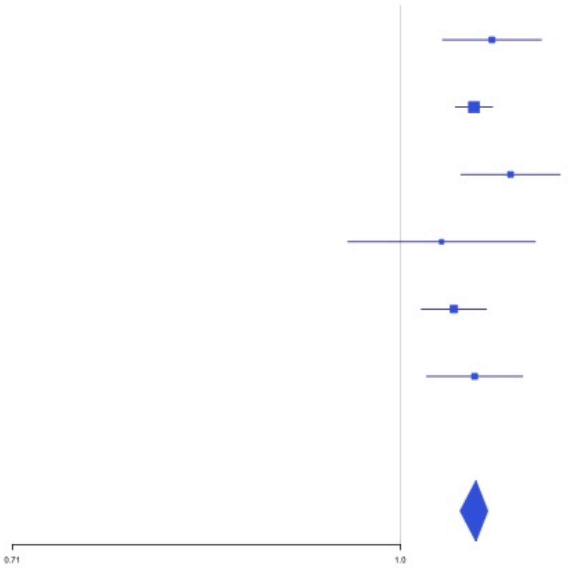

**Supplementary Figure 1.85** – rs2084408 (chr23:154346709) in locus 135 (BRCC3)

| Study | Cases | Controls | OR |
|---|---|---|---|
| INVENT-2019_EA | 8705 | 52578 | 0.94 |
| MVP_EA | 18952 | 411746 | 0.92 |
| MVP_AA | 5521 | 103791 | 0.95 |
| MVP_HIS | 1188 | 45634 | 0.93 |
| FinnGen_EA | 9176 | 209616 | 0.96 |
| EGP_EA | 4122 | 181553 | 0.94 |
| **Discovery** | **47664** | **1004918** | **0.94** |

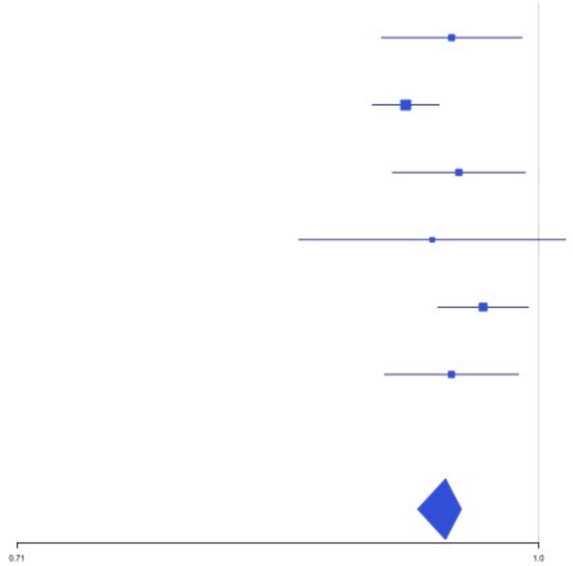
