## Supplemental Figure 2 for "Cross-Ancestry Investigation of Venous Thromboembolism Genomic Predictors"

### Supplementary Figure 2.
### Forest plots of Combined analysis lead variants.

2.1 rs12121476 (chr1:9335327) in locus 1 (H6PD ;SPSB1) . . . . 4
2.2 rs551176418 (chr1:27107263) in locus 2 (ARID1A) . . . . 4
2.3 rs6695572 (chr1:77945635) in locus 3 (AK5) . . . . 5
2.4 rs3832016 (chr1:109818158) in locus 4 (CELSR2) . . . . 5
2.5 rs3767812 (chr1:118155620) in locus 5 (TENT5C) . . . . 6
2.6 rs1267881263 (chr1:150496127) in locus 6 (FALEC ;ADAMTSL4) . . . . 6
2.7 rs905938 (chr1:154991389) in locus 7 (DCST2) . . . . 7
2.8 rs3557 (chr1:161188893) in locus 8 (FCER1G) . . . . 7
2.9 rs6025 (chr1:169519049) in locus 9 (F5) . . . . 8
2.10 rs6695940 (chr1:173882980) in locus 10 (SERPINC1) . . . . 8
2.11 rs143410348 (chr1:196809316) in locus 12 (CFHR1 ;CFHR4) . . . . 9
2.12 rs2842700 (chr1:207282149) in locus 13 (C4BPA) . . . . 9
2.13 rs3811444 (chr1:248039451) in locus 14 (TRIM58) . . . . 10
2.14 rs78475244 (chr2:65086804) in locus 16 (LINC01800) . . . . 10
2.15 rs1867312 (chr2:68619981) in locus 17 (PLEK) . . . . 11
2.16 rs1518760 (chr2:128140500) in locus 18 (MAP3K2) . . . . 11
2.17 rs1912849 (chr2:188310118) in locus 19 (CALCRL) . . . . 12
2.18 rs78872368 (chr2:198545250) in locus 20 (RFTN2 ;MARS2) . . . . 12
2.19 rs715 (chr2:211543055) in locus 21 (CPS1) . . . . 13
2.20 rs68066031 (chr2:224880498) in locus 22 (SERPINE2) . . . . 13
2.21 rs13084580 (chr3:39188182) in locus 24 (CSRNP1) . . . . 14
2.22 rs11130326 (chr3:52771920) in locus 25 (NEK4) . . . . 14
2.23 rs529326506 (chr3:90503764) in locus 26 (EPHA3 ;NONE) . . . . 15
2.24 rs28479320 (chr3:93752529) in locus 27 (ARL13B) . . . . 15
2.25 rs900399 (chr3:156798732) in locus 30 (LINC02029 ;LINC00880) . . . . 16
2.26 rs710446 (chr3:186459927) in locus 32 (KNG1) . . . . 16
2.27 rs3796159 (chr3:194790434) in locus 33 (XXYLT1) . . . . 17
2.28 rs9654093 (chr4:7903763) in locus 34 (AFAP1) . . . . 17
2.29 rs781656 (chr4:57778645) in locus 35 (REST) . . . . 18
2.30 rs6826579 (chr4:83785031) in locus 36 (SEC31A) . . . . 18
2.31 rs2014912 (chr4:86715670) in locus 37 (ARHGAP24) . . . . 19
2.32 rs2066864 (chr4:155525695) in locus 39 (FGG) . . . . 19
2.33 rs3756011 (chr4:187206249) in locus 40 (F11) . . . . 20
2.34 rs16867574 (chr5:38708554) in locus 42 (OSMR-AS1) . . . . 20
2.35 rs7730244 (chr5:72957088) in locus 43 (ARHGEF28) . . . . 21
2.36 rs750465397 (chr5:96289036) in locus 44 (LNPEP) . . . . 21
2.37 rs147133967 (chr5:132426851) in locus 45 (HSPA4) . . . . 22
2.38 rs214059 (chr6:25536937) in locus 46 (CARMIL1) . . . . 22
2.39 rs2394251 (chr6:29943688) in locus 47 (HCG9) . . . . 23
2.40 rs9268863 (chr6:32430289) in locus 48 (HLA-DRA ;HLA-DRB5) . . . . 23
2.41 rs145294670 (chr6:34622561) in locus 49 (ILRUN) . . . . 24
2.42 rs9386182 (chr6:147691069) in locus 52 (STXBP5) . . . . 24
2.43 rs1513275 (chr7:28259233) in locus 53 (JAZF1-AS1) . . . . 25
2.44 rs42038 (chr7:92243719) in locus 54 (CDK6) . . . . 25
2.45 rs10099512 (chr8:9178821) in locus 57 (LOC101929128 ;LOC157273) . . . . 26
2.46 rs2048528 (chr8:23373680) in locus 59 (ENTPD4 ;SLC25A37) . . . . 26
2.47 rs2685412 (chr8:27810603) in locus 60 (SCARA5) . . . . 27
2.48 rs2915595 (chr8:30402817) in locus 61 (RBPMS) . . . . 27
2.49 rs6993770 (chr8:106581528) in locus 62 (ZFPM2) . . . . 28
2.50 rs4236786 (chr8:108291878) in locus 63 (ANGPT1) . . . . 28

2.51 rs10820606 (chr9:99192919) in locus 65 (ZNF367;HABP4) . . . 29
2.52 rs505922 (chr9:136149229) in locus 66 (ABO) . . . 29
2.53 rs1243187 (chr10:21907016) in locus 68 (MLLT10) . . . 30
2.54 rs4272700 (chr10:27881308) in locus 69 (RAB18;MKX) . . . 30
2.55 rs17490626 (chr10:71218646) in locus 72 (TSPAN15) . . . 31
2.56 rs16937003 (chr10:80938499) in locus 73 (ZMIZ1) . . . 31
2.57 rs57866767 (chr10:96023077) in locus 74 (PLCE1) . . . 32
2.58 rs10886430 (chr10:121010256) in locus 75 (GRK5) . . . 32
2.59 rs2030291 (chr11:16251251) in locus 77 (SOX6) . . . 33
2.60 rs11032074 (chr11:32993887) in locus 78 (QSER1) . . . 33
2.61 rs1799963 (chr11:46761055) in locus 79 (F2) . . . 34
2.62 rs141798115 (chr11:56875074) in locus 80 (OR5AK4P;LRRC55) . . . 34
2.63 rs4354705 (chr11:60088159) in locus 81 (MS4A4A;MS4A6E) . . . 35
2.64 rs174551 (chr11:61573684) in locus 82 (FADS1) . . . 35
2.65 rs2846027 (chr11:114003415) in locus 84 (ZBTB16) . . . 36
2.66 rs35257264 (chr11:126296816) in locus 85 (ST3GAL4) . . . 36
2.67 rs7107568 (chr11:130779668) in locus 86 (SNX19) . . . 37
2.68 rs1558519 (chr12:6153738) in locus 87 (VWF) . . . 37
2.69 rs7311483 (chr12:9053661) in locus 88 (A2ML1;PHC1) . . . 38
2.70 rs6580981 (chr12:54723028) in locus 89 (COPZ1) . . . 38
2.71 rs142351376 (chr12:104136288) in locus 90 (STAB2) . . . 39
2.72 rs3184504 (chr12:111884608) in locus 91 (SH2B3) . . . 39
2.73 rs2851435 (chr12:123712416) in locus 92 (MPHOSPH9) . . . 40
2.74 rs3211752 (chr13:113787459) in locus 94 (F10) . . . 40
2.75 rs11158204 (chr14:58844526) in locus 95 (ARID4A;TOMM20L) . . . 41
2.76 rs2127869 (chr14:65794352) in locus 96 (LINC02324;MIR4708) . . . 41
2.77 rs7152641 (chr14:92213967) in locus 97 (CATSPERB;TC2N) . . . 42
2.78 rs28929474 (chr14:94844947) in locus 98 (SERPINA1) . . . 42
2.79 rs28519971 (chr14:103089808) in locus 99 (RCOR1) . . . 43
2.80 rs1247656151 (chr15:43677975) in locus 100 (TUBGCP4) . . . 43
2.81 rs7183672 (chr15:96101018) in locus 103 (LINC00924;LOC105369212) . . . 44
2.82 rs71376077 (chr16:15738114) in locus 105 (NDE1) . . . 44
2.83 rs12443808 (chr16:30996871) in locus 106 (HSD3B7) . . . 45
2.84 rs7197453 (chr16:72079127) in locus 107 (DHODH;HP) . . . 45
2.85 rs77246010 (chr16:75429853) in locus 108 (CFDP1) . . . 46
2.86 rs12445050 (chr16:81870969) in locus 109 (PLCG2) . . . 46
2.87 rs8049403 (chr16:85778651) in locus 110 (C16orf74) . . . 47
2.88 rs28634651 (chr16:88553198) in locus 111 (ZFPM1) . . . 47
2.89 rs6503222 (chr17:1977862) in locus 112 (SMG6) . . . 48
2.90 rs434473 (chr17:6904934) in locus 113 (ALOX12) . . . 48
2.91 rs71138827 (chr17:27833678) in locus 114 (TAOK1) . . . 49
2.92 rs62054822 (chr17:43927708) in locus 115 (MAPT-AS1) . . . 49
2.93 rs1801690 (chr17:64208285) in locus 116 (APOH) . . . 50
2.94 rs573431210 (chr17:67276383) in locus 117 (ABCA5) . . . 50
2.95 rs59277920 (chr19:6077231) in locus 118 (RFX2) . . . 51
2.96 rs8110055 (chr19:10739143) in locus 119 (SLC44A2) . . . 51
2.97 rs889139 (chr19:33889369) in locus 120 (PEPD) . . . 52
2.98 rs2545774 (chr19:41287674) in locus 121 (RAB4B) . . . 52
2.99 rs8108474 (chr19:46301479) in locus 122 (RSPH6A) . . . 53
2.100 rs2638282 (chr19:49213833) in locus 123 (FUT2;MAMSTR) . . . 53
2.101 rs1654425 (chr19:55538980) in locus 124 (GP6) . . . 54
2.102 rs2631628 (chr19:56645250) in locus 125 (ZNF787;ZNF444) . . . 54
2.103 rs117390891 (chr20:23168526) in locus 126 (LINC00656;NXT1) . . . 55

2.104rs6060288 (chr20:33772243) in locus 127 (MMP24-AS1-EDEM2) . . . . . . . . . . . . . 55
2.105rs3070580 (chr22:33165231) in locus 129 (SYN3) . . . . . . . . . . . . . . . . . . . . . . . 56
2.106rs9611844 (chr22:43115776) in locus 130 (A4GALT) . . . . . . . . . . . . . . . . . . . . . 56
2.107rs12388118 (chr23:11380742) in locus 131 (ARHGAP6) . . . . . . . . . . . . . . . . . . . 57
2.108rs6632109 (chr23:34894638) in locus 132 (TMEM47;FAM47B) . . . . . . . . . . . . . . . 57
2.109rs3002416 (chr23:39710195) in locus 133 (MIR1587;BCOR) . . . . . . . . . . . . . . . . 58
2.110rs6048 (chr23:138633280) in locus 134 (F9) . . . . . . . . . . . . . . . . . . . . . . . . . . 58
2.111rs2084408 (chr23:154346709) in locus 135 (BRCC3) . . . . . . . . . . . . . . . . . . . . . 59

**Supplementary Figure 2.1** – rs12121476 (chr1:9335327) in locus 1 (H6PD;SPSB1)

| Study | Cases | Controls | OR |
|---|---|---|---|
| INVENT-2019_EA | 15572 | 115280 | 1.09 |
| INVENT-2019_AA | 799 | 14353 | 0.99 |
| MVP_EA | 18952 | 411746 | 1.04 |
| MVP_AA | 5521 | 103791 | 1.05 |
| MVP_HIS | 1188 | 45634 | 1.13 |
| FinnGen_EA | 9176 | 209616 | 1.01 |
| EGP_EA | 4122 | 181553 | 1.03 |
| UKB_EA | 18385 | 91834 | 1.04 |
| MGB_AA | 225 | 1481 | 0.94 |
| MGB_EA | 2967 | 27957 | 1.02 |
| MGB_HIS | 100 | 1352 | 0.8 |
| UPenn_EA | 380 | 6210 | 0.99 |
| UPenn_AA | 203 | 1719 | 0.91 |
| FARIVE_EA | 545 | 532 | 1.11 |
| BioME_EA | 200 | 8007 | 0.92 |
| BioME_AA | 382 | 6074 | 1.1 |
| BioME_HIS | 397 | 8992 | 1.09 |
| RETROVE_EA | 400 | 400 | 1.07 |
| GAIT2_EA | 85 | 850 | 0.8 |
| BBJ_EAS | 507 | 178366 | 1.02 |
| MESA_EA | 94 | 2341 | 0.84 |
| MESA_AA | 72 | 1511 | 1.01 |
| MESA_HIS | 35 | 1389 | 1.57 |
| **Overall** | **80307** | **1420988** | **1.04** |

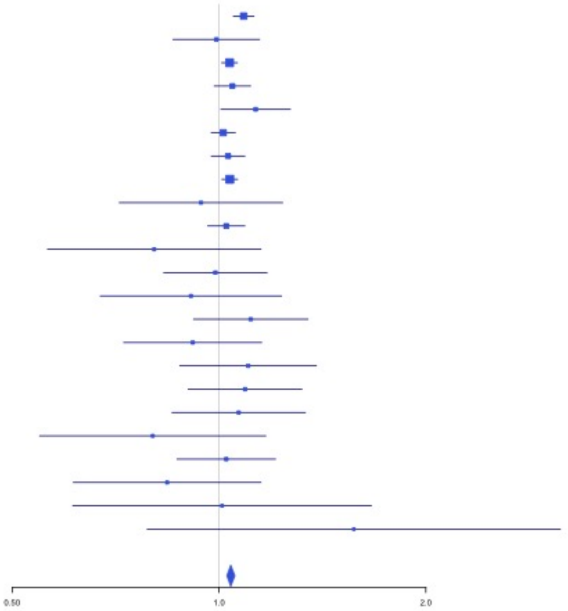


**Supplementary Figure 2.2** – rs551176418 (chr1:27107263) in locus 2 (ARID1A)

| Study | Cases | Controls | OR |
|---|---|---|---|
| MVP_EA | 18952 | 411746 | 1.08 |
| MVP_AA | 5521 | 103791 | 1.08 |
| MVP_HIS | 1188 | 45634 | 1.11 |
| FinnGen_EA | 9176 | 209616 | 1.1 |
| UKB_EA | 18385 | 91834 | 1.07 |
| UPenn_EA | 380 | 6210 | 0.95 |
| UPenn_AA | 203 | 1719 | 0.58 |
| FARIVE_EA | 545 | 532 | 1.14 |
| BioME_AA | 382 | 6074 | 1.69 |
| BioME_HIS | 397 | 8992 | 0.92 |
| BBJ_EAS | 507 | 178366 | 0.51 |
| MARTHA12_EA | 893 | 3414 | 0.96 |
| **Overall** | **56529** | **1067928** | **1.08** |

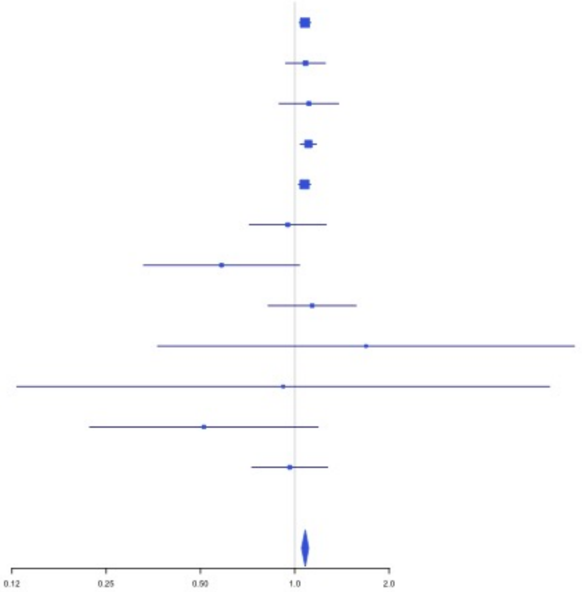

**Supplementary Figure 2.3** – rs6695572 (chr1:77945635) in locus 3 (AK5)

| Study | Cases | Controls | OR |
|---|---|---|---|
| INVENT-2019_EA | 15572 | 115280 | 1.05 |
| INVENT-2019_AA | 799 | 14353 | 1.06 |
| MVP_EA | 18952 | 411746 | 1.03 |
| MVP_AA | 5521 | 103791 | 1.06 |
| MVP_HIS | 1188 | 45634 | 1.02 |
| FinnGen_EA | 9176 | 209616 | 1.05 |
| EGP_EA | 4122 | 181553 | 1.03 |
| UKB_EA | 18385 | 91834 | 1.05 |
| MGB_AA | 225 | 1481 | 1.14 |
| MGB_EA | 2967 | 27957 | 0.98 |
| MGB_HIS | 100 | 1352 | 1.2 |
| UPenn_EA | 380 | 6210 | 1.12 |
| UPenn_AA | 203 | 1719 | 1.1 |
| FARIVE_EA | 545 | 532 | 0.96 |
| BioME_EA | 200 | 8007 | 1.08 |
| BioME_AA | 382 | 6074 | 1 |
| BioME_HIS | 397 | 8992 | 1.12 |
| RETROVE_EA | 400 | 400 | 0.87 |
| GAIT2_EA | 85 | 850 | 0.97 |
| BBJ_EAS | 507 | 178366 | 1.12 |
| MESA_EA | 94 | 2341 | 1.11 |
| MESA_AA | 72 | 1511 | 1.1 |
| MESA_HIS | 35 | 1389 | 1.07 |
| **Overall** | **80307** | **1420988** | **1.04** |

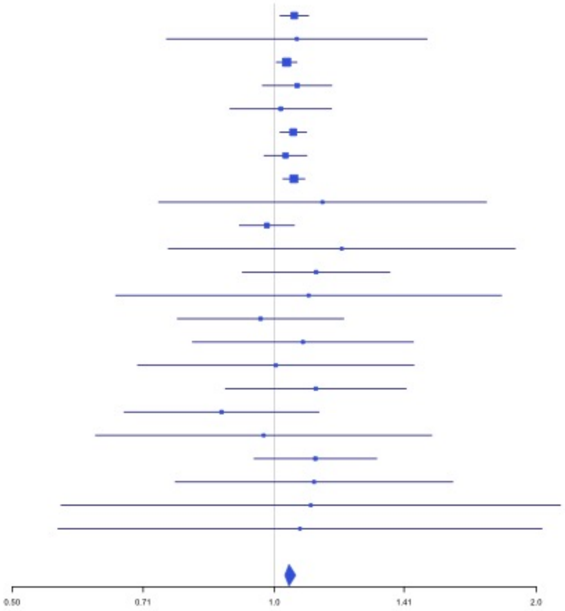


**Supplementary Figure 2.4** – rs3832016 (chr1:109818158) in locus 4 (CELSR2)

| Study | Cases | Controls | OR |
|---|---|---|---|
| INVENT-2019_EA | 15572 | 115280 | 0.98 |
| INVENT-2019_AA | 799 | 14353 | 0.99 |
| MVP_EA | 18952 | 411746 | 0.96 |
| MVP_AA | 5521 | 103791 | 0.95 |
| MVP_HIS | 1188 | 45634 | 0.93 |
| FinnGen_EA | 9176 | 209616 | 0.95 |
| EGP_EA | 4122 | 181553 | 0.99 |
| UKB_EA | 18385 | 91834 | 0.94 |
| UKB_AA | 280 | 1046 | 1.04 |
| UKB_SAS | 189 | 1269 | 0.86 |
| MGB_AA | 225 | 1481 | 0.84 |
| MGB_EA | 2967 | 27957 | 0.97 |
| MGB_HIS | 100 | 1352 | 0.76 |
| UPenn_EA | 380 | 6210 | 1 |
| UPenn_AA | 203 | 1719 | 0.78 |
| FARIVE_EA | 545 | 532 | 1.05 |
| MARTHA12_EA | 893 | 3414 | 0.99 |
| **Overall** | **79497** | **1218787** | **0.96** |

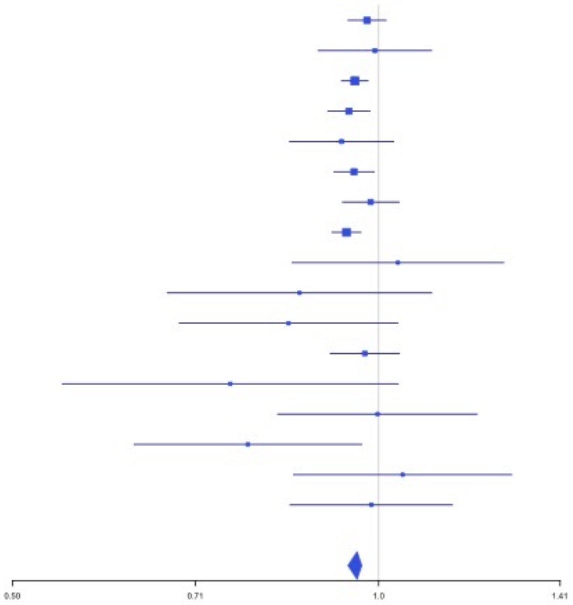

**Supplementary Figure 2.5** – rs3767812 (chr1:118155620) in locus 5 (TENT5C)

| Study | Cases | Controls | OR |
|---|---|---|---|
| INVENT-2019_EA | 15572 | 115280 | 1.04 |
| INVENT-2019_AA | 799 | 14353 | 1.14 |
| MVP_EA | 18952 | 411746 | 1.06 |
| MVP_AA | 5521 | 103791 | 1.08 |
| MVP_HIS | 1188 | 45634 | 1.05 |
| FinnGen_EA | 9176 | 209616 | 1.04 |
| EGP_EA | 4122 | 181553 | 1 |
| UKB_EA | 18385 | 91834 | 1.04 |
| UKB_AA | 280 | 1046 | 1.07 |
| UKB_SAS | 189 | 1269 | 0.9 |
| MGB_AA | 225 | 1481 | 1.17 |
| MGB_EA | 2967 | 27957 | 1.01 |
| MGB_HIS | 100 | 1352 | 1.29 |
| UPenn_EA | 380 | 6210 | 1.09 |
| UPenn_AA | 203 | 1719 | 1 |
| FARIVE_EA | 545 | 532 | 1.26 |
| BioME_EA | 200 | 8007 | 0.95 |
| BioME_AA | 382 | 6074 | 0.98 |
| BioME_HIS | 397 | 8992 | 0.91 |
| RETROVE_EA | 400 | 400 | 1.24 |
| GAIT2_EA | 85 | 850 | 0.93 |
| BBJ_EAS | 507 | 178366 | 1.09 |
| MESA_EA | 94 | 2341 | 0.73 |
| MESA_AA | 72 | 1511 | 0.82 |
| MESA_HIS | 35 | 1389 | 1.07 |
| **Overall** | **80776** | **1423303** | **1.05** |

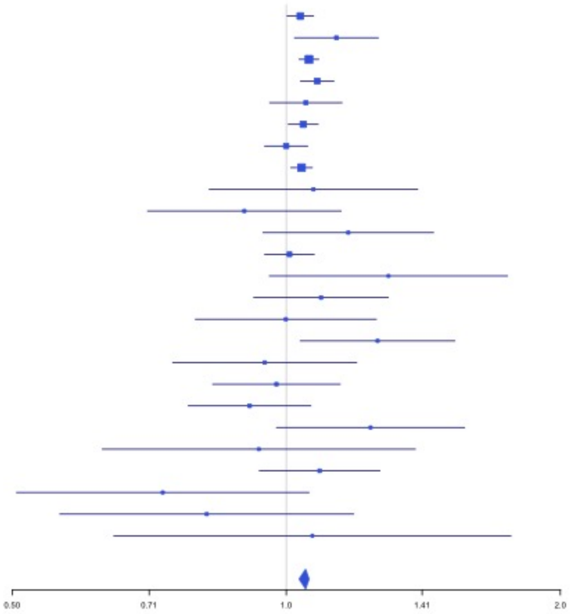


**Supplementary Figure 2.6** – rs1267881263 (chr1:150496127) in locus 6 (FALEC;ADAMTSL4)

| Study | Cases | Controls | OR |
|---|---|---|---|
| MVP_EA | 18952 | 411746 | 1.06 |
| MVP_AA | 5521 | 103791 | 1.02 |
| MVP_HIS | 1188 | 45634 | 1.04 |
| UKB_EA | 18385 | 91834 | 1.04 |
| UKB_AA | 280 | 1046 | 0.85 |
| UKB_SAS | 189 | 1269 | 0.95 |
| UPenn_EA | 380 | 6210 | 1.01 |
| UPenn_AA | 203 | 1719 | 0.99 |
| FARIVE_EA | 545 | 532 | 0.98 |
| MARTHA12_EA | 893 | 3414 | 1.21 |
| **Overall** | **46536** | **667195** | **1.04** |

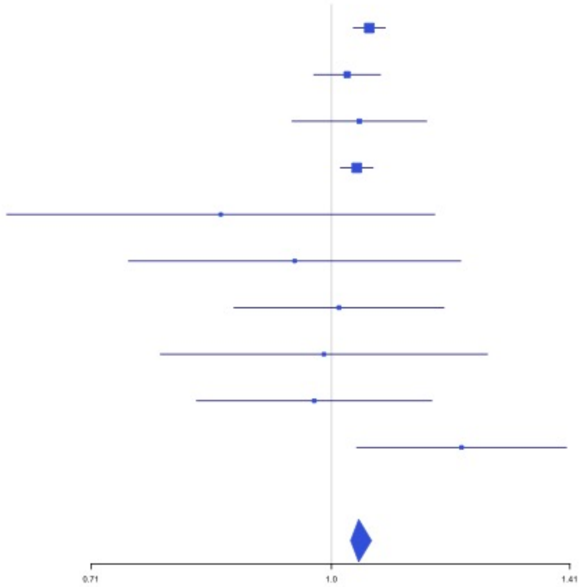

**Supplementary Figure 2.7** – rs905938 (chr1:154991389) in locus 7 (DCST2)

| Study | Cases | Controls | OR |
|---|---|---|---|
| INVENT-2019_EA | 15572 | 115280 | 0.98 |
| INVENT-2019_AA | 799 | 14353 | 0.98 |
| MVP_EA | 18952 | 411746 | 0.97 |
| MVP_AA | 5521 | 103791 | 1 |
| MVP_HIS | 1188 | 45634 | 1.04 |
| FinnGen_EA | 9176 | 209616 | 0.95 |
| EGP_EA | 4122 | 181553 | 0.94 |
| UKB_EA | 18385 | 91834 | 0.96 |
| UKB_AA | 280 | 1046 | 1.05 |
| MGB_AA | 225 | 1481 | 0.89 |
| MGB_EA | 2967 | 27957 | 0.94 |
| MGB_HIS | 100 | 1352 | 0.68 |
| UPenn_EA | 380 | 6210 | 1.14 |
| UPenn_AA | 203 | 1719 | 0.96 |
| FARIVE_EA | 545 | 532 | 0.89 |
| BioME_EA | 200 | 8007 | 1.09 |
| BioME_AA | 382 | 6074 | 1.03 |
| BioME_HIS | 397 | 8992 | 0.98 |
| RETROVE_EA | 400 | 400 | 0.74 |
| GAIT2_EA | 85 | 850 | 0.61 |
| BBJ_EAS | 507 | 178366 | 0.99 |
| MESA_EA | 94 | 2341 | 0.79 |
| MESA_AA | 72 | 1511 | 0.81 |
| MESA_HIS | 35 | 1389 | 1.22 |
| MARTHA12_EA | 893 | 3414 | 1.1 |
| **Overall** | **81480** | **1425448** | **0.97** |

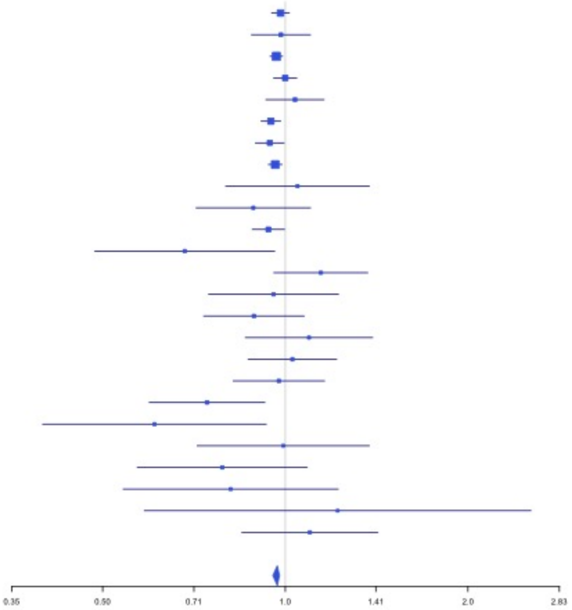


**Supplementary Figure 2.8** – rs3557 (chr1:161188893) in locus 8 (FCER1G)

| Study | Cases | Controls | OR |
|---|---|---|---|
| INVENT-2019_EA | 15572 | 115280 | 1.05 |
| INVENT-2019_AA | 799 | 14353 | 0.96 |
| MVP_EA | 18952 | 411746 | 1.09 |
| MVP_AA | 5521 | 103791 | 1.21 |
| FinnGen_EA | 9176 | 209616 | 1.04 |
| EGP_EA | 4122 | 181553 | 1.01 |
| UKB_EA | 18385 | 91834 | 1.05 |
| MGB_AA | 225 | 1481 | 0.99 |
| MGB_EA | 2967 | 27957 | 1.01 |
| MGB_HIS | 100 | 1352 | 0.87 |
| UPenn_EA | 380 | 6210 | 1.12 |
| UPenn_AA | 203 | 1719 | 1.26 |
| FARIVE_EA | 545 | 532 | 1.26 |
| BioME_EA | 200 | 8007 | 1.14 |
| BioME_AA | 382 | 6074 | 0.87 |
| BioME_HIS | 397 | 8992 | 1.08 |
| RETROVE_EA | 400 | 400 | 1.42 |
| GAIT2_EA | 85 | 850 | 0.6 |
| BBJ_EAS | 507 | 178366 | 1.78 |
| MESA_EA | 94 | 2341 | 1.17 |
| MESA_AA | 72 | 1511 | 1.22 |
| MESA_HIS | 35 | 1389 | 3.34 |
| MARTHA12_EA | 893 | 3414 | 1.37 |
| **Overall** | **80012** | **1378768** | **1.07** |

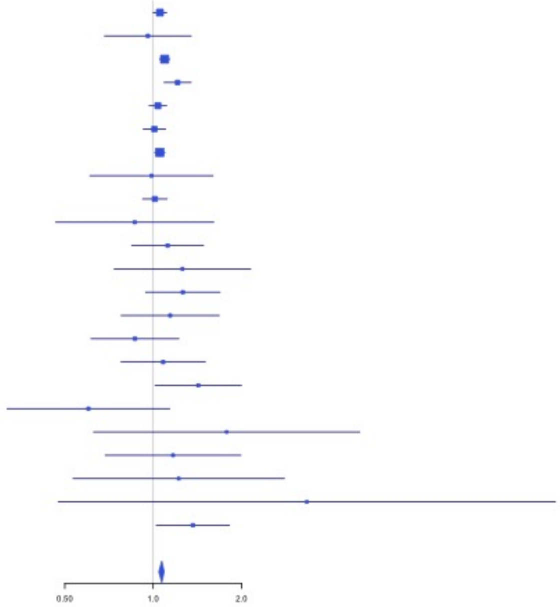

**Supplementary Figure 2.9** – rs6025 (chr1:169519049) in locus 9 (F5)

| Study | Cases | Controls | OR |
|---|---|---|---|
| INVENT-2019_EA | 15572 | 115280 | 2.39 |
| MVP_EA | 18952 | 411746 | 3.12 |
| MVP_AA | 5521 | 103791 | 2.61 |
| FinnGen_EA | 9176 | 209616 | 3.24 |
| EGP_EA | 4122 | 181553 | 3.07 |
| UKB_EA | 18385 | 91834 | 3.14 |
| MGB_EA | 2967 | 27957 | 1.98 |
| UPenn_EA | 380 | 6210 | 2.09 |
| UPenn_AA | 203 | 1719 | 3.96 |
| FARIVE_EA | 545 | 532 | 2.45 |
| BioME_EA | 200 | 8007 | 2.72 |
| BioME_AA | 382 | 6074 | 4.27 |
| BioME_HIS | 397 | 8992 | 1.78 |
| RETROVE_EA | 400 | 400 | 2.27 |
| MESA_EA | 94 | 2341 | 1.85 |
| **Overall** | **77296** | **1176052** | **2.96** |

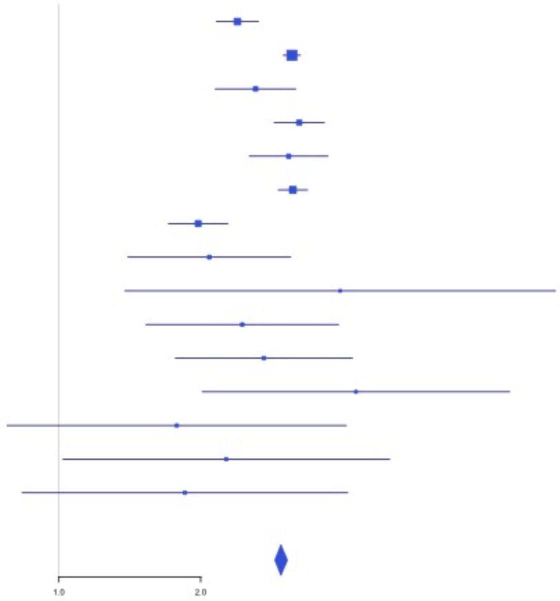


**Supplementary Figure 2.10** – rs6695940 (chr1:173882980) in locus 10 (SERPINC1)

| Study | Cases | Controls | OR |
|---|---|---|---|
| INVENT-2019_EA | 15572 | 115280 | 0.95 |
| INVENT-2019_AA | 799 | 14353 | 1 |
| MVP_EA | 18952 | 411746 | 0.97 |
| MVP_AA | 5521 | 103791 | 0.93 |
| MVP_HIS | 1188 | 45634 | 0.95 |
| FinnGen_EA | 9176 | 209616 | 0.92 |
| UKB_EA | 18385 | 91834 | 0.96 |
| UKB_AA | 280 | 1046 | 0.82 |
| MGB_AA | 225 | 1481 | 0.76 |
| MGB_EA | 2967 | 27957 | 1.01 |
| MGB_HIS | 100 | 1352 | 0.86 |
| UPenn_EA | 380 | 6210 | 0.92 |
| UPenn_AA | 203 | 1719 | 0.71 |
| FARIVE_EA | 545 | 532 | 0.78 |
| BioME_EA | 200 | 8007 | 1.13 |
| BioME_AA | 382 | 6074 | 1.03 |
| BioME_HIS | 397 | 8992 | 1.03 |
| RETROVE_EA | 400 | 400 | 0.96 |
| GAIT2_EA | 85 | 850 | 0.86 |
| MESA_EA | 94 | 2341 | 0.7 |
| MESA_AA | 72 | 1511 | 1.08 |
| MESA_HIS | 35 | 1389 | 0.62 |
| **Overall** | **75958** | **1062115** | **0.95** |

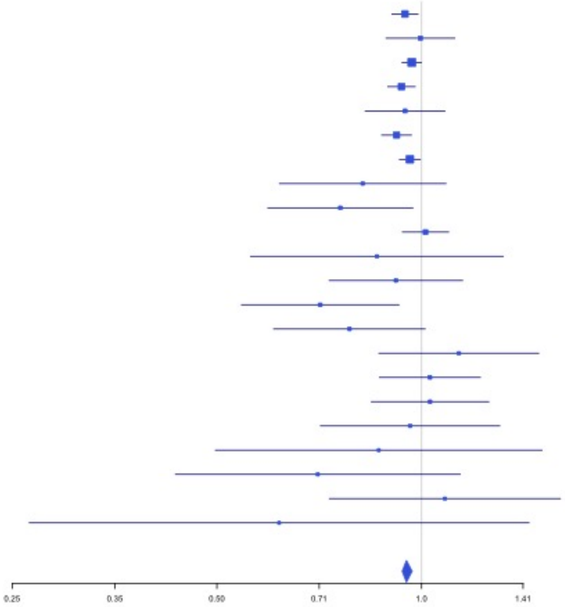

**Supplementary Figure 2.11** – rs143410348 (chr1:196809316) in locus 12 (CFHR1;CFHR4)

| Study | Cases | Controls | OR |
|---|---|---|---|
| INVENT-2019_EA | 15572 | 115280 | 1.02 |
| INVENT-2019_AA | 799 | 14353 | 1.03 |
| MVP_EA | 18952 | 411746 | 1.05 |
| MVP_AA | 5521 | 103791 | 1.04 |
| MVP_HIS | 1188 | 45634 | 1.08 |
| EGP_EA | 4122 | 181553 | 1.06 |
| UKB_EA | 18385 | 91834 | 1.04 |
| UKB_AA | 280 | 1046 | 1.25 |
| UKB_SAS | 189 | 1269 | 1.07 |
| UPenn_EA | 380 | 6210 | 1.16 |
| UPenn_AA | 203 | 1719 | 0.86 |
| FARIVE_EA | 545 | 532 | 1.02 |
| BBJ_EAS | 507 | 178366 | 0.99 |
| **Overall** | **66643** | **1153333** | **1.04** |

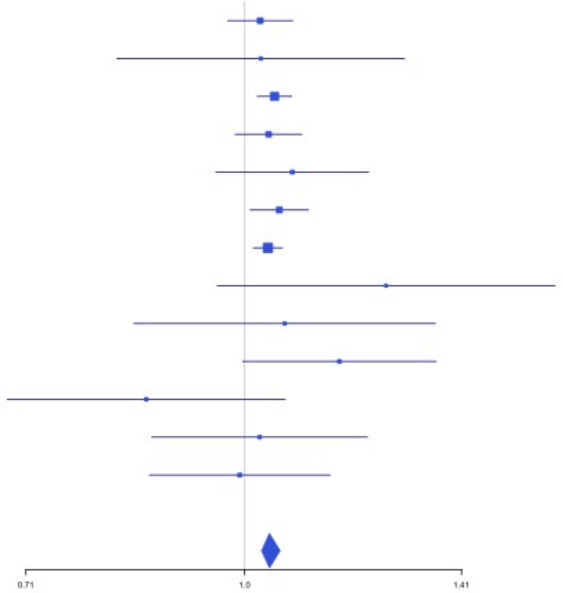


**Supplementary Figure 2.12** – rs2842700 (chr1:207282149) in locus 13 (C4BPA)

| Study | Cases | Controls | OR |
|---|---|---|---|
| INVENT-2019_EA | 15572 | 115280 | 1.18 |
| INVENT-2019_AA | 799 | 14353 | 1.04 |
| MVP_EA | 18952 | 411746 | 1.16 |
| MVP_AA | 5521 | 103791 | 1.07 |
| MVP_HIS | 1188 | 45634 | 1.13 |
| FinnGen_EA | 9176 | 209616 | 1.13 |
| EGP_EA | 4122 | 181553 | 1.02 |
| UKB_EA | 18385 | 91834 | 1.09 |
| UKB_AA | 280 | 1046 | 1.06 |
| MGB_AA | 225 | 1481 | 0.9 |
| MGB_EA | 2967 | 27957 | 1.07 |
| MGB_HIS | 100 | 1352 | 0.92 |
| UPenn_EA | 380 | 6210 | 1.15 |
| UPenn_AA | 203 | 1719 | 0.86 |
| FARIVE_EA | 545 | 532 | 1.45 |
| BioME_EA | 200 | 8007 | 0.9 |
| BioME_AA | 382 | 6074 | 0.98 |
| BioME_HIS | 397 | 8992 | 1 |
| RETROVE_EA | 400 | 400 | 1.12 |
| GAIT2_EA | 85 | 850 | 1.54 |
| MESA_EA | 94 | 2341 | 1.32 |
| MESA_AA | 72 | 1511 | 0.93 |
| MESA_HIS | 35 | 1389 | 1.46 |
| **Overall** | **80080** | **1243668** | **1.11** |

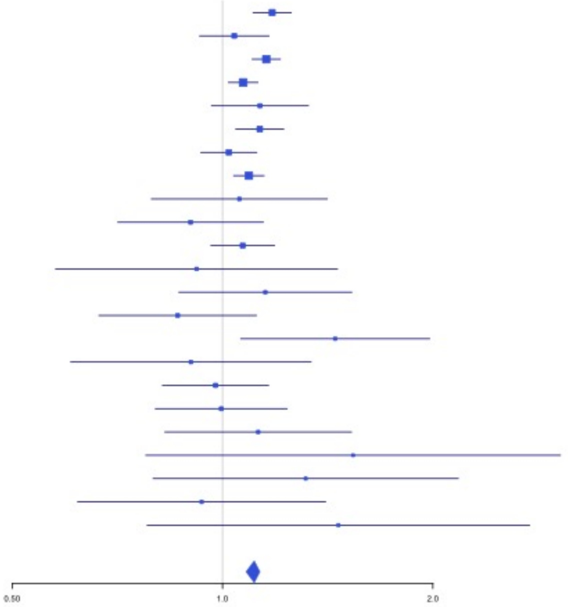

**Supplementary Figure 2.13** – rs3811444 (chr1:248039451) in locus 14 (TRIM58)

| Study | Cases | Controls | OR |
|---|---|---|---|
| INVENT-2019_EA | 15572 | 115280 | 0.99 |
| INVENT-2019_AA | 799 | 14353 | 1.02 |
| MVP_EA | 18952 | 411746 | 0.94 |
| MVP_AA | 5521 | 103791 | 1 |
| FinnGen_EA | 9176 | 209616 | 0.97 |
| EGP_EA | 4122 | 181553 | 0.95 |
| UKB_EA | 18385 | 91834 | 0.97 |
| UKB_SAS | 189 | 1269 | 0.89 |
| MGB_AA | 225 | 1481 | 1.47 |
| MGB_EA | 2967 | 27957 | 0.97 |
| MGB_HIS | 100 | 1352 | 0.81 |
| UPenn_EA | 380 | 6210 | 0.86 |
| UPenn_AA | 203 | 1719 | 0.71 |
| FARIVE_EA | 545 | 532 | 0.85 |
| BioME_EA | 200 | 8007 | 0.76 |
| BioME_AA | 382 | 6074 | 0.85 |
| BioME_HIS | 397 | 8992 | 0.93 |
| RETROVE_EA | 400 | 400 | 0.85 |
| GAIT2_EA | 85 | 850 | 1.09 |
| BBJ_EAS | 507 | 178366 | 1.07 |
| MESA_EA | 94 | 2341 | 0.8 |
| MESA_AA | 72 | 1511 | 0.78 |
| MESA_HIS | 35 | 1389 | 0.89 |
| MARTHA12_EA | 893 | 3414 | 0.92 |
| **Overall** | **80201** | **1380037** | **0.96** |

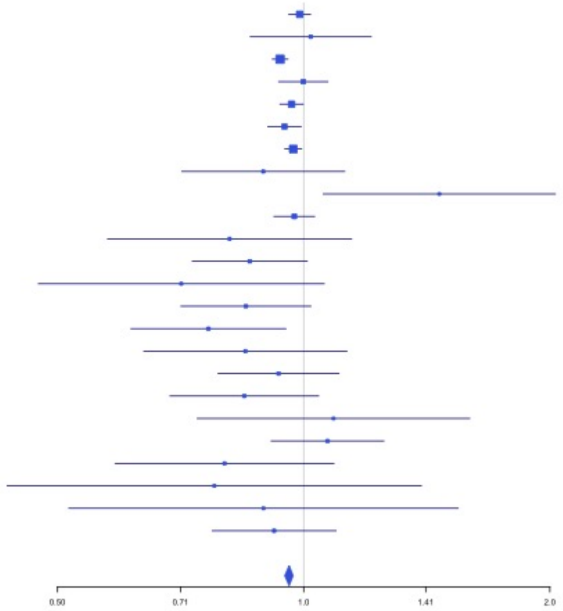


**Supplementary Figure 2.14** – rs78475244 (chr2:65086804) in locus 16 (LINC01800)

| Study | Cases | Controls | OR |
|---|---|---|---|
| INVENT-2019_EA | 15572 | 115280 | 0.94 |
| INVENT-2019_AA | 799 | 14353 | 1.06 |
| MVP_EA | 18952 | 411746 | 0.93 |
| MVP_AA | 5521 | 103791 | 0.89 |
| MVP_HIS | 1188 | 45634 | 0.95 |
| FinnGen_EA | 9176 | 209616 | 0.94 |
| EGP_EA | 4122 | 181553 | 0.96 |
| UKB_EA | 18385 | 91834 | 0.94 |
| MGB_AA | 225 | 1481 | 0.73 |
| MGB_EA | 2967 | 27957 | 0.91 |
| MGB_HIS | 100 | 1352 | 1.5 |
| UPenn_EA | 380 | 6210 | 0.88 |
| UPenn_AA | 203 | 1719 | 0.59 |
| FARIVE_EA | 545 | 532 | 1.29 |
| BioME_EA | 200 | 8007 | 0.68 |
| BioME_AA | 382 | 6074 | 0.89 |
| BioME_HIS | 397 | 8992 | 0.73 |
| RETROVE_EA | 400 | 400 | 0.85 |
| GAIT2_EA | 85 | 850 | 0.7 |
| BBJ_EAS | 507 | 178366 | 0.86 |
| MESA_EA | 94 | 2341 | 0.46 |
| MESA_AA | 72 | 1511 | 0.73 |
| MESA_HIS | 35 | 1389 | 1.47 |
| **Overall** | **80307** | **1420988** | **0.93** |

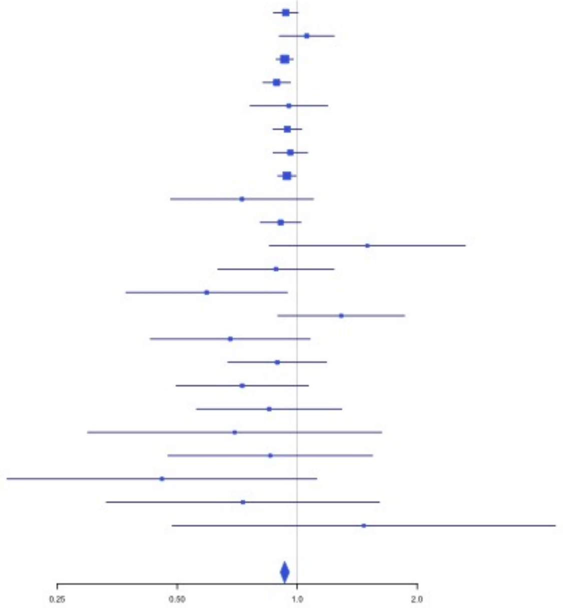

**Supplementary Figure 2.15** – rs1867312 (chr2:68619981) in locus 17 (PLEK)

| Study | Cases | Controls | OR |
|---|---|---|---|
| INVENT-2019_EA | 15572 | 115280 | 0.96 |
| INVENT-2019_AA | 799 | 14353 | 0.92 |
| MVP_EA | 18952 | 411746 | 0.96 |
| MVP_AA | 5521 | 103791 | 0.94 |
| MVP_HIS | 1188 | 45634 | 0.94 |
| FinnGen_EA | 9176 | 209616 | 0.95 |
| EGP_EA | 4122 | 181553 | 0.98 |
| UKB_EA | 18385 | 91834 | 0.93 |
| UKB_AA | 280 | 1046 | 0.95 |
| UKB_SAS | 189 | 1269 | 0.94 |
| MGB_AA | 225 | 1481 | 0.88 |
| MGB_EA | 2967 | 27957 | 0.94 |
| MGB_HIS | 100 | 1352 | 0.76 |
| UPenn_EA | 380 | 6210 | 0.91 |
| UPenn_AA | 203 | 1719 | 1.17 |
| FARIVE_EA | 545 | 532 | 0.92 |
| BioME_EA | 200 | 8007 | 0.83 |
| BioME_AA | 382 | 6074 | 0.95 |
| BioME_HIS | 397 | 8992 | 0.92 |
| RETROVE_EA | 400 | 400 | 0.88 |
| GAIT2_EA | 85 | 850 | 0.81 |
| BBJ_EAS | 507 | 178366 | 1.02 |
| MESA_EA | 94 | 2341 | 1.17 |
| MESA_AA | 72 | 1511 | 0.95 |
| MESA_HIS | 35 | 1389 | 0.91 |
| MARTHA12_EA | 893 | 3414 | 0.88 |
| **Overall** | **81669** | **1426717** | **0.95** |

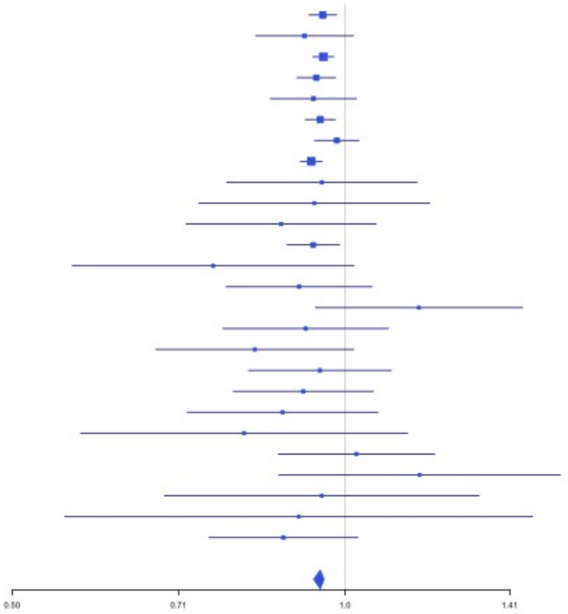


**Supplementary Figure 2.16** – rs1518760 (chr2:128140500) in locus 18 (MAP3K2)

| Study | Cases | Controls | OR |
|---|---|---|---|
| INVENT-2019_EA | 15572 | 115280 | 0.95 |
| INVENT-2019_AA | 799 | 14353 | 0.93 |
| MVP_EA | 18952 | 411746 | 0.92 |
| MVP_AA | 5521 | 103791 | 0.9 |
| MVP_HIS | 1188 | 45634 | 0.9 |
| FinnGen_EA | 9176 | 209616 | 0.95 |
| EGP_EA | 4122 | 181553 | 0.92 |
| UKB_EA | 18385 | 91834 | 0.95 |
| UKB_AA | 280 | 1046 | 0.8 |
| UKB_SAS | 189 | 1269 | 0.93 |
| MGB_AA | 225 | 1481 | 0.87 |
| MGB_EA | 2967 | 27957 | 0.95 |
| MGB_HIS | 100 | 1352 | 0.98 |
| UPenn_EA | 380 | 6210 | 0.81 |
| UPenn_AA | 203 | 1719 | 0.97 |
| FARIVE_EA | 545 | 532 | 0.92 |
| BioME_EA | 200 | 8007 | 1.07 |
| BioME_AA | 382 | 6074 | 0.83 |
| BioME_HIS | 397 | 8992 | 0.8 |
| RETROVE_EA | 400 | 400 | 0.88 |
| GAIT2_EA | 85 | 850 | 0.99 |
| BBJ_EAS | 507 | 178366 | 0.4 |
| MESA_EA | 94 | 2341 | 1.13 |
| MESA_AA | 72 | 1511 | 0.96 |
| MESA_HIS | 35 | 1389 | 1.08 |
| MARTHA12_EA | 893 | 3414 | 0.89 |
| **Overall** | **81669** | **1426717** | **0.93** |

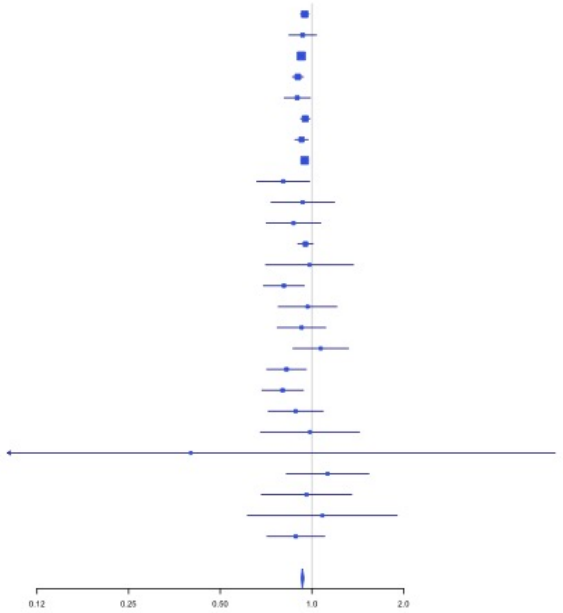

**Supplementary Figure 2.17** – rs1912849 (chr2:188310118) in locus 19 (CALCRL)

| Study | Cases | Controls | OR |
|---|---|---|---|
| INVENT-2019_EA | 15572 | 115280 | 1.03 |
| INVENT-2019_AA | 799 | 14353 | 1.01 |
| MVP_EA | 18952 | 411746 | 1.05 |
| MVP_AA | 5521 | 103791 | 1.05 |
| MVP_HIS | 1188 | 45634 | 1.02 |
| FinnGen_EA | 9176 | 209616 | 1.02 |
| EGP_EA | 4122 | 181553 | 1.04 |
| UKB_EA | 18385 | 91834 | 1.03 |
| UKB_AA | 280 | 1046 | 0.96 |
| UKB_SAS | 189 | 1269 | 1.18 |
| MGB_AA | 225 | 1481 | 1.29 |
| MGB_EA | 2967 | 27957 | 1.04 |
| MGB_HIS | 100 | 1352 | 1.2 |
| UPenn_EA | 380 | 6210 | 1.04 |
| UPenn_AA | 203 | 1719 | 1.12 |
| FARIVE_EA | 545 | 532 | 1.01 |
| BioME_EA | 200 | 8007 | 1.12 |
| BioME_AA | 382 | 6074 | 1.14 |
| BioME_HIS | 397 | 8992 | 1.1 |
| RETROVE_EA | 400 | 400 | 1.19 |
| GAIT2_EA | 85 | 850 | 1.18 |
| BBJ_EAS | 507 | 178366 | 1.05 |
| MESA_EA | 94 | 2341 | 1.19 |
| MESA_AA | 72 | 1511 | 1.05 |
| MESA_HIS | 35 | 1389 | 0.77 |
| MARTHA12_EA | 893 | 3414 | 1.08 |
| **Overall** | **81669** | **1426717** | **1.04** |

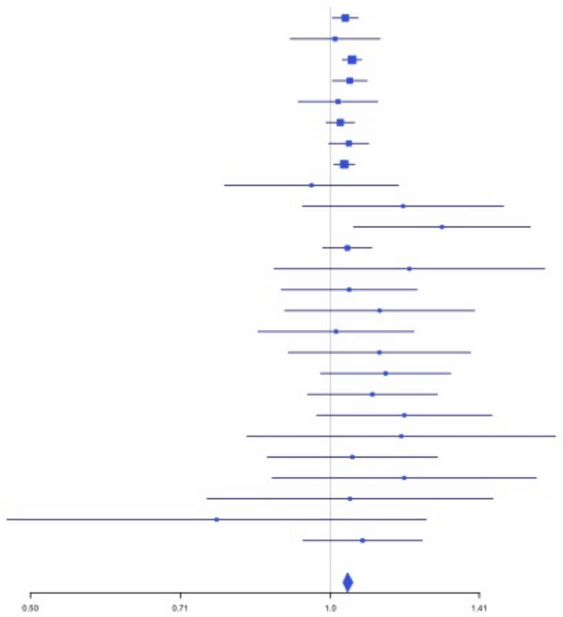


**Supplementary Figure 2.18** – rs78872368 (chr2:198545250) in locus 20 (RFTN2;MARS2)

| Study | Cases | Controls | OR |
|---|---|---|---|
| INVENT-2019_EA | 15572 | 115280 | 0.95 |
| INVENT-2019_AA | 799 | 14353 | 0.84 |
| MVP_EA | 18952 | 411746 | 0.96 |
| MVP_AA | 5521 | 103791 | 1.06 |
| MVP_HIS | 1188 | 45634 | 0.98 |
| FinnGen_EA | 9176 | 209616 | 0.96 |
| EGP_EA | 4122 | 181553 | 0.97 |
| UKB_EA | 18385 | 91834 | 0.96 |
| MGB_AA | 225 | 1481 | 0.99 |
| MGB_EA | 2967 | 27957 | 0.95 |
| MGB_HIS | 100 | 1352 | 1.08 |
| UPenn_EA | 380 | 6210 | 0.84 |
| UPenn_AA | 203 | 1719 | 0.87 |
| FARIVE_EA | 545 | 532 | 1.01 |
| BioME_EA | 200 | 8007 | 0.97 |
| BioME_AA | 382 | 6074 | 1 |
| BioME_HIS | 397 | 8992 | 0.79 |
| RETROVE_EA | 400 | 400 | 0.95 |
| GAIT2_EA | 85 | 850 | 0.85 |
| MESA_EA | 94 | 2341 | 0.88 |
| MESA_AA | 72 | 1511 | 0.89 |
| MESA_HIS | 35 | 1389 | 0.79 |
| MARTHA12_EA | 893 | 3414 | 0.98 |
| **Overall** | **80693** | **1246036** | **0.96** |

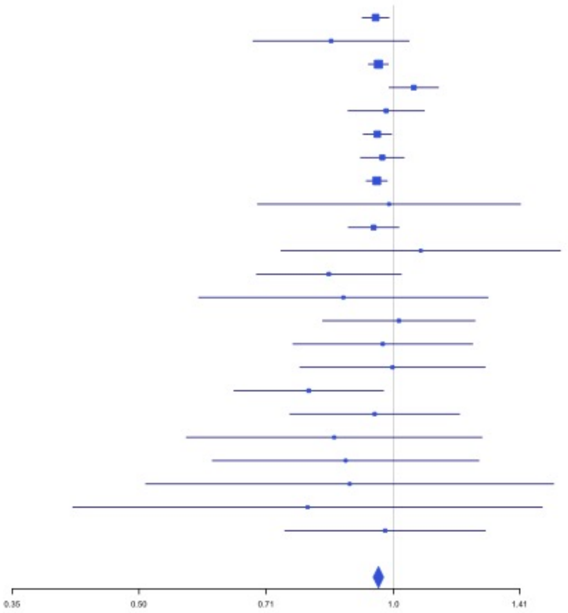

**Supplementary Figure 2.19** – rs715 (chr2:211543055) in locus 21 (CPS1)

| Study | Cases | Controls | OR |
|---|---|---|---|
| INVENT-2019_EA | 15572 | 115280 | 0.96 |
| INVENT-2019_AA | 799 | 14353 | 1 |
| MVP_EA | 18952 | 411746 | 0.95 |
| MVP_AA | 5521 | 103791 | 0.98 |
| MVP_HIS | 1188 | 45634 | 1.05 |
| FinnGen_EA | 9176 | 209616 | 0.93 |
| EGP_EA | 4122 | 181553 | 0.96 |
| UKB_EA | 18385 | 91834 | 0.97 |
| UKB_SAS | 189 | 1269 | 1.08 |
| MGB_AA | 225 | 1481 | 1.17 |
| MGB_EA | 2967 | 27957 | 1.01 |
| MGB_HIS | 100 | 1352 | 0.96 |
| UPenn_EA | 380 | 6210 | 1.1 |
| UPenn_AA | 203 | 1719 | 1.04 |
| FARIVE_EA | 545 | 532 | 1.12 |
| BioME_EA | 200 | 8007 | 0.95 |
| BioME_AA | 382 | 6074 | 1 |
| BioME_HIS | 397 | 8992 | 0.91 |
| RETROVE_EA | 400 | 400 | 1.02 |
| GAIT2_EA | 85 | 850 | 0.7 |
| BBJ_EAS | 507 | 178366 | 0.86 |
| MESA_EA | 94 | 2341 | 0.92 |
| MESA_AA | 72 | 1511 | 0.7 |
| MESA_HIS | 35 | 1389 | 0.9 |
| **Overall** | **80496** | **1422257** | **0.96** |

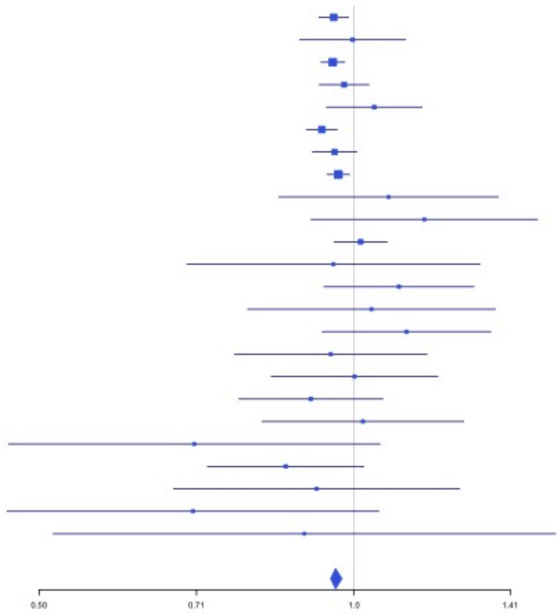


**Supplementary Figure 2.20** – rs68066031 (chr2:224880498) in locus 22 (SERPINE2)

| Study | Cases | Controls | OR |
|---|---|---|---|
| INVENT-2019_EA | 15572 | 115280 | 0.94 |
| INVENT-2019_AA | 799 | 14353 | 0.9 |
| MVP_EA | 18952 | 411746 | 0.94 |
| MVP_AA | 5521 | 103791 | 0.94 |
| MVP_HIS | 1188 | 45634 | 0.92 |
| FinnGen_EA | 9176 | 209616 | 0.95 |
| EGP_EA | 4122 | 181553 | 0.98 |
| UKB_EA | 18385 | 91834 | 0.96 |
| MGB_AA | 225 | 1481 | 0.87 |
| MGB_EA | 2967 | 27957 | 0.95 |
| MGB_HIS | 100 | 1352 | 0.81 |
| UPenn_EA | 380 | 6210 | 1 |
| UPenn_AA | 203 | 1719 | 0.68 |
| FARIVE_EA | 545 | 532 | 0.98 |
| BioME_EA | 200 | 8007 | 0.84 |
| BioME_AA | 382 | 6074 | 1.18 |
| BioME_HIS | 397 | 8992 | 0.93 |
| RETROVE_EA | 400 | 400 | 1.02 |
| GAIT2_EA | 85 | 850 | 1.25 |
| MESA_EA | 94 | 2341 | 1.07 |
| MESA_AA | 72 | 1511 | 0.97 |
| MESA_HIS | 35 | 1389 | 2.39 |
| **Overall** | **79800** | **1242622** | **0.95** |

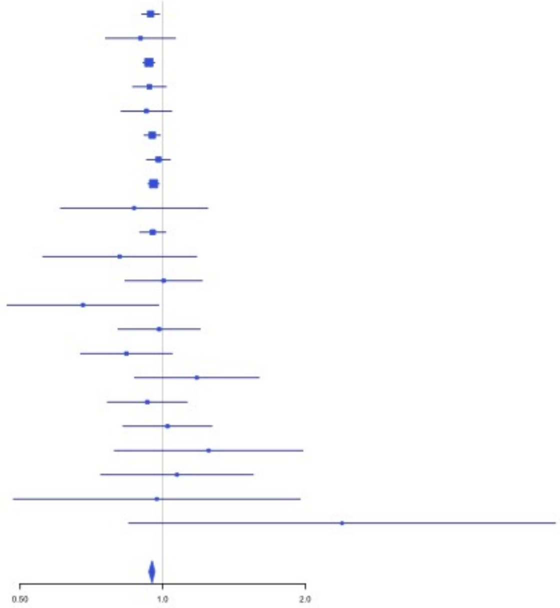

**Supplementary Figure 2.21** – rs13084580 (chr3:39188182) in locus 24 (CSRNP1)

| Study | Cases | Controls | OR |
|---|---|---|---|
| INVENT-2019_EA | 15572 | 115280 | 1.08 |
| INVENT-2019_AA | 799 | 14353 | 1.07 |
| MVP_EA | 18952 | 411746 | 1.1 |
| MVP_AA | 5521 | 103791 | 1.08 |
| MVP_HIS | 1188 | 45634 | 1.01 |
| FinnGen_EA | 9176 | 209616 | 1.12 |
| EGP_EA | 4122 | 181553 | 1.05 |
| UKB_EA | 18385 | 91834 | 1.05 |
| MGB_AA | 225 | 1481 | 1.02 |
| MGB_EA | 2967 | 27957 | 1.07 |
| MGB_HIS | 100 | 1352 | 1.13 |
| UPenn_EA | 380 | 6210 | 1.02 |
| UPenn_AA | 203 | 1719 | 1.01 |
| FARIVE_EA | 545 | 532 | 1.32 |
| BioME_EA | 200 | 8007 | 1.09 |
| BioME_AA | 382 | 6074 | 1.18 |
| BioME_HIS | 397 | 8992 | 0.99 |
| RETROVE_EA | 400 | 400 | 1.44 |
| GAIT2_EA | 85 | 850 | 0.98 |
| BBJ_EAS | 507 | 178366 | 0.88 |
| MESA_EA | 94 | 2341 | 1.04 |
| MESA_AA | 72 | 1511 | 1.59 |
| MESA_HIS | 35 | 1389 | 0.78 |
| MARTHA12_EA | 893 | 3414 | 1.2 |
| **Overall** | **81200** | **1424402** | **1.08** |

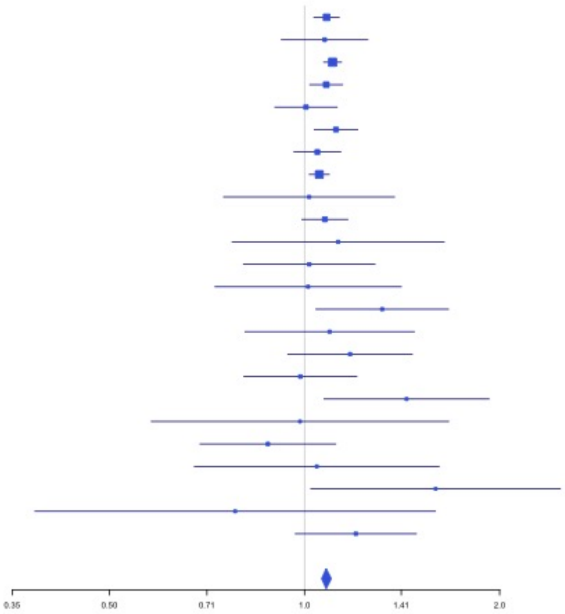


**Supplementary Figure 2.22** – rs11130326 (chr3:52771920) in locus 25 (NEK4)

| Study | Cases | Controls | OR |
|---|---|---|---|
| INVENT-2019_EA | 15572 | 115280 | 1.05 |
| INVENT-2019_AA | 799 | 14353 | 0.93 |
| MVP_EA | 18952 | 411746 | 1.04 |
| MVP_AA | 5521 | 103791 | 1.05 |
| MVP_HIS | 1188 | 45634 | 0.96 |
| FinnGen_EA | 9176 | 209616 | 1.05 |
| UPenn_EA | 380 | 6210 | 0.94 |
| UPenn_AA | 203 | 1719 | 1.08 |
| FARIVE_EA | 545 | 532 | 0.95 |
| MESA_AA | 72 | 1511 | 1.13 |
| MESA_HIS | 35 | 1389 | 0.93 |
| MARTHA12_EA | 893 | 3414 | 0.99 |
| **Overall** | **53336** | **915195** | **1.04** |

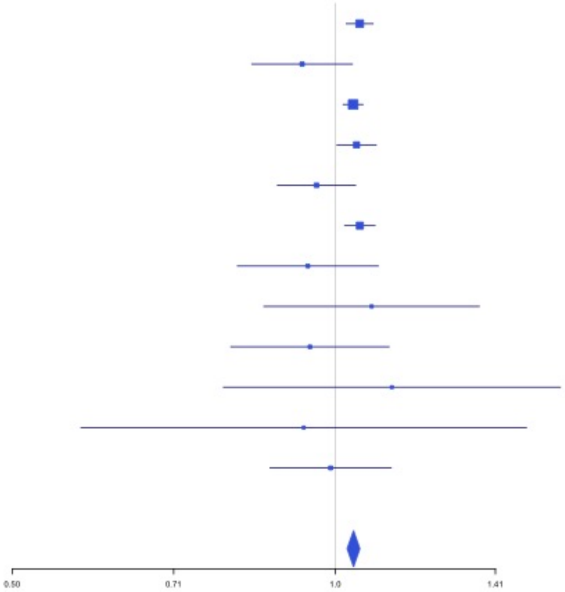

**Supplementary Figure 2.23** – rs529326506 (chr3:90503764) in locus 26 (EPHA3;NONE)

| Study | Cases | Controls | OR |
|---|---|---|---|
| MVP_AA | 5521 | 103791 | 1.5 |
| MVP_HIS | 1188 | 45634 | 1.93 |
| MGB_AA | 225 | 1481 | 1.15 |
| UPenn_AA | 203 | 1719 | 0.77 |
| BioME_AA | 382 | 6074 | 1.53 |
| BioME_HIS | 397 | 8992 | 1.73 |
| MESA_AA | 72 | 1511 | 1.64 |
| **Overall** | **7988** | **169202** | **1.48** |

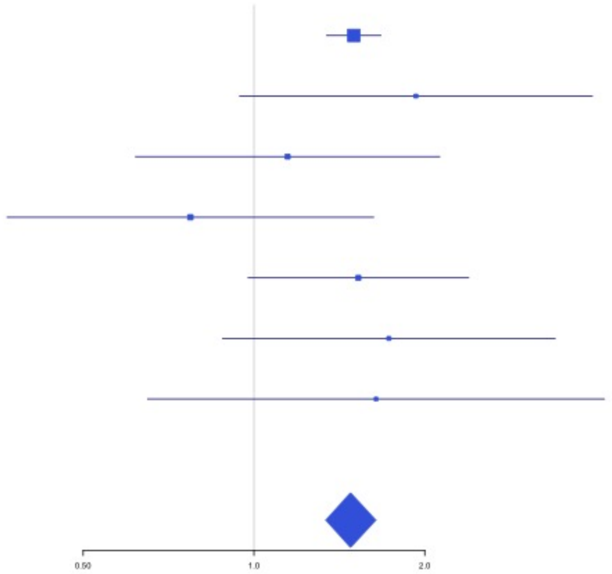


**Supplementary Figure 2.24** – rs28479320 (chr3:93752529) in locus 27 (ARL13B)

| Study | Cases | Controls | OR |
|---|---|---|---|
| INVENT-2019_EA | 15572 | 115280 | 0.94 |
| INVENT-2019_AA | 799 | 14353 | 0.86 |
| MVP_EA | 18952 | 411746 | 0.83 |
| MVP_AA | 5521 | 103791 | 0.87 |
| MVP_HIS | 1188 | 45634 | 0.87 |
| FinnGen_EA | 9176 | 209616 | 0.91 |
| EGP_EA | 4122 | 181553 | 1.07 |
| UKB_EA | 18385 | 91834 | 0.87 |
| MGB_AA | 225 | 1481 | 0.74 |
| MGB_EA | 2967 | 27957 | 0.86 |
| MGB_HIS | 100 | 1352 | 1.08 |
| UPenn_EA | 380 | 6210 | 1.08 |
| UPenn_AA | 203 | 1719 | 0.76 |
| FARIVE_EA | 545 | 532 | 0.78 |
| BioME_EA | 200 | 8007 | 0.76 |
| BioME_AA | 382 | 6074 | 0.76 |
| BioME_HIS | 397 | 8992 | 0.93 |
| RETROVE_EA | 400 | 400 | 0.68 |
| GAIT2_EA | 85 | 850 | 0.7 |
| BBJ_EAS | 507 | 178366 | 0.85 |
| MESA_EA | 94 | 2341 | 0.55 |
| MESA_AA | 72 | 1511 | 1.39 |
| MESA_HIS | 35 | 1389 | 0.87 |
| **Overall** | **80307** | **1420988** | **0.87** |

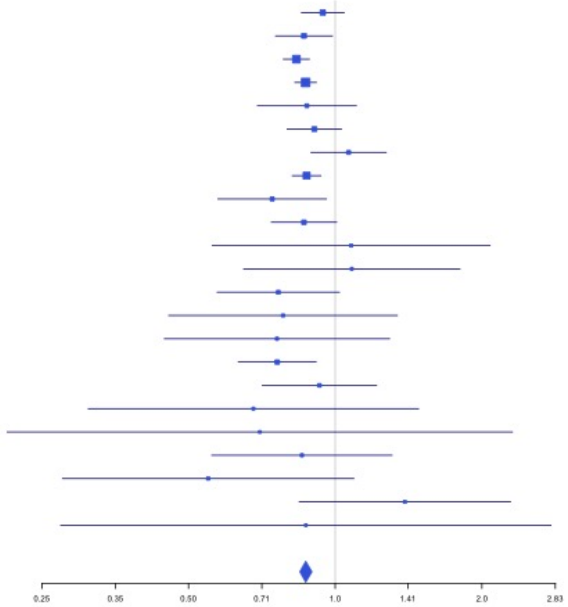

**Supplementary Figure 2.25** – rs900399 (chr3:156798732) in locus 30 (LINC02029;LINC00880)

| Study | Cases | Controls | OR |
|---|---|---|---|
| INVENT-2019_EA | 15572 | 115280 | 1.04 |
| INVENT-2019_AA | 799 | 14353 | 0.98 |
| MVP_EA | 18952 | 411746 | 1.03 |
| MVP_AA | 5521 | 103791 | 1.04 |
| MVP_HIS | 1188 | 45634 | 1.07 |
| FinnGen_EA | 9176 | 209616 | 1.06 |
| EGP_EA | 4122 | 181553 | 1.03 |
| UKB_EA | 18385 | 91834 | 1.03 |
| UKB_AA | 280 | 1046 | 1.13 |
| UKB_SAS | 189 | 1269 | 1.08 |
| MGB_AA | 225 | 1481 | 0.92 |
| MGB_EA | 2967 | 27957 | 1.07 |
| MGB_HIS | 100 | 1352 | 1.12 |
| UPenn_EA | 380 | 6210 | 1.04 |
| UPenn_AA | 203 | 1719 | 1 |
| FARIVE_EA | 545 | 532 | 1 |
| BioME_EA | 200 | 8007 | 1.09 |
| BioME_AA | 382 | 6074 | 1.07 |
| BioME_HIS | 397 | 8992 | 0.93 |
| RETROVE_EA | 400 | 400 | 1.01 |
| GAIT2_EA | 85 | 850 | 1.2 |
| BBJ_EAS | 507 | 178366 | 1.05 |
| MESA_EA | 94 | 2341 | 1.21 |
| MESA_AA | 72 | 1511 | 1.11 |
| MESA_HIS | 35 | 1389 | 1.01 |
| MARTHA12_EA | 893 | 3414 | 1.11 |
| **Overall** | **81669** | **1426717** | **1.04** |

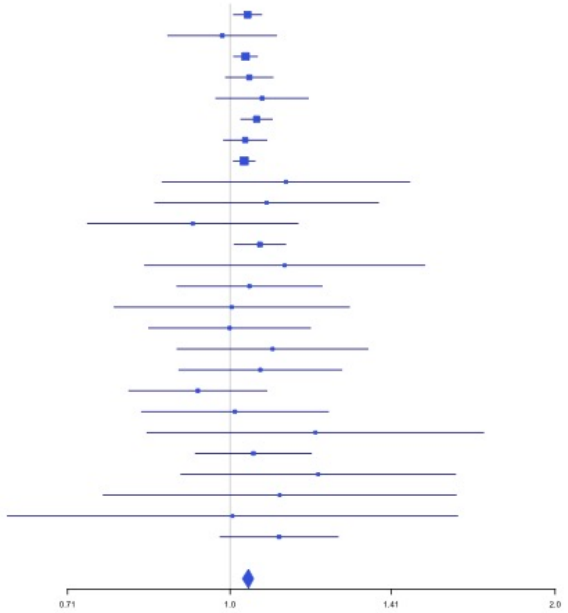


**Supplementary Figure 2.26** – rs710446 (chr3:186459927) in locus 32 (KNG1)

| Study | Cases | Controls | OR |
|---|---|---|---|
| INVENT-2019_EA | 15572 | 115280 | 0.94 |
| INVENT-2019_AA | 799 | 14353 | 0.95 |
| MVP_EA | 18952 | 411746 | 0.97 |
| MVP_AA | 5521 | 103791 | 0.98 |
| MVP_HIS | 1188 | 45634 | 0.95 |
| FinnGen_EA | 9176 | 209616 | 0.94 |
| EGP_EA | 4122 | 181553 | 0.96 |
| UKB_EA | 18385 | 91834 | 0.96 |
| UKB_AA | 280 | 1046 | 1.08 |
| UKB_SAS | 189 | 1269 | 0.97 |
| MGB_AA | 225 | 1481 | 0.9 |
| MGB_EA | 2967 | 27957 | 0.95 |
| MGB_HIS | 100 | 1352 | 1.06 |
| UPenn_EA | 380 | 6210 | 0.91 |
| UPenn_AA | 203 | 1719 | 1.03 |
| FARIVE_EA | 545 | 532 | 0.83 |
| BioME_EA | 200 | 8007 | 1.04 |
| BioME_AA | 382 | 6074 | 1.08 |
| BioME_HIS | 397 | 8992 | 0.96 |
| RETROVE_EA | 400 | 400 | 0.86 |
| GAIT2_EA | 85 | 850 | 1.04 |
| BBJ_EAS | 507 | 178366 | 0.98 |
| MESA_EA | 94 | 2341 | 1.15 |
| MESA_AA | 72 | 1511 | 1.34 |
| MESA_HIS | 35 | 1389 | 1.01 |
| MARTHA12_EA | 893 | 3414 | 0.85 |
| **Overall** | **81669** | **1426717** | **0.96** |

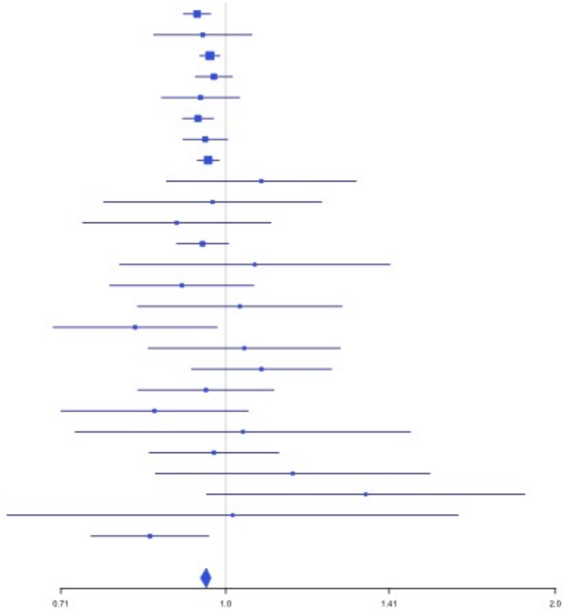

**Supplementary Figure 2.27** – rs3796159 (chr3:194790434) in locus 33 (XXYLT1)

| Study | Cases | Controls | OR |
|---|---|---|---|
| INVENT-2019_EA | 15572 | 115280 | 1.06 |
| INVENT-2019_AA | 799 | 14353 | 1.09 |
| MVP_EA | 18952 | 411746 | 1.07 |
| MVP_AA | 5521 | 103791 | 1.03 |
| FinnGen_EA | 9176 | 209616 | 1.04 |
| UKB_EA | 18385 | 91834 | 1.05 |
| UKB_AA | 280 | 1046 | 1.13 |
| UKB_SAS | 189 | 1269 | 1.06 |
| MGB_AA | 225 | 1481 | 1.1 |
| MGB_EA | 2967 | 27957 | 1 |
| MGB_HIS | 100 | 1352 | 0.92 |
| UPenn_EA | 380 | 6210 | 1.38 |
| UPenn_AA | 203 | 1719 | 0.91 |
| FARIVE_EA | 545 | 532 | 1.14 |
| BioME_EA | 200 | 8007 | 0.87 |
| BioME_AA | 382 | 6074 | 1.13 |
| BioME_HIS | 397 | 8992 | 1.1 |
| RETROVE_EA | 400 | 400 | 1.05 |
| GAIT2_EA | 85 | 850 | 0.93 |
| BBJ_EAS | 507 | 178366 | 0.98 |
| MESA_EA | 94 | 2341 | 1.09 |
| MESA_AA | 72 | 1511 | 1.07 |
| MESA_HIS | 35 | 1389 | 0.98 |
| **Overall** | **75466** | **1196116** | **1.06** |

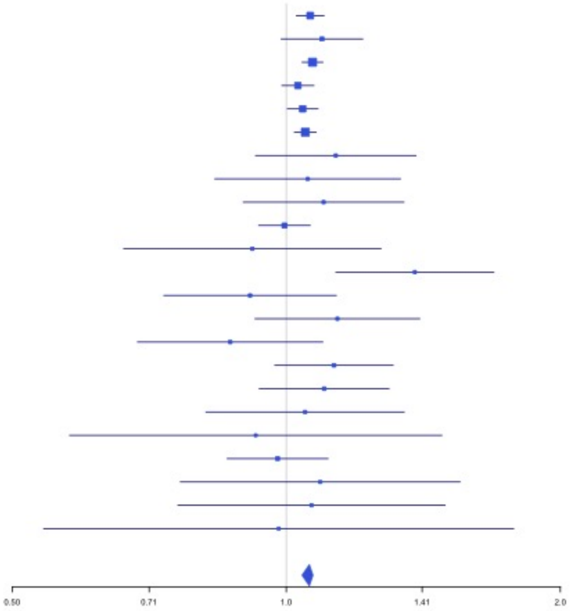


**Supplementary Figure 2.28** – rs9654093 (chr4:7903763) in locus 34 (AFAP1)

| Study | Cases | Controls | OR |
|---|---|---|---|
| INVENT-2019_EA | 15572 | 115280 | 1.05 |
| INVENT-2019_AA | 799 | 14353 | 1.14 |
| MVP_EA | 18952 | 411746 | 1.03 |
| MVP_AA | 5521 | 103791 | 1.05 |
| MVP_HIS | 1188 | 45634 | 1 |
| FinnGen_EA | 9176 | 209616 | 1.04 |
| EGP_EA | 4122 | 181553 | 1.08 |
| UKB_EA | 18385 | 91834 | 1.06 |
| UKB_AA | 280 | 1046 | 1.14 |
| MGB_AA | 225 | 1481 | 0.86 |
| MGB_EA | 2967 | 27957 | 1.04 |
| MGB_HIS | 100 | 1352 | 1.07 |
| UPenn_EA | 380 | 6210 | 0.97 |
| UPenn_AA | 203 | 1719 | 0.96 |
| FARIVE_EA | 545 | 532 | 1.06 |
| BioME_EA | 200 | 8007 | 1.32 |
| BioME_AA | 382 | 6074 | 1.21 |
| BioME_HIS | 397 | 8992 | 1.1 |
| RETROVE_EA | 400 | 400 | 1.09 |
| GAIT2_EA | 85 | 850 | 1.52 |
| BBJ_EAS | 507 | 178366 | 1.11 |
| MESA_EA | 94 | 2341 | 1.37 |
| MESA_AA | 72 | 1511 | 0.86 |
| MESA_HIS | 35 | 1389 | 0.9 |
| **Overall** | **80587** | **1422034** | **1.05** |

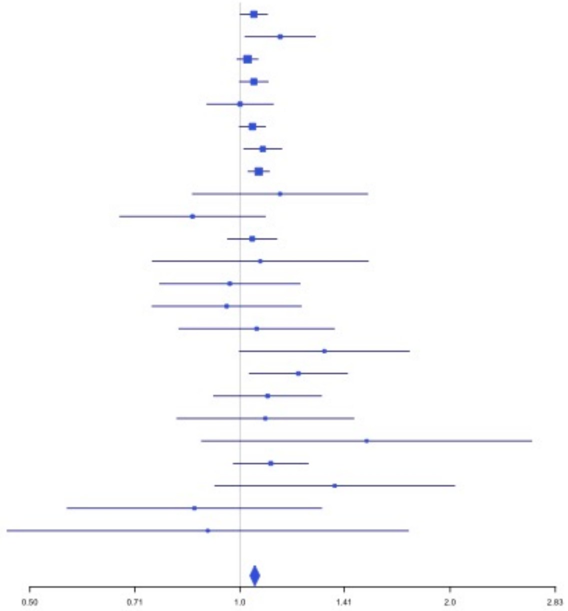

**Supplementary Figure 2.29** – rs781656 (chr4:57778645) in locus 35 (REST)

| Study | Cases | Controls | OR |
|---|---|---|---|
| INVENT-2019_EA | 15572 | 115280 | 1.04 |
| INVENT-2019_AA | 799 | 14353 | 1.11 |
| MVP_EA | 18952 | 411746 | 1.04 |
| MVP_AA | 5521 | 103791 | 1.06 |
| MVP_HIS | 1188 | 45634 | 1.04 |
| FinnGen_EA | 9176 | 209616 | 1.01 |
| EGP_EA | 4122 | 181553 | 1.02 |
| UKB_EA | 18385 | 91834 | 1.05 |
| UKB_SAS | 189 | 1269 | 1.4 |
| MGB_AA | 225 | 1481 | 1.04 |
| MGB_EA | 2967 | 27957 | 1.01 |
| MGB_HIS | 100 | 1352 | 1.18 |
| UPenn_EA | 380 | 6210 | 1.01 |
| UPenn_AA | 203 | 1719 | 1.24 |
| FARIVE_EA | 545 | 532 | 1.13 |
| BioME_EA | 200 | 8007 | 1.08 |
| BioME_AA | 382 | 6074 | 1.04 |
| BioME_HIS | 397 | 8992 | 0.98 |
| RETROVE_EA | 400 | 400 | 0.98 |
| GAIT2_EA | 85 | 850 | 0.7 |
| BBJ_EAS | 507 | 178366 | 0.95 |
| MESA_EA | 94 | 2341 | 1.06 |
| MESA_AA | 72 | 1511 | 1.14 |
| MESA_HIS | 35 | 1389 | 0.91 |
| MARTHA12_EA | 893 | 3414 | 1.29 |
| **Overall** | **81389** | **1425671** | **1.04** |

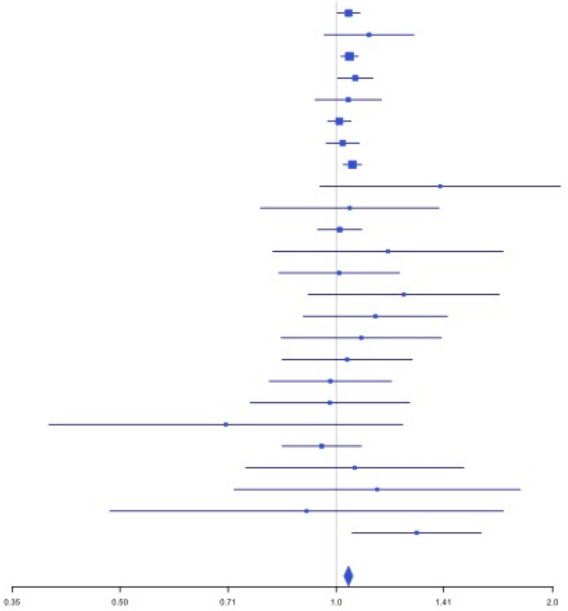


**Supplementary Figure 2.30** – rs6826579 (chr4:83785031) in locus 36 (SEC31A)

| Study | Cases | Controls | OR |
|---|---|---|---|
| INVENT-2019_EA | 15572 | 115280 | 1.05 |
| INVENT-2019_AA | 799 | 14353 | 1.07 |
| MVP_EA | 18952 | 411746 | 1.05 |
| MVP_AA | 5521 | 103791 | 1.03 |
| MVP_HIS | 1188 | 45634 | 0.99 |
| FinnGen_EA | 9176 | 209616 | 1.06 |
| EGP_EA | 4122 | 181553 | 1.05 |
| UKB_EA | 18385 | 91834 | 1.02 |
| UKB_SAS | 189 | 1269 | 0.89 |
| MGB_AA | 225 | 1481 | 1.11 |
| MGB_EA | 2967 | 27957 | 1 |
| MGB_HIS | 100 | 1352 | 1.08 |
| UPenn_EA | 380 | 6210 | 0.93 |
| UPenn_AA | 203 | 1719 | 1.12 |
| FARIVE_EA | 545 | 532 | 1 |
| BioME_EA | 200 | 8007 | 1.3 |
| BioME_AA | 382 | 6074 | 1.05 |
| BioME_HIS | 397 | 8992 | 0.92 |
| RETROVE_EA | 400 | 400 | 1.09 |
| GAIT2_EA | 85 | 850 | 0.96 |
| BBJ_EAS | 507 | 178366 | 1.05 |
| MESA_EA | 94 | 2341 | 1.29 |
| MESA_AA | 72 | 1511 | 1.48 |
| MESA_HIS | 35 | 1389 | 1.33 |
| **Overall** | **80496** | **1422257** | **1.04** |

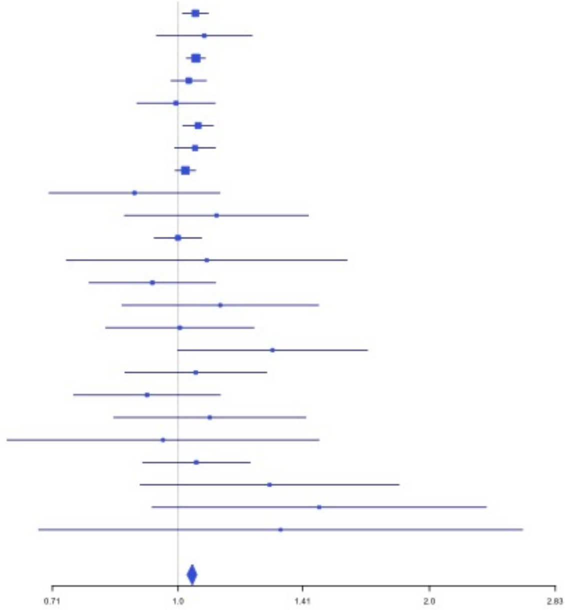

**Supplementary Figure 2.31** – rs2014912 (chr4:86715670) in locus 37 (ARHGAP24)

| Study | Cases | Controls | OR |
|---|---|---|---|
| INVENT-2019_EA | 15572 | 115280 | 0.97 |
| INVENT-2019_AA | 799 | 14353 | 0.92 |
| MVP_EA | 18952 | 411746 | 0.95 |
| MVP_AA | 5521 | 103791 | 0.96 |
| MVP_HIS | 1188 | 45634 | 0.83 |
| FinnGen_EA | 9176 | 209616 | 0.94 |
| EGP_EA | 4122 | 181553 | 0.93 |
| UKB_EA | 18385 | 91834 | 0.95 |
| MGB_AA | 225 | 1481 | 0.77 |
| MGB_EA | 2967 | 27957 | 0.93 |
| MGB_HIS | 100 | 1352 | 1.11 |
| UPenn_EA | 380 | 6210 | 1.06 |
| UPenn_AA | 203 | 1719 | 1.13 |
| FARIVE_EA | 545 | 532 | 0.84 |
| BioME_EA | 200 | 8007 | 0.86 |
| BioME_AA | 382 | 6074 | 1.01 |
| BioME_HIS | 397 | 8992 | 0.87 |
| RETROVE_EA | 400 | 400 | 0.98 |
| GAIT2_EA | 85 | 850 | 0.75 |
| BBJ_EAS | 507 | 178366 | 0.87 |
| MESA_EA | 94 | 2341 | 0.75 |
| MESA_AA | 72 | 1511 | 1.11 |
| MESA_HIS | 35 | 1389 | 1.79 |
| **Overall** | **80307** | **1420988** | **0.95** |

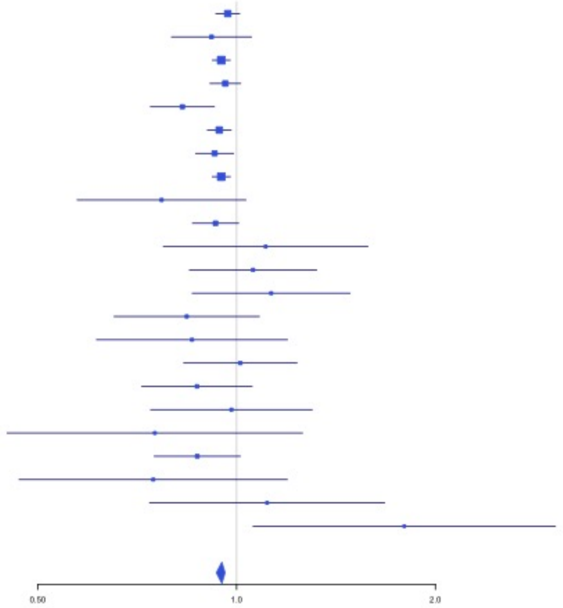


**Supplementary Figure 2.32** – rs2066864 (chr4:155525695) in locus 39 (FGG)

| Study | Cases | Controls | OR |
|---|---|---|---|
| INVENT-2019_EA | 15572 | 115280 | 1.2 |
| INVENT-2019_AA | 799 | 14353 | 1.17 |
| MVP_EA | 18952 | 411746 | 1.26 |
| MVP_AA | 5521 | 103791 | 1.19 |
| MVP_HIS | 1188 | 45634 | 1.26 |
| FinnGen_EA | 9176 | 209616 | 1.24 |
| EGP_EA | 4122 | 181553 | 1.18 |
| UKB_EA | 18385 | 91834 | 1.22 |
| UKB_AA | 280 | 1046 | 1.14 |
| UKB_SAS | 189 | 1269 | 1.14 |
| MGB_AA | 225 | 1481 | 1.23 |
| MGB_EA | 2967 | 27957 | 1.19 |
| UPenn_EA | 380 | 6210 | 1.36 |
| UPenn_AA | 203 | 1719 | 1.38 |
| FARIVE_EA | 545 | 532 | 1.51 |
| BioME_EA | 200 | 8007 | 1.3 |
| BioME_AA | 382 | 6074 | 1.16 |
| BioME_HIS | 397 | 8992 | 1.16 |
| RETROVE_EA | 400 | 400 | 1.46 |
| GAIT2_EA | 85 | 850 | 1.06 |
| BBJ_EAS | 507 | 178366 | 1.19 |
| MESA_EA | 94 | 2341 | 0.98 |
| MESA_AA | 72 | 1511 | 1.2 |
| MESA_HIS | 35 | 1389 | 0.73 |
| MARTHA12_EA | 893 | 3414 | 1.46 |
| **Overall** | **81569** | **1425365** | **1.23** |

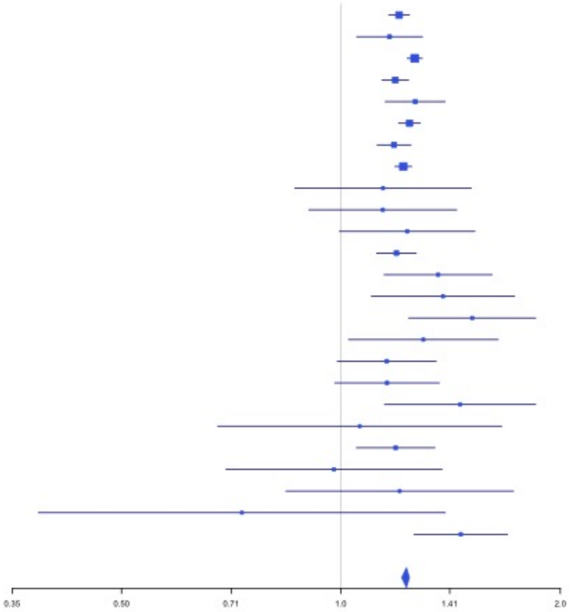

**Supplementary Figure 2.33** – rs3756011 (chr4:187206249) in locus 40 (F11)

| Study | Cases | Controls | OR |
|---|---|---|---|
| INVENT-2019_EA | 15572 | 115280 | 1.2 |
| INVENT-2019_AA | 799 | 14353 | 1.15 |
| MVP_EA | 18952 | 411746 | 1.26 |
| MVP_AA | 5521 | 103791 | 1.18 |
| MVP_HIS | 1188 | 45634 | 1.2 |
| FinnGen_EA | 9176 | 209616 | 1.23 |
| EGP_EA | 4122 | 181553 | 1.25 |
| UKB_EA | 18385 | 91834 | 1.18 |
| UKB_AA | 280 | 1046 | 1.17 |
| UKB_SAS | 189 | 1269 | 1.01 |
| MGB_AA | 225 | 1481 | 1.3 |
| MGB_EA | 2967 | 27957 | 1.18 |
| MGB_HIS | 100 | 1352 | 0.95 |
| UPenn_EA | 380 | 6210 | 1.19 |
| UPenn_AA | 203 | 1719 | 1.05 |
| FARIVE_EA | 545 | 532 | 1.47 |
| BioME_EA | 200 | 8007 | 1.03 |
| BioME_AA | 382 | 6074 | 1.12 |
| BioME_HIS | 397 | 8992 | 1.14 |
| RETROVE_EA | 400 | 400 | 1.4 |
| GAIT2_EA | 85 | 850 | 1.53 |
| BBJ_EAS | 507 | 178366 | 1.14 |
| MESA_EA | 94 | 2341 | 0.99 |
| MESA_AA | 72 | 1511 | 1.17 |
| MESA_HIS | 35 | 1389 | 1.22 |
| **Overall** | **80776** | **1423303** | **1.21** |

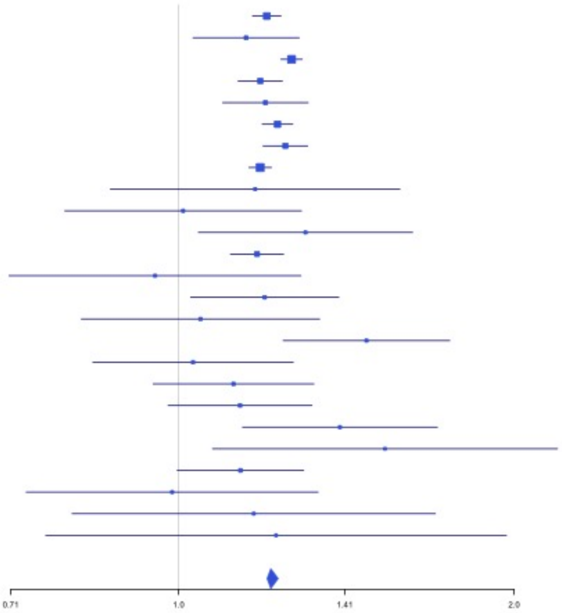


**Supplementary Figure 2.34** – rs16867574 (chr5:38708554) in locus 42 (OSMR-AS1)

| Study | Cases | Controls | OR |
|---|---|---|---|
| INVENT-2019_EA | 15572 | 115280 | 0.93 |
| INVENT-2019_AA | 799 | 14353 | 1.14 |
| MVP_EA | 18952 | 411746 | 0.95 |
| MVP_AA | 5521 | 103791 | 0.94 |
| MVP_HIS | 1188 | 45634 | 0.95 |
| FinnGen_EA | 9176 | 209616 | 0.99 |
| EGP_EA | 4122 | 181553 | 0.93 |
| UKB_EA | 18385 | 91834 | 0.95 |
| UKB_SAS | 189 | 1269 | 0.96 |
| MGB_AA | 225 | 1481 | 1.02 |
| MGB_EA | 2967 | 27957 | 0.97 |
| MGB_HIS | 100 | 1352 | 0.85 |
| UPenn_EA | 380 | 6210 | 1.01 |
| UPenn_AA | 203 | 1719 | 0.85 |
| FARIVE_EA | 545 | 532 | 0.95 |
| BioME_EA | 200 | 8007 | 0.96 |
| BioME_AA | 382 | 6074 | 0.95 |
| BioME_HIS | 397 | 8992 | 1.05 |
| RETROVE_EA | 400 | 400 | 0.86 |
| GAIT2_EA | 85 | 850 | 1.37 |
| BBJ_EAS | 507 | 178366 | 0.98 |
| MESA_EA | 94 | 2341 | 1.13 |
| MESA_AA | 72 | 1511 | 0.96 |
| MESA_HIS | 35 | 1389 | 0.8 |
| **Overall** | **80496** | **1422257** | **0.95** |

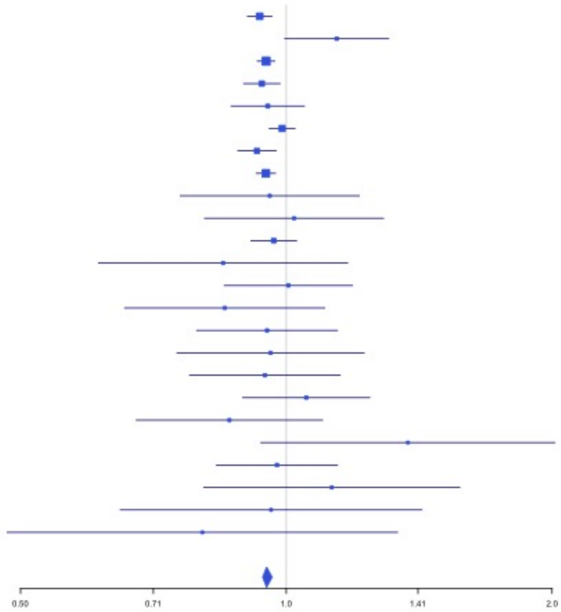

**Supplementary Figure 2.35** – rs7730244 (chr5:72957088) in locus 43 (ARHGEF28)

| Study | Cases | Controls | OR |
|---|---|---|---|
| INVENT-2019_EA | 15572 | 115280 | 0.98 |
| INVENT-2019_AA | 799 | 14353 | 0.95 |
| MVP_EA | 18952 | 411746 | 0.96 |
| MVP_AA | 5521 | 103791 | 0.98 |
| MVP_HIS | 1188 | 45634 | 0.93 |
| FinnGen_EA | 9176 | 209616 | 0.95 |
| EGP_EA | 4122 | 181553 | 0.98 |
| UKB_EA | 18385 | 91834 | 0.97 |
| UKB_AA | 280 | 1046 | 1.15 |
| UKB_SAS | 189 | 1269 | 0.89 |
| MGB_AA | 225 | 1481 | 1.03 |
| MGB_EA | 2967 | 27957 | 1 |
| UPenn_EA | 380 | 6210 | 1.02 |
| UPenn_AA | 203 | 1719 | 1.02 |
| FARIVE_EA | 545 | 532 | 0.9 |
| BioME_EA | 200 | 8007 | 1.02 |
| BioME_AA | 382 | 6074 | 0.86 |
| BioME_HIS | 397 | 8992 | 1 |
| RETROVE_EA | 400 | 400 | 1.06 |
| GAIT2_EA | 85 | 850 | 1.18 |
| BBJ_EAS | 507 | 178366 | 0.89 |
| MESA_EA | 94 | 2341 | 1.19 |
| MESA_AA | 72 | 1511 | 1.03 |
| MESA_HIS | 35 | 1389 | 1.35 |
| **Overall** | **80676** | **1421951** | **0.97** |

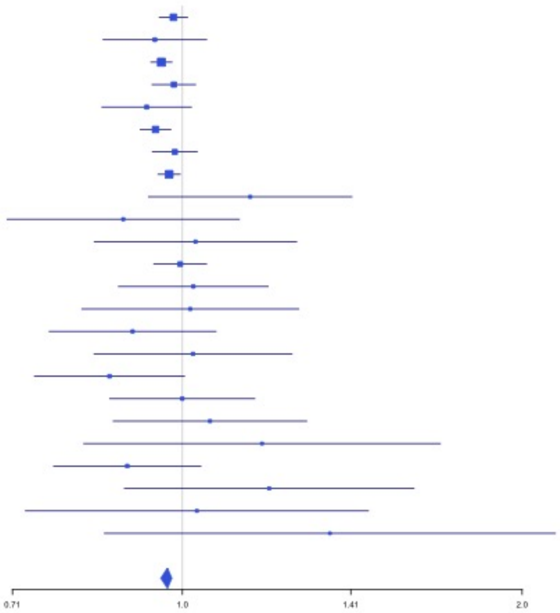


**Supplementary Figure 2.36** – rs750465397 (chr5:96289036) in locus 44 (LNPEP)

| Study | Cases | Controls | OR |
|---|---|---|---|
| INVENT-2019_EA | 15572 | 115280 | 0.97 |
| INVENT-2019_AA | 799 | 14353 | 1.05 |
| MVP_EA | 18952 | 411746 | 0.96 |
| MVP_AA | 5521 | 103791 | 0.96 |
| MVP_HIS | 1188 | 45634 | 0.97 |
| FinnGen_EA | 9176 | 209616 | 0.96 |
| EGP_EA | 4122 | 181553 | 0.97 |
| UKB_EA | 18385 | 91834 | 0.97 |
| UKB_AA | 280 | 1046 | 1.03 |
| UKB_SAS | 189 | 1269 | 1 |
| MGB_AA | 225 | 1481 | 1.07 |
| MGB_EA | 2967 | 27957 | 0.98 |
| MGB_HIS | 100 | 1352 | 0.87 |
| UPenn_EA | 380 | 6210 | 1.16 |
| UPenn_AA | 203 | 1719 | 0.77 |
| FARIVE_EA | 545 | 532 | 0.97 |
| BioME_EA | 200 | 8007 | 1.07 |
| BioME_AA | 382 | 6074 | 1 |
| BioME_HIS | 397 | 8992 | 0.99 |
| BBJ_EAS | 507 | 178366 | 1.01 |
| MARTHA12_EA | 893 | 3414 | 0.81 |
| **Overall** | **80983** | **1420226** | **0.97** |

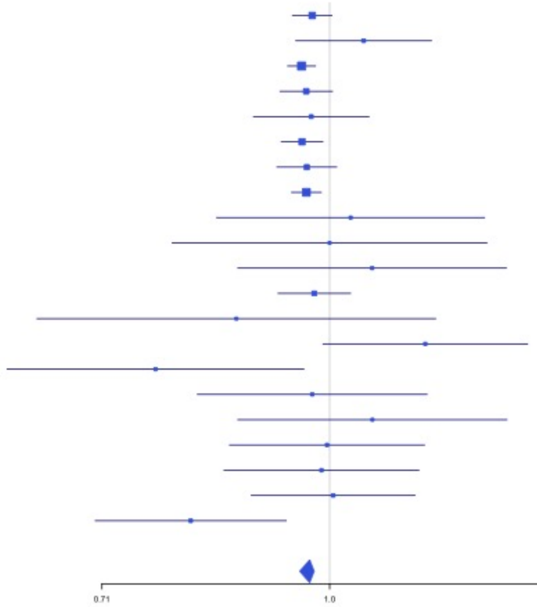

**Supplementary Figure 2.37** – rs147133967 (chr5:132426851) in locus 45 (HSPA4)

| Study | Cases | Controls | OR |
|---|---|---|---|
| INVENT-2019_EA | 15572 | 115280 | 0.96 |
| MVP_EA | 18952 | 411746 | 0.9 |
| MVP_AA | 5521 | 103791 | 1.14 |
| MVP_HIS | 1188 | 45634 | 1.04 |
| FinnGen_EA | 9176 | 209616 | 0.93 |
| EGP_EA | 4122 | 181553 | 0.96 |
| UKB_EA | 18385 | 91834 | 0.94 |
| MGB_AA | 225 | 1481 | 0.86 |
| MGB_EA | 2967 | 27957 | 0.94 |
| MGB_HIS | 100 | 1352 | 2.75 |
| UPenn_EA | 380 | 6210 | 1.11 |
| UPenn_AA | 203 | 1719 | 0.83 |
| FARIVE_EA | 545 | 532 | 1.12 |
| BioME_EA | 200 | 8007 | 0.92 |
| BioME_AA | 382 | 6074 | 0.49 |
| BioME_HIS | 397 | 8992 | 0.96 |
| **Overall** | **78315** | **1221778** | **0.94** |

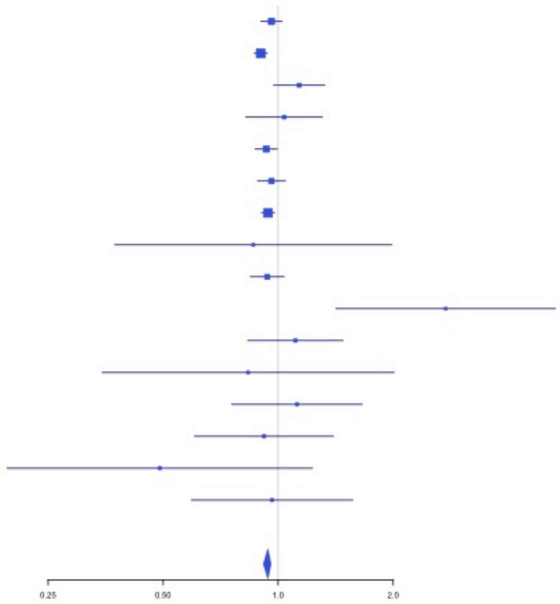


**Supplementary Figure 2.38** – rs214059 (chr6:25536937) in locus 46 (CARMIL1)

| Study | Cases | Controls | OR |
|---|---|---|---|
| INVENT-2019_EA | 15572 | 115280 | 1.01 |
| INVENT-2019_AA | 799 | 14353 | 0.95 |
| MVP_EA | 18952 | 411746 | 1.04 |
| MVP_AA | 5521 | 103791 | 1.04 |
| MVP_HIS | 1188 | 45634 | 1.05 |
| FinnGen_EA | 9176 | 209616 | 1.06 |
| EGP_EA | 4122 | 181553 | 1.02 |
| UKB_EA | 18385 | 91834 | 1.04 |
| UKB_AA | 280 | 1046 | 0.94 |
| UKB_SAS | 189 | 1269 | 1.08 |
| MGB_AA | 225 | 1481 | 1.05 |
| MGB_EA | 2967 | 27957 | 1.06 |
| MGB_HIS | 100 | 1352 | 0.95 |
| UPenn_EA | 380 | 6210 | 0.99 |
| UPenn_AA | 203 | 1719 | 0.93 |
| FARIVE_EA | 545 | 532 | 1.15 |
| BioME_EA | 200 | 8007 | 1.24 |
| BioME_AA | 382 | 6074 | 1.01 |
| BioME_HIS | 397 | 8992 | 1.09 |
| RETROVE_EA | 400 | 400 | 1.13 |
| GAIT2_EA | 85 | 850 | 0.9 |
| BBJ_EAS | 507 | 178366 | 1.05 |
| MESA_EA | 94 | 2341 | 1.01 |
| MESA_AA | 72 | 1511 | 0.82 |
| MESA_HIS | 35 | 1389 | 1.36 |
| MARTHA12_EA | 893 | 3414 | 1.17 |
| **Overall** | **81669** | **1426717** | **1.04** |

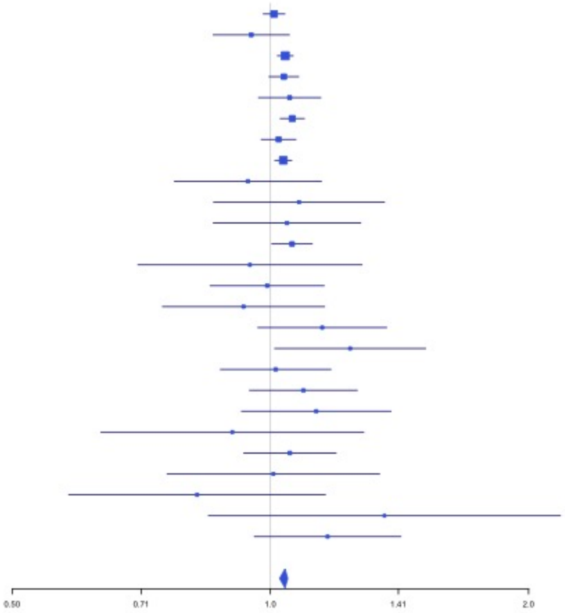

**Supplementary Figure 2.39** – rs2394251 (chr6:29943688) in locus 47 (HCG9)

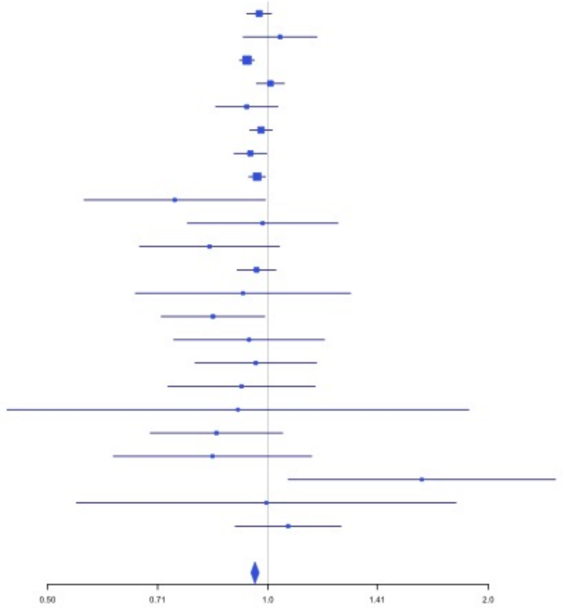


| Study | Cases | Controls | OR |
|---|---|---|---|
| INVENT-2019_EA | 15572 | 115280 | 0.97 |
| INVENT-2019_AA | 799 | 14353 | 1.04 |
| MVP_EA | 18952 | 411746 | 0.94 |
| MVP_AA | 5521 | 103791 | 1.01 |
| MVP_HIS | 1188 | 45634 | 0.94 |
| FinnGen_EA | 9176 | 209616 | 0.98 |
| EGP_EA | 4122 | 181553 | 0.95 |
| UKB_EA | 18385 | 91834 | 0.97 |
| UKB_AA | 280 | 1046 | 0.75 |
| UKB_SAS | 189 | 1269 | 0.98 |
| MGB_AA | 225 | 1481 | 0.83 |
| MGB_EA | 2967 | 27957 | 0.96 |
| MGB_HIS | 100 | 1352 | 0.92 |
| UPenn_EA | 380 | 6210 | 0.84 |
| UPenn_AA | 203 | 1719 | 0.94 |
| FARIVE_EA | 545 | 532 | 0.96 |
| RETROVE_EA | 400 | 400 | 0.92 |
| GAIT2_EA | 85 | 850 | 0.91 |
| BBJ_EAS | 507 | 178366 | 0.85 |
| MESA_EA | 94 | 2341 | 0.84 |
| MESA_AA | 72 | 1511 | 1.62 |
| MESA_HIS | 35 | 1389 | 0.99 |
| MARTHA12_EA | 893 | 3414 | 1.07 |
| **Overall** | **80690** | **1403644** | **0.96** |

**Supplementary Figure 2.40** – rs9268863 (chr6:32430289) in locus 48 (HLA-DRA ;HLA-DRB5)

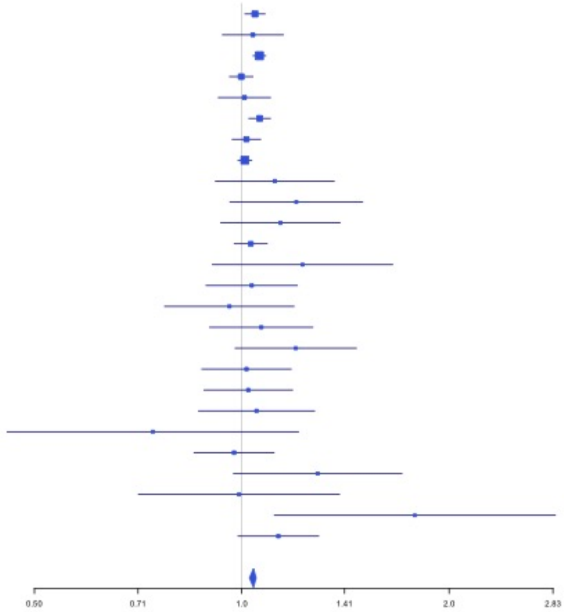


| Study | Cases | Controls | OR |
|---|---|---|---|
| INVENT-2019_EA | 15572 | 115280 | 1.05 |
| INVENT-2019_AA | 799 | 14353 | 1.04 |
| MVP_EA | 18952 | 411746 | 1.06 |
| MVP_AA | 5521 | 103791 | 1 |
| MVP_HIS | 1188 | 45634 | 1.01 |
| FinnGen_EA | 9176 | 209616 | 1.06 |
| EGP_EA | 4122 | 181553 | 1.02 |
| UKB_EA | 18385 | 91834 | 1.01 |
| UKB_AA | 280 | 1046 | 1.12 |
| UKB_SAS | 189 | 1269 | 1.2 |
| MGB_AA | 225 | 1481 | 1.14 |
| MGB_EA | 2967 | 27957 | 1.03 |
| MGB_HIS | 100 | 1352 | 1.23 |
| UPenn_EA | 380 | 6210 | 1.03 |
| UPenn_AA | 203 | 1719 | 0.96 |
| FARIVE_EA | 545 | 532 | 1.07 |
| BioME_EA | 200 | 8007 | 1.2 |
| BioME_AA | 382 | 6074 | 1.02 |
| BioME_HIS | 397 | 8992 | 1.02 |
| RETROVE_EA | 400 | 400 | 1.05 |
| GAIT2_EA | 85 | 850 | 0.74 |
| BBJ_EAS | 507 | 178366 | 0.97 |
| MESA_EA | 94 | 2341 | 1.29 |
| MESA_AA | 72 | 1511 | 0.99 |
| MESA_HIS | 35 | 1389 | 1.78 |
| MARTHA12_EA | 893 | 3414 | 1.13 |
| **Overall** | **81669** | **1426717** | **1.04** |

**Supplementary Figure 2.41** – rs145294670 (chr6:34622561) in locus 49 (ILRUN)

| Study | Cases | Controls | OR |
|---|---|---|---|
| INVENT-2019_EA | 15572 | 115280 | 1.03 |
| MVP_EA | 18952 | 411746 | 1.09 |
| MVP_AA | 5521 | 103791 | 1.17 |
| MVP_HIS | 1188 | 45634 | 1.02 |
| FinnGen_EA | 9176 | 209616 | 1.05 |
| EGP_EA | 4122 | 181553 | 1.05 |
| UKB_EA | 18385 | 91834 | 1.02 |
| MGB_AA | 225 | 1481 | 1.29 |
| MGB_EA | 2967 | 27957 | 1.07 |
| MGB_HIS | 100 | 1352 | 0.44 |
| UPenn_EA | 380 | 6210 | 1.06 |
| UPenn_AA | 203 | 1719 | 1.72 |
| FARIVE_EA | 545 | 532 | 0.93 |
| BioME_EA | 200 | 8007 | 1.17 |
| BioME_AA | 382 | 6074 | 0.95 |
| BioME_HIS | 397 | 8992 | 0.97 |
| BBJ_EAS | 507 | 178366 | 0.35 |
| MARTHA12_EA | 893 | 3414 | 0.96 |
| **Overall** | **79715** | **1403558** | **1.05** |

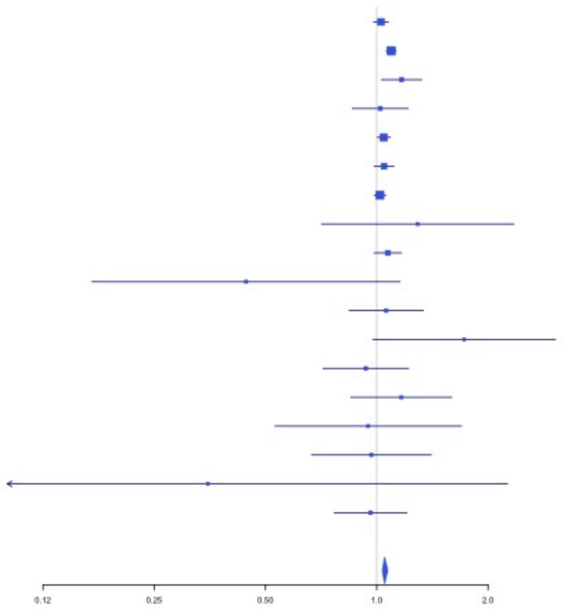


**Supplementary Figure 2.42** – rs9386182 (chr6:147691069) in locus 52 (STXBP5)

| Study | Cases | Controls | OR |
|---|---|---|---|
| INVENT-2019_EA | 15572 | 115280 | 0.94 |
| INVENT-2019_AA | 799 | 14353 | 0.91 |
| MVP_EA | 18952 | 411746 | 0.94 |
| MVP_AA | 5521 | 103791 | 0.94 |
| MVP_HIS | 1188 | 45634 | 0.91 |
| FinnGen_EA | 9176 | 209616 | 0.98 |
| UKB_EA | 18385 | 91834 | 0.94 |
| UKB_SAS | 189 | 1269 | 0.88 |
| MGB_AA | 225 | 1481 | 0.87 |
| MGB_EA | 2967 | 27957 | 0.95 |
| MGB_HIS | 100 | 1352 | 0.94 |
| UPenn_EA | 380 | 6210 | 1.03 |
| UPenn_AA | 203 | 1719 | 1.07 |
| FARIVE_EA | 545 | 532 | 1.01 |
| BioME_EA | 200 | 8007 | 1.03 |
| BioME_AA | 382 | 6074 | 0.94 |
| BioME_HIS | 397 | 8992 | 0.98 |
| BBJ_EAS | 507 | 178366 | 0.9 |
| MESA_AA | 72 | 1511 | 0.89 |
| MESA_HIS | 35 | 1389 | 0.64 |
| MARTHA12_EA | 893 | 3414 | 0.97 |
| **Overall** | **76688** | **1240527** | **0.95** |

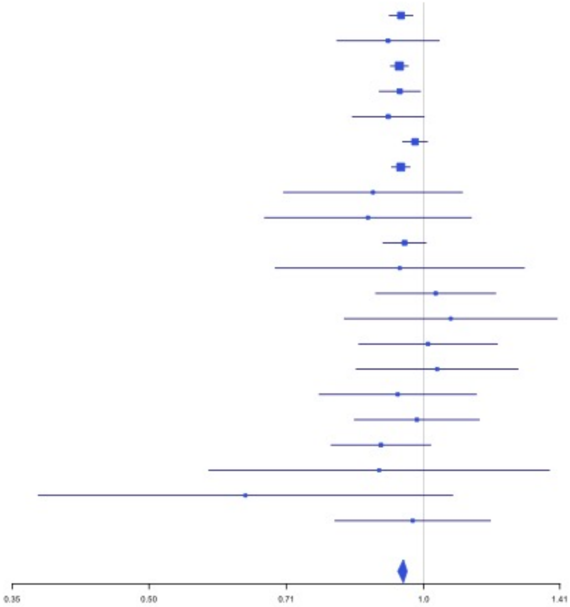

**Supplementary Figure 2.43** – rs1513275 (chr7:28259233) in locus 53 (JAZF1-AS1)

| Study | Cases | Controls | OR |
|---|---|---|---|
| INVENT-2019_EA | 15572 | 115280 | 1.05 |
| INVENT-2019_AA | 799 | 14353 | 1 |
| MVP_EA | 18952 | 411746 | 1.05 |
| MVP_AA | 5521 | 103791 | 1.03 |
| MVP_HIS | 1188 | 45634 | 1 |
| FinnGen_EA | 9176 | 209616 | 1.09 |
| EGP_EA | 4122 | 181553 | 1.01 |
| UKB_EA | 18385 | 91834 | 1.05 |
| UKB_AA | 280 | 1046 | 0.98 |
| UKB_SAS | 189 | 1269 | 0.89 |
| MGB_AA | 225 | 1481 | 0.93 |
| MGB_EA | 2967 | 27957 | 1.02 |
| MGB_HIS | 100 | 1352 | 0.76 |
| UPenn_EA | 380 | 6210 | 1.1 |
| UPenn_AA | 203 | 1719 | 1.14 |
| FARIVE_EA | 545 | 532 | 1.09 |
| BioME_EA | 200 | 8007 | 1.13 |
| BioME_AA | 382 | 6074 | 1.06 |
| BioME_HIS | 397 | 8992 | 1.03 |
| RETROVE_EA | 400 | 400 | 1.07 |
| GAIT2_EA | 85 | 850 | 1.23 |
| BBJ_EAS | 507 | 178366 | 0.93 |
| MESA_EA | 94 | 2341 | 1.3 |
| MESA_AA | 72 | 1511 | 1.08 |
| MESA_HIS | 35 | 1389 | 0.99 |
| **Overall** | **80776** | **1423303** | **1.05** |

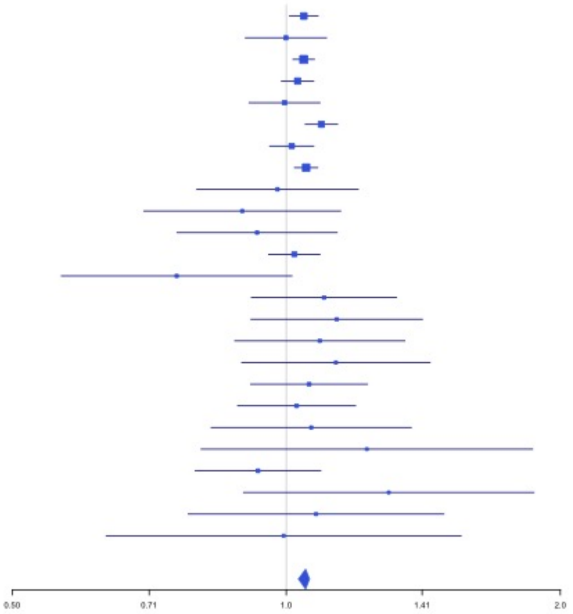


**Supplementary Figure 2.44** – rs42038 (chr7:92243719) in locus 54 (CDK6)

| Study | Cases | Controls | OR |
|---|---|---|---|
| INVENT-2019_EA | 15572 | 115280 | 1.04 |
| INVENT-2019_AA | 799 | 14353 | 1.03 |
| MVP_EA | 18952 | 411746 | 1.04 |
| MVP_AA | 5521 | 103791 | 1.02 |
| MVP_HIS | 1188 | 45634 | 1.13 |
| FinnGen_EA | 9176 | 209616 | 1.04 |
| EGP_EA | 4122 | 181553 | 1.05 |
| UKB_EA | 18385 | 91834 | 1.02 |
| UKB_AA | 280 | 1046 | 0.96 |
| MGB_AA | 225 | 1481 | 1.22 |
| MGB_EA | 2967 | 27957 | 1.06 |
| MGB_HIS | 100 | 1352 | 1.12 |
| UPenn_EA | 380 | 6210 | 1.11 |
| UPenn_AA | 203 | 1719 | 1.09 |
| FARIVE_EA | 545 | 532 | 1 |
| BioME_EA | 200 | 8007 | 1.19 |
| BioME_AA | 382 | 6074 | 0.99 |
| BioME_HIS | 397 | 8992 | 1.04 |
| RETROVE_EA | 400 | 400 | 0.92 |
| GAIT2_EA | 85 | 850 | 0.85 |
| BBJ_EAS | 507 | 178366 | 0.87 |
| MESA_EA | 94 | 2341 | 1.15 |
| MESA_AA | 72 | 1511 | 1.14 |
| MESA_HIS | 35 | 1389 | 1.03 |
| MARTHA12_EA | 893 | 3414 | 1 |
| **Overall** | **81480** | **1425448** | **1.04** |

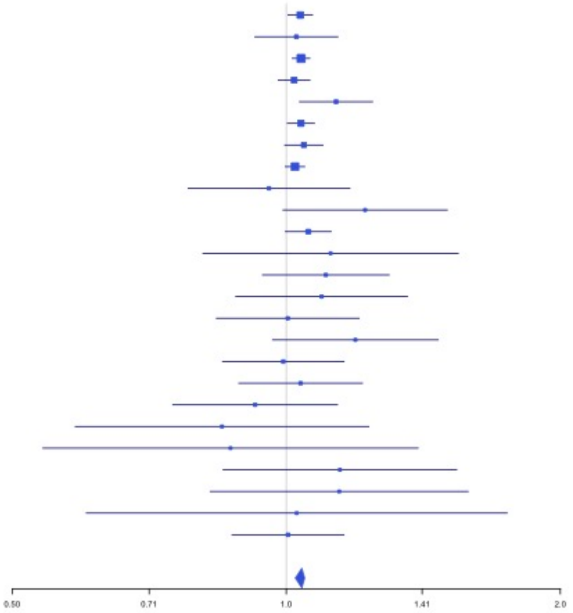

**Supplementary Figure 2.45** – rs10099512 (chr8:9178821) in locus 57 (LOC101929128 ;LOC157273)

| Study | Cases | Controls | OR |
|---|---|---|---|
| INVENT-2019_EA | 15572 | 115280 | 1.05 |
| INVENT-2019_AA | 799 | 14353 | 1.15 |
| MVP_EA | 18952 | 411746 | 1.03 |
| MVP_AA | 5521 | 103791 | 1.02 |
| MVP_HIS | 1188 | 45634 | 1.05 |
| FinnGen_EA | 9176 | 209616 | 1.06 |
| EGP_EA | 4122 | 181553 | 1.2 |
| UKB_EA | 18385 | 91834 | 1.11 |
| UKB_AA | 280 | 1046 | 1.07 |
| MGB_AA | 225 | 1481 | 0.74 |
| MGB_EA | 2967 | 27957 | 1.13 |
| MGB_HIS | 100 | 1352 | 1.19 |
| UPenn_EA | 380 | 6210 | 1.21 |
| UPenn_AA | 203 | 1719 | 0.96 |
| FARIVE_EA | 545 | 532 | 1.01 |
| BioME_EA | 200 | 8007 | 0.97 |
| BioME_AA | 382 | 6074 | 1.07 |
| BioME_HIS | 397 | 8992 | 1.16 |
| RETROVE_EA | 400 | 400 | 0.97 |
| GAIT2_EA | 85 | 850 | 0.97 |
| BBJ_EAS | 507 | 178366 | 1.22 |
| MESA_EA | 94 | 2341 | 1.13 |
| MESA_AA | 72 | 1511 | 1.39 |
| MESA_HIS | 35 | 1389 | 1.31 |
| MARTHA12_EA | 893 | 3414 | 0.94 |
| **Overall** | **81480** | **1425448** | **1.06** |

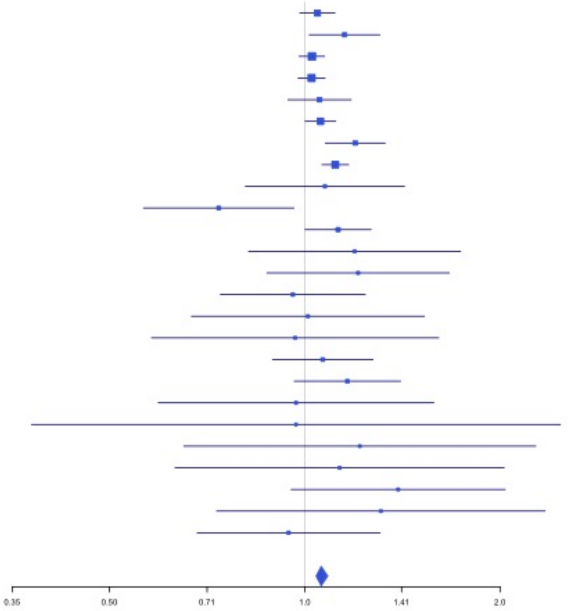


**Supplementary Figure 2.46** – rs2048528 (chr8:23373680) in locus 59 (ENTPD4 ;SLC25A37)

| Study | Cases | Controls | OR |
|---|---|---|---|
| INVENT-2019_EA | 15572 | 115280 | 0.97 |
| INVENT-2019_AA | 799 | 14353 | 0.95 |
| MVP_EA | 18952 | 411746 | 0.97 |
| MVP_AA | 5521 | 103791 | 0.98 |
| MVP_HIS | 1188 | 45634 | 0.9 |
| FinnGen_EA | 9176 | 209616 | 0.98 |
| EGP_EA | 4122 | 181553 | 0.95 |
| UKB_EA | 18385 | 91834 | 0.96 |
| UKB_AA | 280 | 1046 | 1.2 |
| UKB_SAS | 189 | 1269 | 1.13 |
| MGB_AA | 225 | 1481 | 1.01 |
| MGB_EA | 2967 | 27957 | 0.97 |
| MGB_HIS | 100 | 1352 | 0.73 |
| UPenn_EA | 380 | 6210 | 1.07 |
| UPenn_AA | 203 | 1719 | 0.99 |
| FARIVE_EA | 545 | 532 | 0.85 |
| BioME_EA | 200 | 8007 | 0.89 |
| BioME_AA | 382 | 6074 | 0.98 |
| BioME_HIS | 397 | 8992 | 0.86 |
| RETROVE_EA | 400 | 400 | 0.95 |
| GAIT2_EA | 85 | 850 | 0.88 |
| BBJ_EAS | 507 | 178366 | 0.89 |
| MESA_EA | 94 | 2341 | 0.71 |
| MESA_AA | 72 | 1511 | 1.03 |
| MESA_HIS | 35 | 1389 | 1.24 |
| **Overall** | **80776** | **1423303** | **0.97** |

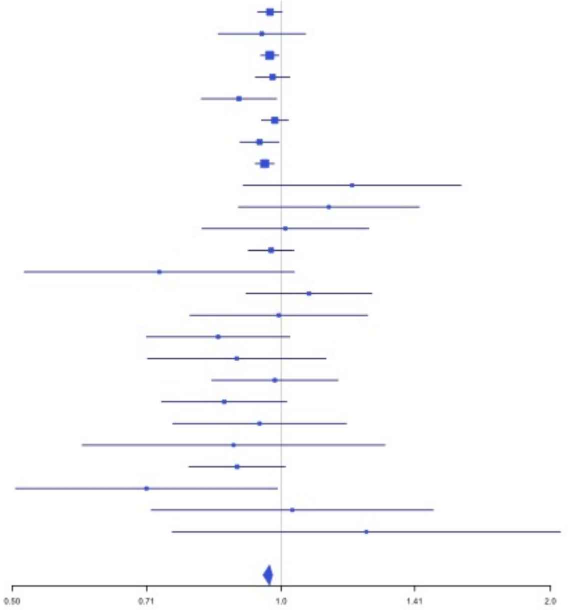

**Supplementary Figure 2.47** – rs2685412 (chr8:27810603) in locus 60 (SCARA5)

| Study | Cases | Controls | OR |
|---|---|---|---|
| INVENT-2019_EA | 15572 | 115280 | 0.97 |
| INVENT-2019_AA | 799 | 14353 | 0.95 |
| MVP_EA | 18952 | 411746 | 0.92 |
| MVP_AA | 5521 | 103791 | 0.98 |
| MVP_HIS | 1188 | 45634 | 0.92 |
| FinnGen_EA | 9176 | 209616 | 0.96 |
| EGP_EA | 4122 | 181553 | 0.91 |
| UKB_EA | 18385 | 91834 | 0.95 |
| UKB_AA | 280 | 1046 | 0.94 |
| MGB_AA | 225 | 1481 | 1.04 |
| MGB_EA | 2967 | 27957 | 0.92 |
| MGB_HIS | 100 | 1352 | 0.97 |
| UPenn_EA | 380 | 6210 | 0.93 |
| UPenn_AA | 203 | 1719 | 1.25 |
| FARIVE_EA | 545 | 532 | 0.93 |
| BioME_EA | 200 | 8007 | 0.94 |
| BioME_AA | 382 | 6074 | 0.93 |
| BioME_HIS | 397 | 8992 | 0.89 |
| RETROVE_EA | 400 | 400 | 0.87 |
| GAIT2_EA | 85 | 850 | 1.24 |
| MESA_AA | 72 | 1511 | 0.98 |
| MESA_HIS | 35 | 1389 | 0.94 |
| **Overall** | **79986** | **1241327** | **0.95** |

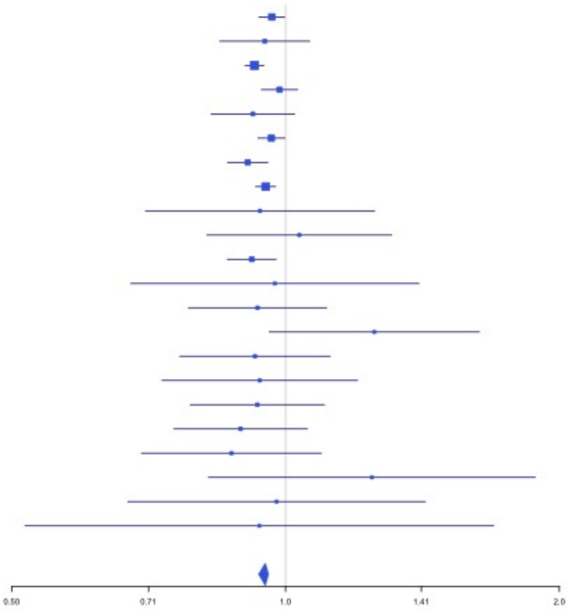


**Supplementary Figure 2.48** – rs2915595 (chr8:30402817) in locus 61 (RBPMS)

| Study | Cases | Controls | OR |
|---|---|---|---|
| INVENT-2019_EA | 15572 | 115280 | 1.03 |
| INVENT-2019_AA | 799 | 14353 | 1.03 |
| MVP_EA | 18952 | 411746 | 1.03 |
| MVP_AA | 5521 | 103791 | 1.02 |
| MVP_HIS | 1188 | 45634 | 1.1 |
| FinnGen_EA | 9176 | 209616 | 1.05 |
| EGP_EA | 4122 | 181553 | 1.02 |
| UKB_EA | 18385 | 91834 | 1.04 |
| UKB_SAS | 189 | 1269 | 0.89 |
| MGB_AA | 225 | 1481 | 1.44 |
| MGB_EA | 2967 | 27957 | 1.11 |
| MGB_HIS | 100 | 1352 | 0.9 |
| UPenn_EA | 380 | 6210 | 1.16 |
| UPenn_AA | 203 | 1719 | 1.11 |
| FARIVE_EA | 545 | 532 | 1.02 |
| BioME_EA | 200 | 8007 | 0.9 |
| BioME_AA | 382 | 6074 | 1.12 |
| BioME_HIS | 397 | 8992 | 1.04 |
| RETROVE_EA | 400 | 400 | 1.05 |
| GAIT2_EA | 85 | 850 | 1.15 |
| BBJ_EAS | 507 | 178366 | 0.97 |
| MESA_EA | 94 | 2341 | 1.16 |
| MESA_AA | 72 | 1511 | 0.55 |
| MESA_HIS | 35 | 1389 | 0.42 |
| **Overall** | **80496** | **1422257** | **1.04** |

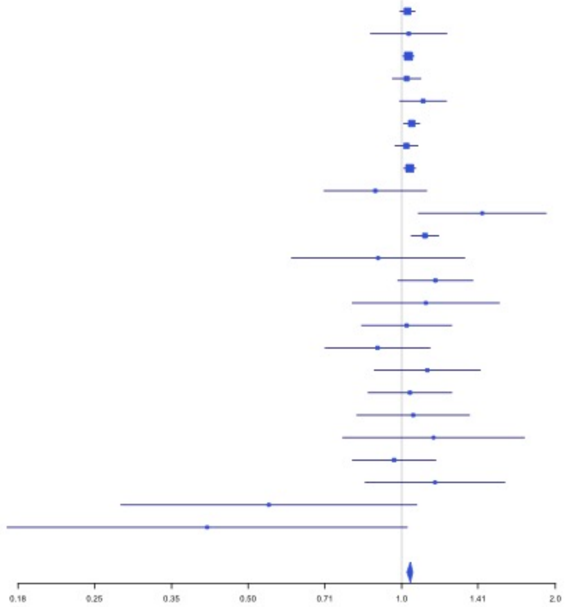

**Supplementary Figure 2.49** – rs6993770 (chr8:106581528) in locus 62 (ZFPM2)

| Study | Cases | Controls | OR |
|---|---|---|---|
| INVENT-2019_EA | 15572 | 115280 | 1.07 |
| INVENT-2019_AA | 799 | 14353 | 1.08 |
| MVP_EA | 18952 | 411746 | 1.1 |
| MVP_AA | 5521 | 103791 | 1.1 |
| MVP_HIS | 1188 | 45634 | 1.06 |
| FinnGen_EA | 9176 | 209616 | 1.05 |
| EGP_EA | 4122 | 181553 | 1.05 |
| UKB_EA | 18385 | 91834 | 1.08 |
| UKB_AA | 280 | 1046 | 0.95 |
| UKB_SAS | 189 | 1269 | 1 |
| MGB_AA | 225 | 1481 | 1.06 |
| MGB_EA | 2967 | 27957 | 1.05 |
| MGB_HIS | 100 | 1352 | 1.04 |
| UPenn_EA | 380 | 6210 | 0.99 |
| UPenn_AA | 203 | 1719 | 1.1 |
| FARIVE_EA | 545 | 532 | 1.08 |
| BioME_EA | 200 | 8007 | 1.09 |
| BioME_AA | 382 | 6074 | 1.07 |
| BioME_HIS | 397 | 8992 | 1.2 |
| RETROVE_EA | 400 | 400 | 1.35 |
| GAIT2_EA | 85 | 850 | 1.09 |
| BBJ_EAS | 507 | 178366 | 1.09 |
| MESA_EA | 94 | 2341 | 0.98 |
| MESA_AA | 72 | 1511 | 1.19 |
| MESA_HIS | 35 | 1389 | 0.94 |
| MARTHA12_EA | 893 | 3414 | 1 |
| **Overall** | **81669** | **1426717** | **1.08** |

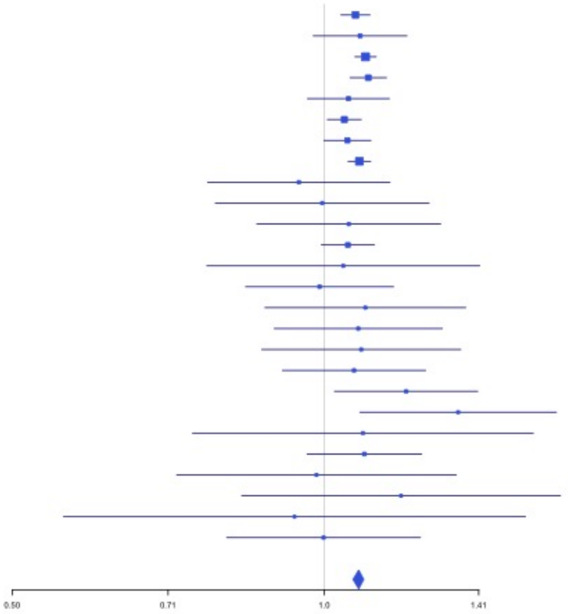


**Supplementary Figure 2.50** – rs4236786 (chr8:108291878) in locus 63 (ANGPT1)

| Study | Cases | Controls | OR |
|---|---|---|---|
| INVENT-2019_EA | 15572 | 115280 | 1.04 |
| INVENT-2019_AA | 799 | 14353 | 1.01 |
| MVP_EA | 18952 | 411746 | 1.07 |
| MVP_AA | 5521 | 103791 | 0.99 |
| MVP_HIS | 1188 | 45634 | 0.95 |
| FinnGen_EA | 9176 | 209616 | 1.01 |
| EGP_EA | 4122 | 181553 | 1.02 |
| UKB_EA | 18385 | 91834 | 1.04 |
| UKB_SAS | 189 | 1269 | 0.96 |
| MGB_AA | 225 | 1481 | 0.89 |
| MGB_EA | 2967 | 27957 | 1.02 |
| MGB_HIS | 100 | 1352 | 0.99 |
| UPenn_EA | 380 | 6210 | 0.93 |
| UPenn_AA | 203 | 1719 | 1.33 |
| FARIVE_EA | 545 | 532 | 1.12 |
| BioME_EA | 200 | 8007 | 1 |
| BioME_AA | 382 | 6074 | 0.99 |
| BioME_HIS | 397 | 8992 | 1.04 |
| BBJ_EAS | 507 | 178366 | 1.08 |
| MESA_AA | 72 | 1511 | 1.08 |
| MESA_HIS | 35 | 1389 | 1.24 |
| **Overall** | **79917** | **1418666** | **1.04** |

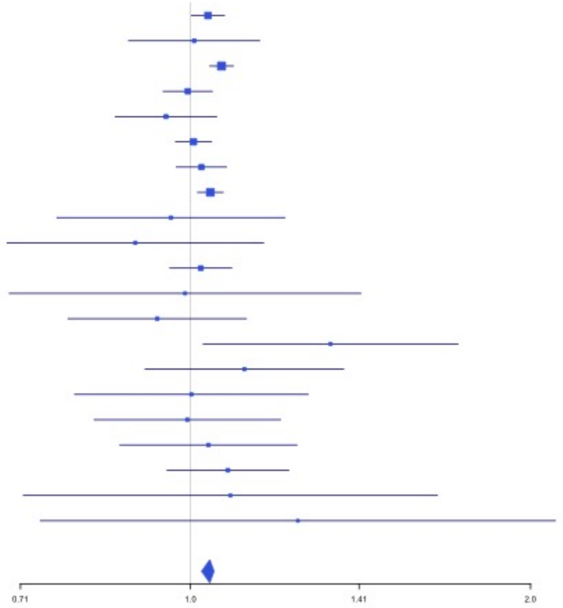

**Supplementary Figure 2.51** – rs10820606 (chr9:99192919) in locus 65 (ZNF367;HABP4)

| Study | Cases | Controls | OR |
|---|---|---|---|
| INVENT-2019_EA | 15572 | 115280 | 1.03 |
| INVENT-2019_AA | 799 | 14353 | 0.97 |
| MVP_EA | 18952 | 411746 | 1.07 |
| MVP_AA | 5521 | 103791 | 1.07 |
| MVP_HIS | 1188 | 45634 | 1.09 |
| UKB_EA | 18385 | 91834 | 1.03 |
| UKB_AA | 280 | 1046 | 1.09 |
| MGB_AA | 225 | 1481 | 1.1 |
| MGB_EA | 2967 | 27957 | 1.1 |
| MGB_HIS | 100 | 1352 | 0.89 |
| UPenn_EA | 380 | 6210 | 1.1 |
| UPenn_AA | 203 | 1719 | 0.64 |
| FARIVE_EA | 545 | 532 | 1.16 |
| BioME_EA | 200 | 8007 | 1.32 |
| BioME_AA | 382 | 6074 | 1.07 |
| BioME_HIS | 397 | 8992 | 1.18 |
| MESA_AA | 72 | 1511 | 0.84 |
| MESA_HIS | 35 | 1389 | 1.14 |
| **Overall** | **66203** | **848908** | **1.05** |

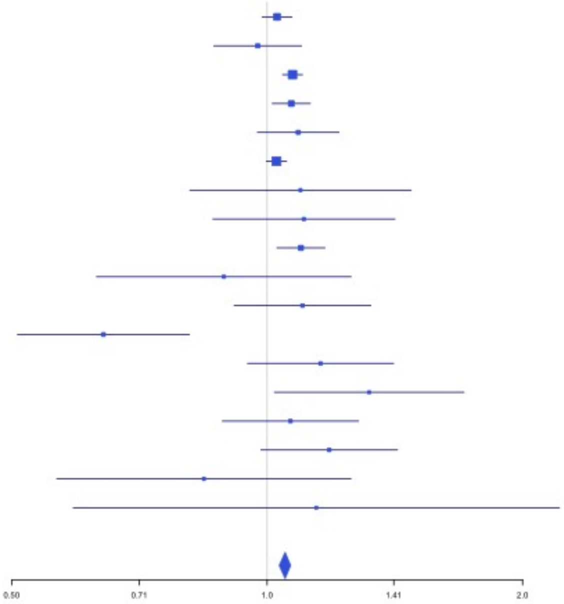


**Supplementary Figure 2.52** – rs505922 (chr9:136149229) in locus 66 (ABO)

| Study | Cases | Controls | OR |
|---|---|---|---|
| INVENT-2019_EA | 15572 | 115280 | 0.74 |
| INVENT-2019_AA | 799 | 14353 | 0.82 |
| MVP_EA | 18952 | 411746 | 0.71 |
| MVP_AA | 5521 | 103791 | 0.75 |
| MVP_HIS | 1188 | 45634 | 0.7 |
| FinnGen_EA | 9176 | 209616 | 0.78 |
| EGP_EA | 4122 | 181553 | 0.82 |
| UKB_EA | 18385 | 91834 | 0.76 |
| UKB_AA | 280 | 1046 | 0.93 |
| UKB_SAS | 189 | 1269 | 0.72 |
| MGB_AA | 225 | 1481 | 0.86 |
| MGB_EA | 2967 | 27957 | 0.82 |
| MGB_HIS | 100 | 1352 | 0.9 |
| UPenn_EA | 380 | 6210 | 0.77 |
| UPenn_AA | 203 | 1719 | 0.86 |
| FARIVE_EA | 545 | 532 | 0.57 |
| BioME_EA | 200 | 8007 | 0.79 |
| BioME_AA | 382 | 6074 | 0.82 |
| BioME_HIS | 397 | 8992 | 0.85 |
| RETROVE_EA | 400 | 400 | 0.65 |
| GAIT2_EA | 85 | 850 | 0.54 |
| BBJ_EAS | 507 | 178366 | 0.87 |
| MESA_EA | 94 | 2341 | 0.83 |
| MESA_AA | 72 | 1511 | 1.04 |
| MESA_HIS | 35 | 1389 | 0.86 |
| MARTHA12_EA | 893 | 3414 | 0.57 |
| **Overall** | **81669** | **1426717** | **0.75** |

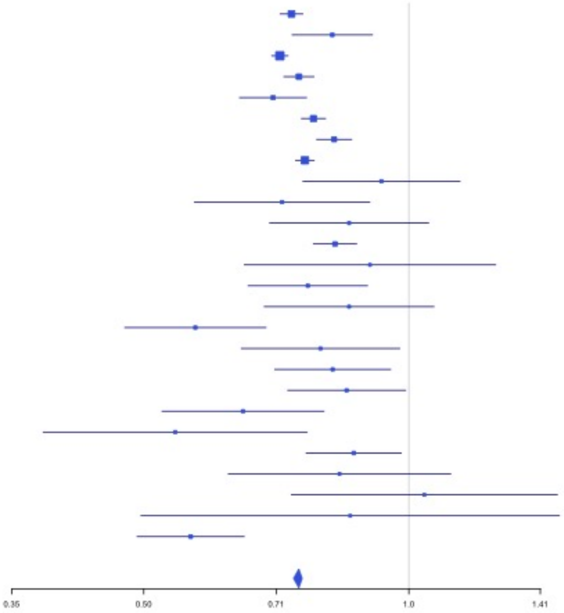

**Supplementary Figure 2.53** – rs1243187 (chr10:21907016) in locus 68 (MLLT10)

| Study | Cases | Controls | OR |
|---|---|---|---|
| INVENT-2019_EA | 15572 | 115280 | 0.96 |
| INVENT-2019_AA | 799 | 14353 | 1.01 |
| MVP_EA | 18952 | 411746 | 0.97 |
| MVP_AA | 5521 | 103791 | 0.99 |
| MVP_HIS | 1188 | 45634 | 1.03 |
| FinnGen_EA | 9176 | 209616 | 0.98 |
| EGP_EA | 4122 | 181553 | 0.98 |
| UKB_EA | 18385 | 91834 | 0.95 |
| UKB_SAS | 189 | 1269 | 0.88 |
| MGB_AA | 225 | 1481 | 0.61 |
| MGB_EA | 2967 | 27957 | 0.94 |
| MGB_HIS | 100 | 1352 | 1.28 |
| UPenn_EA | 380 | 6210 | 0.97 |
| UPenn_AA | 203 | 1719 | 0.83 |
| FARIVE_EA | 545 | 532 | 0.96 |
| BioME_EA | 200 | 8007 | 0.91 |
| BioME_AA | 382 | 6074 | 1.08 |
| BioME_HIS | 397 | 8992 | 0.98 |
| RETROVE_EA | 400 | 400 | 1.1 |
| GAIT2_EA | 85 | 850 | 1.3 |
| BBJ_EAS | 507 | 178366 | 1.27 |
| MESA_EA | 94 | 2341 | 1.15 |
| MESA_AA | 72 | 1511 | 0.64 |
| MESA_HIS | 35 | 1389 | 1.27 |
| **Overall** | **80496** | **1422257** | **0.97** |

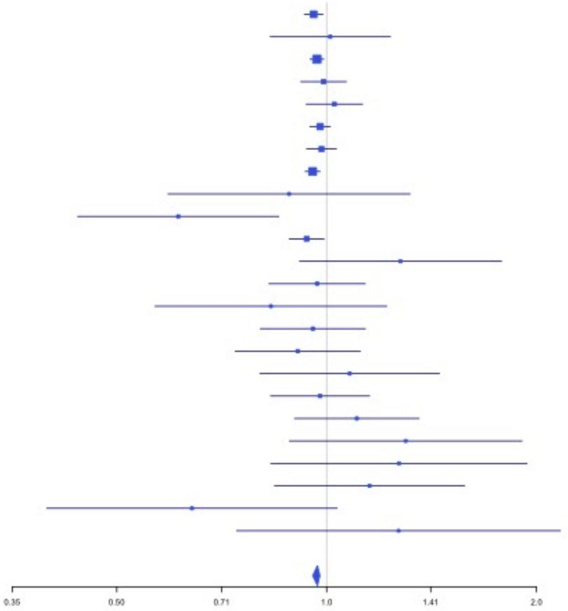


**Supplementary Figure 2.54** – rs4272700 (chr10:27881308) in locus 69 (RAB18;MKX)

| Study | Cases | Controls | OR |
|---|---|---|---|
| INVENT-2019_EA | 15572 | 115280 | 1.02 |
| INVENT-2019_AA | 799 | 14353 | 1.03 |
| MVP_EA | 18952 | 411746 | 1.03 |
| MVP_AA | 5521 | 103791 | 1.19 |
| MVP_HIS | 1188 | 45634 | 1.11 |
| FinnGen_EA | 9176 | 209616 | 1.05 |
| EGP_EA | 4122 | 181553 | 1.07 |
| UKB_EA | 18385 | 91834 | 1.02 |
| UKB_SAS | 189 | 1269 | 0.94 |
| MGB_AA | 225 | 1481 | 1.47 |
| MGB_EA | 2967 | 27957 | 1.06 |
| MGB_HIS | 100 | 1352 | 0.8 |
| UPenn_EA | 380 | 6210 | 1.03 |
| UPenn_AA | 203 | 1719 | 1.3 |
| FARIVE_EA | 545 | 532 | 1.26 |
| BioME_EA | 200 | 8007 | 1.1 |
| BioME_AA | 382 | 6074 | 1.28 |
| BioME_HIS | 397 | 8992 | 1.11 |
| RETROVE_EA | 400 | 400 | 1.26 |
| GAIT2_EA | 85 | 850 | 0.8 |
| BBJ_EAS | 507 | 178366 | 1 |
| MESA_EA | 94 | 2341 | 0.96 |
| MESA_AA | 72 | 1511 | 1.46 |
| MESA_HIS | 35 | 1389 | 1.5 |
| **Overall** | **80496** | **1422257** | **1.04** |

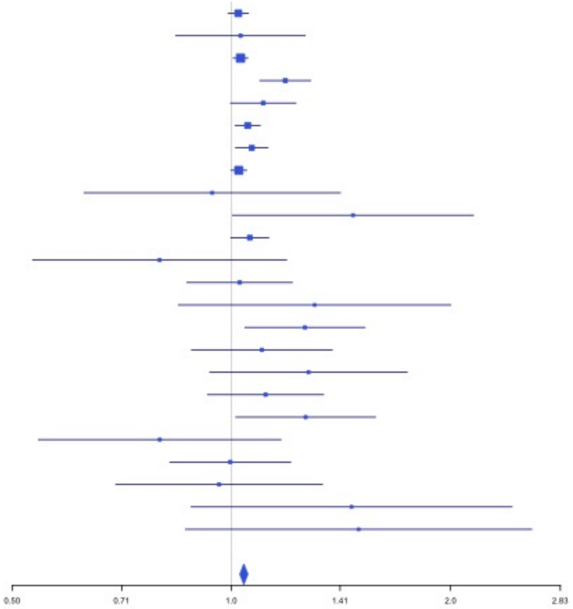

**Supplementary Figure 2.55** – rs17490626 (chr10:71218646) in locus 72 (TSPAN15)

| Study | Cases | Controls | OR |
|---|---|---|---|
| INVENT-2019_EA | 15572 | 115280 | 0.85 |
| INVENT-2019_AA | 799 | 14353 | 0.84 |
| MVP_EA | 18952 | 411746 | 0.75 |
| MVP_AA | 5521 | 103791 | 0.82 |
| MVP_HIS | 1188 | 45634 | 0.86 |
| FinnGen_EA | 9176 | 209616 | 0.83 |
| EGP_EA | 4122 | 181553 | 0.83 |
| UKB_EA | 18385 | 91834 | 0.82 |
| MGB_AA | 225 | 1481 | 0.99 |
| MGB_EA | 2967 | 27957 | 0.83 |
| MGB_HIS | 100 | 1352 | 0.77 |
| UPenn_EA | 380 | 6210 | 0.98 |
| UPenn_AA | 203 | 1719 | 1.07 |
| FARIVE_EA | 545 | 532 | 0.85 |
| BioME_EA | 200 | 8007 | 0.67 |
| BioME_AA | 382 | 6074 | 0.54 |
| BioME_HIS | 397 | 8992 | 0.81 |
| RETROVE_EA | 400 | 400 | 0.72 |
| GAIT2_EA | 85 | 850 | 1.13 |
| BBJ_EAS | 507 | 178366 | 0.39 |
| MESA_EA | 94 | 2341 | 0.93 |
| MESA_AA | 72 | 1511 | 0.33 |
| MESA_HIS | 35 | 1389 | 1.07 |
| **Overall** | **80307** | **1420988** | **0.81** |

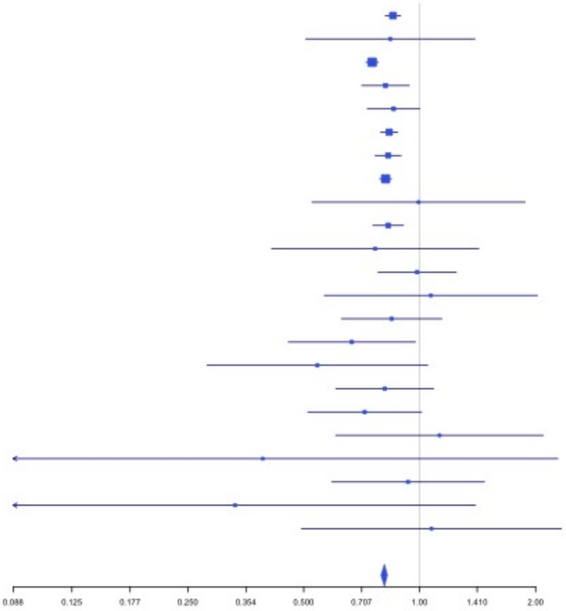


**Supplementary Figure 2.56** – rs16937003 (chr10:80938499) in locus 73 (ZMIZ1)

| Study | Cases | Controls | OR |
|---|---|---|---|
| INVENT-2019_EA | 15572 | 115280 | 1.15 |
| INVENT-2019_AA | 799 | 14353 | 1.03 |
| MVP_EA | 18952 | 411746 | 1.13 |
| MVP_AA | 5521 | 103791 | 1.24 |
| MVP_HIS | 1188 | 45634 | 1.04 |
| FinnGen_EA | 9176 | 209616 | 1.18 |
| EGP_EA | 4122 | 181553 | 1.11 |
| UKB_EA | 18385 | 91834 | 1.06 |
| MGB_AA | 225 | 1481 | 1.17 |
| MGB_EA | 2967 | 27957 | 1.08 |
| MGB_HIS | 100 | 1352 | 0.75 |
| UPenn_EA | 380 | 6210 | 0.76 |
| UPenn_AA | 203 | 1719 | 1.14 |
| FARIVE_EA | 545 | 532 | 1.8 |
| BioME_EA | 200 | 8007 | 1.13 |
| BioME_AA | 382 | 6074 | 0.78 |
| BioME_HIS | 397 | 8992 | 1.21 |
| RETROVE_EA | 400 | 400 | 1 |
| GAIT2_EA | 85 | 850 | 1.74 |
| BBJ_EAS | 507 | 178366 | 1.07 |
| MESA_EA | 94 | 2341 | 0.36 |
| MESA_AA | 72 | 1511 | 2.25 |
| MESA_HIS | 35 | 1389 | 0.38 |
| **Overall** | **80307** | **1420988** | **1.12** |

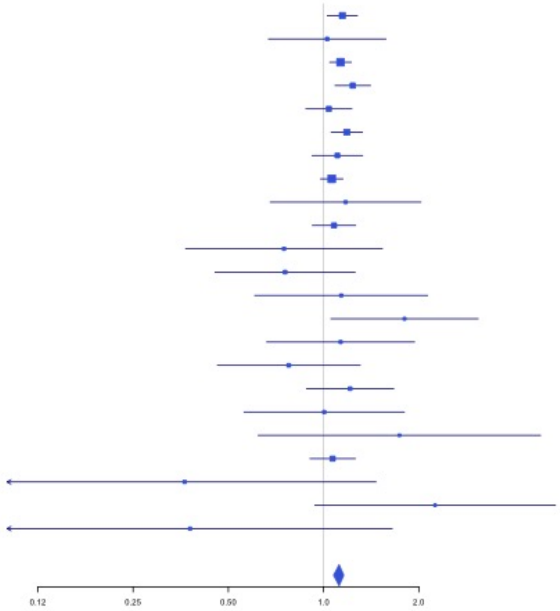

**Supplementary Figure 2.57** – rs57866767 (chr10:96023077) in locus 74 (PLCE1)

| Study | Cases | Controls | OR |
|---|---|---|---|
| INVENT-2019_EA | 15572 | 115280 | 0.97 |
| INVENT-2019_AA | 799 | 14353 | 0.86 |
| MVP_EA | 18952 | 411746 | 0.96 |
| MVP_AA | 5521 | 103791 | 0.95 |
| MVP_HIS | 1188 | 45634 | 1.02 |
| FinnGen_EA | 9176 | 209616 | 0.96 |
| EGP_EA | 4122 | 181553 | 0.96 |
| UKB_EA | 18385 | 91834 | 0.96 |
| UKB_AA | 280 | 1046 | 1.04 |
| UKB_SAS | 189 | 1269 | 1 |
| MGB_AA | 225 | 1481 | 0.97 |
| MGB_EA | 2967 | 27957 | 0.96 |
| MGB_HIS | 100 | 1352 | 1.11 |
| UPenn_EA | 380 | 6210 | 0.92 |
| UPenn_AA | 203 | 1719 | 0.99 |
| FARIVE_EA | 545 | 532 | 0.91 |
| BioME_EA | 200 | 8007 | 0.85 |
| BioME_AA | 382 | 6074 | 0.96 |
| BioME_HIS | 397 | 8992 | 0.85 |
| RETROVE_EA | 400 | 400 | 1.03 |
| GAIT2_EA | 85 | 850 | 0.91 |
| BBJ_EAS | 507 | 178366 | 0.9 |
| MESA_EA | 94 | 2341 | 0.91 |
| MESA_AA | 72 | 1511 | 1.18 |
| MESA_HIS | 35 | 1389 | 1.01 |
| MARTHA12_EA | 893 | 3414 | 0.95 |
| **Overall** | **81669** | **1426717** | **0.96** |

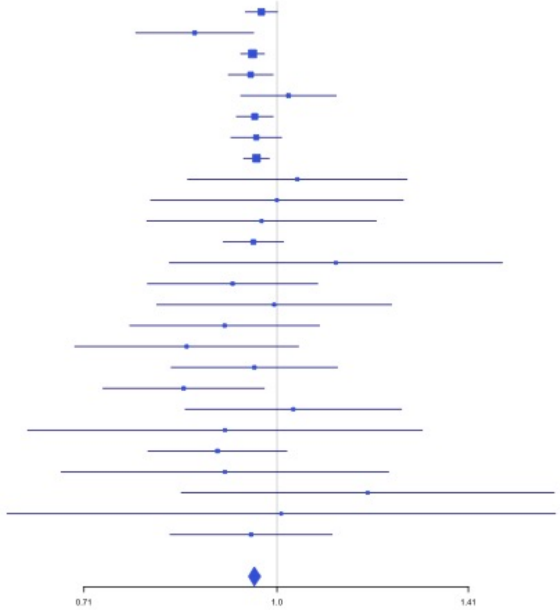


**Supplementary Figure 2.58** – rs10886430 (chr10:121010256) in locus 75 (GRK5)

| Study | Cases | Controls | OR |
|---|---|---|---|
| INVENT-2019_EA | 15572 | 115280 | 0.9 |
| INVENT-2019_AA | 799 | 14353 | 1.43 |
| MVP_EA | 18952 | 411746 | 0.86 |
| MVP_AA | 5521 | 103791 | 0.89 |
| MVP_HIS | 1188 | 45634 | 0.83 |
| FinnGen_EA | 9176 | 209616 | 0.91 |
| EGP_EA | 4122 | 181553 | 0.92 |
| UKB_EA | 18385 | 91834 | 0.89 |
| MGB_AA | 225 | 1481 | 1.11 |
| MGB_EA | 2967 | 27957 | 0.93 |
| MGB_HIS | 100 | 1352 | 1.21 |
| UPenn_EA | 380 | 6210 | 0.95 |
| UPenn_AA | 203 | 1719 | 0.66 |
| FARIVE_EA | 545 | 532 | 1.04 |
| BioME_EA | 200 | 8007 | 0.76 |
| BioME_AA | 382 | 6074 | 1.16 |
| BioME_HIS | 397 | 8992 | 0.92 |
| RETROVE_EA | 400 | 400 | 0.88 |
| GAIT2_EA | 85 | 850 | 0.88 |
| BBJ_EAS | 507 | 178366 | 3.05 |
| MESA_EA | 94 | 2341 | 0.75 |
| MESA_AA | 72 | 1511 | 0.33 |
| MESA_HIS | 35 | 1389 | 1.71 |
| **Overall** | **80307** | **1420988** | **0.89** |

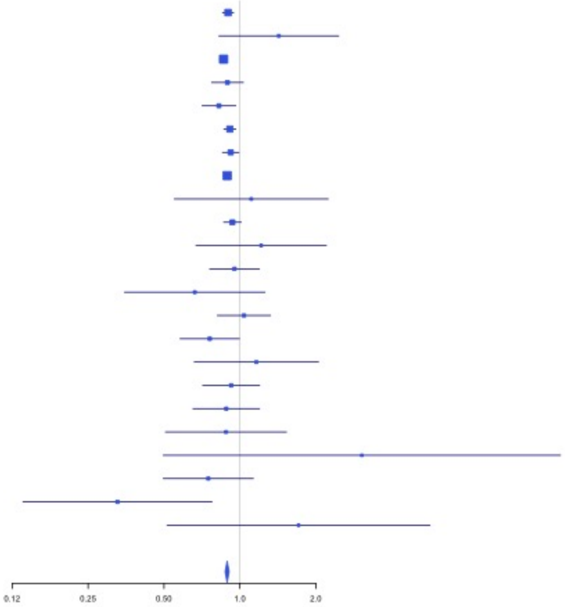

**Supplementary Figure 2.59** – rs2030291 (chr11:16251251) in locus 77 (SOX6)

| Study | Cases | Controls | OR |
|---|---|---|---|
| INVENT-2019_EA | 15572 | 115280 | 0.96 |
| INVENT-2019_AA | 799 | 14353 | 0.95 |
| MVP_EA | 18952 | 411746 | 0.98 |
| MVP_AA | 5521 | 103791 | 0.99 |
| MVP_HIS | 1188 | 45634 | 0.94 |
| FinnGen_EA | 9176 | 209616 | 0.93 |
| EGP_EA | 4122 | 181553 | 0.94 |
| UKB_EA | 18385 | 91834 | 0.98 |
| UKB_AA | 280 | 1046 | 0.97 |
| UKB_SAS | 189 | 1269 | 0.98 |
| MGB_AA | 225 | 1481 | 1.19 |
| MGB_EA | 2967 | 27957 | 0.97 |
| MGB_HIS | 100 | 1352 | 0.98 |
| UPenn_EA | 380 | 6210 | 0.83 |
| UPenn_AA | 203 | 1719 | 0.79 |
| FARIVE_EA | 545 | 532 | 0.93 |
| BioME_EA | 200 | 8007 | 0.89 |
| BioME_AA | 382 | 6074 | 1.01 |
| BioME_HIS | 397 | 8992 | 1.05 |
| RETROVE_EA | 400 | 400 | 1.11 |
| GAIT2_EA | 85 | 850 | 0.95 |
| BBJ_EAS | 507 | 178366 | 0.94 |
| MESA_EA | 94 | 2341 | 0.94 |
| MESA_AA | 72 | 1511 | 0.89 |
| MESA_HIS | 35 | 1389 | 0.74 |
| MARTHA12_EA | 893 | 3414 | 0.89 |
| **Overall** | **81669** | **1426717** | **0.97** |

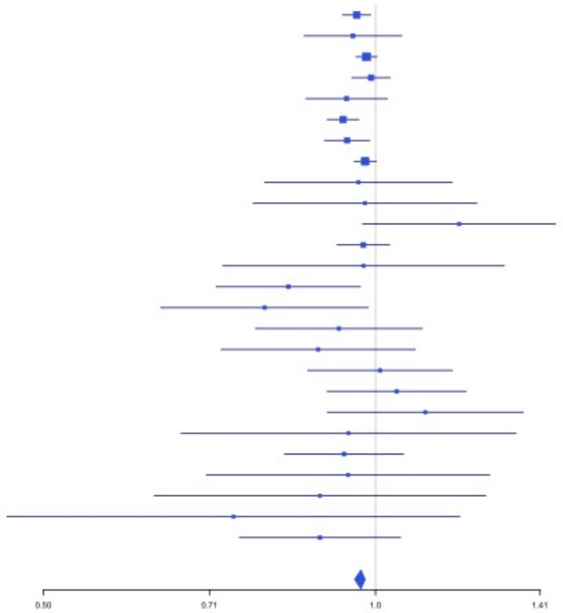


**Supplementary Figure 2.60** – rs11032074 (chr11:32993887) in locus 78 (QSER1)

| Study | Cases | Controls | OR |
|---|---|---|---|
| INVENT-2019_EA | 15572 | 115280 | 1.06 |
| INVENT-2019_AA | 799 | 14353 | 0.95 |
| MVP_EA | 18952 | 411746 | 1.05 |
| MVP_AA | 5521 | 103791 | 1.04 |
| MVP_HIS | 1188 | 45634 | 1.07 |
| FinnGen_EA | 9176 | 209616 | 1.06 |
| EGP_EA | 4122 | 181553 | 1.04 |
| UKB_EA | 18385 | 91834 | 1.04 |
| UKB_AA | 280 | 1046 | 1.22 |
| MGB_AA | 225 | 1481 | 0.82 |
| MGB_EA | 2967 | 27957 | 1.01 |
| MGB_HIS | 100 | 1352 | 0.96 |
| UPenn_EA | 380 | 6210 | 0.89 |
| UPenn_AA | 203 | 1719 | 1.04 |
| FARIVE_EA | 545 | 532 | 0.97 |
| BioME_EA | 200 | 8007 | 1.01 |
| BioME_AA | 382 | 6074 | 1.18 |
| BioME_HIS | 397 | 8992 | 0.99 |
| RETROVE_EA | 400 | 400 | 1.1 |
| GAIT2_EA | 85 | 850 | 0.86 |
| BBJ_EAS | 507 | 178366 | 1.22 |
| MESA_EA | 94 | 2341 | 0.83 |
| MESA_AA | 72 | 1511 | 0.78 |
| MESA_HIS | 35 | 1389 | 1.23 |
| **Overall** | **80587** | **1422034** | **1.05** |

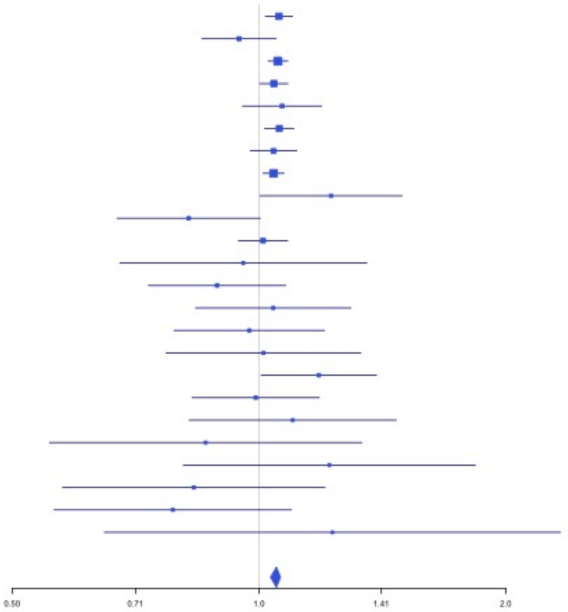

**Supplementary Figure 2.61** – rs1799963 (chr11:46761055) in locus 79 (F2)

| Study | Cases | Controls | OR |
|---|---|---|---|
| INVENT-2019_EA | 15572 | 115280 | 1.6 |
| MVP_EA | 18952 | 411746 | 2.08 |
| MVP_AA | 5521 | 103791 | 1.73 |
| MVP_HIS | 1188 | 45634 | 1.58 |
| FinnGen_EA | 9176 | 209616 | 3.42 |
| EGP_EA | 4122 | 181553 | 1.94 |
| UKB_EA | 18385 | 91834 | 1.93 |
| MGB_EA | 2967 | 27957 | 1.42 |
| MGB_HIS | 100 | 1352 | 1.55 |
| UPenn_EA | 380 | 6210 | 0.94 |
| UPenn_AA | 203 | 1719 | 2.52 |
| FARIVE_EA | 545 | 532 | 3.09 |
| BioME_EA | 200 | 8007 | 1.4 |
| BioME_AA | 382 | 6074 | 0.58 |
| BioME_HIS | 397 | 8992 | 1.76 |
| RETROVE_EA | 400 | 400 | 1.49 |
| GAIT2_EA | 85 | 850 | 2.76 |
| BBJ_EAS | 507 | 178366 | 0.44 |
| MESA_EA | 94 | 2341 | 1.22 |
| MARTHA12_EA | 893 | 3414 | 3.82 |
| **Overall** | **80069** | **1405668** | **1.97** |

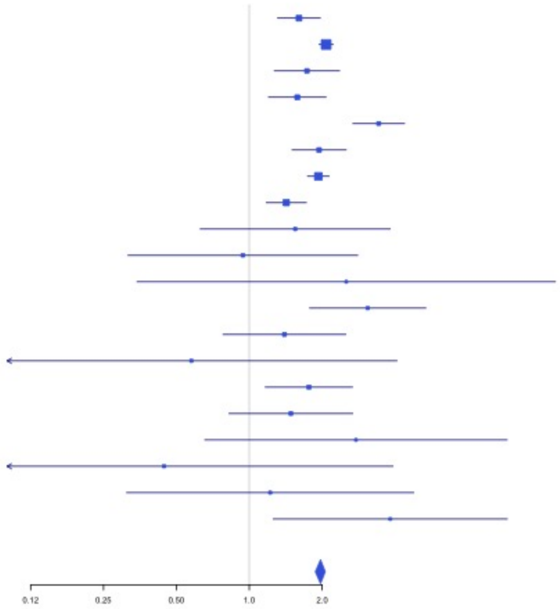


**Supplementary Figure 2.62** – rs141798115 (chr11:56875074) in locus 80 (OR5AK4P ;LRRC55)

| Study | Cases | Controls | OR |
|---|---|---|---|
| INVENT-2019_EA | 15572 | 115280 | 1.09 |
| MVP_EA | 18952 | 411746 | 1.37 |
| MVP_AA | 5521 | 103791 | 1.01 |
| MVP_HIS | 1188 | 45634 | 0.79 |
| FinnGen_EA | 9176 | 209616 | 1.4 |
| EGP_EA | 4122 | 181553 | 1.37 |
| UKB_EA | 18385 | 91834 | 1.19 |
| MGB_EA | 2967 | 27957 | 1.12 |
| MGB_HIS | 100 | 1352 | 1.39 |
| UPenn_EA | 380 | 6210 | 1.04 |
| UPenn_AA | 203 | 1719 | 1.12 |
| FARIVE_EA | 545 | 532 | 1.7 |
| BioME_EA | 200 | 8007 | 0.54 |
| BioME_AA | 382 | 6074 | 0.63 |
| BioME_HIS | 397 | 8992 | 1.33 |
| RETROVE_EA | 400 | 400 | 0.57 |
| GAIT2_EA | 85 | 850 | 1.33 |
| MESA_EA | 94 | 2341 | 0.42 |
| MARTHA12_EA | 893 | 3414 | 0.99 |
| **Overall** | **79562** | **1227302** | **1.26** |

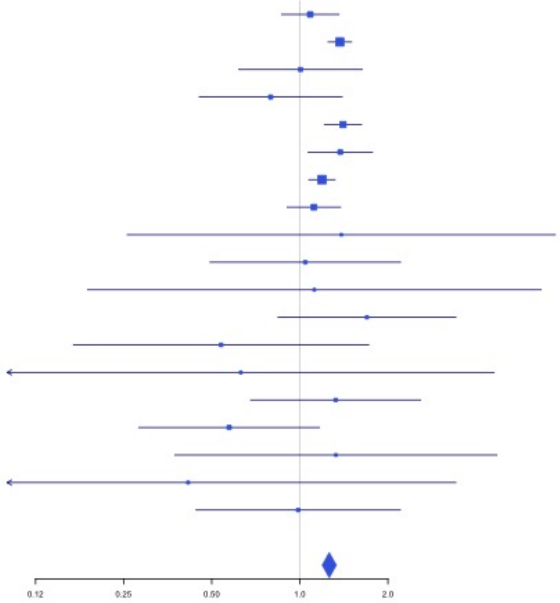

**Supplementary Figure 2.63** – rs4354705 (chr11:60088159) in locus 81 (MS4A4A ;MS4A6E)

| Study | Cases | Controls | OR |
|---|---|---|---|
| INVENT-2019_EA | 15572 | 115280 | 1.02 |
| INVENT-2019_AA | 799 | 14353 | 1.03 |
| MVP_EA | 18952 | 411746 | 1.04 |
| MVP_AA | 5521 | 103791 | 1.02 |
| MVP_HIS | 1188 | 45634 | 1.02 |
| FinnGen_EA | 9176 | 209616 | 1.01 |
| EGP_EA | 4122 | 181553 | 1.05 |
| UKB_EA | 18385 | 91834 | 1.03 |
| UKB_AA | 280 | 1046 | 1.1 |
| UKB_SAS | 189 | 1269 | 1.03 |
| MGB_AA | 225 | 1481 | 0.97 |
| MGB_EA | 2967 | 27957 | 1.07 |
| MGB_HIS | 100 | 1352 | 1 |
| UPenn_EA | 380 | 6210 | 1.02 |
| UPenn_AA | 203 | 1719 | 1.14 |
| FARIVE_EA | 545 | 532 | 0.98 |
| BioME_EA | 200 | 8007 | 0.86 |
| BioME_AA | 382 | 6074 | 1.09 |
| BioME_HIS | 397 | 8992 | 0.96 |
| RETROVE_EA | 400 | 400 | 1.13 |
| GAIT2_EA | 85 | 850 | 1.06 |
| BBJ_EAS | 507 | 178366 | 1.07 |
| MESA_EA | 94 | 2341 | 1.13 |
| MESA_AA | 72 | 1511 | 0.57 |
| MESA_HIS | 35 | 1389 | 0.65 |
| MARTHA12_EA | 893 | 3414 | 1.09 |
| **Overall** | **81669** | **1426717** | **1.03** |

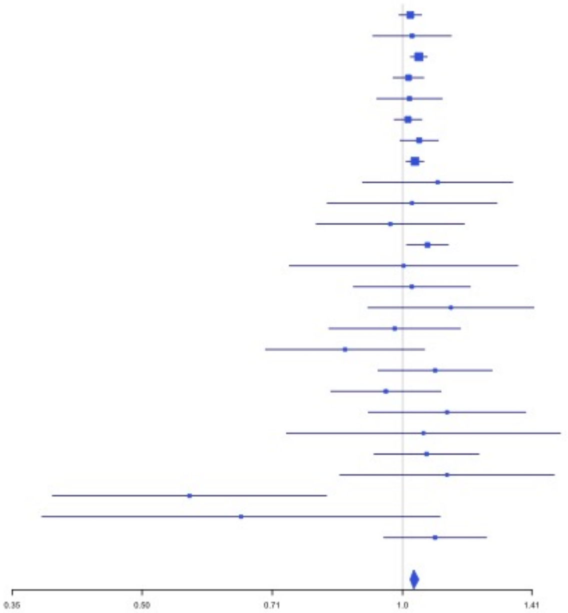


**Supplementary Figure 2.64** – rs174551 (chr11:61573684) in locus 82 (FADS1)

| Study | Cases | Controls | OR |
|---|---|---|---|
| INVENT-2019_EA | 15572 | 115280 | 1.05 |
| INVENT-2019_AA | 799 | 14353 | 0.97 |
| MVP_EA | 18952 | 411746 | 1.09 |
| MVP_AA | 5521 | 103791 | 1.05 |
| MVP_HIS | 1188 | 45634 | 1.08 |
| FinnGen_EA | 9176 | 209616 | 1.06 |
| UKB_EA | 18385 | 91834 | 1.06 |
| MGB_AA | 225 | 1481 | 1.14 |
| MGB_EA | 2967 | 27957 | 1.06 |
| MGB_HIS | 100 | 1352 | 0.95 |
| UPenn_EA | 380 | 6210 | 1.04 |
| UPenn_AA | 203 | 1719 | 1.2 |
| FARIVE_EA | 545 | 532 | 1.18 |
| BBJ_EAS | 507 | 178366 | 1.11 |
| MESA_AA | 72 | 1511 | 2.18 |
| MESA_HIS | 35 | 1389 | 1.31 |
| MARTHA12_EA | 893 | 3414 | 1.22 |
| **Overall** | **75520** | **1216185** | **1.07** |

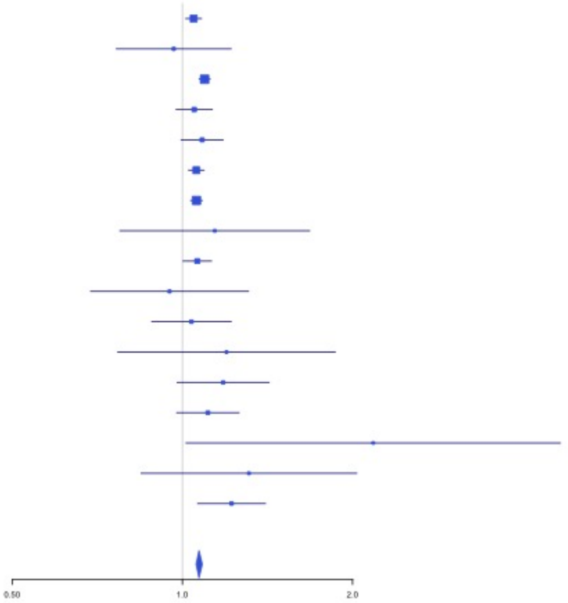

**Supplementary Figure 2.65** – rs2846027 (chr11:114003415) in locus 84 (ZBTB16)

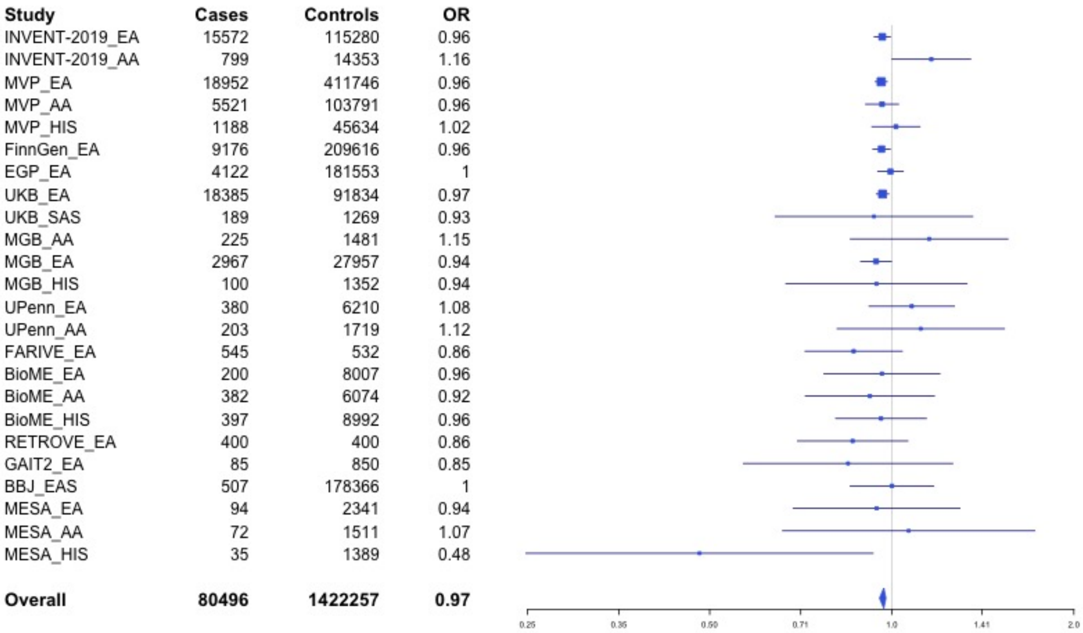

| Study | Cases | Controls | OR |
|---|---|---|---|
| INVENT-2019_EA | 15572 | 115280 | 0.96 |
| INVENT-2019_AA | 799 | 14353 | 1.16 |
| MVP_EA | 18952 | 411746 | 0.96 |
| MVP_AA | 5521 | 103791 | 0.96 |
| MVP_HIS | 1188 | 45634 | 1.02 |
| FinnGen_EA | 9176 | 209616 | 0.96 |
| EGP_EA | 4122 | 181553 | 1 |
| UKB_EA | 18385 | 91834 | 0.97 |
| UKB_SAS | 189 | 1269 | 0.93 |
| MGB_AA | 225 | 1481 | 1.15 |
| MGB_EA | 2967 | 27957 | 0.94 |
| MGB_HIS | 100 | 1352 | 0.94 |
| UPenn_EA | 380 | 6210 | 1.08 |
| UPenn_AA | 203 | 1719 | 1.12 |
| FARIVE_EA | 545 | 532 | 0.86 |
| BioME_EA | 200 | 8007 | 0.96 |
| BioME_AA | 382 | 6074 | 0.92 |
| BioME_HIS | 397 | 8992 | 0.96 |
| RETROVE_EA | 400 | 400 | 0.86 |
| GAIT2_EA | 85 | 850 | 0.85 |
| BBJ_EAS | 507 | 178366 | 1 |
| MESA_EA | 94 | 2341 | 0.94 |
| MESA_AA | 72 | 1511 | 1.07 |
| MESA_HIS | 35 | 1389 | 0.48 |
| **Overall** | **80496** | **1422257** | **0.97** |



**Supplementary Figure 2.66** – rs35257264 (chr11:126296816) in locus 85 (ST3GAL4)

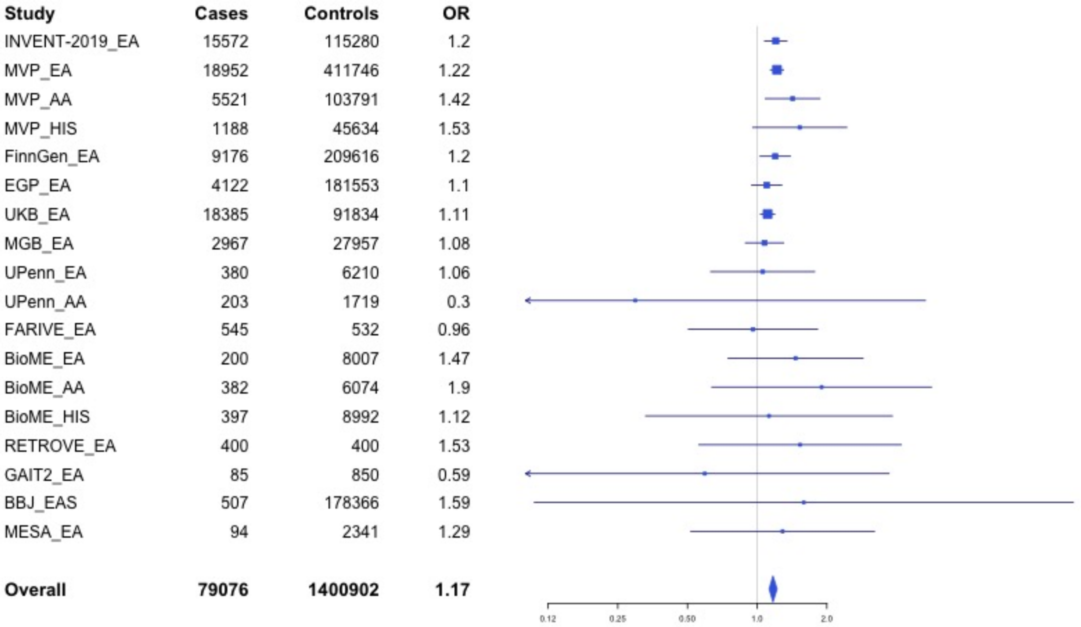

| Study | Cases | Controls | OR |
|---|---|---|---|
| INVENT-2019_EA | 15572 | 115280 | 1.2 |
| MVP_EA | 18952 | 411746 | 1.22 |
| MVP_AA | 5521 | 103791 | 1.42 |
| MVP_HIS | 1188 | 45634 | 1.53 |
| FinnGen_EA | 9176 | 209616 | 1.2 |
| EGP_EA | 4122 | 181553 | 1.1 |
| UKB_EA | 18385 | 91834 | 1.11 |
| MGB_EA | 2967 | 27957 | 1.08 |
| UPenn_EA | 380 | 6210 | 1.06 |
| UPenn_AA | 203 | 1719 | 0.3 |
| FARIVE_EA | 545 | 532 | 0.96 |
| BioME_EA | 200 | 8007 | 1.47 |
| BioME_AA | 382 | 6074 | 1.9 |
| BioME_HIS | 397 | 8992 | 1.12 |
| RETROVE_EA | 400 | 400 | 1.53 |
| GAIT2_EA | 85 | 850 | 0.59 |
| BBJ_EAS | 507 | 178366 | 1.59 |
| MESA_EA | 94 | 2341 | 1.29 |
| **Overall** | **79076** | **1400902** | **1.17** |

**Supplementary Figure 2.67** – rs7107568 (chr11:130779668) in locus 86 (SNX19)

| Study | Cases | Controls | OR |
|---|---|---|---|
| INVENT-2019_EA | 15572 | 115280 | 0.95 |
| INVENT-2019_AA | 799 | 14353 | 0.98 |
| MVP_EA | 18952 | 411746 | 0.98 |
| MVP_AA | 5521 | 103791 | 0.97 |
| MVP_HIS | 1188 | 45634 | 0.89 |
| FinnGen_EA | 9176 | 209616 | 0.98 |
| EGP_EA | 4122 | 181553 | 0.97 |
| UKB_EA | 18385 | 91834 | 0.97 |
| UKB_AA | 280 | 1046 | 0.8 |
| UKB_SAS | 189 | 1269 | 1.15 |
| MGB_AA | 225 | 1481 | 0.88 |
| MGB_EA | 2967 | 27957 | 0.96 |
| MGB_HIS | 100 | 1352 | 1.08 |
| UPenn_EA | 380 | 6210 | 0.83 |
| UPenn_AA | 203 | 1719 | 0.99 |
| FARIVE_EA | 545 | 532 | 0.93 |
| BioME_EA | 200 | 8007 | 0.91 |
| BioME_AA | 382 | 6074 | 0.99 |
| BioME_HIS | 397 | 8992 | 1.04 |
| RETROVE_EA | 400 | 400 | 1 |
| GAIT2_EA | 85 | 850 | 0.88 |
| BBJ_EAS | 507 | 178366 | 1 |
| MESA_EA | 94 | 2341 | 1.05 |
| MESA_AA | 72 | 1511 | 0.99 |
| MESA_HIS | 35 | 1389 | 1.01 |
| MARTHA12_EA | 893 | 3414 | 0.99 |
| **Overall** | **81669** | **1426717** | **0.97** |

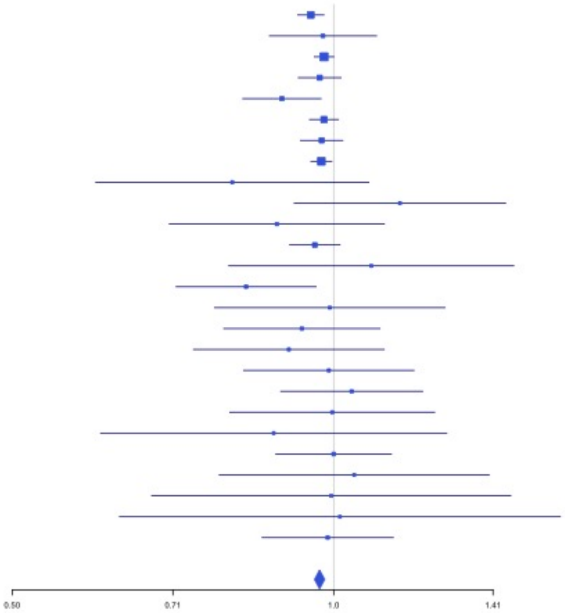


**Supplementary Figure 2.68** – rs1558519 (chr12:6153738) in locus 87 (VWF)

| Study | Cases | Controls | OR |
|---|---|---|---|
| INVENT-2019_EA | 15572 | 115280 | 0.92 |
| INVENT-2019_AA | 799 | 14353 | 0.99 |
| MVP_EA | 18952 | 411746 | 0.92 |
| MVP_AA | 5521 | 103791 | 0.96 |
| MVP_HIS | 1188 | 45634 | 1.01 |
| FinnGen_EA | 9176 | 209616 | 0.94 |
| EGP_EA | 4122 | 181553 | 0.96 |
| UKB_EA | 18385 | 91834 | 0.94 |
| UKB_AA | 280 | 1046 | 0.98 |
| UKB_SAS | 189 | 1269 | 0.83 |
| MGB_AA | 225 | 1481 | 1.08 |
| MGB_EA | 2967 | 27957 | 0.98 |
| MGB_HIS | 100 | 1352 | 0.78 |
| UPenn_EA | 380 | 6210 | 1.08 |
| UPenn_AA | 203 | 1719 | 1.11 |
| FARIVE_EA | 545 | 532 | 0.86 |
| BioME_EA | 200 | 8007 | 0.87 |
| BioME_AA | 382 | 6074 | 0.95 |
| BioME_HIS | 397 | 8992 | 1.02 |
| RETROVE_EA | 400 | 400 | 0.9 |
| GAIT2_EA | 85 | 850 | 1.02 |
| BBJ_EAS | 507 | 178366 | 1.04 |
| MESA_EA | 94 | 2341 | 0.82 |
| MESA_AA | 72 | 1511 | 1.39 |
| MESA_HIS | 35 | 1389 | 0.7 |
| MARTHA12_EA | 893 | 3414 | 0.96 |
| **Overall** | **81669** | **1426717** | **0.94** |

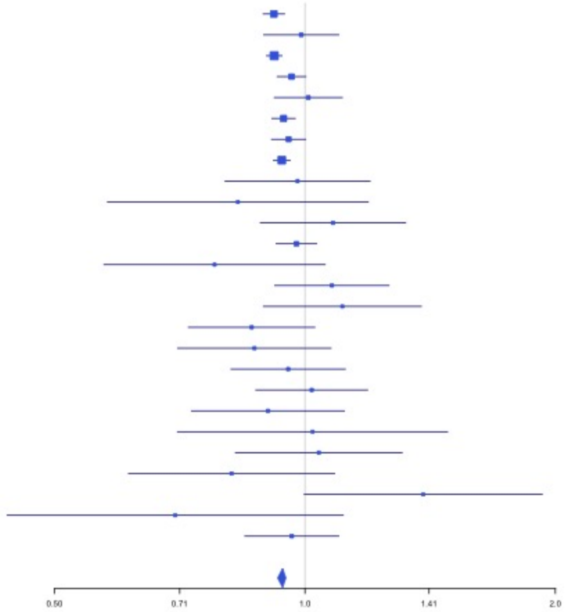

**Supplementary Figure 2.69** – rs7311483 (chr12:9053661) in locus 88 (A2ML1 ;PHC1)

| Study | Cases | Controls | OR |
|---|---|---|---|
| INVENT-2019_EA | 15572 | 115280 | 0.98 |
| INVENT-2019_AA | 799 | 14353 | 0.99 |
| MVP_EA | 18952 | 411746 | 0.95 |
| MVP_AA | 5521 | 103791 | 0.92 |
| MVP_HIS | 1188 | 45634 | 0.91 |
| FinnGen_EA | 9176 | 209616 | 0.95 |
| EGP_EA | 4122 | 181553 | 0.95 |
| UKB_EA | 18385 | 91834 | 0.97 |
| UKB_SAS | 189 | 1269 | 0.75 |
| MGB_AA | 225 | 1481 | 0.76 |
| MGB_EA | 2967 | 27957 | 0.97 |
| MGB_HIS | 100 | 1352 | 1 |
| UPenn_EA | 380 | 6210 | 1.05 |
| UPenn_AA | 203 | 1719 | 0.91 |
| FARIVE_EA | 545 | 532 | 0.94 |
| BioME_EA | 200 | 8007 | 0.85 |
| BioME_AA | 382 | 6074 | 0.89 |
| BioME_HIS | 397 | 8992 | 0.98 |
| RETROVE_EA | 400 | 400 | 1.05 |
| GAIT2_EA | 85 | 850 | 1.12 |
| BBJ_EAS | 507 | 178366 | 0.99 |
| MESA_EA | 94 | 2341 | 0.88 |
| MESA_AA | 72 | 1511 | 0.9 |
| MESA_HIS | 35 | 1389 | 1.46 |
| **Overall** | **80496** | **1422257** | **0.96** |

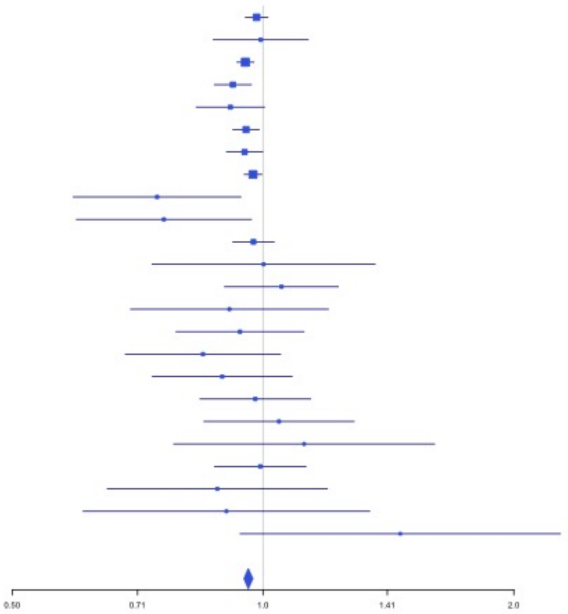


**Supplementary Figure 2.70** – rs6580981 (chr12:54723028) in locus 89 (COPZ1)

| Study | Cases | Controls | OR |
|---|---|---|---|
| INVENT-2019_EA | 15572 | 115280 | 0.96 |
| INVENT-2019_AA | 799 | 14353 | 1.03 |
| MVP_EA | 18952 | 411746 | 0.94 |
| MVP_AA | 5521 | 103791 | 0.97 |
| MVP_HIS | 1188 | 45634 | 0.95 |
| FinnGen_EA | 9176 | 209616 | 0.97 |
| EGP_EA | 4122 | 181553 | 0.98 |
| UKB_EA | 18385 | 91834 | 0.94 |
| UKB_SAS | 189 | 1269 | 1.1 |
| MGB_AA | 225 | 1481 | 1.1 |
| MGB_HIS | 100 | 1352 | 0.95 |
| UPenn_EA | 380 | 6210 | 0.99 |
| UPenn_AA | 203 | 1719 | 1 |
| FARIVE_EA | 545 | 532 | 0.9 |
| BioME_EA | 200 | 8007 | 1.01 |
| BioME_AA | 382 | 6074 | 1.14 |
| BioME_HIS | 397 | 8992 | 0.96 |
| RETROVE_EA | 400 | 400 | 0.82 |
| GAIT2_EA | 85 | 850 | 0.89 |
| BBJ_EAS | 507 | 178366 | 0.89 |
| MESA_EA | 94 | 2341 | 0.83 |
| MESA_AA | 72 | 1511 | 1.04 |
| MESA_HIS | 35 | 1389 | 0.88 |
| MARTHA12_EA | 893 | 3414 | 0.9 |
| **Overall** | **78422** | **1397714** | **0.95** |

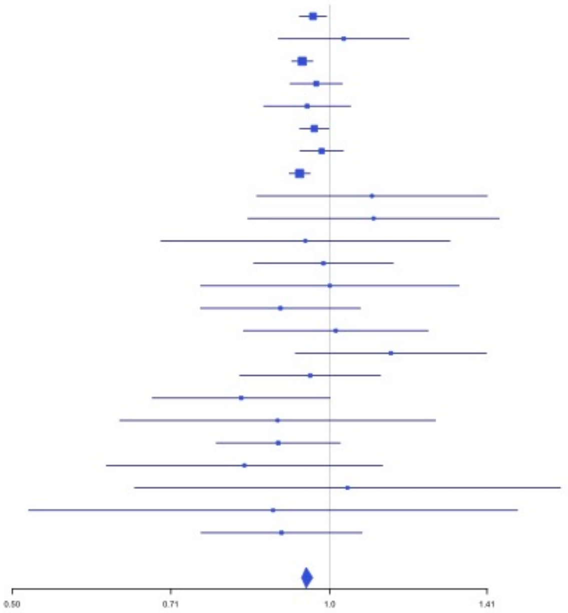

**Supplementary Figure 2.71** – rs142351376 (chr12:104136288) in locus 90 (STAB2)

| Study | Cases | Controls | OR |
|---|---|---|---|
| MVP_EA | 18952 | 411746 | 1.78 |
| FinnGen_EA | 9176 | 209616 | 1.67 |
| EGP_EA | 4122 | 181553 | 1.91 |
| UPenn_EA | 380 | 6210 | 0.99 |
| **Overall** | **32630** | **809125** | **1.68** |

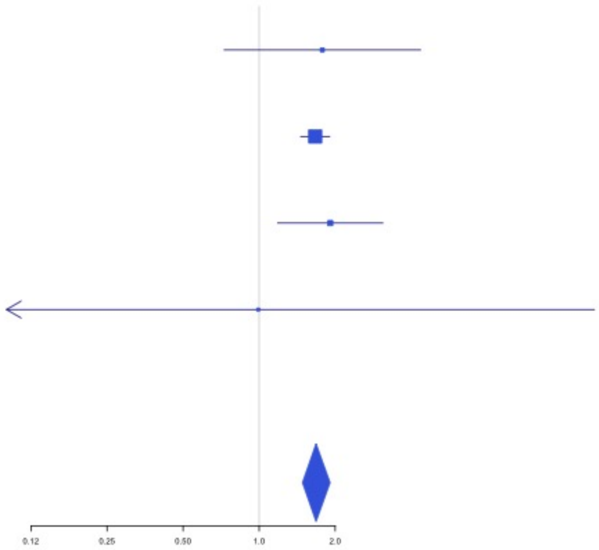


**Supplementary Figure 2.72** – rs3184504 (chr12:111884608) in locus 91 (SH2B3)

| Study | Cases | Controls | OR |
|---|---|---|---|
| INVENT-2019_EA | 15572 | 115280 | 1.04 |
| INVENT-2019_AA | 799 | 14353 | 0.99 |
| MVP_EA | 18952 | 411746 | 1.05 |
| MVP_AA | 5521 | 103791 | 1.02 |
| MVP_HIS | 1188 | 45634 | 1.05 |
| FinnGen_EA | 9176 | 209616 | 1.04 |
| EGP_EA | 4122 | 181553 | 1.05 |
| UKB_EA | 18385 | 91834 | 1.06 |
| MGB_AA | 225 | 1481 | 0.79 |
| MGB_EA | 2967 | 27957 | 0.99 |
| MGB_HIS | 100 | 1352 | 1.01 |
| UPenn_EA | 380 | 6210 | 1.05 |
| UPenn_AA | 203 | 1719 | 1.3 |
| FARIVE_EA | 545 | 532 | 0.95 |
| BioME_EA | 200 | 8007 | 0.99 |
| BioME_AA | 382 | 6074 | 0.94 |
| BioME_HIS | 397 | 8992 | 1.14 |
| RETROVE_EA | 400 | 400 | 0.98 |
| GAIT2_EA | 85 | 850 | 0.79 |
| BBJ_EAS | 507 | 178366 | 0.35 |
| MESA_EA | 94 | 2341 | 1.06 |
| MESA_AA | 72 | 1511 | 0.63 |
| MESA_HIS | 35 | 1389 | 1.08 |
| MARTHA12_EA | 893 | 3414 | 1.13 |
| **Overall** | **81200** | **1424402** | **1.05** |

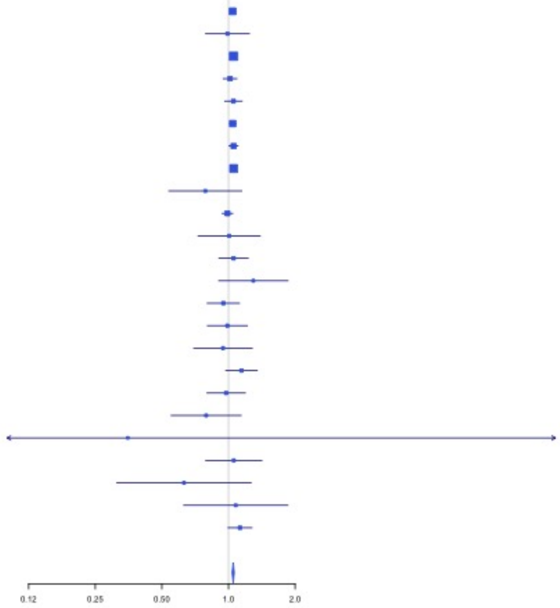

**Supplementary Figure 2.73** – rs2851435 (chr12:123712416) in locus 92 (MPHOSPH9)

| Study | Cases | Controls | OR |
|---|---|---|---|
| INVENT-2019_EA | 15572 | 115280 | 1.05 |
| INVENT-2019_AA | 799 | 14353 | 1.06 |
| MVP_EA | 18952 | 411746 | 1.06 |
| MVP_AA | 5521 | 103791 | 1.02 |
| MVP_HIS | 1188 | 45634 | 1.04 |
| FinnGen_EA | 9176 | 209616 | 1.02 |
| EGP_EA | 4122 | 181553 | 1.02 |
| UKB_EA | 18385 | 91834 | 1.06 |
| UKB_AA | 280 | 1046 | 1.04 |
| UKB_SAS | 189 | 1269 | 1.01 |
| MGB_AA | 225 | 1481 | 1.05 |
| MGB_EA | 2967 | 27957 | 1 |
| MGB_HIS | 100 | 1352 | 1.17 |
| UPenn_EA | 380 | 6210 | 1.22 |
| UPenn_AA | 203 | 1719 | 0.98 |
| FARIVE_EA | 545 | 532 | 1 |
| BioME_EA | 200 | 8007 | 1.06 |
| BioME_AA | 382 | 6074 | 1.09 |
| BioME_HIS | 397 | 8992 | 1.1 |
| RETROVE_EA | 400 | 400 | 1.14 |
| GAIT2_EA | 85 | 850 | 0.94 |
| MESA_EA | 94 | 2341 | 1.23 |
| MESA_AA | 72 | 1511 | 1.33 |
| MESA_HIS | 35 | 1389 | 0.71 |
| MARTHA12_EA | 893 | 3414 | 1.11 |
| **Overall** | **81162** | **1248351** | **1.05** |

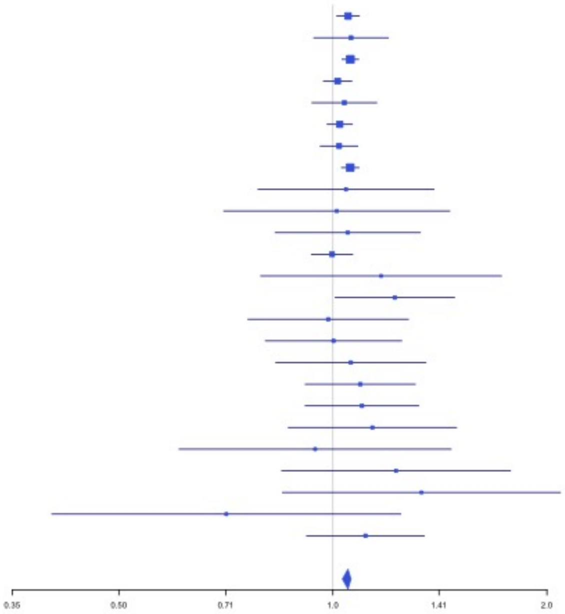


**Supplementary Figure 2.74** – rs3211752 (chr13:113787459) in locus 94 (F10)

| Study | Cases | Controls | OR |
|---|---|---|---|
| INVENT-2019_EA | 15572 | 115280 | 0.95 |
| INVENT-2019_AA | 799 | 14353 | 1.07 |
| MVP_EA | 18952 | 411746 | 0.95 |
| MVP_AA | 5521 | 103791 | 1 |
| MVP_HIS | 1188 | 45634 | 0.98 |
| FinnGen_EA | 9176 | 209616 | 0.94 |
| EGP_EA | 4122 | 181553 | 0.94 |
| UKB_EA | 18385 | 91834 | 0.96 |
| UKB_SAS | 189 | 1269 | 0.91 |
| MGB_AA | 225 | 1481 | 1.03 |
| MGB_EA | 2967 | 27957 | 0.98 |
| MGB_HIS | 100 | 1352 | 0.79 |
| UPenn_EA | 380 | 6210 | 0.93 |
| UPenn_AA | 203 | 1719 | 0.86 |
| FARIVE_EA | 545 | 532 | 1.03 |
| BioME_EA | 200 | 8007 | 0.96 |
| BioME_AA | 382 | 6074 | 1.04 |
| BioME_HIS | 397 | 8992 | 0.98 |
| RETROVE_EA | 400 | 400 | 0.97 |
| GAIT2_EA | 85 | 850 | 0.8 |
| MESA_EA | 94 | 2341 | 0.72 |
| MESA_AA | 72 | 1511 | 1.06 |
| MESA_HIS | 35 | 1389 | 0.69 |
| **Overall** | **79989** | **1243891** | **0.95** |

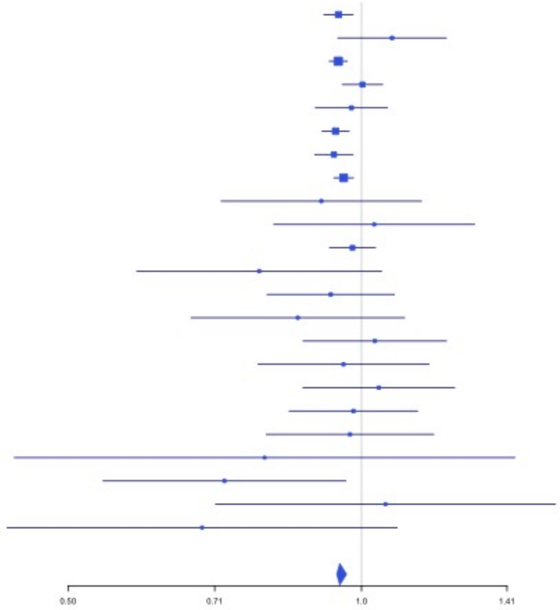

**Supplementary Figure 2.75** – rs11158204 (chr14:58844526) in locus 95 (ARID4A ;TOMM20L)

| Study | Cases | Controls | OR |
|---|---|---|---|
| INVENT-2019_EA | 15572 | 115280 | 0.92 |
| INVENT-2019_AA | 799 | 14353 | 0.94 |
| MVP_EA | 18952 | 411746 | 0.98 |
| MVP_AA | 5521 | 103791 | 0.99 |
| MVP_HIS | 1188 | 45634 | 0.99 |
| FinnGen_EA | 9176 | 209616 | 0.97 |
| EGP_EA | 4122 | 181553 | 0.94 |
| UKB_EA | 18385 | 91834 | 0.95 |
| UKB_AA | 280 | 1046 | 0.92 |
| MGB_AA | 225 | 1481 | 0.96 |
| MGB_EA | 2967 | 27957 | 0.91 |
| MGB_HIS | 100 | 1352 | 0.99 |
| UPenn_EA | 380 | 6210 | 1.19 |
| UPenn_AA | 203 | 1719 | 1.05 |
| FARIVE_EA | 545 | 532 | 0.81 |
| BioME_EA | 200 | 8007 | 0.84 |
| BioME_AA | 382 | 6074 | 1.02 |
| BioME_HIS | 397 | 8992 | 1.07 |
| RETROVE_EA | 400 | 400 | 1 |
| GAIT2_EA | 85 | 850 | 1.04 |
| BBJ_EAS | 507 | 178366 | 1.02 |
| MESA_EA | 94 | 2341 | 0.92 |
| MESA_AA | 72 | 1511 | 0.79 |
| MESA_HIS | 35 | 1389 | 0.99 |
| MARTHA12_EA | 893 | 3414 | 0.93 |
| **Overall** | **81480** | **1425448** | **0.96** |

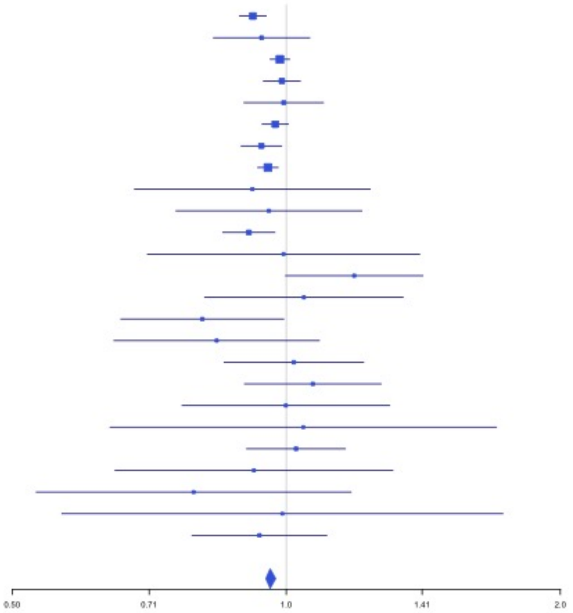


**Supplementary Figure 2.76** – rs2127869 (chr14:65794352) in locus 96 (LINC02324 ;MIR4708)

| Study | Cases | Controls | OR |
|---|---|---|---|
| INVENT-2019_EA | 15572 | 115280 | 0.97 |
| INVENT-2019_AA | 799 | 14353 | 1.04 |
| MVP_EA | 18952 | 411746 | 0.95 |
| MVP_AA | 5521 | 103791 | 0.98 |
| MVP_HIS | 1188 | 45634 | 0.93 |
| FinnGen_EA | 9176 | 209616 | 0.98 |
| EGP_EA | 4122 | 181553 | 0.99 |
| UKB_EA | 18385 | 91834 | 0.97 |
| UKB_AA | 280 | 1046 | 0.91 |
| UKB_SAS | 189 | 1269 | 1.25 |
| MGB_AA | 225 | 1481 | 1.17 |
| MGB_EA | 2967 | 27957 | 0.96 |
| MGB_HIS | 100 | 1352 | 1.26 |
| UPenn_EA | 380 | 6210 | 1.03 |
| UPenn_AA | 203 | 1719 | 1.08 |
| FARIVE_EA | 545 | 532 | 1.04 |
| BioME_EA | 200 | 8007 | 0.78 |
| BioME_AA | 382 | 6074 | 1.01 |
| BioME_HIS | 397 | 8992 | 0.89 |
| RETROVE_EA | 400 | 400 | 1.11 |
| GAIT2_EA | 85 | 850 | 0.86 |
| BBJ_EAS | 507 | 178366 | 0.91 |
| MESA_EA | 94 | 2341 | 0.92 |
| MESA_AA | 72 | 1511 | 1.02 |
| MESA_HIS | 35 | 1389 | 1.19 |
| **Overall** | **80776** | **1423303** | **0.97** |

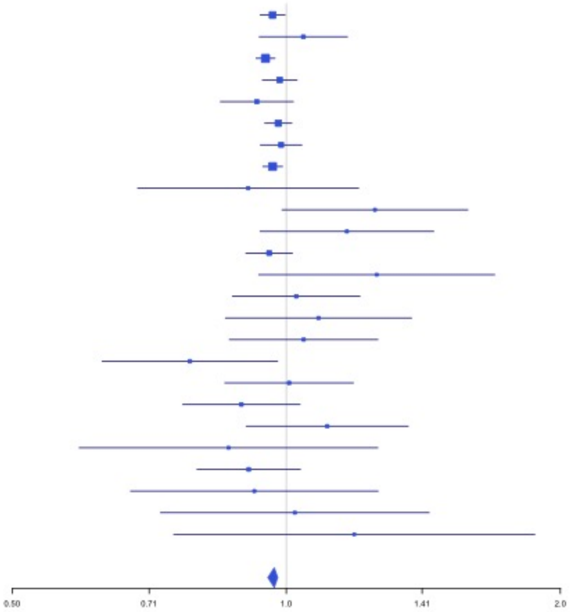

**Supplementary Figure 2.77** – rs7152641 (chr14:92213967) in locus 97 (CATSPERB;TC2N)

| Study | Cases | Controls | OR |
|---|---|---|---|
| INVENT-2019_EA | 15572 | 115280 | 0.95 |
| INVENT-2019_AA | 799 | 14353 | 0.94 |
| MVP_EA | 18952 | 411746 | 0.94 |
| MVP_AA | 5521 | 103791 | 0.95 |
| MVP_HIS | 1188 | 45634 | 0.9 |
| FinnGen_EA | 9176 | 209616 | 0.94 |
| EGP_EA | 4122 | 181553 | 0.9 |
| UKB_EA | 18385 | 91834 | 0.95 |
| UKB_AA | 280 | 1046 | 1 |
| UKB_SAS | 189 | 1269 | 0.92 |
| MGB_AA | 225 | 1481 | 0.82 |
| MGB_EA | 2967 | 27957 | 1.01 |
| MGB_HIS | 100 | 1352 | 1.03 |
| UPenn_EA | 380 | 6210 | 0.98 |
| UPenn_AA | 203 | 1719 | 1.07 |
| FARIVE_EA | 545 | 532 | 0.97 |
| BioME_EA | 200 | 8007 | 0.84 |
| BioME_AA | 382 | 6074 | 0.89 |
| BioME_HIS | 397 | 8992 | 0.91 |
| RETROVE_EA | 400 | 400 | 0.99 |
| GAIT2_EA | 85 | 850 | 1.02 |
| BBJ_EAS | 507 | 178366 | 0.95 |
| MESA_EA | 94 | 2341 | 0.82 |
| MESA_AA | 72 | 1511 | 0.84 |
| MESA_HIS | 35 | 1389 | 1.24 |
| **Overall** | **80776** | **1423303** | **0.95** |

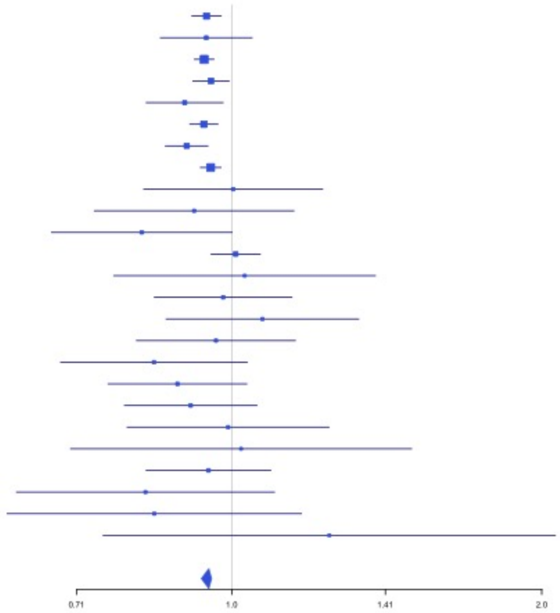


**Supplementary Figure 2.78** – rs28929474 (chr14:94844947) in locus 98 (SERPINA1)

| Study | Cases | Controls | OR |
|---|---|---|---|
| INVENT-2019_EA | 15572 | 115280 | 1.1 |
| MVP_EA | 18952 | 411746 | 1.09 |
| MVP_AA | 5521 | 103791 | 1.28 |
| MVP_HIS | 1188 | 45634 | 1.01 |
| FinnGen_EA | 9176 | 209616 | 1.2 |
| EGP_EA | 4122 | 181553 | 0.99 |
| UKB_EA | 18385 | 91834 | 1.2 |
| MGB_EA | 2967 | 27957 | 1.08 |
| UPenn_AA | 203 | 1719 | 0.45 |
| FARIVE_EA | 545 | 532 | 0.86 |
| BioME_EA | 200 | 8007 | 1.38 |
| BioME_AA | 382 | 6074 | 0.92 |
| BioME_HIS | 397 | 8992 | 0.97 |
| RETROVE_EA | 400 | 400 | 2.36 |
| GAIT2_EA | 85 | 850 | 4.05 |
| MESA_EA | 94 | 2341 | 0.25 |
| MARTHA12_EA | 893 | 3414 | 0.77 |
| **Overall** | **79082** | **1219740** | **1.13** |

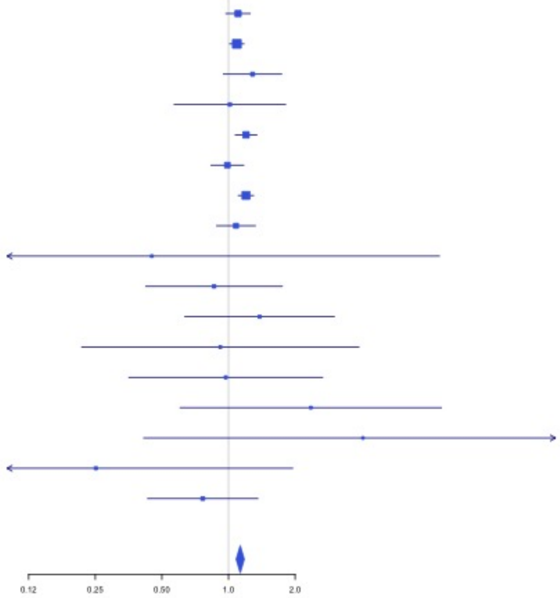

**Supplementary Figure 2.79** – rs28519971 (chr14:103089808) in locus 99 (RCOR1)

| Study | Cases | Controls | OR |
|---|---|---|---|
| INVENT-2019_EA | 15572 | 115280 | 0.97 |
| INVENT-2019_AA | 799 | 14353 | 0.89 |
| MVP_EA | 18952 | 411746 | 0.97 |
| MVP_AA | 5521 | 103791 | 0.95 |
| MVP_HIS | 1188 | 45634 | 0.86 |
| FinnGen_EA | 9176 | 209616 | 0.96 |
| EGP_EA | 4122 | 181553 | 0.94 |
| UKB_EA | 18385 | 91834 | 0.96 |
| UKB_AA | 280 | 1046 | 1.03 |
| UKB_SAS | 189 | 1269 | 0.97 |
| MGB_AA | 225 | 1481 | 1.13 |
| MGB_EA | 2967 | 27957 | 0.97 |
| MGB_HIS | 100 | 1352 | 0.96 |
| UPenn_EA | 380 | 6210 | 1.06 |
| UPenn_AA | 203 | 1719 | 1.02 |
| FARIVE_EA | 545 | 532 | 0.99 |
| BioME_EA | 200 | 8007 | 0.98 |
| BioME_AA | 382 | 6074 | 0.91 |
| BioME_HIS | 397 | 8992 | 1.12 |
| BBJ_EAS | 507 | 178366 | 1.11 |
| MESA_AA | 72 | 1511 | 1.07 |
| MESA_HIS | 35 | 1389 | 1.11 |
| **Overall** | **80197** | **1419712** | **0.96** |

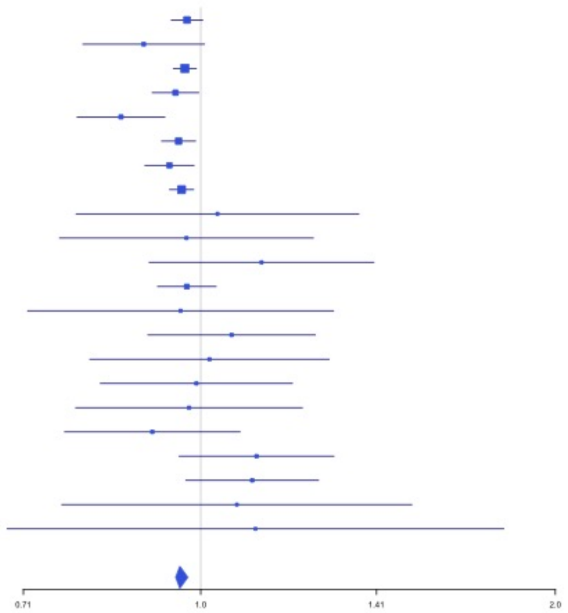


**Supplementary Figure 2.80** – rs1247656151 (chr15:43677975) in locus 100 (TUBGCP4)

| Study | Cases | Controls | OR |
|---|---|---|---|
| INVENT-2019_EA | 15572 | 115280 | 1.1 |
| MVP_EA | 18952 | 411746 | 1.16 |
| MVP_AA | 5521 | 103791 | 0.95 |
| MVP_HIS | 1188 | 45634 | 1.52 |
| FinnGen_EA | 9176 | 209616 | 1.15 |
| EGP_EA | 4122 | 181553 | 1.1 |
| UKB_EA | 18385 | 91834 | 1.2 |
| MGB_AA | 225 | 1481 | 0.63 |
| MGB_EA | 2967 | 27957 | 1.2 |
| MGB_HIS | 100 | 1352 | 1.2 |
| UPenn_EA | 380 | 6210 | 1.31 |
| UPenn_AA | 203 | 1719 | 0.87 |
| FARIVE_EA | 545 | 532 | 1.84 |
| BioME_EA | 200 | 8007 | 1.36 |
| BioME_AA | 382 | 6074 | 0.5 |
| BioME_HIS | 397 | 8992 | 1.63 |
| BBJ_EAS | 507 | 178366 | 0.18 |
| MARTHA12_EA | 893 | 3414 | 1.11 |
| **Overall** | **79715** | **1403558** | **1.16** |

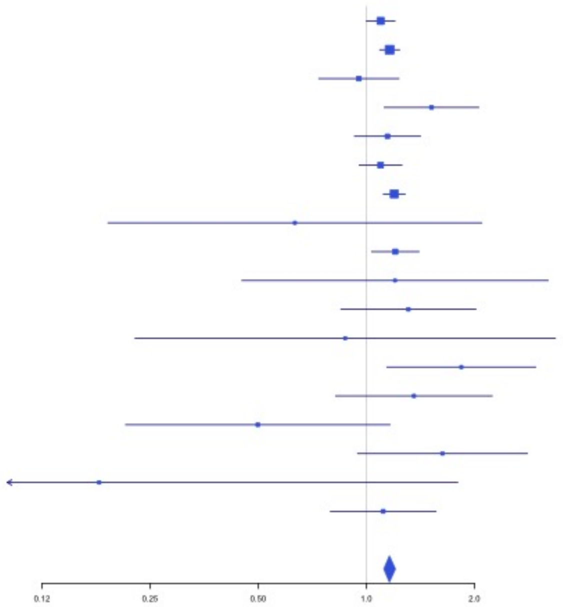

**Supplementary Figure 2.81** – rs7183672 (chr15:96101018) in locus 103 (LINC00924 ;LOC105369212)

| Study | Cases | Controls | OR |
|---|---|---|---|
| INVENT-2019_EA | 15572 | 115280 | 0.98 |
| INVENT-2019_AA | 799 | 14353 | 0.98 |
| MVP_EA | 18952 | 411746 | 0.96 |
| MVP_AA | 5521 | 103791 | 0.97 |
| MVP_HIS | 1188 | 45634 | 1.02 |
| FinnGen_EA | 9176 | 209616 | 0.98 |
| EGP_EA | 4122 | 181553 | 0.97 |
| UKB_EA | 18385 | 91834 | 0.95 |
| UKB_SAS | 189 | 1269 | 0.93 |
| MGB_AA | 225 | 1481 | 1.06 |
| MGB_EA | 2967 | 27957 | 1.03 |
| MGB_HIS | 100 | 1352 | 1.06 |
| UPenn_EA | 380 | 6210 | 0.99 |
| UPenn_AA | 203 | 1719 | 1.25 |
| FARIVE_EA | 545 | 532 | 1.03 |
| BioME_EA | 200 | 8007 | 0.96 |
| BioME_AA | 382 | 6074 | 1.06 |
| BioME_HIS | 397 | 8992 | 0.93 |
| RETROVE_EA | 400 | 400 | 0.95 |
| GAIT2_EA | 85 | 850 | 0.73 |
| BBJ_EAS | 507 | 178366 | 1.03 |
| MESA_EA | 94 | 2341 | 1.03 |
| MESA_AA | 72 | 1511 | 1.27 |
| MESA_HIS | 35 | 1389 | 1.34 |
| **Overall** | **80496** | **1422257** | **0.96** |

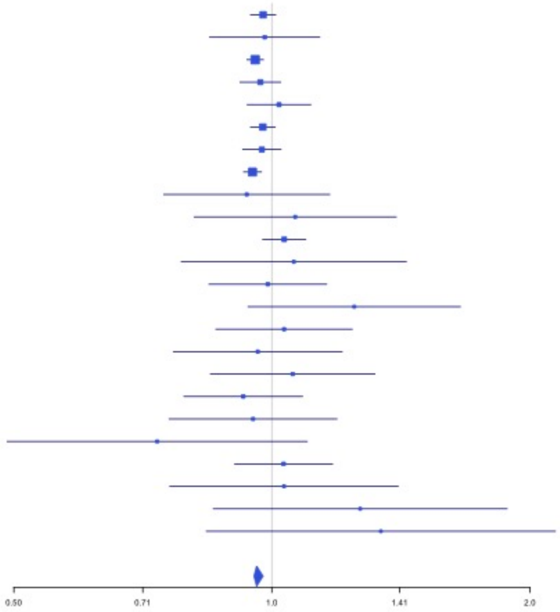


**Supplementary Figure 2.82** – rs71376077 (chr16:15738114) in locus 105 (NDE1)

| Study | Cases | Controls | OR |
|---|---|---|---|
| INVENT-2019_EA | 15572 | 115280 | 1.09 |
| INVENT-2019_AA | 799 | 14353 | 1.14 |
| MVP_EA | 18952 | 411746 | 1.15 |
| MVP_HIS | 1188 | 45634 | 1.1 |
| FinnGen_EA | 9176 | 209616 | 1.18 |
| EGP_EA | 4122 | 181553 | 1.1 |
| UKB_EA | 18385 | 91834 | 1.17 |
| MGB_AA | 225 | 1481 | 1.08 |
| MGB_EA | 2967 | 27957 | 0.9 |
| MGB_HIS | 100 | 1352 | 0.76 |
| UPenn_EA | 380 | 6210 | 1.26 |
| UPenn_AA | 203 | 1719 | 1.21 |
| FARIVE_EA | 545 | 532 | 0.42 |
| BioME_EA | 200 | 8007 | 1.13 |
| BioME_AA | 382 | 6074 | 1.44 |
| BioME_HIS | 397 | 8992 | 0.92 |
| RETROVE_EA | 400 | 400 | 1.48 |
| GAIT2_EA | 85 | 850 | 0.39 |
| BBJ_EAS | 507 | 178366 | 1.22 |
| MESA_EA | 94 | 2341 | 2.24 |
| MESA_AA | 72 | 1511 | 1.61 |
| MESA_HIS | 35 | 1389 | 1.54 |
| **Overall** | **74786** | **1317197** | **1.15** |

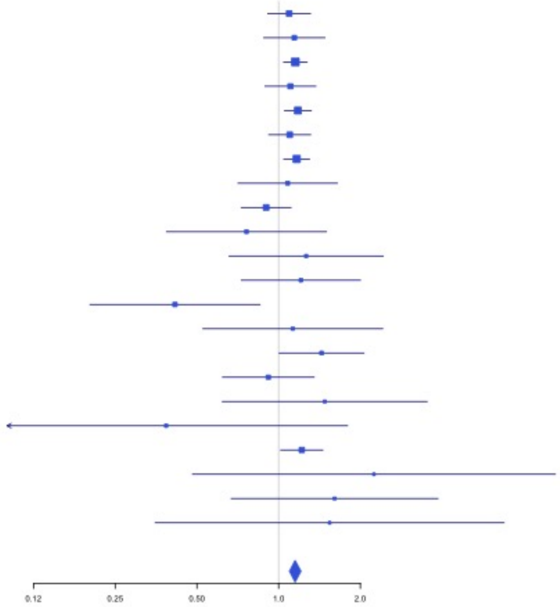

**Supplementary Figure 2.83** – rs12443808 (chr16:30996871) in locus 106 (HSD3B7)

| Study | Cases | Controls | OR |
|---|---|---|---|
| INVENT-2019_EA | 15572 | 115280 | 1.06 |
| INVENT-2019_AA | 799 | 14353 | 1.09 |
| MVP_EA | 18952 | 411746 | 1.06 |
| MVP_AA | 5521 | 103791 | 1.05 |
| MVP_HIS | 1188 | 45634 | 1.09 |
| FinnGen_EA | 9176 | 209616 | 1.05 |
| EGP_EA | 4122 | 181553 | 1.02 |
| UKB_EA | 18385 | 91834 | 1.03 |
| UKB_AA | 280 | 1046 | 0.93 |
| UKB_SAS | 189 | 1269 | 1.05 |
| MGB_AA | 225 | 1481 | 1.08 |
| MGB_EA | 2967 | 27957 | 1.02 |
| MGB_HIS | 100 | 1352 | 0.85 |
| UPenn_EA | 380 | 6210 | 0.96 |
| UPenn_AA | 203 | 1719 | 1.1 |
| FARIVE_EA | 545 | 532 | 1.12 |
| RETROVE_EA | 400 | 400 | 1.09 |
| GAIT2_EA | 85 | 850 | 0.71 |
| BBJ_EAS | 507 | 178366 | 1.09 |
| MESA_EA | 94 | 2341 | 0.91 |
| MESA_AA | 72 | 1511 | 0.85 |
| MESA_HIS | 35 | 1389 | 0.61 |
| MARTHA12_EA | 893 | 3414 | 1.14 |
| **Overall** | **80690** | **1403644** | **1.05** |

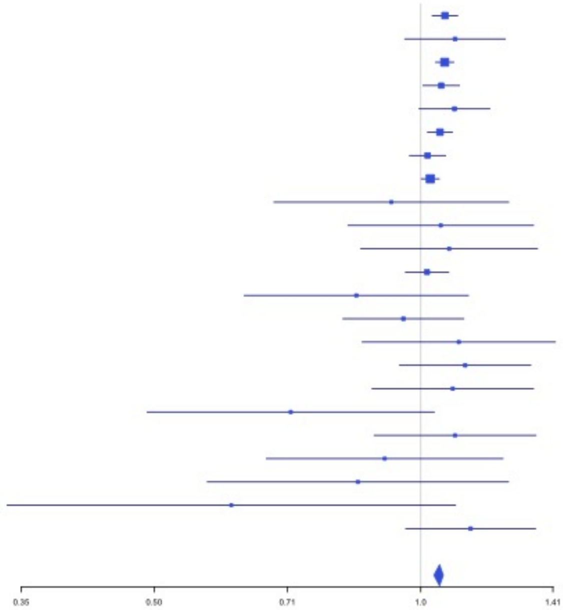


**Supplementary Figure 2.84** – rs7197453 (chr16:72079127) in locus 107 (DHODH ;HP)

| Study | Cases | Controls | OR |
|---|---|---|---|
| INVENT-2019_EA | 15572 | 115280 | 1.02 |
| INVENT-2019_AA | 799 | 14353 | 1.01 |
| MVP_EA | 18952 | 411746 | 1.04 |
| MVP_AA | 5521 | 103791 | 1.04 |
| MVP_HIS | 1188 | 45634 | 1.02 |
| FinnGen_EA | 9176 | 209616 | 1.03 |
| EGP_EA | 4122 | 181553 | 1.05 |
| UKB_EA | 18385 | 91834 | 1.03 |
| UKB_AA | 280 | 1046 | 1.12 |
| UKB_SAS | 189 | 1269 | 1.03 |
| MGB_AA | 225 | 1481 | 1.16 |
| MGB_EA | 2967 | 27957 | 1.01 |
| MGB_HIS | 100 | 1352 | 1.06 |
| UPenn_EA | 380 | 6210 | 1.01 |
| UPenn_AA | 203 | 1719 | 1.05 |
| FARIVE_EA | 545 | 532 | 1.05 |
| BioME_EA | 200 | 8007 | 1.1 |
| BioME_AA | 382 | 6074 | 0.98 |
| BioME_HIS | 397 | 8992 | 1.01 |
| RETROVE_EA | 400 | 400 | 0.96 |
| GAIT2_EA | 85 | 850 | 1.61 |
| BBJ_EAS | 507 | 178366 | 0.99 |
| MESA_EA | 94 | 2341 | 1.35 |
| MESA_AA | 72 | 1511 | 0.91 |
| MESA_HIS | 35 | 1389 | 0.94 |
| MARTHA12_EA | 893 | 3414 | 1.04 |
| **Overall** | **81669** | **1426717** | **1.03** |

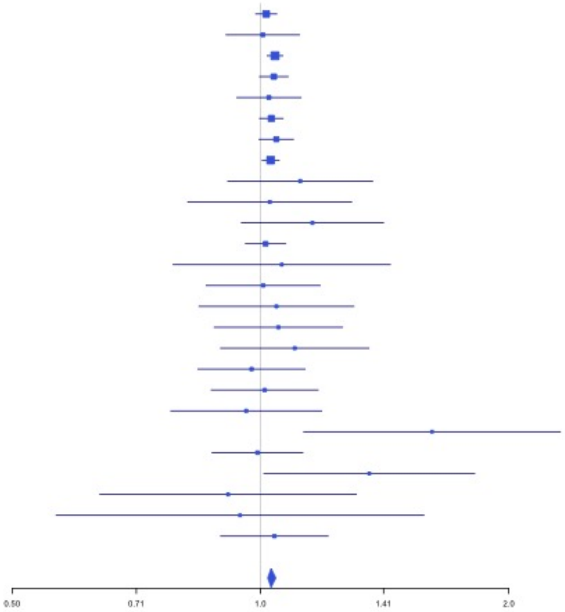

**Supplementary Figure 2.85** – rs77246010 (chr16:75429853) in locus 108 (CFDP1)

| Study | Cases | Controls | OR |
|---|---|---|---|
| INVENT-2019_EA | 15572 | 115280 | 1.07 |
| INVENT-2019_AA | 799 | 14353 | 1.13 |
| MVP_EA | 18952 | 411746 | 1.04 |
| MVP_AA | 5521 | 103791 | 1.01 |
| MVP_HIS | 1188 | 45634 | 1 |
| UKB_EA | 18385 | 91834 | 1.05 |
| UKB_AA | 280 | 1046 | 1.02 |
| UKB_SAS | 189 | 1269 | 1.06 |
| UPenn_EA | 380 | 6210 | 1.05 |
| UPenn_AA | 203 | 1719 | 0.95 |
| FARIVE_EA | 545 | 532 | 1 |
| BBJ_EAS | 507 | 178366 | 1.04 |
| MESA_AA | 72 | 1511 | 1.09 |
| MESA_HIS | 35 | 1389 | 1.12 |
| **Overall** | **62628** | **974680** | **1.04** |

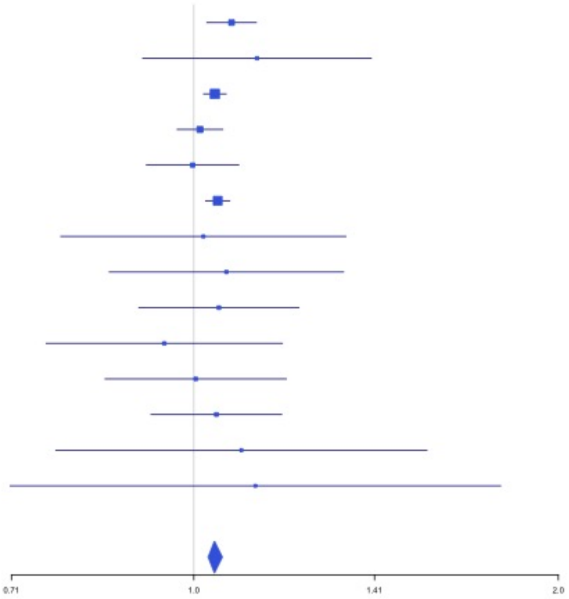


**Supplementary Figure 2.86** – rs12445050 (chr16:81870969) in locus 109 (PLCG2)

| Study | Cases | Controls | OR |
|---|---|---|---|
| INVENT-2019_EA | 15572 | 115280 | 1.1 |
| INVENT-2019_AA | 799 | 14353 | 0.82 |
| MVP_HIS | 1188 | 45634 | 1.28 |
| FinnGen_EA | 9176 | 209616 | 1.1 |
| EGP_EA | 4122 | 181553 | 1.07 |
| UKB_EA | 18385 | 91834 | 1.12 |
| MGB_AA | 225 | 1481 | 1.28 |
| MGB_EA | 2967 | 27957 | 1.04 |
| MGB_HIS | 100 | 1352 | 0.83 |
| UPenn_EA | 380 | 6210 | 1.09 |
| UPenn_AA | 203 | 1719 | 1.24 |
| FARIVE_EA | 545 | 532 | 1.44 |
| BioME_EA | 200 | 8007 | 0.9 |
| BioME_AA | 382 | 6074 | 0.99 |
| BioME_HIS | 397 | 8992 | 1 |
| RETROVE_EA | 400 | 400 | 1.54 |
| GAIT2_EA | 85 | 850 | 1.63 |
| BBJ_EAS | 507 | 178366 | 1.37 |
| MESA_EA | 94 | 2341 | 1.13 |
| MESA_AA | 72 | 1511 | 0.83 |
| MESA_HIS | 35 | 1389 | 1.28 |
| **Overall** | **55834** | **905451** | **1.11** |

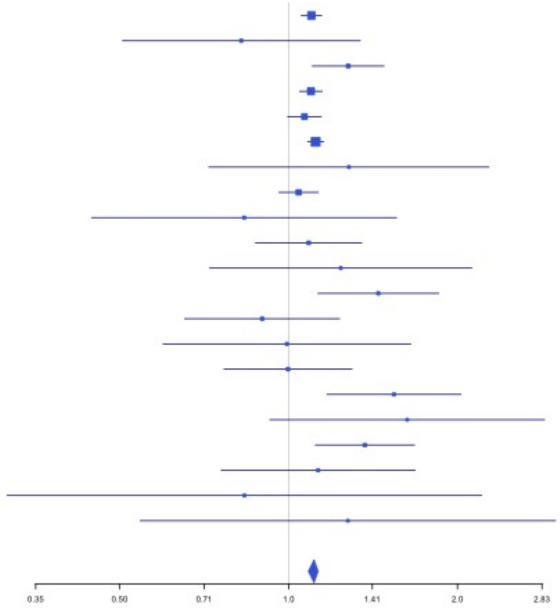

**Supplementary Figure 2.87** – rs8049403 (chr16:85778651) in locus 110 (C16orf74)

| Study | Cases | Controls | OR |
|---|---|---|---|
| INVENT-2019_EA | 15572 | 115280 | 1.03 |
| INVENT-2019_AA | 799 | 14353 | 1.27 |
| MVP_EA | 18952 | 411746 | 1.12 |
| MVP_HIS | 1188 | 45634 | 1.13 |
| FinnGen_EA | 9176 | 209616 | 1.23 |
| EGP_EA | 4122 | 181553 | 1.45 |
| UKB_EA | 18385 | 91834 | 1.15 |
| MGB_AA | 225 | 1481 | 1.13 |
| MGB_EA | 2967 | 27957 | 1.09 |
| MGB_HIS | 100 | 1352 | 1.06 |
| UPenn_EA | 380 | 6210 | 0.32 |
| UPenn_AA | 203 | 1719 | 1.29 |
| FARIVE_EA | 545 | 532 | 1.17 |
| BioME_EA | 200 | 8007 | 1.31 |
| BioME_AA | 382 | 6074 | 0.97 |
| BioME_HIS | 397 | 8992 | 0.8 |
| RETROVE_EA | 400 | 400 | 1.54 |
| GAIT2_EA | 85 | 850 | 2.01 |
| MESA_EA | 94 | 2341 | 1.16 |
| MESA_AA | 72 | 1511 | 0.8 |
| MESA_HIS | 35 | 1389 | 1.24 |
| **Overall** | **74279** | **1138831** | **1.15** |

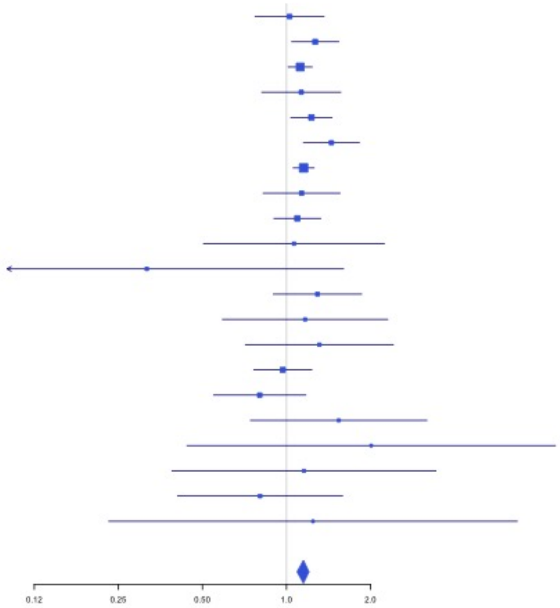


**Supplementary Figure 2.88** – rs28634651 (chr16:88553198) in locus 111 (ZFPM1)

| Study | Cases | Controls | OR |
|---|---|---|---|
| INVENT-2019_EA | 15572 | 115280 | 1.04 |
| INVENT-2019_AA | 799 | 14353 | 1.12 |
| MVP_EA | 18952 | 411746 | 1.06 |
| MVP_HIS | 1188 | 45634 | 1.05 |
| FinnGen_EA | 9176 | 209616 | 1.07 |
| UKB_EA | 18385 | 91834 | 1.04 |
| UKB_AA | 280 | 1046 | 1.2 |
| UKB_SAS | 189 | 1269 | 1.28 |
| MGB_AA | 225 | 1481 | 1.02 |
| MGB_EA | 2967 | 27957 | 1.03 |
| MGB_HIS | 100 | 1352 | 0.88 |
| UPenn_EA | 380 | 6210 | 1.04 |
| UPenn_AA | 203 | 1719 | 1.09 |
| FARIVE_EA | 545 | 532 | 1.16 |
| BioME_EA | 200 | 8007 | 1 |
| BioME_AA | 382 | 6074 | 0.98 |
| BioME_HIS | 397 | 8992 | 0.85 |
| RETROVE_EA | 400 | 400 | 1 |
| GAIT2_EA | 85 | 850 | 0.96 |
| MESA_EA | 94 | 2341 | 0.8 |
| MESA_AA | 72 | 1511 | 1.02 |
| MESA_HIS | 35 | 1389 | 0.59 |
| **Overall** | **70626** | **959593** | **1.05** |

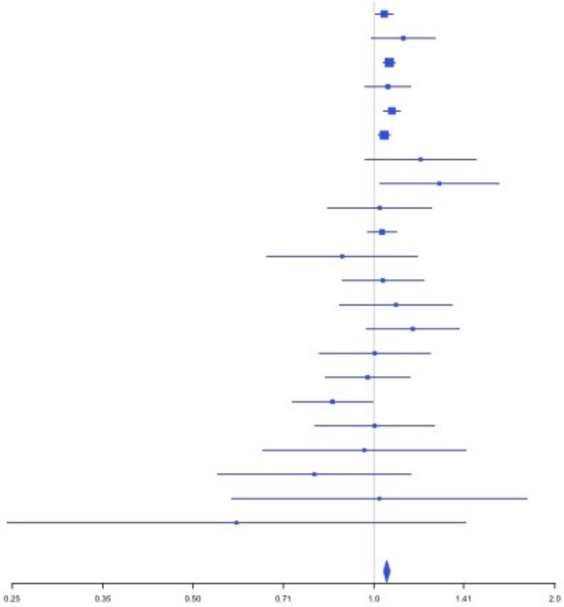

**Supplementary Figure 2.89** – rs6503222 (chr17:1977862) in locus 112 (SMG6)

| Study | Cases | Controls | OR |
|---|---|---|---|
| INVENT-2019_EA | 15572 | 115280 | 1.06 |
| INVENT-2019_AA | 799 | 14353 | 0.95 |
| MVP_EA | 18952 | 411746 | 1.06 |
| MVP_AA | 5521 | 103791 | 1.03 |
| MVP_HIS | 1188 | 45634 | 1.09 |
| FinnGen_EA | 9176 | 209616 | 1.03 |
| EGP_EA | 4122 | 181553 | 1.06 |
| UKB_EA | 18385 | 91834 | 1.04 |
| UKB_SAS | 189 | 1269 | 1.18 |
| MGB_AA | 225 | 1481 | 1.05 |
| MGB_EA | 2967 | 27957 | 1.06 |
| MGB_HIS | 100 | 1352 | 1.36 |
| UPenn_EA | 380 | 6210 | 0.93 |
| UPenn_AA | 203 | 1719 | 0.94 |
| FARIVE_EA | 545 | 532 | 1.13 |
| BioME_EA | 200 | 8007 | 0.95 |
| BioME_AA | 382 | 6074 | 1.11 |
| BioME_HIS | 397 | 8992 | 1.16 |
| RETROVE_EA | 400 | 400 | 1.01 |
| GAIT2_EA | 85 | 850 | 1.04 |
| BBJ_EAS | 507 | 178366 | 1.1 |
| MESA_EA | 94 | 2341 | 1.02 |
| MESA_AA | 72 | 1511 | 1.07 |
| MESA_HIS | 35 | 1389 | 0.7 |
| **Overall** | **80496** | **1422257** | **1.05** |

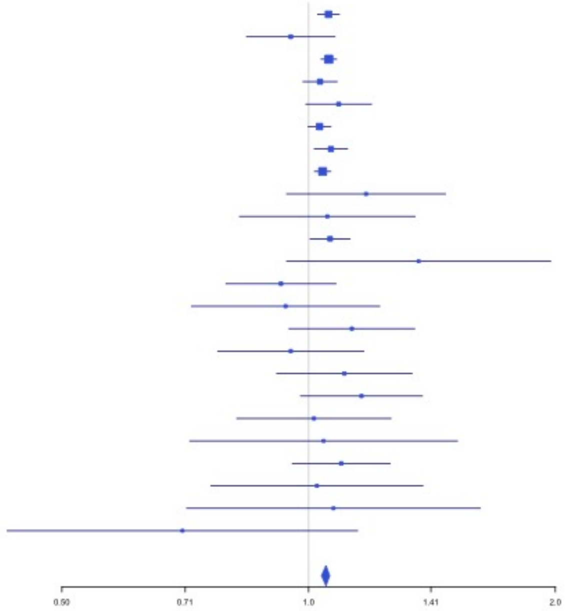


**Supplementary Figure 2.90** – rs434473 (chr17:6904934) in locus 113 (ALOX12)

| Study | Cases | Controls | OR |
|---|---|---|---|
| INVENT-2019_EA | 15572 | 115280 | 0.96 |
| INVENT-2019_AA | 799 | 14353 | 0.97 |
| MVP_EA | 18952 | 411746 | 0.96 |
| MVP_AA | 5521 | 103791 | 0.95 |
| MVP_HIS | 1188 | 45634 | 0.99 |
| FinnGen_EA | 9176 | 209616 | 0.99 |
| EGP_EA | 4122 | 181553 | 0.99 |
| UKB_EA | 18385 | 91834 | 0.96 |
| UKB_SAS | 189 | 1269 | 0.98 |
| MGB_AA | 225 | 1481 | 0.81 |
| MGB_EA | 2967 | 27957 | 1 |
| MGB_HIS | 100 | 1352 | 1.26 |
| UPenn_EA | 380 | 6210 | 0.9 |
| UPenn_AA | 203 | 1719 | 0.77 |
| FARIVE_EA | 545 | 532 | 0.92 |
| BioME_EA | 200 | 8007 | 0.87 |
| BioME_AA | 382 | 6074 | 0.95 |
| BioME_HIS | 397 | 8992 | 0.99 |
| RETROVE_EA | 400 | 400 | 0.92 |
| GAIT2_EA | 85 | 850 | 0.83 |
| BBJ_EAS | 507 | 178366 | 1.06 |
| MESA_EA | 94 | 2341 | 1.34 |
| MESA_AA | 72 | 1511 | 0.97 |
| MESA_HIS | 35 | 1389 | 1.04 |
| MARTHA12_EA | 893 | 3414 | 1.03 |
| **Overall** | **81389** | **1425671** | **0.97** |

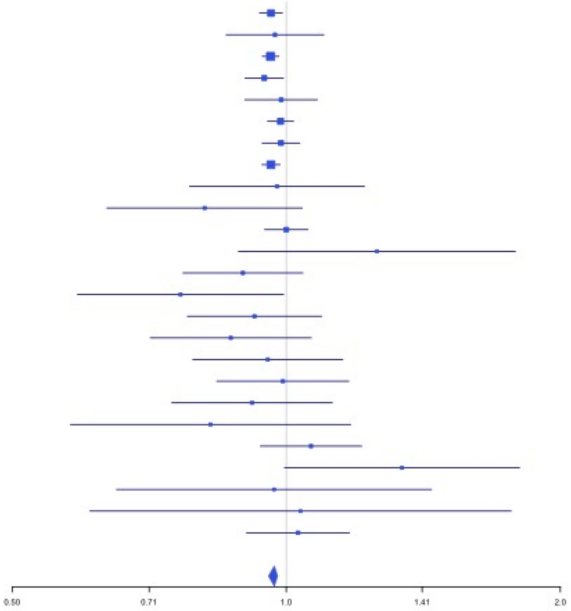

**Supplementary Figure 2.91** – rs71138827 (chr17:27833678) in locus 114 (TAOK1)

| Study | Cases | Controls | OR |
|---|---|---|---|
| INVENT-2019_EA | 15572 | 115280 | 1.04 |
| INVENT-2019_AA | 799 | 14353 | 0.96 |
| MVP_EA | 18952 | 411746 | 1.02 |
| MVP_AA | 5521 | 103791 | 1.07 |
| MVP_HIS | 1188 | 45634 | 0.95 |
| FinnGen_EA | 9176 | 209616 | 1.05 |
| EGP_EA | 4122 | 181553 | 1.03 |
| UKB_EA | 18385 | 91834 | 1.04 |
| UKB_SAS | 189 | 1269 | 1.01 |
| MGB_AA | 225 | 1481 | 1.31 |
| MGB_EA | 2967 | 27957 | 1 |
| MGB_HIS | 100 | 1352 | 1.33 |
| UPenn_EA | 380 | 6210 | 1.16 |
| UPenn_AA | 203 | 1719 | 0.95 |
| FARIVE_EA | 545 | 532 | 0.96 |
| BioME_EA | 200 | 8007 | 1.03 |
| BioME_AA | 382 | 6074 | 1.2 |
| BioME_HIS | 397 | 8992 | 0.99 |
| BBJ_EAS | 507 | 178366 | 1.04 |
| **Overall** | **79810** | **1415766** | **1.03** |

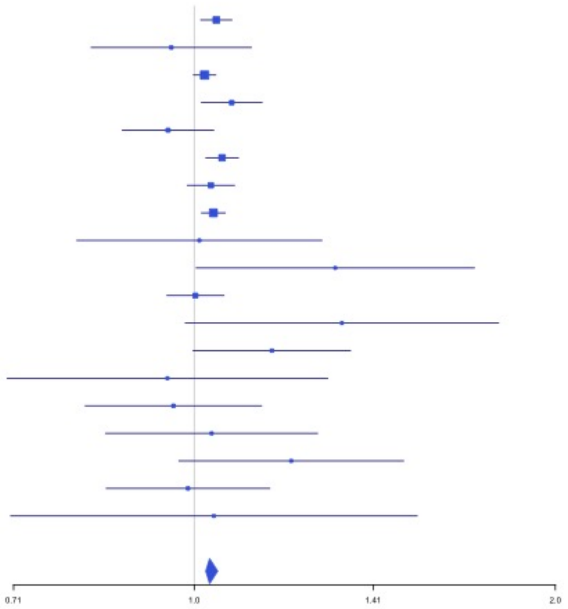


**Supplementary Figure 2.92** – rs62054822 (chr17:43927708) in locus 115 (MAPT-AS1)

| Study | Cases | Controls | OR |
|---|---|---|---|
| INVENT-2019_EA | 15572 | 115280 | 0.96 |
| INVENT-2019_AA | 799 | 14353 | 0.94 |
| MVP_EA | 18952 | 411746 | 0.95 |
| MVP_AA | 5521 | 103791 | 0.9 |
| MVP_HIS | 1188 | 45634 | 0.89 |
| FinnGen_EA | 9176 | 209616 | 0.97 |
| EGP_EA | 4122 | 181553 | 0.97 |
| UKB_EA | 18385 | 91834 | 0.96 |
| MGB_AA | 225 | 1481 | 0.97 |
| MGB_EA | 2967 | 27957 | 0.93 |
| MGB_HIS | 100 | 1352 | 0.84 |
| UPenn_EA | 380 | 6210 | 0.97 |
| UPenn_AA | 203 | 1719 | 0.84 |
| FARIVE_EA | 545 | 532 | 0.97 |
| BioME_EA | 200 | 8007 | 0.92 |
| BioME_AA | 382 | 6074 | 0.81 |
| BioME_HIS | 397 | 8992 | 0.88 |
| RETROVE_EA | 400 | 400 | 0.81 |
| GAIT2_EA | 85 | 850 | 1.31 |
| BBJ_EAS | 507 | 178366 | 3.2 |
| MESA_EA | 94 | 2341 | 1 |
| MESA_AA | 72 | 1511 | 0.69 |
| MESA_HIS | 35 | 1389 | 1.8 |
| MARTHA12_EA | 893 | 3414 | 0.85 |
| **Overall** | **81200** | **1424402** | **0.95** |

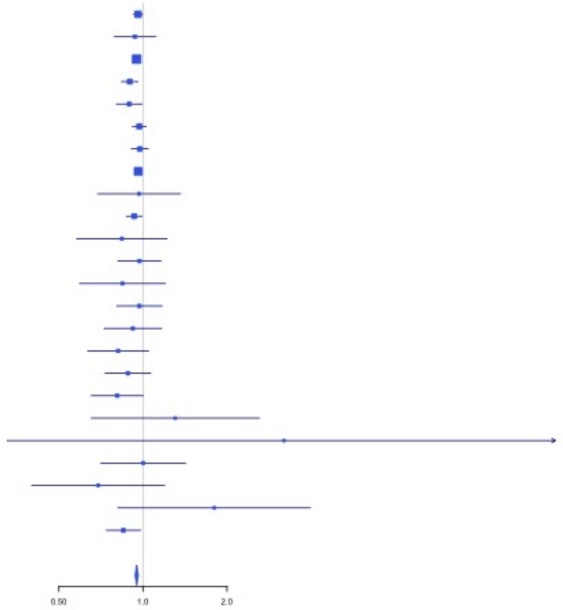

**Supplementary Figure 2.93** – rs1801690 (chr17:64208285) in locus 116 (APOH)

| Study | Cases | Controls | OR |
|---|---|---|---|
| INVENT-2019_EA | 15572 | 115280 | 1.03 |
| MVP_EA | 18952 | 411746 | 1.11 |
| MVP_AA | 5521 | 103791 | 1.01 |
| MVP_HIS | 1188 | 45634 | 0.95 |
| FinnGen_EA | 9176 | 209616 | 1.11 |
| EGP_EA | 4122 | 181553 | 1.04 |
| UKB_EA | 18385 | 91834 | 1.04 |
| MGB_AA | 225 | 1481 | 1.13 |
| MGB_EA | 2967 | 27957 | 0.97 |
| MGB_HIS | 100 | 1352 | 1.42 |
| UPenn_EA | 380 | 6210 | 1.04 |
| UPenn_AA | 203 | 1719 | 1.56 |
| FARIVE_EA | 545 | 532 | 1.3 |
| BioME_EA | 200 | 8007 | 0.93 |
| BioME_AA | 382 | 6074 | 2.42 |
| BioME_HIS | 397 | 8992 | 0.97 |
| RETROVE_EA | 400 | 400 | 1.46 |
| GAIT2_EA | 85 | 850 | 1.14 |
| BBJ_EAS | 507 | 178366 | 1.02 |
| MARTHA12_EA | 893 | 3414 | 1.33 |
| **Overall** | **80200** | **1404808** | **1.07** |

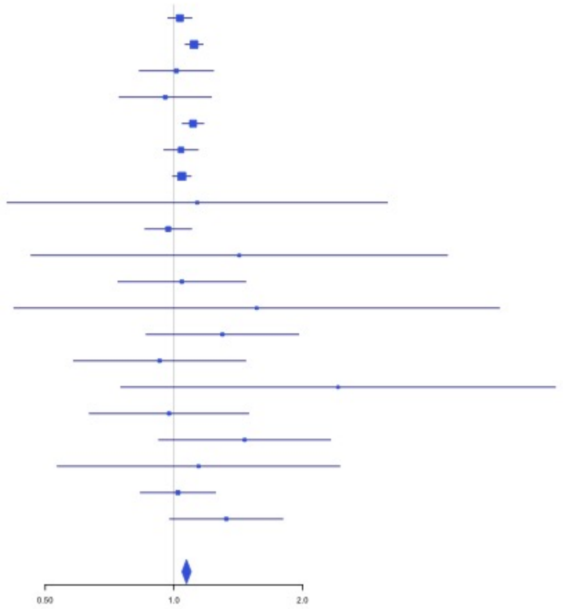


**Supplementary Figure 2.94** – rs573431210 (chr17:67276383) in locus 117 (ABCA5)

| Study | Cases | Controls | OR |
|---|---|---|---|
| INVENT-2019_EA | 15572 | 115280 | 1.14 |
| MVP_EA | 18952 | 411746 | 0.88 |
| MVP_AA | 5521 | 103791 | 0.76 |
| MVP_HIS | 1188 | 45634 | 0.94 |
| FinnGen_EA | 9176 | 209616 | 0.95 |
| EGP_EA | 4122 | 181553 | 0.69 |
| UKB_EA | 18385 | 91834 | 0.82 |
| MGB_EA | 2967 | 27957 | 0.93 |
| MGB_HIS | 100 | 1352 | 0.98 |
| UPenn_EA | 380 | 6210 | 1.24 |
| UPenn_AA | 203 | 1719 | 2.23 |
| FARIVE_EA | 545 | 532 | 0.52 |
| BioME_EA | 200 | 8007 | 1.19 |
| BioME_AA | 382 | 6074 | 0.82 |
| BioME_HIS | 397 | 8992 | 0.67 |
| MARTHA12_EA | 893 | 3414 | 0.71 |
| **Overall** | **78983** | **1223711** | **0.87** |

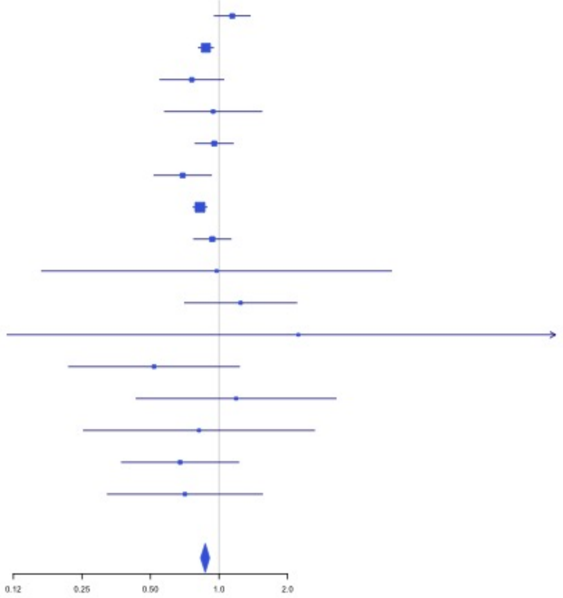

**Supplementary Figure 2.95** – rs59277920 (chr19:6077231) in locus 118 (RFX2)

| Study | Cases | Controls | OR |
|---|---|---|---|
| INVENT-2019_EA | 15572 | 115280 | 0.97 |
| INVENT-2019_AA | 799 | 14353 | 0.92 |
| MVP_EA | 18952 | 411746 | 0.92 |
| MVP_AA | 5521 | 103791 | 0.93 |
| MVP_HIS | 1188 | 45634 | 0.97 |
| FinnGen_EA | 9176 | 209616 | 0.98 |
| EGP_EA | 4122 | 181553 | 0.94 |
| UKB_EA | 18385 | 91834 | 0.97 |
| UKB_AA | 280 | 1046 | 0.92 |
| MGB_AA | 225 | 1481 | 1.04 |
| MGB_EA | 2967 | 27957 | 0.97 |
| MGB_HIS | 100 | 1352 | 1.02 |
| UPenn_EA | 380 | 6210 | 0.97 |
| UPenn_AA | 203 | 1719 | 0.81 |
| FARIVE_EA | 545 | 532 | 0.79 |
| BioME_EA | 200 | 8007 | 1.02 |
| BioME_AA | 382 | 6074 | 1.05 |
| BioME_HIS | 397 | 8992 | 1.04 |
| RETROVE_EA | 400 | 400 | 0.88 |
| GAIT2_EA | 85 | 850 | 0.81 |
| BBJ_EAS | 507 | 178366 | 1.06 |
| MESA_EA | 94 | 2341 | 0.61 |
| MESA_AA | 72 | 1511 | 1.16 |
| MESA_HIS | 35 | 1389 | 0.66 |
| **Overall** | **80587** | **1422034** | **0.95** |

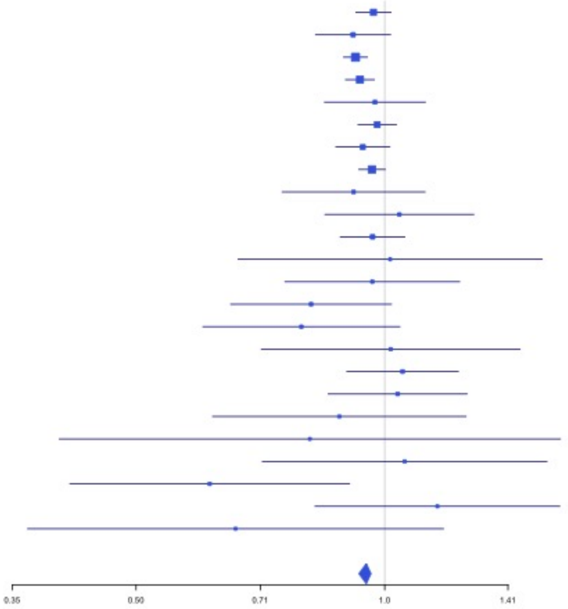


**Supplementary Figure 2.96** – rs8110055 (chr19:10739143) in locus 119 (SLC44A2)

| Study | Cases | Controls | OR |
|---|---|---|---|
| INVENT-2019_EA | 15572 | 115280 | 0.9 |
| INVENT-2019_AA | 799 | 14353 | 0.96 |
| MVP_EA | 18952 | 411746 | 0.88 |
| MVP_AA | 5521 | 103791 | 0.86 |
| MVP_HIS | 1188 | 45634 | 0.78 |
| FinnGen_EA | 9176 | 209616 | 0.87 |
| EGP_EA | 4122 | 181553 | 0.93 |
| UKB_EA | 18385 | 91834 | 0.9 |
| UKB_SAS | 189 | 1269 | 1.24 |
| MGB_AA | 225 | 1481 | 0.97 |
| MGB_EA | 2967 | 27957 | 0.93 |
| MGB_HIS | 100 | 1352 | 0.89 |
| UPenn_EA | 380 | 6210 | 1.07 |
| UPenn_AA | 203 | 1719 | 0.76 |
| FARIVE_EA | 545 | 532 | 0.84 |
| BioME_EA | 200 | 8007 | 0.89 |
| BioME_AA | 382 | 6074 | 1.02 |
| BioME_HIS | 397 | 8992 | 0.93 |
| RETROVE_EA | 400 | 400 | 0.72 |
| GAIT2_EA | 85 | 850 | 0.67 |
| BBJ_EAS | 507 | 178366 | 0.93 |
| MESA_EA | 94 | 2341 | 0.92 |
| MESA_AA | 72 | 1511 | 1.29 |
| MESA_HIS | 35 | 1389 | 0.69 |
| MARTHA12_EA | 893 | 3414 | 0.87 |
| **Overall** | **81389** | **1425671** | **0.89** |

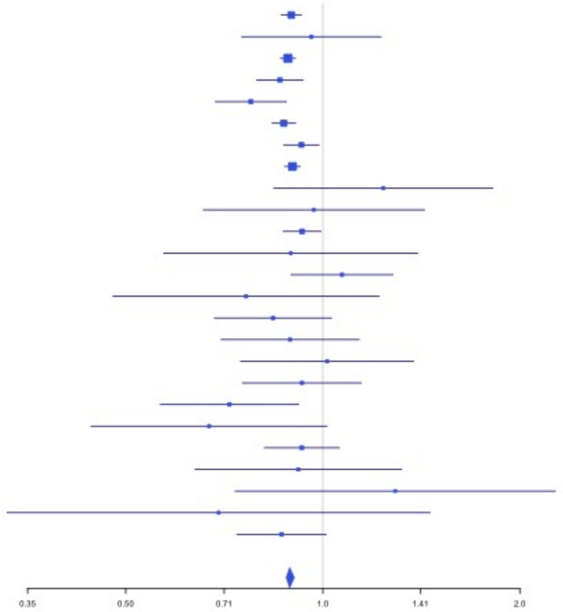

**Supplementary Figure 2.97** – rs889139 (chr19:33889369) in locus 120 (PEPD)

| Study | Cases | Controls | OR |
|---|---|---|---|
| INVENT-2019_EA | 15572 | 115280 | 0.97 |
| INVENT-2019_AA | 799 | 14353 | 1.01 |
| MVP_EA | 18952 | 411746 | 0.98 |
| MVP_AA | 5521 | 103791 | 0.97 |
| MVP_HIS | 1188 | 45634 | 0.94 |
| FinnGen_EA | 9176 | 209616 | 0.97 |
| EGP_EA | 4122 | 181553 | 0.97 |
| UKB_EA | 18385 | 91834 | 0.95 |
| UKB_AA | 280 | 1046 | 1.05 |
| UKB_SAS | 189 | 1269 | 1.01 |
| MGB_AA | 225 | 1481 | 1.02 |
| MGB_EA | 2967 | 27957 | 1 |
| MGB_HIS | 100 | 1352 | 0.99 |
| UPenn_EA | 380 | 6210 | 1.05 |
| UPenn_AA | 203 | 1719 | 1.14 |
| FARIVE_EA | 545 | 532 | 0.85 |
| RETROVE_EA | 400 | 400 | 1.08 |
| GAIT2_EA | 85 | 850 | 1.14 |
| BBJ_EAS | 507 | 178366 | 0.81 |
| MESA_EA | 94 | 2341 | 0.94 |
| MESA_AA | 72 | 1511 | 0.84 |
| MESA_HIS | 35 | 1389 | 0.72 |
| MARTHA12_EA | 893 | 3414 | 1.02 |
| **Overall** | **80690** | **1403644** | **0.97** |

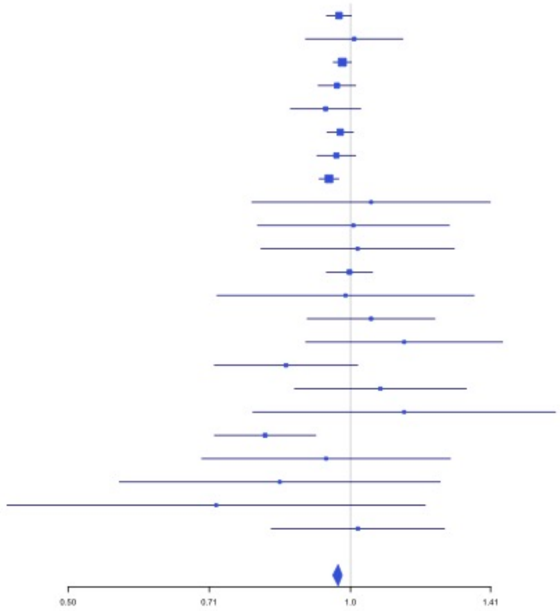


**Supplementary Figure 2.98** – rs2545774 (chr19:41287674) in locus 121 (RAB4B)

| Study | Cases | Controls | OR |
|---|---|---|---|
| INVENT-2019_EA | 15572 | 115280 | 0.95 |
| INVENT-2019_AA | 799 | 14353 | 0.95 |
| MVP_EA | 18952 | 411746 | 0.97 |
| MVP_AA | 5521 | 103791 | 1 |
| MVP_HIS | 1188 | 45634 | 1.02 |
| FinnGen_EA | 9176 | 209616 | 0.94 |
| EGP_EA | 4122 | 181553 | 0.93 |
| UKB_EA | 18385 | 91834 | 0.97 |
| UKB_SAS | 189 | 1269 | 0.86 |
| MGB_AA | 225 | 1481 | 1 |
| MGB_EA | 2967 | 27957 | 0.96 |
| MGB_HIS | 100 | 1352 | 1.08 |
| UPenn_EA | 380 | 6210 | 0.93 |
| UPenn_AA | 203 | 1719 | 1.07 |
| FARIVE_EA | 545 | 532 | 0.78 |
| BioME_EA | 200 | 8007 | 0.88 |
| BioME_AA | 382 | 6074 | 1.13 |
| BioME_HIS | 397 | 8992 | 0.99 |
| RETROVE_EA | 400 | 400 | 1.26 |
| GAIT2_EA | 85 | 850 | 1 |
| BBJ_EAS | 507 | 178366 | 1.06 |
| MESA_EA | 94 | 2341 | 0.9 |
| MESA_AA | 72 | 1511 | 0.7 |
| MESA_HIS | 35 | 1389 | 0.7 |
| **Overall** | **80496** | **1422257** | **0.96** |

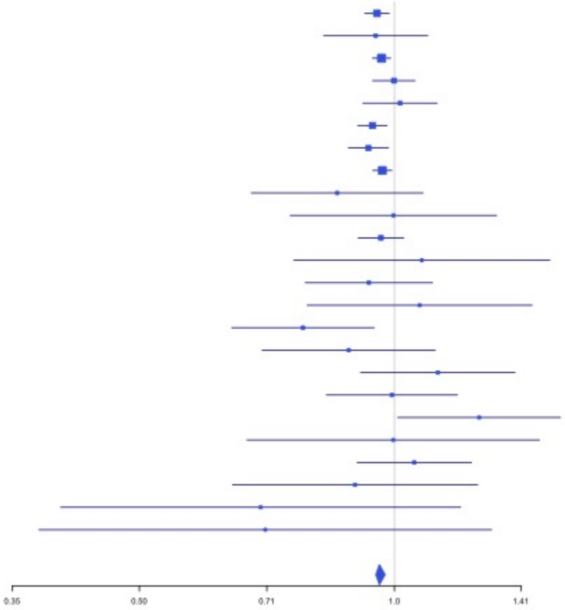

**Supplementary Figure 2.99** – rs8108474 (chr19:46301479) in locus 122 (RSPH6A)

| Study | Cases | Controls | OR |
|---|---|---|---|
| INVENT-2019_EA | 15572 | 115280 | 1.05 |
| INVENT-2019_AA | 799 | 14353 | 1.04 |
| MVP_EA | 18952 | 411746 | 1.04 |
| MVP_AA | 5521 | 103791 | 1.03 |
| MVP_HIS | 1188 | 45634 | 1.08 |
| FinnGen_EA | 9176 | 209616 | 1.05 |
| EGP_EA | 4122 | 181553 | 1 |
| UKB_EA | 18385 | 91834 | 1.05 |
| UKB_AA | 280 | 1046 | 1.01 |
| UKB_SAS | 189 | 1269 | 1.17 |
| MGB_AA | 225 | 1481 | 1 |
| MGB_EA | 2967 | 27957 | 1.04 |
| MGB_HIS | 100 | 1352 | 0.95 |
| UPenn_EA | 380 | 6210 | 0.93 |
| UPenn_AA | 203 | 1719 | 1.07 |
| FARIVE_EA | 545 | 532 | 0.88 |
| BioME_EA | 200 | 8007 | 1.04 |
| BioME_AA | 382 | 6074 | 0.93 |
| BioME_HIS | 397 | 8992 | 0.91 |
| RETROVE_EA | 400 | 400 | 1.01 |
| GAIT2_EA | 85 | 850 | 1.02 |
| BBJ_EAS | 507 | 178366 | 0.93 |
| MESA_EA | 94 | 2341 | 1.01 |
| MESA_AA | 72 | 1511 | 1.2 |
| MESA_HIS | 35 | 1389 | 1.15 |
| **Overall** | **80776** | **1423303** | **1.04** |

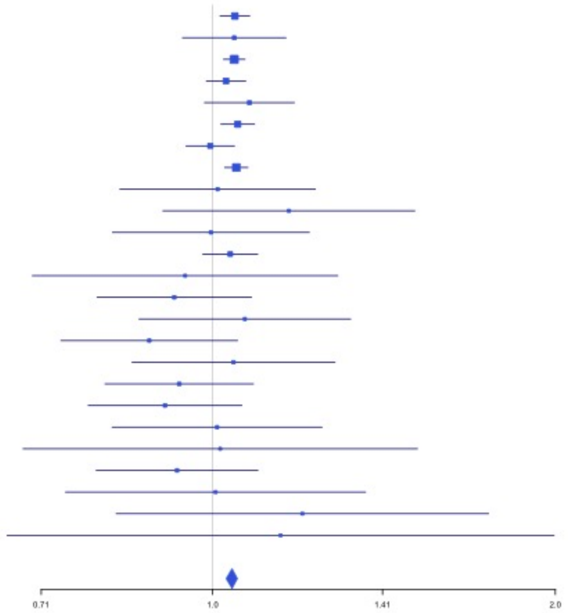


**Supplementary Figure 2.100** – rs2638282 (chr19:49213833) in locus 123 (FUT2;MAMSTR)

| Study | Cases | Controls | OR |
|---|---|---|---|
| INVENT-2019_EA | 15572 | 115280 | 1.04 |
| INVENT-2019_AA | 799 | 14353 | 1.07 |
| MVP_EA | 18952 | 411746 | 1.04 |
| MVP_AA | 5521 | 103791 | 1.06 |
| MVP_HIS | 1188 | 45634 | 1.01 |
| FinnGen_EA | 9176 | 209616 | 1.04 |
| EGP_EA | 4122 | 181553 | 1.05 |
| UKB_EA | 18385 | 91834 | 1.05 |
| UKB_AA | 280 | 1046 | 1.09 |
| UKB_SAS | 189 | 1269 | 0.95 |
| MGB_AA | 225 | 1481 | 1.01 |
| MGB_EA | 2967 | 27957 | 1.02 |
| MGB_HIS | 100 | 1352 | 1.18 |
| UPenn_EA | 380 | 6210 | 1.08 |
| UPenn_AA | 203 | 1719 | 0.86 |
| FARIVE_EA | 545 | 532 | 0.99 |
| BioME_EA | 200 | 8007 | 1 |
| BioME_AA | 382 | 6074 | 1.06 |
| BioME_HIS | 397 | 8992 | 1.06 |
| RETROVE_EA | 400 | 400 | 1.11 |
| GAIT2_EA | 85 | 850 | 1.21 |
| BBJ_EAS | 507 | 178366 | 10.41 |
| MESA_EA | 94 | 2341 | 1.08 |
| MESA_AA | 72 | 1511 | 1.11 |
| MESA_HIS | 35 | 1389 | 0.82 |
| MARTHA12_EA | 893 | 3414 | 1.06 |
| **Overall** | **81669** | **1426717** | **1.04** |

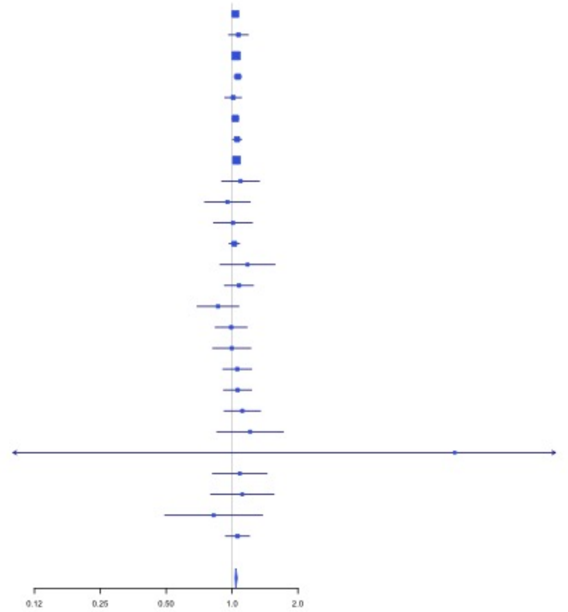

**Supplementary Figure 2.101** – rs1654425 (chr19:55538980) in locus 124 (GP6)

| Study | Cases | Controls | OR |
|---|---|---|---|
| INVENT-2019_EA | 15572 | 115280 | 0.93 |
| INVENT-2019_AA | 799 | 14353 | 0.83 |
| MVP_EA | 18952 | 411746 | 0.9 |
| MVP_AA | 5521 | 103791 | 0.95 |
| MVP_HIS | 1188 | 45634 | 0.86 |
| FinnGen_EA | 9176 | 209616 | 0.92 |
| EGP_EA | 4122 | 181553 | 0.93 |
| UKB_EA | 18385 | 91834 | 0.93 |
| UKB_SAS | 189 | 1269 | 0.7 |
| MGB_AA | 225 | 1481 | 0.79 |
| MGB_EA | 2967 | 27957 | 0.89 |
| MGB_HIS | 100 | 1352 | 1.04 |
| UPenn_EA | 380 | 6210 | 0.97 |
| UPenn_AA | 203 | 1719 | 0.84 |
| FARIVE_EA | 545 | 532 | 0.78 |
| BioME_EA | 200 | 8007 | 1.06 |
| BioME_AA | 382 | 6074 | 1.09 |
| BioME_HIS | 397 | 8992 | 0.87 |
| RETROVE_EA | 400 | 400 | 0.67 |
| GAIT2_EA | 85 | 850 | 1.09 |
| BBJ_EAS | 507 | 178366 | 1.3 |
| MESA_EA | 94 | 2341 | 0.82 |
| MESA_AA | 72 | 1511 | 1.07 |
| MESA_HIS | 35 | 1389 | 0.71 |
| MARTHA12_EA | 893 | 3414 | 0.96 |
| **Overall** | **81389** | **1425671** | **0.92** |

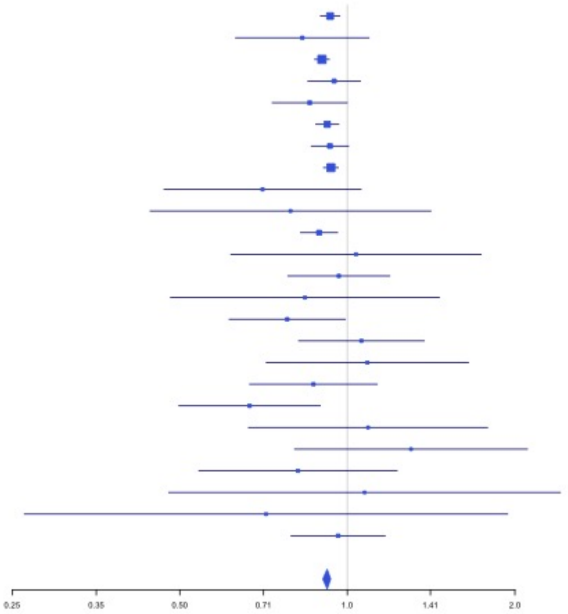


**Supplementary Figure 2.102** – rs2631628 (chr19:56645250) in locus 125 (ZNF787;ZNF444)

| Study | Cases | Controls | OR |
|---|---|---|---|
| INVENT-2019_AA | 799 | 14353 | 0.92 |
| MVP_EA | 18952 | 411746 | 0.9 |
| MVP_AA | 5521 | 103791 | 0.89 |
| MVP_HIS | 1188 | 45634 | 0.91 |
| FinnGen_EA | 9176 | 209616 | 0.9 |
| EGP_EA | 4122 | 181553 | 0.83 |
| UKB_EA | 18385 | 91834 | 0.87 |
| MGB_AA | 225 | 1481 | 1.23 |
| MGB_HIS | 100 | 1352 | 0.98 |
| UPenn_EA | 380 | 6210 | 0.5 |
| UPenn_AA | 203 | 1719 | 0.95 |
| BioME_EA | 200 | 8007 | 0.96 |
| BioME_AA | 382 | 6074 | 1.1 |
| BioME_HIS | 397 | 8992 | 0.9 |
| BBJ_EAS | 507 | 178366 | 1.02 |
| MESA_EA | 94 | 2341 | 0.78 |
| MESA_AA | 72 | 1511 | 1.21 |
| MESA_HIS | 35 | 1389 | 0.82 |
| **Overall** | **60738** | **1275969** | **0.9** |

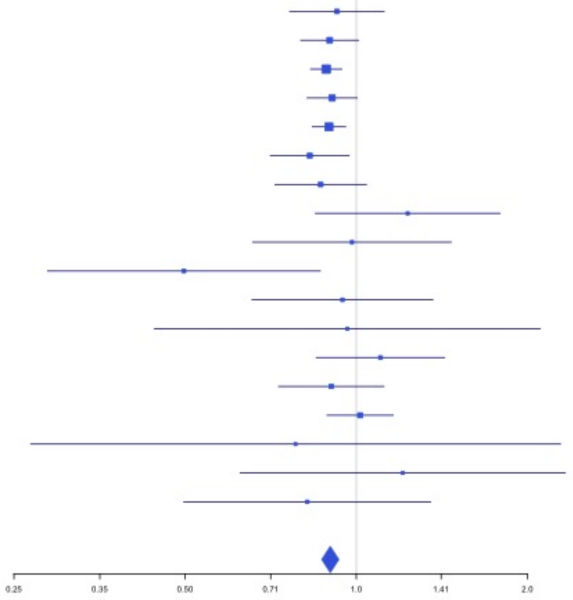

**Supplementary Figure 2.103** – rs117390891 (chr20:23168526) in locus 126 (LINC00656;NXT1)

| Study | Cases | Controls | OR |
|---|---|---|---|
| INVENT-2019_EA | 15572 | 115280 | 0.93 |
| INVENT-2019_AA | 799 | 14353 | 1.1 |
| MVP_EA | 18952 | 411746 | 0.9 |
| MVP_AA | 5521 | 103791 | 0.91 |
| MVP_HIS | 1188 | 45634 | 0.84 |
| FinnGen_EA | 9176 | 209616 | 0.91 |
| EGP_EA | 4122 | 181553 | 0.94 |
| UKB_EA | 18385 | 91834 | 0.93 |
| UKB_SAS | 189 | 1269 | 0.9 |
| MGB_AA | 225 | 1481 | 1.04 |
| MGB_EA | 2967 | 27957 | 1.03 |
| MGB_HIS | 100 | 1352 | 0.74 |
| UPenn_EA | 380 | 6210 | 0.96 |
| UPenn_AA | 203 | 1719 | 1.07 |
| FARIVE_EA | 545 | 532 | 0.92 |
| BioME_EA | 200 | 8007 | 1.08 |
| BioME_AA | 382 | 6074 | 0.97 |
| BioME_HIS | 397 | 8992 | 0.96 |
| RETROVE_EA | 400 | 400 | 0.9 |
| GAIT2_EA | 85 | 850 | 1.05 |
| BBJ_EAS | 507 | 178366 | 0.85 |
| MESA_EA | 94 | 2341 | 0.63 |
| MESA_AA | 72 | 1511 | 0.89 |
| MESA_HIS | 35 | 1389 | 1.58 |
| **Overall** | **80496** | **1422257** | **0.93** |

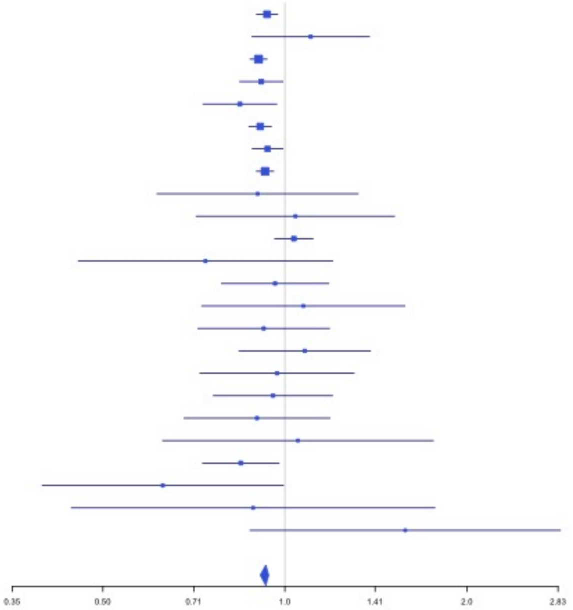


**Supplementary Figure 2.104** – rs6060288 (chr20:33772243) in locus 127 (MMP24-AS1-EDEM2)

| Study | Cases | Controls | OR |
|---|---|---|---|
| INVENT-2019_EA | 15572 | 115280 | 1.1 |
| INVENT-2019_AA | 799 | 14353 | 1.02 |
| MVP_EA | 18952 | 411746 | 1.15 |
| MVP_AA | 5521 | 103791 | 1.1 |
| MVP_HIS | 1188 | 45634 | 1.06 |
| FinnGen_EA | 9176 | 209616 | 1.12 |
| EGP_EA | 4122 | 181553 | 1.09 |
| UKB_EA | 18385 | 91834 | 1.11 |
| UKB_AA | 280 | 1046 | 1.24 |
| UKB_SAS | 189 | 1269 | 0.84 |
| MGB_AA | 225 | 1481 | 0.97 |
| MGB_EA | 2967 | 27957 | 1.05 |
| MGB_HIS | 100 | 1352 | 0.95 |
| UPenn_EA | 380 | 6210 | 0.87 |
| UPenn_AA | 203 | 1719 | 1.06 |
| FARIVE_EA | 545 | 532 | 1.24 |
| BioME_EA | 200 | 8007 | 1 |
| BioME_AA | 382 | 6074 | 1.04 |
| BioME_HIS | 397 | 8992 | 1.1 |
| RETROVE_EA | 400 | 400 | 1.12 |
| GAIT2_EA | 85 | 850 | 1.26 |
| BBJ_EAS | 507 | 178366 | 1.08 |
| MESA_EA | 94 | 2341 | 1.24 |
| MESA_AA | 72 | 1511 | 1.07 |
| MESA_HIS | 35 | 1389 | 1.11 |
| MARTHA12_EA | 893 | 3414 | 1.28 |
| **Overall** | **81669** | **1426717** | **1.12** |

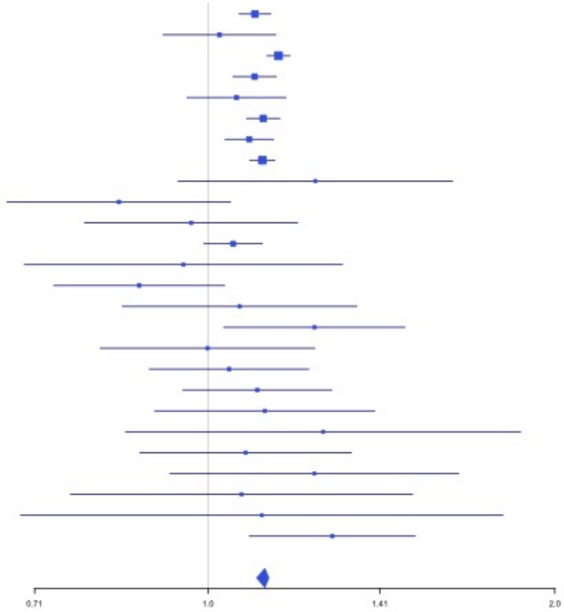

**Supplementary Figure 2.105** – rs3070580 (chr22:33165231) in locus 129 (SYN3)

| Study | Cases | Controls | OR |
|---|---|---|---|
| INVENT-2019_EA | 15572 | 115280 | 1.04 |
| INVENT-2019_AA | 799 | 14353 | 1.21 |
| MVP_EA | 18952 | 411746 | 1.05 |
| MVP_HIS | 1188 | 45634 | 1.05 |
| FinnGen_EA | 9176 | 209616 | 1.04 |
| EGP_EA | 4122 | 181553 | 1.08 |
| UKB_EA | 18385 | 91834 | 1.02 |
| UKB_SAS | 189 | 1269 | 0.87 |
| UPenn_EA | 380 | 6210 | 1.21 |
| UPenn_AA | 203 | 1719 | 0.62 |
| FARIVE_EA | 545 | 532 | 1.15 |
| BBJ_EAS | 507 | 178366 | 1.13 |
| **Overall** | **70018** | **1258112** | **1.04** |

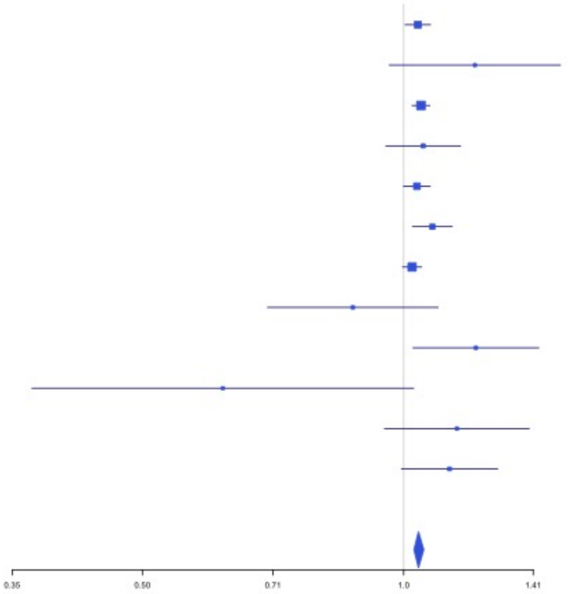


**Supplementary Figure 2.106** – rs9611844 (chr22:43115776) in locus 130 (A4GALT)

| Study | Cases | Controls | OR |
|---|---|---|---|
| INVENT-2019_EA | 15572 | 115280 | 1.04 |
| INVENT-2019_AA | 799 | 14353 | 1.08 |
| MVP_EA | 18952 | 411746 | 1.11 |
| MVP_AA | 5521 | 103791 | 1.15 |
| MVP_HIS | 1188 | 45634 | 1.11 |
| FinnGen_EA | 9176 | 209616 | 1.08 |
| EGP_EA | 4122 | 181553 | 1.17 |
| UKB_EA | 18385 | 91834 | 1.06 |
| MGB_AA | 225 | 1481 | 0.82 |
| MGB_EA | 2967 | 27957 | 1.03 |
| MGB_HIS | 100 | 1352 | 1.41 |
| UPenn_EA | 380 | 6210 | 0.89 |
| UPenn_AA | 203 | 1719 | 0.94 |
| FARIVE_EA | 545 | 532 | 1.15 |
| BioME_EA | 200 | 8007 | 1.07 |
| BioME_AA | 382 | 6074 | 1.19 |
| BioME_HIS | 397 | 8992 | 1.02 |
| RETROVE_EA | 400 | 400 | 1.15 |
| GAIT2_EA | 85 | 850 | 1.89 |
| BBJ_EAS | 507 | 178366 | 1.01 |
| MESA_EA | 94 | 2341 | 0.89 |
| MESA_AA | 72 | 1511 | 0.7 |
| MESA_HIS | 35 | 1389 | 1.16 |
| **Overall** | **80307** | **1420988** | **1.09** |

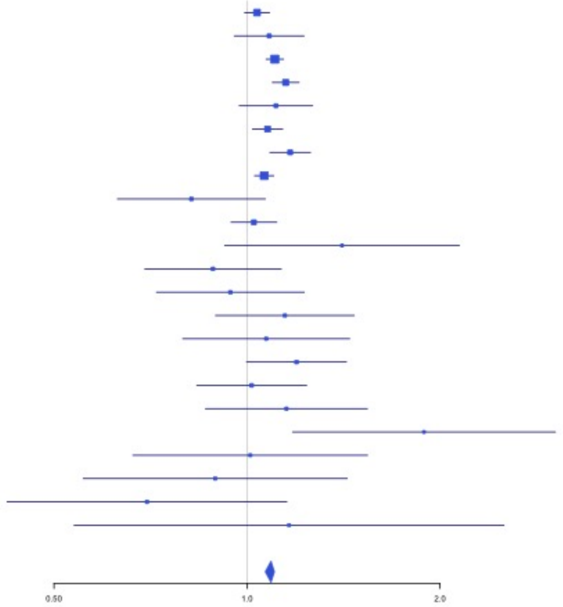

**Supplementary Figure 2.107** – rs12388118 (chr23:11380742) in locus 131 (ARHGAP6)

| Study | Cases | Controls | OR |
|---|---|---|---|
| INVENT-2019_EA | 8705 | 52578 | 1.07 |
| MVP_EA | 18952 | 411746 | 1.06 |
| MVP_AA | 5521 | 103791 | 1.02 |
| MVP_HIS | 1188 | 45634 | 0.98 |
| EGP_EA | 4122 | 181553 | 1 |
| UKBB_EA | 280 | 1046 | 1.05 |
| FARIVE_EA | 545 | 532 | 1.23 |
| BBJ_EAS | 507 | 178366 | 1.01 |
| MGB_AA | 225 | 1481 | 1.06 |
| MGB_EA | 2967 | 27957 | 1.05 |
| MGB_HIS | 100 | 1352 | 1.11 |
| **Overall** | **43112** | **1006036** | **1.05** |

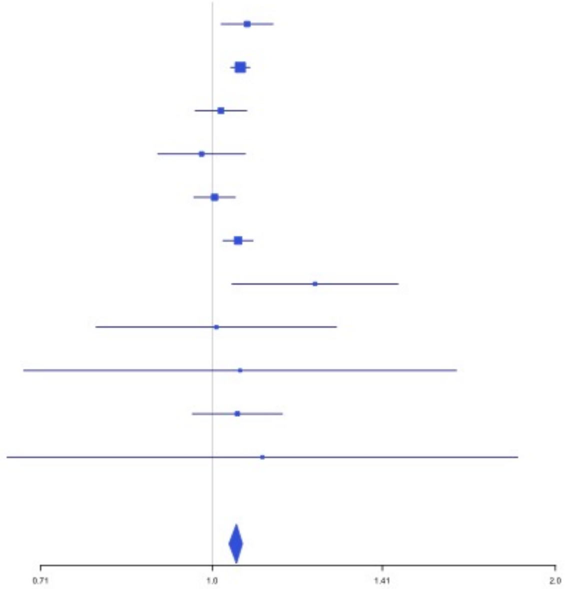


**Supplementary Figure 2.108** – rs6632109 (chr23:34894638) in locus 132 (TMEM47;FAM47B)

| Study | Cases | Controls | OR |
|---|---|---|---|
| INVENT-2019_EA | 8705 | 52578 | 1.04 |
| MVP_EA | 18952 | 411746 | 1.04 |
| MVP_AA | 5521 | 103791 | 1.09 |
| MVP_HIS | 1188 | 45634 | 1.01 |
| FinnGen_EA | 9176 | 209616 | 1.04 |
| EGP_EA | 4122 | 181553 | 1.05 |
| UKBB_EA | 280 | 1046 | 1.01 |
| UKBB_SAS | 189 | 1269 | 1 |
| FARIVE_EA | 545 | 532 | 0.84 |
| RETROVE_EA | 400 | 400 | 1.09 |
| GAIT2_EA | 85 | 850 | 1.03 |
| BBJ_EAS | 507 | 178366 | 1 |
| MGB_AA | 225 | 1481 | 1.09 |
| MGB_EA | 2967 | 27957 | 1.05 |
| MGB_HIS | 100 | 1352 | 1.03 |
| **Overall** | **52962** | **1218171** | **1.04** |

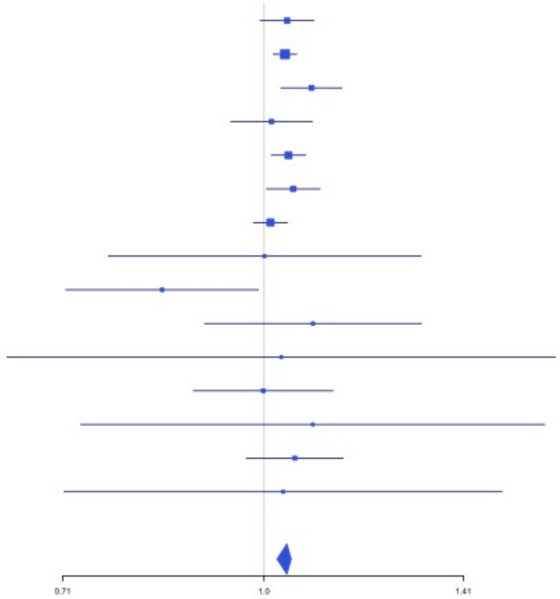

**Supplementary Figure 2.109** – rs3002416 (chr23:39710195) in locus 133 (MIR1587;BCOR)

| Study | Cases | Controls | OR |
|---|---|---|---|
| INVENT-2019_EA | 8705 | 52578 | 0.91 |
| MVP_EA | 18952 | 411746 | 0.94 |
| MVP_AA | 5521 | 103791 | 0.91 |
| MVP_HIS | 1188 | 45634 | 1 |
| FinnGen_EA | 9176 | 209616 | 0.96 |
| EGP_EA | 4122 | 181553 | 0.98 |
| UKBB_EA | 280 | 1046 | 0.93 |
| UKBB_SAS | 189 | 1269 | 0.93 |
| FARIVE_EA | 545 | 532 | 0.77 |
| RETROVE_EA | 400 | 400 | 0.87 |
| GAIT2_EA | 85 | 850 | 1.28 |
| BBJ_EAS | 507 | 178366 | 0.99 |
| MGB_AA | 225 | 1481 | 0.86 |
| MGB_EA | 2967 | 27957 | 0.98 |
| MGB_HIS | 100 | 1352 | 0.96 |
| **Overall** | **52962** | **1218171** | **0.94** |

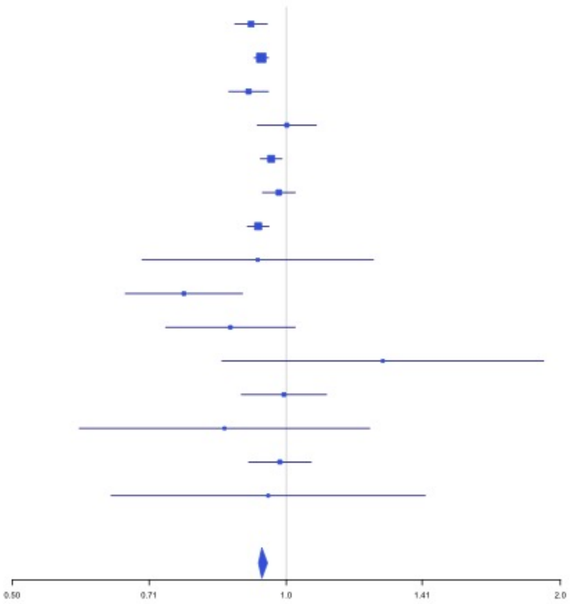


**Supplementary Figure 2.110** – rs6048 (chr23:138633280) in locus 134 (F9)

| Study | Cases | Controls | OR |
|---|---|---|---|
| INVENT-2019_EA | 8705 | 52578 | 1.09 |
| MVP_EA | 18952 | 411746 | 1.07 |
| MVP_AA | 5521 | 103791 | 1.1 |
| MVP_HIS | 1188 | 45634 | 1.04 |
| FinnGen_EA | 9176 | 209616 | 1.05 |
| EGP_EA | 4122 | 181553 | 1.07 |
| UKBB_EA | 280 | 1046 | 1.07 |
| FARIVE_EA | 545 | 532 | 1.07 |
| RETROVE_EA | 400 | 400 | 1.22 |
| GAIT2_EA | 85 | 850 | 1.29 |
| BBJ_EAS | 507 | 178366 | 0.87 |
| MGB_AA | 225 | 1481 | 0.83 |
| MGB_EA | 2967 | 27957 | 1.02 |
| MGB_HIS | 100 | 1352 | 1.55 |
| **Overall** | **52773** | **1216902** | **1.07** |

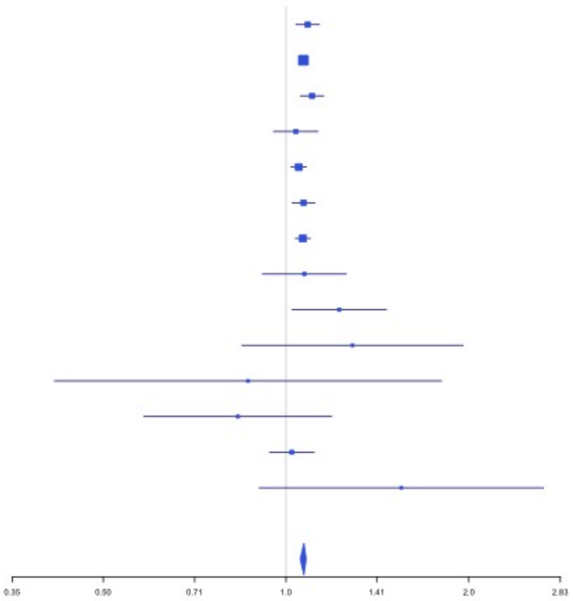

**Supplementary Figure 2.111** – rs2084408 (chr23:154346709) in locus 135 (BRCC3)

| Study | Cases | Controls | OR |
|---|---|---|---|
| INVENT-2019_EA | 8705 | 52578 | 0.94 |
| MVP_EA | 18952 | 411746 | 0.92 |
| MVP_AA | 5521 | 103791 | 0.95 |
| MVP_HIS | 1188 | 45634 | 0.93 |
| FinnGen_EA | 9176 | 209616 | 0.96 |
| EGP_EA | 4122 | 181553 | 0.94 |
| UKBB_AA | 18385 | 91834 | 0.57 |
| UKBB_EA | 280 | 1046 | 0.95 |
| UKBB_SAS | 189 | 1269 | 0.87 |
| FARIVE_EA | 545 | 532 | 0.91 |
| RETROVE_EA | 400 | 400 | 0.92 |
| GAIT2_EA | 85 | 850 | 0.68 |
| BBJ_EAS | 507 | 178366 | 0.96 |
| MGB_AA | 225 | 1481 | 1.01 |
| MGB_EA | 2967 | 27957 | 0.97 |
| MGB_HIS | 100 | 1352 | 1.03 |
| **Overall** | **71347** | **1310005** | **0.94** |

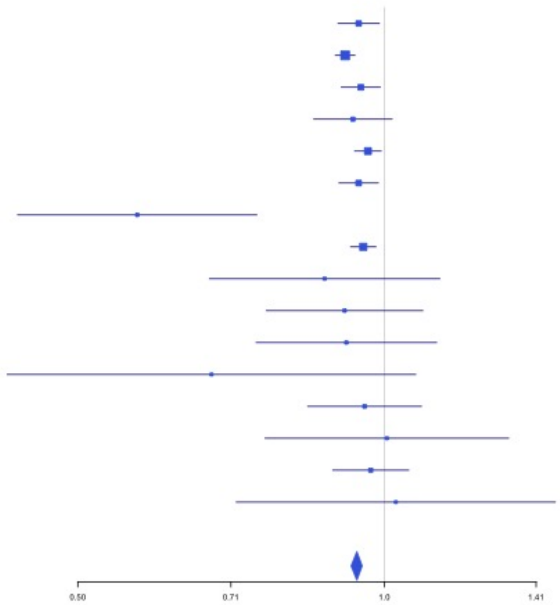
