## Supplemental Figure 3 for "Cross-Ancestry Investigation of Venous Thromboembolism Genomic Predictors"

### Supplementary Figure 3.
### Forest plots of European analysis lead variants.


3.1 rs9442571 (chr1:9349611) in locus 1 (H6PD;SPSB1) . . . 3
3.2 rs551176418 (chr1:27107263) in locus 2 (ARID1A) . . . 3
3.3 rs12753797 (chr1:77981959) in locus 3 (AK5) . . . 4
3.4 rs4970834 (chr1:109814880) in locus 4 (CELSR2) . . . 4
3.5 rs1775815 (chr1:118148830) in locus 5 (TENT5C) . . . 5
3.6 rs1267881263 (chr1:150496127) in locus 6 (FALEC;ADAMTSL4) . . . 5
3.7 rs76798800 (chr1:154994978) in locus 7 (DCST2) . . . 6
3.8 rs3557 (chr1:161188893) in locus 8 (FCER1G) . . . 6
3.9 rs6025 (chr1:169519049) in locus 9 (F5) . . . 7
3.10 rs4540639 (chr1:192104320) in locus 11 (LINC01680;RGS18) . . . 7
3.11 rs2842700 (chr1:207282149) in locus 13 (C4BPA) . . . 8
3.12 rs3811444 (chr1:248039451) in locus 14 (TRIM58) . . . 8
3.13 rs1867312 (chr2:68619981) in locus 17 (PLEK) . . . 9
3.14 rs1158867 (chr2:128177377) in locus 18 (PROC) . . . 9
3.15 rs1912846 (chr2:188325444) in locus 19 (CALCRL;TFPI) . . . 10
3.16 rs78872368 (chr2:198545250) in locus 20 (RFTN2;MARS2) . . . 10
3.17 rs1047891 (chr2:211540507) in locus 21 (CPS1) . . . 11
3.18 rs13412535 (chr2:224874874) in locus 22 (SERPINE2) . . . 11
3.19 rs13084580 (chr3:39188182) in locus 24 (CSRNP1) . . . 12
3.20 rs11130326 (chr3:52771920) in locus 25 (NEK4) . . . 12
3.21 rs9866664 (chr3:90437988) in locus 26 (EPHA3;NONE) . . . 13
3.22 rs28479320 (chr3:93752529) in locus 27 (ARL13B) . . . 13
3.23 rs9854955 (chr3:156795525) in locus 30 (LEKR1;LINC00880) . . . 14
3.24 rs710446 (chr3:186459927) in locus 32 (KNG1) . . . 14
3.25 rs3796159 (chr3:194790434) in locus 33 (XXYLT1) . . . 15
3.26 rs72976751 (chr4:86725959) in locus 37 (ARHGAP24) . . . 15
3.27 rs35225200 (chr4:103146888) in locus 38 (BANK1;SLC39A8) . . . 16
3.28 rs2066864 (chr4:155525695) in locus 39 (FGG) . . . 16
3.29 rs3756011 (chr4:187206249) in locus 40 (F11) . . . 17
3.30 rs112367053 (chr5:28379046) in locus 41 (LINC02103;LSP1P3) . . . 17
3.31 rs16867574 (chr5:38708554) in locus 42 (OSMR-AS1) . . . 18
3.32 rs4235677 (chr5:72925091) in locus 43 (ARHGEF28) . . . 18
3.33 rs251339 (chr5:96235038) in locus 44 (ERAP2) . . . 19
3.34 rs147133967 (chr5:132426851) in locus 45 (HSPA4) . . . 19
3.35 rs214059 (chr6:25536937) in locus 46 (CARMIL1) . . . 20
3.36 rs2394251 (chr6:29943688) in locus 47 (HCG9) . . . 20
3.37 rs28383307 (chr6:32586755) in locus 48 (HLA-DRB1;HLA-DQA1) . . . 21
3.38 rs145294670 (chr6:34622561) in locus 49 (ILRUN) . . . 21
3.39 rs2754251 (chr6:88385949) in locus 51 (AKIRIN2) . . . 22
3.40 rs9390460 (chr6:147694334) in locus 52 (STXBP5) . . . 22
3.41 rs1513275 (chr7:28259233) in locus 53 (JAZF1-AS1) . . . 23
3.42 rs370720 (chr8:9168146) in locus 57 (LOC101929128;LOC157273) . . . 23
3.43 rs2726950 (chr8:27806764) in locus 60 (SCARA5) . . . 24
3.44 rs6993770 (chr8:106581528) in locus 62 (ZFPM2) . . . 24
3.45 rs1491336705 (chr8:108295235) in locus 63 (ANGPT1) . . . 25
3.46 rs10820606 (chr9:99192919) in locus 65 (ZNF367;HABP4) . . . 25
3.47 rs687289 (chr9:136137106) in locus 66 (ABO) . . . 26
3.48 rs1243184 (chr10:21931937) in locus 68 (MLLT10) . . . 26
3.49 rs10763665 (chr10:28771491) in locus 70 (LINC02652) . . . 27
3.50 rs78707713 (chr10:71245276) in locus 72 (TSPAN15) . . . 27

3.51 rs57866767 (chr10:96023077) in locus 74 (PLCE1) . . . . 28
3.52 rs10886430 (chr10:121010256) in locus 75 (GRK5) . . . . 28
3.53 rs7122100 (chr11:10732560) in locus 76 (IRAG1;CTR9) . . . . 29
3.54 rs12800654 (chr11:16243320) in locus 77 (SOX6) . . . . 29
3.55 rs4755476 (chr11:32980428) in locus 78 (QSER1) . . . . 30
3.56 rs1799963 (chr11:46761055) in locus 79 (F2) . . . . 30
3.57 rs141798115 (chr11:56875074) in locus 80 (OR5AK4P;LRRC55) . . . . 31
3.58 rs55777218 (chr11:60070946) in locus 81 (MS4A4A) . . . . 31
3.59 rs174551 (chr11:61573684) in locus 82 (FADS1) . . . . 32
3.60 rs1145656 (chr11:73305859) in locus 83 (FAM168A) . . . . 32
3.61 rs1997547 (chr11:114002744) in locus 84 (ZBTB16) . . . . 33
3.62 rs35257264 (chr11:126296816) in locus 85 (ST3GAL4) . . . . 33
3.63 rs7135039 (chr12:6160614) in locus 87 (VWF) . . . . 34
3.64 rs7311483 (chr12:9053661) in locus 88 (A2ML1;PHC1) . . . . 34
3.65 rs4759076 (chr12:54729872) in locus 89 (COPZ1) . . . . 35
3.66 rs142351376 (chr12:104136288) in locus 90 (STAB2) . . . . 35
3.67 rs56750693 (chr12:112144586) in locus 91 (ACAD10) . . . . 36
3.68 rs2851436 (chr12:123667354) in locus 92 (MPHOSPH9) . . . . 36
3.69 rs3211752 (chr13:113787459) in locus 94 (F10) . . . . 37
3.70 rs8004077 (chr14:58846731) in locus 95 (ARID4A;TOMM20L) . . . . 37
3.71 rs2127869 (chr14:65794352) in locus 96 (LINC02324;MIR4708) . . . . 38
3.72 rs61990092 (chr14:92289301) in locus 97 (TC2N) . . . . 38
3.73 rs28929474 (chr14:94844947) in locus 98 (SERPINA1) . . . . 39
3.74 rs4906229 (chr14:103057511) in locus 99 (LINC02323;RCOR1) . . . . 39
3.75 rs141866277 (chr15:43950699) in locus 100 (PPIP5K1P1-CATSPER2) . . . . 40
3.76 rs340009 (chr15:60899639) in locus 101 (RORA-AS1) . . . . 40
3.77 rs7183672 (chr15:96101018) in locus 103 (LINC00924;LOC105369212) . . . . 41
3.78 rs12443808 (chr16:30996871) in locus 106 (HSD3B7) . . . . 41
3.79 rs1346057617 (chr16:75434183) in locus 108 (CFDP1) . . . . 42
3.80 rs62044142 (chr16:81868968) in locus 109 (PLCG2) . . . . 42
3.81 rs28634651 (chr16:88553198) in locus 111 (ZFPM1) . . . . 43
3.82 rs4790311 (chr17:1979188) in locus 112 (SMG6) . . . . 43
3.83 rs11078659 (chr17:6903944) in locus 113 (ALOX12-AS1) . . . . 44
3.84 rs536327 (chr17:27857698) in locus 114 (TAOK1) . . . . 44
3.85 rs9468 (chr17:44101563) in locus 115 (MAPT) . . . . 45
3.86 rs1801690 (chr17:64208285) in locus 116 (APOH) . . . . 45
3.87 rs573431210 (chr17:67276383) in locus 117 (ABCA5) . . . . 46
3.88 rs8109681 (chr19:10738836) in locus 119 (SLC44A2) . . . . 46
3.89 rs2545774 (chr19:41287674) in locus 121 (RAB4B) . . . . 47
3.90 rs8108474 (chr19:46301479) in locus 122 (RSPH6A) . . . . 47
3.91 rs646327 (chr19:49209851) in locus 123 (FUT2) . . . . 48
3.92 rs1654425 (chr19:55538980) in locus 124 (GP6) . . . . 48
3.93 rs117390891 (chr20:23168526) in locus 126 (LINC00656;NXT1) . . . . 49
3.94 rs6087685 (chr20:33777612) in locus 127 (MMP24-AS1-EDEM2) . . . . 49
3.95 rs3070580 (chr22:33165231) in locus 129 (SYN3) . . . . 50
3.96 rs9611844 (chr22:43115776) in locus 130 (A4GALT) . . . . 50
3.97 rs12388118 (chr23:11380742) in locus 131 (ARHGAP6) . . . . 51
3.98 rs3002416 (chr23:39710195) in locus 133 (MIR1587;BCOR) . . . . 51
3.99 rs6048 (chr23:138633280) in locus 134 (F9) . . . . 52
3.100 rs4898406 (chr23:154273269) in locus 135 (FUNDC2) . . . . 52

**Supplementary Figure 3.1** – rs9442571 (chr1:9349611) in locus 1 (H6PD ;SPSB1)

| Study | Cases | Controls | OR |
|---|---|---|---|
| INVENT-2019_EA | 15572 | 115280 | 1.11 |
| MVP_EA | 18952 | 411746 | 1.05 |
| FinnGen_EA | 9176 | 209616 | 1.05 |
| EGP_EA | 4122 | 181553 | 1.03 |
| UKB_EA | 18385 | 91834 | 1.04 |
| MGB_EA | 2967 | 27957 | 1.01 |
| UPenn_EA | 380 | 6210 | 1.03 |
| FARIVE_EA | 545 | 532 | 0.91 |
| BioME_EA | 200 | 8007 | 0.98 |
| RETROVE_EA | 400 | 400 | 1.76 |
| GAIT2_EA | 85 | 850 | 1.84 |
| MESA_EA | 94 | 2341 | 0.86 |
| **European** | **70878** | **1056326** | **1.05** |

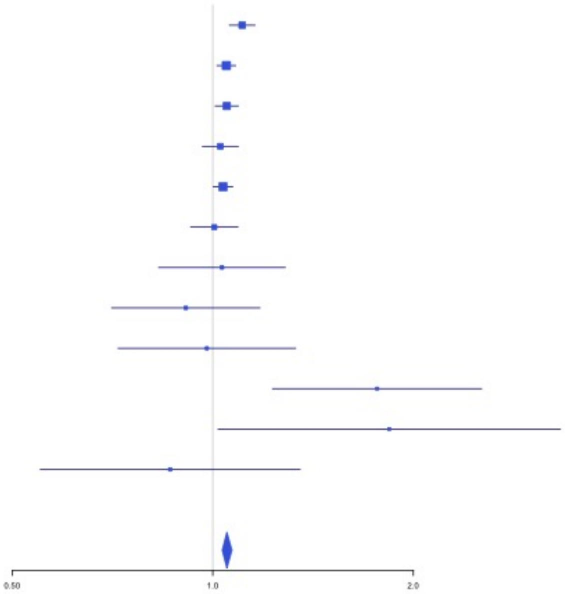


**Supplementary Figure 3.2** – rs551176418 (chr1:27107263) in locus 2 (ARID1A)

| Study | Cases | Controls | OR |
|---|---|---|---|
| MVP_EA | 18952 | 411746 | 1.08 |
| FinnGen_EA | 9176 | 209616 | 1.1 |
| UKB_EA | 18385 | 91834 | 1.07 |
| UPenn_EA | 380 | 6210 | 0.95 |
| FARIVE_EA | 545 | 532 | 1.14 |
| MARTHA12_EA | 893 | 3414 | 0.96 |
| **European** | **48331** | **723352** | **1.08** |

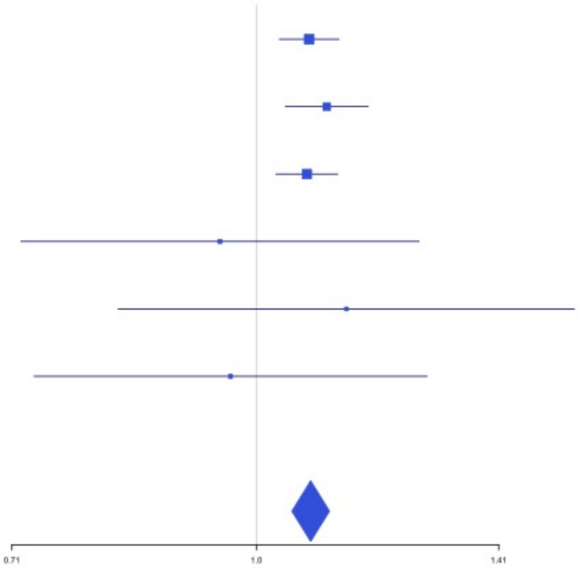

**Supplementary Figure 3.3** – rs12753797 (chr1:77981959) in locus 3 (AK5)

| Study | Cases | Controls | OR |
|---|---|---|---|
| INVENT-2019_EA | 15572 | 115280 | 1.06 |
| MVP_EA | 18952 | 411746 | 1.03 |
| FinnGen_EA | 9176 | 209616 | 1.05 |
| EGP_EA | 4122 | 181553 | 1.02 |
| UKB_EA | 18385 | 91834 | 1.06 |
| MGB_EA | 2967 | 27957 | 0.98 |
| UPenn_EA | 380 | 6210 | 1.17 |
| FARIVE_EA | 545 | 532 | 0.96 |
| BioME_EA | 200 | 8007 | 1.03 |
| RETROVE_EA | 400 | 400 | 0.93 |
| GAIT2_EA | 85 | 850 | 1.16 |
| MESA_EA | 94 | 2341 | 1.17 |
| **European** | **70878** | **1056326** | **1.04** |

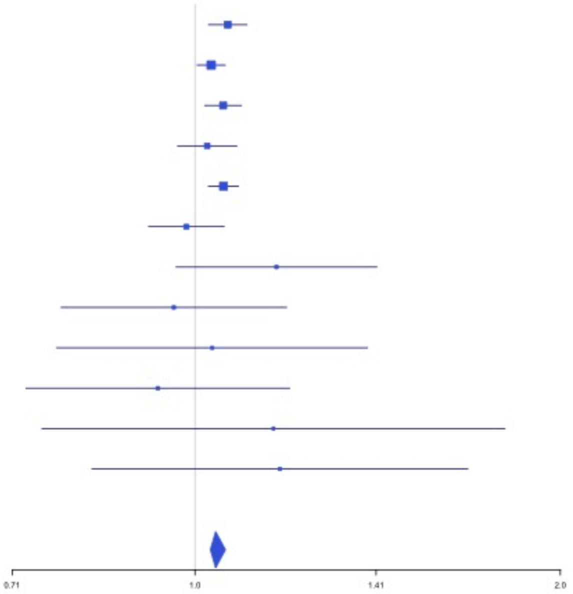


**Supplementary Figure 3.4** – rs4970834 (chr1:109814880) in locus 4 (CELSR2)

| Study | Cases | Controls | OR |
|---|---|---|---|
| INVENT-2019_EA | 15572 | 115280 | 1.05 |
| MVP_EA | 18952 | 411746 | 1.05 |
| FinnGen_EA | 9176 | 209616 | 1.06 |
| EGP_EA | 4122 | 181553 | 1.01 |
| UKB_EA | 18385 | 91834 | 1.06 |
| MGB_EA | 2967 | 27957 | 0.99 |
| UPenn_EA | 380 | 6210 | 1.01 |
| FARIVE_EA | 545 | 532 | 0.97 |
| BioME_EA | 200 | 8007 | 1.03 |
| RETROVE_EA | 400 | 400 | 1.27 |
| GAIT2_EA | 85 | 850 | 1.56 |
| MESA_EA | 94 | 2341 | 1.22 |
| MARTHA12_EA | 893 | 3414 | 1.02 |
| **European** | **71771** | **1059740** | **1.05** |

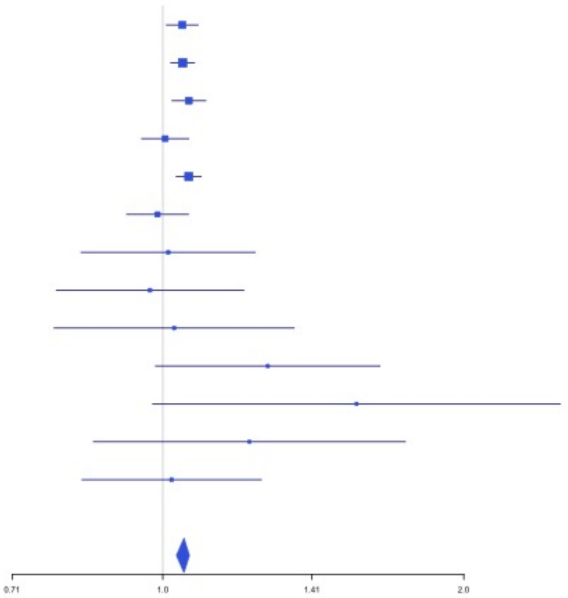

**Supplementary Figure 3.5** – rs1775815 (chr1:118148830) in locus 5 (TENT5C)

| Study | Cases | Controls | OR |
|---|---|---|---|
| INVENT-2019_EA | 15572 | 115280 | 1.04 |
| MVP_EA | 18952 | 411746 | 1.05 |
| FinnGen_EA | 9176 | 209616 | 1.02 |
| EGP_EA | 4122 | 181553 | 1.05 |
| UKB_EA | 18385 | 91834 | 1.03 |
| MGB_EA | 2967 | 27957 | 1.06 |
| UPenn_EA | 380 | 6210 | 1.02 |
| FARIVE_EA | 545 | 532 | 1.25 |
| BioME_EA | 200 | 8007 | 0.94 |
| RETROVE_EA | 400 | 400 | 1.16 |
| GAIT2_EA | 85 | 850 | 0.88 |
| MESA_EA | 94 | 2341 | 1.04 |
| **European** | **70878** | **1056326** | **1.04** |

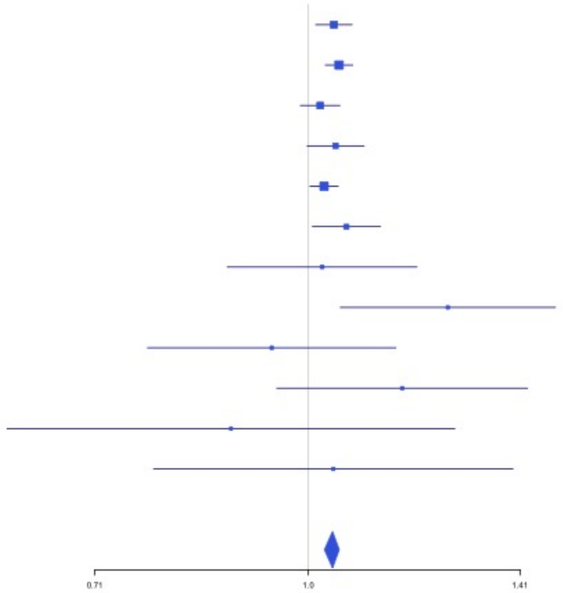


**Supplementary Figure 3.6** – rs1267881263 (chr1:150496127) in locus 6 (FALEC;ADAMTSL4)

| Study | Cases | Controls | OR |
|---|---|---|---|
| MVP_EA | 18952 | 411746 | 1.06 |
| UKB_EA | 18385 | 91834 | 1.04 |
| UPenn_EA | 380 | 6210 | 1.01 |
| FARIVE_EA | 545 | 532 | 0.98 |
| MARTHA12_EA | 893 | 3414 | 1.21 |
| **European** | **39155** | **513736** | **1.05** |

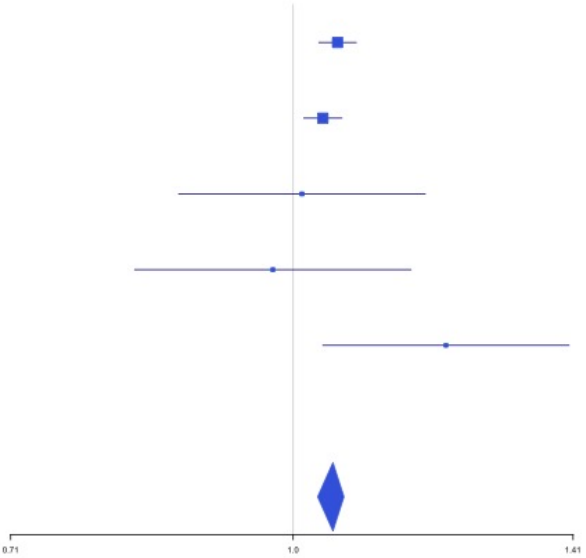

**Supplementary Figure 3.7** – rs76798800 (chr1:154994978) in locus 7 (DCST2)

| Study | Cases | Controls | OR |
|---|---|---|---|
| INVENT-2019_EA | 15572 | 115280 | 1.02 |
| MVP_EA | 18952 | 411746 | 1.03 |
| FinnGen_EA | 9176 | 209616 | 1.05 |
| EGP_EA | 4122 | 181553 | 1.06 |
| UKB_EA | 18385 | 91834 | 1.04 |
| MGB_EA | 2967 | 27957 | 1.07 |
| UPenn_EA | 380 | 6210 | 0.87 |
| FARIVE_EA | 545 | 532 | 1.14 |
| BioME_EA | 200 | 8007 | 0.9 |
| RETROVE_EA | 400 | 400 | 1.38 |
| GAIT2_EA | 85 | 850 | 1.7 |
| MESA_EA | 94 | 2341 | 1.25 |
| MARTHA12_EA | 893 | 3414 | 0.91 |
| **European** | **71771** | **1059740** | **1.04** |

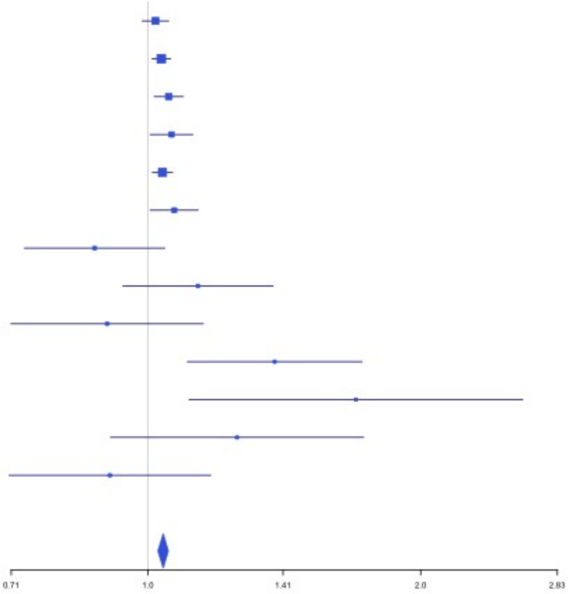


**Supplementary Figure 3.8** – rs3557 (chr1:161188893) in locus 8 (FCER1G)

| Study | Cases | Controls | OR |
|---|---|---|---|
| INVENT-2019_EA | 15572 | 115280 | 1.05 |
| MVP_EA | 18952 | 411746 | 1.09 |
| FinnGen_EA | 9176 | 209616 | 1.04 |
| EGP_EA | 4122 | 181553 | 1.01 |
| UKB_EA | 18385 | 91834 | 1.05 |
| MGB_EA | 2967 | 27957 | 1.01 |
| UPenn_EA | 380 | 6210 | 1.12 |
| FARIVE_EA | 545 | 532 | 1.26 |
| BioME_EA | 200 | 8007 | 1.14 |
| RETROVE_EA | 400 | 400 | 1.42 |
| GAIT2_EA | 85 | 850 | 0.6 |
| MESA_EA | 94 | 2341 | 1.17 |
| MARTHA12_EA | 893 | 3414 | 1.37 |
| **European** | **71771** | **1059740** | **1.06** |

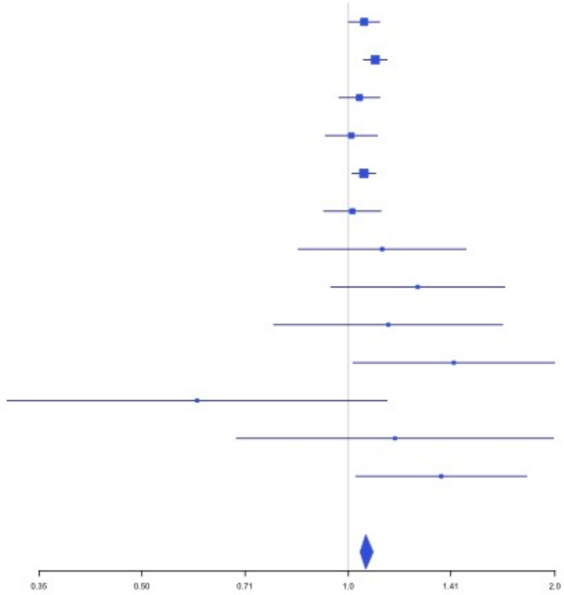

**Supplementary Figure 3.9** – rs6025 (chr1:169519049) in locus 9 (F5)

| Study | Cases | Controls | OR |
|---|---|---|---|
| INVENT-2019_EA | 15572 | 115280 | 2.39 |
| MVP_EA | 18952 | 411746 | 3.12 |
| FinnGen_EA | 9176 | 209616 | 3.24 |
| EGP_EA | 4122 | 181553 | 3.07 |
| UKB_EA | 18385 | 91834 | 3.14 |
| MGB_EA | 2967 | 27957 | 1.98 |
| UPenn_EA | 380 | 6210 | 2.09 |
| FARIVE_EA | 545 | 532 | 2.45 |
| BioME_EA | 200 | 8007 | 2.72 |
| RETROVE_EA | 400 | 400 | 2.27 |
| MESA_EA | 94 | 2341 | 1.85 |
| **European** | **70793** | **1055476** | **2.98** |

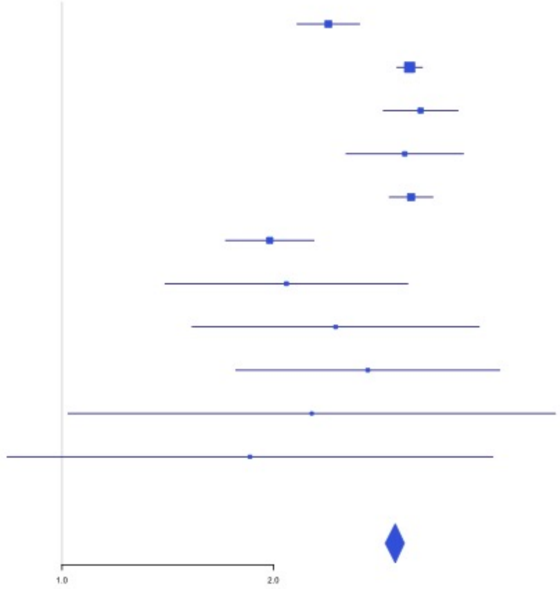


**Supplementary Figure 3.10** – rs4540639 (chr1:192104320) in locus 11 (LINC01680;RGS18)

| Study | Cases | Controls | OR |
|---|---|---|---|
| INVENT-2019_EA | 15572 | 115280 | 1.03 |
| MVP_EA | 18952 | 411746 | 1.05 |
| FinnGen_EA | 9176 | 209616 | 1.05 |
| EGP_EA | 4122 | 181553 | 1.04 |
| UKB_EA | 18385 | 91834 | 1.02 |
| MGB_EA | 2967 | 27957 | 1.02 |
| UPenn_EA | 380 | 6210 | 1.07 |
| FARIVE_EA | 545 | 532 | 1.06 |
| BioME_EA | 200 | 8007 | 0.85 |
| RETROVE_EA | 400 | 400 | 1.15 |
| GAIT2_EA | 85 | 850 | 0.97 |
| MESA_EA | 94 | 2341 | 1.27 |
| **European** | **70878** | **1056326** | **1.04** |

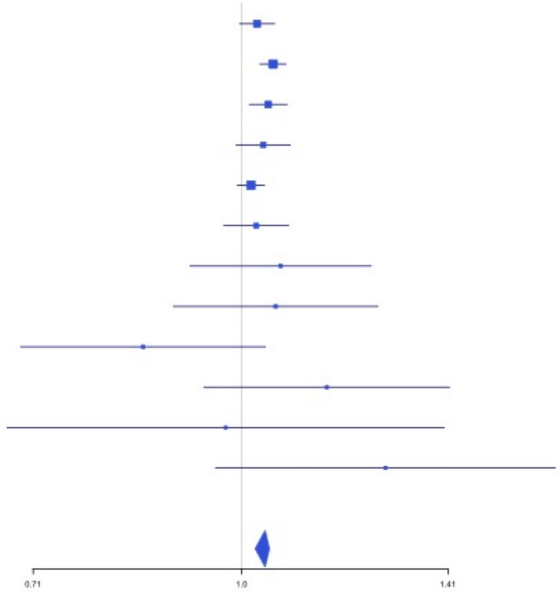

**Supplementary Figure 3.11** – rs2842700 (chr1:207282149) in locus 13 (C4BPA)

| Study | Cases | Controls | OR |
|---|---|---|---|
| INVENT-2019_EA | 15572 | 115280 | 1.18 |
| MVP_EA | 18952 | 411746 | 1.16 |
| FinnGen_EA | 9176 | 209616 | 1.13 |
| EGP_EA | 4122 | 181553 | 1.02 |
| UKB_EA | 18385 | 91834 | 1.09 |
| MGB_EA | 2967 | 27957 | 1.07 |
| UPenn_EA | 380 | 6210 | 1.15 |
| FARIVE_EA | 545 | 532 | 1.45 |
| BioME_EA | 200 | 8007 | 0.9 |
| RETROVE_EA | 400 | 400 | 1.12 |
| GAIT2_EA | 85 | 850 | 1.54 |
| MESA_EA | 94 | 2341 | 1.32 |
| **European** | **70878** | **1056326** | **1.12** |

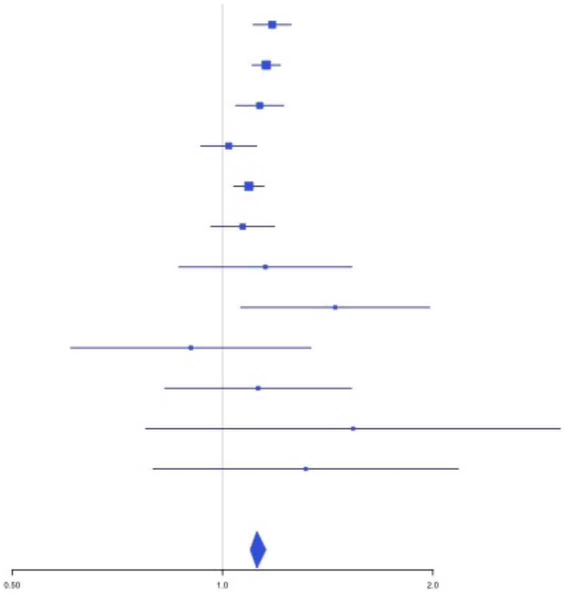


**Supplementary Figure 3.12** – rs3811444 (chr1:248039451) in locus 14 (TRIM58)

| Study | Cases | Controls | OR |
|---|---|---|---|
| INVENT-2019_EA | 15572 | 115280 | 0.99 |
| MVP_EA | 18952 | 411746 | 0.94 |
| FinnGen_EA | 9176 | 209616 | 0.97 |
| EGP_EA | 4122 | 181553 | 0.95 |
| UKB_EA | 18385 | 91834 | 0.97 |
| MGB_EA | 2967 | 27957 | 0.97 |
| UPenn_EA | 380 | 6210 | 0.86 |
| FARIVE_EA | 545 | 532 | 0.85 |
| BioME_EA | 200 | 8007 | 0.76 |
| RETROVE_EA | 400 | 400 | 0.85 |
| GAIT2_EA | 85 | 850 | 1.09 |
| MESA_EA | 94 | 2341 | 0.8 |
| MARTHA12_EA | 893 | 3414 | 0.92 |
| **European** | **71771** | **1059740** | **0.96** |

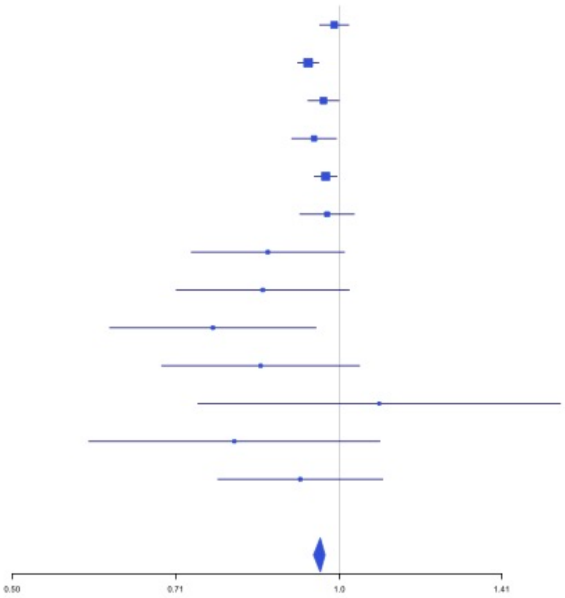

**Supplementary Figure 3.13** – rs1867312 (chr2:68619981) in locus 17 (PLEK)

| Study | Cases | Controls | OR |
|---|---|---|---|
| INVENT-2019_EA | 15572 | 115280 | 0.96 |
| MVP_EA | 18952 | 411746 | 0.96 |
| FinnGen_EA | 9176 | 209616 | 0.95 |
| EGP_EA | 4122 | 181553 | 0.98 |
| UKB_EA | 18385 | 91834 | 0.93 |
| MGB_EA | 2967 | 27957 | 0.94 |
| UPenn_EA | 380 | 6210 | 0.91 |
| FARIVE_EA | 545 | 532 | 0.92 |
| BioME_EA | 200 | 8007 | 0.83 |
| RETROVE_EA | 400 | 400 | 0.88 |
| GAIT2_EA | 85 | 850 | 0.81 |
| MESA_EA | 94 | 2341 | 1.17 |
| MARTHA12_EA | 893 | 3414 | 0.88 |
| **European** | **71771** | **1059740** | **0.95** |

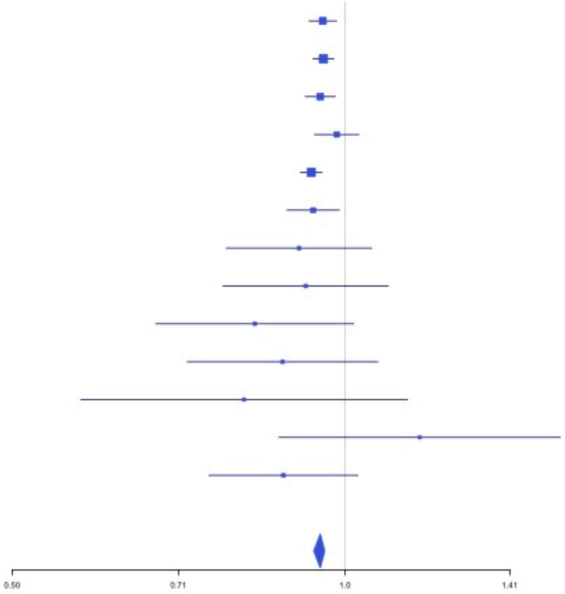


**Supplementary Figure 3.14** – rs1158867 (chr2:128177377) in locus 18 (PROC)

| Study | Cases | Controls | OR |
|---|---|---|---|
| INVENT-2019_EA | 15572 | 115280 | 0.95 |
| MVP_EA | 18952 | 411746 | 0.92 |
| FinnGen_EA | 9176 | 209616 | 0.94 |
| EGP_EA | 4122 | 181553 | 0.92 |
| UKB_EA | 18385 | 91834 | 0.95 |
| MGB_EA | 2967 | 27957 | 0.96 |
| UPenn_EA | 380 | 6210 | 0.82 |
| FARIVE_EA | 545 | 532 | 0.82 |
| BioME_EA | 200 | 8007 | 1.04 |
| RETROVE_EA | 400 | 400 | 0.86 |
| GAIT2_EA | 85 | 850 | 1.04 |
| MESA_EA | 94 | 2341 | 1.26 |
| MARTHA12_EA | 893 | 3414 | 0.9 |
| **European** | **71771** | **1059740** | **0.94** |

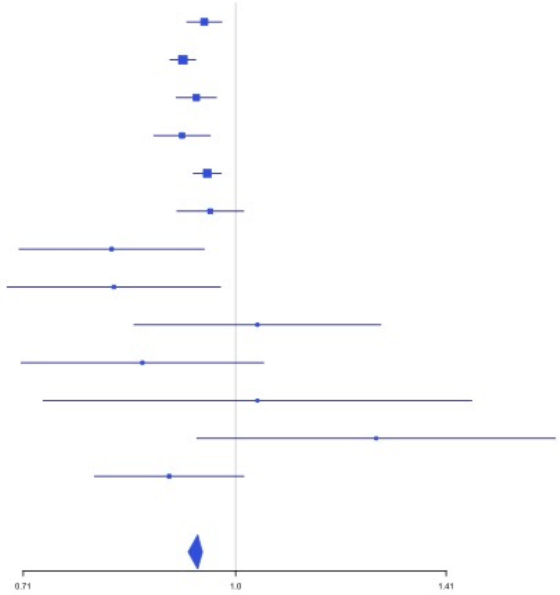

**Supplementary Figure 3.15** – rs1912846 (chr2:188325444) in locus 19 (CALCRL ;TFPI)

| Study | Cases | Controls | OR |
|---|---|---|---|
| INVENT-2019_EA | 15572 | 115280 | 0.97 |
| MVP_EA | 18952 | 411746 | 0.95 |
| FinnGen_EA | 9176 | 209616 | 0.98 |
| EGP_EA | 4122 | 181553 | 0.96 |
| UKB_EA | 18385 | 91834 | 0.97 |
| MGB_EA | 2967 | 27957 | 0.96 |
| UPenn_EA | 380 | 6210 | 0.95 |
| FARIVE_EA | 545 | 532 | 1 |
| BioME_EA | 200 | 8007 | 0.91 |
| RETROVE_EA | 400 | 400 | 0.87 |
| GAIT2_EA | 85 | 850 | 0.86 |
| MESA_EA | 94 | 2341 | 0.85 |
| MARTHA12_EA | 893 | 3414 | 0.93 |
| **European** | **71771** | **1059740** | **0.96** |

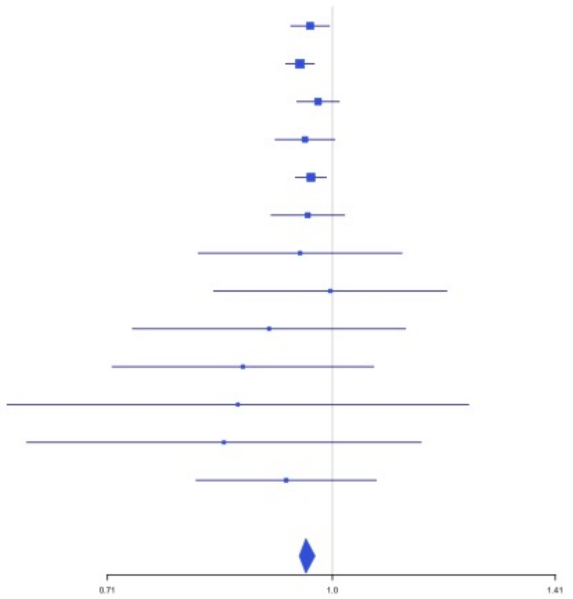


**Supplementary Figure 3.16** – rs78872368 (chr2:198545250) in locus 20 (RFTN2 ;MARS2)

| Study | Cases | Controls | OR |
|---|---|---|---|
| INVENT-2019_EA | 15572 | 115280 | 0.95 |
| MVP_EA | 18952 | 411746 | 0.96 |
| FinnGen_EA | 9176 | 209616 | 0.96 |
| EGP_EA | 4122 | 181553 | 0.97 |
| UKB_EA | 18385 | 91834 | 0.96 |
| MGB_EA | 2967 | 27957 | 0.95 |
| UPenn_EA | 380 | 6210 | 0.84 |
| FARIVE_EA | 545 | 532 | 1.01 |
| BioME_EA | 200 | 8007 | 0.97 |
| RETROVE_EA | 400 | 400 | 0.95 |
| GAIT2_EA | 85 | 850 | 0.85 |
| MESA_EA | 94 | 2341 | 0.88 |
| MARTHA12_EA | 893 | 3414 | 0.98 |
| **European** | **71771** | **1059740** | **0.96** |

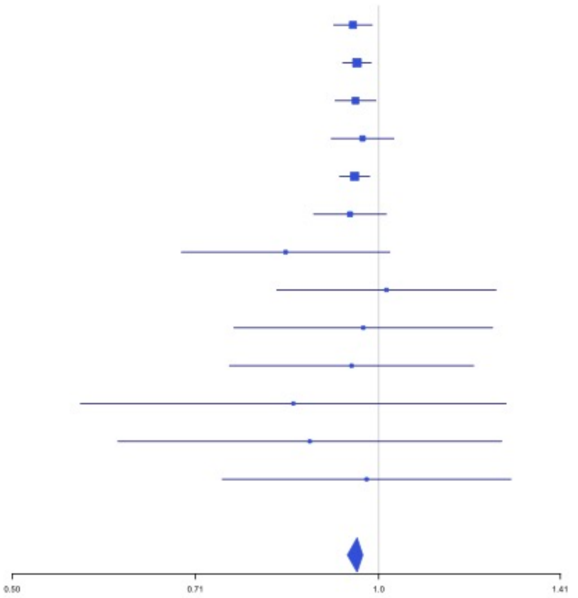

**Supplementary Figure 3.17** – rs1047891 (chr2:211540507) in locus 21 (CPS1)

| Study | Cases | Controls | OR |
|---|---|---|---|
| INVENT-2019_EA | 15572 | 115280 | 1.04 |
| MVP_EA | 18952 | 411746 | 1.05 |
| FinnGen_EA | 9176 | 209616 | 1.06 |
| EGP_EA | 4122 | 181553 | 1.05 |
| UKB_EA | 18385 | 91834 | 1.04 |
| MGB_EA | 2967 | 27957 | 0.99 |
| UPenn_EA | 380 | 6210 | 0.88 |
| FARIVE_EA | 545 | 532 | 0.84 |
| BioME_EA | 200 | 8007 | 1.08 |
| RETROVE_EA | 400 | 400 | 0.96 |
| GAIT2_EA | 85 | 850 | 1.43 |
| MESA_EA | 94 | 2341 | 1.02 |
| **European** | **70878** | **1056326** | **1.04** |

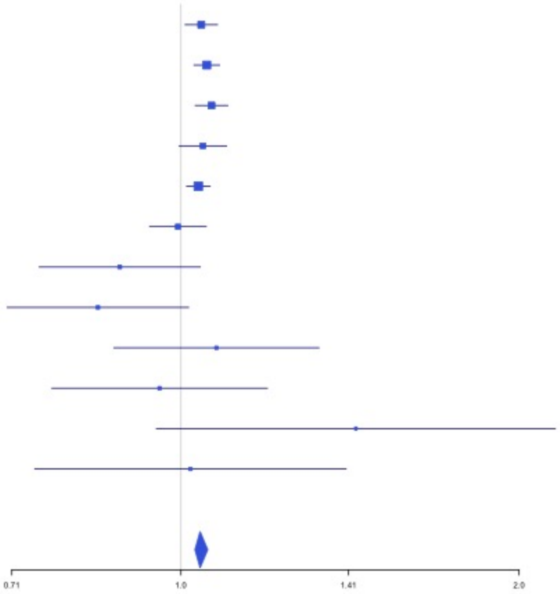


**Supplementary Figure 3.18** – rs13412535 (chr2:224874874) in locus 22 (SERPINE2)

| Study | Cases | Controls | OR |
|---|---|---|---|
| INVENT-2019_EA | 15572 | 115280 | 1.07 |
| MVP_EA | 18952 | 411746 | 1.07 |
| FinnGen_EA | 9176 | 209616 | 1.05 |
| EGP_EA | 4122 | 181553 | 1.02 |
| UKB_EA | 18385 | 91834 | 1.05 |
| MGB_EA | 2967 | 27957 | 1.05 |
| UPenn_EA | 380 | 6210 | 0.99 |
| FARIVE_EA | 545 | 532 | 1.01 |
| BioME_EA | 200 | 8007 | 1.2 |
| RETROVE_EA | 400 | 400 | 0.98 |
| GAIT2_EA | 85 | 850 | 0.79 |
| MESA_EA | 94 | 2341 | 0.95 |
| **European** | **70878** | **1056326** | **1.05** |

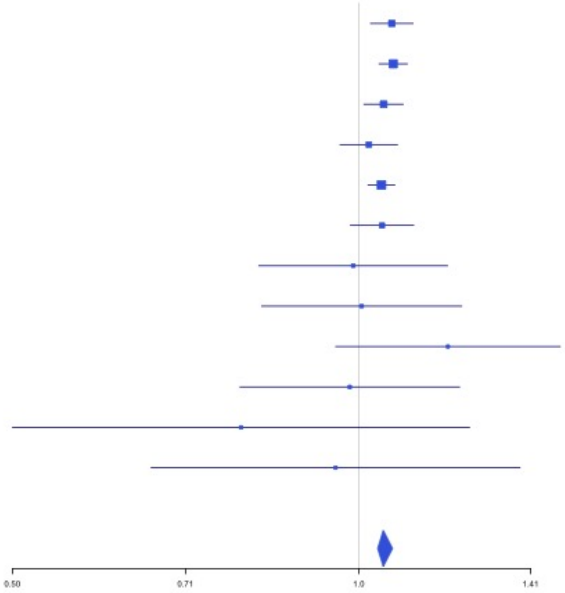

**Supplementary Figure 3.19** – rs13084580 (chr3:39188182) in locus 24 (CSRNP1)

| Study | Cases | Controls | OR |
|---|---|---|---|
| INVENT-2019_EA | 15572 | 115280 | 1.08 |
| MVP_EA | 18952 | 411746 | 1.1 |
| FinnGen_EA | 9176 | 209616 | 1.12 |
| EGP_EA | 4122 | 181553 | 1.05 |
| UKB_EA | 18385 | 91834 | 1.05 |
| MGB_EA | 2967 | 27957 | 1.07 |
| UPenn_EA | 380 | 6210 | 1.02 |
| FARIVE_EA | 545 | 532 | 1.32 |
| BioME_EA | 200 | 8007 | 1.09 |
| RETROVE_EA | 400 | 400 | 1.44 |
| GAIT2_EA | 85 | 850 | 0.98 |
| MESA_EA | 94 | 2341 | 1.04 |
| MARTHA12_EA | 893 | 3414 | 1.2 |
| **European** | **71771** | **1059740** | **1.08** |

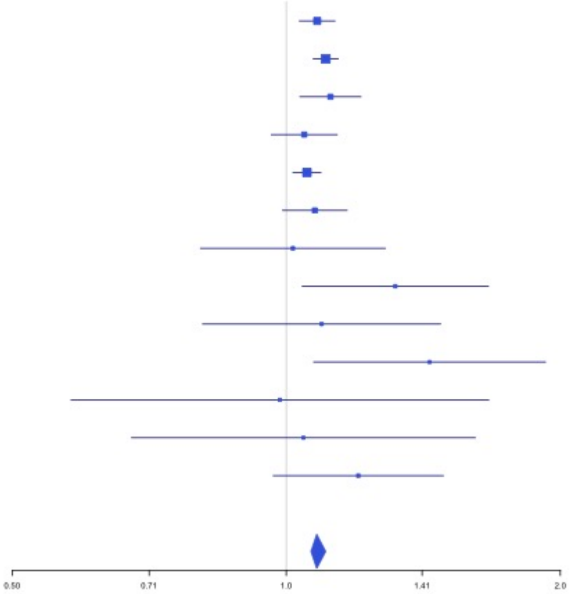


**Supplementary Figure 3.20** – rs11130326 (chr3:52771920) in locus 25 (NEK4)

| Study | Cases | Controls | OR |
|---|---|---|---|
| INVENT-2019_EA | 15572 | 115280 | 1.05 |
| MVP_EA | 18952 | 411746 | 1.04 |
| FinnGen_EA | 9176 | 209616 | 1.05 |
| UPenn_EA | 380 | 6210 | 0.94 |
| FARIVE_EA | 545 | 532 | 0.95 |
| MARTHA12_EA | 893 | 3414 | 0.99 |
| **European** | **45518** | **746798** | **1.04** |

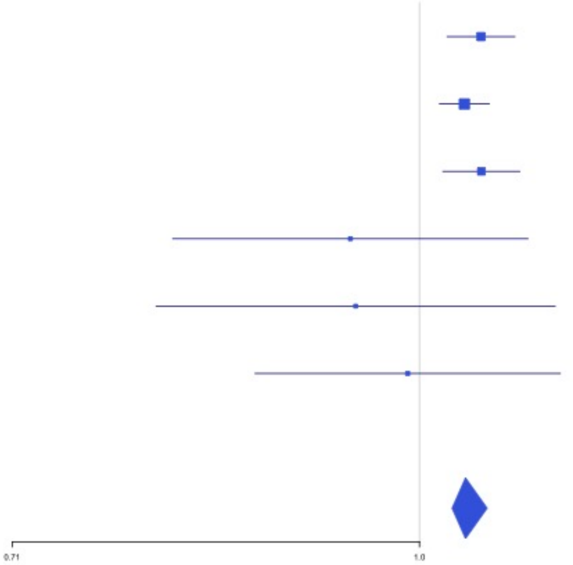

**Supplementary Figure 3.21** – rs9866664 (chr3:90437988) in locus 26 (EPHA3;NONE)

| Study | Cases | Controls | OR |
|---|---|---|---|
| INVENT-2019_EA | 15572 | 115280 | 1.05 |
| MVP_EA | 18952 | 411746 | 1.06 |
| FinnGen_EA | 9176 | 209616 | 1.05 |
| EGP_EA | 4122 | 181553 | 1.06 |
| UKB_EA | 18385 | 91834 | 1.04 |
| MGB_EA | 2967 | 27957 | 1.02 |
| UPenn_EA | 380 | 6210 | 0.96 |
| FARIVE_EA | 545 | 532 | 1.21 |
| BioME_EA | 200 | 8007 | 1.02 |
| RETROVE_EA | 400 | 400 | 1.02 |
| GAIT2_EA | 85 | 850 | 0.89 |
| MESA_EA | 94 | 2341 | 1.27 |
| **European** | **70878** | **1056326** | **1.05** |

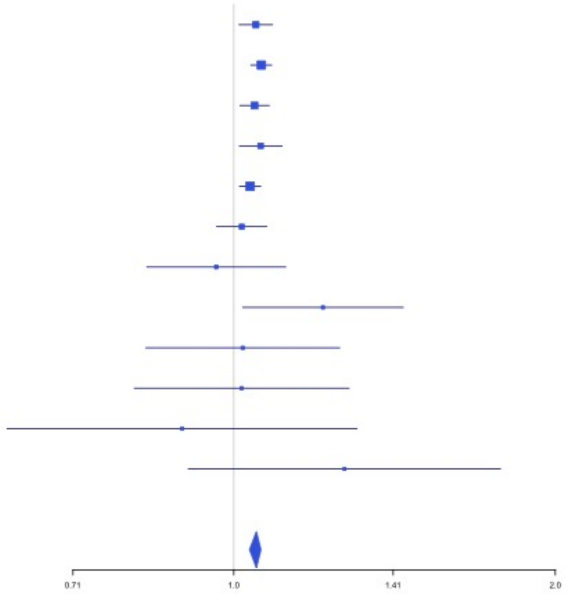


**Supplementary Figure 3.22** – rs28479320 (chr3:93752529) in locus 27 (ARL13B)

| Study | Cases | Controls | OR |
|---|---|---|---|
| INVENT-2019_EA | 15572 | 115280 | 0.94 |
| MVP_EA | 18952 | 411746 | 0.83 |
| FinnGen_EA | 9176 | 209616 | 0.91 |
| EGP_EA | 4122 | 181553 | 1.07 |
| UKB_EA | 18385 | 91834 | 0.87 |
| MGB_EA | 2967 | 27957 | 0.86 |
| UPenn_EA | 380 | 6210 | 1.08 |
| FARIVE_EA | 545 | 532 | 0.78 |
| BioME_EA | 200 | 8007 | 0.76 |
| RETROVE_EA | 400 | 400 | 0.68 |
| GAIT2_EA | 85 | 850 | 0.7 |
| MESA_EA | 94 | 2341 | 0.55 |
| **European** | **70878** | **1056326** | **0.87** |

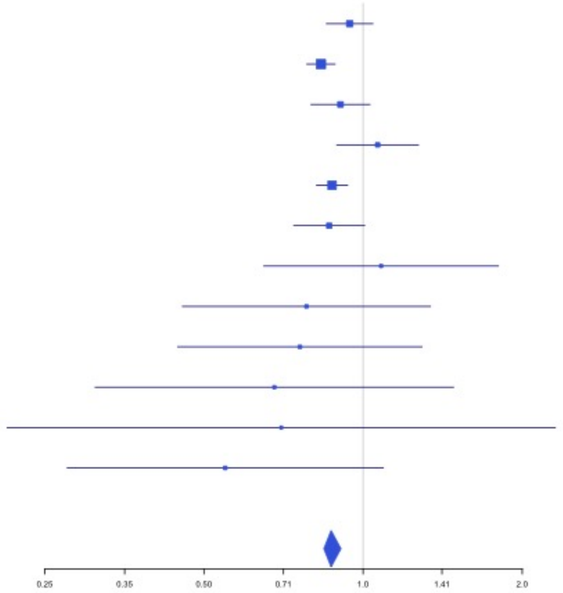

**Supplementary Figure 3.23** – rs9854955 (chr3:156795525) in locus 30 (LEKR1 ;LINC00880)

| Study | Cases | Controls | OR |
|---|---|---|---|
| INVENT-2019_EA | 15572 | 115280 | 1.04 |
| MVP_EA | 18952 | 411746 | 1.03 |
| FinnGen_EA | 9176 | 209616 | 1.06 |
| EGP_EA | 4122 | 181553 | 1.04 |
| UKB_EA | 18385 | 91834 | 1.03 |
| MGB_EA | 2967 | 27957 | 1.07 |
| UPenn_EA | 380 | 6210 | 1.02 |
| FARIVE_EA | 545 | 532 | 1.03 |
| BioME_EA | 200 | 8007 | 1.04 |
| RETROVE_EA | 400 | 400 | 1.06 |
| GAIT2_EA | 85 | 850 | 1.15 |
| MESA_EA | 94 | 2341 | 1.2 |
| MARTHA12_EA | 893 | 3414 | 1.15 |
| **European** | **71771** | **1059740** | **1.04** |

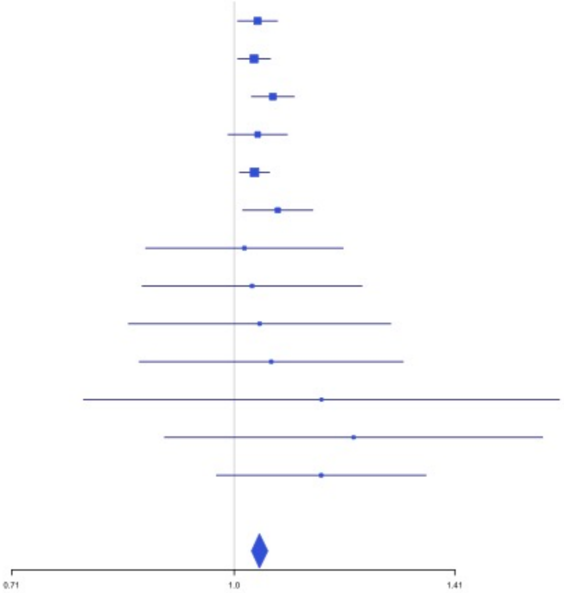


**Supplementary Figure 3.24** – rs710446 (chr3:186459927) in locus 32 (KNG1)

| Study | Cases | Controls | OR |
|---|---|---|---|
| INVENT-2019_EA | 15572 | 115280 | 0.94 |
| MVP_EA | 18952 | 411746 | 0.97 |
| FinnGen_EA | 9176 | 209616 | 0.94 |
| EGP_EA | 4122 | 181553 | 0.96 |
| UKB_EA | 18385 | 91834 | 0.96 |
| MGB_EA | 2967 | 27957 | 0.95 |
| UPenn_EA | 380 | 6210 | 0.91 |
| FARIVE_EA | 545 | 532 | 0.83 |
| BioME_EA | 200 | 8007 | 1.04 |
| RETROVE_EA | 400 | 400 | 0.86 |
| GAIT2_EA | 85 | 850 | 1.04 |
| MESA_EA | 94 | 2341 | 1.15 |
| MARTHA12_EA | 893 | 3414 | 0.85 |
| **European** | **71771** | **1059740** | **0.96** |

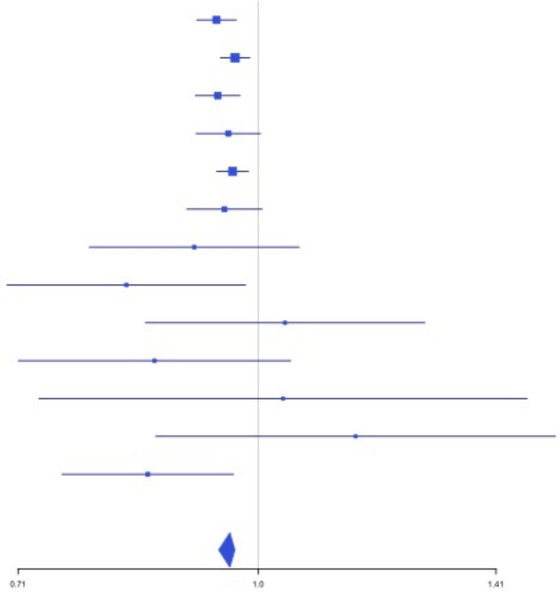

**Supplementary Figure 3.25** – rs3796159 (chr3:194790434) in locus 33 (XXYLT1)

| Study | Cases | Controls | OR |
|---|---|---|---|
| INVENT-2019_EA | 15572 | 115280 | 1.06 |
| MVP_EA | 18952 | 411746 | 1.07 |
| FinnGen_EA | 9176 | 209616 | 1.04 |
| UKB_EA | 18385 | 91834 | 1.05 |
| MGB_EA | 2967 | 27957 | 1 |
| UPenn_EA | 380 | 6210 | 1.38 |
| FARIVE_EA | 545 | 532 | 1.14 |
| BioME_EA | 200 | 8007 | 0.87 |
| RETROVE_EA | 400 | 400 | 1.05 |
| GAIT2_EA | 85 | 850 | 0.93 |
| MESA_EA | 94 | 2341 | 1.09 |
| **European** | **66756** | **874773** | **1.05** |

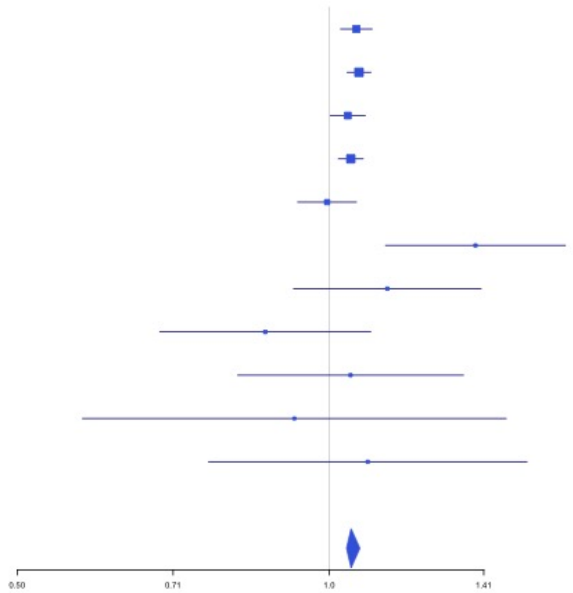


**Supplementary Figure 3.26** – rs72976751 (chr4:86725959) in locus 37 (ARHGAP24)

| Study | Cases | Controls | OR |
|---|---|---|---|
| INVENT-2019_EA | 15572 | 115280 | 1.05 |
| MVP_EA | 18952 | 411746 | 1.05 |
| FinnGen_EA | 9176 | 209616 | 1.06 |
| EGP_EA | 4122 | 181553 | 1.12 |
| UKB_EA | 18385 | 91834 | 1.07 |
| MGB_EA | 2967 | 27957 | 1.04 |
| UPenn_EA | 380 | 6210 | 0.9 |
| FARIVE_EA | 545 | 532 | 1.13 |
| BioME_EA | 200 | 8007 | 1.18 |
| RETROVE_EA | 400 | 400 | 1.02 |
| GAIT2_EA | 85 | 850 | 1.45 |
| MESA_EA | 94 | 2341 | 1.28 |
| **European** | **70878** | **1056326** | **1.06** |

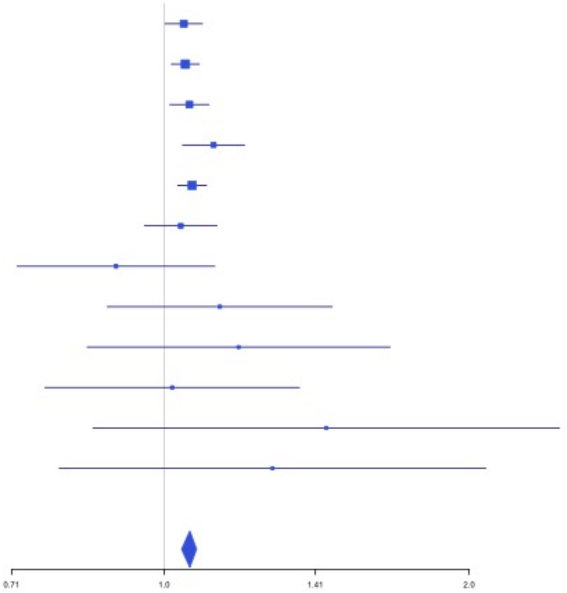

**Supplementary Figure 3.27** – rs35225200 (chr4:103146888) in locus 38 (BANK1;SLC39A8)

| Study | Cases | Controls | OR |
|---|---|---|---|
| INVENT-2019_EA | 15572 | 115280 | 0.98 |
| MVP_EA | 18952 | 411746 | 0.91 |
| FinnGen_EA | 9176 | 209616 | 1 |
| EGP_EA | 4122 | 181553 | 0.94 |
| UKB_EA | 18385 | 91834 | 0.92 |
| MGB_EA | 2967 | 27957 | 0.98 |
| UPenn_EA | 380 | 6210 | 0.84 |
| FARIVE_EA | 545 | 532 | 1.33 |
| BioME_EA | 200 | 8007 | 1.15 |
| RETROVE_EA | 400 | 400 | 0.72 |
| GAIT2_EA | 85 | 850 | 0.56 |
| MESA_EA | 94 | 2341 | 1.38 |
| MARTHA12_EA | 893 | 3414 | 1.01 |
| **European** | **71771** | **1059740** | **0.94** |

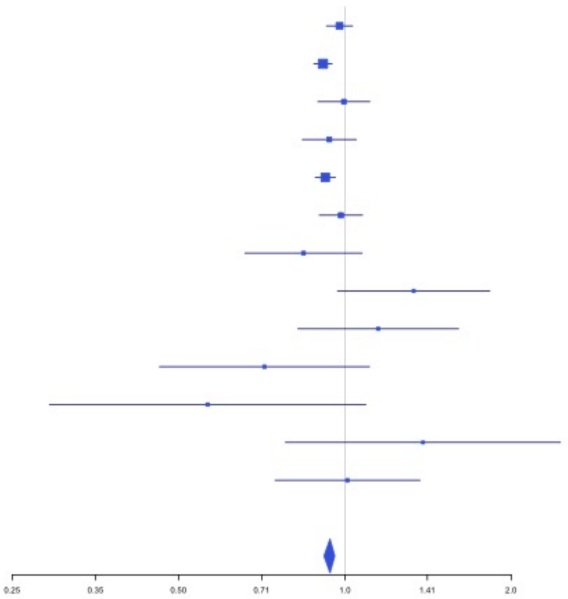


**Supplementary Figure 3.28** – rs2066864 (chr4:155525695) in locus 39 (FGG)

| Study | Cases | Controls | OR |
|---|---|---|---|
| INVENT-2019_EA | 15572 | 115280 | 1.2 |
| MVP_EA | 18952 | 411746 | 1.26 |
| FinnGen_EA | 9176 | 209616 | 1.24 |
| EGP_EA | 4122 | 181553 | 1.18 |
| UKB_EA | 18385 | 91834 | 1.22 |
| MGB_EA | 2967 | 27957 | 1.19 |
| UPenn_EA | 380 | 6210 | 1.36 |
| FARIVE_EA | 545 | 532 | 1.51 |
| BioME_EA | 200 | 8007 | 1.3 |
| RETROVE_EA | 400 | 400 | 1.46 |
| GAIT2_EA | 85 | 850 | 1.06 |
| MESA_EA | 94 | 2341 | 0.98 |
| MARTHA12_EA | 893 | 3414 | 1.46 |
| **European** | **71771** | **1059740** | **1.23** |

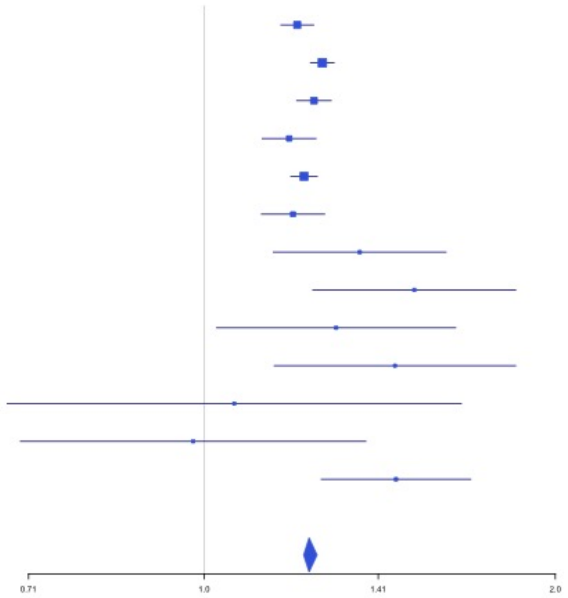

**Supplementary Figure 3.29** – rs3756011 (chr4:187206249) in locus 40 (F11)

| Study | Cases | Controls | OR |
|---|---|---|---|
| INVENT-2019_EA | 15572 | 115280 | 1.2 |
| MVP_EA | 18952 | 411746 | 1.26 |
| FinnGen_EA | 9176 | 209616 | 1.23 |
| EGP_EA | 4122 | 181553 | 1.25 |
| UKB_EA | 18385 | 91834 | 1.18 |
| MGB_EA | 2967 | 27957 | 1.18 |
| UPenn_EA | 380 | 6210 | 1.19 |
| FARIVE_EA | 545 | 532 | 1.47 |
| BioME_EA | 200 | 8007 | 1.03 |
| RETROVE_EA | 400 | 400 | 1.4 |
| GAIT2_EA | 85 | 850 | 1.53 |
| MESA_EA | 94 | 2341 | 0.99 |
| **European** | **70878** | **1056326** | **1.22** |

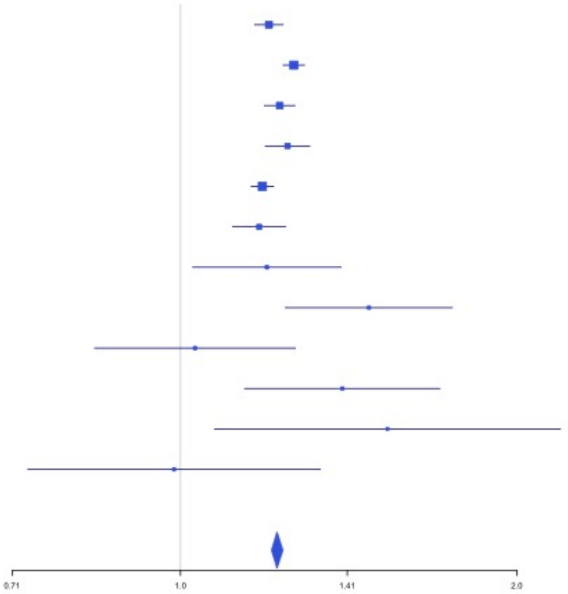


**Supplementary Figure 3.30** – rs112367053 (chr5:28379046) in locus 41 (LINC02103;LSP1P3)

| Study | Cases | Controls | OR |
|---|---|---|---|
| INVENT-2019_EA | 15572 | 115280 | 1.08 |
| FinnGen_EA | 9176 | 209616 | 1.05 |
| EGP_EA | 4122 | 181553 | 1.03 |
| MGB_EA | 2967 | 27957 | 1.13 |
| BioME_EA | 200 | 8007 | 0.95 |
| **European** | **32037** | **542413** | **1.06** |

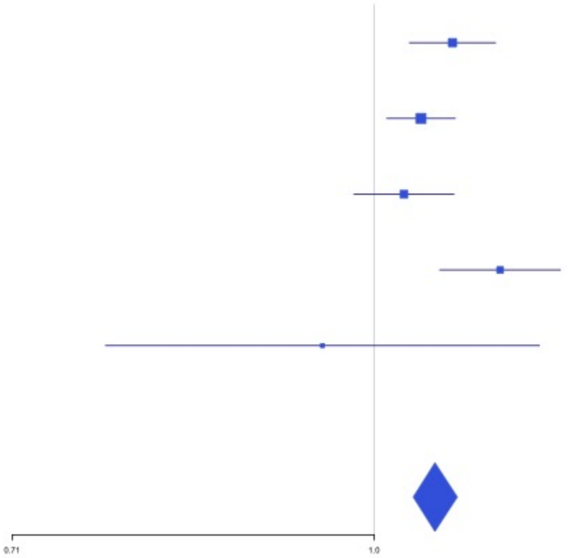

**Supplementary Figure 3.31** – rs16867574 (chr5:38708554) in locus 42 (OSMR-AS1)

| Study | Cases | Controls | OR |
|---|---|---|---|
| INVENT-2019_EA | 15572 | 115280 | 0.93 |
| MVP_EA | 18952 | 411746 | 0.95 |
| FinnGen_EA | 9176 | 209616 | 0.99 |
| EGP_EA | 4122 | 181553 | 0.93 |
| UKB_EA | 18385 | 91834 | 0.95 |
| MGB_EA | 2967 | 27957 | 0.97 |
| UPenn_EA | 380 | 6210 | 1.01 |
| FARIVE_EA | 545 | 532 | 0.95 |
| BioME_EA | 200 | 8007 | 0.96 |
| RETROVE_EA | 400 | 400 | 0.86 |
| GAIT2_EA | 85 | 850 | 1.37 |
| MESA_EA | 94 | 2341 | 1.13 |
| **European** | **70878** | **1056326** | **0.95** |

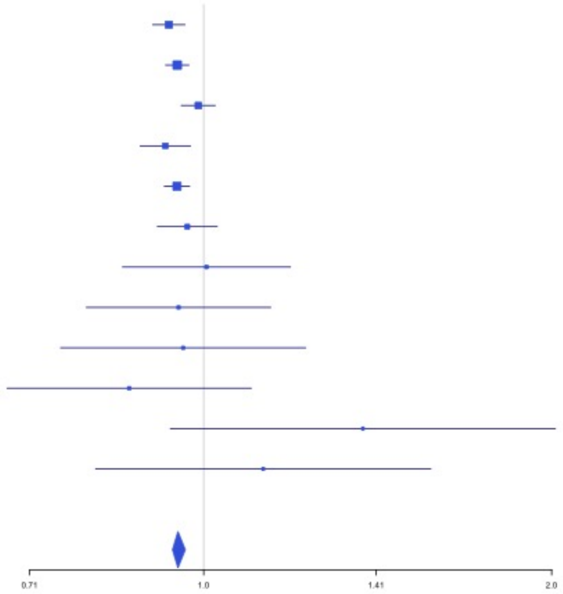


**Supplementary Figure 3.32** – rs4235677 (chr5:72925091) in locus 43 (ARHGEF28)

| Study | Cases | Controls | OR |
|---|---|---|---|
| INVENT-2019_EA | 15572 | 115280 | 0.97 |
| MVP_EA | 18952 | 411746 | 0.96 |
| FinnGen_EA | 9176 | 209616 | 0.94 |
| EGP_EA | 4122 | 181553 | 1 |
| UKB_EA | 18385 | 91834 | 0.97 |
| MGB_EA | 2967 | 27957 | 1 |
| UPenn_EA | 380 | 6210 | 0.98 |
| FARIVE_EA | 545 | 532 | 0.93 |
| BioME_EA | 200 | 8007 | 1.02 |
| RETROVE_EA | 400 | 400 | 1.01 |
| GAIT2_EA | 85 | 850 | 1.1 |
| MESA_EA | 94 | 2341 | 1.2 |
| **European** | **70878** | **1056326** | **0.97** |

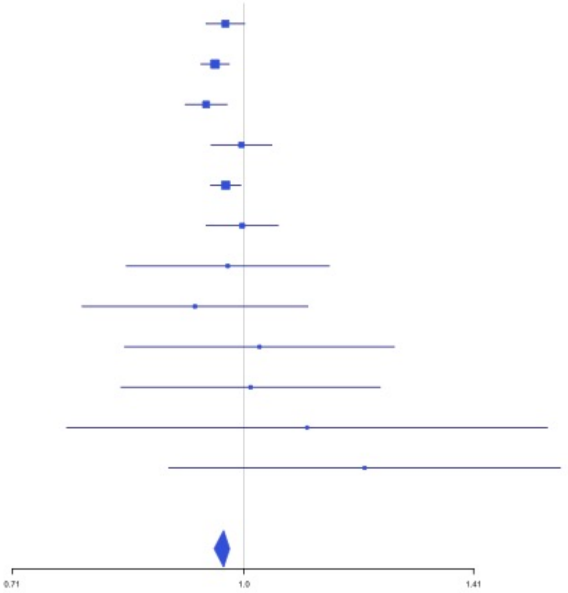

**Supplementary Figure 3.33** – rs251339 (chr5:96235038) in locus 44 (ERAP2)

| Study | Cases | Controls | OR |
|---|---|---|---|
| INVENT-2019_EA | 15572 | 115280 | 0.97 |
| MVP_EA | 18952 | 411746 | 0.96 |
| FinnGen_EA | 9176 | 209616 | 0.94 |
| EGP_EA | 4122 | 181553 | 0.99 |
| UKB_EA | 18385 | 91834 | 0.97 |
| MGB_EA | 2967 | 27957 | 0.96 |
| UPenn_EA | 380 | 6210 | 1.11 |
| FARIVE_EA | 545 | 532 | 1.03 |
| BioME_EA | 200 | 8007 | 0.94 |
| RETROVE_EA | 400 | 400 | 1.29 |
| GAIT2_EA | 85 | 850 | 0.84 |
| MESA_EA | 94 | 2341 | 1.07 |
| MARTHA12_EA | 893 | 3414 | 0.86 |
| **European** | **71771** | **1059740** | **0.96** |

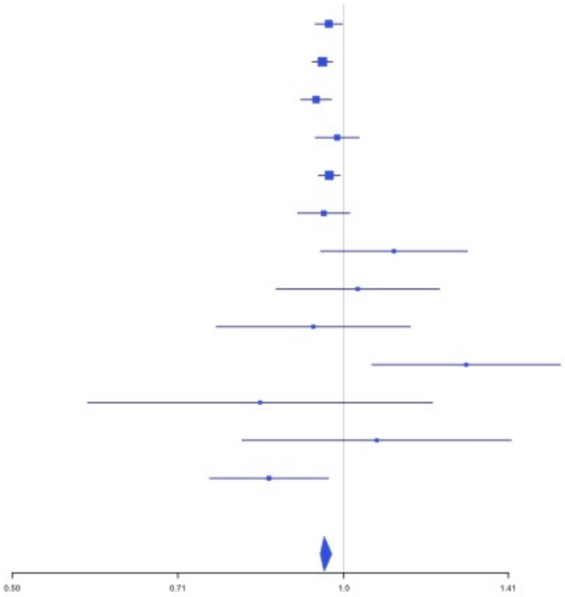


**Supplementary Figure 3.34** – rs147133967 (chr5:132426851) in locus 45 (HSPA4)

| Study | Cases | Controls | OR |
|---|---|---|---|
| INVENT-2019_EA | 15572 | 115280 | 0.96 |
| MVP_EA | 18952 | 411746 | 0.9 |
| FinnGen_EA | 9176 | 209616 | 0.93 |
| EGP_EA | 4122 | 181553 | 0.96 |
| UKB_EA | 18385 | 91834 | 0.94 |
| MGB_EA | 2967 | 27957 | 0.94 |
| UPenn_EA | 380 | 6210 | 1.11 |
| FARIVE_EA | 545 | 532 | 1.12 |
| BioME_EA | 200 | 8007 | 0.92 |
| **European** | **70299** | **1052735** | **0.93** |

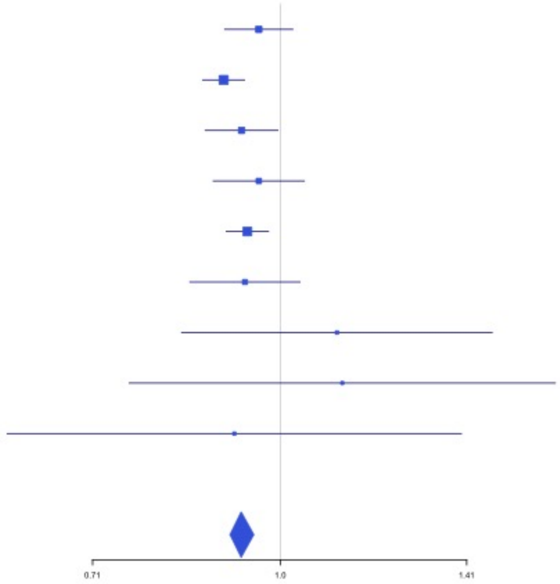

**Supplementary Figure 3.35** – rs214059 (chr6:25536937) in locus 46 (CARMIL1)

| Study | Cases | Controls | OR |
|---|---|---|---|
| INVENT-2019_EA | 15572 | 115280 | 1.01 |
| MVP_EA | 18952 | 411746 | 1.04 |
| FinnGen_EA | 9176 | 209616 | 1.06 |
| EGP_EA | 4122 | 181553 | 1.02 |
| UKB_EA | 18385 | 91834 | 1.04 |
| MGB_EA | 2967 | 27957 | 1.06 |
| UPenn_EA | 380 | 6210 | 0.99 |
| FARIVE_EA | 545 | 532 | 1.15 |
| BioME_EA | 200 | 8007 | 1.24 |
| RETROVE_EA | 400 | 400 | 1.13 |
| GAIT2_EA | 85 | 850 | 0.9 |
| MESA_EA | 94 | 2341 | 1.01 |
| MARTHA12_EA | 893 | 3414 | 1.17 |
| **European** | **71771** | **1059740** | **1.04** |

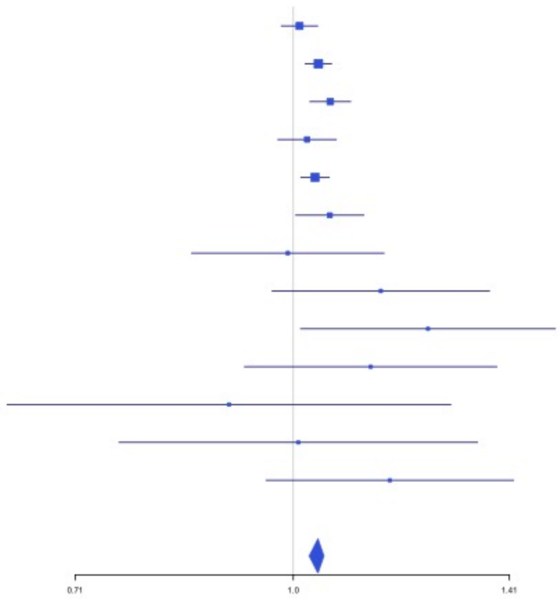


**Supplementary Figure 3.36** – rs2394251 (chr6:29943688) in locus 47 (HCG9)

| Study | Cases | Controls | OR |
|---|---|---|---|
| INVENT-2019_EA | 15572 | 115280 | 0.97 |
| MVP_EA | 18952 | 411746 | 0.94 |
| FinnGen_EA | 9176 | 209616 | 0.98 |
| EGP_EA | 4122 | 181553 | 0.95 |
| UKB_EA | 18385 | 91834 | 0.97 |
| MGB_EA | 2967 | 27957 | 0.96 |
| UPenn_EA | 380 | 6210 | 0.84 |
| FARIVE_EA | 545 | 532 | 0.96 |
| RETROVE_EA | 400 | 400 | 0.92 |
| GAIT2_EA | 85 | 850 | 0.91 |
| MESA_EA | 94 | 2341 | 0.84 |
| MARTHA12_EA | 893 | 3414 | 1.07 |
| **European** | **71571** | **1051733** | **0.96** |

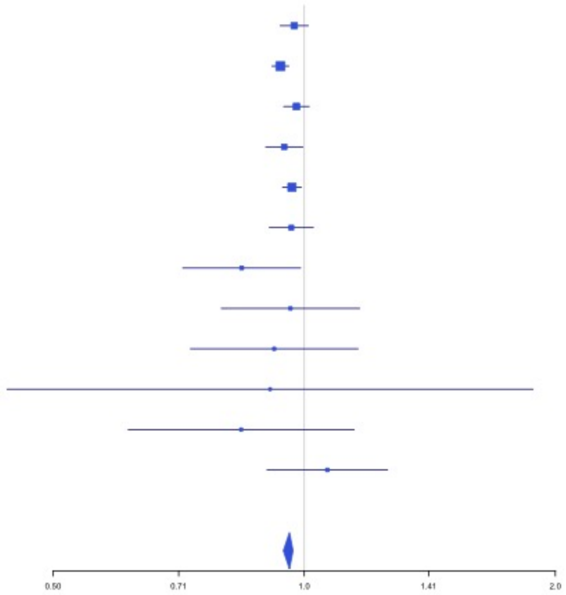

**Supplementary Figure 3.37** – rs28383307 (chr6:32586755) in locus 48 (HLA-DRB1;HLA-DQA1)

| Study | Cases | Controls | OR |
|---|---|---|---|
| INVENT-2019_EA | 15572 | 115280 | 0.95 |
| MVP_EA | 18952 | 411746 | 0.94 |
| FinnGen_EA | 9176 | 209616 | 0.91 |
| UKB_EA | 18385 | 91834 | 0.97 |
| MGB_EA | 2967 | 27957 | 0.9 |
| UPenn_EA | 380 | 6210 | 0.98 |
| FARIVE_EA | 545 | 532 | 0.79 |
| BioME_EA | 200 | 8007 | 0.94 |
| **European** | **66177** | **871182** | **0.95** |

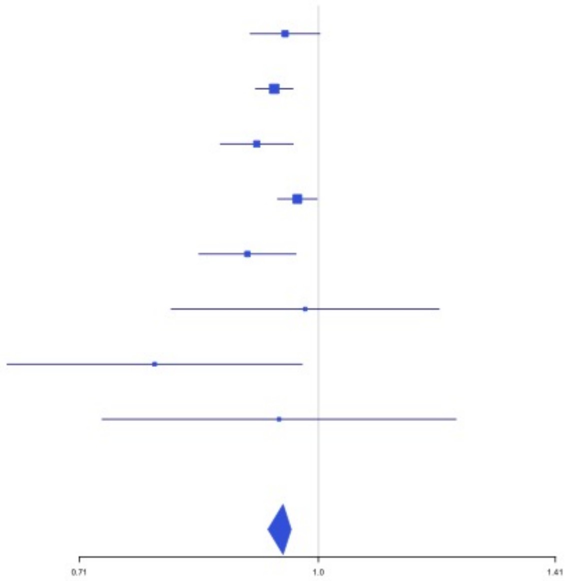


**Supplementary Figure 3.38** – rs145294670 (chr6:34622561) in locus 49 (ILRUN)

| Study | Cases | Controls | OR |
|---|---|---|---|
| INVENT-2019_EA | 15572 | 115280 | 1.03 |
| MVP_EA | 18952 | 411746 | 1.09 |
| FinnGen_EA | 9176 | 209616 | 1.05 |
| EGP_EA | 4122 | 181553 | 1.05 |
| UKB_EA | 18385 | 91834 | 1.02 |
| MGB_EA | 2967 | 27957 | 1.07 |
| UPenn_EA | 380 | 6210 | 1.06 |
| FARIVE_EA | 545 | 532 | 0.93 |
| BioME_EA | 200 | 8007 | 1.17 |
| MARTHA12_EA | 893 | 3414 | 0.96 |
| **European** | **71192** | **1056149** | **1.05** |

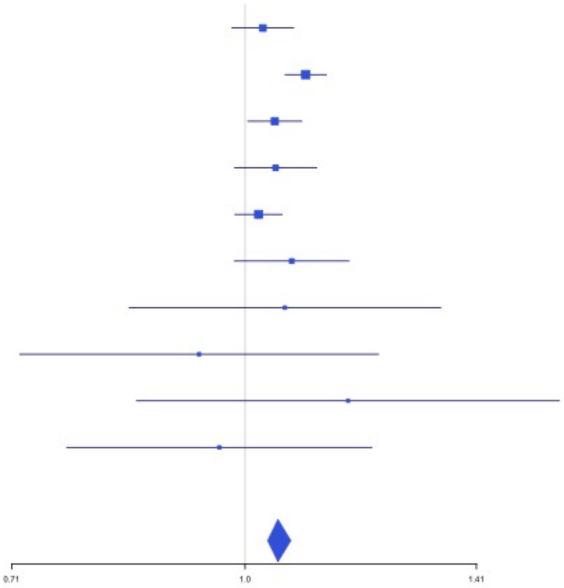

**Supplementary Figure 3.39** – rs2754251 (chr6:88385949) in locus 51 (AKIRIN2)

| Study | Cases | Controls | OR |
|---|---|---|---|
| INVENT-2019_EA | 15572 | 115280 | 1.04 |
| MVP_EA | 18952 | 411746 | 1.09 |
| FinnGen_EA | 9176 | 209616 | 1.08 |
| EGP_EA | 4122 | 181553 | 0.99 |
| UKB_EA | 18385 | 91834 | 1.08 |
| MGB_EA | 2967 | 27957 | 1.12 |
| UPenn_EA | 380 | 6210 | 1.12 |
| FARIVE_EA | 545 | 532 | 1.18 |
| BioME_EA | 200 | 8007 | 1.09 |
| RETROVE_EA | 400 | 400 | 1.03 |
| GAIT2_EA | 85 | 850 | 0.43 |
| MESA_EA | 94 | 2341 | 1.63 |
| **European** | **70878** | **1056326** | **1.07** |

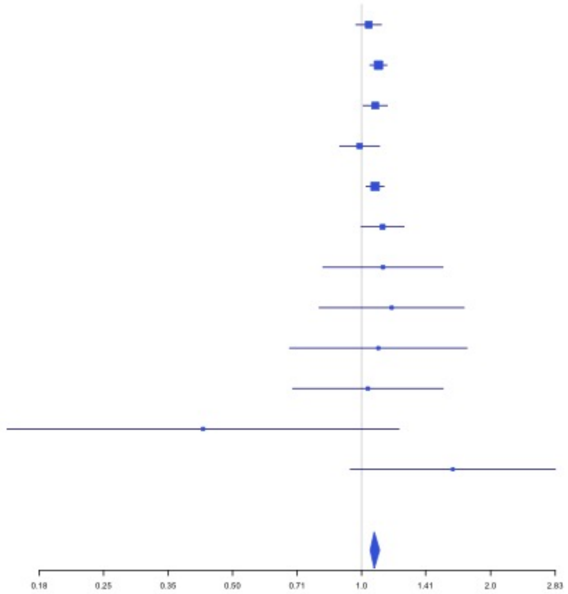


**Supplementary Figure 3.40** – rs9390460 (chr6:147694334) in locus 52 (STXBP5)

| Study | Cases | Controls | OR |
|---|---|---|---|
| INVENT-2019_EA | 15572 | 115280 | 0.94 |
| MVP_EA | 18952 | 411746 | 0.94 |
| FinnGen_EA | 9176 | 209616 | 0.98 |
| EGP_EA | 4122 | 181553 | 0.98 |
| UKB_EA | 18385 | 91834 | 0.95 |
| MGB_EA | 2967 | 27957 | 0.96 |
| UPenn_EA | 380 | 6210 | 1.03 |
| FARIVE_EA | 545 | 532 | 1.01 |
| BioME_EA | 200 | 8007 | 1.06 |
| RETROVE_EA | 400 | 400 | 0.95 |
| GAIT2_EA | 85 | 850 | 0.73 |
| MESA_EA | 94 | 2341 | 0.85 |
| MARTHA12_EA | 893 | 3414 | 0.98 |
| **European** | **71771** | **1059740** | **0.95** |

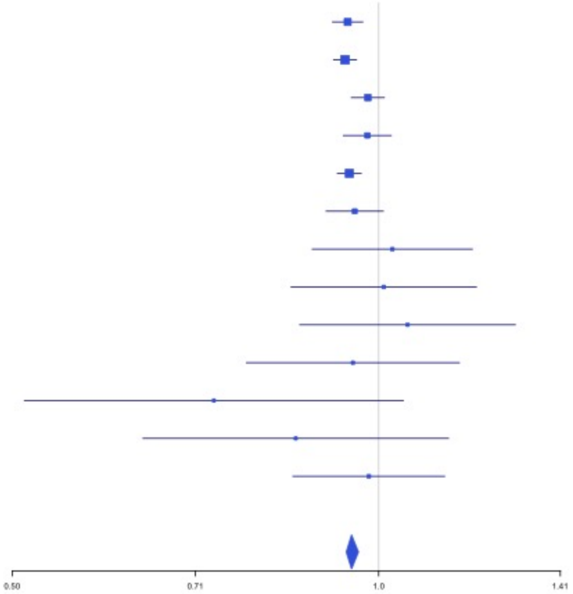

**Supplementary Figure 3.41** – rs1513275 (chr7:28259233) in locus 53 (JAZF1-AS1)

| Study | Cases | Controls | OR |
|---|---|---|---|
| INVENT-2019_EA | 15572 | 115280 | 1.05 |
| MVP_EA | 18952 | 411746 | 1.05 |
| FinnGen_EA | 9176 | 209616 | 1.09 |
| EGP_EA | 4122 | 181553 | 1.01 |
| UKB_EA | 18385 | 91834 | 1.05 |
| MGB_EA | 2967 | 27957 | 1.02 |
| UPenn_EA | 380 | 6210 | 1.1 |
| FARIVE_EA | 545 | 532 | 1.09 |
| BioME_EA | 200 | 8007 | 1.13 |
| RETROVE_EA | 400 | 400 | 1.07 |
| GAIT2_EA | 85 | 850 | 1.23 |
| MESA_EA | 94 | 2341 | 1.3 |
| **European** | **70878** | **1056326** | **1.05** |

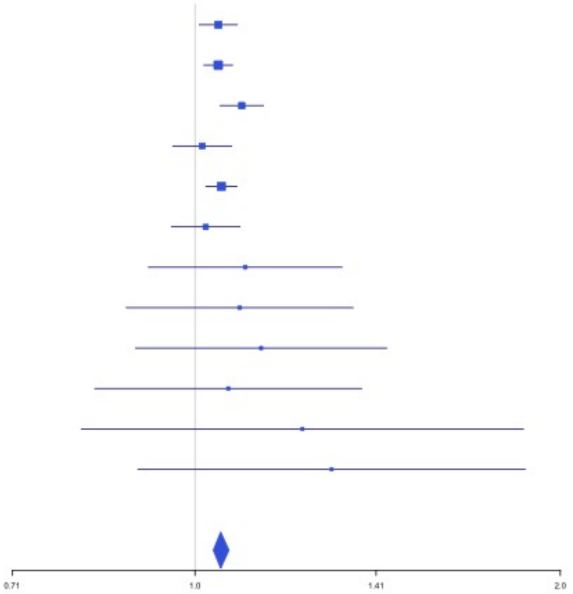


**Supplementary Figure 3.42** – rs370720 (chr8:9168146) in locus 57 (LOC101929128 ;LOC157273)

| Study | Cases | Controls | OR |
|---|---|---|---|
| INVENT-2019_EA | 15572 | 115280 | 1.04 |
| MVP_EA | 18952 | 411746 | 1.04 |
| FinnGen_EA | 9176 | 209616 | 1.06 |
| EGP_EA | 4122 | 181553 | 1.14 |
| UKB_EA | 18385 | 91834 | 1.12 |
| MGB_EA | 2967 | 27957 | 1.11 |
| UPenn_EA | 380 | 6210 | 1.15 |
| FARIVE_EA | 545 | 532 | 1.11 |
| BioME_EA | 200 | 8007 | 1.04 |
| RETROVE_EA | 400 | 400 | 0.94 |
| GAIT2_EA | 85 | 850 | 0.97 |
| MESA_EA | 94 | 2341 | 1.09 |
| **European** | **70878** | **1056326** | **1.08** |

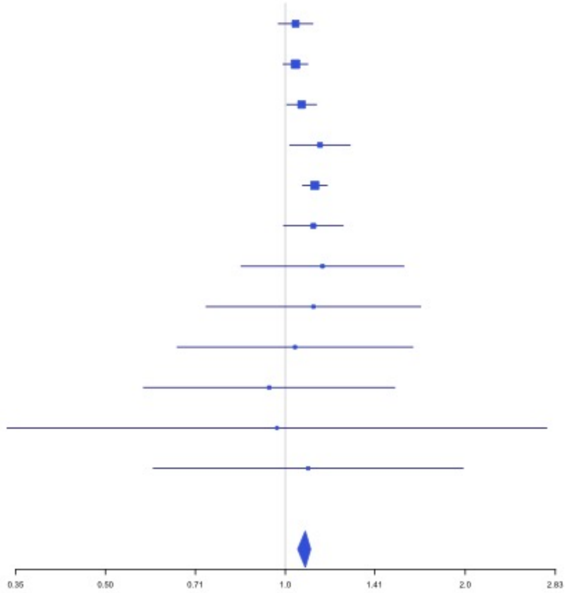

**Supplementary Figure 3.43** – rs2726950 (chr8:27806764) in locus 60 (SCARA5)

| Study | Cases | Controls | OR |
|---|---|---|---|
| INVENT-2019_EA | 15572 | 115280 | 0.96 |
| MVP_EA | 18952 | 411746 | 0.92 |
| FinnGen_EA | 9176 | 209616 | 0.98 |
| EGP_EA | 4122 | 181553 | 0.91 |
| UKB_EA | 18385 | 91834 | 0.95 |
| MGB_EA | 2967 | 27957 | 0.92 |
| UPenn_EA | 380 | 6210 | 0.92 |
| FARIVE_EA | 545 | 532 | 0.91 |
| BioME_EA | 200 | 8007 | 0.96 |
| RETROVE_EA | 400 | 400 | 0.88 |
| GAIT2_EA | 85 | 850 | 1.19 |
| MESA_EA | 94 | 2341 | 1.06 |
| **European** | **70878** | **1056326** | **0.94** |

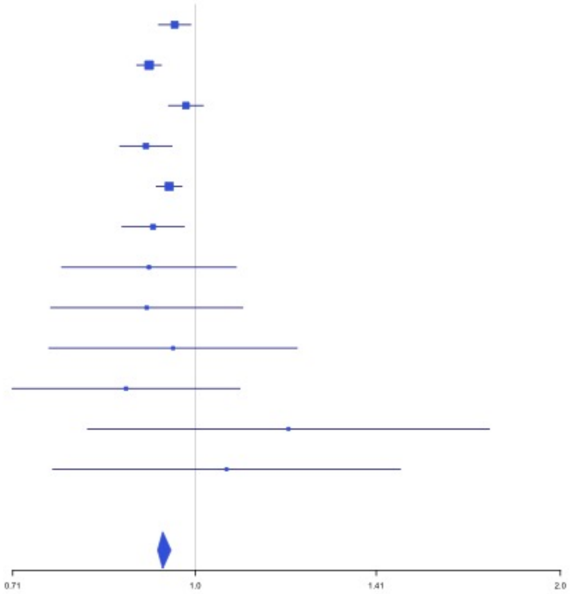


**Supplementary Figure 3.44** – rs6993770 (chr8:106581528) in locus 62 (ZFPM2)

| Study | Cases | Controls | OR |
|---|---|---|---|
| INVENT-2019_EA | 15572 | 115280 | 1.07 |
| MVP_EA | 18952 | 411746 | 1.1 |
| FinnGen_EA | 9176 | 209616 | 1.05 |
| EGP_EA | 4122 | 181553 | 1.05 |
| UKB_EA | 18385 | 91834 | 1.08 |
| MGB_EA | 2967 | 27957 | 1.05 |
| UPenn_EA | 380 | 6210 | 0.99 |
| FARIVE_EA | 545 | 532 | 1.08 |
| BioME_EA | 200 | 8007 | 1.09 |
| RETROVE_EA | 400 | 400 | 1.35 |
| GAIT2_EA | 85 | 850 | 1.09 |
| MESA_EA | 94 | 2341 | 0.98 |
| MARTHA12_EA | 893 | 3414 | 1 |
| **European** | **71771** | **1059740** | **1.08** |

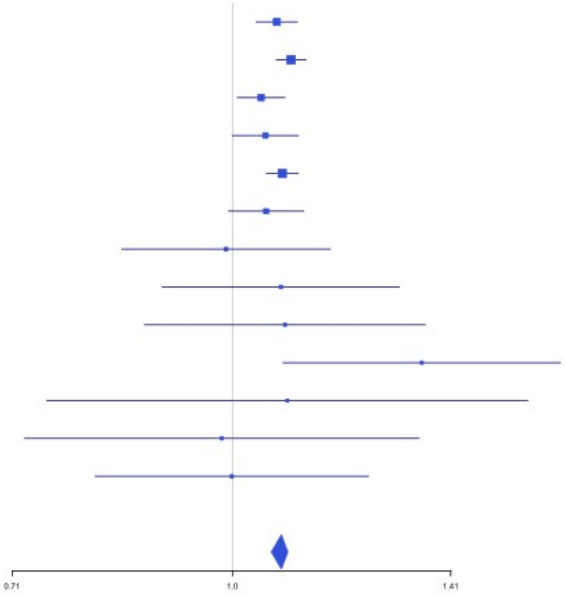

**Supplementary Figure 3.45** – rs1491336705 (chr8:108295235) in locus 63 (ANGPT1)

| Study | Cases | Controls | OR |
|---|---|---|---|
| MVP_EA | 18952 | 411746 | 0.93 |
| UKB_EA | 18385 | 91834 | 0.96 |
| MGB_EA | 2967 | 27957 | 0.98 |
| UPenn_EA | 380 | 6210 | 1.05 |
| FARIVE_EA | 545 | 532 | 0.9 |
| BioME_EA | 200 | 8007 | 1.02 |
| **European** | **41429** | **546286** | **0.95** |

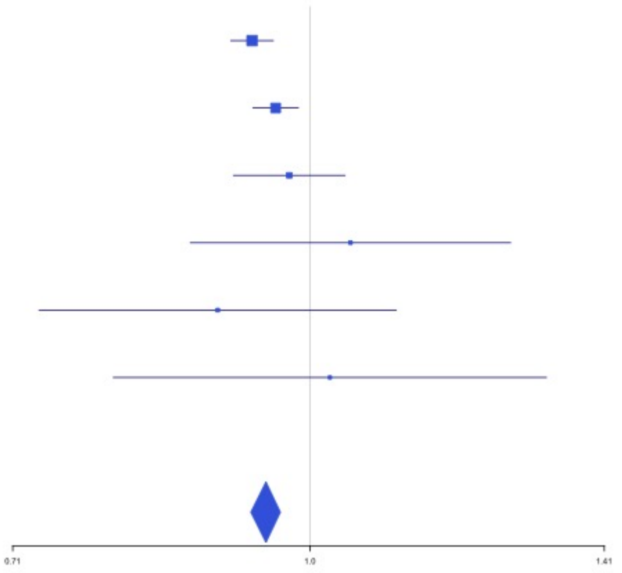


**Supplementary Figure 3.46** – rs10820606 (chr9:99192919) in locus 65 (ZNF367;HABP4)

| Study | Cases | Controls | OR |
|---|---|---|---|
| INVENT-2019_EA | 15572 | 115280 | 1.03 |
| MVP_EA | 18952 | 411746 | 1.07 |
| UKB_EA | 18385 | 91834 | 1.03 |
| MGB_EA | 2967 | 27957 | 1.1 |
| UPenn_EA | 380 | 6210 | 1.1 |
| FARIVE_EA | 545 | 532 | 1.16 |
| BioME_EA | 200 | 8007 | 1.32 |
| **European** | **57001** | **661566** | **1.05** |

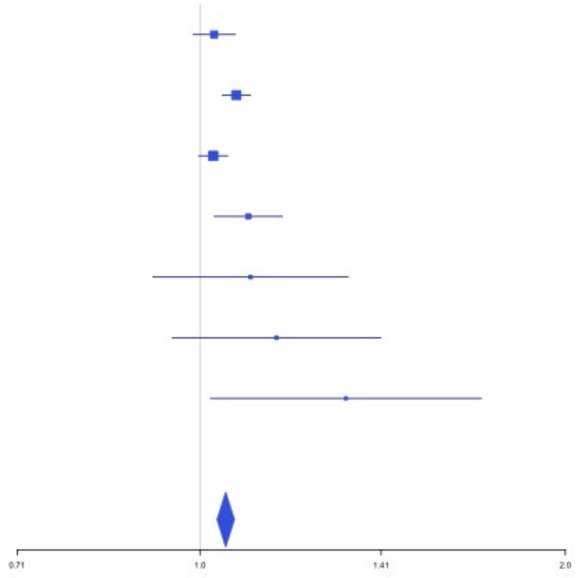

**Supplementary Figure 3.47** – rs687289 (chr9:136137106) in locus 66 (ABO)

| Study | Cases | Controls | OR |
|---|---|---|---|
| INVENT-2019_EA | 15572 | 115280 | 1.36 |
| MVP_EA | 18952 | 411746 | 1.4 |
| FinnGen_EA | 9176 | 209616 | 1.28 |
| EGP_EA | 4122 | 181553 | 1.22 |
| UKB_EA | 18385 | 91834 | 1.32 |
| MGB_EA | 2967 | 27957 | 1.21 |
| UPenn_EA | 380 | 6210 | 1.28 |
| FARIVE_EA | 545 | 532 | 1.71 |
| BioME_EA | 200 | 8007 | 1.26 |
| RETROVE_EA | 400 | 400 | 1.56 |
| GAIT2_EA | 85 | 850 | 1.84 |
| MESA_EA | 94 | 2341 | 1.18 |
| MARTHA12_EA | 893 | 3414 | 1.72 |
| **European** | **71771** | **1059740** | **1.34** |

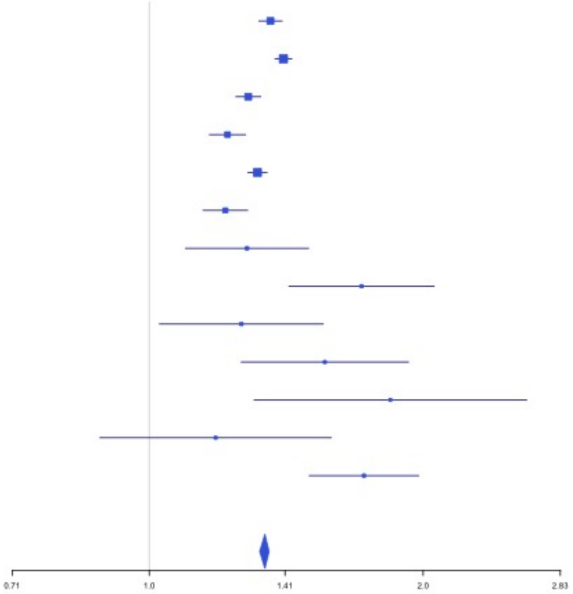


**Supplementary Figure 3.48** – rs1243184 (chr10:21931937) in locus 68 (MLLT10)

| Study | Cases | Controls | OR |
|---|---|---|---|
| INVENT-2019_EA | 15572 | 115280 | 0.95 |
| MVP_EA | 18952 | 411746 | 0.97 |
| FinnGen_EA | 9176 | 209616 | 0.98 |
| EGP_EA | 4122 | 181553 | 0.98 |
| UKB_EA | 18385 | 91834 | 0.96 |
| MGB_EA | 2967 | 27957 | 0.94 |
| UPenn_EA | 380 | 6210 | 0.95 |
| FARIVE_EA | 545 | 532 | 0.97 |
| BioME_EA | 200 | 8007 | 0.93 |
| RETROVE_EA | 400 | 400 | 1.11 |
| GAIT2_EA | 85 | 850 | 1.28 |
| MESA_EA | 94 | 2341 | 1.19 |
| **European** | **70878** | **1056326** | **0.96** |

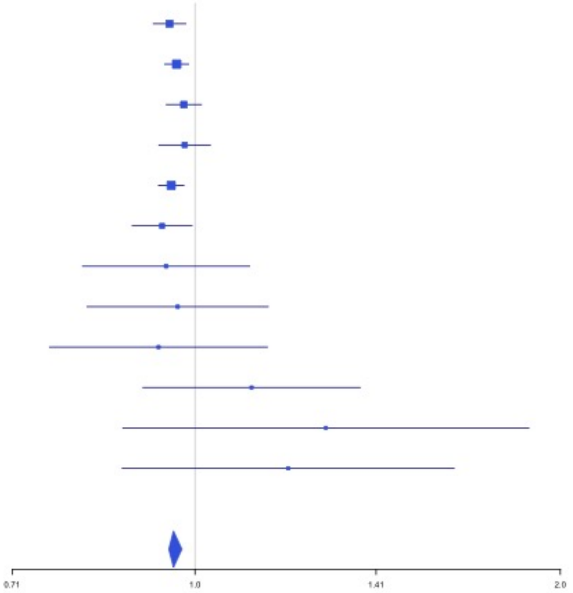

**Supplementary Figure 3.49** – rs10763665 (chr10:28771491) in locus 70 (LINC02652)

| Study | Cases | Controls | OR |
|---|---|---|---|
| INVENT-2019_EA | 15572 | 115280 | 0.98 |
| MVP_EA | 18952 | 411746 | 0.98 |
| FinnGen_EA | 9176 | 209616 | 0.95 |
| EGP_EA | 4122 | 181553 | 0.95 |
| UKB_EA | 18385 | 91834 | 0.96 |
| MGB_EA | 2967 | 27957 | 0.95 |
| UPenn_EA | 380 | 6210 | 0.96 |
| FARIVE_EA | 545 | 532 | 1 |
| BioME_EA | 200 | 8007 | 1.05 |
| RETROVE_EA | 400 | 400 | 1.06 |
| GAIT2_EA | 85 | 850 | 1.21 |
| MESA_EA | 94 | 2341 | 0.79 |
| **European** | **70878** | **1056326** | **0.97** |

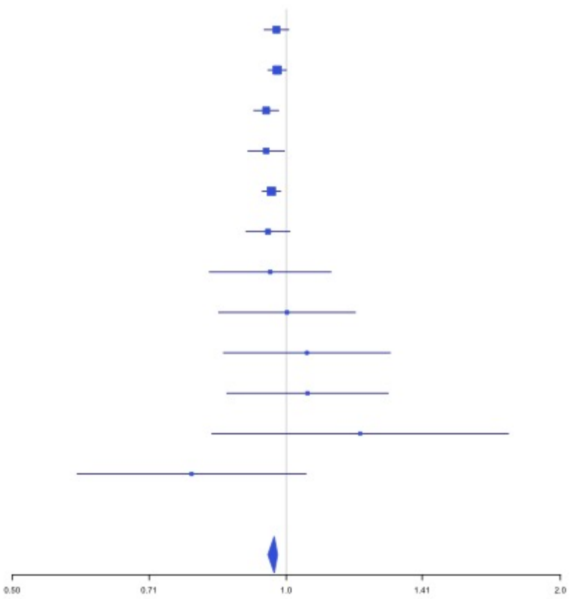


**Supplementary Figure 3.50** – rs78707713 (chr10:71245276) in locus 72 (TSPAN15)

| Study | Cases | Controls | OR |
|---|---|---|---|
| INVENT-2019_EA | 15572 | 115280 | 1.18 |
| MVP_EA | 18952 | 411746 | 1.33 |
| FinnGen_EA | 9176 | 209616 | 1.2 |
| EGP_EA | 4122 | 181553 | 1.21 |
| UKB_EA | 18385 | 91834 | 1.23 |
| MGB_EA | 2967 | 27957 | 1.21 |
| UPenn_EA | 380 | 6210 | 1.01 |
| FARIVE_EA | 545 | 532 | 1.36 |
| BioME_EA | 200 | 8007 | 1.52 |
| RETROVE_EA | 400 | 400 | 1.35 |
| GAIT2_EA | 85 | 850 | 0.79 |
| MESA_EA | 94 | 2341 | 1.09 |
| **European** | **70878** | **1056326** | **1.24** |

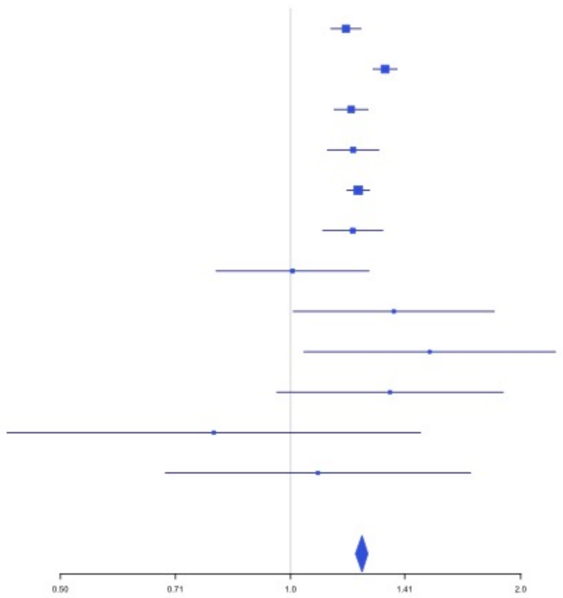

**Supplementary Figure 3.51** – rs57866767 (chr10:96023077) in locus 74 (PLCE1)

| Study | Cases | Controls | OR |
|---|---|---|---|
| INVENT-2019_EA | 15572 | 115280 | 0.97 |
| MVP_EA | 18952 | 411746 | 0.96 |
| FinnGen_EA | 9176 | 209616 | 0.96 |
| EGP_EA | 4122 | 181553 | 0.96 |
| UKB_EA | 18385 | 91834 | 0.96 |
| MGB_EA | 2967 | 27957 | 0.96 |
| UPenn_EA | 380 | 6210 | 0.92 |
| FARIVE_EA | 545 | 532 | 0.91 |
| BioME_EA | 200 | 8007 | 0.85 |
| RETROVE_EA | 400 | 400 | 1.03 |
| GAIT2_EA | 85 | 850 | 0.91 |
| MESA_EA | 94 | 2341 | 0.91 |
| MARTHA12_EA | 893 | 3414 | 0.95 |
| **European** | **71771** | **1059740** | **0.96** |

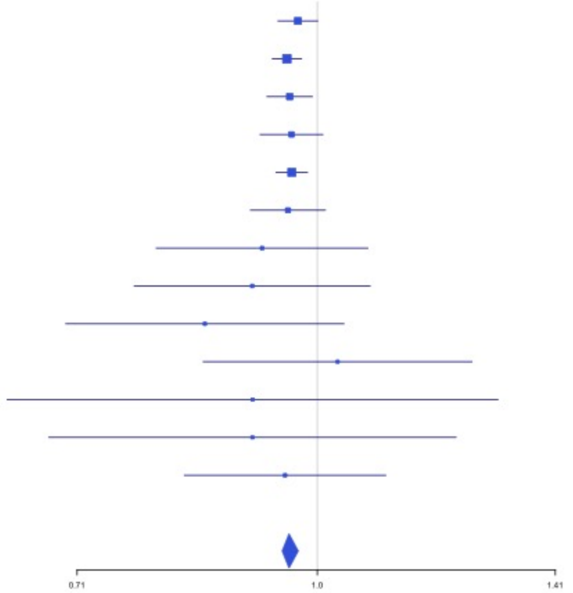


**Supplementary Figure 3.52** – rs10886430 (chr10:121010256) in locus 75 (GRK5)

| Study | Cases | Controls | OR |
|---|---|---|---|
| INVENT-2019_EA | 15572 | 115280 | 0.9 |
| MVP_EA | 18952 | 411746 | 0.86 |
| FinnGen_EA | 9176 | 209616 | 0.91 |
| EGP_EA | 4122 | 181553 | 0.92 |
| UKB_EA | 18385 | 91834 | 0.89 |
| MGB_EA | 2967 | 27957 | 0.93 |
| UPenn_EA | 380 | 6210 | 0.95 |
| FARIVE_EA | 545 | 532 | 1.04 |
| BioME_EA | 200 | 8007 | 0.76 |
| RETROVE_EA | 400 | 400 | 0.88 |
| GAIT2_EA | 85 | 850 | 0.88 |
| MESA_EA | 94 | 2341 | 0.75 |
| **European** | **70878** | **1056326** | **0.89** |

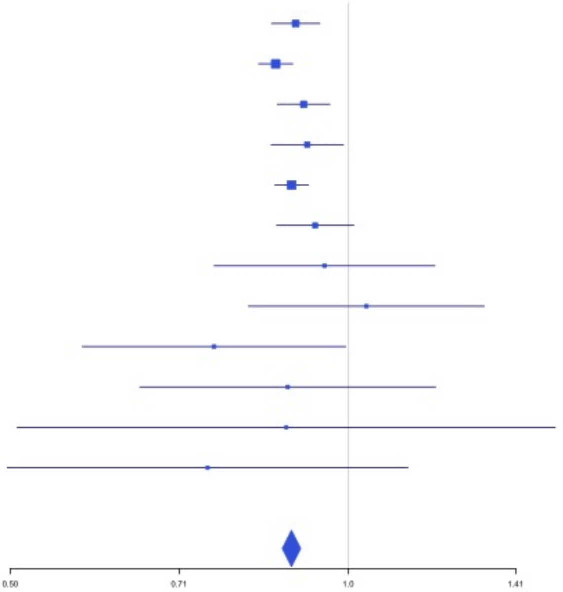

**Supplementary Figure 3.53** – rs7122100 (chr11:10732560) in locus 76 (IRAG1;CTR9)

| Study | Cases | Controls | OR |
|---|---|---|---|
| INVENT-2019_EA | 15572 | 115280 | 1.03 |
| MVP_EA | 18952 | 411746 | 1.05 |
| EGP_EA | 4122 | 181553 | 1.05 |
| UKB_EA | 18385 | 91834 | 1.04 |
| MGB_EA | 2967 | 27957 | 1.07 |
| UPenn_EA | 380 | 6210 | 0.98 |
| FARIVE_EA | 545 | 532 | 0.94 |
| BioME_EA | 200 | 8007 | 1.01 |
| RETROVE_EA | 400 | 400 | 1.07 |
| GAIT2_EA | 85 | 850 | 0.86 |
| MESA_EA | 94 | 2341 | 1.11 |
| MARTHA12_EA | 893 | 3414 | 1.05 |
| **European** | **62595** | **850124** | **1.04** |

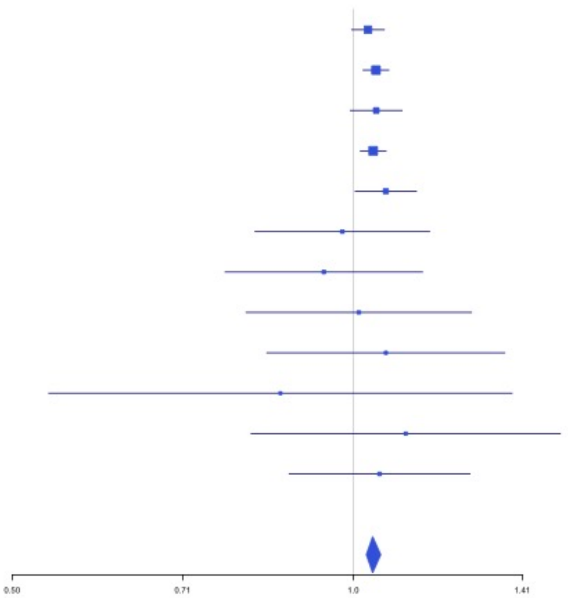


**Supplementary Figure 3.54** – rs12800654 (chr11:16243320) in locus 77 (SOX6)

| Study | Cases | Controls | OR |
|---|---|---|---|
| INVENT-2019_EA | 15572 | 115280 | 0.94 |
| MVP_EA | 18952 | 411746 | 0.94 |
| FinnGen_EA | 9176 | 209616 | 0.97 |
| EGP_EA | 4122 | 181553 | 0.93 |
| UKB_EA | 18385 | 91834 | 0.95 |
| MGB_EA | 2967 | 27957 | 0.98 |
| UPenn_EA | 380 | 6210 | 0.78 |
| FARIVE_EA | 545 | 532 | 0.91 |
| BioME_EA | 200 | 8007 | 0.93 |
| RETROVE_EA | 400 | 400 | 0.92 |
| GAIT2_EA | 85 | 850 | 1.04 |
| MESA_EA | 94 | 2341 | 0.82 |
| **European** | **70878** | **1056326** | **0.95** |

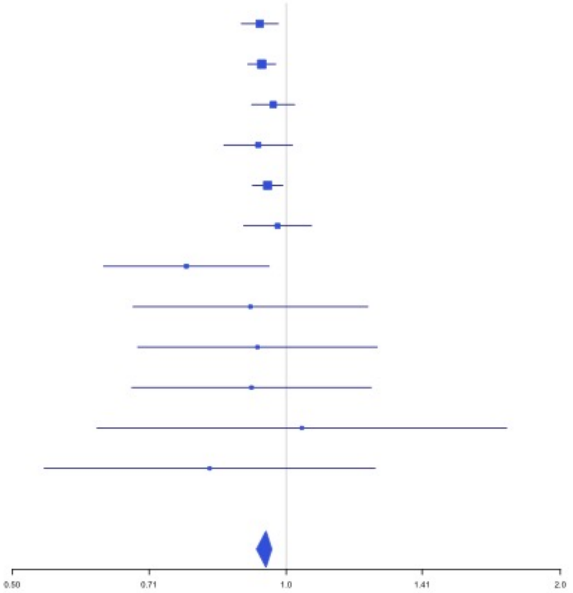

**Supplementary Figure 3.55** – rs4755476 (chr11:32980428) in locus 78 (QSER1)

| Study | Cases | Controls | OR |
|---|---|---|---|
| INVENT-2019_EA | 15572 | 115280 | 1.07 |
| MVP_EA | 18952 | 411746 | 1.05 |
| FinnGen_EA | 9176 | 209616 | 1.08 |
| EGP_EA | 4122 | 181553 | 1.05 |
| UKB_EA | 18385 | 91834 | 1.05 |
| MGB_EA | 2967 | 27957 | 1.01 |
| UPenn_EA | 380 | 6210 | 0.9 |
| FARIVE_EA | 545 | 532 | 1 |
| BioME_EA | 200 | 8007 | 0.95 |
| RETROVE_EA | 400 | 400 | 1.11 |
| GAIT2_EA | 85 | 850 | 0.88 |
| MESA_EA | 94 | 2341 | 0.91 |
| **European** | **70878** | **1056326** | **1.05** |

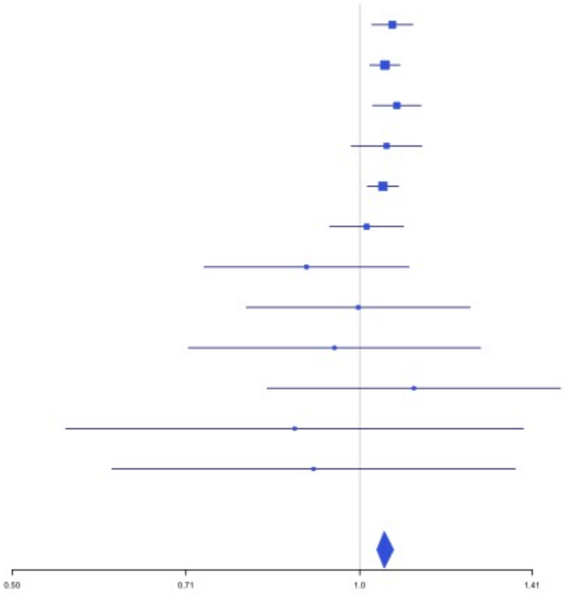


**Supplementary Figure 3.56** – rs1799963 (chr11:46761055) in locus 79 (F2)

| Study | Cases | Controls | OR |
|---|---|---|---|
| INVENT-2019_EA | 15572 | 115280 | 1.6 |
| MVP_EA | 18952 | 411746 | 2.08 |
| FinnGen_EA | 9176 | 209616 | 3.42 |
| EGP_EA | 4122 | 181553 | 1.94 |
| UKB_EA | 18385 | 91834 | 1.93 |
| MGB_EA | 2967 | 27957 | 1.42 |
| UPenn_EA | 380 | 6210 | 0.94 |
| FARIVE_EA | 545 | 532 | 3.09 |
| BioME_EA | 200 | 8007 | 1.4 |
| RETROVE_EA | 400 | 400 | 1.49 |
| GAIT2_EA | 85 | 850 | 2.76 |
| MESA_EA | 94 | 2341 | 1.22 |
| MARTHA12_EA | 893 | 3414 | 3.82 |
| **European** | **71771** | **1059740** | **1.99** |

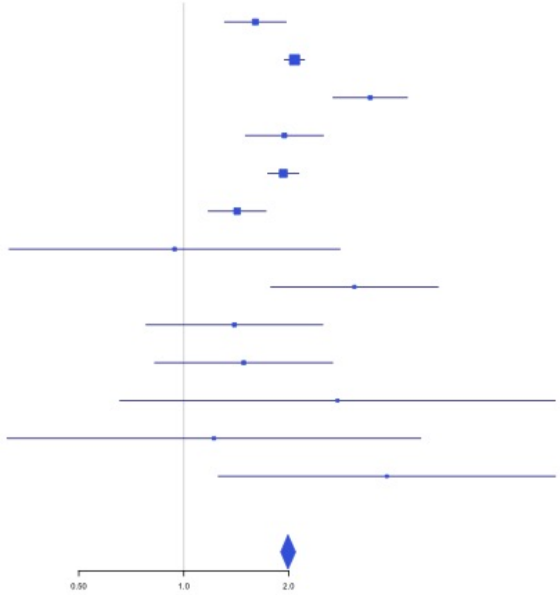

**Supplementary Figure 3.57** – rs141798115 (chr11:56875074) in locus 80 (OR5AK4P ;LRRC55)

| Study | Cases | Controls | OR |
|---|---|---|---|
| INVENT-2019_EA | 15572 | 115280 | 1.09 |
| MVP_EA | 18952 | 411746 | 1.37 |
| FinnGen_EA | 9176 | 209616 | 1.4 |
| EGP_EA | 4122 | 181553 | 1.37 |
| UKB_EA | 18385 | 91834 | 1.19 |
| MGB_EA | 2967 | 27957 | 1.12 |
| UPenn_EA | 380 | 6210 | 1.04 |
| FARIVE_EA | 545 | 532 | 1.7 |
| BioME_EA | 200 | 8007 | 0.54 |
| RETROVE_EA | 400 | 400 | 0.57 |
| GAIT2_EA | 85 | 850 | 1.33 |
| MESA_EA | 94 | 2341 | 0.42 |
| MARTHA12_EA | 893 | 3414 | 0.99 |
| **European** | **71771** | **1059740** | **1.27** |

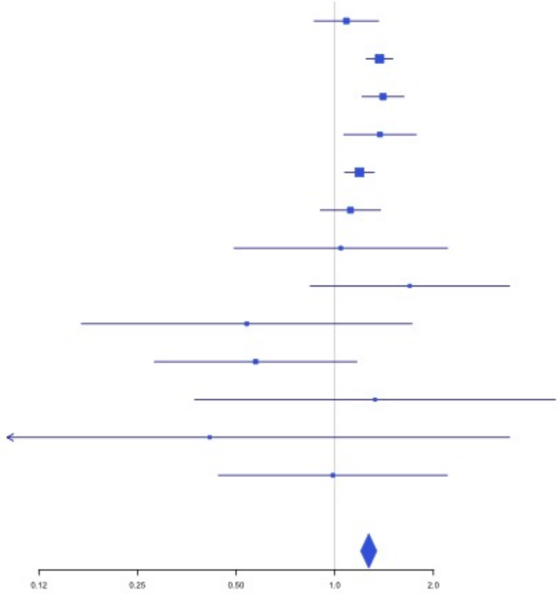


**Supplementary Figure 3.58** – rs55777218 (chr11:60070946) in locus 81 (MS4A4A)

| Study | Cases | Controls | OR |
|---|---|---|---|
| INVENT-2019_EA | 15572 | 115280 | 0.98 |
| MVP_EA | 18952 | 411746 | 0.96 |
| FinnGen_EA | 9176 | 209616 | 0.98 |
| EGP_EA | 4122 | 181553 | 0.95 |
| UKB_EA | 18385 | 91834 | 0.97 |
| MGB_EA | 2967 | 27957 | 0.94 |
| UPenn_EA | 380 | 6210 | 0.99 |
| FARIVE_EA | 545 | 532 | 1.02 |
| BioME_EA | 200 | 8007 | 1.15 |
| RETROVE_EA | 400 | 400 | 0.91 |
| GAIT2_EA | 85 | 850 | 0.96 |
| MESA_EA | 94 | 2341 | 0.88 |
| MARTHA12_EA | 893 | 3414 | 0.9 |
| **European** | **71771** | **1059740** | **0.97** |

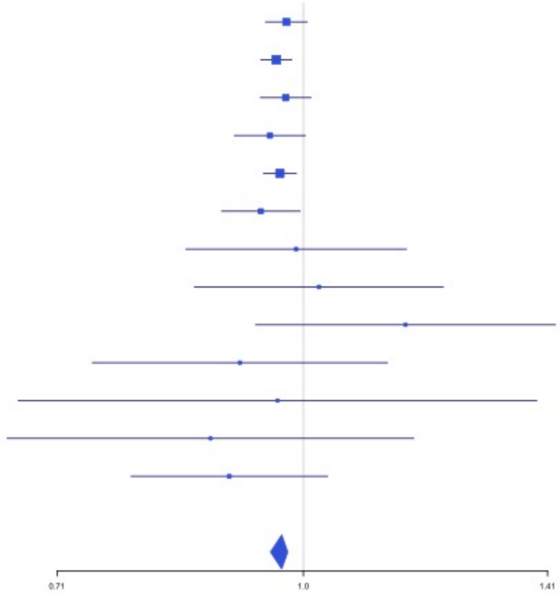

**Supplementary Figure 3.59** – rs174551 (chr11:61573684) in locus 82 (FADS1)

| Study | Cases | Controls | OR |
|---|---|---|---|
| INVENT-2019_EA | 15572 | 115280 | 1.05 |
| MVP_EA | 18952 | 411746 | 1.09 |
| FinnGen_EA | 9176 | 209616 | 1.06 |
| UKB_EA | 18385 | 91834 | 1.06 |
| MGB_EA | 2967 | 27957 | 1.06 |
| UPenn_EA | 380 | 6210 | 1.04 |
| FARIVE_EA | 545 | 532 | 1.18 |
| MARTHA12_EA | 893 | 3414 | 1.22 |
| **European** | **66870** | **866589** | **1.07** |

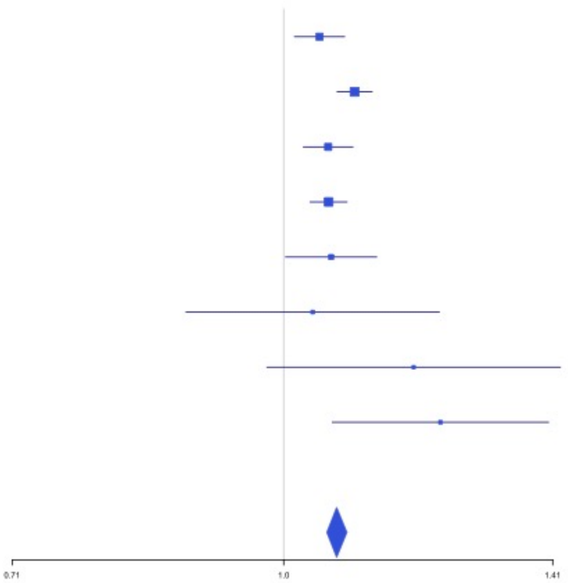

**Supplementary Figure 3.60** – rs1145656 (chr11:73305859) in locus 83 (FAM168A)

| Study | Cases | Controls | OR |
|---|---|---|---|
| INVENT-2019_EA | 15572 | 115280 | 0.97 |
| MVP_EA | 18952 | 411746 | 0.96 |
| FinnGen_EA | 9176 | 209616 | 0.96 |
| EGP_EA | 4122 | 181553 | 0.99 |
| UKB_EA | 18385 | 91834 | 0.95 |
| MGB_EA | 2967 | 27957 | 0.9 |
| UPenn_EA | 380 | 6210 | 1 |
| FARIVE_EA | 545 | 532 | 1.14 |
| BioME_EA | 200 | 8007 | 0.83 |
| RETROVE_EA | 400 | 400 | 0.99 |
| GAIT2_EA | 85 | 850 | 0.85 |
| MESA_EA | 94 | 2341 | 0.88 |
| **European** | **70878** | **1056326** | **0.96** |

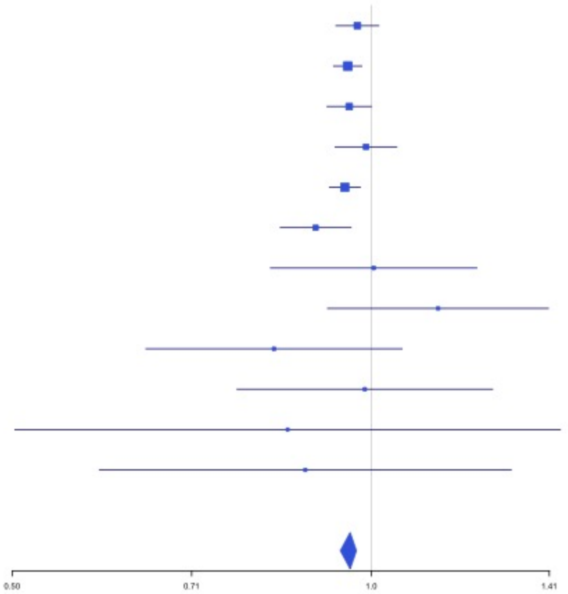

**Supplementary Figure 3.61** – rs1997547 (chr11:114002744) in locus 84 (ZBTB16)

| Study | Cases | Controls | OR |
|---|---|---|---|
| INVENT-2019_EA | 15572 | 115280 | 0.96 |
| MVP_EA | 18952 | 411746 | 0.96 |
| FinnGen_EA | 9176 | 209616 | 0.96 |
| EGP_EA | 4122 | 181553 | 0.99 |
| UKB_EA | 18385 | 91834 | 0.97 |
| MGB_EA | 2967 | 27957 | 0.94 |
| UPenn_EA | 380 | 6210 | 1.08 |
| FARIVE_EA | 545 | 532 | 0.86 |
| BioME_EA | 200 | 8007 | 0.96 |
| RETROVE_EA | 400 | 400 | 0.87 |
| GAIT2_EA | 85 | 850 | 0.85 |
| MESA_EA | 94 | 2341 | 0.94 |
| **European** | **70878** | **1056326** | **0.96** |

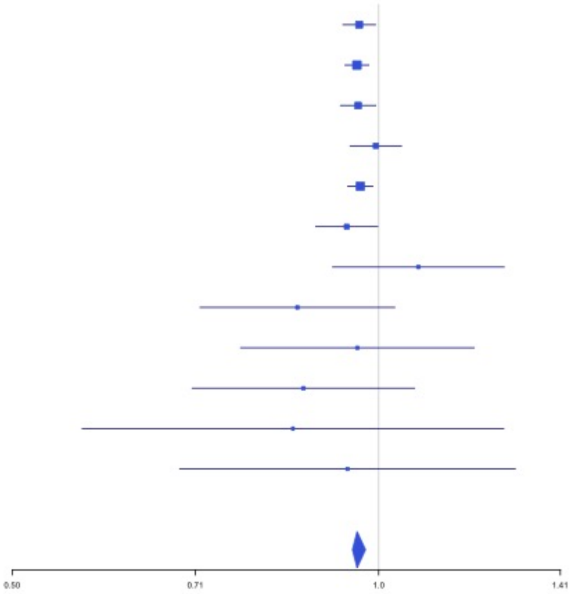


**Supplementary Figure 3.62** – rs35257264 (chr11:126296816) in locus 85 (ST3GAL4)

| Study | Cases | Controls | OR |
|---|---|---|---|
| INVENT-2019_EA | 15572 | 115280 | 1.2 |
| MVP_EA | 18952 | 411746 | 1.22 |
| FinnGen_EA | 9176 | 209616 | 1.2 |
| EGP_EA | 4122 | 181553 | 1.1 |
| UKB_EA | 18385 | 91834 | 1.11 |
| MGB_EA | 2967 | 27957 | 1.08 |
| UPenn_EA | 380 | 6210 | 1.06 |
| FARIVE_EA | 545 | 532 | 0.96 |
| BioME_EA | 200 | 8007 | 1.47 |
| RETROVE_EA | 400 | 400 | 1.53 |
| GAIT2_EA | 85 | 850 | 0.59 |
| MESA_EA | 94 | 2341 | 1.29 |
| **European** | **70878** | **1056326** | **1.16** |

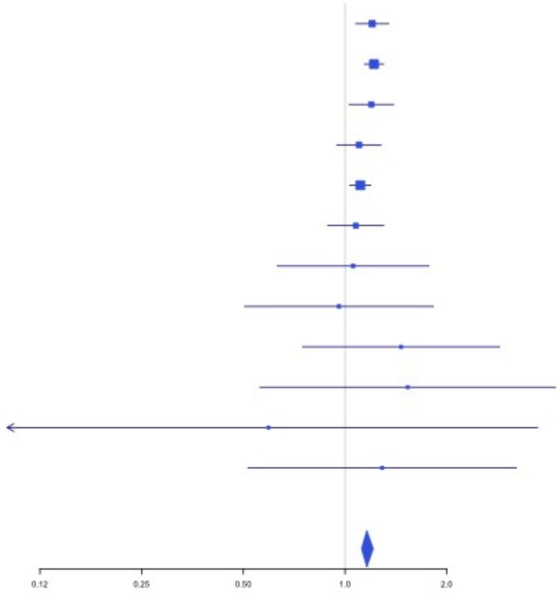

**Supplementary Figure 3.63** – rs7135039 (chr12:6160614) in locus 87 (VWF)

| Study | Cases | Controls | OR |
|---|---|---|---|
| INVENT-2019_EA | 15572 | 115280 | 1.09 |
| MVP_EA | 18952 | 411746 | 1.09 |
| FinnGen_EA | 9176 | 209616 | 1.06 |
| EGP_EA | 4122 | 181553 | 1.05 |
| UKB_EA | 18385 | 91834 | 1.07 |
| MGB_EA | 2967 | 27957 | 1.01 |
| UPenn_EA | 380 | 6210 | 0.99 |
| FARIVE_EA | 545 | 532 | 1.2 |
| BioME_EA | 200 | 8007 | 1.14 |
| RETROVE_EA | 400 | 400 | 1.09 |
| GAIT2_EA | 85 | 850 | 0.89 |
| MESA_EA | 94 | 2341 | 1.27 |
| MARTHA12_EA | 893 | 3414 | 1.04 |
| **European** | **71771** | **1059740** | **1.07** |

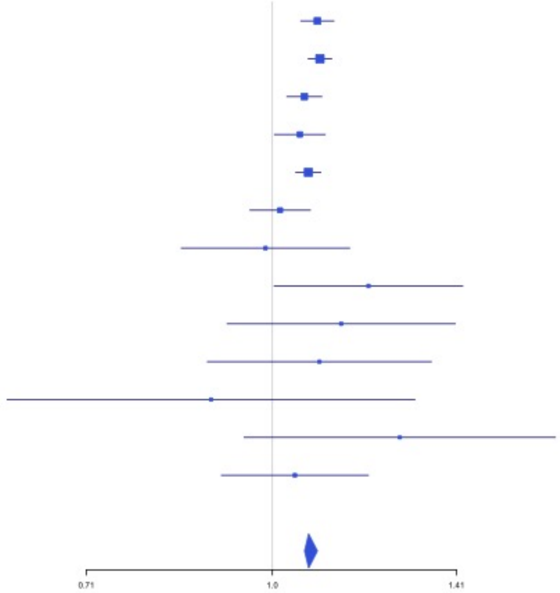


**Supplementary Figure 3.64** – rs7311483 (chr12:9053661) in locus 88 (A2ML1 ;PHC1)

| Study | Cases | Controls | OR |
|---|---|---|---|
| INVENT-2019_EA | 15572 | 115280 | 0.98 |
| MVP_EA | 18952 | 411746 | 0.95 |
| FinnGen_EA | 9176 | 209616 | 0.95 |
| EGP_EA | 4122 | 181553 | 0.95 |
| UKB_EA | 18385 | 91834 | 0.97 |
| MGB_EA | 2967 | 27957 | 0.97 |
| UPenn_EA | 380 | 6210 | 1.05 |
| FARIVE_EA | 545 | 532 | 0.94 |
| BioME_EA | 200 | 8007 | 0.85 |
| RETROVE_EA | 400 | 400 | 1.05 |
| GAIT2_EA | 85 | 850 | 1.12 |
| MESA_EA | 94 | 2341 | 0.88 |
| **European** | **70878** | **1056326** | **0.96** |

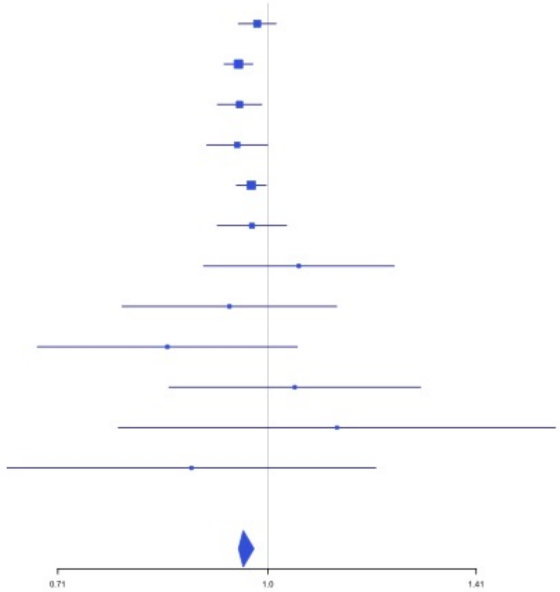

**Supplementary Figure 3.65** – rs4759076 (chr12:54729872) in locus 89 (COPZ1)

| Study | Cases | Controls | OR |
|---|---|---|---|
| INVENT-2019_EA | 15572 | 115280 | 1.04 |
| MVP_EA | 18952 | 411746 | 1.06 |
| FinnGen_EA | 9176 | 209616 | 1.03 |
| EGP_EA | 4122 | 181553 | 1.02 |
| UKB_EA | 18385 | 91834 | 1.07 |
| UPenn_EA | 380 | 6210 | 1.05 |
| FARIVE_EA | 545 | 532 | 1.14 |
| BioME_EA | 200 | 8007 | 0.98 |
| RETROVE_EA | 400 | 400 | 1.22 |
| GAIT2_EA | 85 | 850 | 1.13 |
| MESA_EA | 94 | 2341 | 1.18 |
| MARTHA12_EA | 893 | 3414 | 1.16 |
| **European** | **68804** | **1031783** | **1.05** |

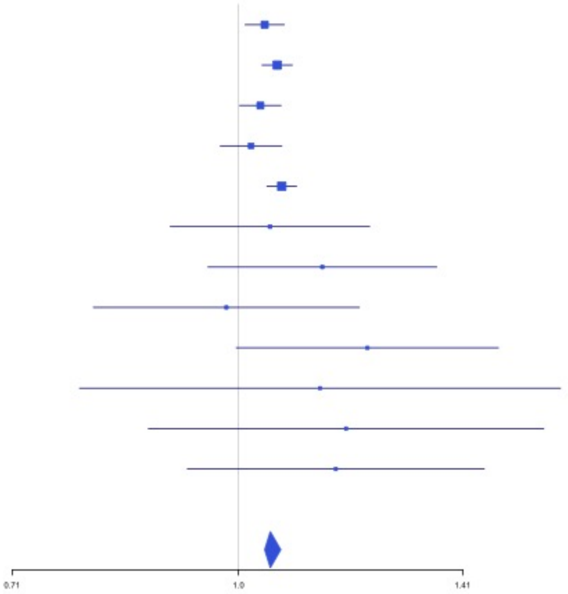


**Supplementary Figure 3.66** – rs142351376 (chr12:104136288) in locus 90 (STAB2)

| Study | Cases | Controls | OR |
|---|---|---|---|
| MVP_EA | 18952 | 411746 | 1.78 |
| FinnGen_EA | 9176 | 209616 | 1.67 |
| EGP_EA | 4122 | 181553 | 1.91 |
| UPenn_EA | 380 | 6210 | 0.99 |
| **European** | **32630** | **809125** | **1.68** |

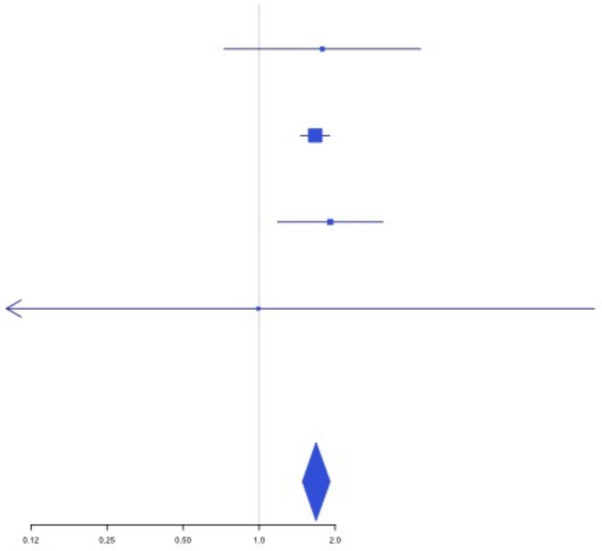

**Supplementary Figure 3.67** – rs56750693 (chr12:112144586) in locus 91 (ACAD10)

| Study | Cases | Controls | OR |
|---|---|---|---|
| INVENT-2019_EA | 15572 | 115280 | 1.06 |
| MVP_EA | 18952 | 411746 | 1.07 |
| UKB_EA | 18385 | 91834 | 1.07 |
| UPenn_EA | 380 | 6210 | 1.03 |
| FARIVE_EA | 545 | 532 | 0.98 |
| MARTHA12_EA | 893 | 3414 | 1.28 |
| **European** | **54727** | **629016** | **1.07** |

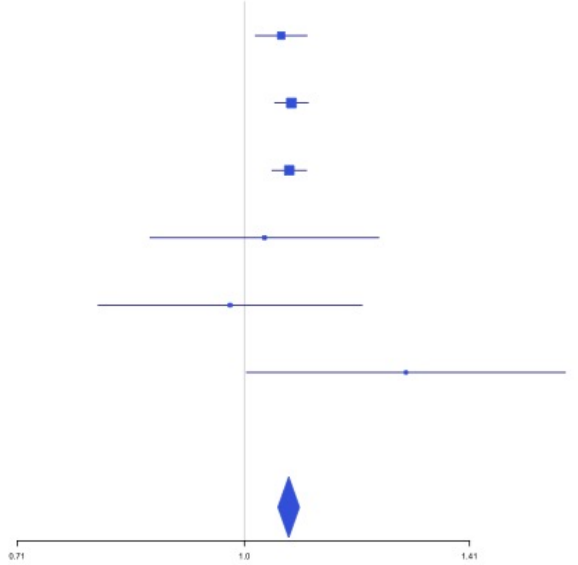


**Supplementary Figure 3.68** – rs2851436 (chr12:123667354) in locus 92 (MPHOSPH9)

| Study | Cases | Controls | OR |
|---|---|---|---|
| INVENT-2019_EA | 15572 | 115280 | 1.05 |
| MVP_EA | 18952 | 411746 | 1.05 |
| FinnGen_EA | 9176 | 209616 | 1.03 |
| EGP_EA | 4122 | 181553 | 1.03 |
| UKB_EA | 18385 | 91834 | 1.06 |
| MGB_EA | 2967 | 27957 | 1 |
| UPenn_EA | 380 | 6210 | 1.2 |
| FARIVE_EA | 545 | 532 | 1.07 |
| BioME_EA | 200 | 8007 | 1.15 |
| RETROVE_EA | 400 | 400 | 1.12 |
| GAIT2_EA | 85 | 850 | 0.91 |
| MESA_EA | 94 | 2341 | 1.2 |
| MARTHA12_EA | 893 | 3414 | 1.09 |
| **European** | **71771** | **1059740** | **1.05** |

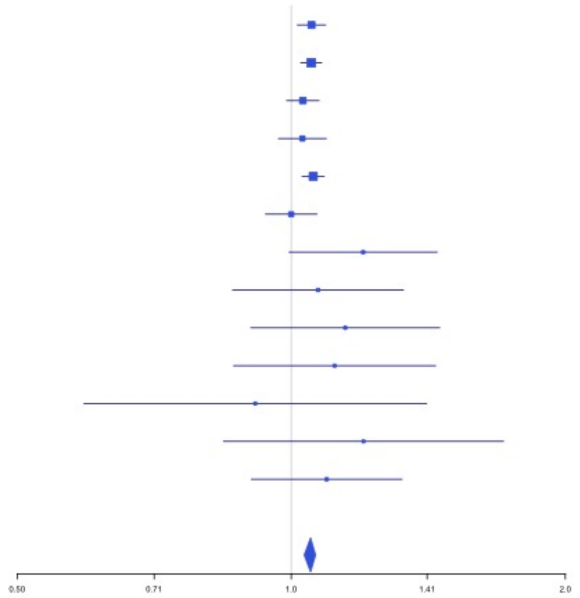

**Supplementary Figure 3.69** – rs3211752 (chr13:113787459) in locus 94 (F10)

| Study | Cases | Controls | OR |
|---|---|---|---|
| INVENT-2019_EA | 15572 | 115280 | 0.95 |
| MVP_EA | 18952 | 411746 | 0.95 |
| FinnGen_EA | 9176 | 209616 | 0.94 |
| EGP_EA | 4122 | 181553 | 0.94 |
| UKB_EA | 18385 | 91834 | 0.96 |
| MGB_EA | 2967 | 27957 | 0.98 |
| UPenn_EA | 380 | 6210 | 0.93 |
| FARIVE_EA | 545 | 532 | 1.03 |
| BioME_EA | 200 | 8007 | 0.96 |
| RETROVE_EA | 400 | 400 | 0.97 |
| GAIT2_EA | 85 | 850 | 0.8 |
| MESA_EA | 94 | 2341 | 0.72 |
| **European** | **70878** | **1056326** | **0.95** |

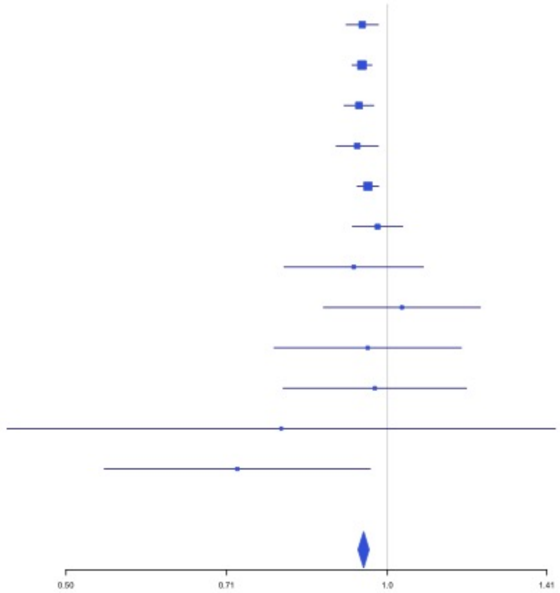


**Supplementary Figure 3.70** – rs8004077 (chr14:58846731) in locus 95 (ARID4A ;TOMM20L)

| Study | Cases | Controls | OR |
|---|---|---|---|
| INVENT-2019_EA | 15572 | 115280 | 1.09 |
| MVP_EA | 18952 | 411746 | 1.02 |
| FinnGen_EA | 9176 | 209616 | 1.03 |
| EGP_EA | 4122 | 181553 | 1.07 |
| UKB_EA | 18385 | 91834 | 1.05 |
| MGB_EA | 2967 | 27957 | 1.1 |
| UPenn_EA | 380 | 6210 | 0.84 |
| FARIVE_EA | 545 | 532 | 1.24 |
| BioME_EA | 200 | 8007 | 1.2 |
| RETROVE_EA | 400 | 400 | 1 |
| GAIT2_EA | 85 | 850 | 0.96 |
| MESA_EA | 94 | 2341 | 1.08 |
| MARTHA12_EA | 893 | 3414 | 1.07 |
| **European** | **71771** | **1059740** | **1.04** |

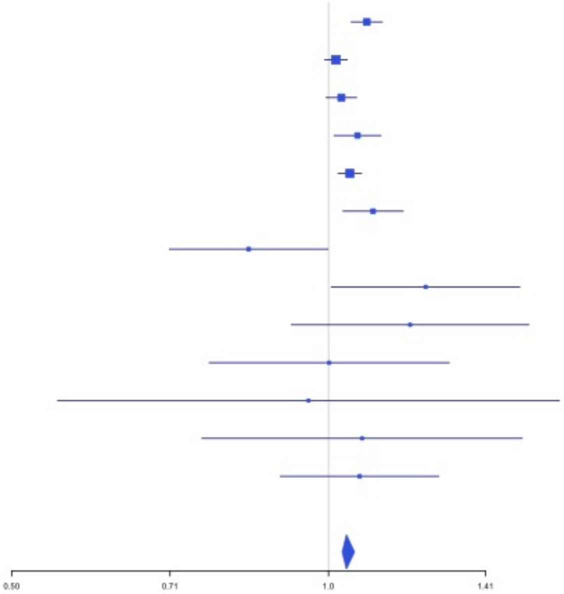

**Supplementary Figure 3.71** – rs2127869 (chr14:65794352) in locus 96 (LINC02324;MIR4708)

| Study | Cases | Controls | OR |
|---|---|---|---|
| INVENT-2019_EA | 15572 | 115280 | 0.97 |
| MVP_EA | 18952 | 411746 | 0.95 |
| FinnGen_EA | 9176 | 209616 | 0.98 |
| EGP_EA | 4122 | 181553 | 0.99 |
| UKB_EA | 18385 | 91834 | 0.97 |
| MGB_EA | 2967 | 27957 | 0.96 |
| UPenn_EA | 380 | 6210 | 1.03 |
| FARIVE_EA | 545 | 532 | 1.04 |
| BioME_EA | 200 | 8007 | 0.78 |
| RETROVE_EA | 400 | 400 | 1.11 |
| GAIT2_EA | 85 | 850 | 0.86 |
| MESA_EA | 94 | 2341 | 0.92 |
| **European** | **70878** | **1056326** | **0.96** |

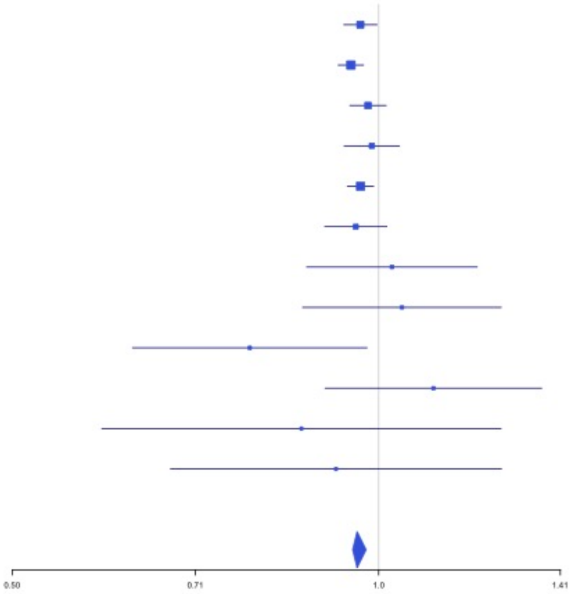


**Supplementary Figure 3.72** – rs61990092 (chr14:92289301) in locus 97 (TC2N)

| Study | Cases | Controls | OR |
|---|---|---|---|
| INVENT-2019_EA | 15572 | 115280 | 1.07 |
| MVP_EA | 18952 | 411746 | 1.07 |
| FinnGen_EA | 9176 | 209616 | 1.06 |
| EGP_EA | 4122 | 181553 | 1.09 |
| UKB_EA | 18385 | 91834 | 1.05 |
| MGB_EA | 2967 | 27957 | 1 |
| UPenn_EA | 380 | 6210 | 1.01 |
| FARIVE_EA | 545 | 532 | 1.01 |
| BioME_EA | 200 | 8007 | 1.04 |
| RETROVE_EA | 400 | 400 | 1.07 |
| GAIT2_EA | 85 | 850 | 1.01 |
| MESA_EA | 94 | 2341 | 1.05 |
| **European** | **70878** | **1056326** | **1.06** |

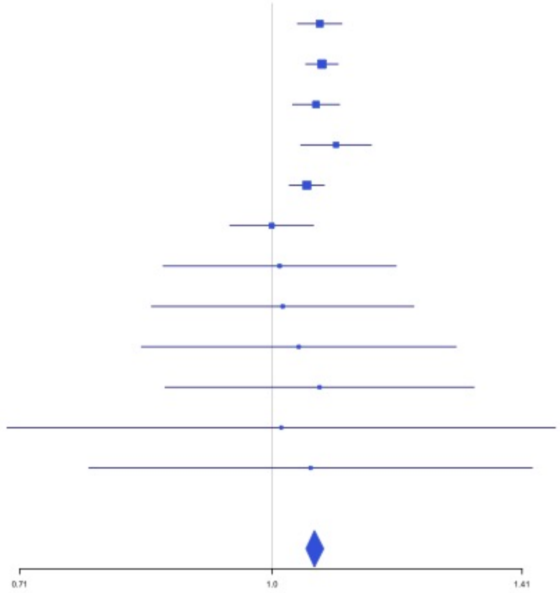

**Supplementary Figure 3.73** – rs28929474 (chr14:94844947) in locus 98 (SERPINA1)

| Study | Cases | Controls | OR |
|---|---|---|---|
| INVENT-2019_EA | 15572 | 115280 | 1.1 |
| MVP_EA | 18952 | 411746 | 1.09 |
| FinnGen_EA | 9176 | 209616 | 1.2 |
| EGP_EA | 4122 | 181553 | 0.99 |
| UKB_EA | 18385 | 91834 | 1.2 |
| MGB_EA | 2967 | 27957 | 1.08 |
| FARIVE_EA | 545 | 532 | 0.86 |
| BioME_EA | 200 | 8007 | 1.38 |
| RETROVE_EA | 400 | 400 | 2.36 |
| GAIT2_EA | 85 | 850 | 4.05 |
| MESA_EA | 94 | 2341 | 0.25 |
| MARTHA12_EA | 893 | 3414 | 0.77 |
| **European** | **71391** | **1053530** | **1.13** |

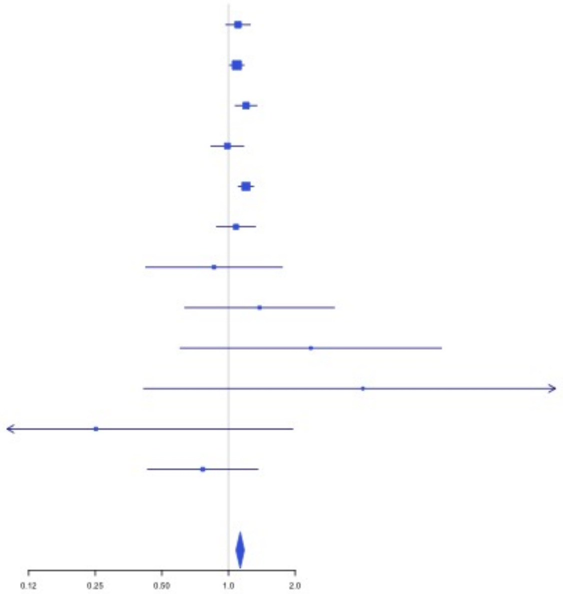


**Supplementary Figure 3.74** – rs4906229 (chr14:103057511) in locus 99 (LINC02323;RCOR1)

| Study | Cases | Controls | OR |
|---|---|---|---|
| INVENT-2019_EA | 15572 | 115280 | 1.02 |
| MVP_EA | 18952 | 411746 | 1.03 |
| FinnGen_EA | 9176 | 209616 | 1.05 |
| EGP_EA | 4122 | 181553 | 1.08 |
| UKB_EA | 18385 | 91834 | 1.05 |
| MGB_EA | 2967 | 27957 | 1.07 |
| UPenn_EA | 380 | 6210 | 0.88 |
| FARIVE_EA | 545 | 532 | 0.88 |
| BioME_EA | 200 | 8007 | 1.02 |
| RETROVE_EA | 400 | 400 | 1.02 |
| GAIT2_EA | 85 | 850 | 1.15 |
| MESA_EA | 94 | 2341 | 0.95 |
| **European** | **70878** | **1056326** | **1.04** |

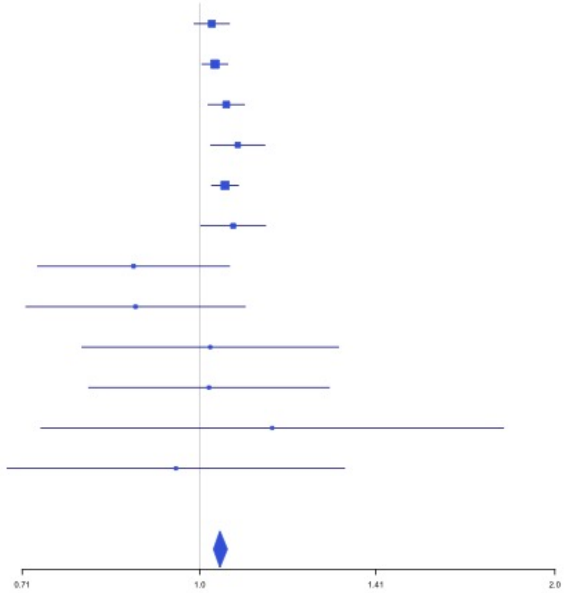

**Supplementary Figure 3.75** – rs141866277 (chr15:43950699) in locus 100 (PPIP5K1P1-CATSPER2)

| Study | Cases | Controls | OR |
|---|---|---|---|
| INVENT-2019_EA | 15572 | 115280 | 1.08 |
| MVP_EA | 18952 | 411746 | 1.2 |
| FinnGen_EA | 9176 | 209616 | 1.23 |
| EGP_EA | 4122 | 181553 | 1.14 |
| UKB_EA | 18385 | 91834 | 1.21 |
| MGB_EA | 2967 | 27957 | 1.2 |
| UPenn_EA | 380 | 6210 | 1.26 |
| FARIVE_EA | 545 | 532 | 1.89 |
| RETROVE_EA | 400 | 400 | 2.33 |
| GAIT2_EA | 85 | 850 | 1.21 |
| MESA_EA | 94 | 2341 | 0.81 |
| MARTHA12_EA | 893 | 3414 | 1.17 |
| **European** | **71571** | **1051733** | **1.18** |

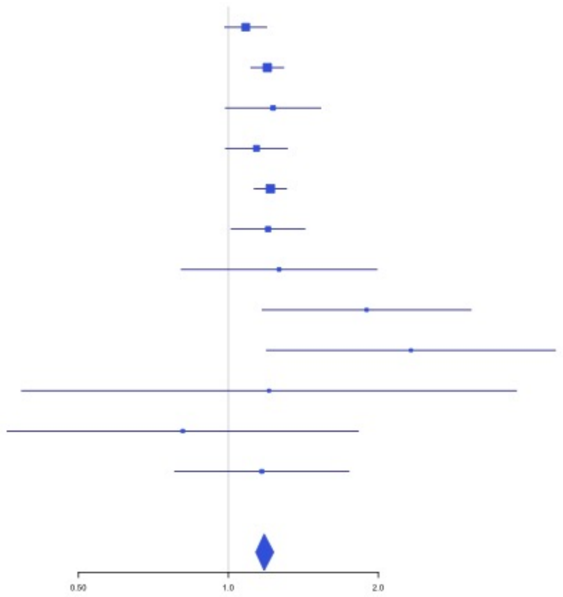


**Supplementary Figure 3.76** – rs340009 (chr15:60899639) in locus 101 (RORA-AS1)

| Study | Cases | Controls | OR |
|---|---|---|---|
| INVENT-2019_EA | 15572 | 115280 | 0.96 |
| MVP_EA | 18952 | 411746 | 0.96 |
| FinnGen_EA | 9176 | 209616 | 0.97 |
| EGP_EA | 4122 | 181553 | 0.95 |
| UKB_EA | 18385 | 91834 | 0.98 |
| MGB_EA | 2967 | 27957 | 0.96 |
| UPenn_EA | 380 | 6210 | 1.1 |
| FARIVE_EA | 545 | 532 | 1.06 |
| BioME_EA | 200 | 8007 | 0.82 |
| RETROVE_EA | 400 | 400 | 0.88 |
| GAIT2_EA | 85 | 850 | 1.12 |
| MESA_EA | 94 | 2341 | 1.18 |
| MARTHA12_EA | 893 | 3414 | 0.95 |
| **European** | **71771** | **1059740** | **0.97** |

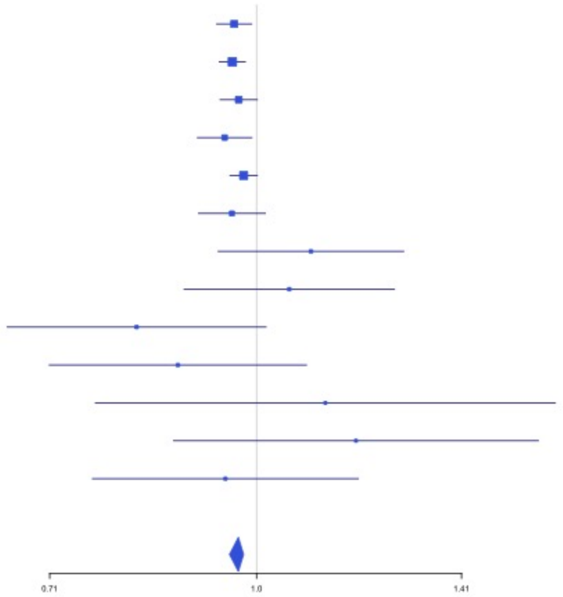

**Supplementary Figure 3.77** – rs7183672 (chr15:96101018) in locus 103 (LINC00924 ;LOC105369212)

| Study | Cases | Controls | OR |
|---|---|---|---|
| INVENT-2019_EA | 15572 | 115280 | 0.98 |
| MVP_EA | 18952 | 411746 | 0.96 |
| FinnGen_EA | 9176 | 209616 | 0.98 |
| EGP_EA | 4122 | 181553 | 0.97 |
| UKB_EA | 18385 | 91834 | 0.95 |
| MGB_EA | 2967 | 27957 | 1.03 |
| UPenn_EA | 380 | 6210 | 0.99 |
| FARIVE_EA | 545 | 532 | 1.03 |
| BioME_EA | 200 | 8007 | 0.96 |
| RETROVE_EA | 400 | 400 | 0.95 |
| GAIT2_EA | 85 | 850 | 0.73 |
| MESA_EA | 94 | 2341 | 1.03 |
| **European** | **70878** | **1056326** | **0.96** |

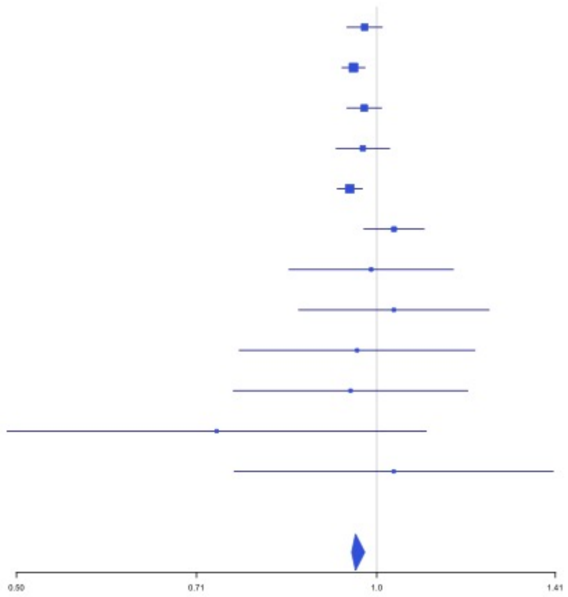


**Supplementary Figure 3.78** – rs12443808 (chr16:30996871) in locus 106 (HSD3B7)

| Study | Cases | Controls | OR |
|---|---|---|---|
| INVENT-2019_EA | 15572 | 115280 | 1.06 |
| MVP_EA | 18952 | 411746 | 1.06 |
| FinnGen_EA | 9176 | 209616 | 1.05 |
| EGP_EA | 4122 | 181553 | 1.02 |
| UKB_EA | 18385 | 91834 | 1.03 |
| MGB_EA | 2967 | 27957 | 1.02 |
| UPenn_EA | 380 | 6210 | 0.96 |
| FARIVE_EA | 545 | 532 | 1.12 |
| RETROVE_EA | 400 | 400 | 1.09 |
| GAIT2_EA | 85 | 850 | 0.71 |
| MESA_EA | 94 | 2341 | 0.91 |
| MARTHA12_EA | 893 | 3414 | 1.14 |
| **European** | **71571** | **1051733** | **1.04** |

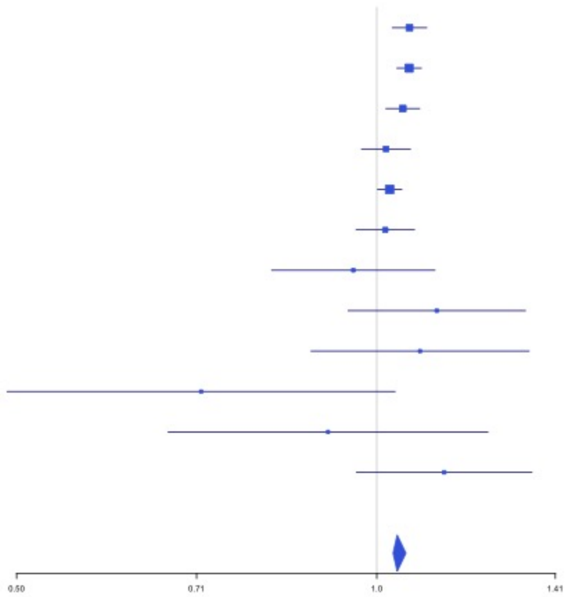

**Supplementary Figure 3.79** – rs1346057617 (chr16:75434183) in locus 108 (CFDP1)

| Study | Cases | Controls | OR |
|---|---|---|---|
| INVENT-2019_EA | 15572 | 115280 | 0.96 |
| MVP_EA | 18952 | 411746 | 0.96 |
| UKB_EA | 18385 | 91834 | 0.95 |
| UPenn_EA | 380 | 6210 | 0.95 |
| FARIVE_EA | 545 | 532 | 0.99 |
| **European** | **53834** | **625602** | **0.96** |

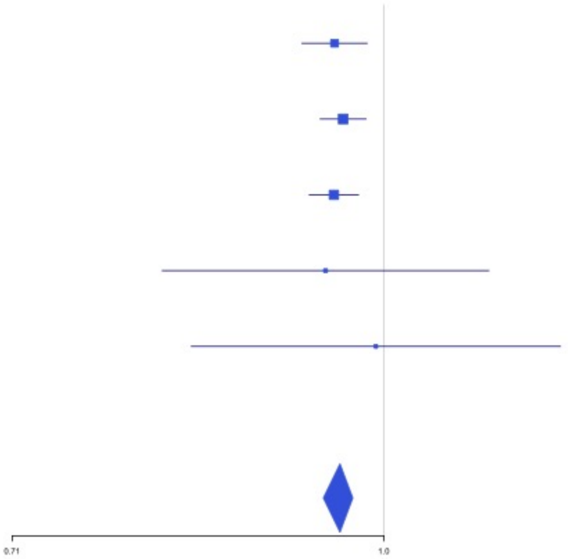


**Supplementary Figure 3.80** – rs62044142 (chr16:81868968) in locus 109 (PLCG2)

| Study | Cases | Controls | OR |
|---|---|---|---|
| INVENT-2019_EA | 15572 | 115280 | 0.91 |
| FinnGen_EA | 9176 | 209616 | 0.92 |
| EGP_EA | 4122 | 181553 | 0.94 |
| UKB_EA | 18385 | 91834 | 0.9 |
| MGB_EA | 2967 | 27957 | 0.94 |
| UPenn_EA | 380 | 6210 | 0.94 |
| FARIVE_EA | 545 | 532 | 0.68 |
| BioME_EA | 200 | 8007 | 1.05 |
| RETROVE_EA | 400 | 400 | 0.64 |
| GAIT2_EA | 85 | 850 | 0.6 |
| MESA_EA | 94 | 2341 | 0.89 |
| **European** | **51926** | **644580** | **0.91** |

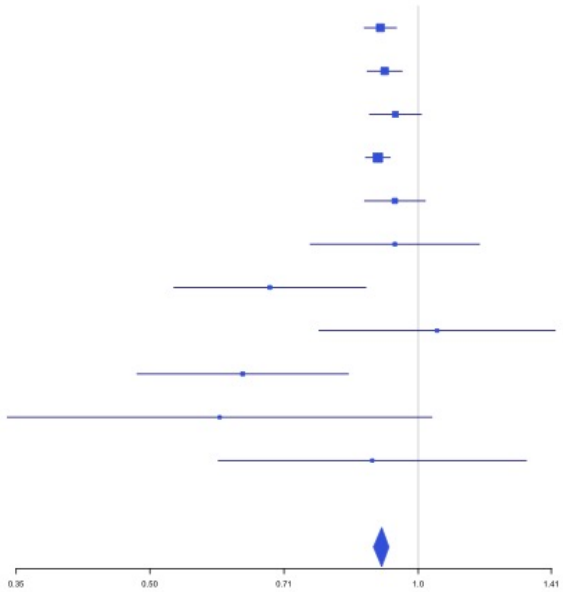

**Supplementary Figure 3.81** – rs28634651 (chr16:88553198) in locus 111 (ZFPM1)

| Study | Cases | Controls | OR |
|---|---|---|---|
| INVENT-2019_EA | 15572 | 115280 | 1.04 |
| MVP_EA | 18952 | 411746 | 1.06 |
| FinnGen_EA | 9176 | 209616 | 1.07 |
| UKB_EA | 18385 | 91834 | 1.04 |
| MGB_EA | 2967 | 27957 | 1.03 |
| UPenn_EA | 380 | 6210 | 1.04 |
| FARIVE_EA | 545 | 532 | 1.16 |
| BioME_EA | 200 | 8007 | 1 |
| RETROVE_EA | 400 | 400 | 1 |
| GAIT2_EA | 85 | 850 | 0.96 |
| MESA_EA | 94 | 2341 | 0.8 |
| **European** | **66756** | **874773** | **1.05** |

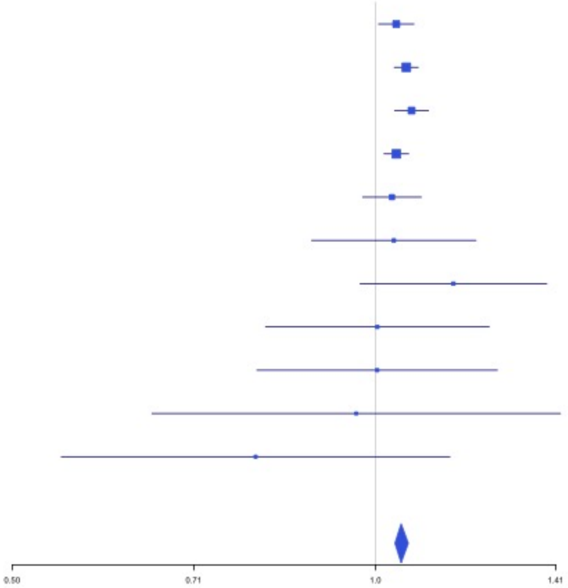


**Supplementary Figure 3.82** – rs4790311 (chr17:1979188) in locus 112 (SMG6)

| Study | Cases | Controls | OR |
|---|---|---|---|
| INVENT-2019_EA | 15572 | 115280 | 0.95 |
| MVP_EA | 18952 | 411746 | 0.95 |
| FinnGen_EA | 9176 | 209616 | 0.97 |
| EGP_EA | 4122 | 181553 | 0.94 |
| UKB_EA | 18385 | 91834 | 0.96 |
| MGB_EA | 2967 | 27957 | 0.94 |
| UPenn_EA | 380 | 6210 | 1.07 |
| FARIVE_EA | 545 | 532 | 0.89 |
| BioME_EA | 200 | 8007 | 1.06 |
| RETROVE_EA | 400 | 400 | 0.98 |
| GAIT2_EA | 85 | 850 | 0.95 |
| MESA_EA | 94 | 2341 | 0.97 |
| **European** | **70878** | **1056326** | **0.95** |

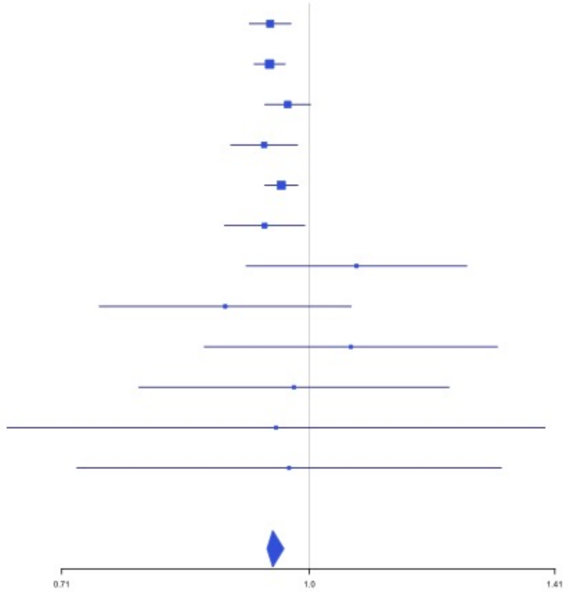

**Supplementary Figure 3.83** – rs11078659 (chr17:6903944) in locus 113 (ALOX12-AS1)

| Study | Cases | Controls | OR |
|---|---|---|---|
| INVENT-2019_EA | 15572 | 115280 | 0.96 |
| MVP_EA | 18952 | 411746 | 0.95 |
| FinnGen_EA | 9176 | 209616 | 0.97 |
| EGP_EA | 4122 | 181553 | 0.99 |
| UKB_EA | 18385 | 91834 | 0.97 |
| MGB_EA | 2967 | 27957 | 1 |
| UPenn_EA | 380 | 6210 | 0.93 |
| FARIVE_EA | 545 | 532 | 0.81 |
| BioME_EA | 200 | 8007 | 0.88 |
| RETROVE_EA | 400 | 400 | 0.98 |
| GAIT2_EA | 85 | 850 | 0.77 |
| MESA_EA | 94 | 2341 | 1.26 |
| MARTHA12_EA | 893 | 3414 | 1.05 |
| **European** | **71771** | **1059740** | **0.97** |

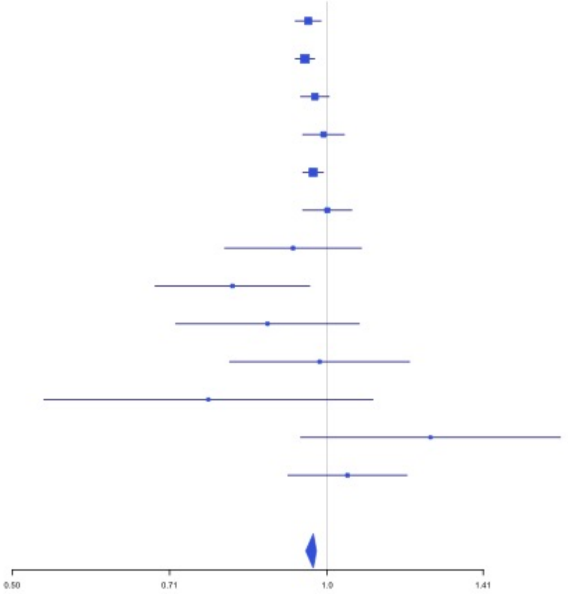


**Supplementary Figure 3.84** – rs536327 (chr17:27857698) in locus 114 (TAOK1)

| Study | Cases | Controls | OR |
|---|---|---|---|
| INVENT-2019_EA | 15572 | 115280 | 1.04 |
| MVP_EA | 18952 | 411746 | 1.02 |
| FinnGen_EA | 9176 | 209616 | 1.05 |
| EGP_EA | 4122 | 181553 | 1.03 |
| UKB_EA | 18385 | 91834 | 1.04 |
| UPenn_EA | 380 | 6210 | 1.15 |
| FARIVE_EA | 545 | 532 | 0.91 |
| BioME_EA | 200 | 8007 | 1.02 |
| RETROVE_EA | 400 | 400 | 1.21 |
| GAIT2_EA | 85 | 850 | 1.15 |
| MESA_EA | 94 | 2341 | 0.92 |
| **European** | **67911** | **1028369** | **1.03** |

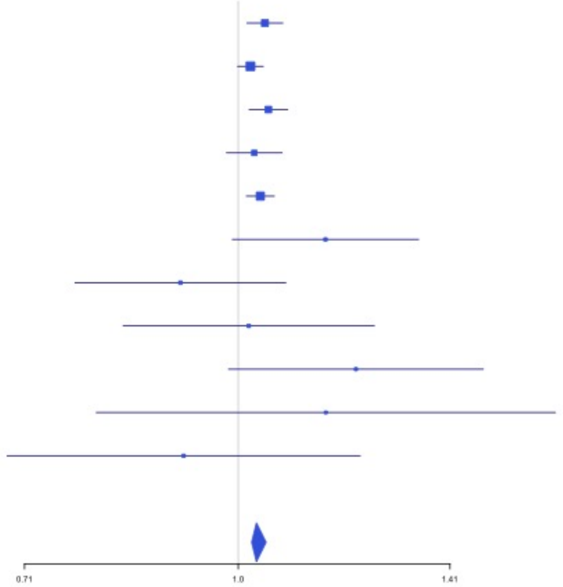

**Supplementary Figure 3.85** – rs9468 (chr17:44101563) in locus 115 (MAPT)

| Study | Cases | Controls | OR |
|---|---|---|---|
| INVENT-2019_EA | 15572 | 115280 | 0.95 |
| MVP_EA | 18952 | 411746 | 0.95 |
| FinnGen_EA | 9176 | 209616 | 0.96 |
| EGP_EA | 4122 | 181553 | 0.97 |
| UKB_EA | 18385 | 91834 | 0.96 |
| MGB_EA | 2967 | 27957 | 0.93 |
| UPenn_EA | 380 | 6210 | 0.97 |
| FARIVE_EA | 545 | 532 | 0.98 |
| BioME_EA | 200 | 8007 | 0.9 |
| RETROVE_EA | 400 | 400 | 0.8 |
| GAIT2_EA | 85 | 850 | 0.82 |
| MESA_EA | 94 | 2341 | 1 |
| MARTHA12_EA | 893 | 3414 | 0.86 |
| **European** | **71771** | **1059740** | **0.95** |

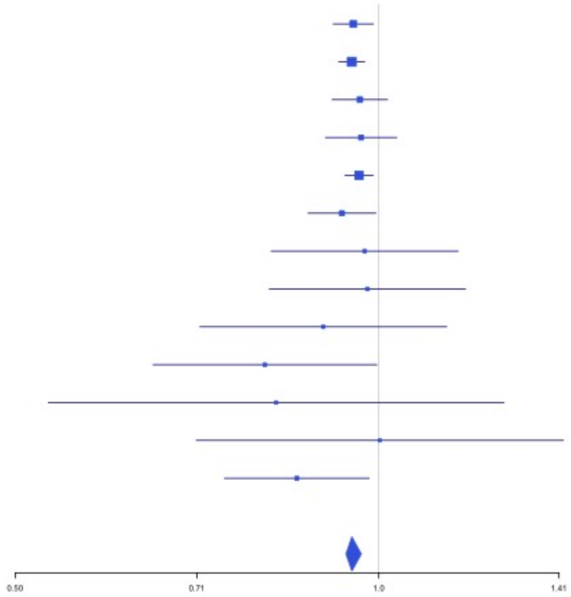


**Supplementary Figure 3.86** – rs1801690 (chr17:64208285) in locus 116 (APOH)

| Study | Cases | Controls | OR |
|---|---|---|---|
| INVENT-2019_EA | 15572 | 115280 | 1.03 |
| MVP_EA | 18952 | 411746 | 1.11 |
| FinnGen_EA | 9176 | 209616 | 1.11 |
| EGP_EA | 4122 | 181553 | 1.04 |
| UKB_EA | 18385 | 91834 | 1.04 |
| MGB_EA | 2967 | 27957 | 0.97 |
| UPenn_EA | 380 | 6210 | 1.04 |
| FARIVE_EA | 545 | 532 | 1.3 |
| BioME_EA | 200 | 8007 | 0.93 |
| RETROVE_EA | 400 | 400 | 1.46 |
| GAIT2_EA | 85 | 850 | 1.14 |
| MARTHA12_EA | 893 | 3414 | 1.33 |
| **European** | **71677** | **1057399** | **1.07** |

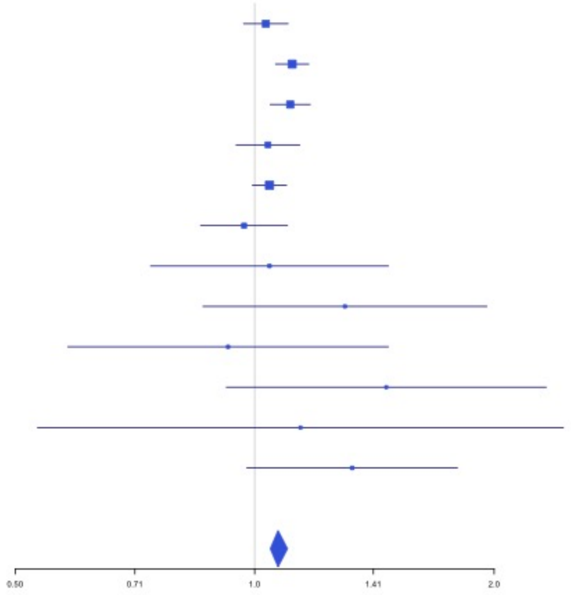

**Supplementary Figure 3.87** – rs573431210 (chr17:67276383) in locus 117 (ABCA5)

| Study | Cases | Controls | OR |
|---|---|---|---|
| INVENT-2019_EA | 15572 | 115280 | 1.14 |
| MVP_EA | 18952 | 411746 | 0.88 |
| FinnGen_EA | 9176 | 209616 | 0.95 |
| EGP_EA | 4122 | 181553 | 0.69 |
| UKB_EA | 18385 | 91834 | 0.82 |
| MGB_EA | 2967 | 27957 | 0.93 |
| UPenn_EA | 380 | 6210 | 1.24 |
| FARIVE_EA | 545 | 532 | 0.52 |
| BioME_EA | 200 | 8007 | 1.19 |
| MARTHA12_EA | 893 | 3414 | 0.71 |
| **European** | **71192** | **1056149** | **0.87** |

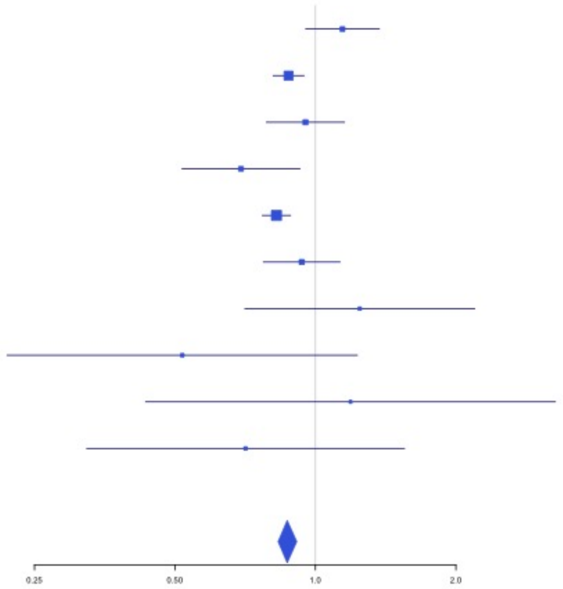


**Supplementary Figure 3.88** – rs8109681 (chr19:10738836) in locus 119 (SLC44A2)

| Study | Cases | Controls | OR |
|---|---|---|---|
| INVENT-2019_EA | 15572 | 115280 | 0.89 |
| MVP_EA | 18952 | 411746 | 0.88 |
| FinnGen_EA | 9176 | 209616 | 0.87 |
| EGP_EA | 4122 | 181553 | 0.93 |
| UKB_EA | 18385 | 91834 | 0.9 |
| MGB_EA | 2967 | 27957 | 0.93 |
| UPenn_EA | 380 | 6210 | 1.07 |
| FARIVE_EA | 545 | 532 | 0.84 |
| BioME_EA | 200 | 8007 | 0.88 |
| RETROVE_EA | 400 | 400 | 0.72 |
| GAIT2_EA | 85 | 850 | 0.68 |
| MESA_EA | 94 | 2341 | 0.91 |
| MARTHA12_EA | 893 | 3414 | 0.86 |
| **European** | **71771** | **1059740** | **0.89** |

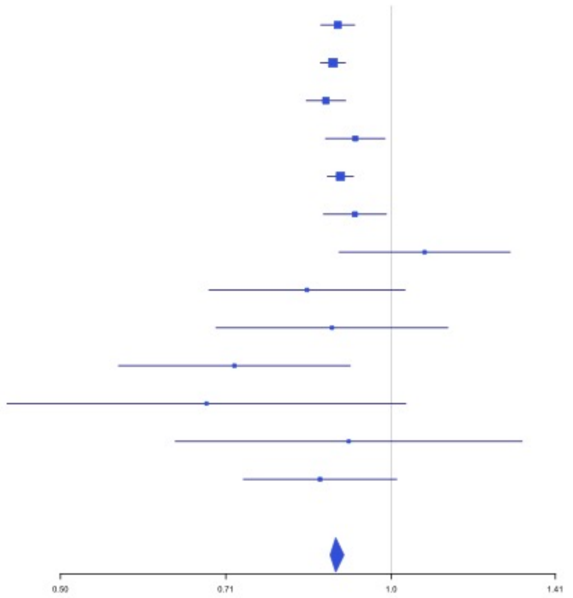

**Supplementary Figure 3.89** – rs2545774 (chr19:41287674) in locus 121 (RAB4B)

| Study | Cases | Controls | OR |
|---|---|---|---|
| INVENT-2019_EA | 15572 | 115280 | 0.95 |
| MVP_EA | 18952 | 411746 | 0.97 |
| FinnGen_EA | 9176 | 209616 | 0.94 |
| EGP_EA | 4122 | 181553 | 0.93 |
| UKB_EA | 18385 | 91834 | 0.97 |
| MGB_EA | 2967 | 27957 | 0.96 |
| UPenn_EA | 380 | 6210 | 0.93 |
| FARIVE_EA | 545 | 532 | 0.78 |
| BioME_EA | 200 | 8007 | 0.88 |
| RETROVE_EA | 400 | 400 | 1.26 |
| GAIT2_EA | 85 | 850 | 1 |
| MESA_EA | 94 | 2341 | 0.9 |
| **European** | **70878** | **1056326** | **0.96** |

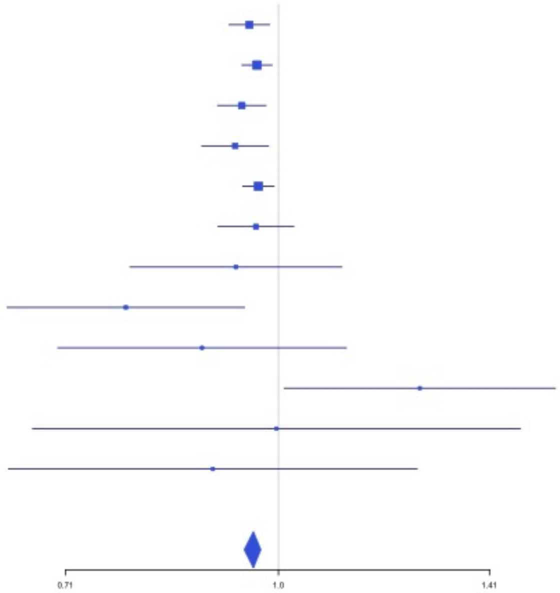


**Supplementary Figure 3.90** – rs8108474 (chr19:46301479) in locus 122 (RSPH6A)

| Study | Cases | Controls | OR |
|---|---|---|---|
| INVENT-2019_EA | 15572 | 115280 | 1.05 |
| MVP_EA | 18952 | 411746 | 1.04 |
| FinnGen_EA | 9176 | 209616 | 1.05 |
| EGP_EA | 4122 | 181553 | 1 |
| UKB_EA | 18385 | 91834 | 1.05 |
| MGB_EA | 2967 | 27957 | 1.04 |
| UPenn_EA | 380 | 6210 | 0.93 |
| FARIVE_EA | 545 | 532 | 0.88 |
| BioME_EA | 200 | 8007 | 1.04 |
| RETROVE_EA | 400 | 400 | 1.01 |
| GAIT2_EA | 85 | 850 | 1.02 |
| MESA_EA | 94 | 2341 | 1.01 |
| **European** | **70878** | **1056326** | **1.04** |

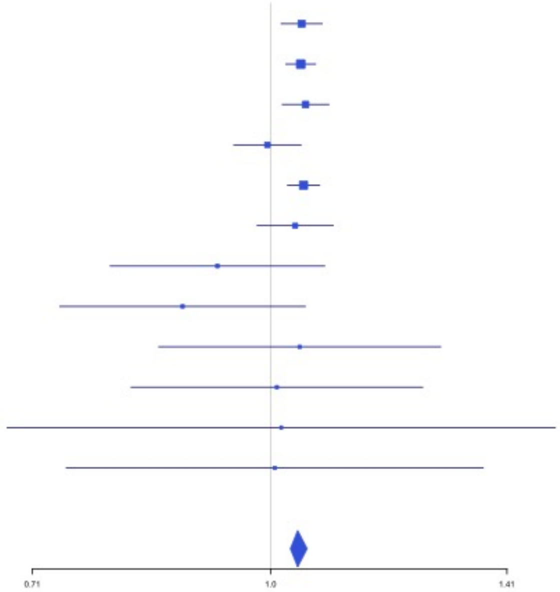

**Supplementary Figure 3.91** – rs646327 (chr19:49209851) in locus 123 (FUT2)

| Study | Cases | Controls | OR |
|---|---|---|---|
| INVENT-2019_EA | 15572 | 115280 | 0.96 |
| MVP_EA | 18952 | 411746 | 0.96 |
| FinnGen_EA | 9176 | 209616 | 0.97 |
| EGP_EA | 4122 | 181553 | 0.95 |
| UKB_EA | 18385 | 91834 | 0.95 |
| MGB_EA | 2967 | 27957 | 0.98 |
| UPenn_EA | 380 | 6210 | 0.96 |
| FARIVE_EA | 545 | 532 | 0.96 |
| BioME_EA | 200 | 8007 | 1 |
| RETROVE_EA | 400 | 400 | 0.88 |
| GAIT2_EA | 85 | 850 | 0.8 |
| MESA_EA | 94 | 2341 | 0.95 |
| MARTHA12_EA | 893 | 3414 | 0.94 |
| **European** | **71771** | **1059740** | **0.96** |

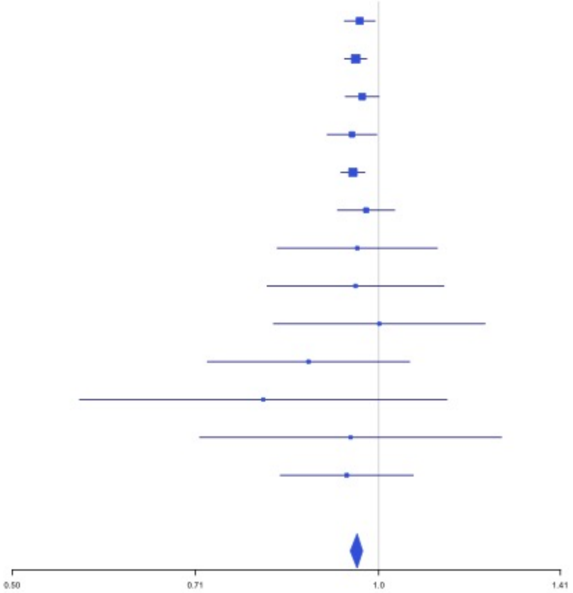


**Supplementary Figure 3.92** – rs1654425 (chr19:55538980) in locus 124 (GP6)

| Study | Cases | Controls | OR |
|---|---|---|---|
| INVENT-2019_EA | 15572 | 115280 | 0.93 |
| MVP_EA | 18952 | 411746 | 0.9 |
| FinnGen_EA | 9176 | 209616 | 0.92 |
| EGP_EA | 4122 | 181553 | 0.93 |
| UKB_EA | 18385 | 91834 | 0.93 |
| MGB_EA | 2967 | 27957 | 0.89 |
| UPenn_EA | 380 | 6210 | 0.97 |
| FARIVE_EA | 545 | 532 | 0.78 |
| BioME_EA | 200 | 8007 | 1.06 |
| RETROVE_EA | 400 | 400 | 0.67 |
| GAIT2_EA | 85 | 850 | 1.09 |
| MESA_EA | 94 | 2341 | 0.82 |
| MARTHA12_EA | 893 | 3414 | 0.96 |
| **European** | **71771** | **1059740** | **0.92** |

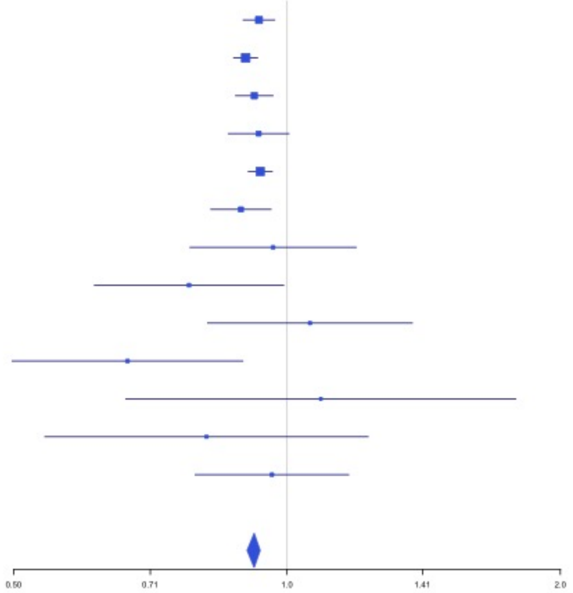

**Supplementary Figure 3.93** – rs117390891 (chr20:23168526) in locus 126 (LINC00656 ;NXT1)

| Study | Cases | Controls | OR |
|---|---|---|---|
| INVENT-2019_EA | 15572 | 115280 | 0.93 |
| MVP_EA | 18952 | 411746 | 0.9 |
| FinnGen_EA | 9176 | 209616 | 0.91 |
| EGP_EA | 4122 | 181553 | 0.94 |
| UKB_EA | 18385 | 91834 | 0.93 |
| MGB_EA | 2967 | 27957 | 1.03 |
| UPenn_EA | 380 | 6210 | 0.96 |
| FARIVE_EA | 545 | 532 | 0.92 |
| BioME_EA | 200 | 8007 | 1.08 |
| RETROVE_EA | 400 | 400 | 0.9 |
| GAIT2_EA | 85 | 850 | 1.05 |
| MESA_EA | 94 | 2341 | 0.63 |
| **European** | **70878** | **1056326** | **0.93** |

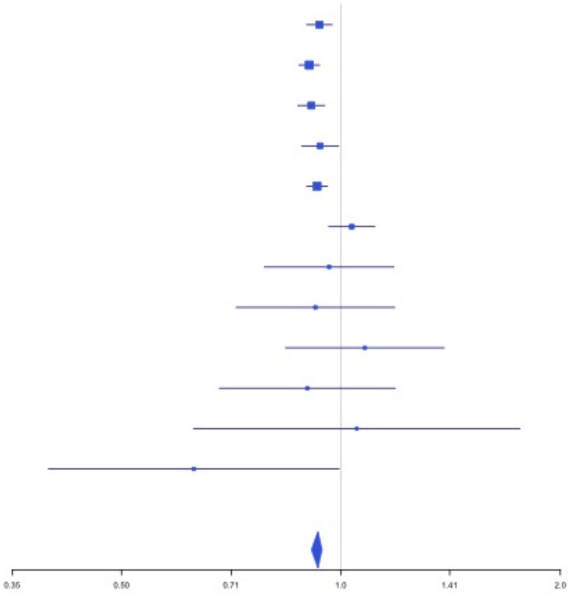


**Supplementary Figure 3.94** – rs6087685 (chr20:33777612) in locus 127 (MMP24-AS1-EDEM2)

| Study | Cases | Controls | OR |
|---|---|---|---|
| INVENT-2019_EA | 15572 | 115280 | 1.1 |
| MVP_EA | 18952 | 411746 | 1.15 |
| FinnGen_EA | 9176 | 209616 | 1.12 |
| EGP_EA | 4122 | 181553 | 1.09 |
| UKB_EA | 18385 | 91834 | 1.11 |
| MGB_EA | 2967 | 27957 | 1.06 |
| UPenn_EA | 380 | 6210 | 0.87 |
| FARIVE_EA | 545 | 532 | 1.24 |
| BioME_EA | 200 | 8007 | 1 |
| RETROVE_EA | 400 | 400 | 1.14 |
| GAIT2_EA | 85 | 850 | 1.29 |
| MESA_EA | 94 | 2341 | 1.23 |
| MARTHA12_EA | 893 | 3414 | 1.28 |
| **European** | **71771** | **1059740** | **1.12** |

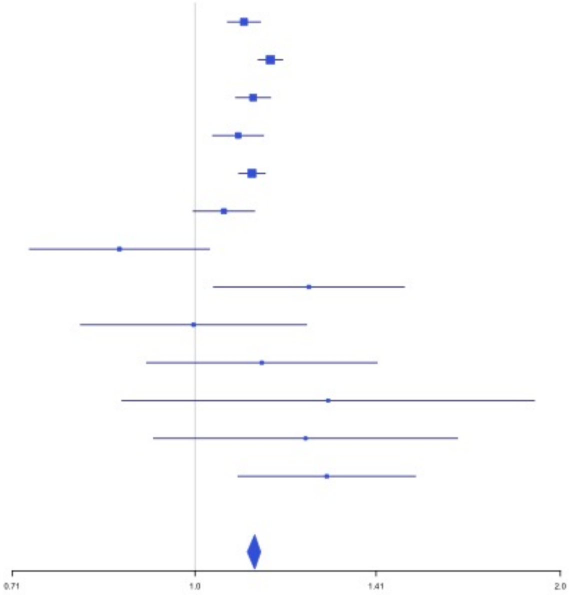

**Supplementary Figure 3.95** – rs3070580 (chr22:33165231) in locus 129 (SYN3)

| Study | Cases | Controls | OR |
|---|---|---|---|
| INVENT-2019_EA | 15572 | 115280 | 1.04 |
| MVP_EA | 18952 | 411746 | 1.05 |
| FinnGen_EA | 9176 | 209616 | 1.04 |
| EGP_EA | 4122 | 181553 | 1.08 |
| UKB_EA | 18385 | 91834 | 1.02 |
| UPenn_EA | 380 | 6210 | 1.21 |
| FARIVE_EA | 545 | 532 | 1.15 |
| **European** | **67132** | **1016771** | **1.04** |

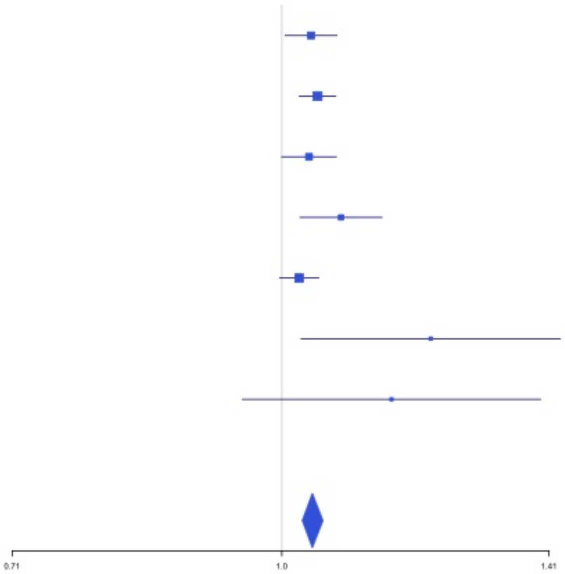


**Supplementary Figure 3.96** – rs9611844 (chr22:43115776) in locus 130 (A4GALT)

| Study | Cases | Controls | OR |
|---|---|---|---|
| INVENT-2019_EA | 15572 | 115280 | 1.04 |
| MVP_EA | 18952 | 411746 | 1.11 |
| FinnGen_EA | 9176 | 209616 | 1.08 |
| EGP_EA | 4122 | 181553 | 1.17 |
| UKB_EA | 18385 | 91834 | 1.06 |
| MGB_EA | 2967 | 27957 | 1.03 |
| UPenn_EA | 380 | 6210 | 0.89 |
| FARIVE_EA | 545 | 532 | 1.15 |
| BioME_EA | 200 | 8007 | 1.07 |
| RETROVE_EA | 400 | 400 | 1.15 |
| GAIT2_EA | 85 | 850 | 1.89 |
| MESA_EA | 94 | 2341 | 0.89 |
| **European** | **70878** | **1056326** | **1.08** |

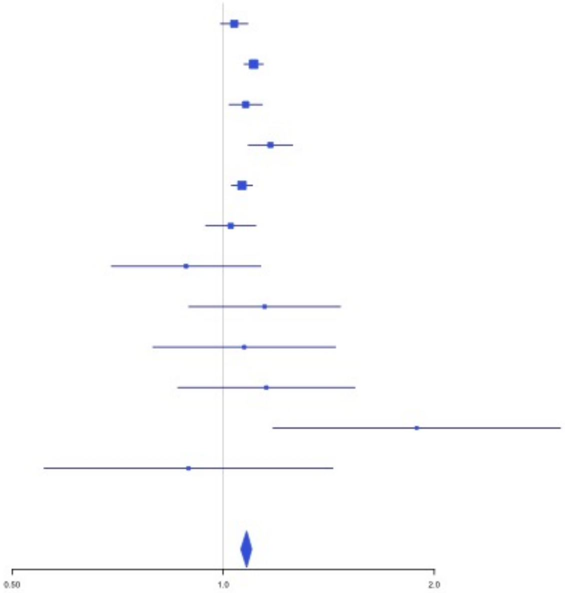

**Supplementary Figure 3.97** – rs12388118 (chr23:11380742) in locus 131 (ARHGAP6)

| Study | Cases | Controls | OR |
|---|---|---|---|
| INVENT-2019_EA | 8705 | 52578 | 1.07 |
| MVP_EA | 18952 | 411746 | 1.06 |
| EGP_EA | 4122 | 181553 | 1 |
| UKBB_EA | 280 | 1046 | 1.05 |
| FARIVE_EA | 545 | 532 | 1.23 |
| MGB_EA | 2967 | 27957 | 1.05 |
| **European** | **35571** | **675412** | **1.05** |

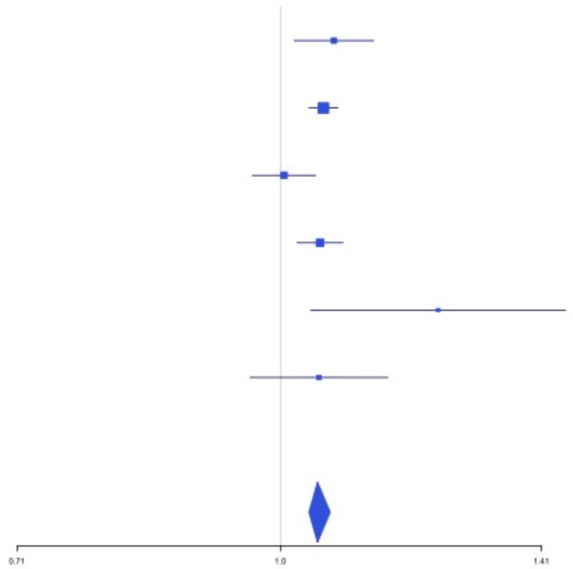


**Supplementary Figure 3.98** – rs3002416 (chr23:39710195) in locus 133 (MIR1587;BCOR)

| Study | Cases | Controls | OR |
|---|---|---|---|
| INVENT-2019_EA | 8705 | 52578 | 0.91 |
| MVP_EA | 18952 | 411746 | 0.94 |
| FinnGen_EA | 9176 | 209616 | 0.96 |
| EGP_EA | 4122 | 181553 | 0.98 |
| UKBB_EA | 280 | 1046 | 0.93 |
| FARIVE_EA | 545 | 532 | 0.77 |
| RETROVE_EA | 400 | 400 | 0.87 |
| GAIT2_EA | 85 | 850 | 1.28 |
| MGB_EA | 2967 | 27957 | 0.98 |
| **European** | **45232** | **886278** | **0.94** |

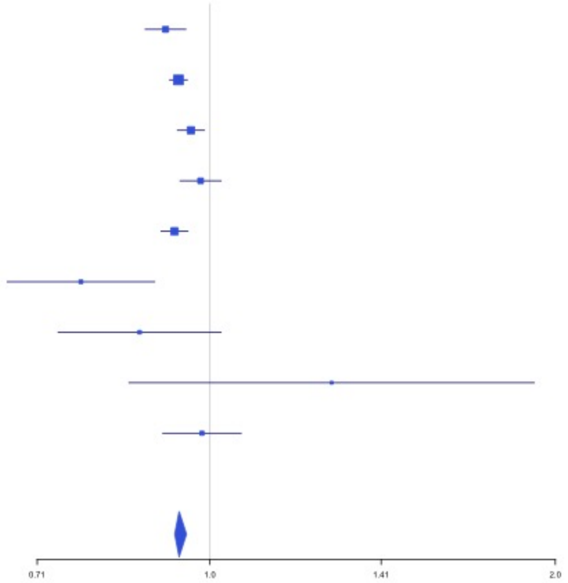

**Supplementary Figure 3.99** – rs6048 (chr23:138633280) in locus 134 (F9)

| Study | Cases | Controls | OR |
|---|---|---|---|
| INVENT-2019_EA | 8705 | 52578 | 1.09 |
| MVP_EA | 18952 | 411746 | 1.07 |
| FinnGen_EA | 9176 | 209616 | 1.05 |
| EGP_EA | 4122 | 181553 | 1.07 |
| UKBB_EA | 280 | 1046 | 1.07 |
| FARIVE_EA | 545 | 532 | 1.07 |
| RETROVE_EA | 400 | 400 | 1.22 |
| GAIT2_EA | 85 | 850 | 1.29 |
| MGB_EA | 2967 | 27957 | 1.02 |
| **European** | **45232** | **886278** | **1.07** |

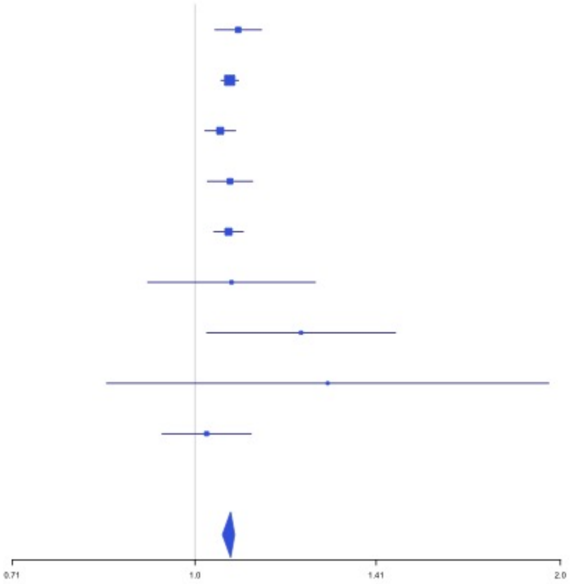


**Supplementary Figure 3.100** – rs4898406 (chr23:154273269) in locus 135 (FUNDC2)

| Study | Cases | Controls | OR |
|---|---|---|---|
| INVENT-2019_EA | 8705 | 52578 | 1.08 |
| MVP_EA | 18952 | 411746 | 1.08 |
| FinnGen_EA | 9176 | 209616 | 1.03 |
| EGP_EA | 4122 | 181553 | 1.06 |
| UKBB_EA | 280 | 1046 | 1.07 |
| FARIVE_EA | 545 | 532 | 1.09 |
| RETROVE_EA | 400 | 400 | 1.14 |
| GAIT2_EA | 85 | 850 | 1.41 |
| MGB_EA | 2967 | 27957 | 1.04 |
| **European** | **45232** | **886278** | **1.07** |

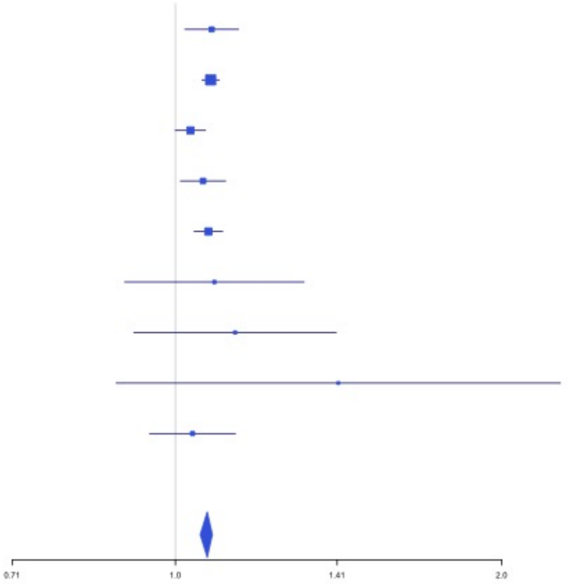
