## Supplemental Figure 4 for "Cross-Ancestry Investigation of Venous Thromboembolism Genomic Predictors"

### Supplementary Figure 4.
### Forest plots of African analysis lead variants.


4.1 rs10800427 (chr1:169272451) in locus 9 (NME7) . . . . . . . . . . . . . . . . . . . . . . . . 2
4.2 rs182293241 (chr2:128029746) in locus 18 (ERCC3) . . . . . . . . . . . . . . . . . . . . . . 2
4.3 rs529326506 (chr3:90503764) in locus 26 (EPHA3;NONE) . . . . . . . . . . . . . . . . . . 3
4.4 rs80133433 (chr3:93791551) in locus 27 (NSUN3) . . . . . . . . . . . . . . . . . . . . . . . 3
4.5 rs2066864 (chr4:155525695) in locus 39 (FGG) . . . . . . . . . . . . . . . . . . . . . . . . 4
4.6 rs56810541 (chr4:187200550) in locus 40 (F11) . . . . . . . . . . . . . . . . . . . . . . . . 4
4.7 rs8176719 (chr9:136132908) in locus 66 (ABO) . . . . . . . . . . . . . . . . . . . . . . . . 5
4.8 rs57950734 (chr12:6145649) in locus 87 (VWF) . . . . . . . . . . . . . . . . . . . . . . . . 5
4.9 rs76668186 (chr16:6686083) in locus 104 (RBFOX1) . . . . . . . . . . . . . . . . . . . . . 6
4.10 rs13306849 (chr20:23030199) in locus 126 (THBD) . . . . . . . . . . . . . . . . . . . . . . 6
4.11 rs1376223093 (chr20:33756333) in locus 127 (MMP24-AS1-EDEM2) . . . . . . . . . . . 7
4.12 rs114102448 (chr21:47523605) in locus 128 (COL6A2) . . . . . . . . . . . . . . . . . . . 7
4.13 rs73174952 (chr22:43111733) in locus 130 (A4GALT) . . . . . . . . . . . . . . . . . . . . 8

**Supplementary Figure 4.1** – rs10800427 (chr1:169272451) in locus 9 (NME7)

| Study | Cases | Controls | OR |
|---|---|---|---|
| MVP_AA | 5521 | 103791 | 1.62 |
| MGB_AA | 225 | 1481 | 1.31 |
| UPenn_AA | 203 | 1719 | 2.88 |
| BioME_AA | 382 | 6074 | 2.54 |
| MESA_AA | 72 | 1511 | 2.7 |
| **African** | **6403** | **114576** | **1.7** |

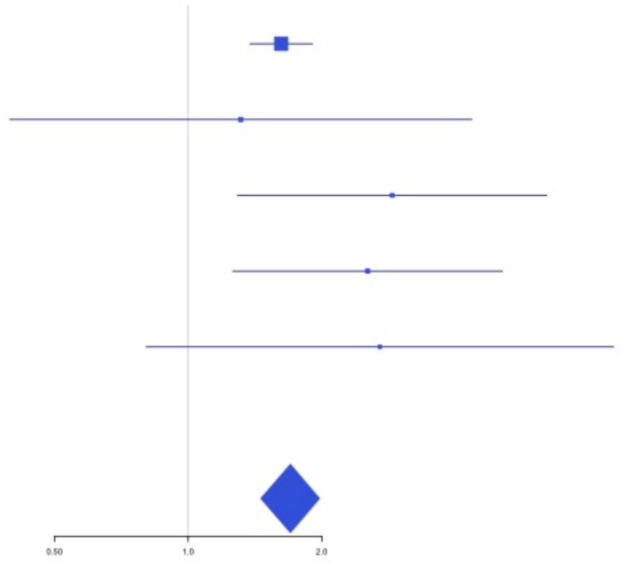


**Supplementary Figure 4.2** – rs182293241 (chr2:128029746) in locus 18 (ERCC3)

| Study | Cases | Controls | OR |
|---|---|---|---|
| MVP_AA | 5521 | 103791 | 1.89 |
| MGB_AA | 225 | 1481 | 1.98 |
| UPenn_AA | 203 | 1719 | 1.26 |
| BioME_AA | 382 | 6074 | 1.89 |
| MESA_AA | 72 | 1511 | 1.32 |
| **African** | **6403** | **114576** | **1.86** |

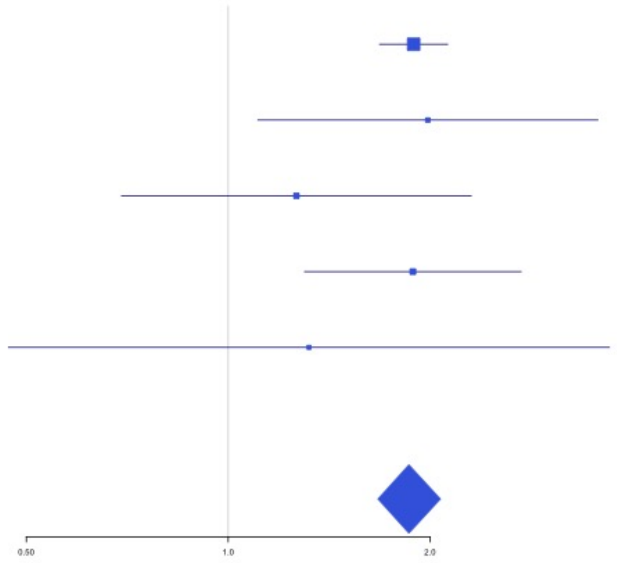

**Supplementary Figure 4.3** – rs529326506 (chr3:90503764) in locus 26 (EPHA3 ;NONE)

| Study | Cases | Controls | OR |
|---|---|---|---|
| MVP_AA | 5521 | 103791 | 1.5 |
| MGB_AA | 225 | 1481 | 1.15 |
| UPenn_AA | 203 | 1719 | 0.77 |
| BioME_AA | 382 | 6074 | 1.53 |
| MESA_AA | 72 | 1511 | 1.64 |
| **African** | **6403** | **114576** | **1.47** |

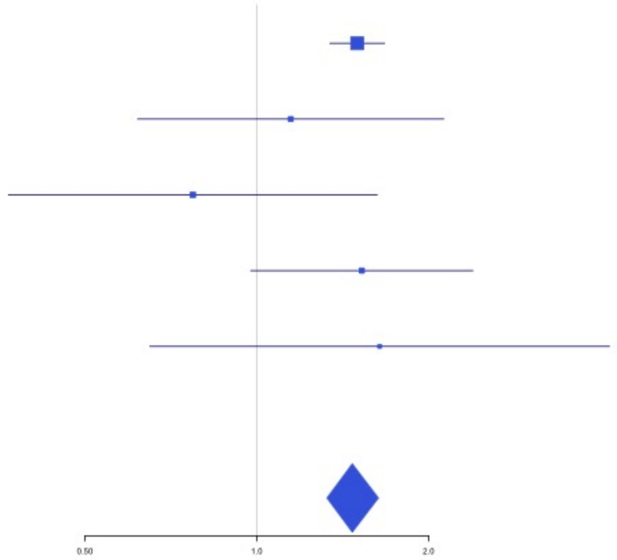


**Supplementary Figure 4.4** – rs80133433 (chr3:93791551) in locus 27 (NSUN3)

| Study | Cases | Controls | OR |
|---|---|---|---|
| MVP_AA | 5521 | 103791 | 1.59 |
| MGB_AA | 225 | 1481 | 1.22 |
| UPenn_AA | 203 | 1719 | 0.8 |
| BioME_AA | 382 | 6074 | 1.33 |
| MESA_AA | 72 | 1511 | 1.58 |
| **African** | **6403** | **114576** | **1.55** |

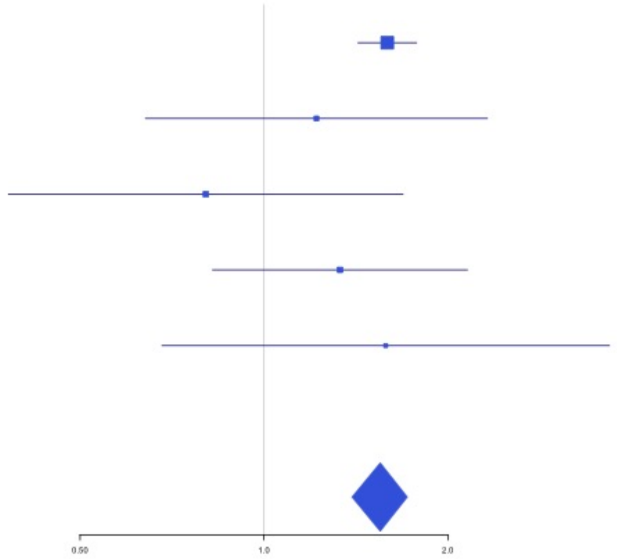

**Supplementary Figure 4.5** – rs2066864 (chr4:155525695) in locus 39 (FGG)

| Study | Cases | Controls | OR |
|---|---|---|---|
| INVENT-2019_AA | 799 | 14353 | 1.17 |
| MVP_AA | 5521 | 103791 | 1.19 |
| UKB_AA | 280 | 1046 | 1.14 |
| MGB_AA | 225 | 1481 | 1.23 |
| UPenn_AA | 203 | 1719 | 1.38 |
| BioME_AA | 382 | 6074 | 1.16 |
| MESA_AA | 72 | 1511 | 1.2 |
| **African** | **7482** | **129975** | **1.19** |

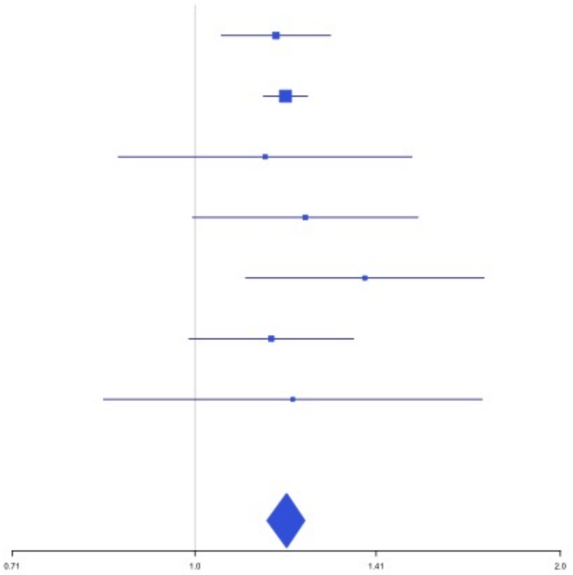


**Supplementary Figure 4.6** – rs56810541 (chr4:187200550) in locus 40 (F11)

| Study | Cases | Controls | OR |
|---|---|---|---|
| INVENT-2019_AA | 799 | 14353 | 0.82 |
| MVP_AA | 5521 | 103791 | 0.84 |
| UKB_AA | 280 | 1046 | 0.87 |
| MGB_AA | 225 | 1481 | 0.76 |
| UPenn_AA | 203 | 1719 | 0.95 |
| BioME_AA | 382 | 6074 | 0.89 |
| MESA_AA | 72 | 1511 | 0.88 |
| **African** | **7482** | **129975** | **0.84** |

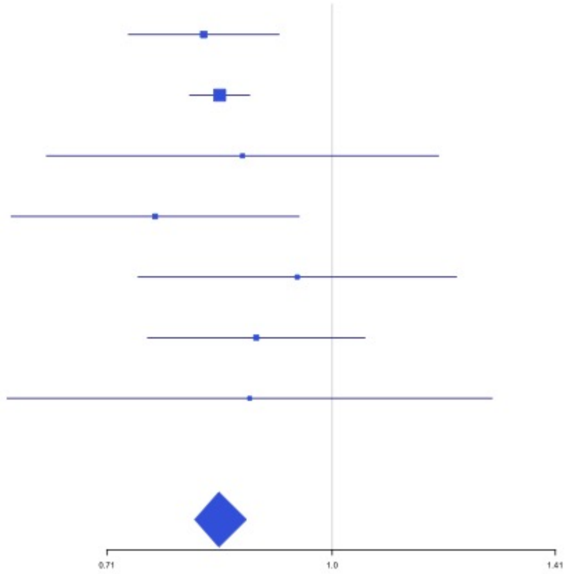

**Supplementary Figure 4.7** – rs8176719 (chr9:136132908) in locus 66 (ABO)

| Study | Cases | Controls | OR |
|---|---|---|---|
| INVENT-2019_AA | 799 | 14353 | 0.77 |
| MVP_AA | 5521 | 103791 | 0.7 |
| UKB_AA | 280 | 1046 | 1.04 |
| MGB_AA | 225 | 1481 | 0.82 |
| UPenn_AA | 203 | 1719 | 0.96 |
| BioME_AA | 382 | 6074 | 0.76 |
| **African** | **7410** | **128464** | **0.73** |

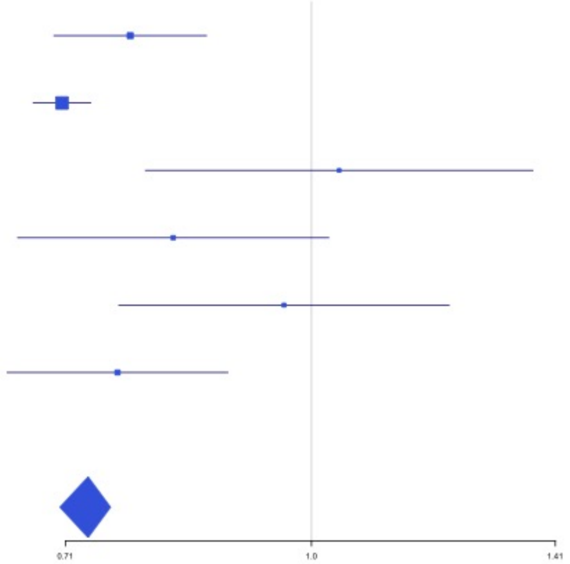


**Supplementary Figure 4.8** – rs57950734 (chr12:6145649) in locus 87 (VWF)

| Study | Cases | Controls | OR |
|---|---|---|---|
| INVENT-2019_AA | 799 | 14353 | 1.1 |
| MVP_AA | 5521 | 103791 | 1.29 |
| MGB_AA | 225 | 1481 | 1.3 |
| UPenn_AA | 203 | 1719 | 1.13 |
| BioME_AA | 382 | 6074 | 1.32 |
| MESA_AA | 72 | 1511 | 1.19 |
| **African** | **7202** | **128929** | **1.26** |

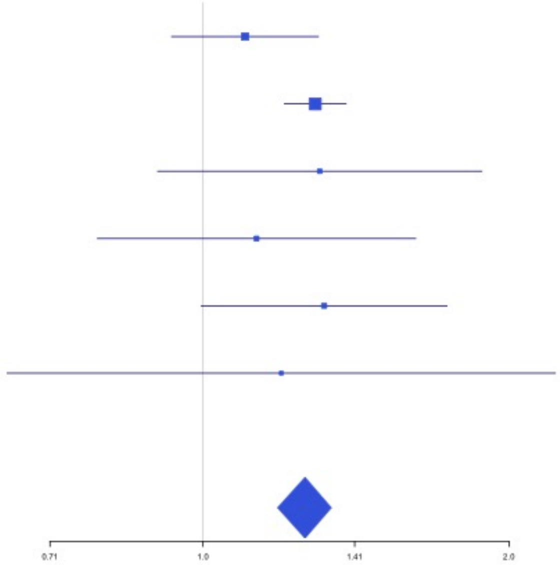

**Supplementary Figure 4.9** – rs76668186 (chr16:6686083) in locus 104 (RBFOX1)

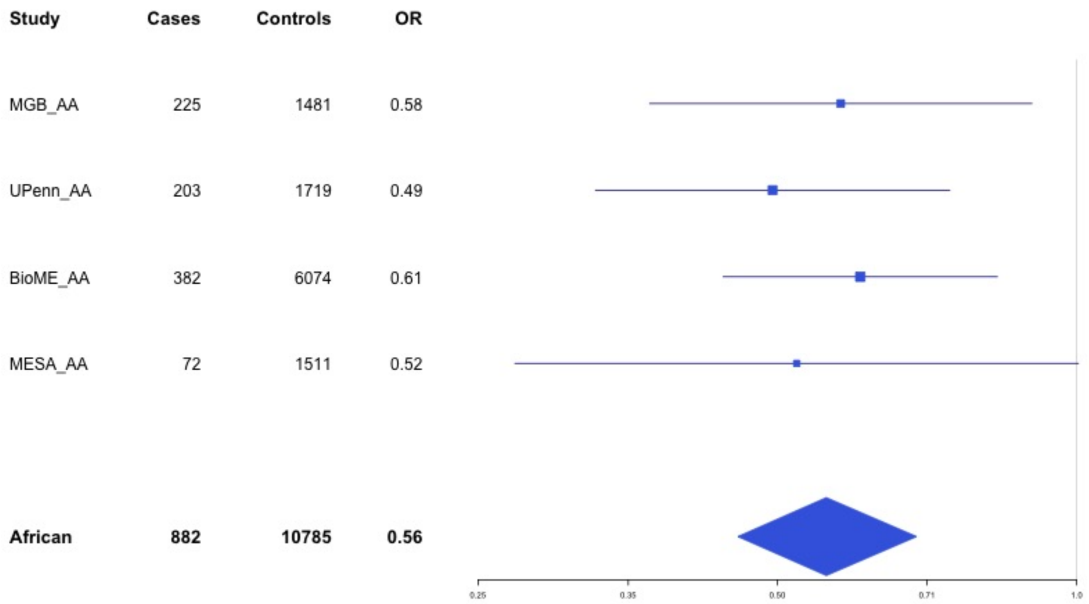


**Supplementary Figure 4.10** – rs13306849 (chr20:23030199) in locus 126 (THBD)

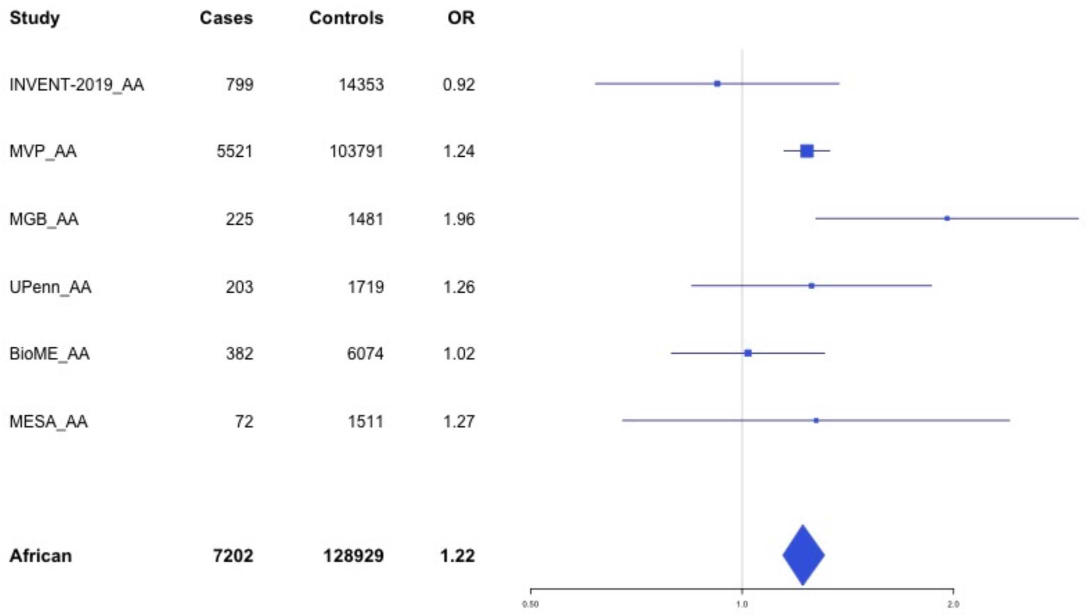

**Supplementary Figure 4.11** – rs1376223093 (chr20:33756333) in locus 127 (MMP24-AS1-EDEM2)

| Study | Cases | Controls | OR |
|---|---|---|---|
| MVP_AA | 5521 | 103791 | 0.88 |
| UKB_AA | 280 | 1046 | 0.8 |
| UPenn_AA | 203 | 1719 | 1.04 |
| **African** | **6004** | **106556** | **0.88** |

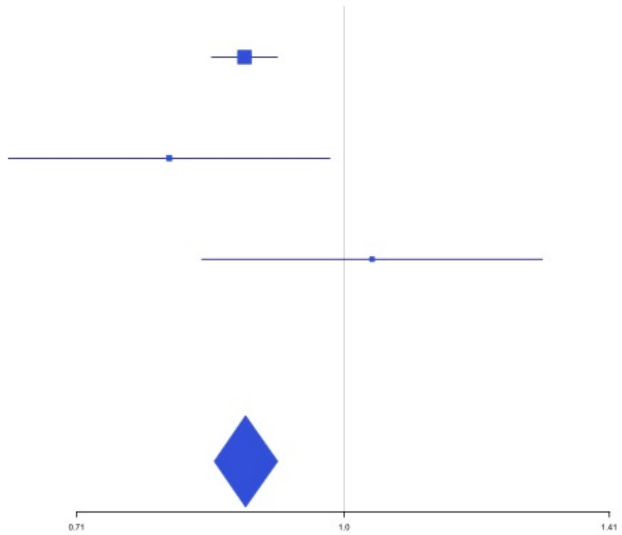


**Supplementary Figure 4.12** – rs114102448 (chr21:47523605) in locus 128 (COL6A2)

| Study | Cases | Controls | OR |
|---|---|---|---|
| MGB_AA | 225 | 1481 | 1.7 |
| UPenn_AA | 203 | 1719 | 1.6 |
| BioME_AA | 382 | 6074 | 3.41 |
| MESA_AA | 72 | 1511 | 2.98 |
| **African** | **882** | **10785** | **2.59** |

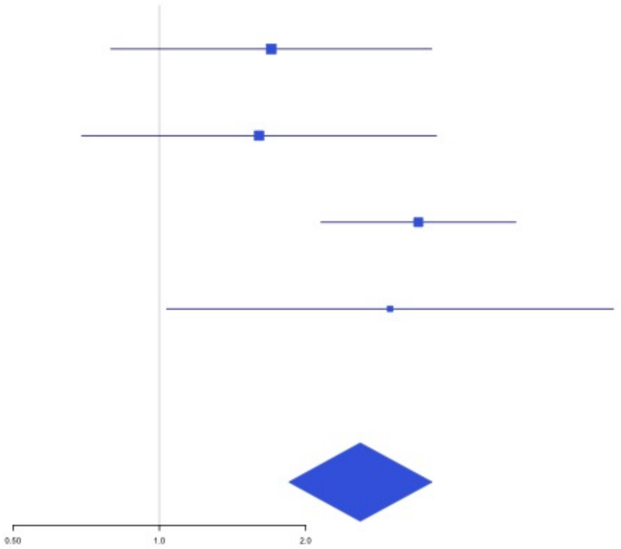

**Supplementary Figure 4.13** – rs73174952 (chr22:43111733) in locus 130 (A4GALT)

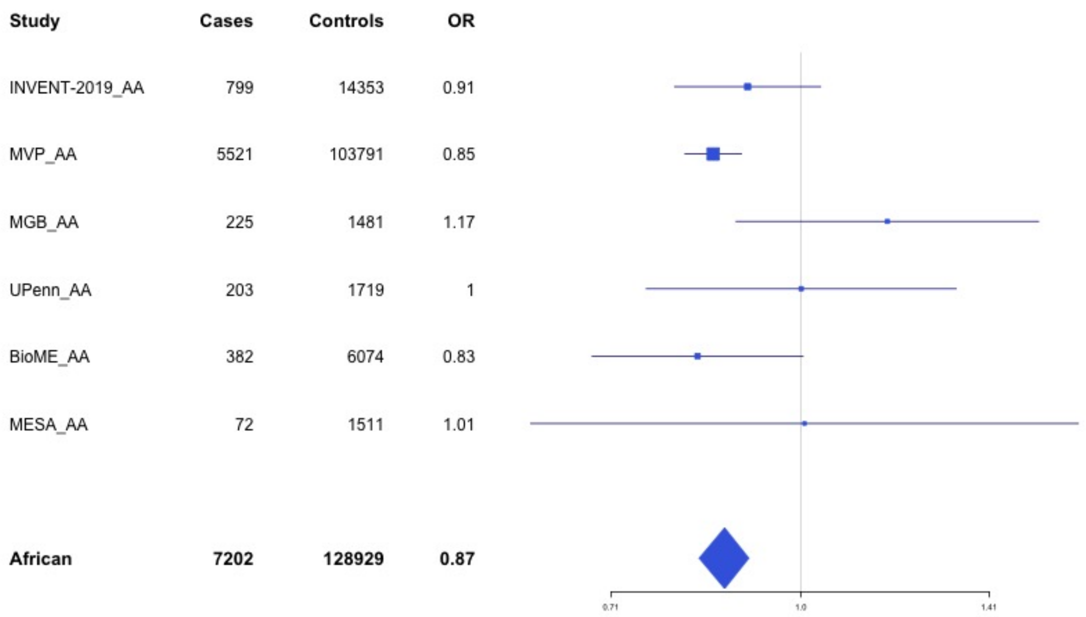
