## Supplemental Figure 5 for "Cross-Ancestry Investigation of Venous Thromboembolism Genomic Predictors"

### Supplementary Figure 5.
### Regional association plots of Discovery analysis lead variants.


5.1 rs9442580 (chr1:9339467) in locus 1 (H6PD ;SPSB1) . . . 3
5.2 rs3767812 (chr1:118155620) in locus 5 (TENT5C) . . . 4
5.3 rs6025 (chr1:169519049) in locus 9 (F5) . . . 5
5.4 rs2842700 (chr1:207282149) in locus 13 (C4BPA) . . . 6
5.5 rs3811444 (chr1:248039451) in locus 14 (TRIM58) . . . 7
5.6 rs545891600 (chr2:5378301) in locus 15 (LINC01249 ;LINC01248) . . . 8
5.7 rs7600986 (chr2:68636923) in locus 17 (PLEK ;FBXO48) . . . 9
5.8 rs182293241 (chr2:128029746) in locus 18 (ERCC3) . . . 10
5.9 rs6719550 (chr2:188272460) in locus 19 (CALCRL) . . . 11
5.10 rs715 (chr2:211543055) in locus 21 (CPS1) . . . 12
5.11 rs13412535 (chr2:224874874) in locus 22 (SERPINE2) . . . 13
5.12 rs2960420 (chr3:12314512) in locus 23 (SYN2 ;PPARG) . . . 14
5.13 rs13084580 (chr3:39188182) in locus 24 (CSRNP1) . . . 15
5.14 rs11130326 (chr3:52771920) in locus 25 (NEK4) . . . 16
5.15 rs562281690 (chr3:90177913) in locus 26 (EPHA3 ;NONE) . . . 17
5.16 rs79324379 (chr3:93712671) in locus 27 (ARL13B) . . . 18
5.17 rs9872572 (chr3:123108613) in locus 28 (ADCY5) . . . 19
5.18 rs62282204 (chr3:138584405) in locus 29 (PIK3CB ;LINC01391) . . . 20
5.19 rs7613621 (chr3:169191186) in locus 31 (MECOM) . . . 21
5.20 rs710446 (chr3:186459927) in locus 32 (KNG1) . . . 22
5.21 rs6797948 (chr3:194784705) in locus 33 (LINC01968 ;XXYLT1) . . . 23
5.22 rs6826579 (chr4:83785031) in locus 36 (SEC31A) . . . 24
5.23 rs17010957 (chr4:86719165) in locus 37 (ARHGAP24) . . . 25
5.24 rs2066864 (chr4:155525695) in locus 39 (FGG) . . . 26
5.25 rs3756011 (chr4:187206249) in locus 40 (F11) . . . 27
5.26 rs16867574 (chr5:38708554) in locus 42 (OSMR-AS1) . . . 28
5.27 rs38032 (chr5:96321887) in locus 44 (LNPEP) . . . 29
5.28 rs9268881 (chr6:32431606) in locus 48 (HLA-DRA ;HLA-DRB5) . . . 30
5.29 rs145294670 (chr6:34622561) in locus 49 (ILRUN) . . . 31
5.30 rs903646797 (chr6:40985346) in locus 50 (LOC101929555) . . . 32
5.31 rs9390460 (chr6:147694334) in locus 52 (STXBP5) . . . 33
5.32 rs42036 (chr7:92241451) in locus 54 (CDK6) . . . 34
5.33 rs538161377 (chr8:4206326) in locus 55 (CSMD1) . . . 35
5.34 rs67694436 (chr8:6654220) in locus 56 (AGPAT5 ;XKR5) . . . 36
5.35 rs7010926 (chr8:10626791) in locus 58 (PINX1) . . . 37
5.36 rs2685417 (chr8:27807434) in locus 60 (SCARA5) . . . 38
5.37 rs6993770 (chr8:106581528) in locus 62 (ZFPM2) . . . 39
5.38 rs77375493 (chr9:5073770) in locus 64 (JAK2) . . . 40
5.39 rs35208412 (chr9:99194509) in locus 65 (ZNF367 ;HABP4) . . . 41
5.40 rs505922 (chr9:136149229) in locus 66 (ABO) . . . 42
5.41 rs1887091 (chr10:14535113) in locus 67 (MIR1265 ;FAM107B) . . . 43
5.42 rs529892067 (chr10:43568540) in locus 71 (MIR5100 ;RET) . . . 44
5.43 rs17490626 (chr10:71218646) in locus 72 (TSPAN15) . . . 45
5.44 rs16937003 (chr10:80938499) in locus 73 (ZMIZ1) . . . 46
5.45 rs2274224 (chr10:96039597) in locus 74 (PLCE1) . . . 47
5.46 rs10886430 (chr10:121010256) in locus 75 (GRK5) . . . 48
5.47 rs11032074 (chr11:32993887) in locus 78 (QSER1) . . . 49
5.48 rs1799963 (chr11:46761055) in locus 79 (F2) . . . 50
5.49 rs141687379 (chr11:56666822) in locus 80 (FADS2B) . . . 51
5.50 rs174551 (chr11:61573684) in locus 82 (FADS1) . . . 52

5.51 rs35257264 (chr11:126296816) in locus 85 (ST3GAL4) . . . . . . . . . . . . . . . . . . . 53
5.52 rs1558519 (chr12:6153738) in locus 87 (VWF) . . . . . . . . . . . . . . . . . . . . . . . . . 54
5.53 rs7311483 (chr12:9053661) in locus 88 (A2ML1;PHC1) . . . . . . . . . . . . . . . . . . . . 55
5.54 rs6580981 (chr12:54723028) in locus 89 (COPZ1) . . . . . . . . . . . . . . . . . . . . . . . 56
5.55 rs142351376 (chr12:104136288) in locus 90 (STAB2) . . . . . . . . . . . . . . . . . . . . 57
5.56 rs3184504 (chr12:111884608) in locus 91 (SH2B3) . . . . . . . . . . . . . . . . . . . . . . 58
5.57 rs75940120 (chr13:109686619) in locus 93 (MYO16) . . . . . . . . . . . . . . . . . . . . . 59
5.58 rs3211752 (chr13:113787459) in locus 94 (F10) . . . . . . . . . . . . . . . . . . . . . . . . 60
5.59 rs57035593 (chr14:92268096) in locus 97 (TC2N) . . . . . . . . . . . . . . . . . . . . . . 61
5.60 rs8013957 (chr14:103140254) in locus 99 (RCOR1) . . . . . . . . . . . . . . . . . . . . . 62
5.61 rs55707100 (chr15:43820717) in locus 100 (MAP1A) . . . . . . . . . . . . . . . . . . . . 63
5.62 rs59442804 (chr15:60899031) in locus 101 (RORA-AS1) . . . . . . . . . . . . . . . . . . 64
5.63 rs182906510 (chr15:67372922) in locus 102 (SMAD3) . . . . . . . . . . . . . . . . . . . 65
5.64 rs12443808 (chr16:30996871) in locus 106 (HSD3B7) . . . . . . . . . . . . . . . . . . . 66
5.65 rs56943275 (chr16:81898152) in locus 109 (PLCG2) . . . . . . . . . . . . . . . . . . . . 67
5.66 rs28634651 (chr16:88553198) in locus 111 (ZFPM1) . . . . . . . . . . . . . . . . . . . . 68
5.67 rs6503222 (chr17:1977862) in locus 112 (SMG6) . . . . . . . . . . . . . . . . . . . . . . . 69
5.68 rs7225756 (chr17:6893691) in locus 113 (ALOX12-AS1) . . . . . . . . . . . . . . . . . . 70
5.69 rs62054822 (chr17:43927708) in locus 115 (MAPT-AS1) . . . . . . . . . . . . . . . . . . 71
5.70 rs142140545 (chr17:64191540) in locus 116 (CEP112;APOH) . . . . . . . . . . . . . . . 72
5.71 rs59277920 (chr19:6077231) in locus 118 (RFX2) . . . . . . . . . . . . . . . . . . . . . . 73
5.72 rs8110055 (chr19:10739143) in locus 119 (SLC44A2) . . . . . . . . . . . . . . . . . . . . 74
5.73 rs34783010 (chr19:46180414) in locus 122 (GIPR) . . . . . . . . . . . . . . . . . . . . . . 75
5.74 rs1688264 (chr19:49209560) in locus 123 (FUT2) . . . . . . . . . . . . . . . . . . . . . . 76
5.75 rs1654425 (chr19:55538980) in locus 124 (GP6) . . . . . . . . . . . . . . . . . . . . . . . 77
5.76 rs2631628 (chr19:56645250) in locus 125 (ZNF787;ZNF444) . . . . . . . . . . . . . . . . 78
5.77 rs79388863 (chr20:23168500) in locus 126 (LINC00656;NXT1) . . . . . . . . . . . . . . . 79
5.78 rs6060288 (chr20:33772243) in locus 127 (MMP24-AS1-EDEM2) . . . . . . . . . . . . . 80
5.79 rs4820093 (chr22:33160208) in locus 129 (SYN3) . . . . . . . . . . . . . . . . . . . . . . 81
5.80 rs9611844 (chr22:43115776) in locus 130 (A4GALT) . . . . . . . . . . . . . . . . . . . . 82
5.81 rs34609587 (chr23:11657363) in locus 131 (ARHGAP6) . . . . . . . . . . . . . . . . . . 83
5.82 rs6632109 (chr23:34894638) in locus 132 (TMEM47;FAM47B) . . . . . . . . . . . . . . . 84
5.83 rs3002416 (chr23:39710195) in locus 133 (MIR1587;BCOR) . . . . . . . . . . . . . . . . 85
5.84 rs6048 (chr23:138633280) in locus 134 (F9) . . . . . . . . . . . . . . . . . . . . . . . . . 86
5.85 rs2084408 (chr23:154346709) in locus 135 (BRCC3) . . . . . . . . . . . . . . . . . . . . 87

**Supplementary Figure 5.1** – rs9442580 (chr1:9339467) in locus 1 (H6PD ;SPSB1)

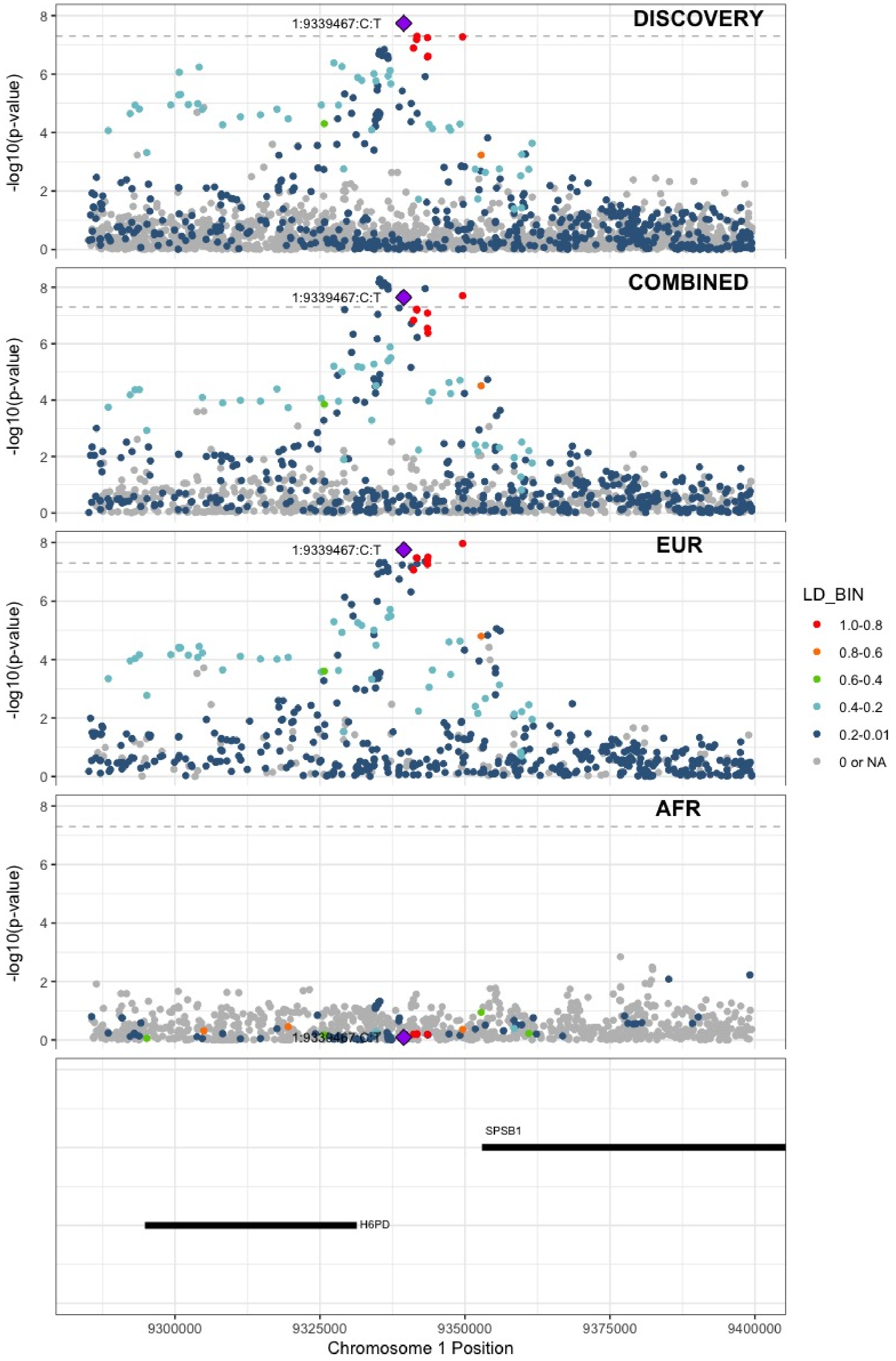

**Supplementary Figure 5.2** – rs3767812 (chr1:118155620) in locus 5 (TENT5C)

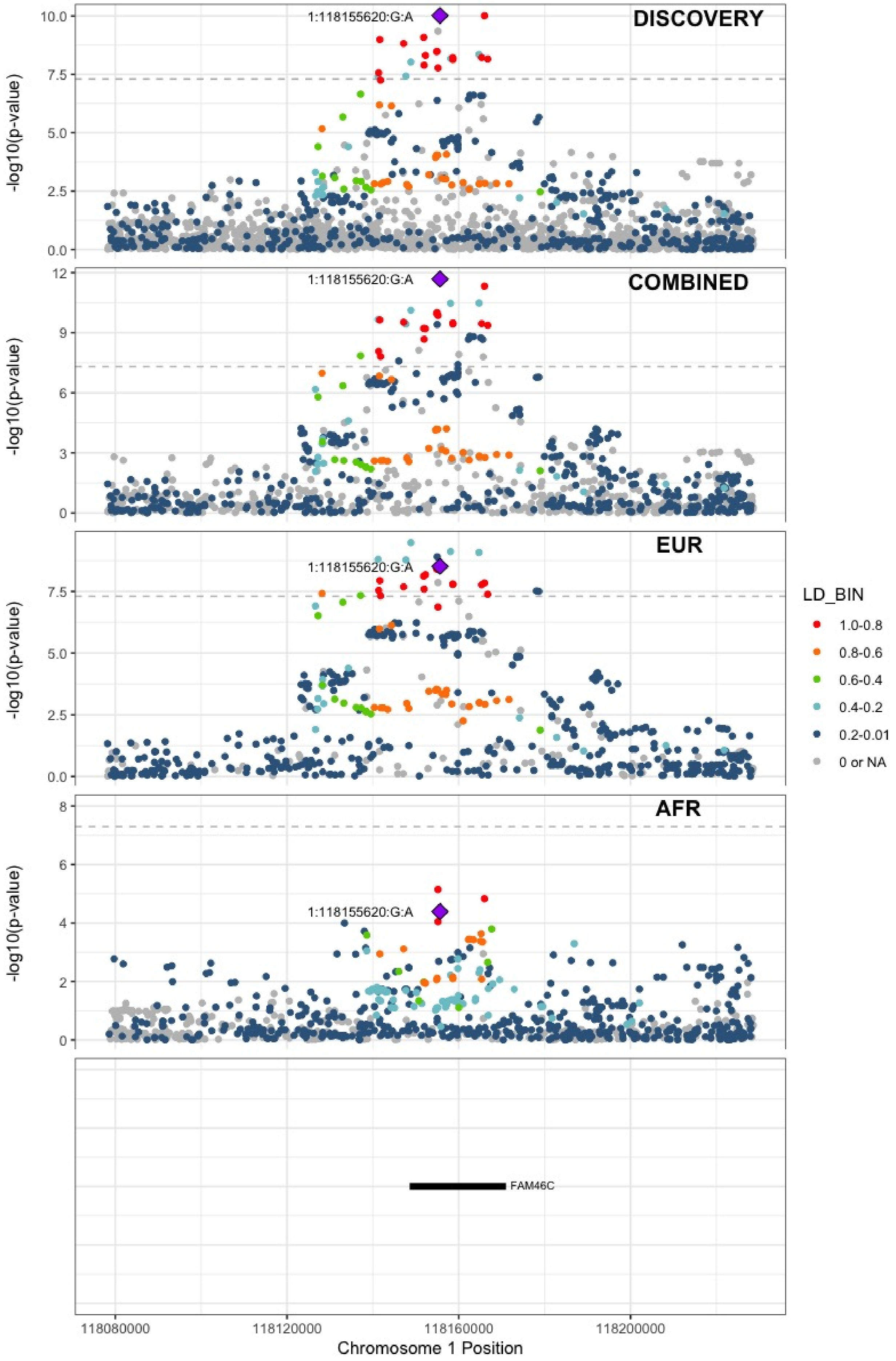

**Supplementary Figure 5.3** – rs6025 (chr1:169519049) in locus 9 (F5)

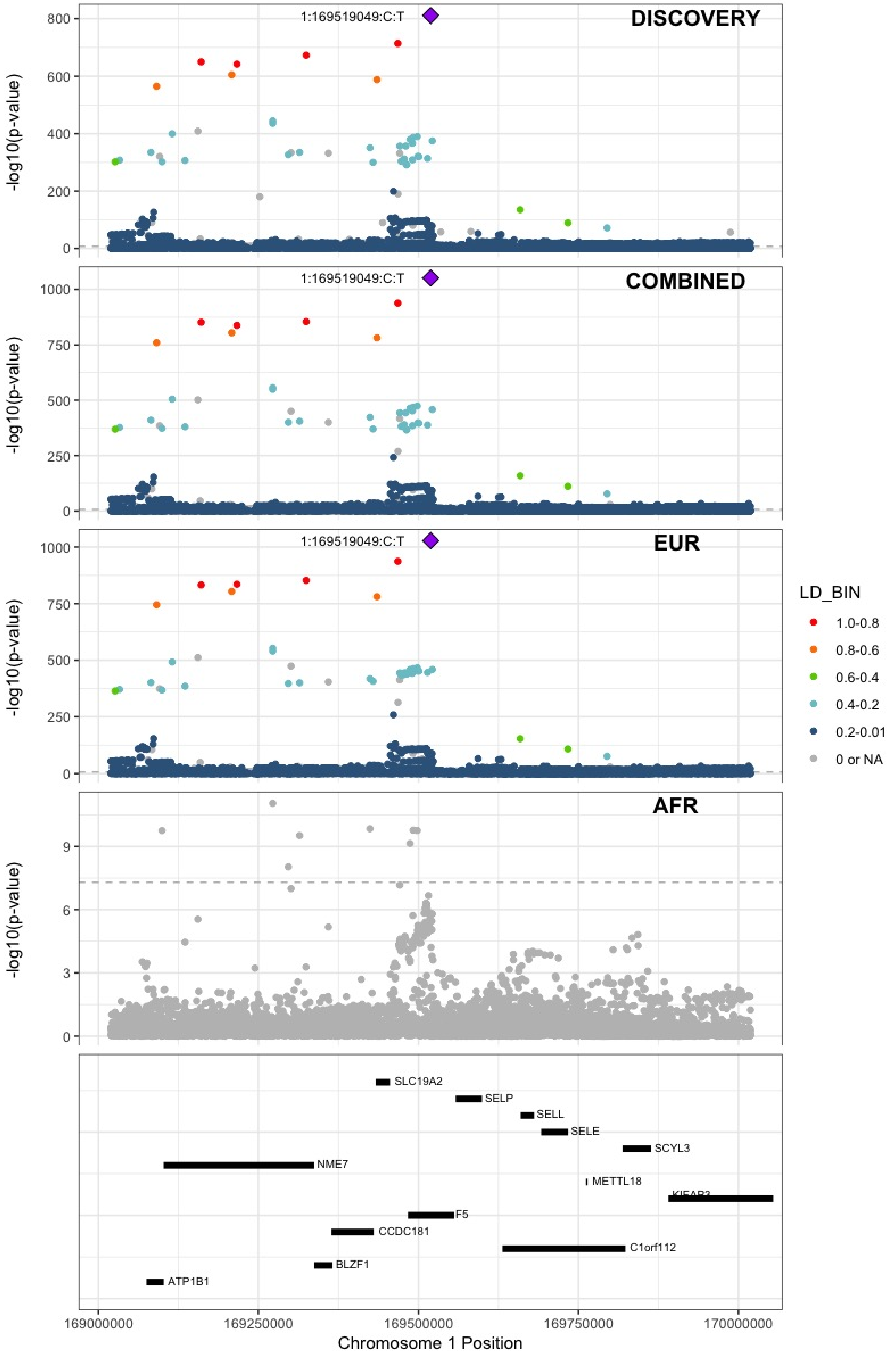

**Supplementary Figure 5.4** – rs2842700 (chr1:207282149) in locus 13 (C4BPA)

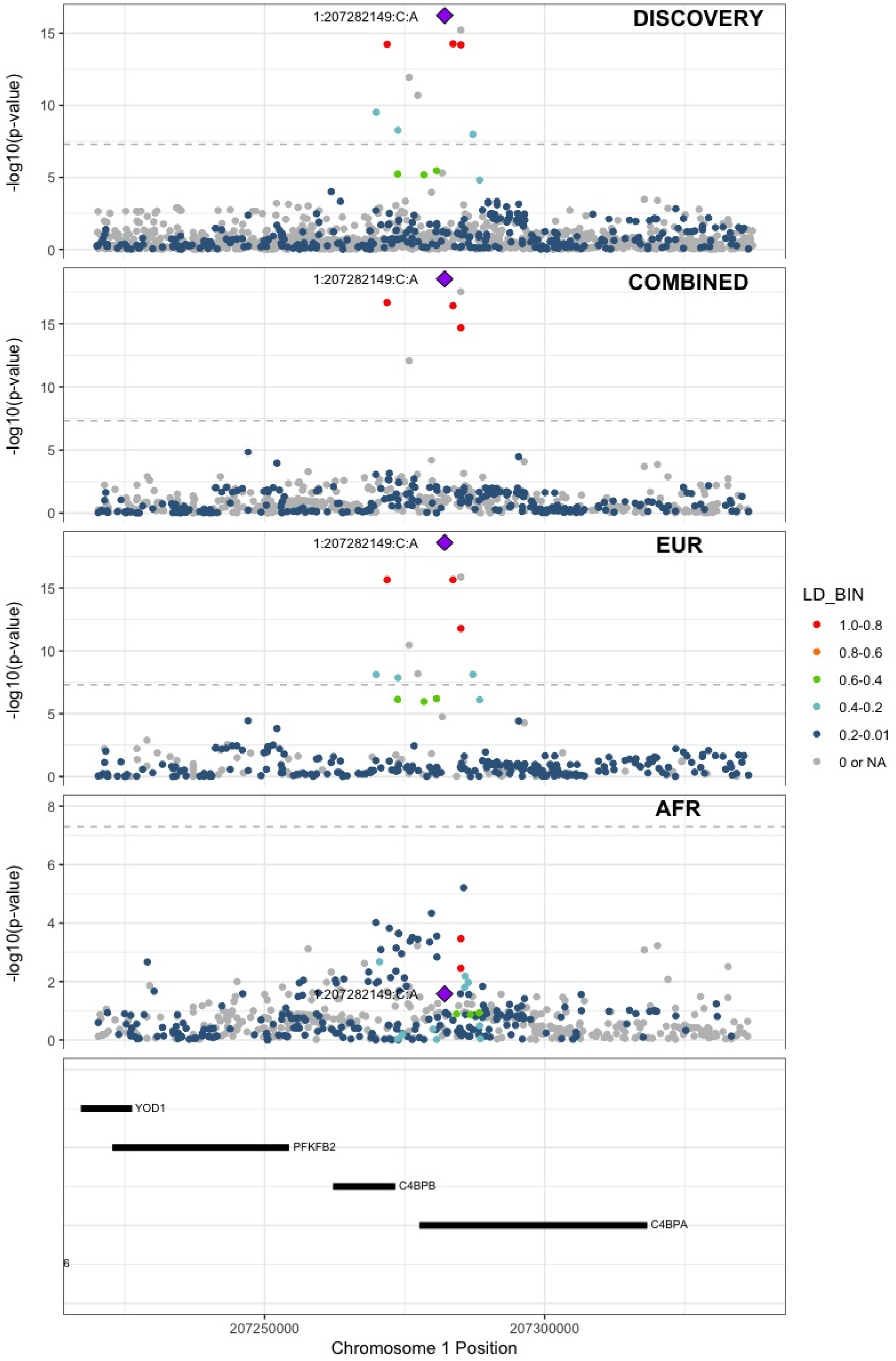

**Supplementary Figure 5.5** – rs3811444 (chr1:248039451) in locus 14 (TRIM58)

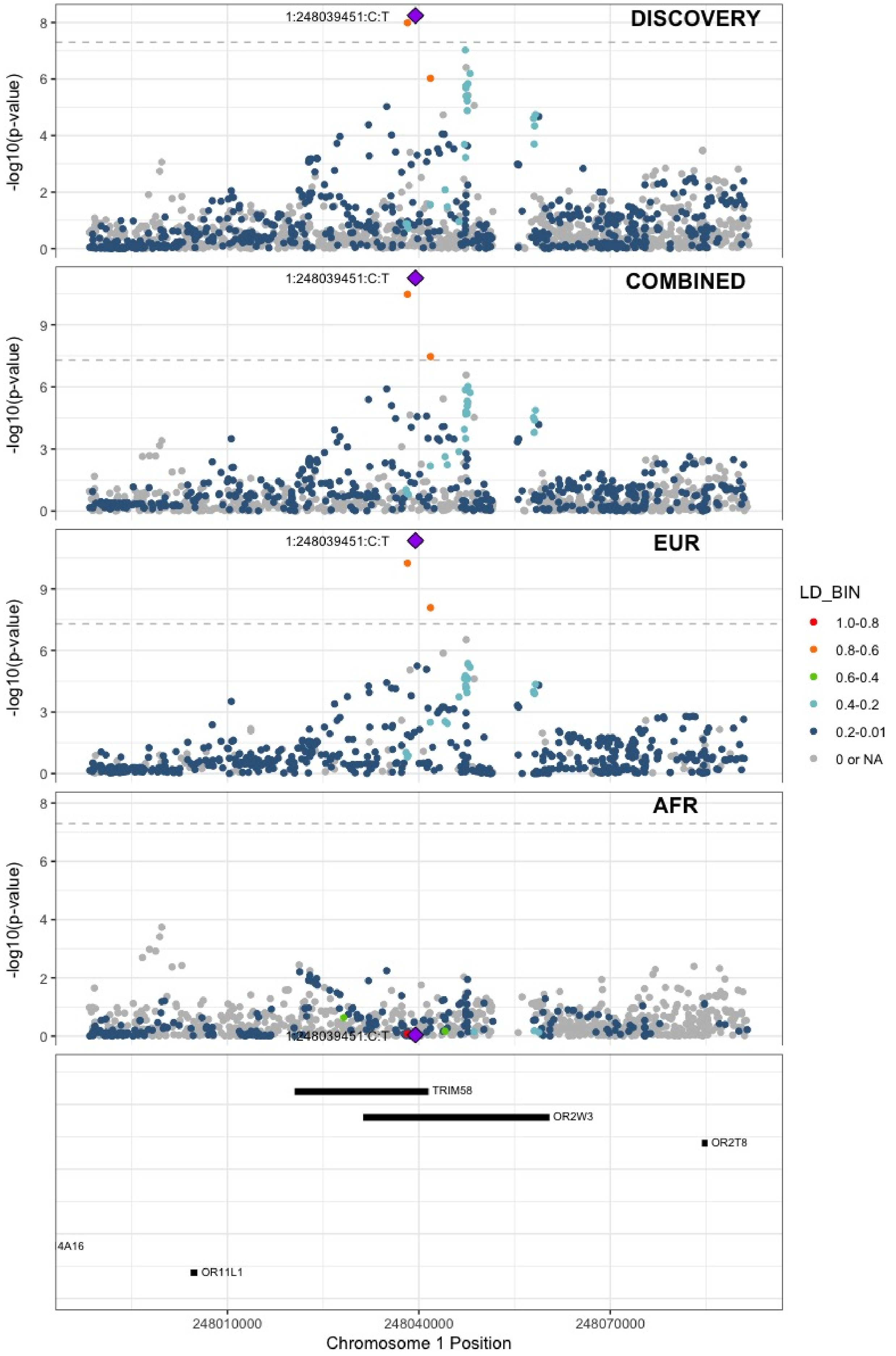

**Supplementary Figure 5.6** – rs545891600 (chr2:5378301) in locus 15 (LINC01249;LINC01248)

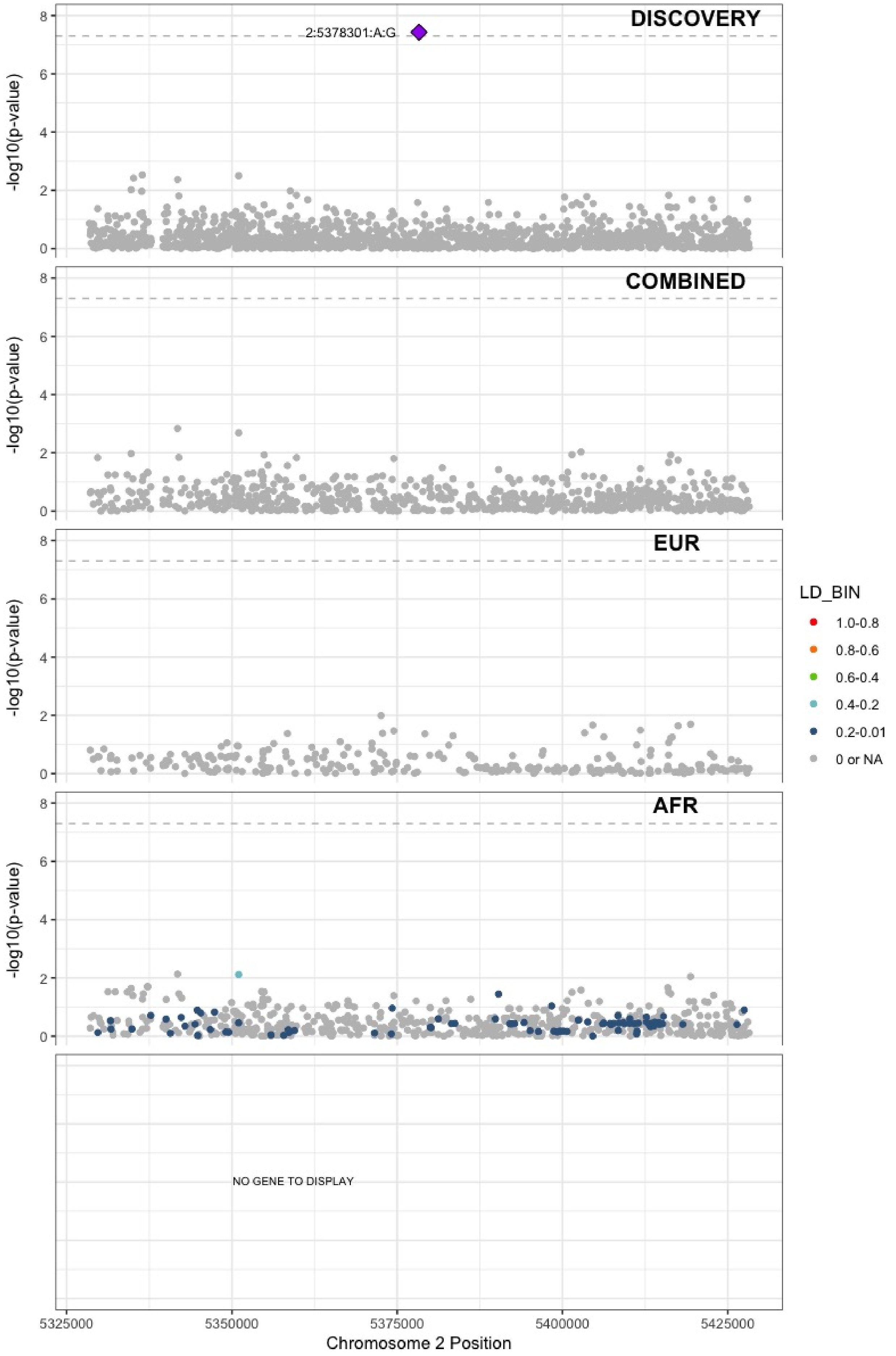

**Supplementary Figure 5.7** – rs7600986 (chr2:68636923) in locus 17 (PLEK ;FBXO48)

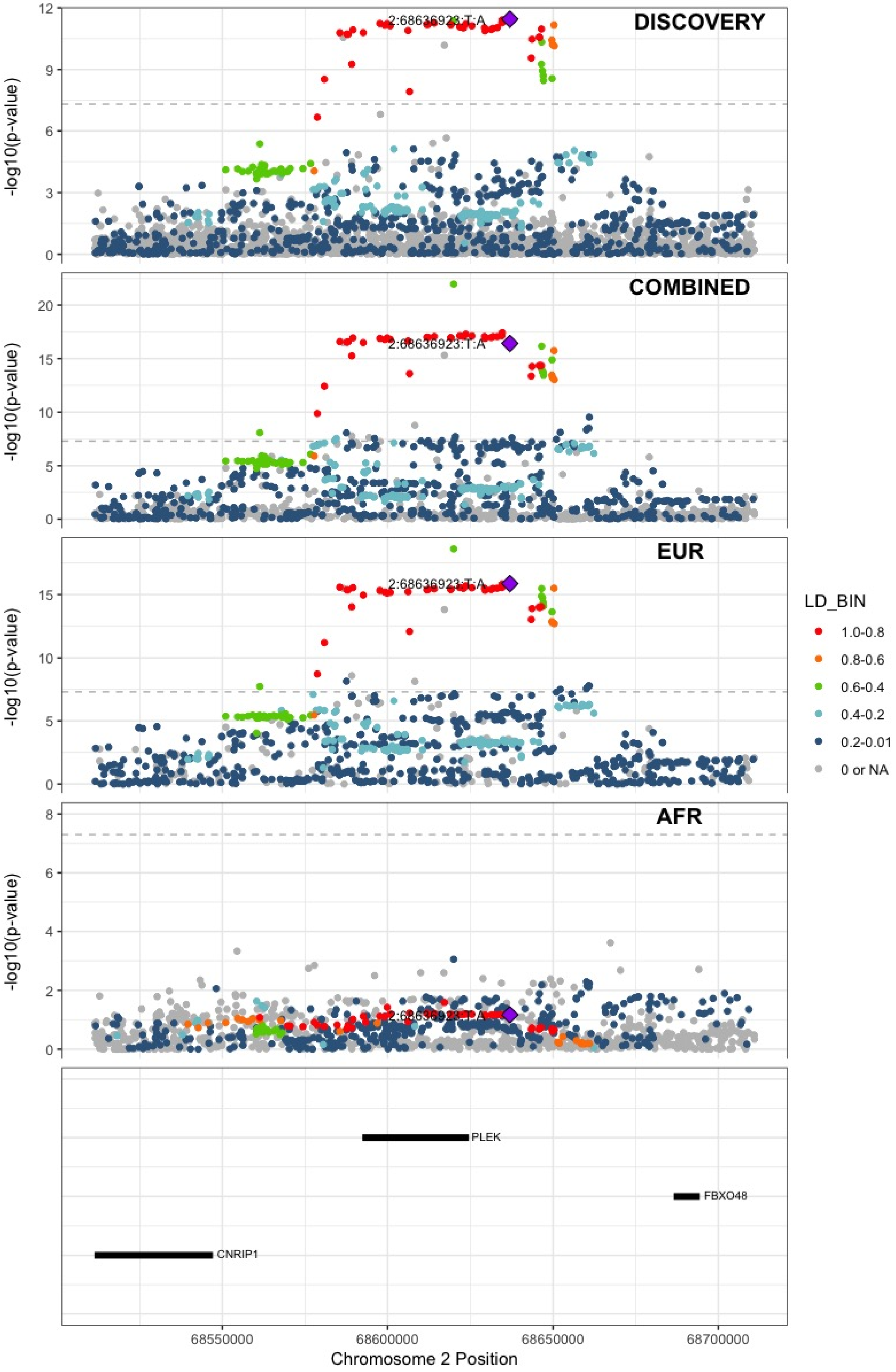

**Supplementary Figure 5.8** – rs182293241 (chr2:128029746) in locus 18 (ERCC3)

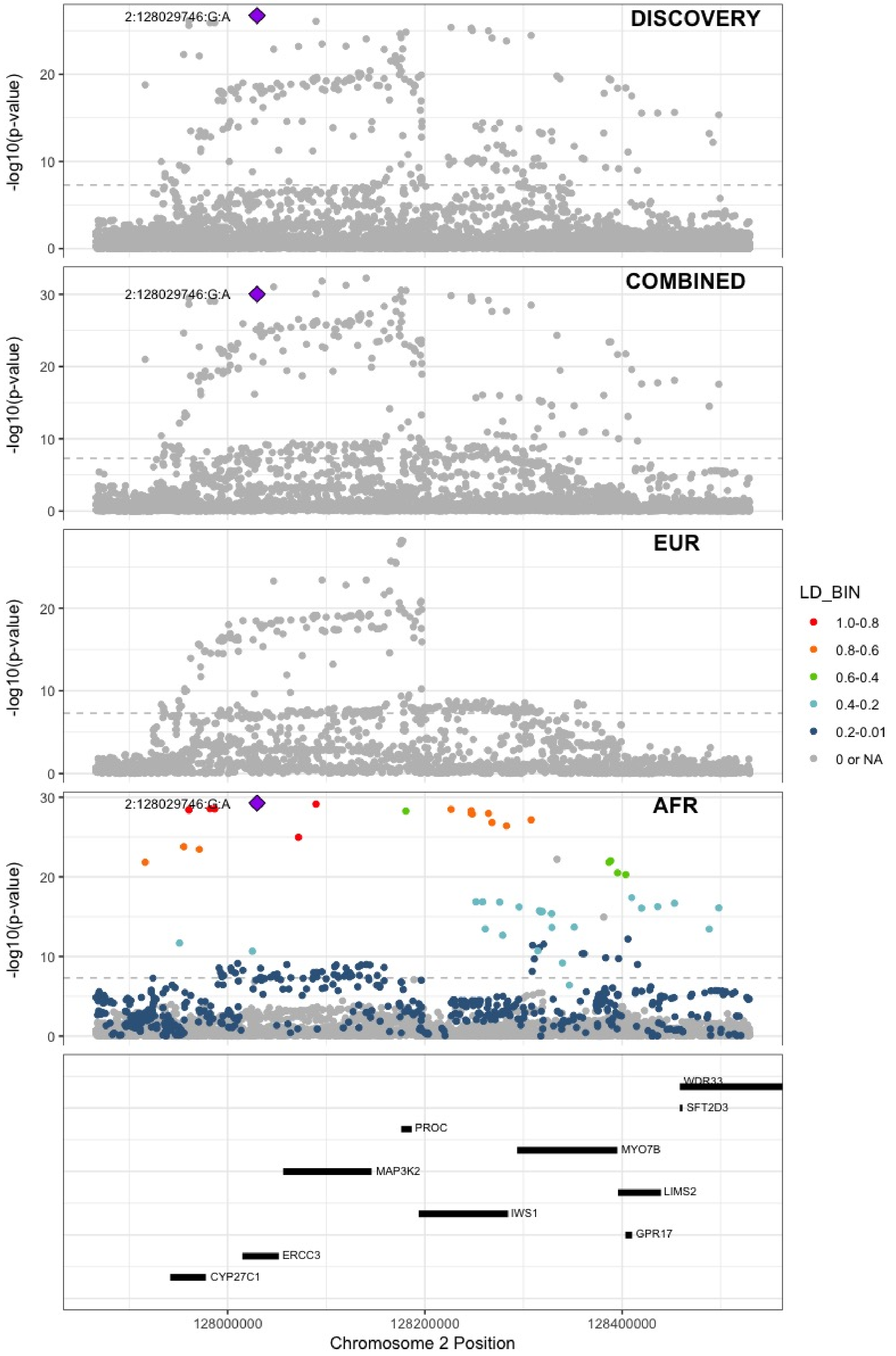

**Supplementary Figure 5.9** – rs6719550 (chr2:188272460) in locus 19 (CALCRL)

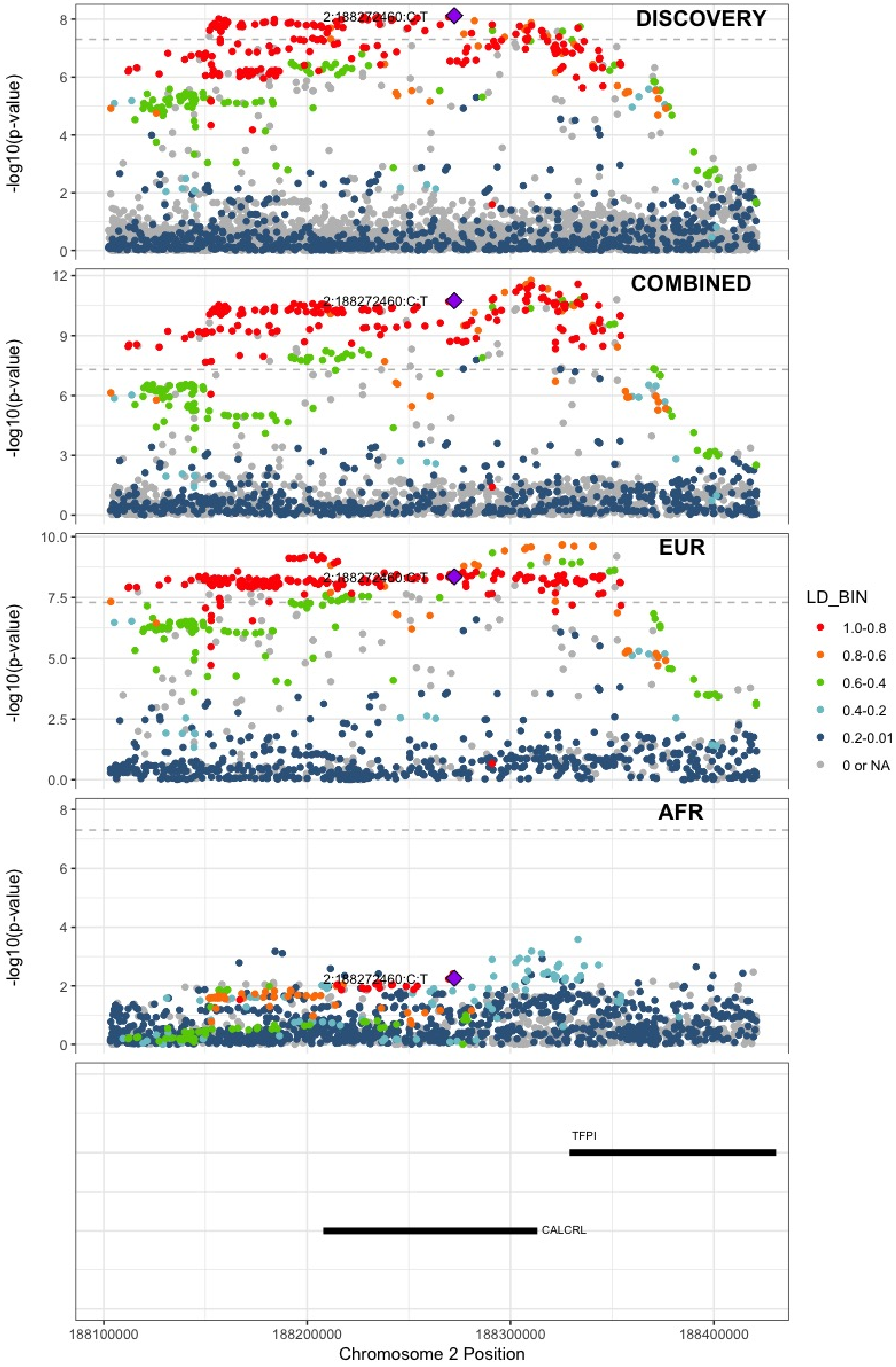

**Supplementary Figure 5.10** – rs715 (chr2:211543055) in locus 21 (CPS1)

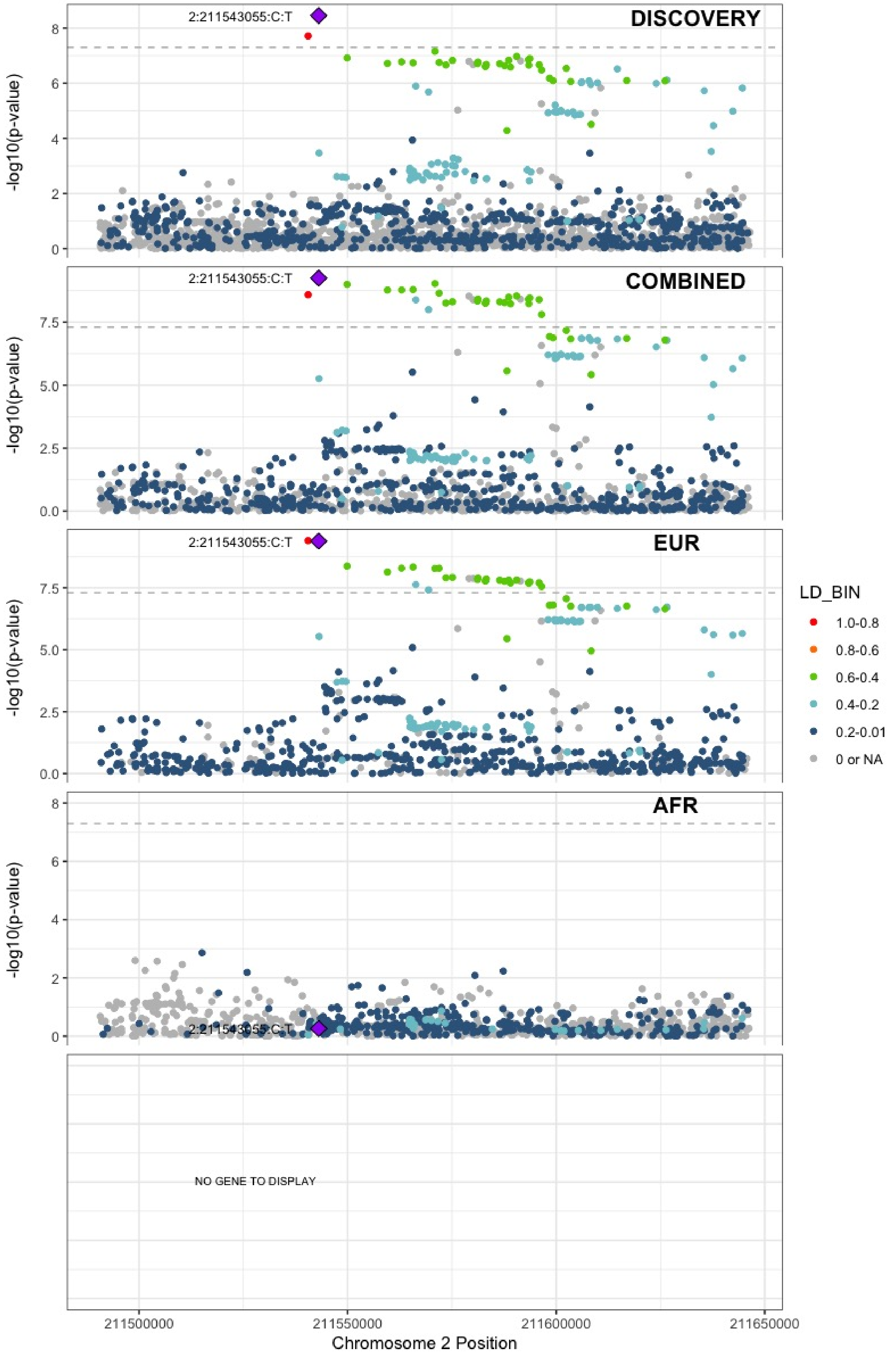

**Supplementary Figure 5.11** – rs13412535 (chr2:224874874) in locus 22 (SERPINE2)

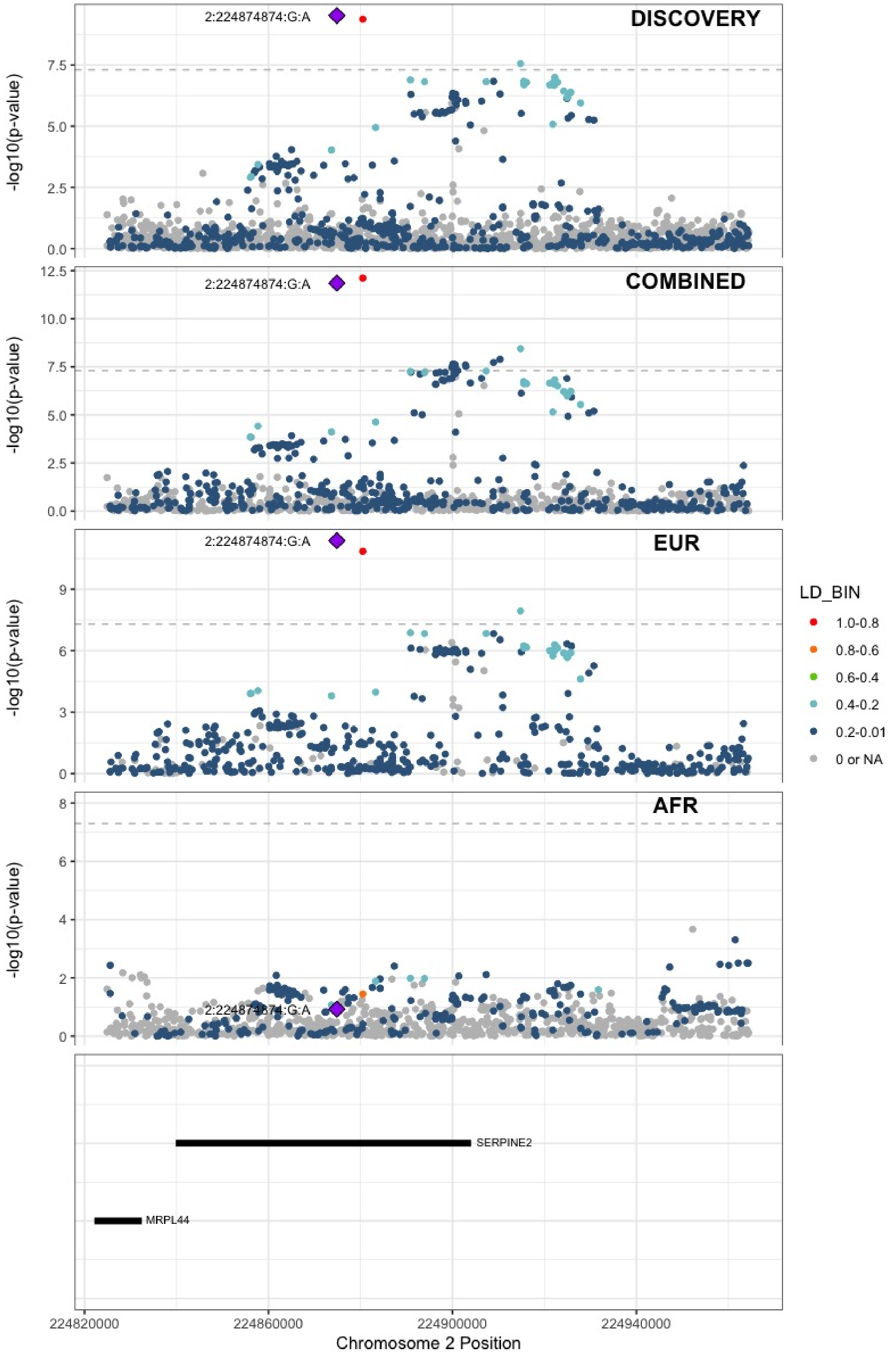

**Supplementary Figure 5.12** – rs2960420 (chr3:12314512) in locus 23 (SYN2;PPARG)

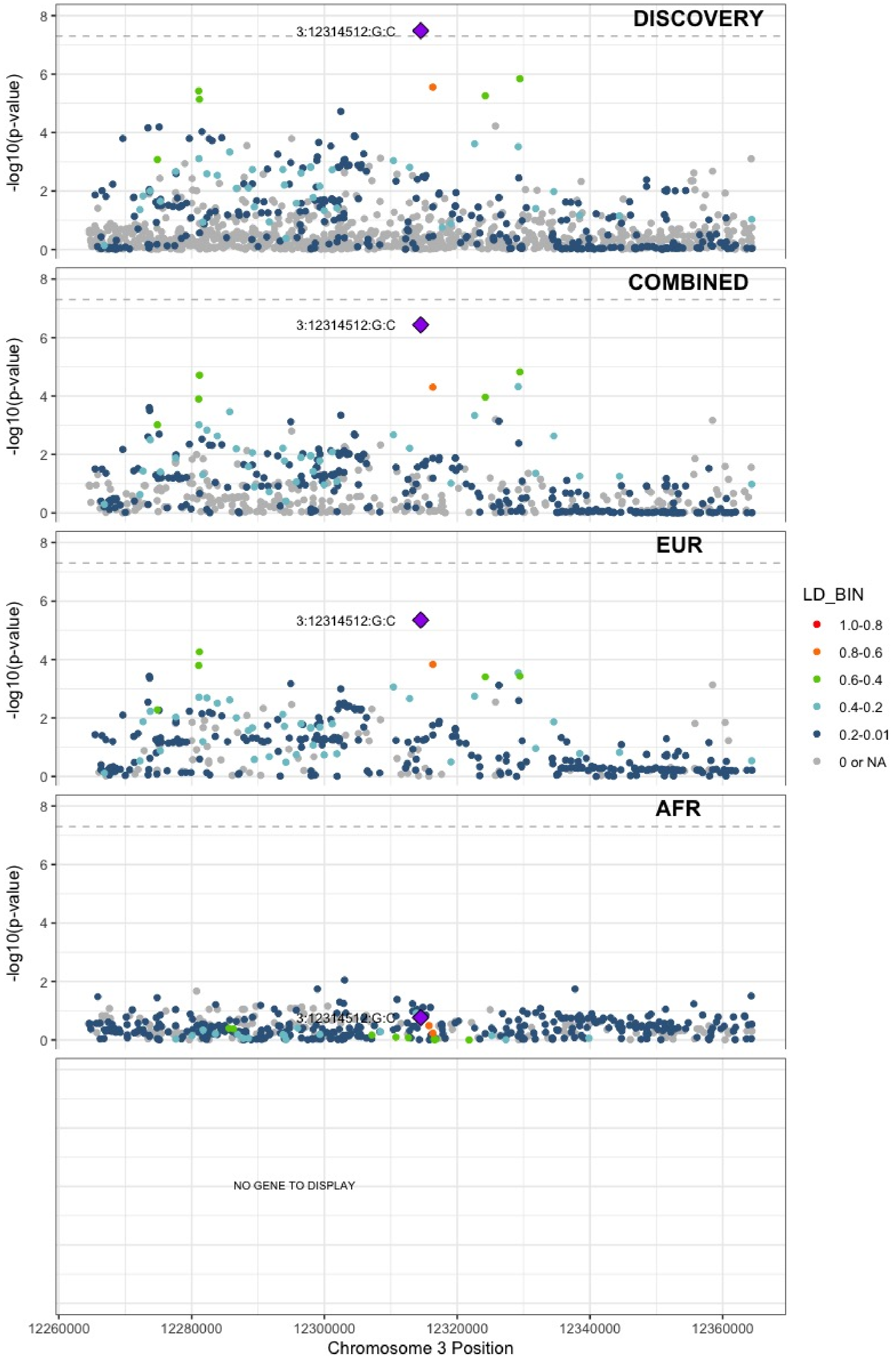

**Supplementary Figure 5.13** – rs13084580 (chr3:39188182) in locus 24 (CSRNP1)

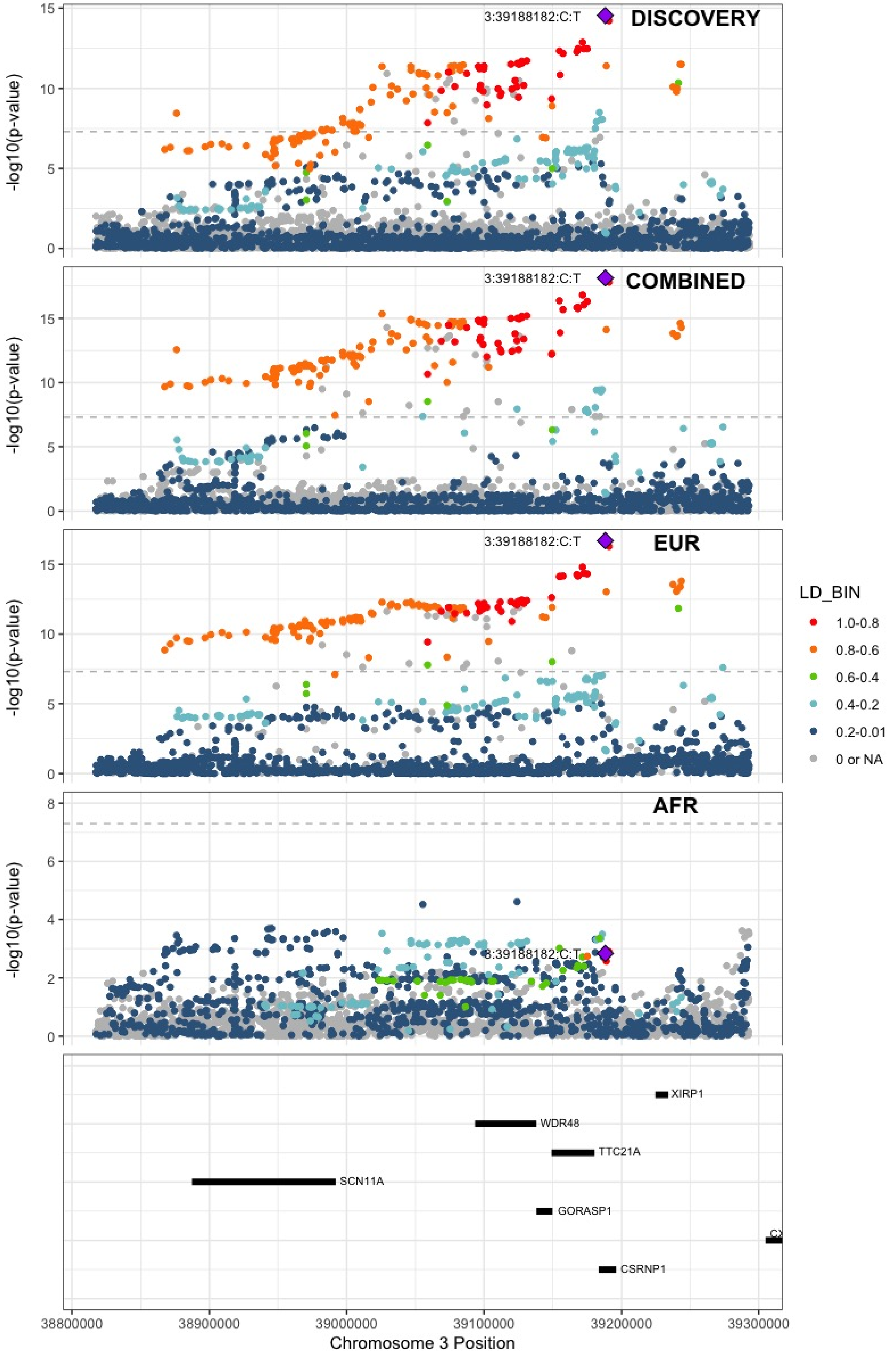

**Supplementary Figure 5.14** – rs11130326 (chr3:52771920) in locus 25 (NEK4)

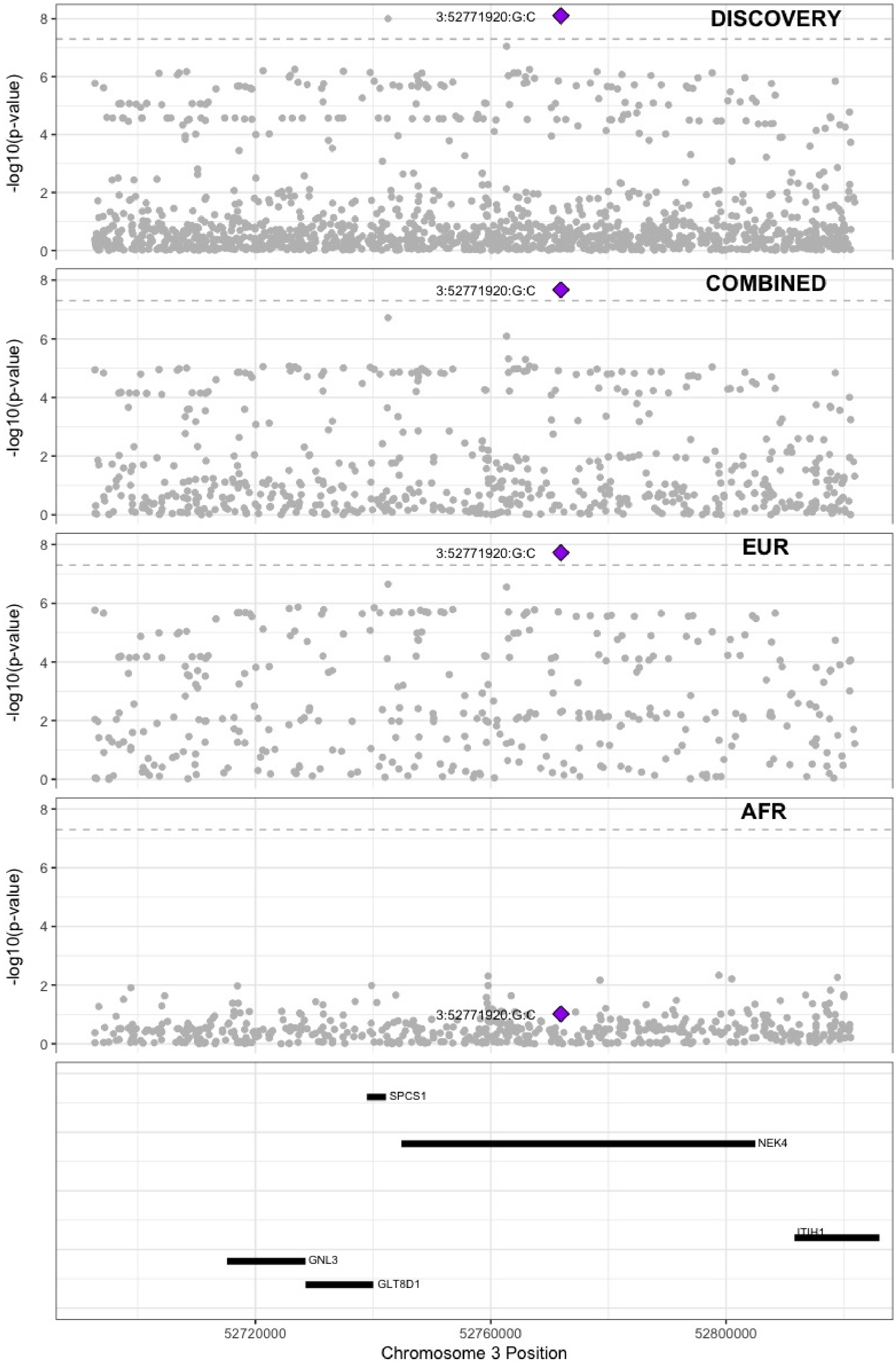

**Supplementary Figure 5.15** – rs562281690 (chr3:90177913) in locus 26 (EPHA3;NONE)

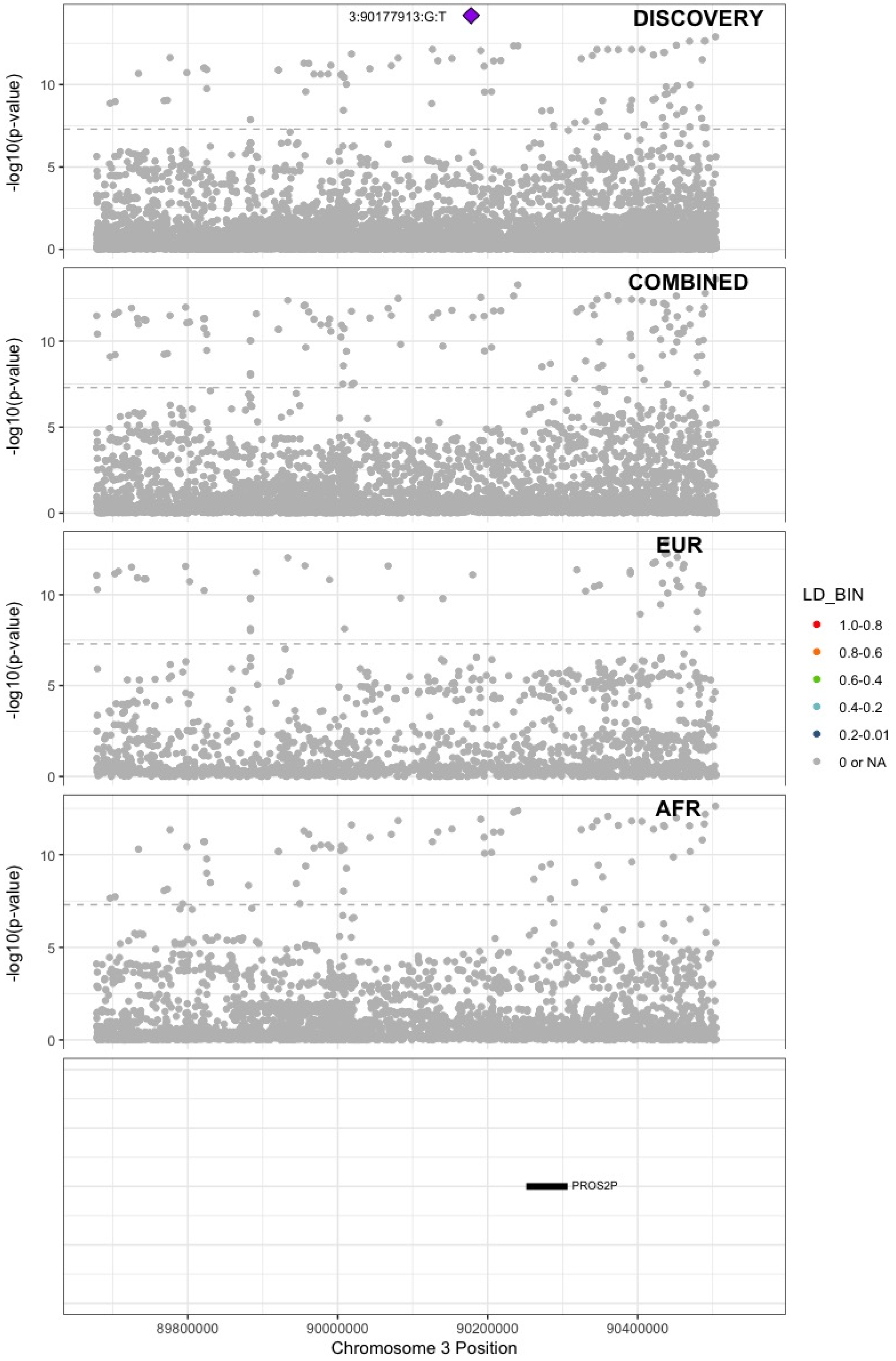

**Supplementary Figure 5.16** – rs79324379 (chr3:93712671) in locus 27 (ARL13B)

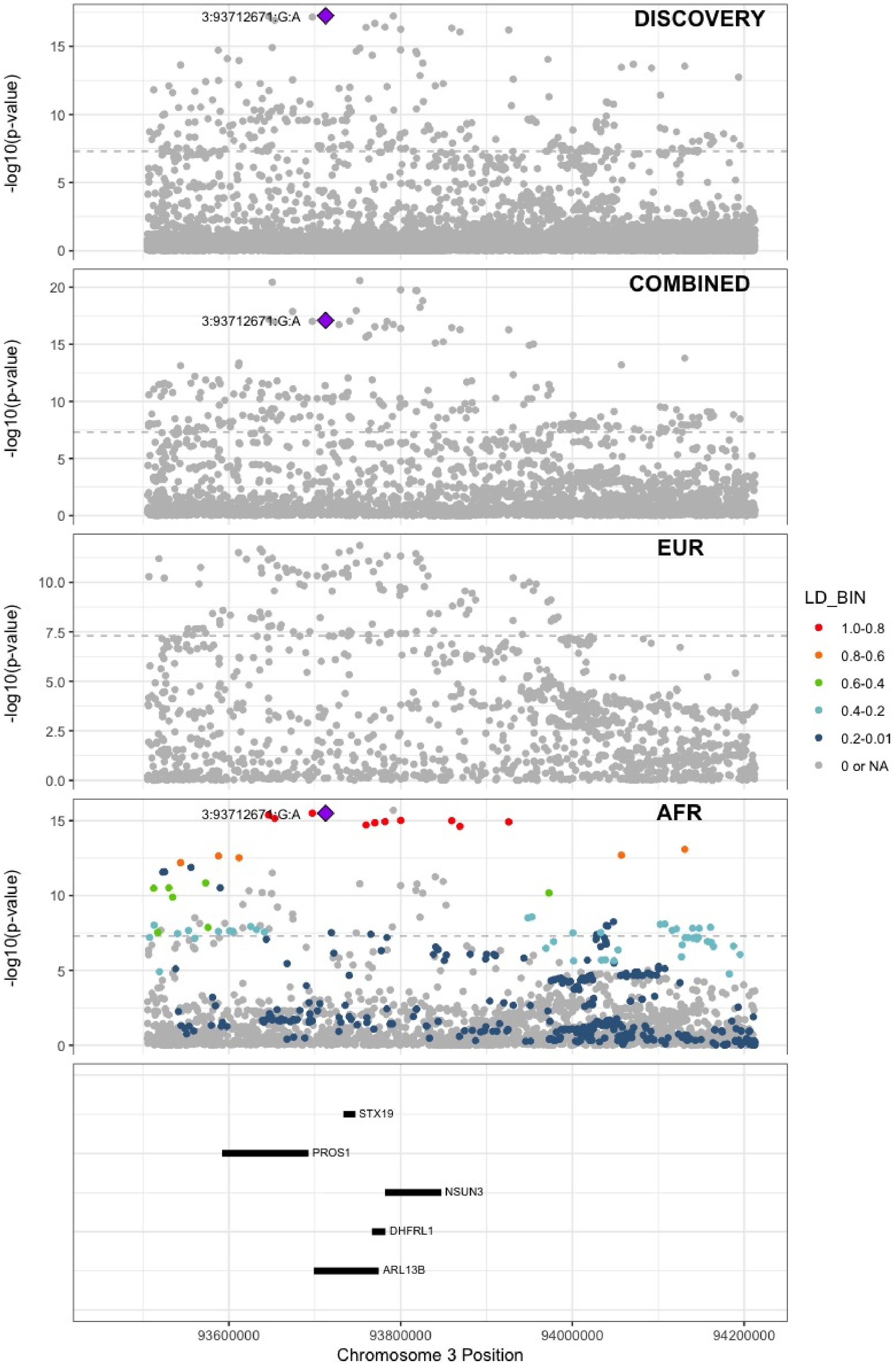

**Supplementary Figure 5.17** – rs9872572 (chr3:123108613) in locus 28 (ADCY5)

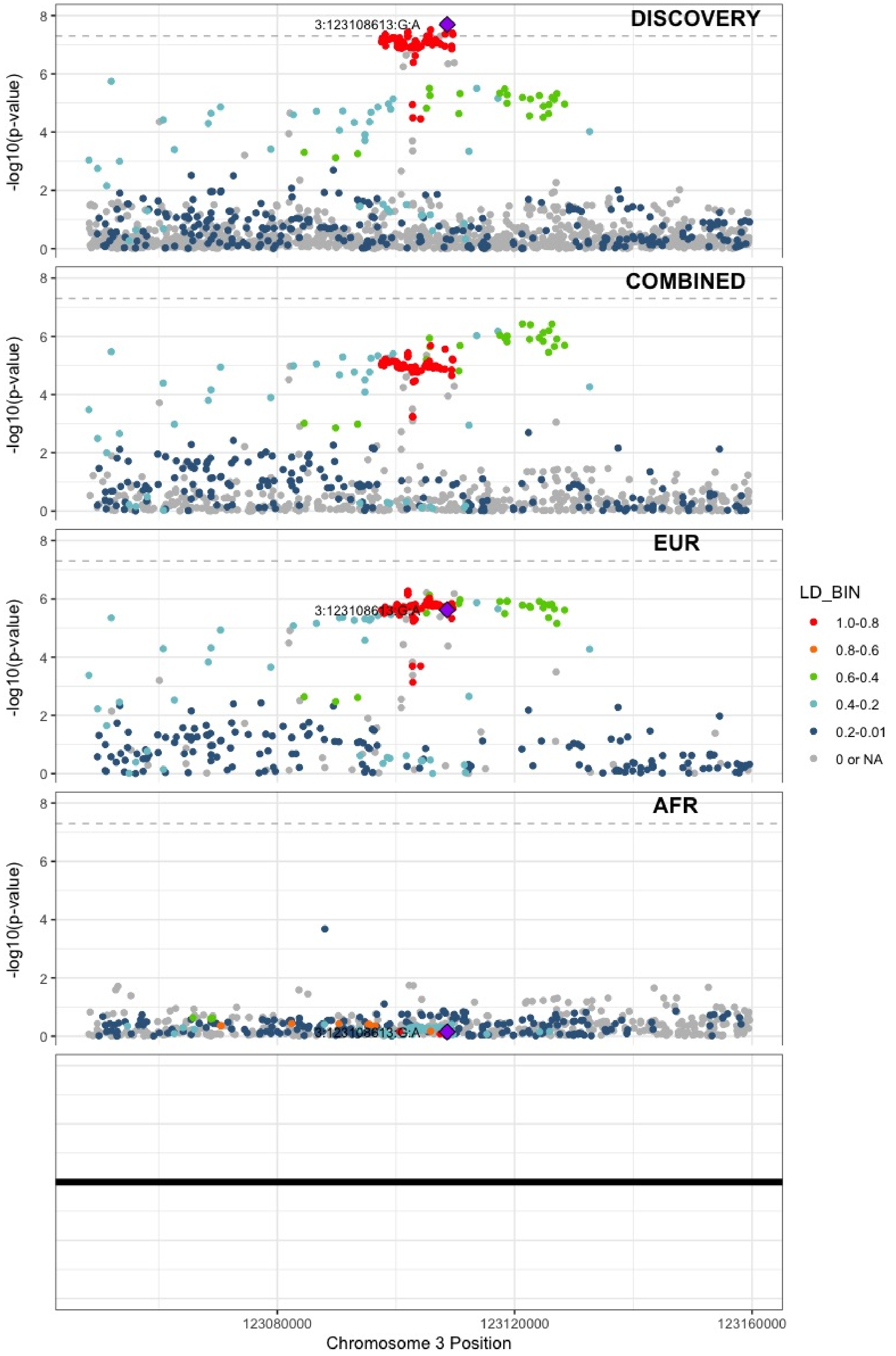

**Supplementary Figure 5.18** – rs62282204 (chr3:138584405) in locus 29 (PIK3CB;LINC01391)

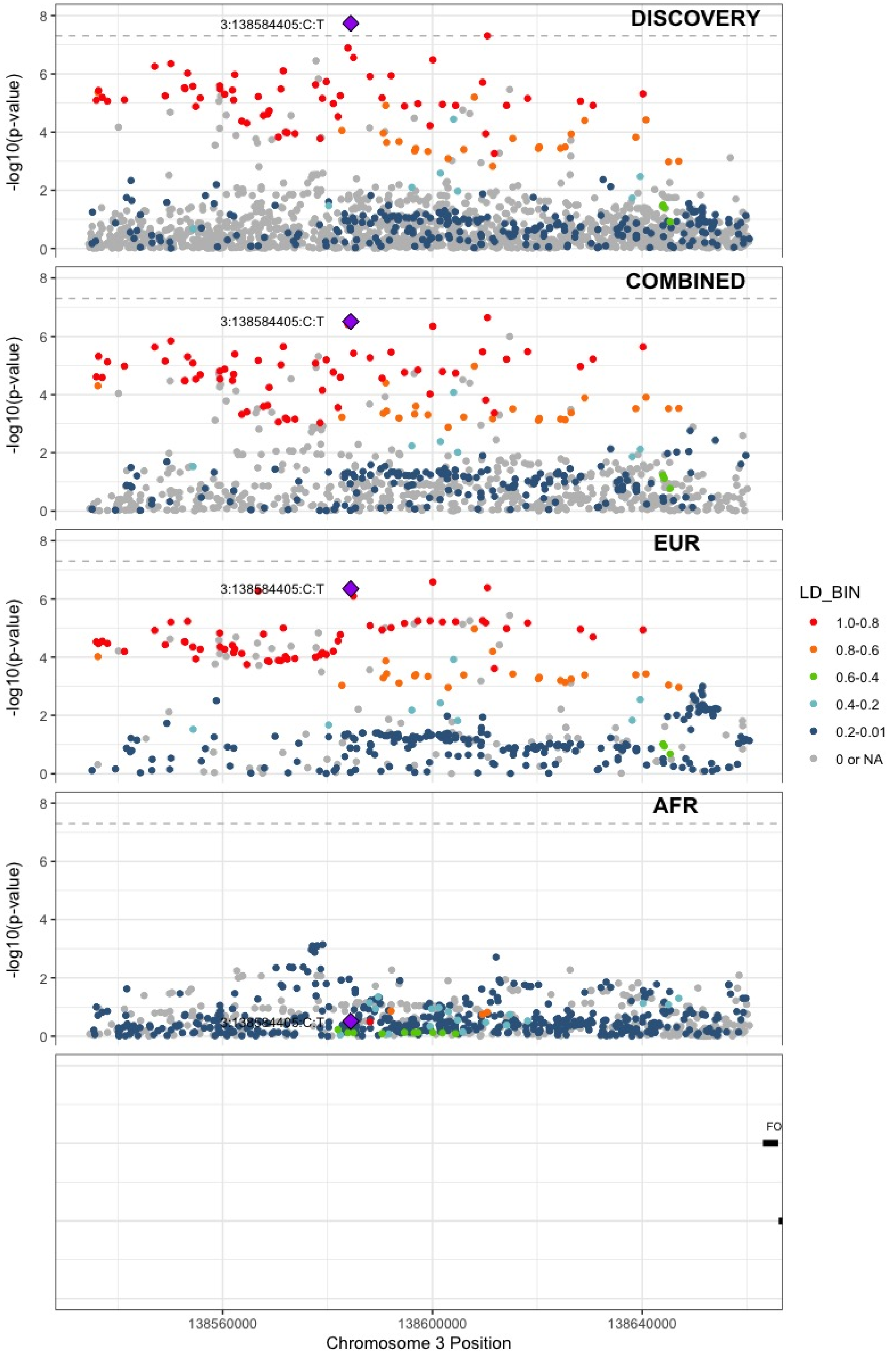

**Supplementary Figure 5.19** – rs7613621 (chr3:169191186) in locus 31 (MECOM)

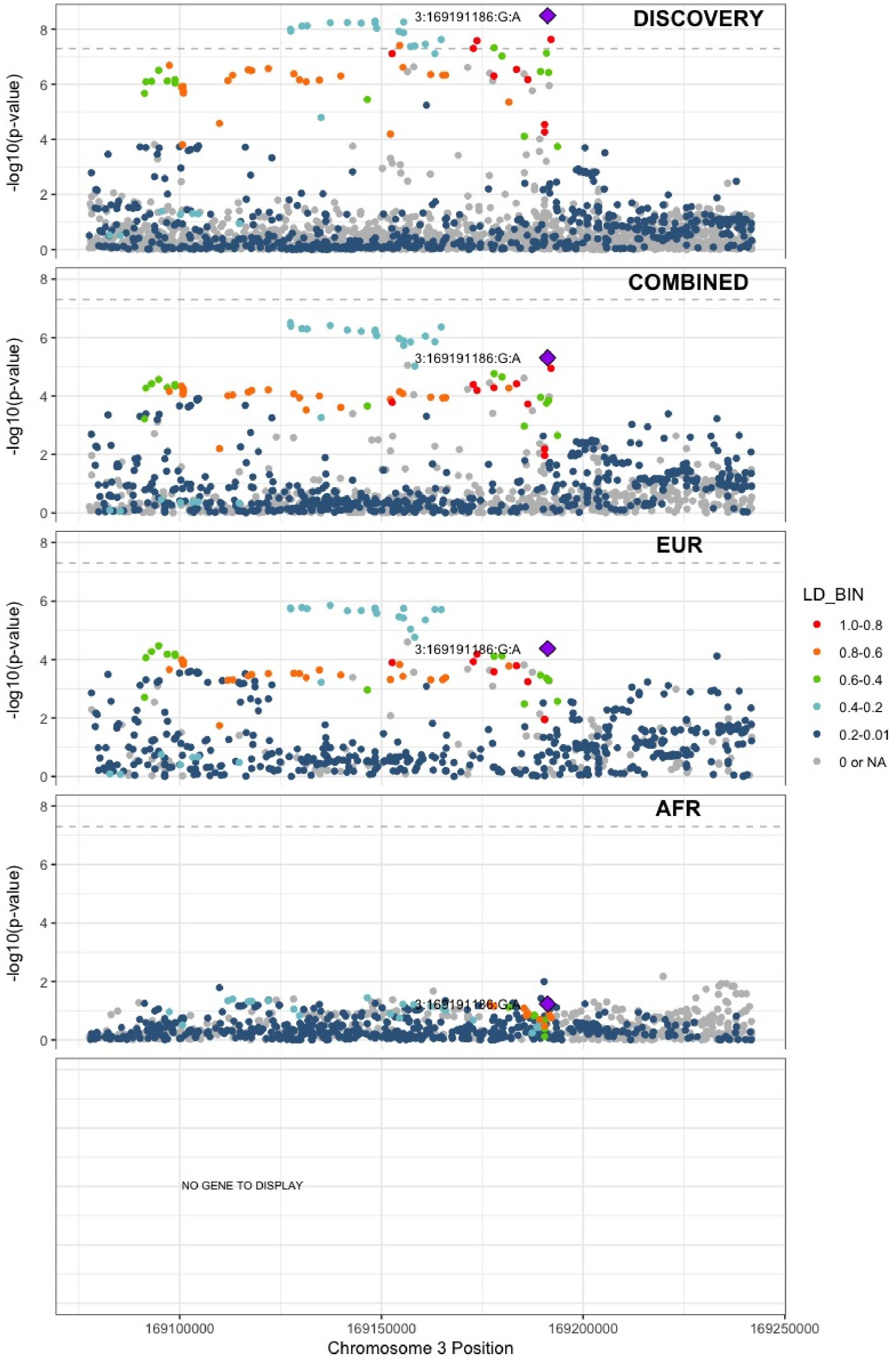

**Supplementary Figure 5.20** – rs710446 (chr3:186459927) in locus 32 (KNG1)

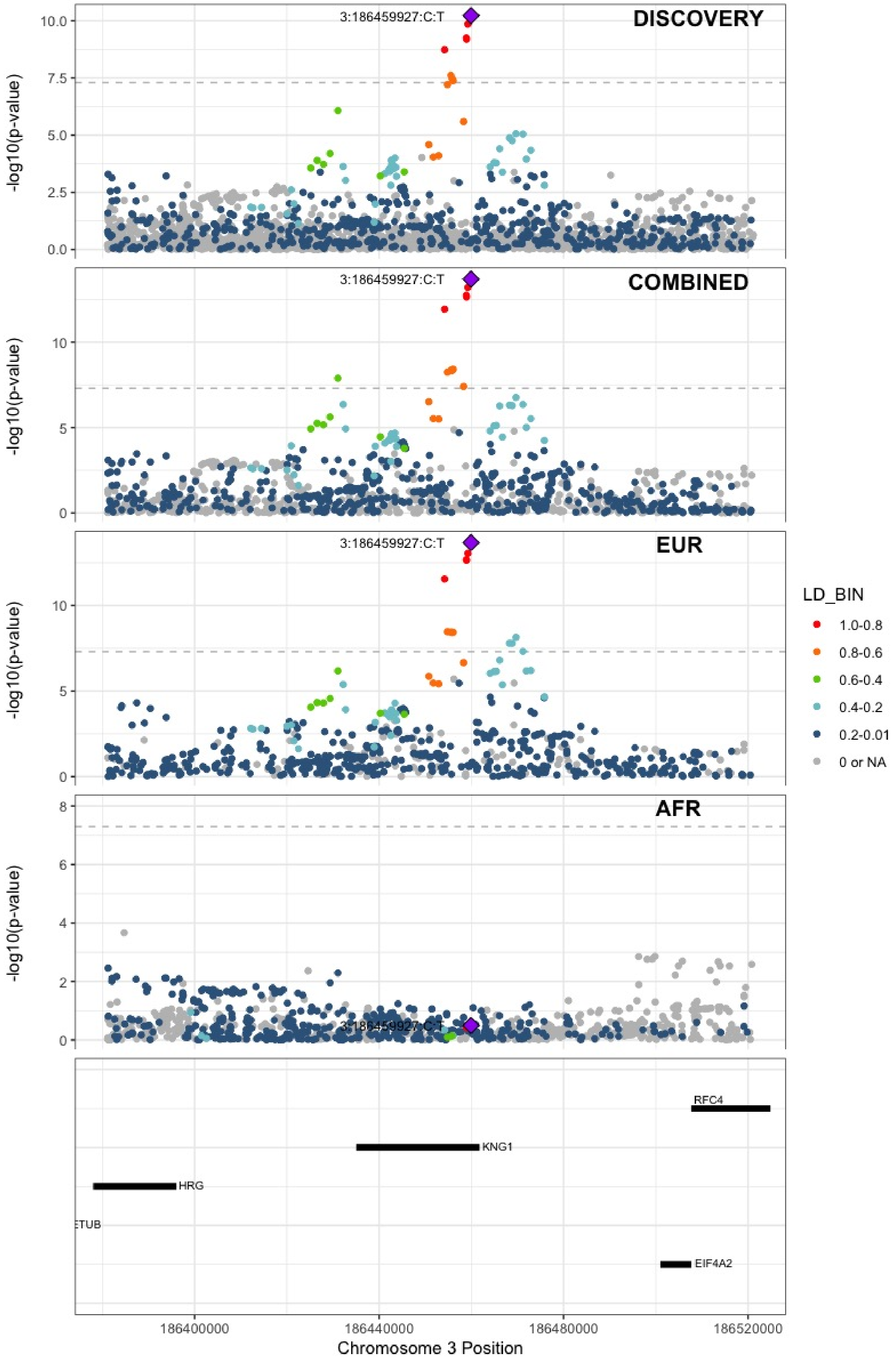

**Supplementary Figure 5.21** – rs6797948 (chr3:194784705) in locus 33 (LINC01968 ;XXYLT1)

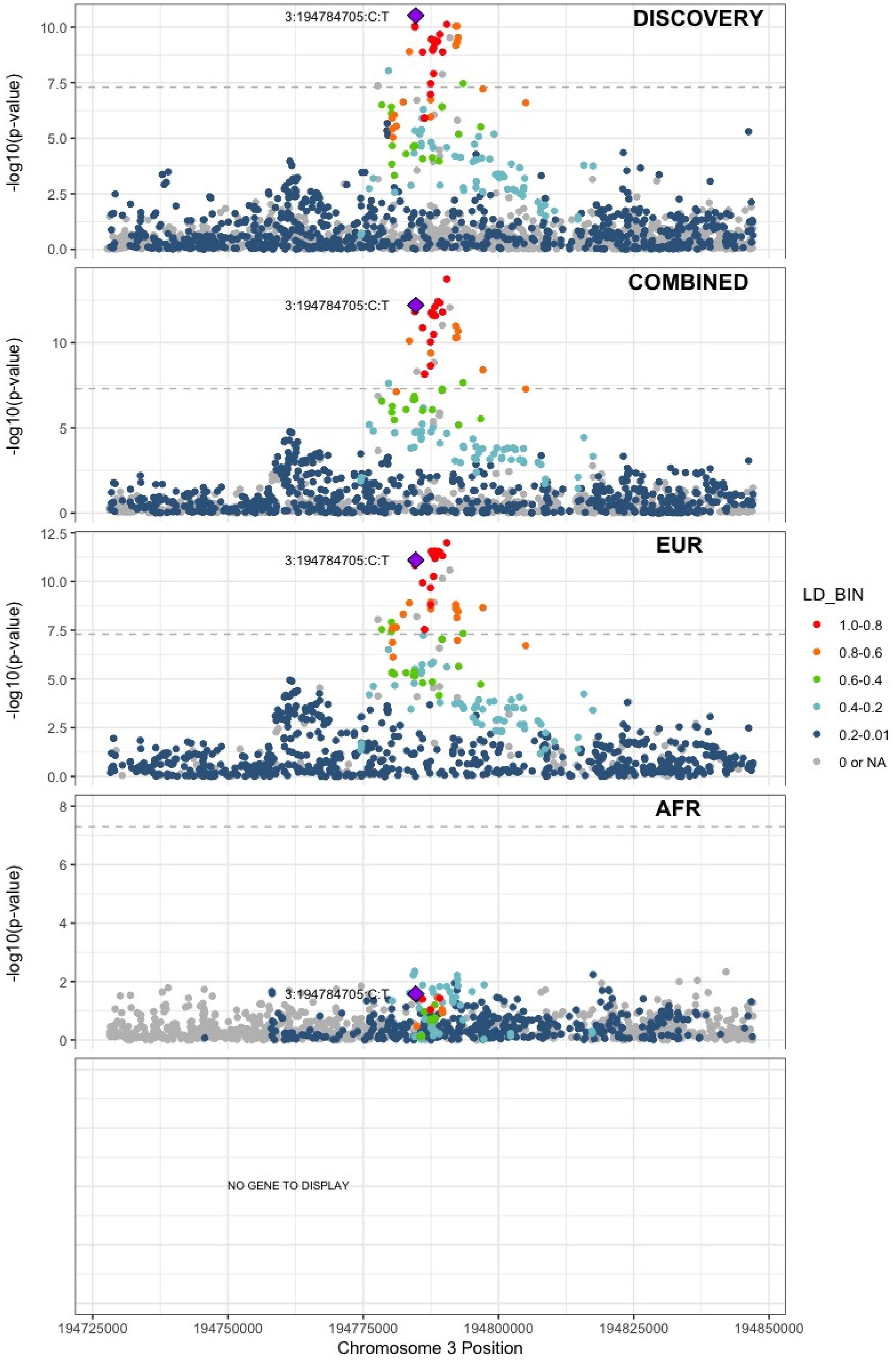

**Supplementary Figure 5.22** – rs6826579 (chr4:83785031) in locus 36 (SEC31A)

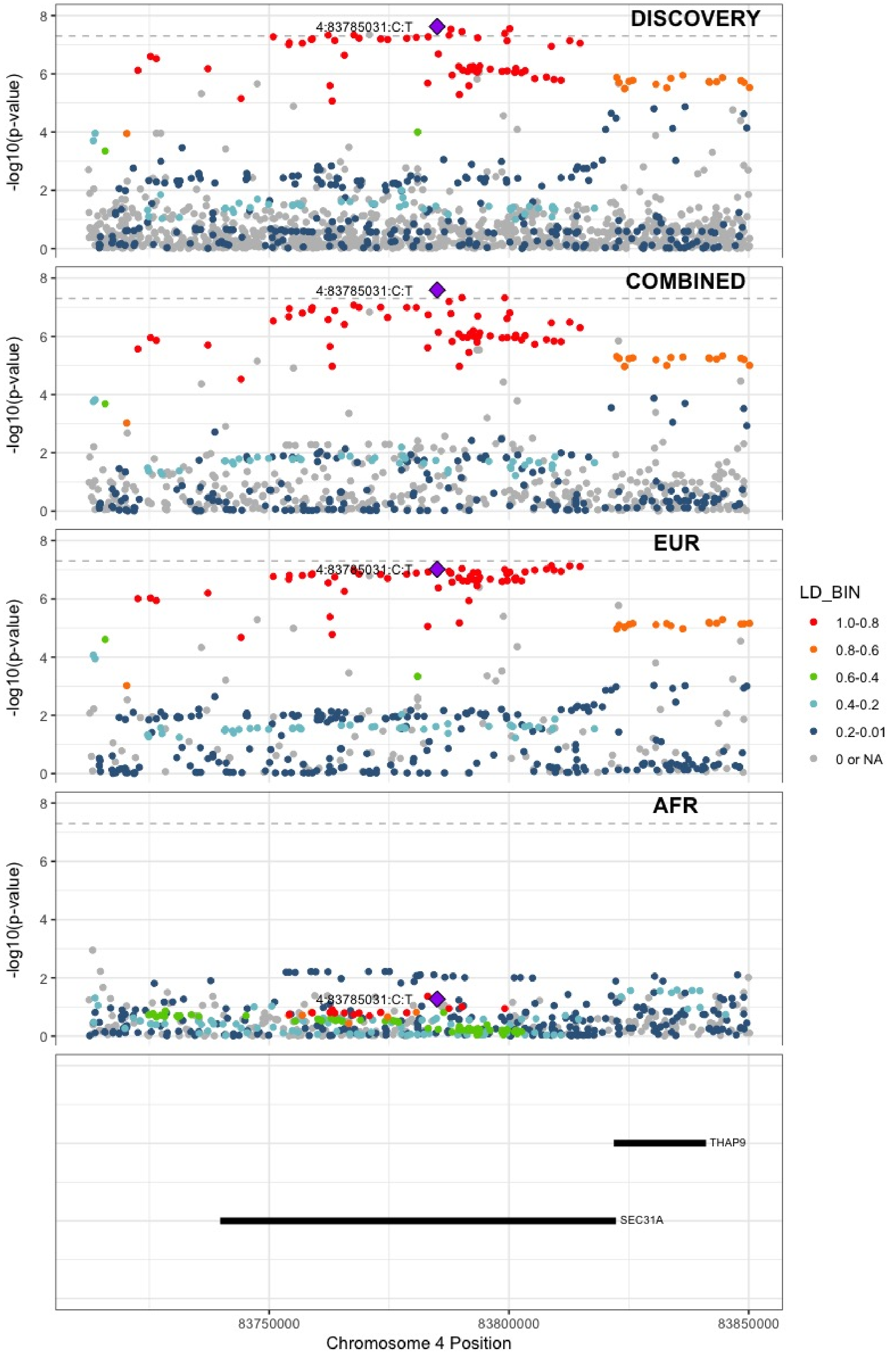

**Supplementary Figure 5.23** – rs17010957 (chr4:86719165) in locus 37 (ARHGAP24)

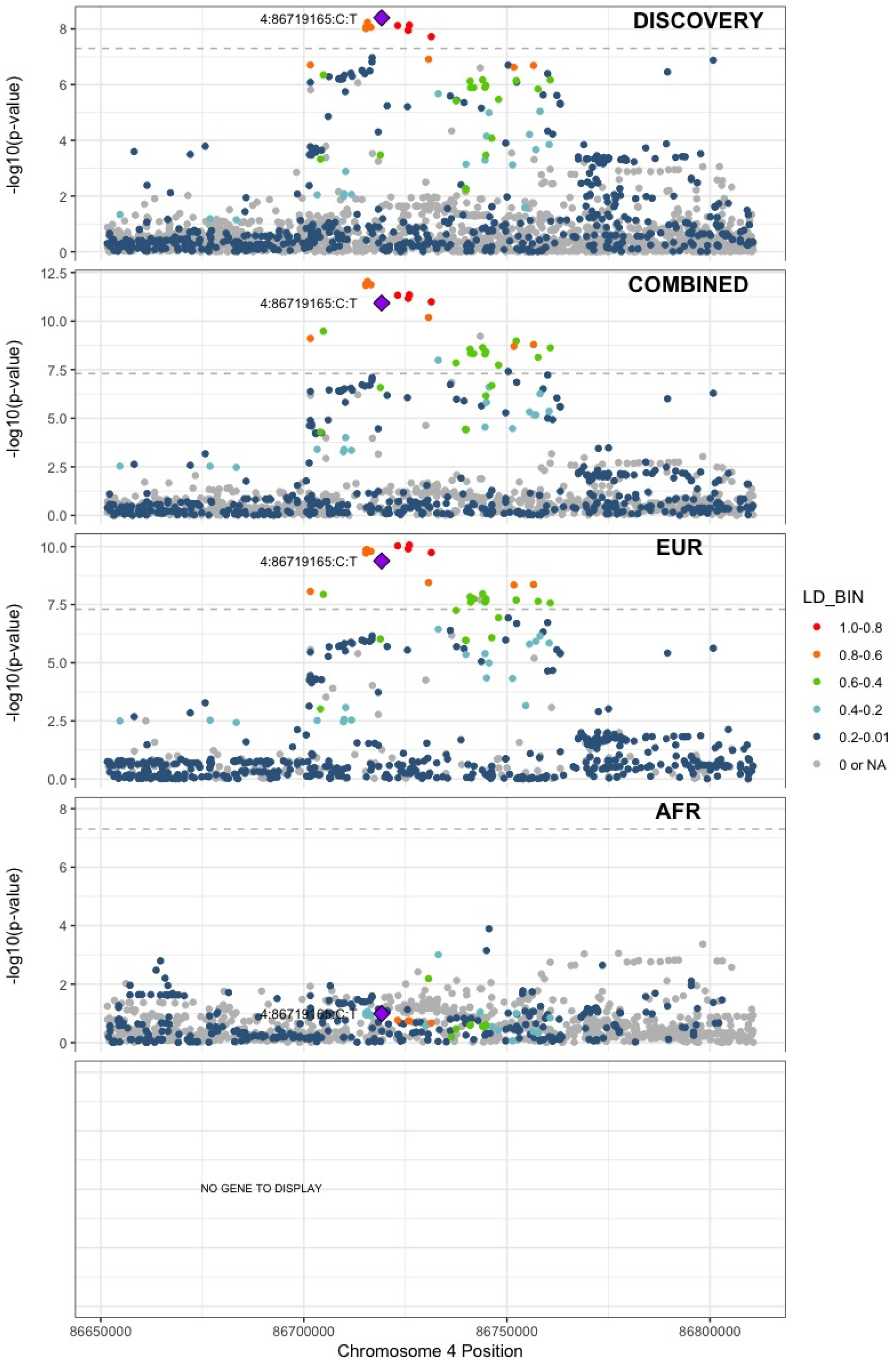

**Supplementary Figure 5.24** – rs2066864 (chr4:155525695) in locus 39 (FGG)

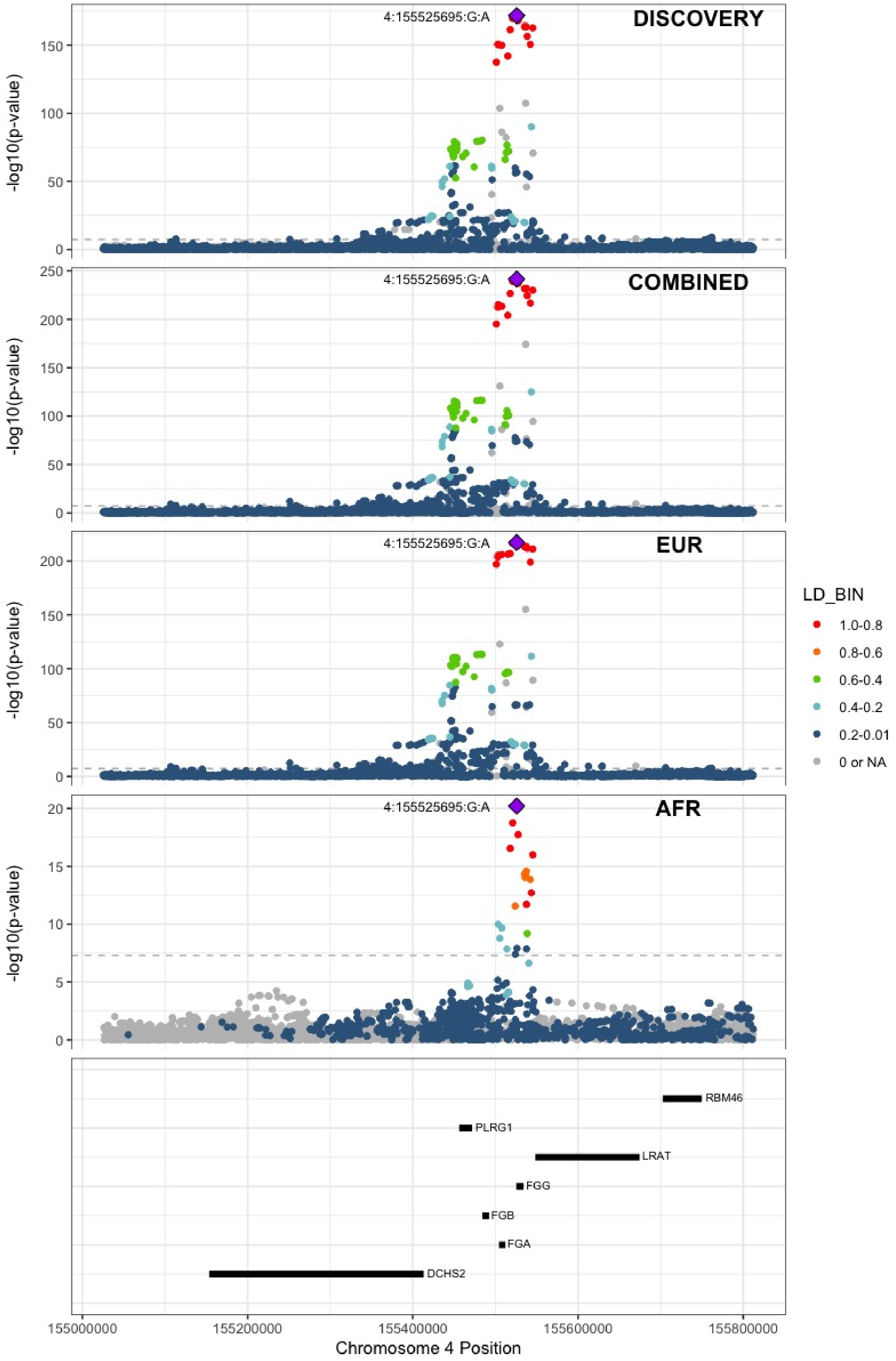

**Supplementary Figure 5.25** – rs3756011 (chr4:187206249) in locus 40 (F11)

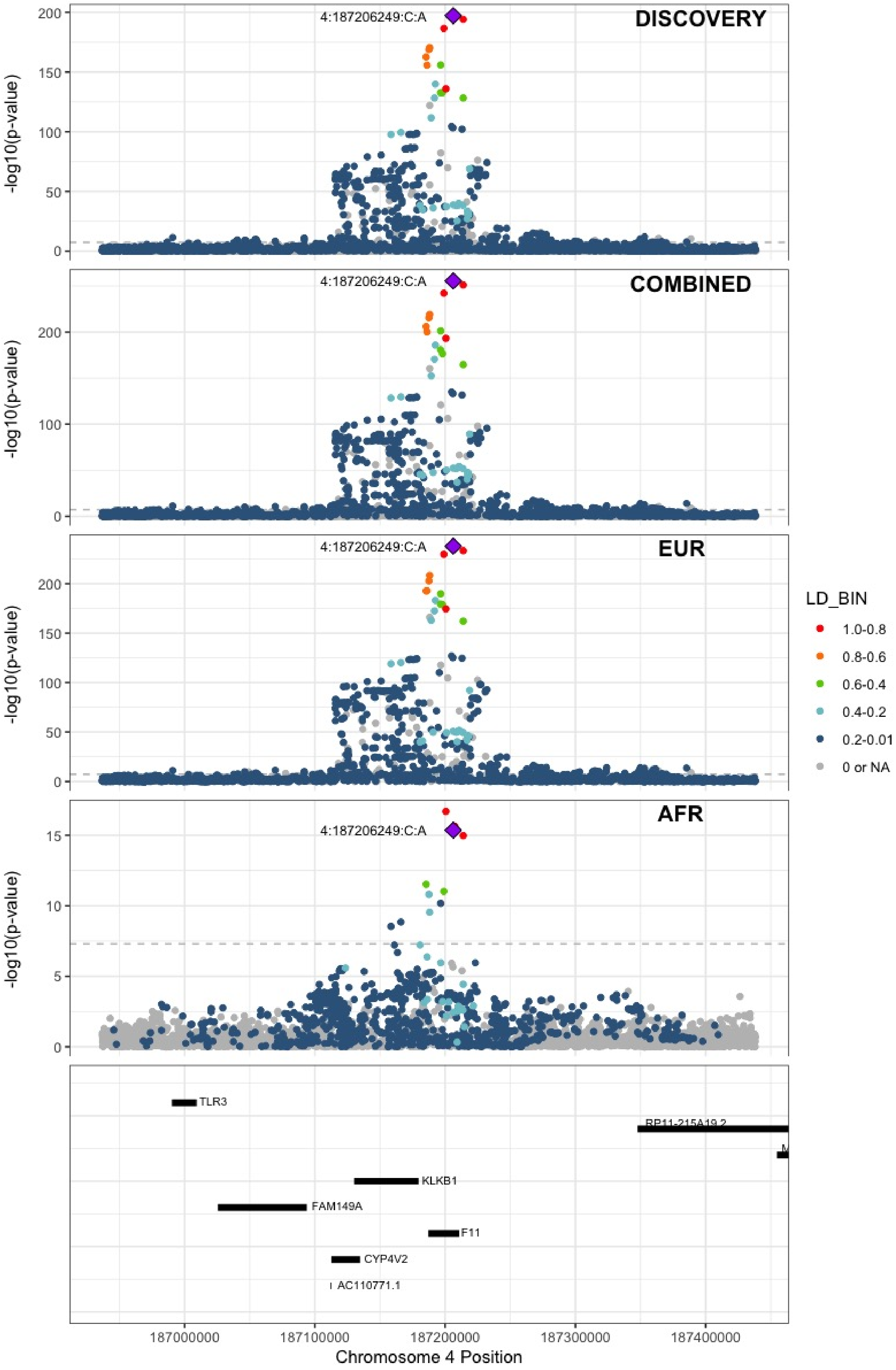

**Supplementary Figure 5.26** – rs16867574 (chr5:38708554) in locus 42 (OSMR-AS1)

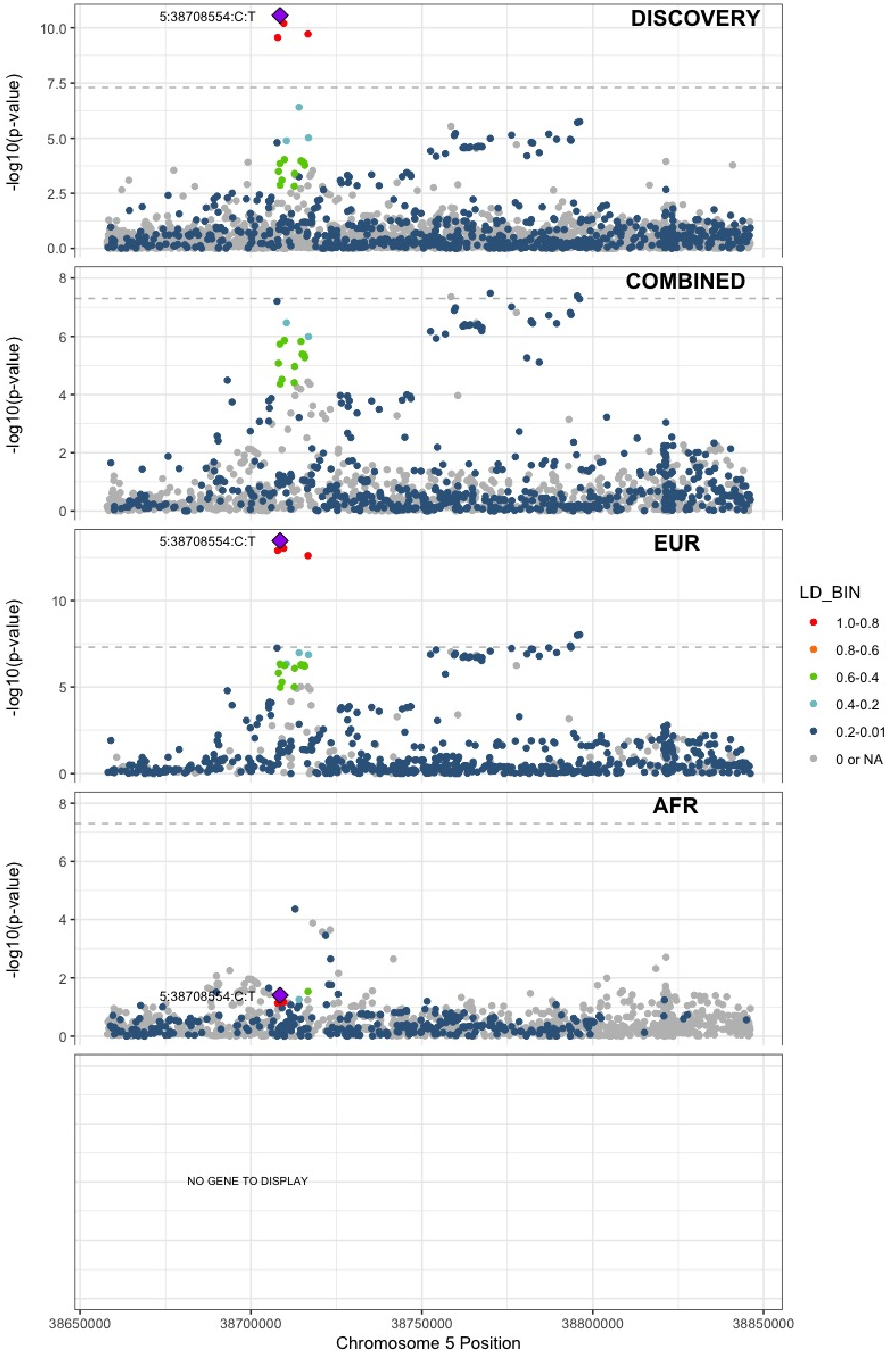

**Supplementary Figure 5.27** – rs38032 (chr5:96321887) in locus 44 (LNPEP)

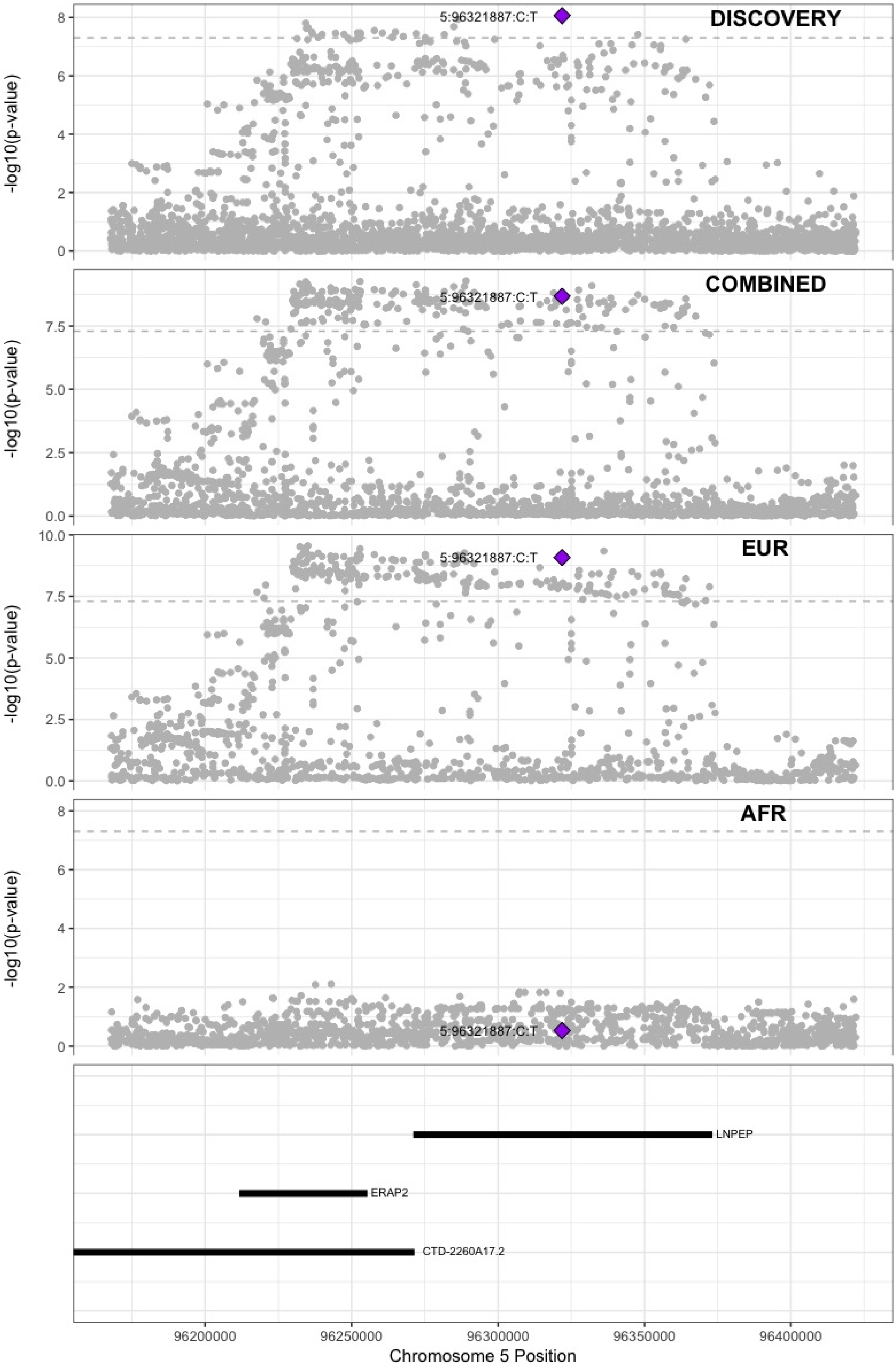

**Supplementary Figure 5.28** – rs9268881 (chr6:32431606) in locus 48 (HLA-DRA ;HLA-DRB5)

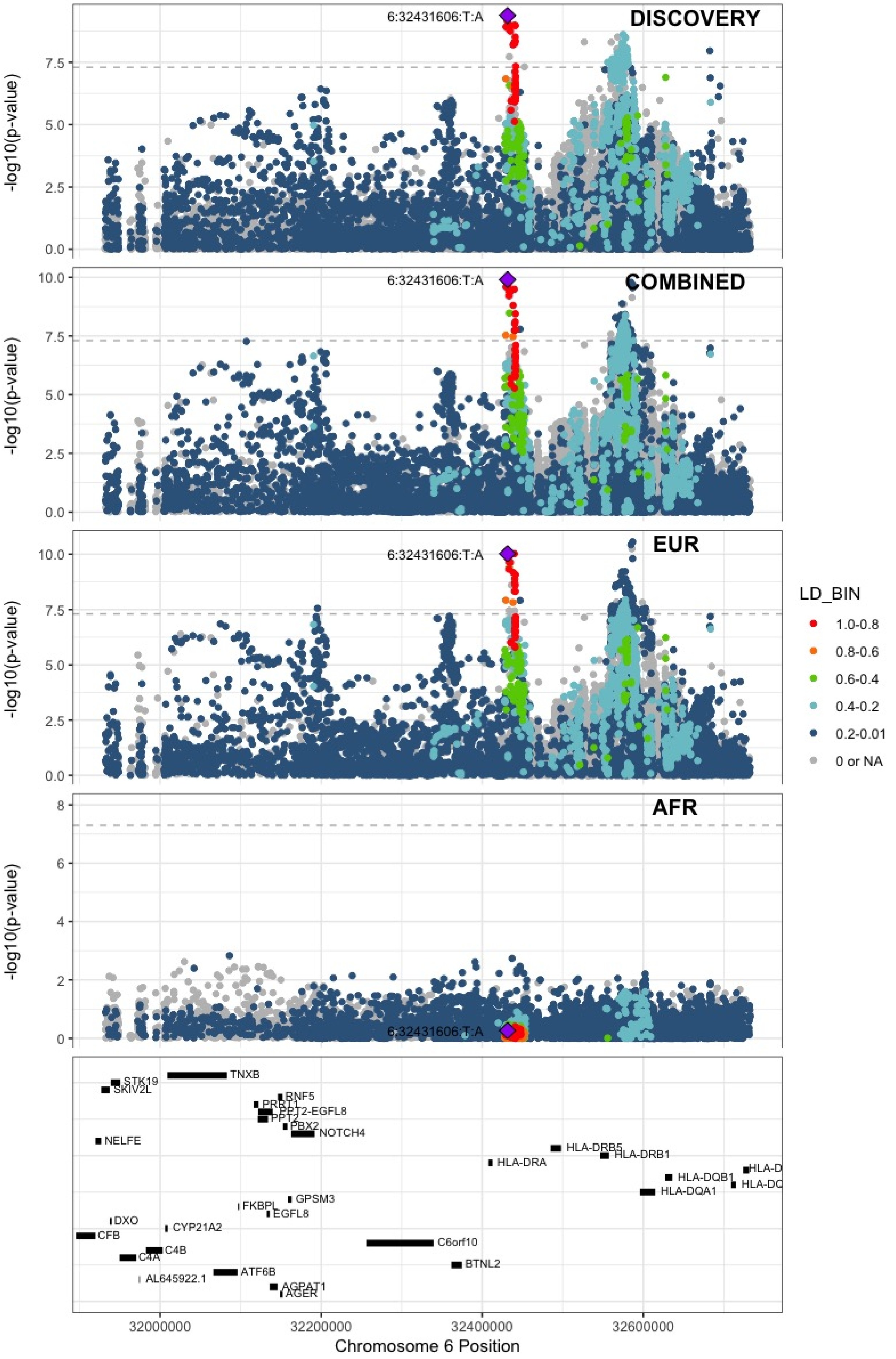

**Supplementary Figure 5.29** – rs145294670 (chr6:34622561) in locus 49 (ILRUN)

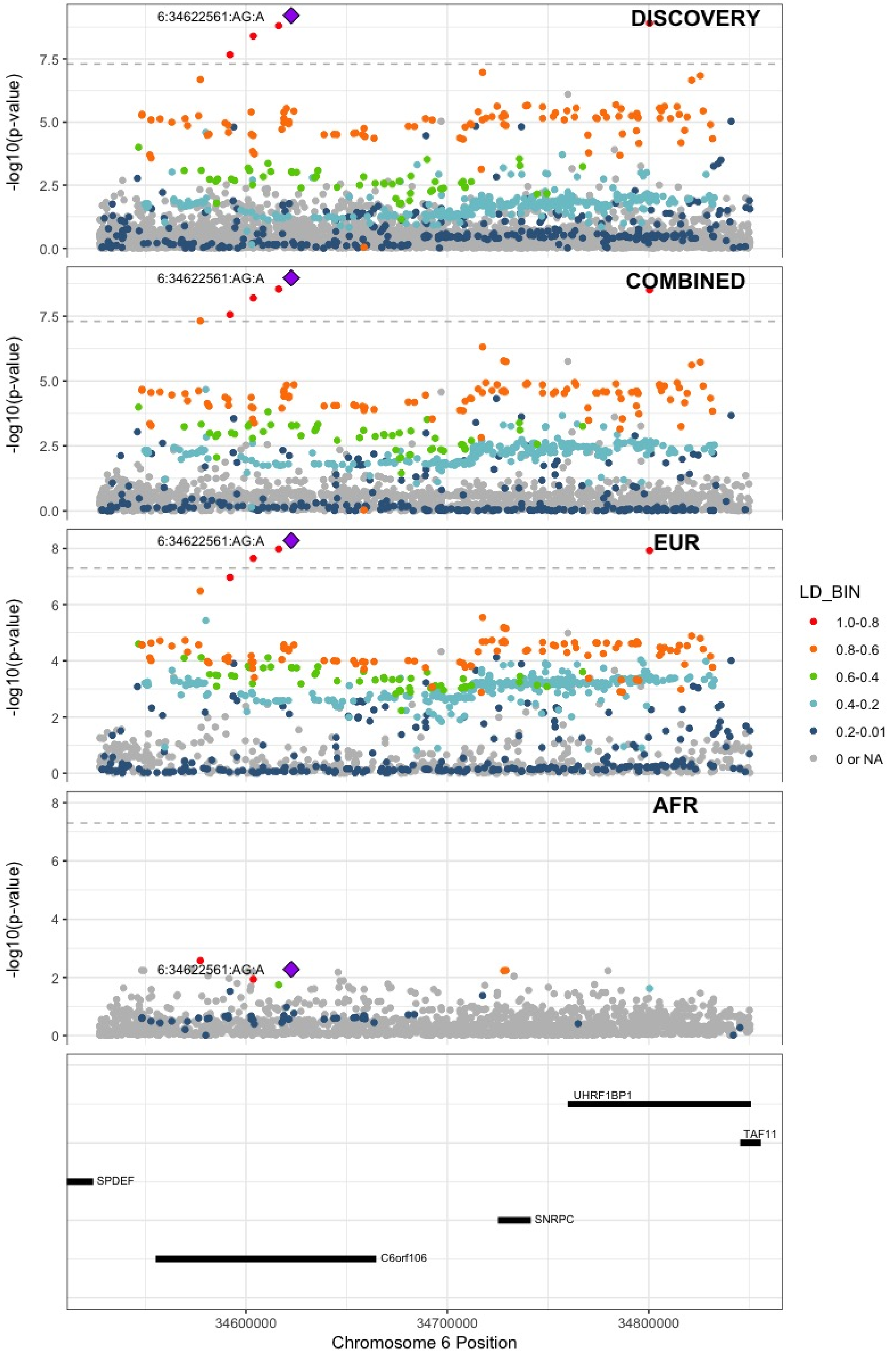

**Supplementary Figure 5.30** – rs903646797 (chr6:40985346) in locus 50 (LOC101929555)

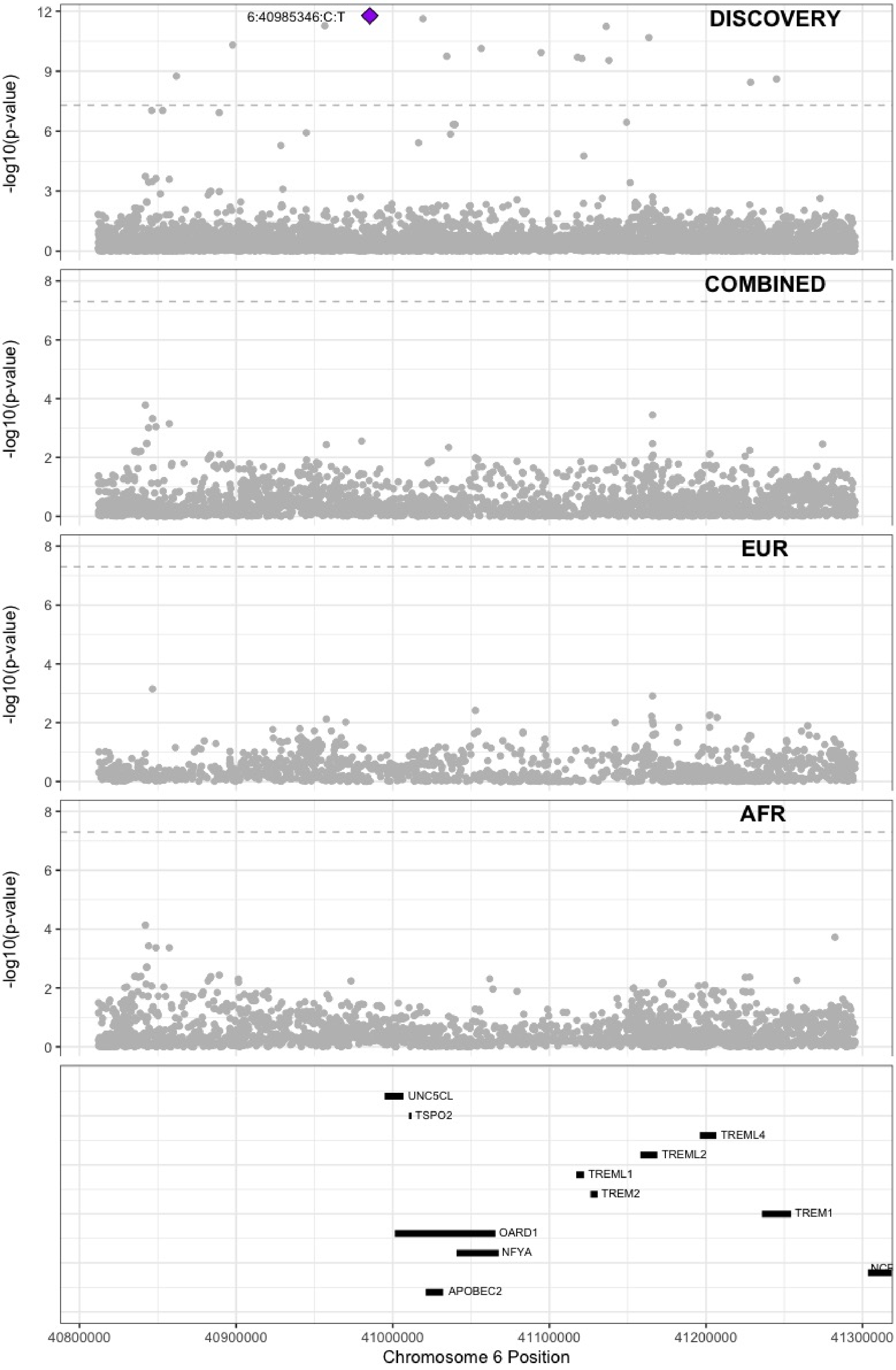

**Supplementary Figure 5.31** – rs9390460 (chr6:147694334) in locus 52 (STXBP5)

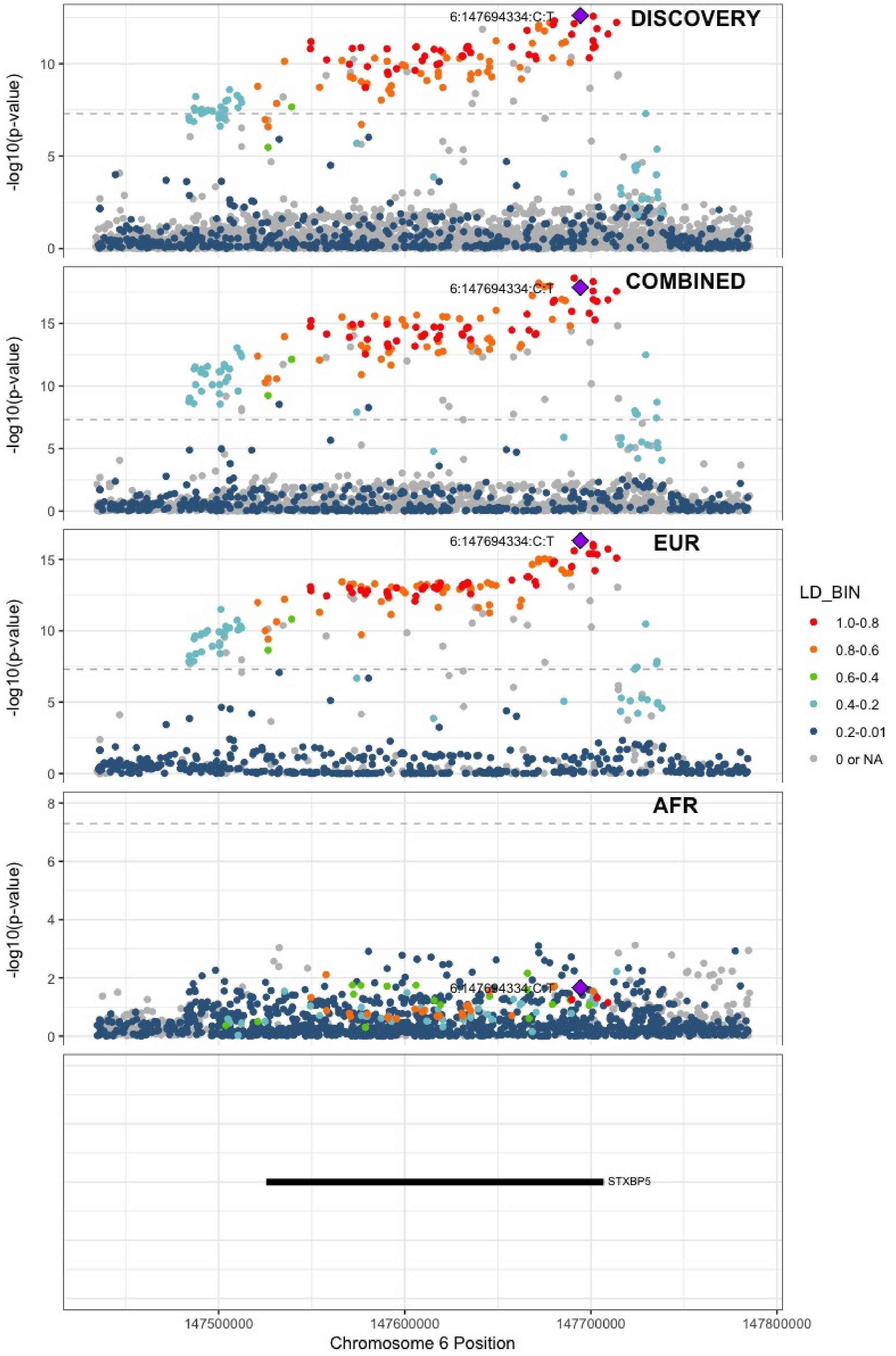

**Supplementary Figure 5.32** – rs42036 (chr7:92241451) in locus 54 (CDK6)

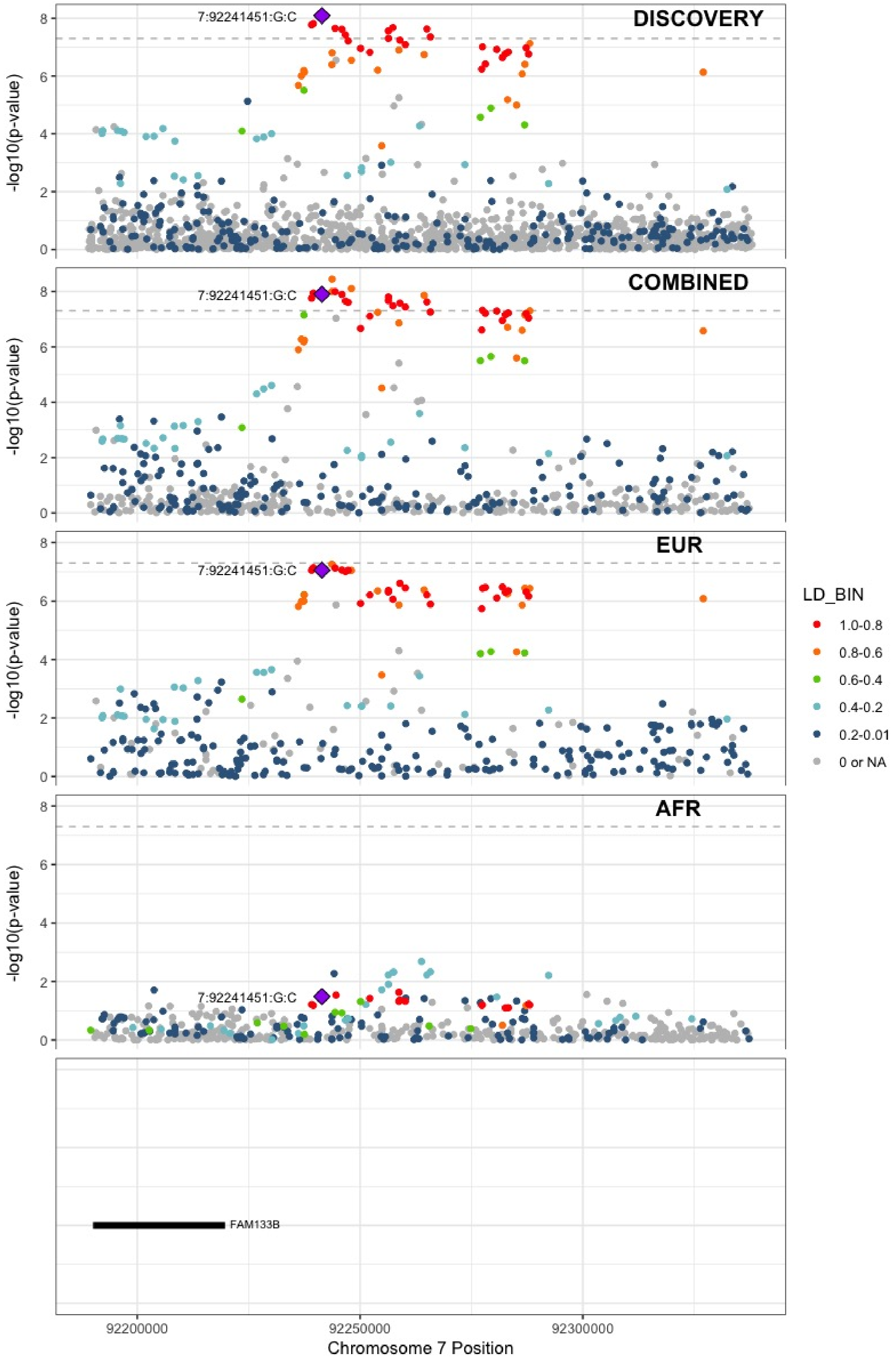

**Supplementary Figure 5.33** – rs538161377 (chr8:4206326) in locus 55 (CSMD1)

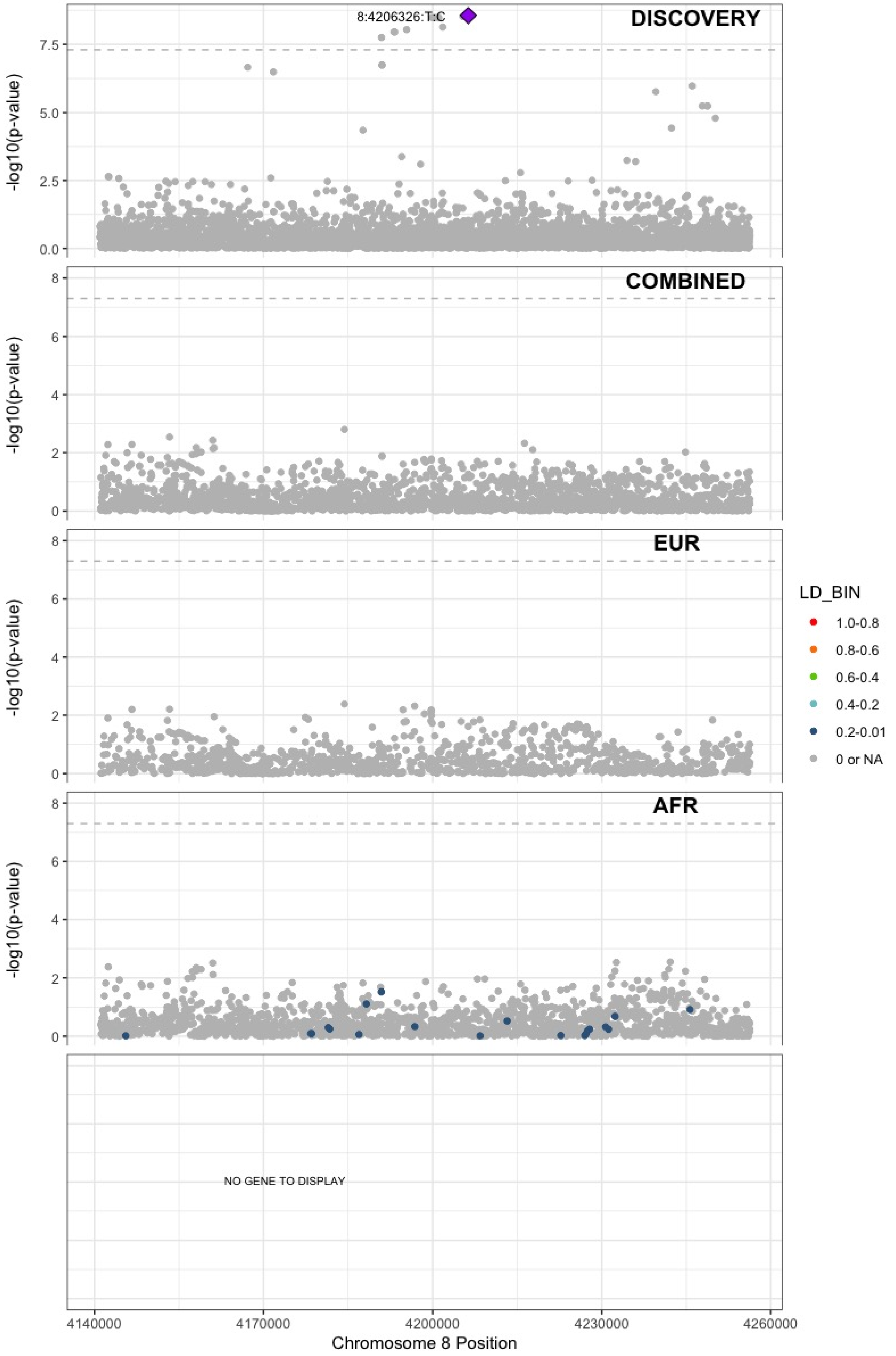

**Supplementary Figure 5.34** – rs67694436 (chr8:6654220) in locus 56 (AGPAT5;XKR5)

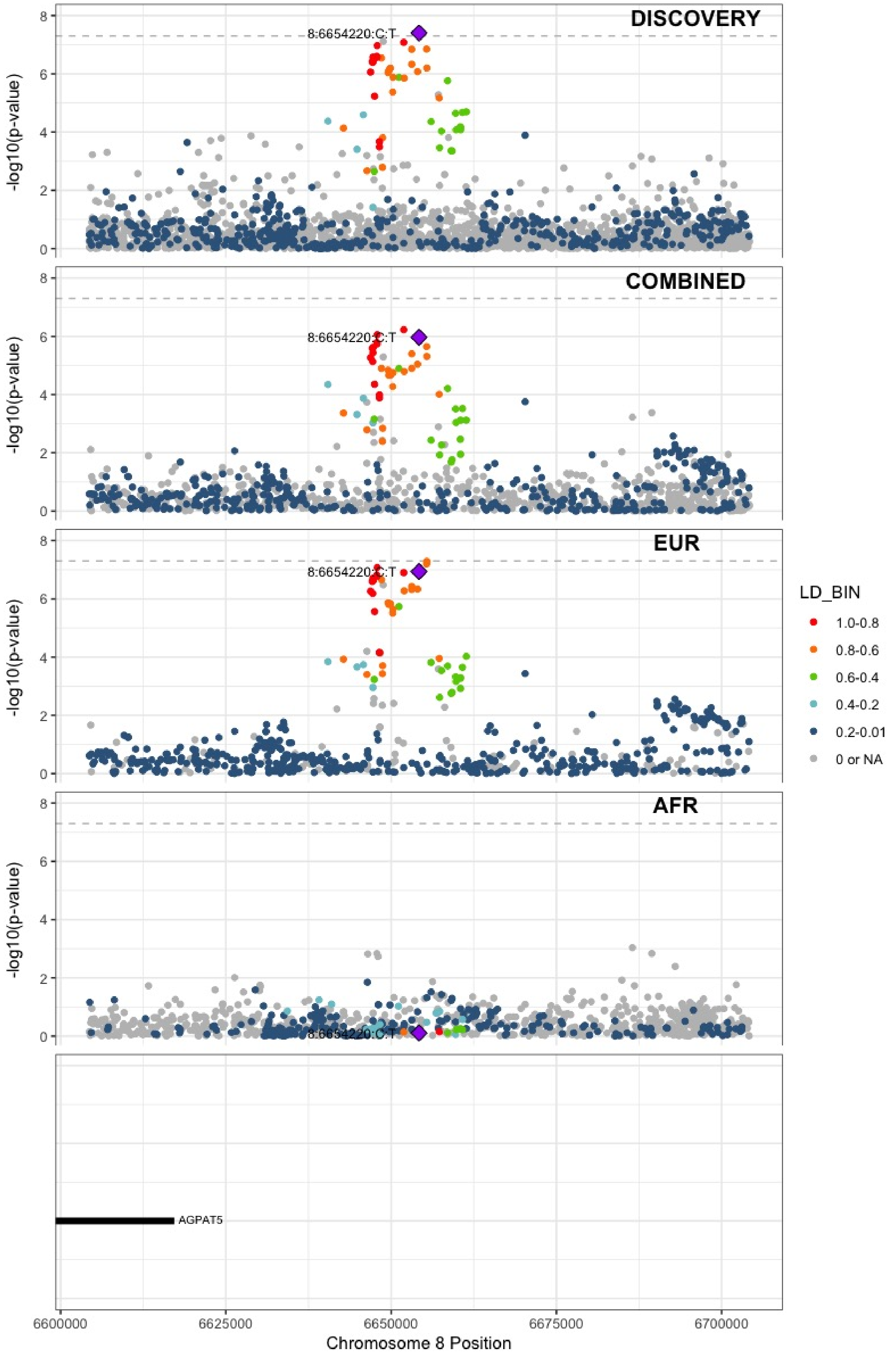

**Supplementary Figure 5.35** – rs7010926 (chr8:10626791) in locus 58 (PINX1)

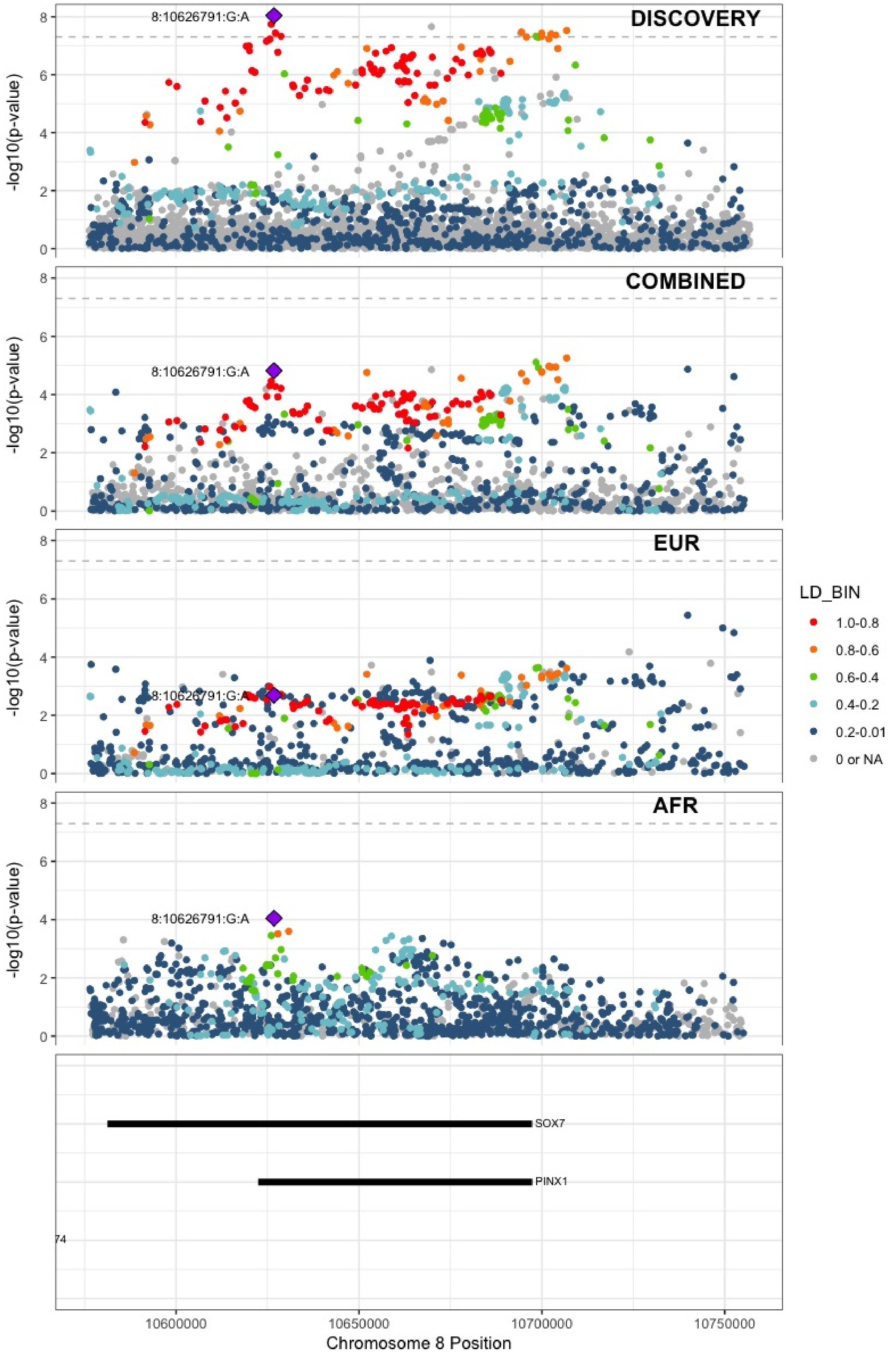

**Supplementary Figure 5.36** – rs2685417 (chr8:27807434) in locus 60 (SCARA5)

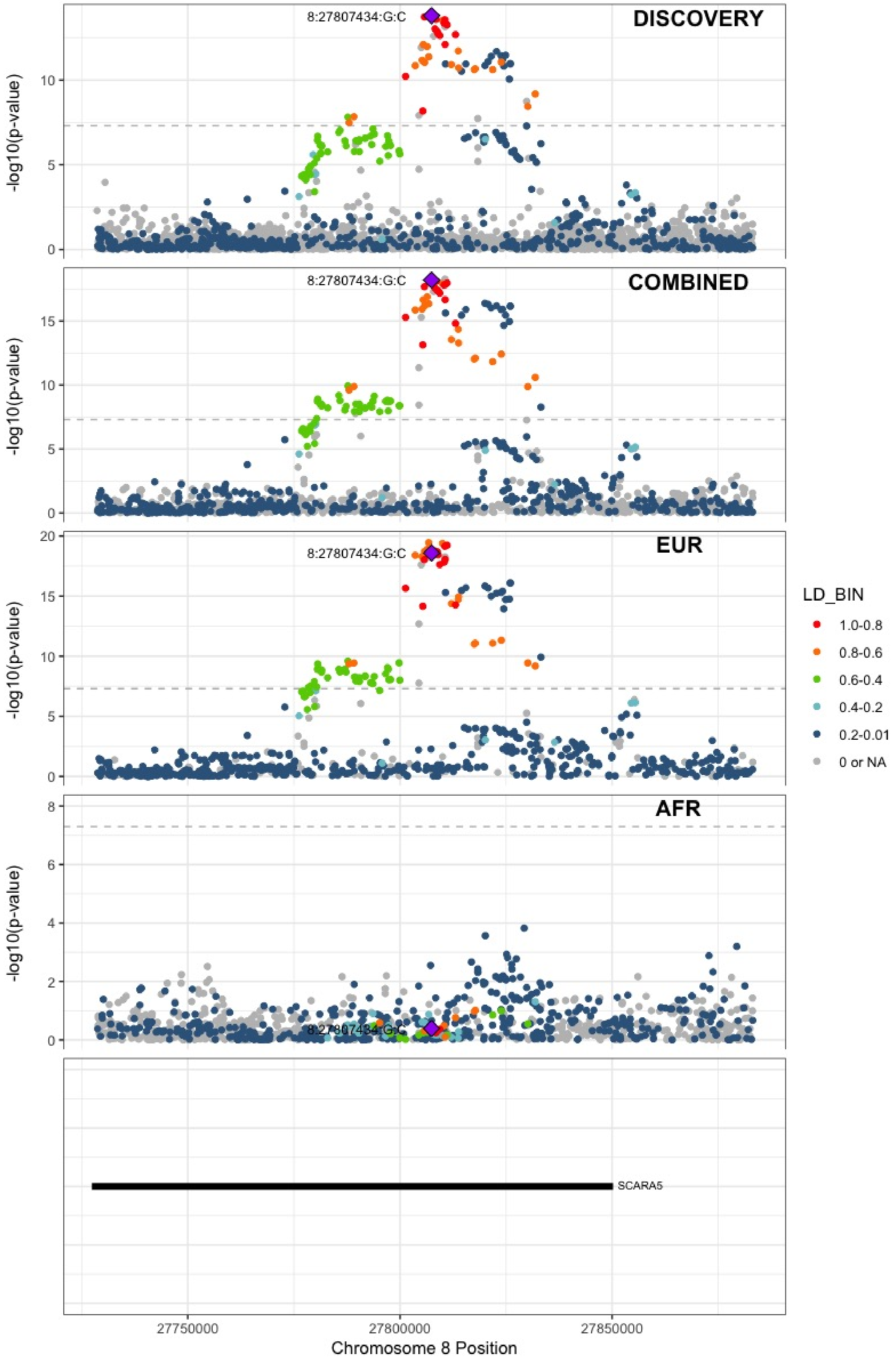

**Supplementary Figure 5.37** – rs6993770 (chr8:106581528) in locus 62 (ZFPM2)

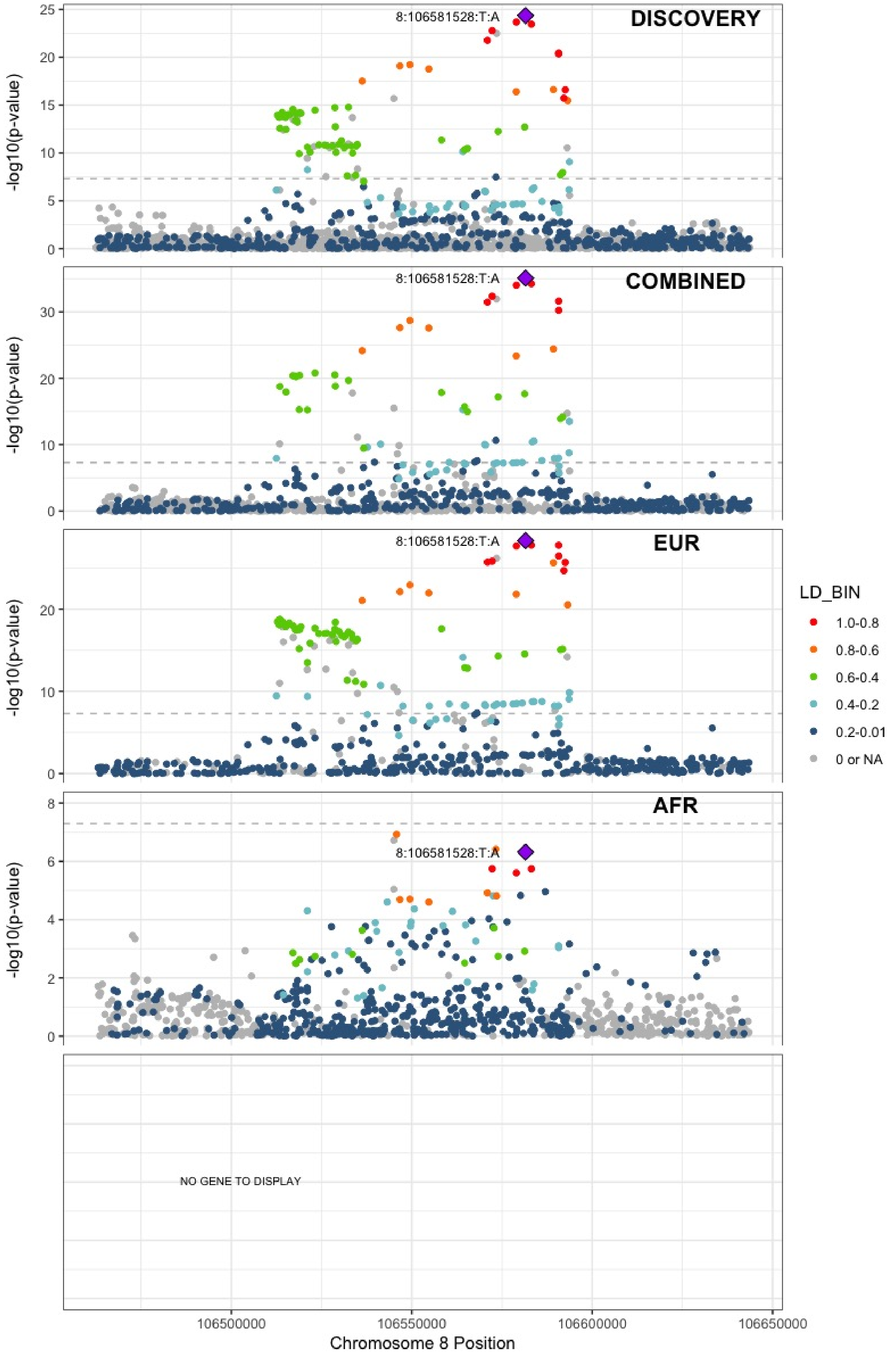

**Supplementary Figure 5.38** – rs77375493 (chr9:5073770) in locus 64 (JAK2)

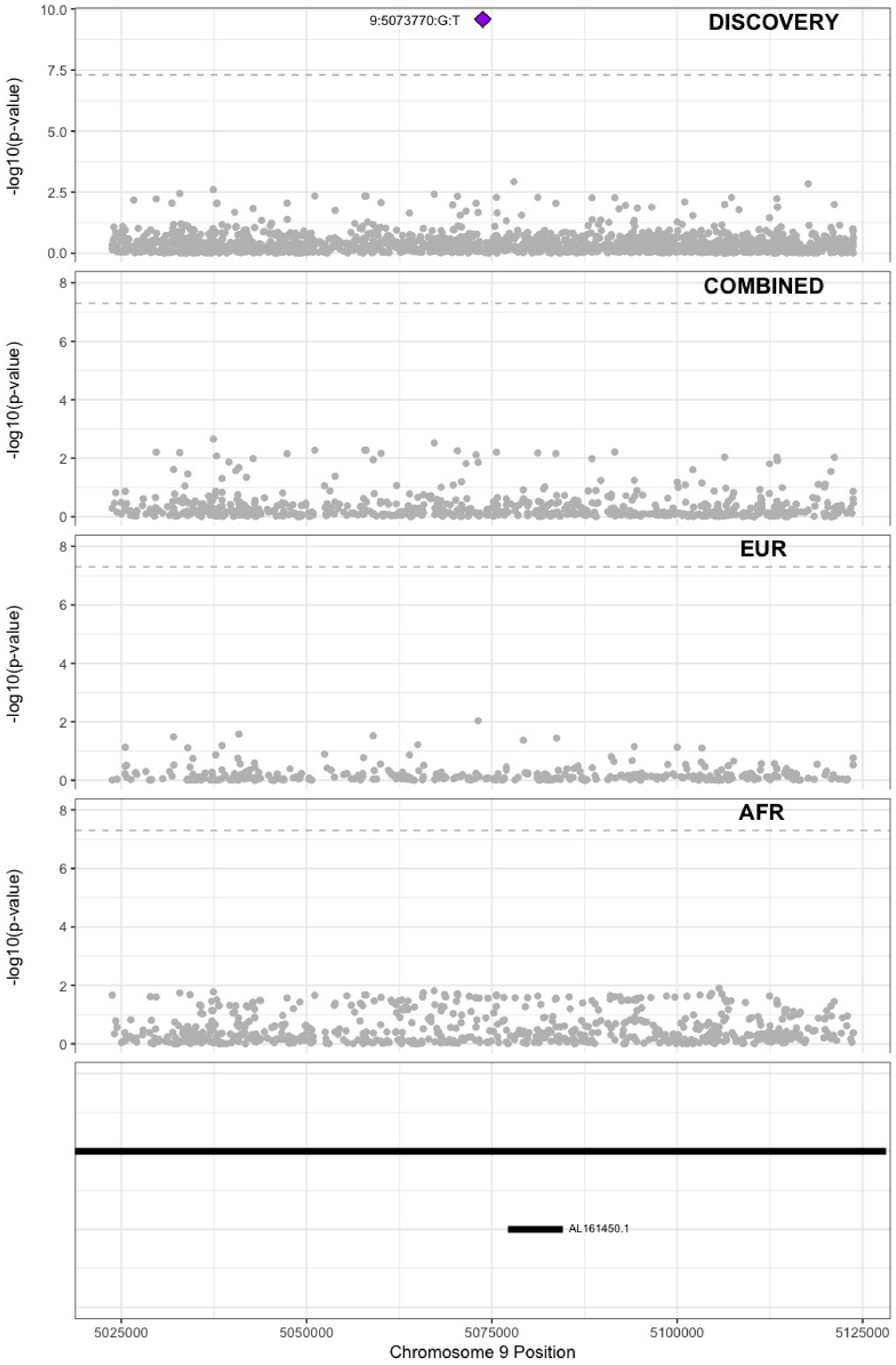

**Supplementary Figure 5.39** – rs35208412 (chr9:99194509) in locus 65 (ZNF367;HABP4)

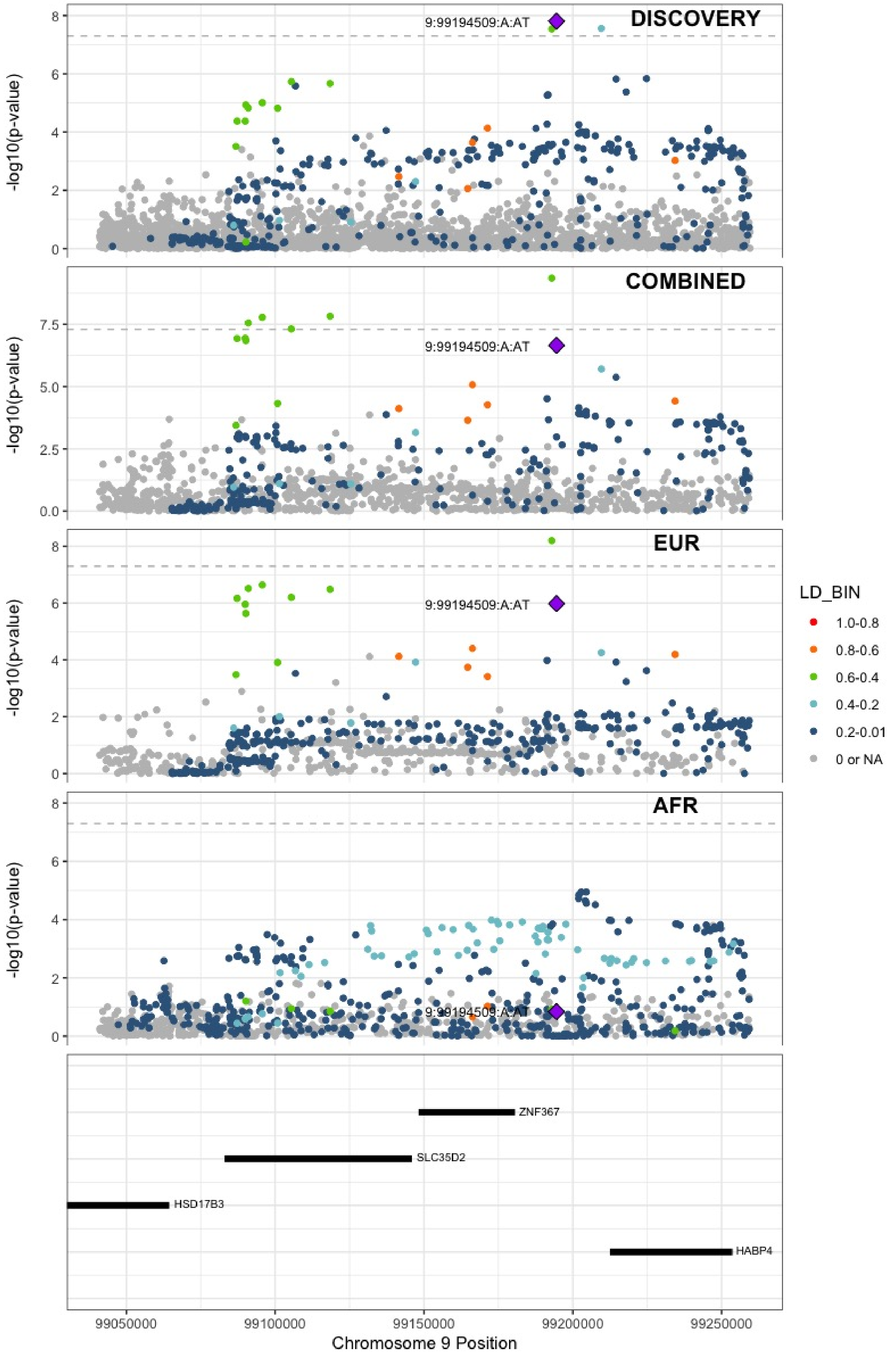

**Supplementary Figure 5.40** – rs505922 (chr9:136149229) in locus 66 (ABO)

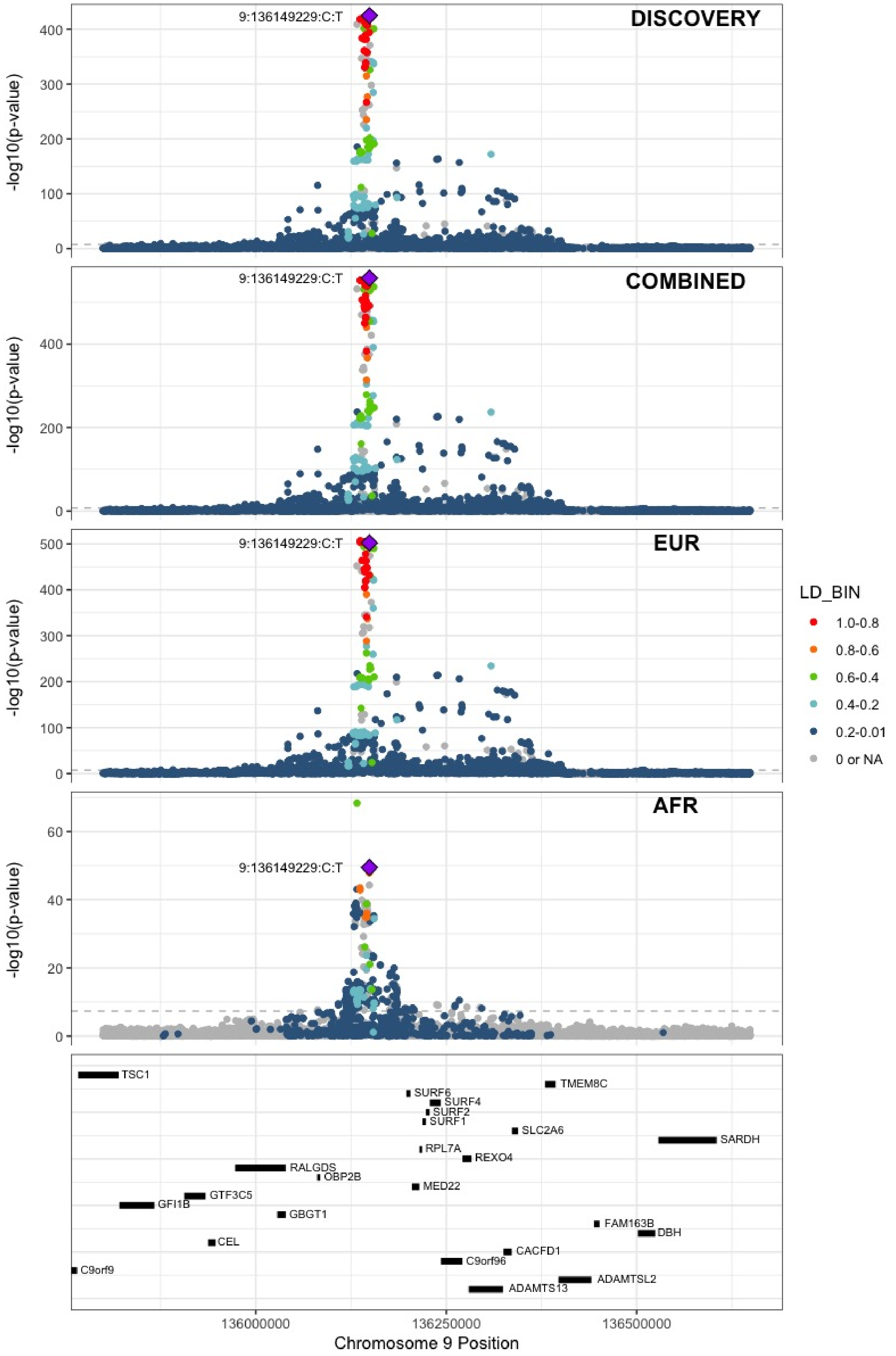

**Supplementary Figure 5.41** – rs1887091 (chr10:14535113) in locus 67 (MIR1265;FAM107B)

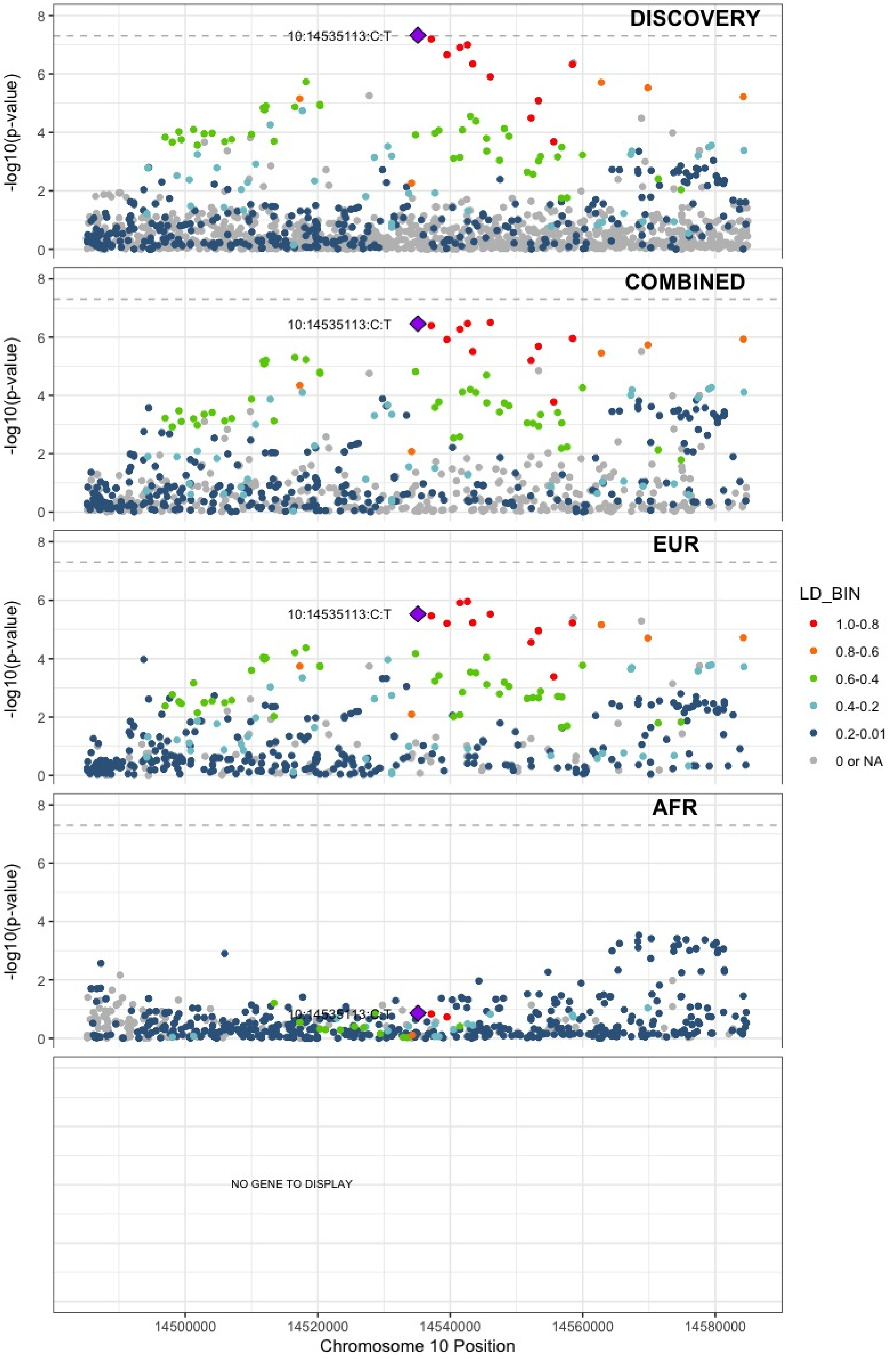

**Supplementary Figure 5.42** – rs529892067 (chr10:43568540) in locus 71 (MIR5100 ;RET)

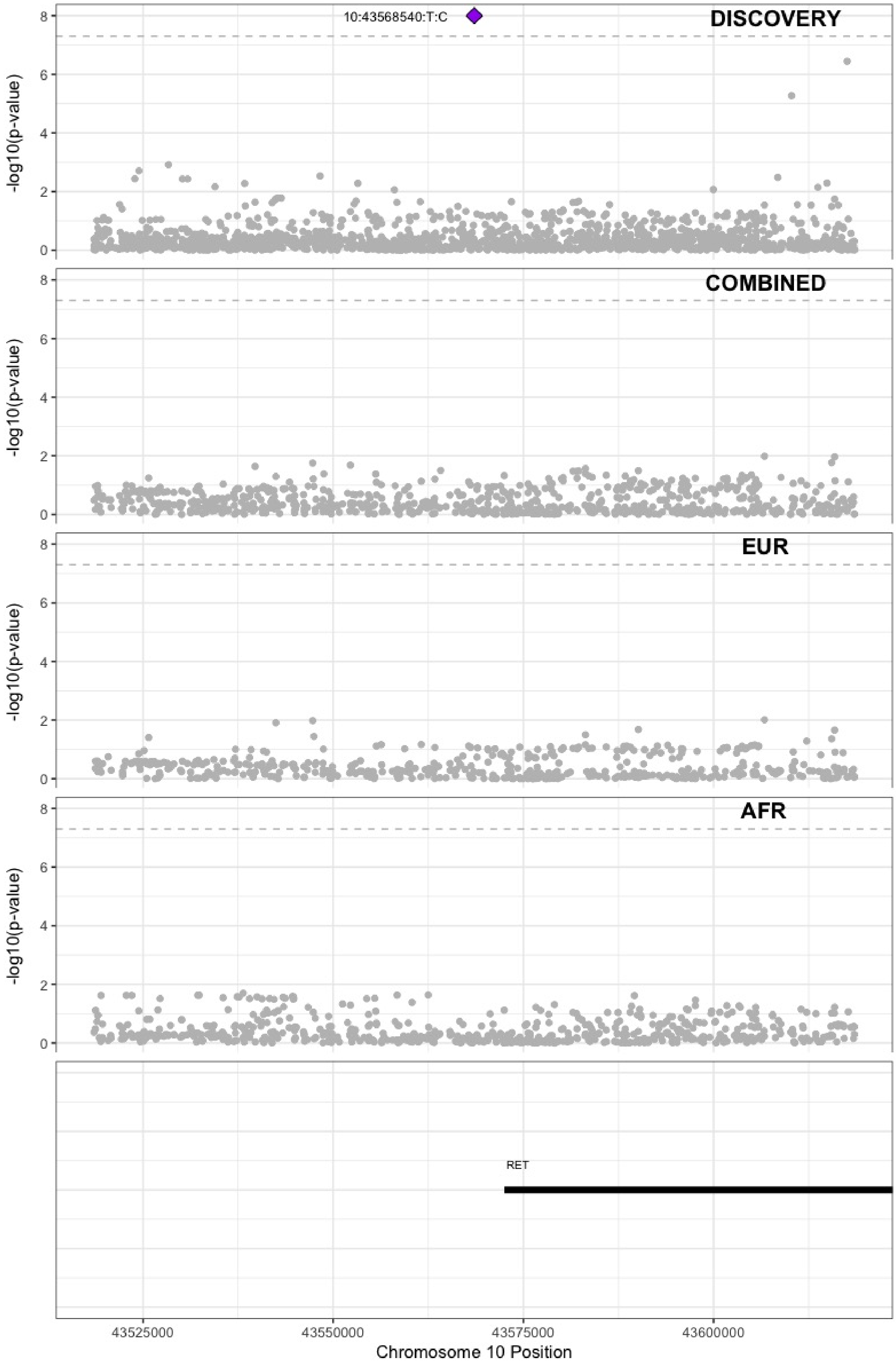

**Supplementary Figure 5.43** – rs17490626 (chr10:71218646) in locus 72 (TSPAN15)

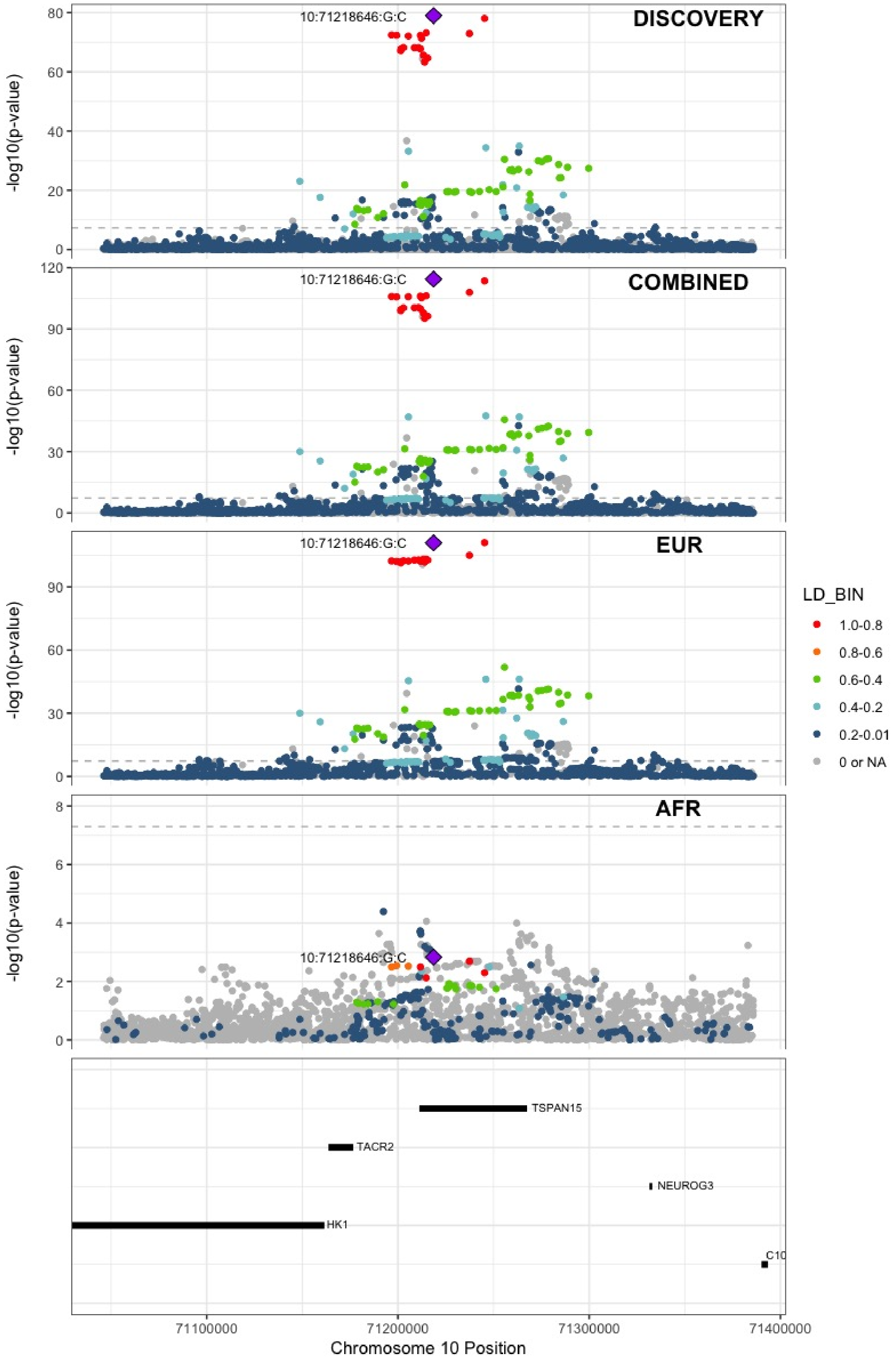

**Supplementary Figure 5.44** – rs16937003 (chr10:80938499) in locus 73 (ZMIZ1)

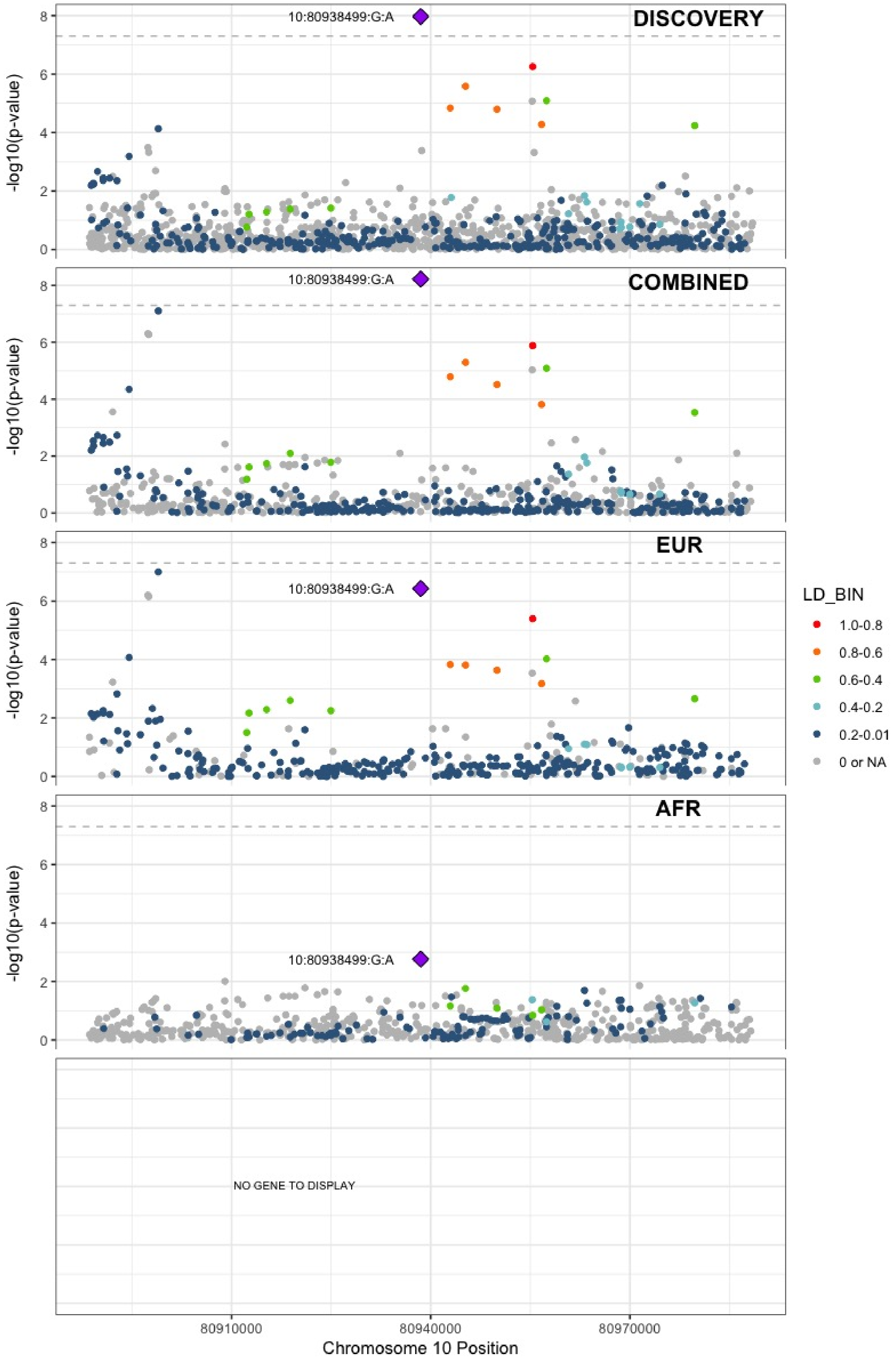

**Supplementary Figure 5.45** – rs2274224 (chr10:96039597) in locus 74 (PLCE1)

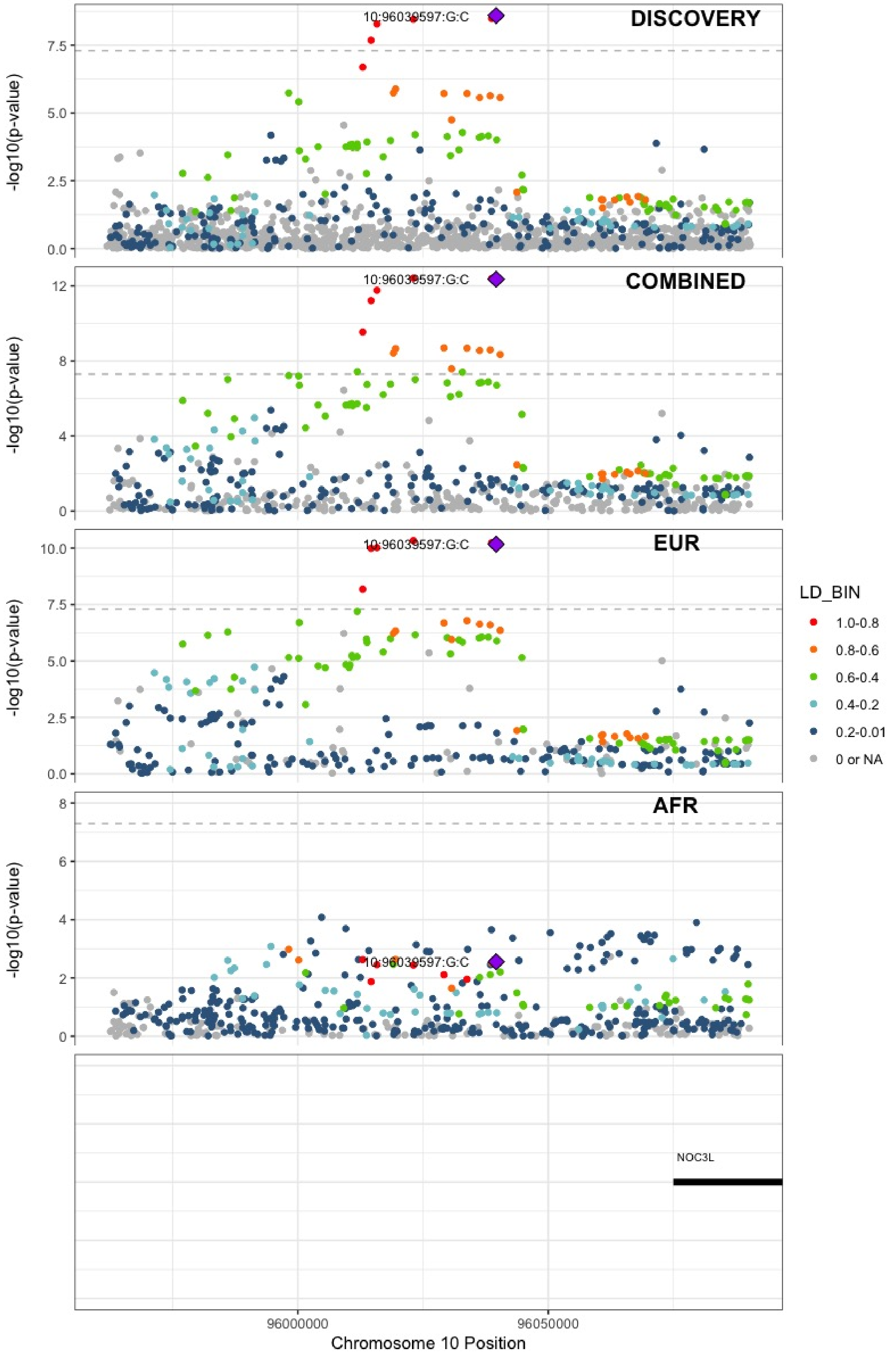

**Supplementary Figure 5.46** – rs10886430 (chr10:121010256) in locus 75 (GRK5)

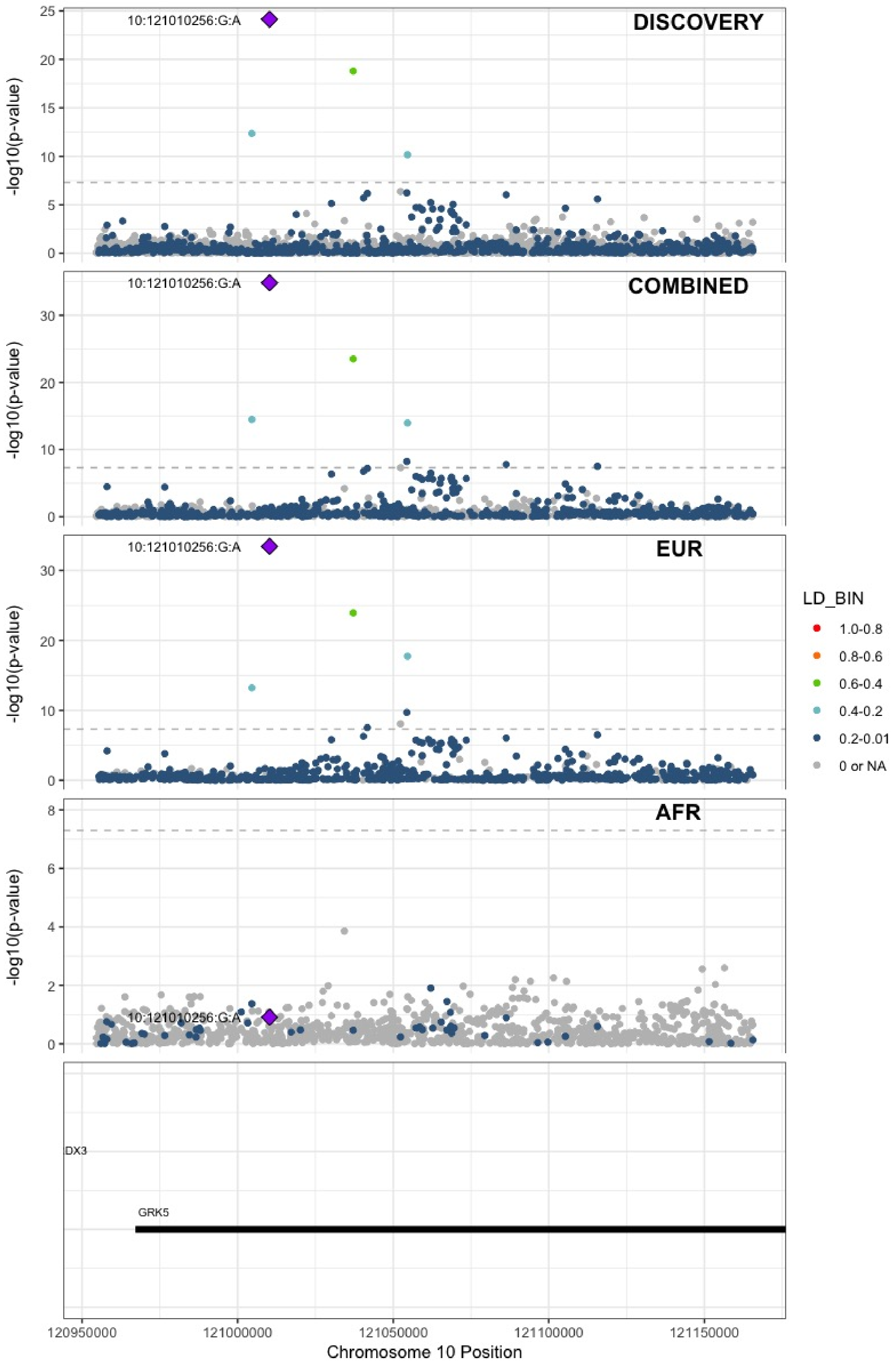

**Supplementary Figure 5.47** – rs11032074 (chr11:32993887) in locus 78 (QSER1)

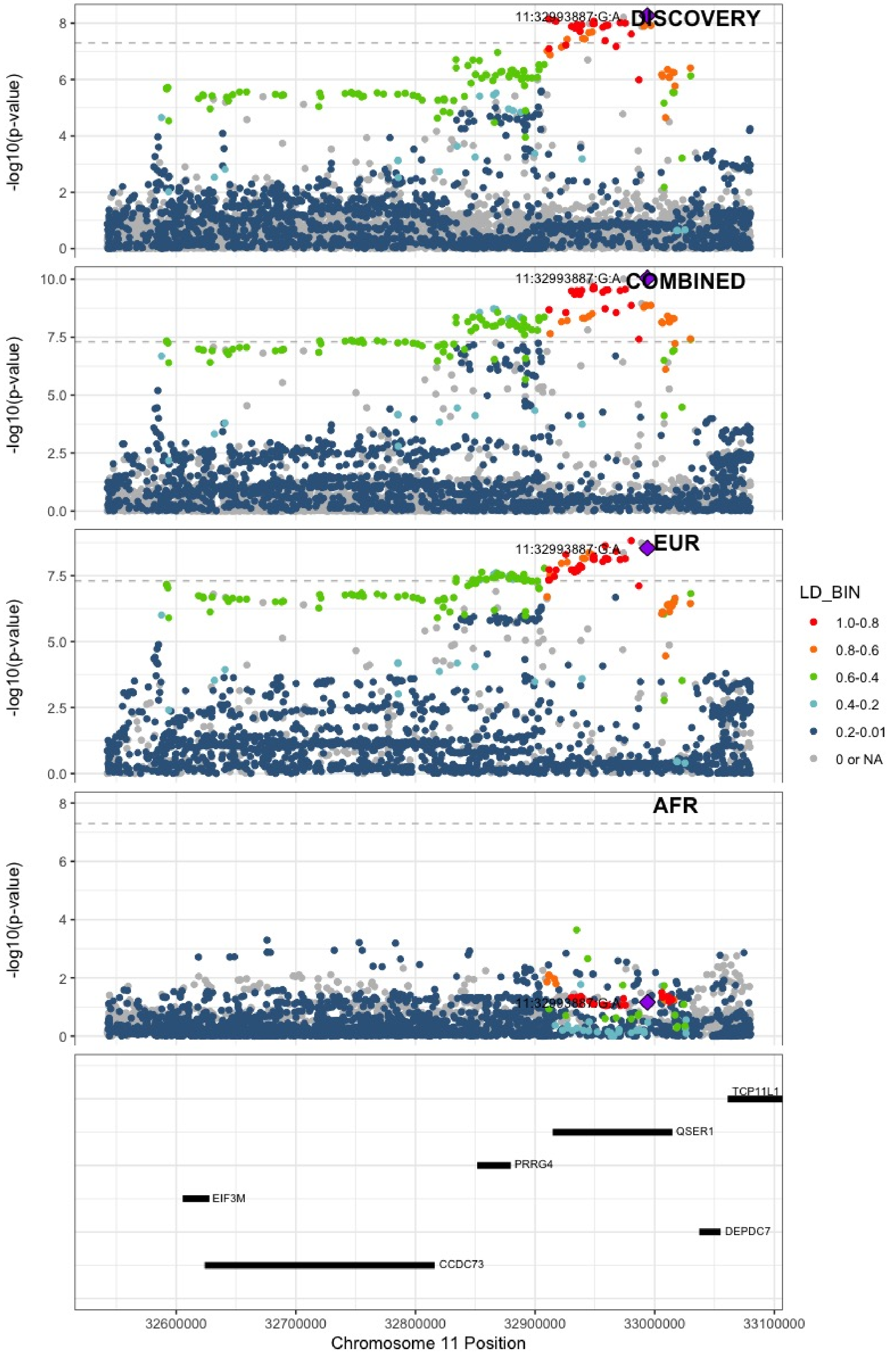

**Supplementary Figure 5.48** – rs1799963 (chr11:46761055) in locus 79 (F2)

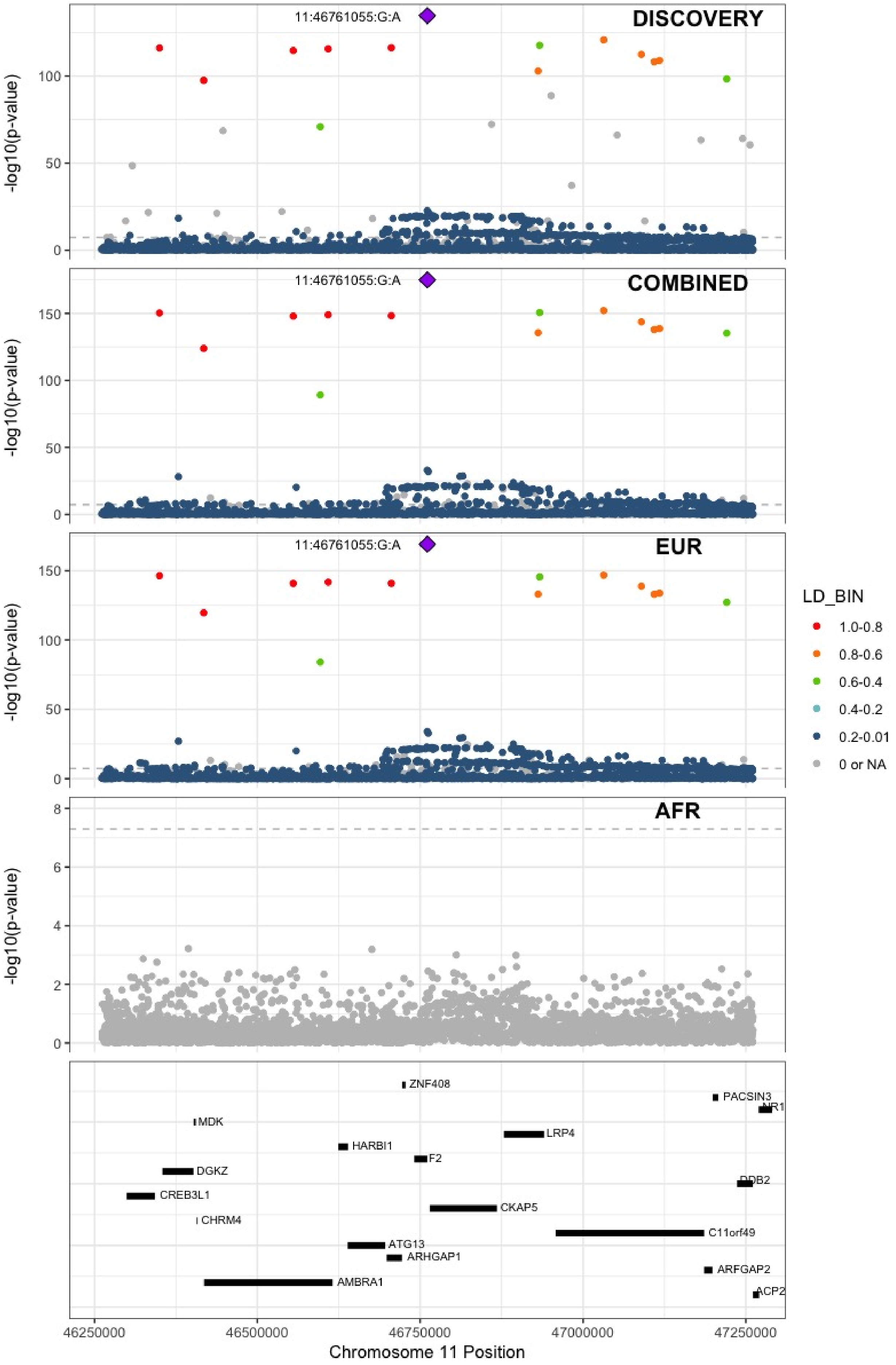

**Supplementary Figure 5.49** – rs141687379 (chr11:56666822) in locus 80 (FADS2B)

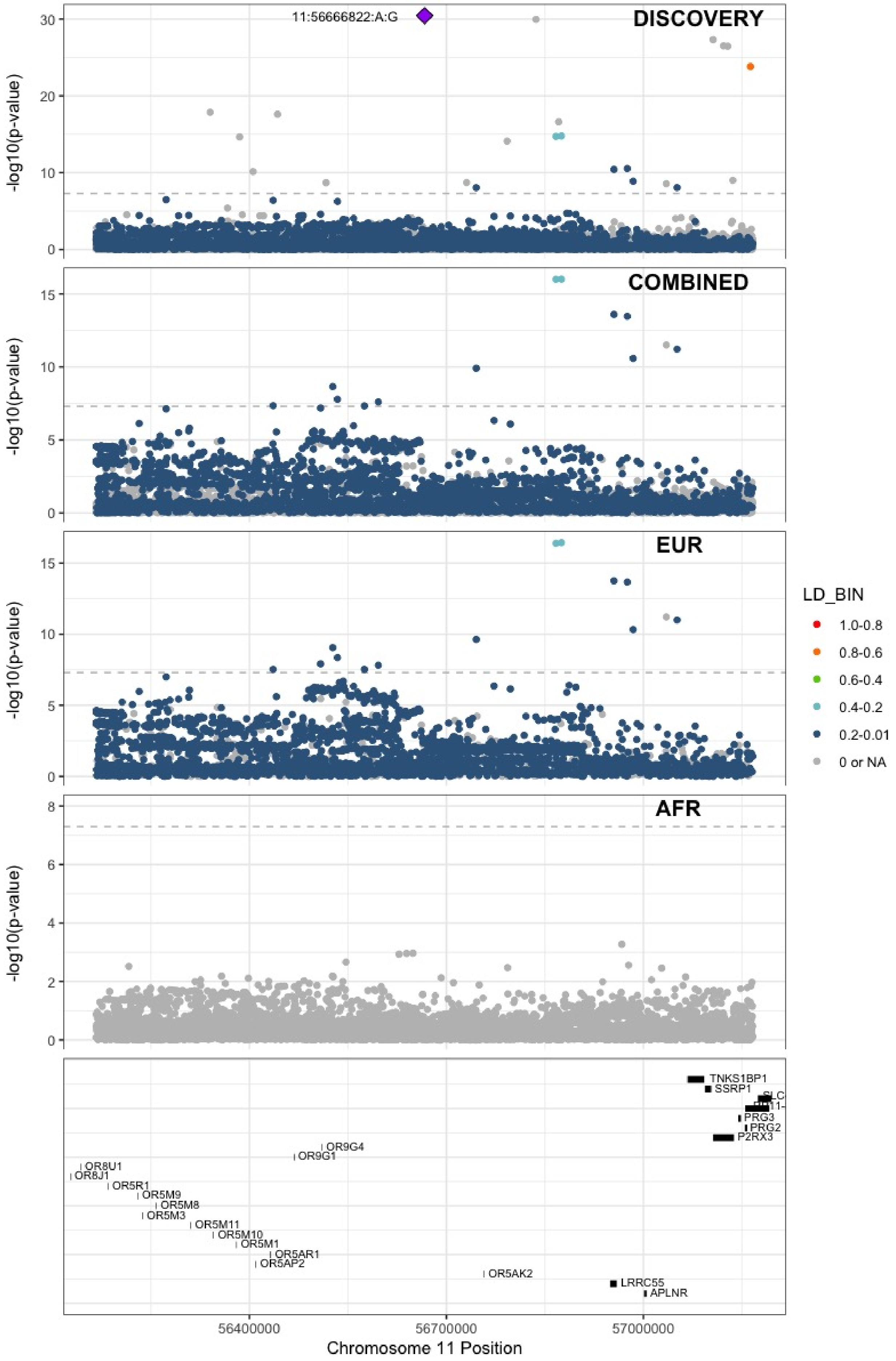

**Supplementary Figure 5.50** – rs174551 (chr11:61573684) in locus 82 (FADS1)

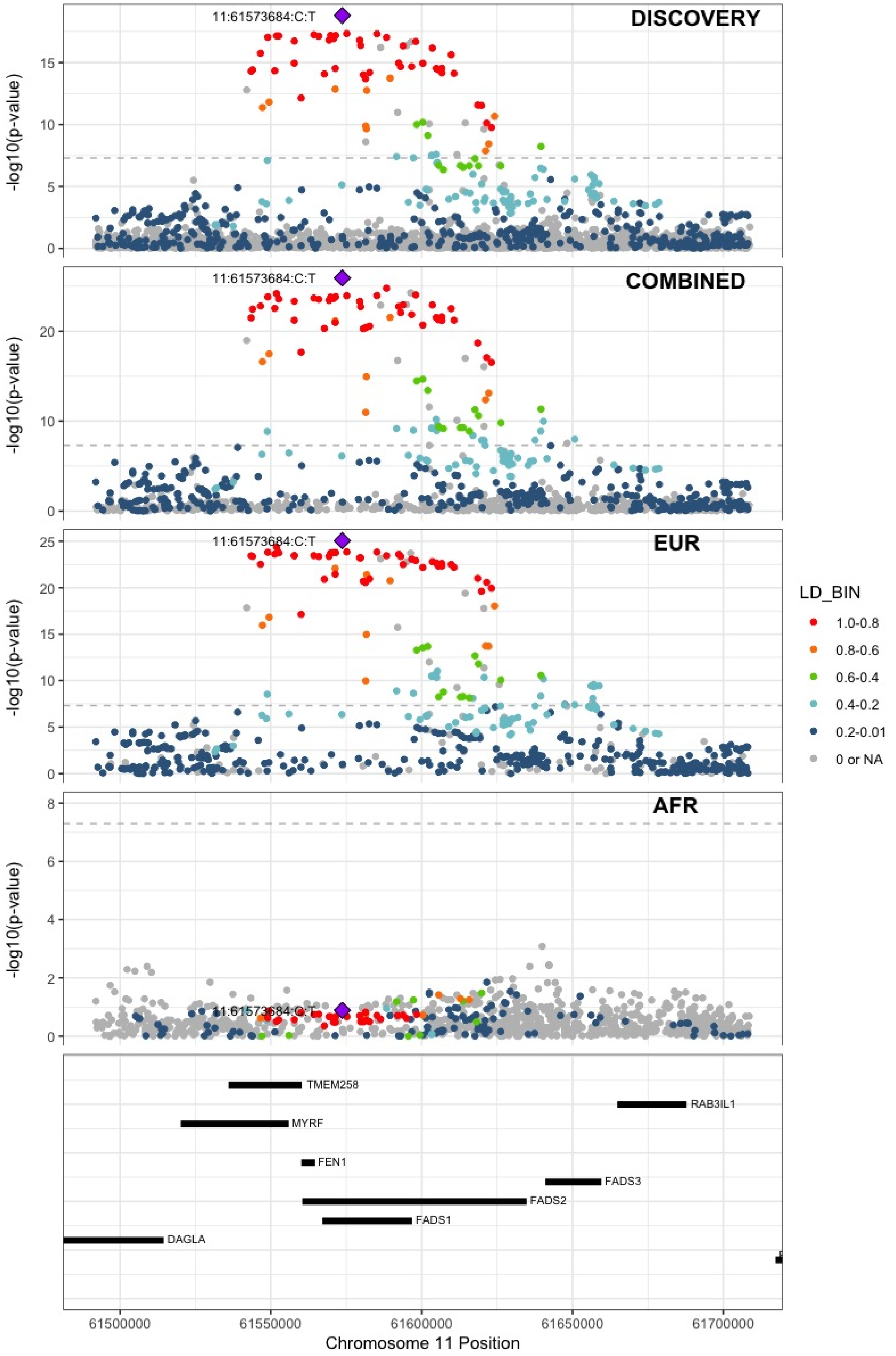

**Supplementary Figure 5.51** – rs35257264 (chr11:126296816) in locus 85 (ST3GAL4)

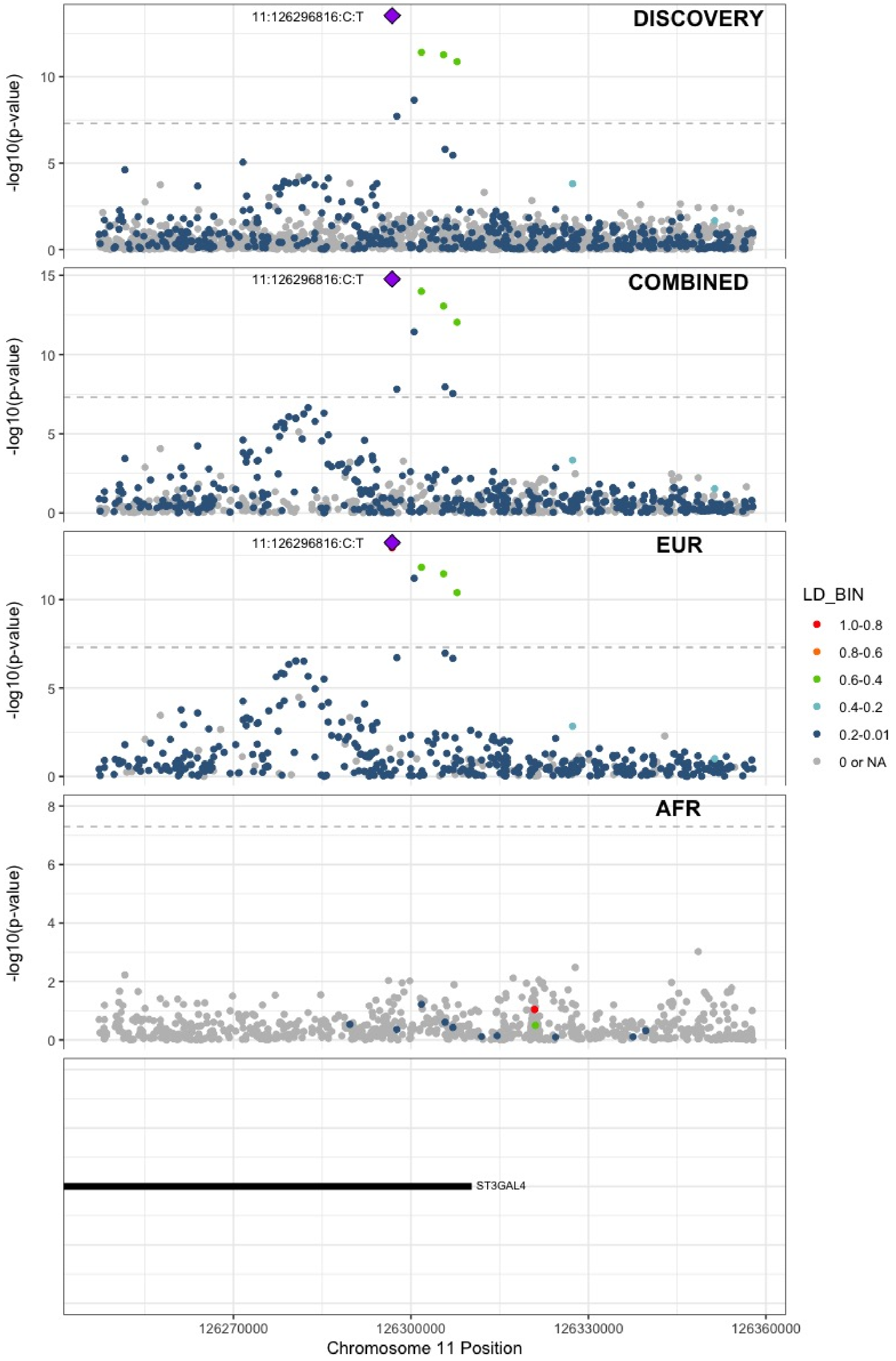

**Supplementary Figure 5.52** – rs1558519 (chr12:6153738) in locus 87 (VWF)

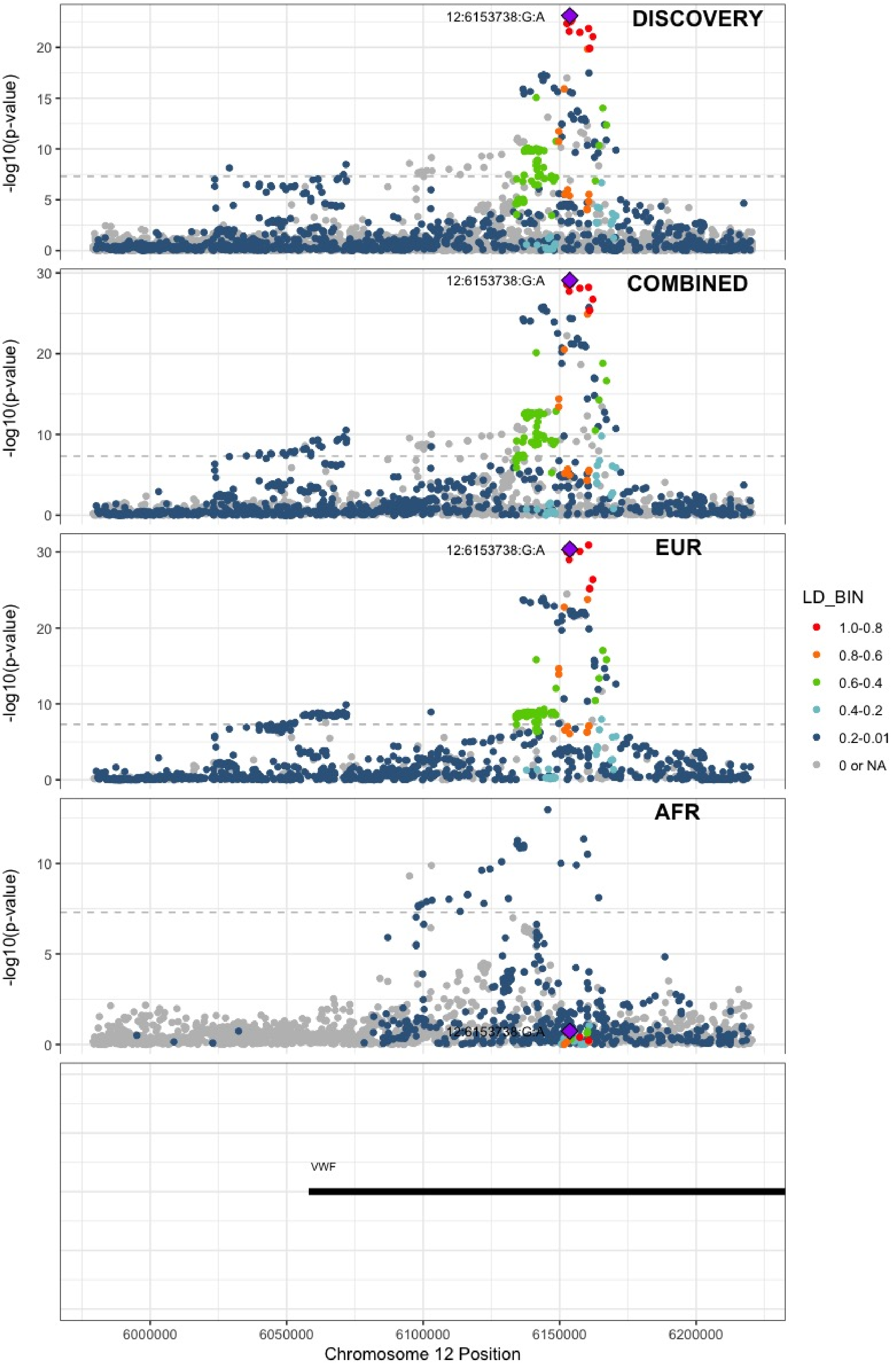

**Supplementary Figure 5.53** – rs7311483 (chr12:9053661) in locus 88 (A2ML1 ;PHC1)

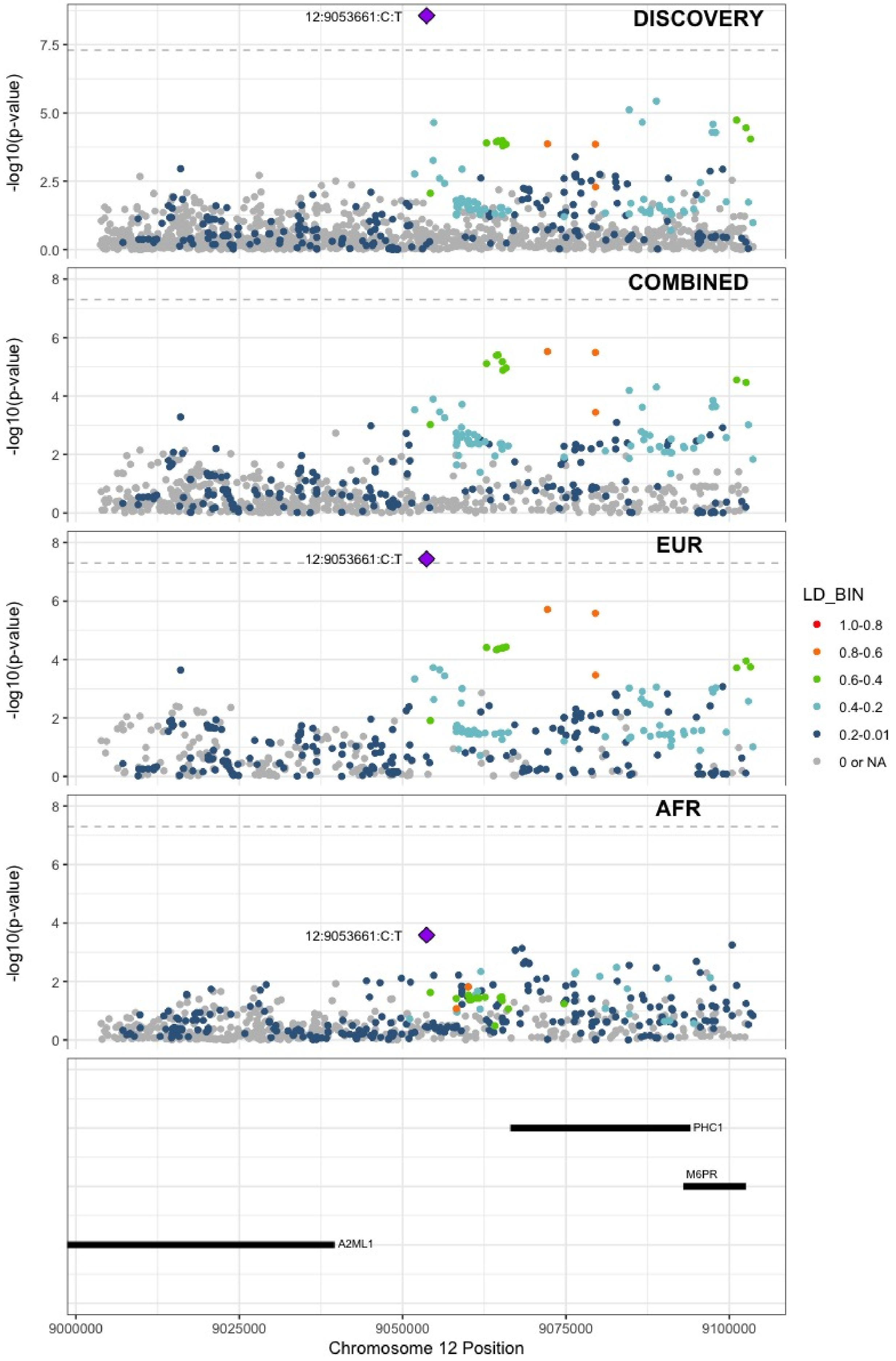

**Supplementary Figure 5.54** – rs6580981 (chr12:54723028) in locus 89 (COPZ1)

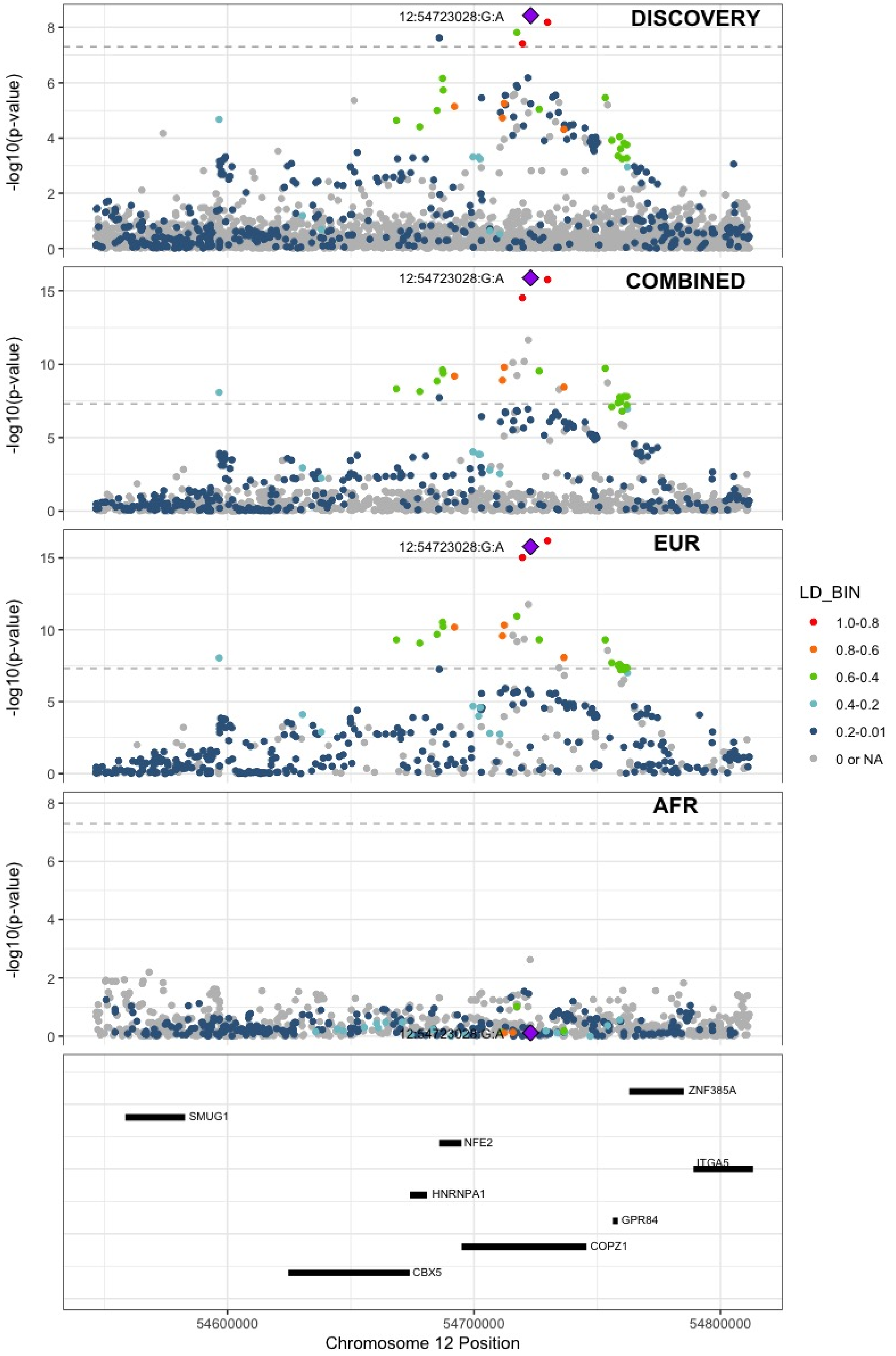

**Supplementary Figure 5.55** – rs142351376 (chr12:104136288) in locus 90 (STAB2)

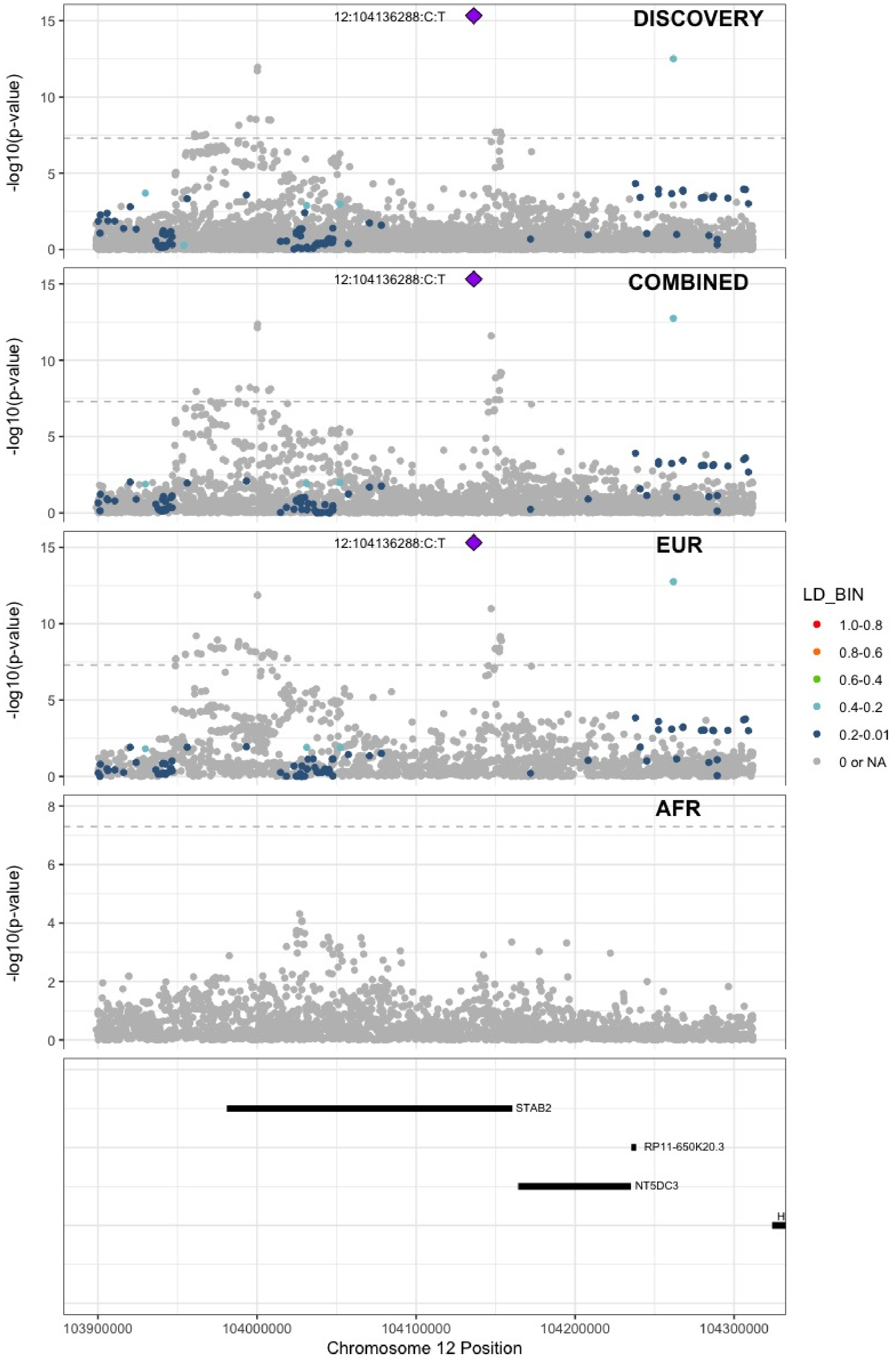

**Supplementary Figure 5.56** – rs3184504 (chr12:111884608) in locus 91 (SH2B3)

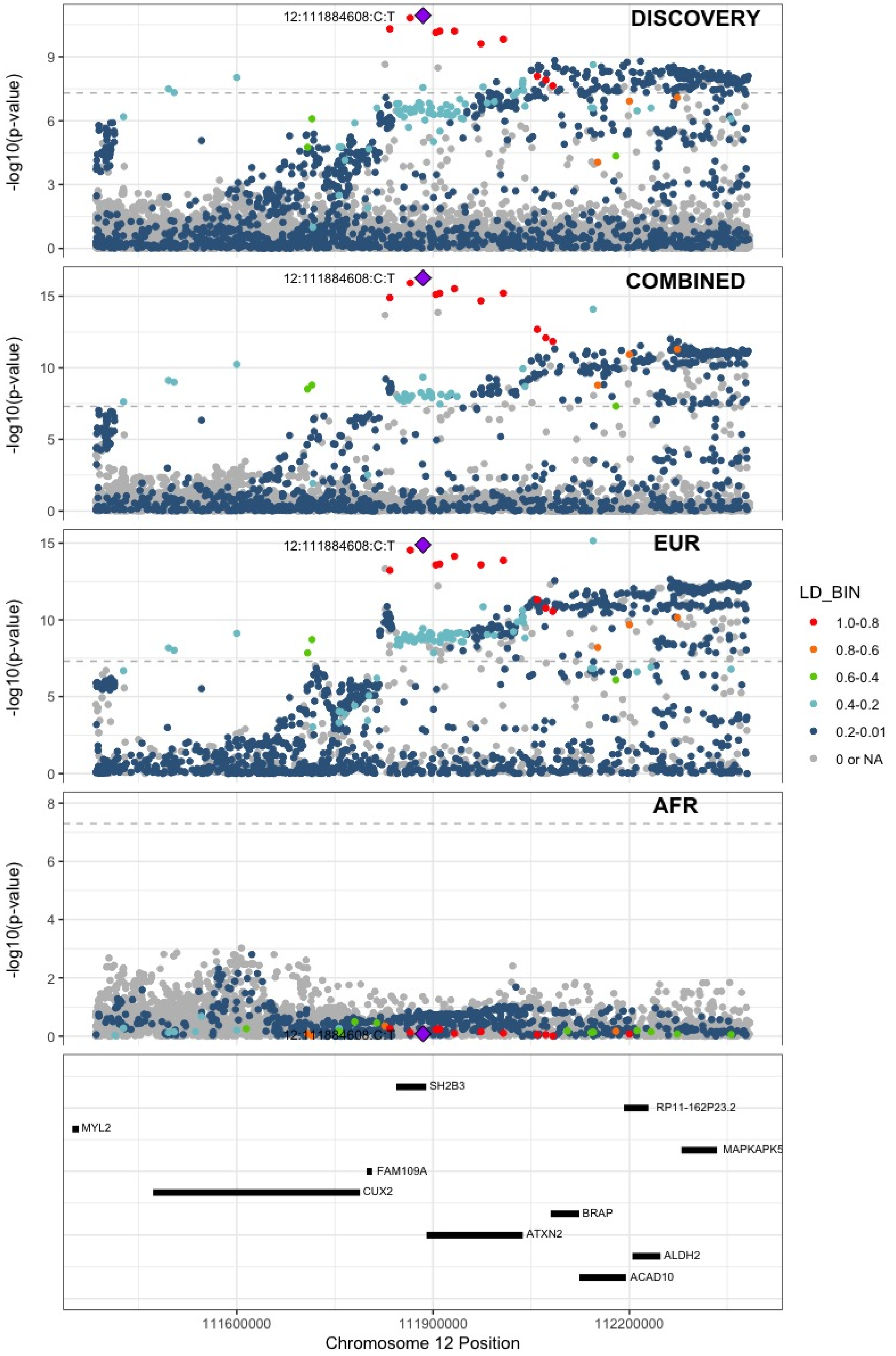

**Supplementary Figure 5.57** – rs75940120 (chr13:109686619) in locus 93 (MYO16)

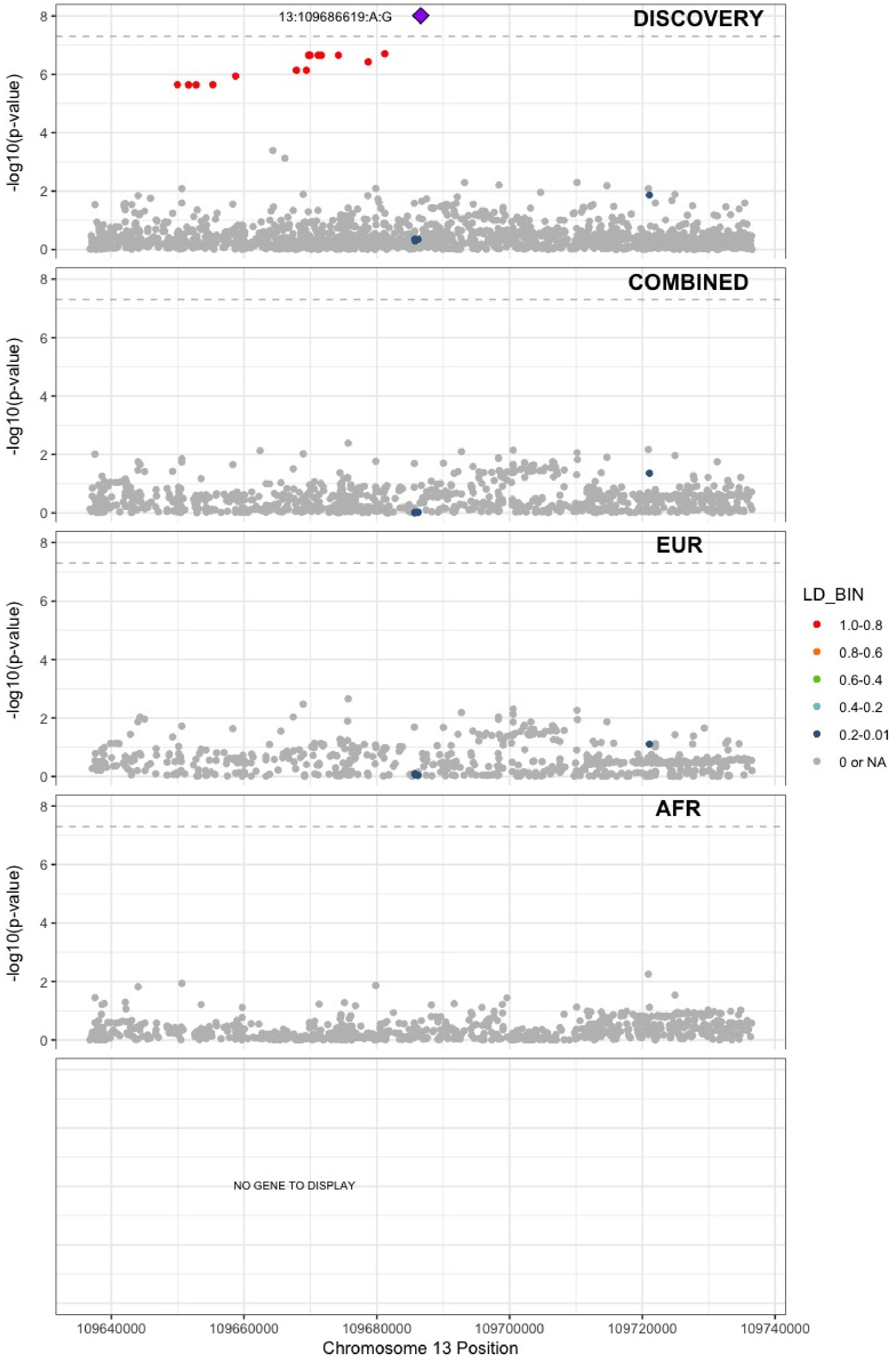

**Supplementary Figure 5.58** – rs3211752 (chr13:113787459) in locus 94 (F10)

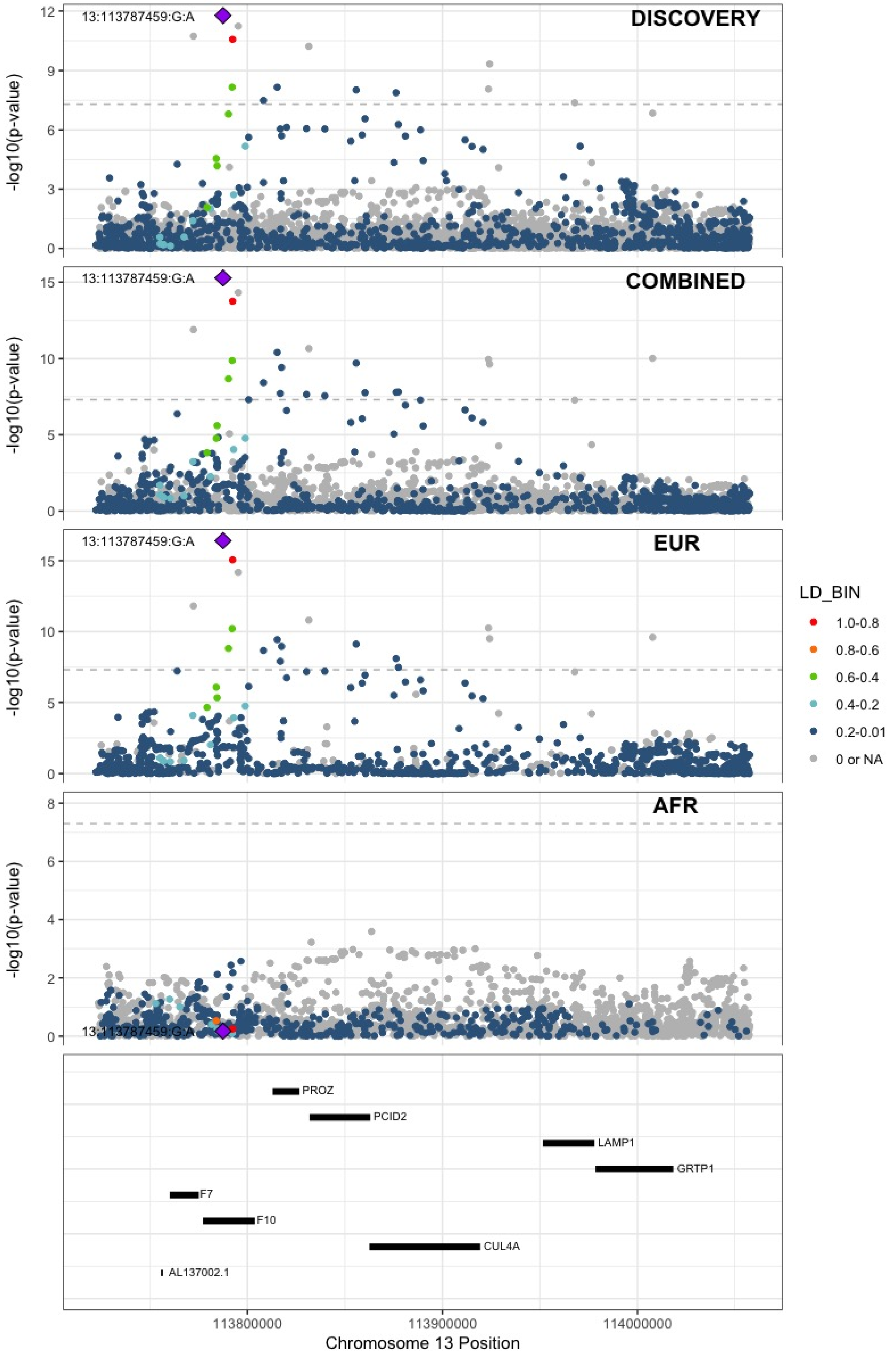

**Supplementary Figure 5.59** – rs57035593 (chr14:92268096) in locus 97 (TC2N)

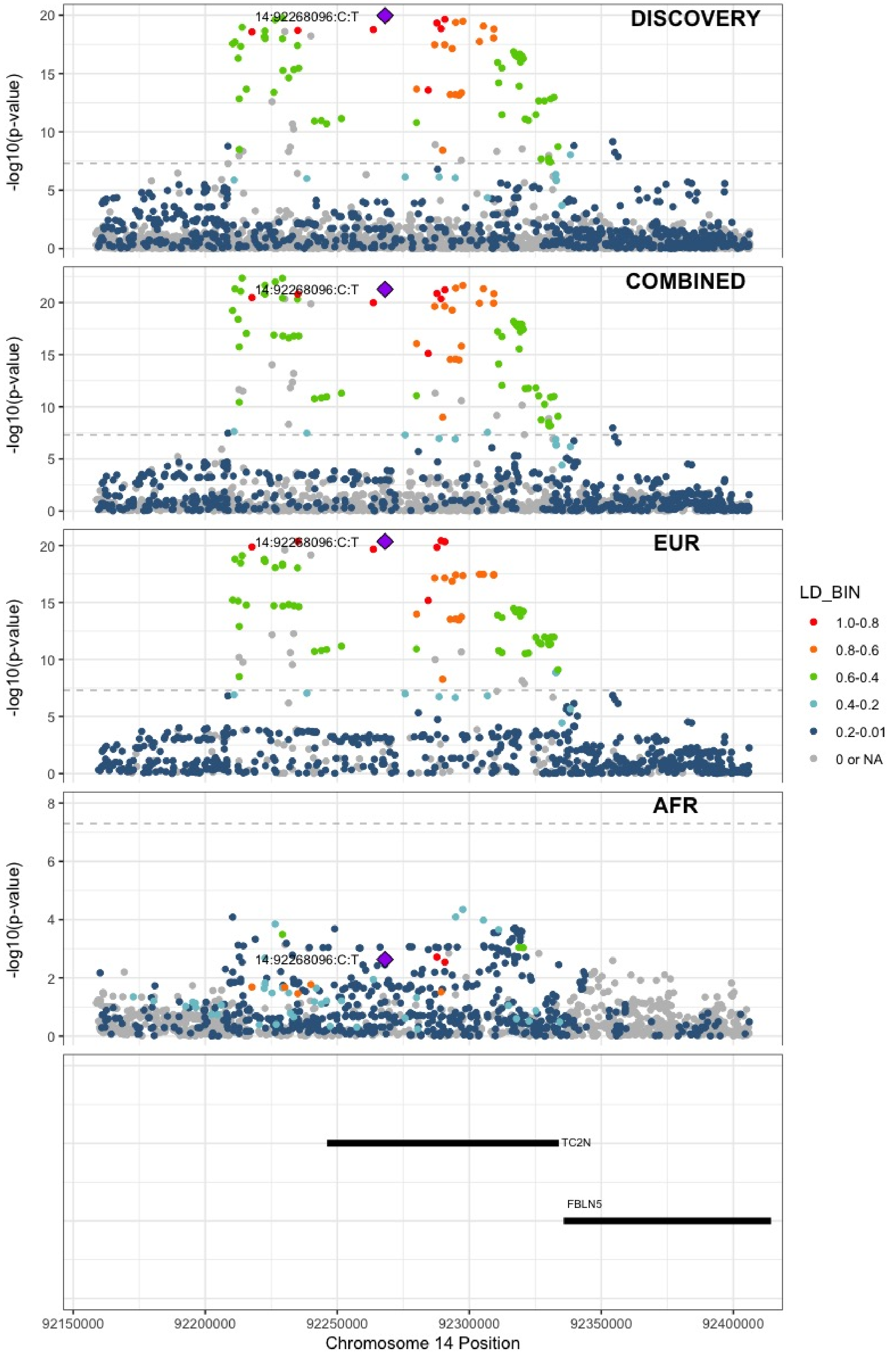

**Supplementary Figure 5.60** – rs8013957 (chr14:103140254) in locus 99 (RCOR1)

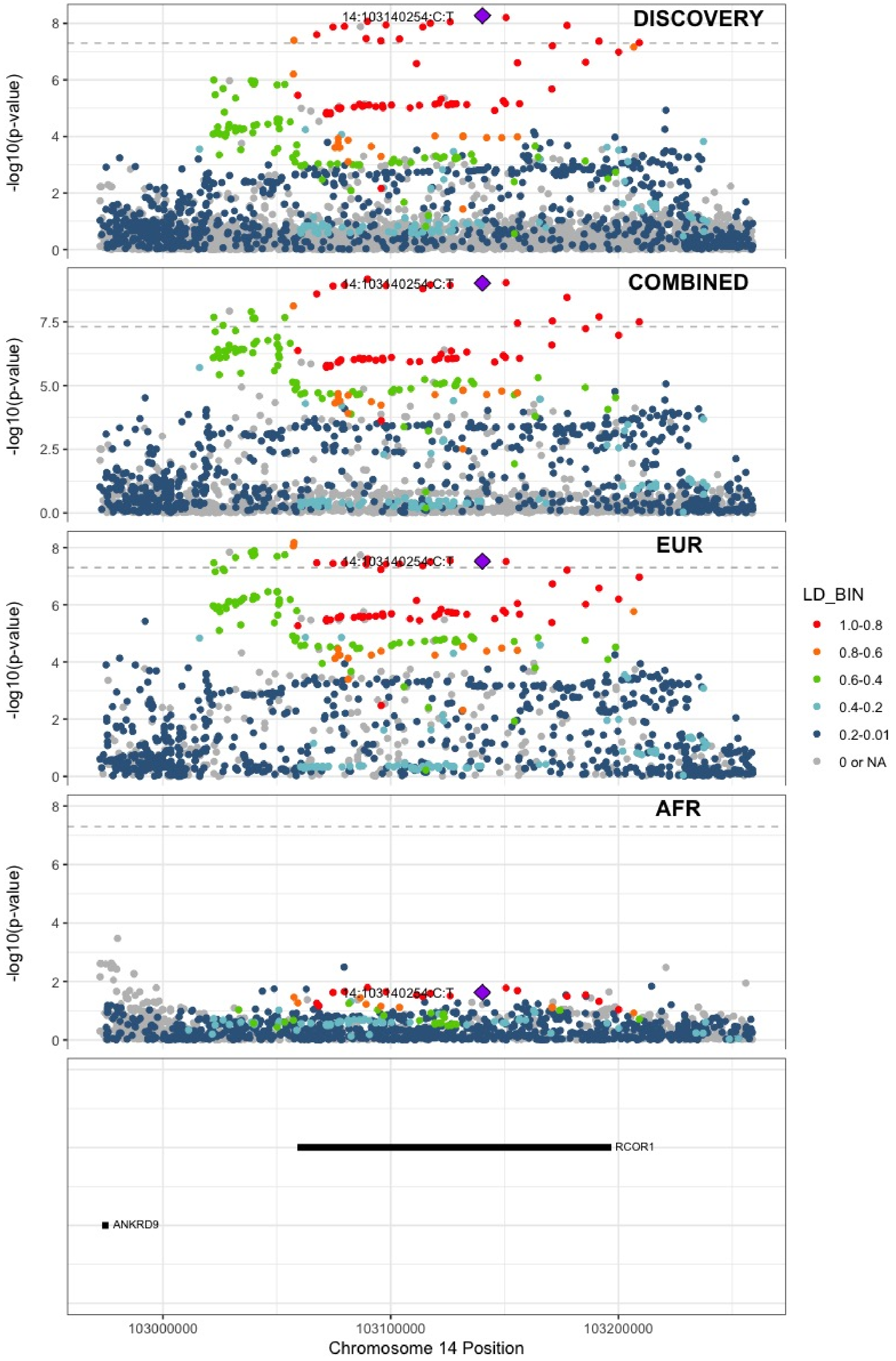

**Supplementary Figure 5.61** – rs55707100 (chr15:43820717) in locus 100 (MAP1A)

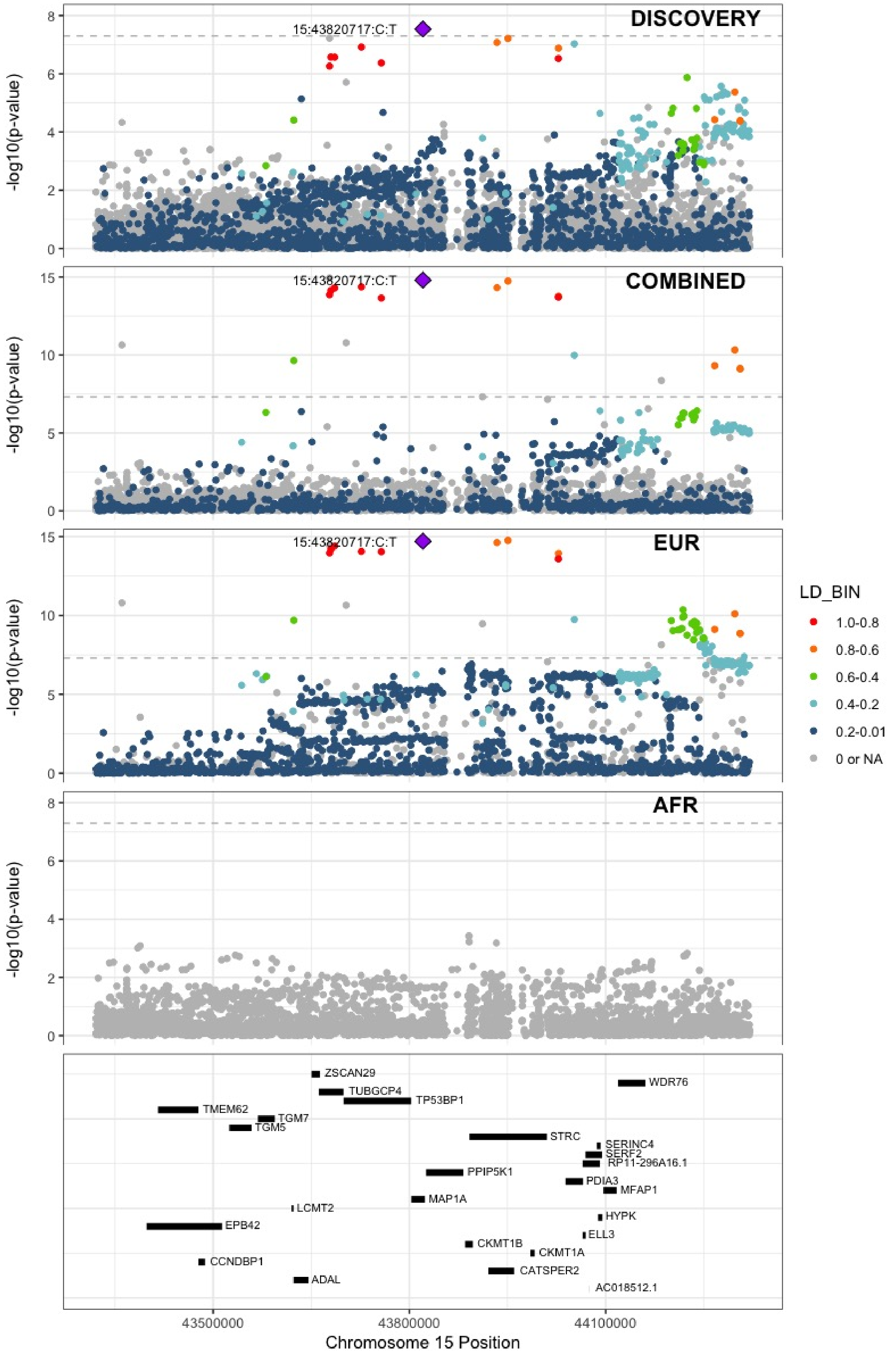

**Supplementary Figure 5.62** – rs59442804 (chr15:60899031) in locus 101 (RORA-AS1)

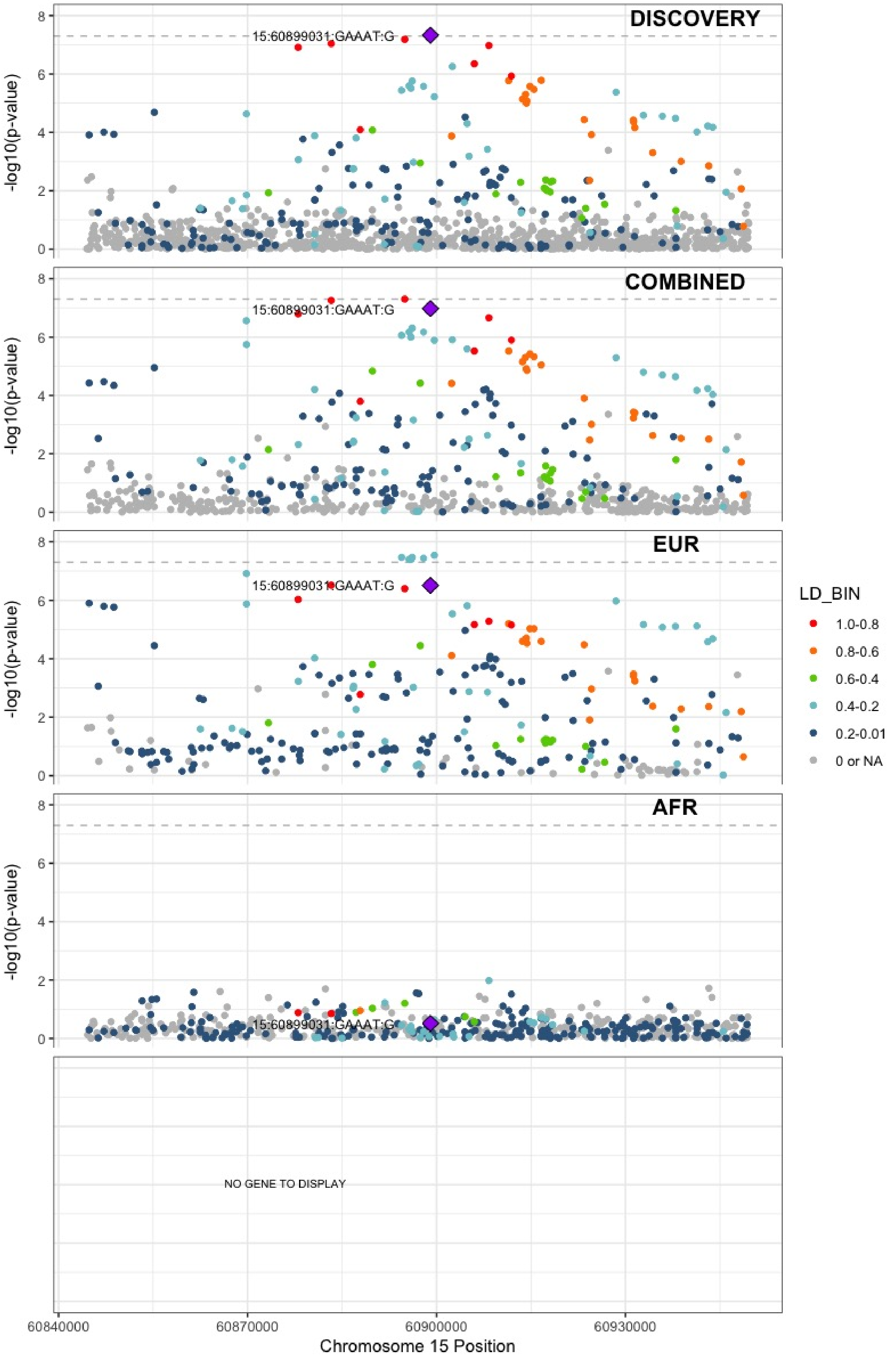

**Supplementary Figure 5.63** – rs182906510 (chr15:67372922) in locus 102 (SMAD3)

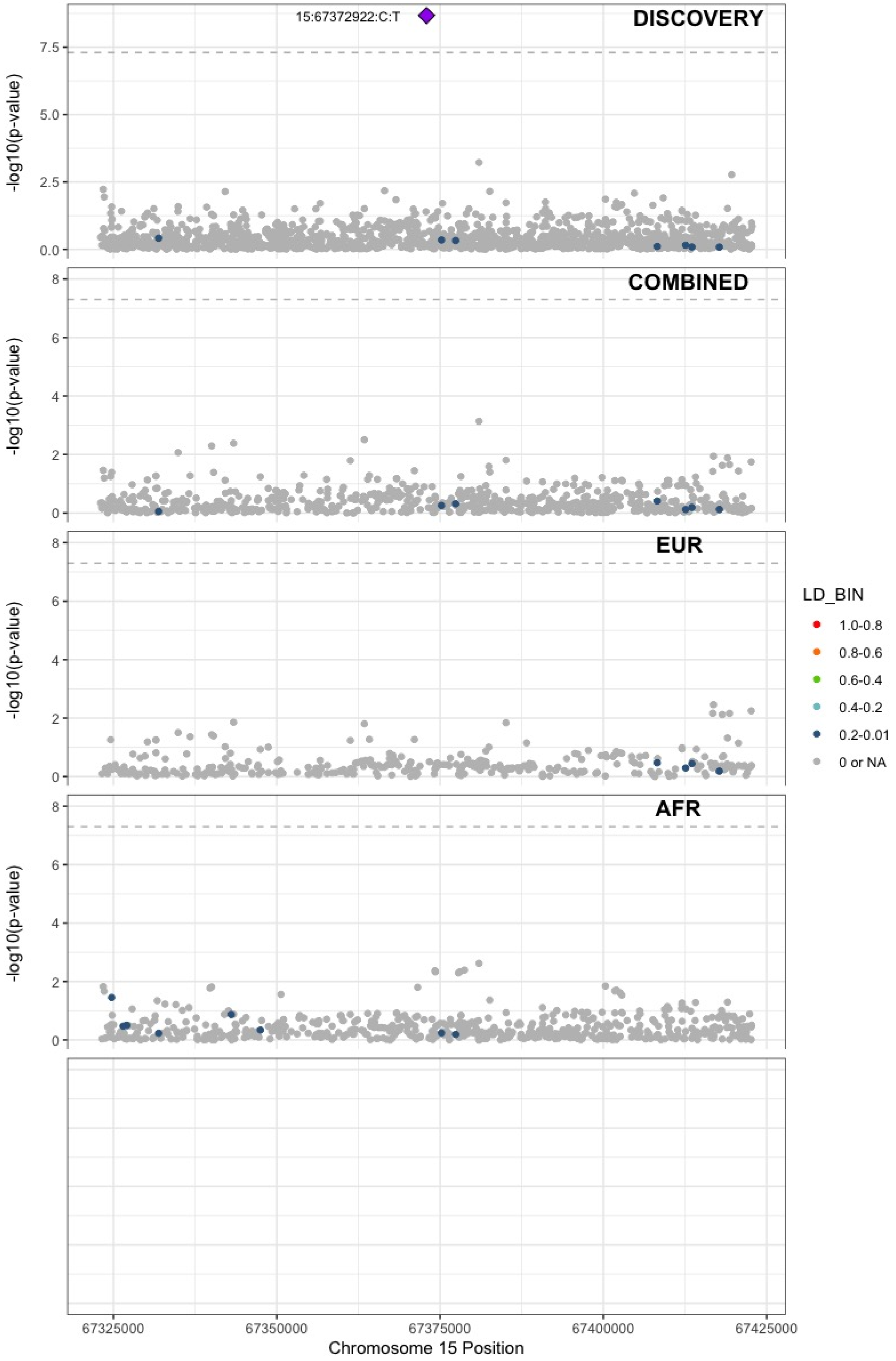

**Supplementary Figure 5.64** – rs12443808 (chr16:30996871) in locus 106 (HSD3B7)

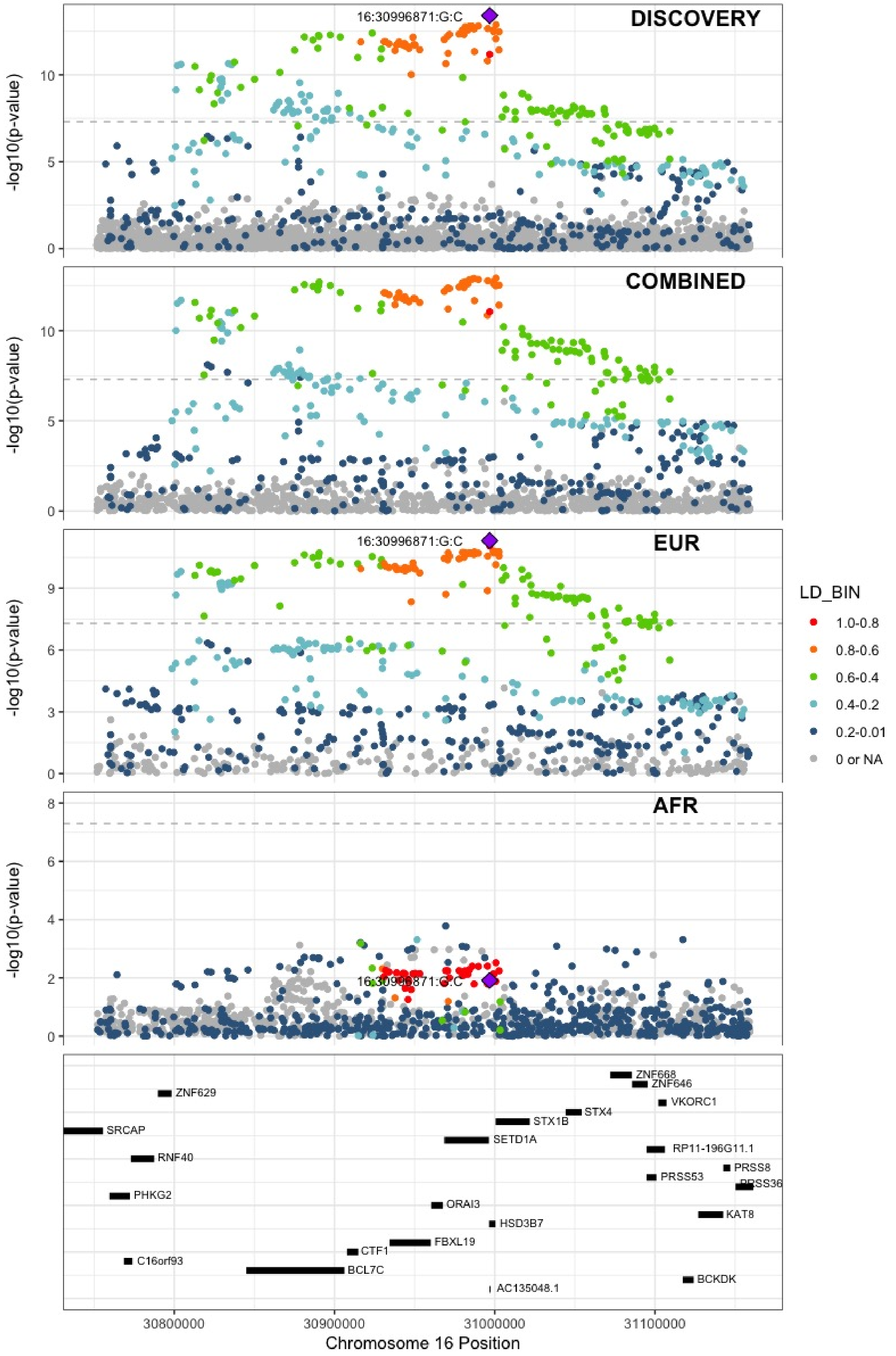

**Supplementary Figure 5.65** – rs56943275 (chr16:81898152) in locus 109 (PLCG2)

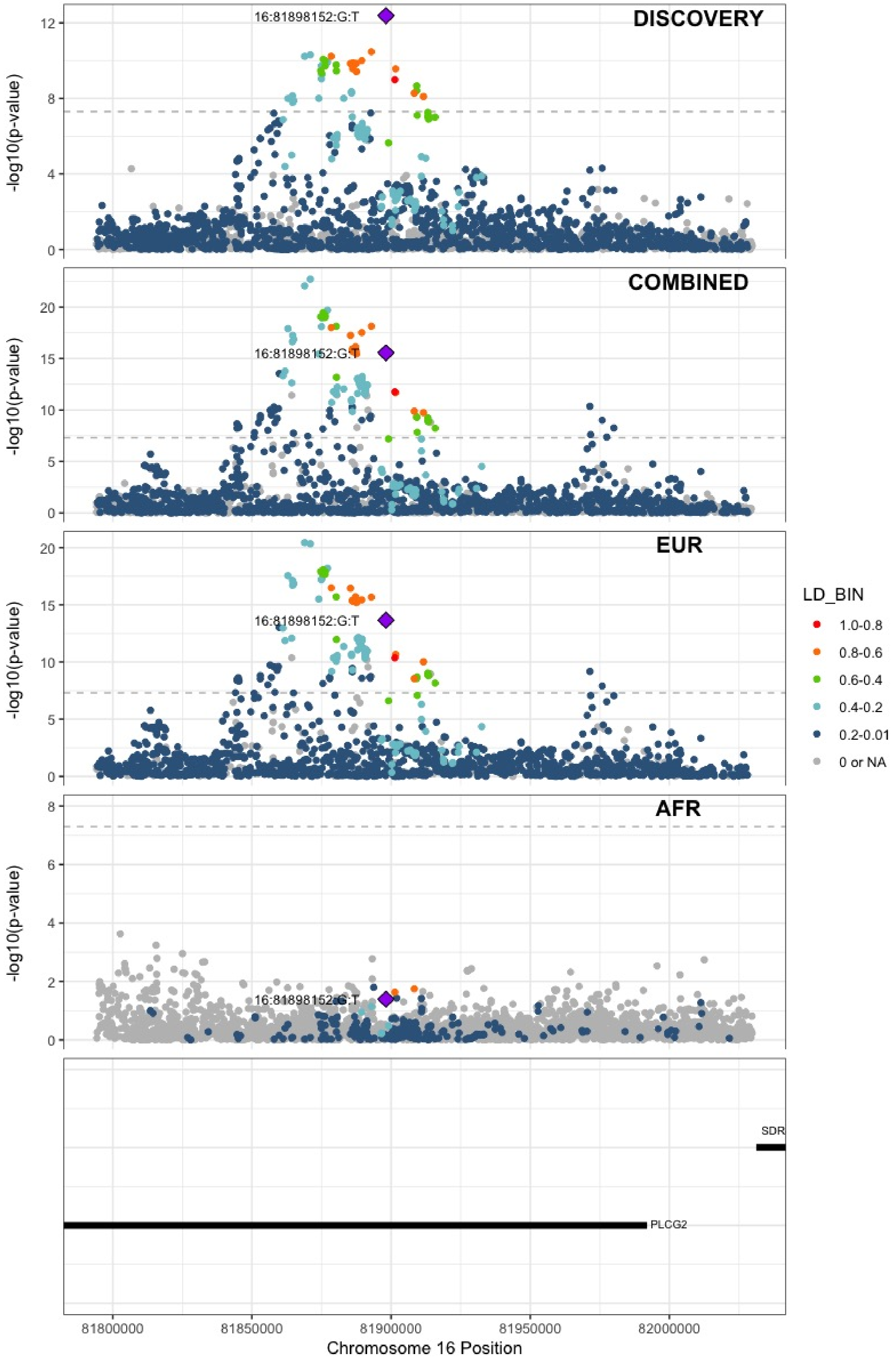

**Supplementary Figure 5.66** – rs28634651 (chr16:88553198) in locus 111 (ZFPM1)

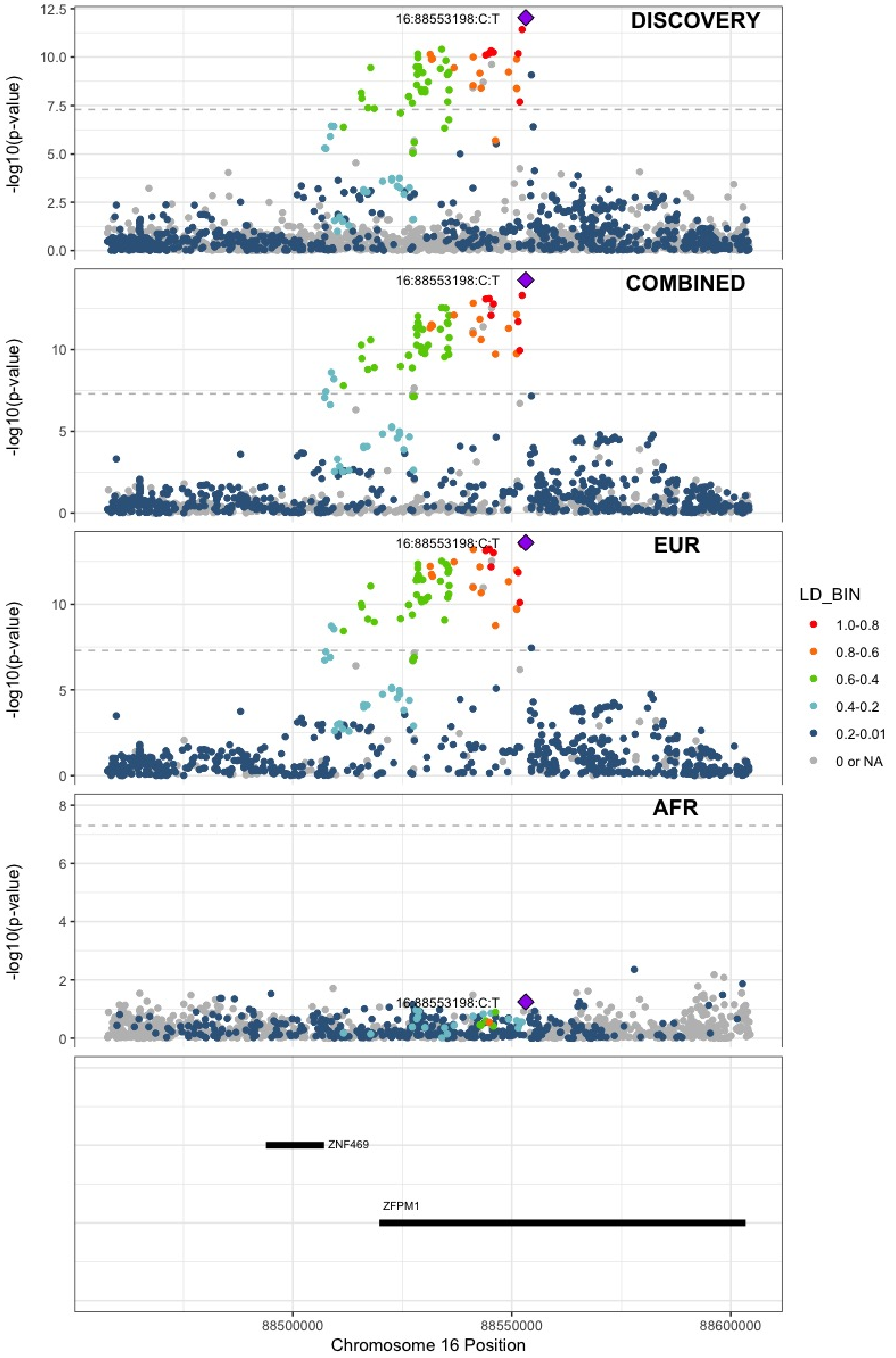

**Supplementary Figure 5.67** – rs6503222 (chr17:1977862) in locus 112 (SMG6)

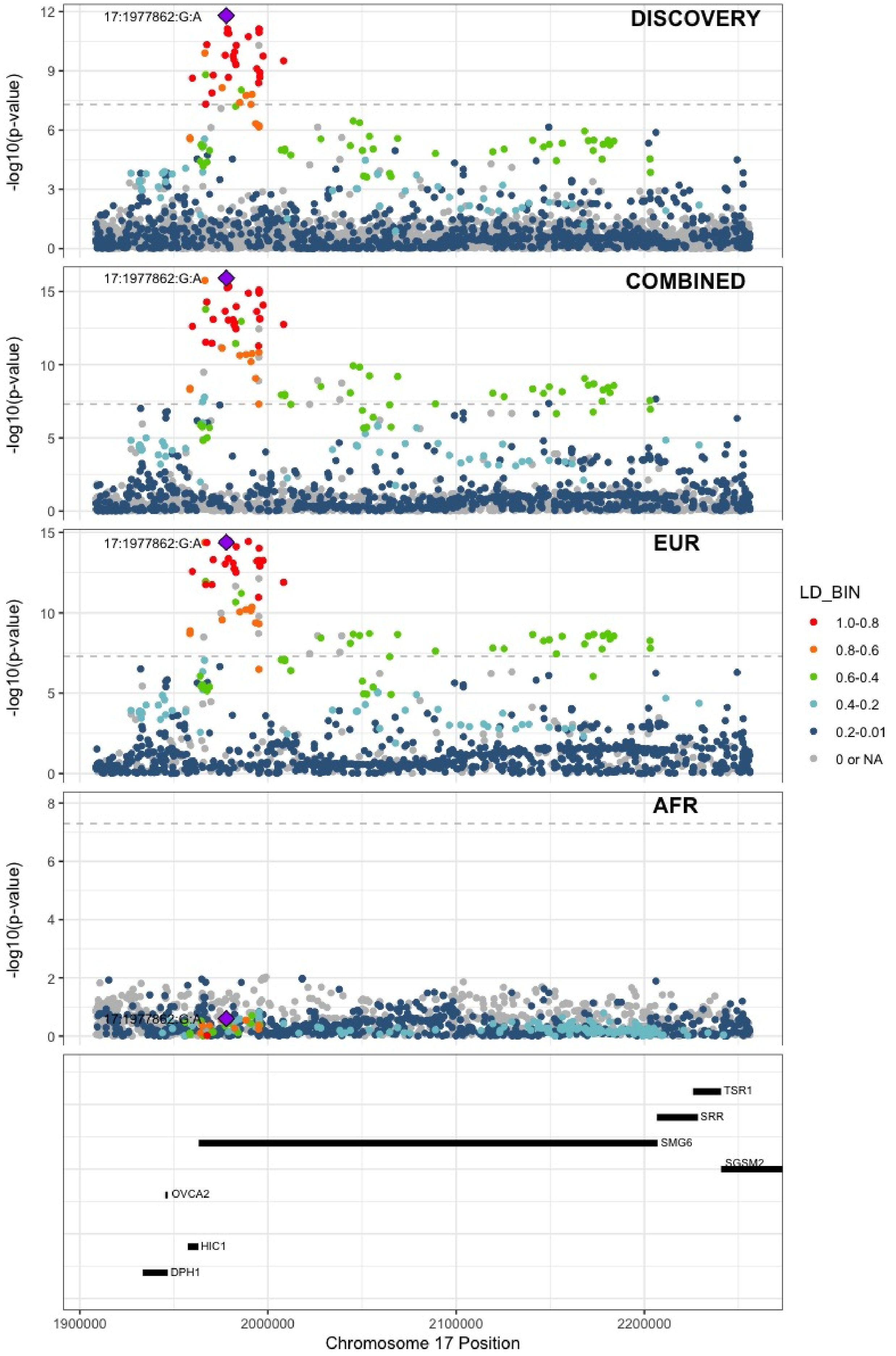

**Supplementary Figure 5.68** – rs7225756 (chr17:6893691) in locus 113 (ALOX12-AS1)

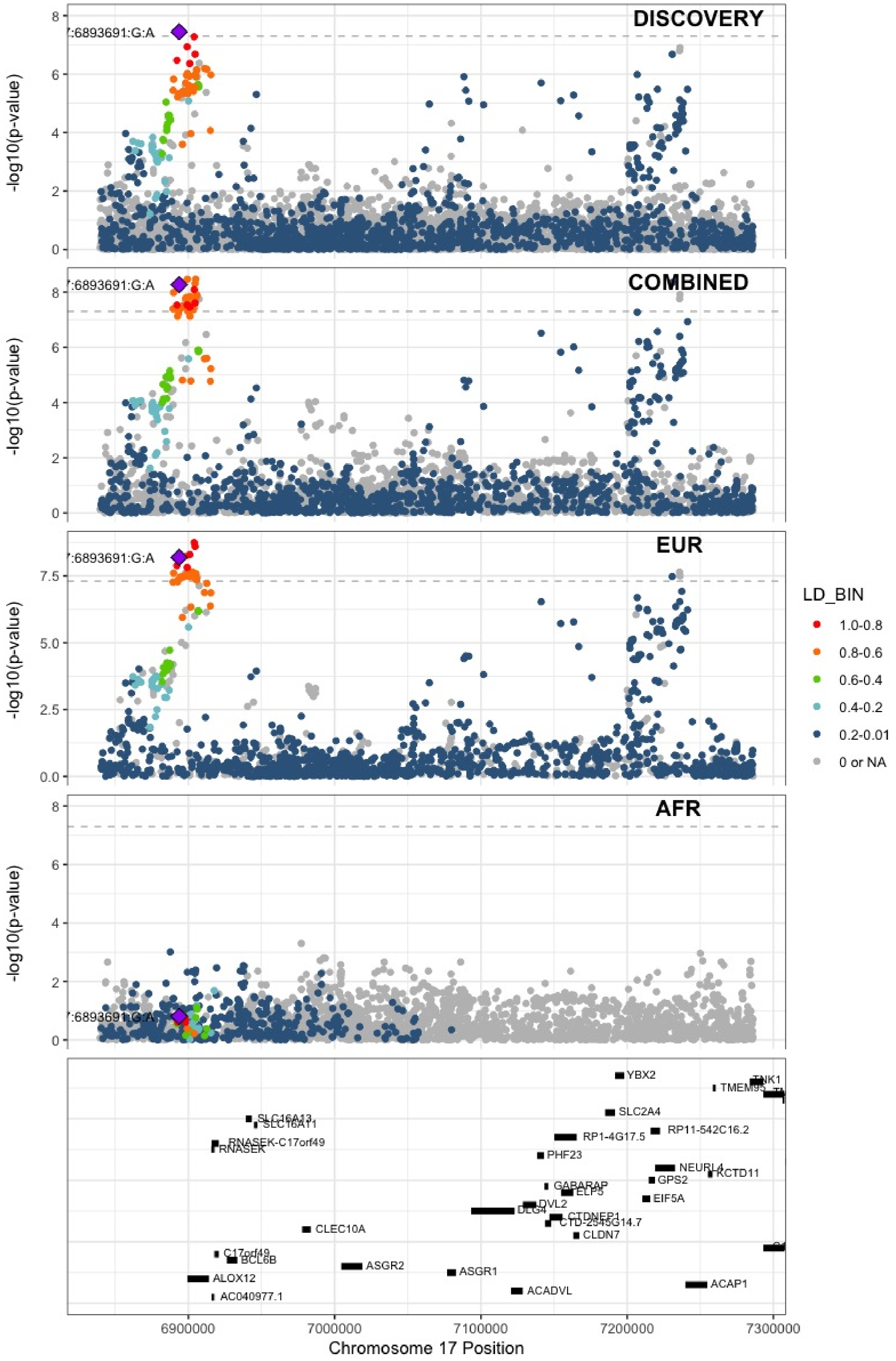

**Supplementary Figure 5.69** – rs62054822 (chr17:43927708) in locus 115 (MAPT-AS1)

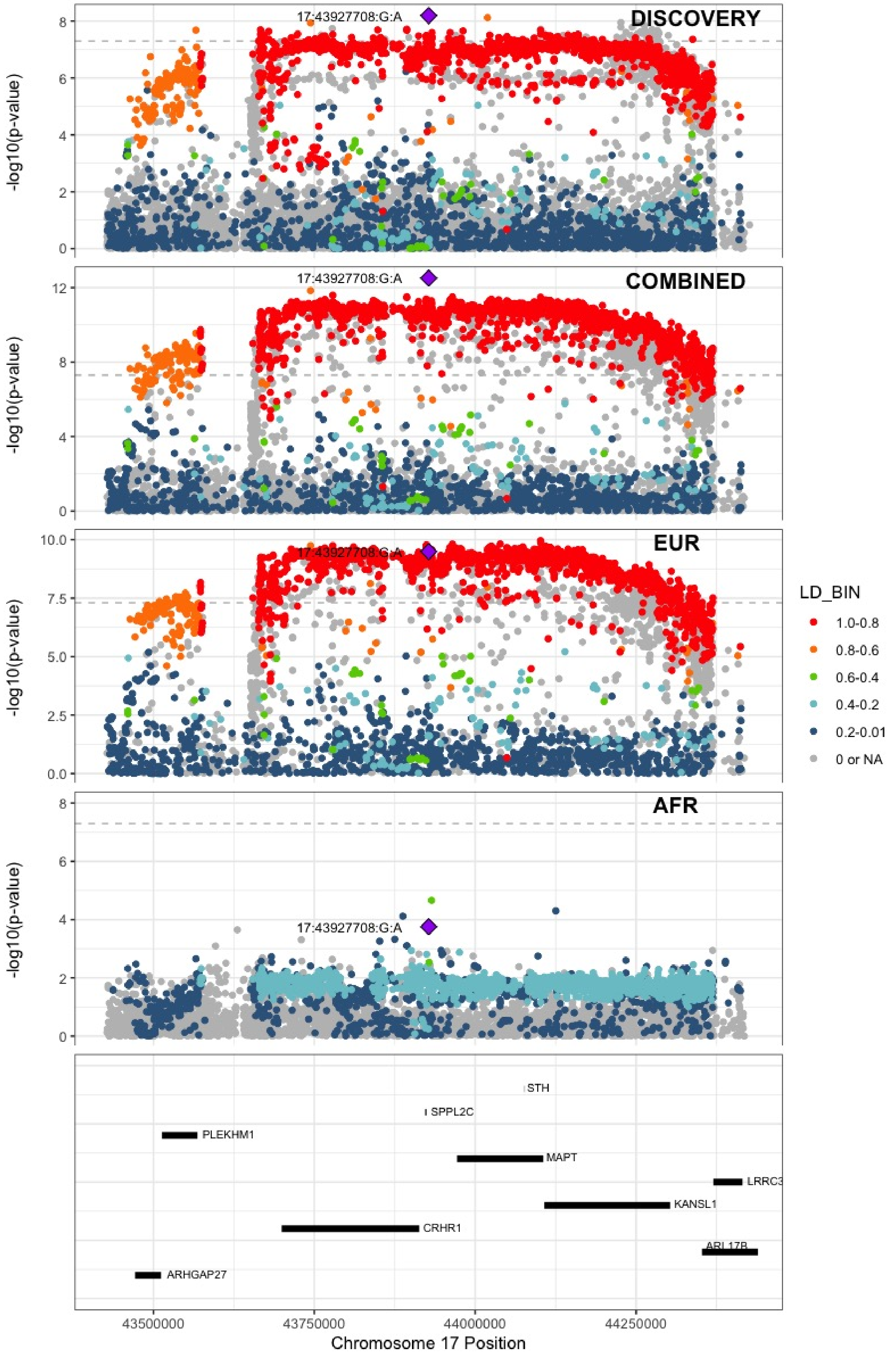

**Supplementary Figure 5.70** – rs142140545 (chr17:64191540) in locus 116 (CEP112;APOH)

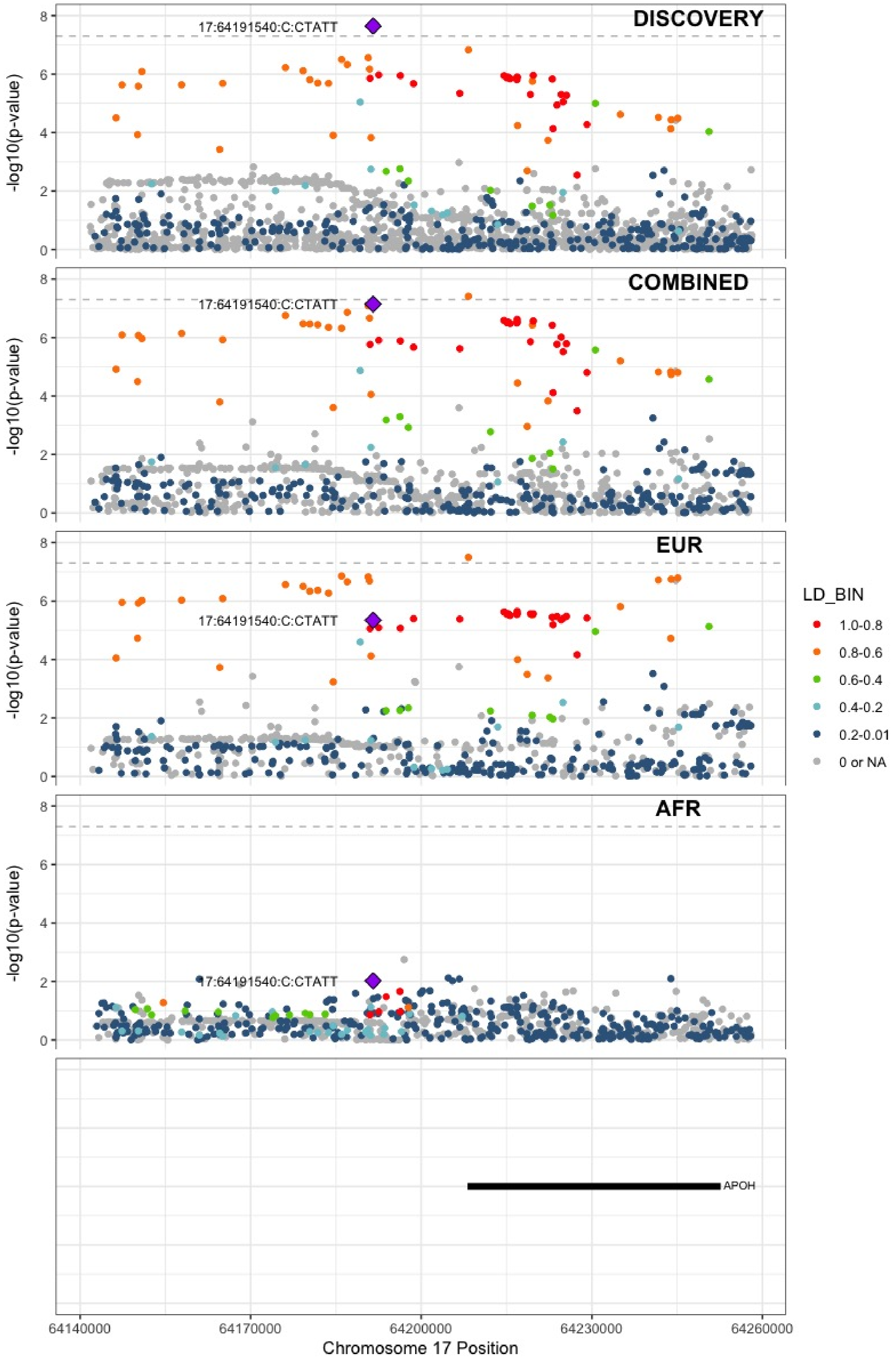

**Supplementary Figure 5.71** – rs59277920 (chr19:6077231) in locus 118 (RFX2)

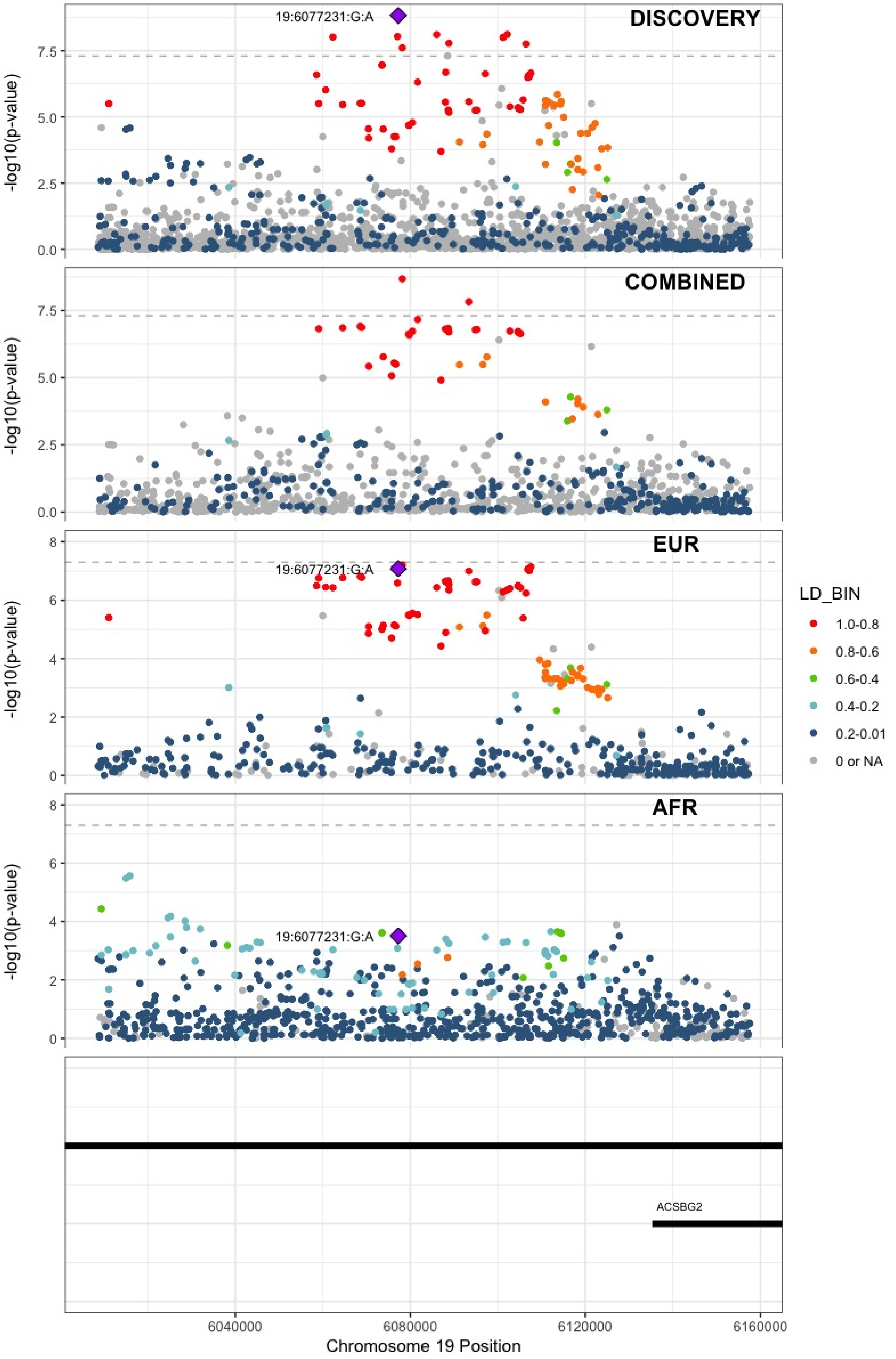

**Supplementary Figure 5.72** – rs8110055 (chr19:10739143) in locus 119 (SLC44A2)

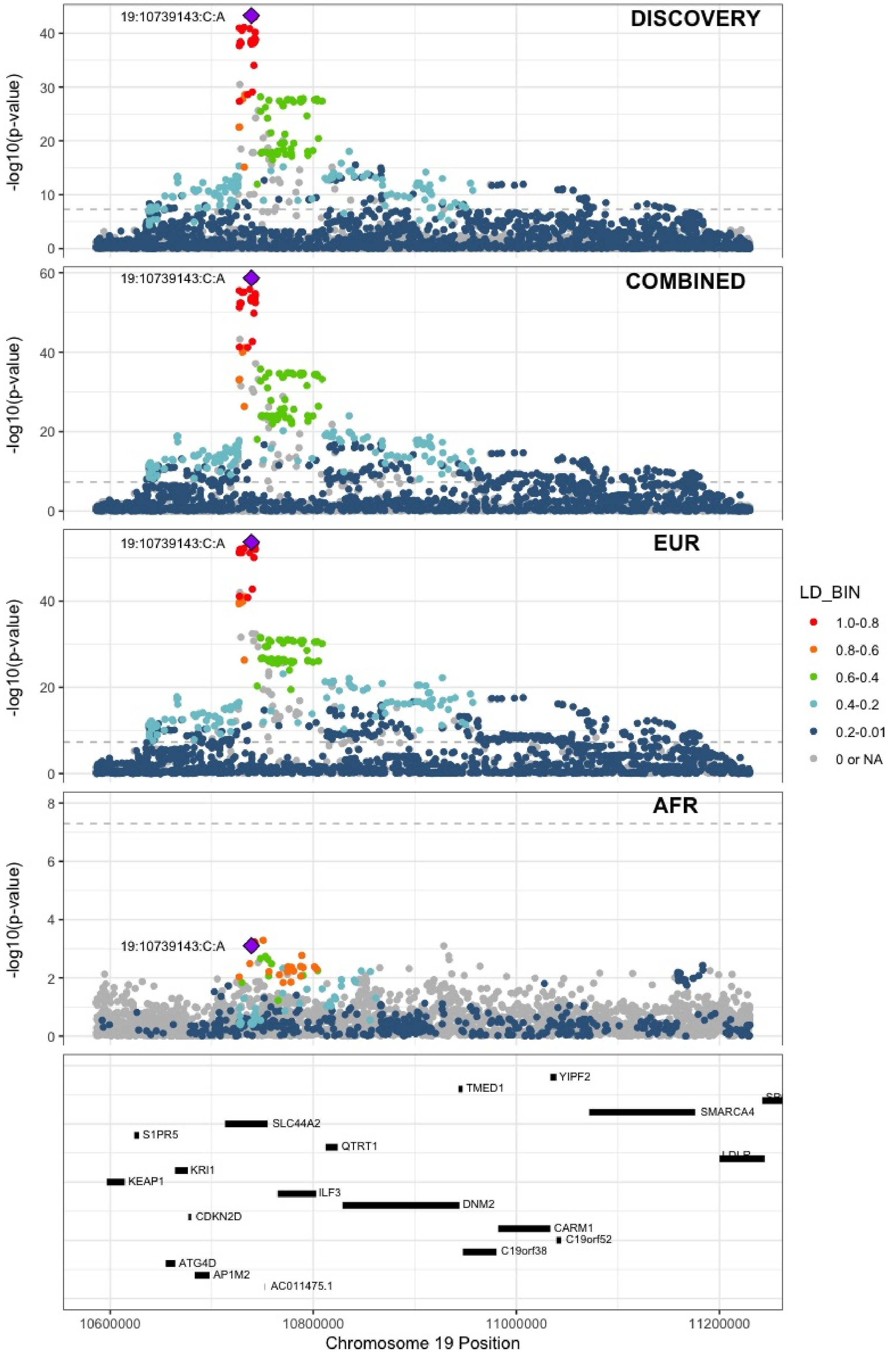

**Supplementary Figure 5.73** – rs34783010 (chr19:46180414) in locus 122 (GIPR)

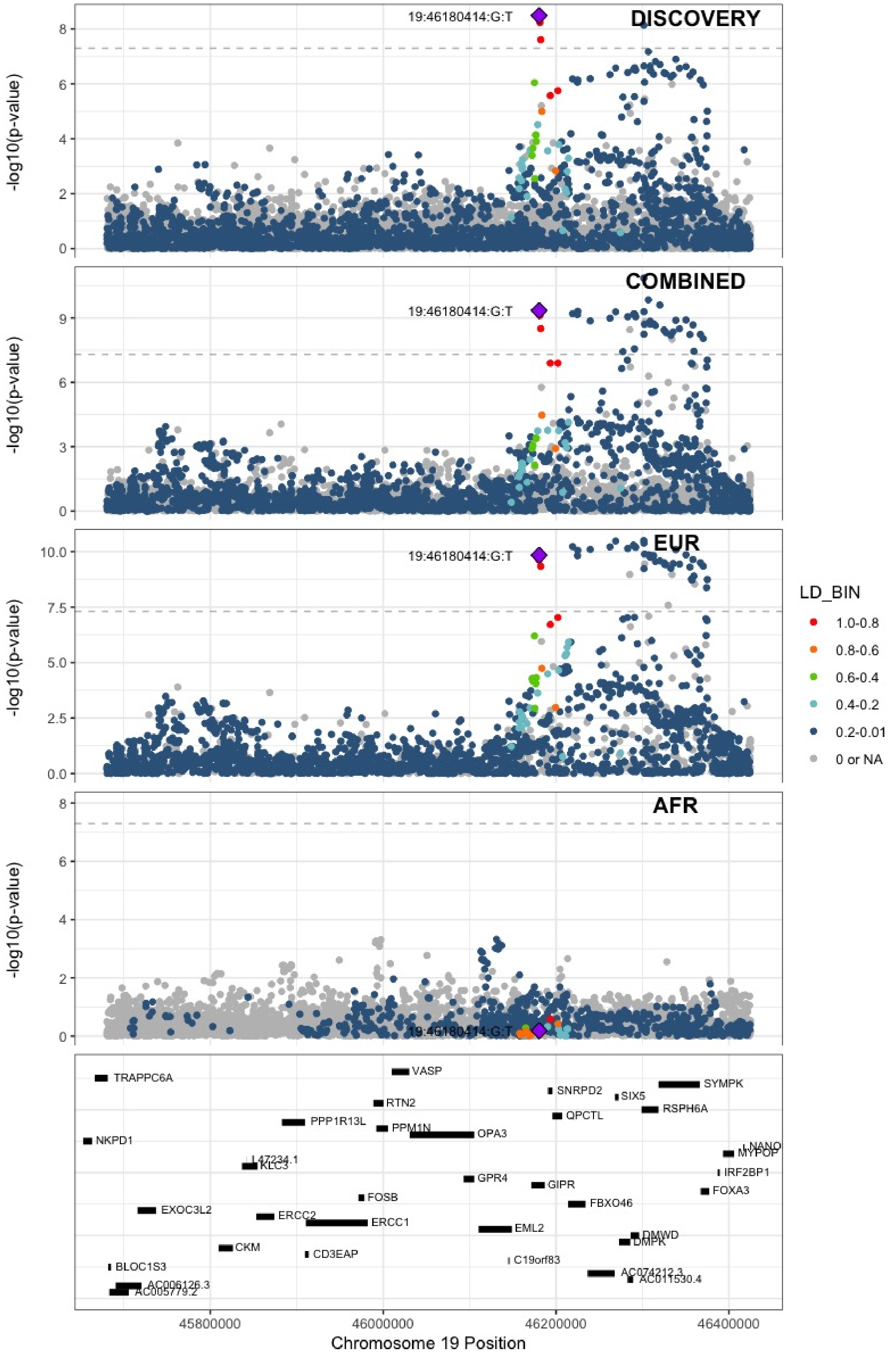

**Supplementary Figure 5.74** – rs1688264 (chr19:49209560) in locus 123 (FUT2)

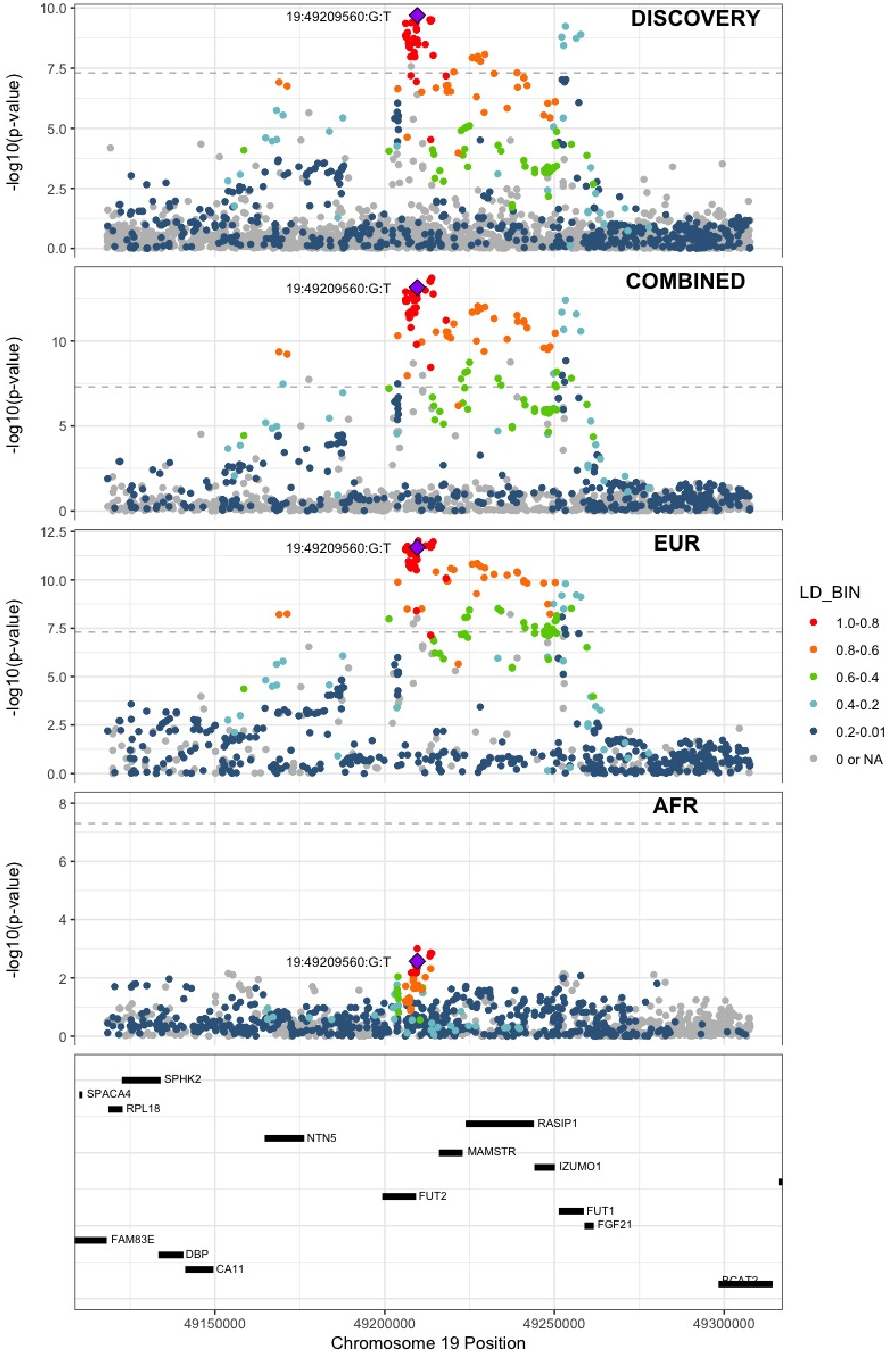

**Supplementary Figure 5.75** – rs1654425 (chr19:55538980) in locus 124 (GP6)

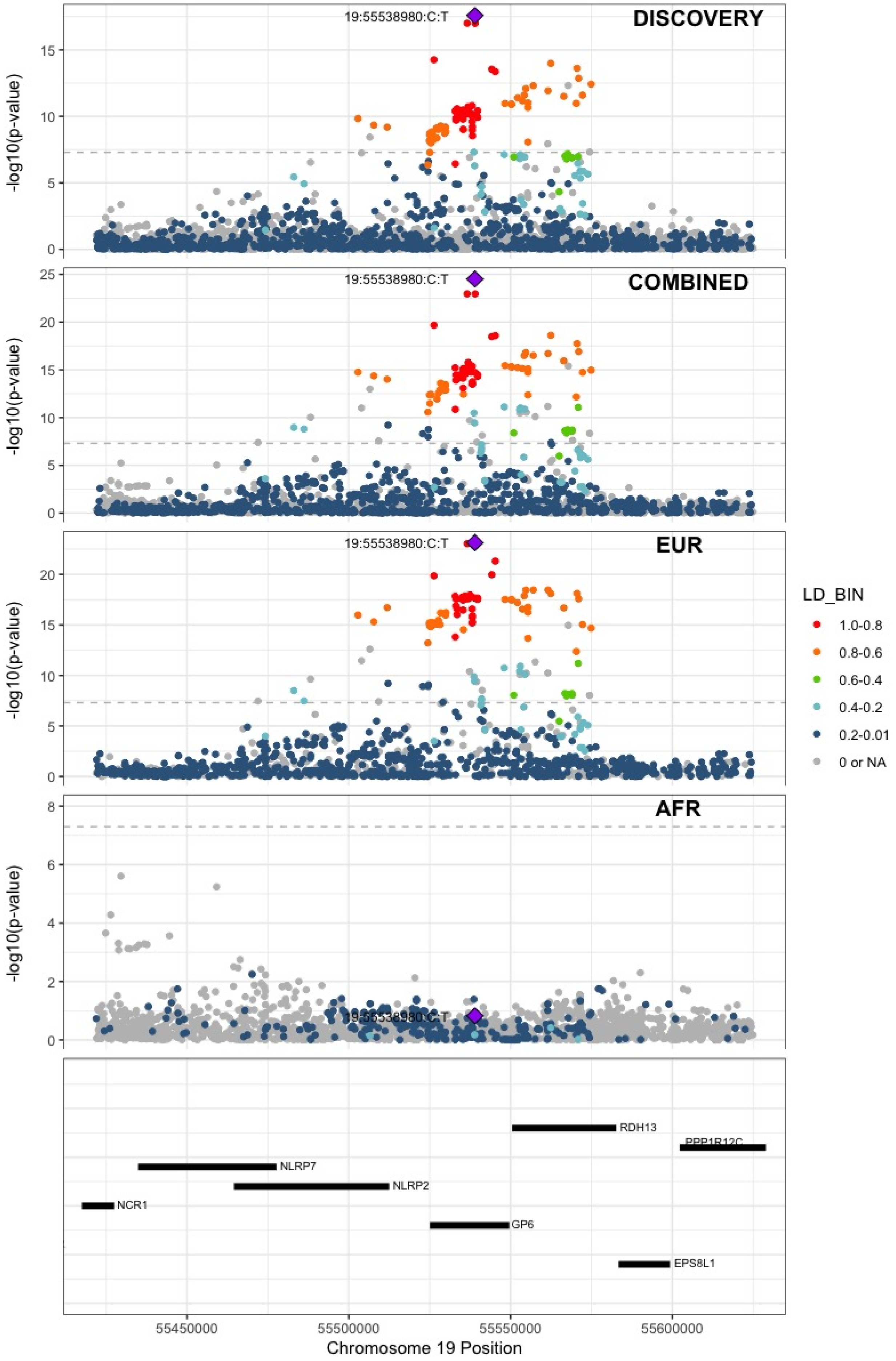

**Supplementary Figure 5.76** – rs2631628 (chr19:56645250) in locus 125 (ZNF787;ZNF444)

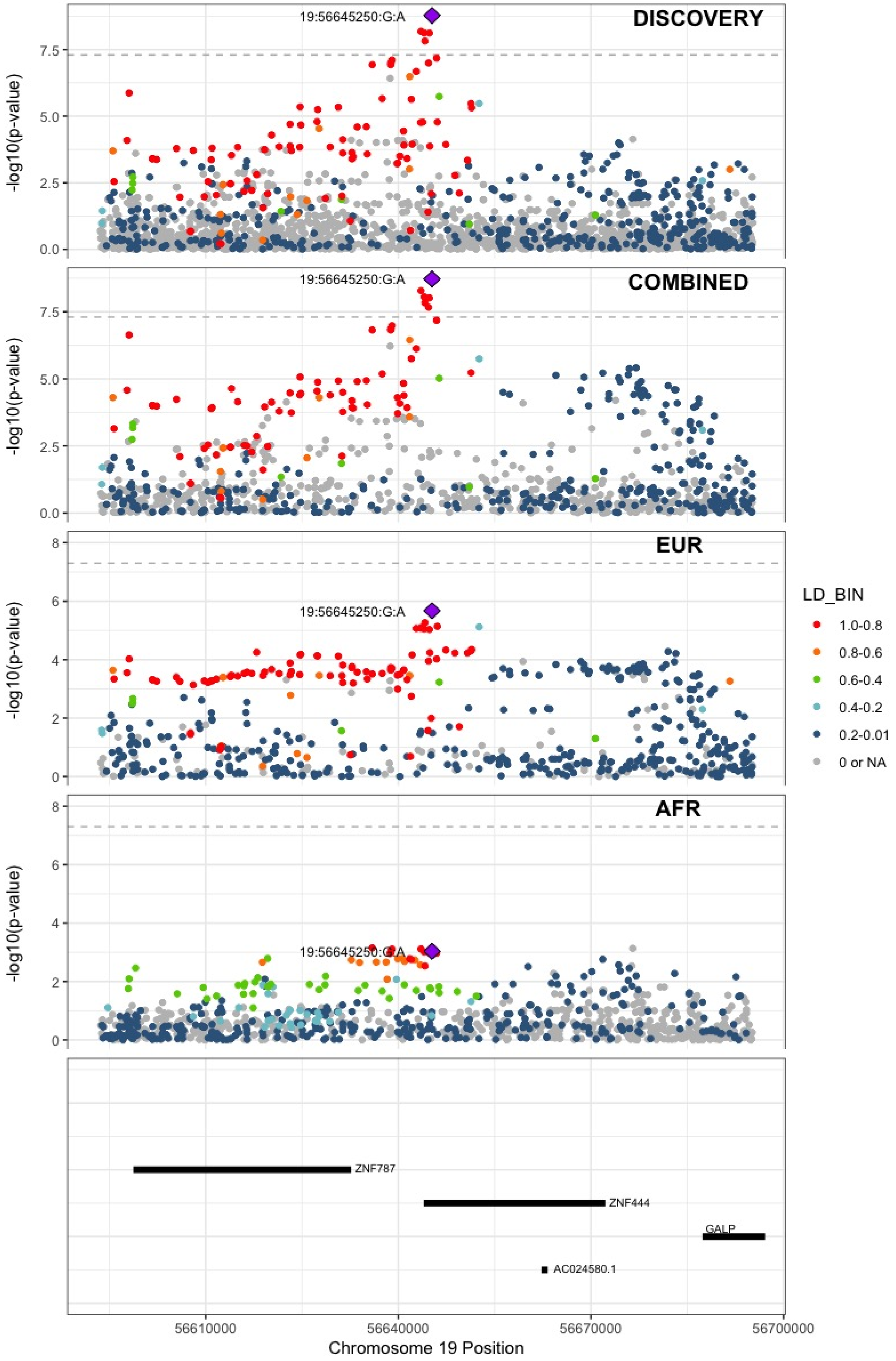

**Supplementary Figure 5.77** – rs79388863 (chr20:23168500) in locus 126 (LINC00656 ;NXT1)

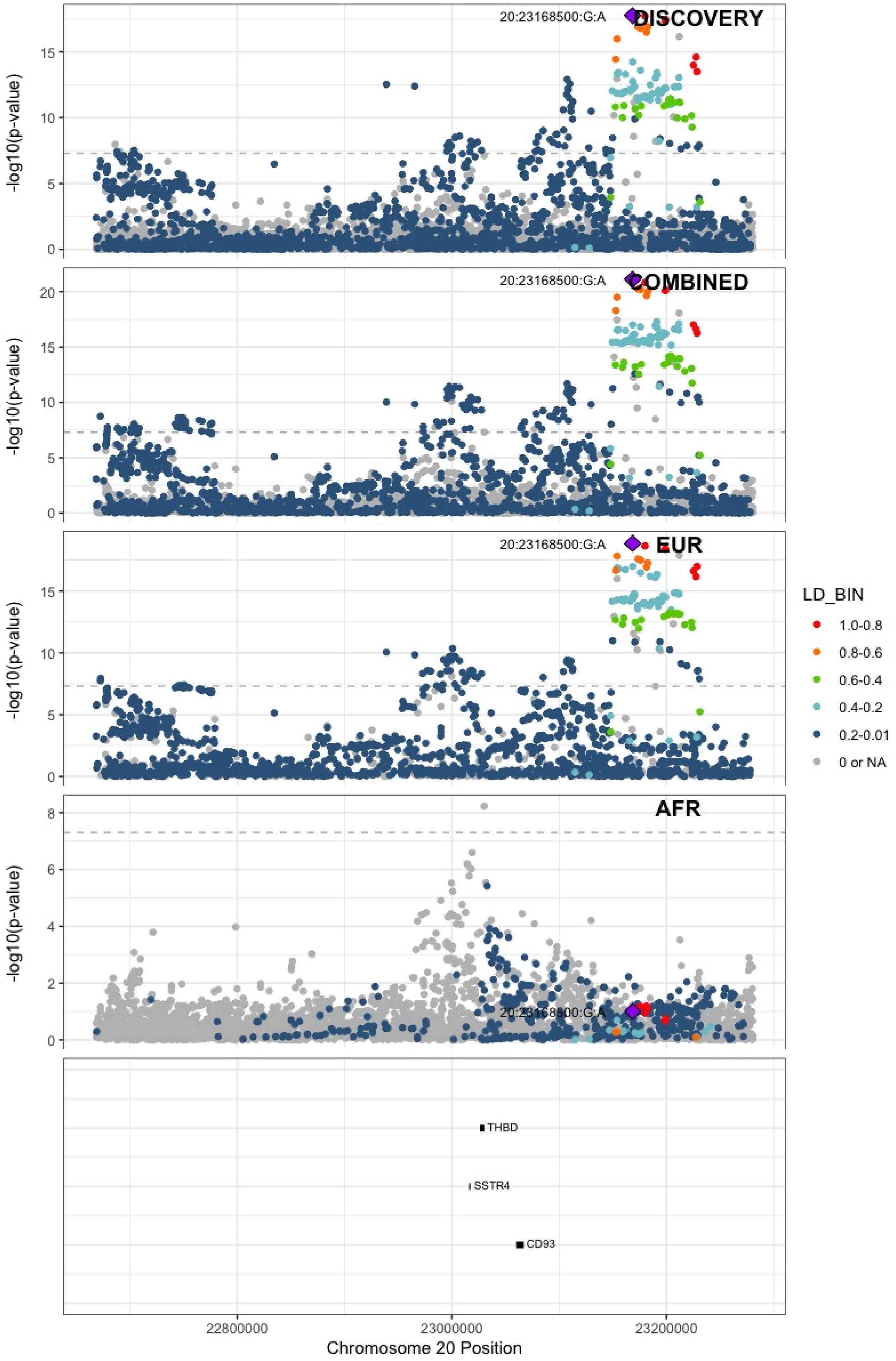

**Supplementary Figure 5.78** – rs6060288 (chr20:33772243) in locus 127 (MMP24-AS1-EDEM2)

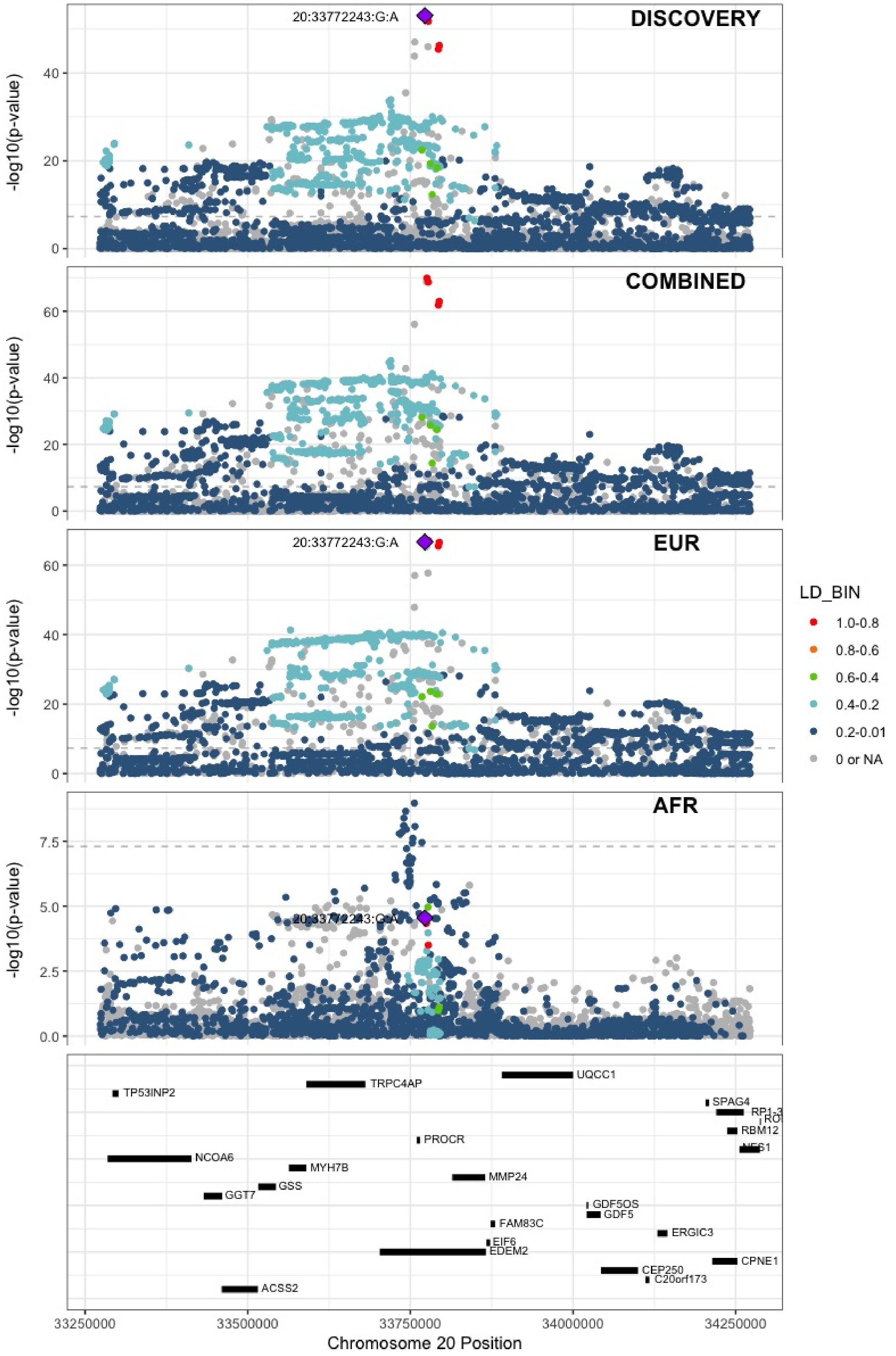

**Supplementary Figure 5.79** – rs4820093 (chr22:33160208) in locus 129 (SYN3)

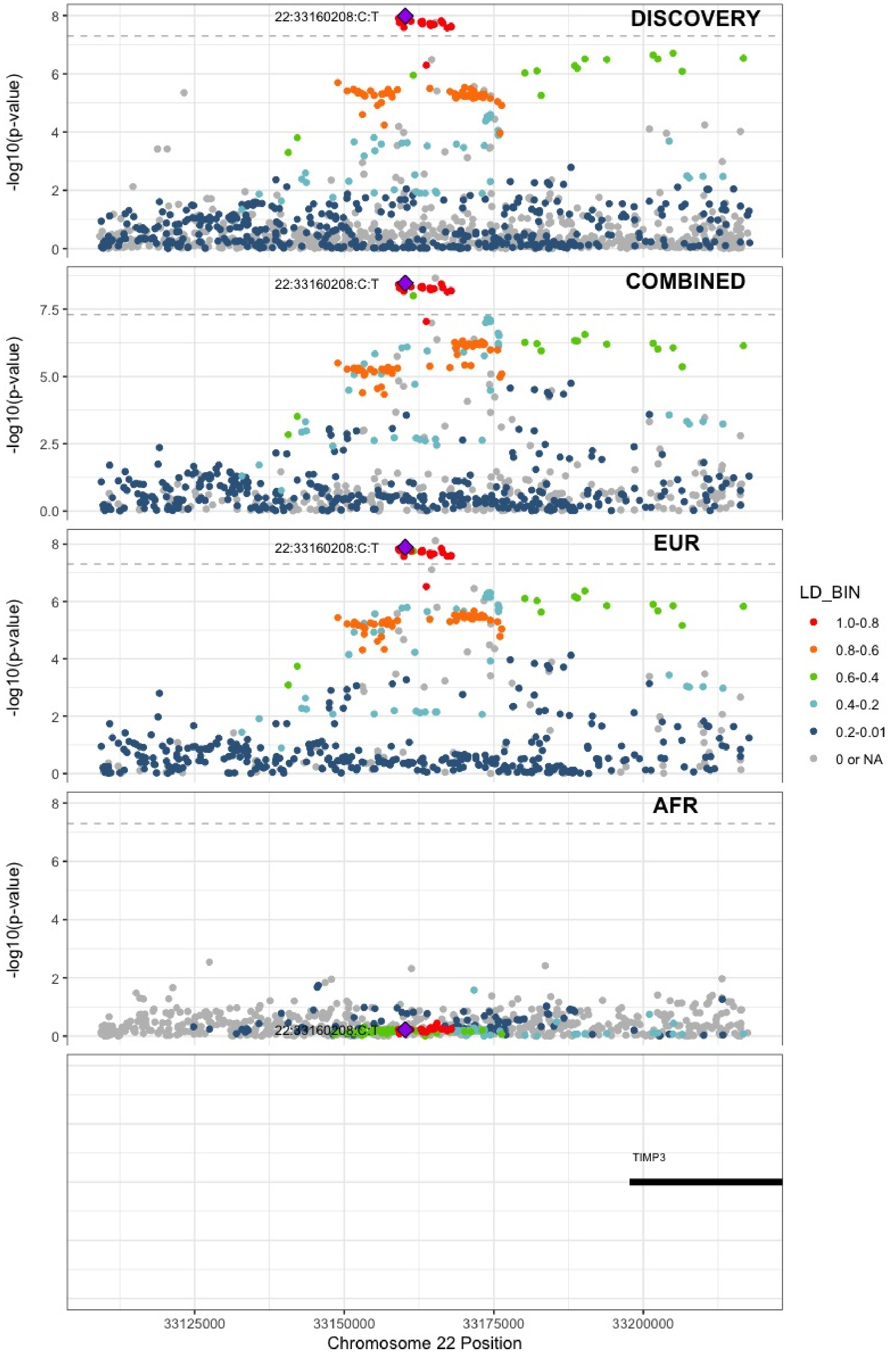

**Supplementary Figure 5.80** – rs9611844 (chr22:43115776) in locus 130 (A4GALT)

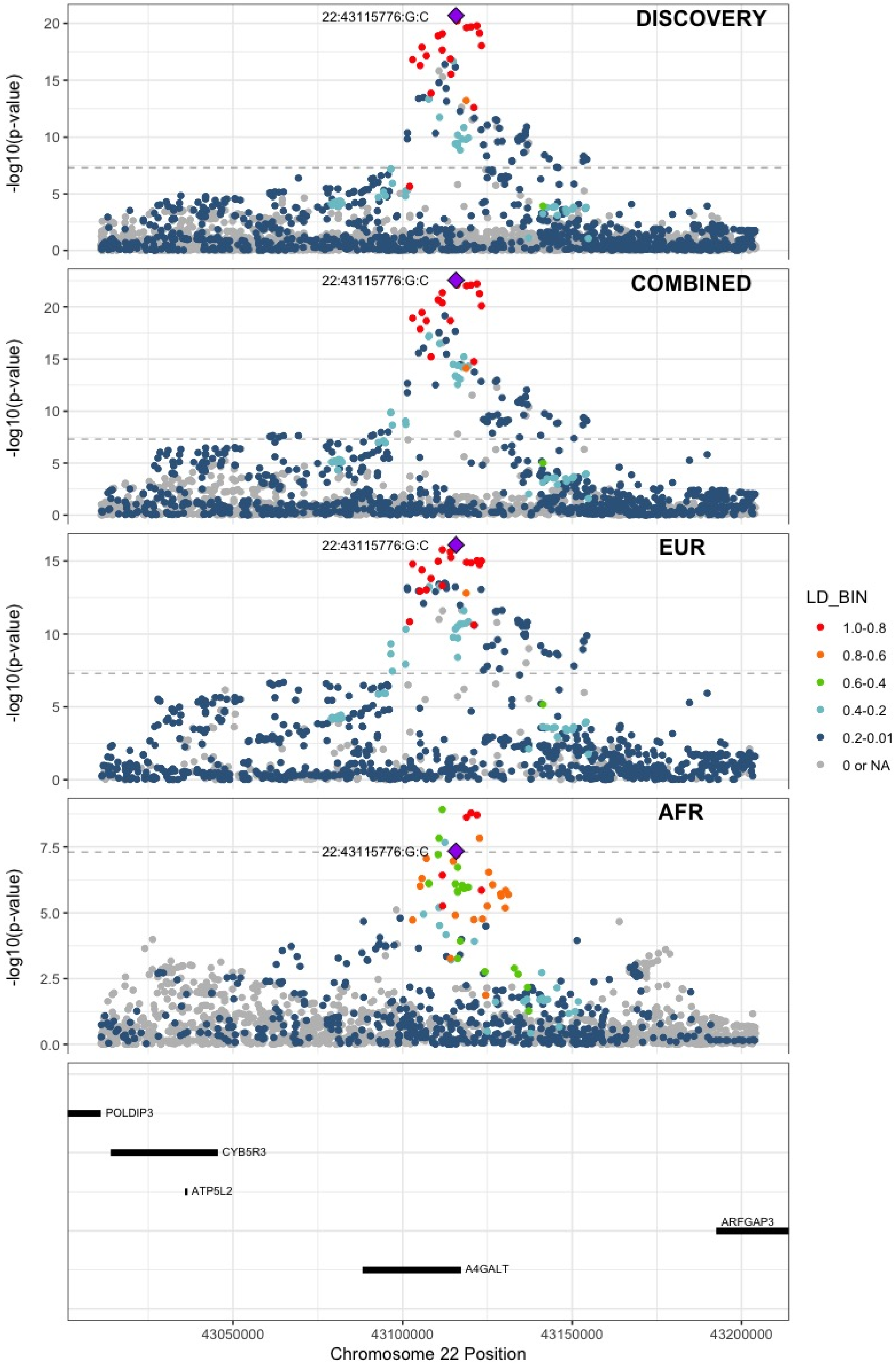

**Supplementary Figure 5.81** – rs34609587 (chr23:11657363) in locus 131 (ARHGAP6)

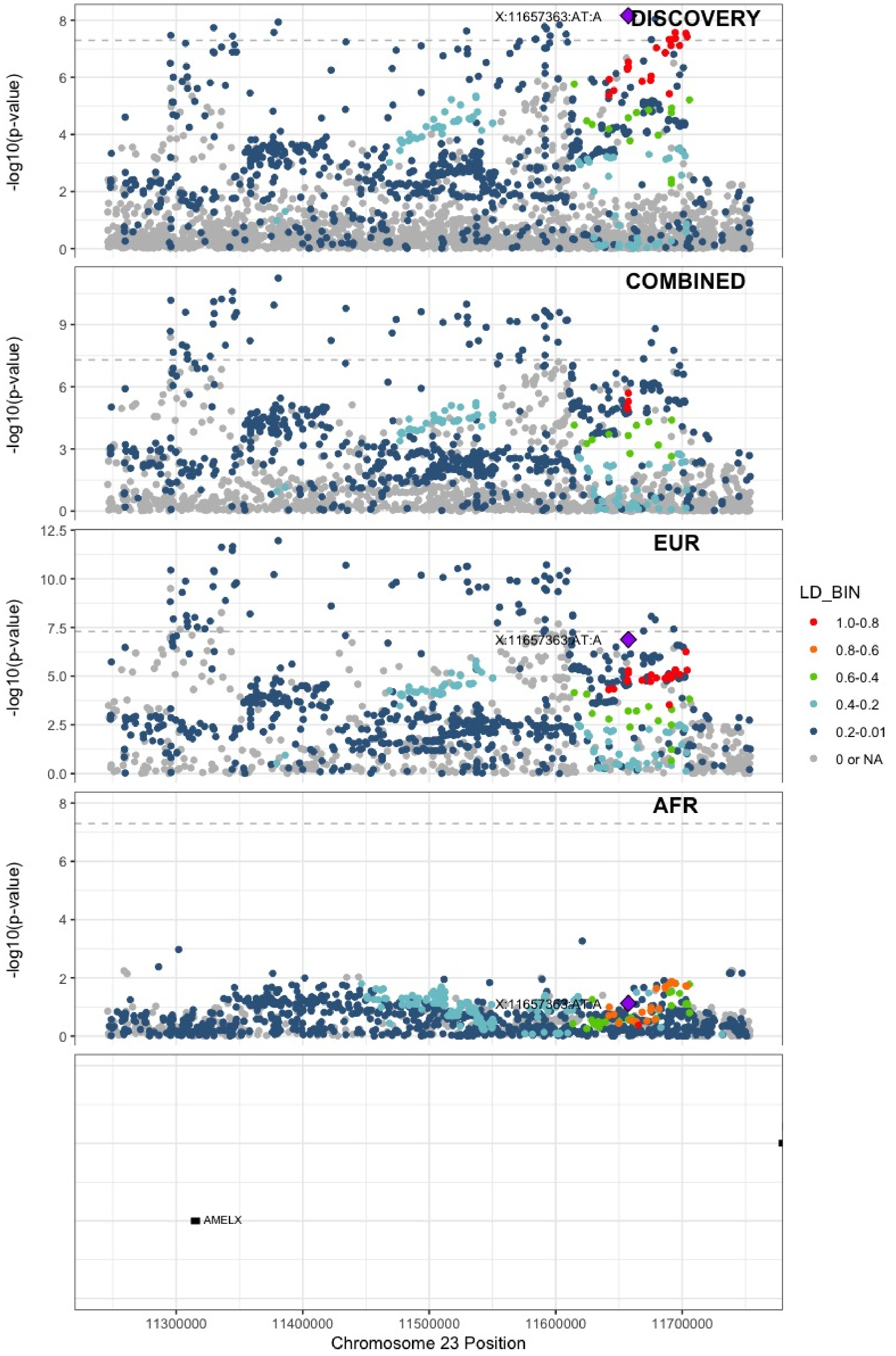

**Supplementary Figure 5.82** – rs6632109 (chr23:34894638) in locus 132 (TMEM47;FAM47B)

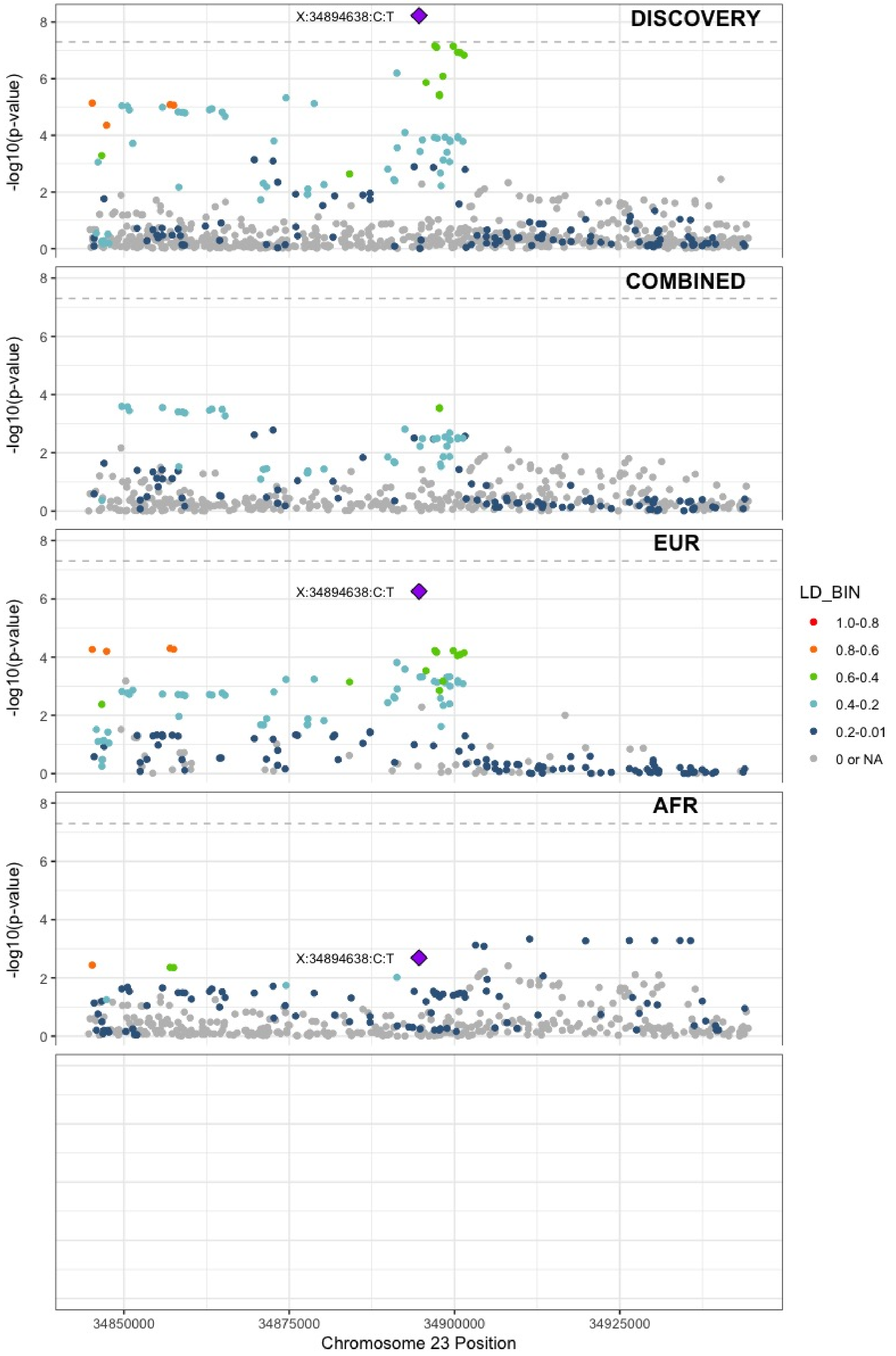

**Supplementary Figure 5.83** – rs3002416 (chr23:39710195) in locus 133 (MIR1587;BCOR)

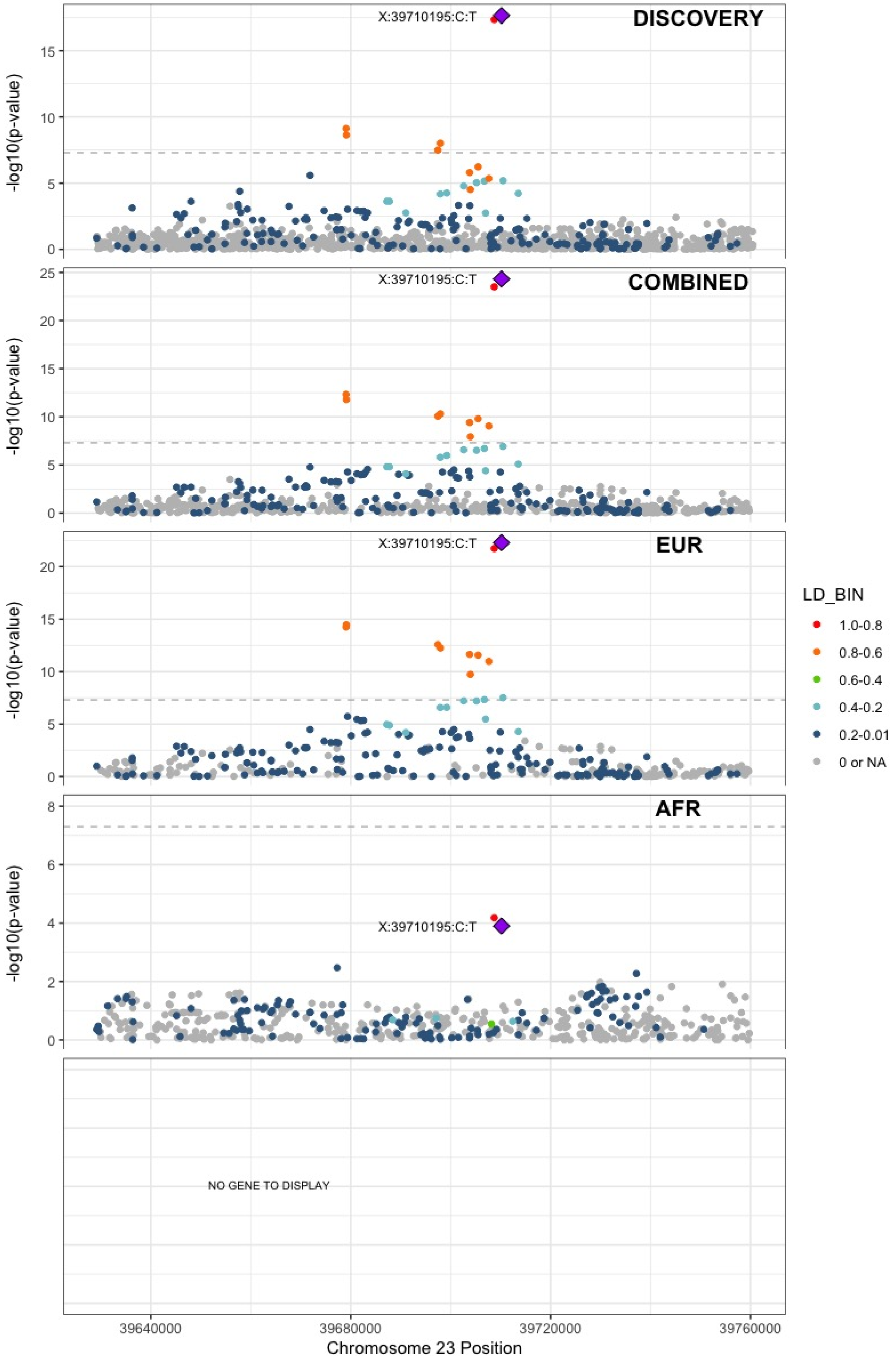

**Supplementary Figure 5.84** – rs6048 (chr23:138633280) in locus 134 (F9)

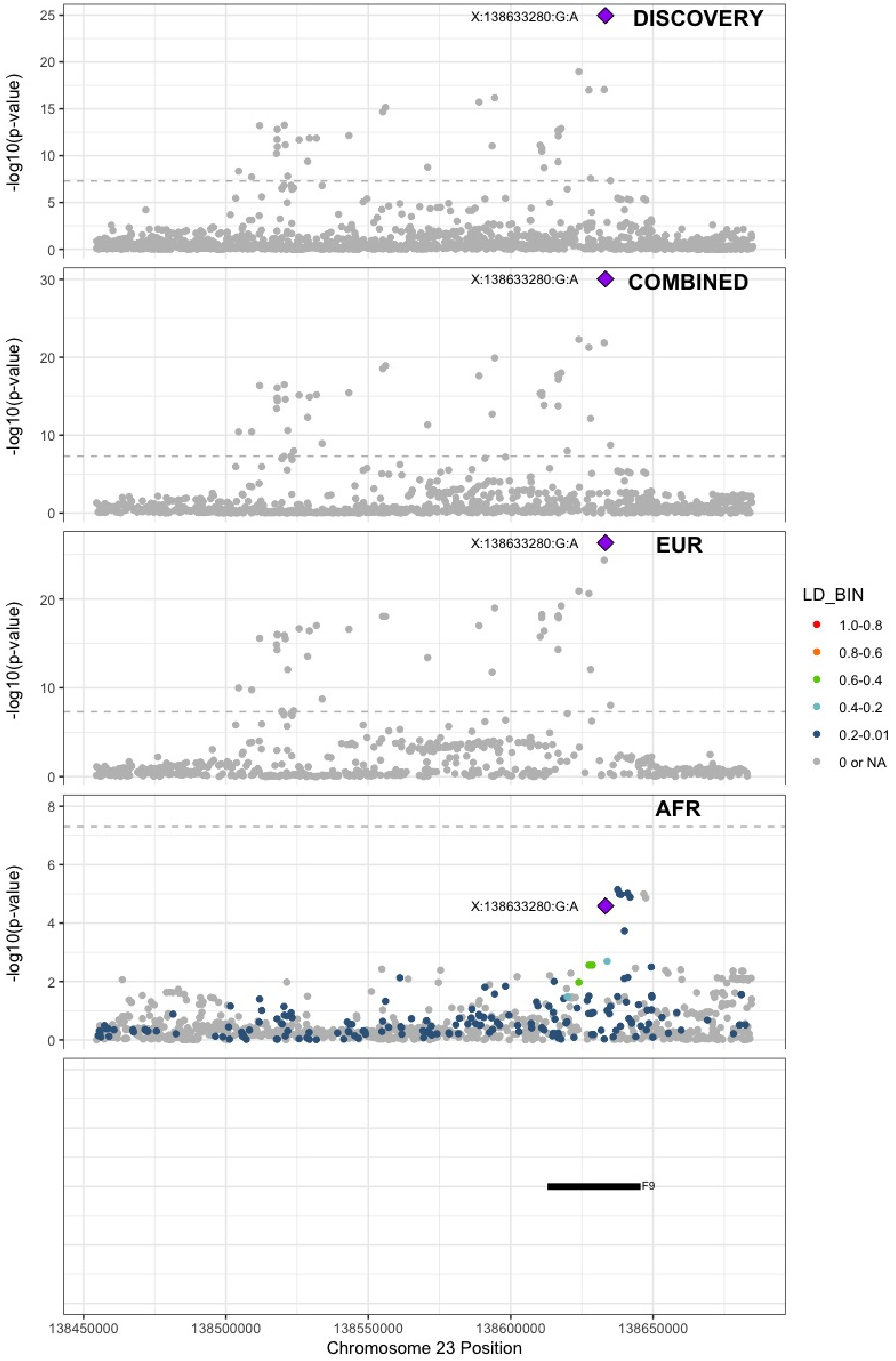

**Supplementary Figure 5.85** – rs2084408 (chr23:154346709) in locus 135 (BRCC3)

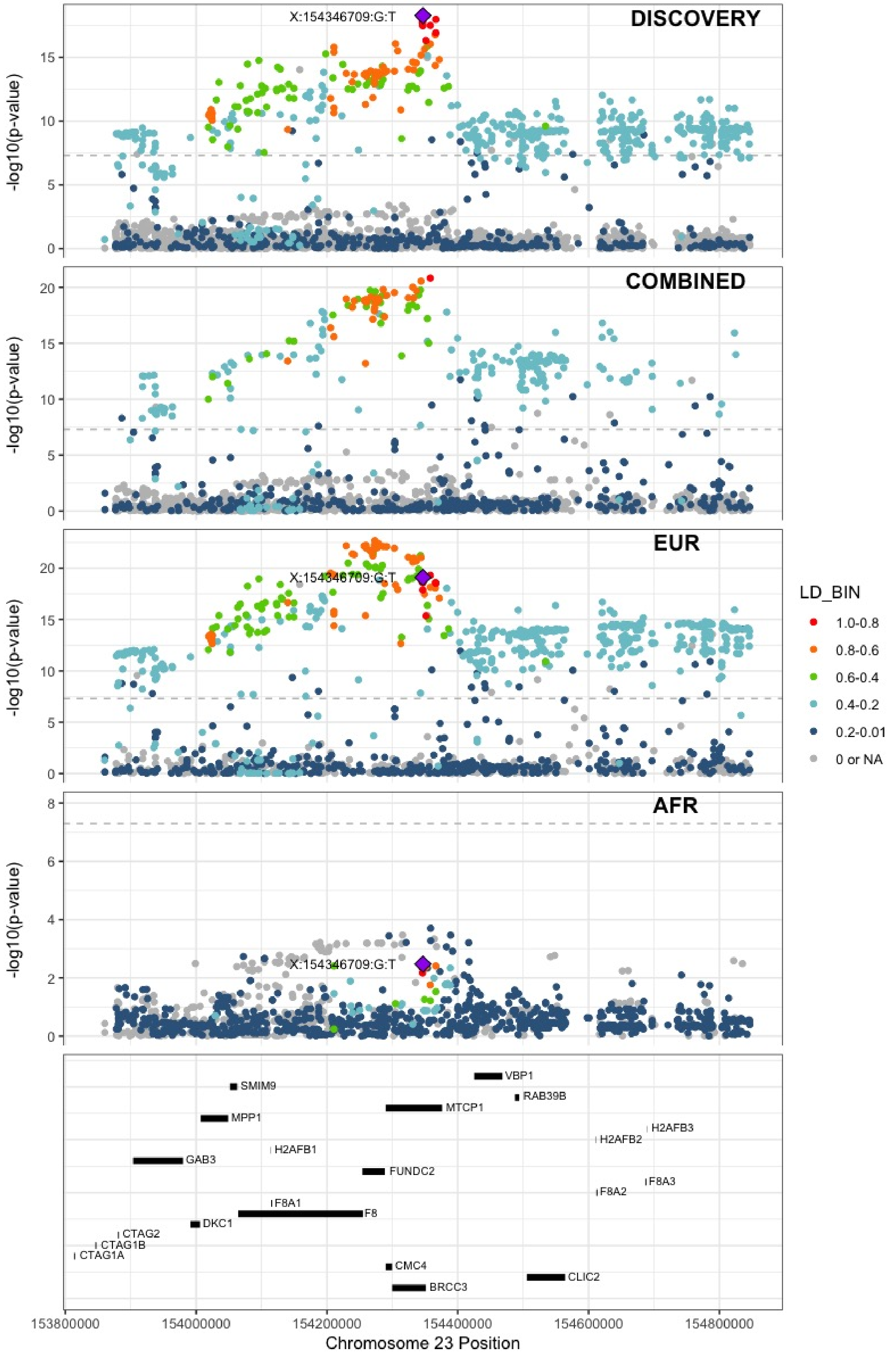
