## Supplemental Figure 6 for "Cross-Ancestry Investigation of Venous Thromboembolism Genomic Predictors"

### Supplementary Figure 6.
### Regional association plots of Combined analysis lead variants.


6.1 rs12121476 (chr1:9335327) in locus 1 (H6PD ;SPSB1) . . . 4
6.2 rs551176418 (chr1:27107263) in locus 2 (ARID1A) . . . 5
6.3 rs6695572 (chr1:77945635) in locus 3 (AK5) . . . 6
6.4 rs3832016 (chr1:109818158) in locus 4 (CELSR2) . . . 7
6.5 rs3767812 (chr1:118155620) in locus 5 (TENT5C) . . . 8
6.6 rs1267881263 (chr1:150496127) in locus 6 (FALEC ;ADAMTSL4) . . . 9
6.7 rs905938 (chr1:154991389) in locus 7 (DCST2) . . . 10
6.8 rs3557 (chr1:161188893) in locus 8 (FCER1G) . . . 11
6.9 rs6025 (chr1:169519049) in locus 9 (F5) . . . 12
6.10 rs6695940 (chr1:173882980) in locus 10 (SERPINC1) . . . 13
6.11 rs143410348 (chr1:196809316) in locus 12 (CFHR1 ;CFHR4) . . . 14
6.12 rs2842700 (chr1:207282149) in locus 13 (C4BPA) . . . 15
6.13 rs3811444 (chr1:248039451) in locus 14 (TRIM58) . . . 16
6.14 rs78475244 (chr2:65086804) in locus 16 (LINC01800) . . . 17
6.15 rs1867312 (chr2:68619981) in locus 17 (PLEK) . . . 18
6.16 rs1518760 (chr2:128140500) in locus 18 (MAP3K2) . . . 19
6.17 rs1912849 (chr2:188310118) in locus 19 (CALCRL) . . . 20
6.18 rs78872368 (chr2:198545250) in locus 20 (RFTN2 ;MARS2) . . . 21
6.19 rs715 (chr2:211543055) in locus 21 (CPS1) . . . 22
6.20 rs68066031 (chr2:224880498) in locus 22 (SERPINE2) . . . 23
6.21 rs13084580 (chr3:39188182) in locus 24 (CSRNP1) . . . 24
6.22 rs11130326 (chr3:52771920) in locus 25 (NEK4) . . . 25
6.23 rs529326506 (chr3:90503764) in locus 26 (EPHA3 ;NONE) . . . 26
6.24 rs28479320 (chr3:93752529) in locus 27 (ARL13B) . . . 27
6.25 rs900399 (chr3:156798732) in locus 30 (LINC02029 ;LINC00880) . . . 28
6.26 rs710446 (chr3:186459927) in locus 32 (KNG1) . . . 29
6.27 rs3796159 (chr3:194790434) in locus 33 (XXYLT1) . . . 30
6.28 rs9654093 (chr4:7903763) in locus 34 (AFAP1) . . . 31
6.29 rs781656 (chr4:57778645) in locus 35 (REST) . . . 32
6.30 rs6826579 (chr4:83785031) in locus 36 (SEC31A) . . . 33
6.31 rs2014912 (chr4:86715670) in locus 37 (ARHGAP24) . . . 34
6.32 rs2066864 (chr4:155525695) in locus 39 (FGG) . . . 35
6.33 rs3756011 (chr4:187206249) in locus 40 (F11) . . . 36
6.34 rs16867574 (chr5:38708554) in locus 42 (OSMR-AS1) . . . 37
6.35 rs7730244 (chr5:72957088) in locus 43 (ARHGEF28) . . . 38
6.36 rs750465397 (chr5:96289036) in locus 44 (LNPEP) . . . 39
6.37 rs147133967 (chr5:132426851) in locus 45 (HSPA4) . . . 40
6.38 rs214059 (chr6:25536937) in locus 46 (CARMIL1) . . . 41
6.39 rs2394251 (chr6:29943688) in locus 47 (HCG9) . . . 42
6.40 rs9268863 (chr6:32430289) in locus 48 (HLA-DRA ;HLA-DRB5) . . . 43
6.41 rs145294670 (chr6:34622561) in locus 49 (ILRUN) . . . 44
6.42 rs9386182 (chr6:147691069) in locus 52 (STXBP5) . . . 45
6.43 rs1513275 (chr7:28259233) in locus 53 (JAZF1-AS1) . . . 46
6.44 rs42038 (chr7:92243719) in locus 54 (CDK6) . . . 47
6.45 rs10099512 (chr8:9178821) in locus 57 (LOC101929128 ;LOC157273) . . . 48
6.46 rs2048528 (chr8:23373680) in locus 59 (ENTPD4 ;SLC25A37) . . . 49
6.47 rs2685412 (chr8:27810603) in locus 60 (SCARA5) . . . 50
6.48 rs2915595 (chr8:30402817) in locus 61 (RBPMS) . . . 51
6.49 rs6993770 (chr8:106581528) in locus 62 (ZFPM2) . . . 52
6.50 rs4236786 (chr8:108291878) in locus 63 (ANGPT1) . . . 53

6.51 rs10820606 (chr9:99192919) in locus 65 (ZNF367;HABP4) . . . 54
6.52 rs505922 (chr9:136149229) in locus 66 (ABO) . . . 55
6.53 rs1243187 (chr10:21907016) in locus 68 (MLLT10) . . . 56
6.54 rs4272700 (chr10:27881308) in locus 69 (RAB18;MKX) . . . 57
6.55 rs17490626 (chr10:71218646) in locus 72 (TSPAN15) . . . 58
6.56 rs16937003 (chr10:80938499) in locus 73 (ZMIZ1) . . . 59
6.57 rs57866767 (chr10:96023077) in locus 74 (PLCE1) . . . 60
6.58 rs10886430 (chr10:121010256) in locus 75 (GRK5) . . . 61
6.59 rs2030291 (chr11:16251251) in locus 77 (SOX6) . . . 62
6.60 rs11032074 (chr11:32993887) in locus 78 (QSER1) . . . 63
6.61 rs1799963 (chr11:46761055) in locus 79 (F2) . . . 64
6.62 rs141798115 (chr11:56875074) in locus 80 (OR5AK4P;LRRC55) . . . 65
6.63 rs4354705 (chr11:60088159) in locus 81 (MS4A4A;MS4A6E) . . . 66
6.64 rs174551 (chr11:61573684) in locus 82 (FADS1) . . . 67
6.65 rs2846027 (chr11:114003415) in locus 84 (ZBTB16) . . . 68
6.66 rs35257264 (chr11:126296816) in locus 85 (ST3GAL4) . . . 69
6.67 rs7107568 (chr11:130779668) in locus 86 (SNX19) . . . 70
6.68 rs1558519 (chr12:6153738) in locus 87 (VWF) . . . 71
6.69 rs7311483 (chr12:9053661) in locus 88 (A2ML1;PHC1) . . . 72
6.70 rs6580981 (chr12:54723028) in locus 89 (COPZ1) . . . 73
6.71 rs142351376 (chr12:104136288) in locus 90 (STAB2) . . . 74
6.72 rs3184504 (chr12:111884608) in locus 91 (SH2B3) . . . 75
6.73 rs2851435 (chr12:123712416) in locus 92 (MPHOSPH9) . . . 76
6.74 rs3211752 (chr13:113787459) in locus 94 (F10) . . . 77
6.75 rs11158204 (chr14:58844526) in locus 95 (ARID4A;TOMM20L) . . . 78
6.76 rs2127869 (chr14:65794352) in locus 96 (LINC02324;MIR4708) . . . 79
6.77 rs7152641 (chr14:92213967) in locus 97 (CATSPERB;TC2N) . . . 80
6.78 rs28929474 (chr14:94844947) in locus 98 (SERPINA1) . . . 81
6.79 rs28519971 (chr14:103089808) in locus 99 (RCOR1) . . . 82
6.80 rs1247656151 (chr15:43677975) in locus 100 (TUBGCP4) . . . 83
6.81 rs7183672 (chr15:96101018) in locus 103 (LINC00924;LOC105369212) . . . 84
6.82 rs71376077 (chr16:15738114) in locus 105 (NDE1) . . . 85
6.83 rs12443808 (chr16:30996871) in locus 106 (HSD3B7) . . . 86
6.84 rs7197453 (chr16:72079127) in locus 107 (DHODH;HP) . . . 87
6.85 rs77246010 (chr16:75429853) in locus 108 (CFDP1) . . . 88
6.86 rs12445050 (chr16:81870969) in locus 109 (PLCG2) . . . 89
6.87 rs8049403 (chr16:85778651) in locus 110 (C16orf74) . . . 90
6.88 rs28634651 (chr16:88553198) in locus 111 (ZFPM1) . . . 91
6.89 rs6503222 (chr17:1977862) in locus 112 (SMG6) . . . 92
6.90 rs434473 (chr17:6904934) in locus 113 (ALOX12) . . . 93
6.91 rs71138827 (chr17:27833678) in locus 114 (TAOK1) . . . 94
6.92 rs62054822 (chr17:43927708) in locus 115 (MAPT-AS1) . . . 95
6.93 rs1801690 (chr17:64208285) in locus 116 (APOH) . . . 96
6.94 rs573431210 (chr17:67276383) in locus 117 (ABCA5) . . . 97
6.95 rs59277920 (chr19:6077231) in locus 118 (RFX2) . . . 98
6.96 rs8110055 (chr19:10739143) in locus 119 (SLC44A2) . . . 99
6.97 rs889139 (chr19:33889369) in locus 120 (PEPD) . . . 100
6.98 rs2545774 (chr19:41287674) in locus 121 (RAB4B) . . . 101
6.99 rs8108474 (chr19:46301479) in locus 122 (RSPH6A) . . . 102
6.100rs2638282 (chr19:49213833) in locus 123 (FUT2;MAMSTR) . . . 103
6.101rs1654425 (chr19:55538980) in locus 124 (GP6) . . . 104
6.102rs2631628 (chr19:56645250) in locus 125 (ZNF787;ZNF444) . . . 105
6.103rs117390891 (chr20:23168526) in locus 126 (LINC00656;NXT1) . . . 106

6.104rs6060288 (chr20:33772243) in locus 127 (MMP24-AS1-EDEM2) . . . . . . . . . . . . . 107
6.105rs3070580 (chr22:33165231) in locus 129 (SYN3) . . . . . . . . . . . . . . . . . . . . . . . 108
6.106rs9611844 (chr22:43115776) in locus 130 (A4GALT) . . . . . . . . . . . . . . . . . . . . . 109
6.107rs12388118 (chr23:11380742) in locus 131 (ARHGAP6) . . . . . . . . . . . . . . . . . . . 110
6.108rs6632109 (chr23:34894638) in locus 132 (TMEM47;FAM47B) . . . . . . . . . . . . . . . 111
6.109rs3002416 (chr23:39710195) in locus 133 (MIR1587;BCOR) . . . . . . . . . . . . . . . . 112
6.110rs6048 (chr23:138633280) in locus 134 (F9) . . . . . . . . . . . . . . . . . . . . . . . . . . 113
6.111rs2084408 (chr23:154346709) in locus 135 (BRCC3) . . . . . . . . . . . . . . . . . . . . . 114

**Supplementary Figure 6.1** – rs12121476 (chr1:9335327) in locus 1 (H6PD ;SPSB1)

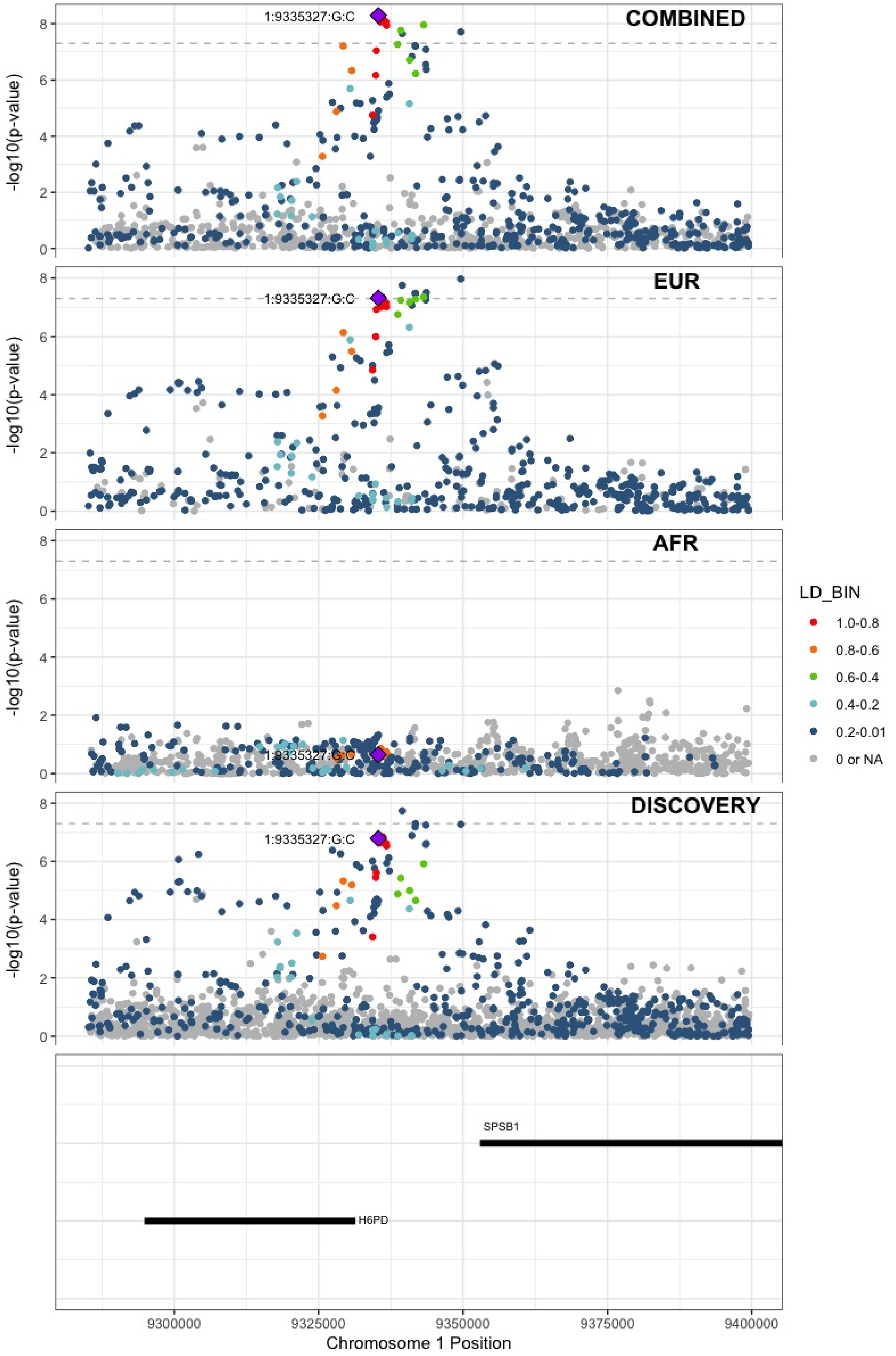

**Supplementary Figure 6.2** – rs551176418 (chr1:27107263) in locus 2 (ARID1A)

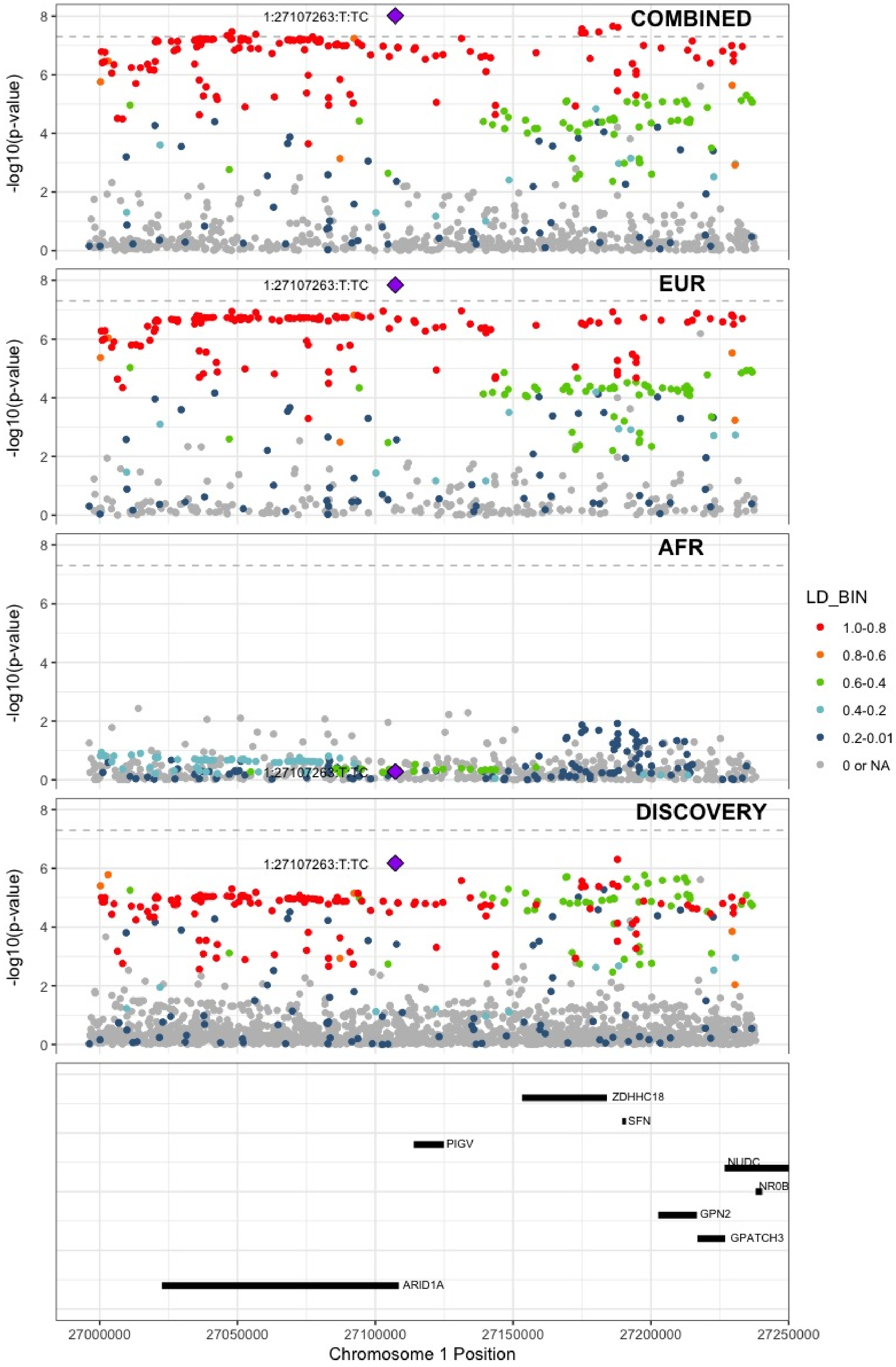

**Supplementary Figure 6.3** – rs6695572 (chr1:77945635) in locus 3 (AK5)

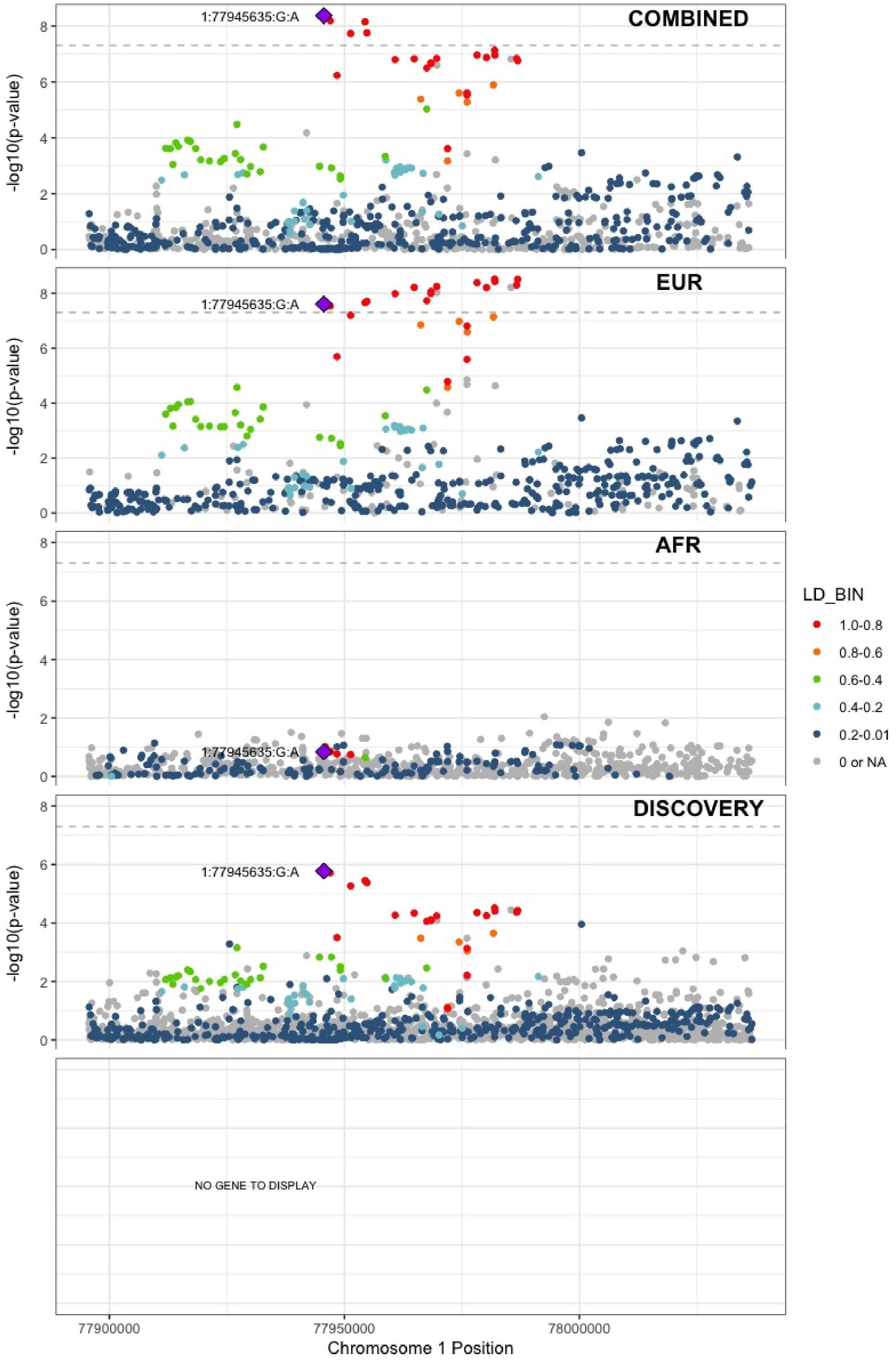

**Supplementary Figure 6.4** – rs3832016 (chr1:109818158) in locus 4 (CELSR2)

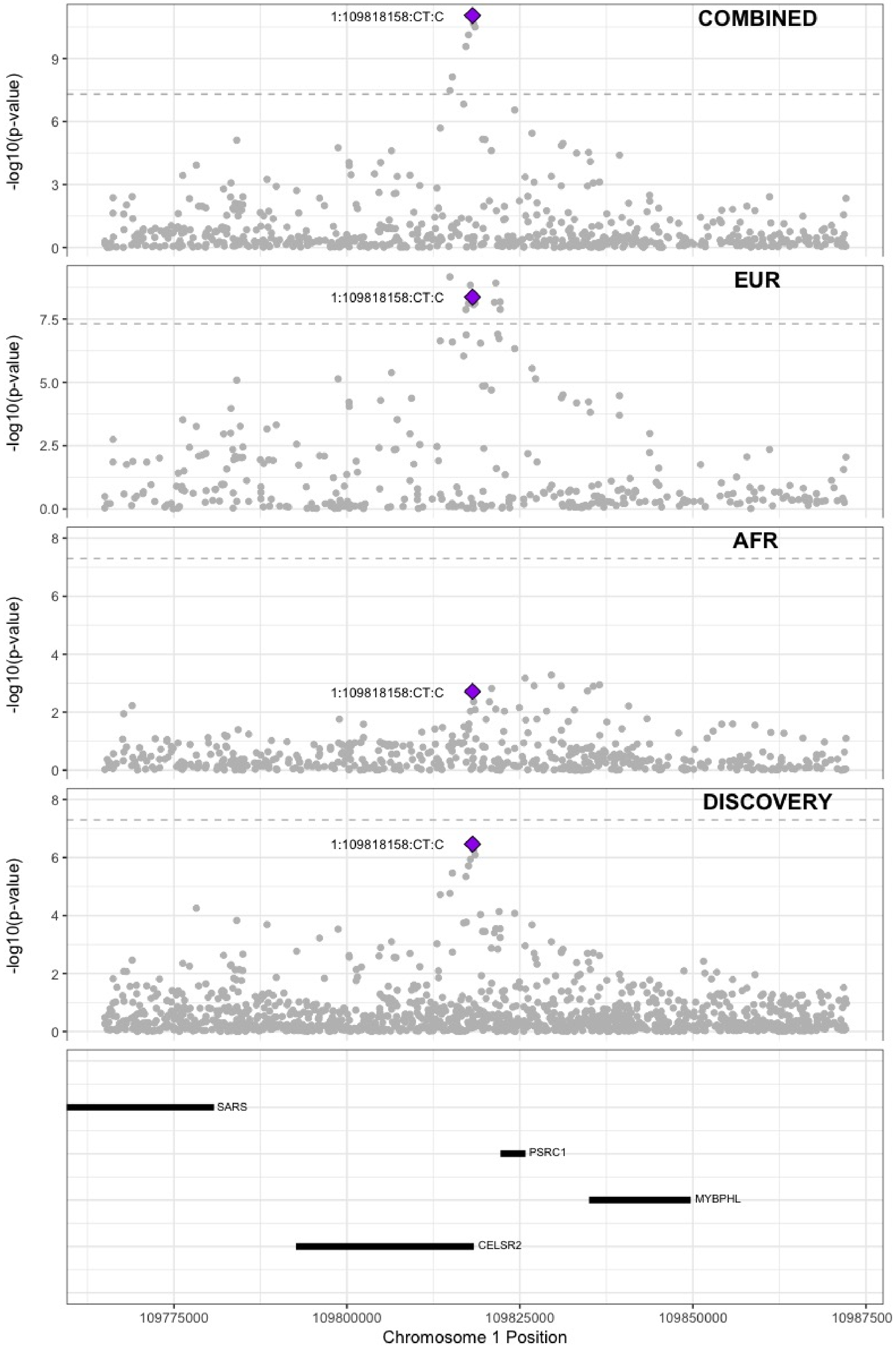

**Supplementary Figure 6.5** – rs3767812 (chr1:118155620) in locus 5 (TENT5C)

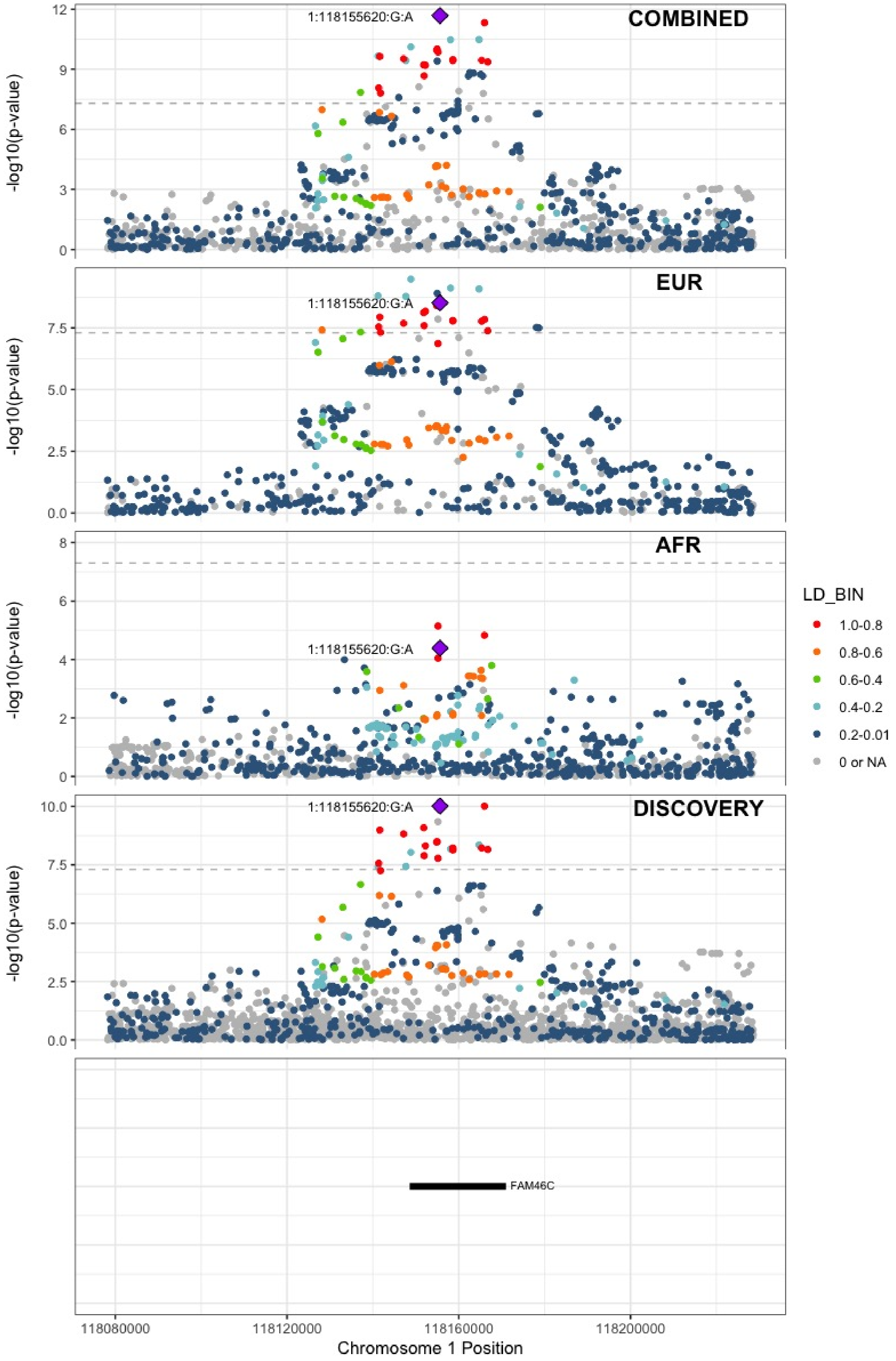

**Supplementary Figure 6.6** – rs1267881263 (chr1:150496127) in locus 6 (FALEC;ADAMTSL4)

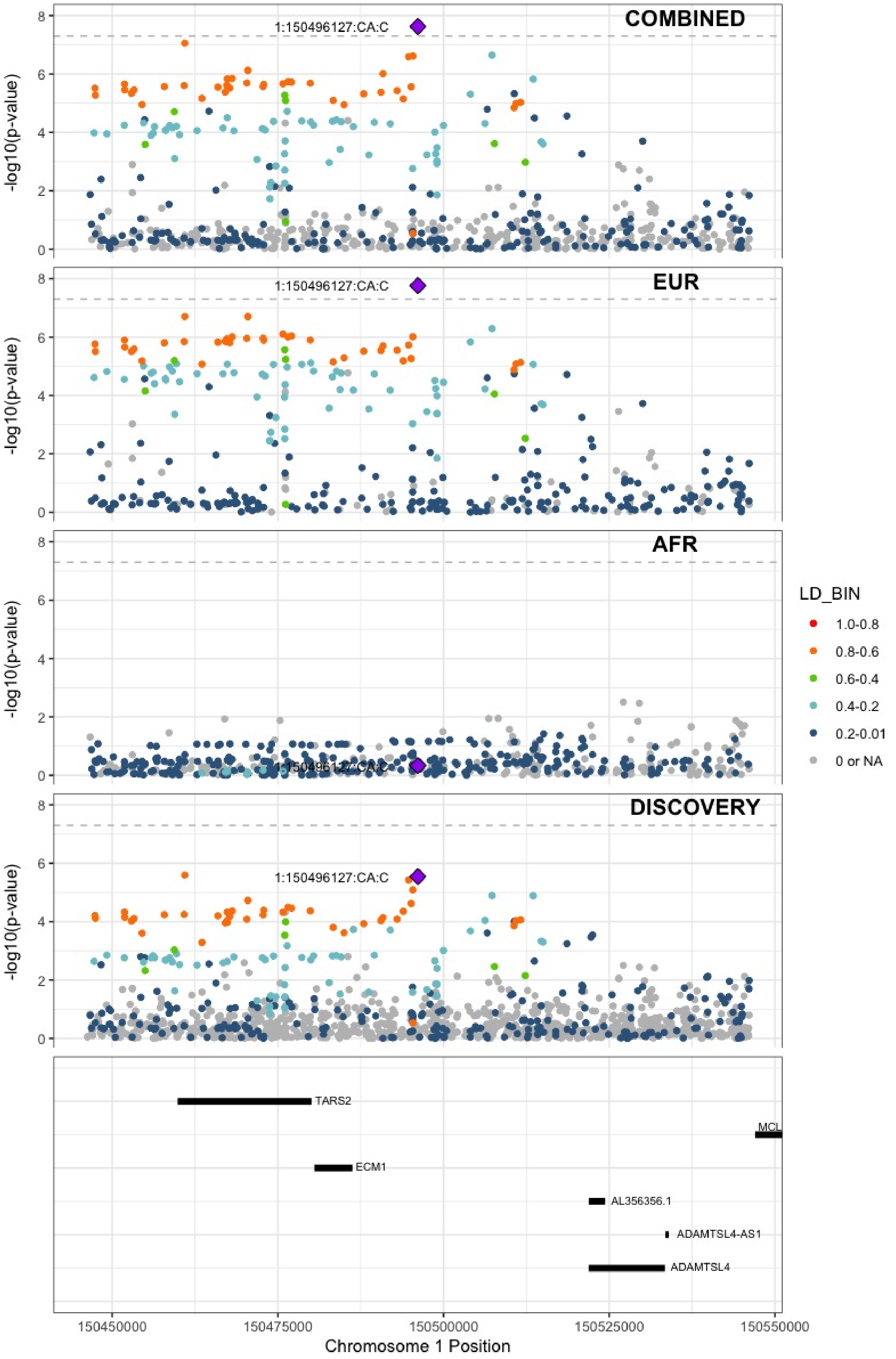

**Supplementary Figure 6.7** – rs905938 (chr1:154991389) in locus 7 (DCST2)

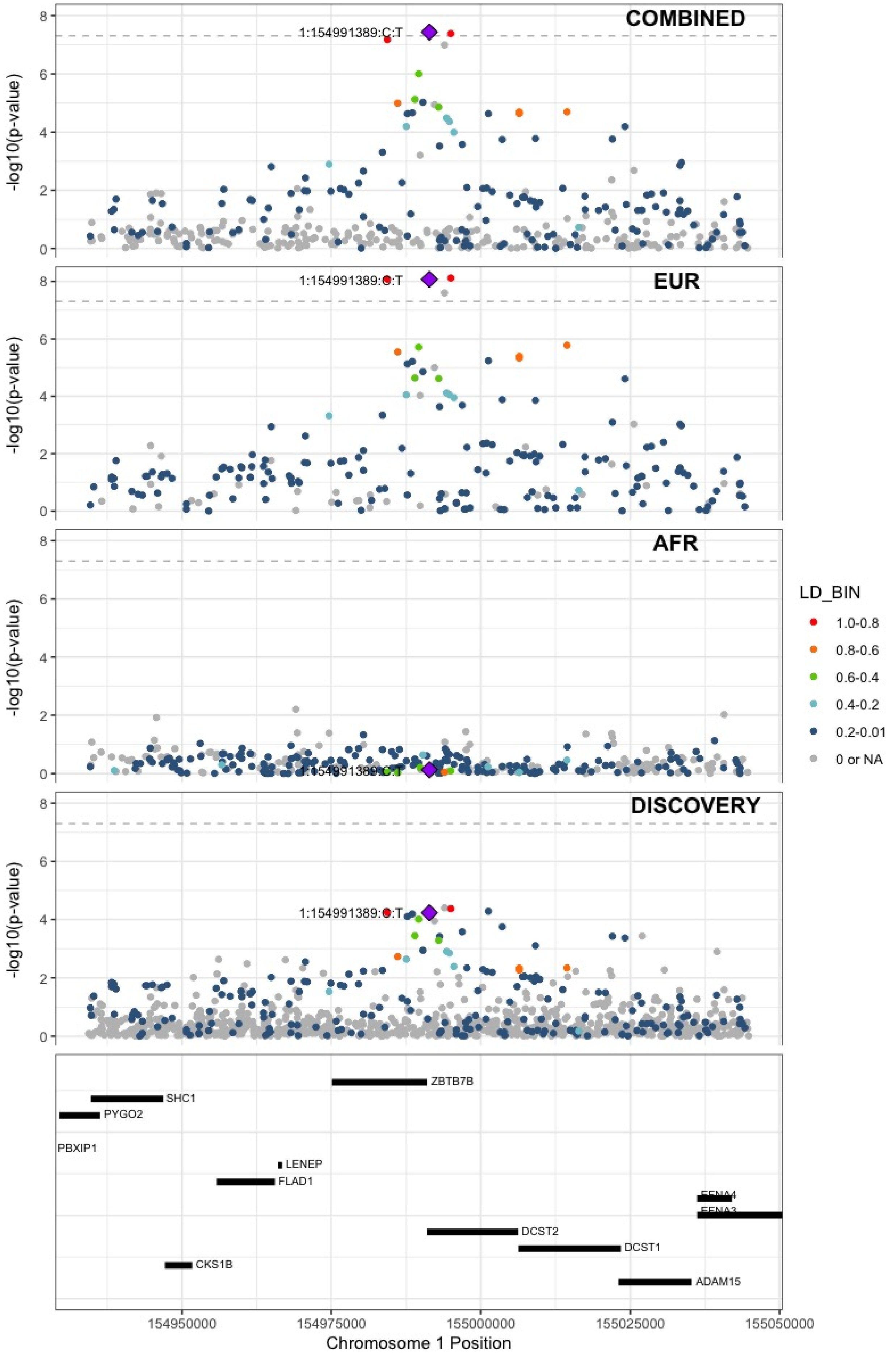

**Supplementary Figure 6.8** – rs3557 (chr1:161188893) in locus 8 (FCER1G)

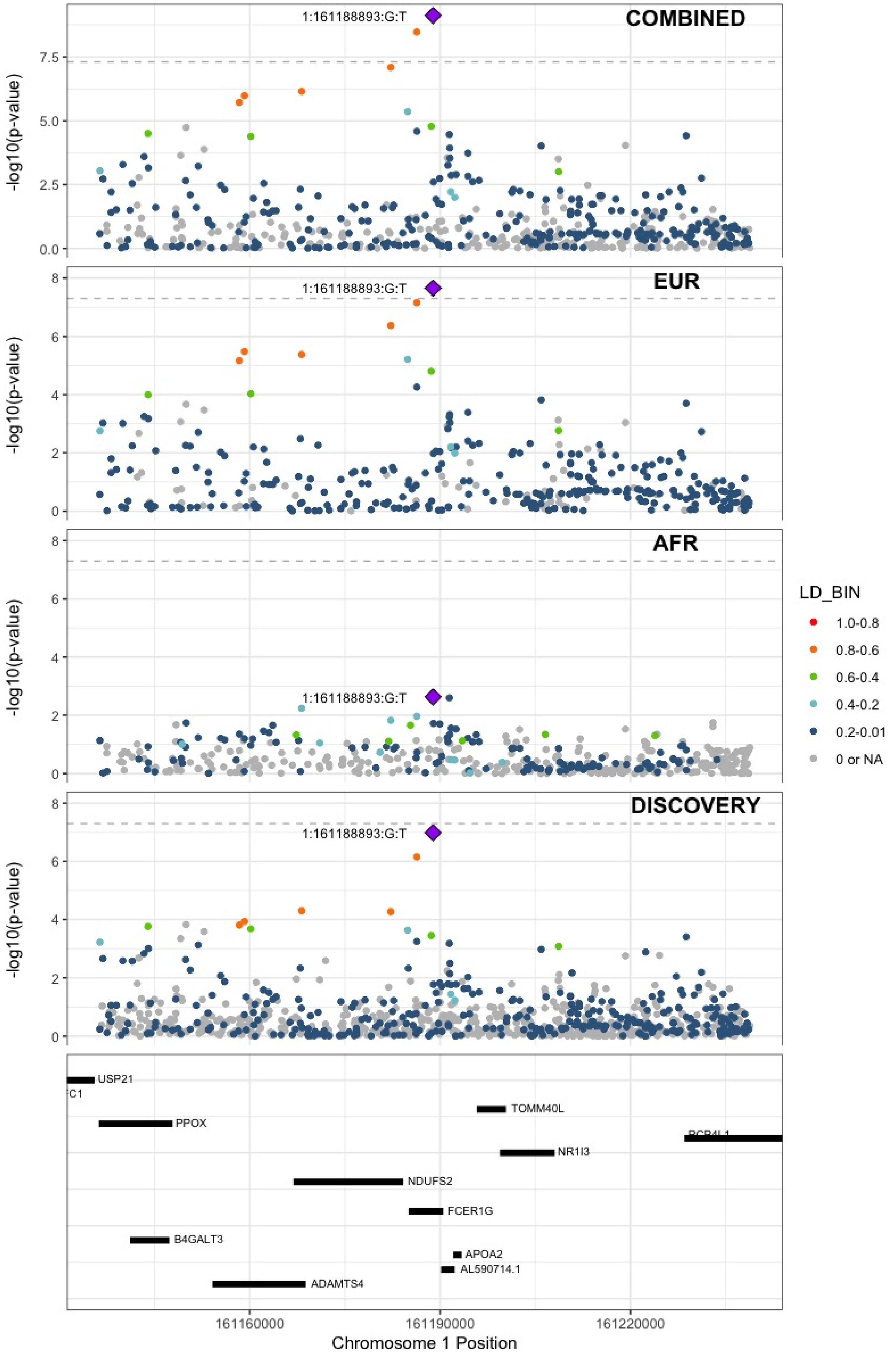

**Supplementary Figure 6.9** – rs6025 (chr1:169519049) in locus 9 (F5)

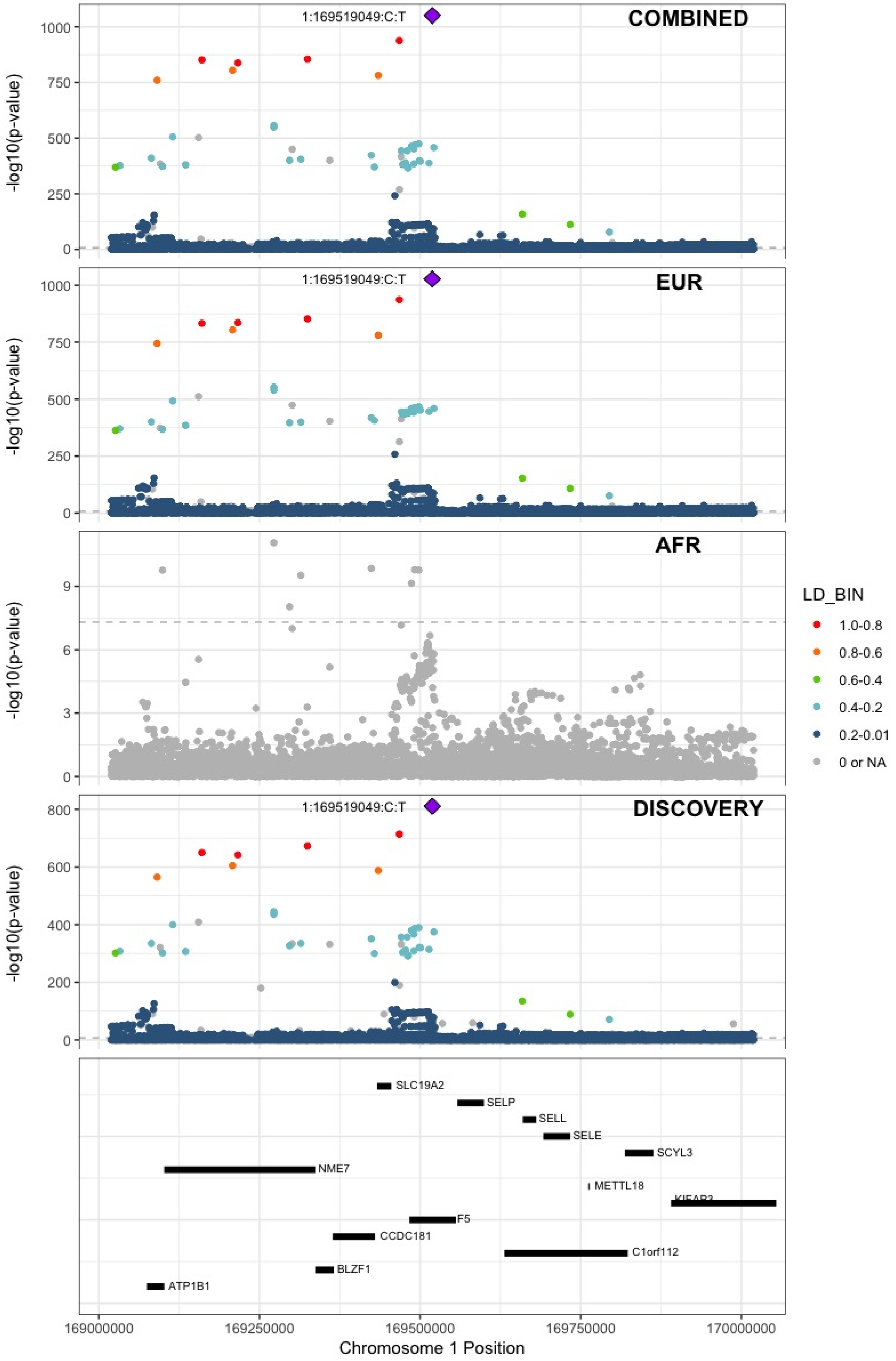

**Supplementary Figure 6.10** – rs6695940 (chr1:173882980) in locus 10 (SERPINC1)

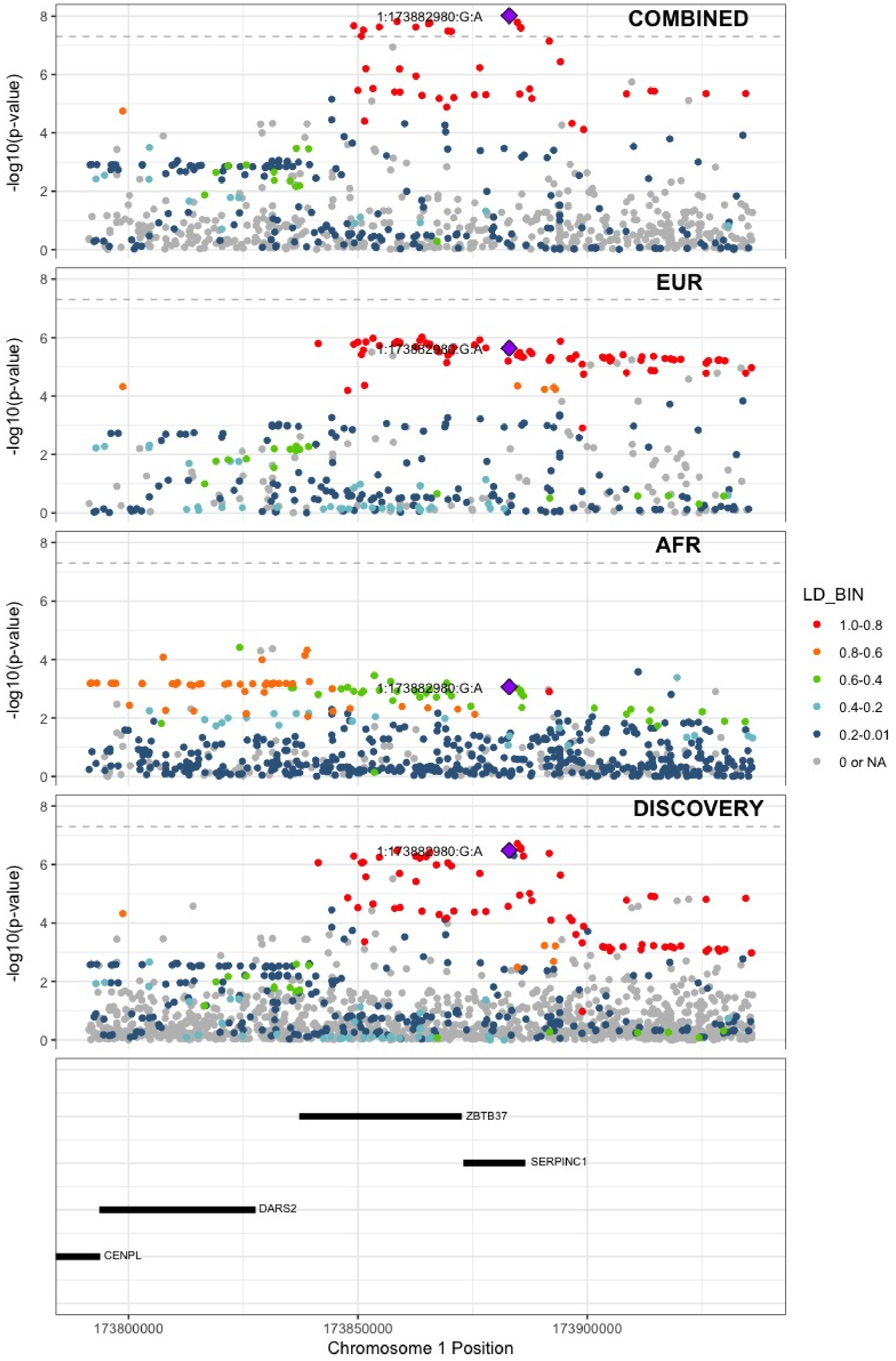

**Supplementary Figure 6.11** – rs143410348 (chr1:196809316) in locus 12 (CFHR1;CFHR4)

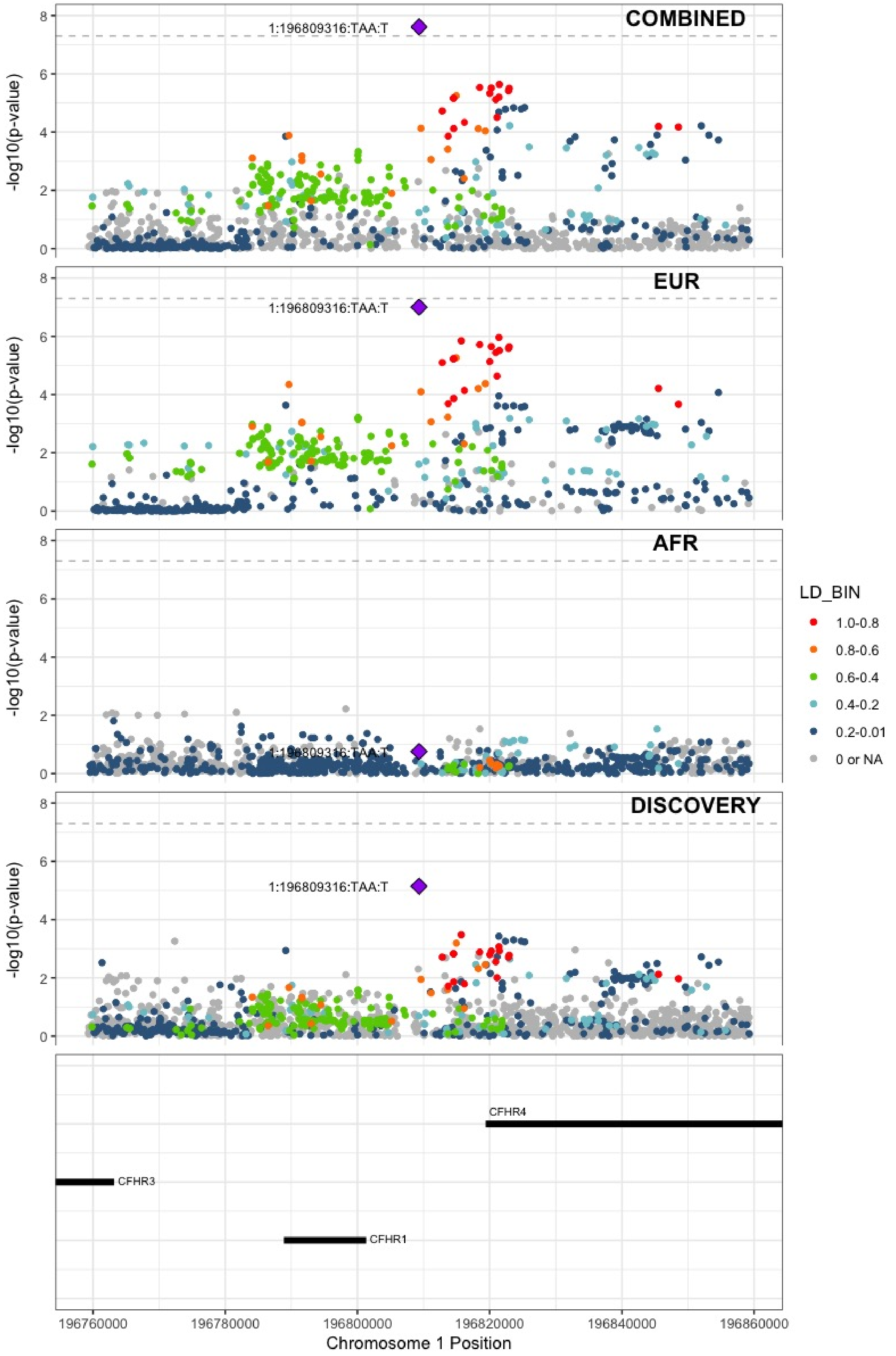

**Supplementary Figure 6.12** – rs2842700 (chr1:207282149) in locus 13 (C4BPA)

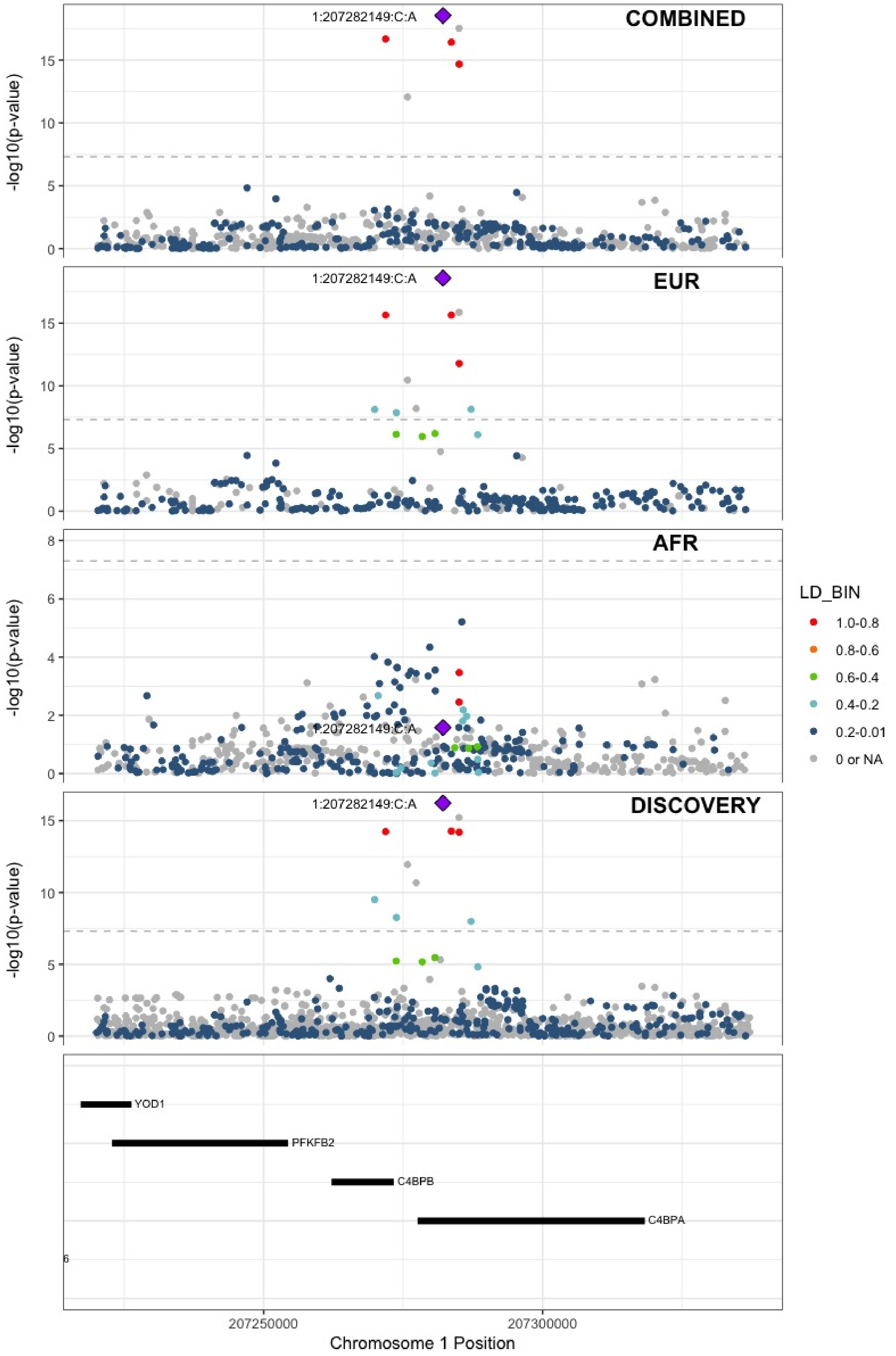

**Supplementary Figure 6.13** – rs3811444 (chr1:248039451) in locus 14 (TRIM58)

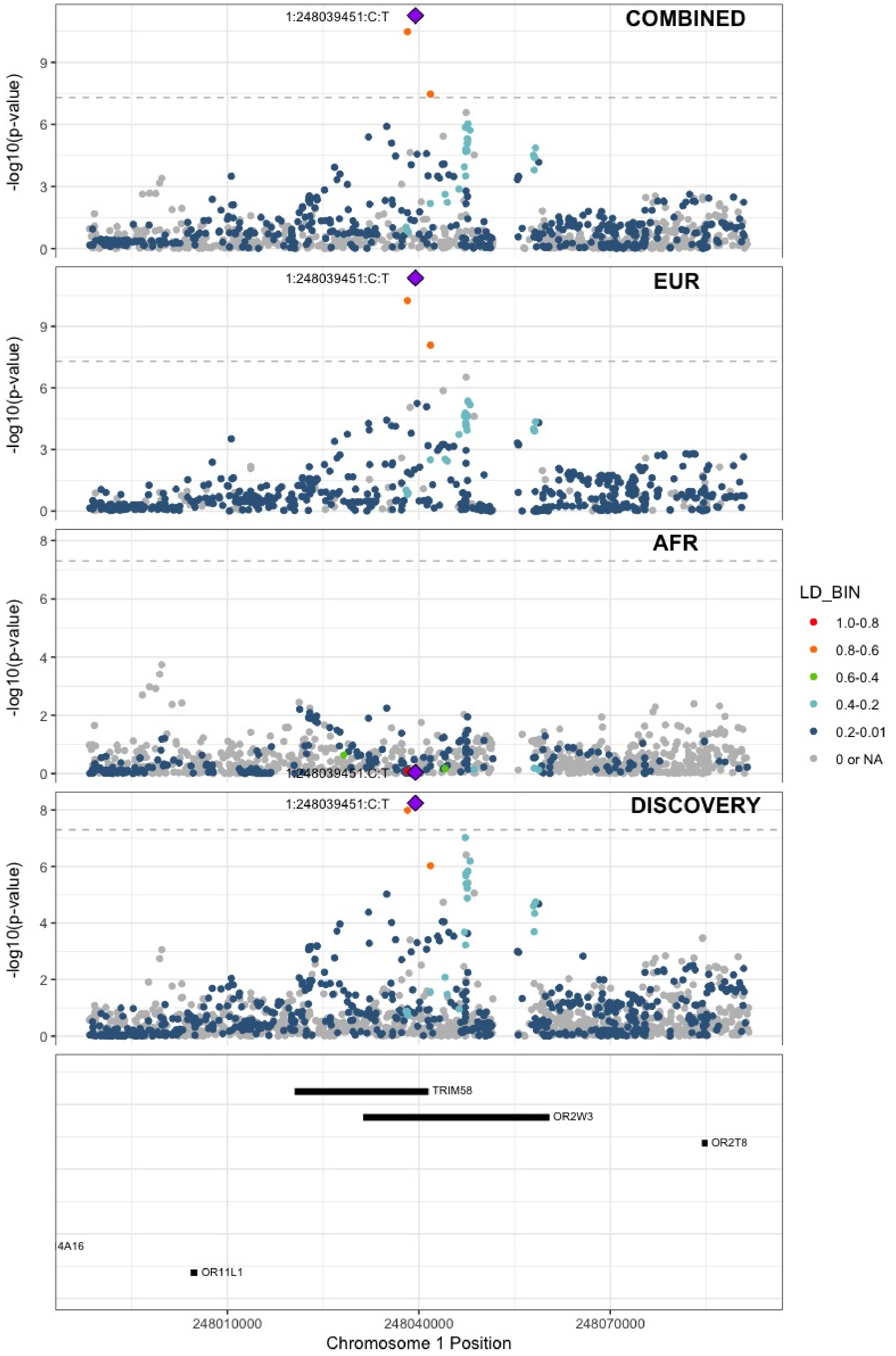

**Supplementary Figure 6.14** – rs78475244 (chr2:65086804) in locus 16 (LINC01800)

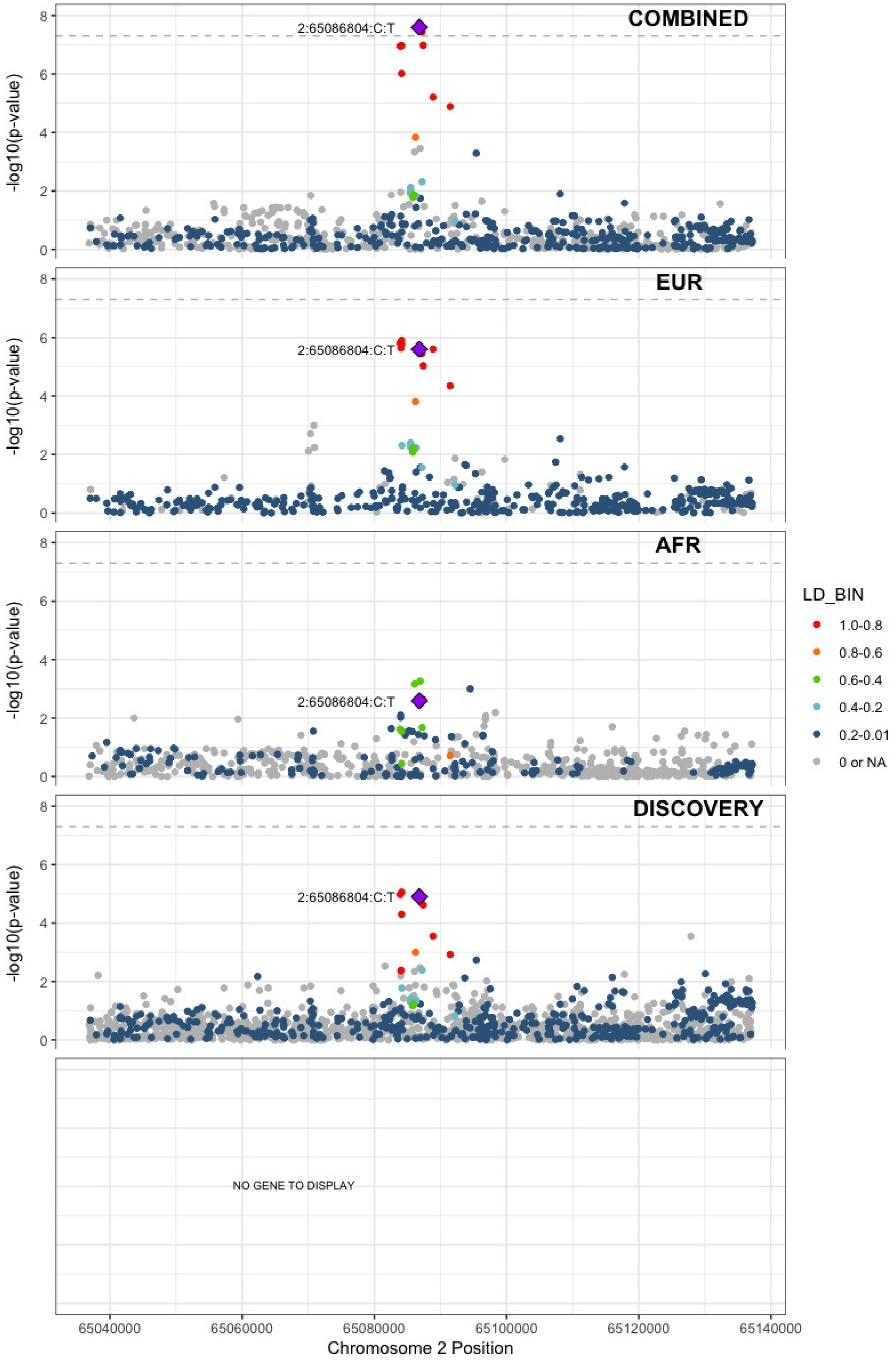

**Supplementary Figure 6.15** – rs1867312 (chr2:68619981) in locus 17 (PLEK)

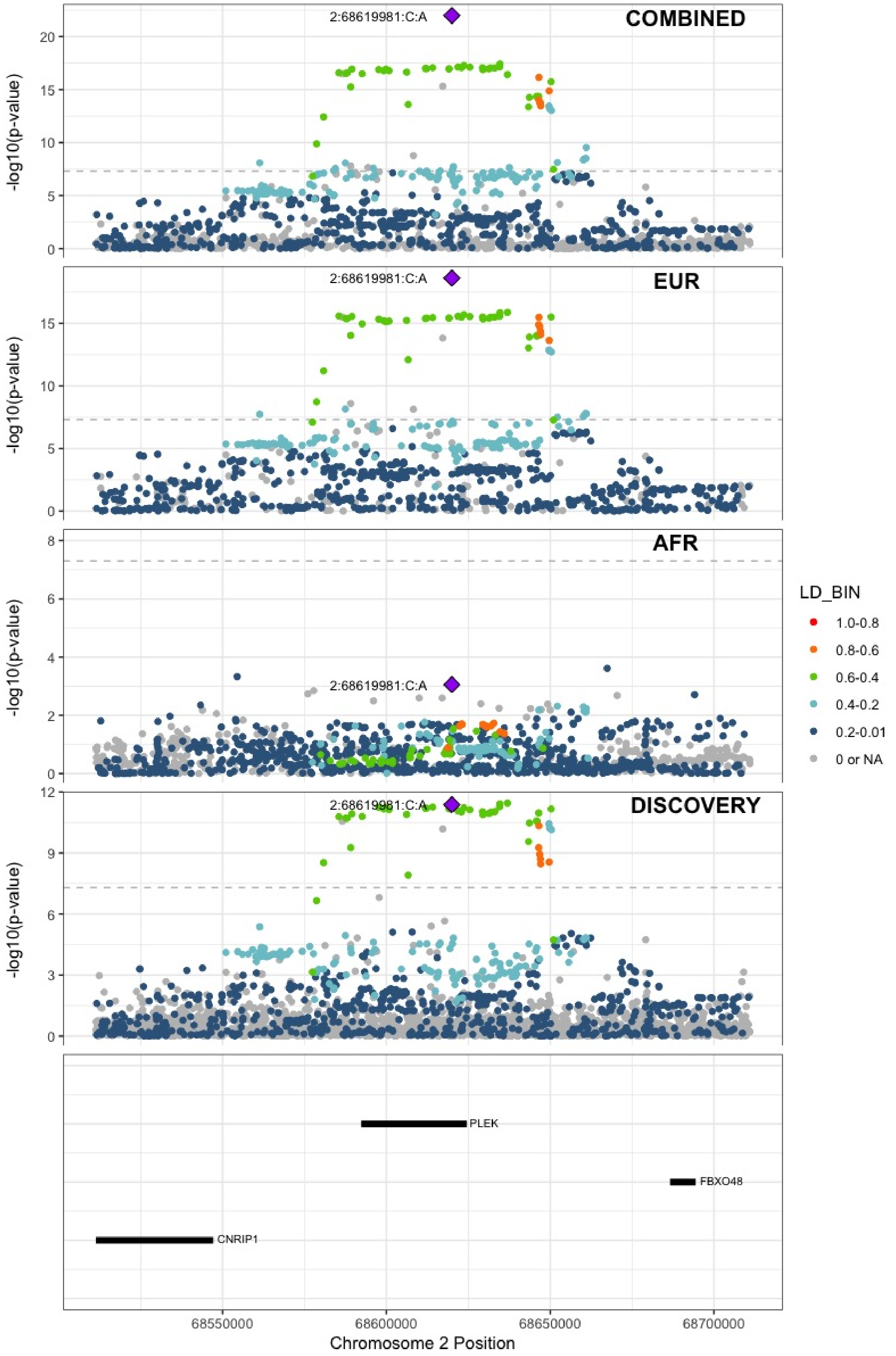

**Supplementary Figure 6.16** – rs1518760 (chr2:128140500) in locus 18 (MAP3K2)

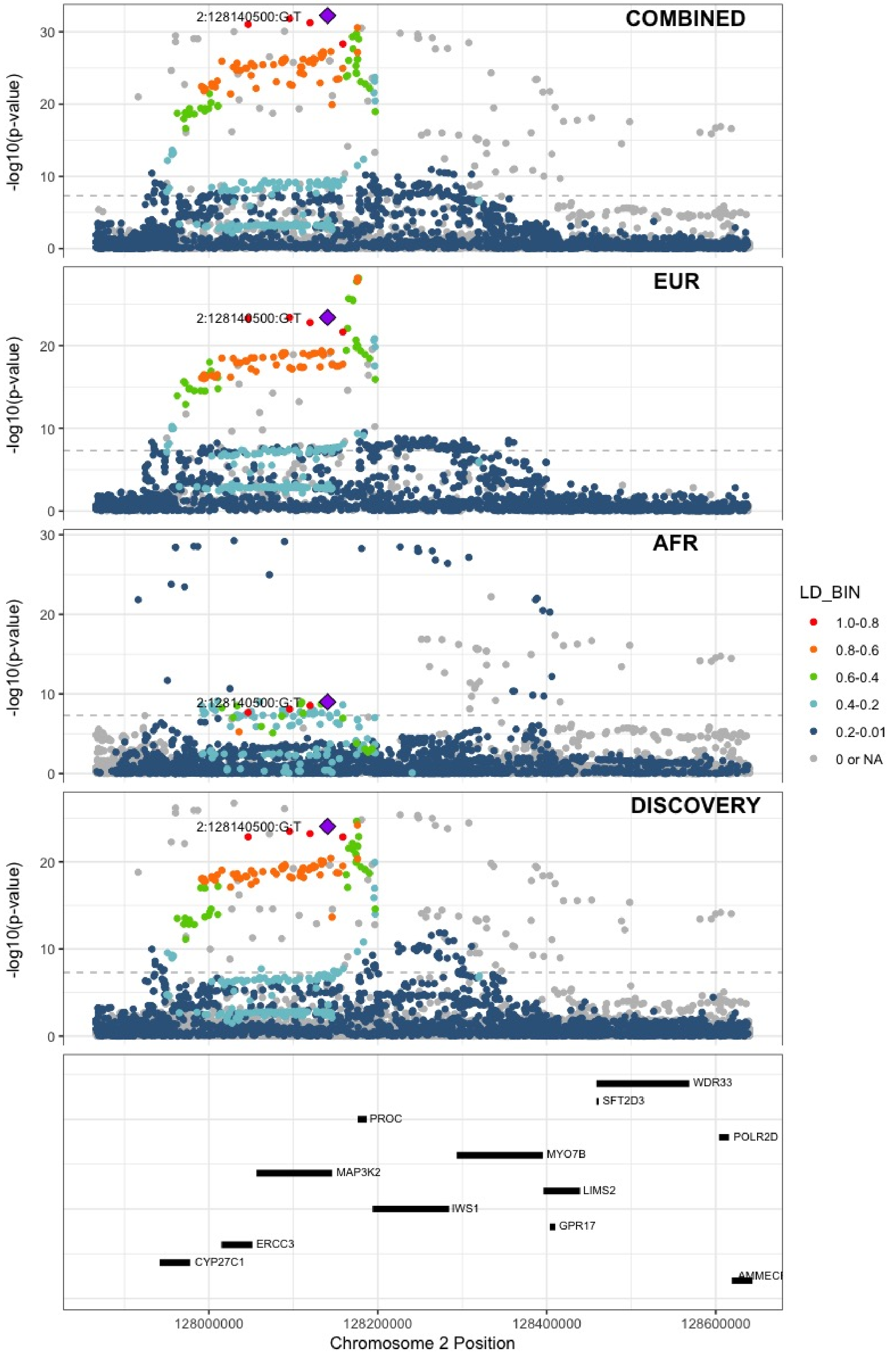

**Supplementary Figure 6.17** – rs1912849 (chr2:188310118) in locus 19 (CALCRL)

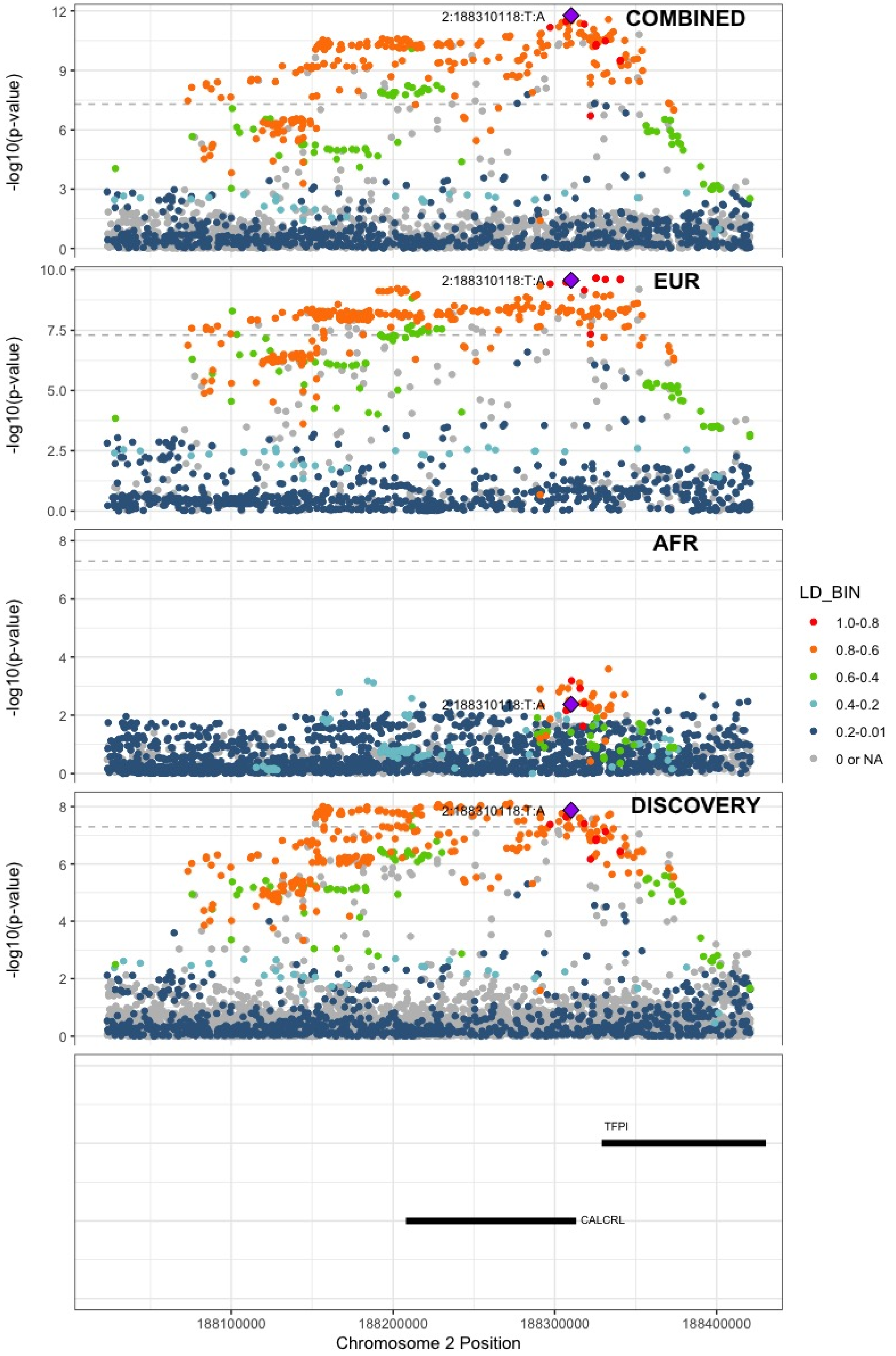

**Supplementary Figure 6.18** – rs78872368 (chr2:198545250) in locus 20 (RFTN2;MARS2)

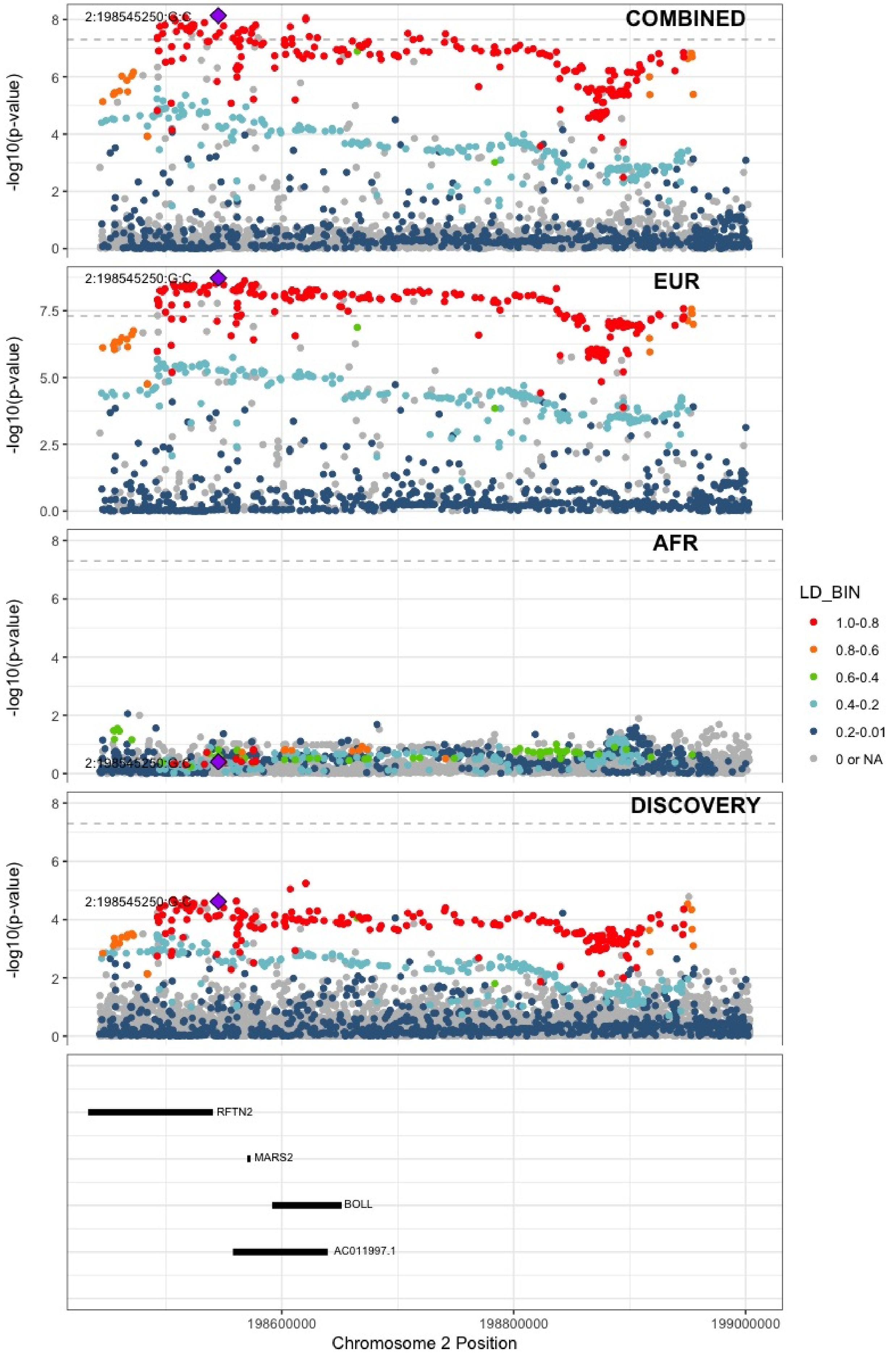

**Supplementary Figure 6.19** – rs715 (chr2:211543055) in locus 21 (CPS1)

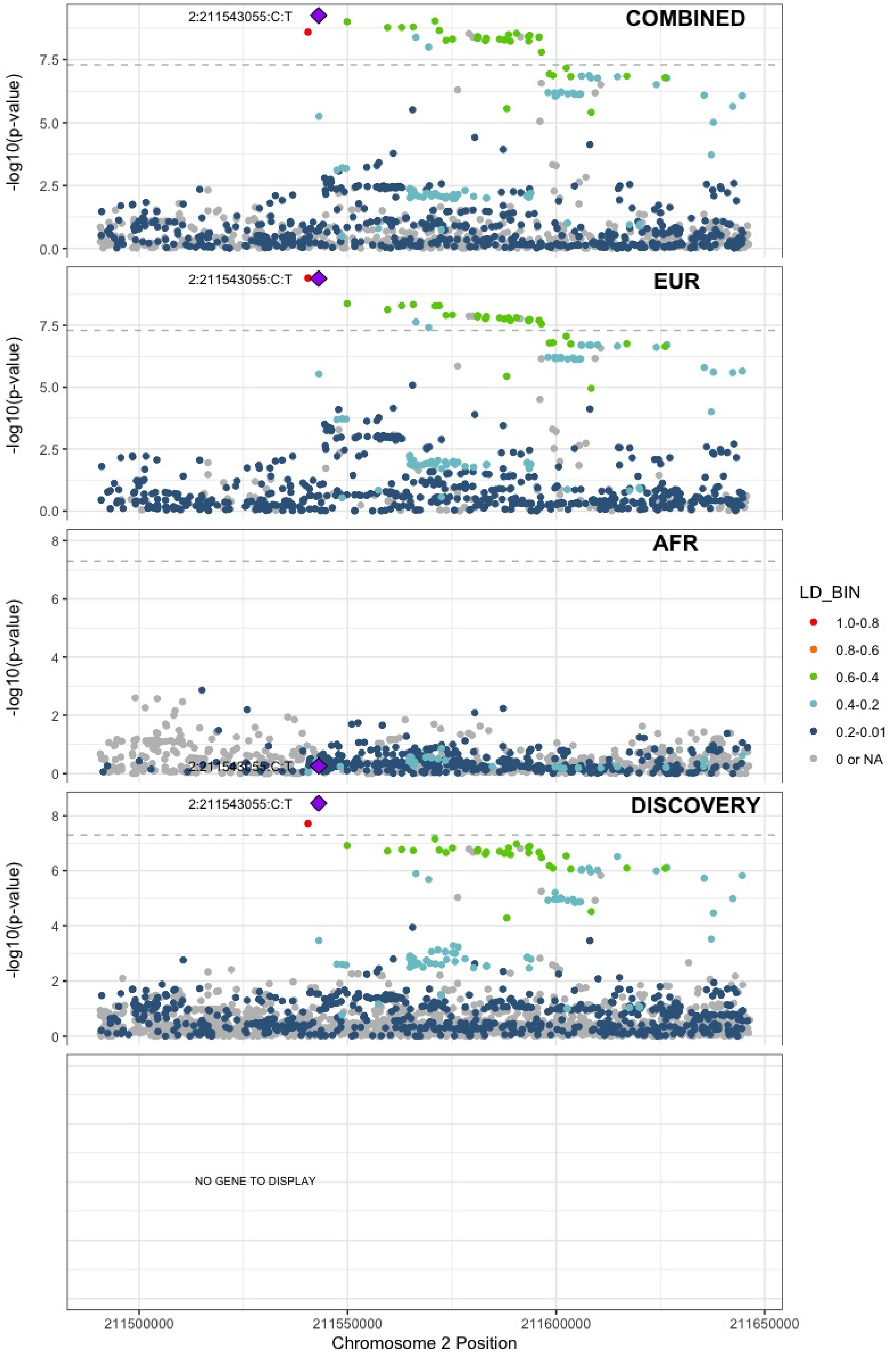

**Supplementary Figure 6.20** – rs68066031 (chr2:224880498) in locus 22 (SERPINE2)

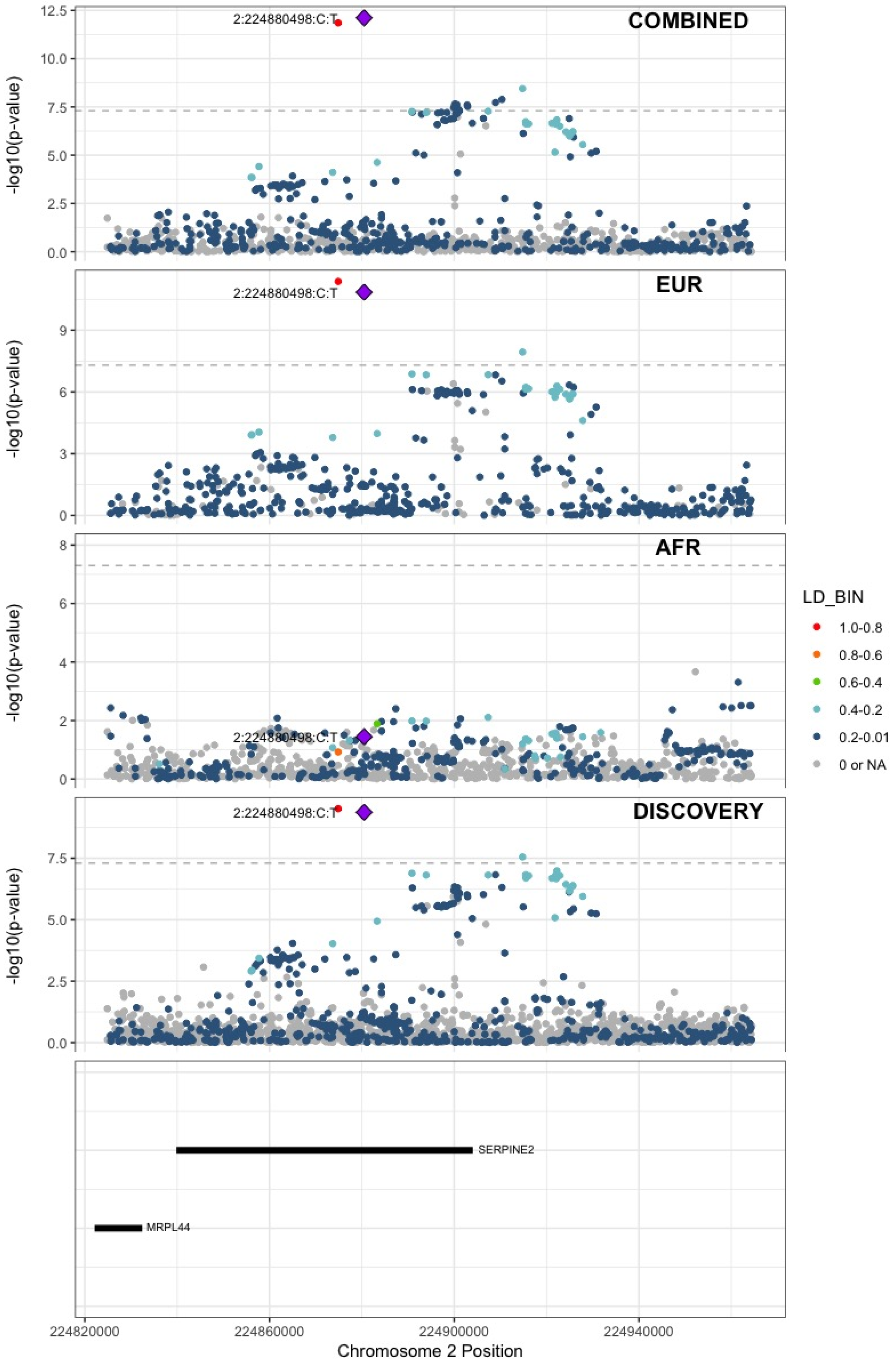

**Supplementary Figure 6.21** – rs13084580 (chr3:39188182) in locus 24 (CSRNP1)

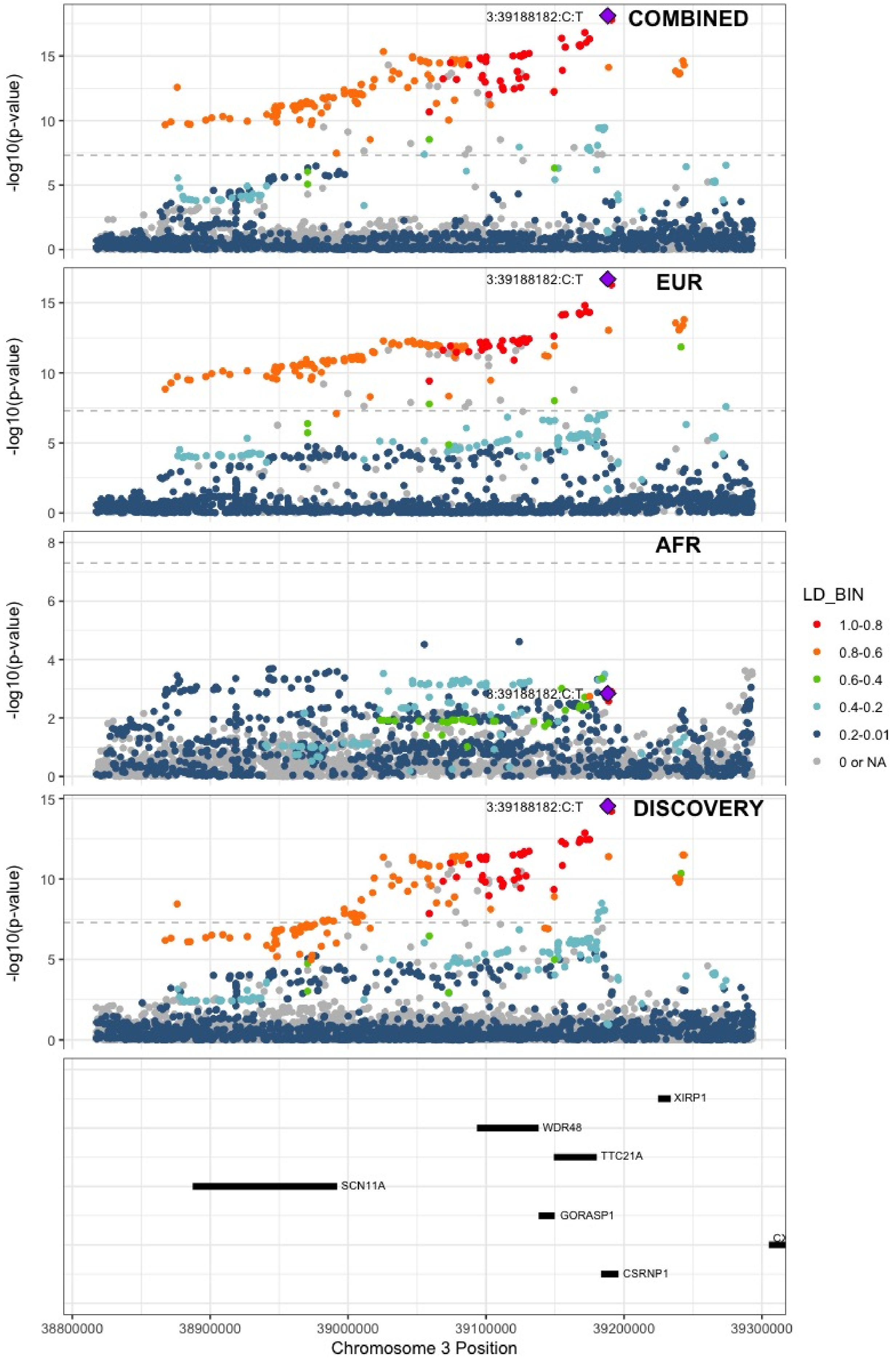

**Supplementary Figure 6.22** – rs11130326 (chr3:52771920) in locus 25 (NEK4)

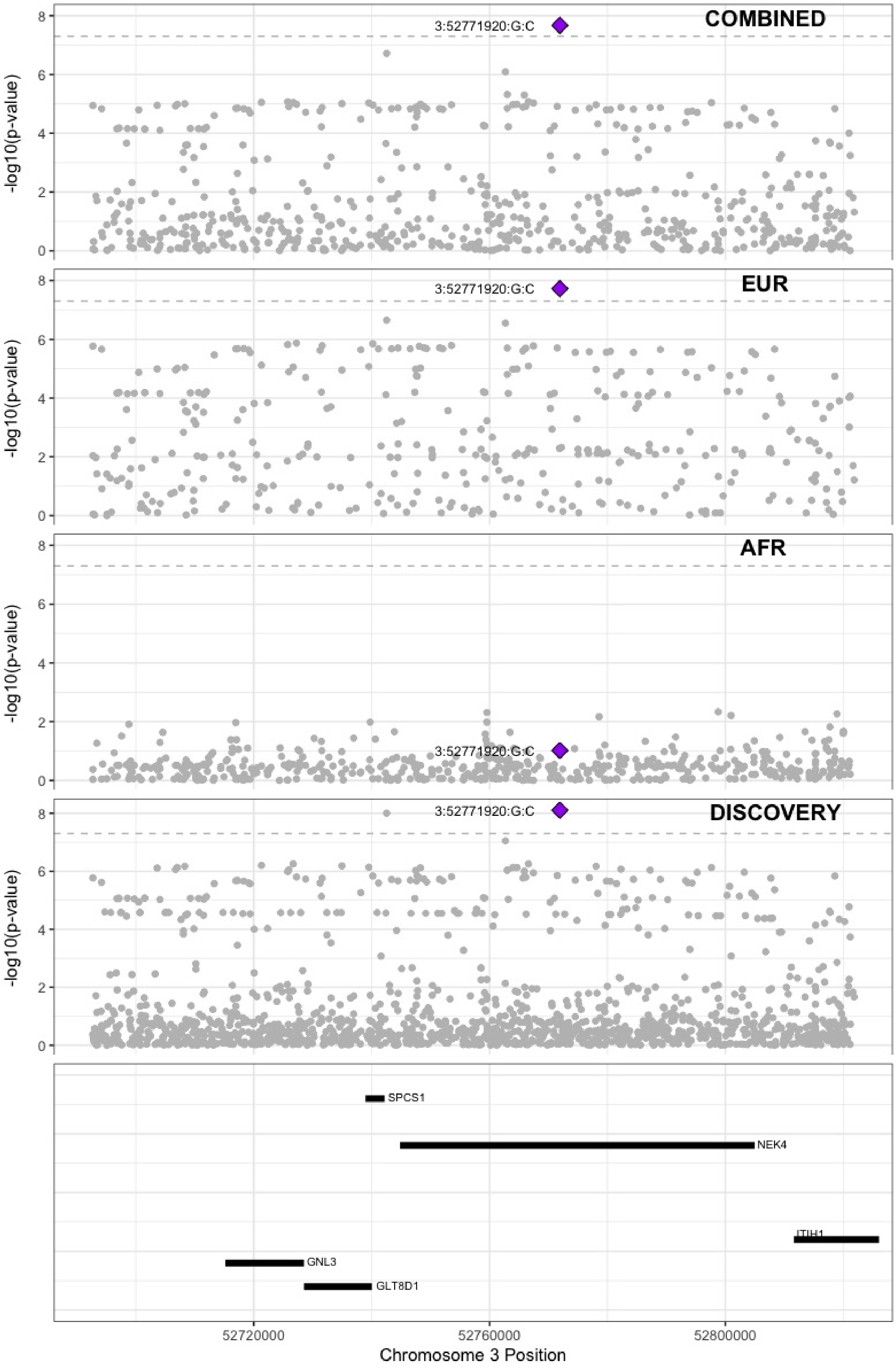

**Supplementary Figure 6.23** – rs529326506 (chr3:90503764) in locus 26 (EPHA3;NONE)

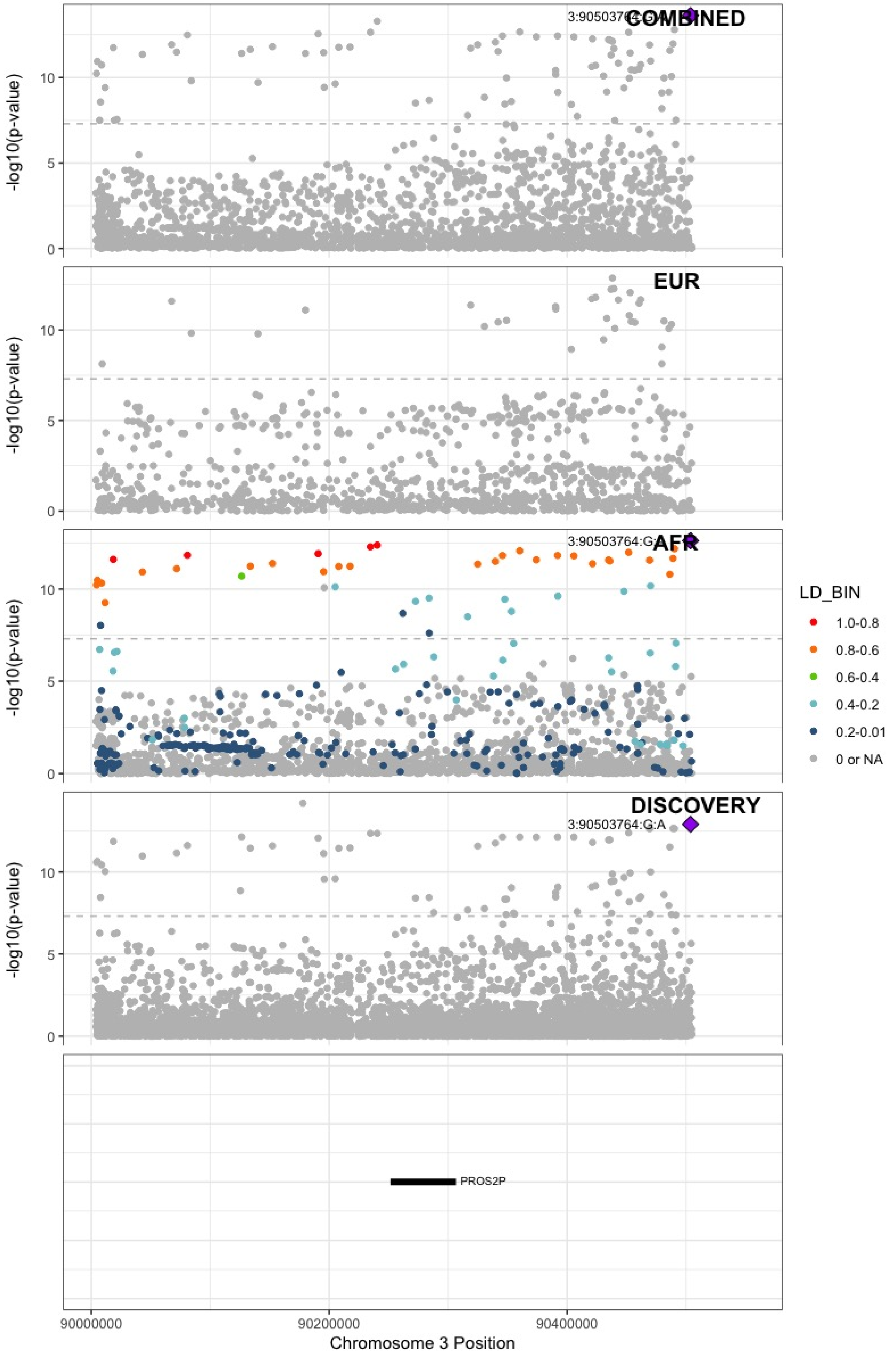

**Supplementary Figure 6.24** – rs28479320 (chr3:93752529) in locus 27 (ARL13B)

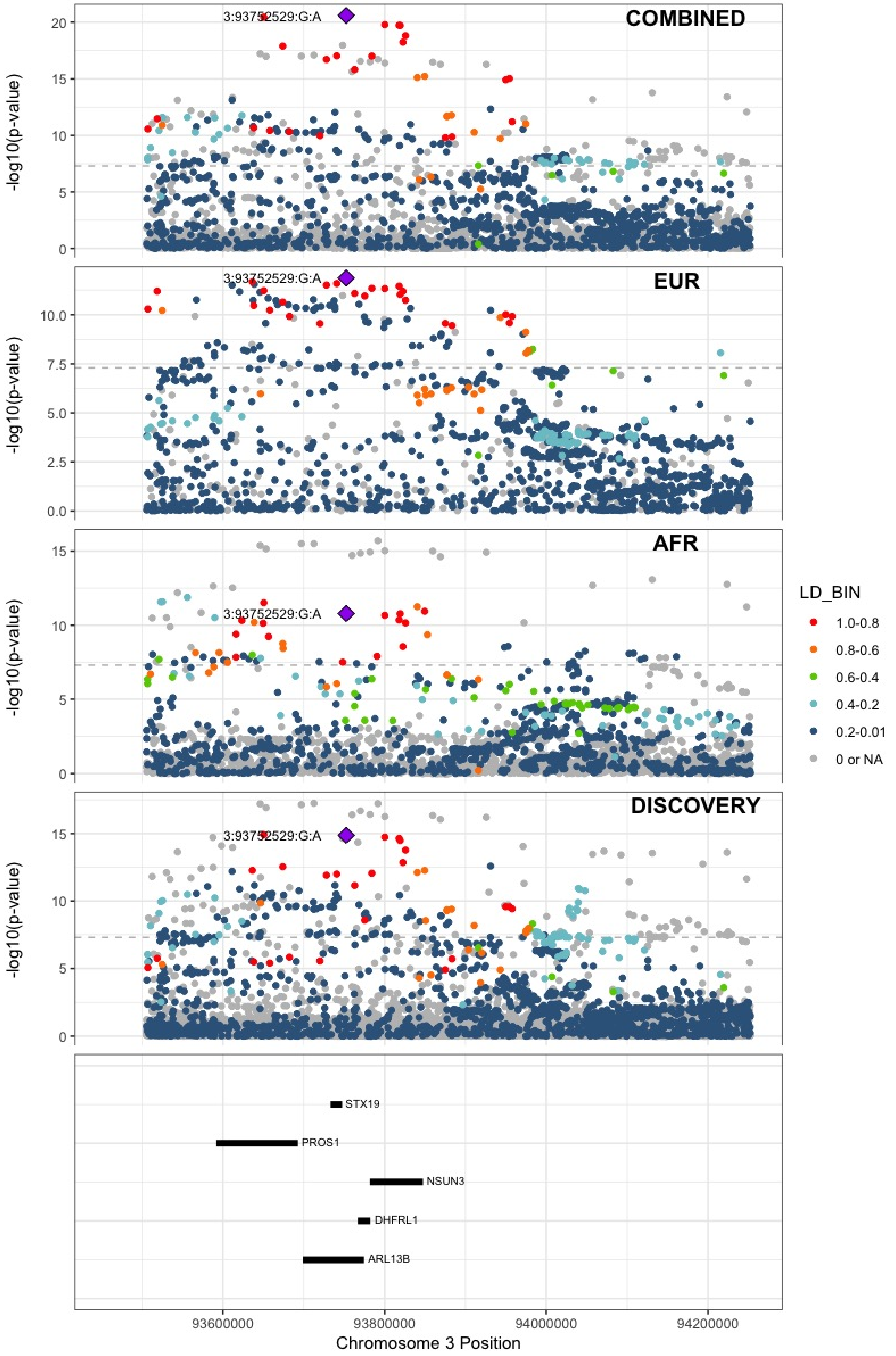

**Supplementary Figure 6.25** – rs900399 (chr3:156798732) in locus 30 (LINC02029;LINC00880)

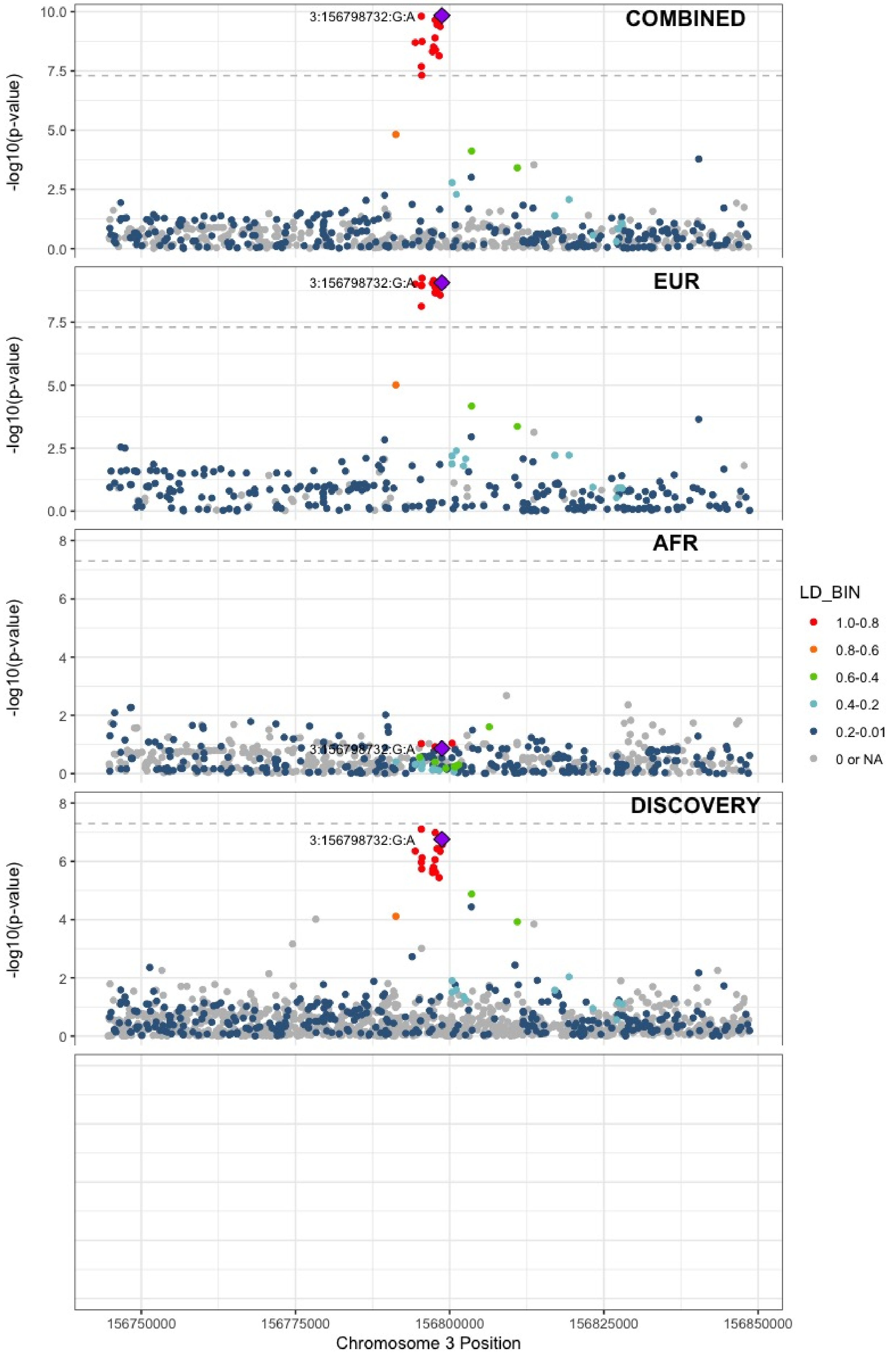

**Supplementary Figure 6.26** – rs710446 (chr3:186459927) in locus 32 (KNG1)

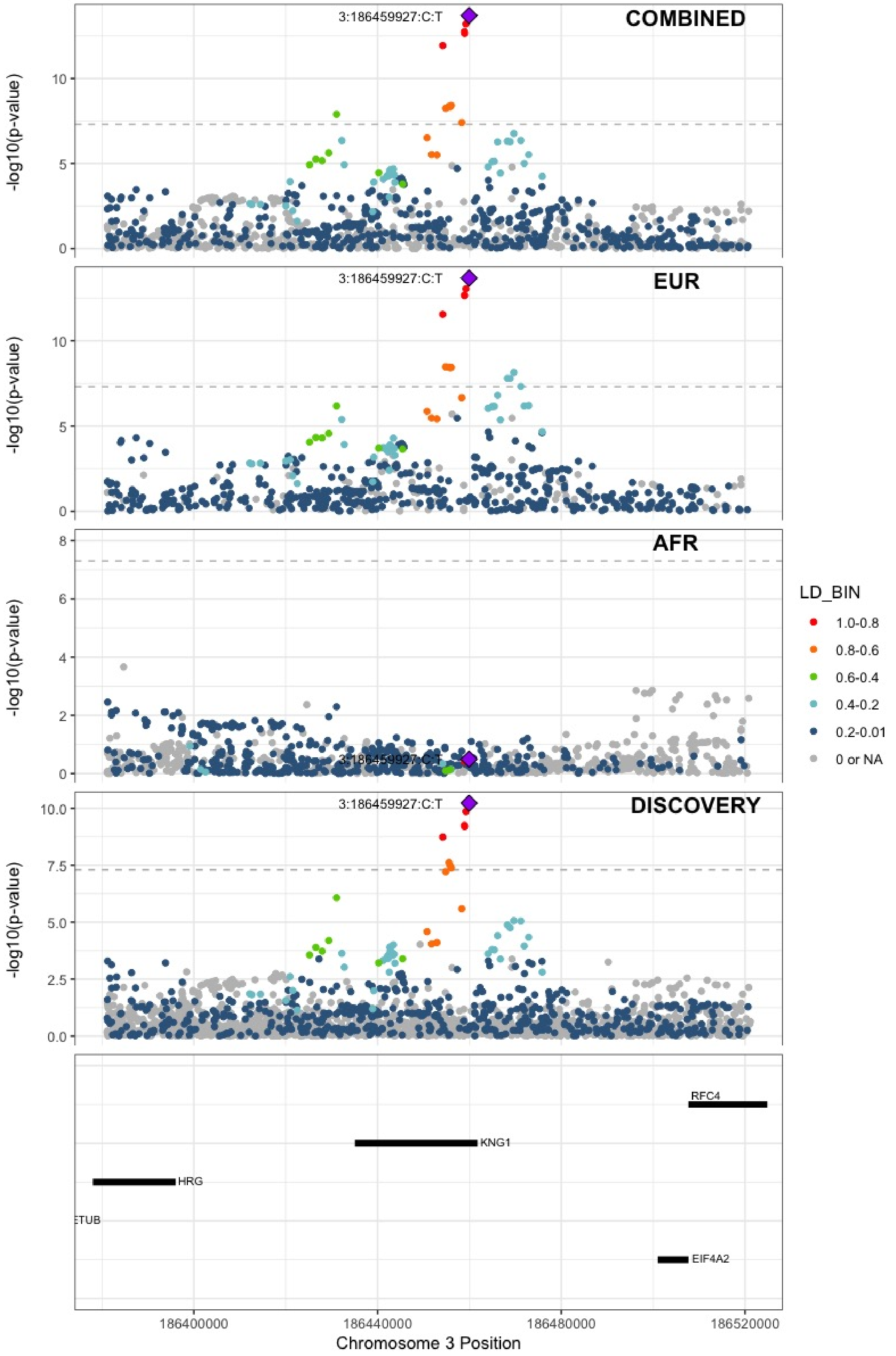

**Supplementary Figure 6.27** – rs3796159 (chr3:194790434) in locus 33 (XXYLT1)

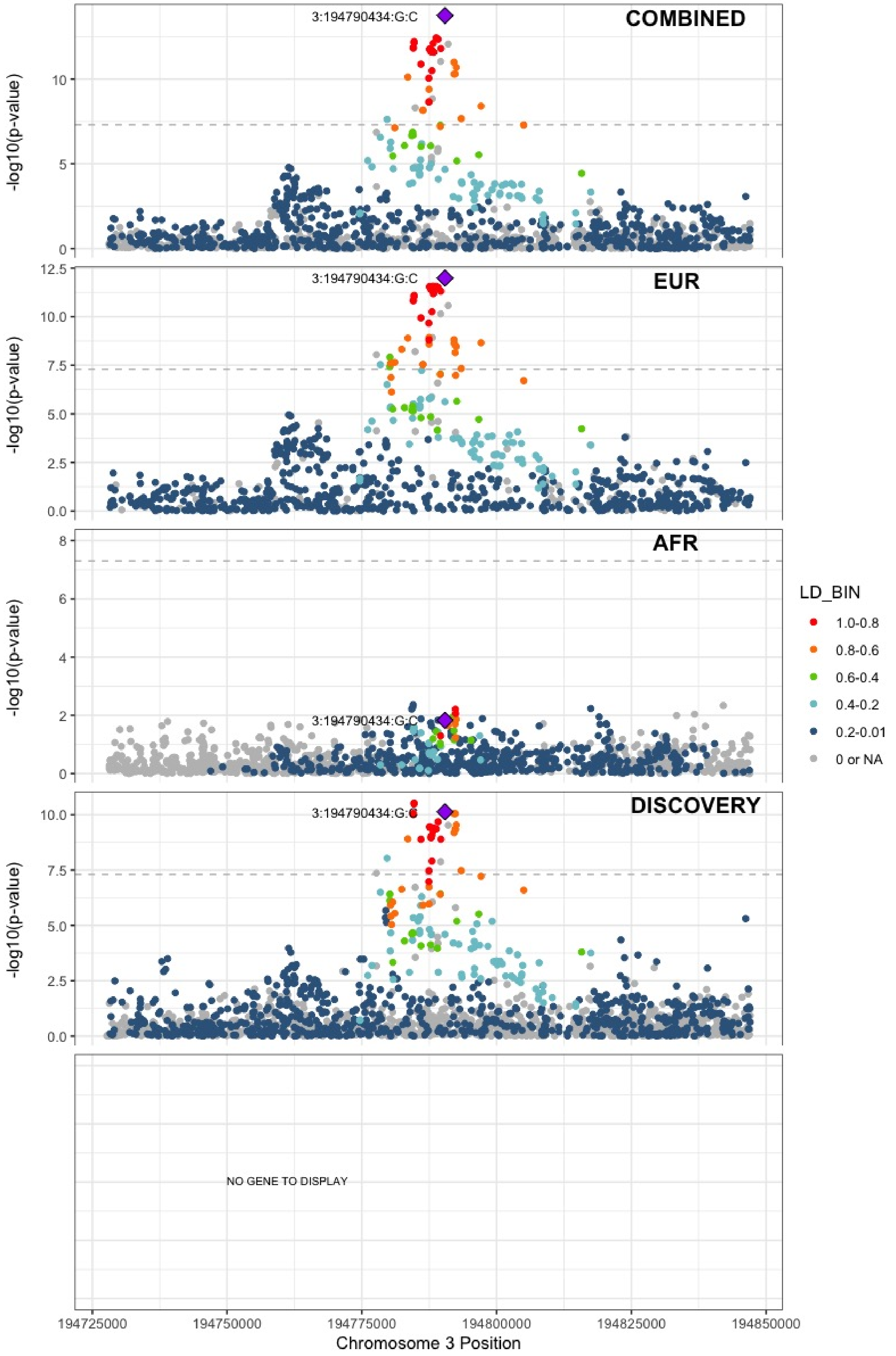

**Supplementary Figure 6.28** – rs9654093 (chr4:7903763) in locus 34 (AFAP1)

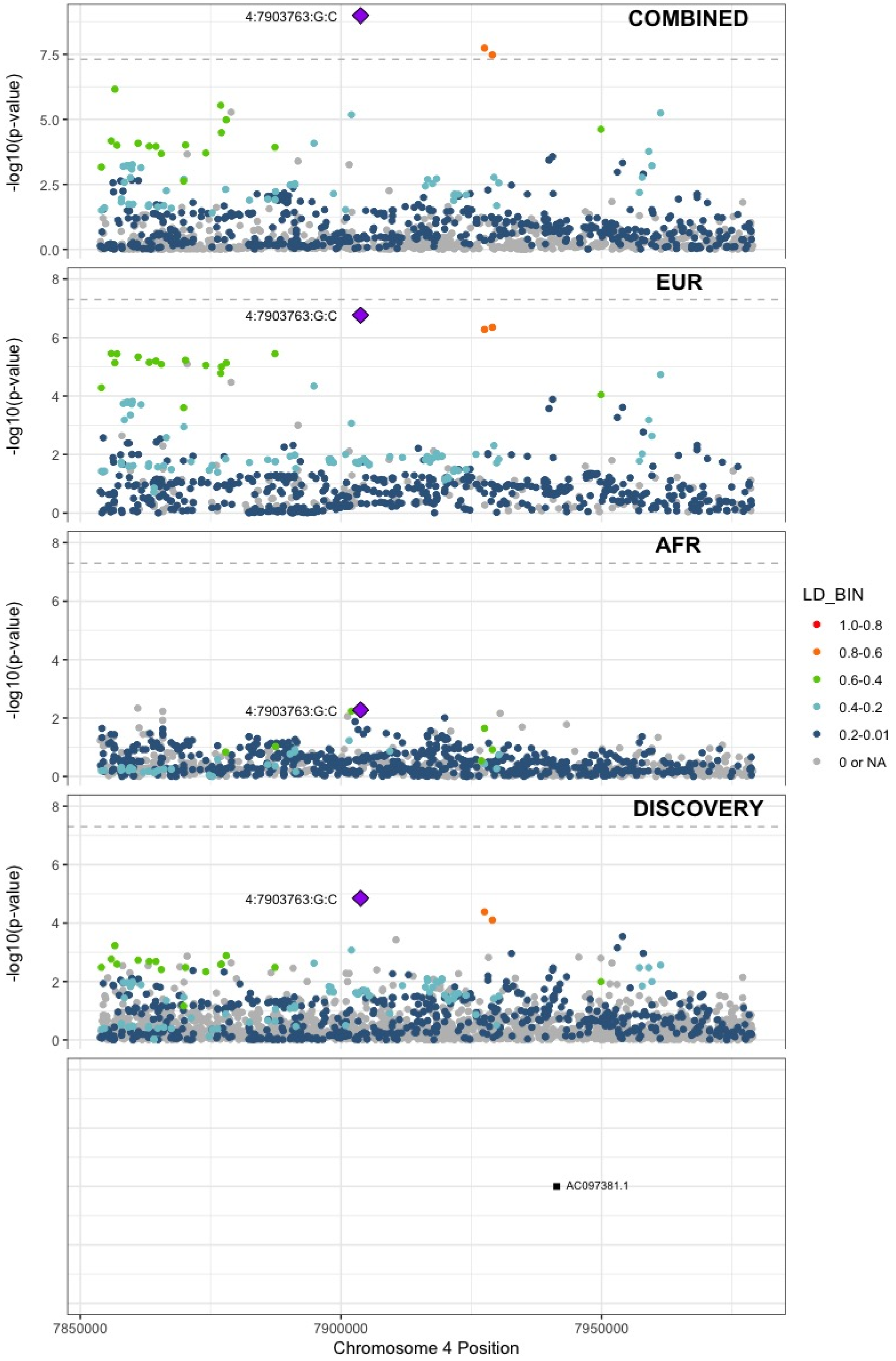

**Supplementary Figure 6.29** – rs781656 (chr4:57778645) in locus 35 (REST)

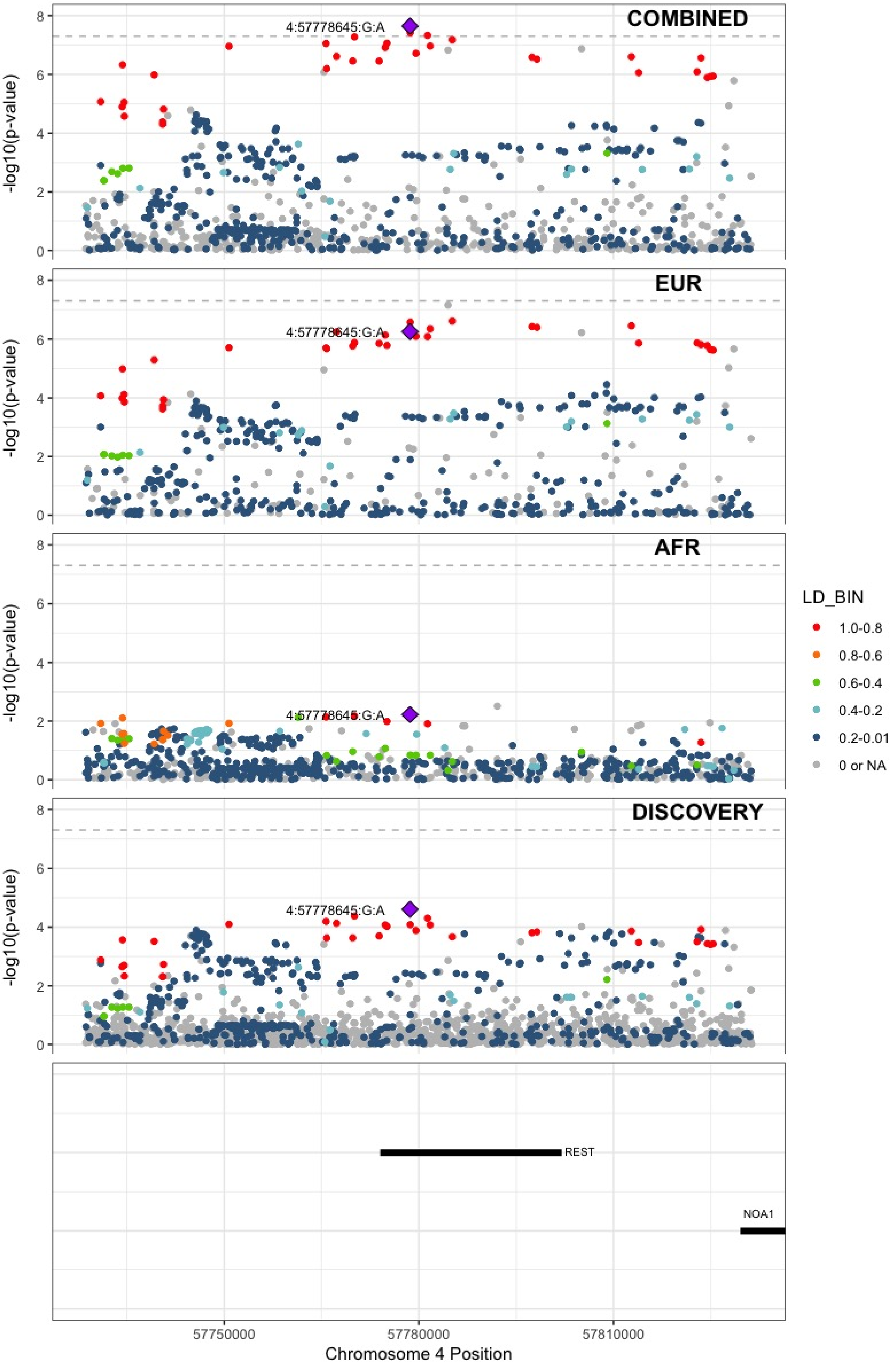

**Supplementary Figure 6.30** – rs6826579 (chr4:83785031) in locus 36 (SEC31A)

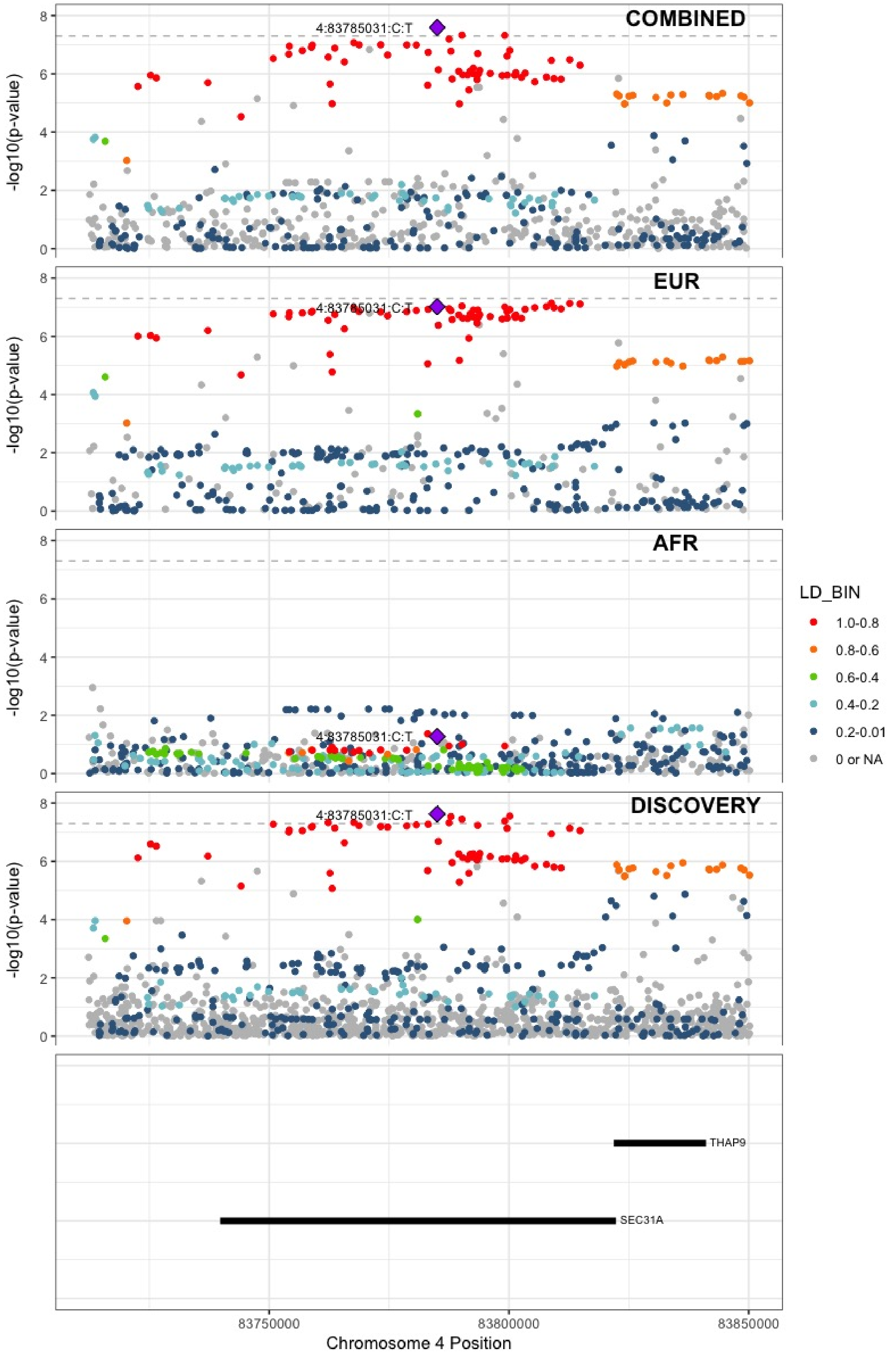

**Supplementary Figure 6.31** – rs2014912 (chr4:86715670) in locus 37 (ARHGAP24)

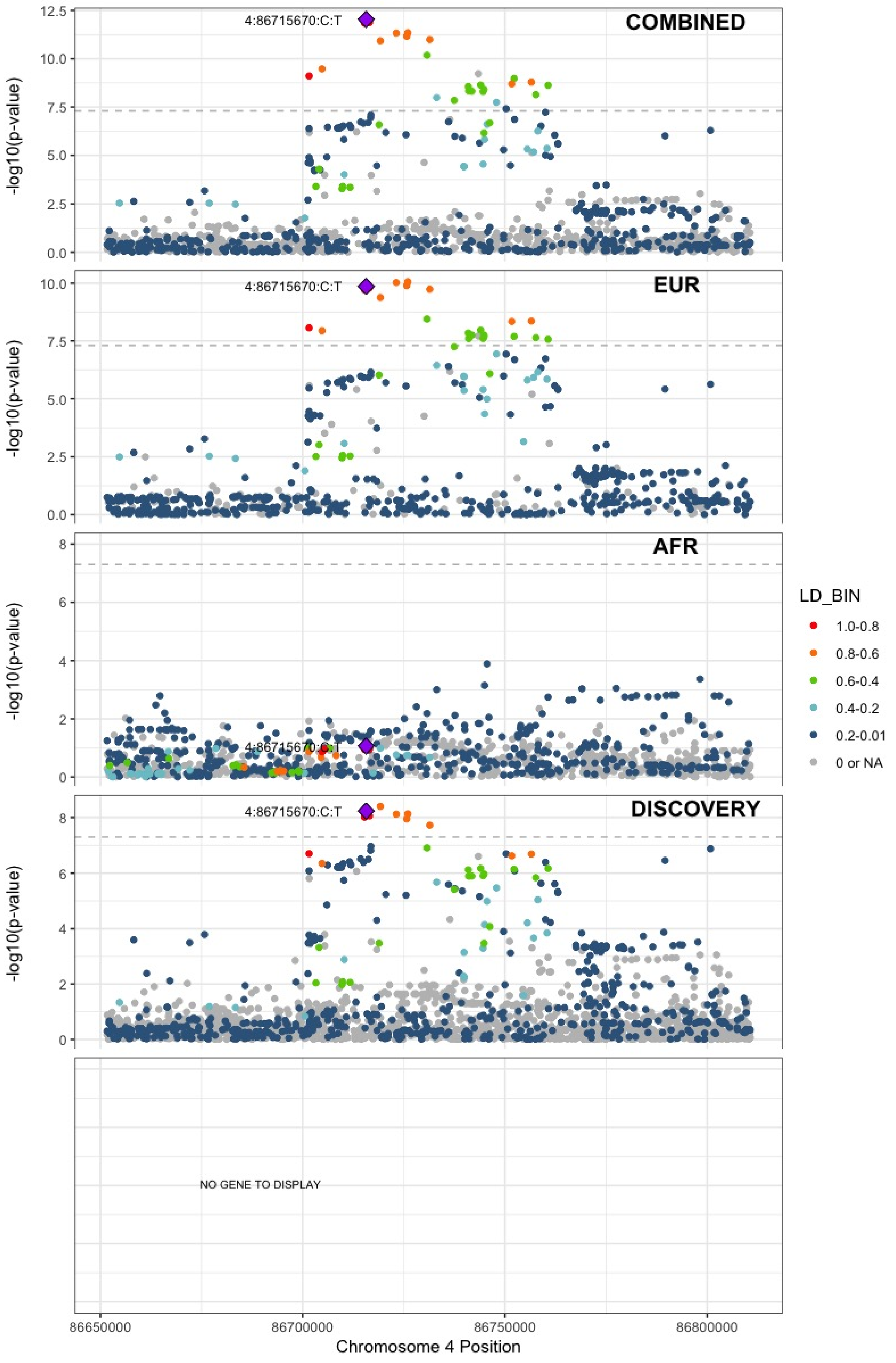

**Supplementary Figure 6.32** – rs2066864 (chr4:155525695) in locus 39 (FGG)

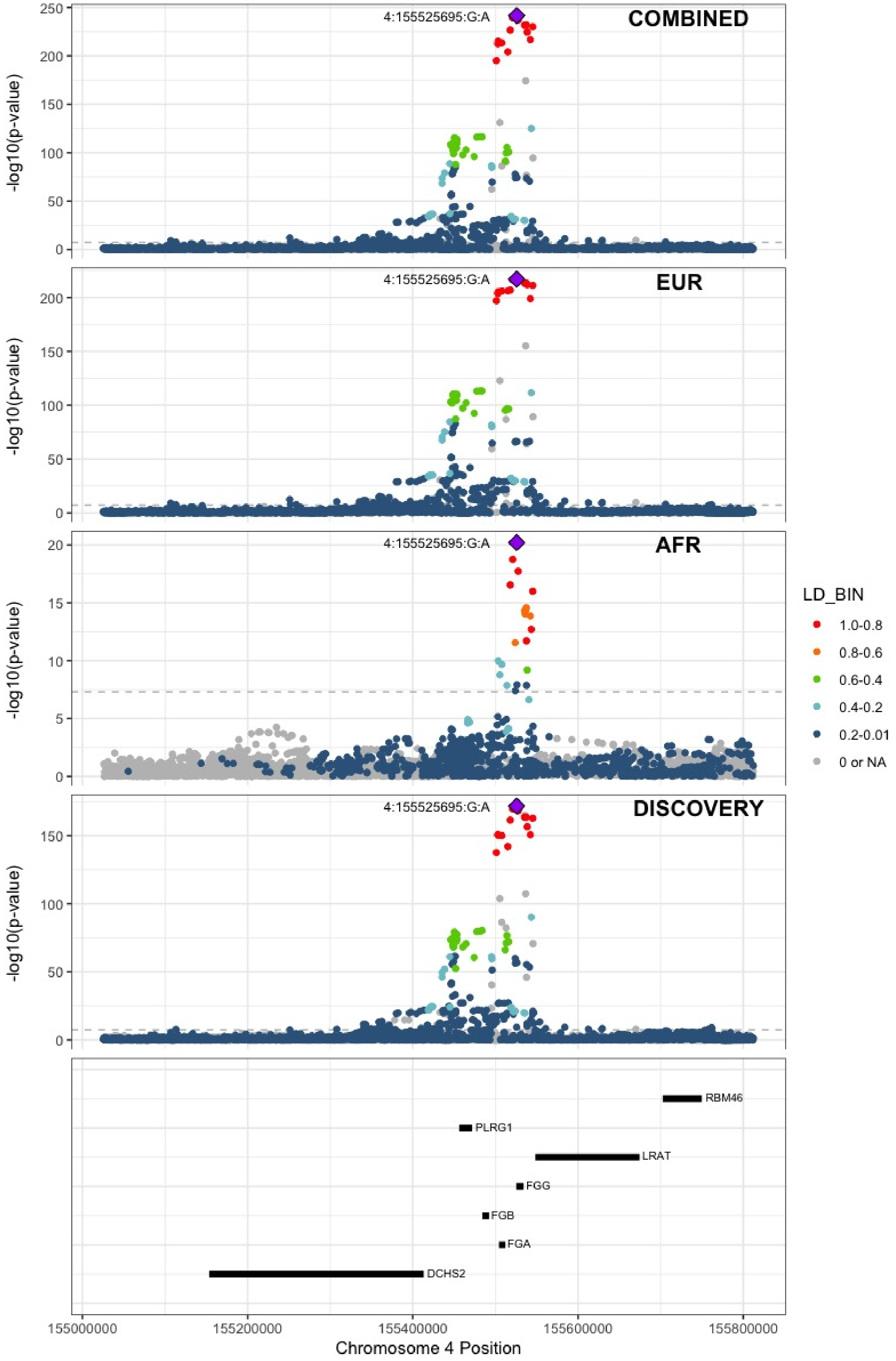

**Supplementary Figure 6.33** – rs3756011 (chr4:187206249) in locus 40 (F11)

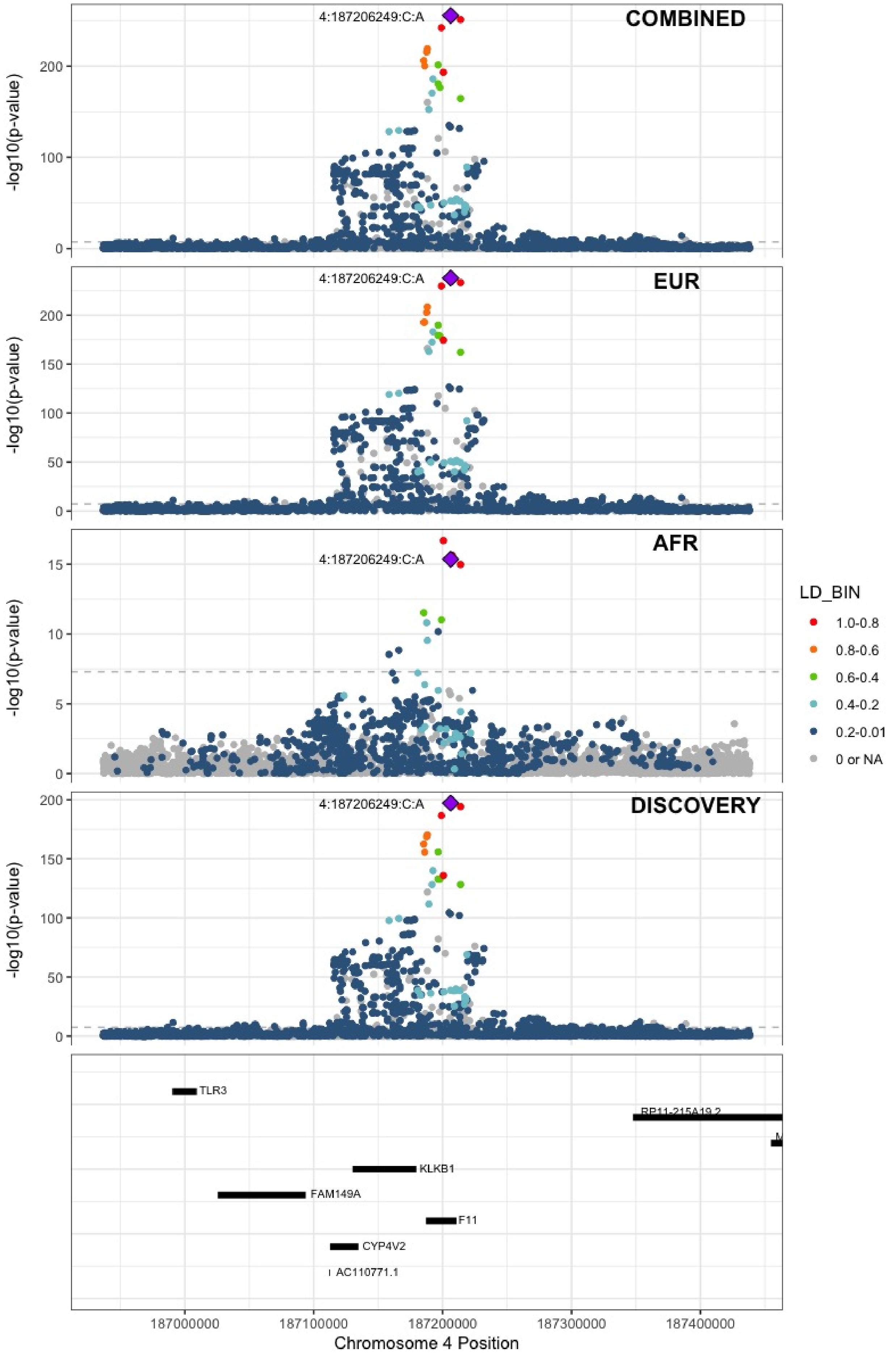

**Supplementary Figure 6.34** – rs16867574 (chr5:38708554) in locus 42 (OSMR-AS1)

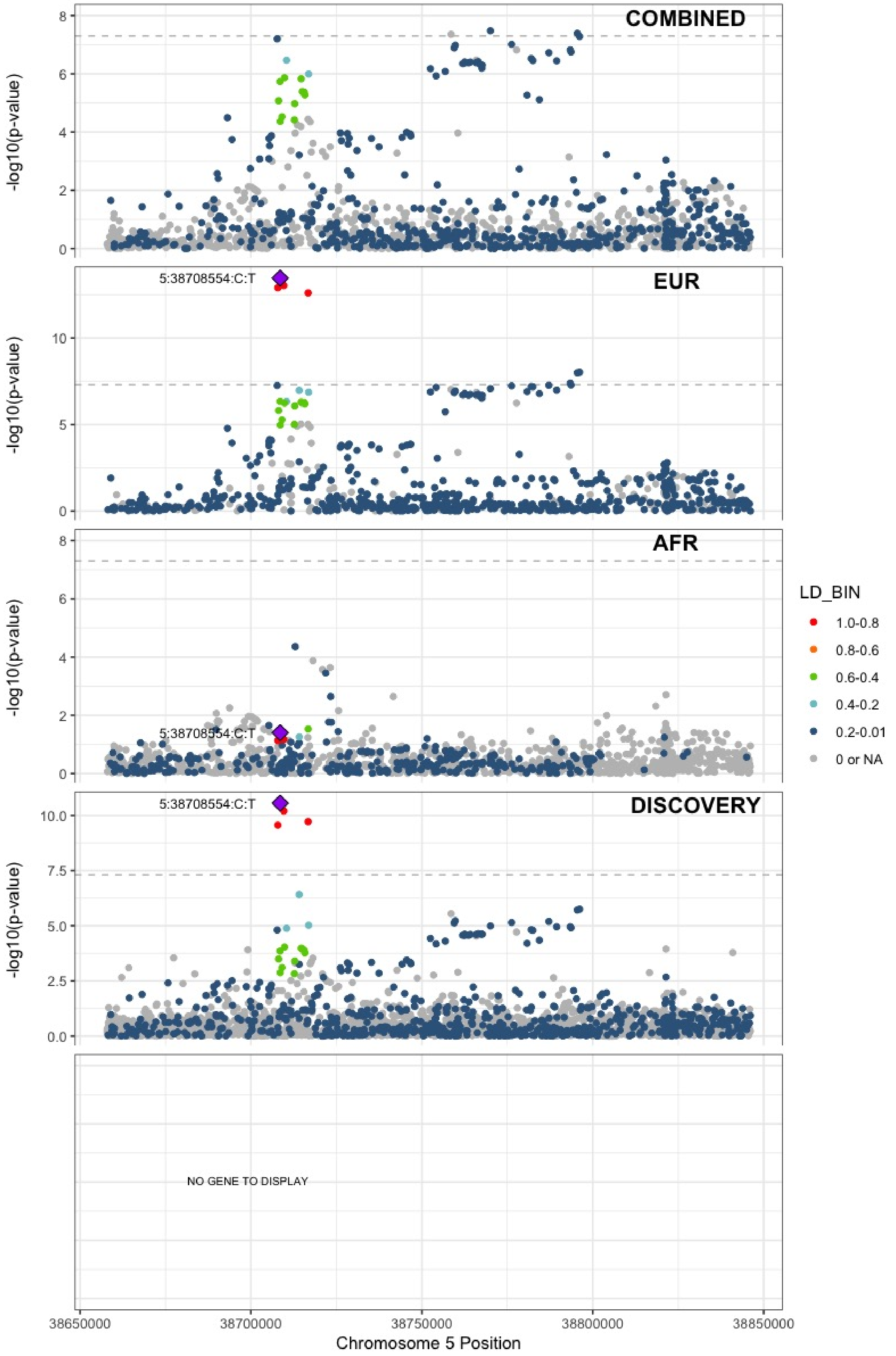

**Supplementary Figure 6.35** – rs7730244 (chr5:72957088) in locus 43 (ARHGEF28)

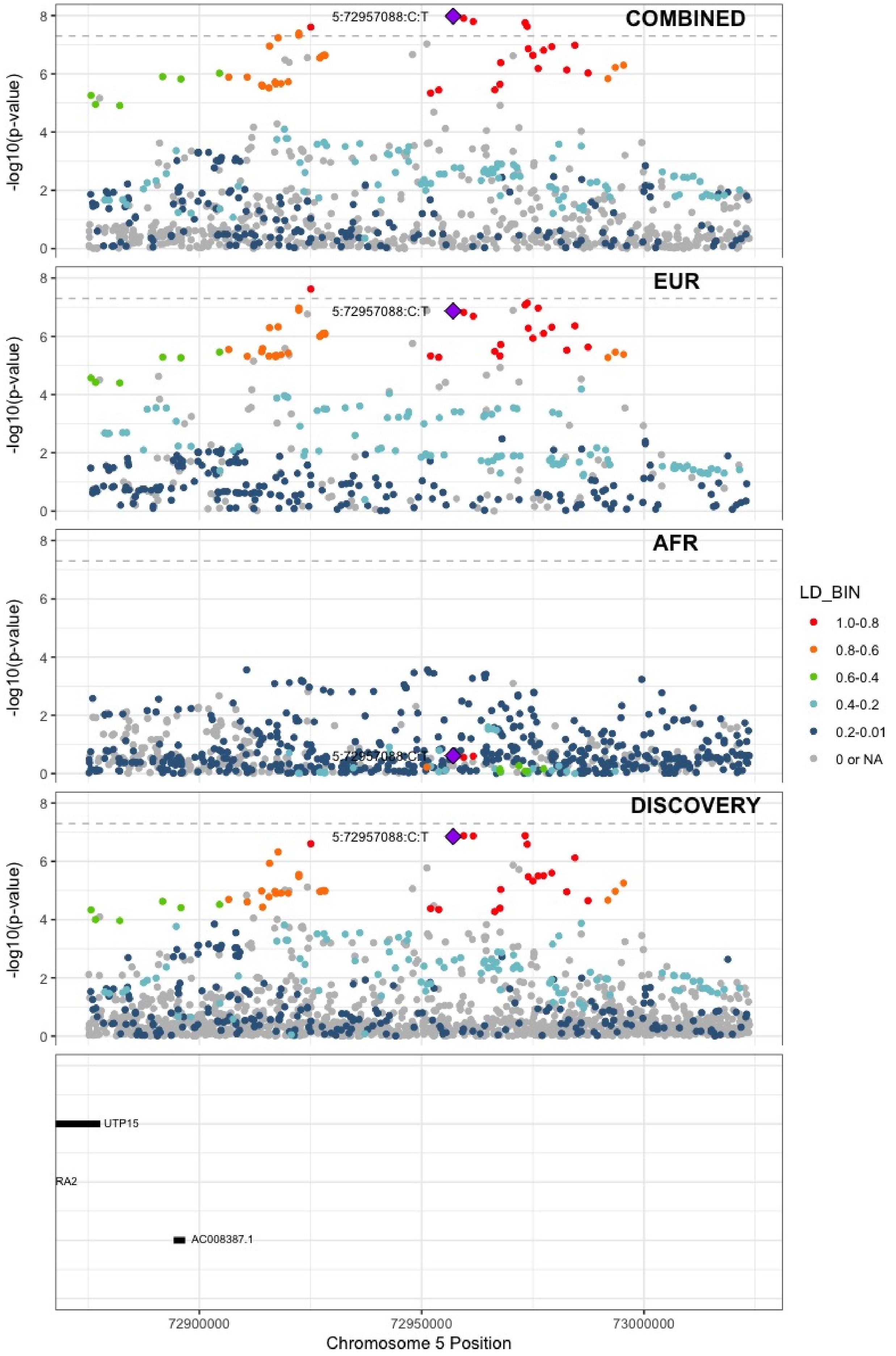

**Supplementary Figure 6.36** – rs750465397 (chr5:96289036) in locus 44 (LNPEP)

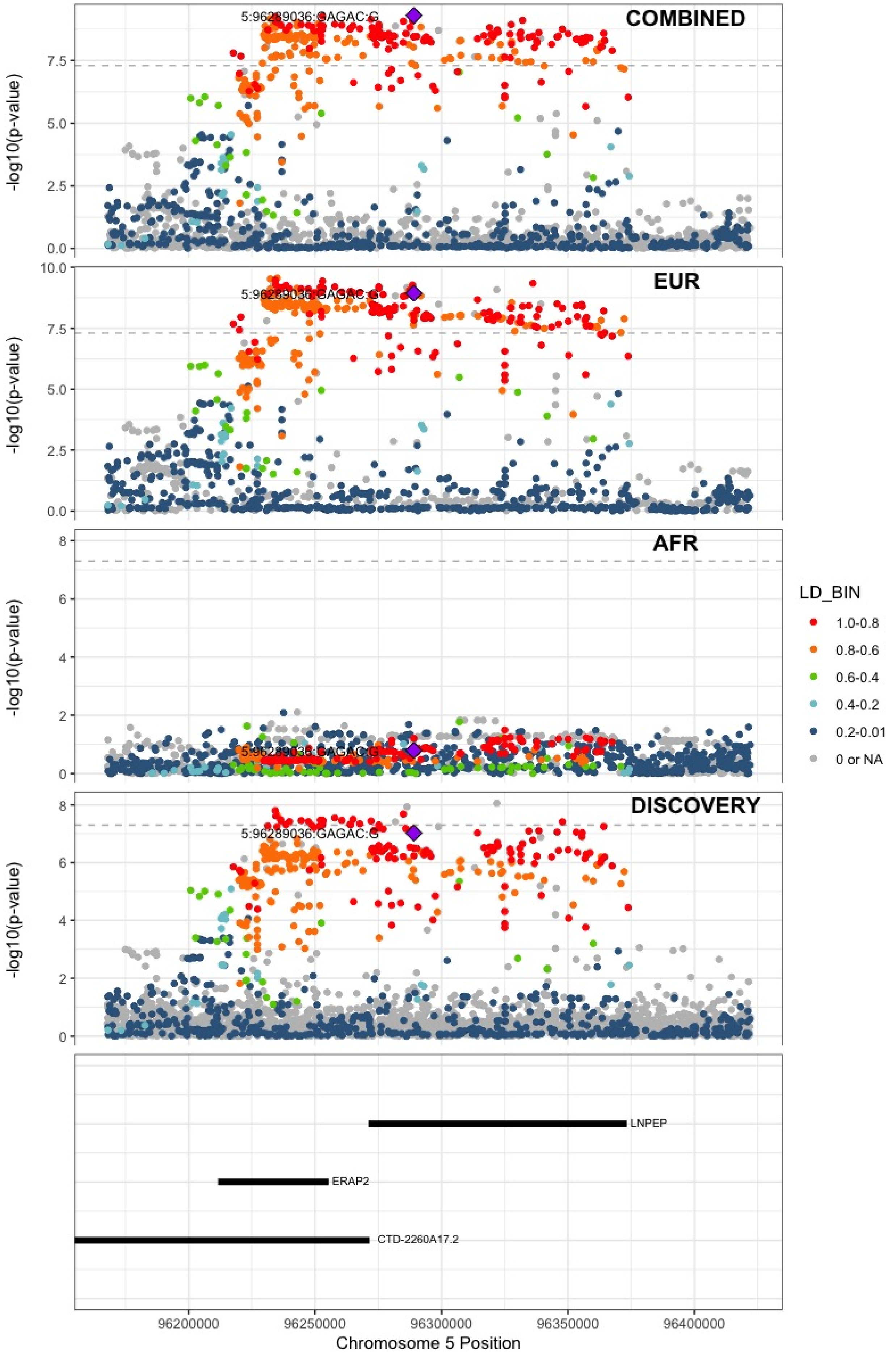

**Supplementary Figure 6.37** – rs147133967 (chr5:132426851) in locus 45 (HSPA4)

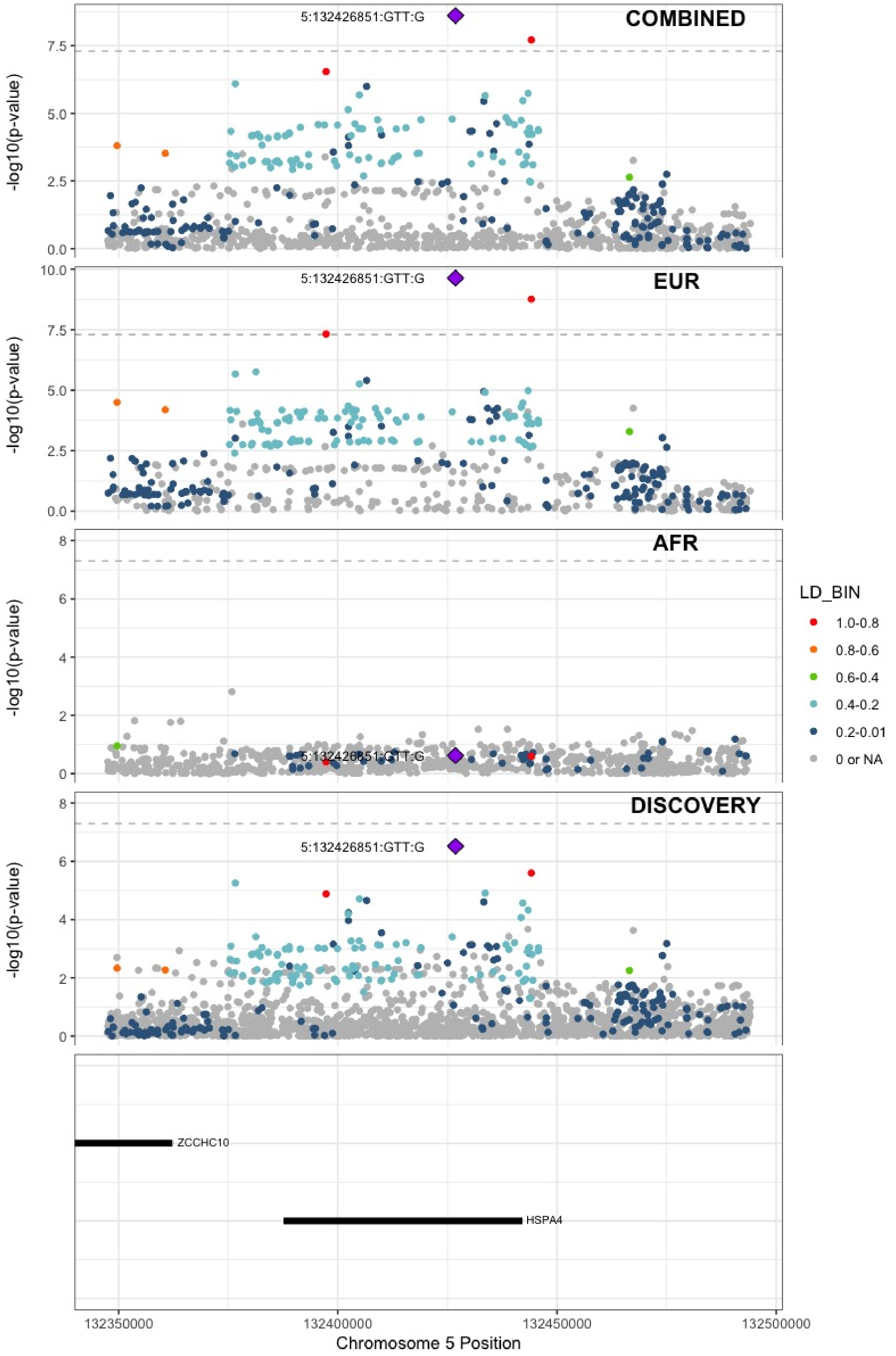

**Supplementary Figure 6.38** – rs214059 (chr6:25536937) in locus 46 (CARMIL1)

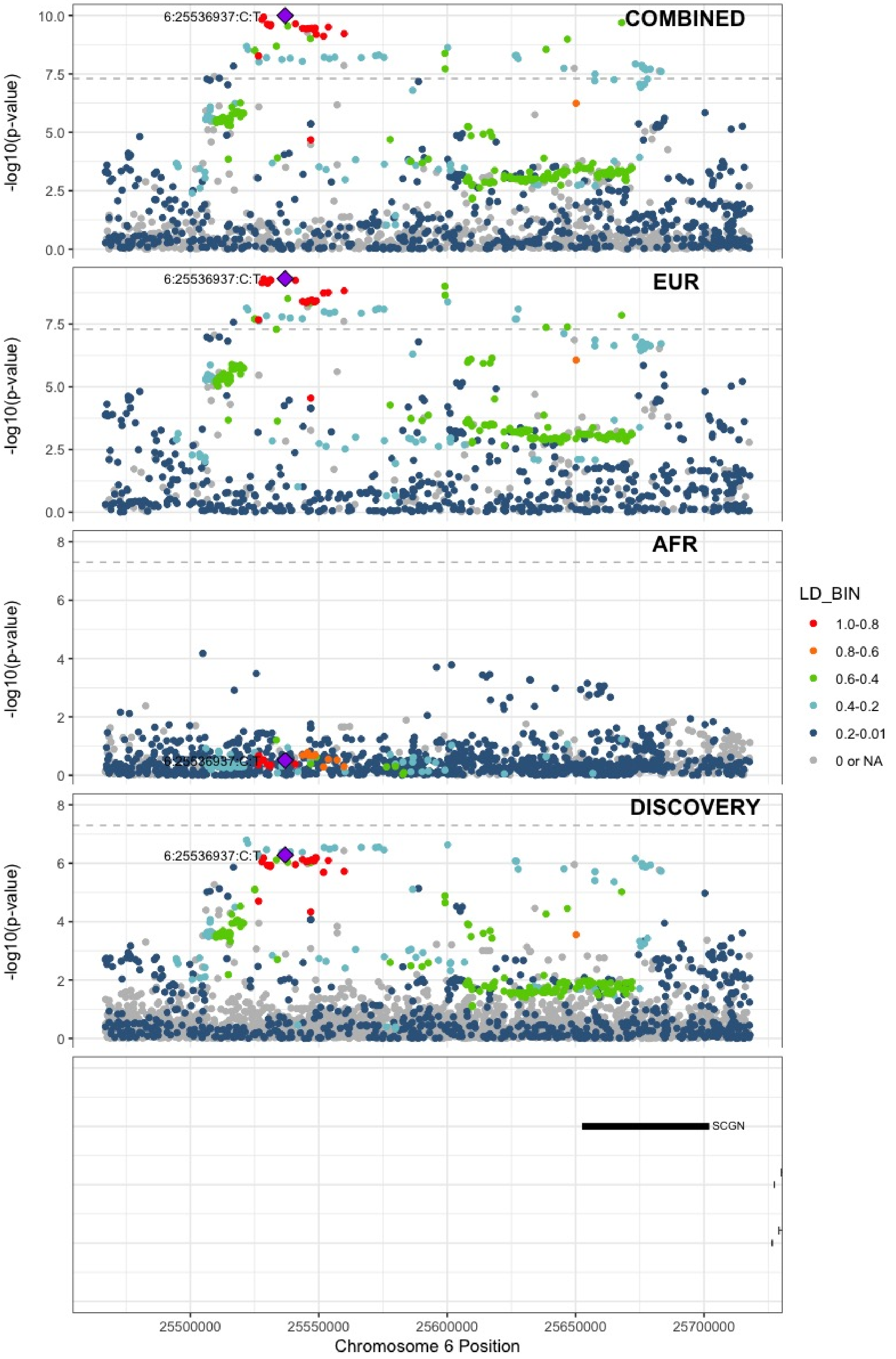

**Supplementary Figure 6.39** – rs2394251 (chr6:29943688) in locus 47 (HCG9)

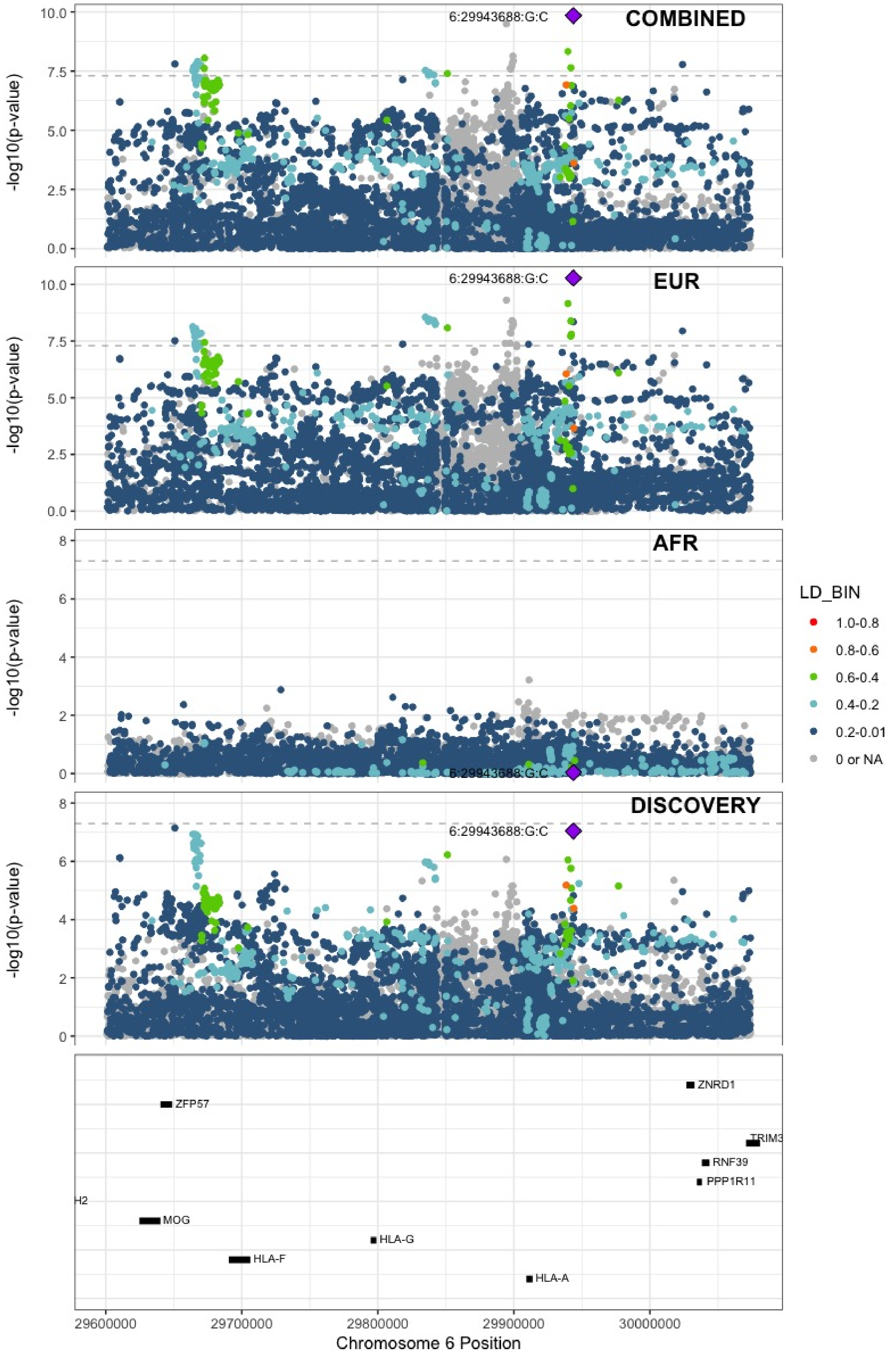

**Supplementary Figure 6.40** – rs9268863 (chr6:32430289) in locus 48 (HLA-DRA ;HLA-DRB5)

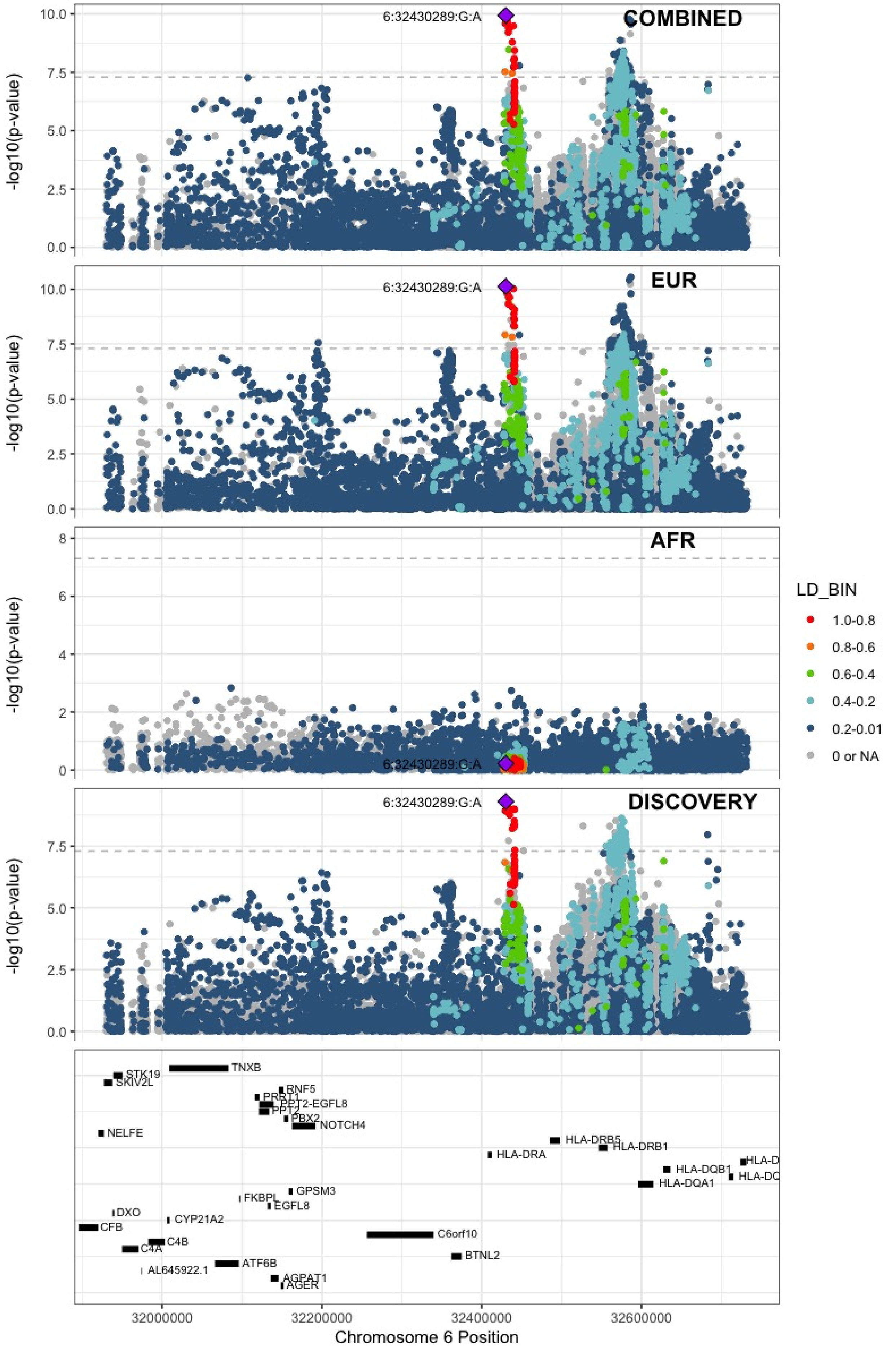

**Supplementary Figure 6.41** – rs145294670 (chr6:34622561) in locus 49 (ILRUN)

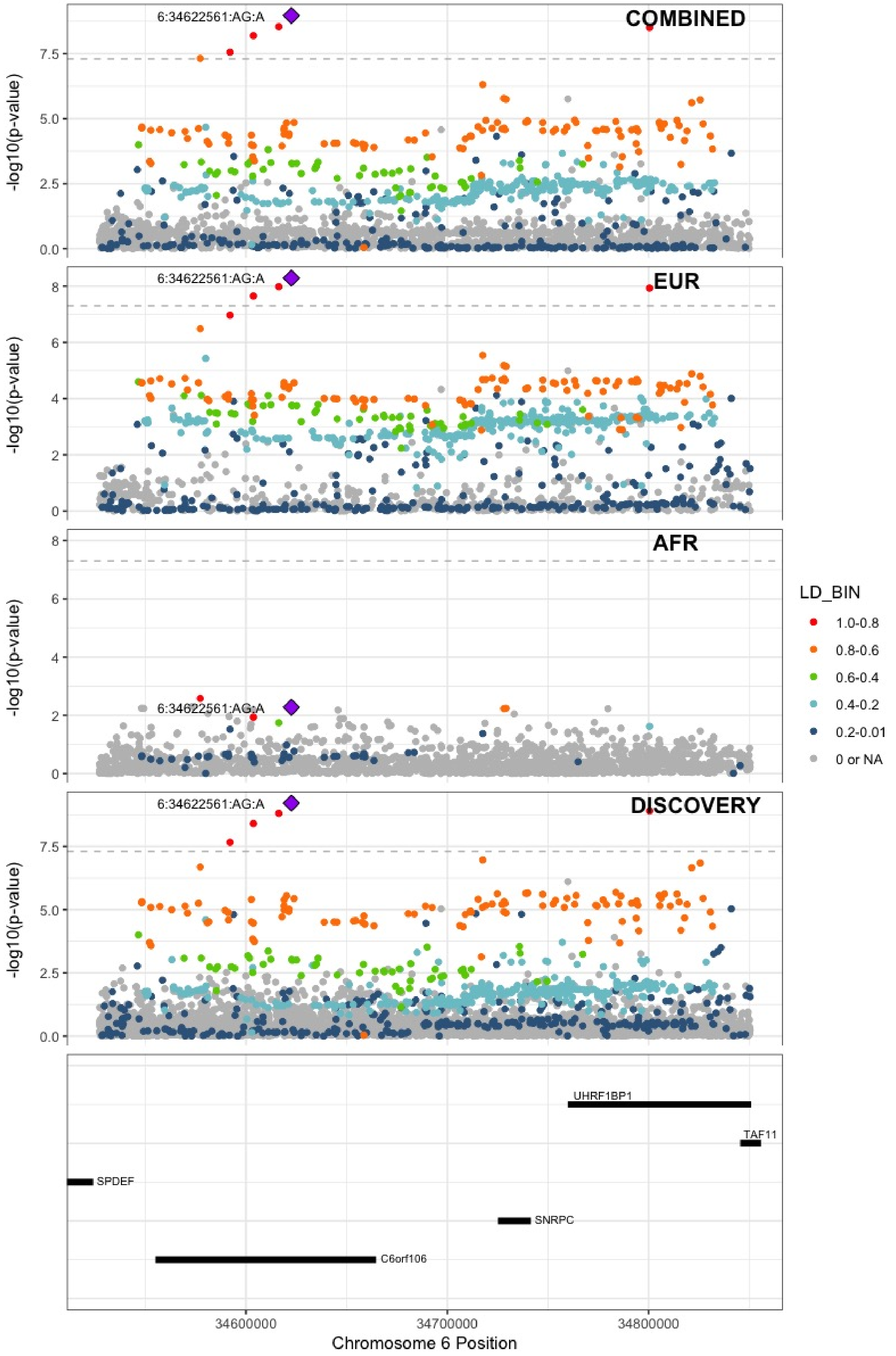

**Supplementary Figure 6.42** – rs9386182 (chr6:147691069) in locus 52 (STXBP5)

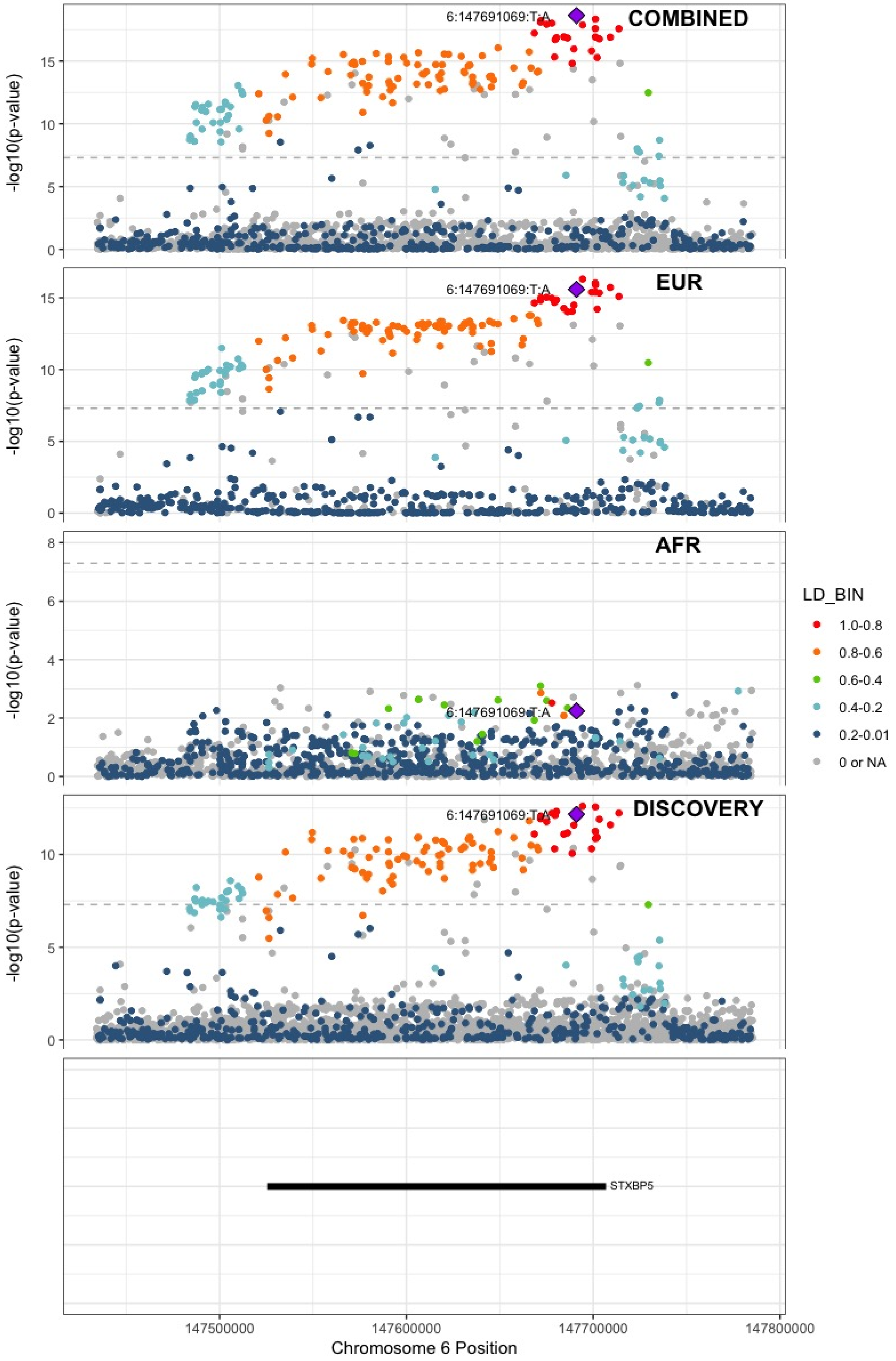

**Supplementary Figure 6.43** – rs1513275 (chr7:28259233) in locus 53 (JAZF1-AS1)

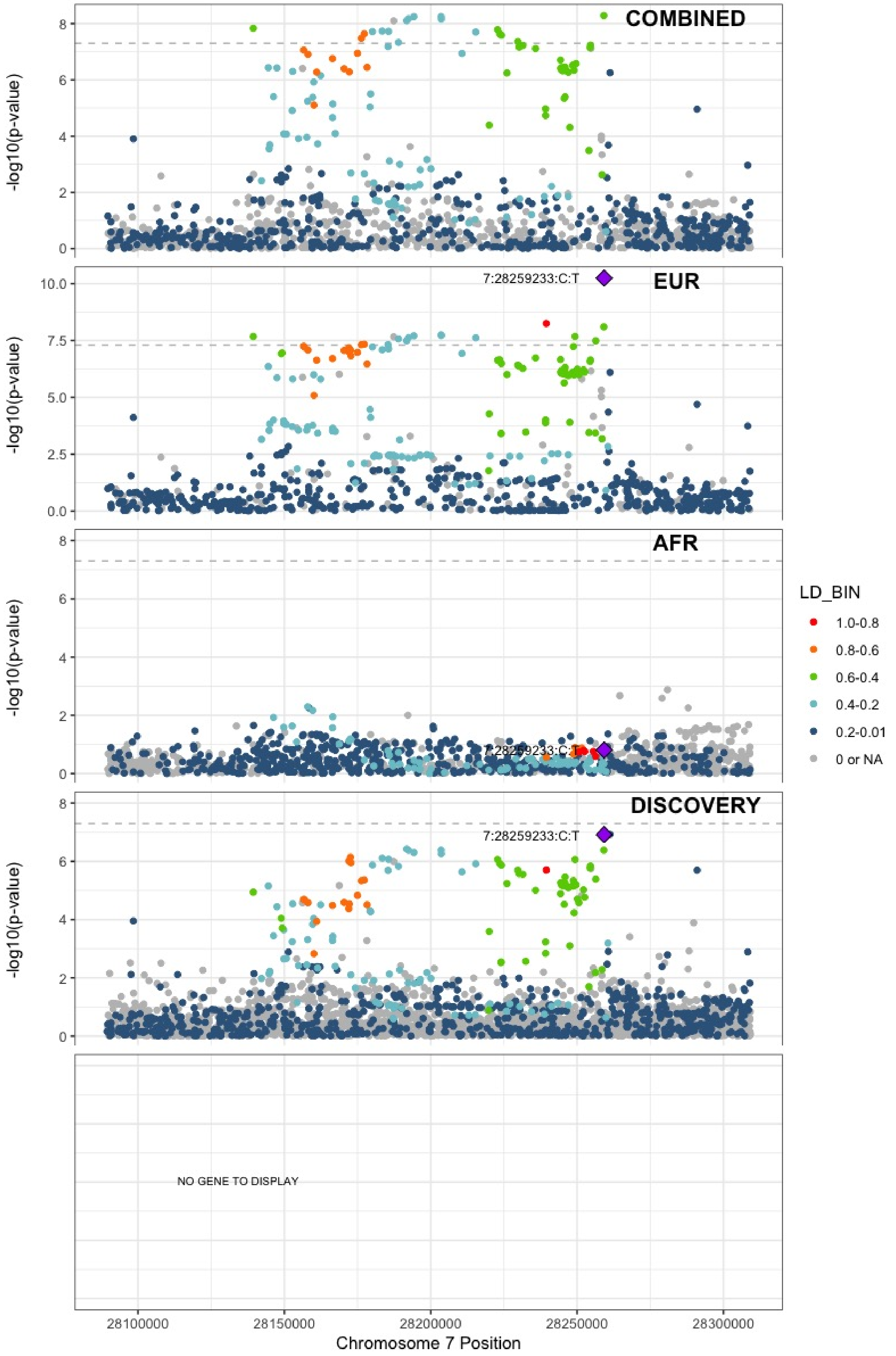

**Supplementary Figure 6.44** – rs42038 (chr7:92243719) in locus 54 (CDK6)

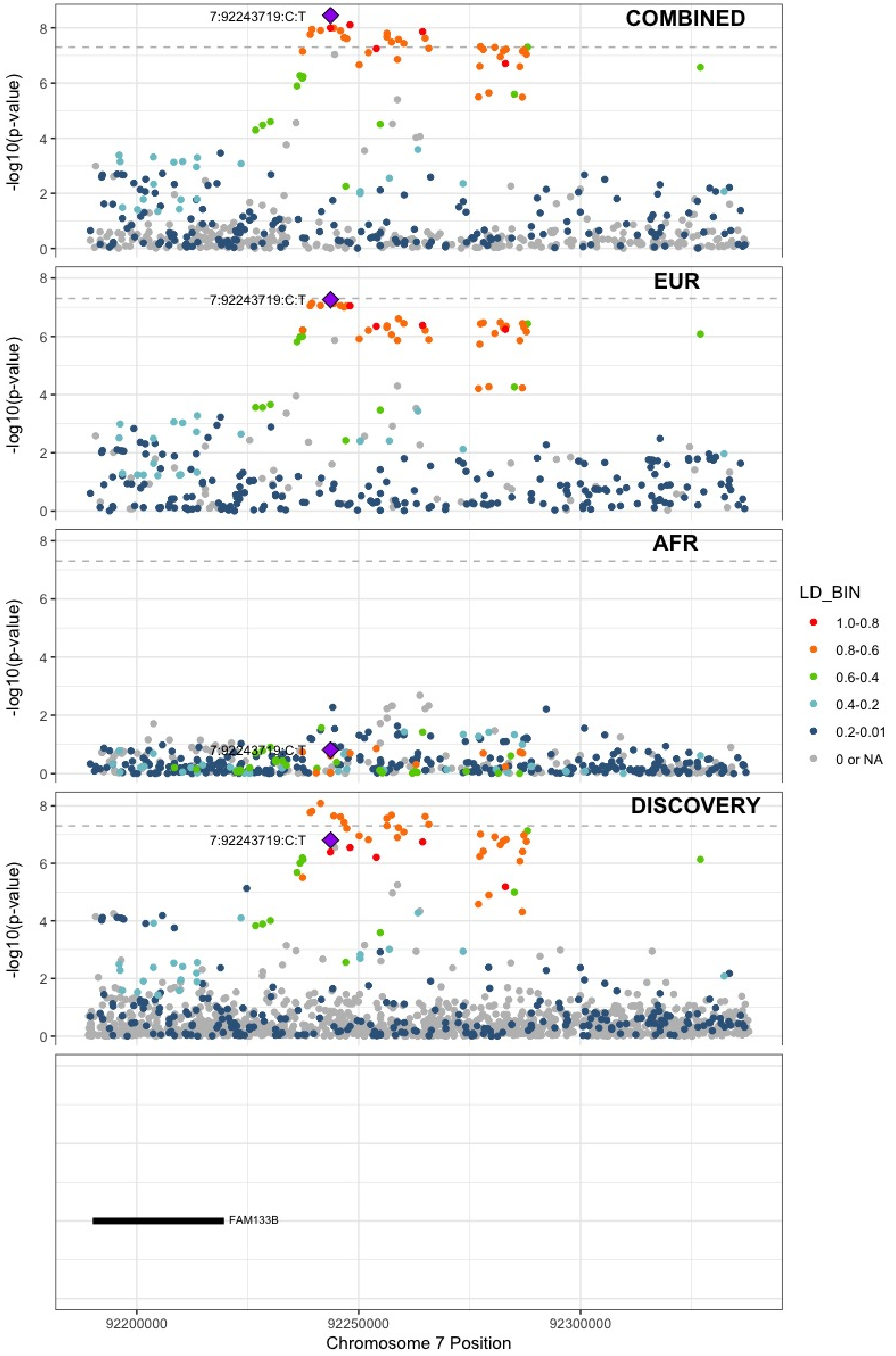

**Supplementary Figure 6.45** – rs10099512 (chr8:9178821) in locus 57 (LOC101929128 ;LOC157273)

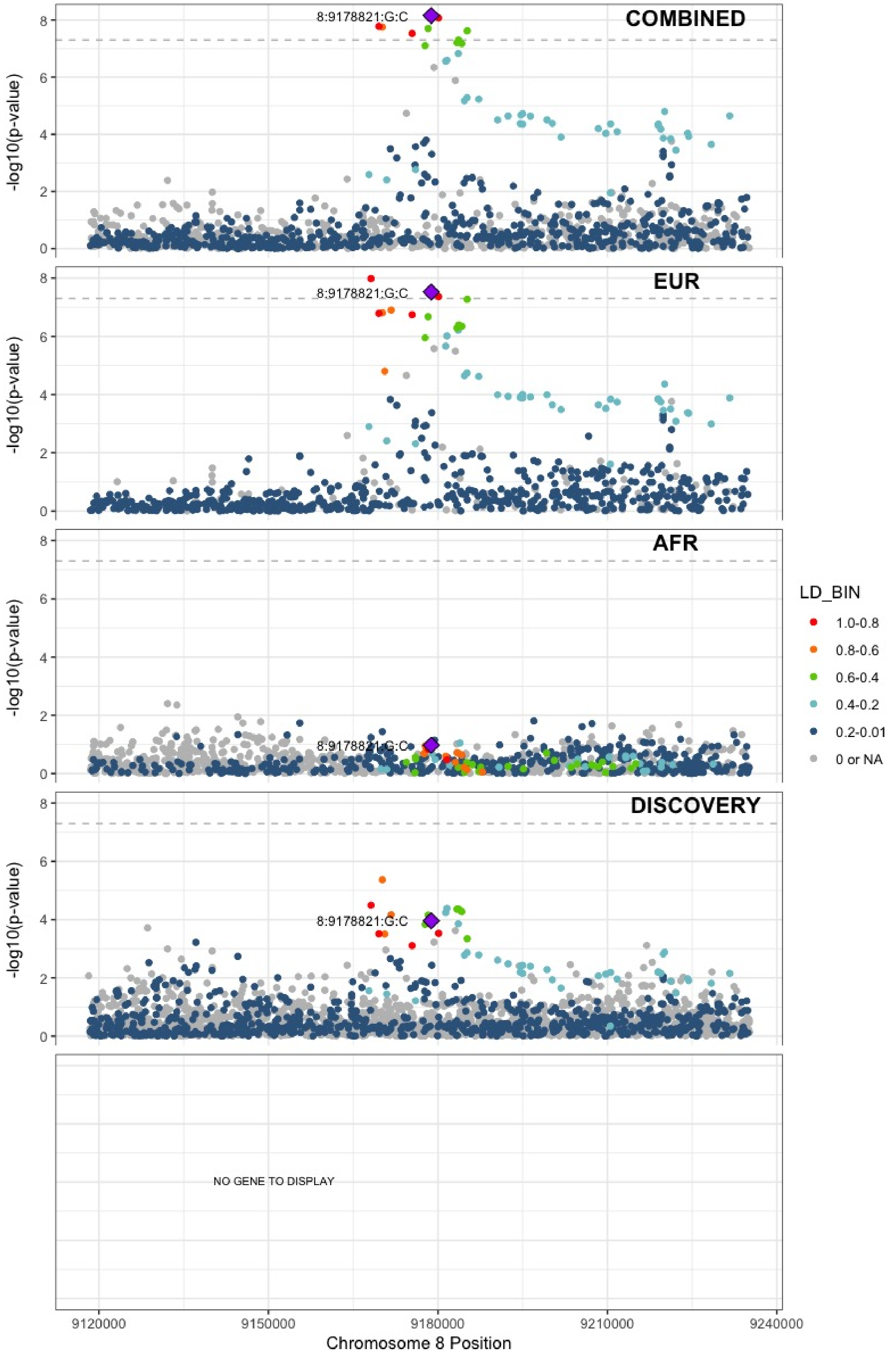

**Supplementary Figure 6.46** – rs2048528 (chr8:23373680) in locus 59 (ENTPD4;SLC25A37)

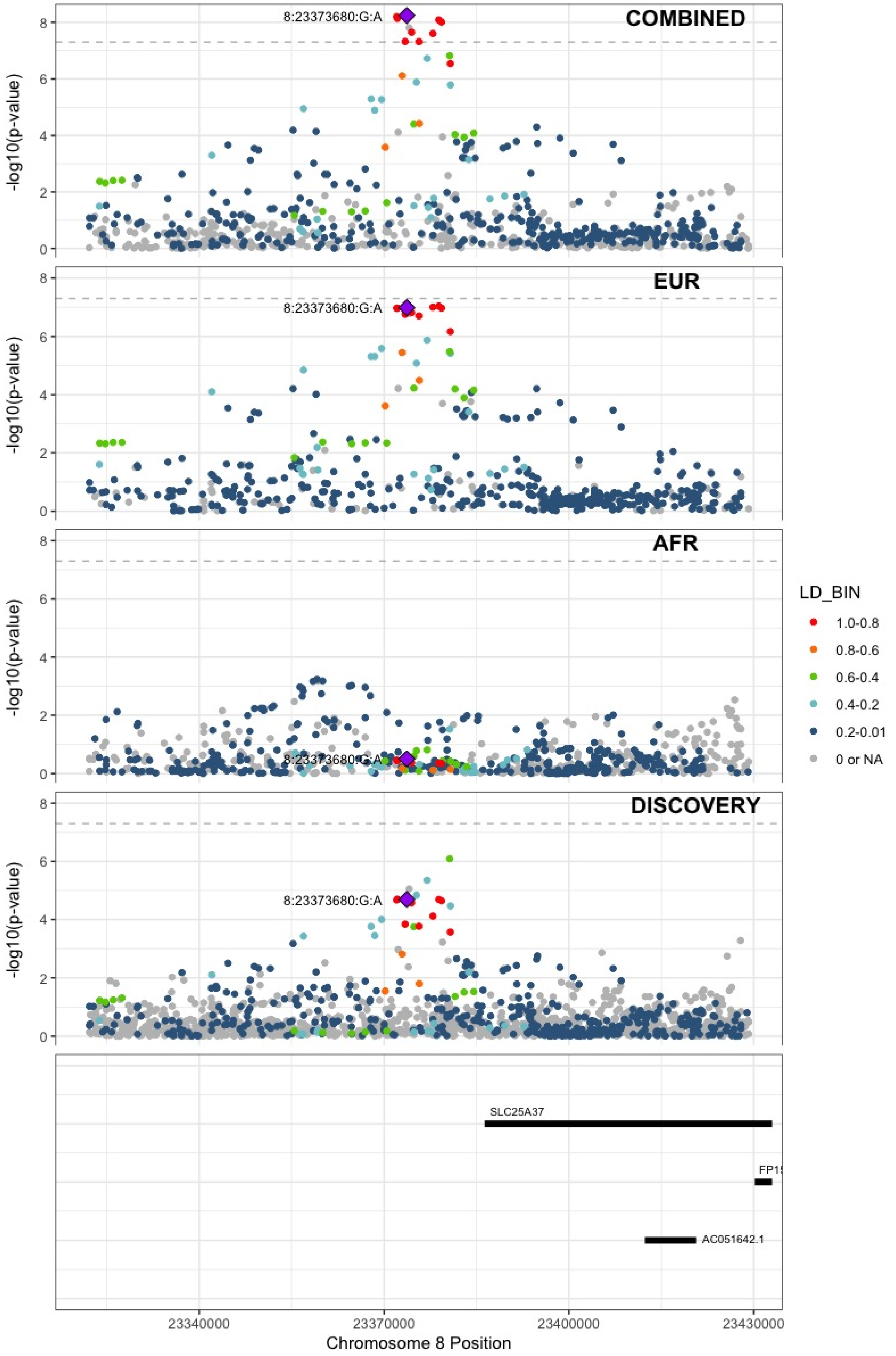

**Supplementary Figure 6.47** – rs2685412 (chr8:27810603) in locus 60 (SCARA5)

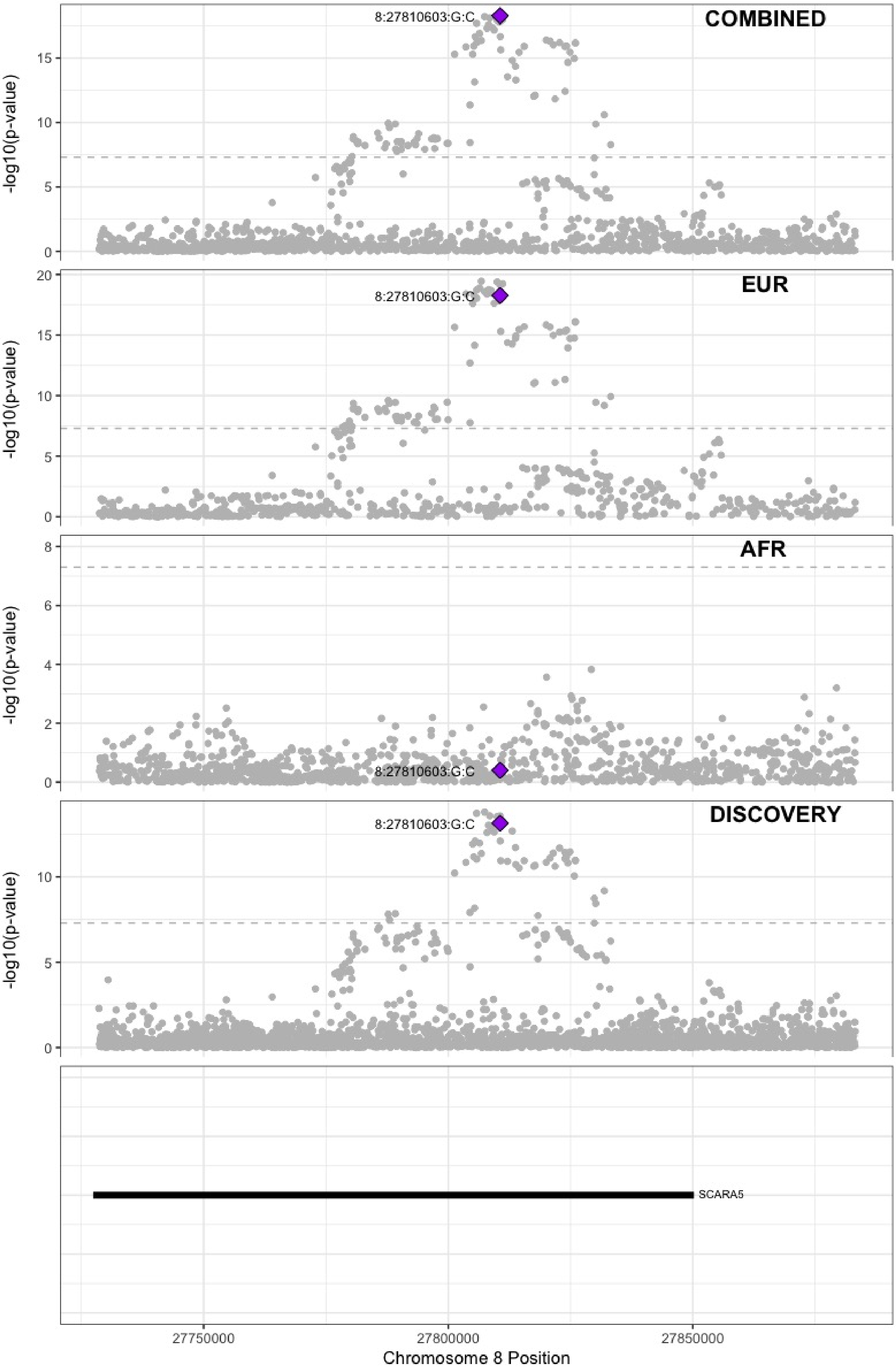

**Supplementary Figure 6.48** – rs2915595 (chr8:30402817) in locus 61 (RBPMS)

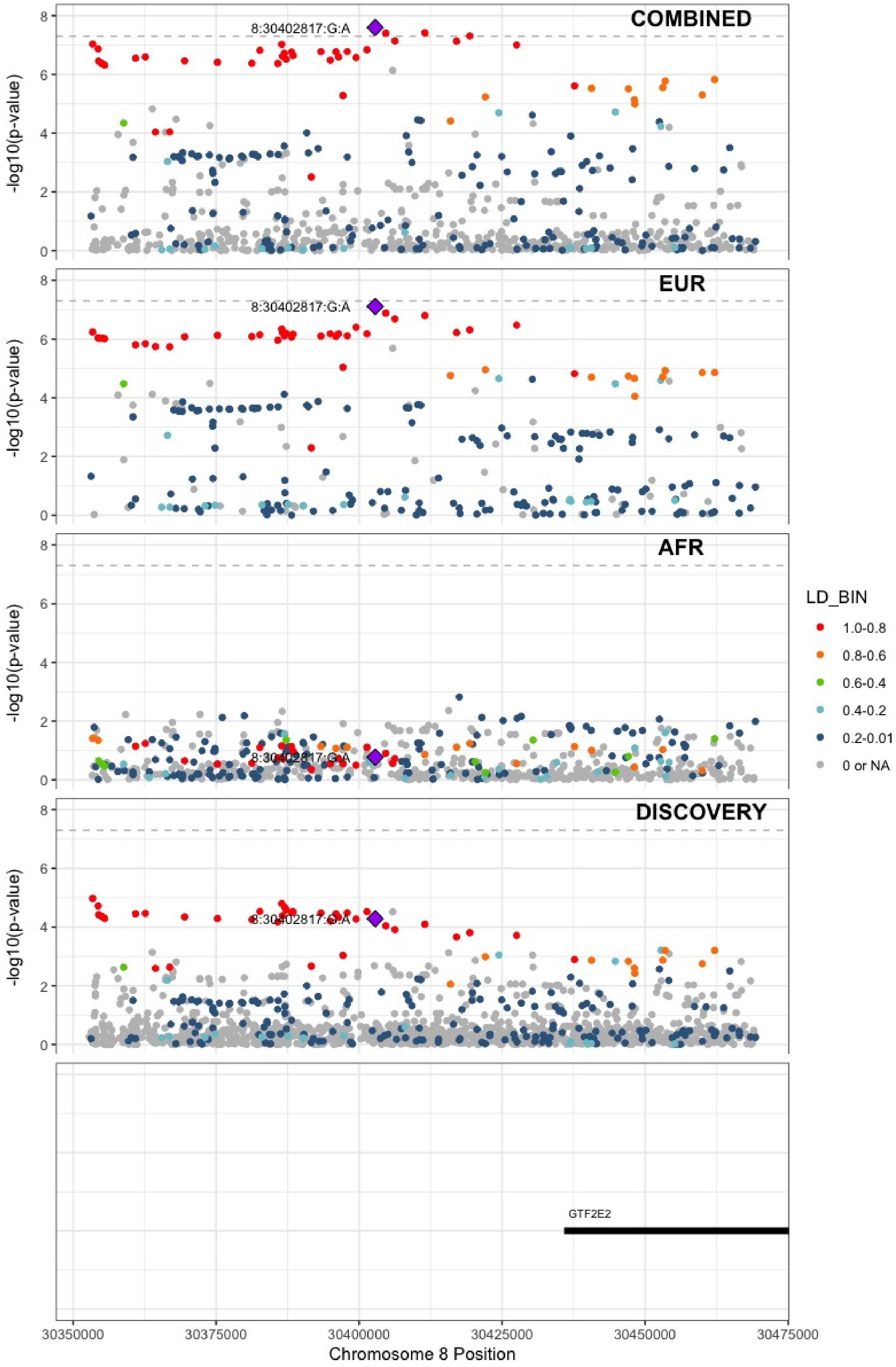

**Supplementary Figure 6.49** – rs6993770 (chr8:106581528) in locus 62 (ZFPM2)

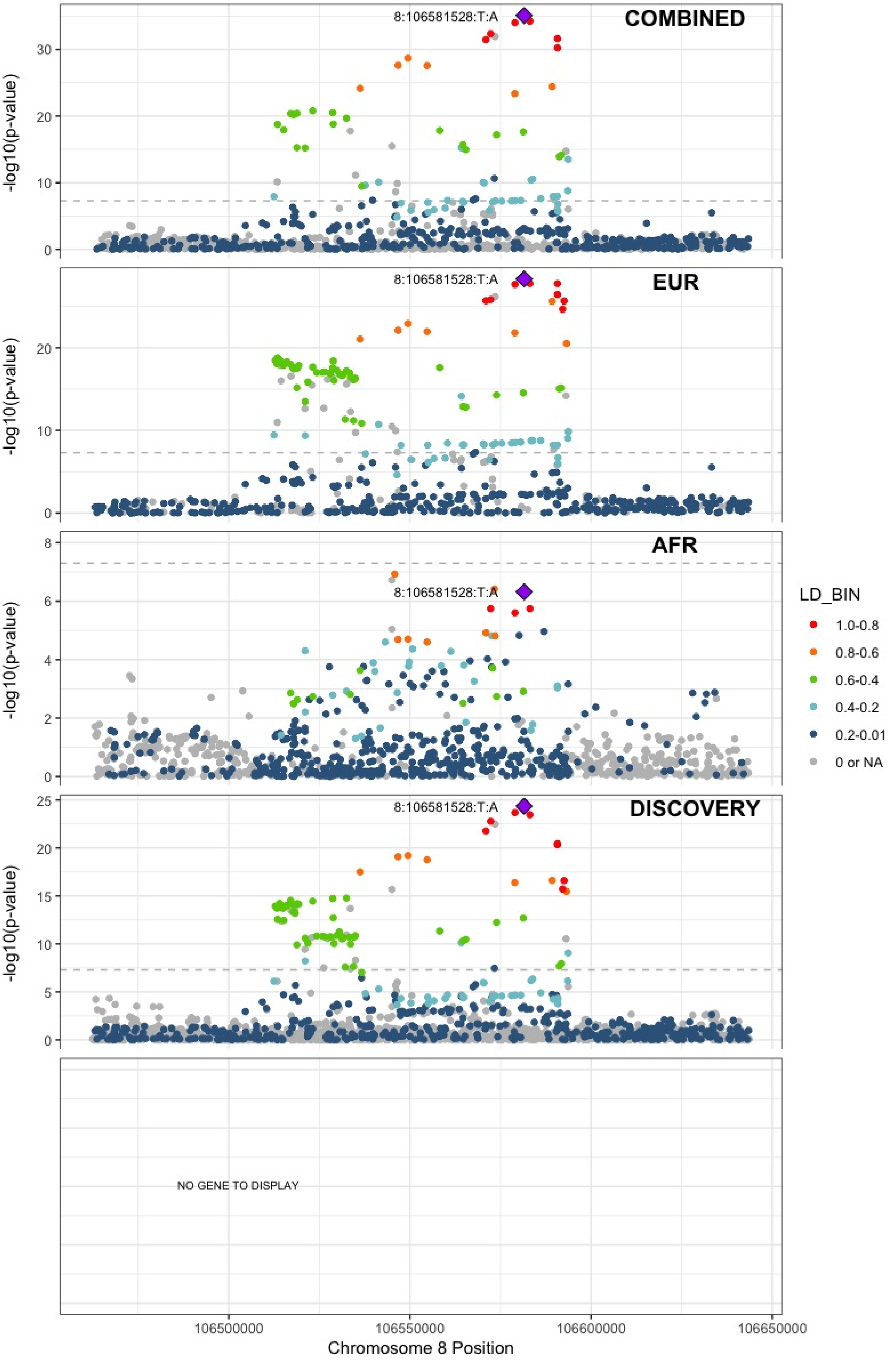

**Supplementary Figure 6.50** – rs4236786 (chr8:108291878) in locus 63 (ANGPT1)

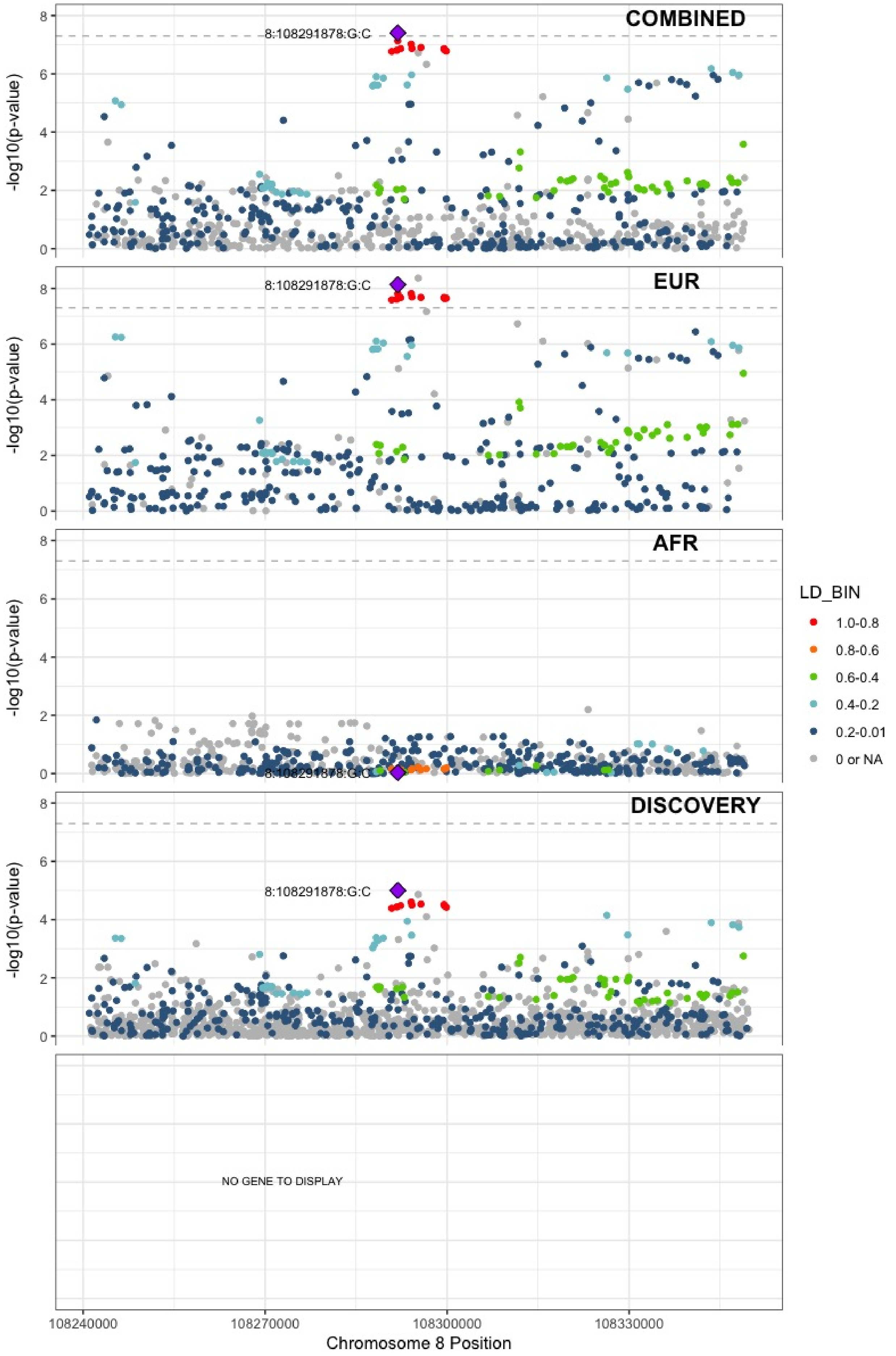

**Supplementary Figure 6.51** – rs10820606 (chr9:99192919) in locus 65 (ZNF367;HABP4)

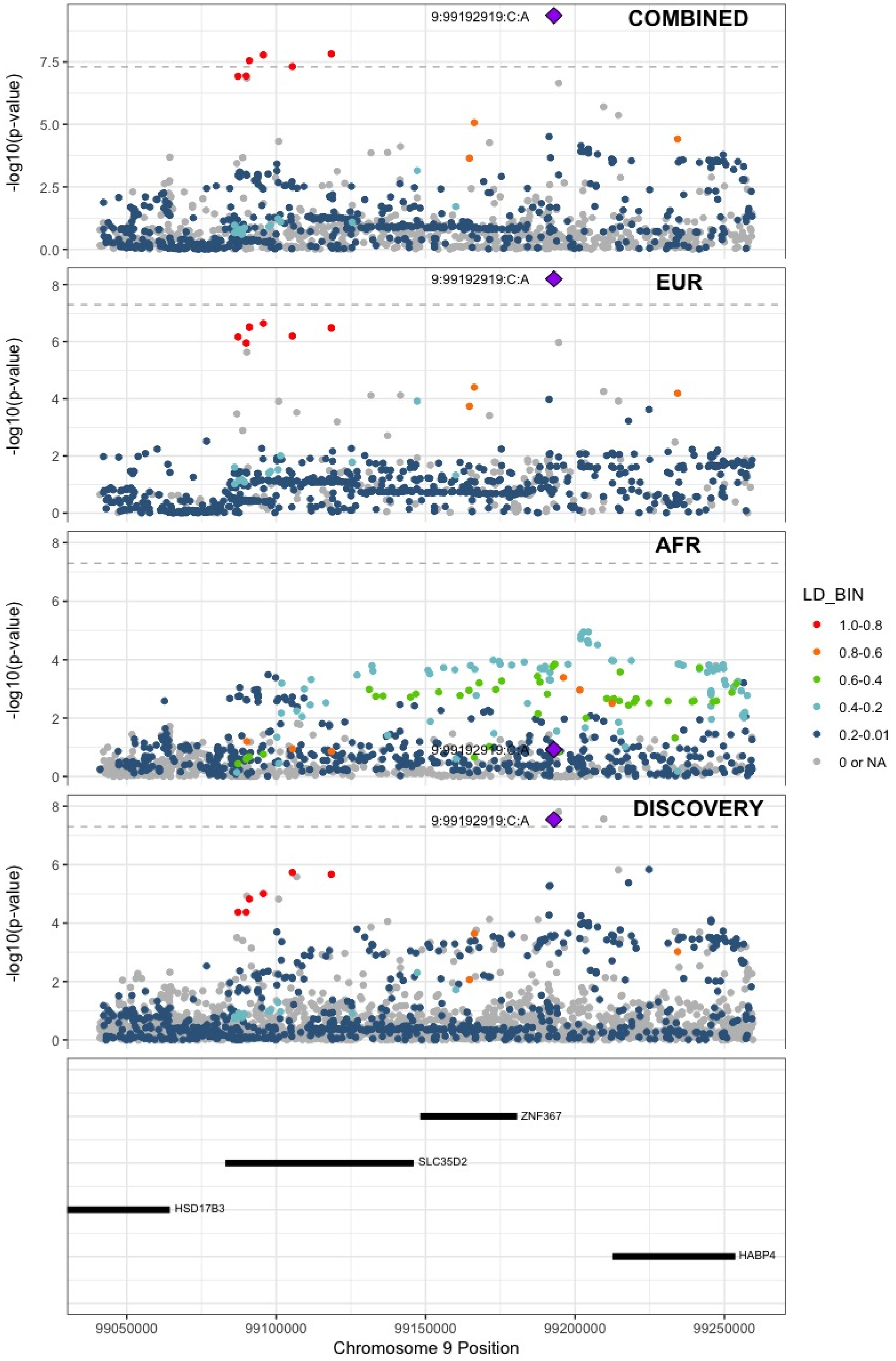

**Supplementary Figure 6.52** – rs505922 (chr9:136149229) in locus 66 (ABO)

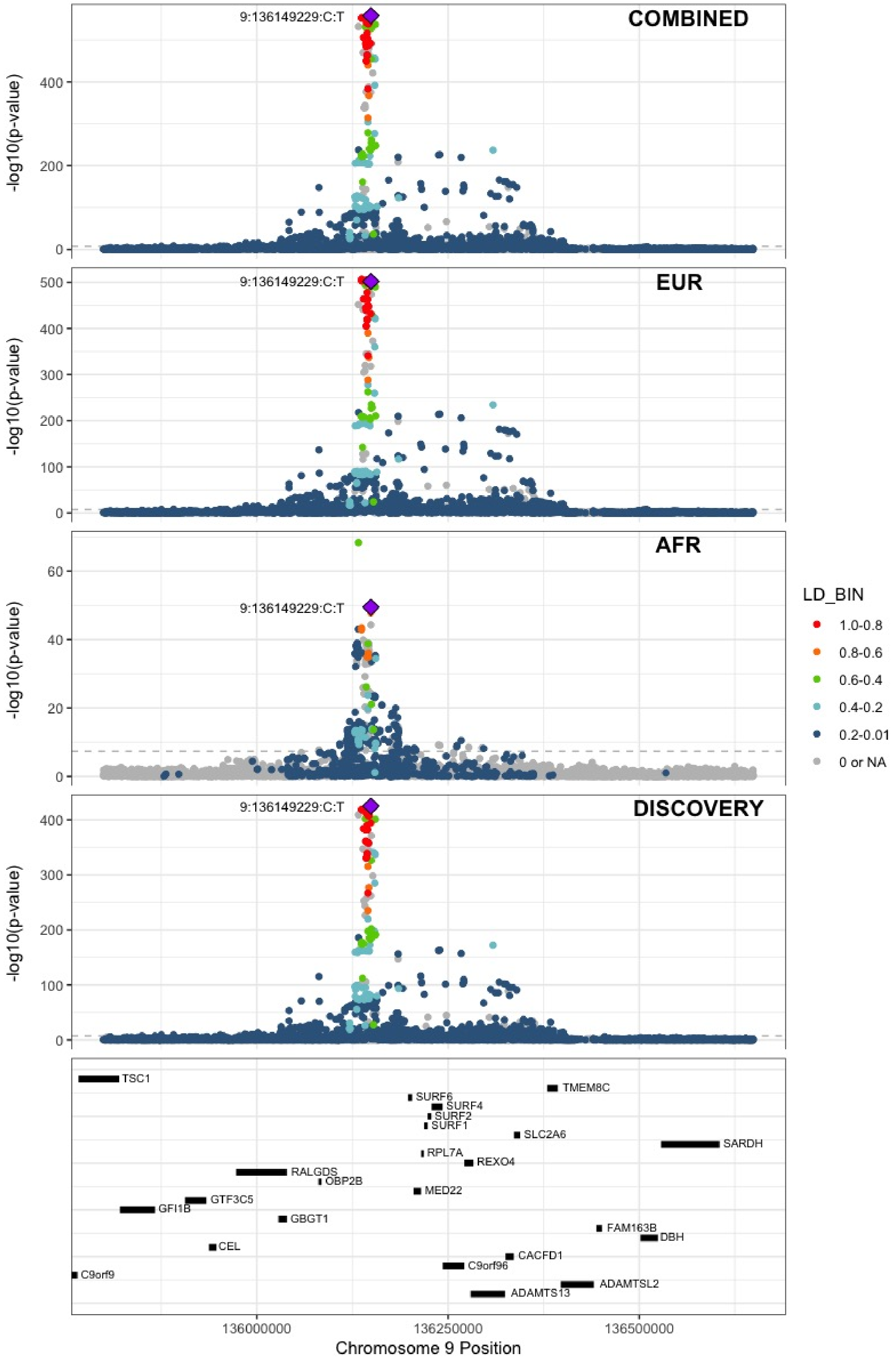

**Supplementary Figure 6.53** – rs1243187 (chr10:21907016) in locus 68 (MLLT10)

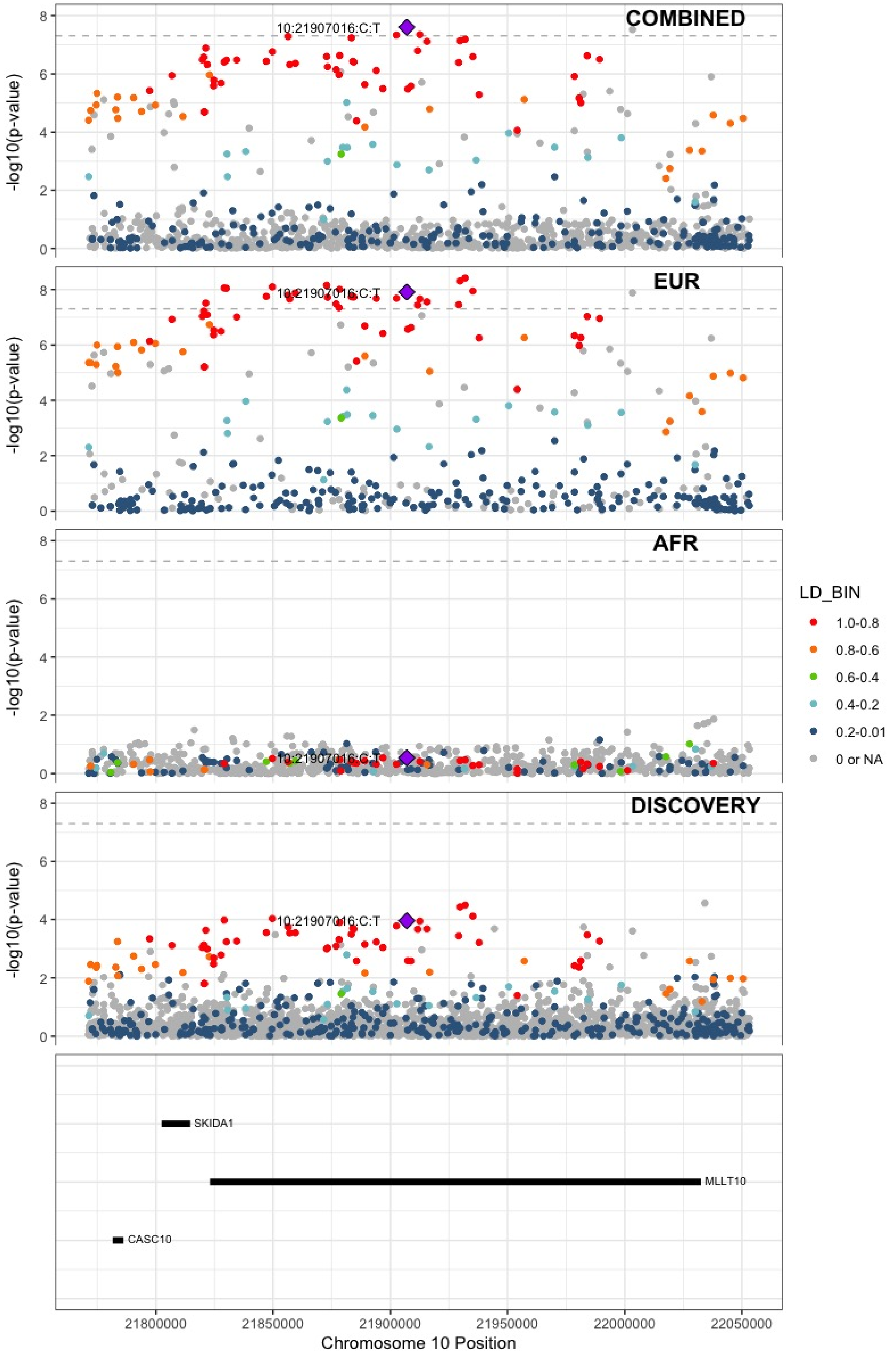

**Supplementary Figure 6.54** – rs4272700 (chr10:27881308) in locus 69 (RAB18 ;MKX)

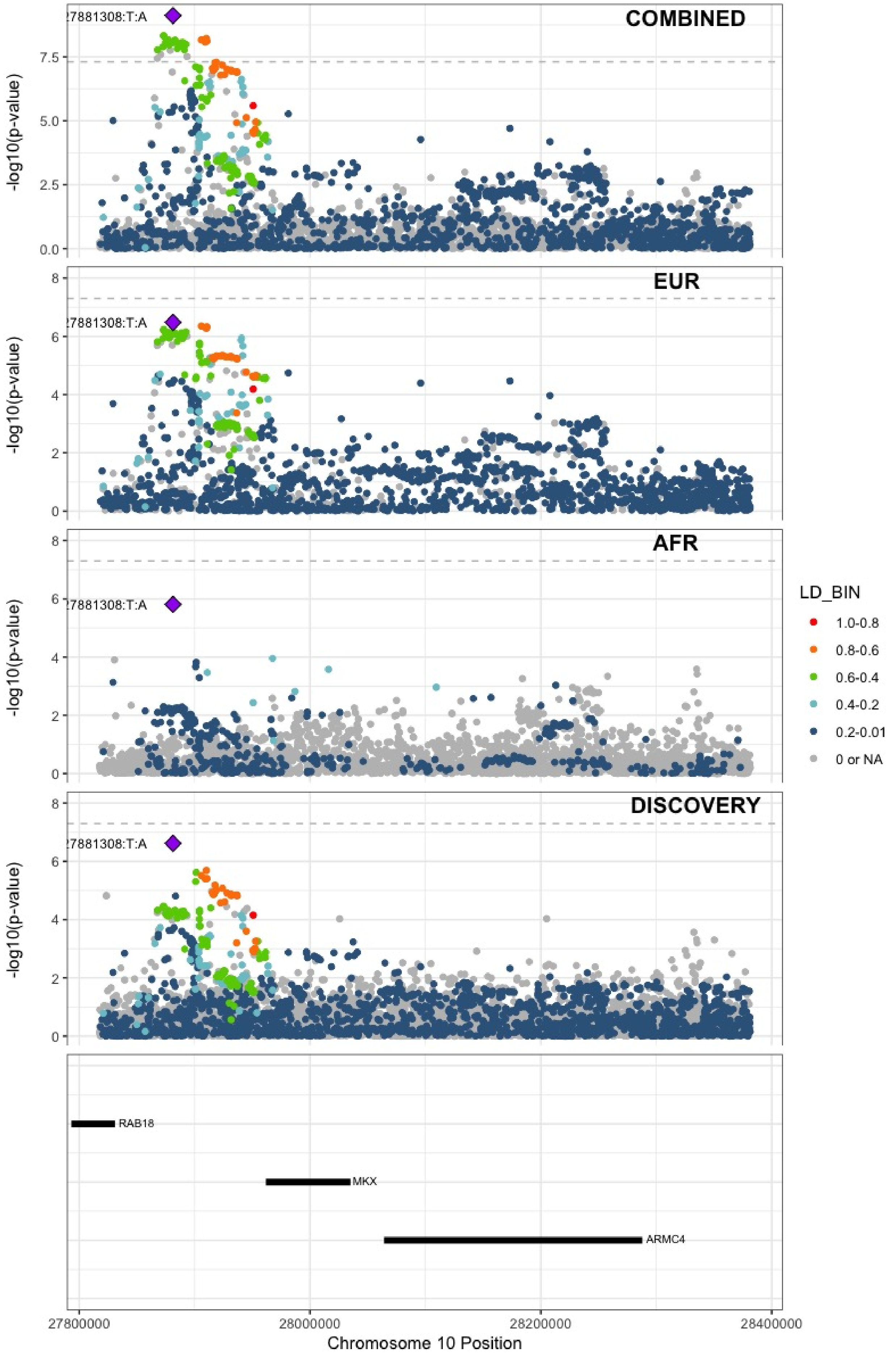

**Supplementary Figure 6.55** – rs17490626 (chr10:71218646) in locus 72 (TSPAN15)

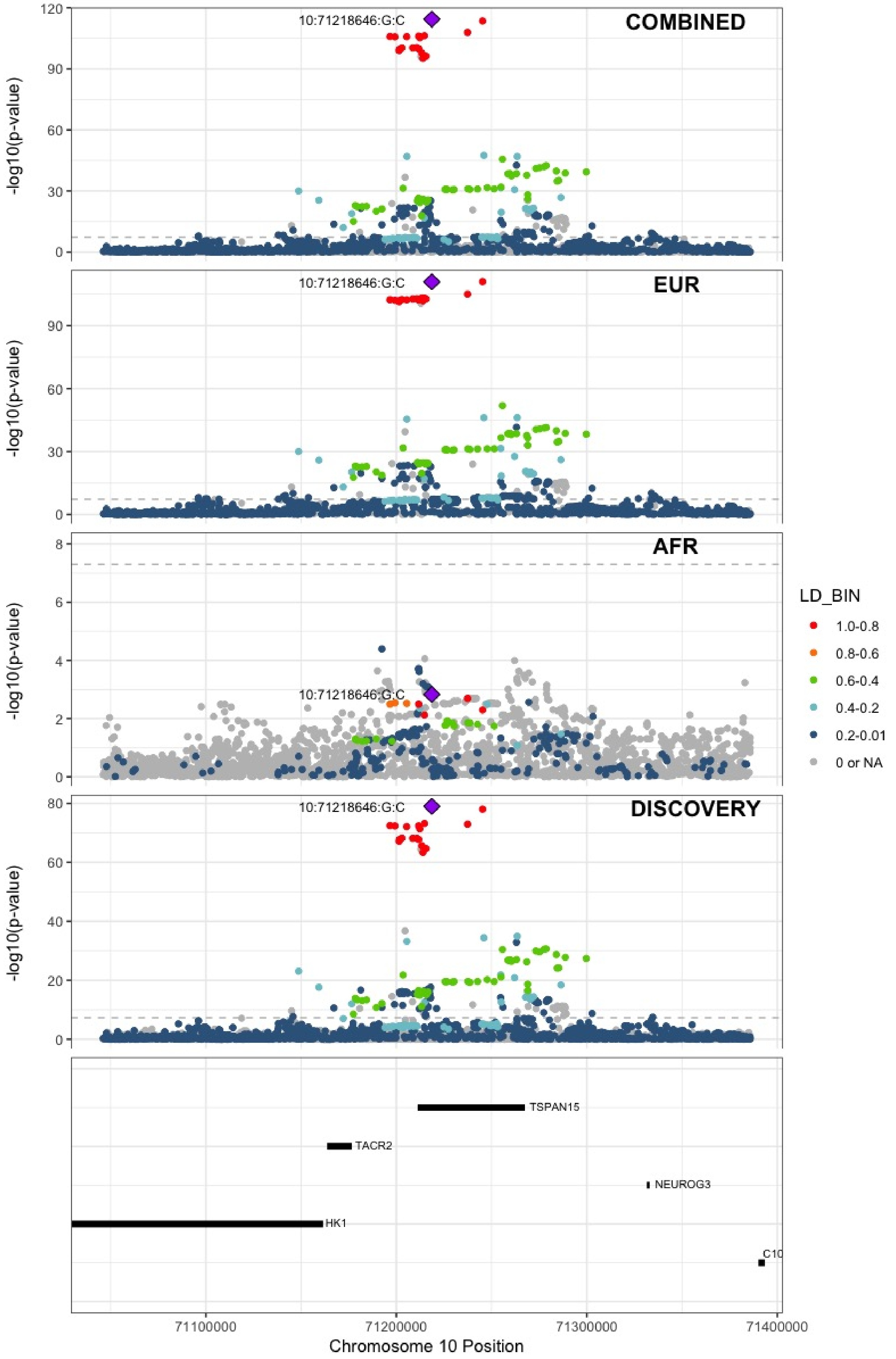

**Supplementary Figure 6.56** – rs16937003 (chr10:80938499) in locus 73 (ZMIZ1)

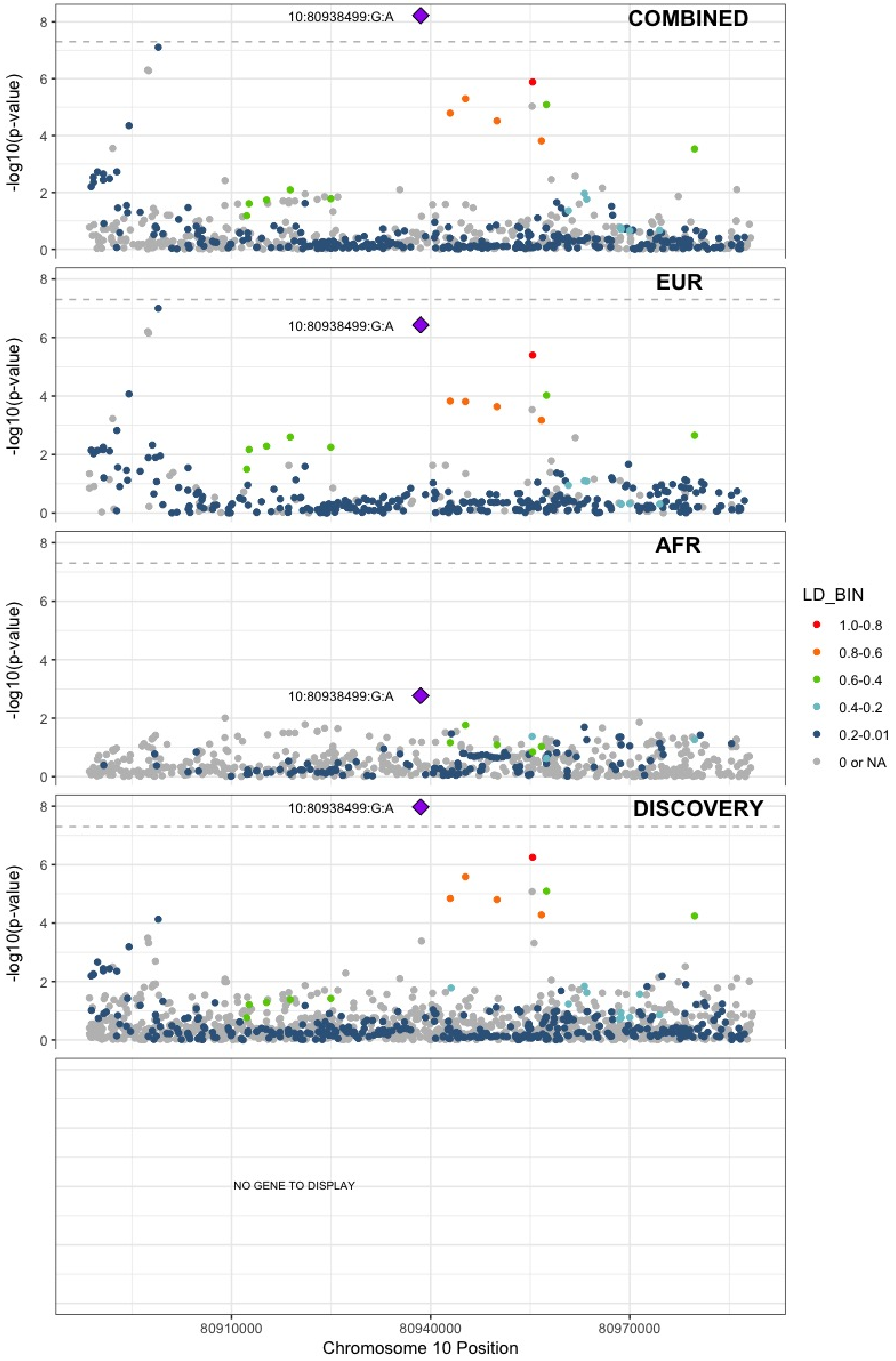

**Supplementary Figure 6.57** – rs57866767 (chr10:96023077) in locus 74 (PLCE1)

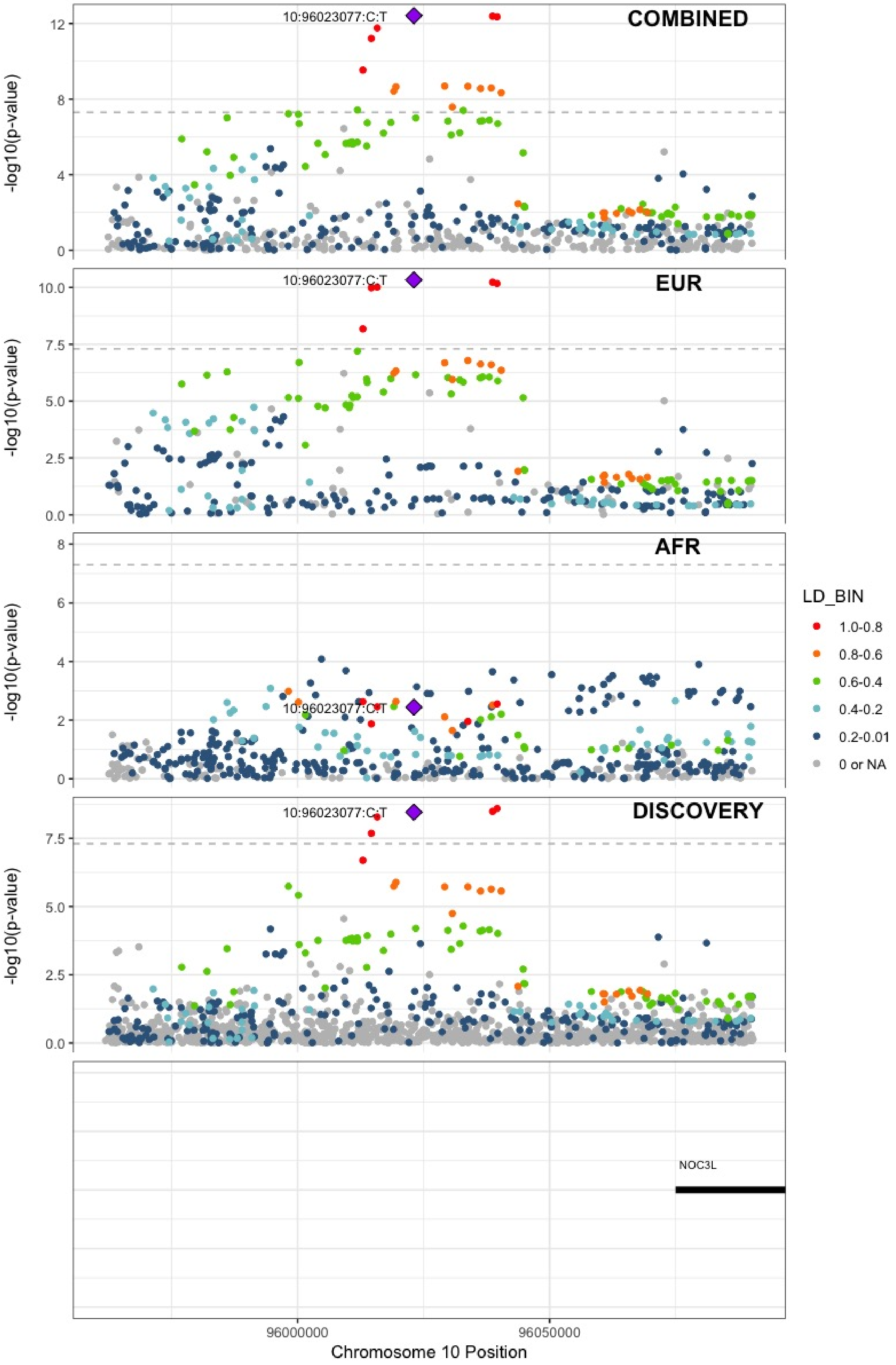

**Supplementary Figure 6.58** – rs10886430 (chr10:121010256) in locus 75 (GRK5)

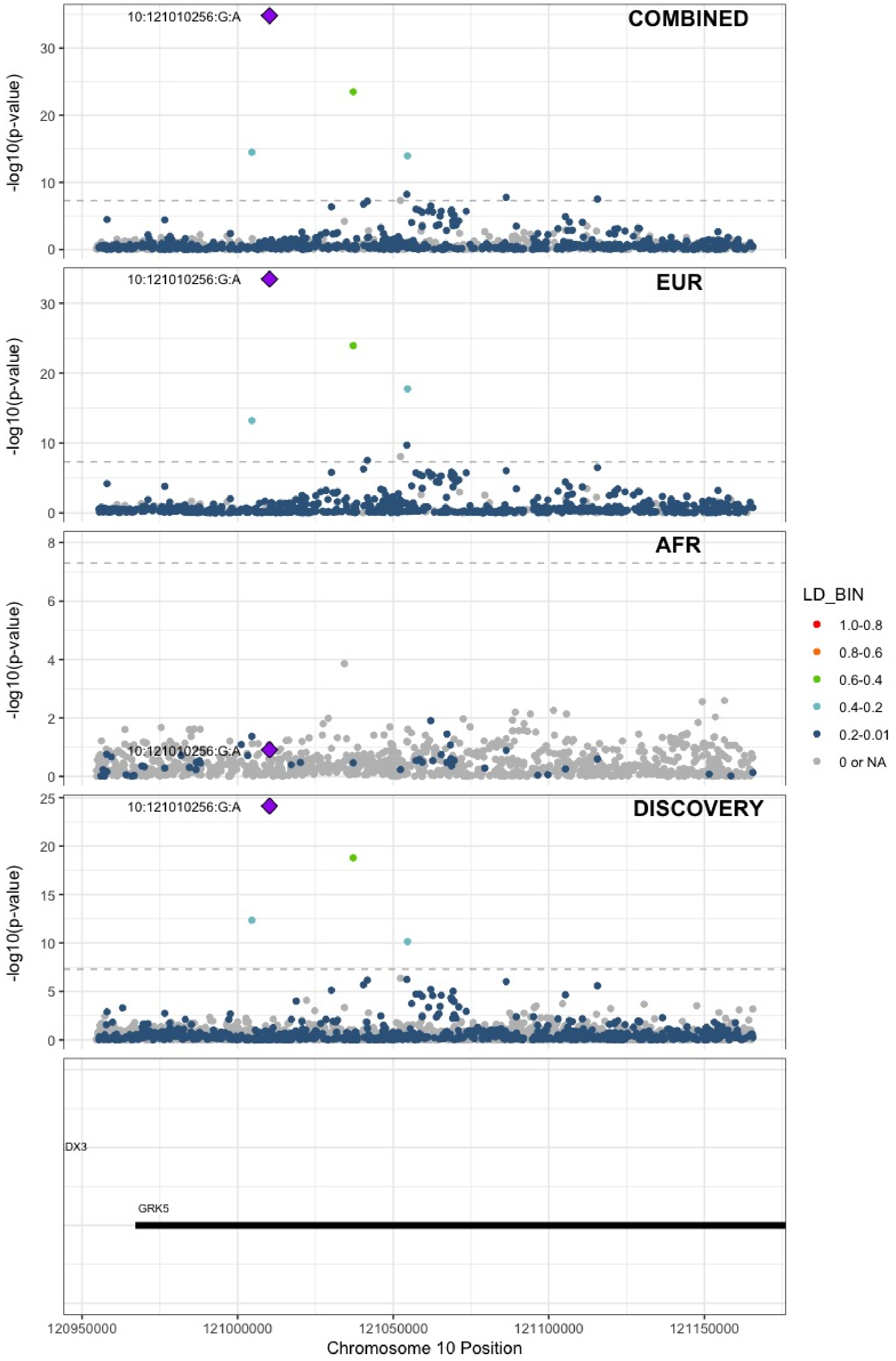

**Supplementary Figure 6.59** – rs2030291 (chr11:16251251) in locus 77 (SOX6)

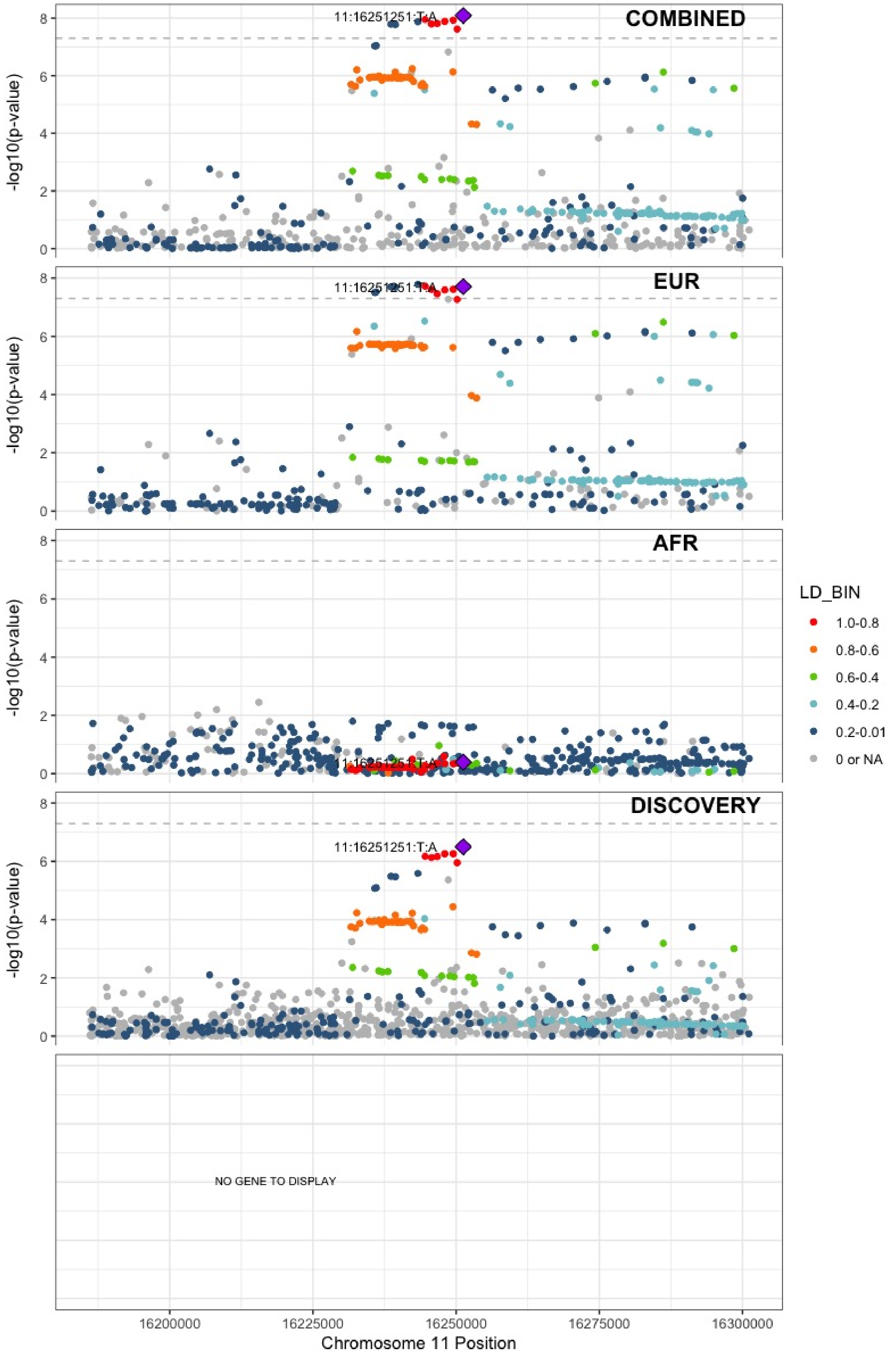

**Supplementary Figure 6.60** – rs11032074 (chr11:32993887) in locus 78 (QSER1)

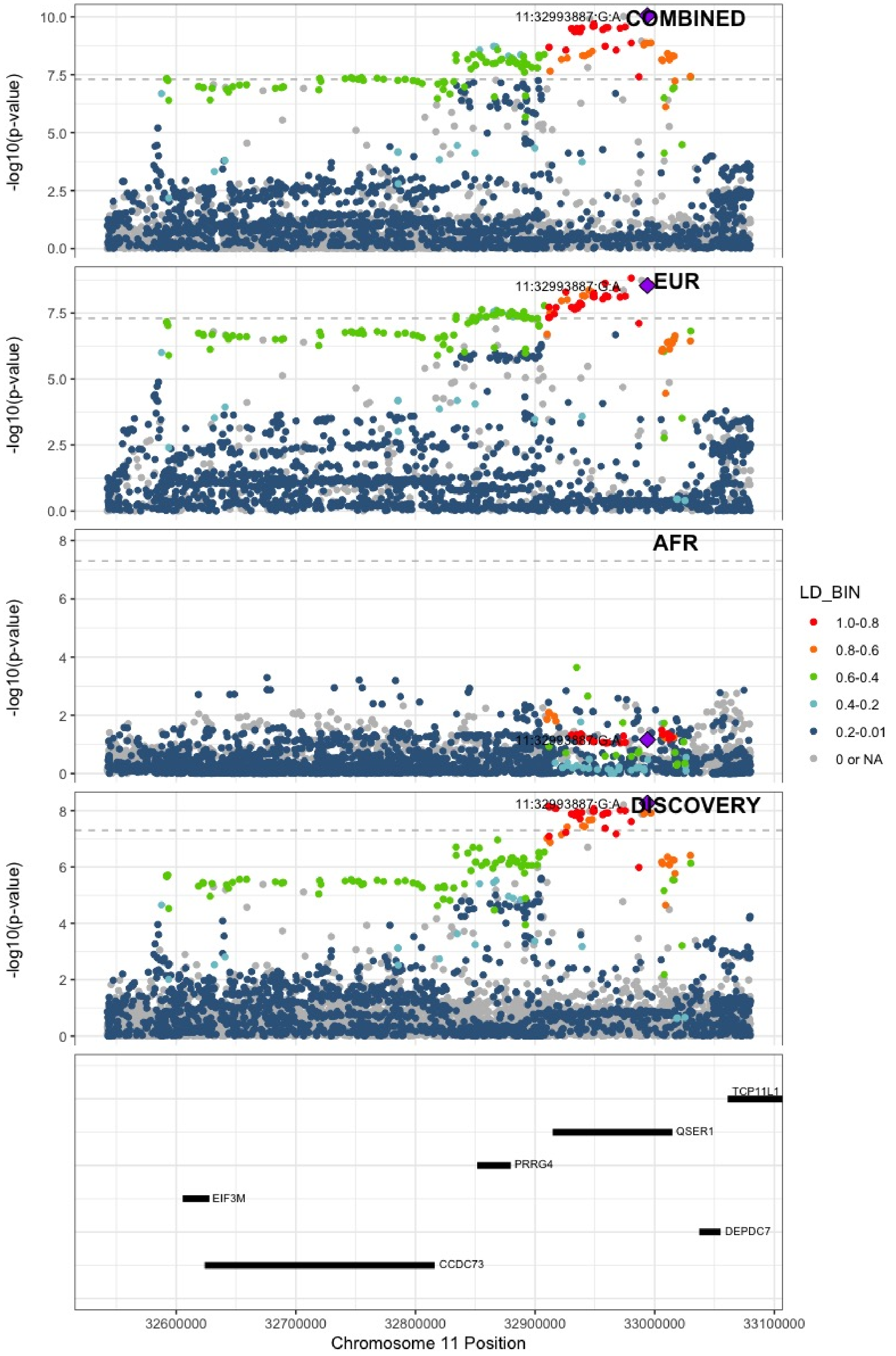

**Supplementary Figure 6.61** – rs1799963 (chr11:46761055) in locus 79 (F2)

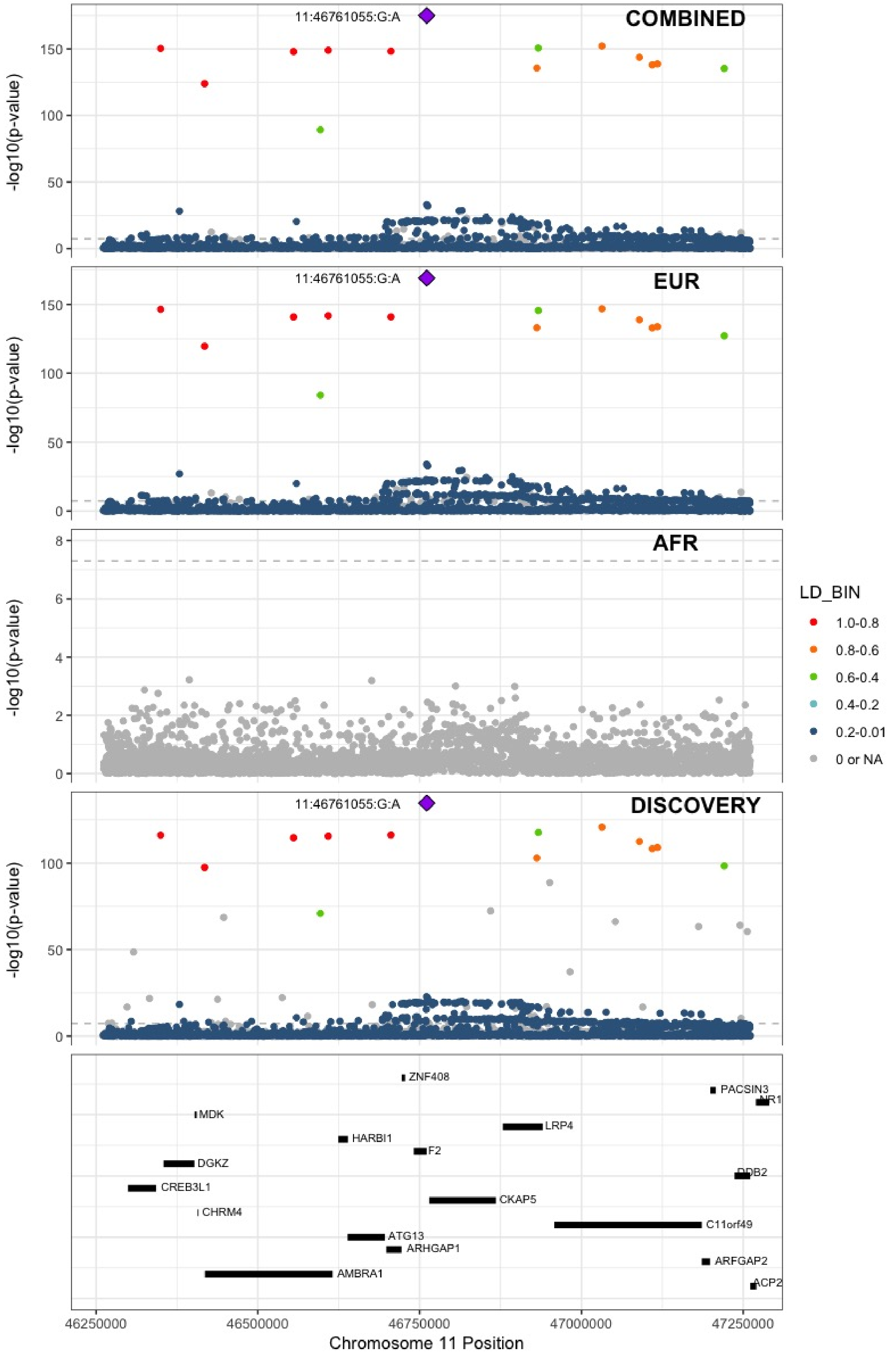

**Supplementary Figure 6.62** – rs141798115 (chr11:56875074) in locus 80 (OR5AK4P ;LRRC55)

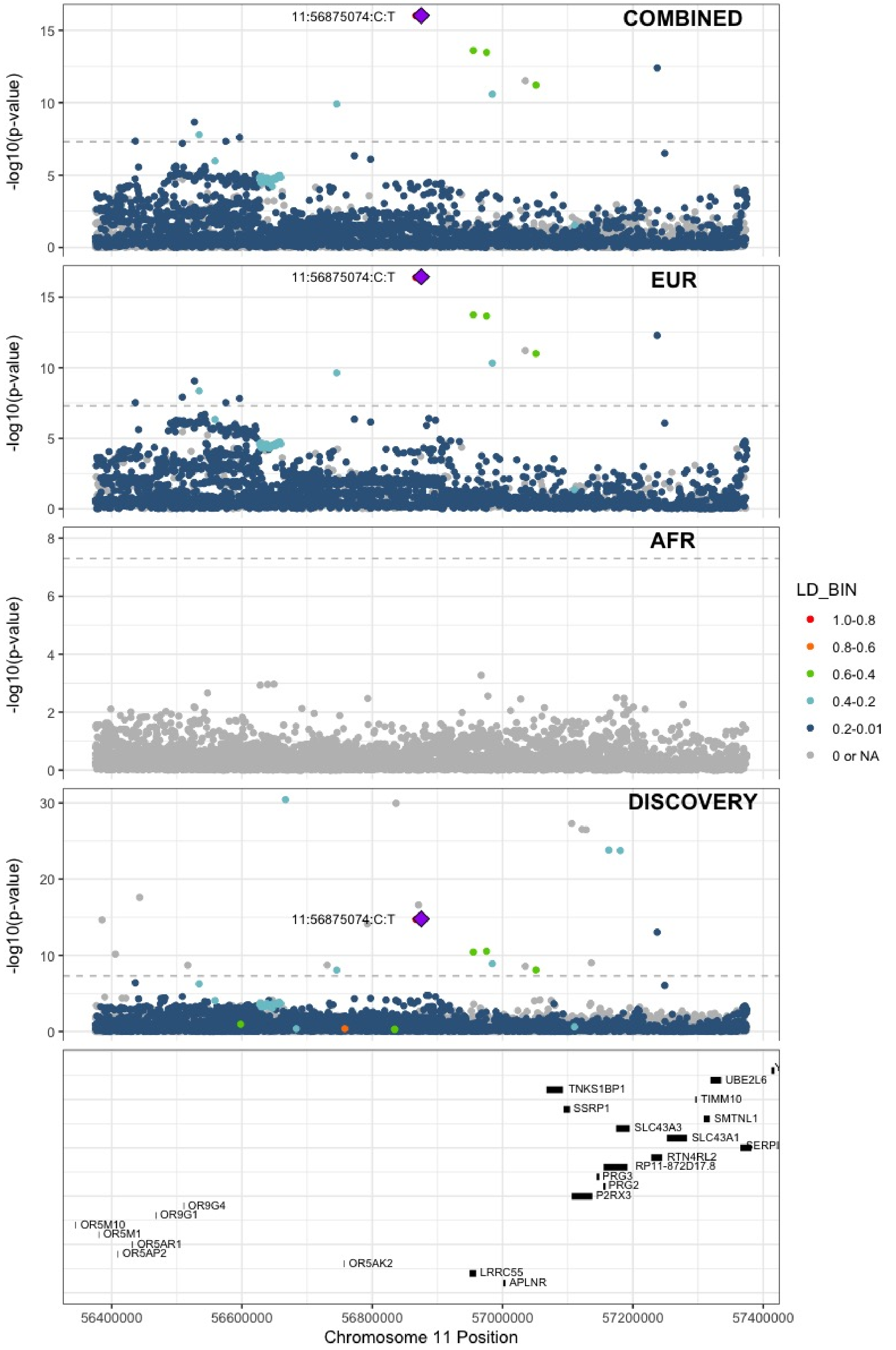

**Supplementary Figure 6.63** – rs4354705 (chr11:60088159) in locus 81 (MS4A4A ;MS4A6E)

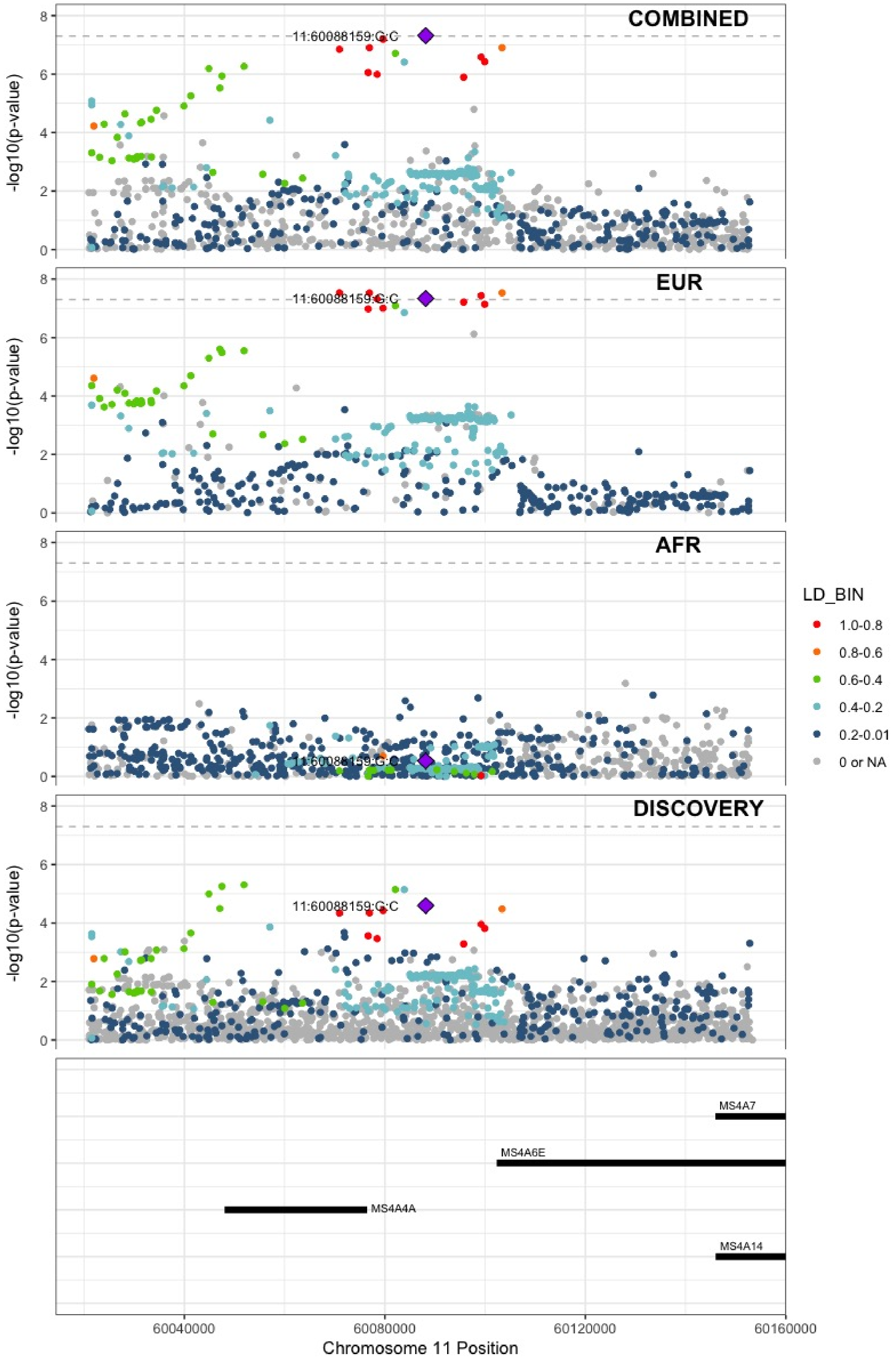

**Supplementary Figure 6.64** – rs174551 (chr11:61573684) in locus 82 (FADS1)

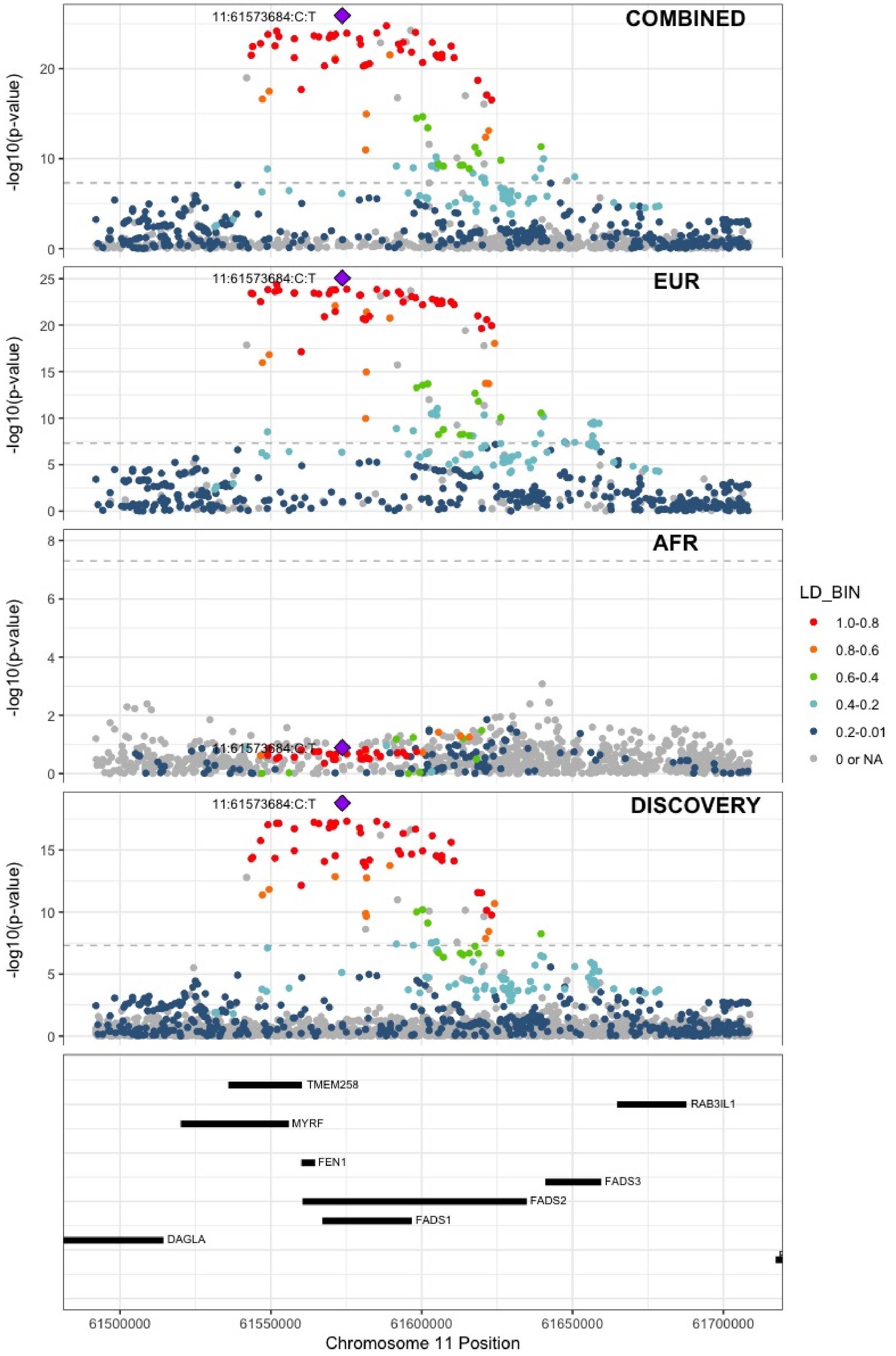

**Supplementary Figure 6.65** – rs2846027 (chr11:114003415) in locus 84 (ZBTB16)

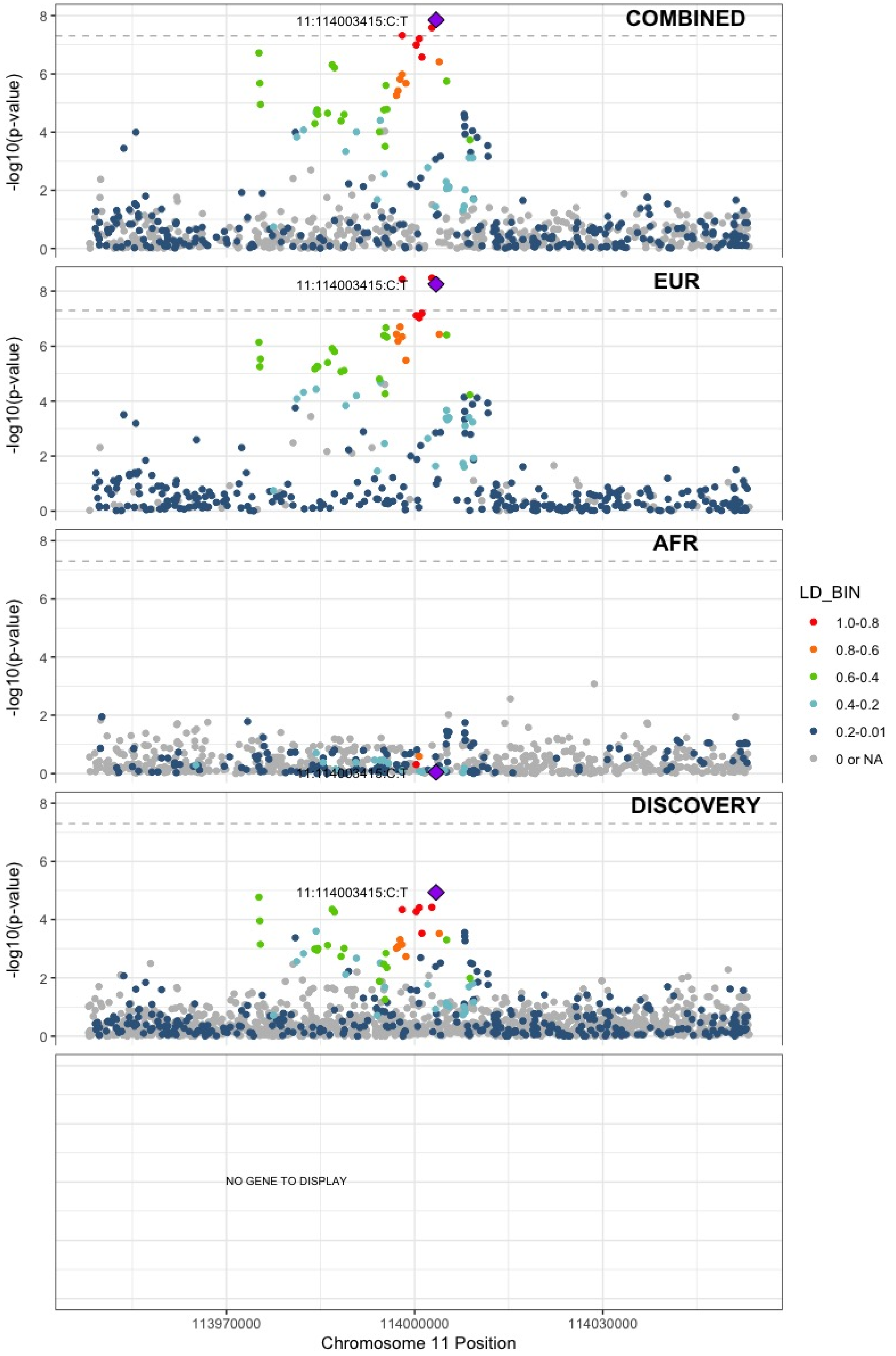

**Supplementary Figure 6.66** – rs35257264 (chr11:126296816) in locus 85 (ST3GAL4)

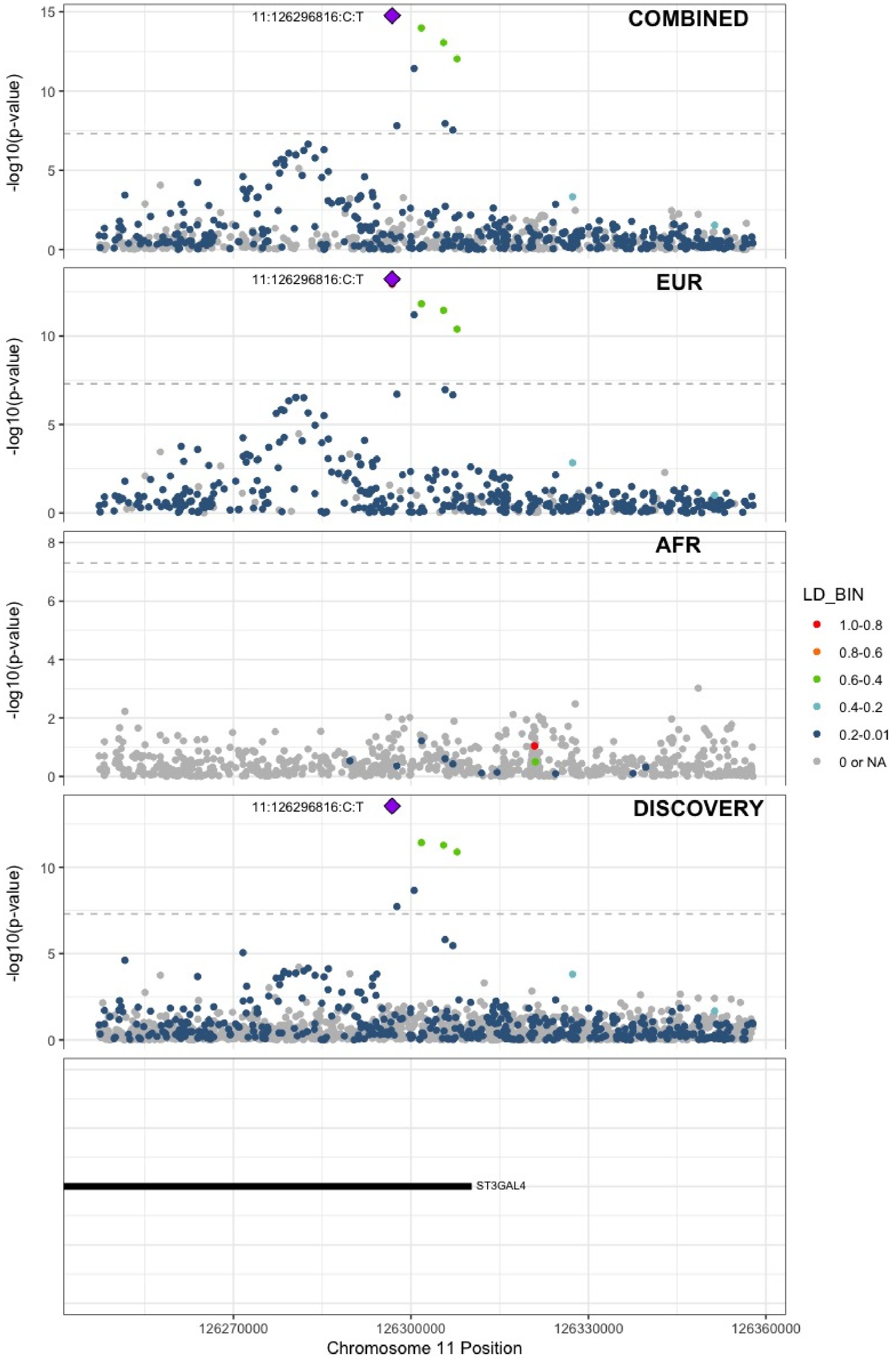

**Supplementary Figure 6.67** – rs7107568 (chr11:130779668) in locus 86 (SNX19)

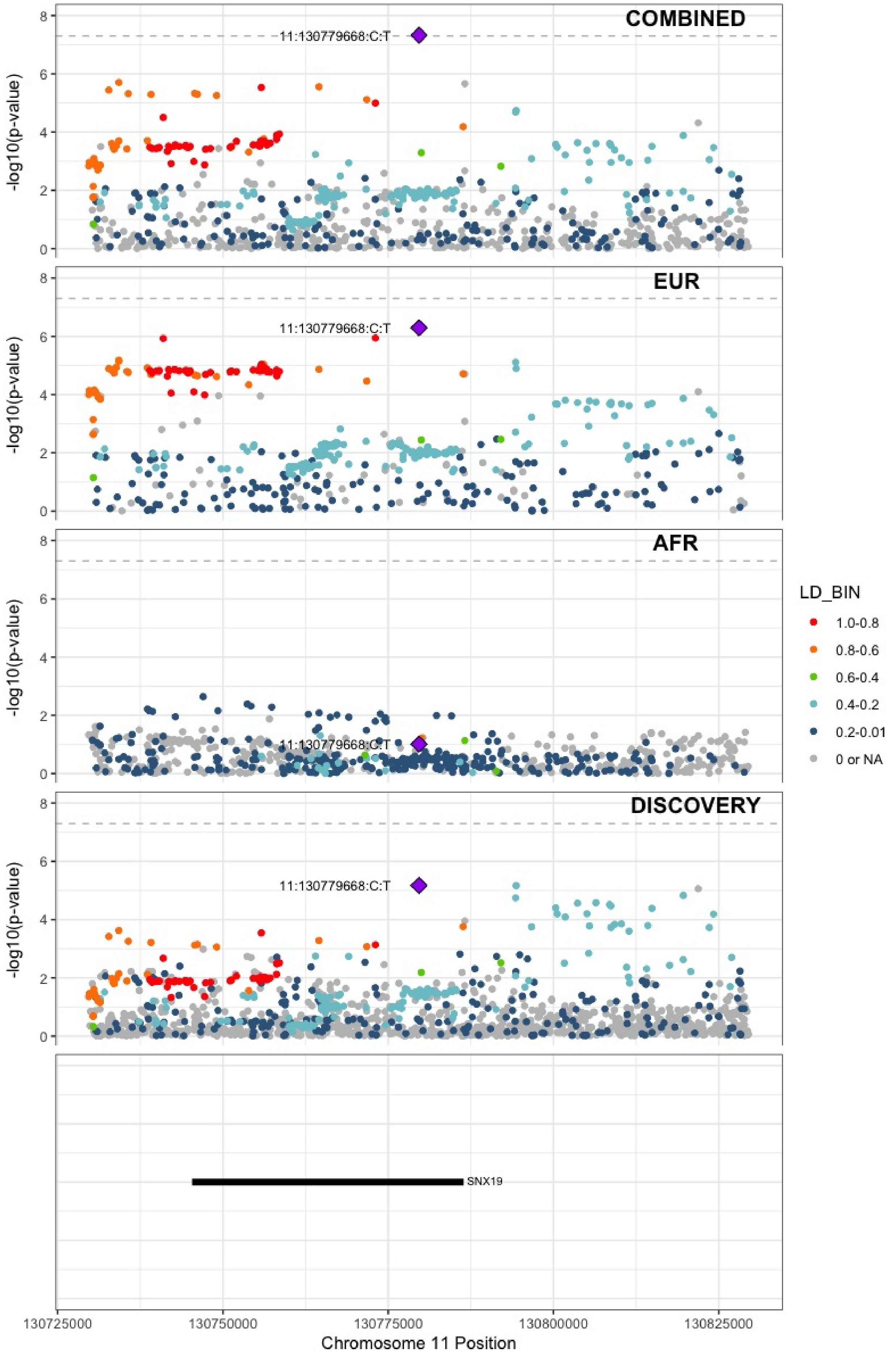

**Supplementary Figure 6.68** – rs1558519 (chr12:6153738) in locus 87 (VWF)

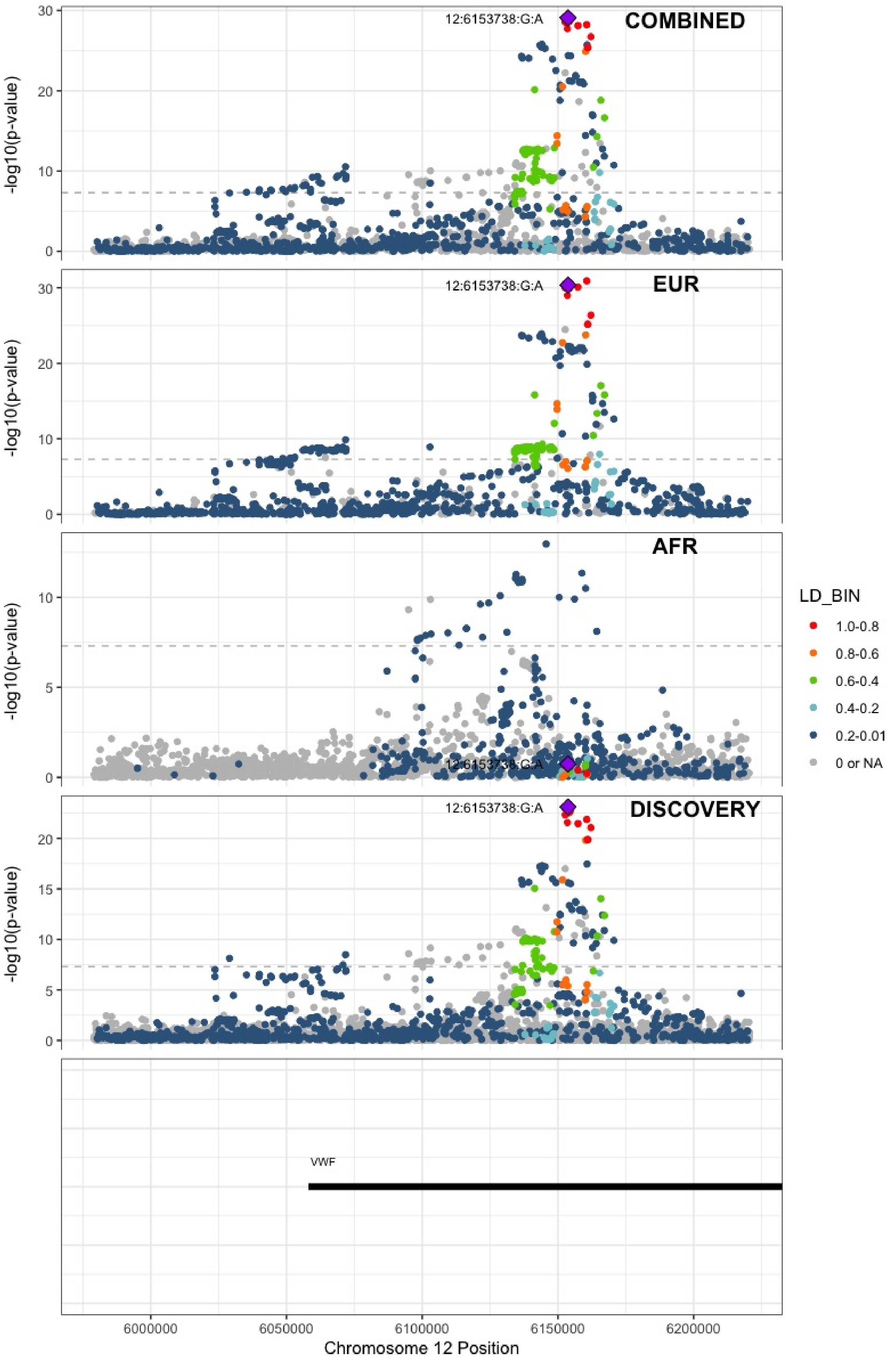

**Supplementary Figure 6.69** – rs7311483 (chr12:9053661) in locus 88 (A2ML1 ;PHC1)

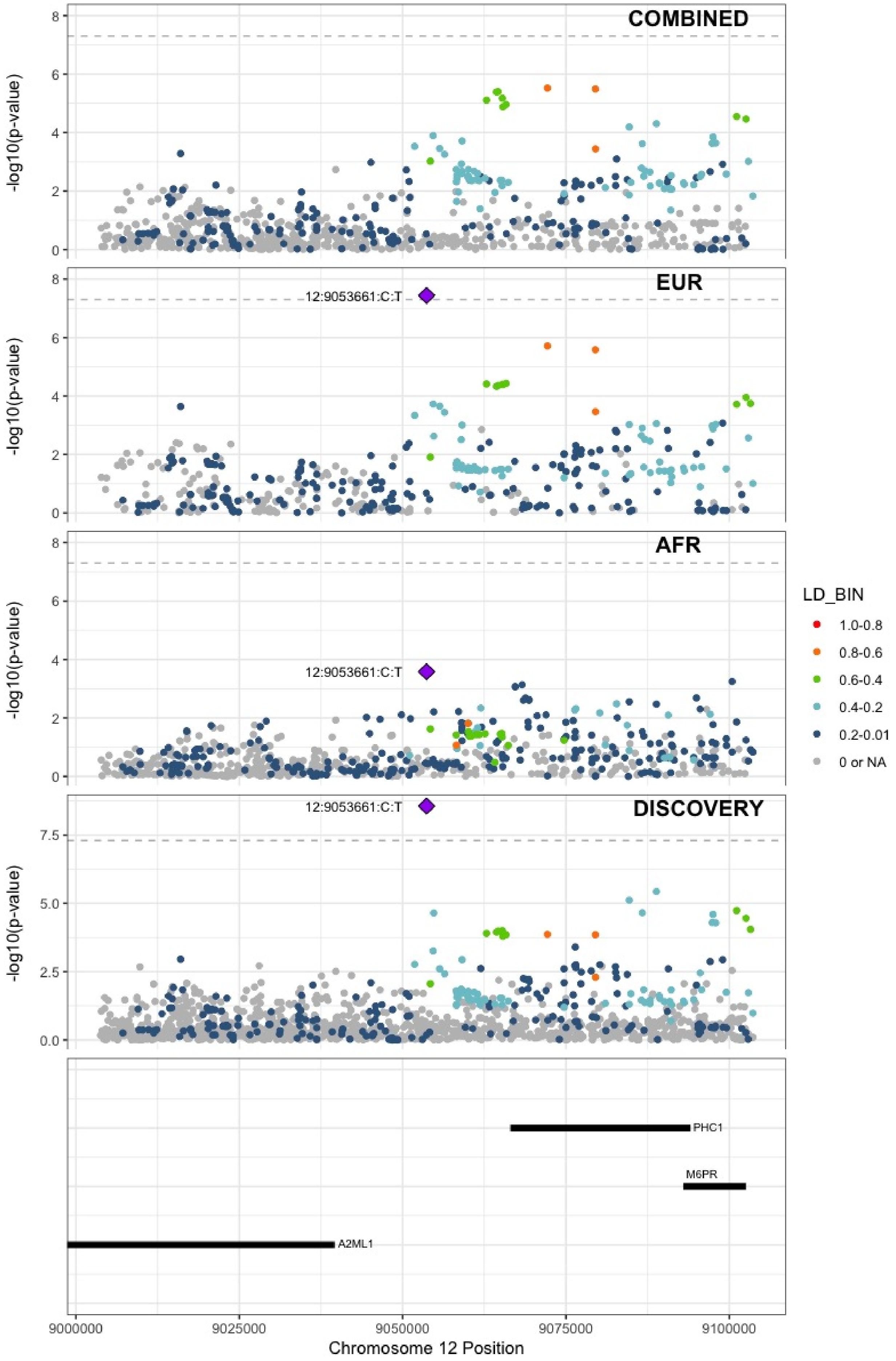

**Supplementary Figure 6.70** – rs6580981 (chr12:54723028) in locus 89 (COPZ1)

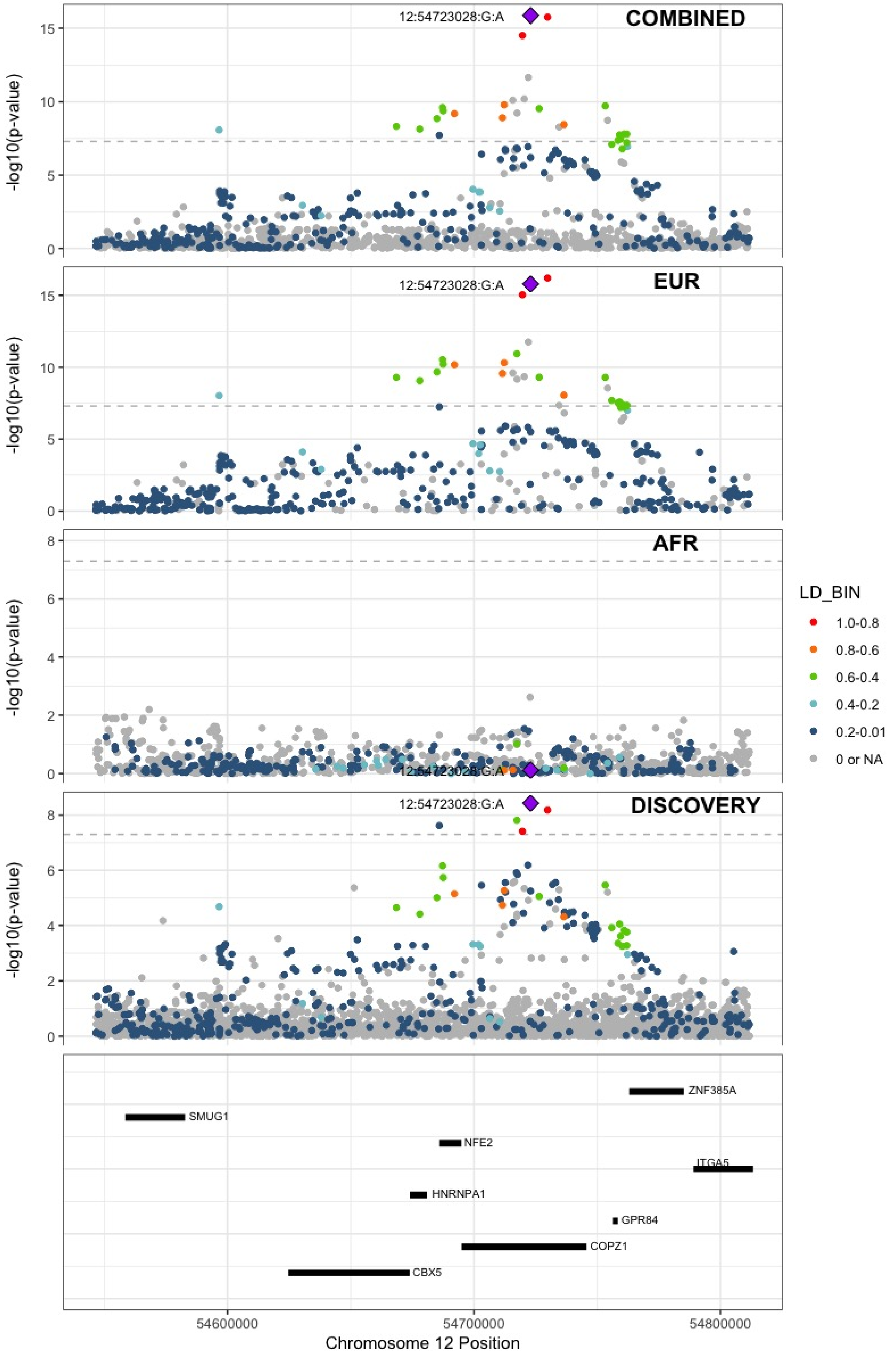

**Supplementary Figure 6.71** – rs142351376 (chr12:104136288) in locus 90 (STAB2)

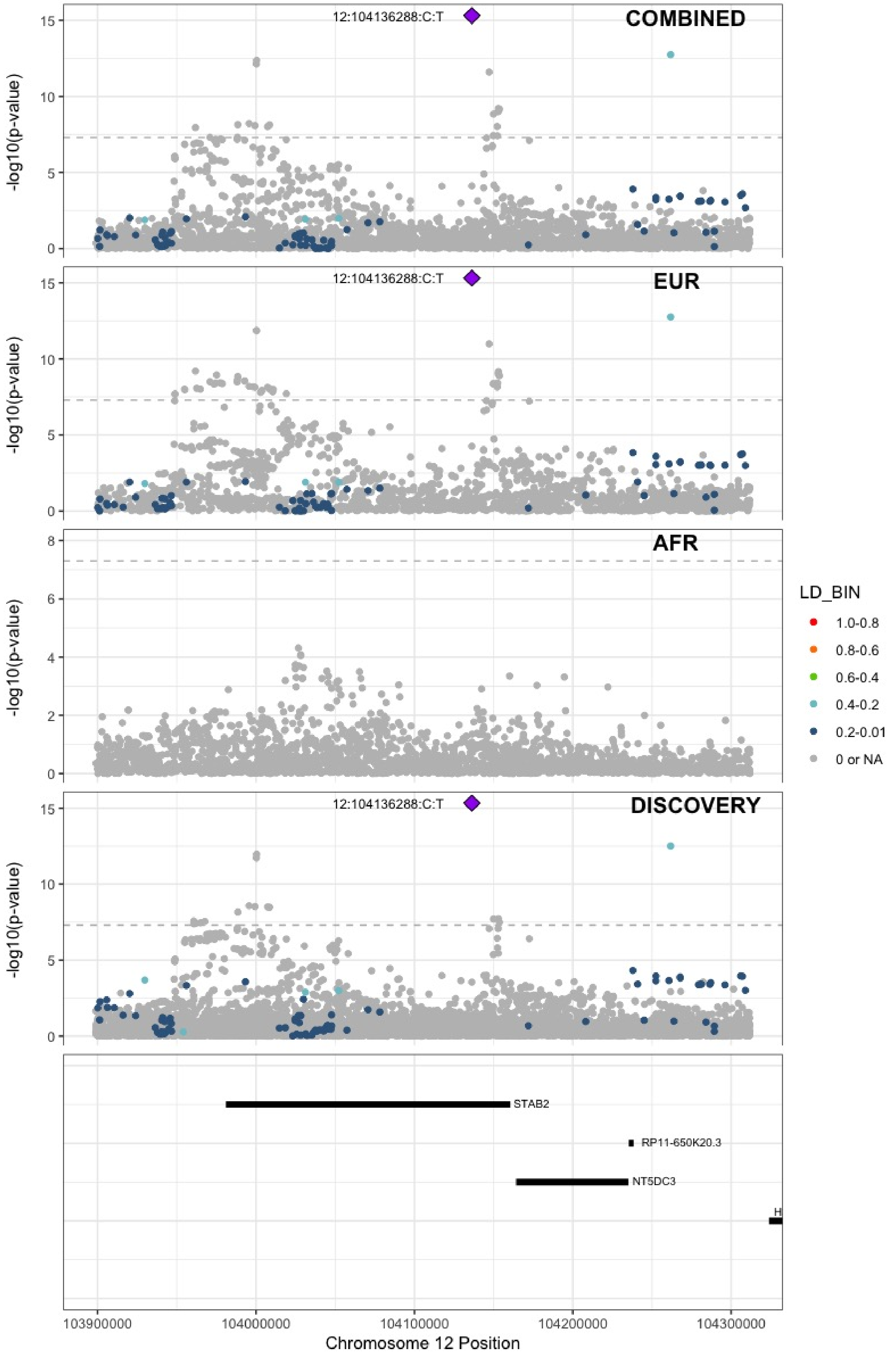

**Supplementary Figure 6.72** – rs3184504 (chr12:111884608) in locus 91 (SH2B3)

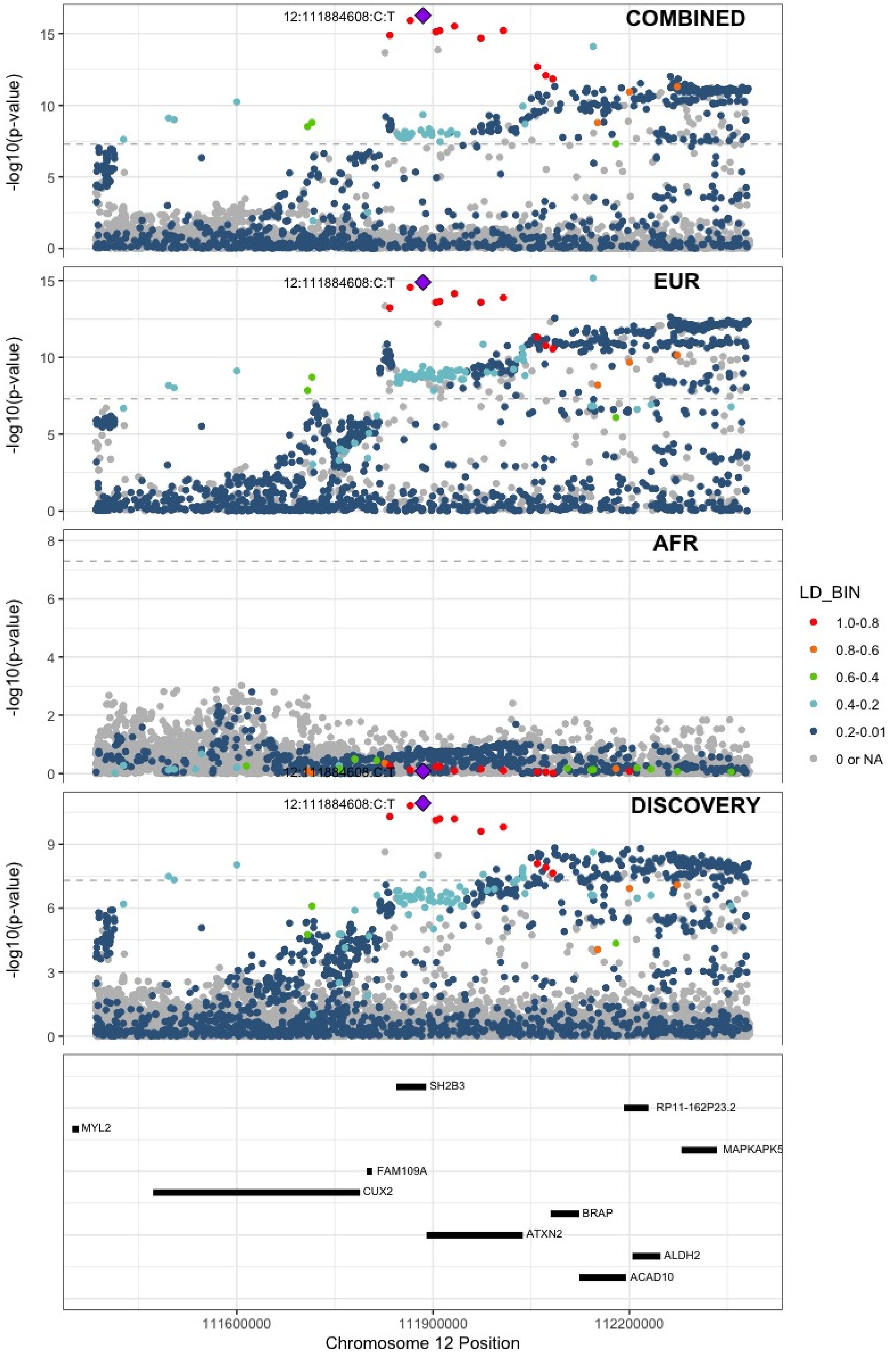

**Supplementary Figure 6.73** – rs2851435 (chr12:123712416) in locus 92 (MPHOSPH9)

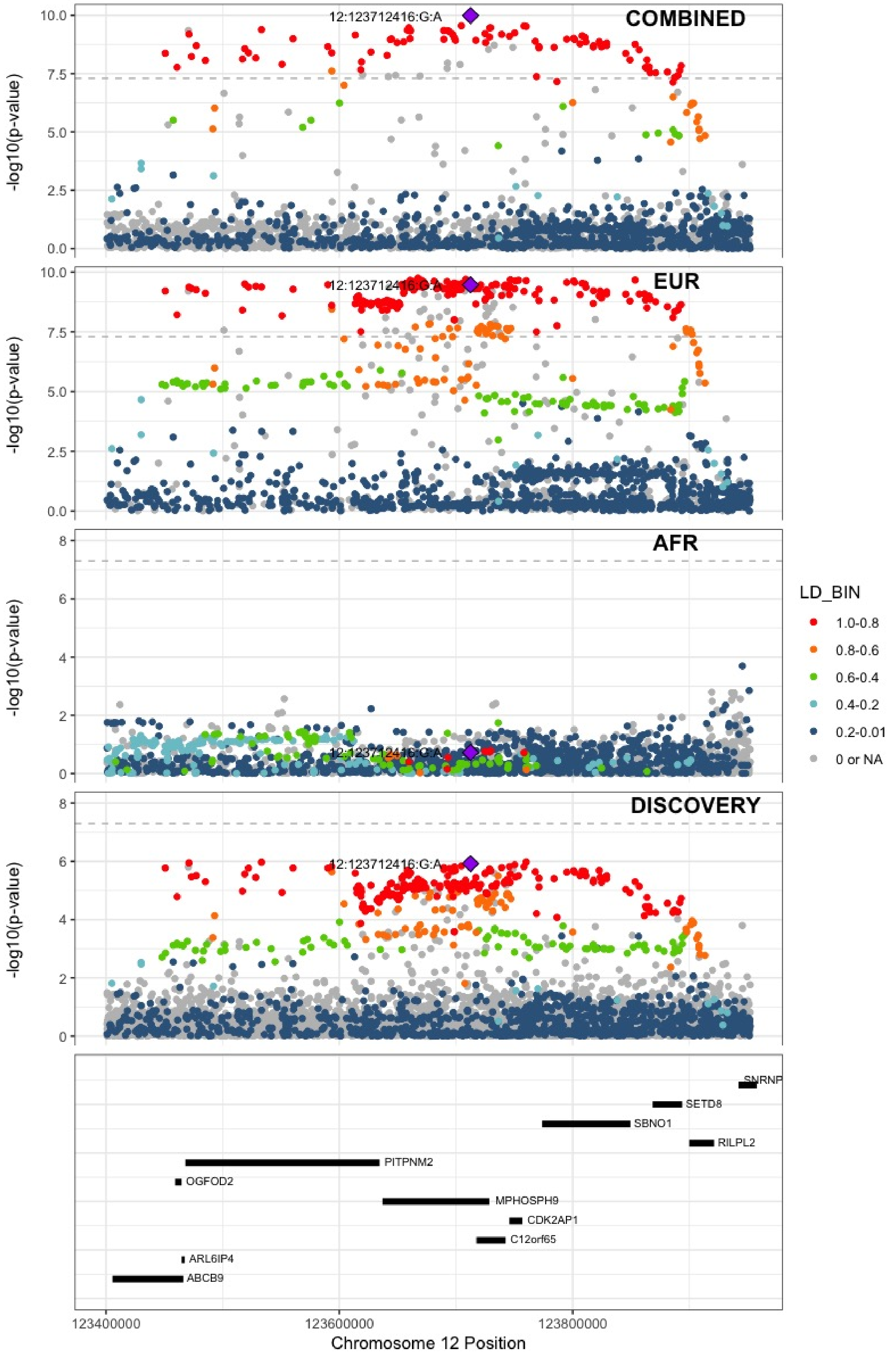

**Supplementary Figure 6.74** – rs3211752 (chr13:113787459) in locus 94 (F10)

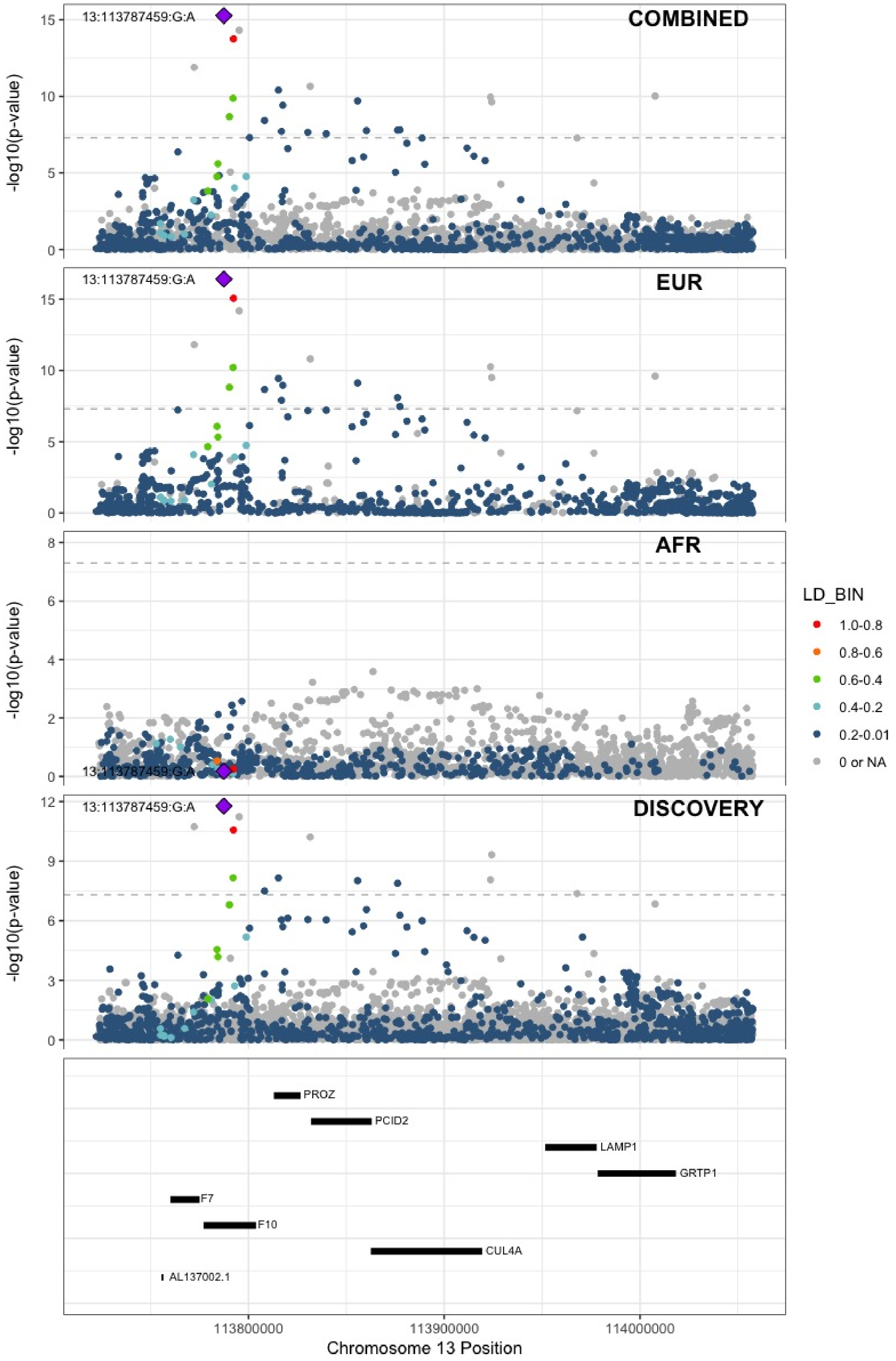

**Supplementary Figure 6.75** – rs11158204 (chr14:58844526) in locus 95 (ARID4A ;TOMM20L)

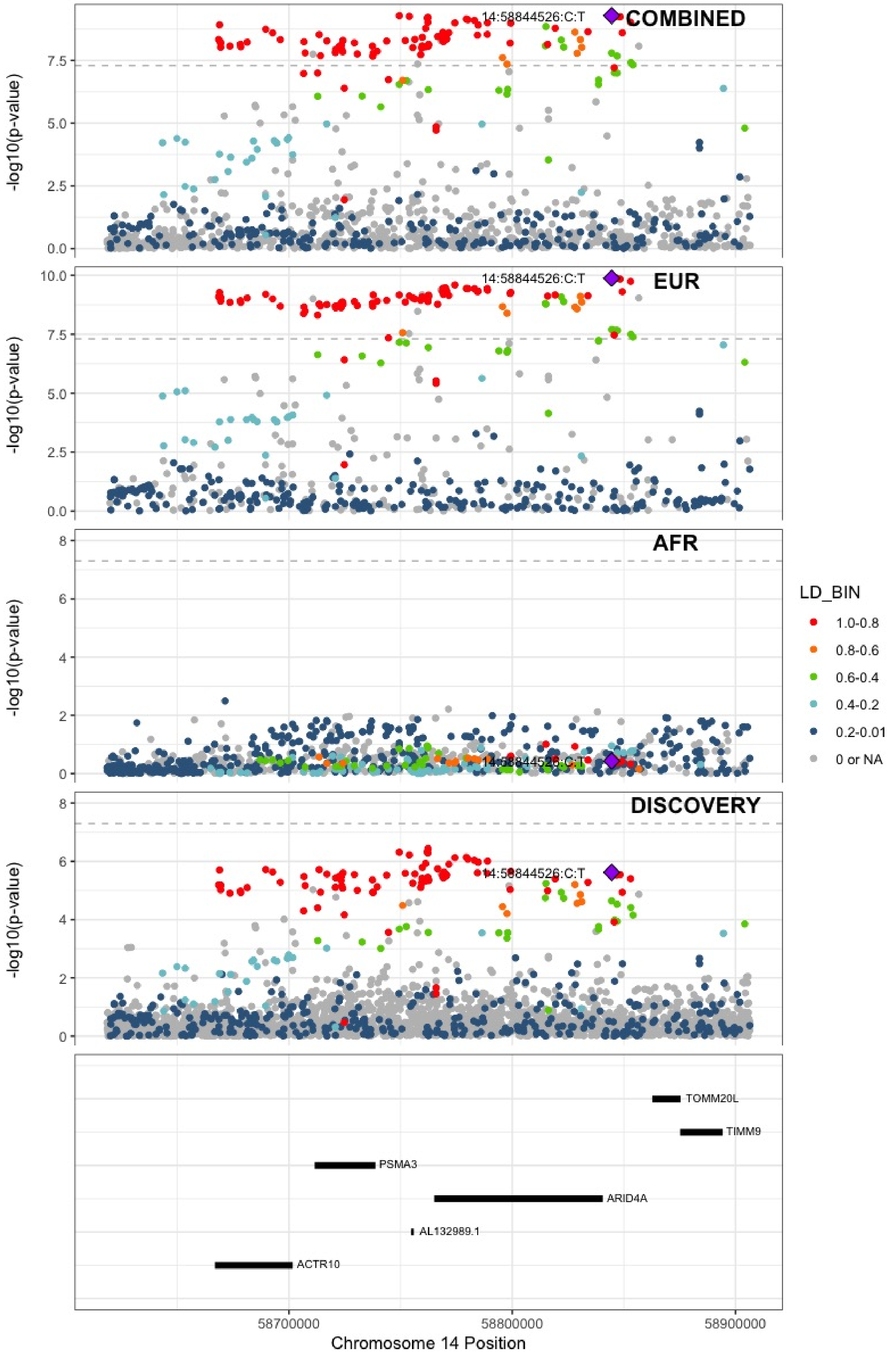

**Supplementary Figure 6.76** – rs2127869 (chr14:65794352) in locus 96 (LINC02324;MIR4708)

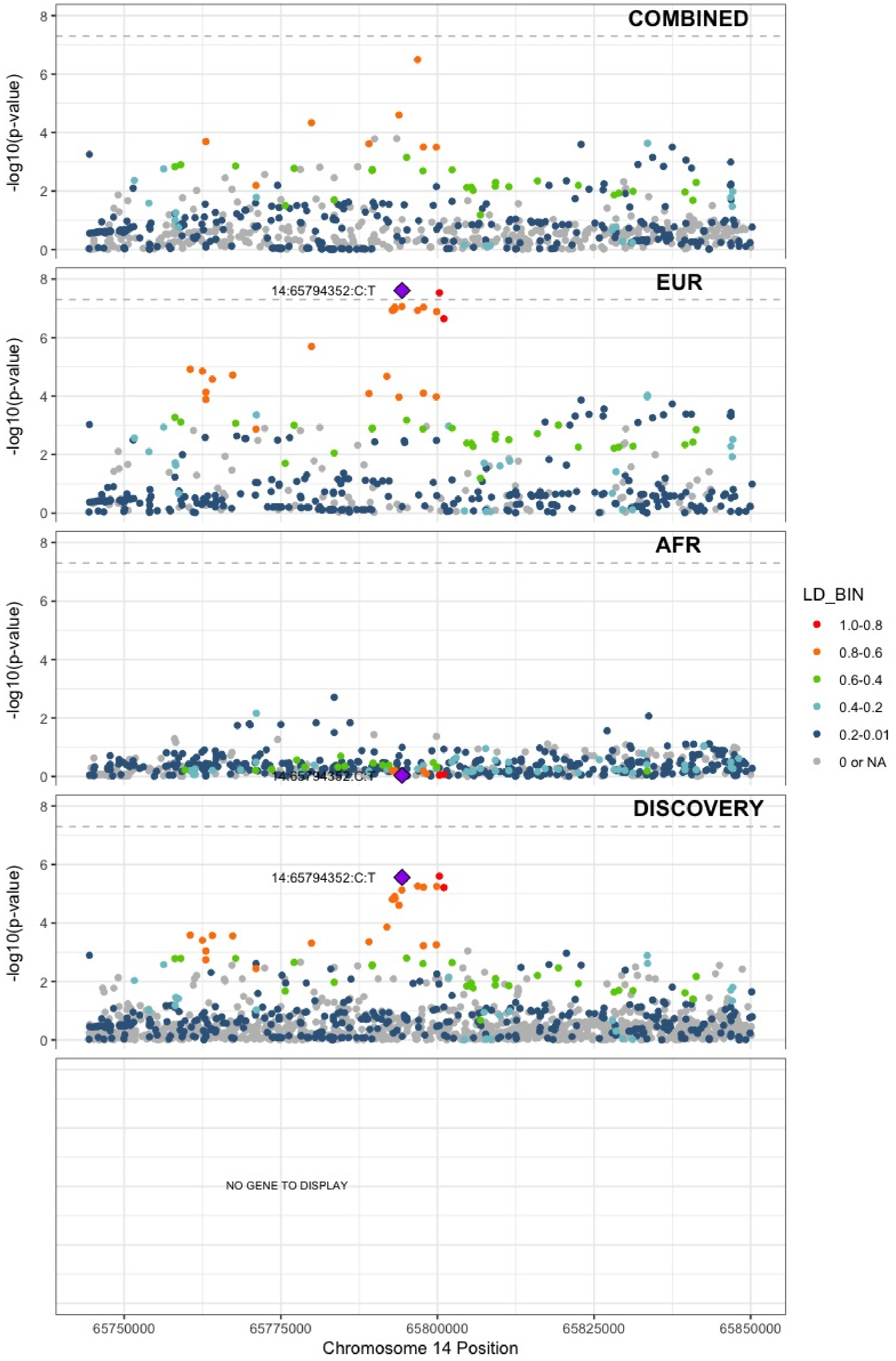

**Supplementary Figure 6.77** – rs7152641 (chr14:92213967) in locus 97 (CATSPERB ;TC2N)

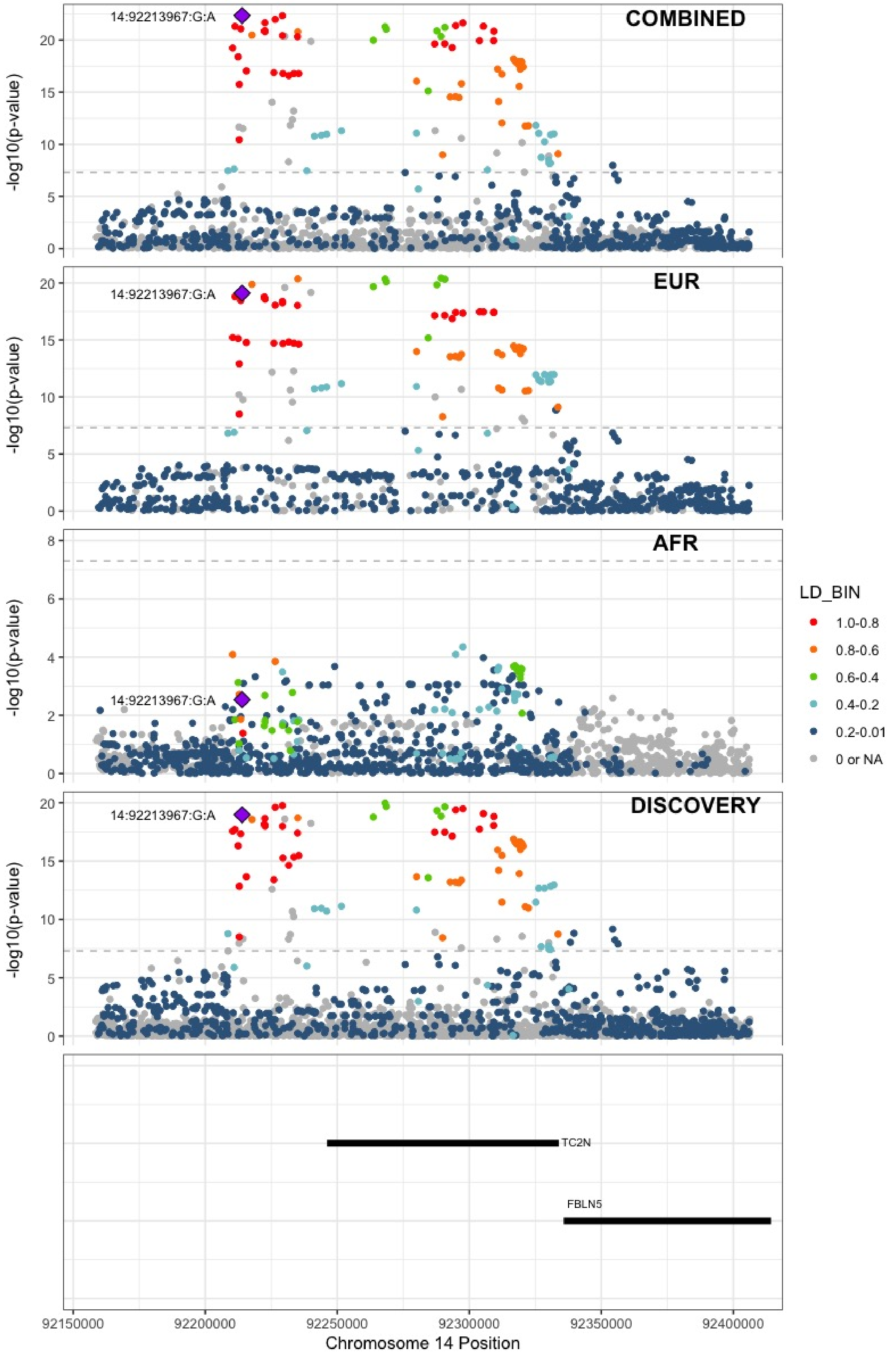

**Supplementary Figure 6.78** – rs28929474 (chr14:94844947) in locus 98 (SERPINA1)

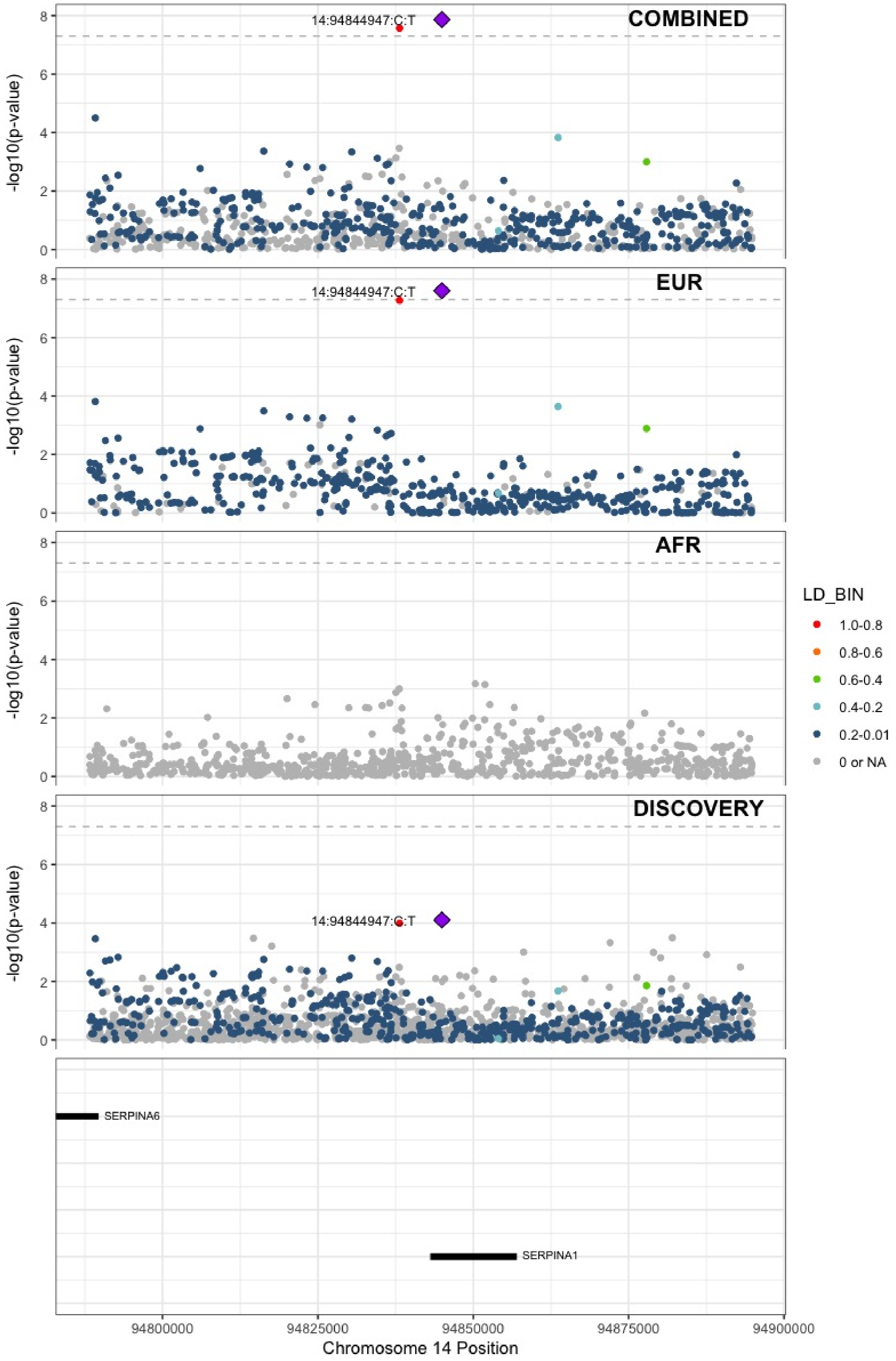

**Supplementary Figure 6.79** – rs28519971 (chr14:103089808) in locus 99 (RCOR1)

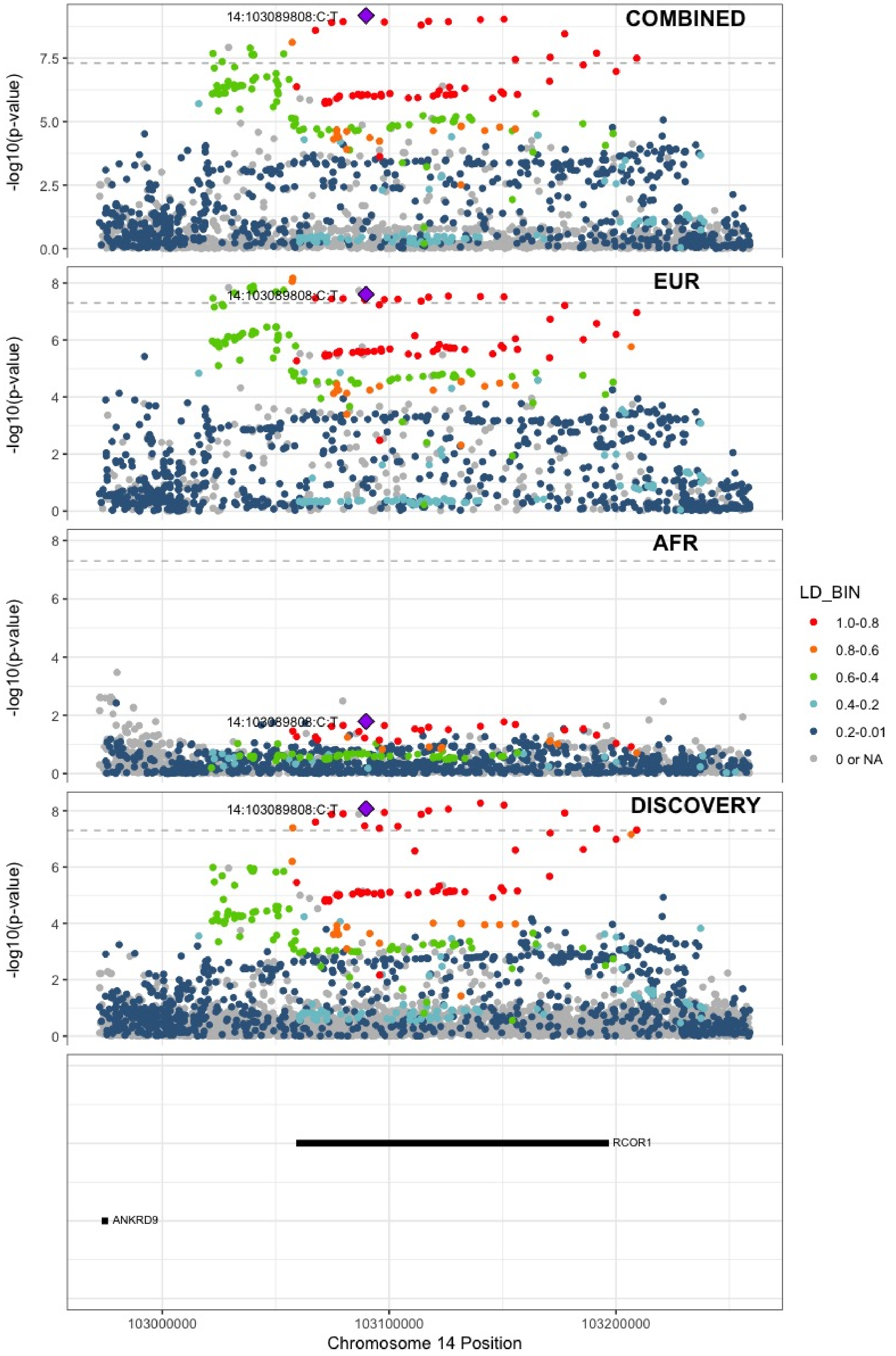

**Supplementary Figure 6.80** – rs1247656151 (chr15:43677975) in locus 100 (TUBGCP4)

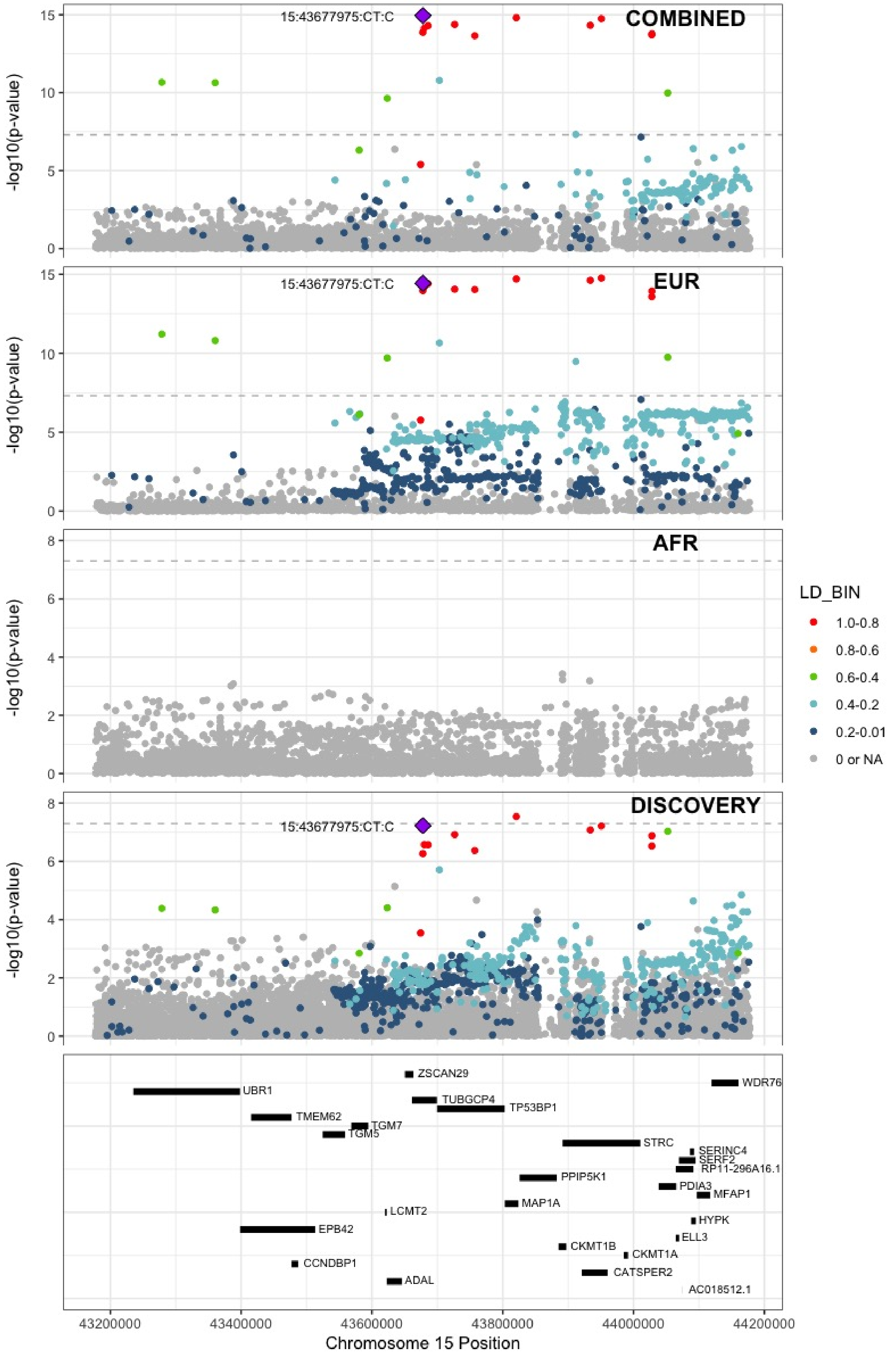

**Supplementary Figure 6.81** – rs7183672 (chr15:96101018) in locus 103 (LINC00924 ;LOC105369212)

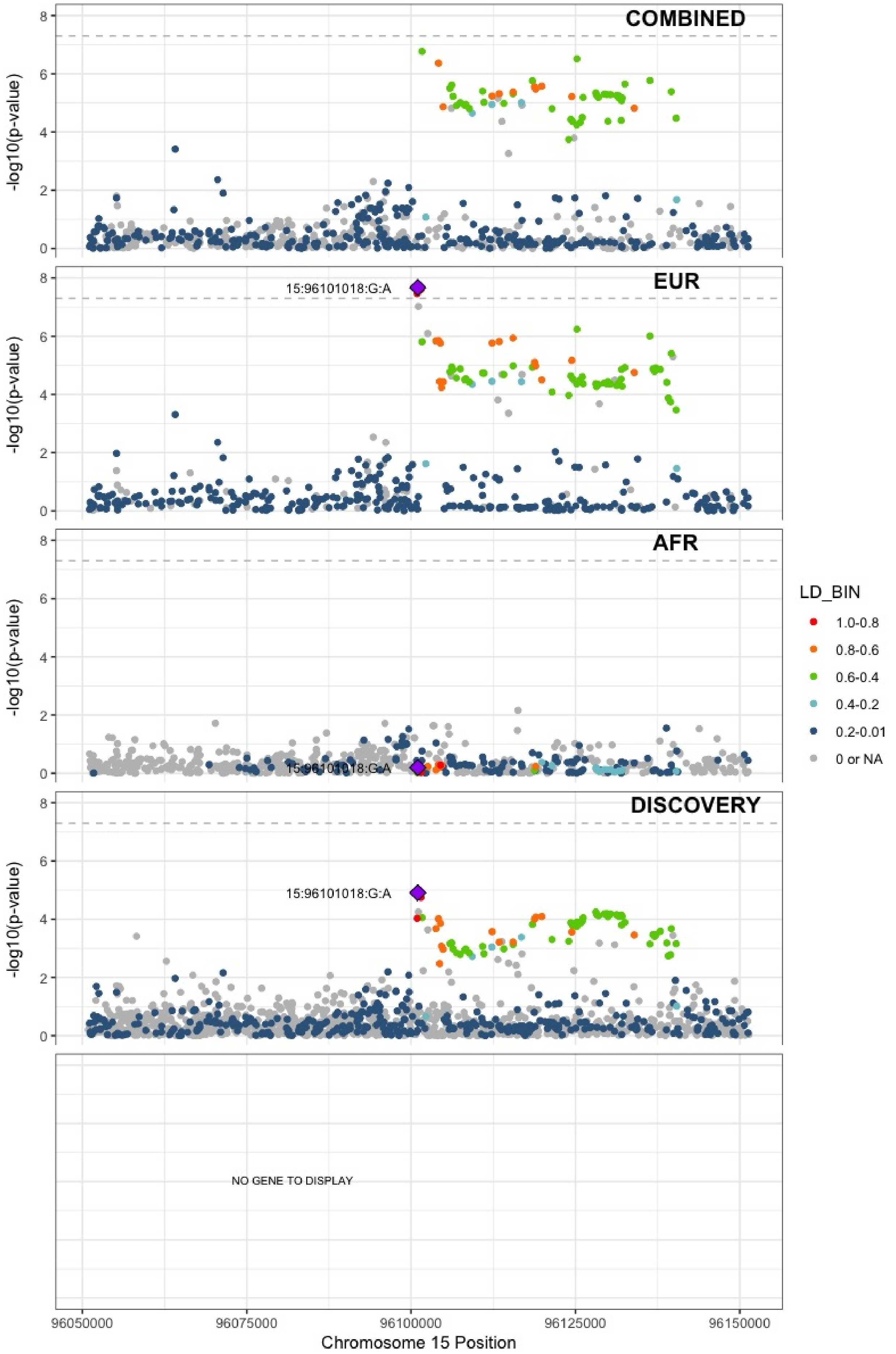

**Supplementary Figure 6.82** – rs71376077 (chr16:15738114) in locus 105 (NDE1)

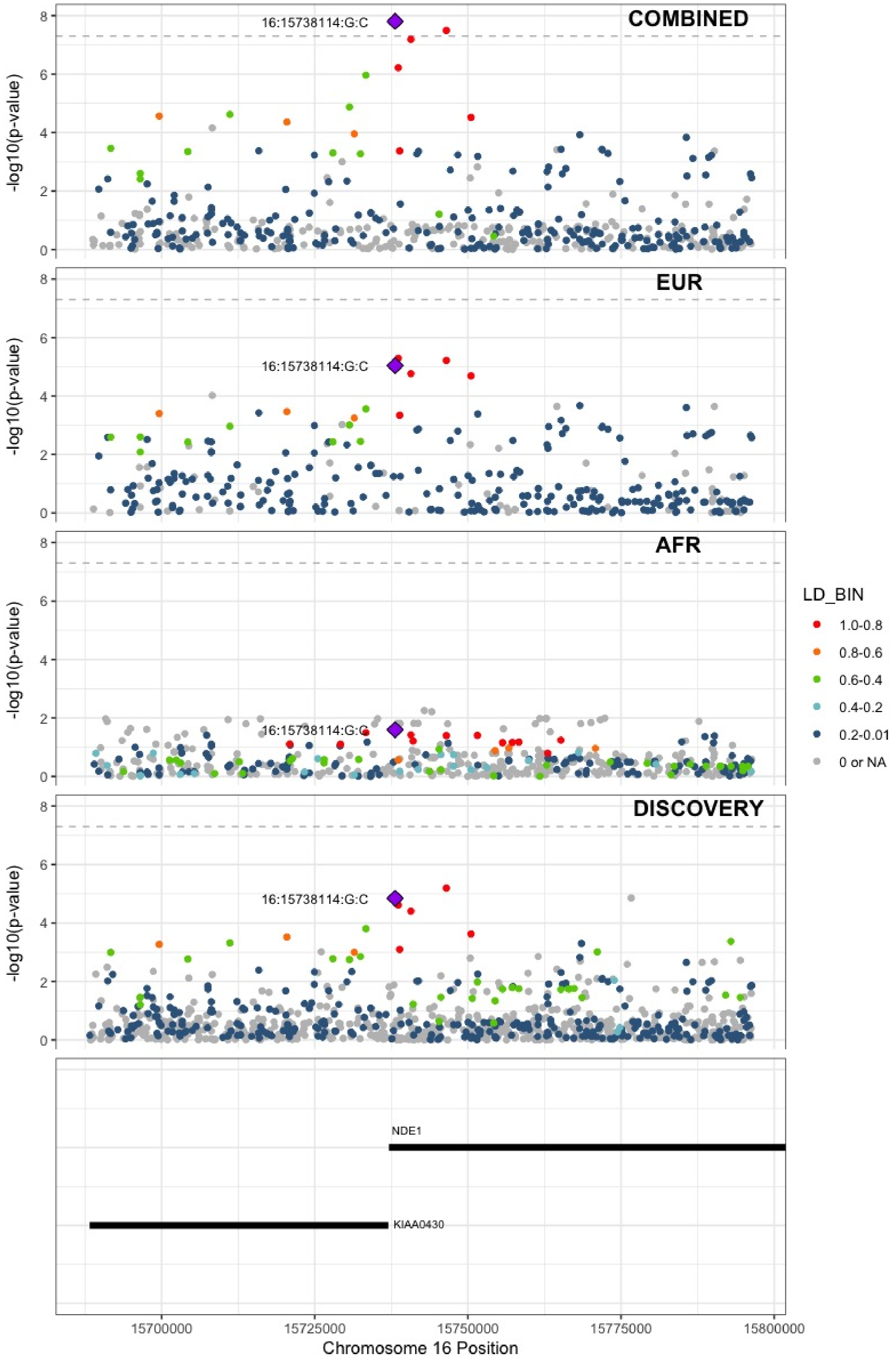

**Supplementary Figure 6.83** – rs12443808 (chr16:30996871) in locus 106 (HSD3B7)

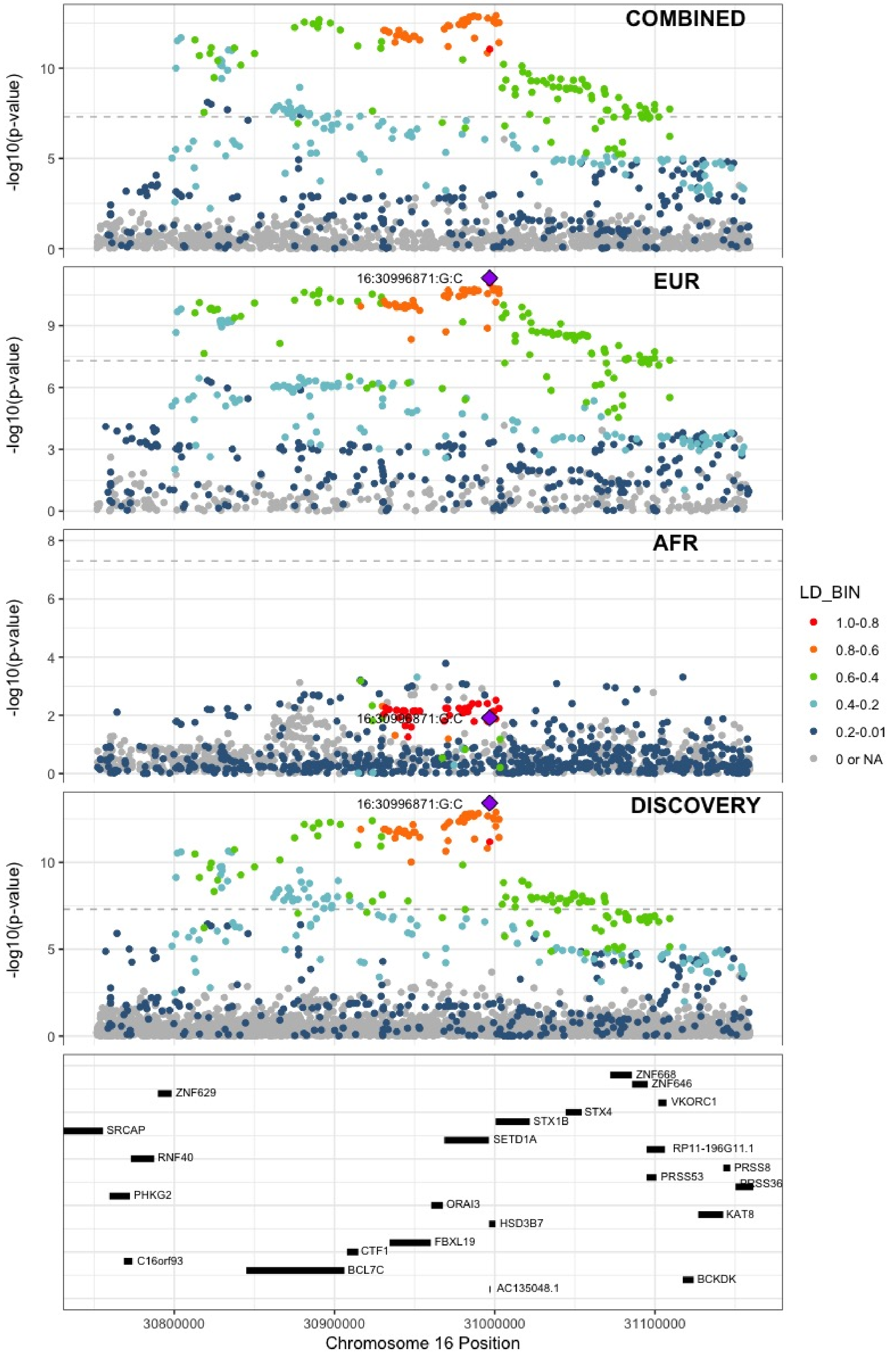

**Supplementary Figure 6.84** – rs7197453 (chr16:72079127) in locus 107 (DHODH ;HP)

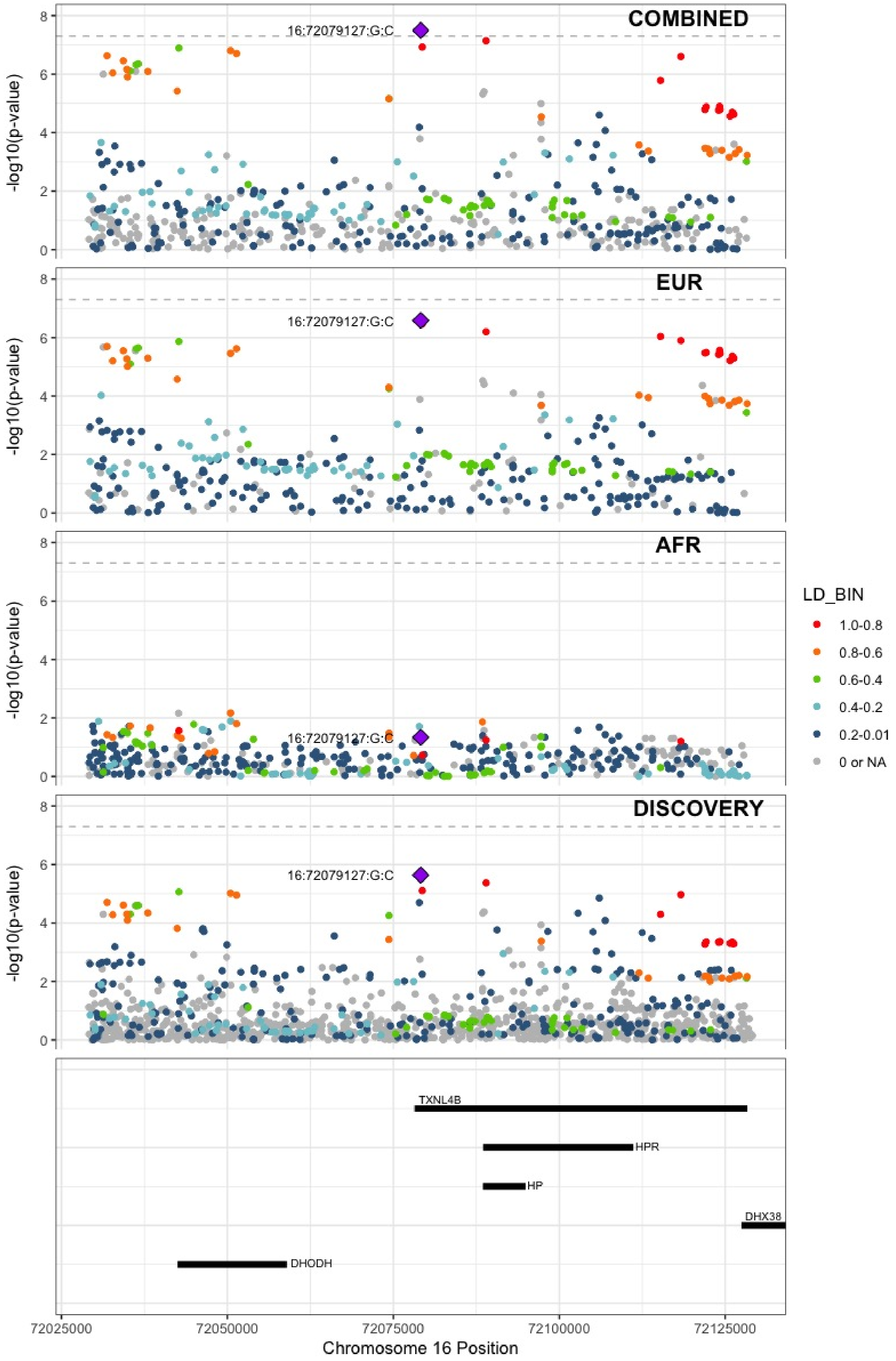

**Supplementary Figure 6.85** – rs77246010 (chr16:75429853) in locus 108 (CFDP1)

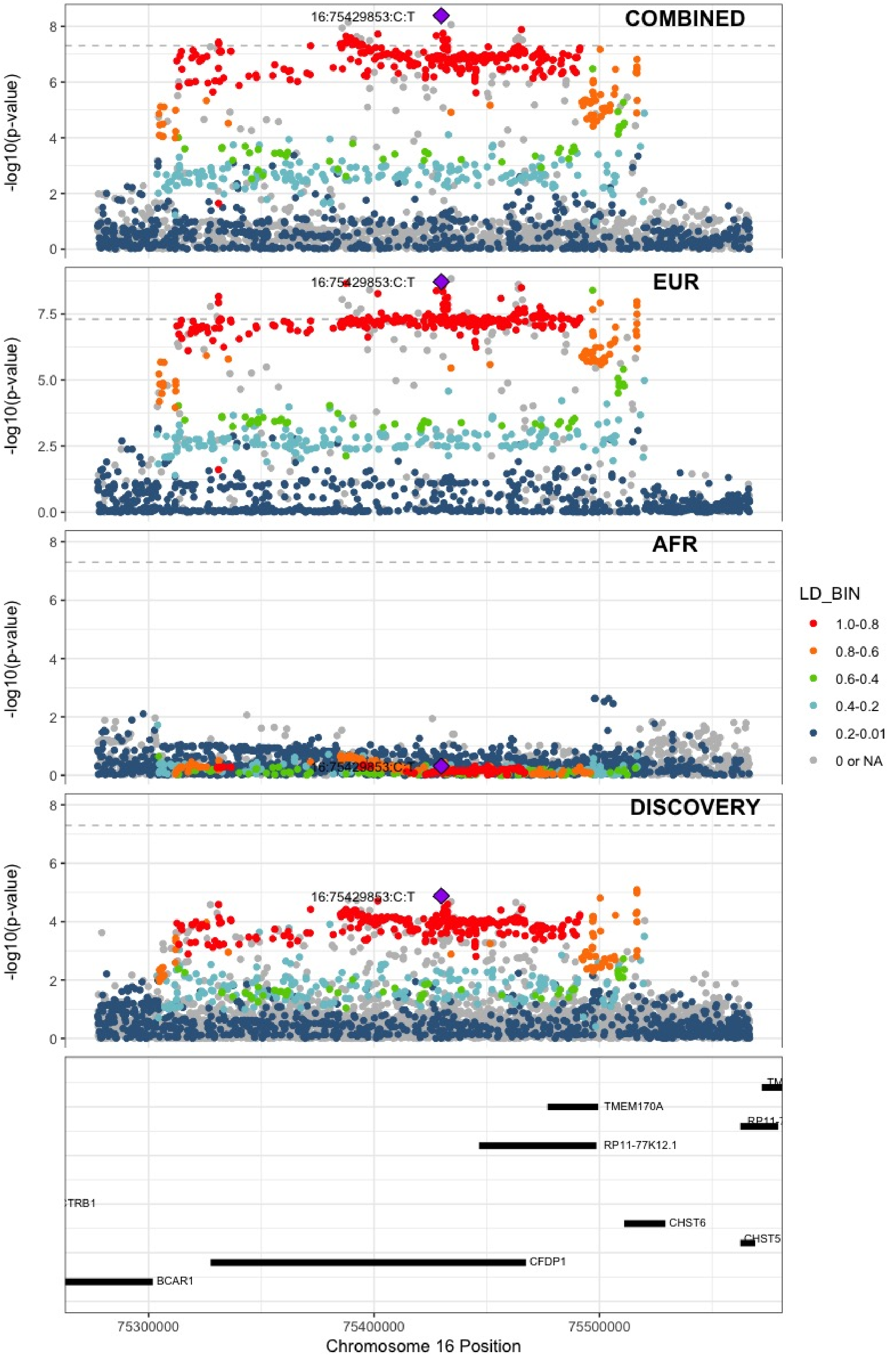

**Supplementary Figure 6.86** – rs12445050 (chr16:81870969) in locus 109 (PLCG2)

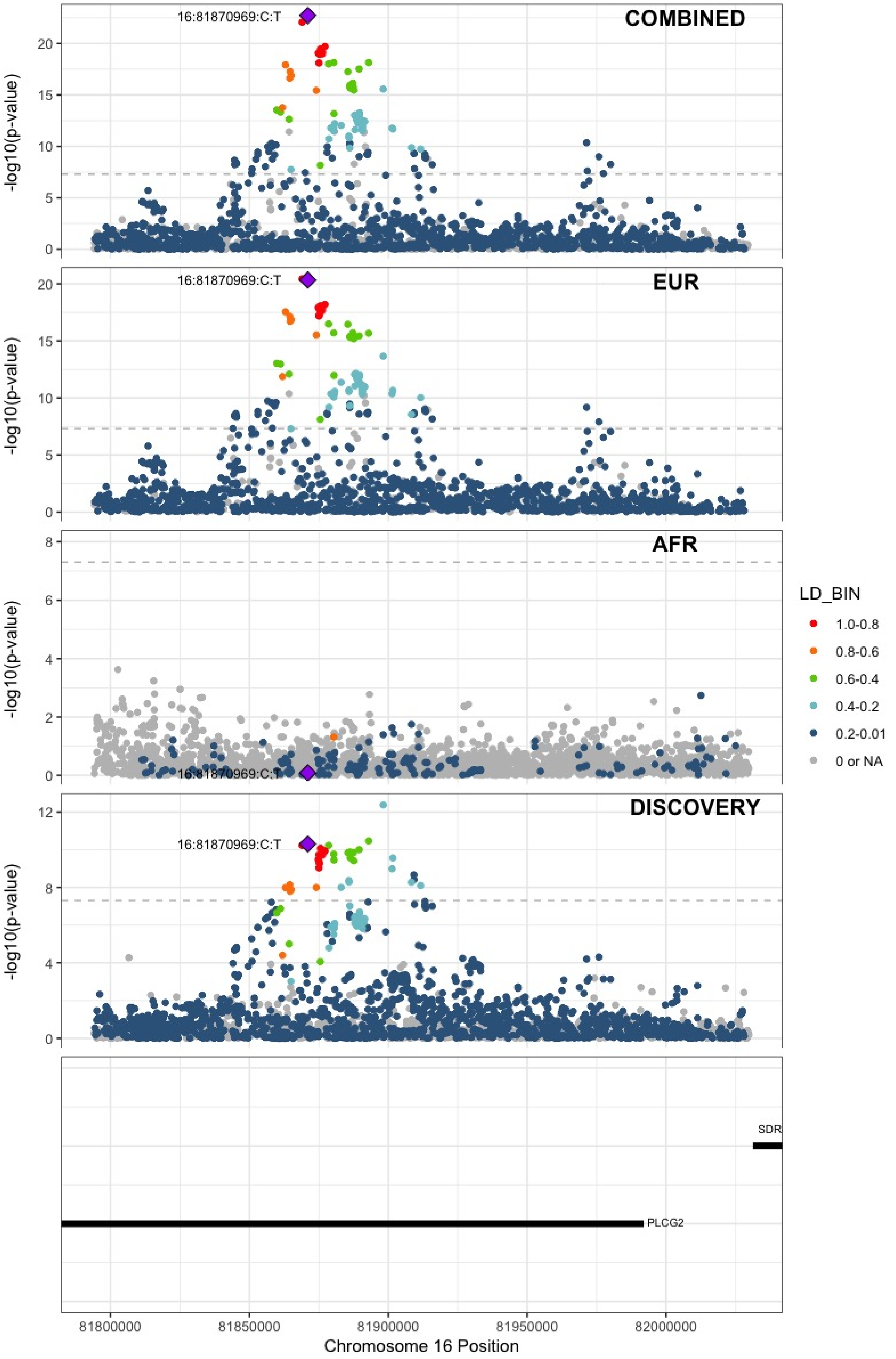

**Supplementary Figure 6.87** – rs8049403 (chr16:85778651) in locus 110 (C16orf74)

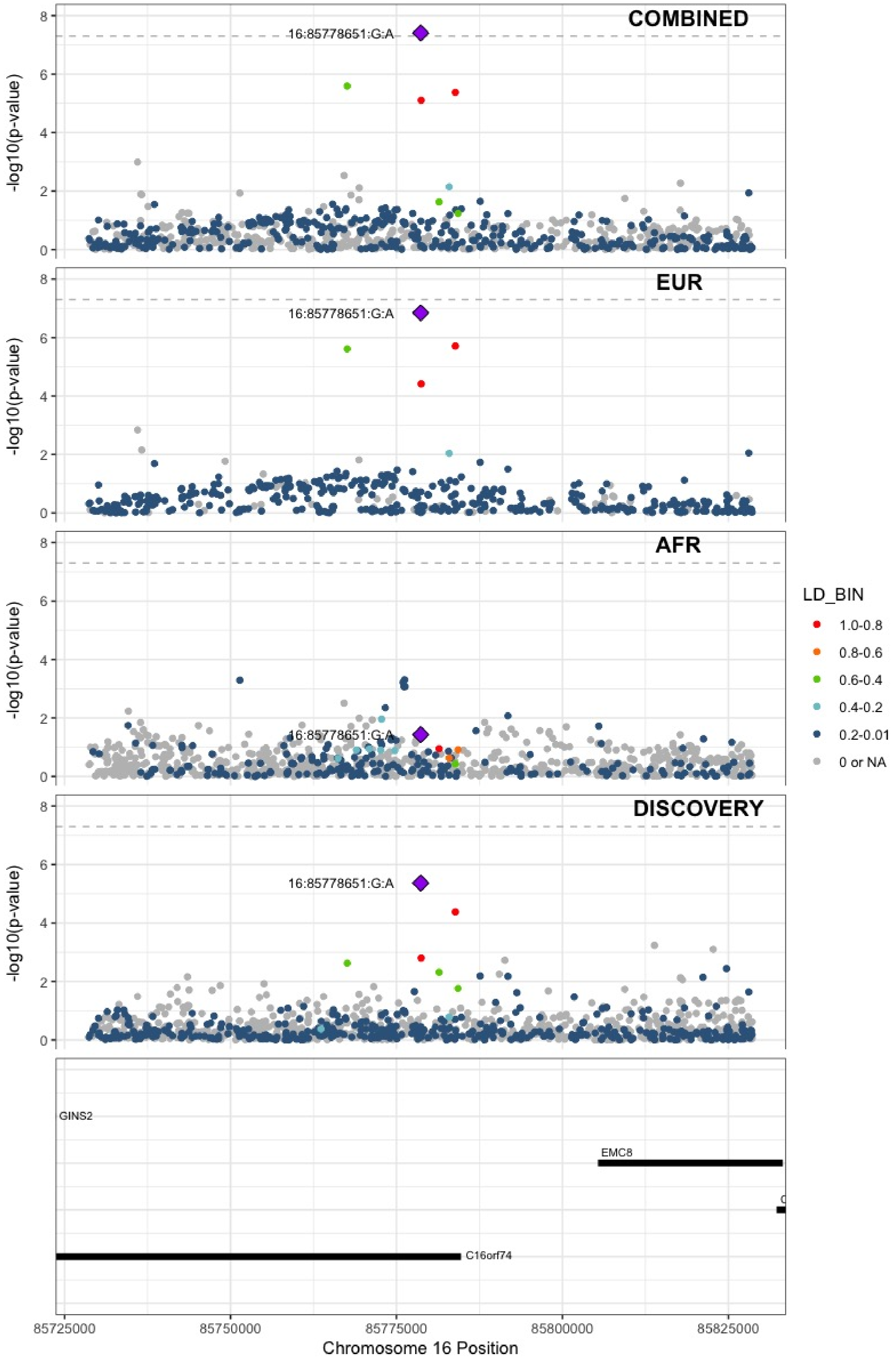

**Supplementary Figure 6.88** – rs28634651 (chr16:88553198) in locus 111 (ZFPM1)

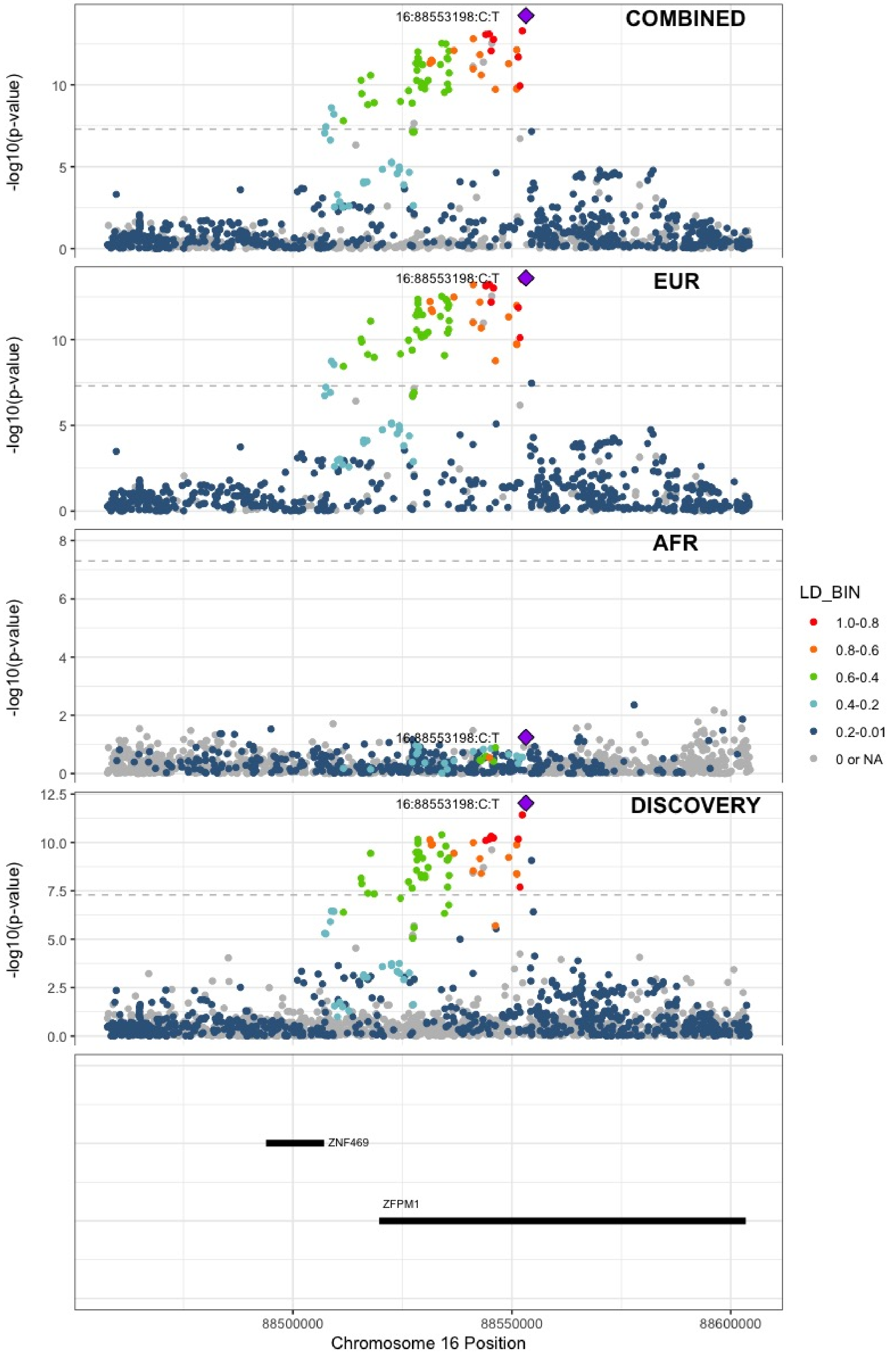

**Supplementary Figure 6.89** – rs6503222 (chr17:1977862) in locus 112 (SMG6)

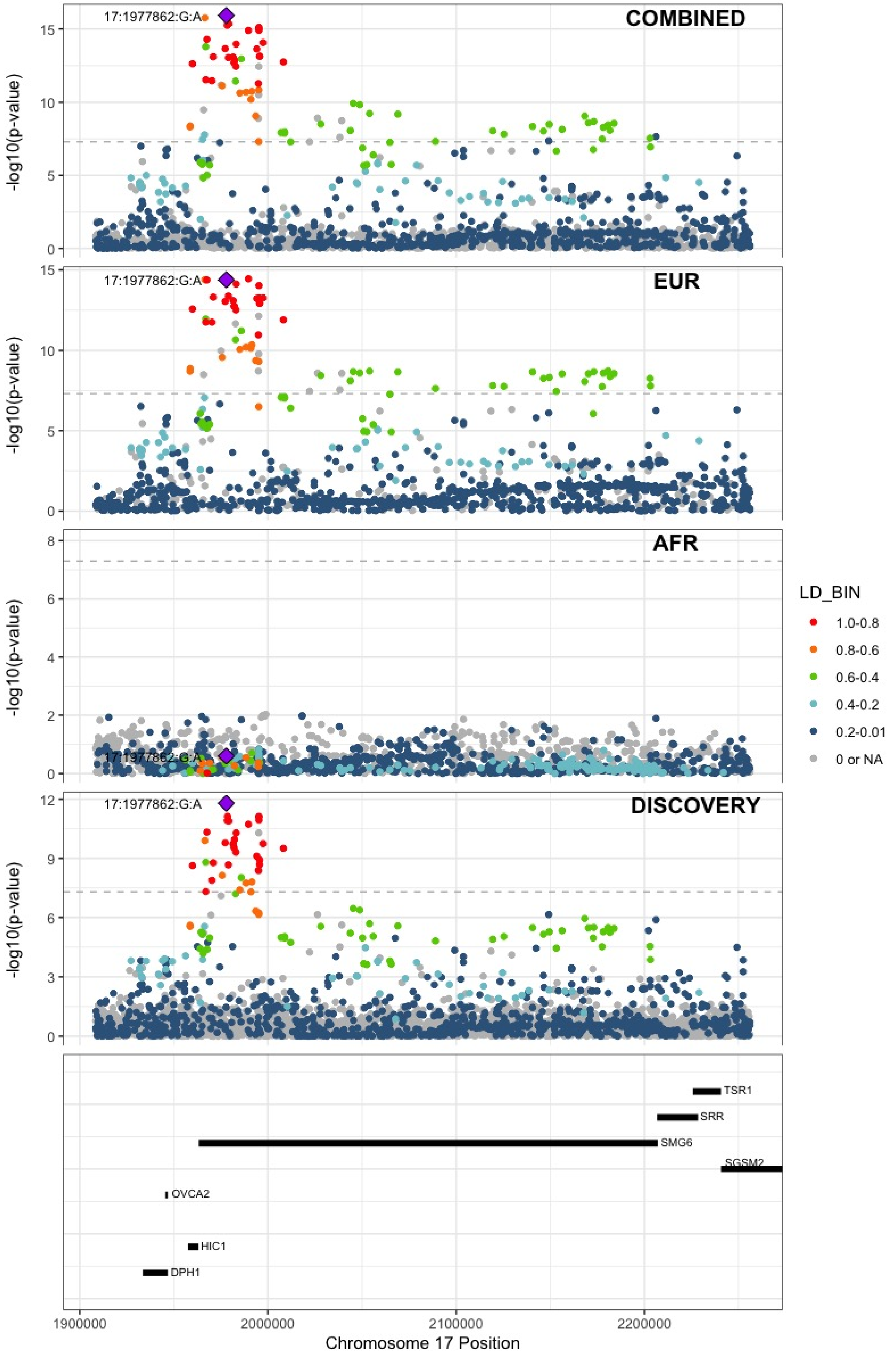

**Supplementary Figure 6.90** – rs434473 (chr17:6904934) in locus 113 (ALOX12)

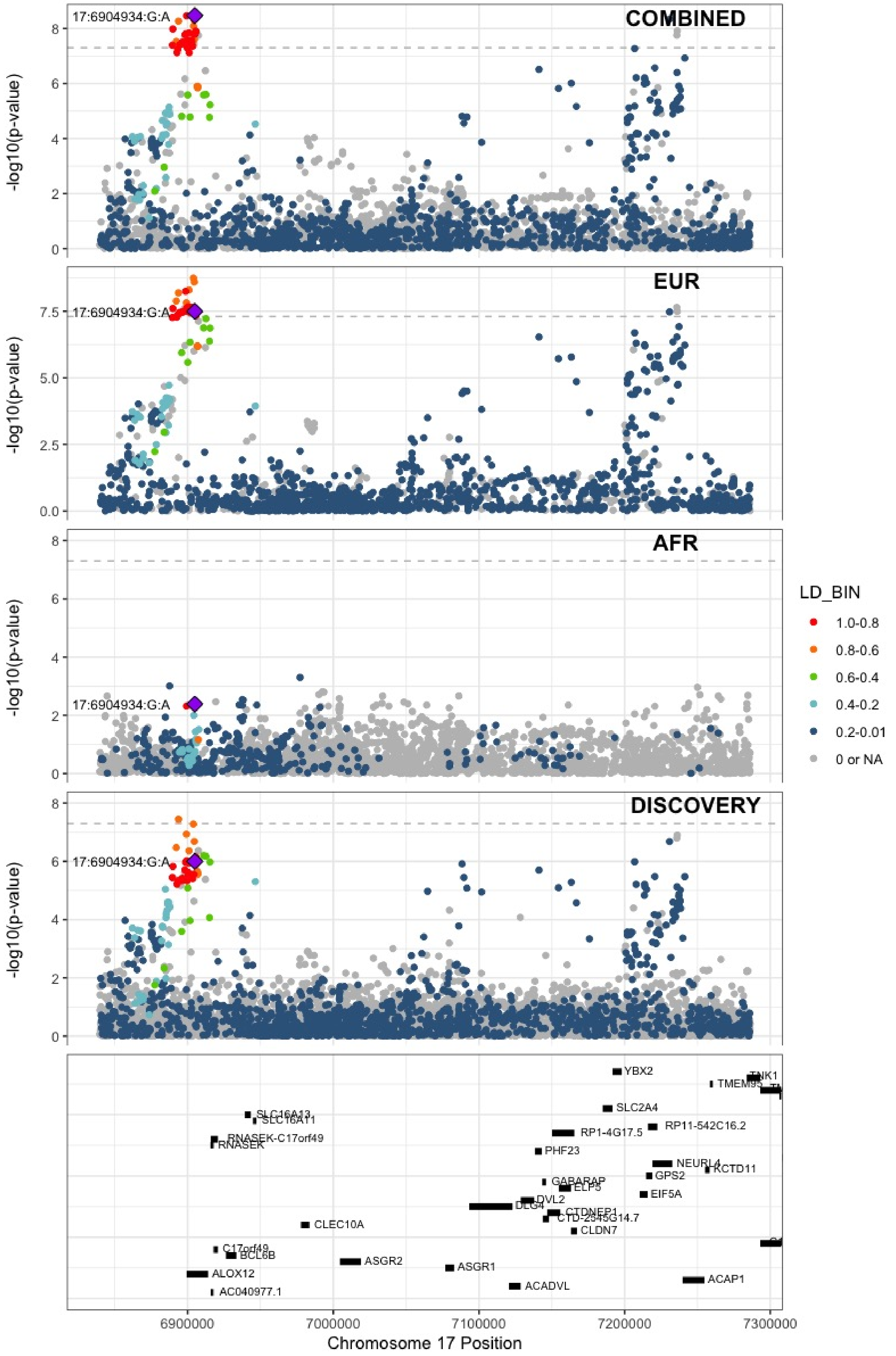

**Supplementary Figure 6.91** – rs71138827 (chr17:27833678) in locus 114 (TAOK1)

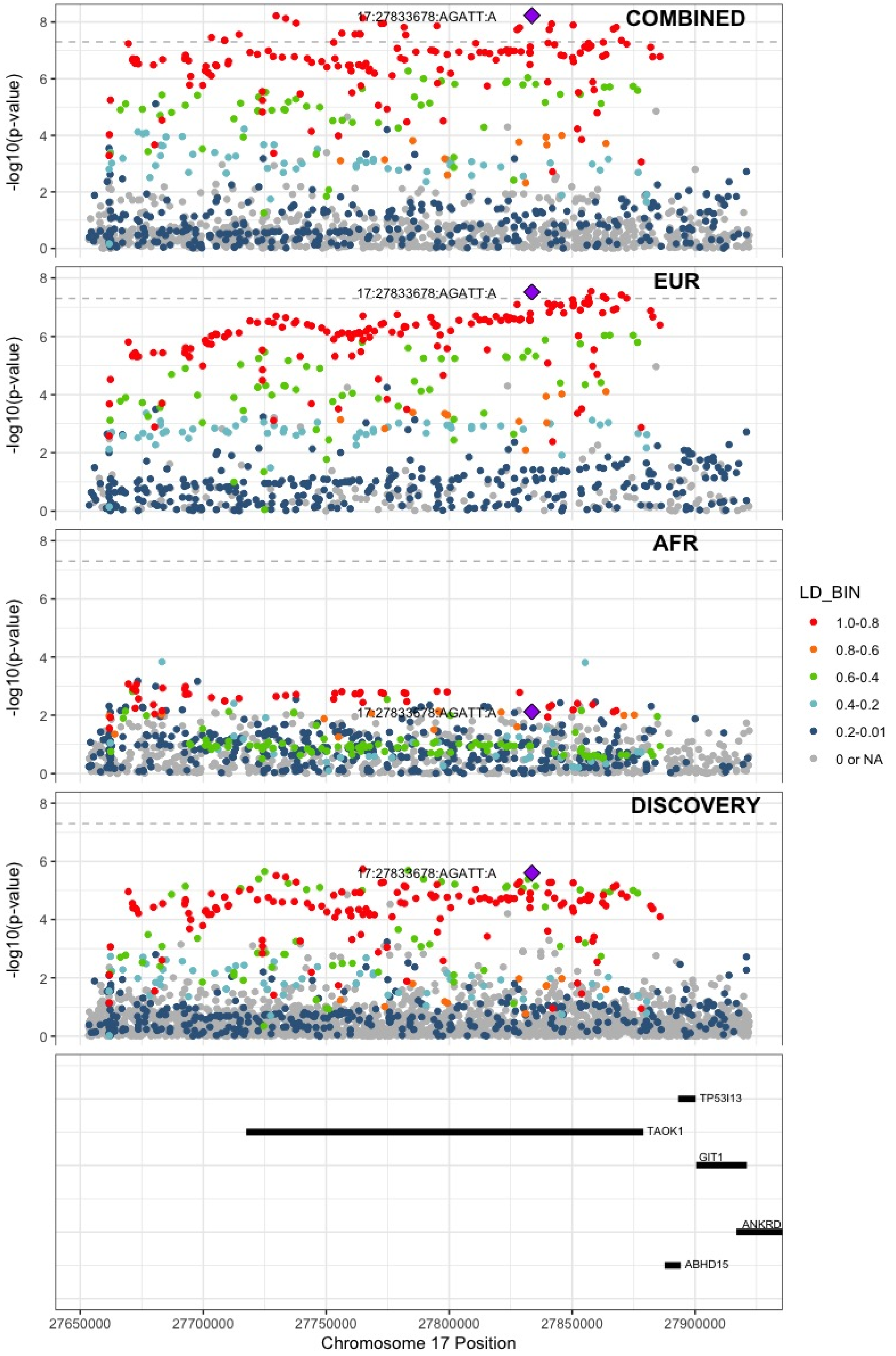

**Supplementary Figure 6.92** – rs62054822 (chr17:43927708) in locus 115 (MAPT-AS1)

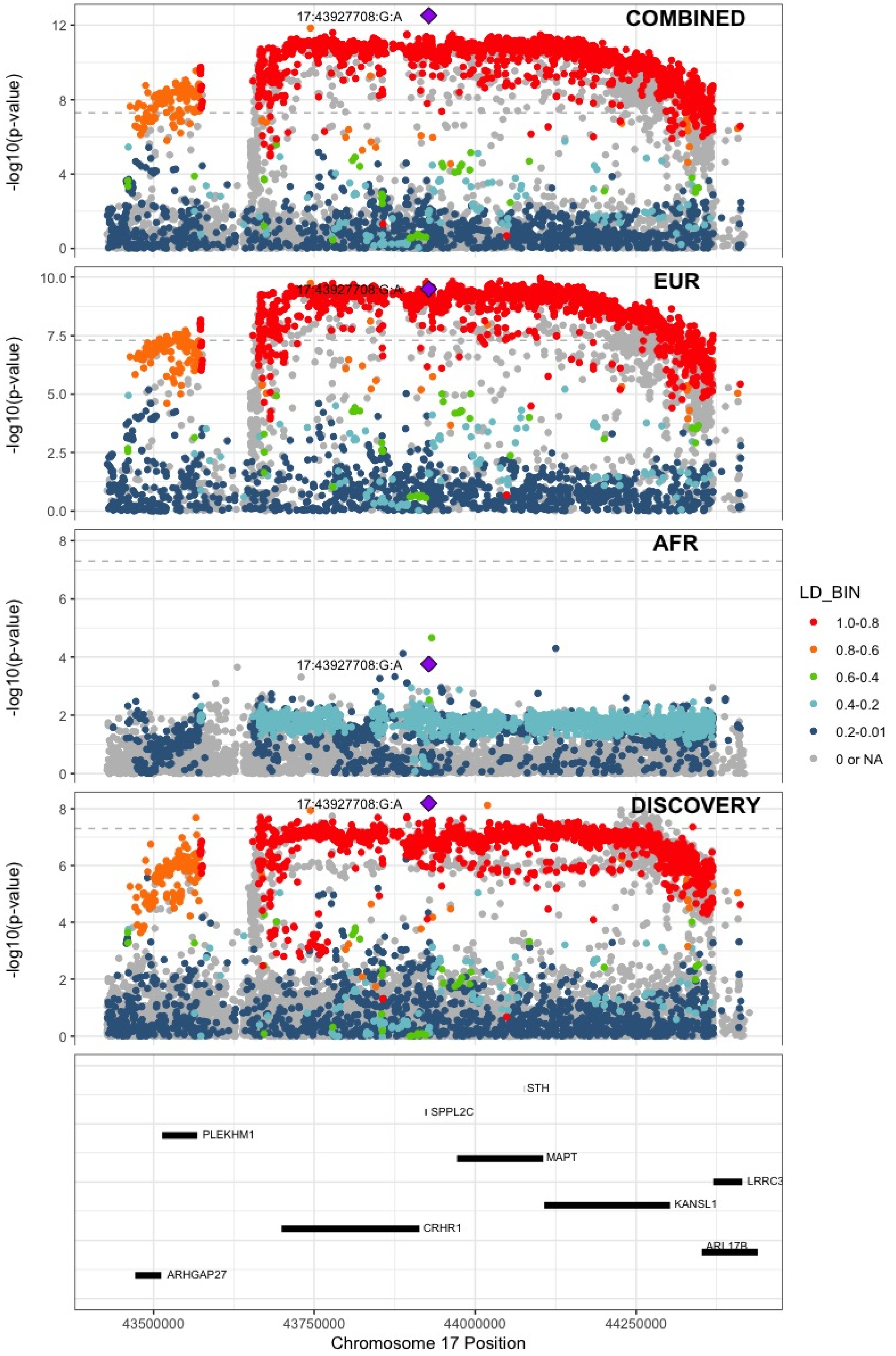

**Supplementary Figure 6.93** – rs1801690 (chr17:64208285) in locus 116 (APOH)

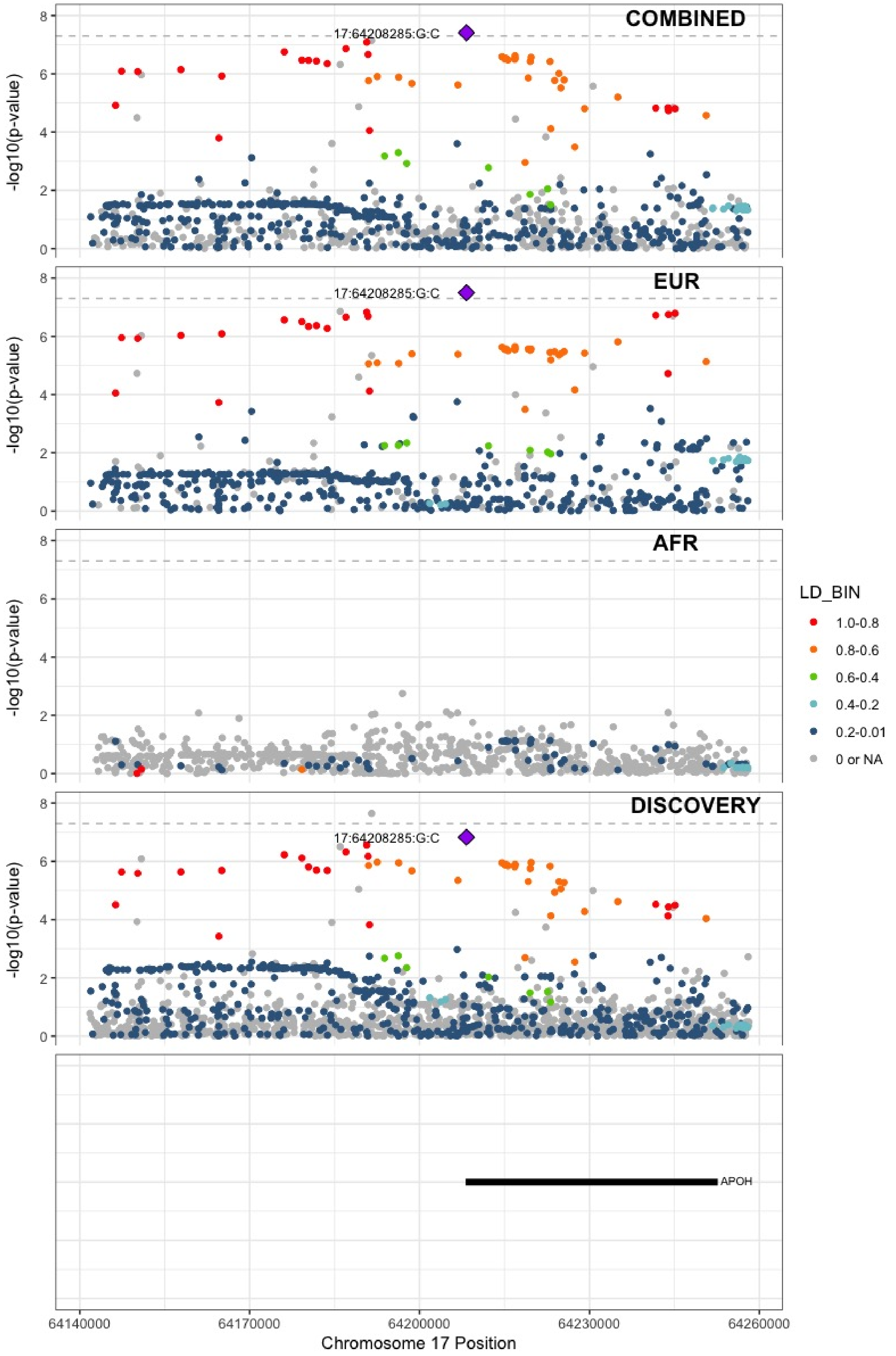

**Supplementary Figure 6.94** – rs573431210 (chr17:67276383) in locus 117 (ABCA5)

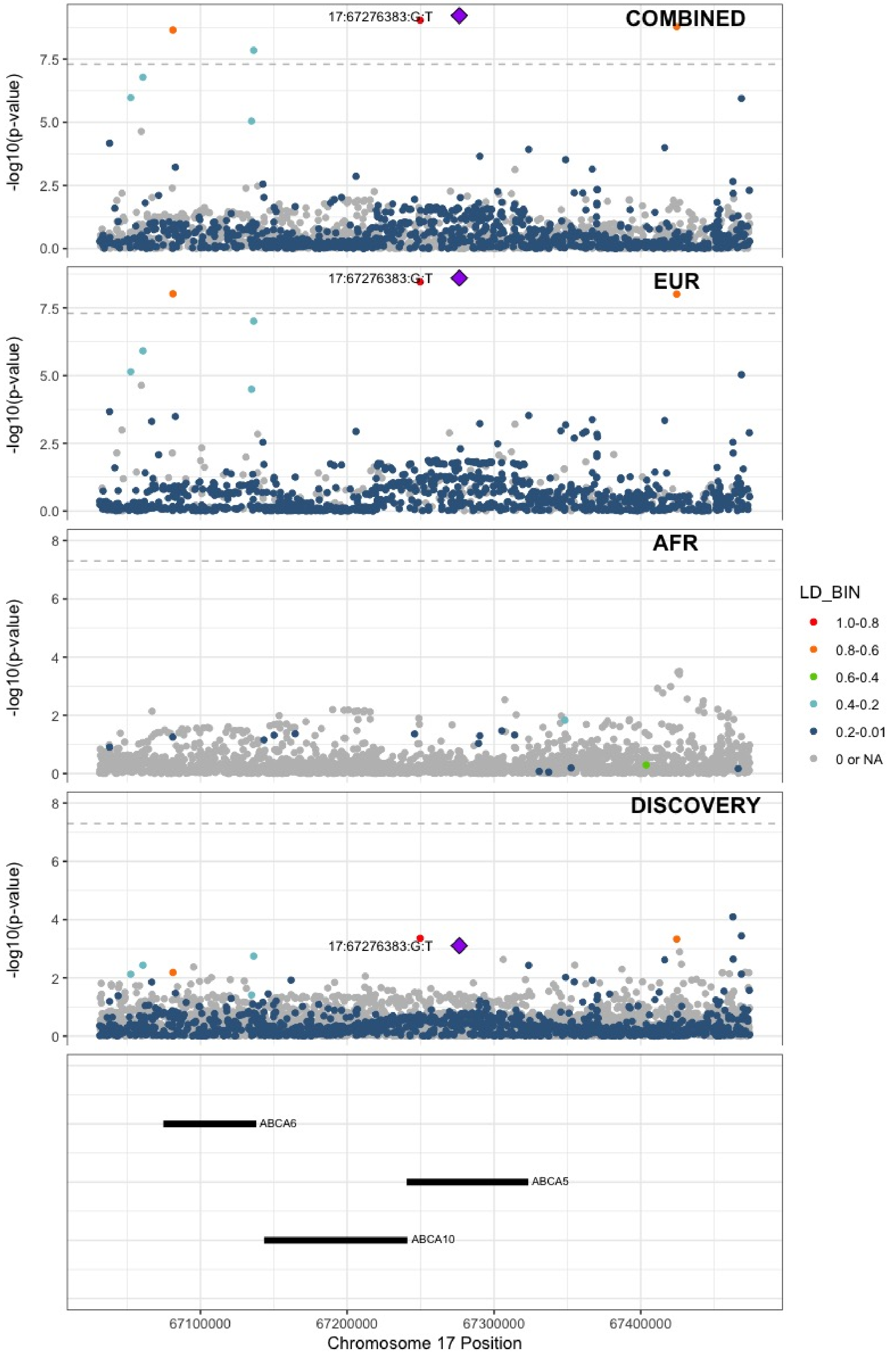

**Supplementary Figure 6.95** – rs59277920 (chr19:6077231) in locus 118 (RFX2)

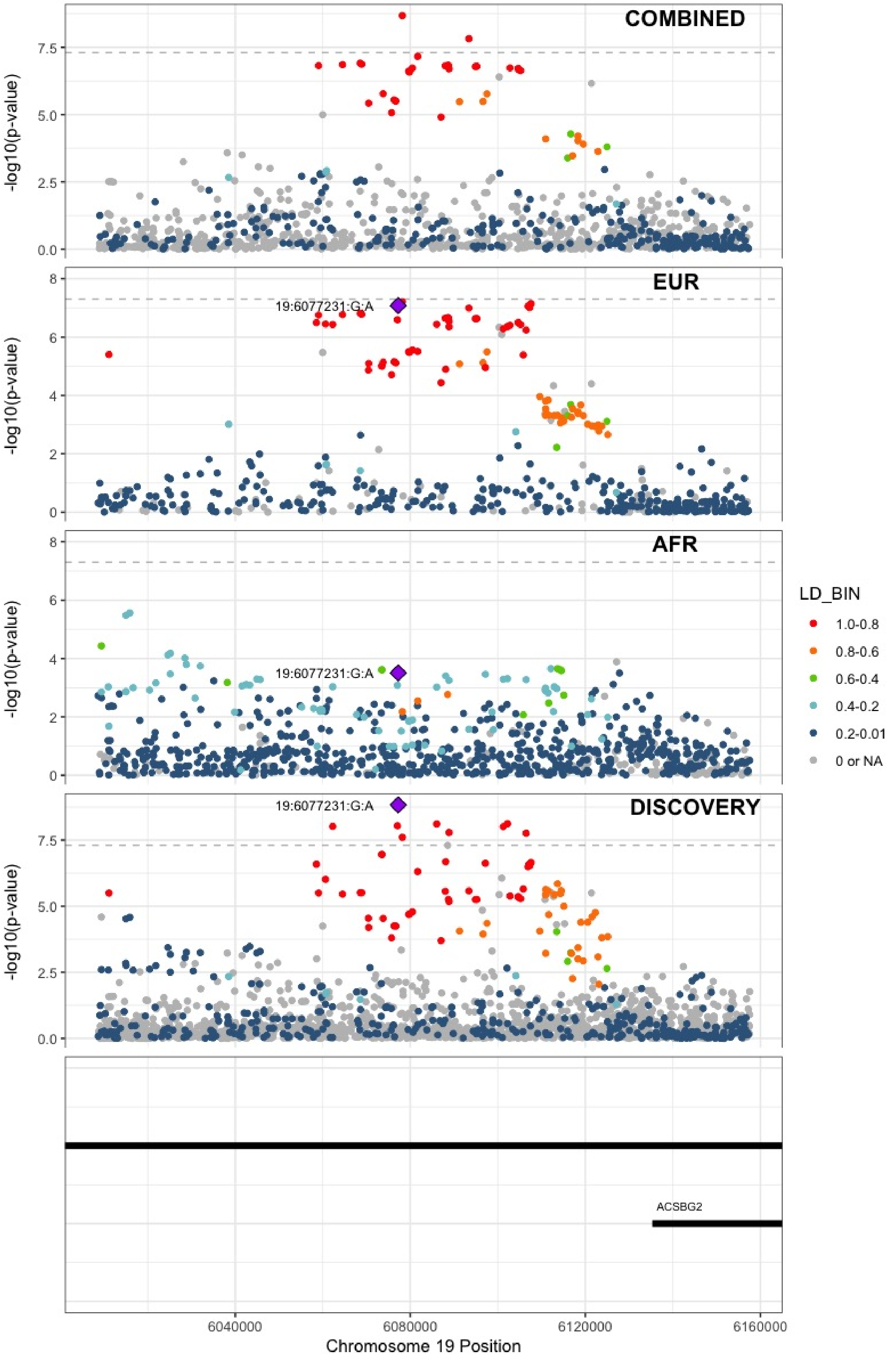

**Supplementary Figure 6.96** – rs8110055 (chr19:10739143) in locus 119 (SLC44A2)

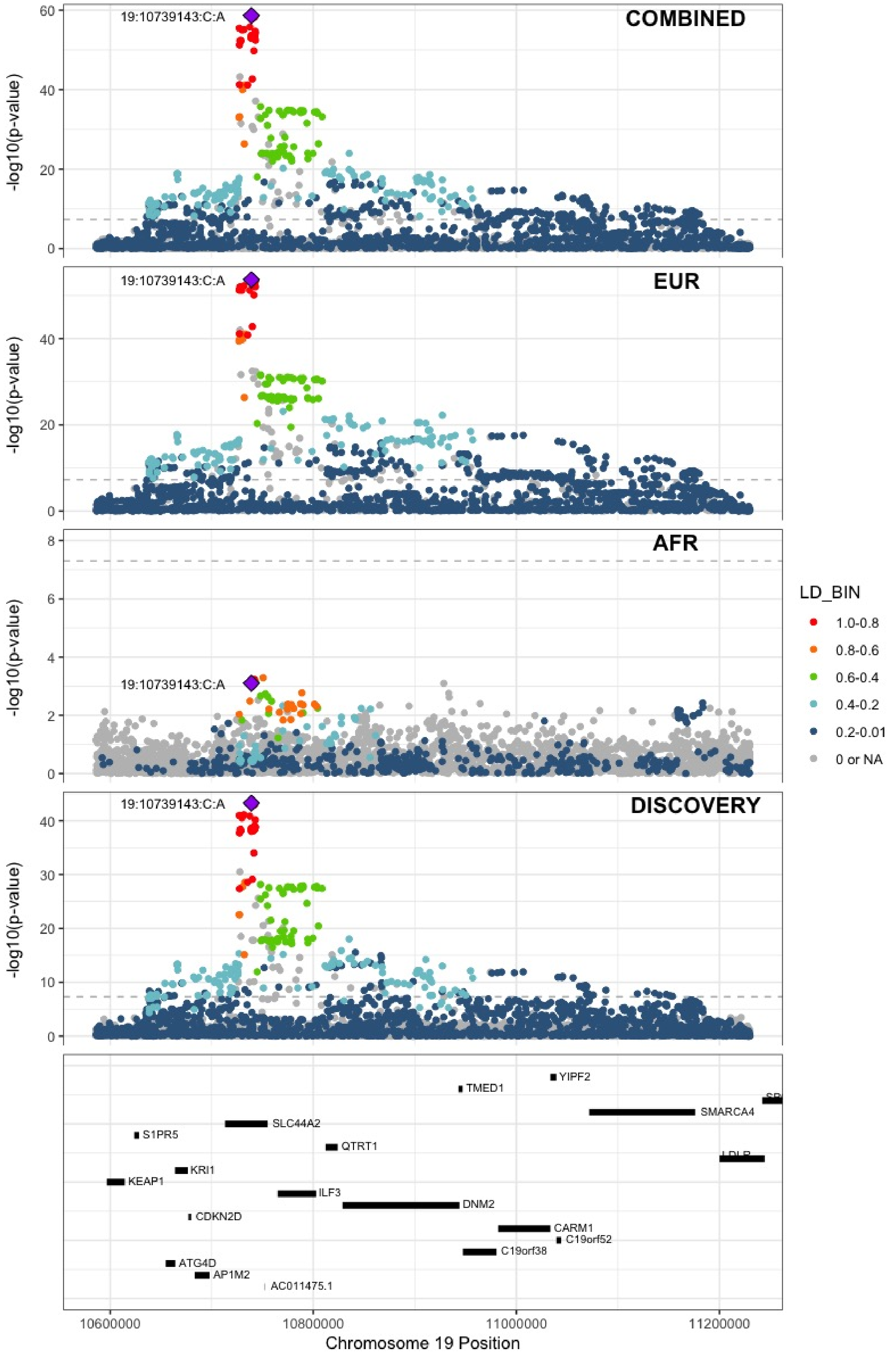

**Supplementary Figure 6.97** – rs889139 (chr19:33889369) in locus 120 (PEPD)

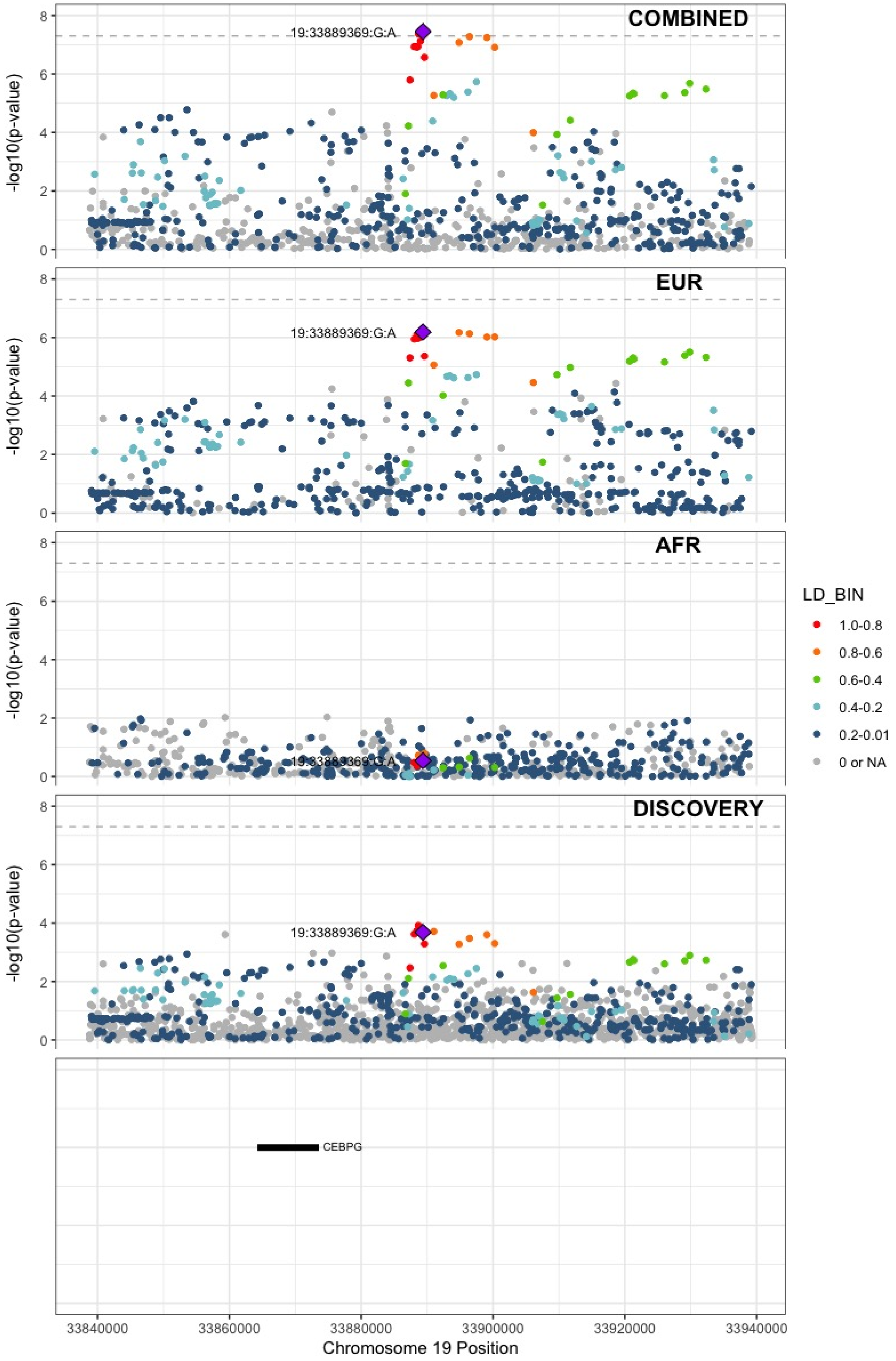

**Supplementary Figure 6.98** – rs2545774 (chr19:41287674) in locus 121 (RAB4B)

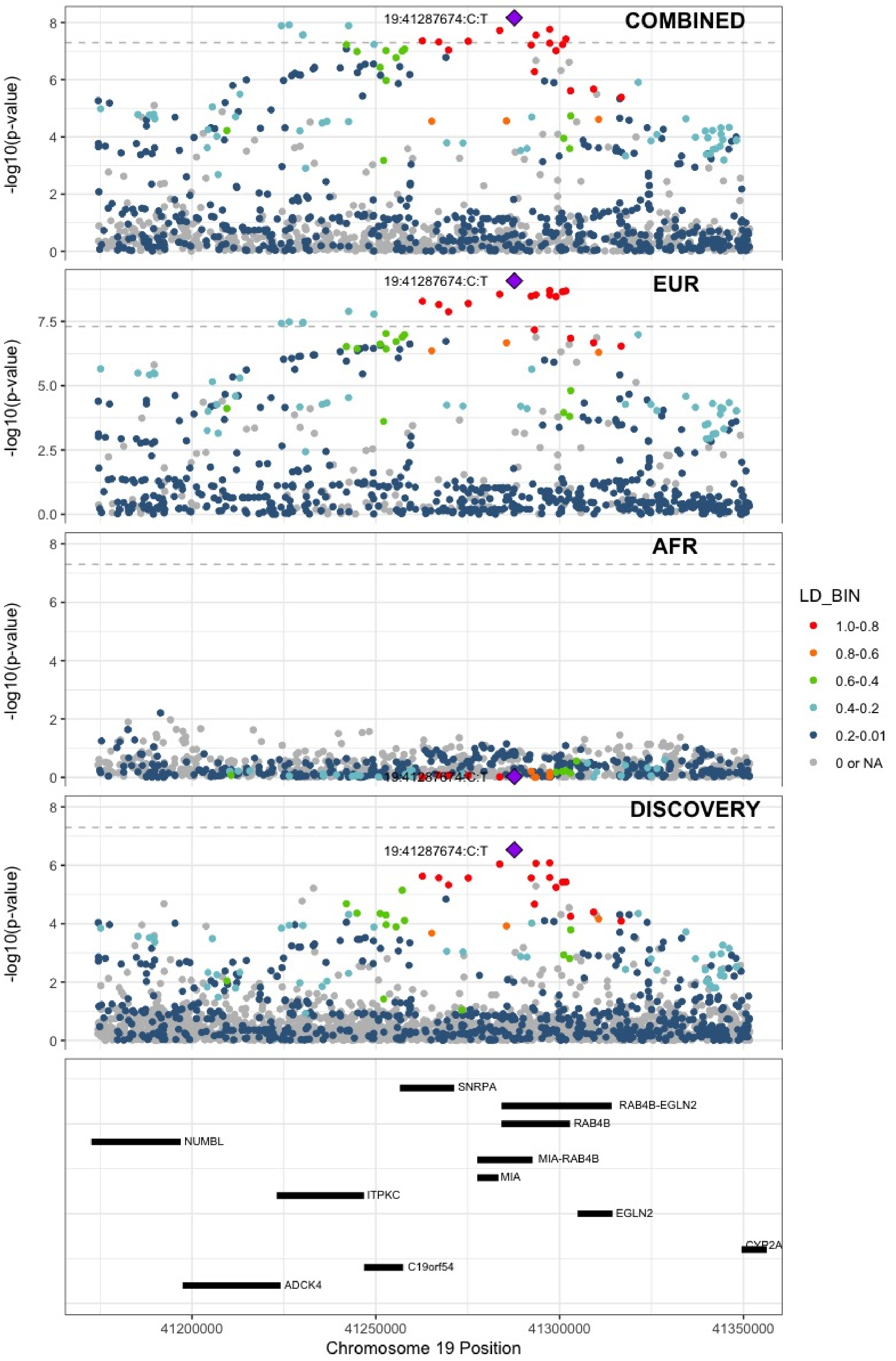

**Supplementary Figure 6.99** – rs8108474 (chr19:46301479) in locus 122 (RSPH6A)

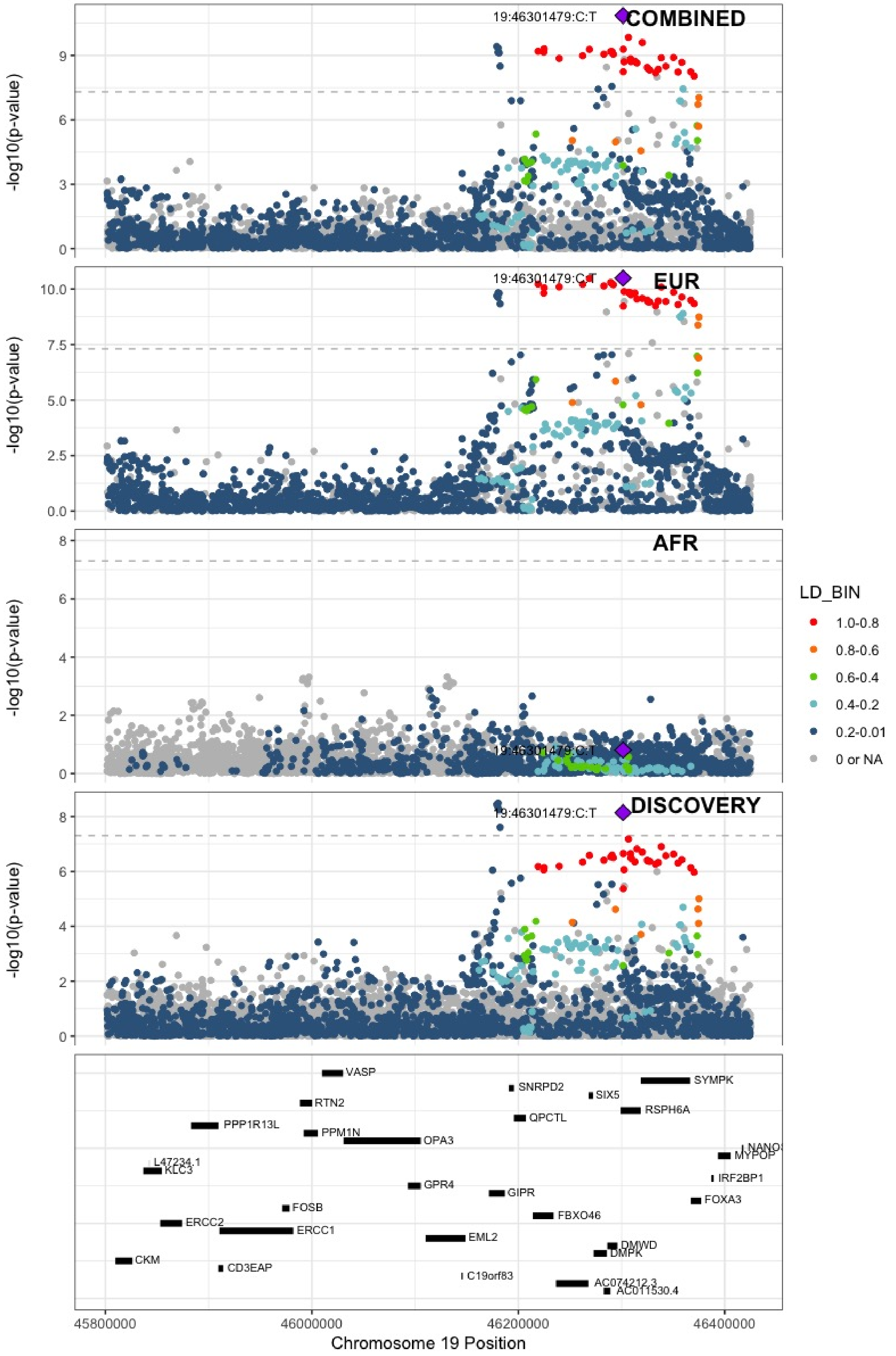

**Supplementary Figure 6.100** – rs2638282 (chr19:49213833) in locus 123 (FUT2;MAMSTR)

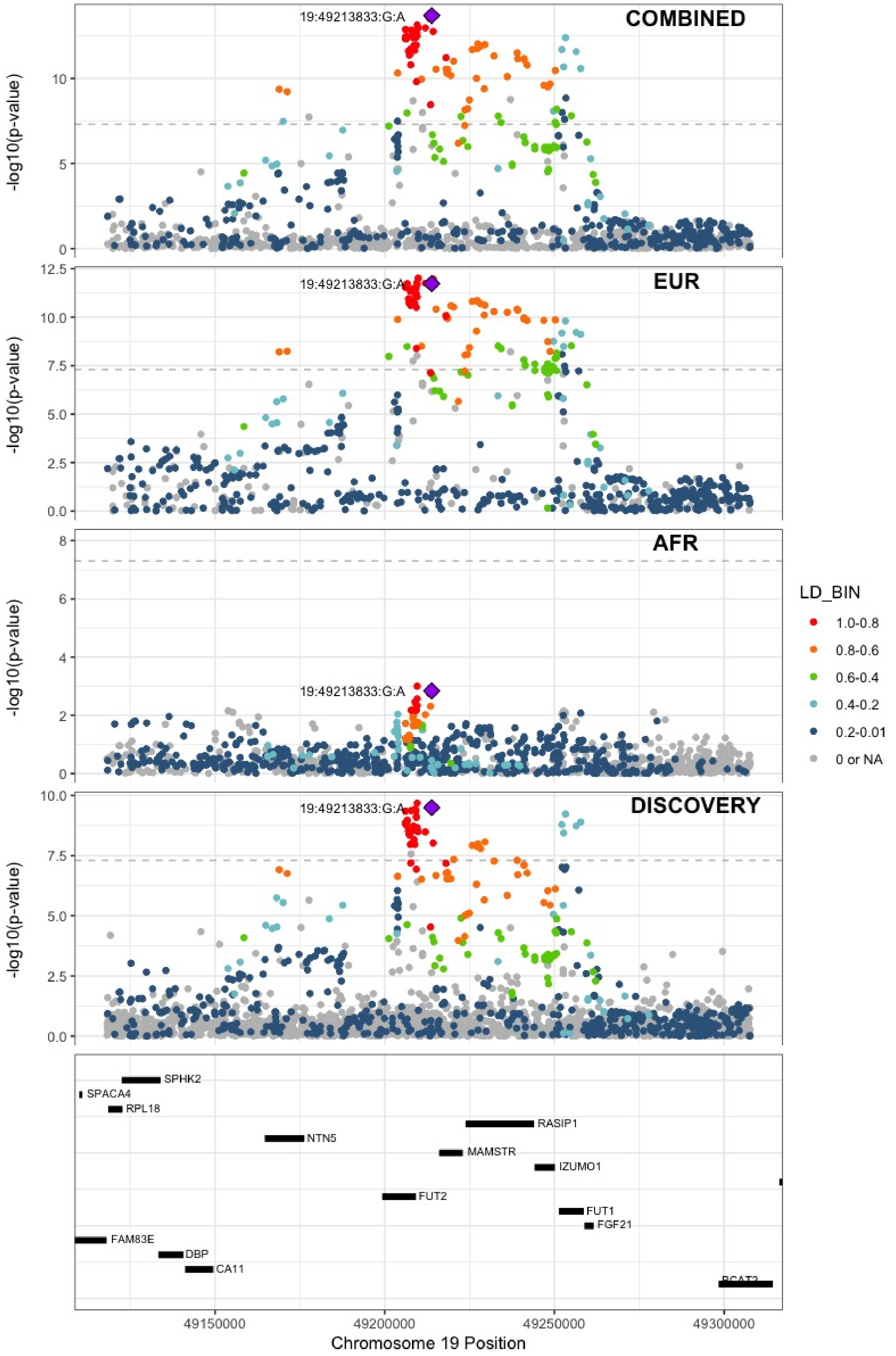

**Supplementary Figure 6.101** – rs1654425 (chr19:55538980) in locus 124 (GP6)

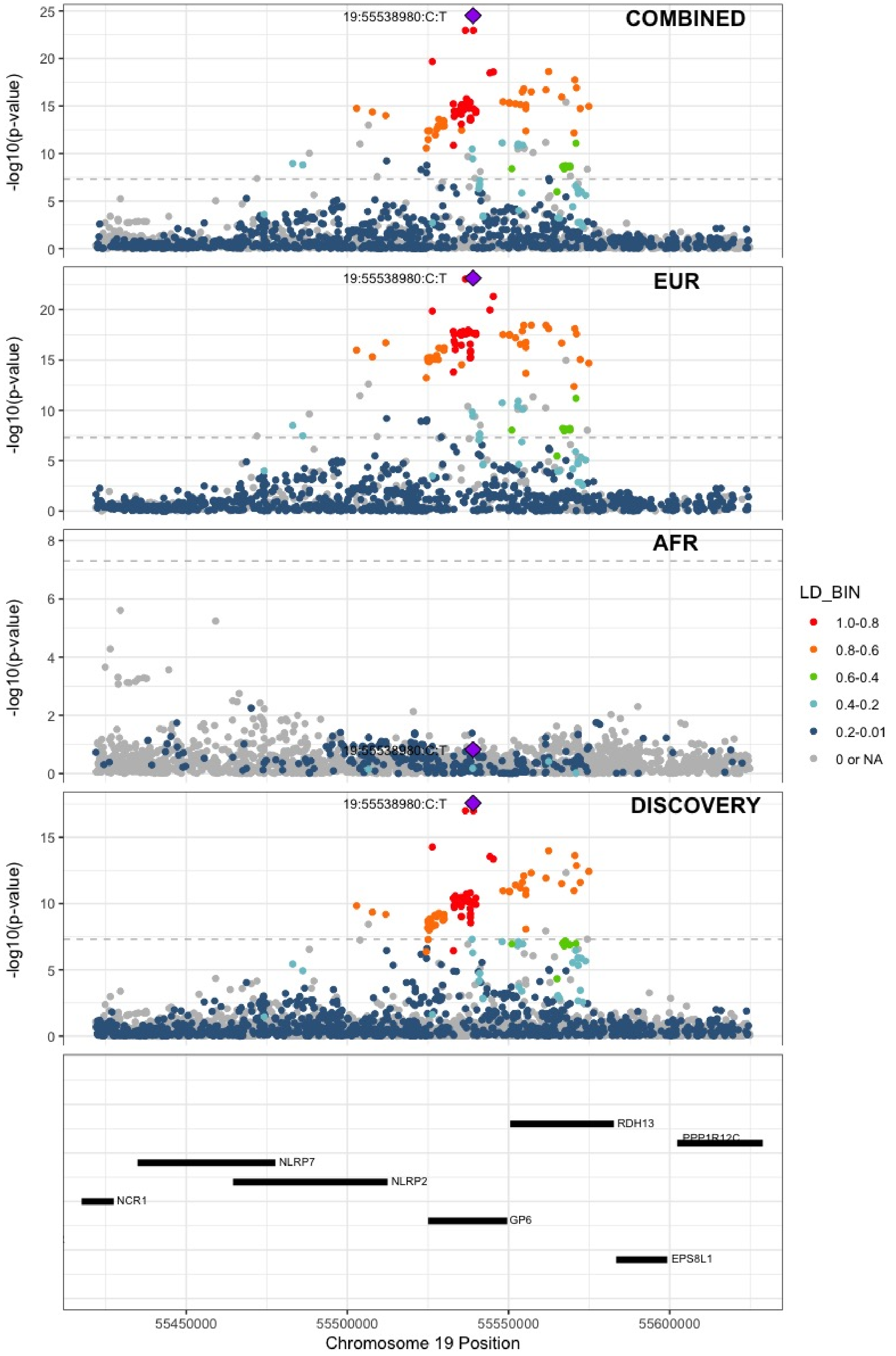

**Supplementary Figure 6.102** – rs2631628 (chr19:56645250) in locus 125 (ZNF787;ZNF444)

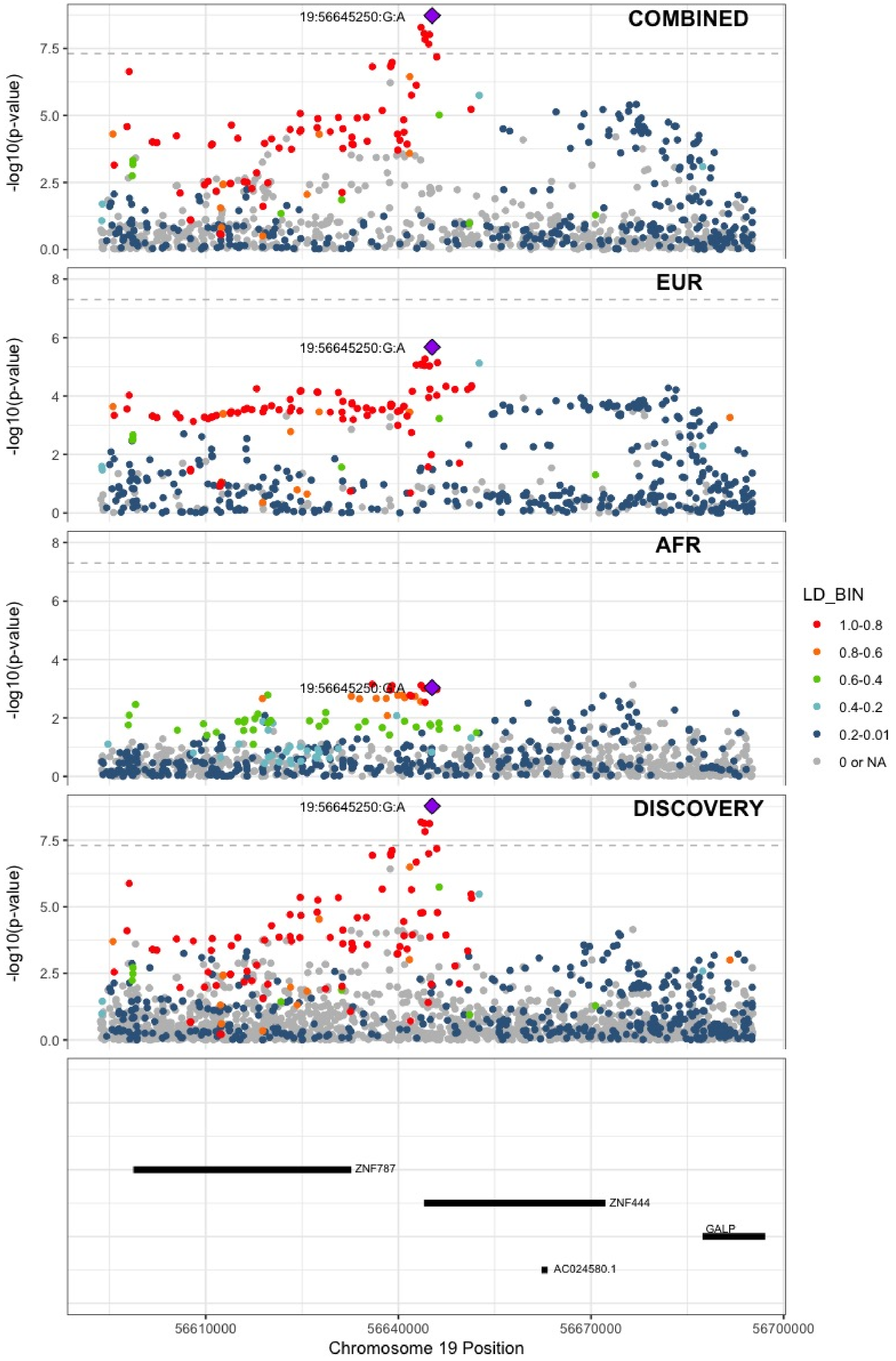

**Supplementary Figure 6.103** – rs117390891 (chr20:23168526) in locus 126 (LINC00656 ;NXT1)

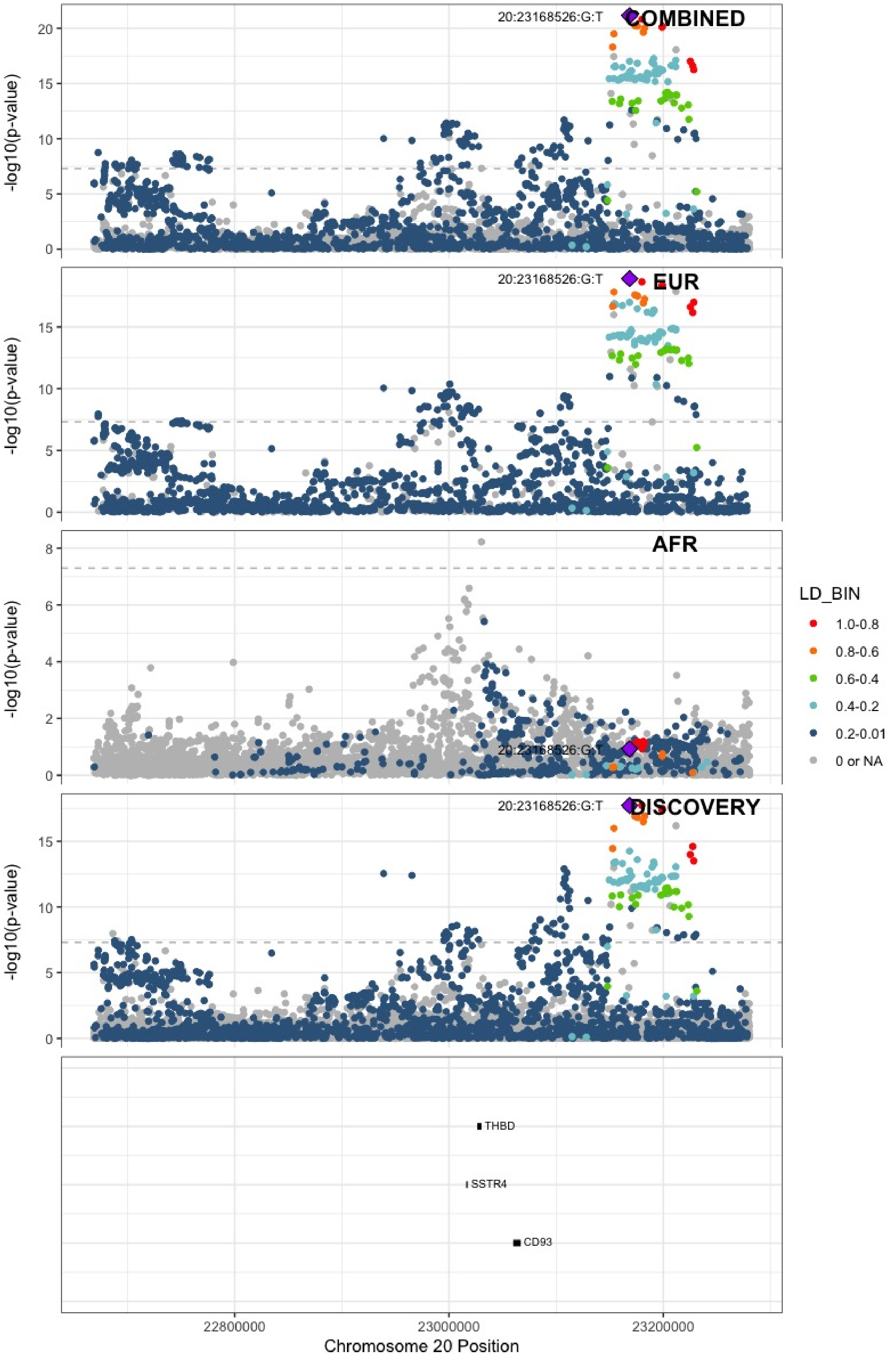

**Supplementary Figure 6.104** – rs6060288 (chr20:33772243) in locus 127 (MMP24-AS1-EDEM2)

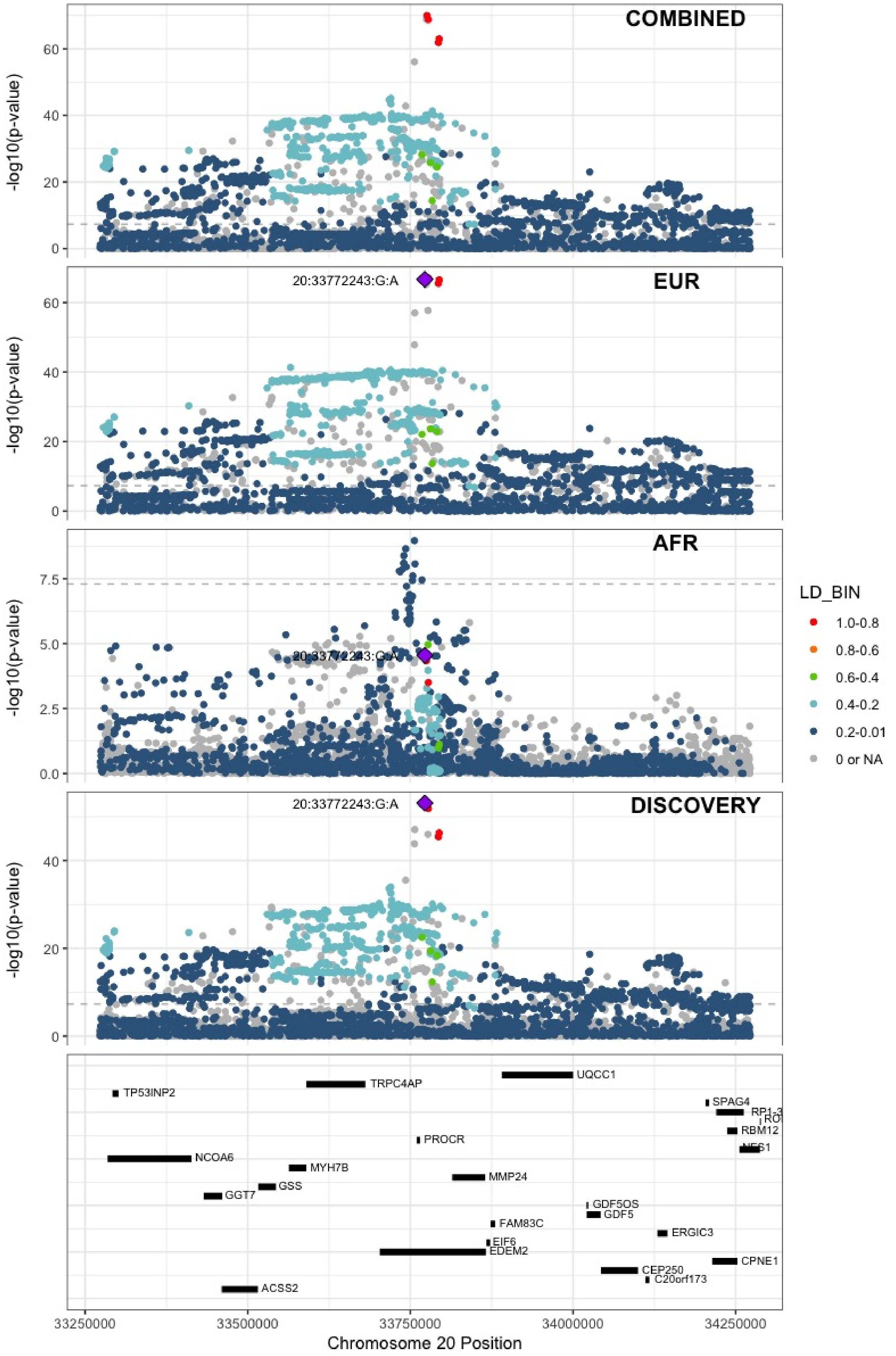

**Supplementary Figure 6.105** – rs3070580 (chr22:33165231) in locus 129 (SYN3)

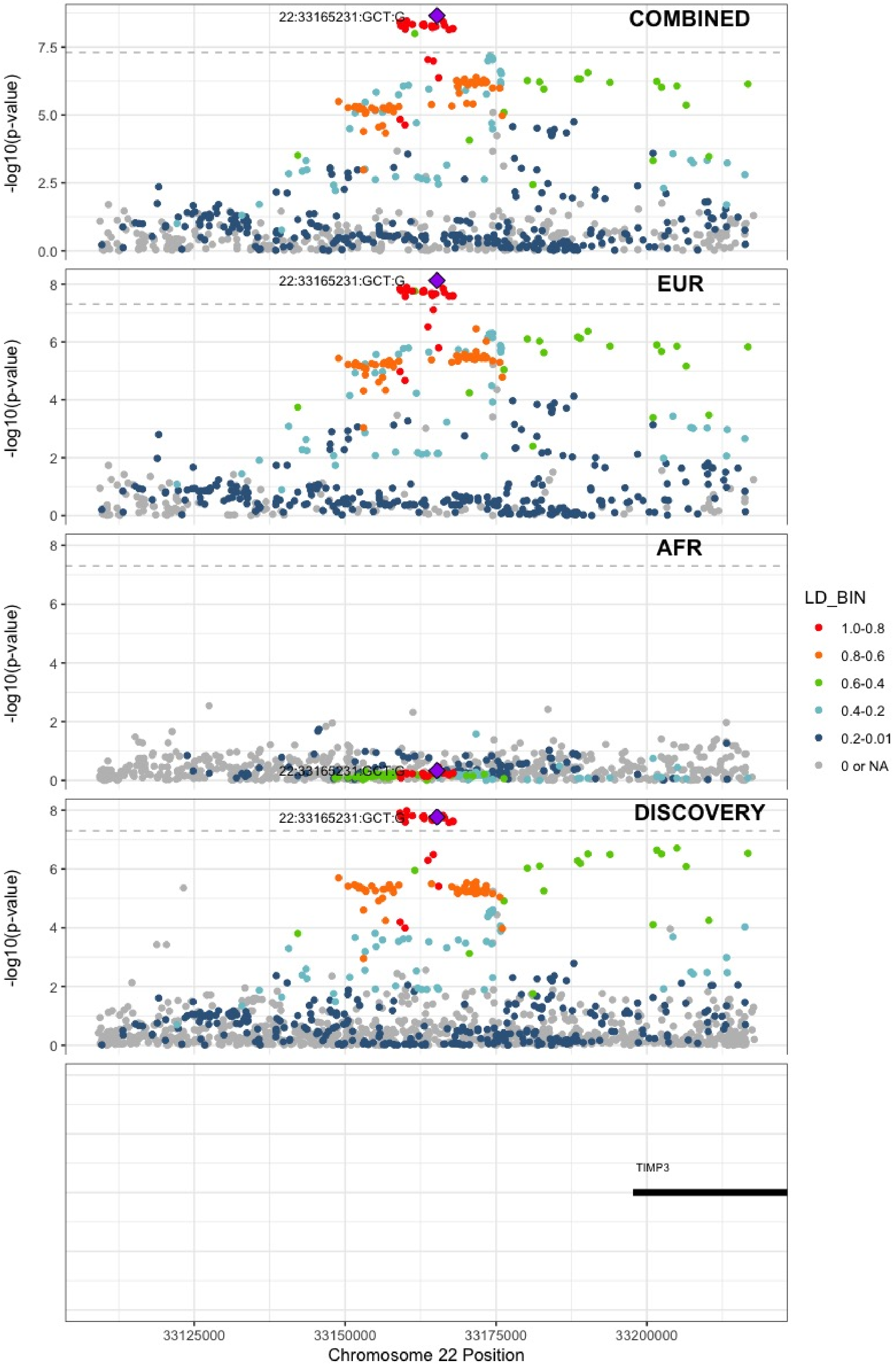

**Supplementary Figure 6.106** – rs9611844 (chr22:43115776) in locus 130 (A4GALT)

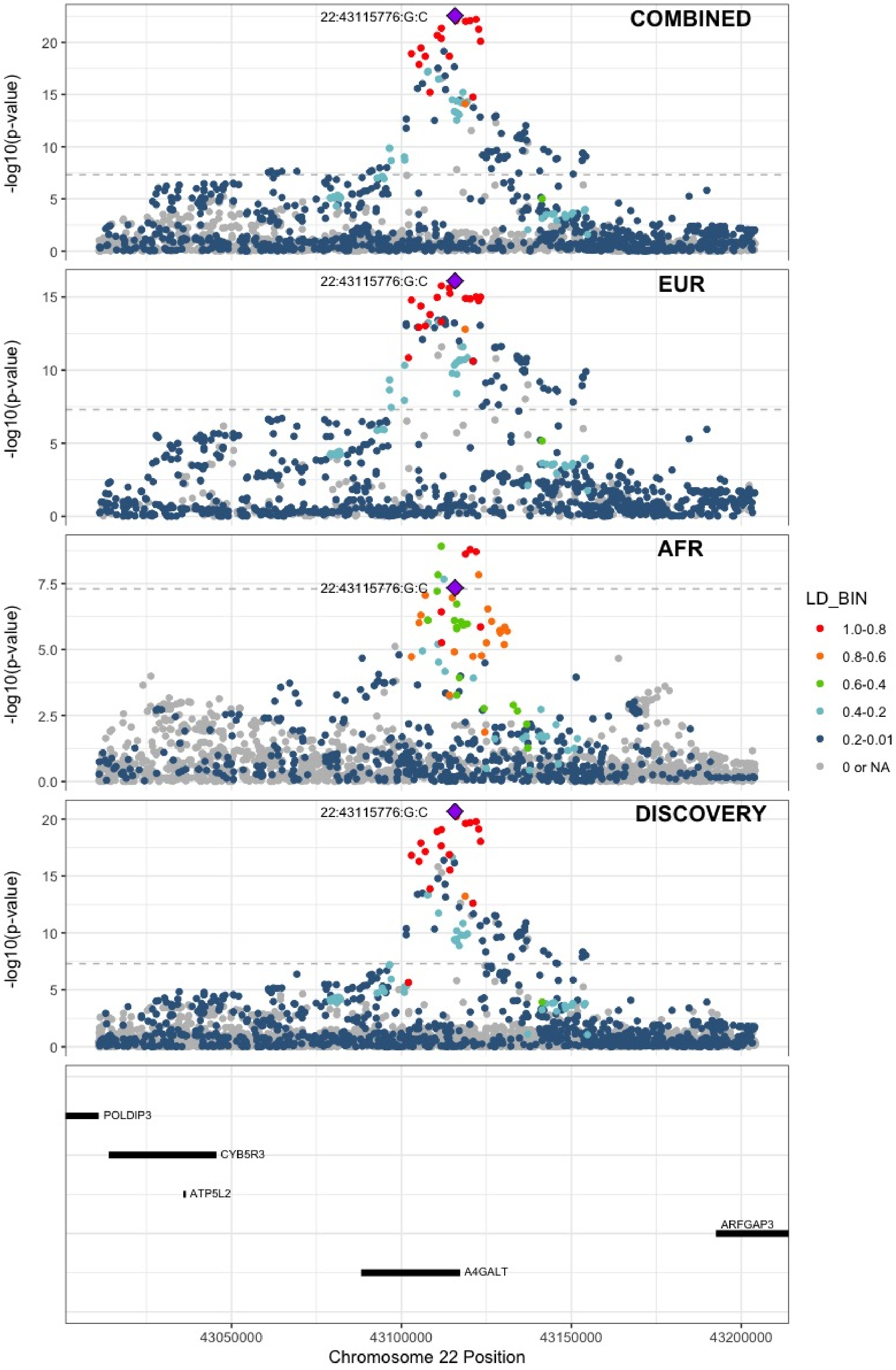

**Supplementary Figure 6.107** – rs12388118 (chr23:11380742) in locus 131 (ARHGAP6)

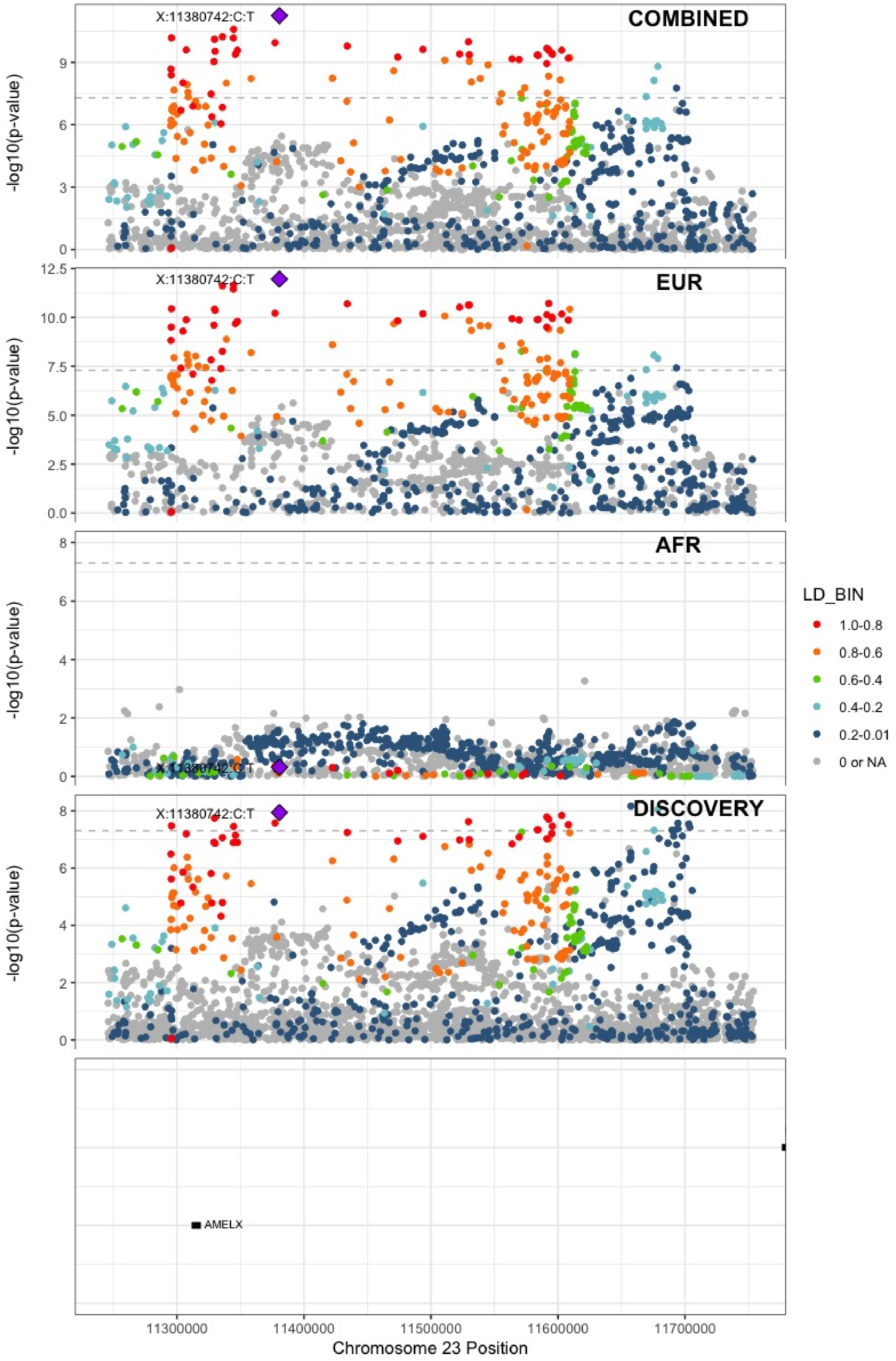

**Supplementary Figure 6.108** – rs6632109 (chr23:34894638) in locus 132 (TMEM47;FAM47B)

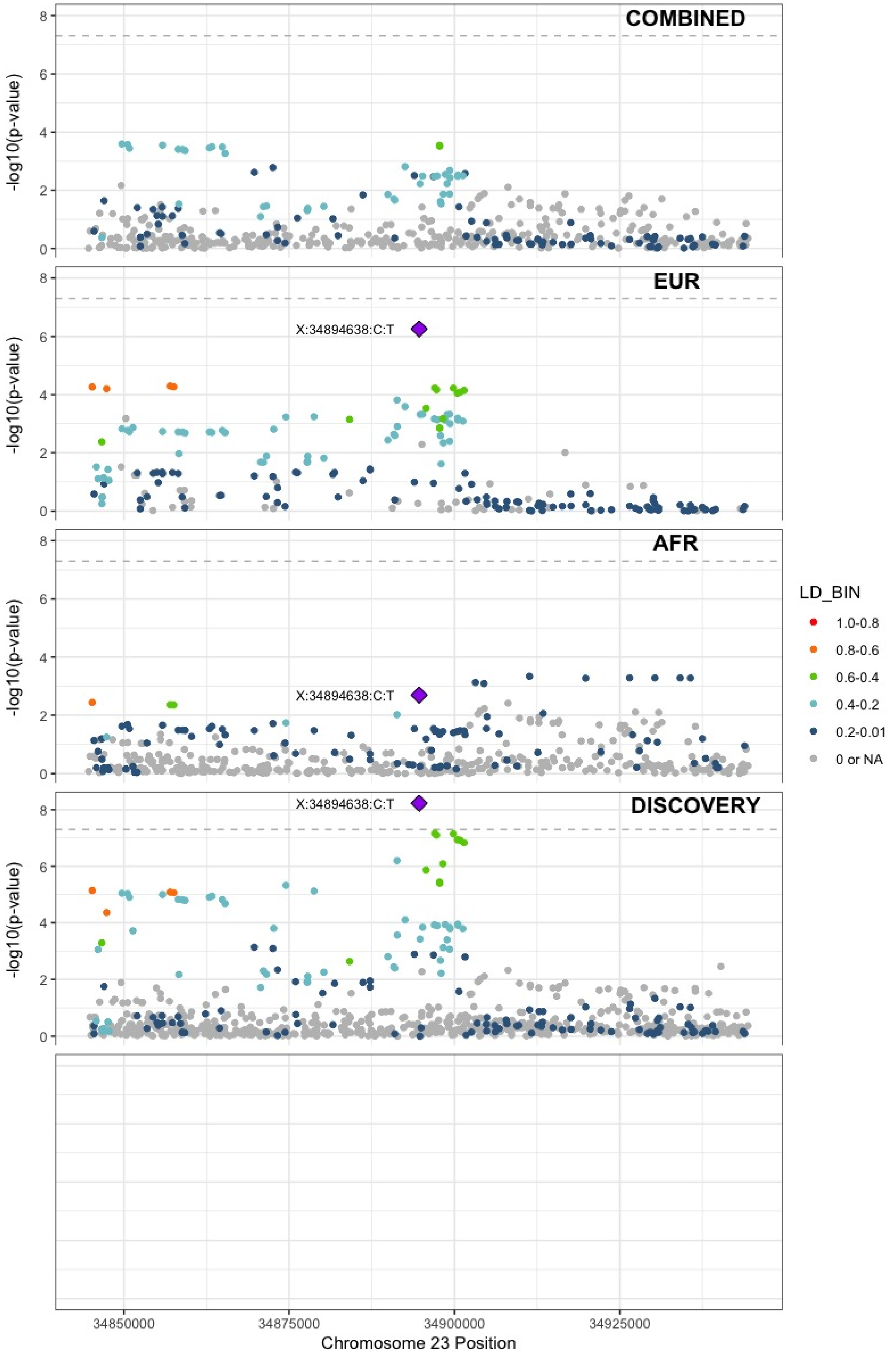

**Supplementary Figure 6.109** – rs3002416 (chr23:39710195) in locus 133 (MIR1587;BCOR)

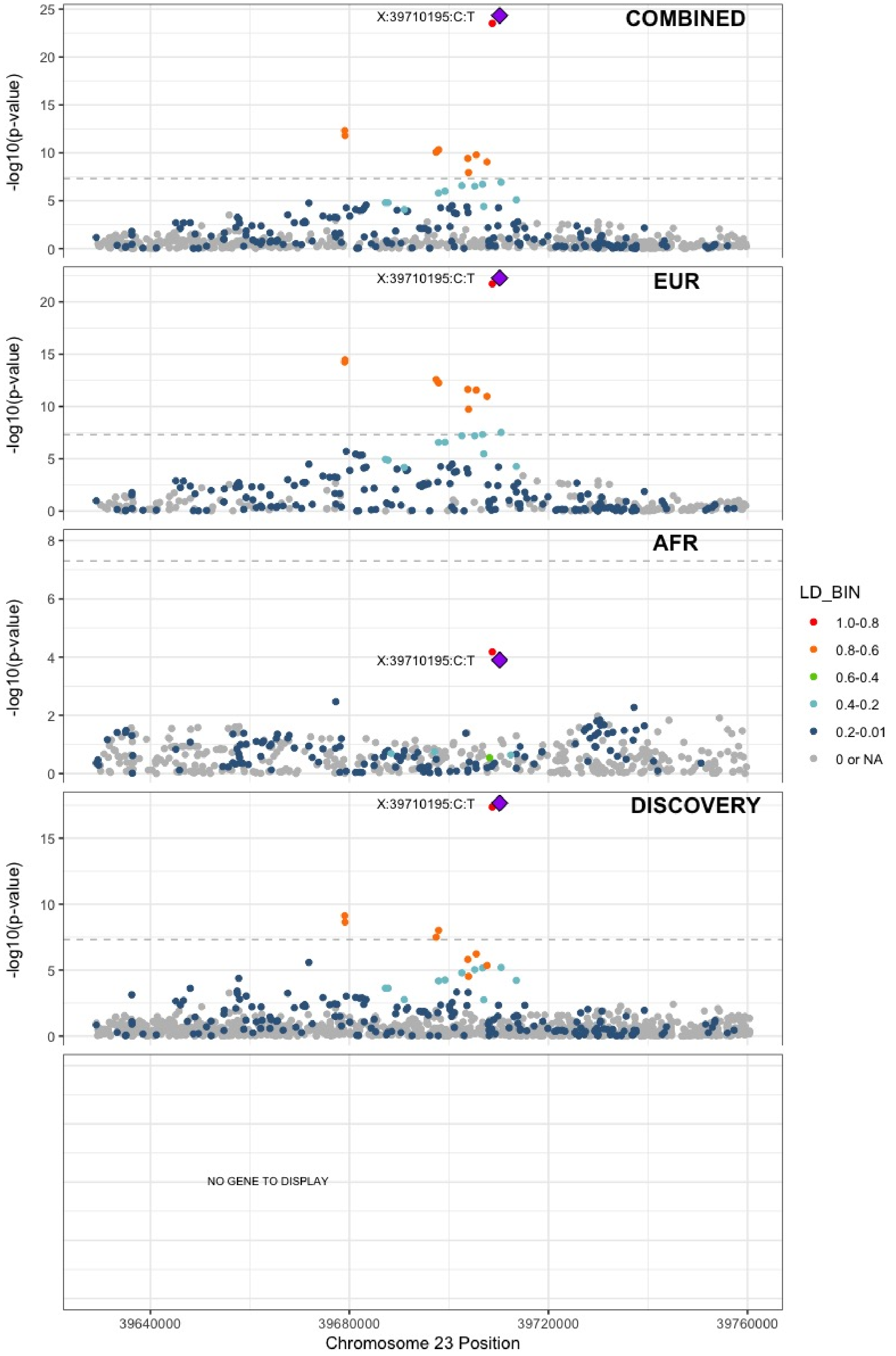

**Supplementary Figure 6.110** – rs6048 (chr23:138633280) in locus 134 (F9)

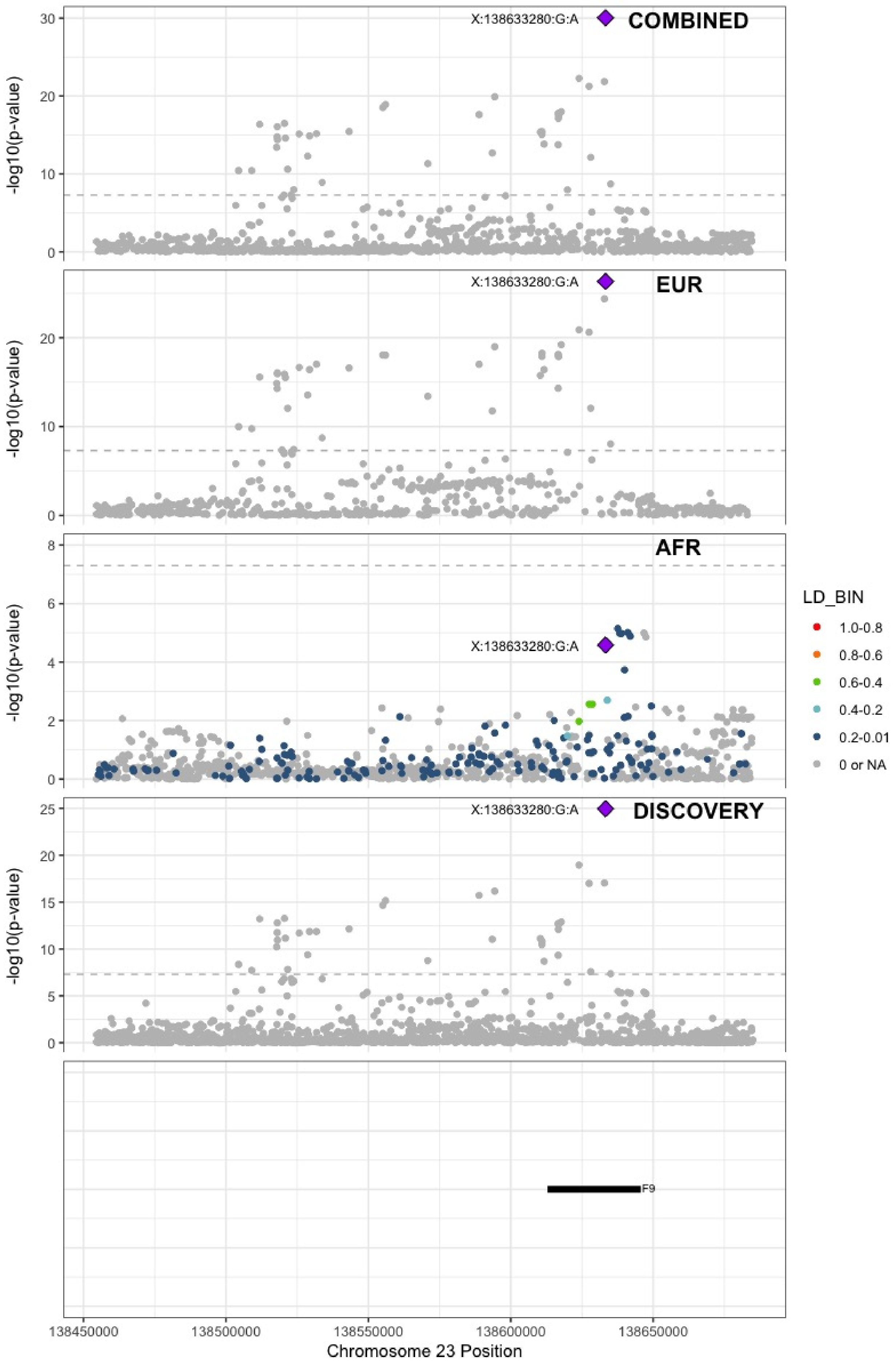

**Supplementary Figure 6.111** – rs2084408 (chr23:154346709) in locus 135 (BRCC3)

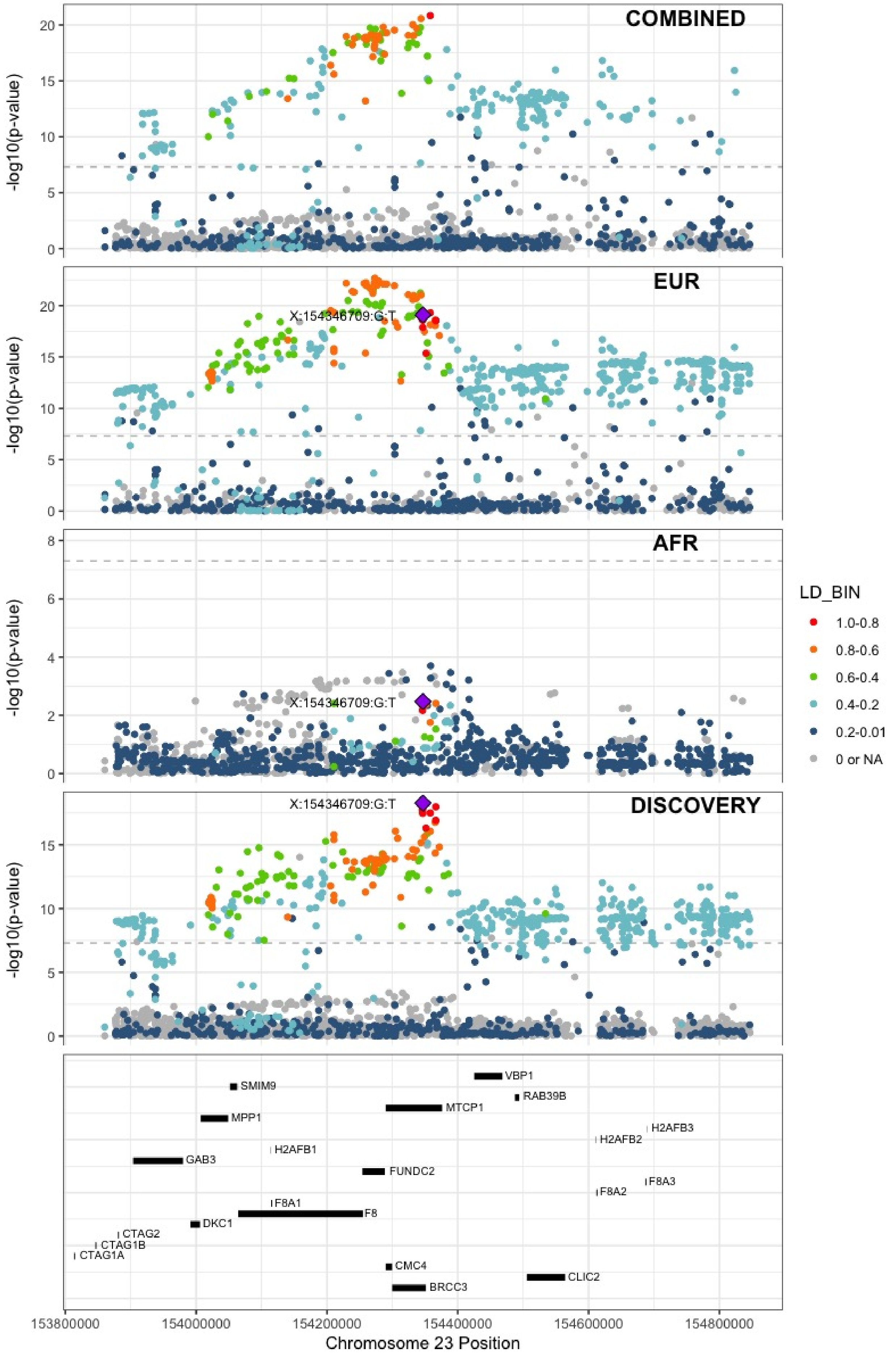
