## Supplemental Figure 7 for "Cross-Ancestry Investigation of Venous Thromboembolism Genomic Predictors"

### Supplementary Figure 7.
### Regional association plots of European analysis lead variants.


7.1 rs9442571 (chr1:9349611) in locus 1 (H6PD ;SPSB1) . . . . . . . . . . . . . . . . . . . . 3
7.2 rs551176418 (chr1:27107263) in locus 2 (ARID1A) . . . . . . . . . . . . . . . . . . . . 4
7.3 rs12753797 (chr1:77981959) in locus 3 (AK5) . . . . . . . . . . . . . . . . . . . . . . 5
7.4 rs4970834 (chr1:109814880) in locus 4 (CELSR2) . . . . . . . . . . . . . . . . . . . . 6
7.5 rs1775815 (chr1:118148830) in locus 5 (TENT5C) . . . . . . . . . . . . . . . . . . . . 7
7.6 rs1267881263 (chr1:150496127) in locus 6 (FALEC ;ADAMTSL4) . . . . . . . . . . . . . 8
7.7 rs76798800 (chr1:154994978) in locus 7 (DCST2) . . . . . . . . . . . . . . . . . . . . 9
7.8 rs3557 (chr1:161188893) in locus 8 (FCER1G) . . . . . . . . . . . . . . . . . . . . . . 10
7.9 rs6025 (chr1:169519049) in locus 9 (F5) . . . . . . . . . . . . . . . . . . . . . . . . 11
7.10 rs4540639 (chr1:192104320) in locus 11 (LINC01680 ;RGS18) . . . . . . . . . . . . . . 12
7.11 rs2842700 (chr1:207282149) in locus 13 (C4BPA) . . . . . . . . . . . . . . . . . . . . 13
7.12 rs3811444 (chr1:248039451) in locus 14 (TRIM58) . . . . . . . . . . . . . . . . . . . 14
7.13 rs1867312 (chr2:68619981) in locus 17 (PLEK) . . . . . . . . . . . . . . . . . . . . . 15
7.14 rs1158867 (chr2:128177377) in locus 18 (PROC) . . . . . . . . . . . . . . . . . . . . 16
7.15 rs1912846 (chr2:188325444) in locus 19 (CALCRL ;TFPI) . . . . . . . . . . . . . . . . 17
7.16 rs78872368 (chr2:198545250) in locus 20 (RFTN2 ;MARS2) . . . . . . . . . . . . . . . . 18
7.17 rs1047891 (chr2:211540507) in locus 21 (CPS1) . . . . . . . . . . . . . . . . . . . . . 19
7.18 rs13412535 (chr2:224874874) in locus 22 (SERPINE2) . . . . . . . . . . . . . . . . . . 20
7.19 rs13084580 (chr3:39188182) in locus 24 (CSRNP1) . . . . . . . . . . . . . . . . . . . 21
7.20 rs11130326 (chr3:52771920) in locus 25 (NEK4) . . . . . . . . . . . . . . . . . . . . 22
7.21 rs9866664 (chr3:90437988) in locus 26 (EPHA3 ;NONE) . . . . . . . . . . . . . . . . . 23
7.22 rs28479320 (chr3:93752529) in locus 27 (ARL13B) . . . . . . . . . . . . . . . . . . . 24
7.23 rs9854955 (chr3:156795525) in locus 30 (LEKR1 ;LINC00880) . . . . . . . . . . . . . . 25
7.24 rs710446 (chr3:186459927) in locus 32 (KNG1) . . . . . . . . . . . . . . . . . . . . . 26
7.25 rs3796159 (chr3:194790434) in locus 33 (XXYLT1) . . . . . . . . . . . . . . . . . . . 27
7.26 rs72976751 (chr4:86725959) in locus 37 (ARHGAP24) . . . . . . . . . . . . . . . . . . 28
7.27 rs35225200 (chr4:103146888) in locus 38 (BANK1 ;SLC39A8) . . . . . . . . . . . . . . 29
7.28 rs2066864 (chr4:155525695) in locus 39 (FGG) . . . . . . . . . . . . . . . . . . . . . 30
7.29 rs3756011 (chr4:187206249) in locus 40 (F11) . . . . . . . . . . . . . . . . . . . . . 31
7.30 rs112367053 (chr5:28379046) in locus 41 (LINC02103 ;LSP1P3) . . . . . . . . . . . . . 32
7.31 rs16867574 (chr5:38708554) in locus 42 (OSMR-AS1) . . . . . . . . . . . . . . . . . . 33
7.32 rs4235677 (chr5:72925091) in locus 43 (ARHGEF28) . . . . . . . . . . . . . . . . . . 34
7.33 rs251339 (chr5:96235038) in locus 44 (ERAP2) . . . . . . . . . . . . . . . . . . . . . 35
7.34 rs147133967 (chr5:132426851) in locus 45 (HSPA4) . . . . . . . . . . . . . . . . . . 36
7.35 rs214059 (chr6:25536937) in locus 46 (CARMIL1) . . . . . . . . . . . . . . . . . . . 37
7.36 rs2394251 (chr6:29943688) in locus 47 (HCG9) . . . . . . . . . . . . . . . . . . . . . 38
7.37 rs28383307 (chr6:32586755) in locus 48 (HLA-DRB1 ;HLA-DQA1) . . . . . . . . . . . . 39
7.38 rs145294670 (chr6:34622561) in locus 49 (ILRUN) . . . . . . . . . . . . . . . . . . . 40
7.39 rs2754251 (chr6:88385949) in locus 51 (AKIRIN2) . . . . . . . . . . . . . . . . . . . 41
7.40 rs9390460 (chr6:147694334) in locus 52 (STXBP5) . . . . . . . . . . . . . . . . . . . 42
7.41 rs1513275 (chr7:28259233) in locus 53 (JAZF1-AS1) . . . . . . . . . . . . . . . . . . 43
7.42 rs370720 (chr8:9168146) in locus 57 (LOC101929128 ;LOC157273) . . . . . . . . . . . 44
7.43 rs2726950 (chr8:27806764) in locus 60 (SCARA5) . . . . . . . . . . . . . . . . . . . . 45
7.44 rs6993770 (chr8:106581528) in locus 62 (ZFPM2) . . . . . . . . . . . . . . . . . . . . 46
7.45 rs1491336705 (chr8:108295235) in locus 63 (ANGPT1) . . . . . . . . . . . . . . . . . 47
7.46 rs10820606 (chr9:99192919) in locus 65 (ZNF367 ;HABP4) . . . . . . . . . . . . . . . . 48
7.47 rs687289 (chr9:136137106) in locus 66 (ABO) . . . . . . . . . . . . . . . . . . . . . . 49
7.48 rs1243184 (chr10:21931937) in locus 68 (MLLT10) . . . . . . . . . . . . . . . . . . . 50
7.49 rs10763665 (chr10:28771491) in locus 70 (LINC02652) . . . . . . . . . . . . . . . . . 51
7.50 rs78707713 (chr10:71245276) in locus 72 (TSPAN15) . . . . . . . . . . . . . . . . . . 52

7.51 rs57866767 (chr10:96023077) in locus 74 (PLCE1) . . . . 53
7.52 rs10886430 (chr10:121010256) in locus 75 (GRK5) . . . . 54
7.53 rs7122100 (chr11:10732560) in locus 76 (IRAG1;CTR9) . . . . 55
7.54 rs12800654 (chr11:16243320) in locus 77 (SOX6) . . . . 56
7.55 rs4755476 (chr11:32980428) in locus 78 (QSER1) . . . . 57
7.56 rs1799963 (chr11:46761055) in locus 79 (F2) . . . . 58
7.57 rs141798115 (chr11:56875074) in locus 80 (OR5AK4P;LRRC55) . . . . 59
7.58 rs55777218 (chr11:60070946) in locus 81 (MS4A4A) . . . . 60
7.59 rs174551 (chr11:61573684) in locus 82 (FADS1) . . . . 61
7.60 rs1145656 (chr11:73305859) in locus 83 (FAM168A) . . . . 62
7.61 rs1997547 (chr11:114002744) in locus 84 (ZBTB16) . . . . 63
7.62 rs35257264 (chr11:126296816) in locus 85 (ST3GAL4) . . . . 64
7.63 rs7135039 (chr12:6160614) in locus 87 (VWF) . . . . 65
7.64 rs7311483 (chr12:9053661) in locus 88 (A2ML1;PHC1) . . . . 66
7.65 rs4759076 (chr12:54729872) in locus 89 (COPZ1) . . . . 67
7.66 rs142351376 (chr12:104136288) in locus 90 (STAB2) . . . . 68
7.67 rs56750693 (chr12:112144586) in locus 91 (ACAD10) . . . . 69
7.68 rs2851436 (chr12:123667354) in locus 92 (MPHOSPH9) . . . . 70
7.69 rs3211752 (chr13:113787459) in locus 94 (F10) . . . . 71
7.70 rs8004077 (chr14:58846731) in locus 95 (ARID4A;TOMM20L) . . . . 72
7.71 rs2127869 (chr14:65794352) in locus 96 (LINC02324;MIR4708) . . . . 73
7.72 rs61990092 (chr14:92289301) in locus 97 (TC2N) . . . . 74
7.73 rs28929474 (chr14:94844947) in locus 98 (SERPINA1) . . . . 75
7.74 rs4906229 (chr14:103057511) in locus 99 (LINC02323;RCOR1) . . . . 76
7.75 rs141866277 (chr15:43950699) in locus 100 (PPIP5K1P1-CATSPER2) . . . . 77
7.76 rs340009 (chr15:60899639) in locus 101 (RORA-AS1) . . . . 78
7.77 rs7183672 (chr15:96101018) in locus 103 (LINC00924;LOC105369212) . . . . 79
7.78 rs12443808 (chr16:30996871) in locus 106 (HSD3B7) . . . . 80
7.79 rs1346057617 (chr16:75434183) in locus 108 (CFDP1) . . . . 81
7.80 rs62044142 (chr16:81868968) in locus 109 (PLCG2) . . . . 82
7.81 rs28634651 (chr16:88553198) in locus 111 (ZFPM1) . . . . 83
7.82 rs4790311 (chr17:1979188) in locus 112 (SMG6) . . . . 84
7.83 rs11078659 (chr17:6903944) in locus 113 (ALOX12-AS1) . . . . 85
7.84 rs536327 (chr17:27857698) in locus 114 (TAOK1) . . . . 86
7.85 rs9468 (chr17:44101563) in locus 115 (MAPT) . . . . 87
7.86 rs1801690 (chr17:64208285) in locus 116 (APOH) . . . . 88
7.87 rs573431210 (chr17:67276383) in locus 117 (ABCA5) . . . . 89
7.88 rs8109681 (chr19:10738836) in locus 119 (SLC44A2) . . . . 90
7.89 rs2545774 (chr19:41287674) in locus 121 (RAB4B) . . . . 91
7.90 rs8108474 (chr19:46301479) in locus 122 (RSPH6A) . . . . 92
7.91 rs646327 (chr19:49209851) in locus 123 (FUT2) . . . . 93
7.92 rs1654425 (chr19:55538980) in locus 124 (GP6) . . . . 94
7.93 rs117390891 (chr20:23168526) in locus 126 (LINC00656;NXT1) . . . . 95
7.94 rs6087685 (chr20:33777612) in locus 127 (MMP24-AS1-EDEM2) . . . . 96
7.95 rs3070580 (chr22:33165231) in locus 129 (SYN3) . . . . 97
7.96 rs9611844 (chr22:43115776) in locus 130 (A4GALT) . . . . 98
7.97 rs12388118 (chr23:11380742) in locus 131 (ARHGAP6) . . . . 99
7.98 rs3002416 (chr23:39710195) in locus 133 (MIR1587;BCOR) . . . . 100
7.99 rs6048 (chr23:138633280) in locus 134 (F9) . . . . 101
7.100rs4898406 (chr23:154273269) in locus 135 (FUNDC2) . . . . 102

**Supplementary Figure 7.1** – rs9442571 (chr1:9349611) in locus 1 (H6PD ;SPSB1)

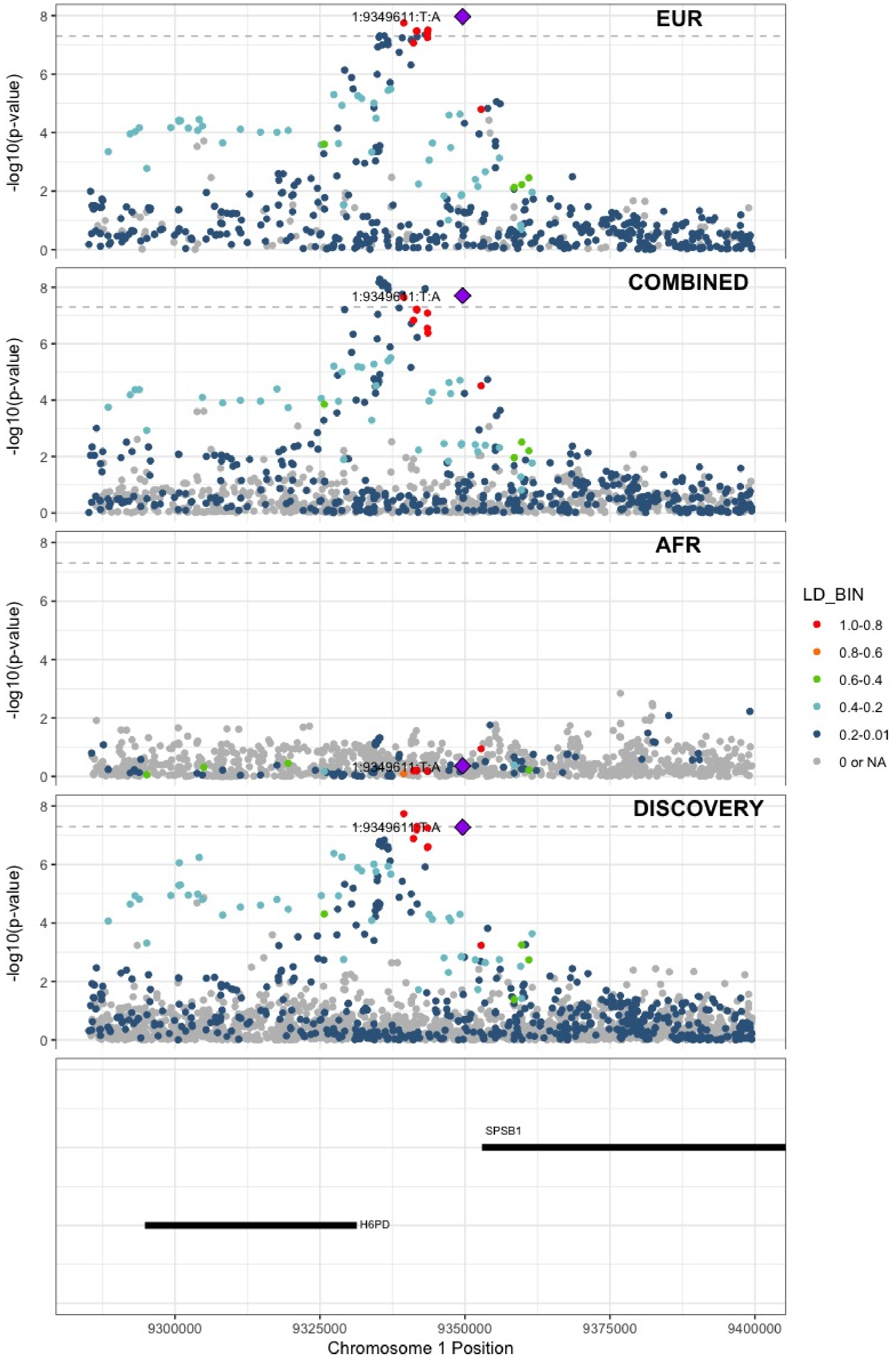

**Supplementary Figure 7.2** – rs551176418 (chr1:27107263) in locus 2 (ARID1A)

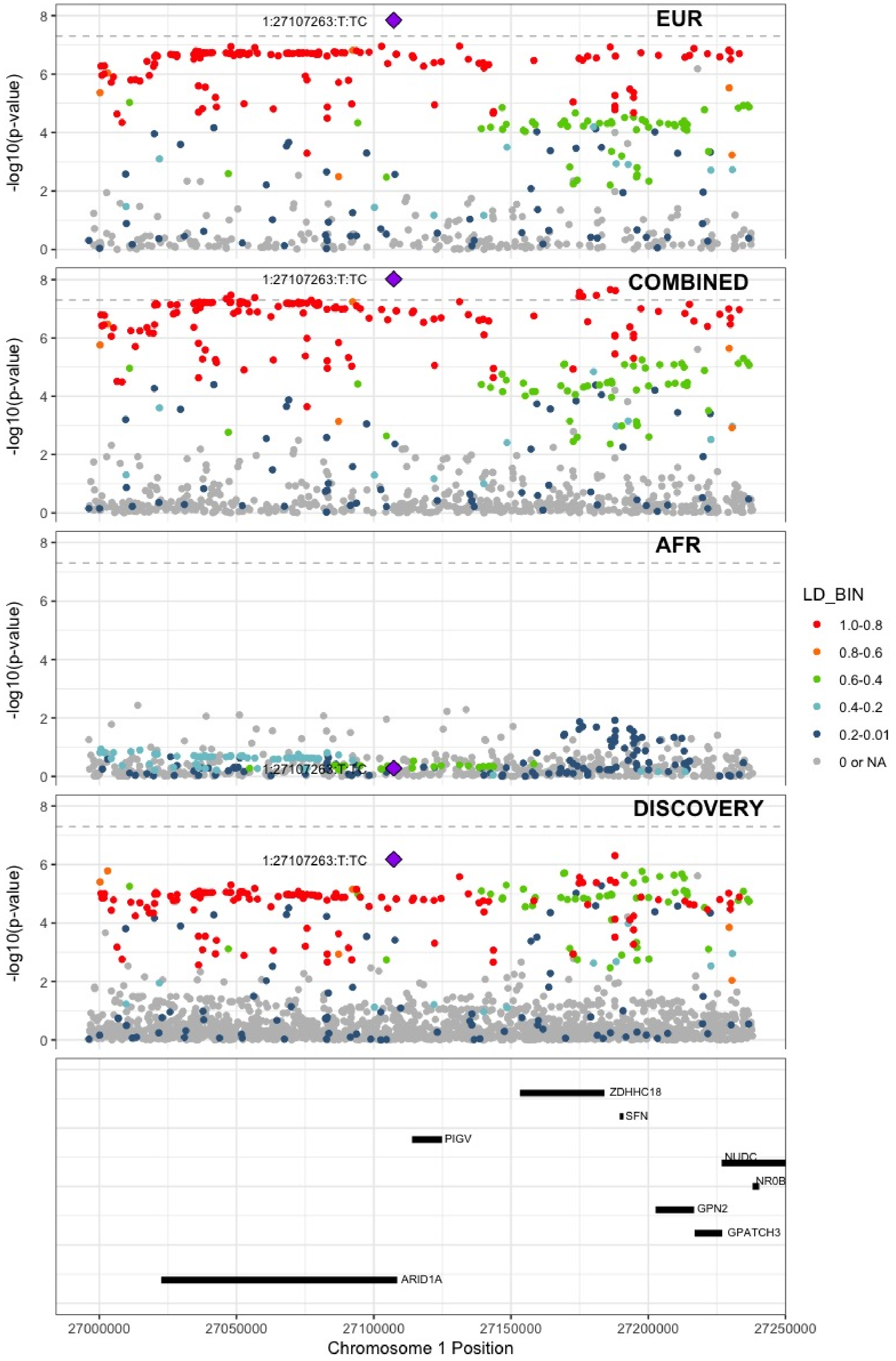

**Supplementary Figure 7.3** – rs12753797 (chr1:77981959) in locus 3 (AK5)

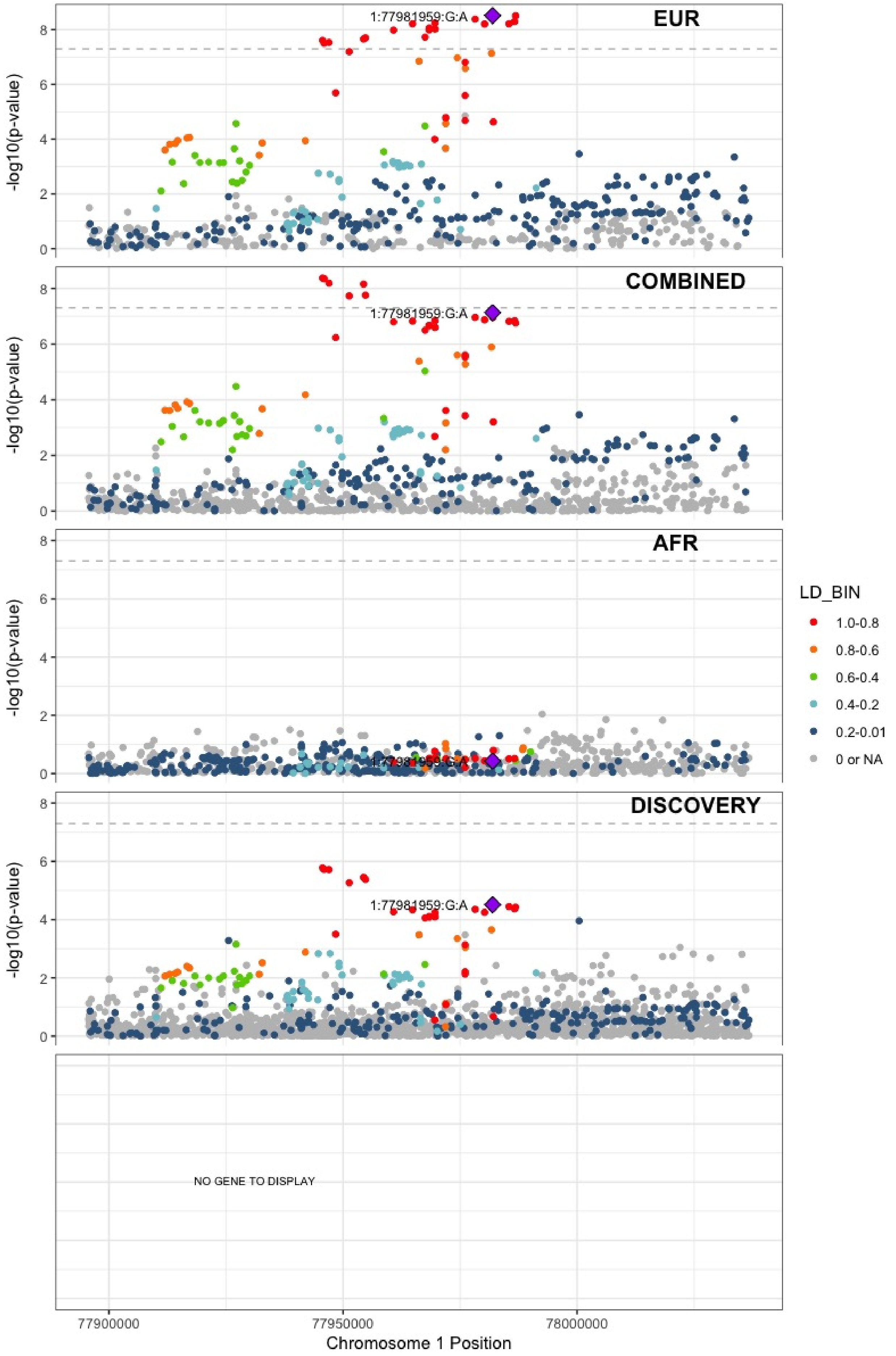

**Supplementary Figure 7.4** – rs4970834 (chr1:109814880) in locus 4 (CELSR2)

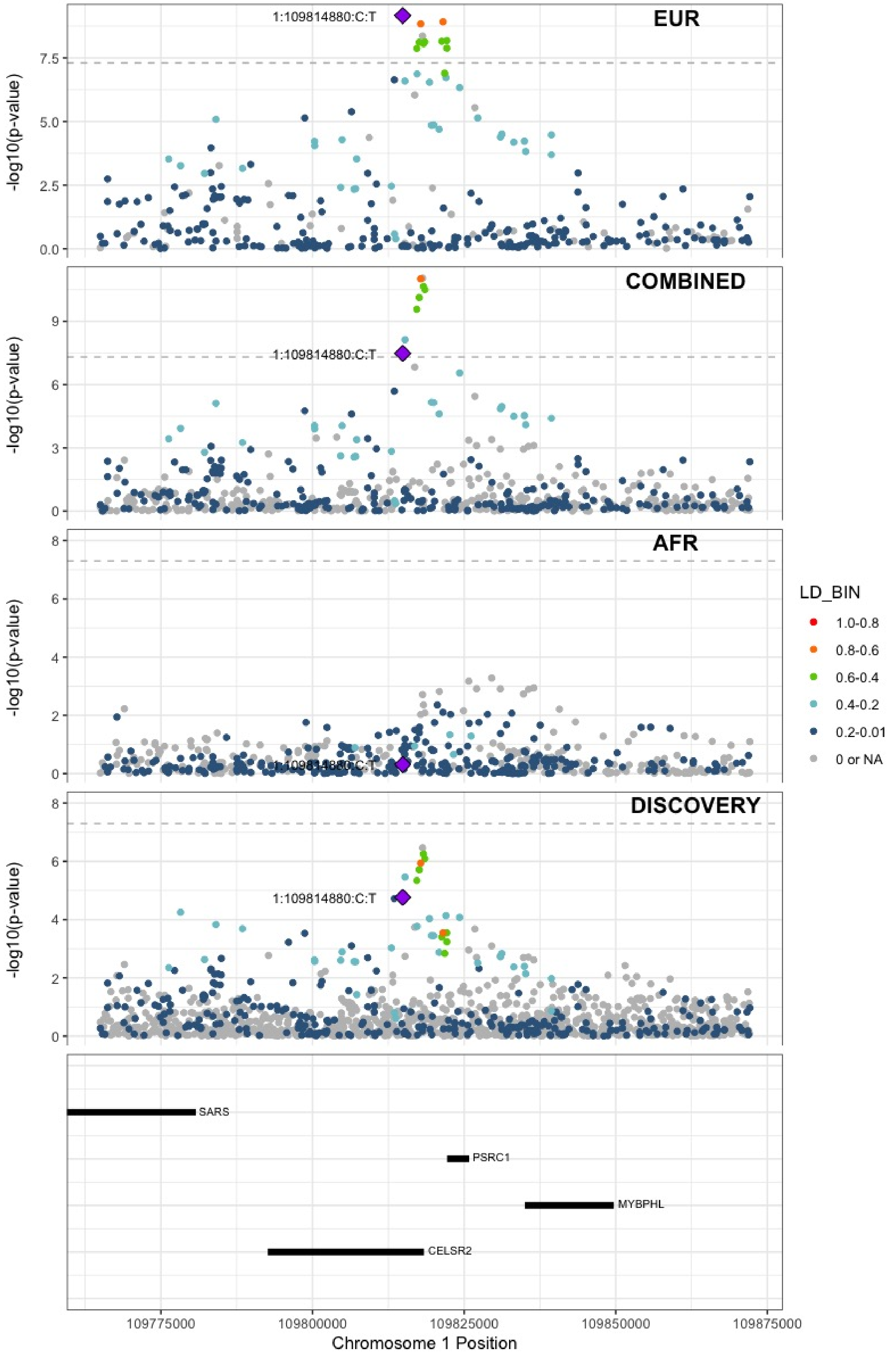

**Supplementary Figure 7.5** – rs1775815 (chr1:118148830) in locus 5 (TENT5C)

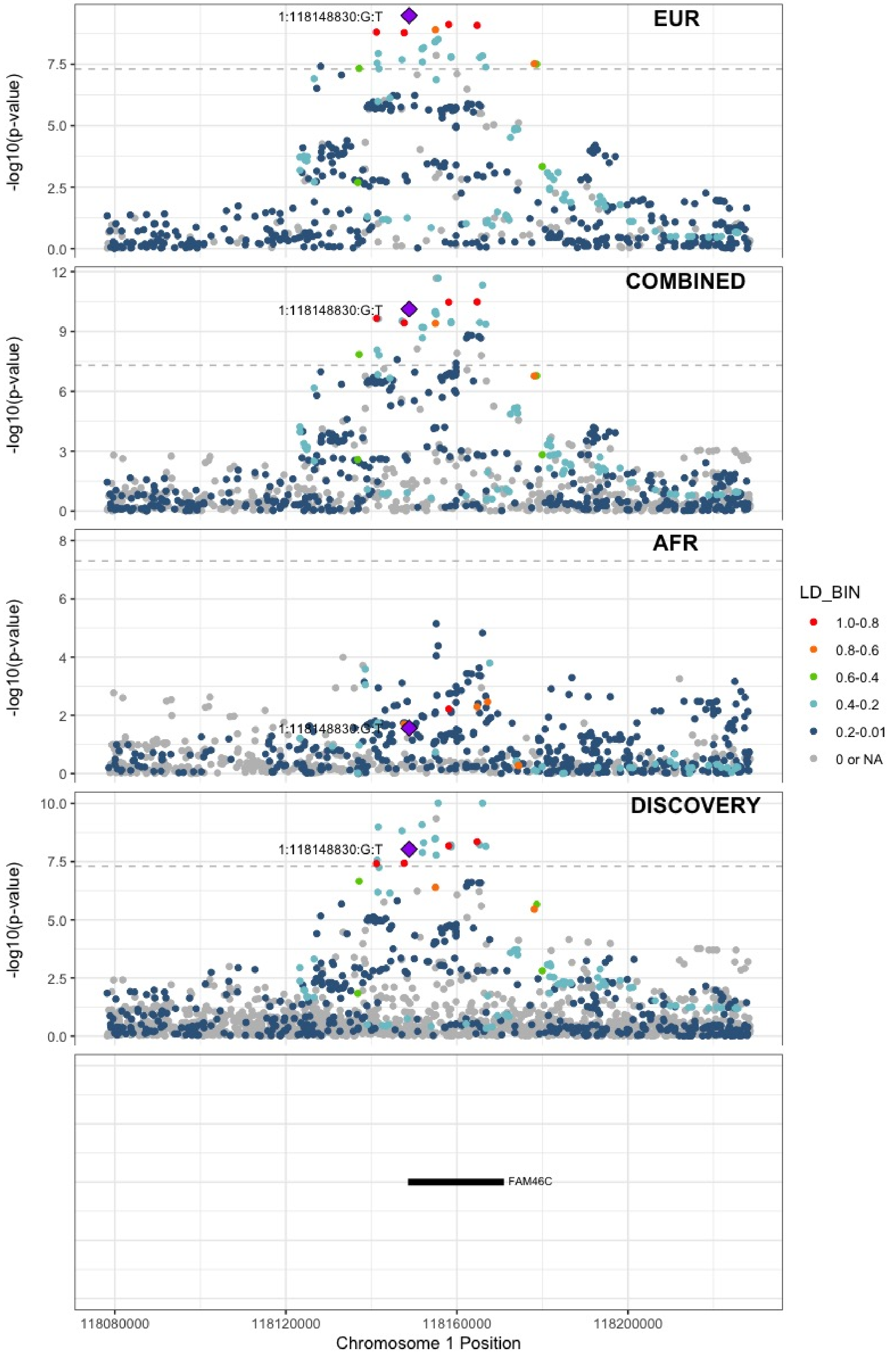

**Supplementary Figure 7.6** – rs1267881263 (chr1:150496127) in locus 6 (FALEC;ADAMTSL4)

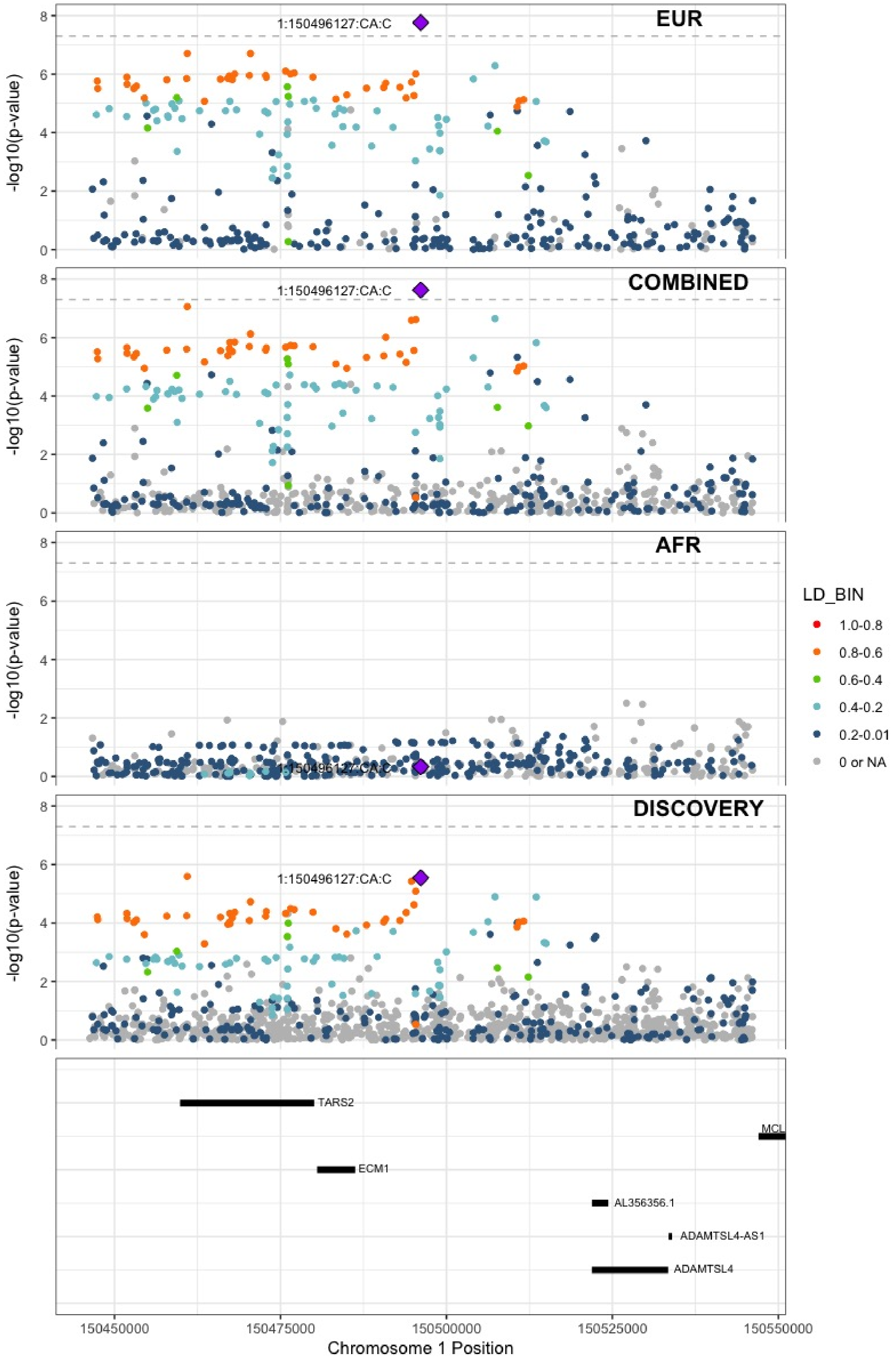

**Supplementary Figure 7.7** – rs76798800 (chr1:154994978) in locus 7 (DCST2)

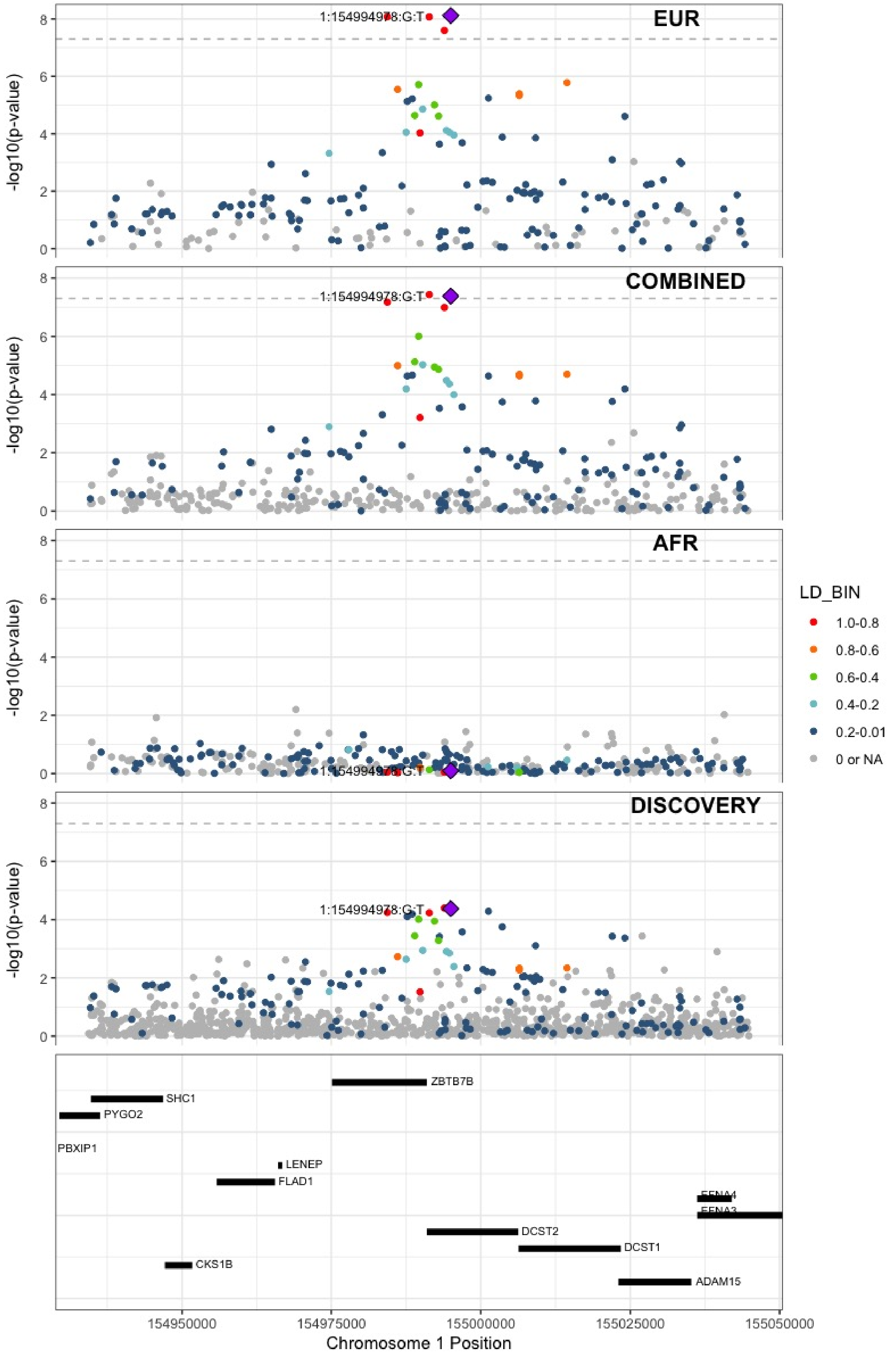

**Supplementary Figure 7.8** – rs3557 (chr1:161188893) in locus 8 (FCER1G)

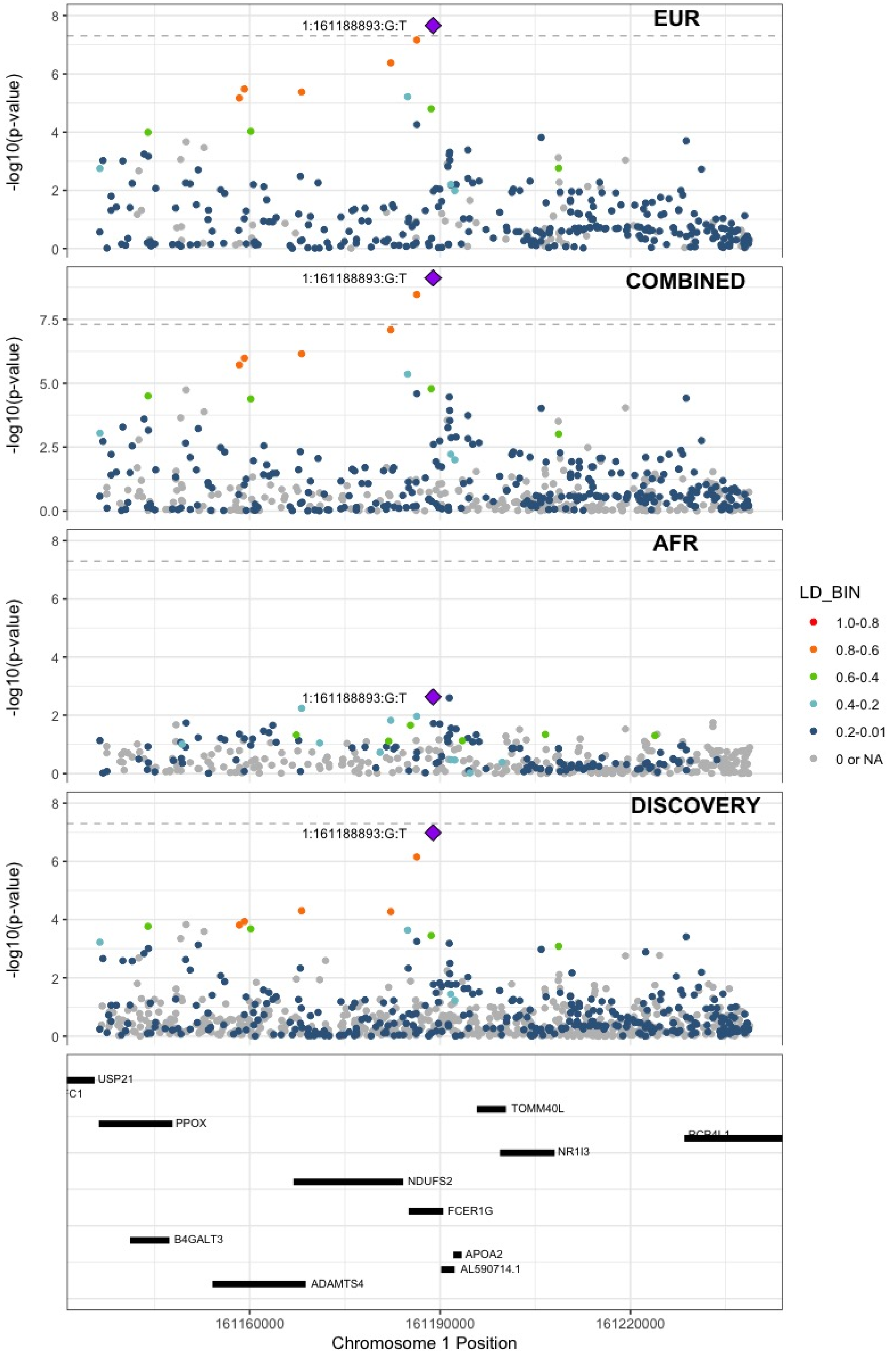

**Supplementary Figure 7.9** – rs6025 (chr1:169519049) in locus 9 (F5)

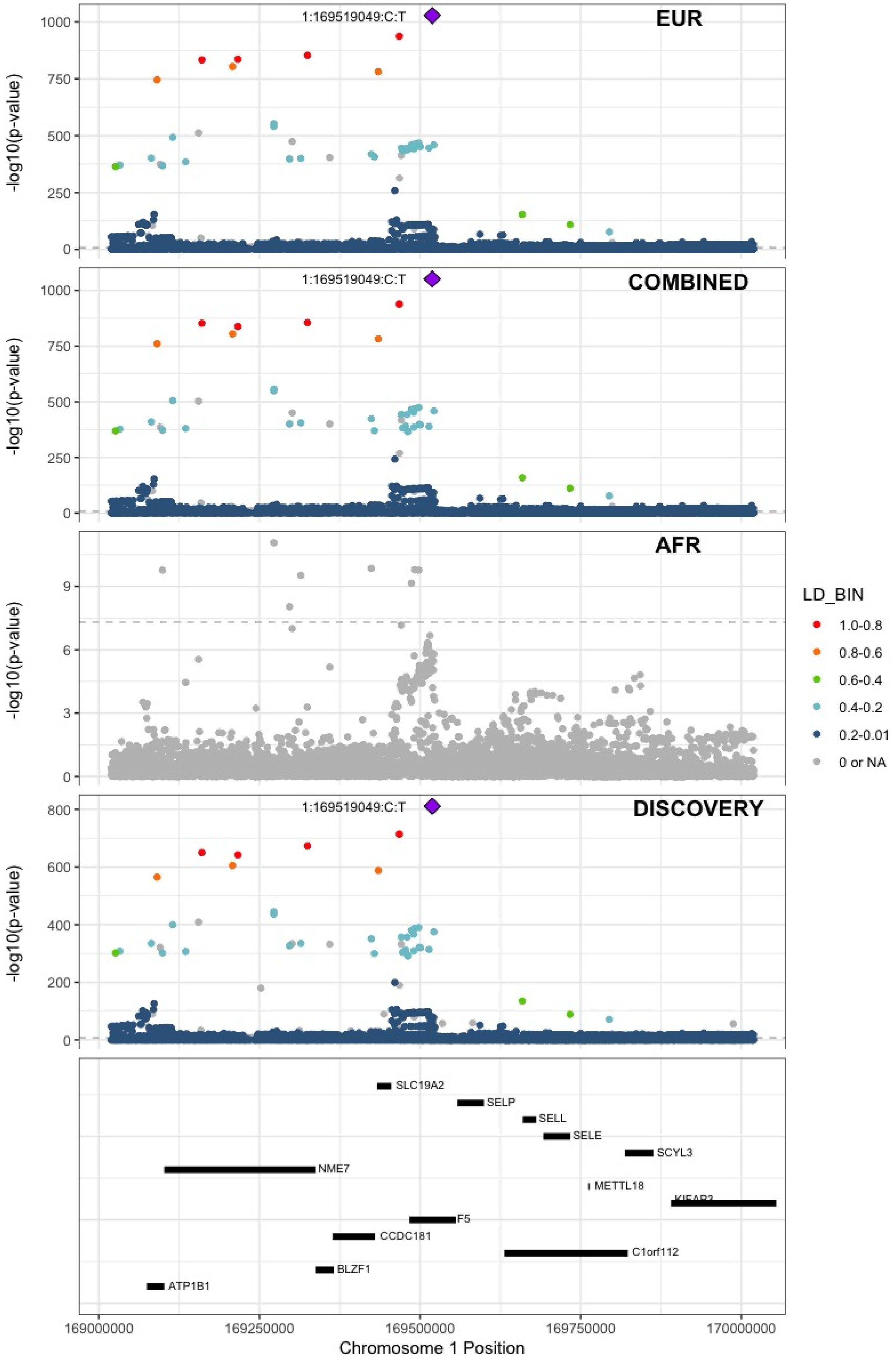

**Supplementary Figure 7.10** – rs4540639 (chr1:192104320) in locus 11 (LINC01680;RGS18)

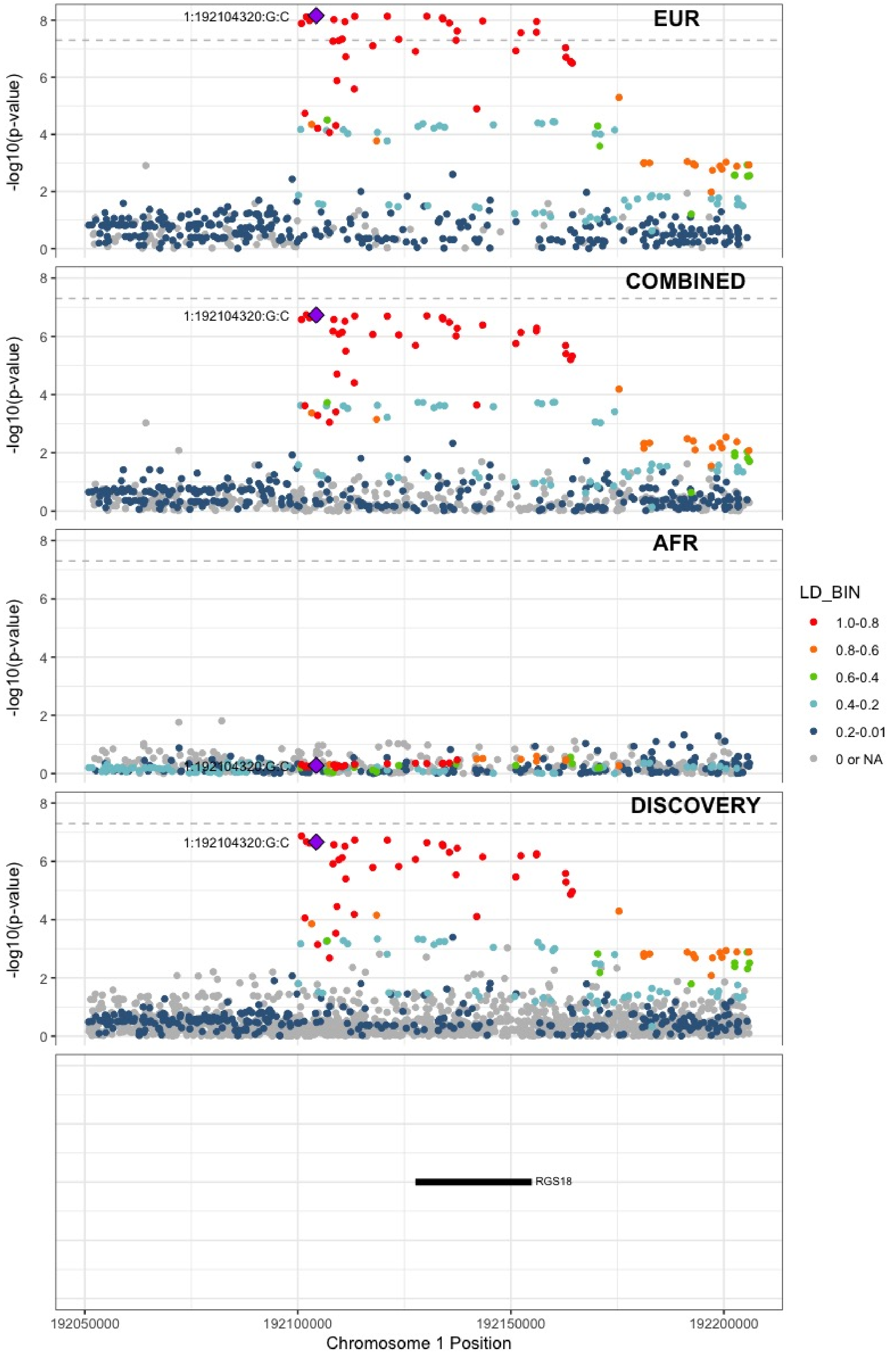

**Supplementary Figure 7.11** – rs2842700 (chr1:207282149) in locus 13 (C4BPA)

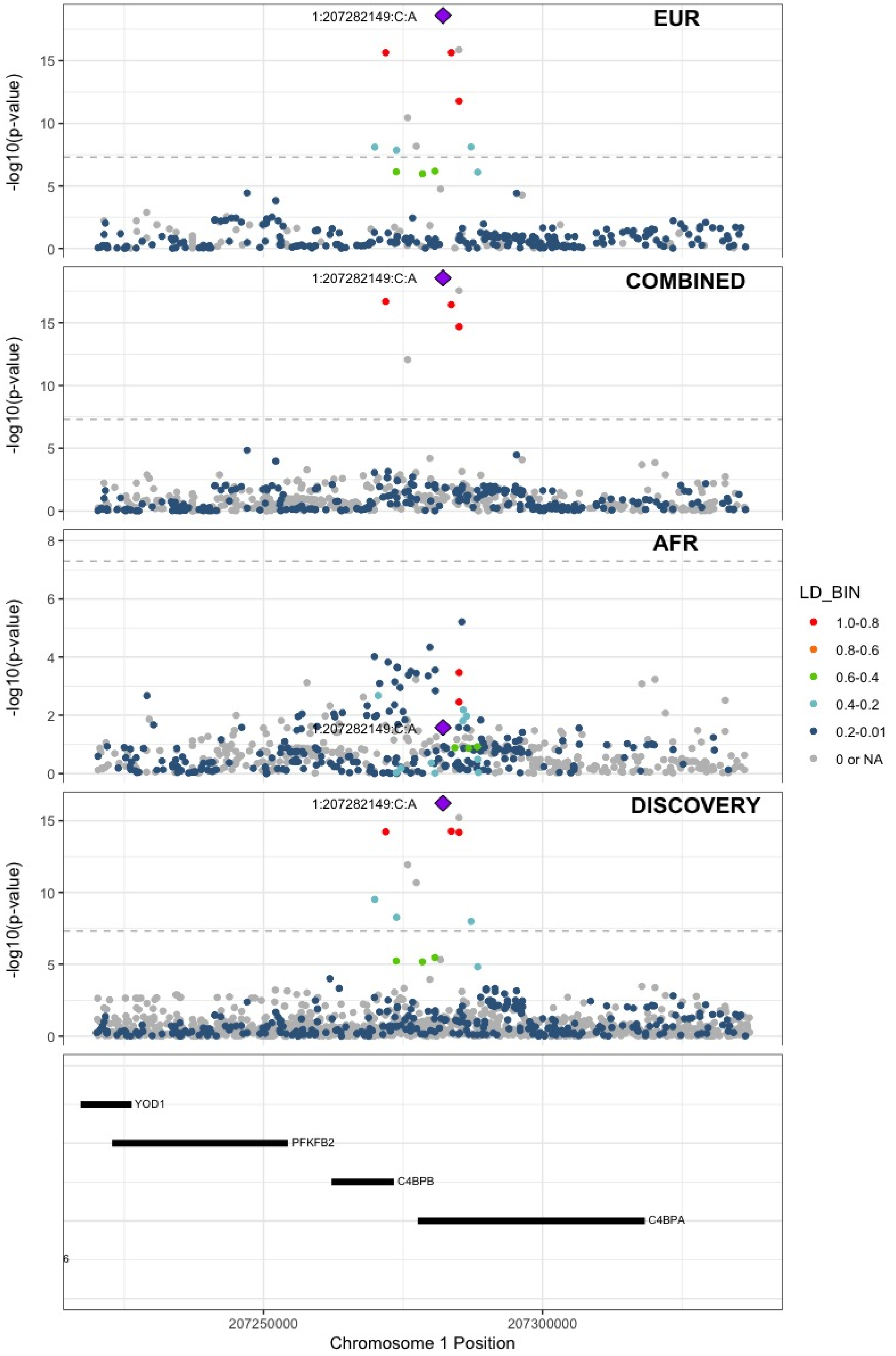

**Supplementary Figure 7.12** – rs3811444 (chr1:248039451) in locus 14 (TRIM58)

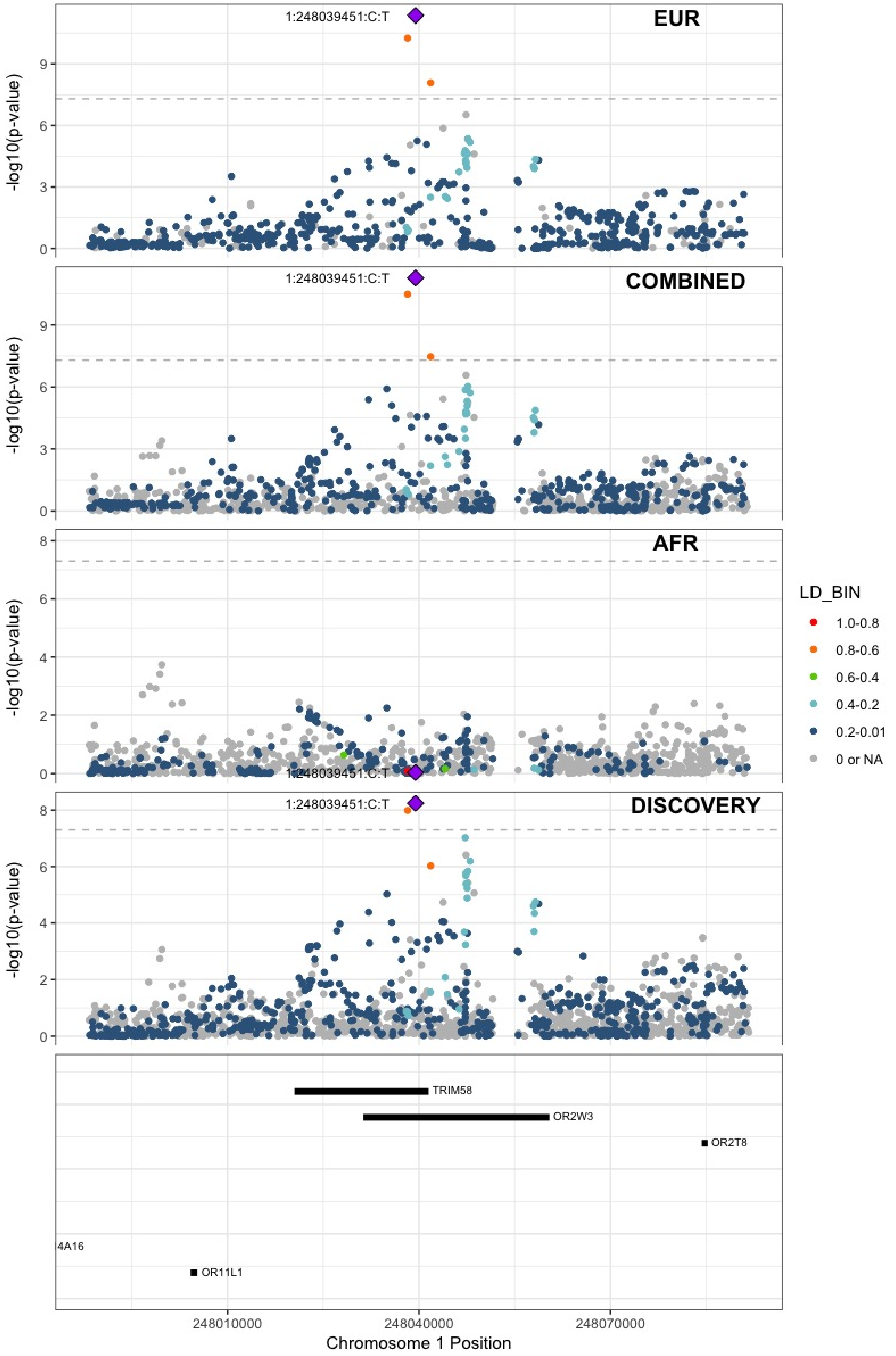

**Supplementary Figure 7.13** – rs1867312 (chr2:68619981) in locus 17 (PLEK)

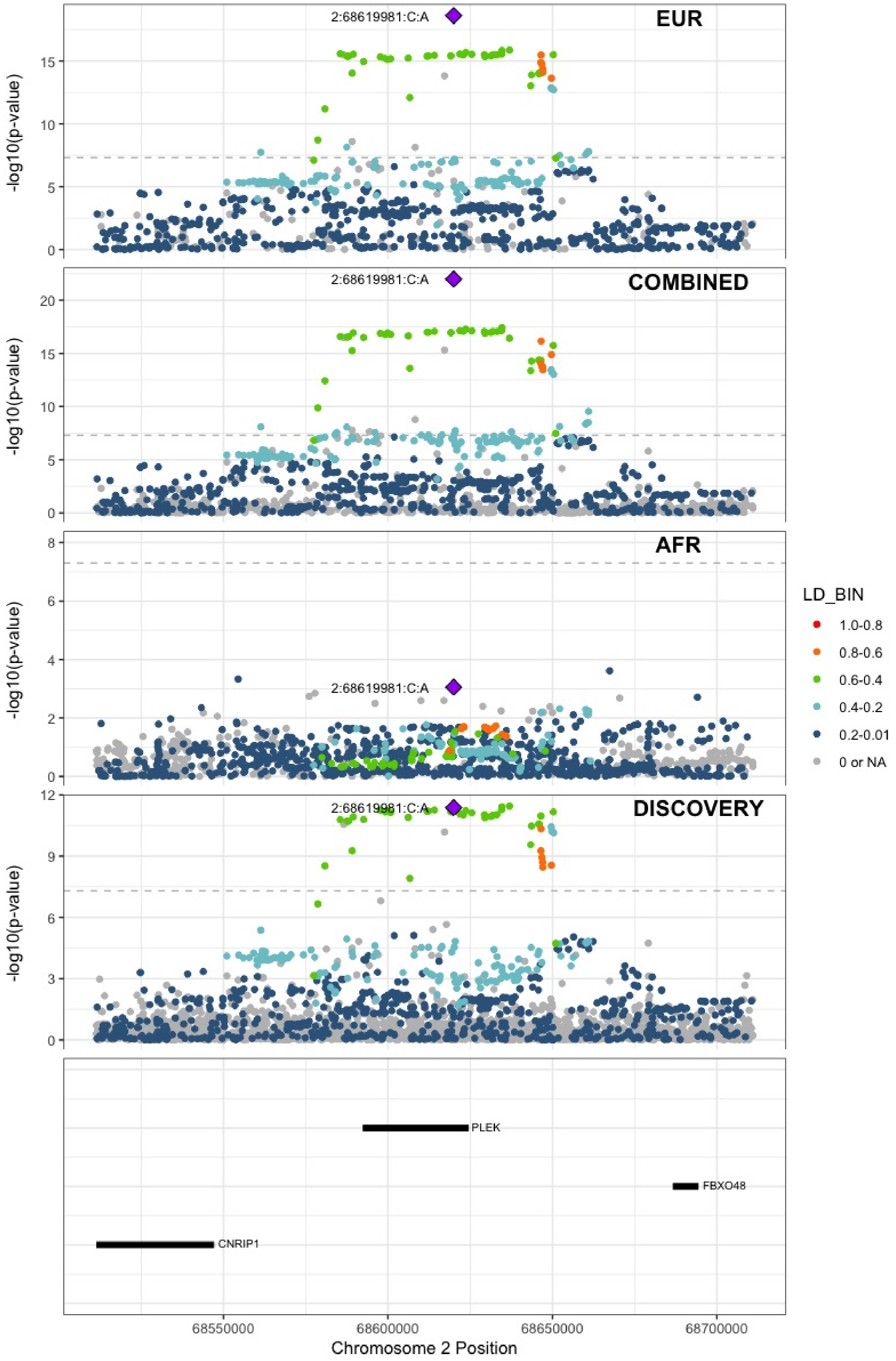

**Supplementary Figure 7.14** – rs1158867 (chr2:128177377) in locus 18 (PROC)

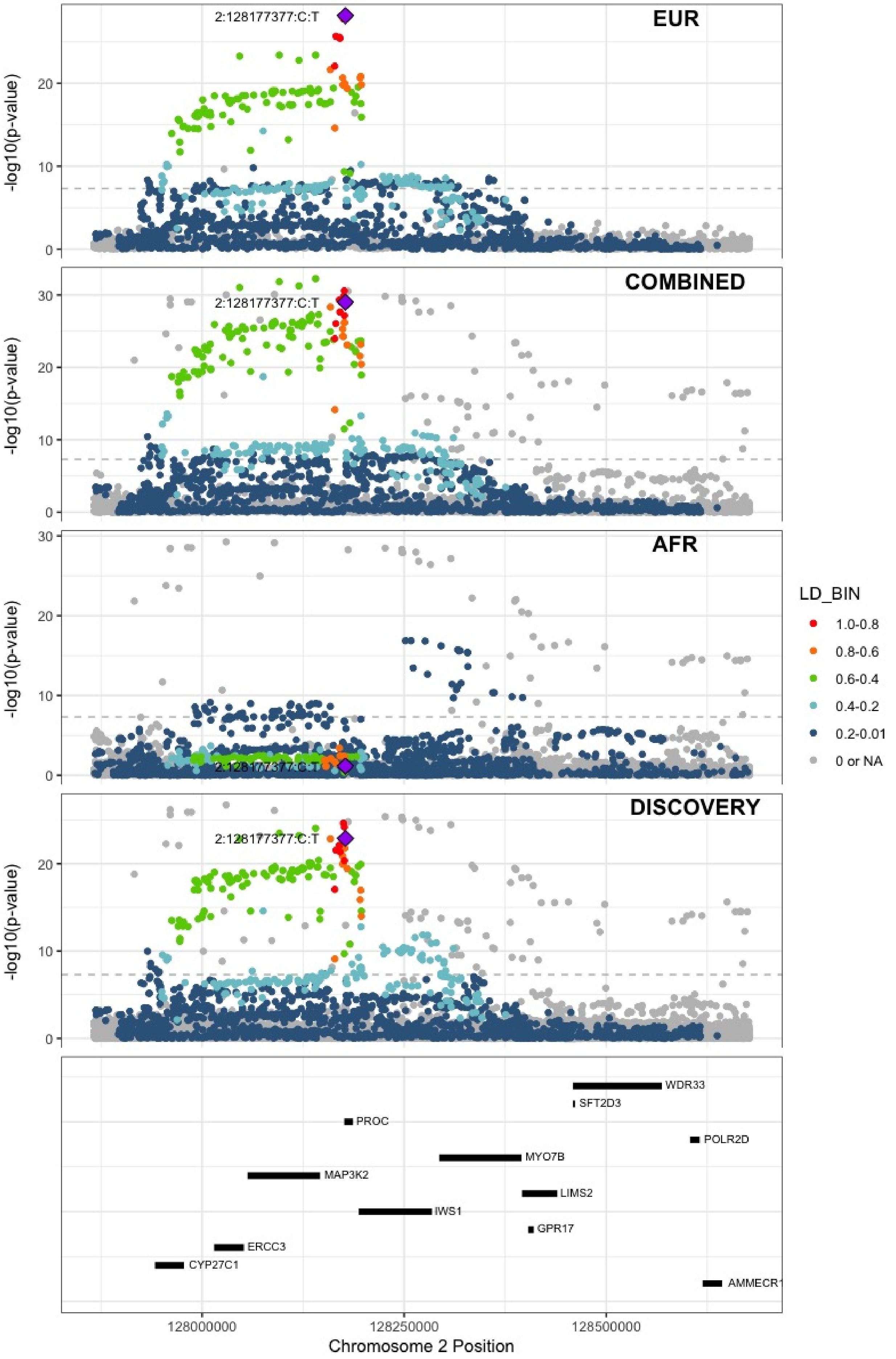

**Supplementary Figure 7.15** – rs1912846 (chr2:188325444) in locus 19 (CALCRL;TFPI)

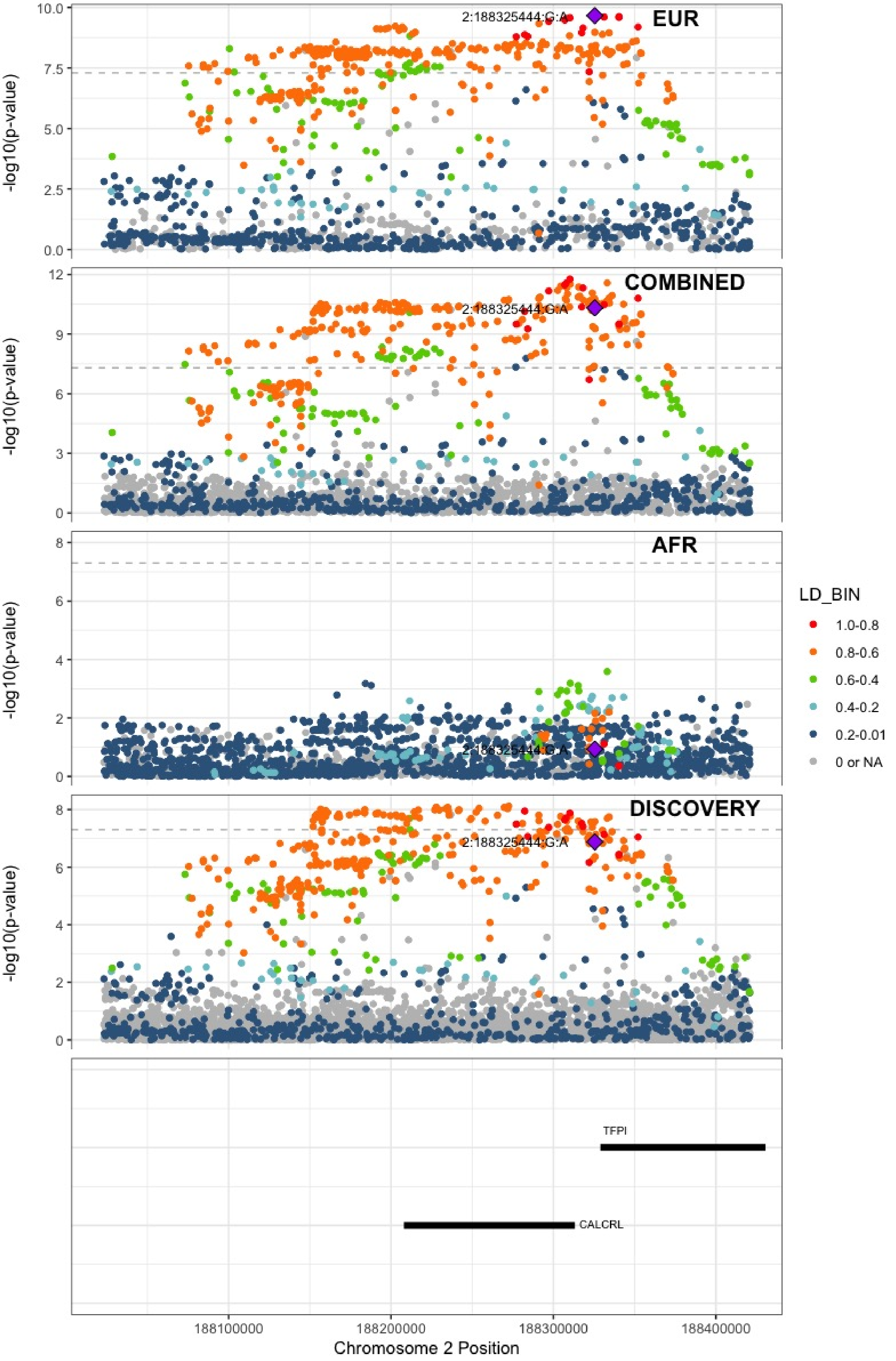

**Supplementary Figure 7.16** – rs78872368 (chr2:198545250) in locus 20 (RFTN2;MARS2)

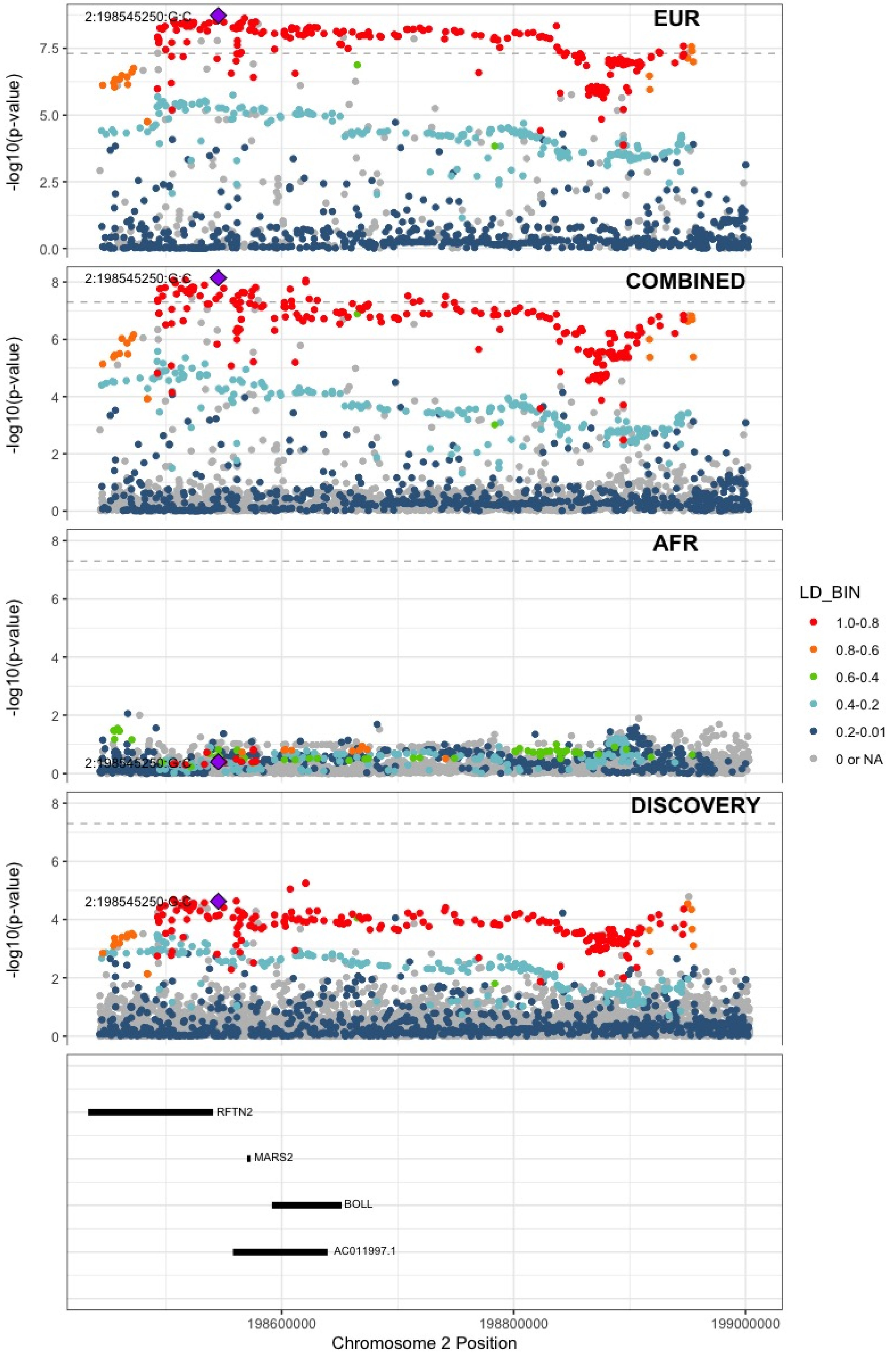

**Supplementary Figure 7.17** – rs1047891 (chr2:211540507) in locus 21 (CPS1)

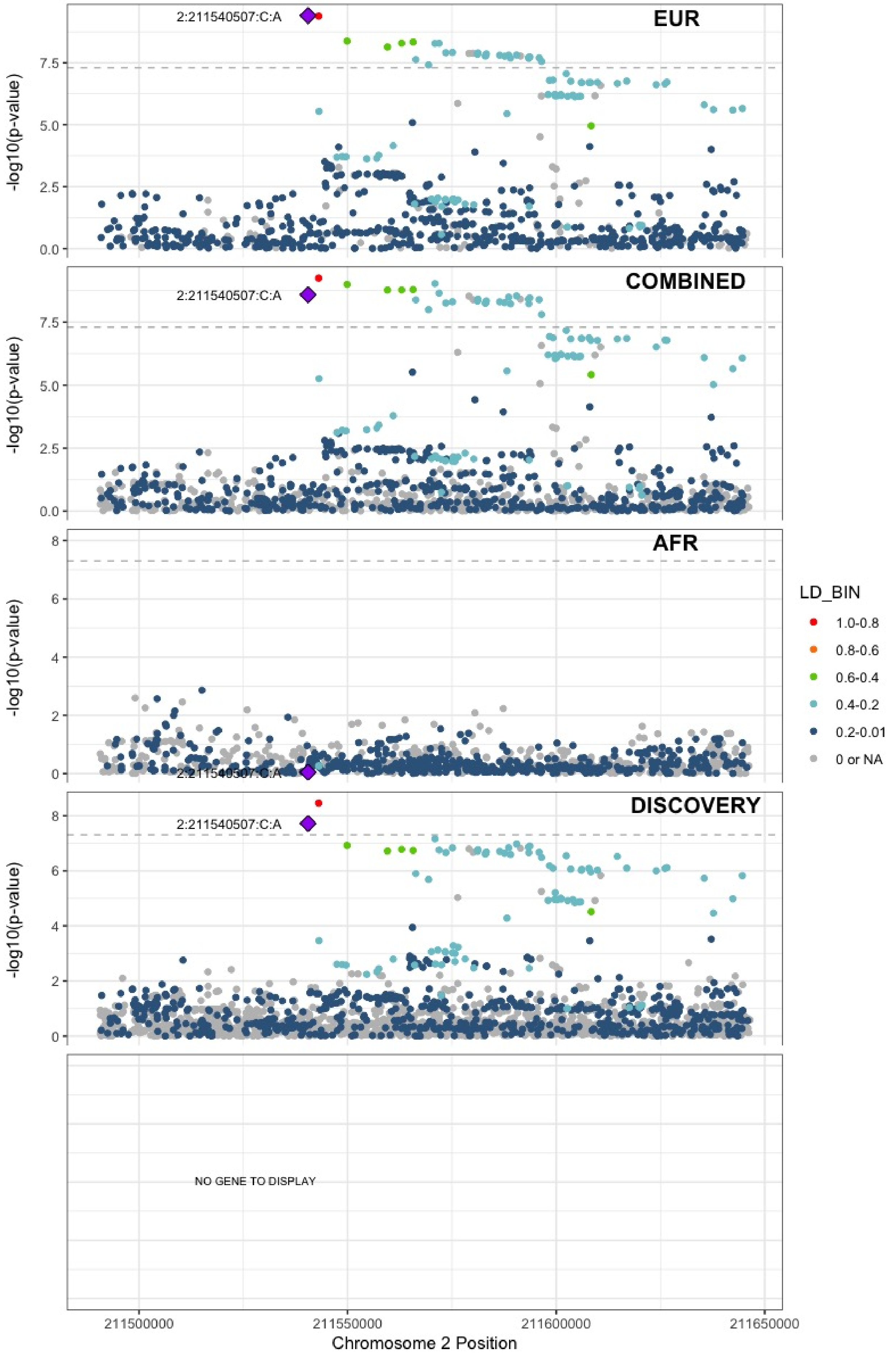

**Supplementary Figure 7.18** – rs13412535 (chr2:224874874) in locus 22 (SERPINE2)

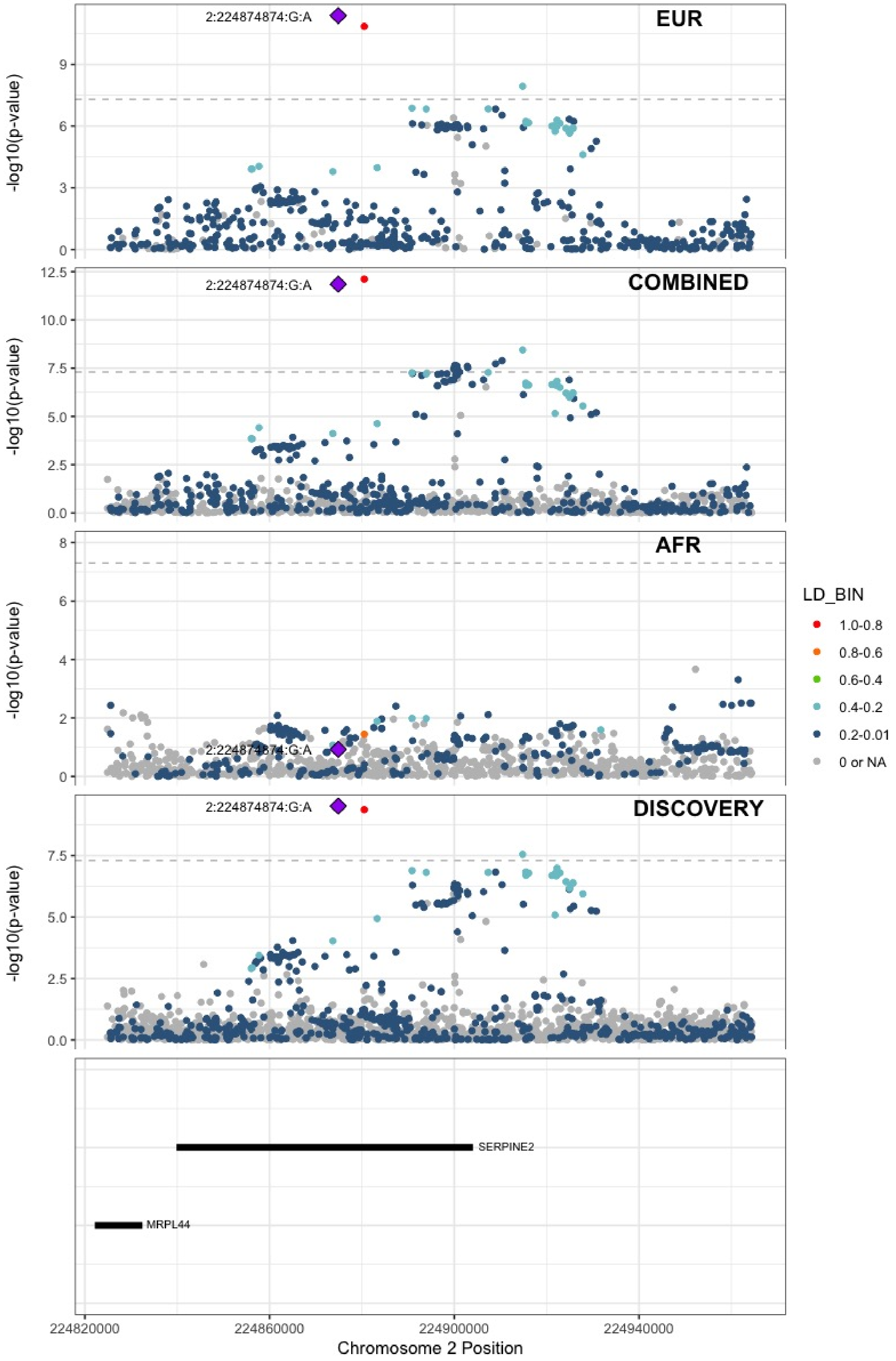

**Supplementary Figure 7.19** – rs13084580 (chr3:39188182) in locus 24 (CSRNP1)

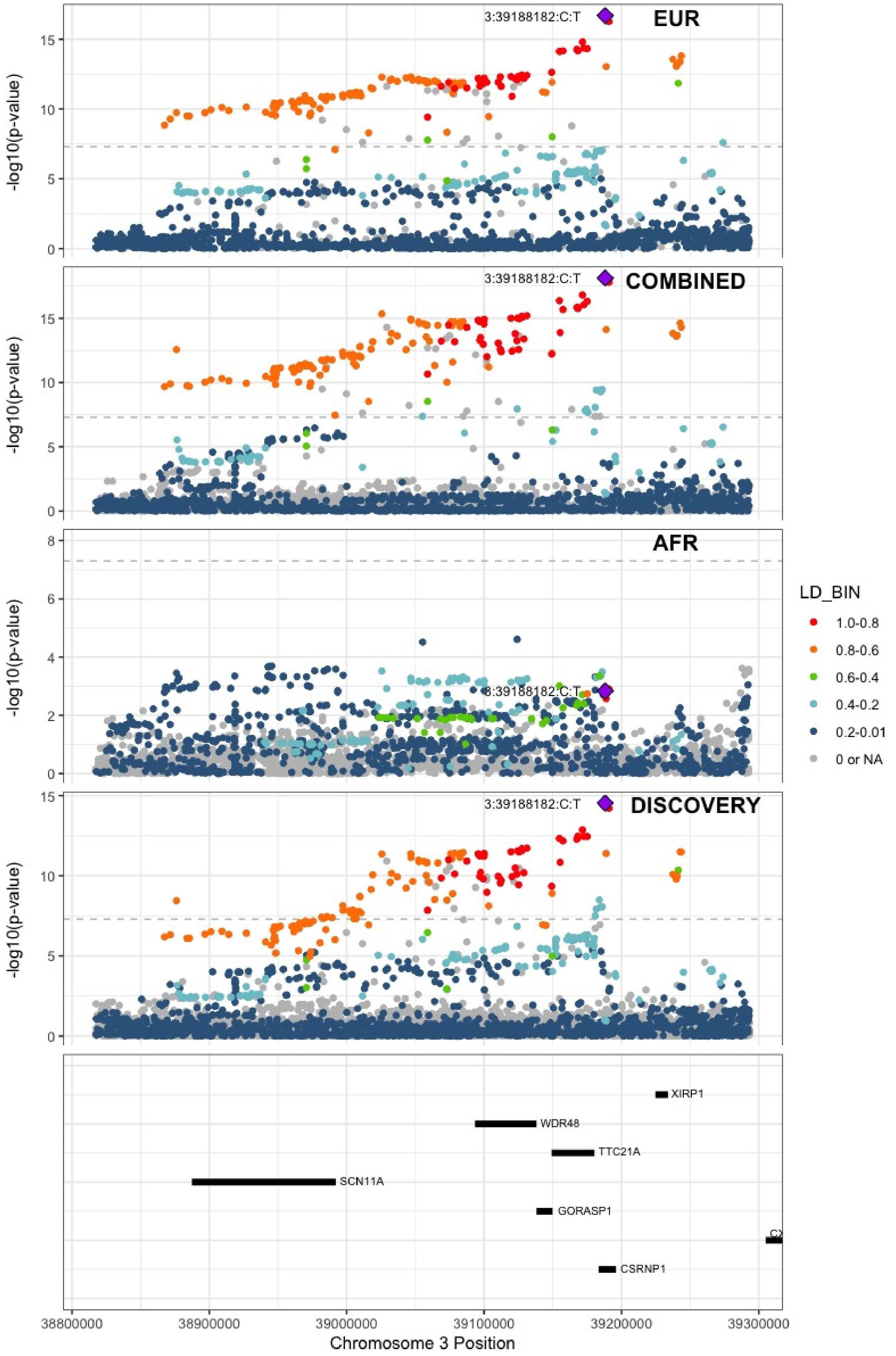

**Supplementary Figure 7.20** – rs11130326 (chr3:52771920) in locus 25 (NEK4)

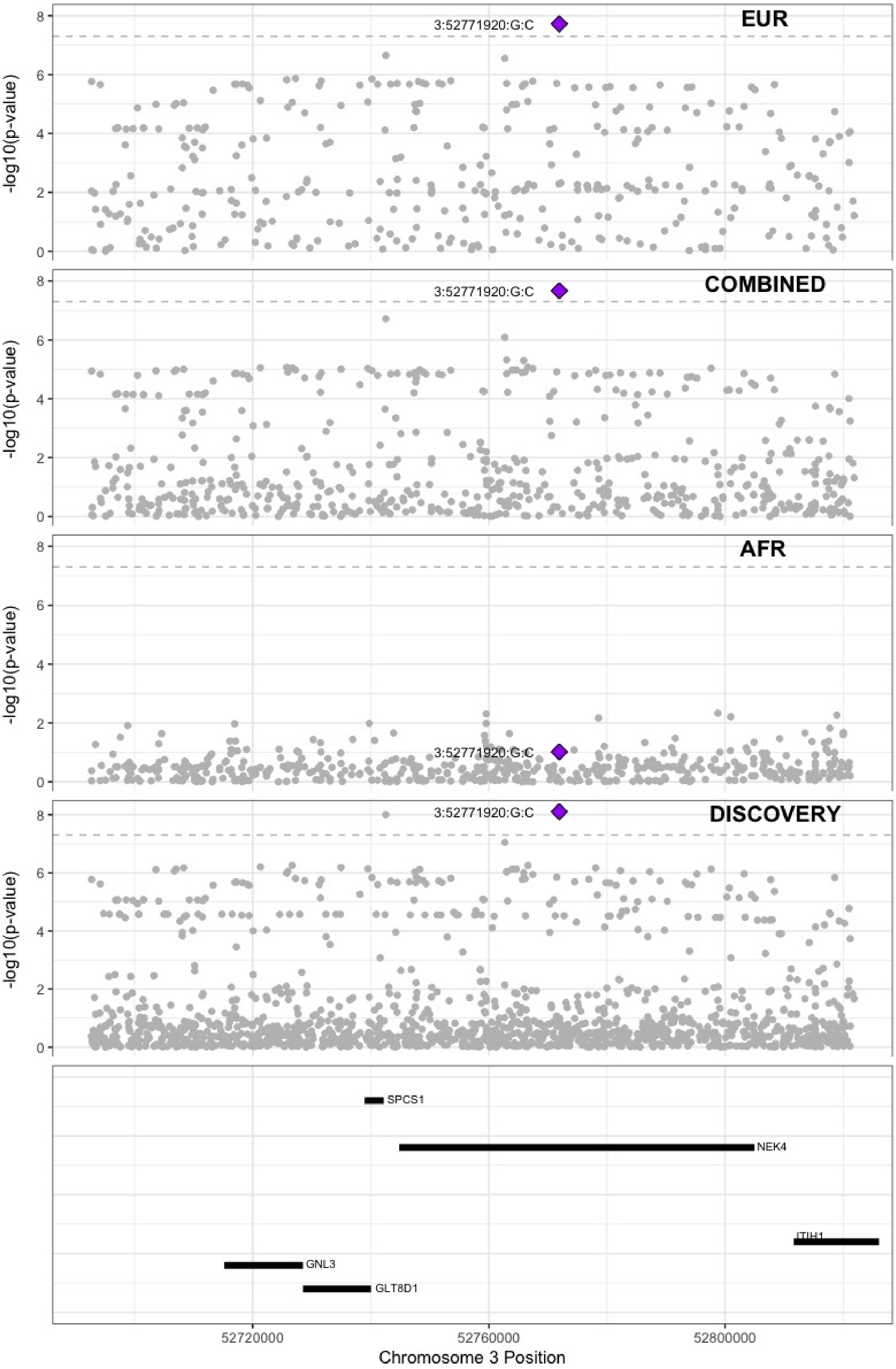

**Supplementary Figure 7.21** – rs9866664 (chr3:90437988) in locus 26 (EPHA3;NONE)

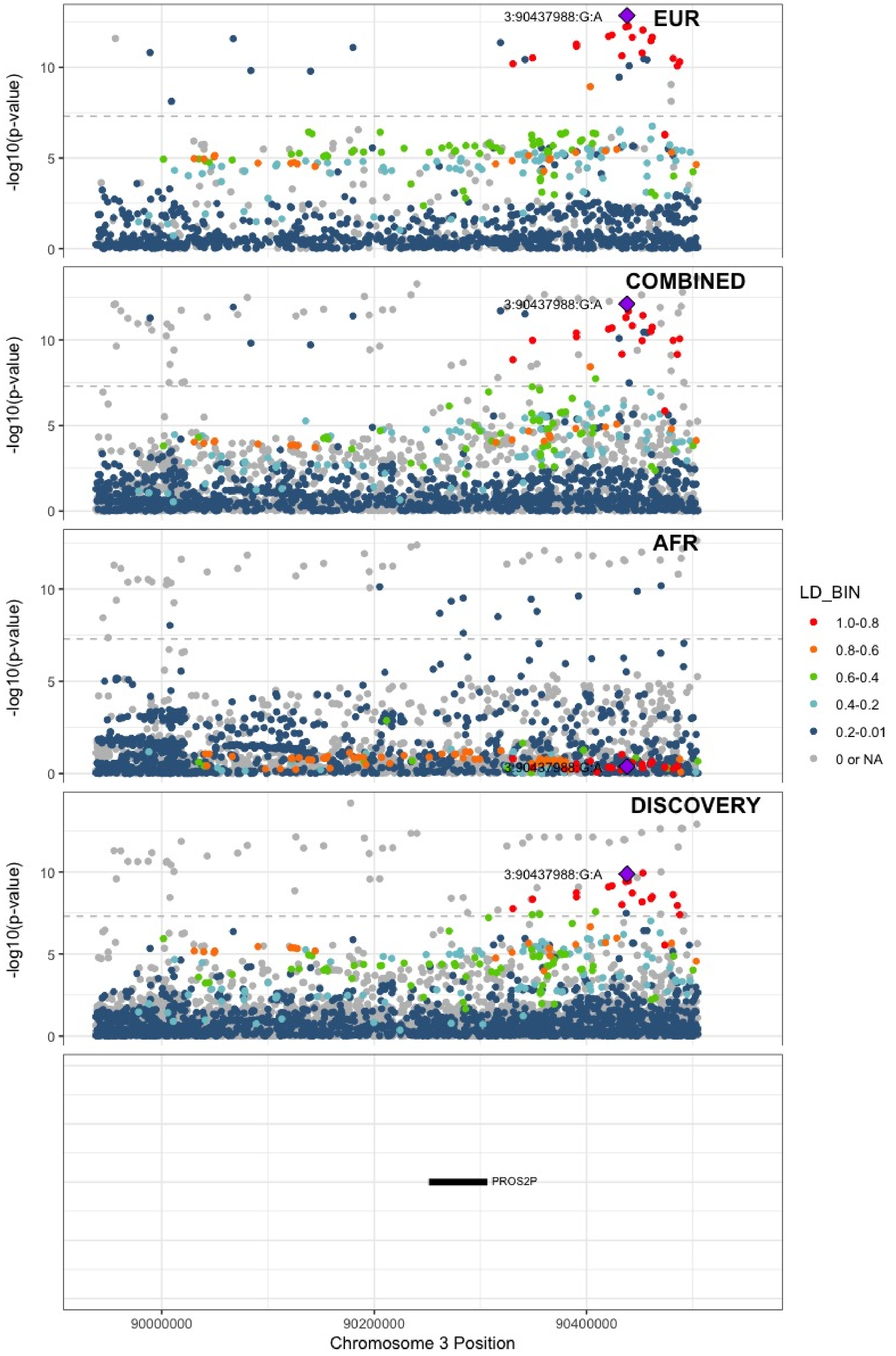

**Supplementary Figure 7.22** – rs28479320 (chr3:93752529) in locus 27 (ARL13B)

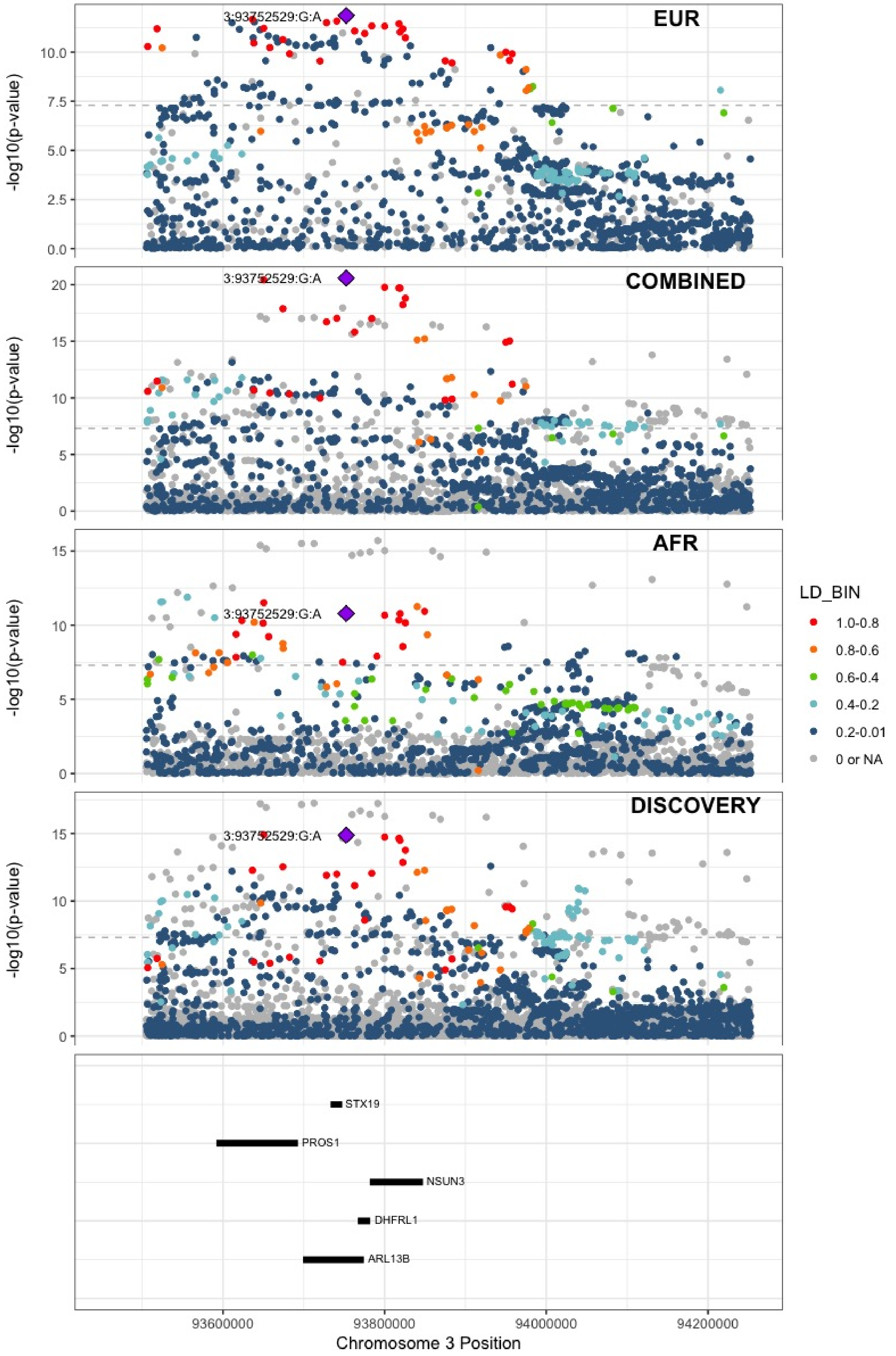

**Supplementary Figure 7.23** – rs9854955 (chr3:156795525) in locus 30 (LEKR1 ;LINC00880)

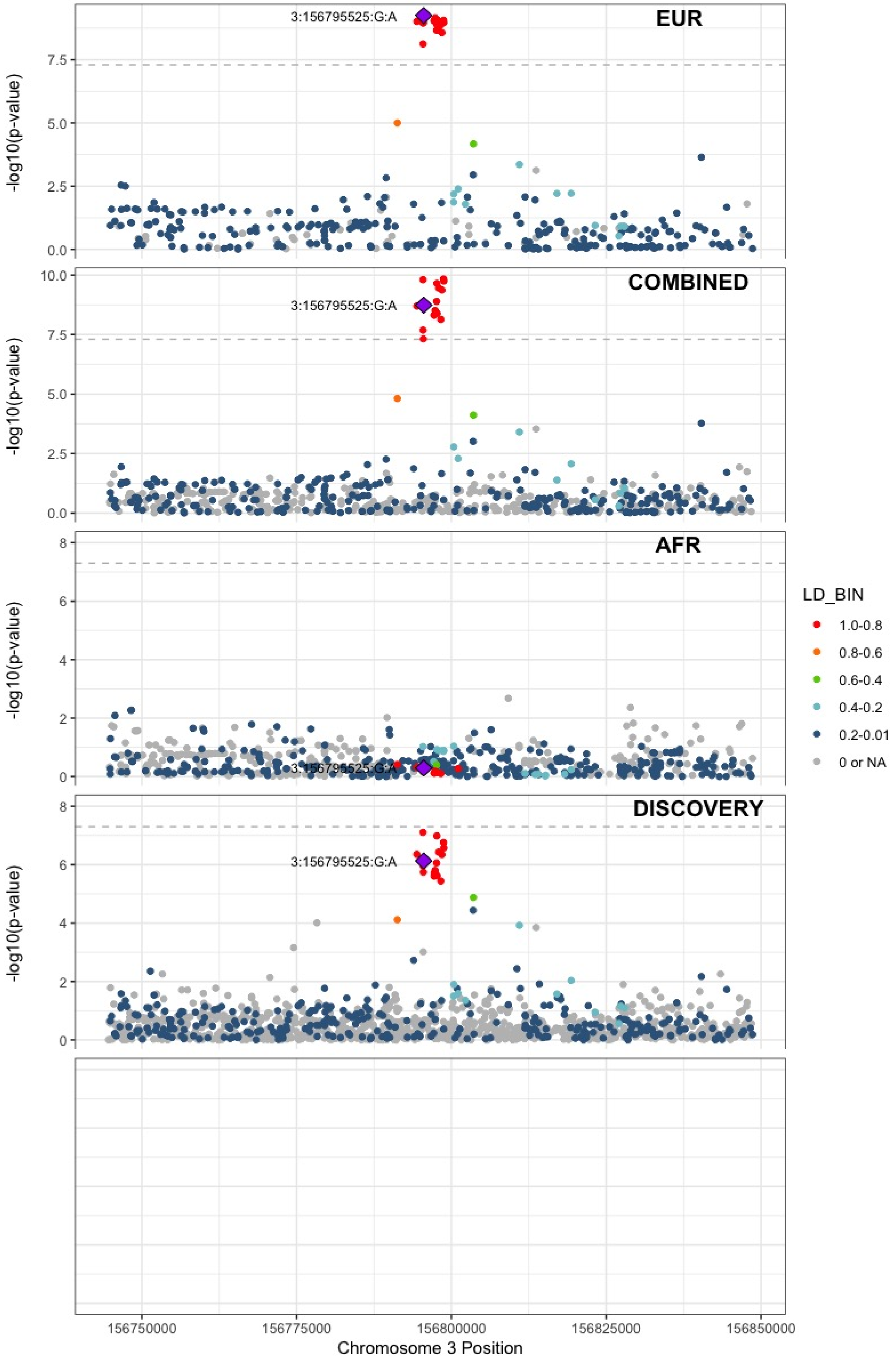

**Supplementary Figure 7.24** – rs710446 (chr3:186459927) in locus 32 (KNG1)

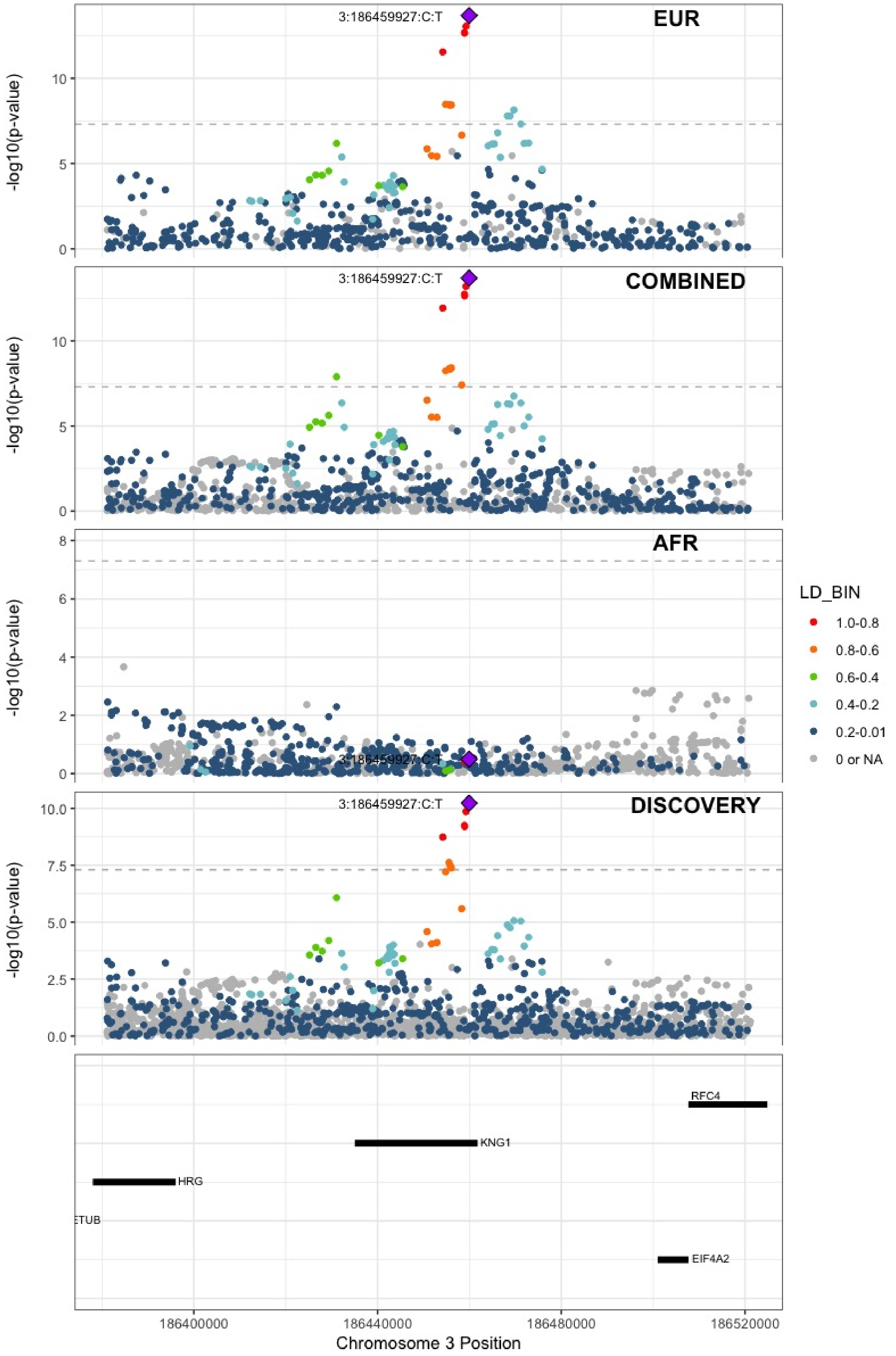

**Supplementary Figure 7.25** – rs3796159 (chr3:194790434) in locus 33 (XXYLT1)

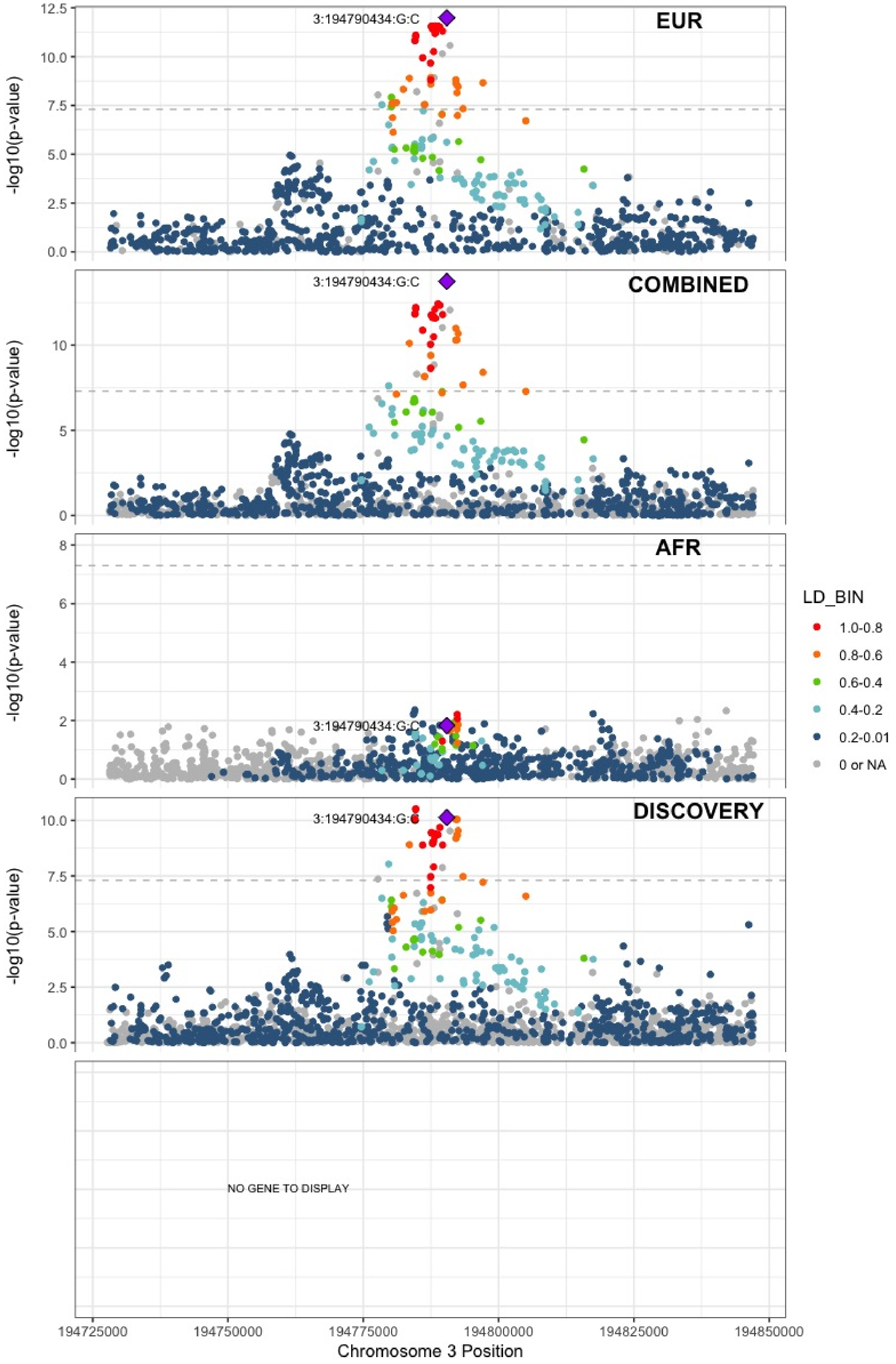

**Supplementary Figure 7.26** – rs72976751 (chr4:86725959) in locus 37 (ARHGAP24)

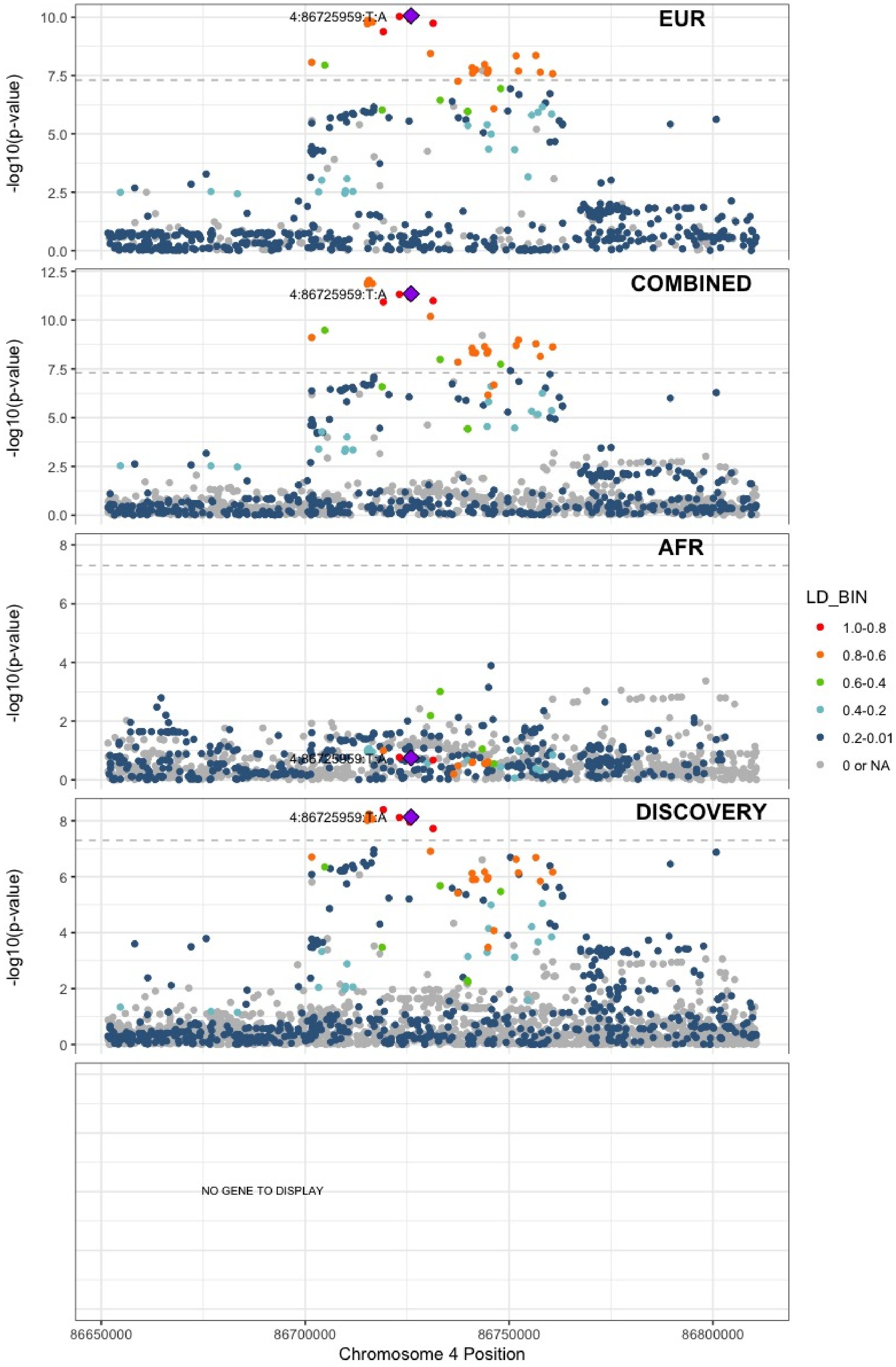

**Supplementary Figure 7.27** – rs35225200 (chr4:103146888) in locus 38 (BANK1;SLC39A8)

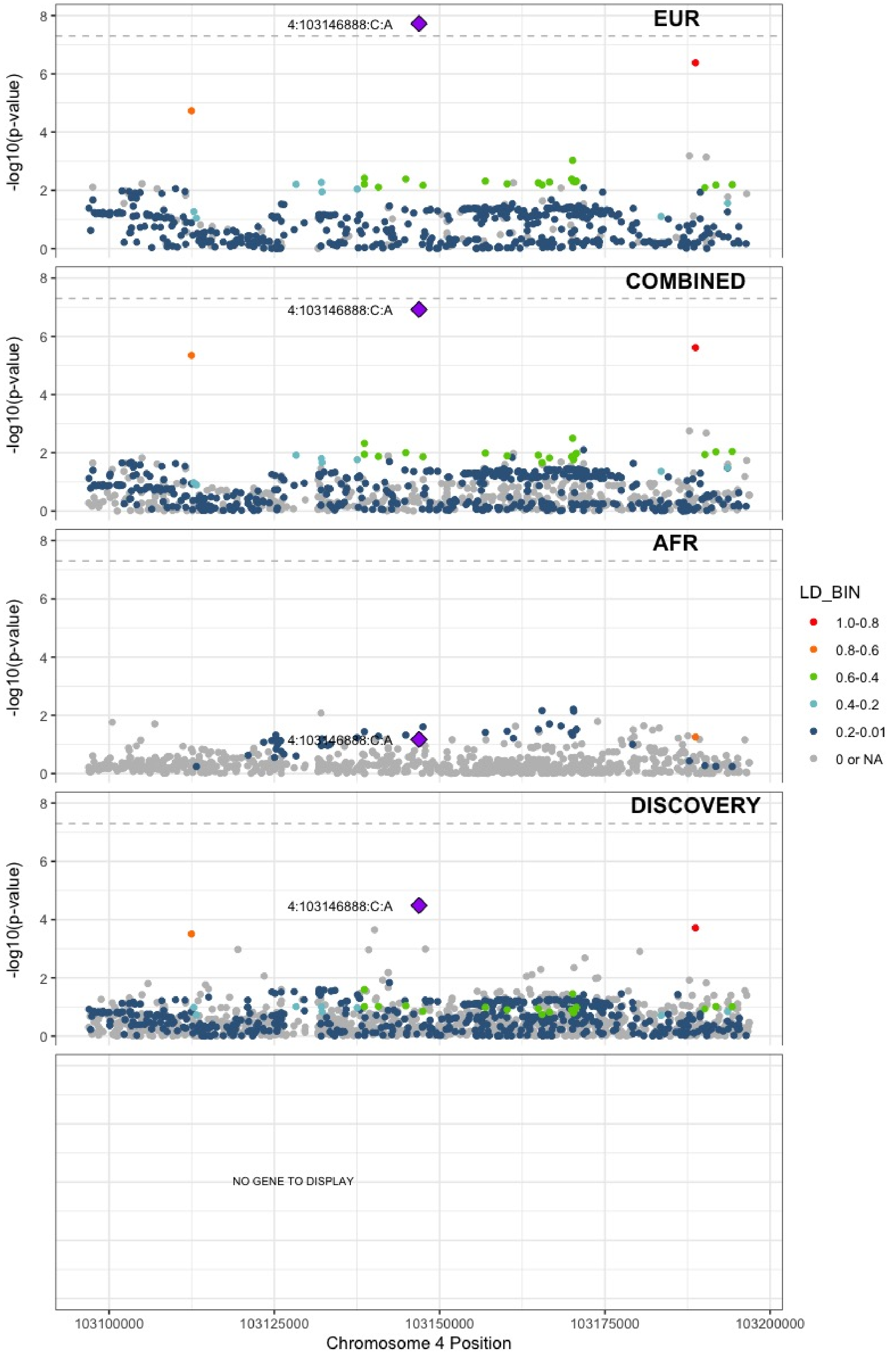

**Supplementary Figure 7.28** – rs2066864 (chr4:155525695) in locus 39 (FGG)

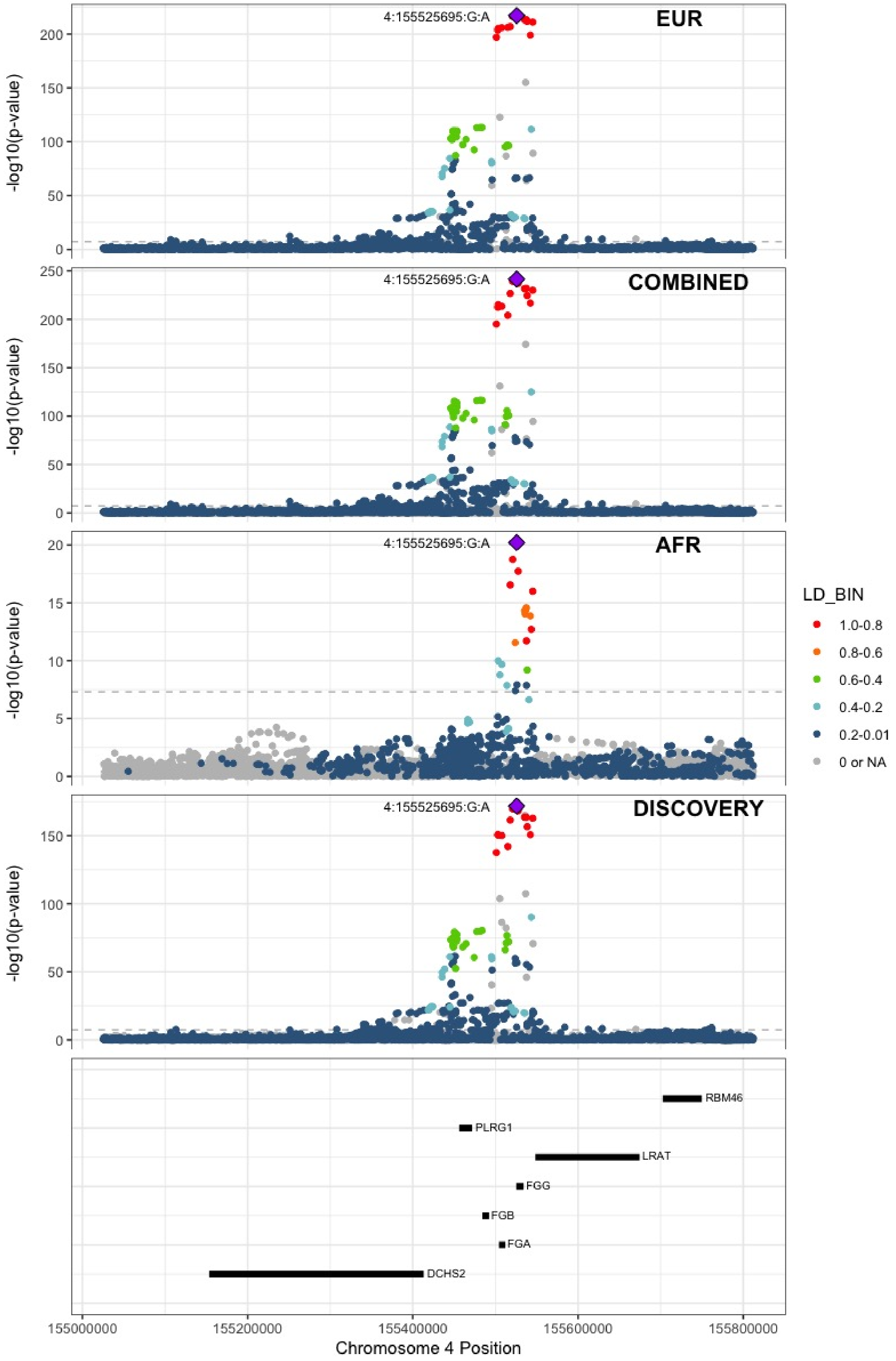

**Supplementary Figure 7.29** – rs3756011 (chr4:187206249) in locus 40 (F11)

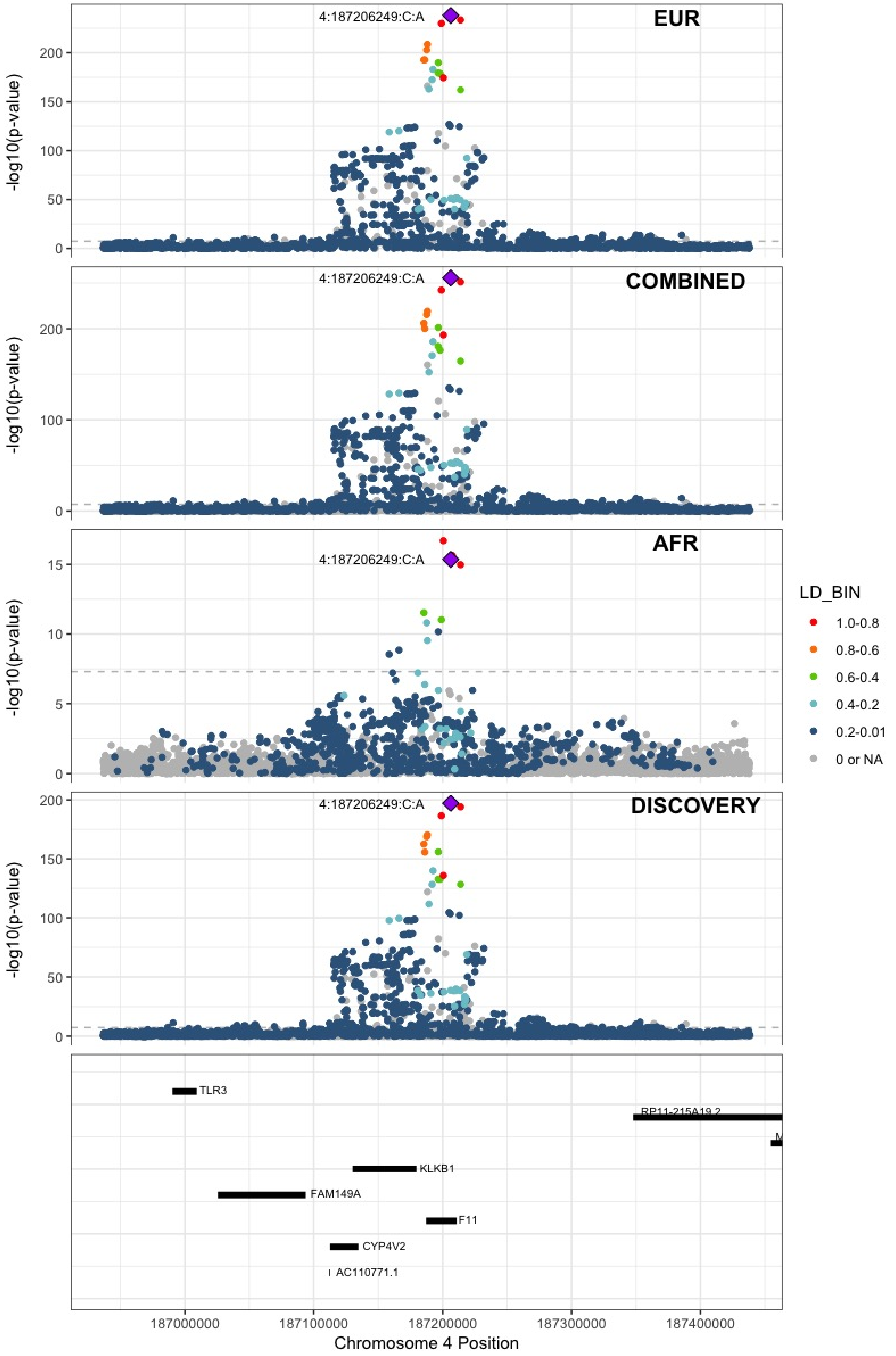

**Supplementary Figure 7.30** – rs112367053 (chr5:28379046) in locus 41 (LINC02103;LSP1P3)

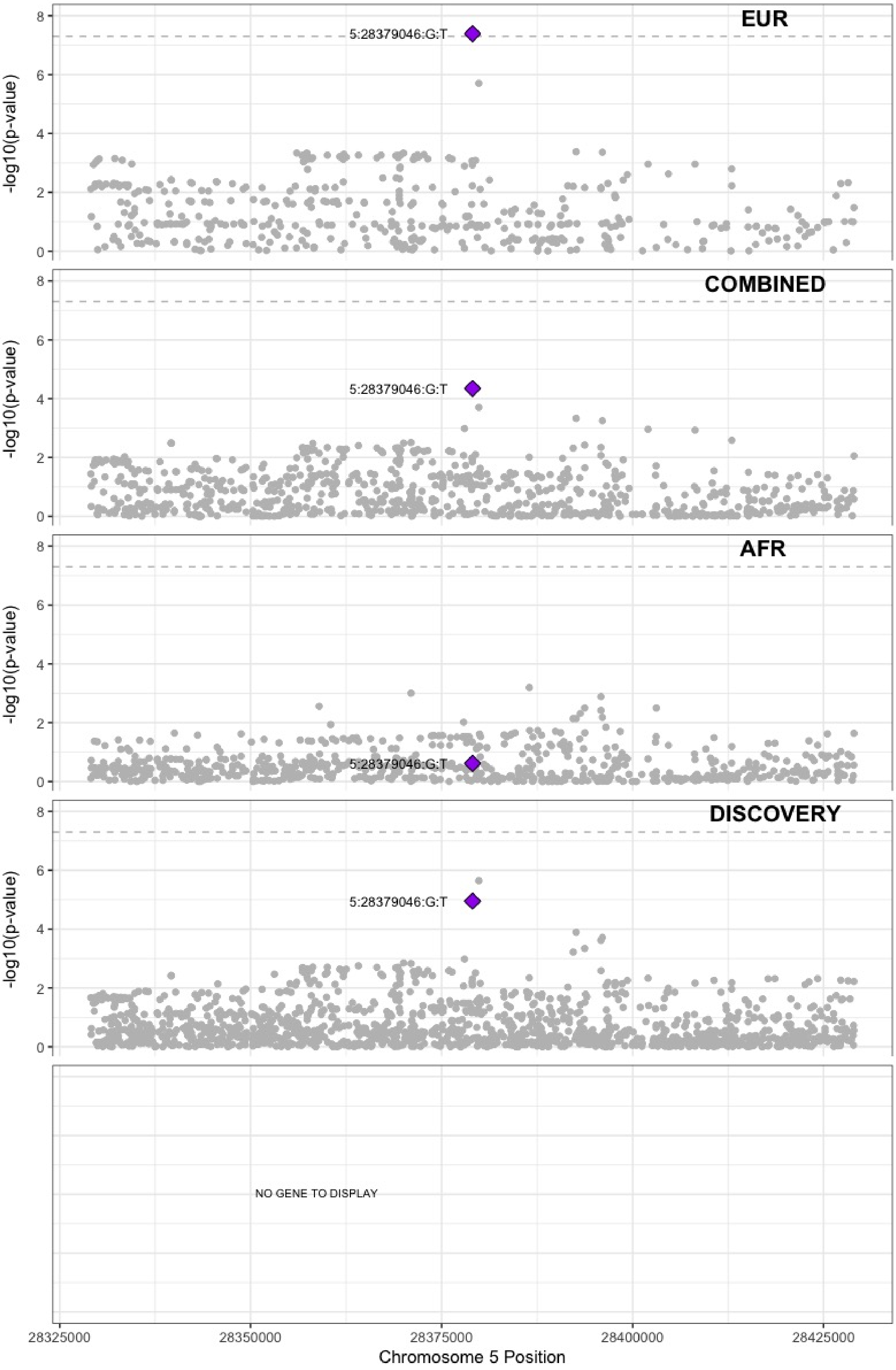

**Supplementary Figure 7.31** – rs16867574 (chr5:38708554) in locus 42 (OSMR-AS1)

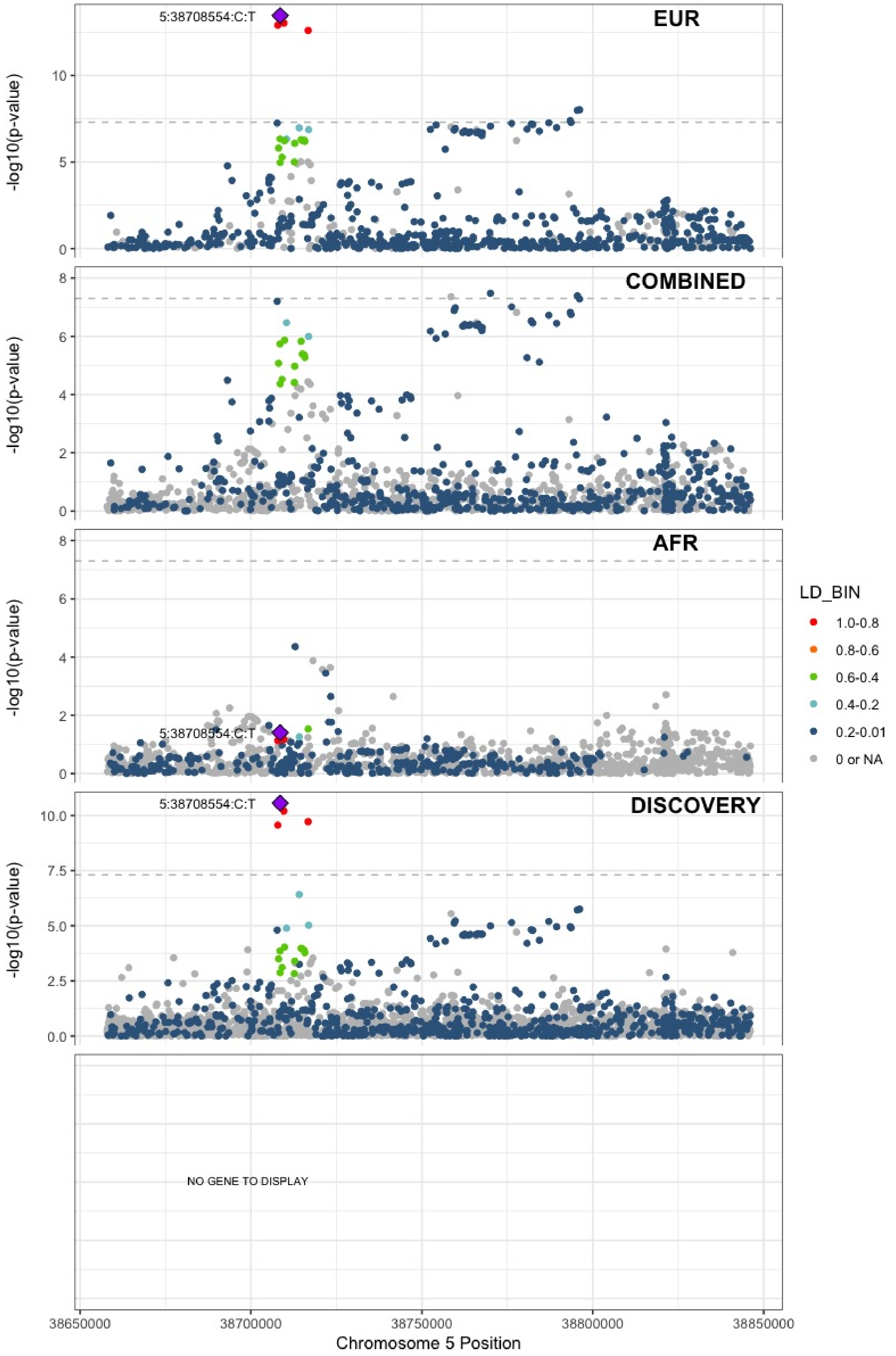

**Supplementary Figure 7.32** – rs4235677 (chr5:72925091) in locus 43 (ARHGEF28)

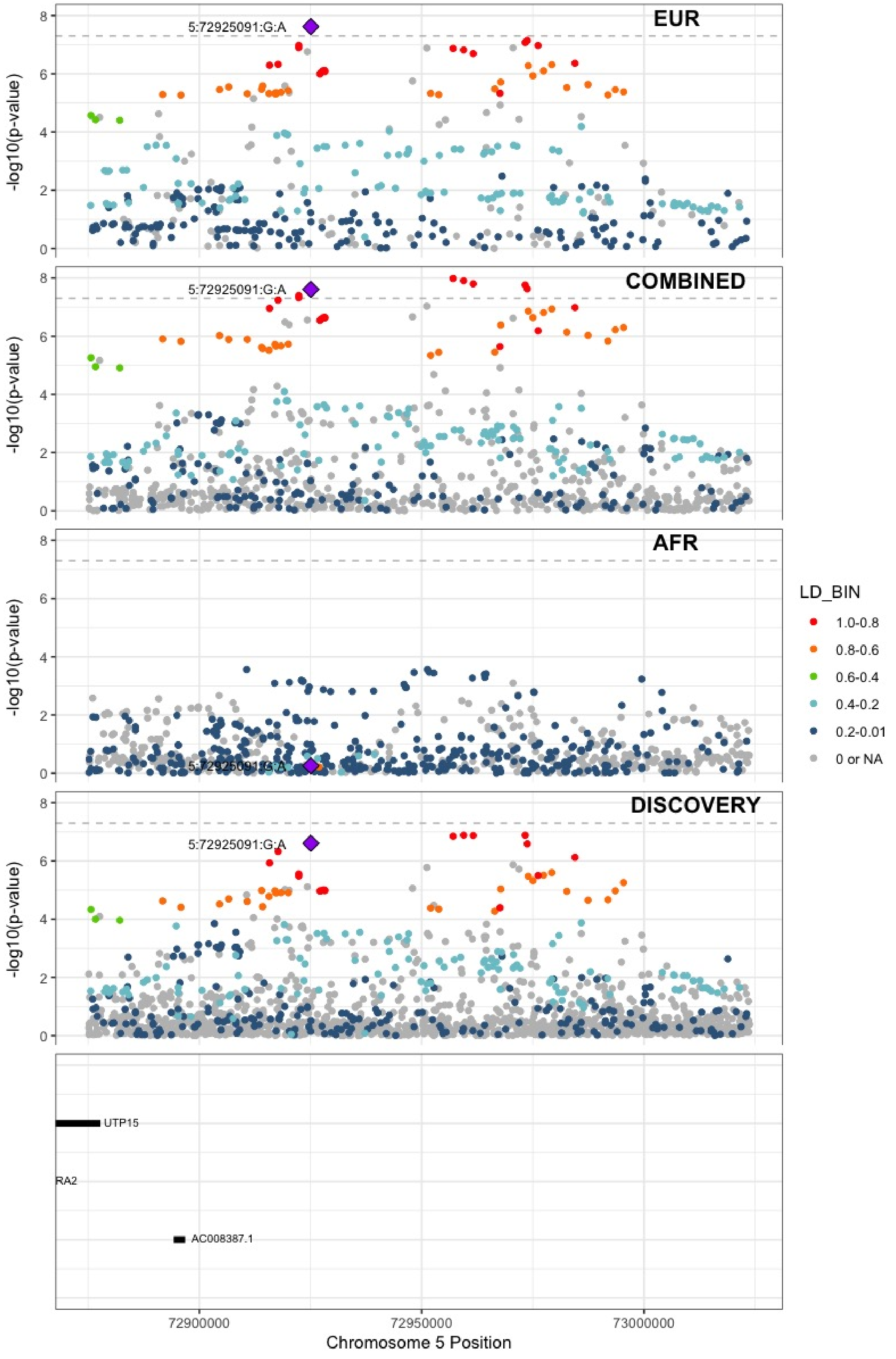

**Supplementary Figure 7.33** – rs251339 (chr5:96235038) in locus 44 (ERAP2)

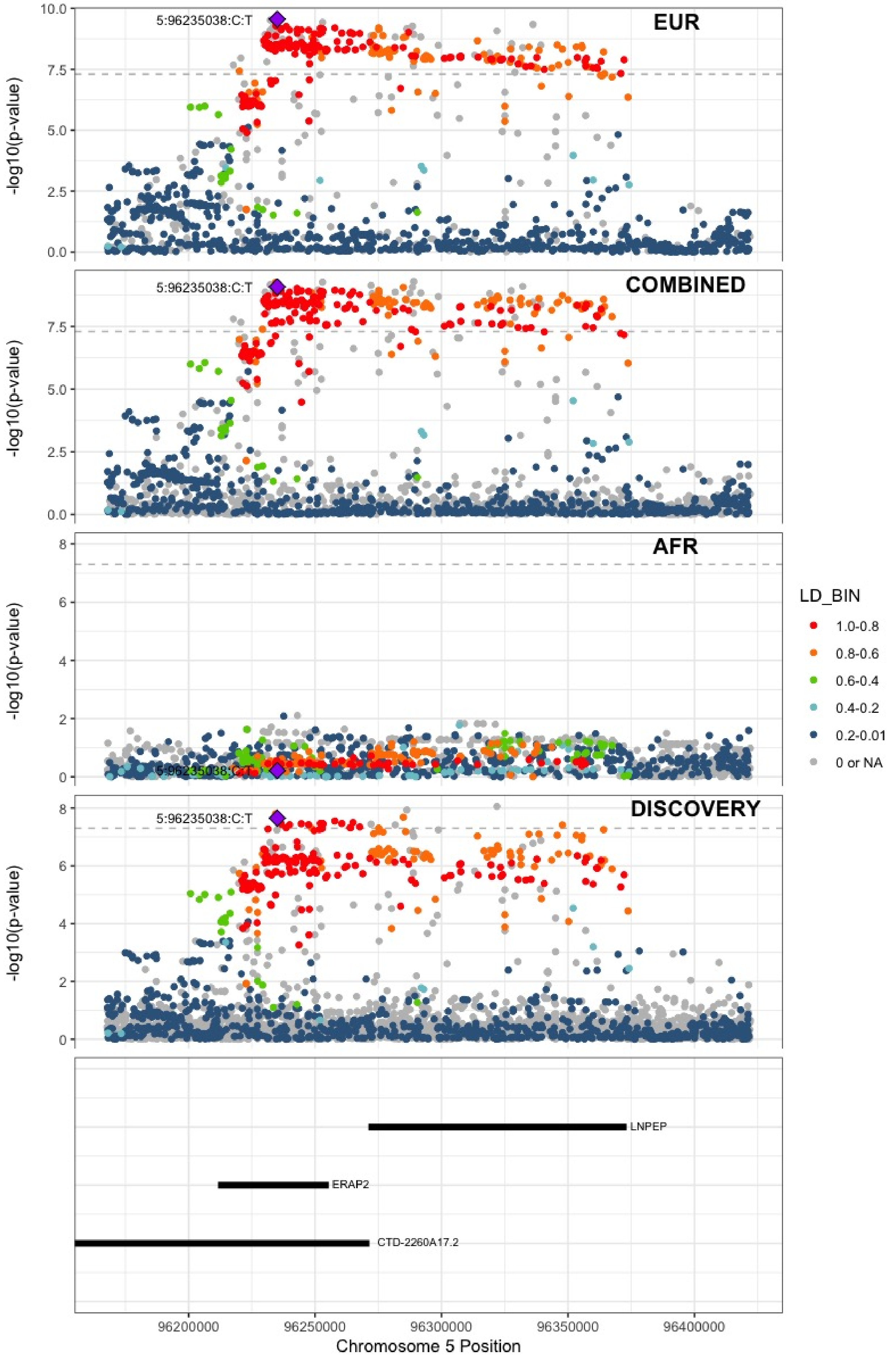

**Supplementary Figure 7.34** – rs147133967 (chr5:132426851) in locus 45 (HSPA4)

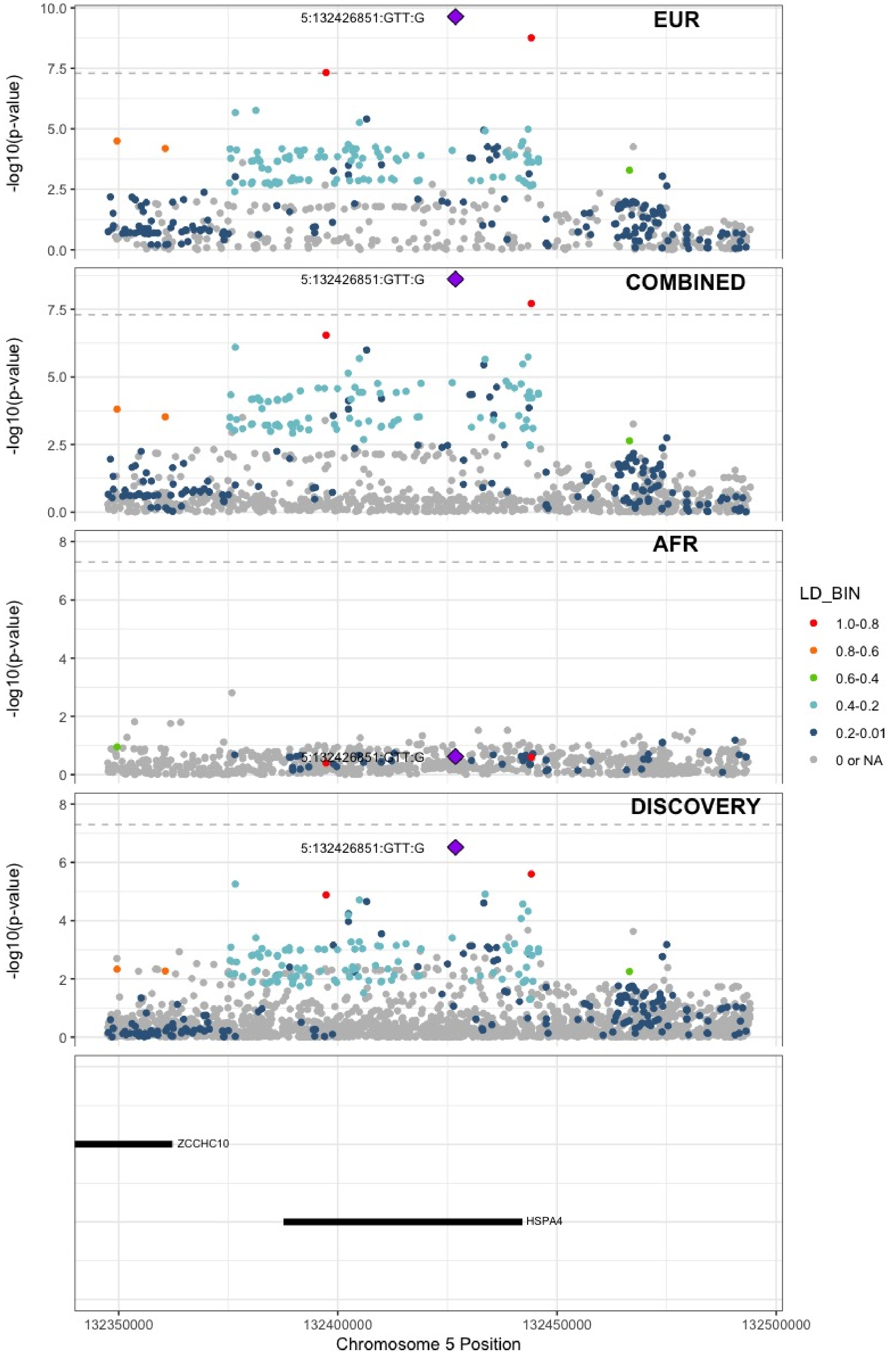

**Supplementary Figure 7.35** – rs214059 (chr6:25536937) in locus 46 (CARMIL1)

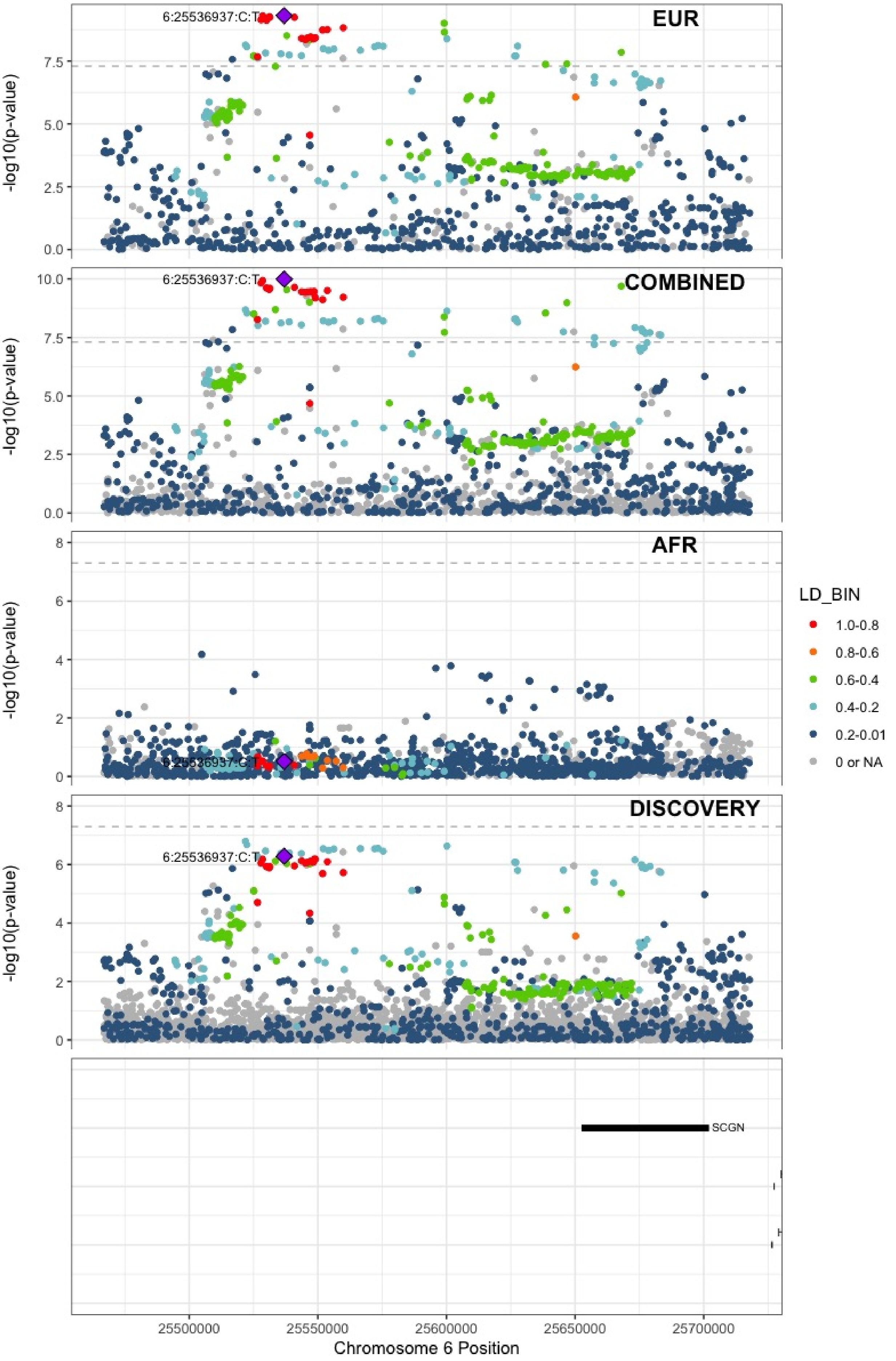

**Supplementary Figure 7.36** – rs2394251 (chr6:29943688) in locus 47 (HCG9)

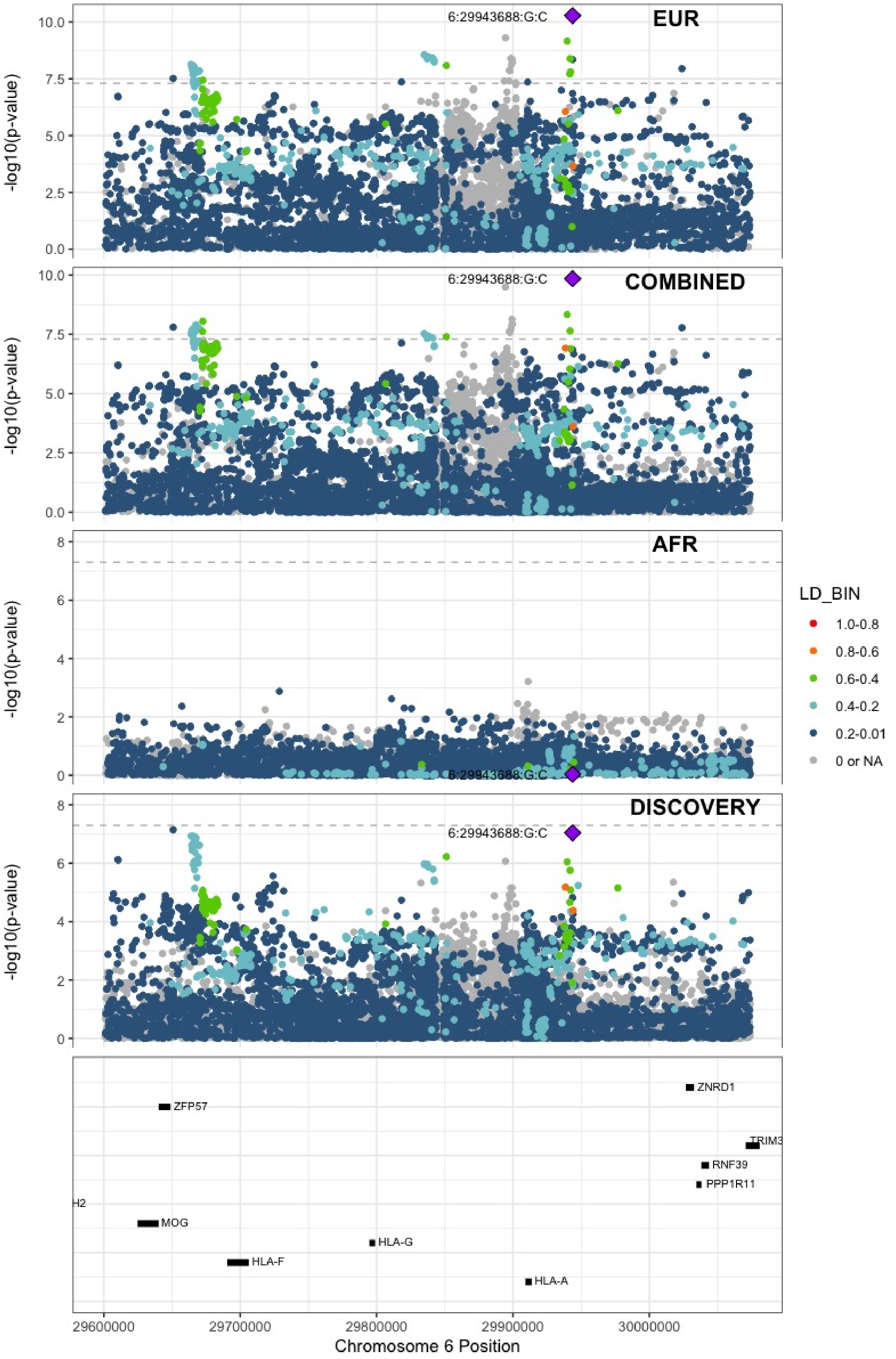

**Supplementary Figure 7.37** – rs28383307 (chr6:32586755) in locus 48 (HLA-DRB1 ;HLA-DQA1)

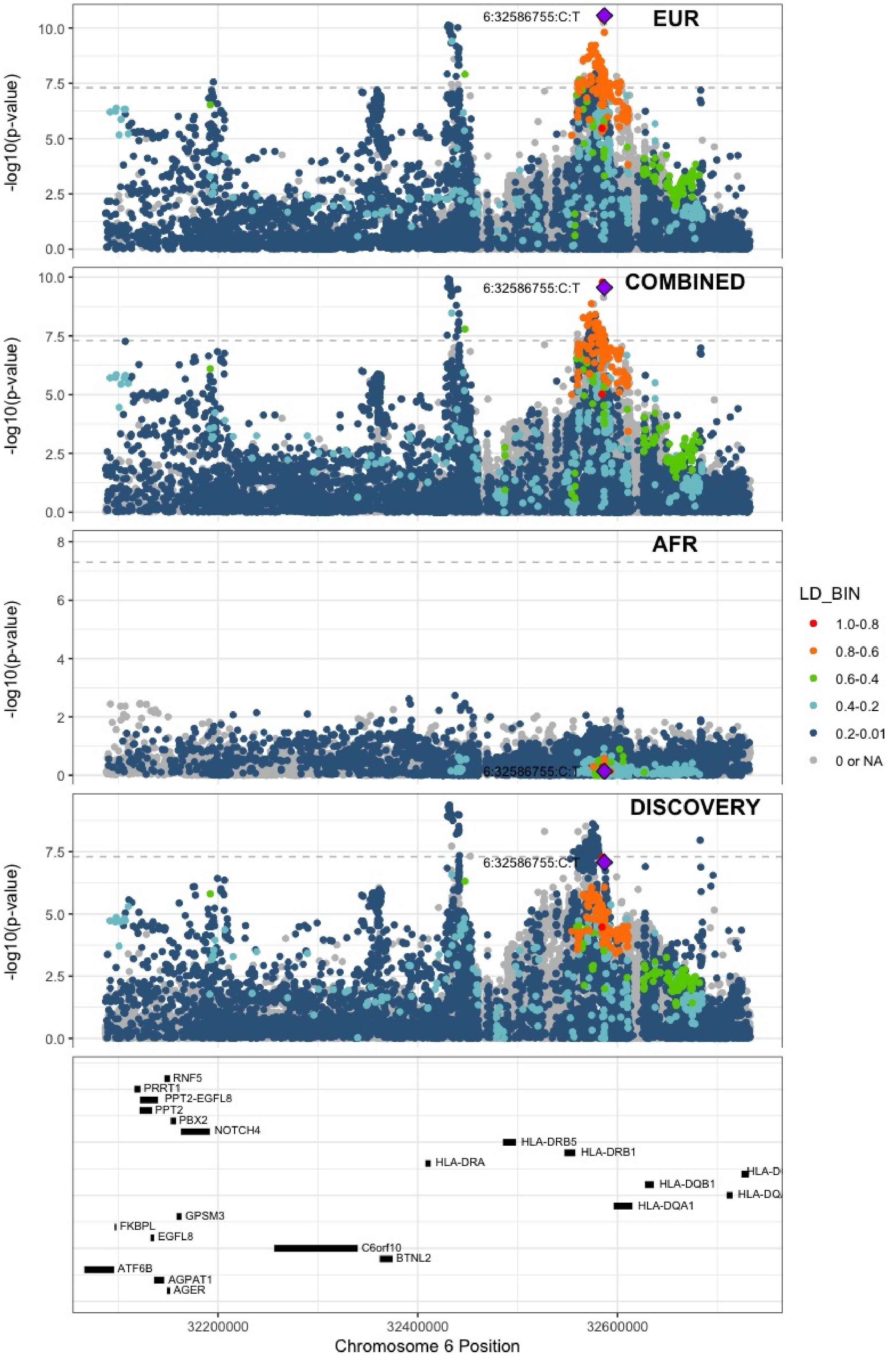

**Supplementary Figure 7.38** – rs145294670 (chr6:34622561) in locus 49 (ILRUN)

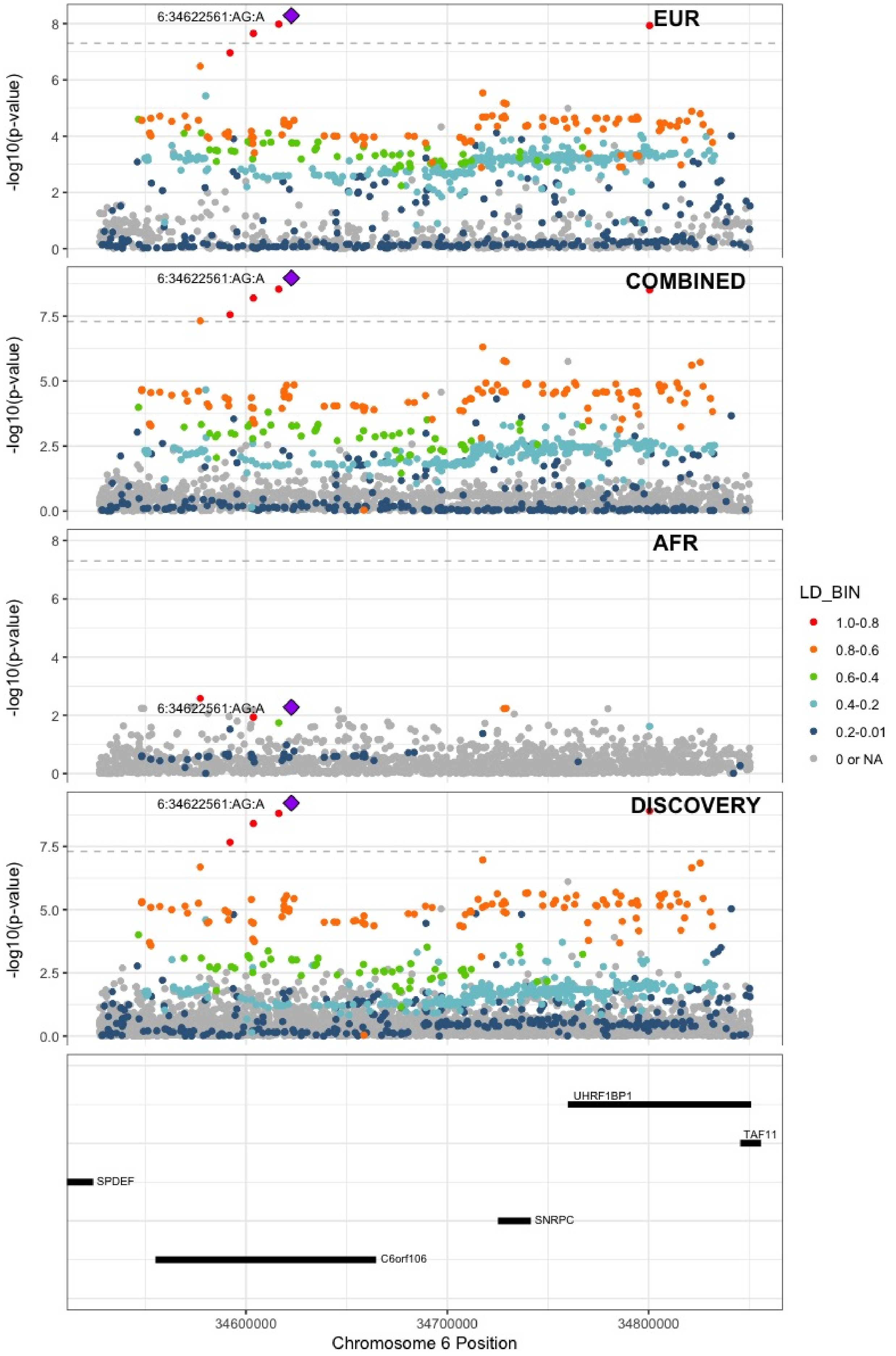

**Supplementary Figure 7.39** – rs2754251 (chr6:88385949) in locus 51 (AKIRIN2)

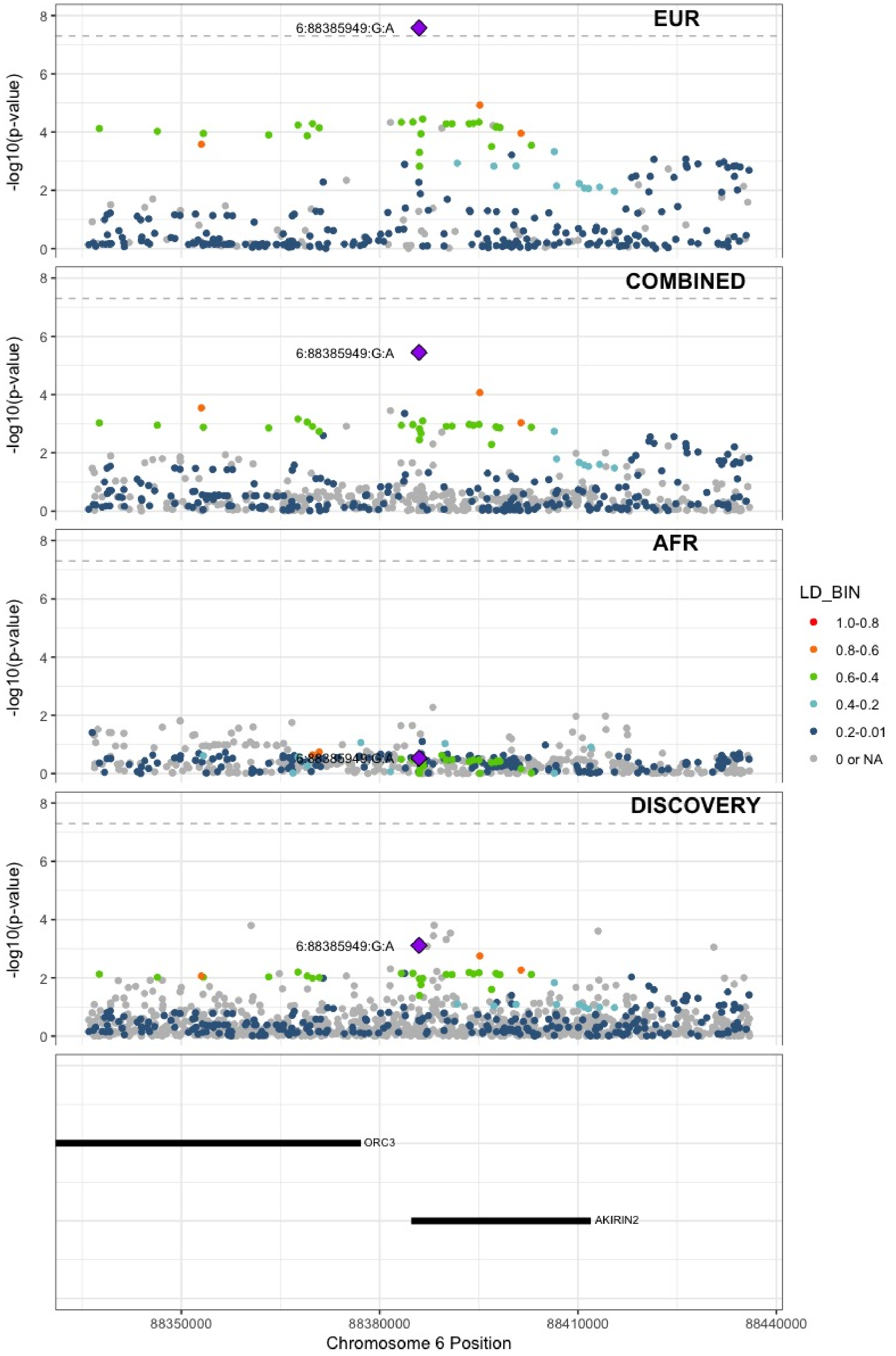

**Supplementary Figure 7.40** – rs9390460 (chr6:147694334) in locus 52 (STXBP5)

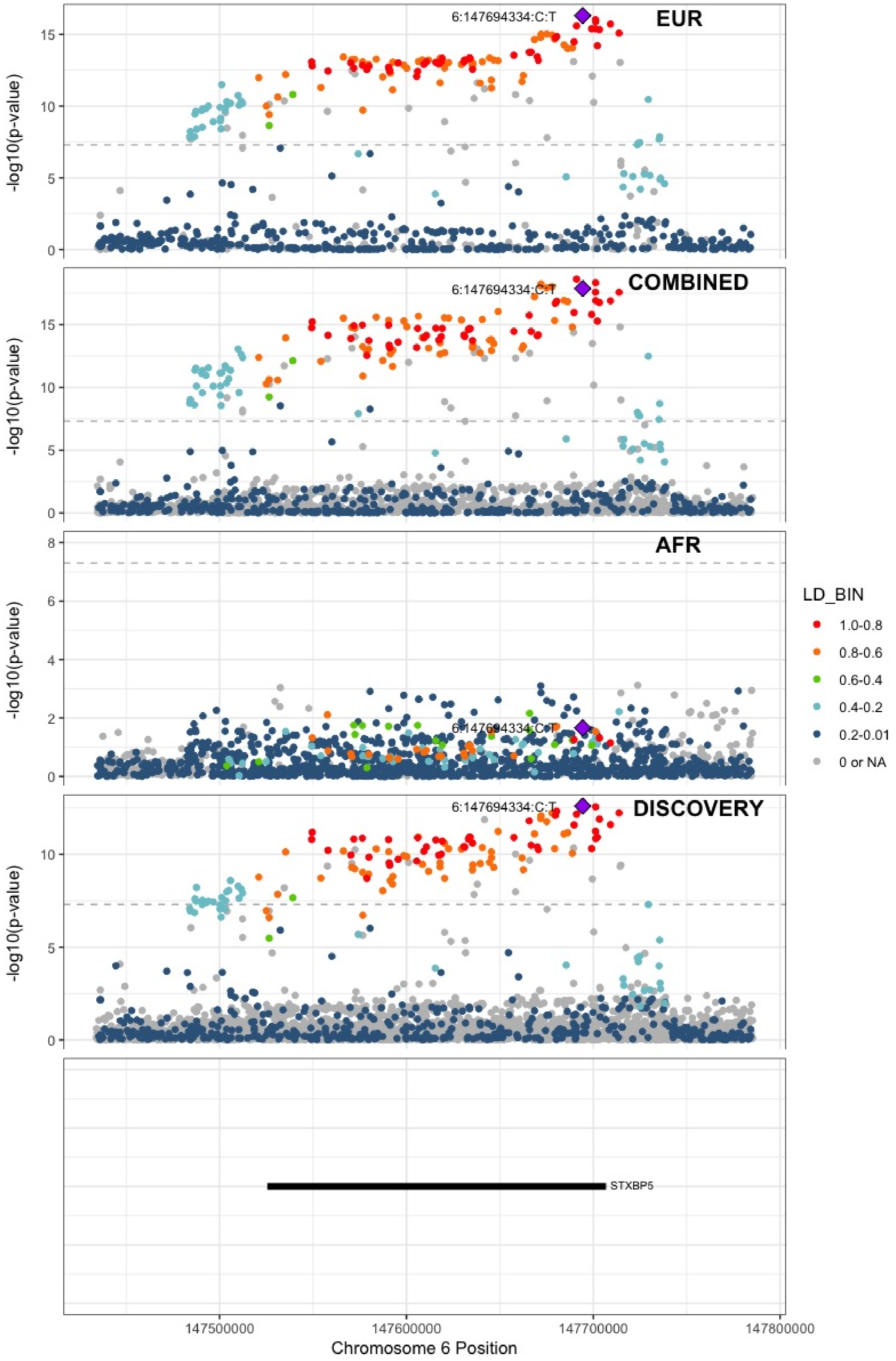

**Supplementary Figure 7.41** – rs1513275 (chr7:28259233) in locus 53 (JAZF1-AS1)

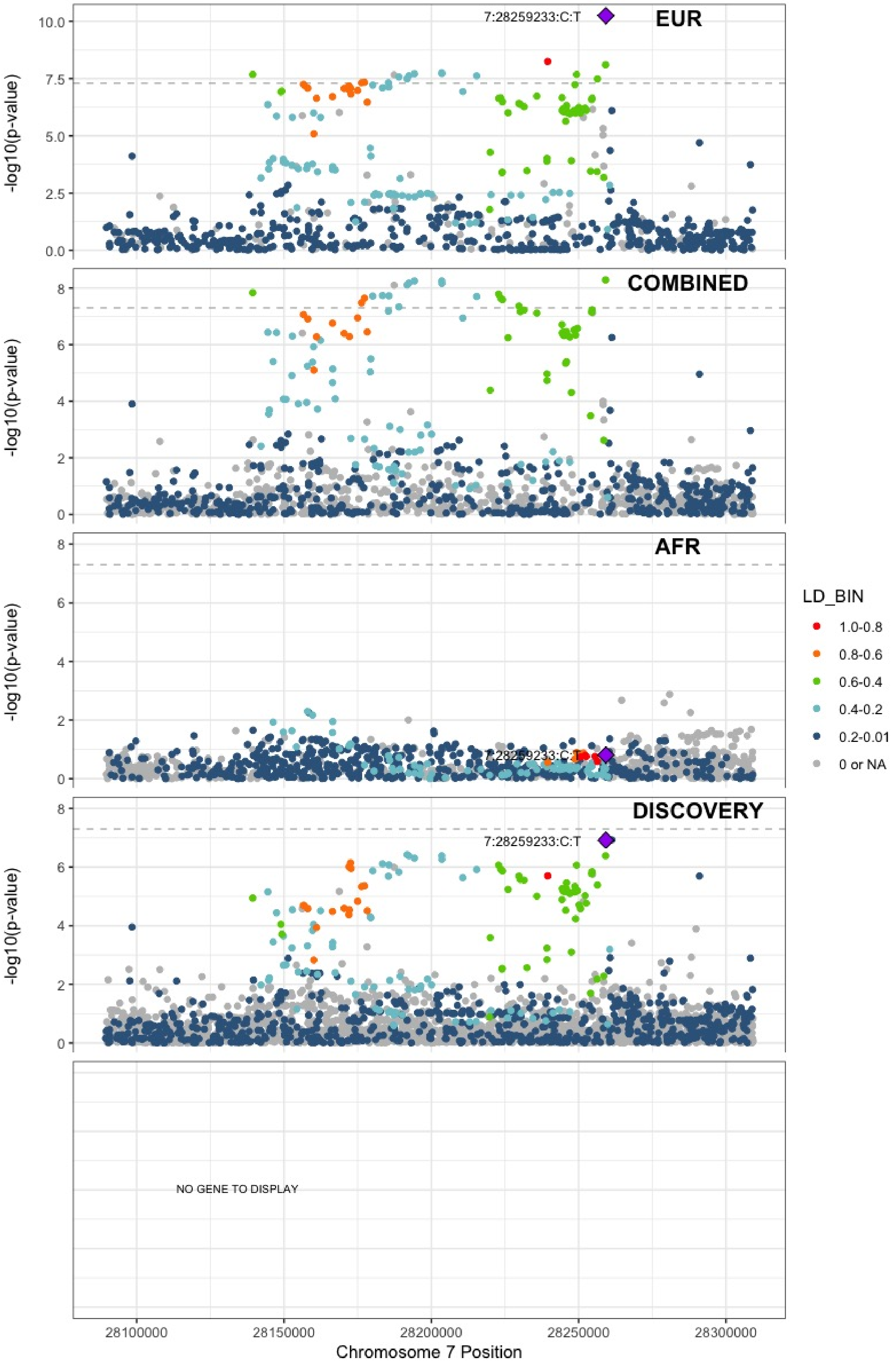

**Supplementary Figure 7.42** – rs370720 (chr8:9168146) in locus 57 (LOC101929128 ;LOC157273)

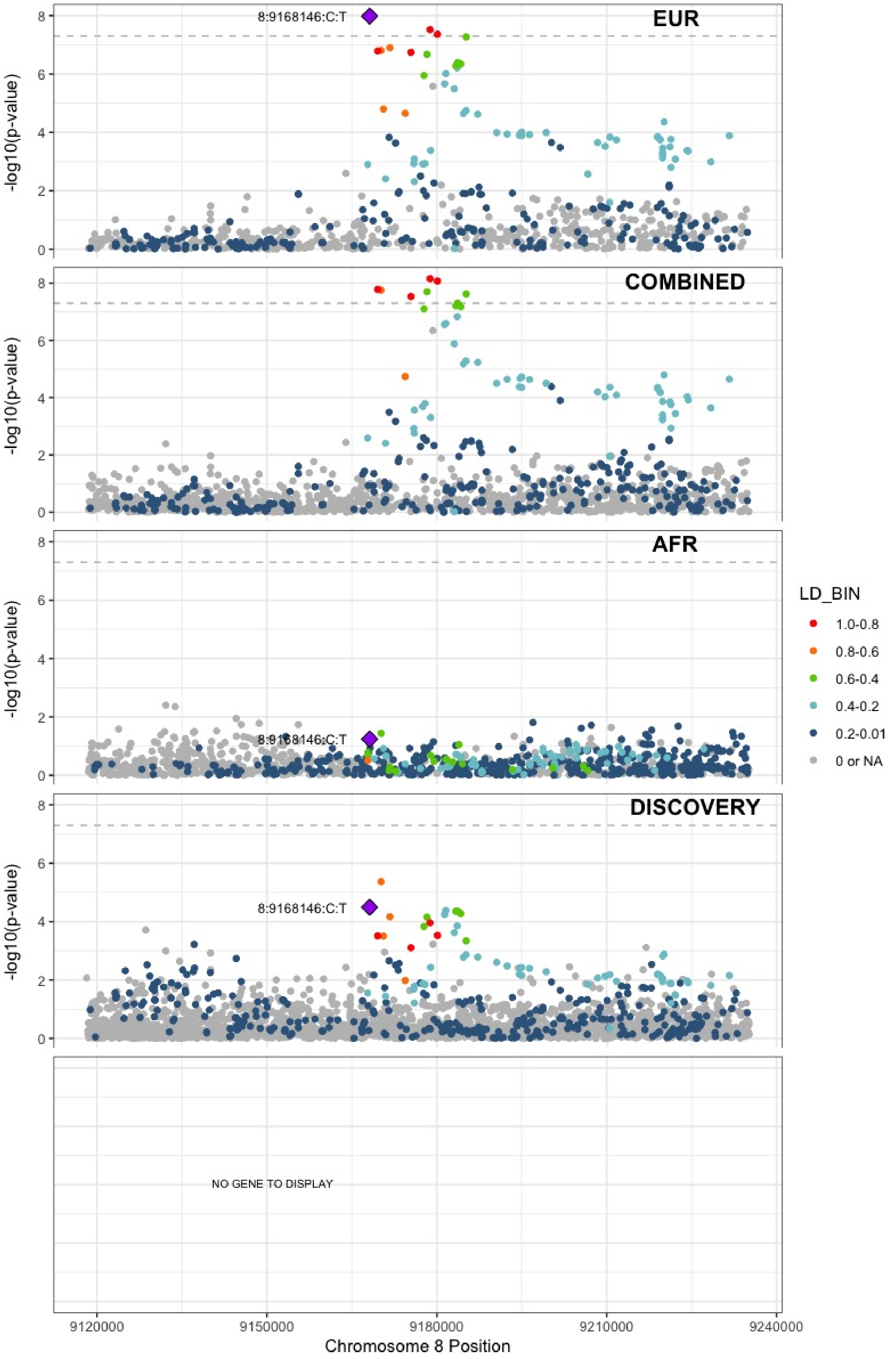

**Supplementary Figure 7.43** – rs2726950 (chr8:27806764) in locus 60 (SCARA5)

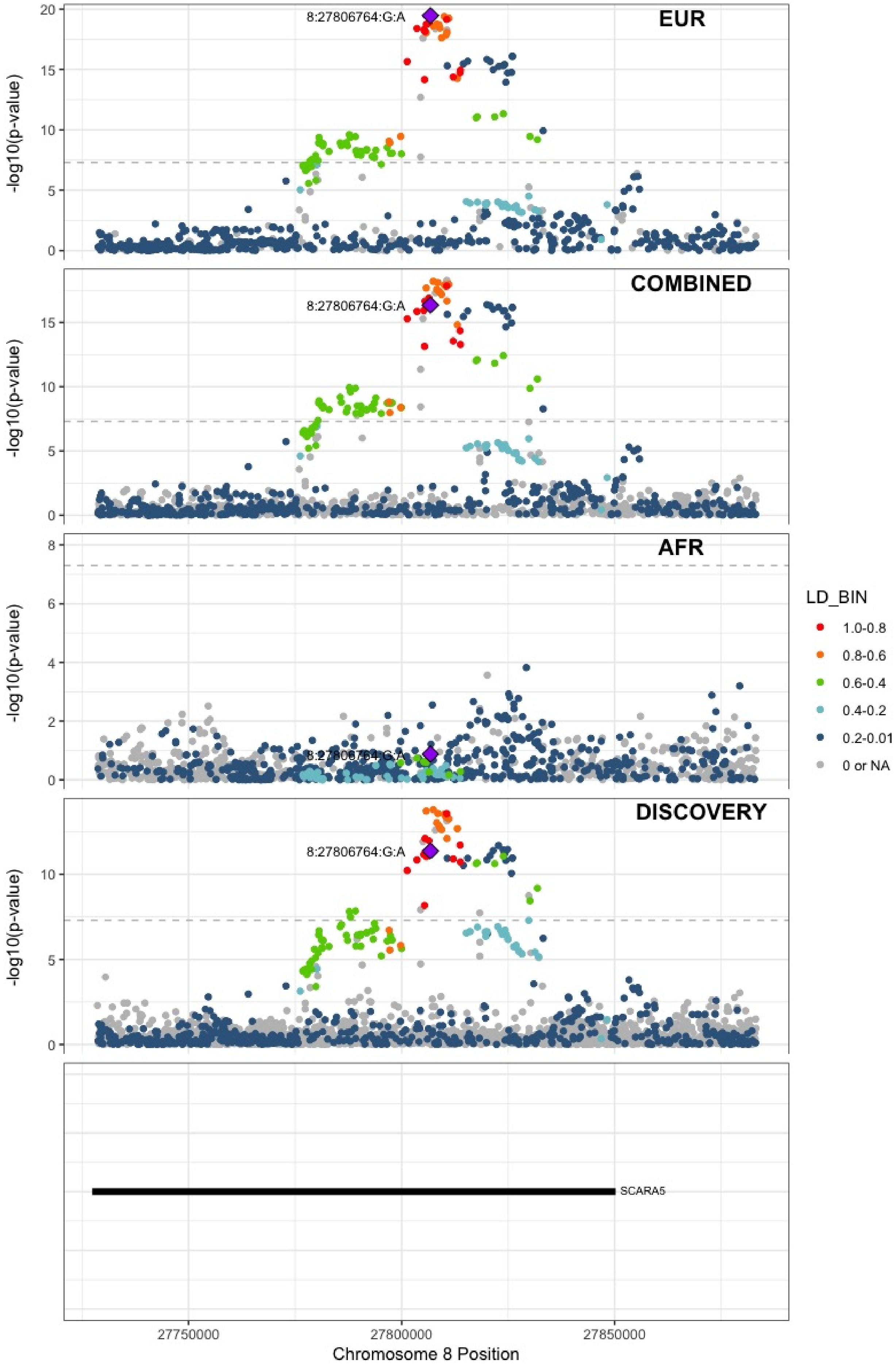

**Supplementary Figure 7.44** – rs6993770 (chr8:106581528) in locus 62 (ZFPM2)

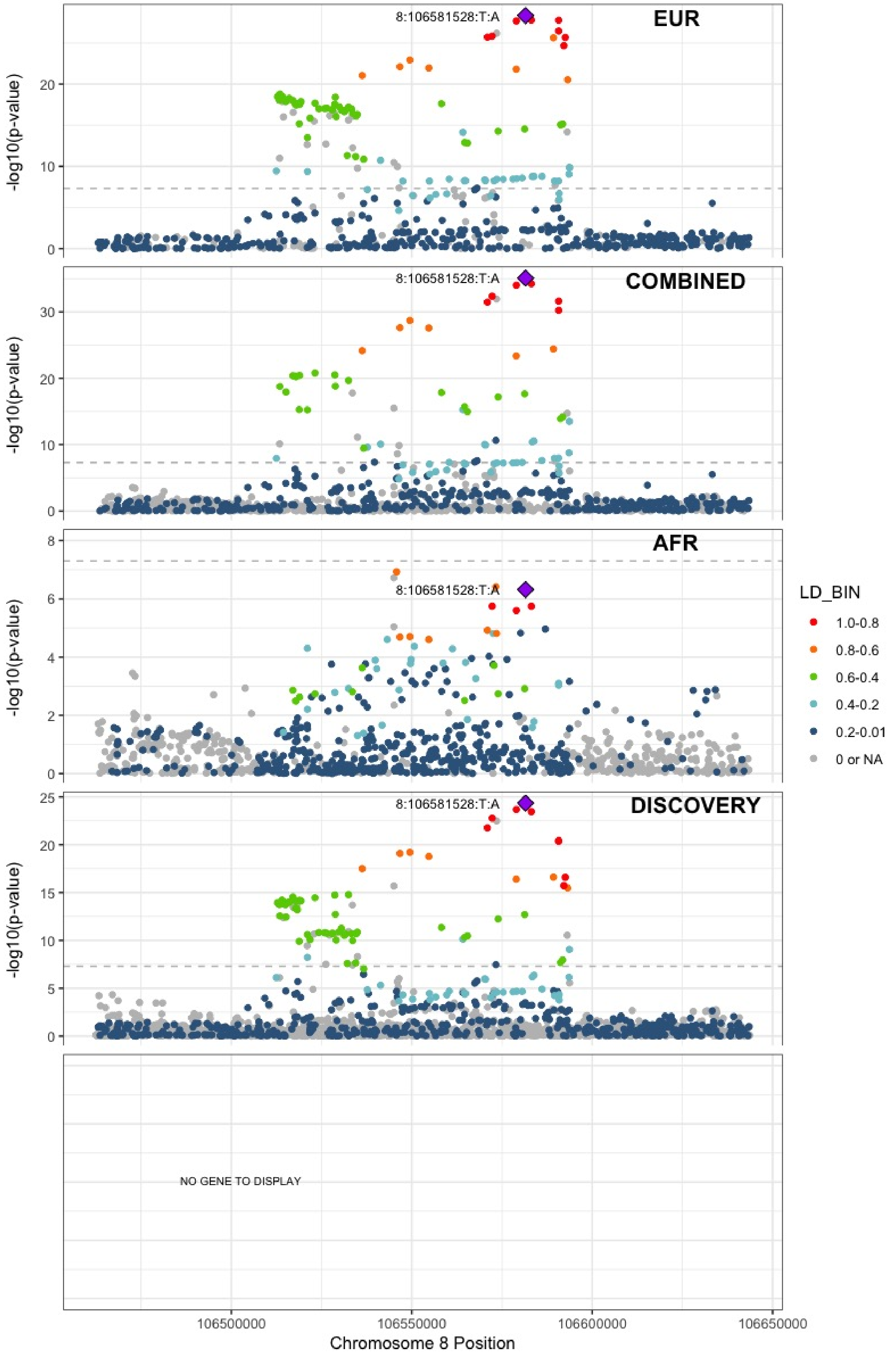

**Supplementary Figure 7.45** – rs1491336705 (chr8:108295235) in locus 63 (ANGPT1)

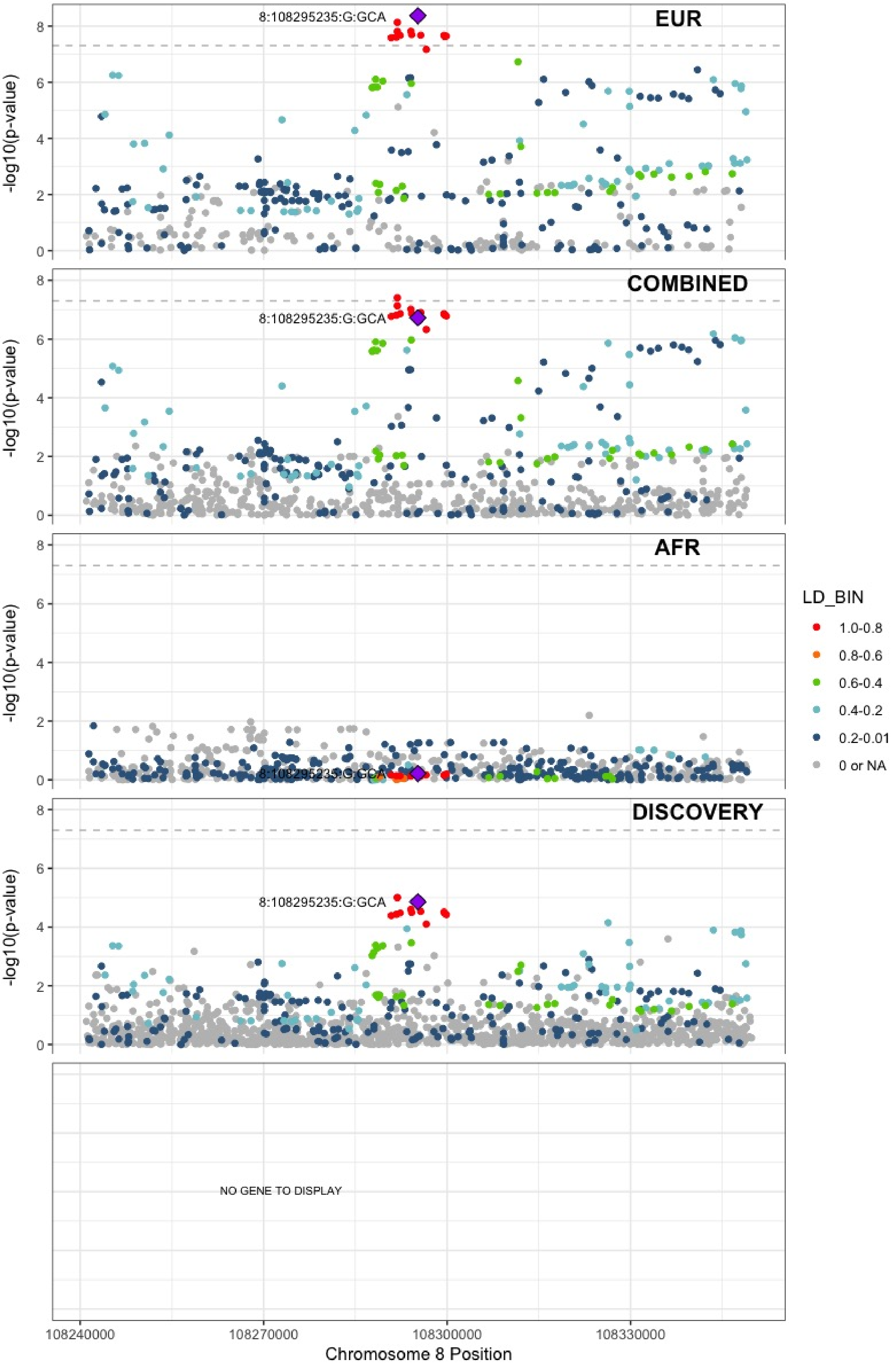

**Supplementary Figure 7.46** – rs10820606 (chr9:99192919) in locus 65 (ZNF367;HABP4)

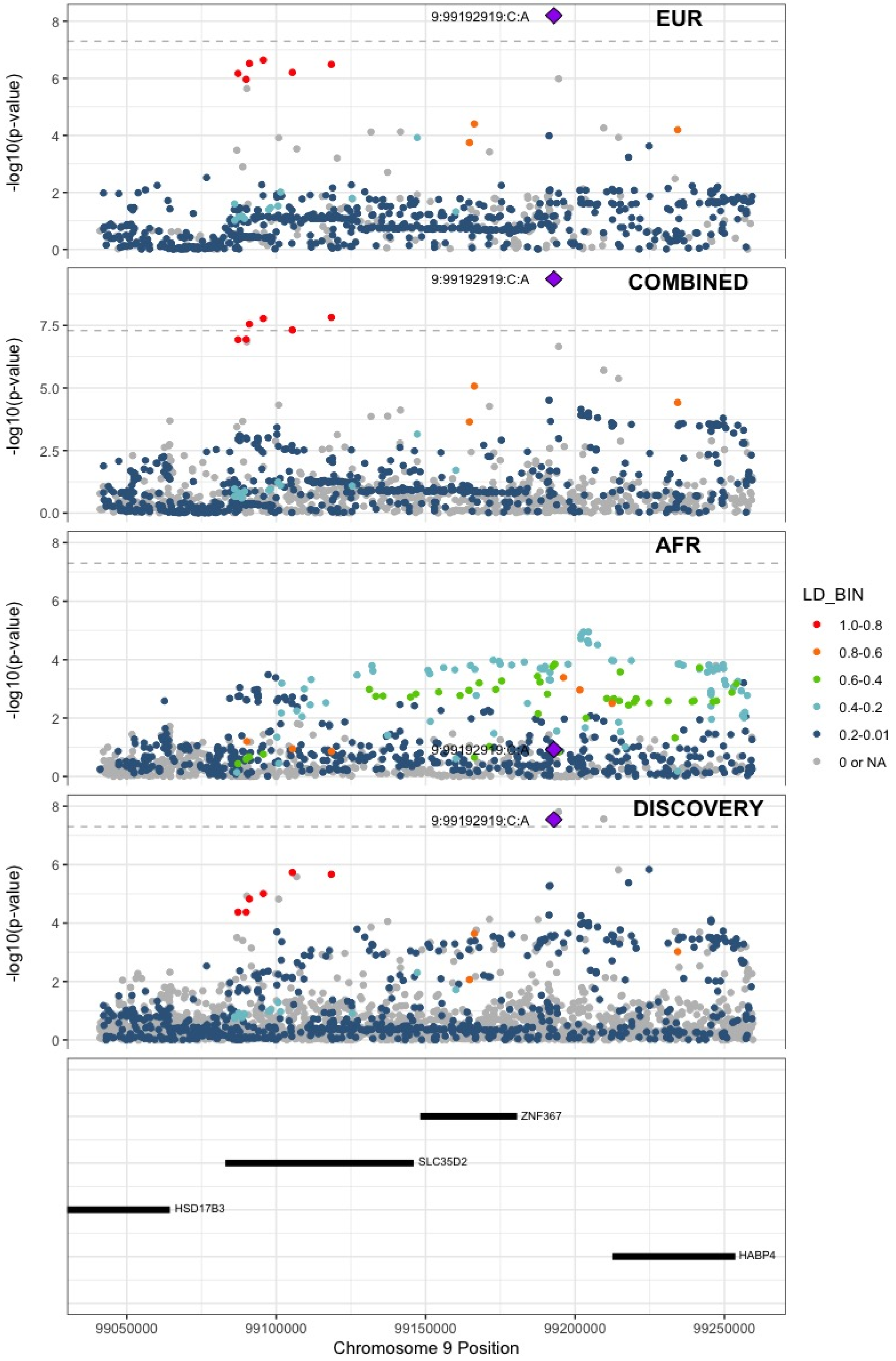

**Supplementary Figure 7.47** – rs687289 (chr9:136137106) in locus 66 (ABO)

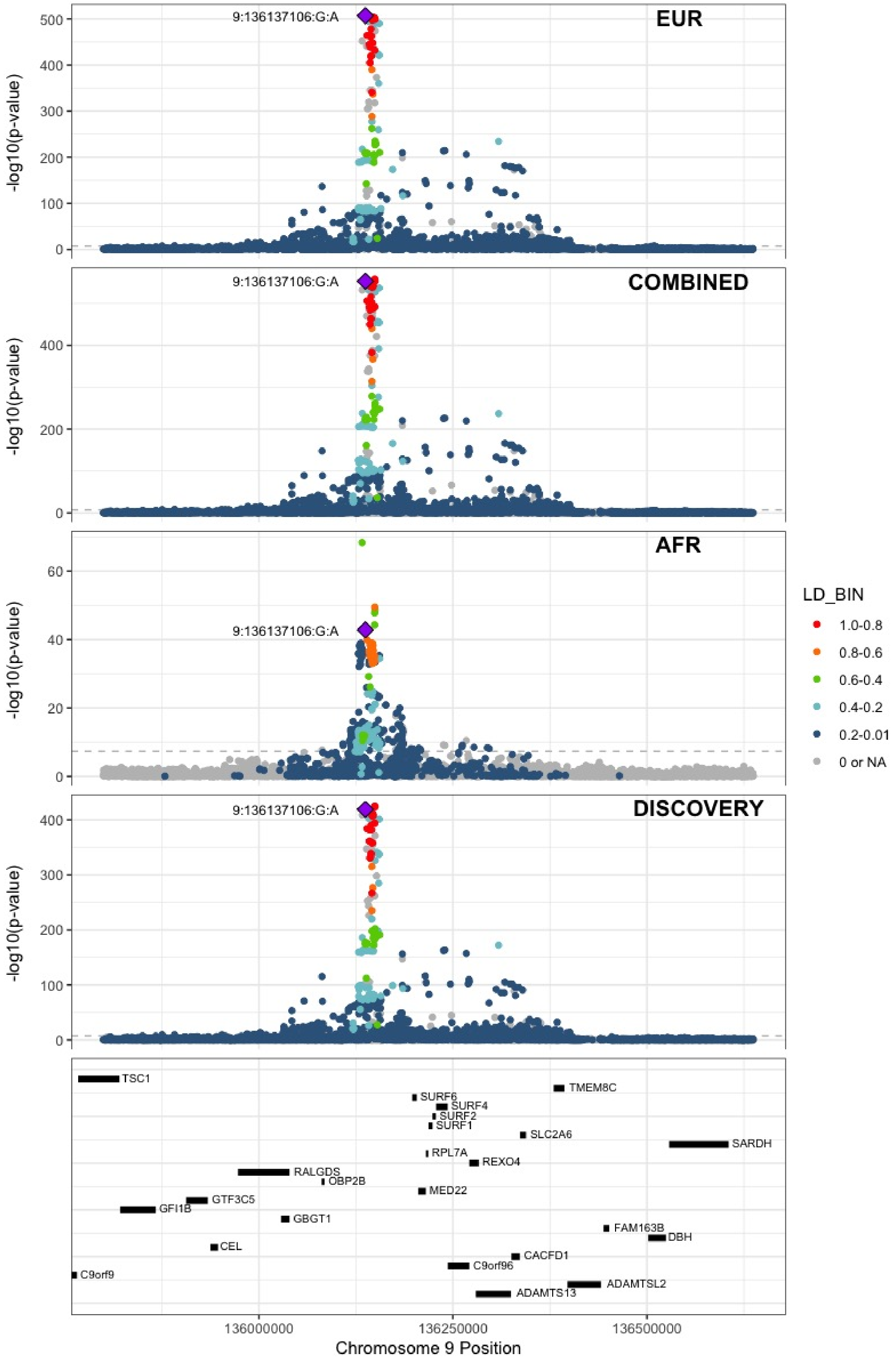

**Supplementary Figure 7.48** – rs1243184 (chr10:21931937) in locus 68 (MLLT10)

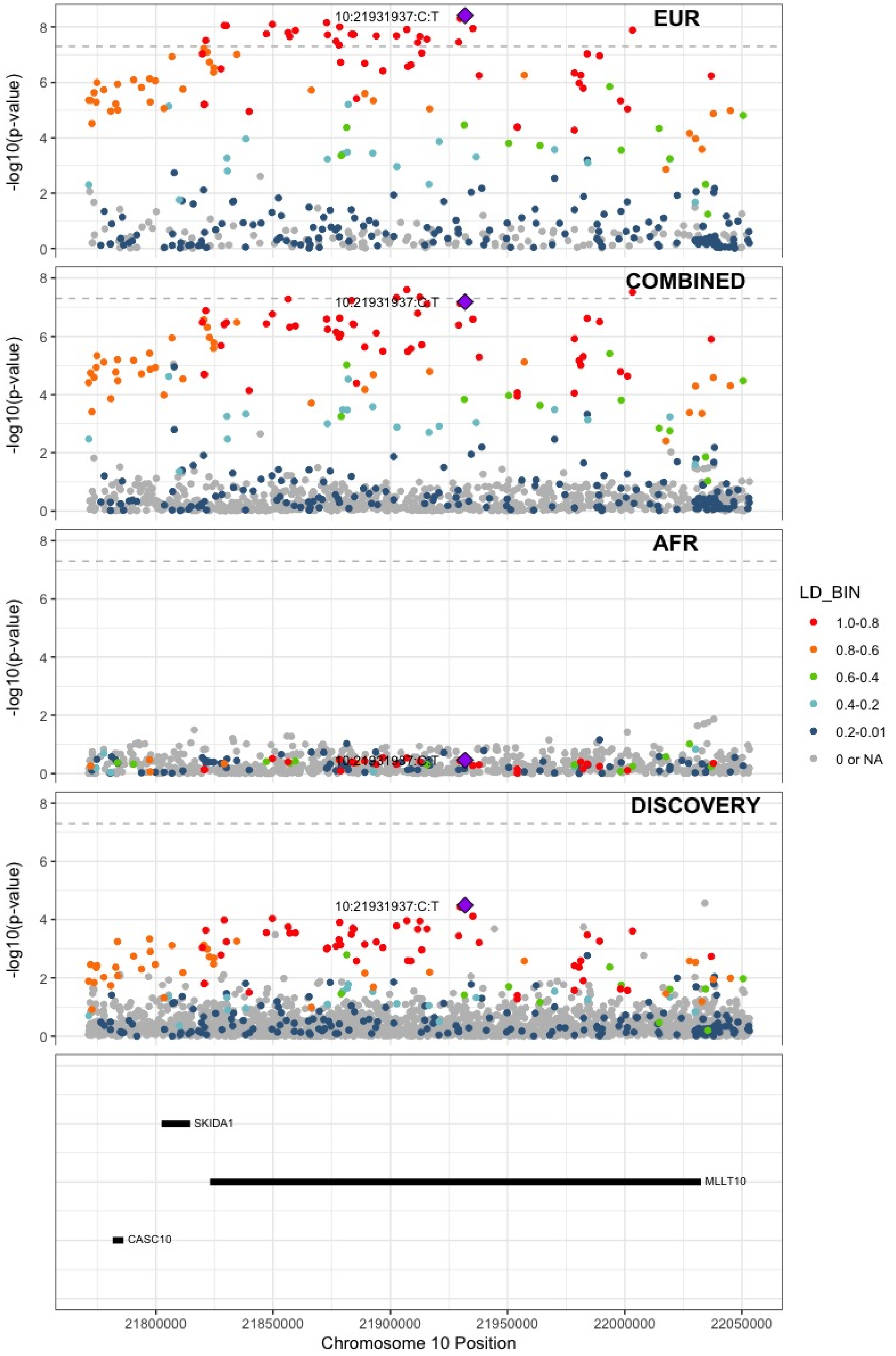

**Supplementary Figure 7.49** – rs10763665 (chr10:28771491) in locus 70 (LINC02652)

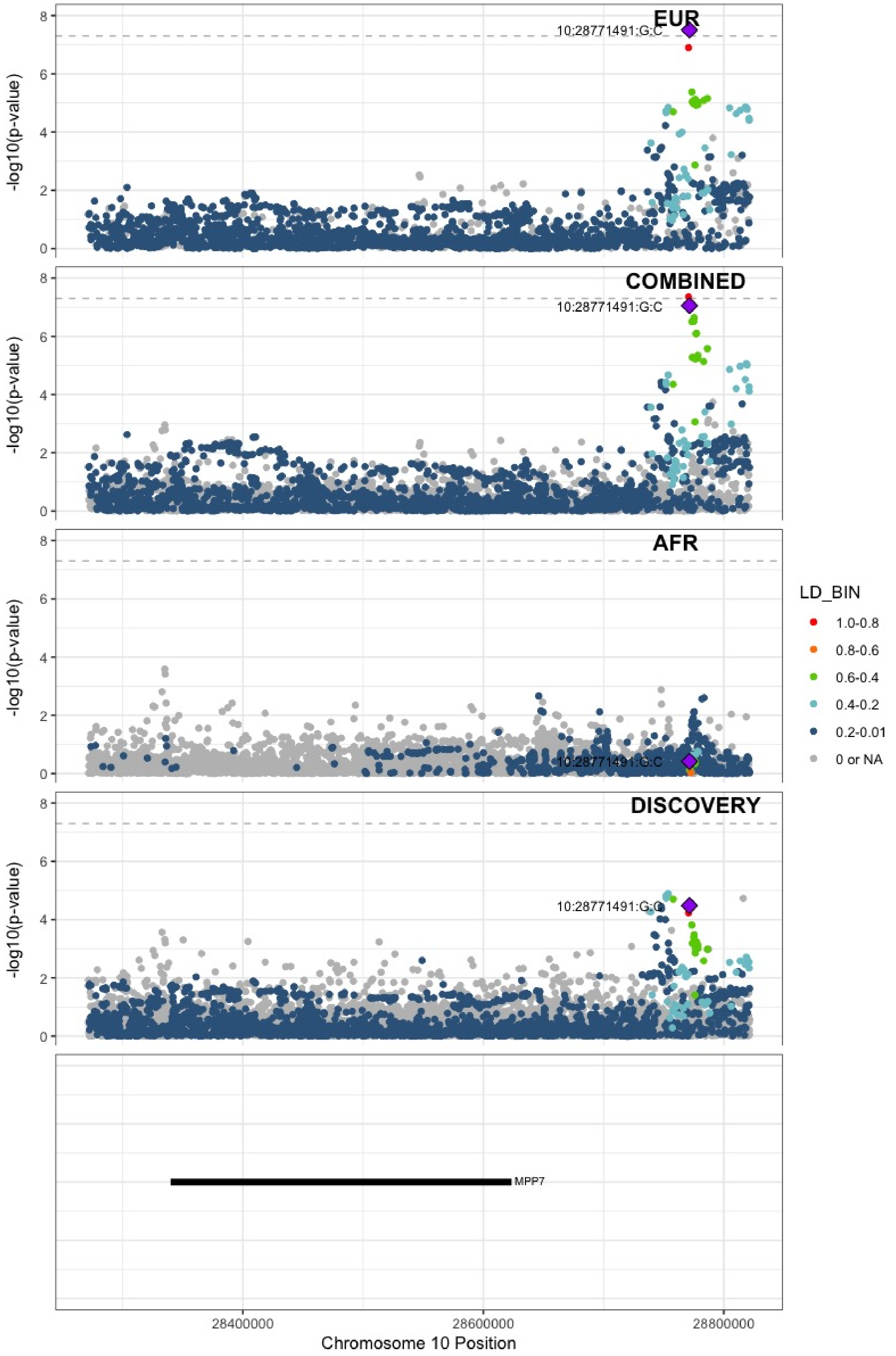

**Supplementary Figure 7.50** – rs78707713 (chr10:71245276) in locus 72 (TSPAN15)

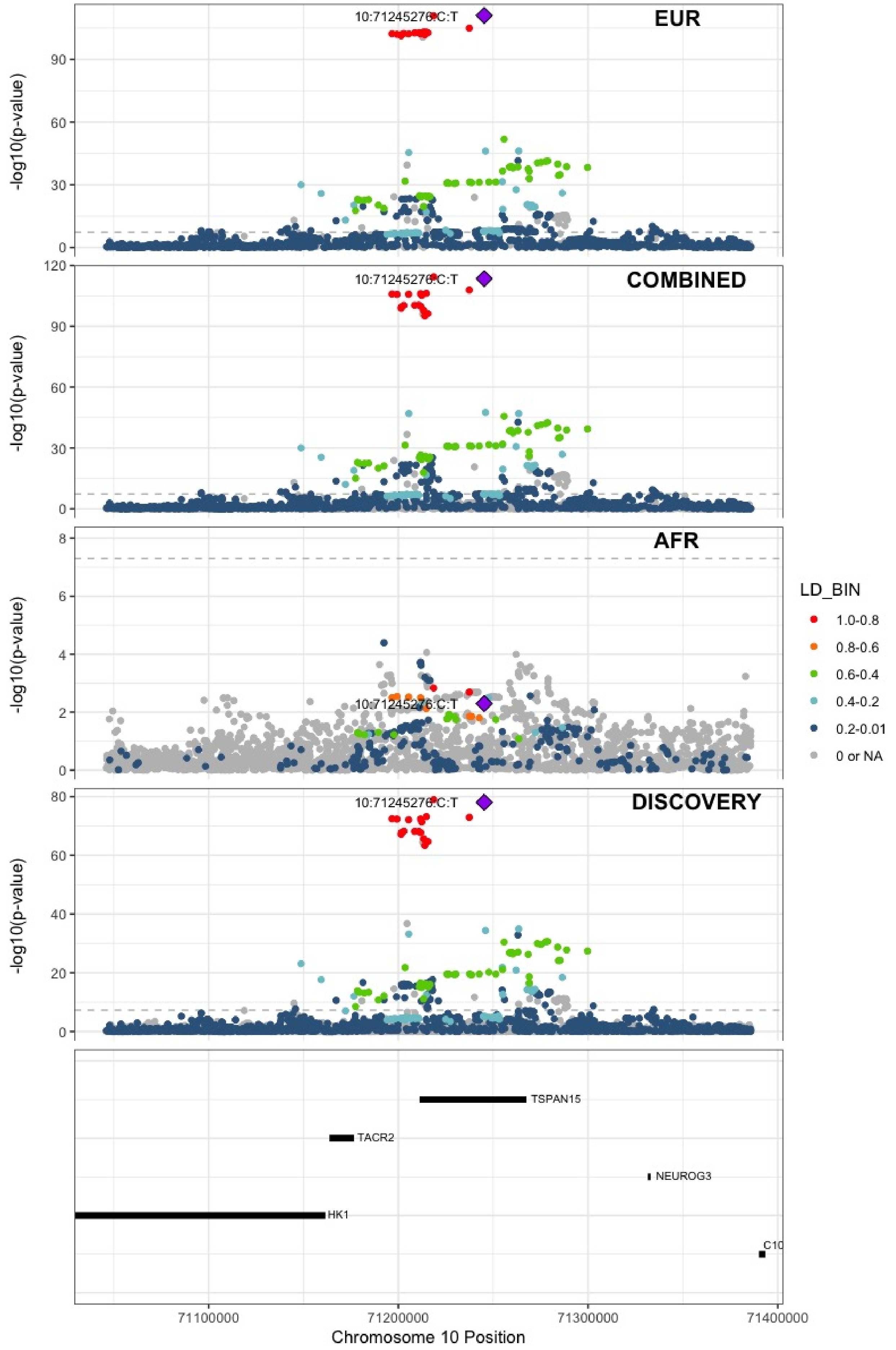

**Supplementary Figure 7.51** – rs57866767 (chr10:96023077) in locus 74 (PLCE1)

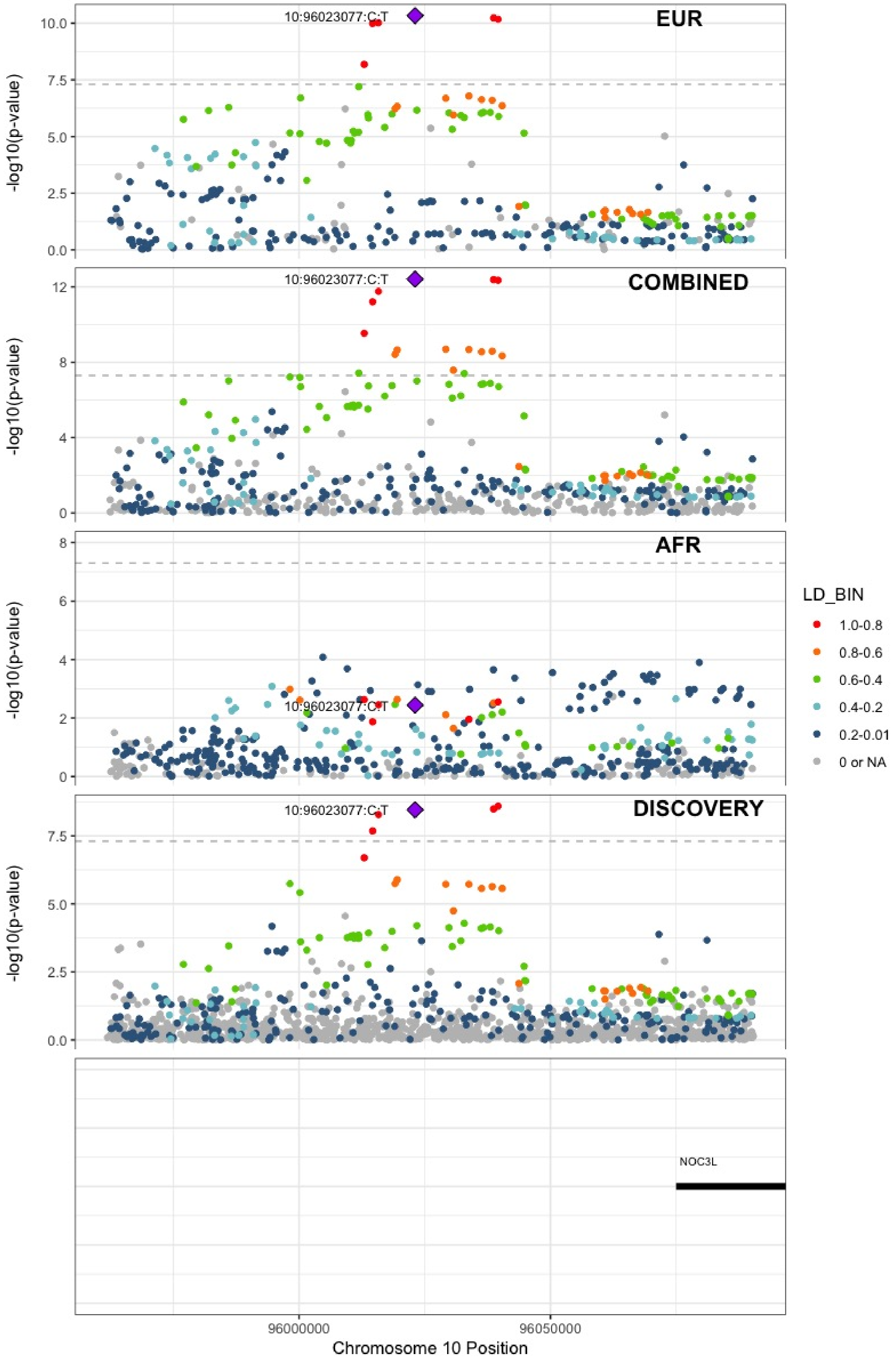

**Supplementary Figure 7.52** – rs10886430 (chr10:121010256) in locus 75 (GRK5)

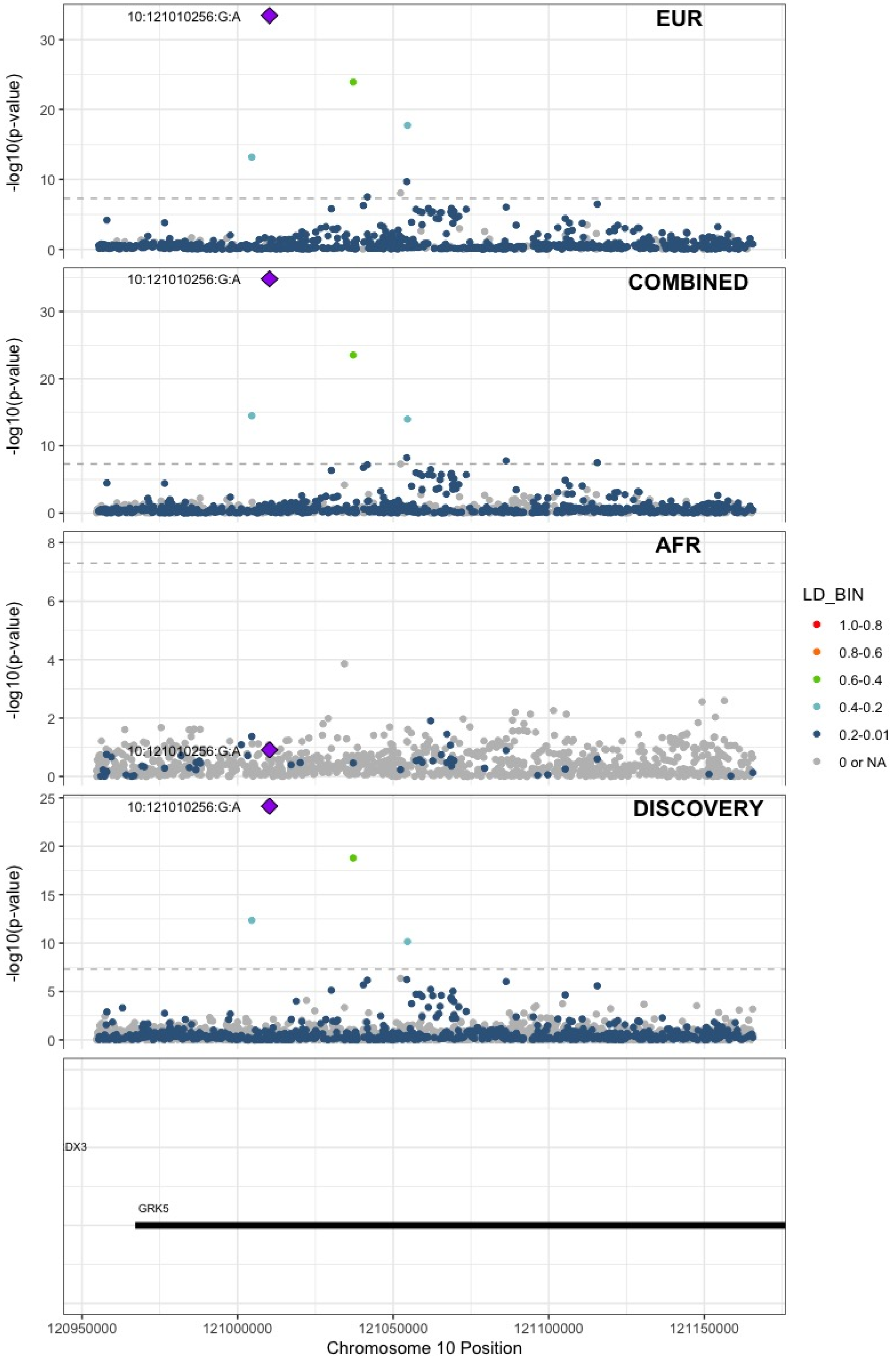

**Supplementary Figure 7.53** – rs7122100 (chr11:10732560) in locus 76 (IRAG1;CTR9)

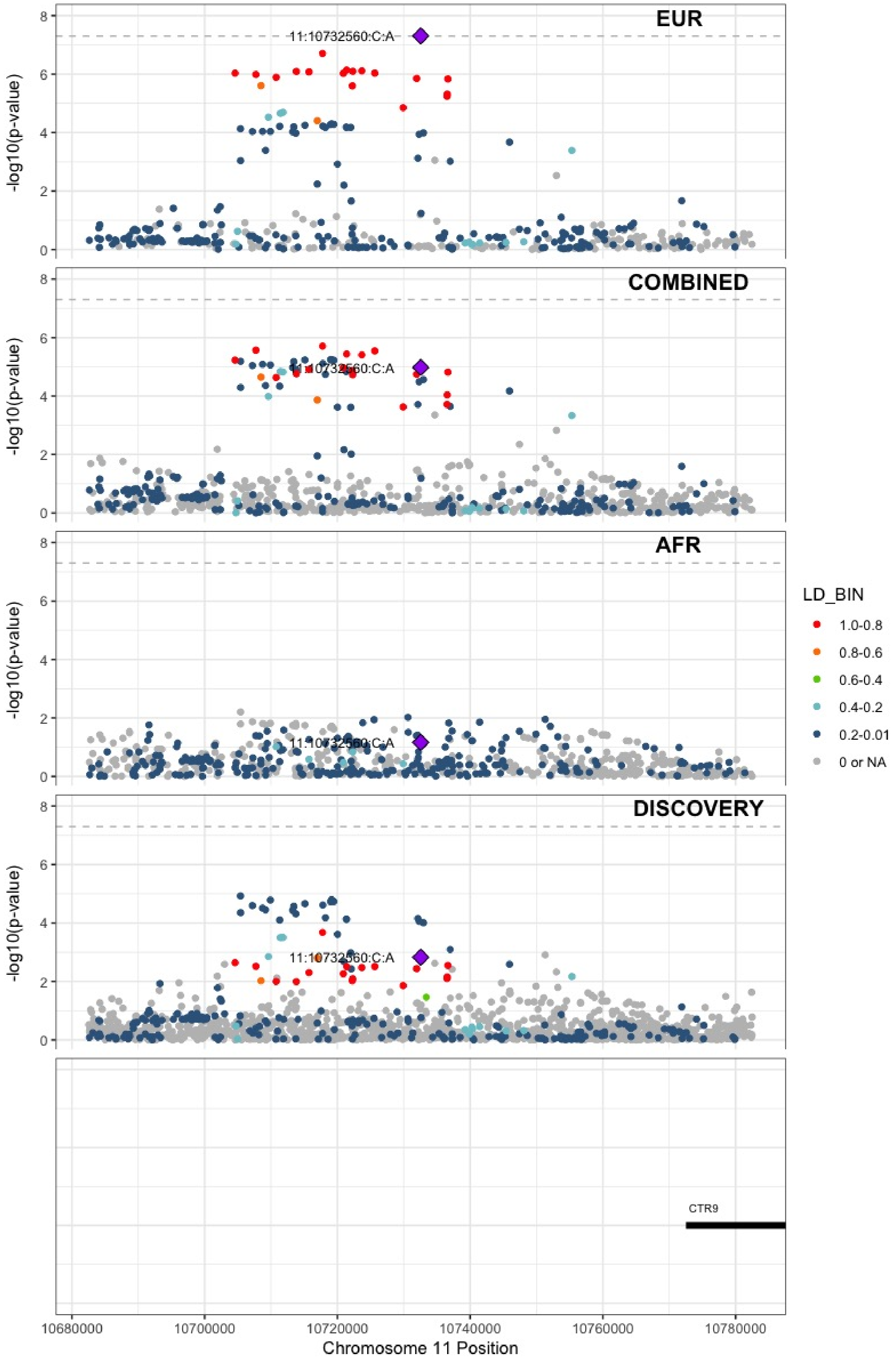

**Supplementary Figure 7.54** – rs12800654 (chr11:16243320) in locus 77 (SOX6)

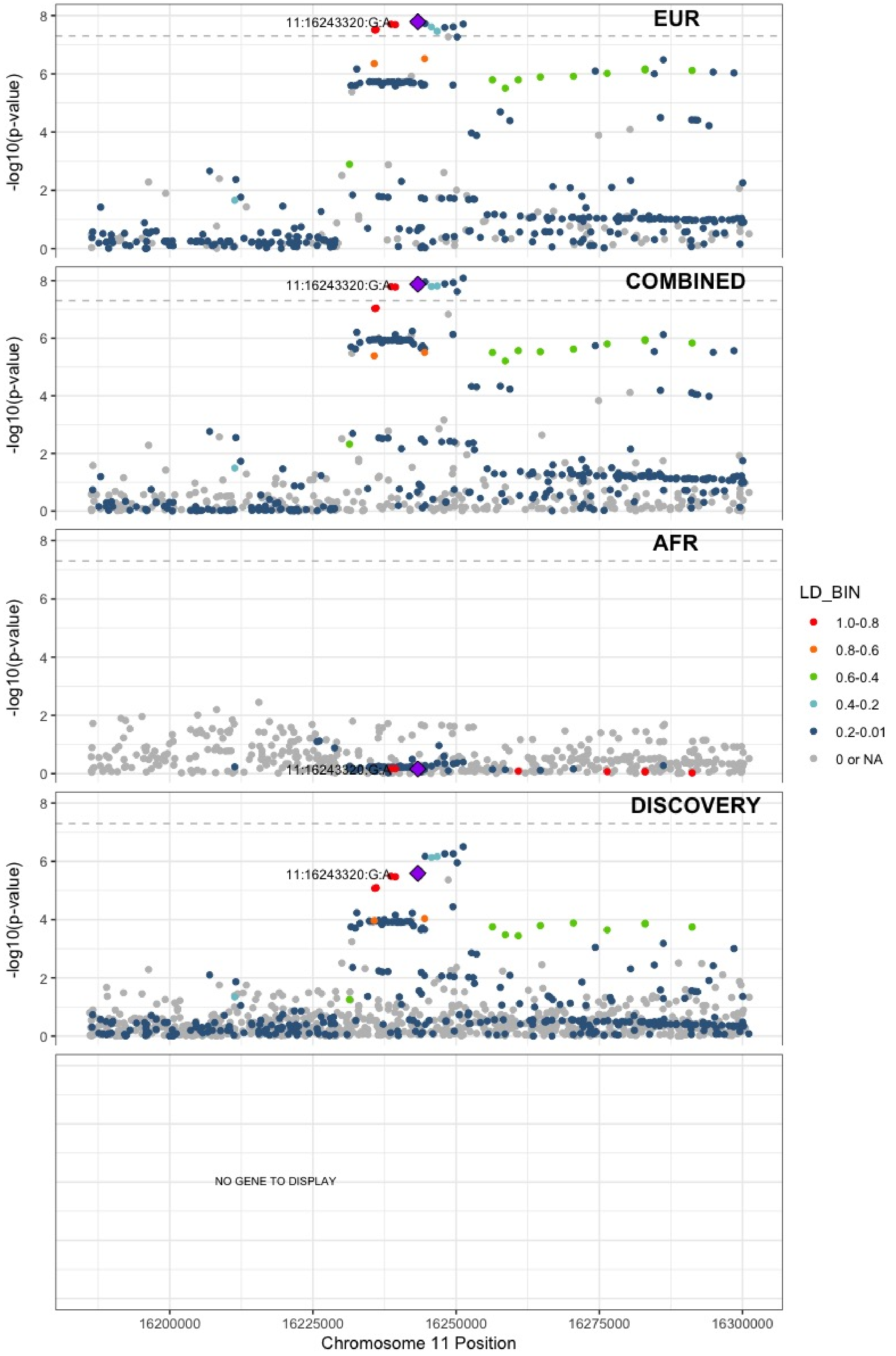

**Supplementary Figure 7.55** – rs4755476 (chr11:32980428) in locus 78 (QSER1)

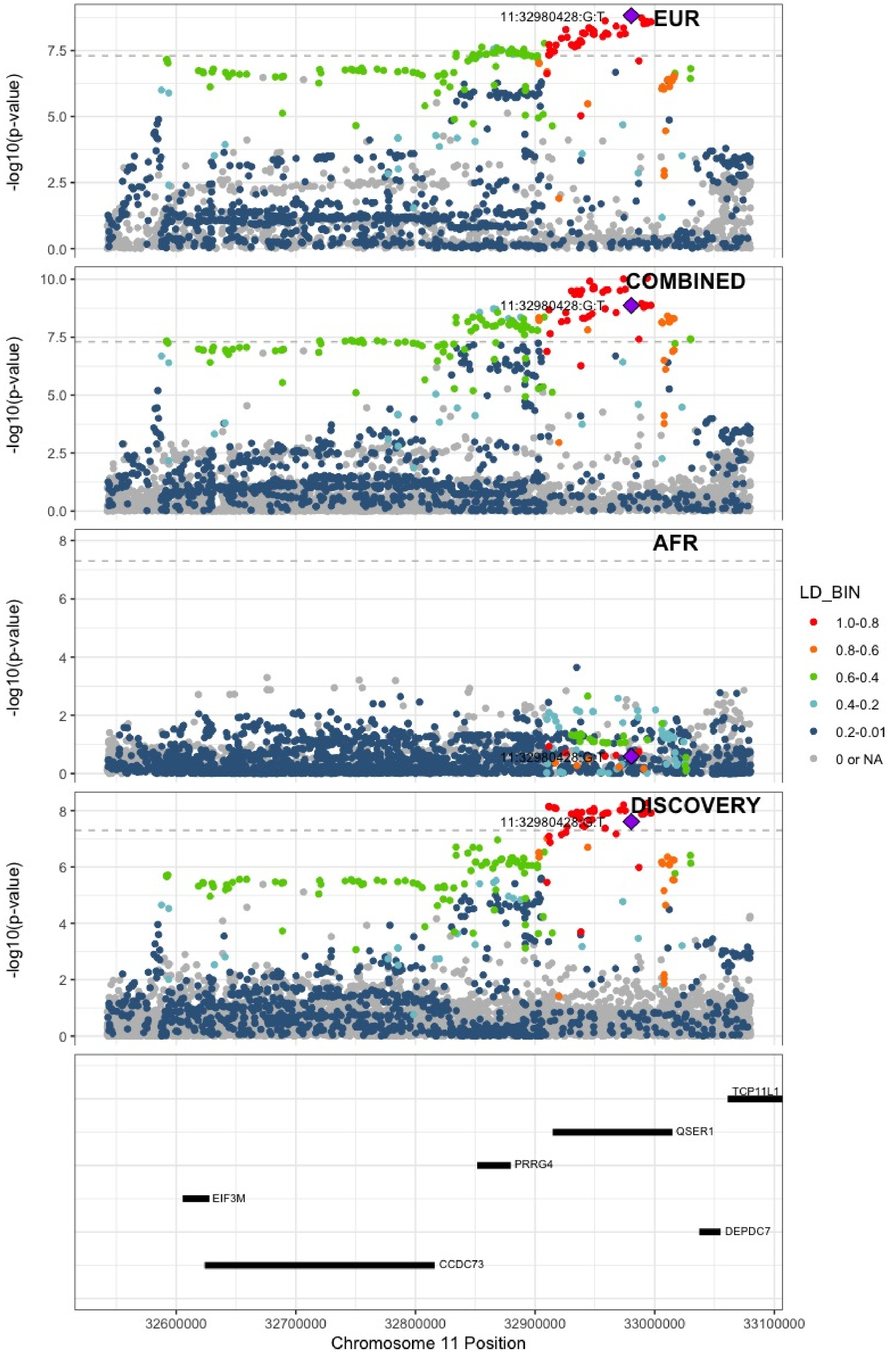

**Supplementary Figure 7.56** – rs1799963 (chr11:46761055) in locus 79 (F2)

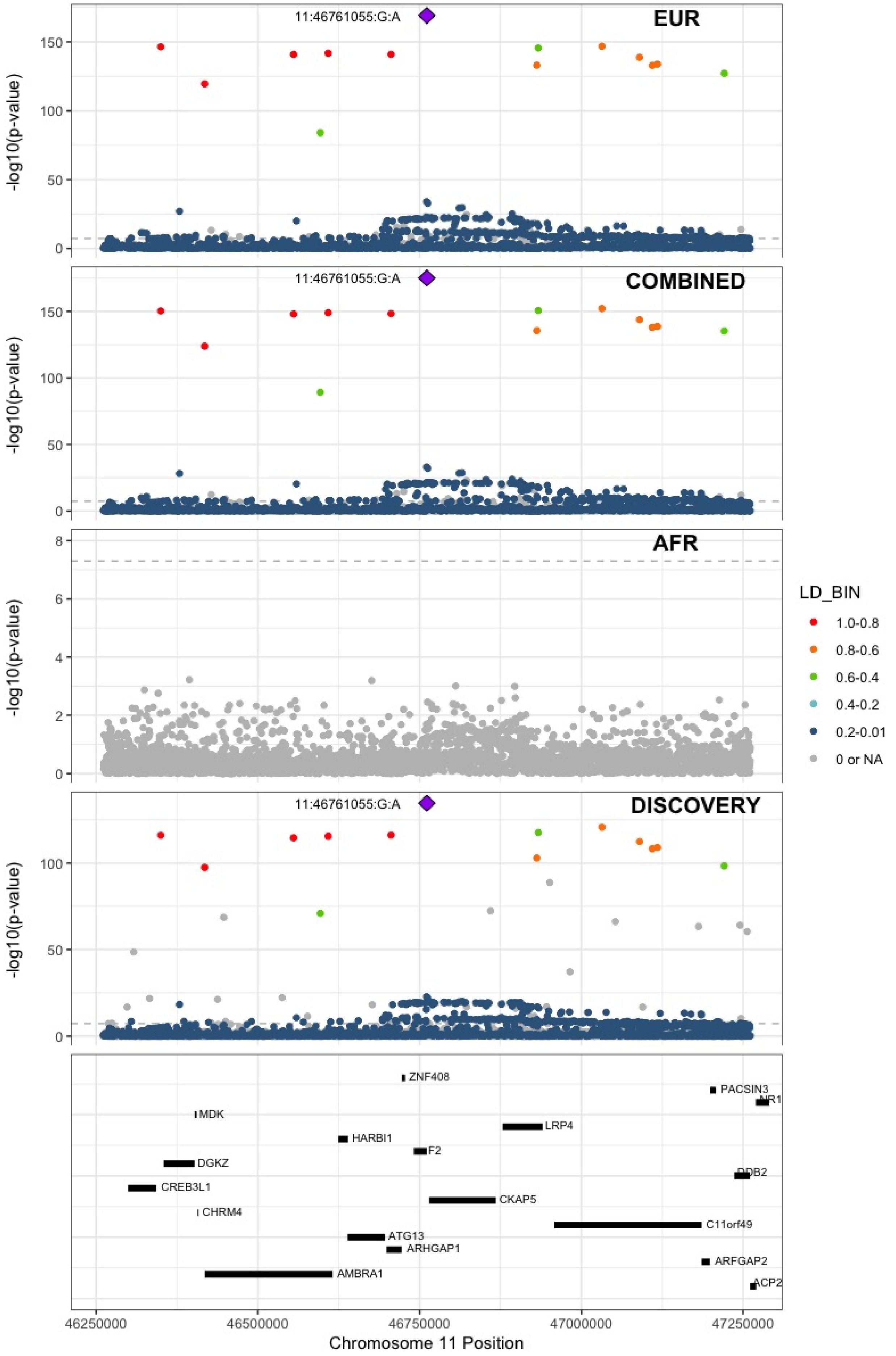

**Supplementary Figure 7.57** – rs141798115 (chr11:56875074) in locus 80 (OR5AK4P ;LRRC55)

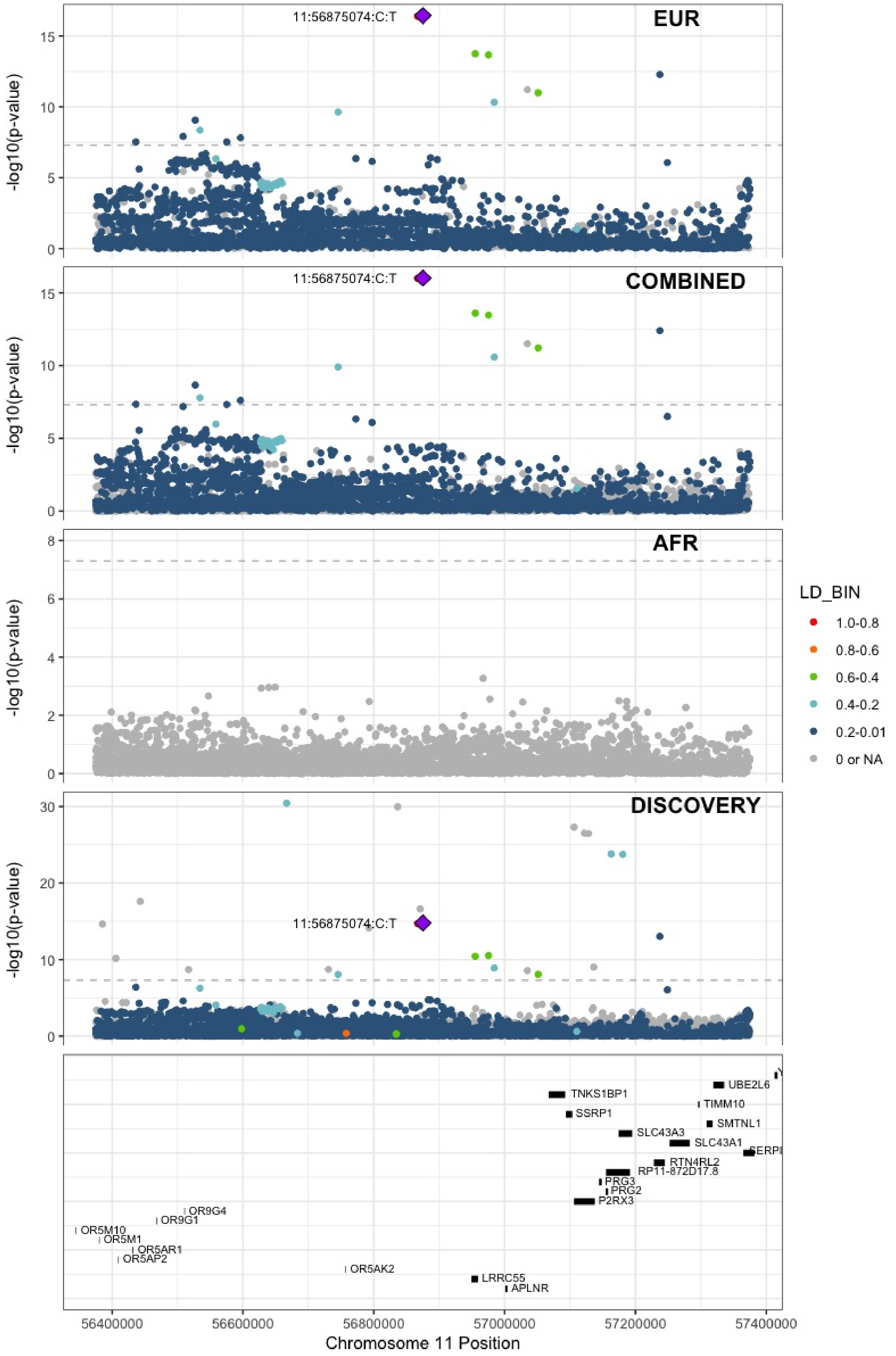

**Supplementary Figure 7.58** – rs55777218 (chr11:60070946) in locus 81 (MS4A4A)

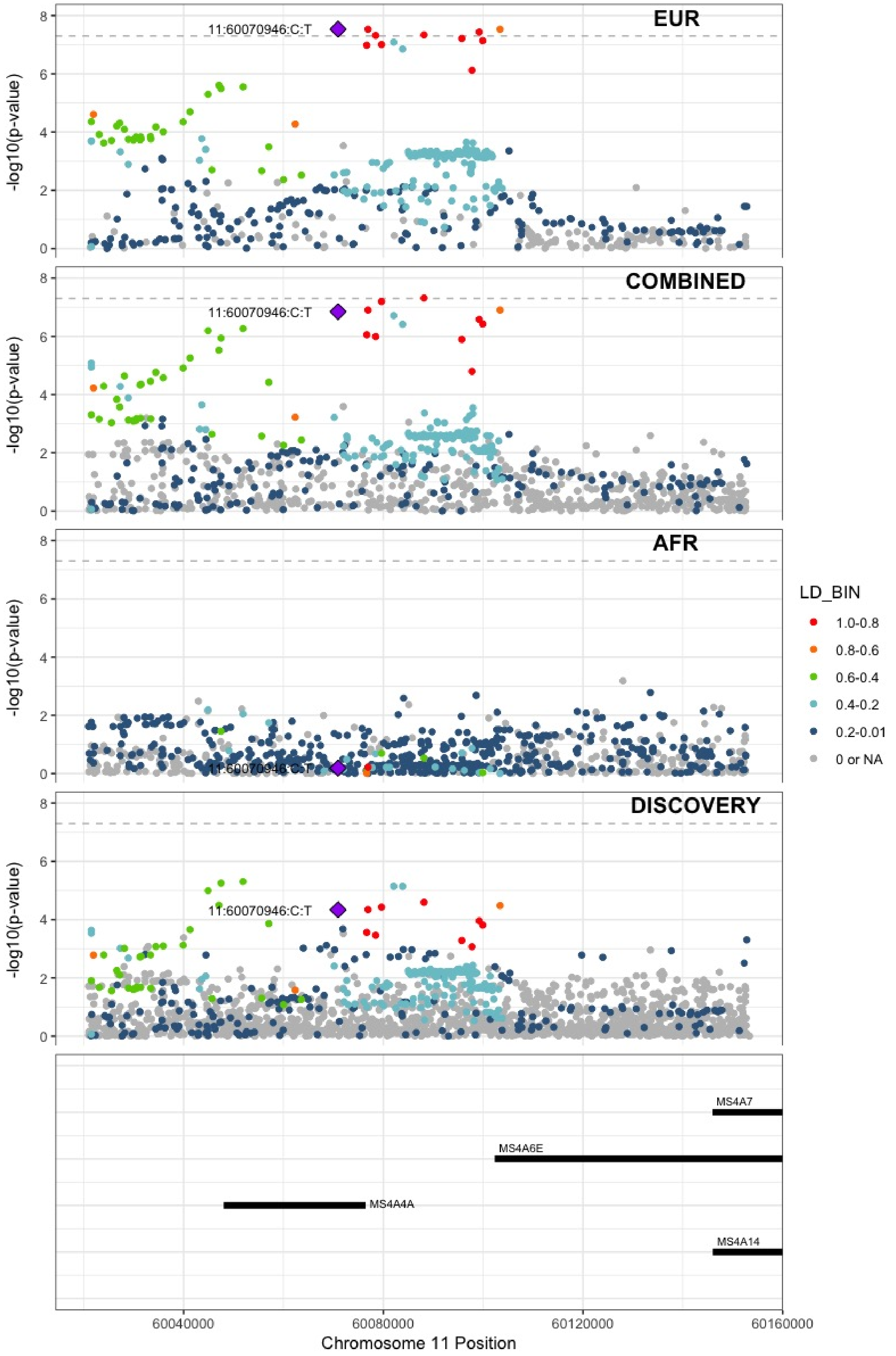

**Supplementary Figure 7.59** – rs174551 (chr11:61573684) in locus 82 (FADS1)

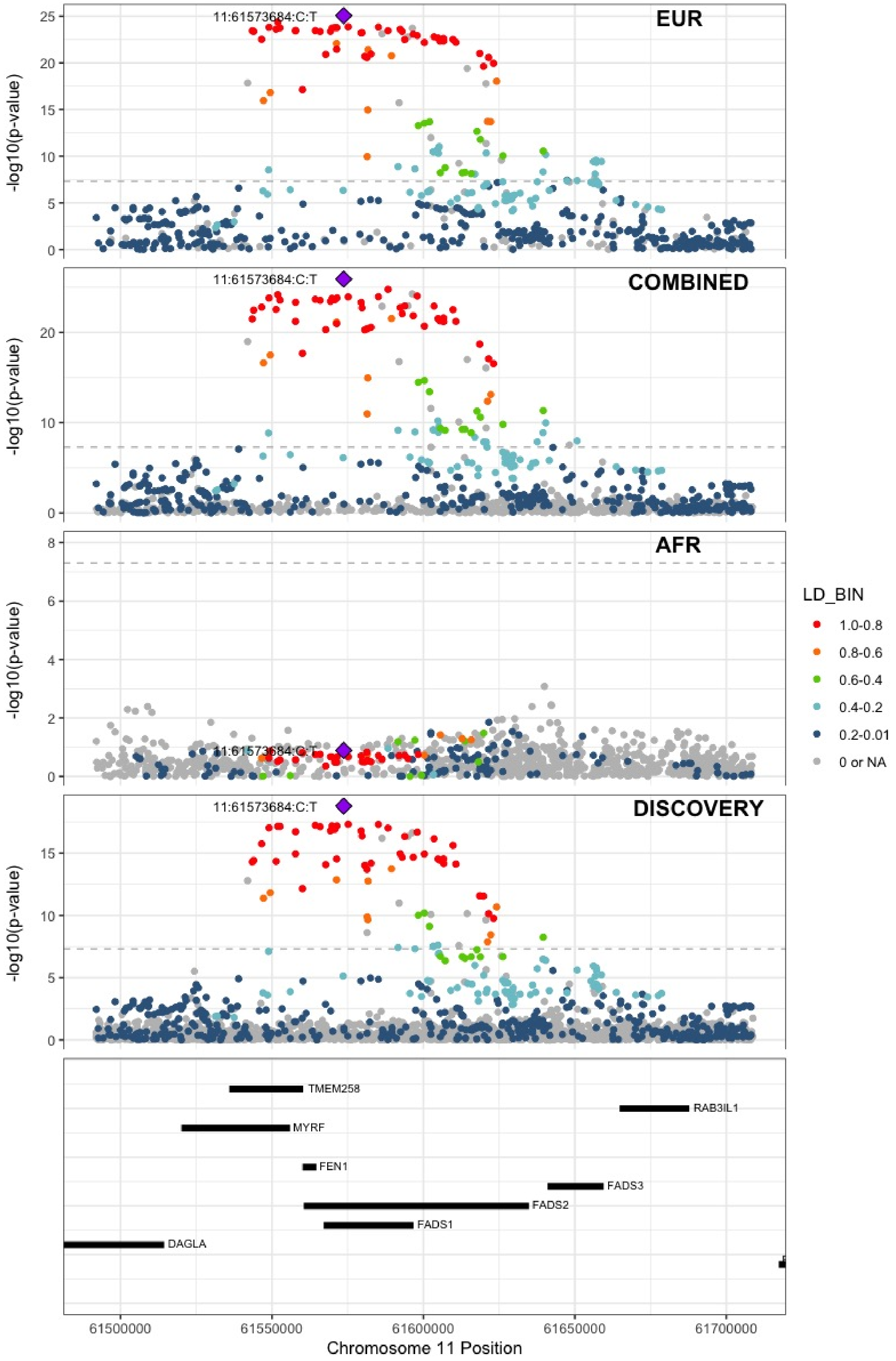

**Supplementary Figure 7.60** – rs1145656 (chr11:73305859) in locus 83 (FAM168A)

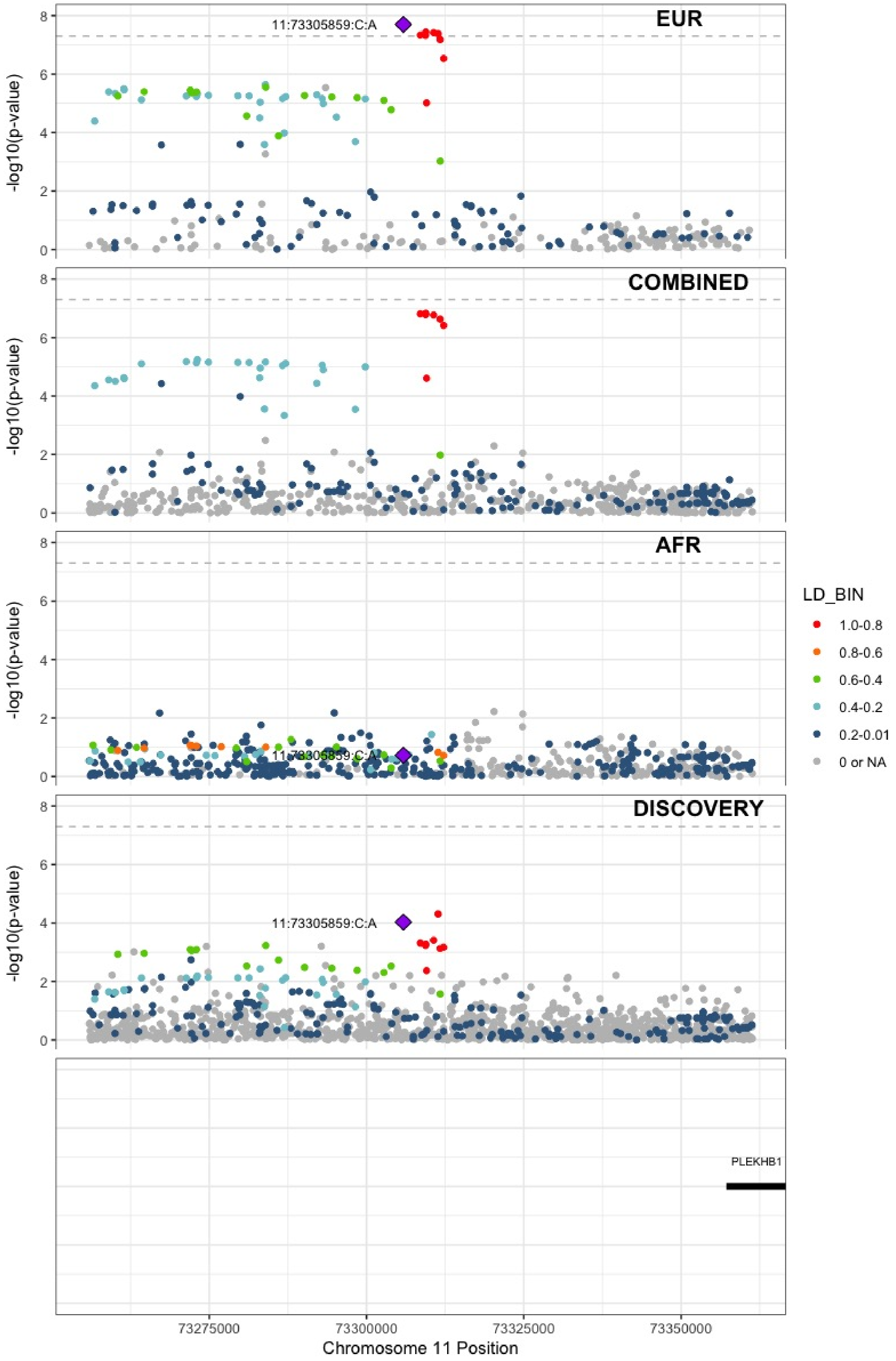

**Supplementary Figure 7.61** – rs1997547 (chr11:114002744) in locus 84 (ZBTB16)

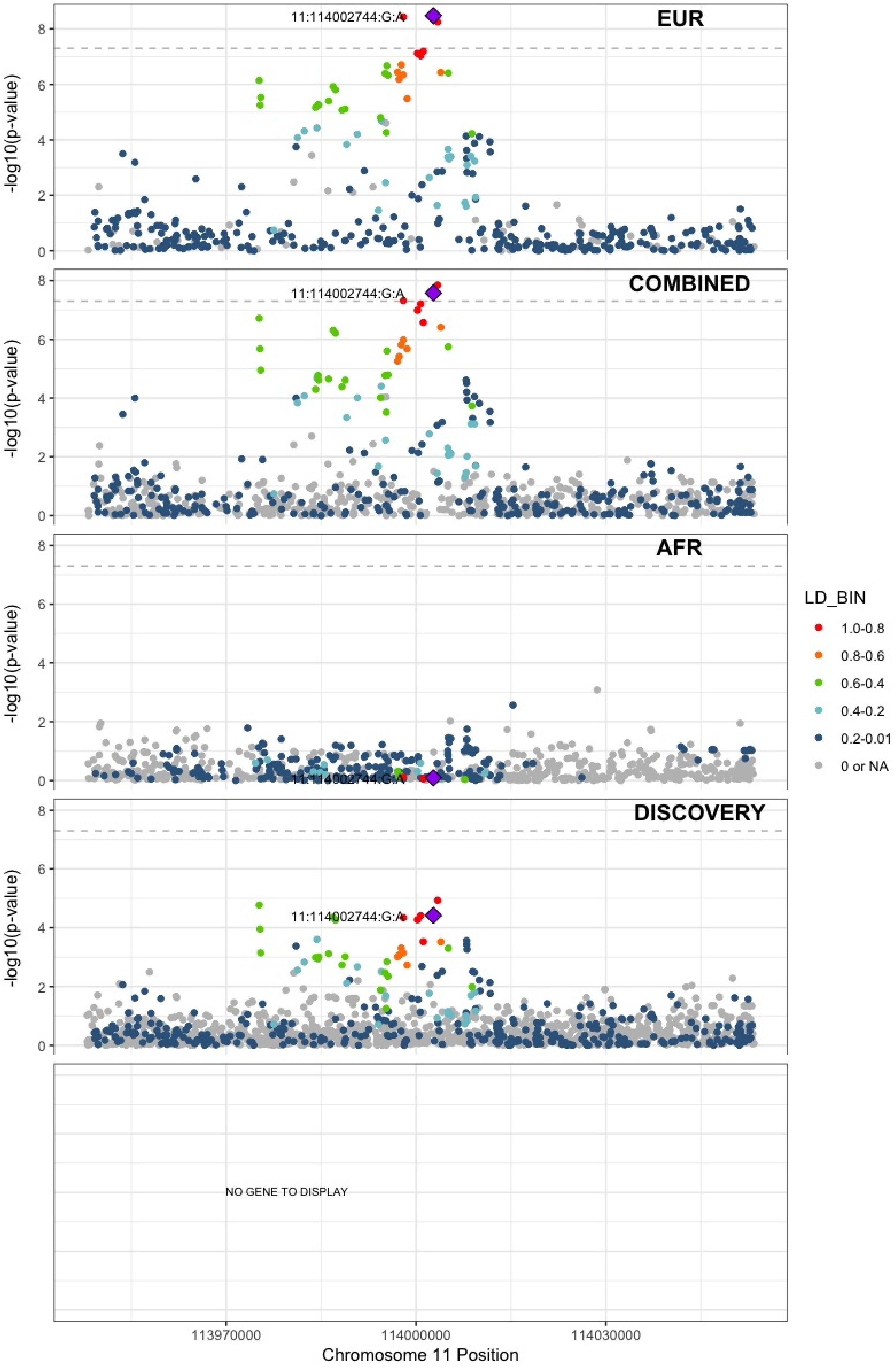

**Supplementary Figure 7.62** – rs35257264 (chr11:126296816) in locus 85 (ST3GAL4)

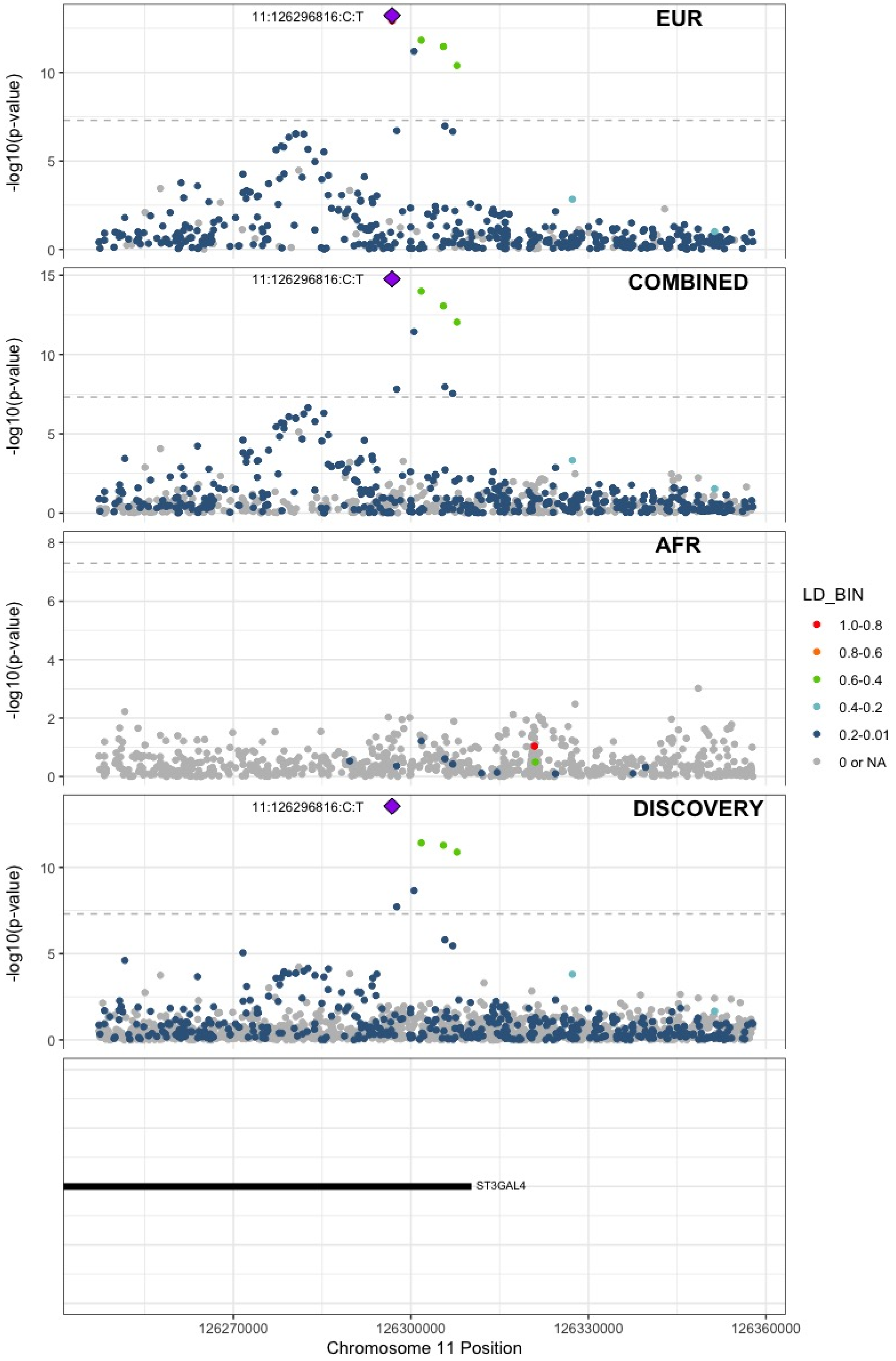

**Supplementary Figure 7.63** – rs7135039 (chr12:6160614) in locus 87 (VWF)

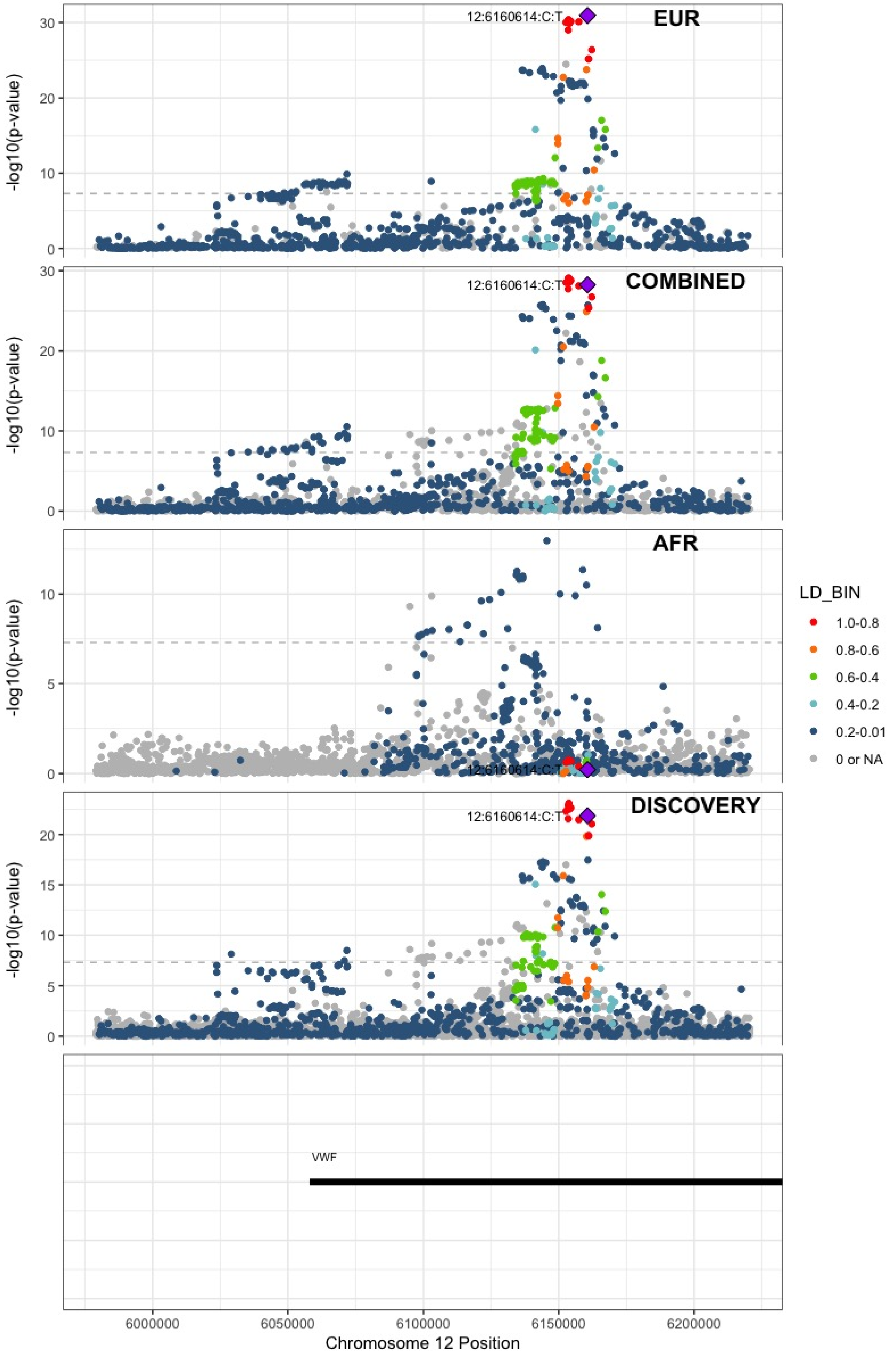

**Supplementary Figure 7.64** – rs7311483 (chr12:9053661) in locus 88 (A2ML1 ;PHC1)

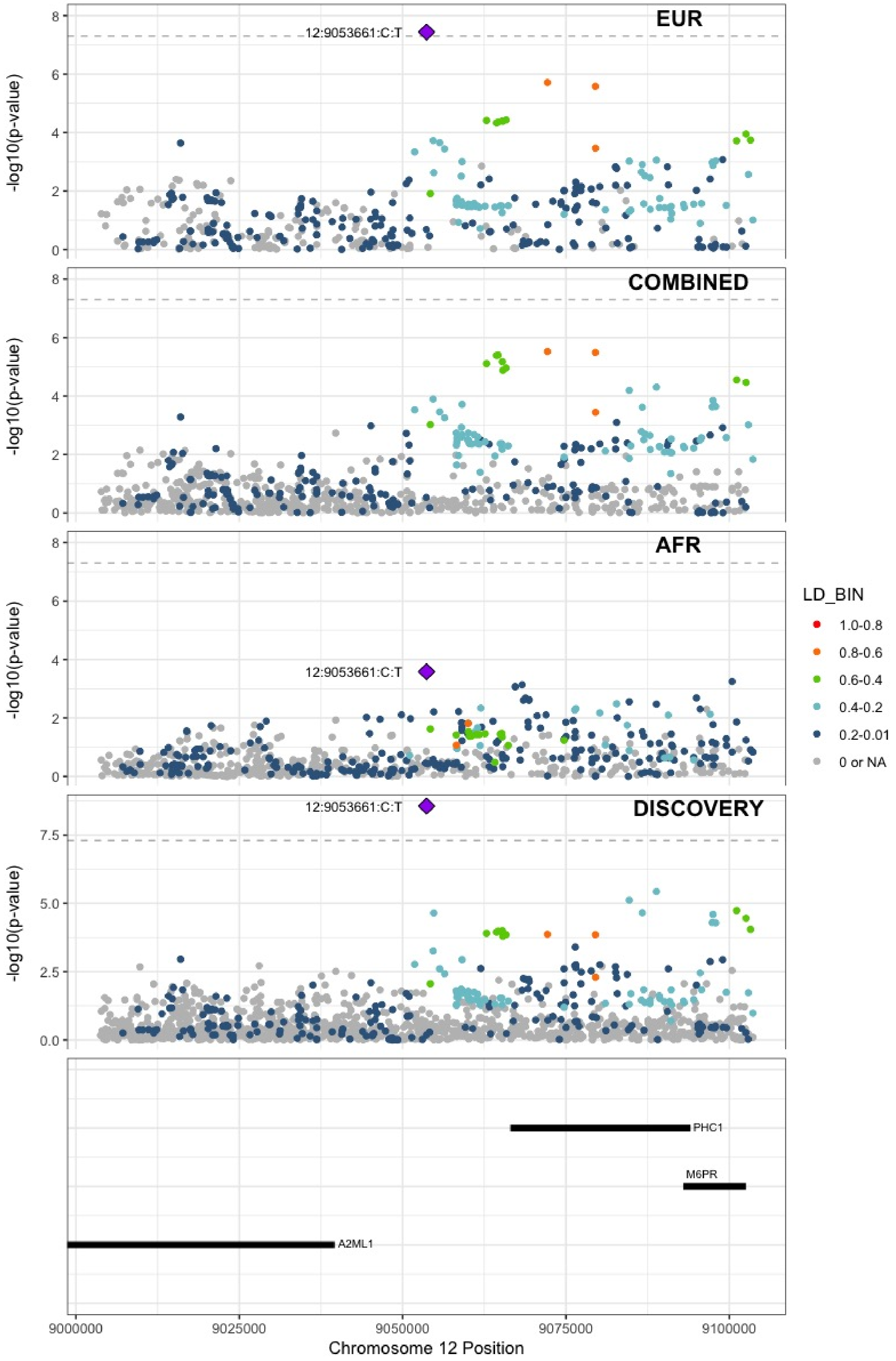

**Supplementary Figure 7.65** – rs4759076 (chr12:54729872) in locus 89 (COPZ1)

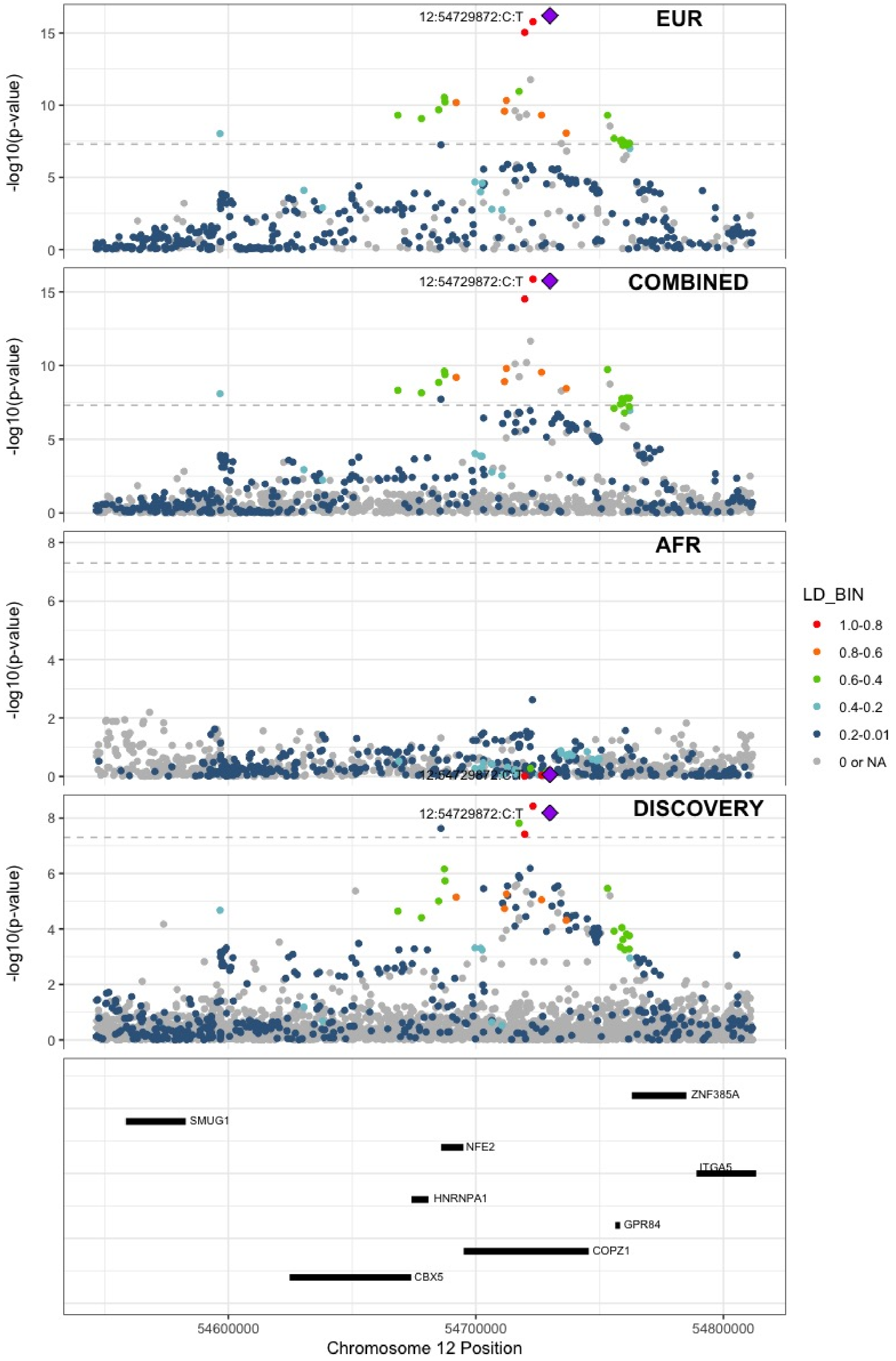

**Supplementary Figure 7.66** – rs142351376 (chr12:104136288) in locus 90 (STAB2)

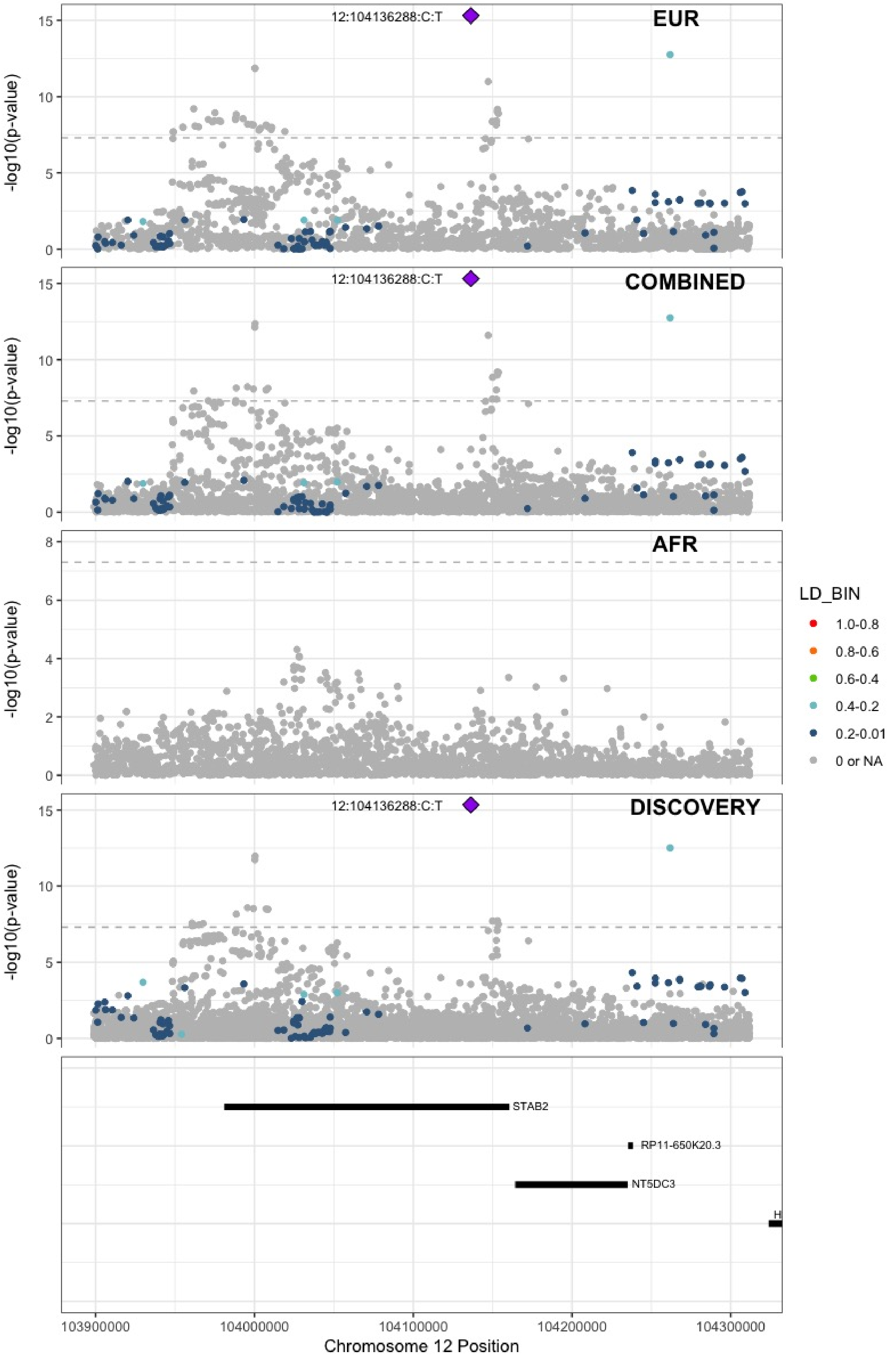

**Supplementary Figure 7.67** – rs56750693 (chr12:112144586) in locus 91 (ACAD10)

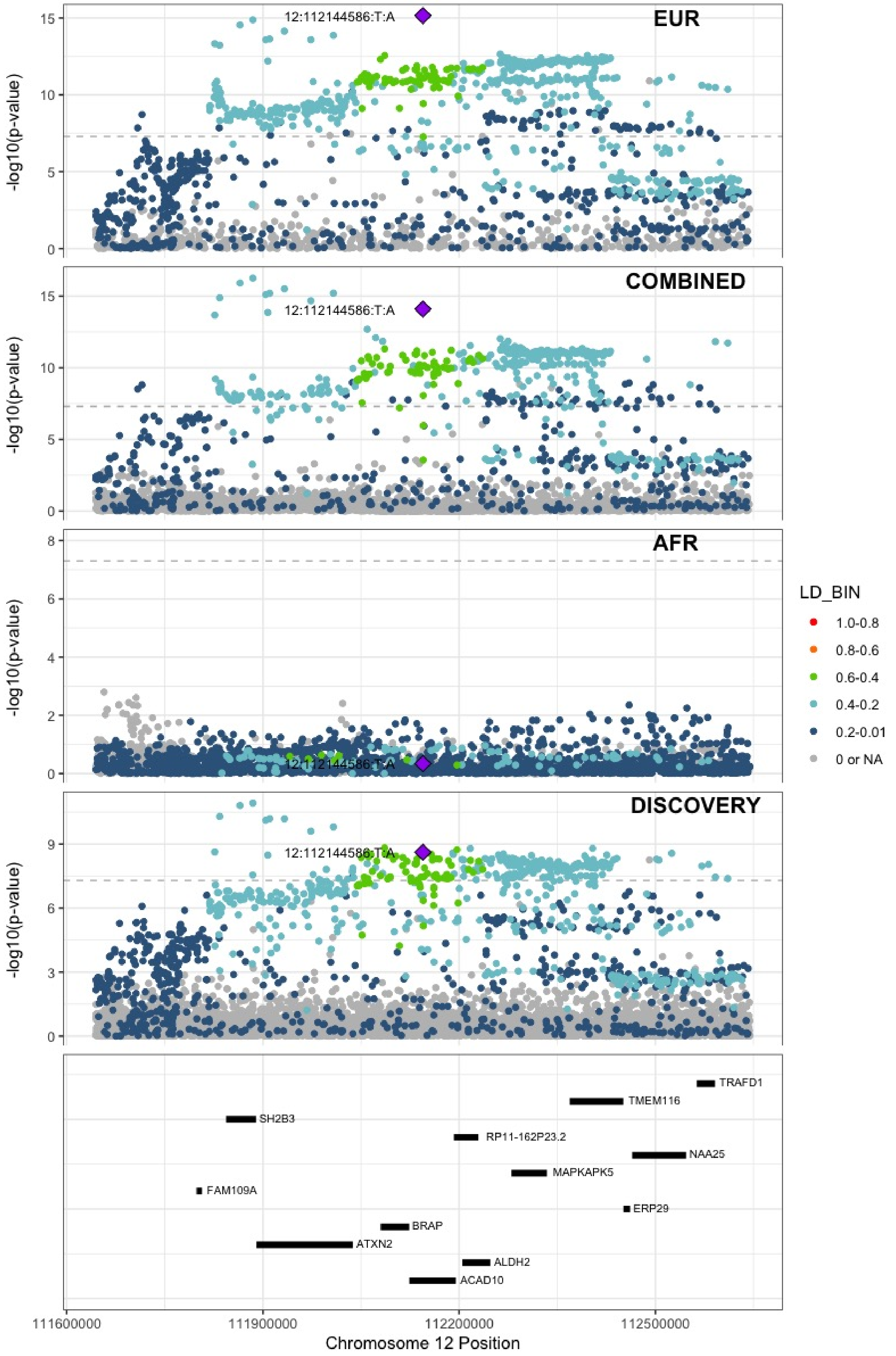

**Supplementary Figure 7.68** – rs2851436 (chr12:123667354) in locus 92 (MPHOSPH9)

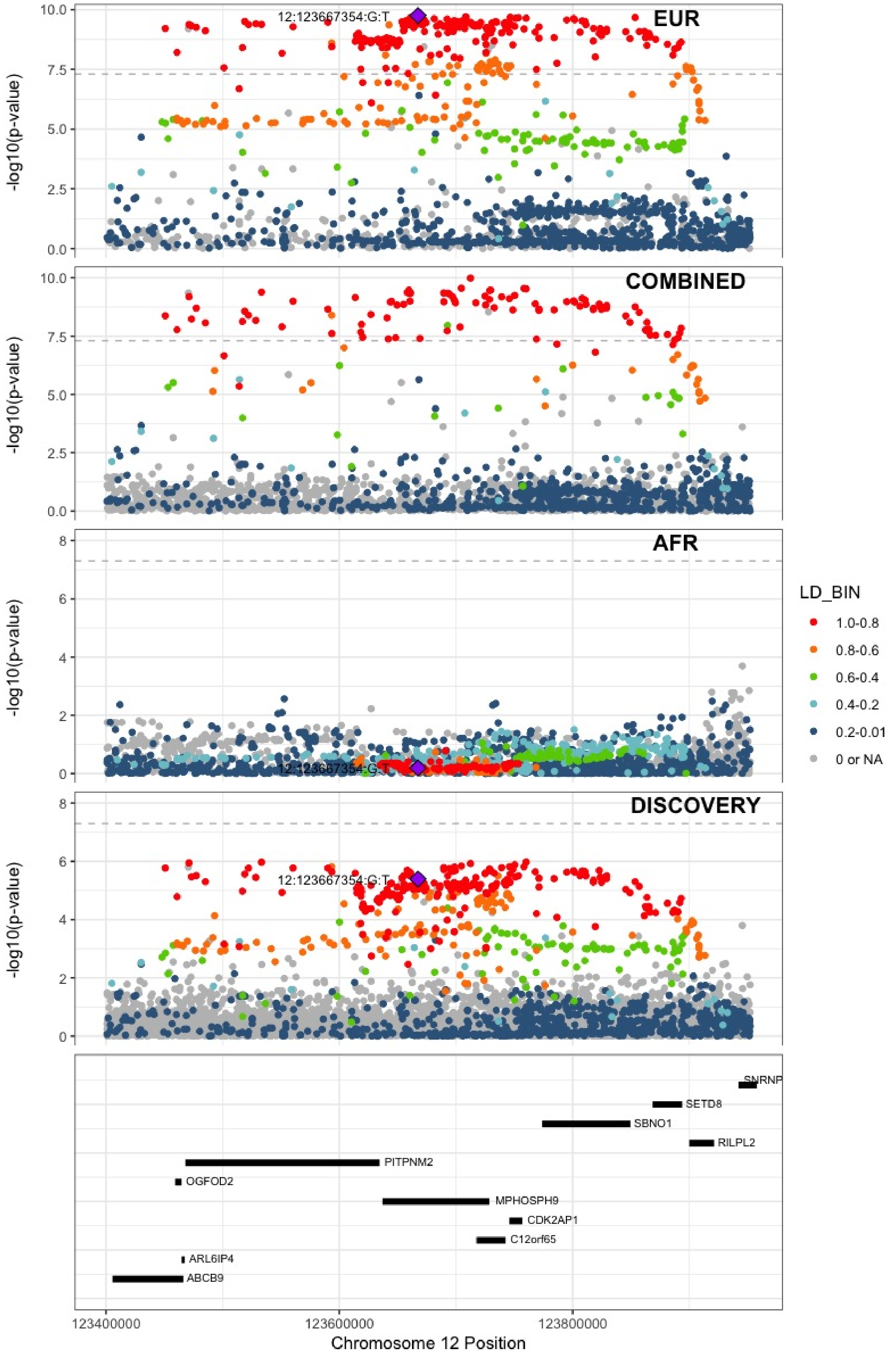

**Supplementary Figure 7.69** – rs3211752 (chr13:113787459) in locus 94 (F10)

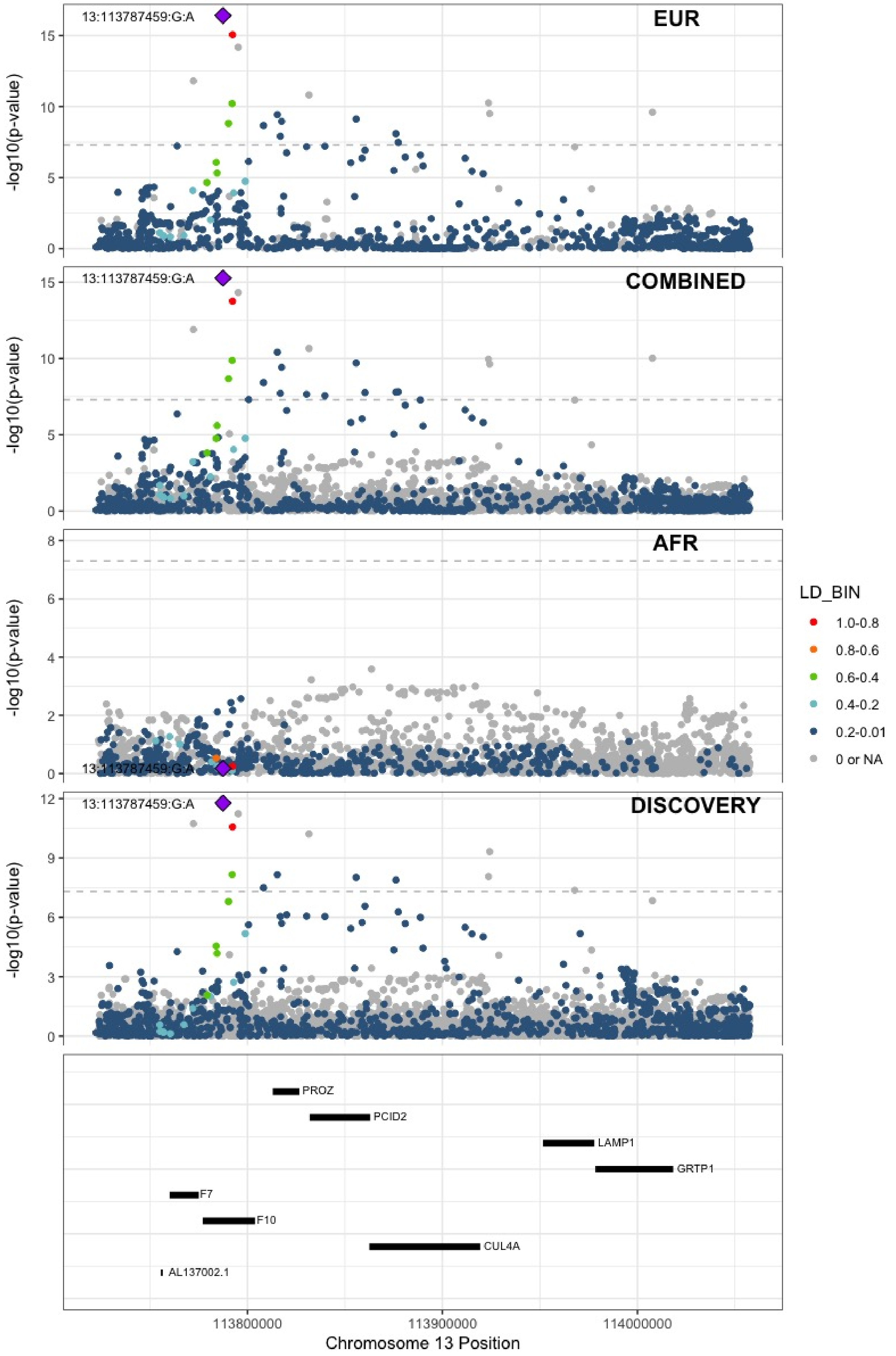

**Supplementary Figure 7.70** – rs8004077 (chr14:58846731) in locus 95 (ARID4A;TOMM20L)

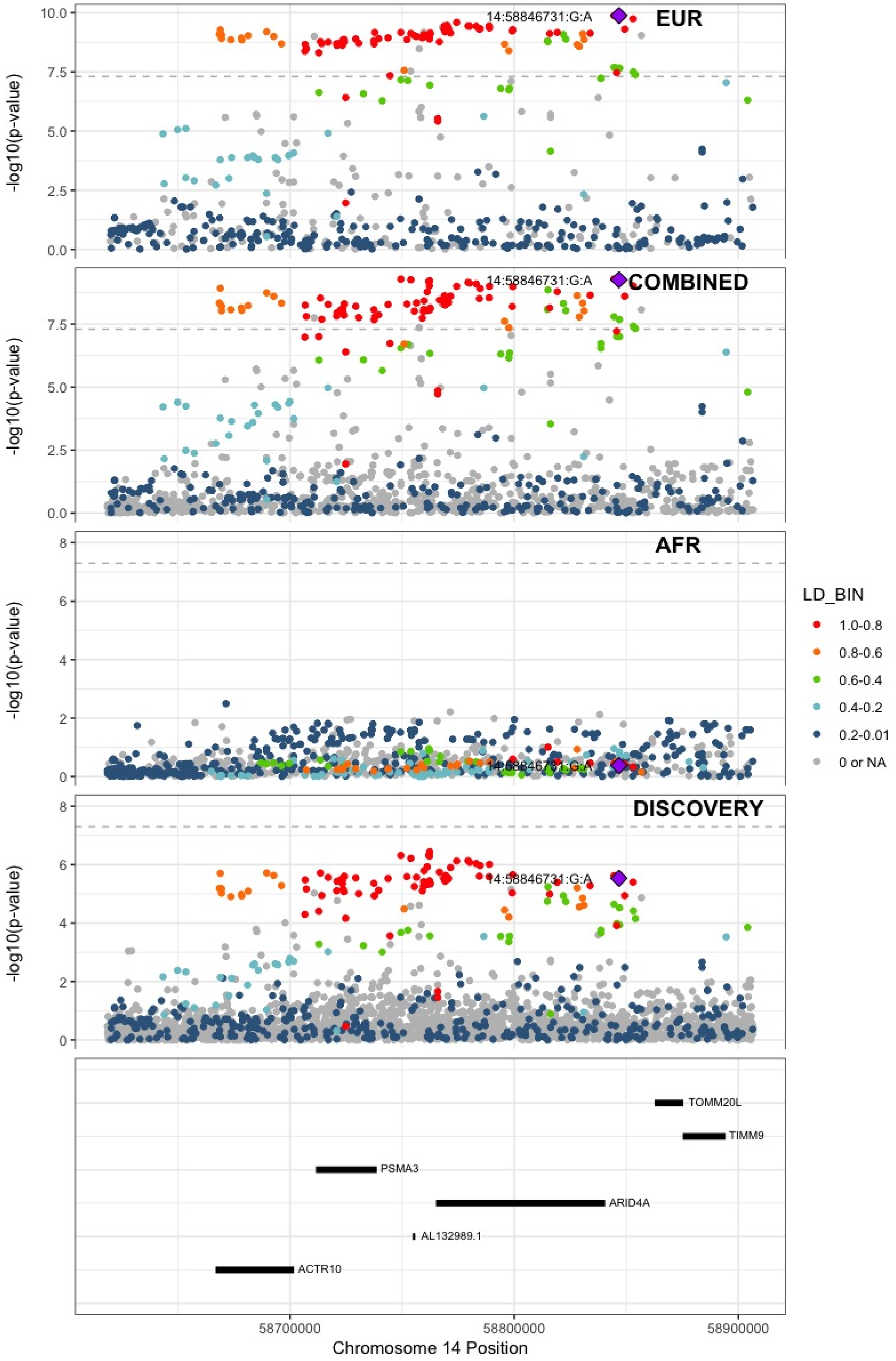

**Supplementary Figure 7.71** – rs2127869 (chr14:65794352) in locus 96 (LINC02324;MIR4708)

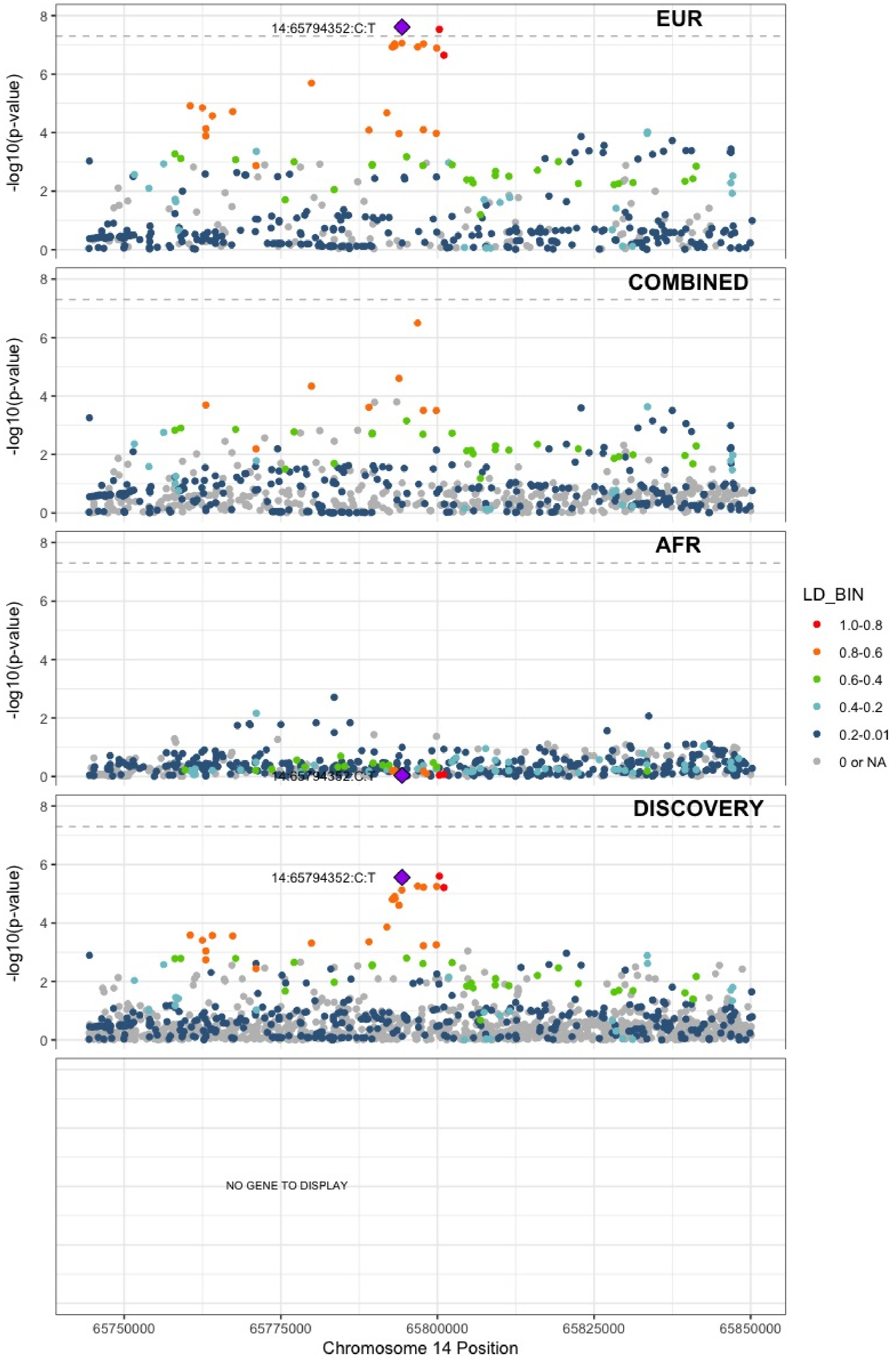

**Supplementary Figure 7.72** – rs61990092 (chr14:92289301) in locus 97 (TC2N)

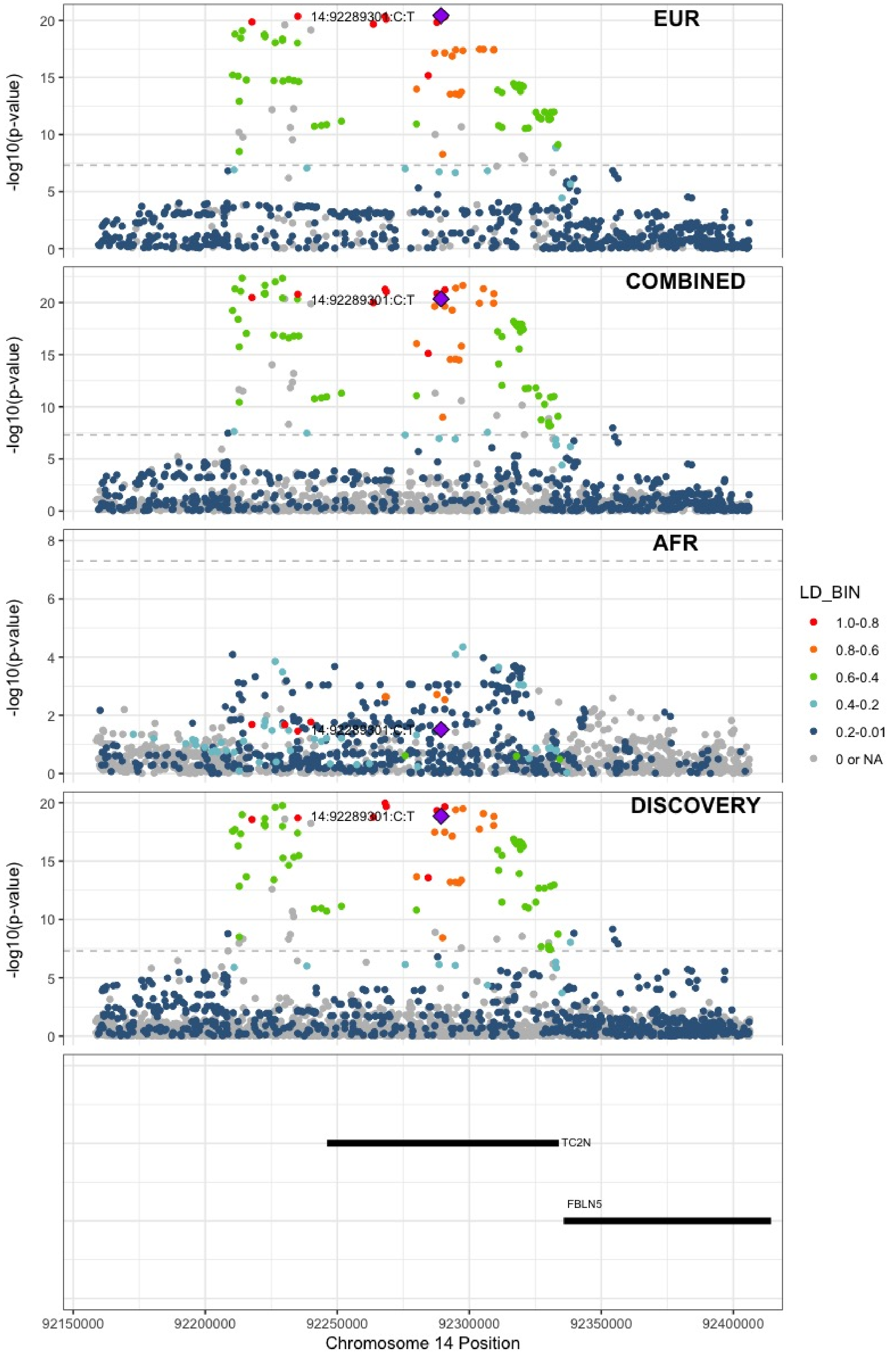

**Supplementary Figure 7.73** – rs28929474 (chr14:94844947) in locus 98 (SERPINA1)

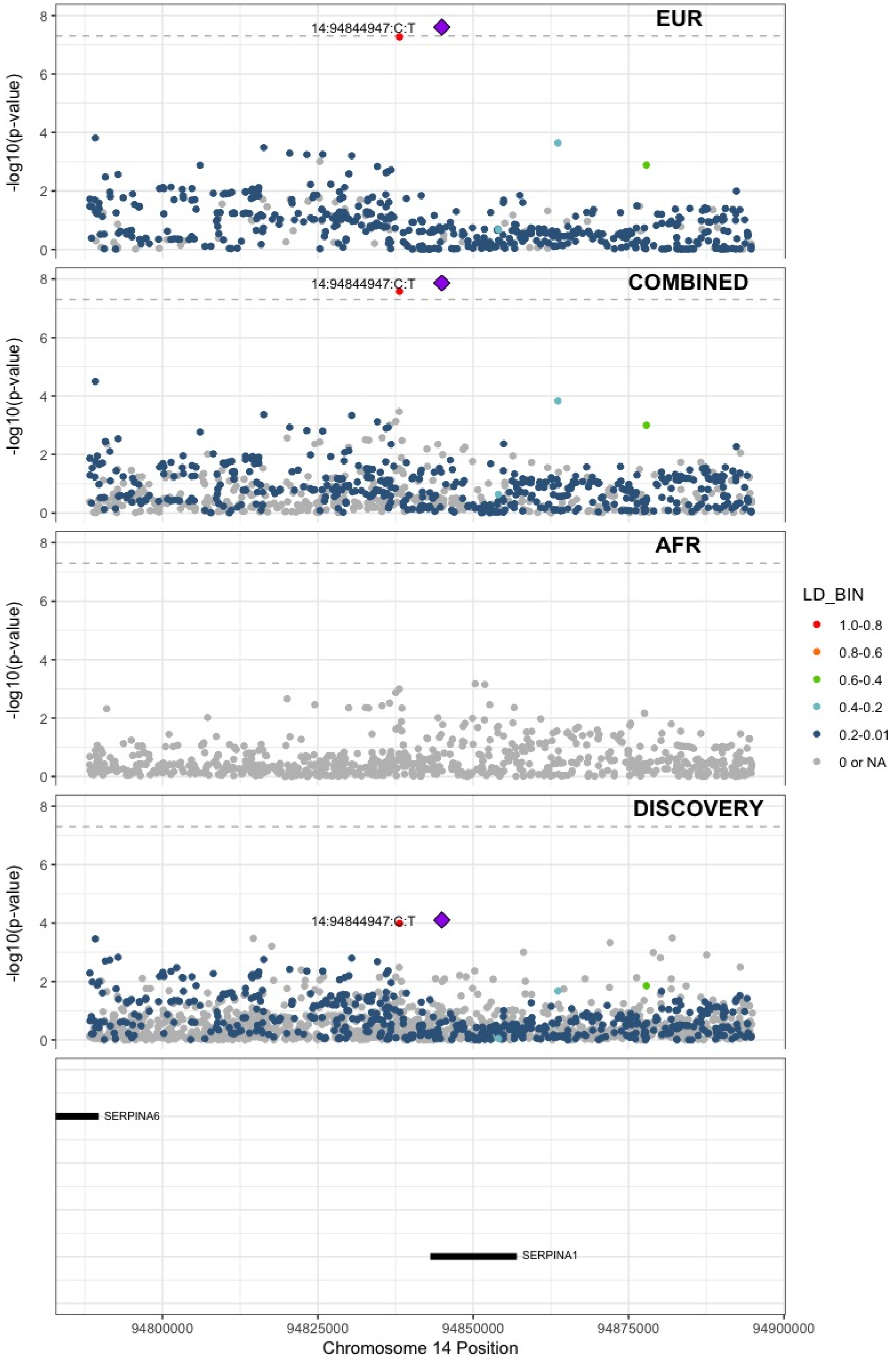

**Supplementary Figure 7.74** – rs4906229 (chr14:103057511) in locus 99 (LINC02323;RCOR1)

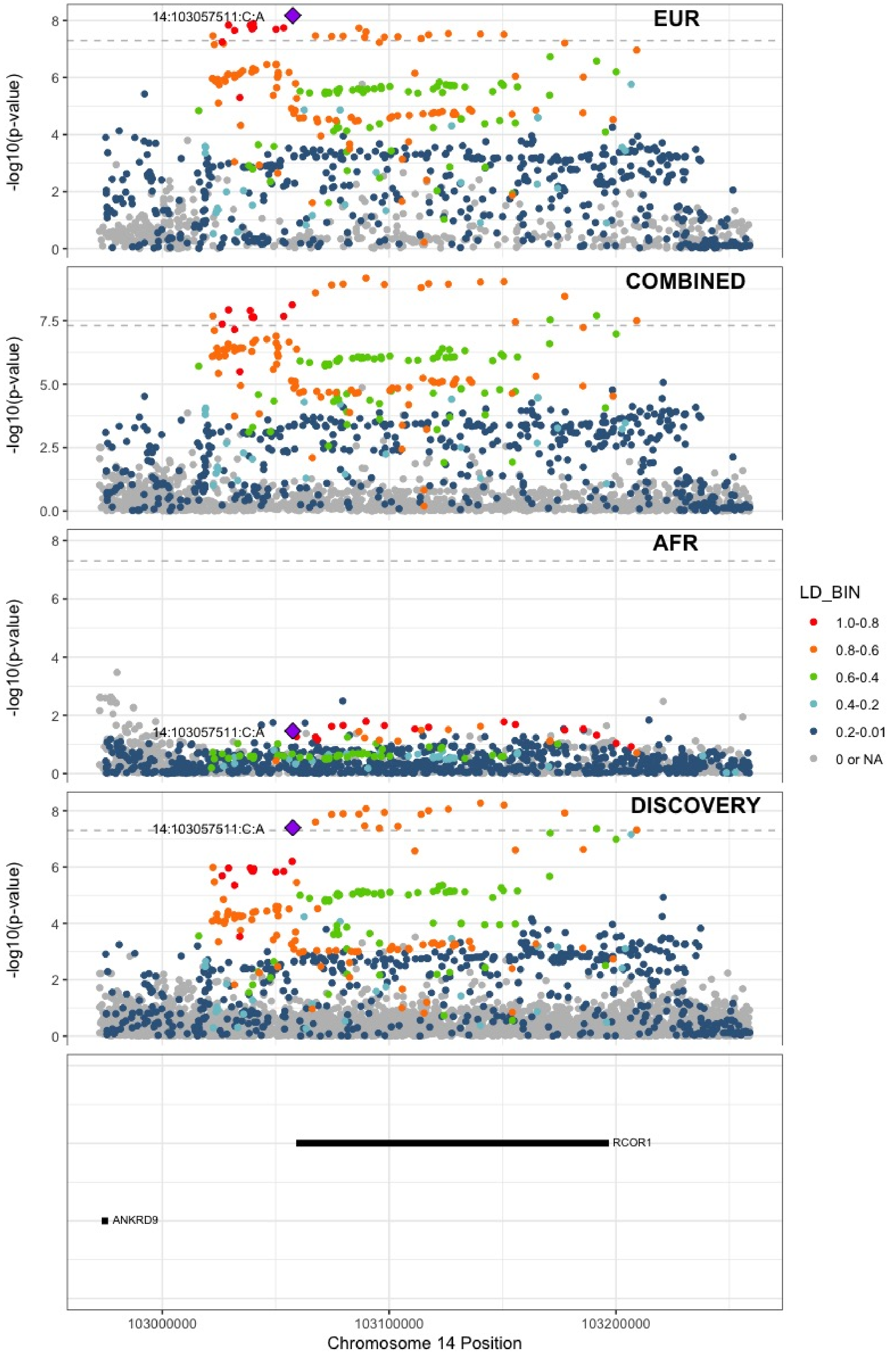

**Supplementary Figure 7.75** – rs141866277 (chr15:43950699) in locus 100 (PPIP5K1P1-CATSPER2)

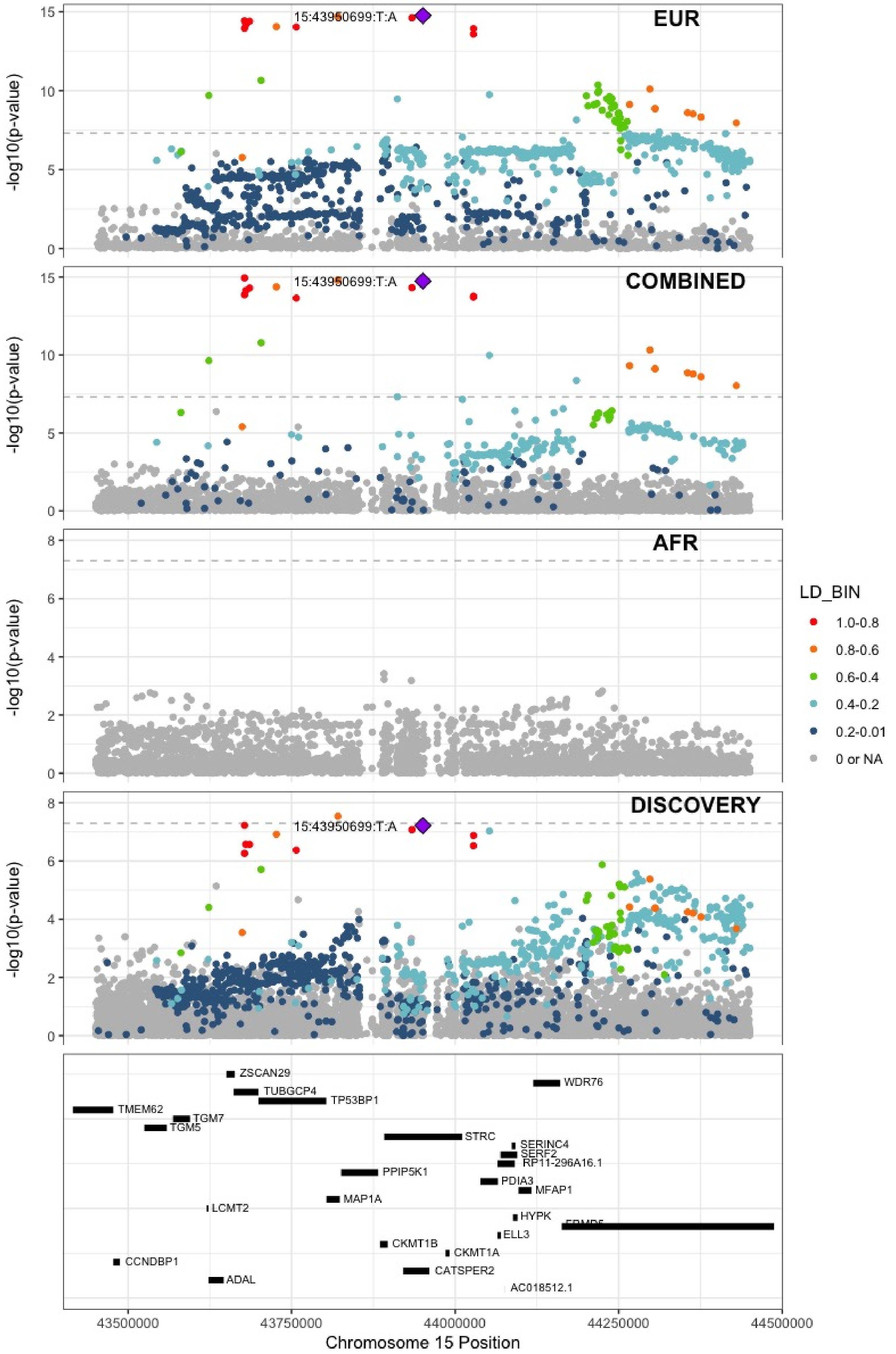

**Supplementary Figure 7.76** – rs340009 (chr15:60899639) in locus 101 (RORA-AS1)

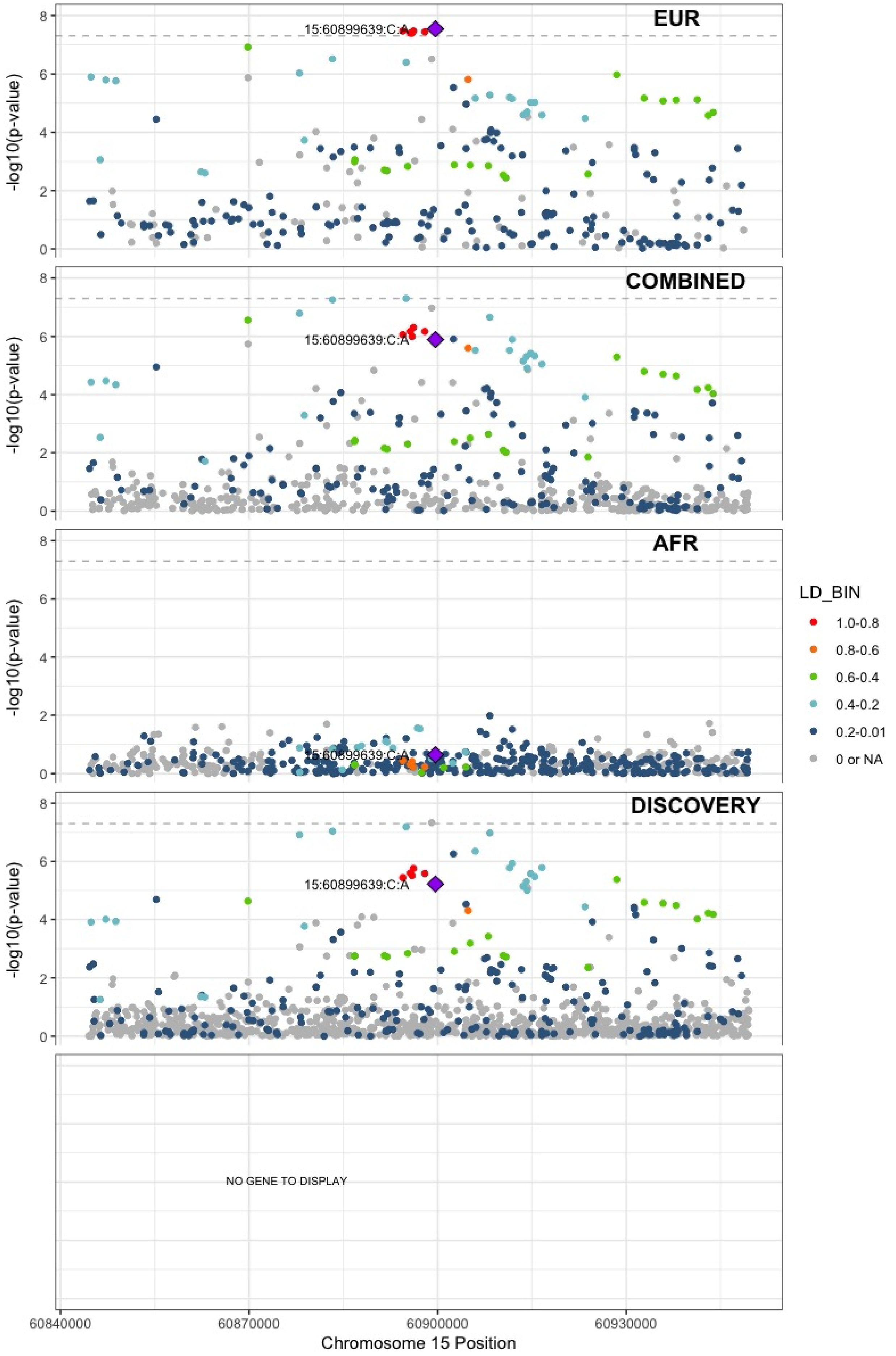

**Supplementary Figure 7.77** – rs7183672 (chr15:96101018) in locus 103 (LINC00924 ;LOC105369212)

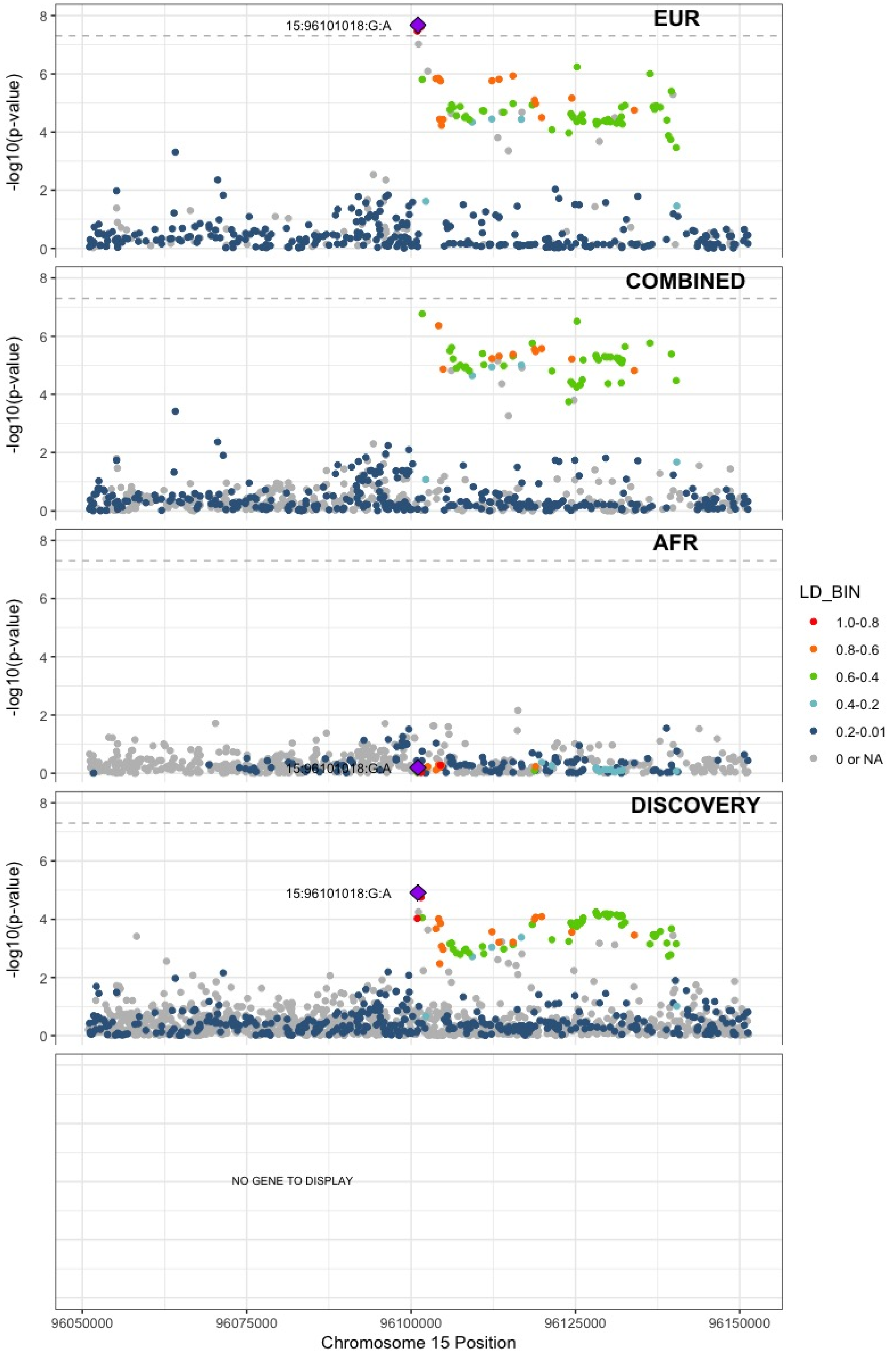

**Supplementary Figure 7.78** – rs12443808 (chr16:30996871) in locus 106 (HSD3B7)

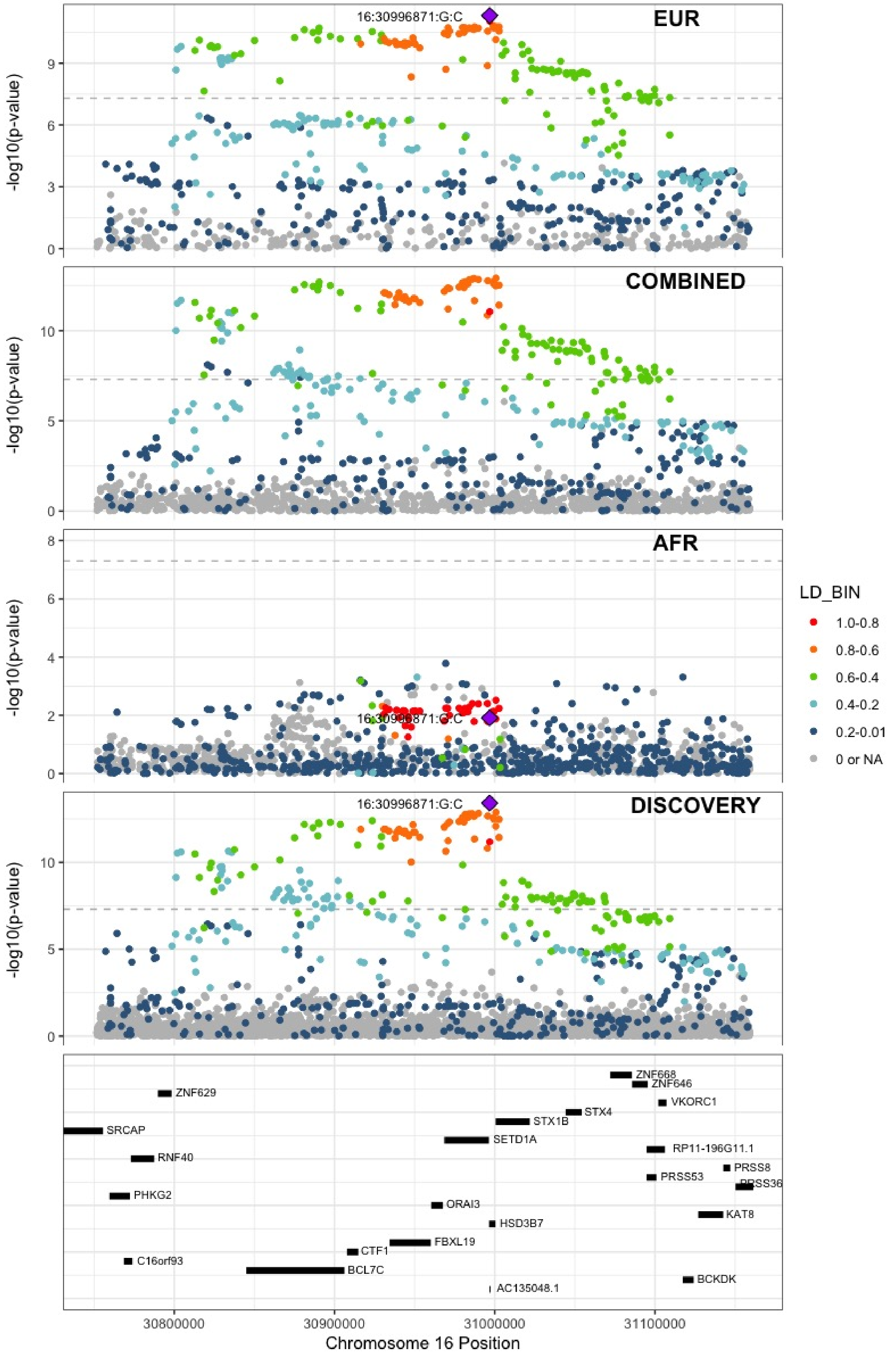

**Supplementary Figure 7.79** – rs1346057617 (chr16:75434183) in locus 108 (CFDP1)

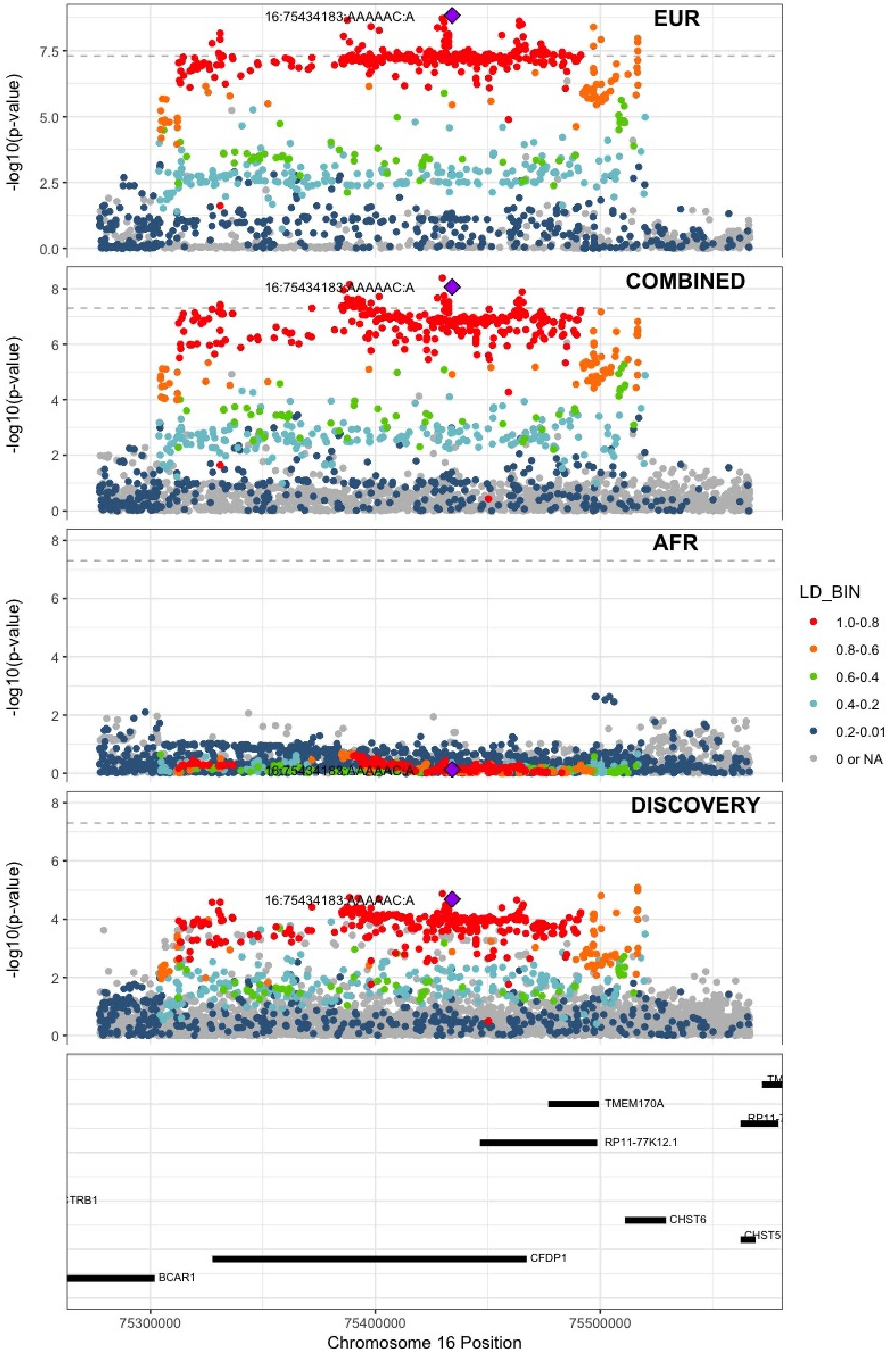

**Supplementary Figure 7.80** – rs62044142 (chr16:81868968) in locus 109 (PLCG2)

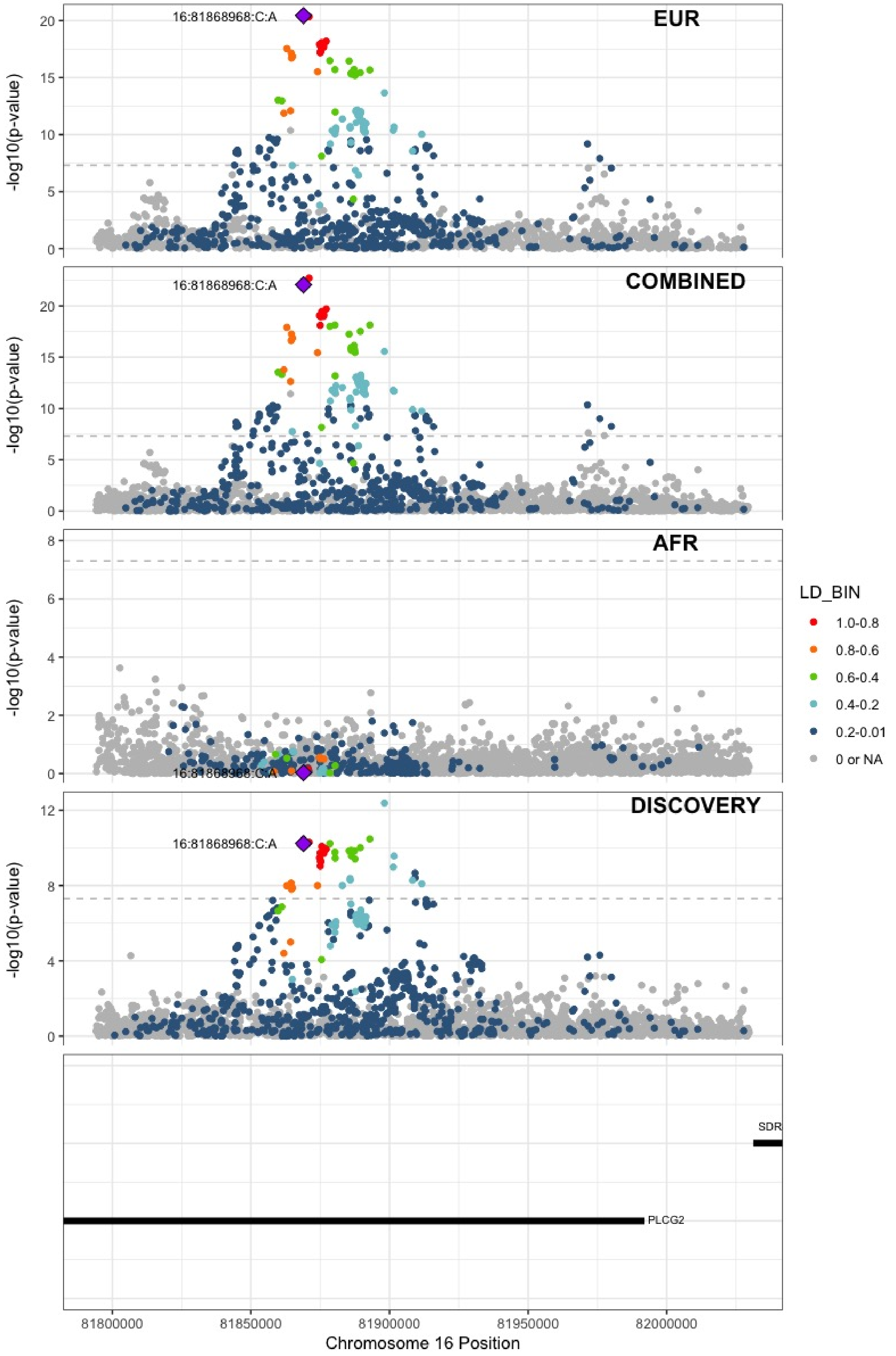

**Supplementary Figure 7.81** – rs28634651 (chr16:88553198) in locus 111 (ZFPM1)

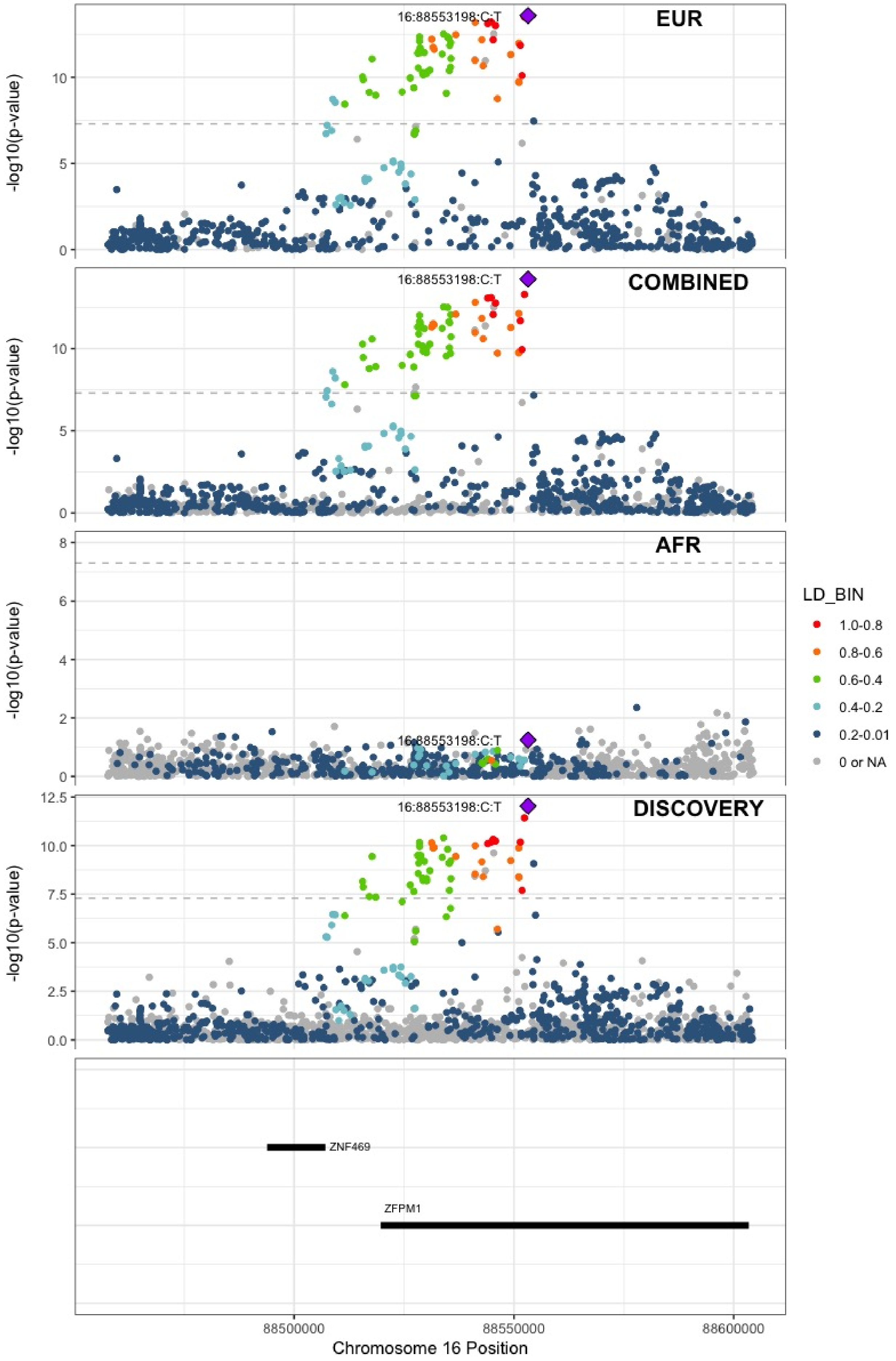

**Supplementary Figure 7.82** – rs4790311 (chr17:1979188) in locus 112 (SMG6)

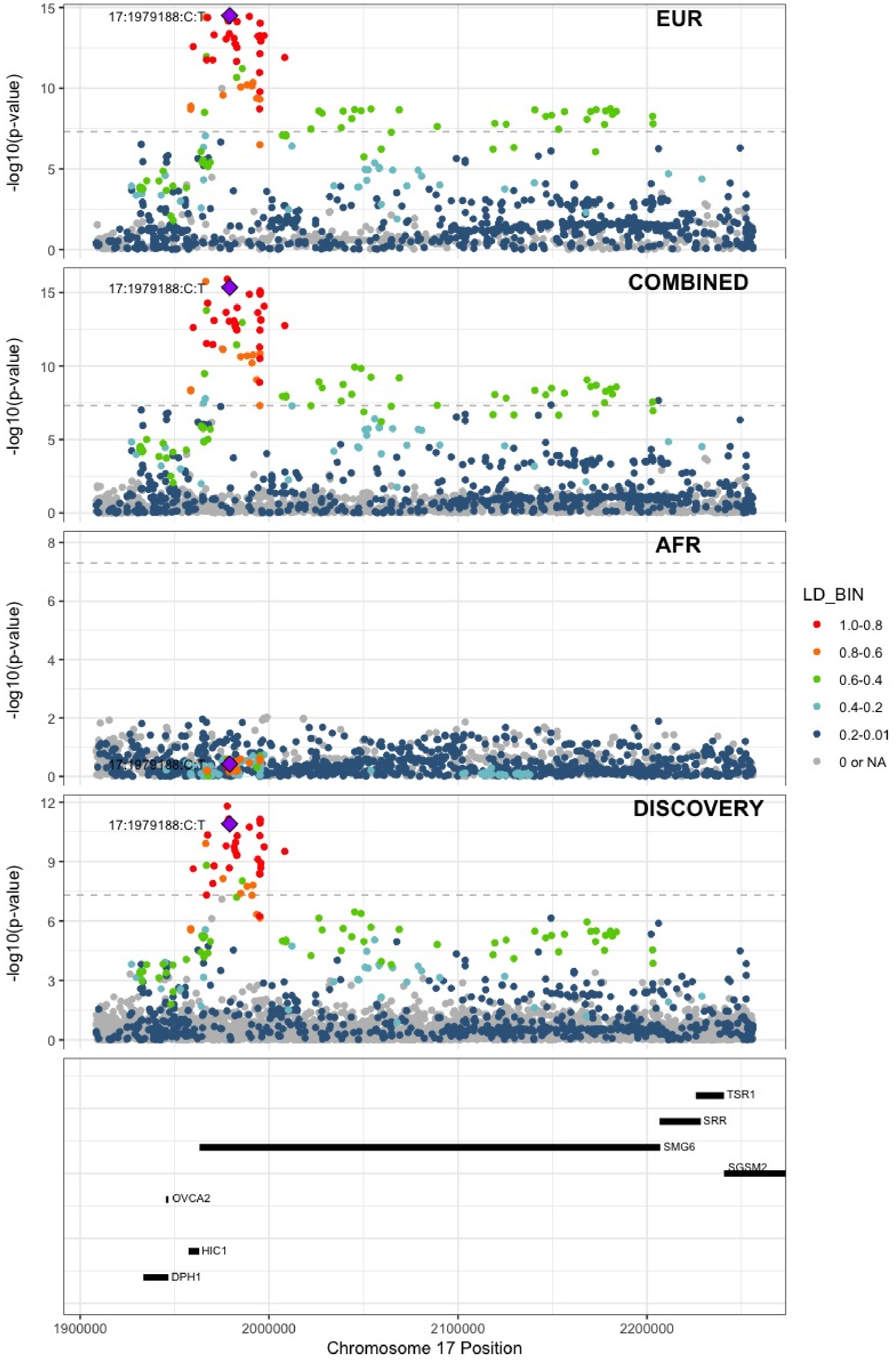

**Supplementary Figure 7.83** – rs11078659 (chr17:6903944) in locus 113 (ALOX12-AS1)

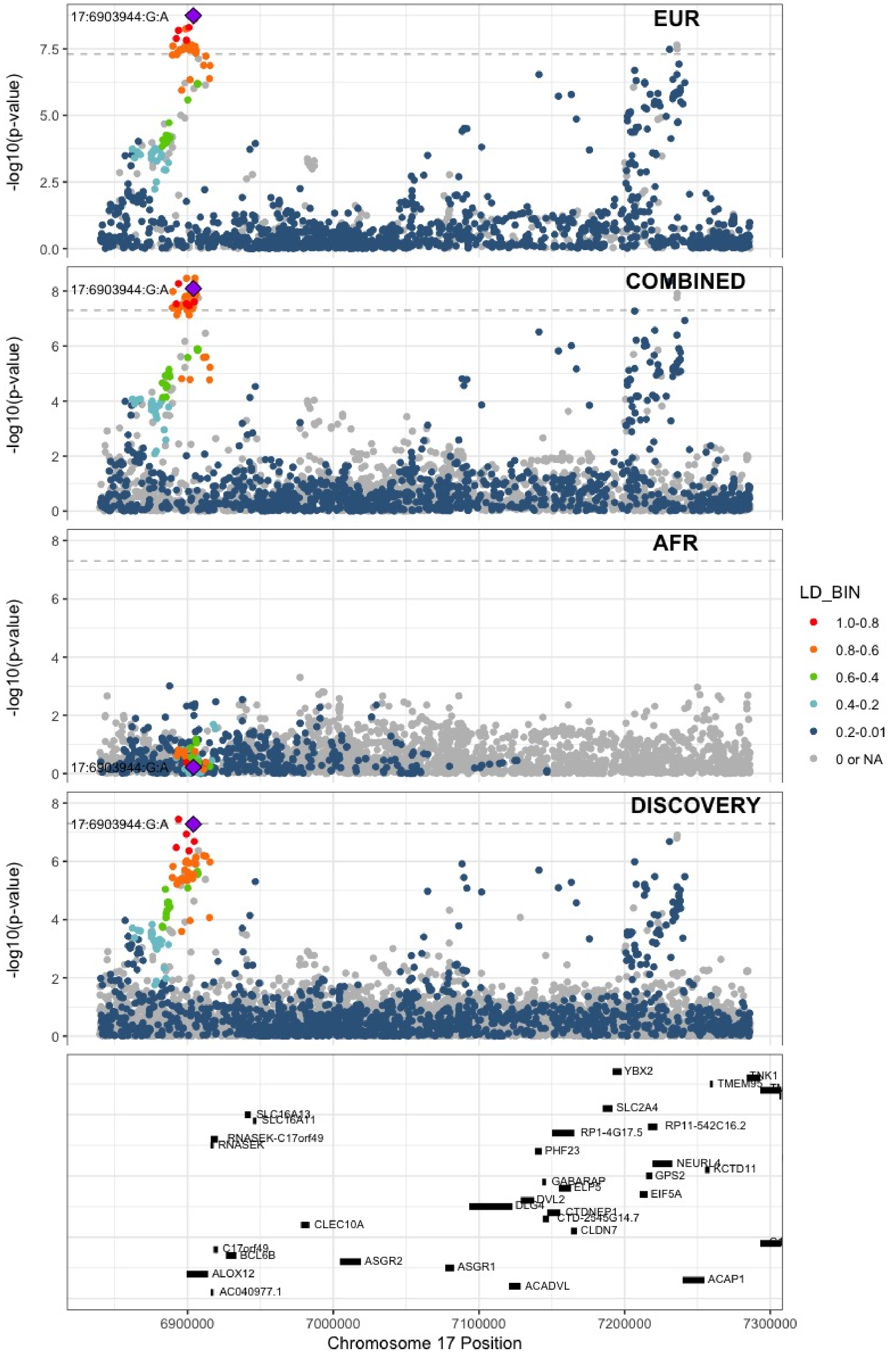

**Supplementary Figure 7.84** – rs536327 (chr17:27857698) in locus 114 (TAOK1)

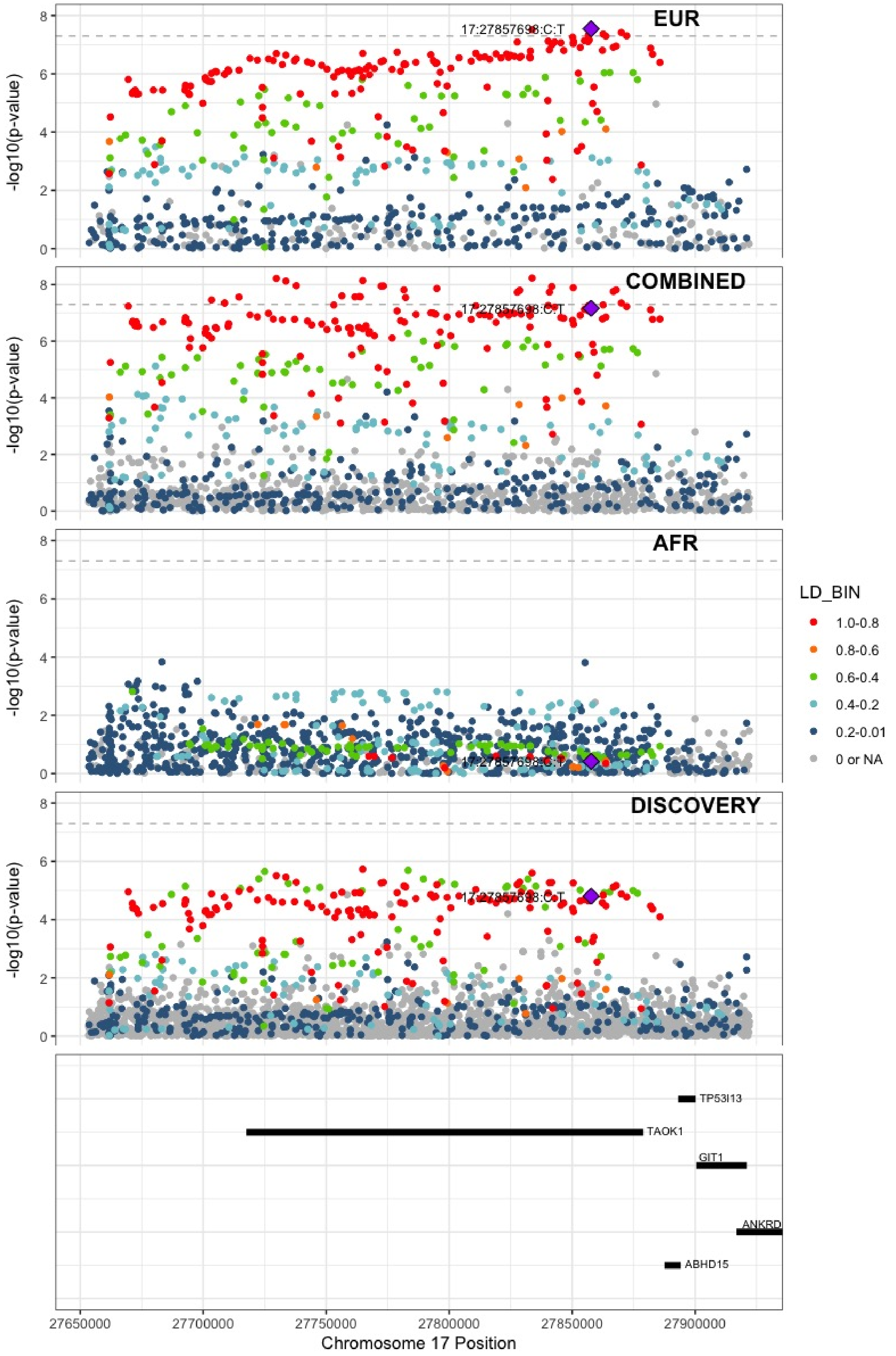

**Supplementary Figure 7.85** – rs9468 (chr17:44101563) in locus 115 (MAPT)

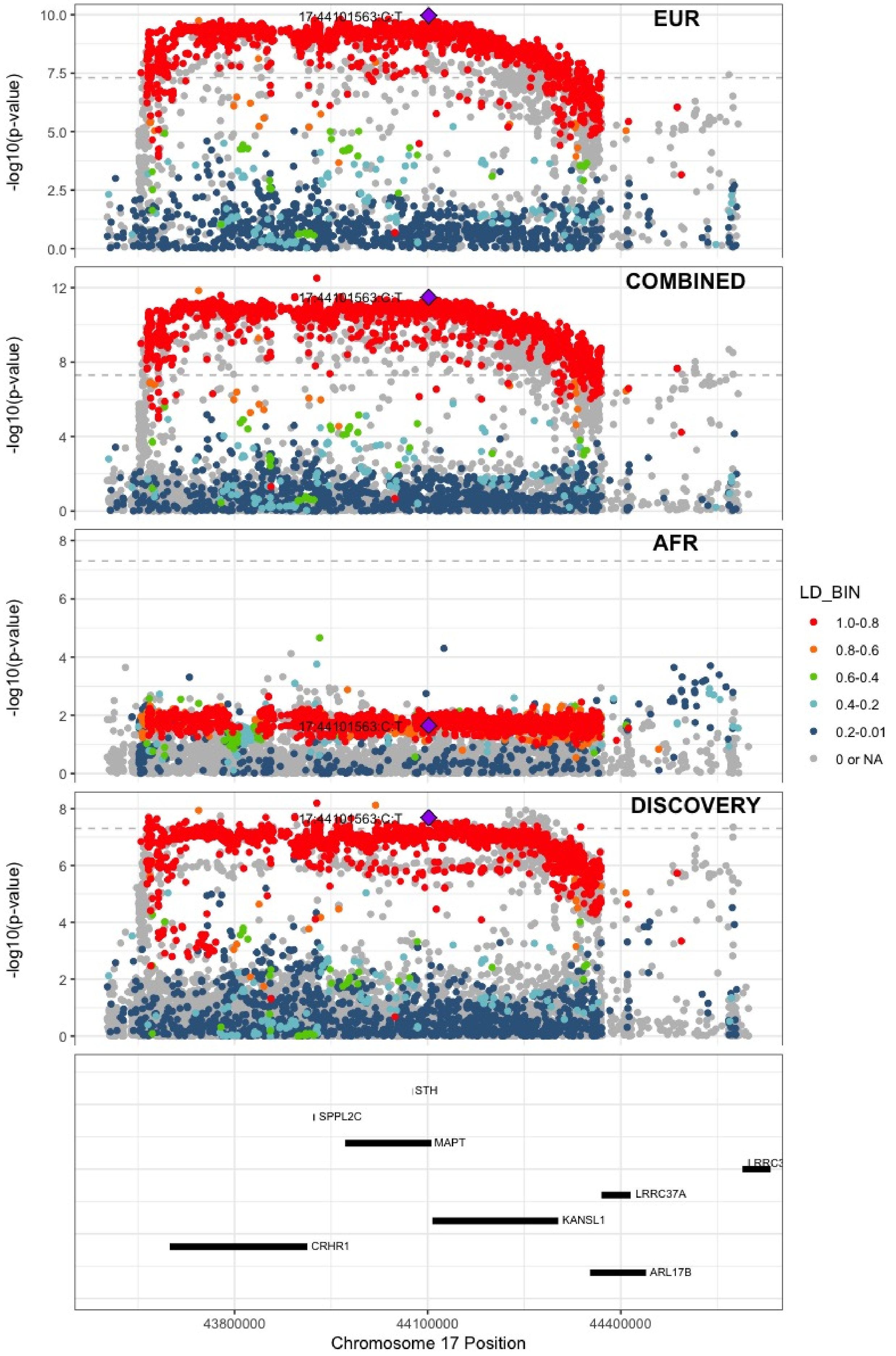

**Supplementary Figure 7.86** – rs1801690 (chr17:64208285) in locus 116 (APOH)

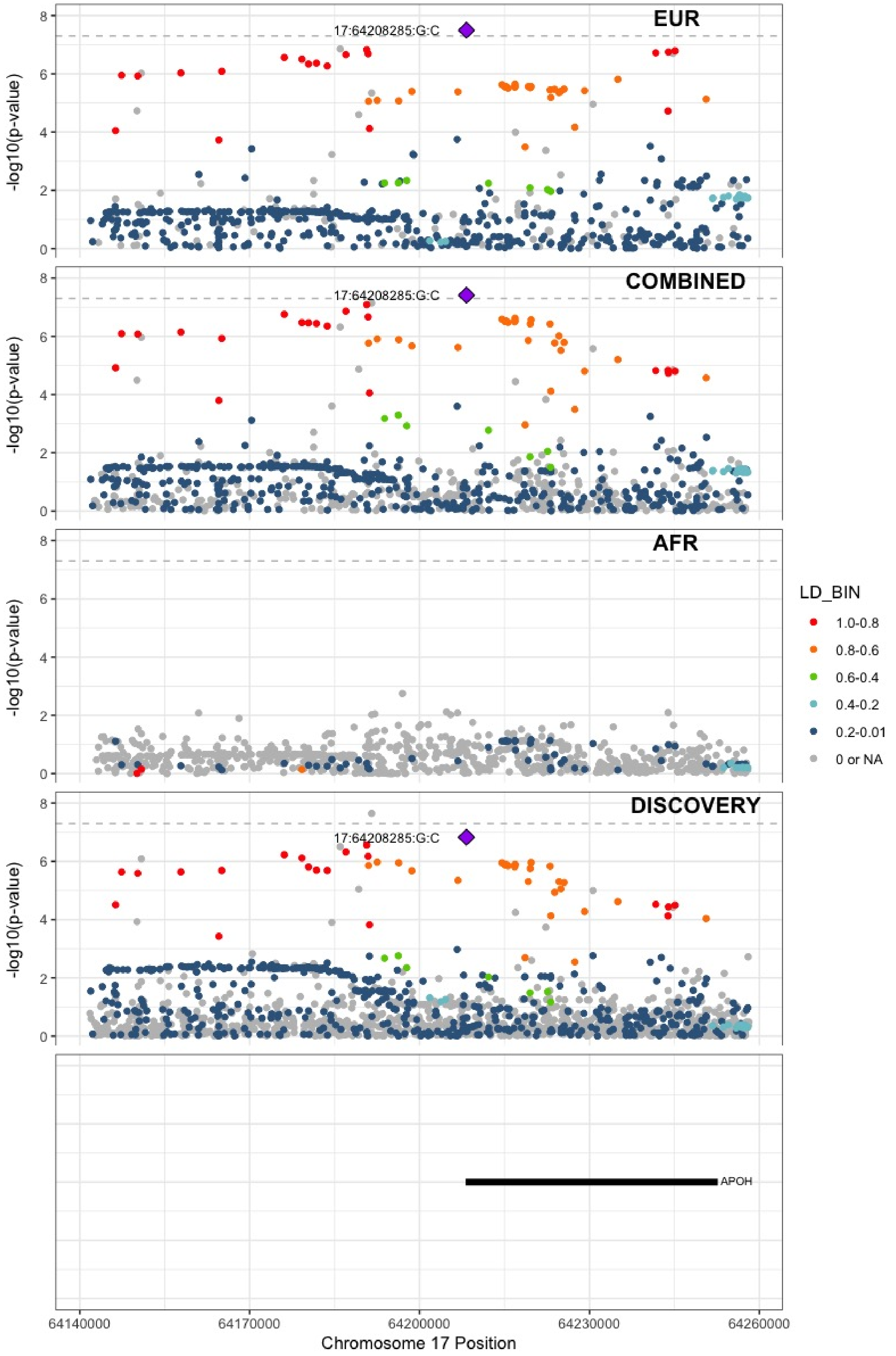

**Supplementary Figure 7.87** – rs573431210 (chr17:67276383) in locus 117 (ABCA5)

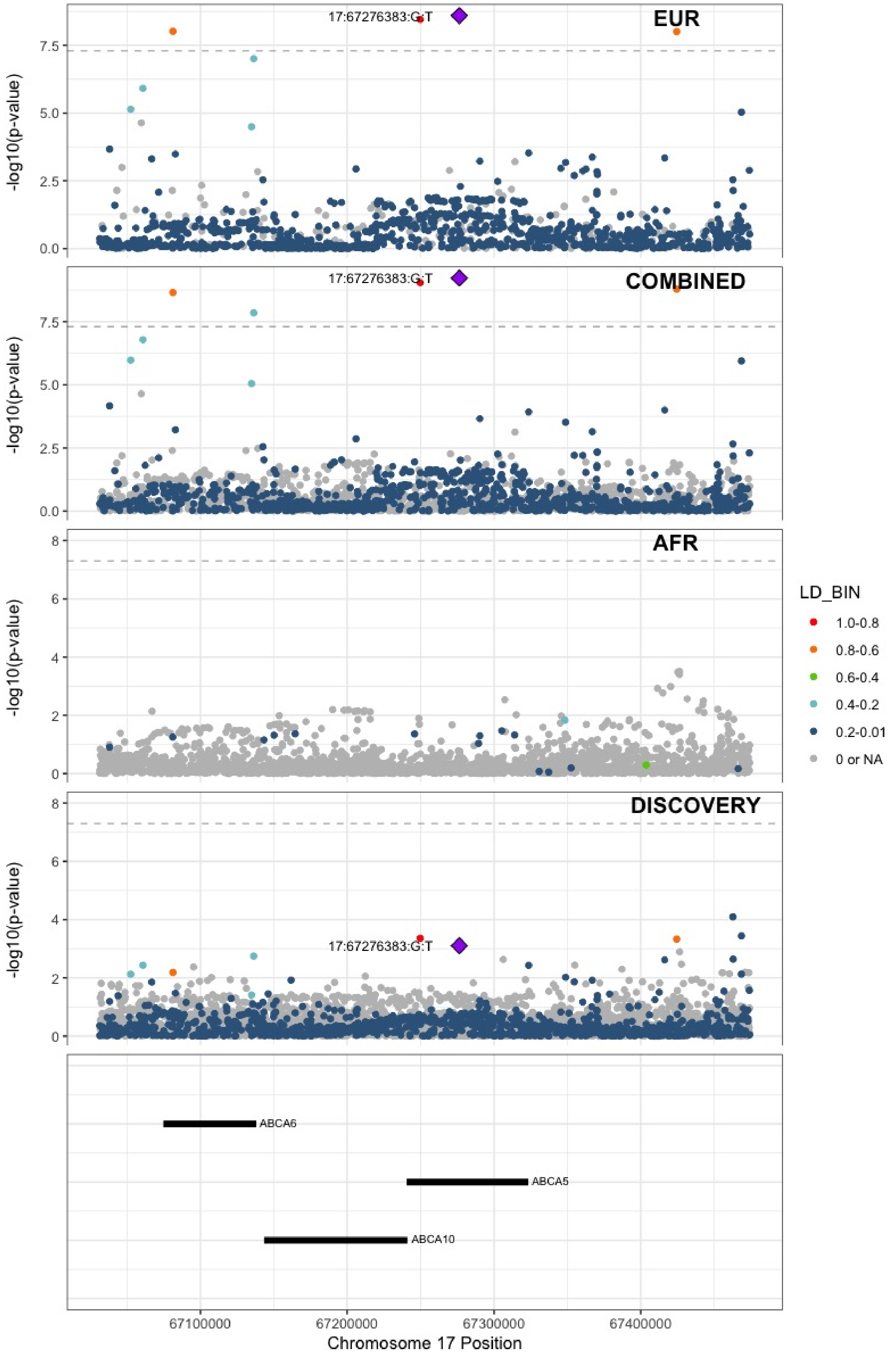

**Supplementary Figure 7.88** – rs8109681 (chr19:10738836) in locus 119 (SLC44A2)

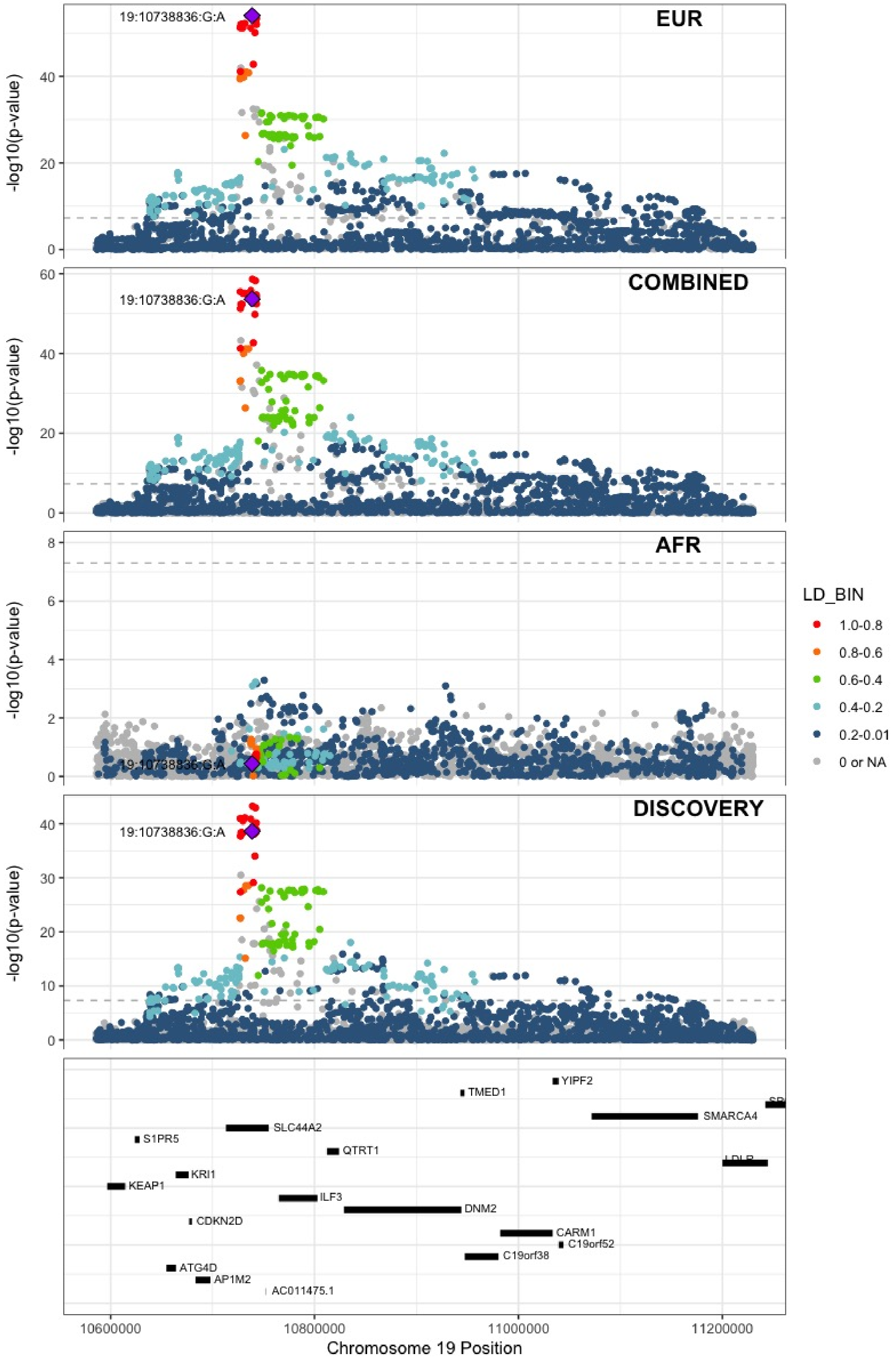

**Supplementary Figure 7.89** – rs2545774 (chr19:41287674) in locus 121 (RAB4B)

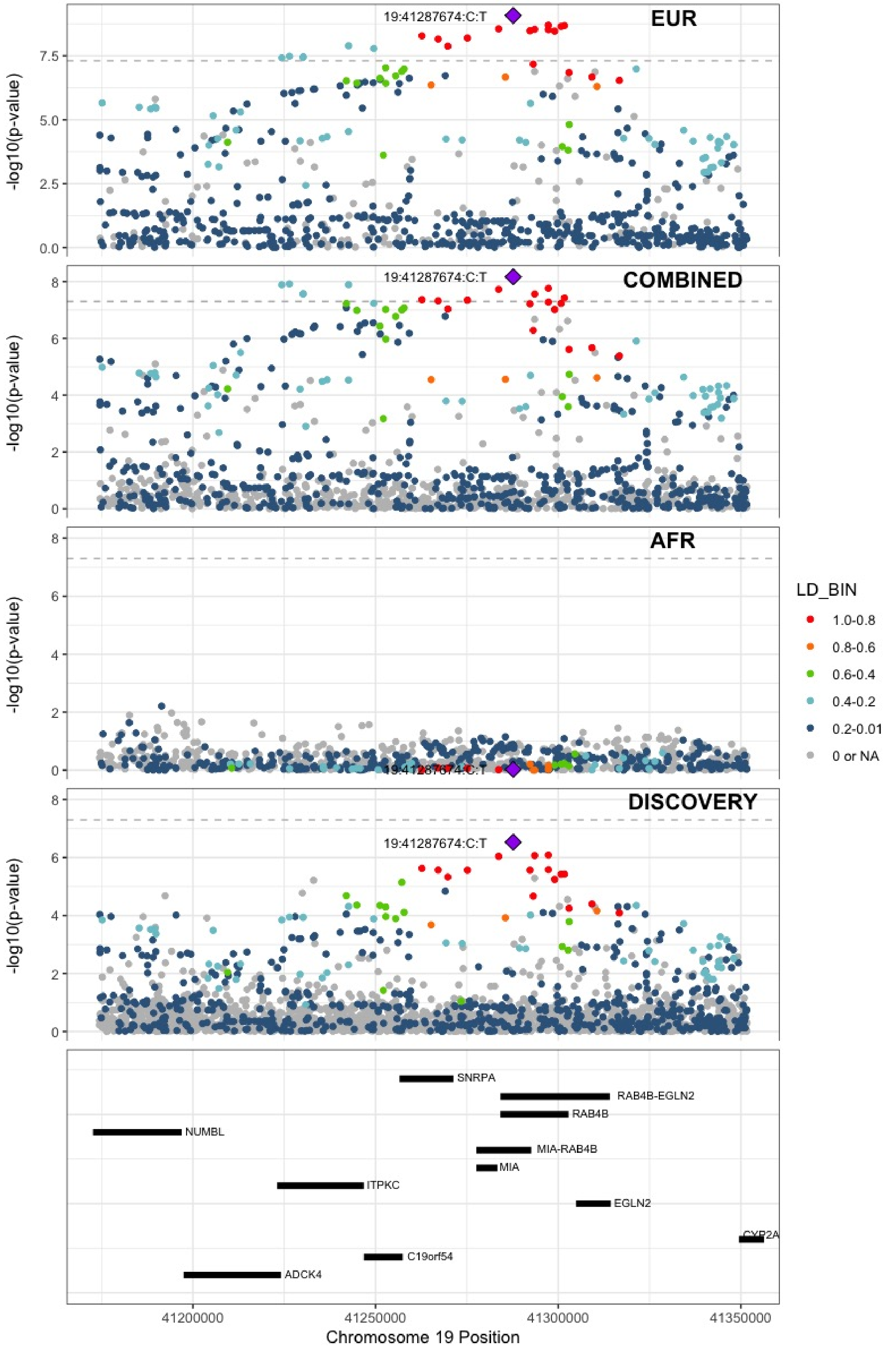

**Supplementary Figure 7.90** – rs8108474 (chr19:46301479) in locus 122 (RSPH6A)

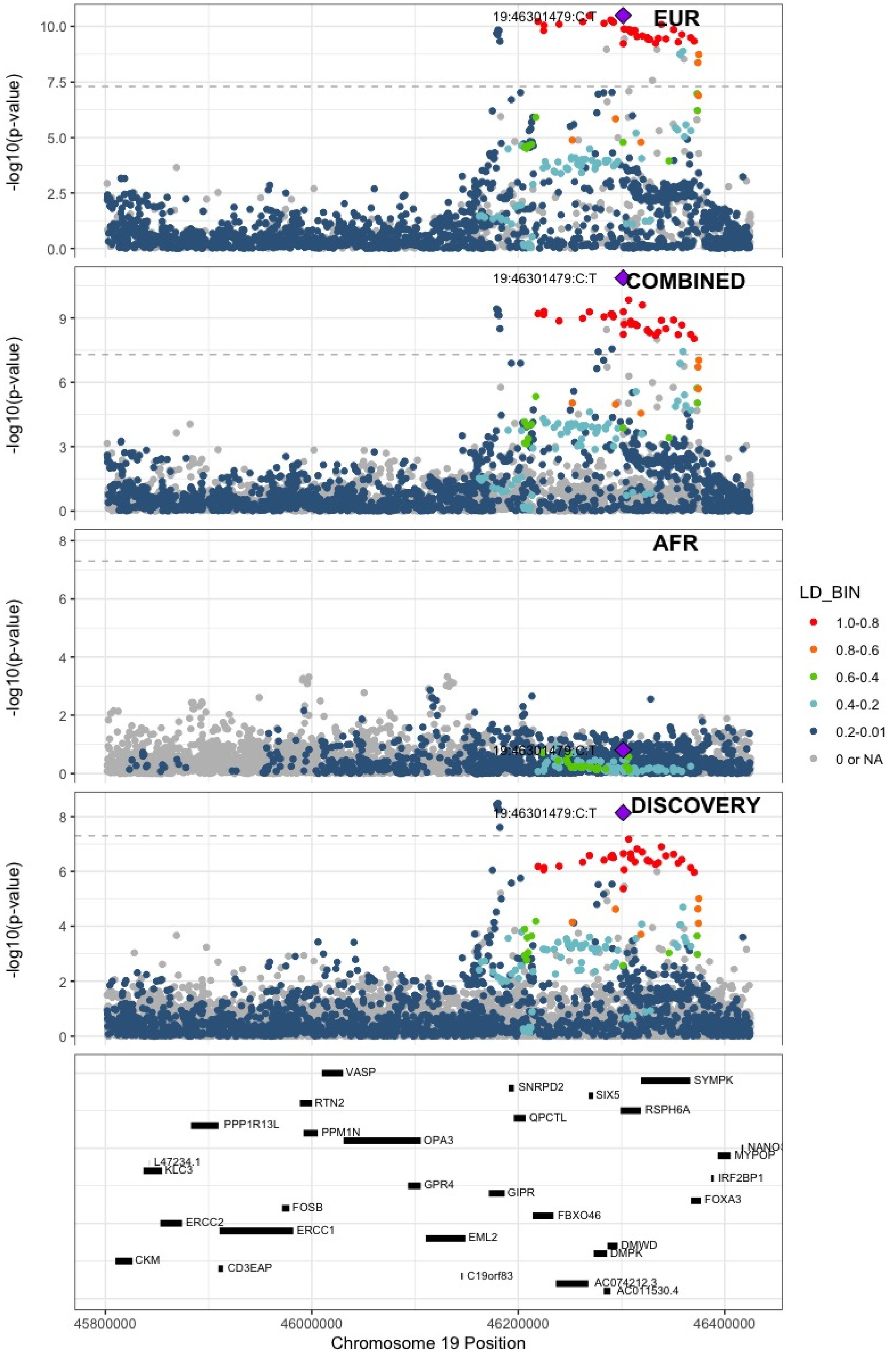

**Supplementary Figure 7.91** – rs646327 (chr19:49209851) in locus 123 (FUT2)

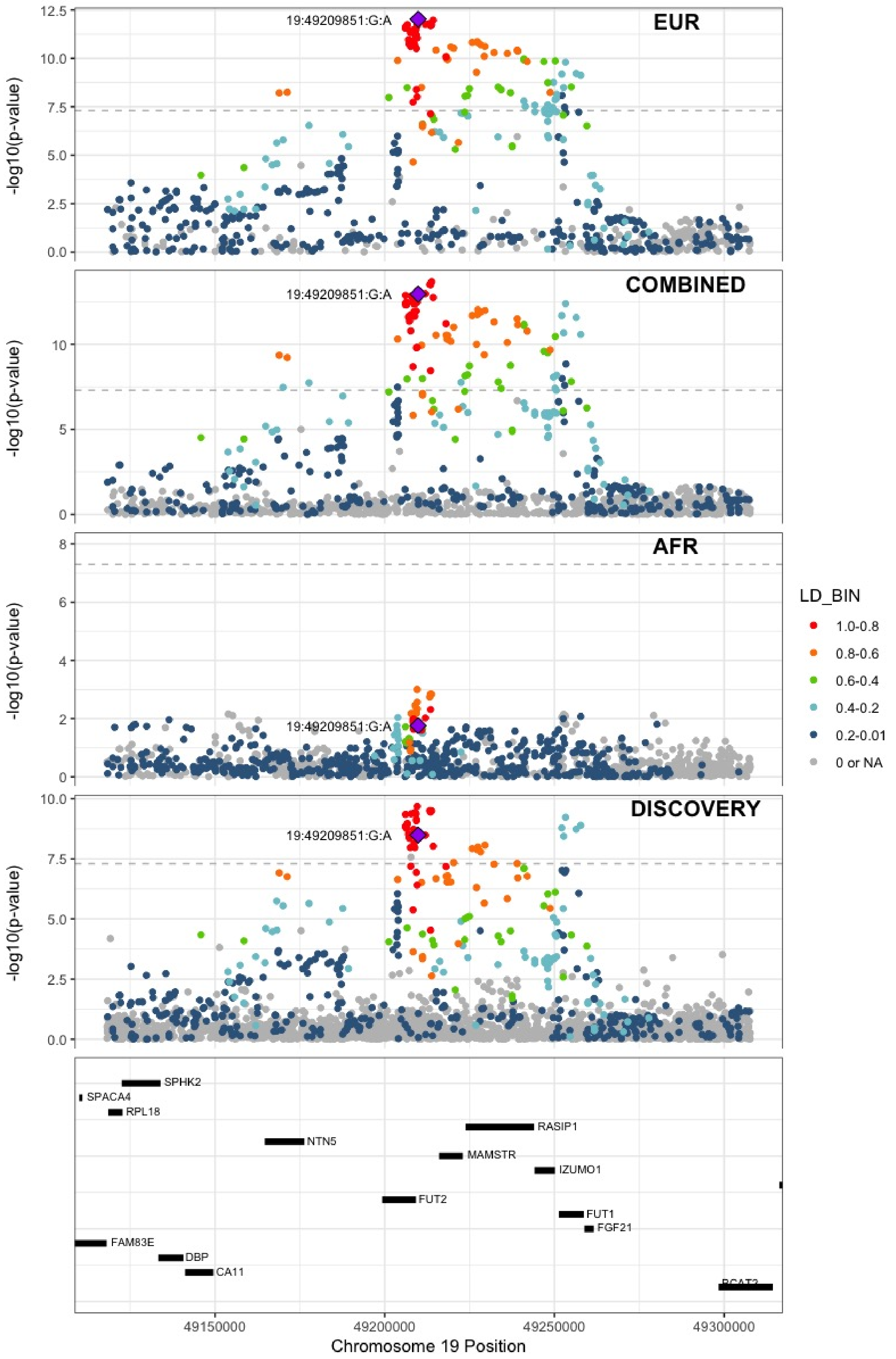

**Supplementary Figure 7.92** – rs1654425 (chr19:55538980) in locus 124 (GP6)

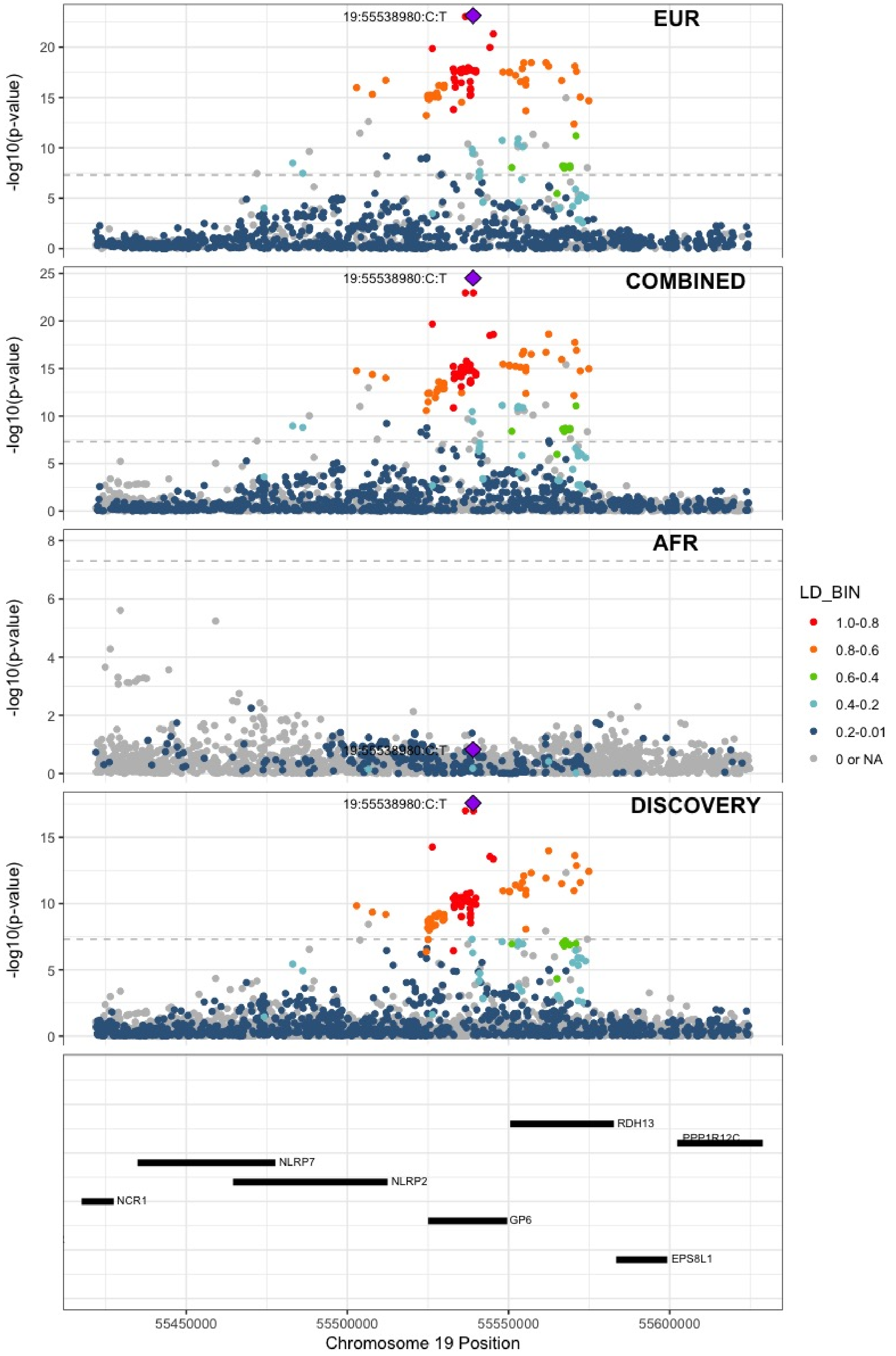

**Supplementary Figure 7.93** – rs117390891 (chr20:23168526) in locus 126 (LINC00656 ;NXT1)

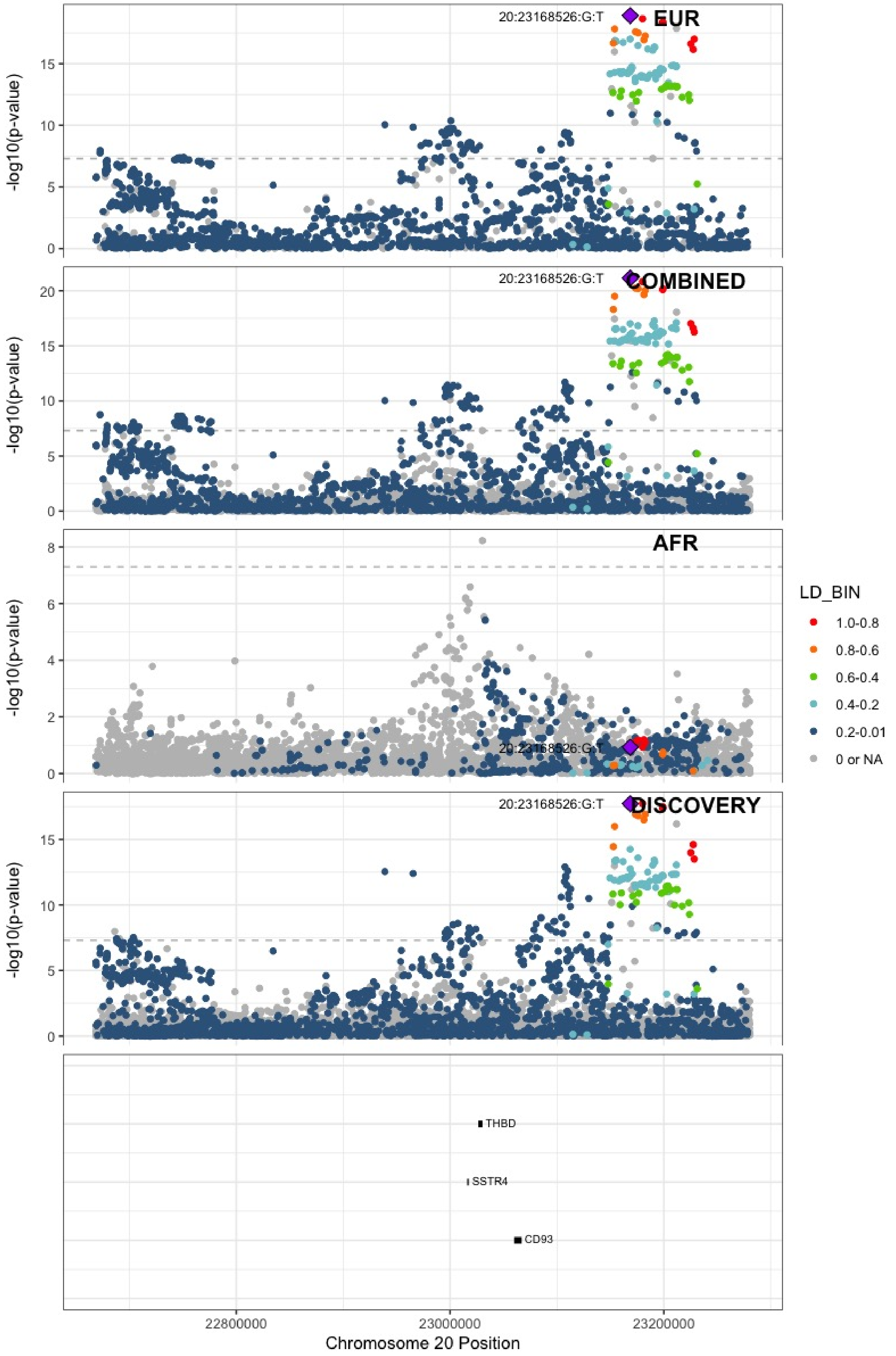

**Supplementary Figure 7.94** – rs6087685 (chr20:33777612) in locus 127 (MMP24-AS1-EDEM2)

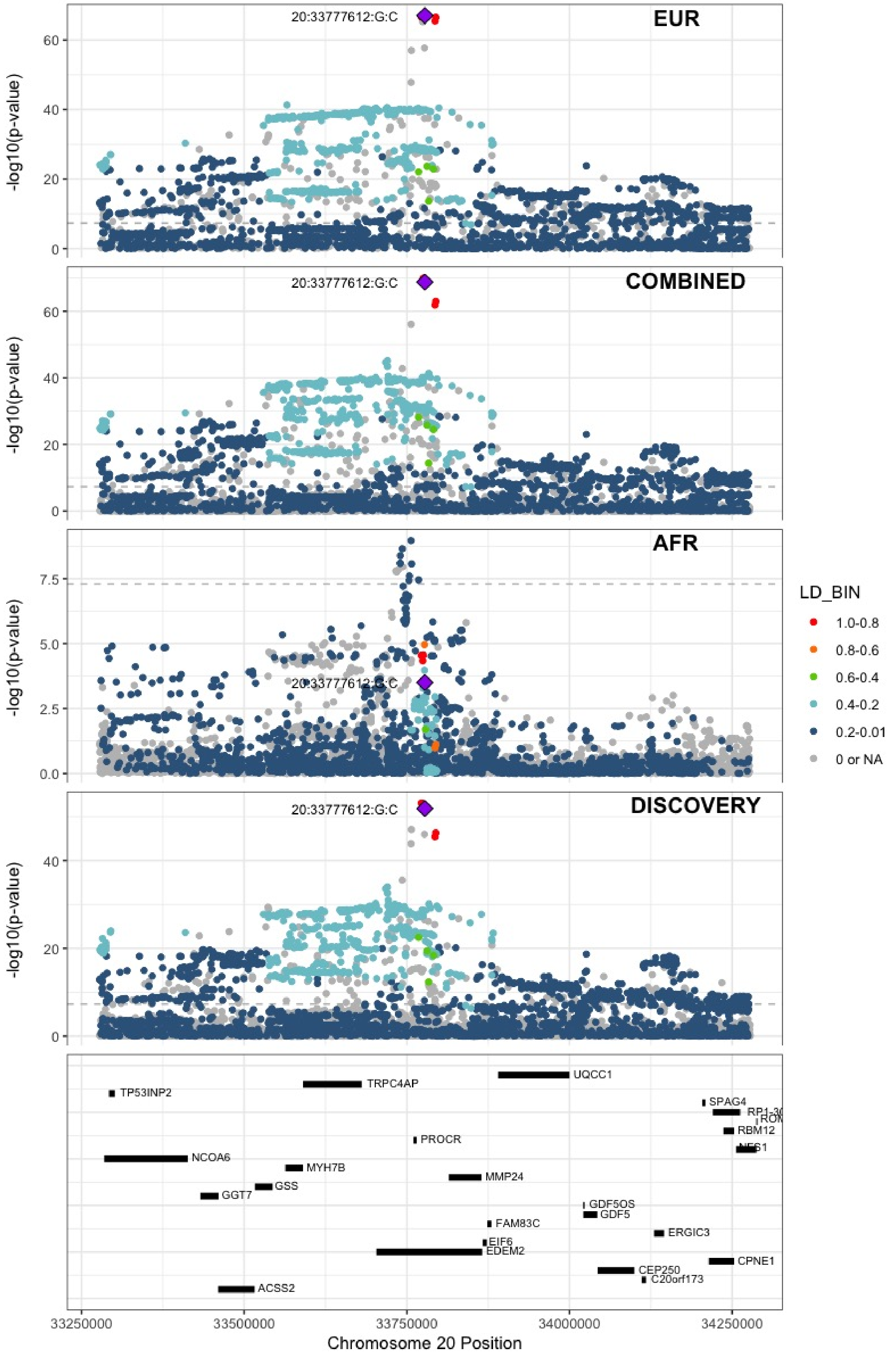

**Supplementary Figure 7.95** – rs3070580 (chr22:33165231) in locus 129 (SYN3)

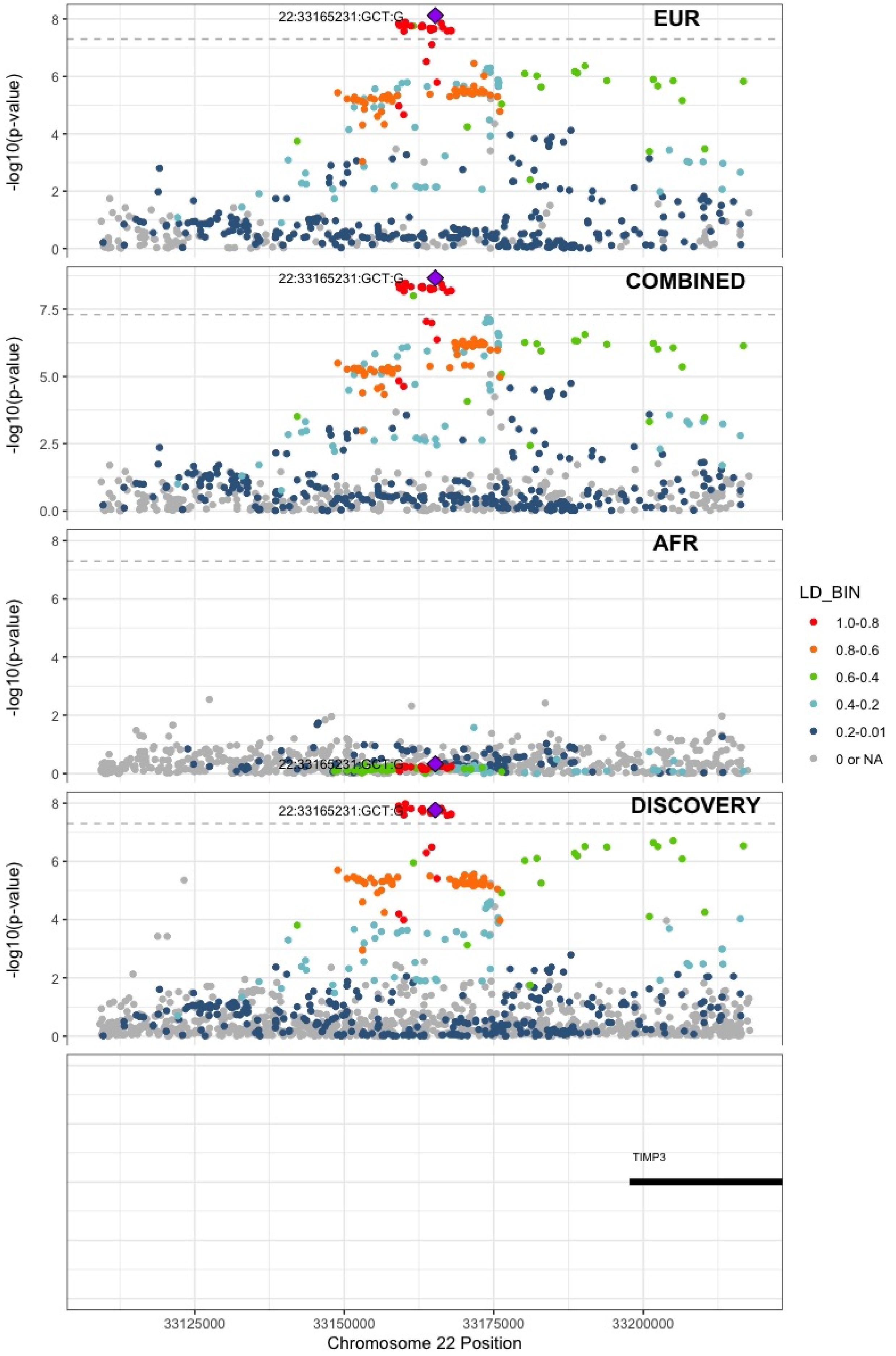

**Supplementary Figure 7.96** – rs9611844 (chr22:43115776) in locus 130 (A4GALT)

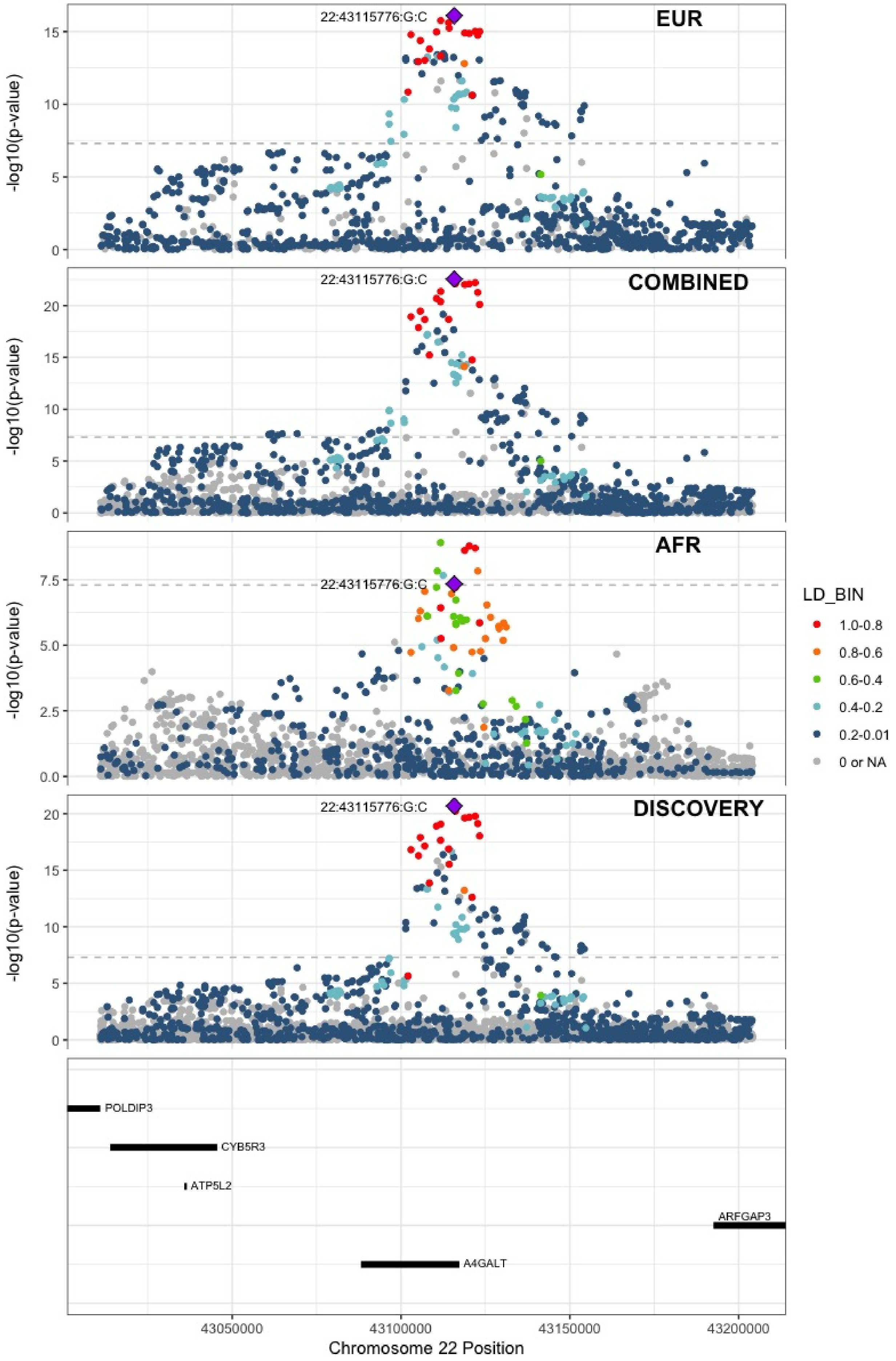

**Supplementary Figure 7.97** – rs12388118 (chr23:11380742) in locus 131 (ARHGAP6)

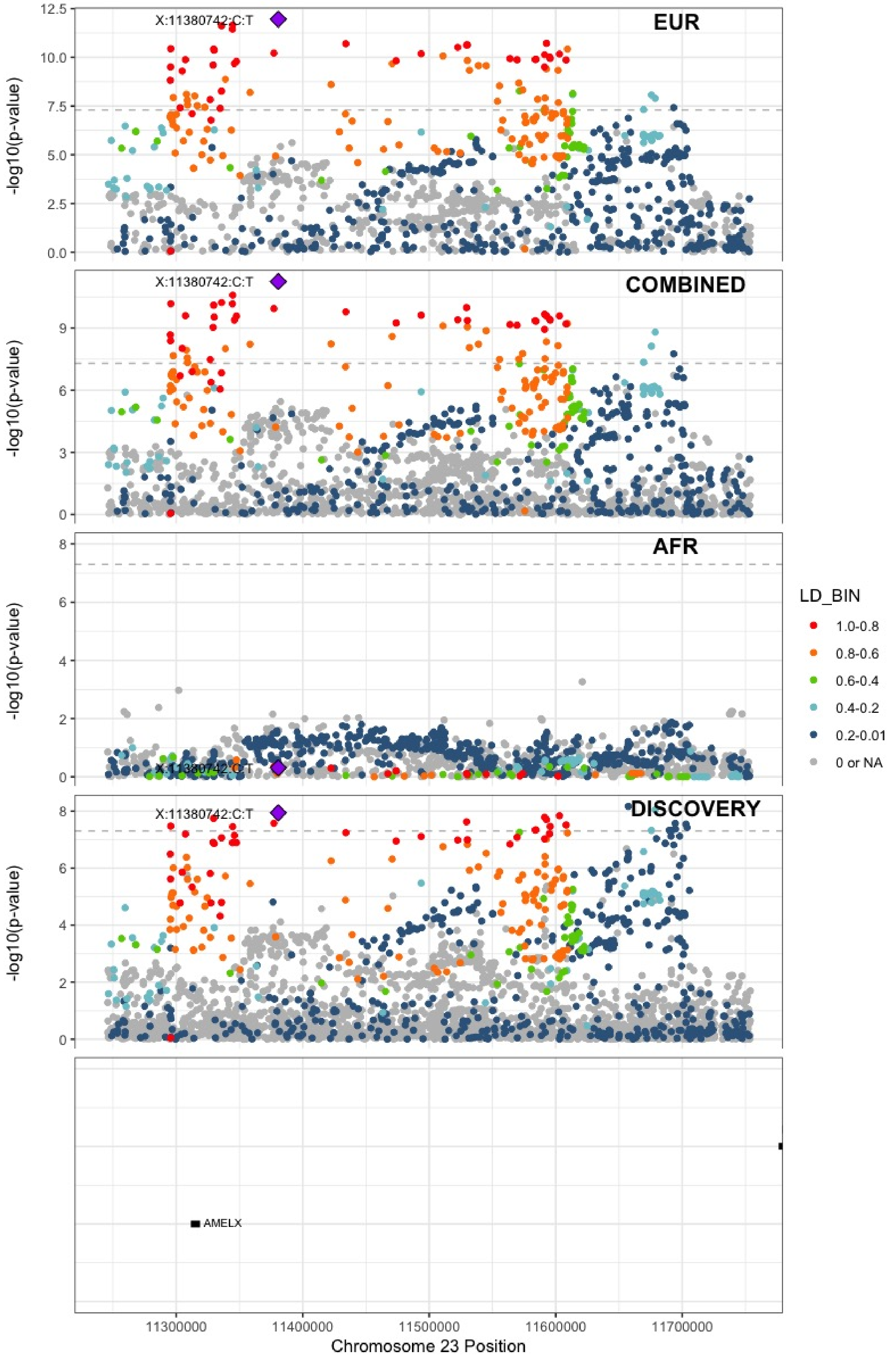

**Supplementary Figure 7.98** – rs3002416 (chr23:39710195) in locus 133 (MIR1587;BCOR)

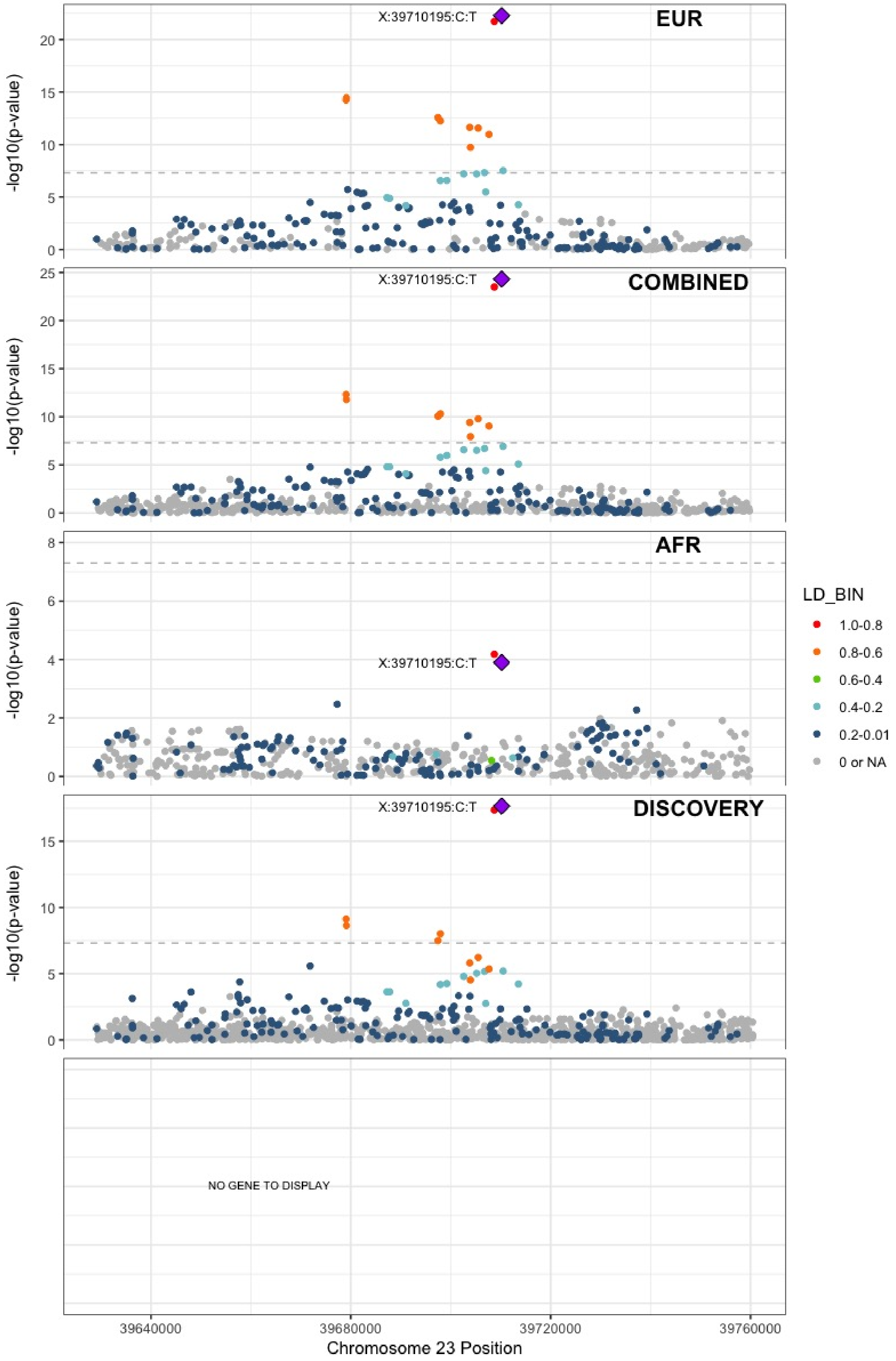

**Supplementary Figure 7.99** – rs6048 (chr23:138633280) in locus 134 (F9)

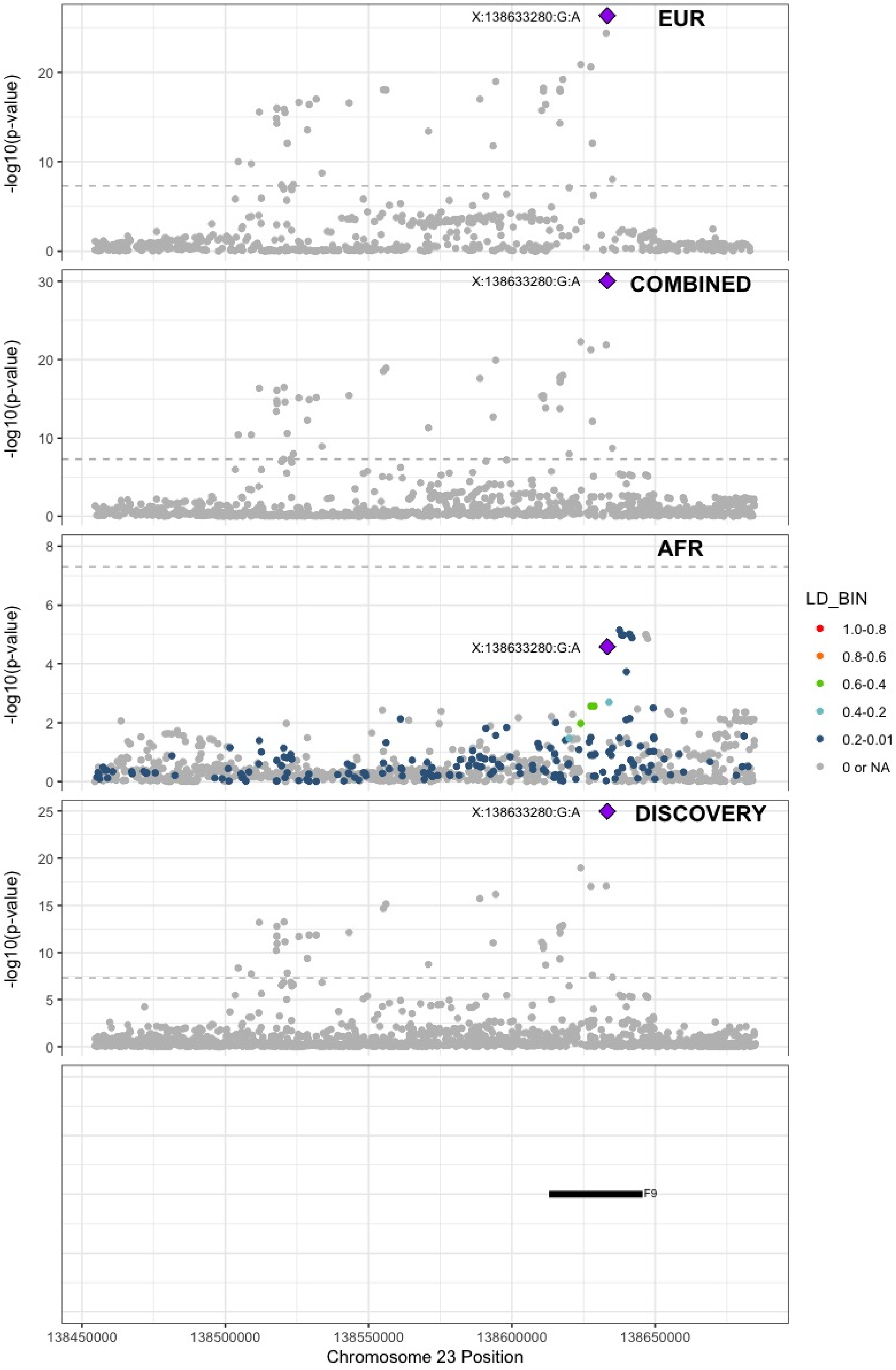

**Supplementary Figure 7.100** – rs4898406 (chr23:154273269) in locus 135 (FUNDC2)

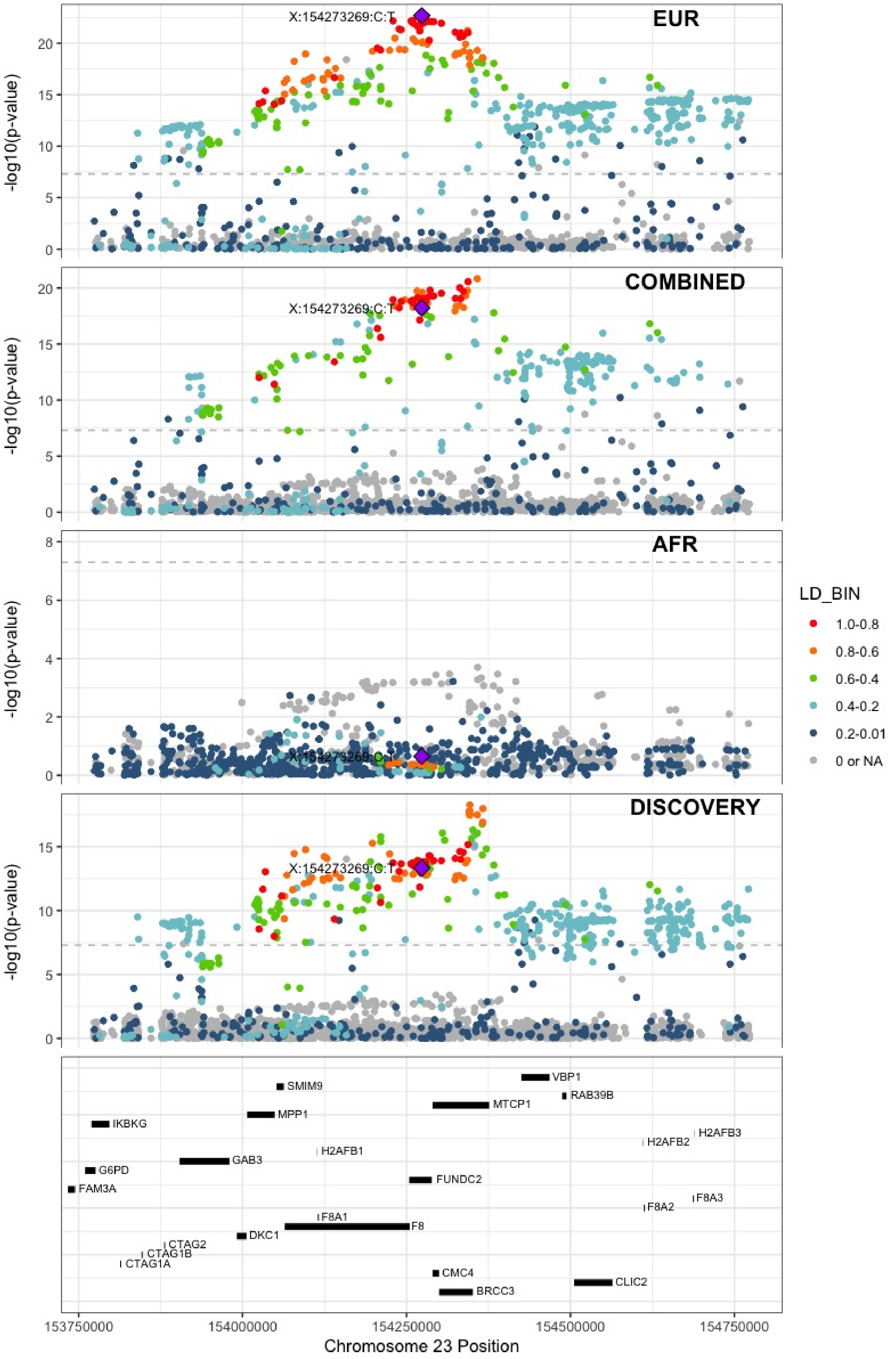
