## Supplemental Figure 8 for "Cross-Ancestry Investigation of Venous Thromboembolism Genomic Predictors"

### Supplementary Figure 8.
### Regional association plots of African analysis lead variants.


8.1 rs10800427 (chr1:169272451) in locus 9 (NME7) . . . . . 2
8.2 rs182293241 (chr2:128029746) in locus 18 (ERCC3) . . . . . 3
8.3 rs529326506 (chr3:90503764) in locus 26 (EPHA3;NONE) . . . . . 4
8.4 rs80133433 (chr3:93791551) in locus 27 (NSUN3) . . . . . 5
8.5 rs2066864 (chr4:155525695) in locus 39 (FGG) . . . . . 6
8.6 rs56810541 (chr4:187200550) in locus 40 (F11) . . . . . 7
8.7 rs8176719 (chr9:136132908) in locus 66 (ABO) . . . . . 8
8.8 rs57950734 (chr12:6145649) in locus 87 (VWF) . . . . . 9
8.9 rs76668186 (chr16:6686083) in locus 104 (RBFOX1) . . . . . 10
8.10 rs13306849 (chr20:23030199) in locus 126 (THBD) . . . . . 11
8.11 rs1376223093 (chr20:33756333) in locus 127 (MMP24-AS1-EDEM2) . . . . . 12
8.12 rs114102448 (chr21:47523605) in locus 128 (COL6A2) . . . . . 13
8.13 rs73174952 (chr22:43111733) in locus 130 (A4GALT) . . . . . 14

**Supplementary Figure 8.1** – rs10800427 (chr1:169272451) in locus 9 (NME7)

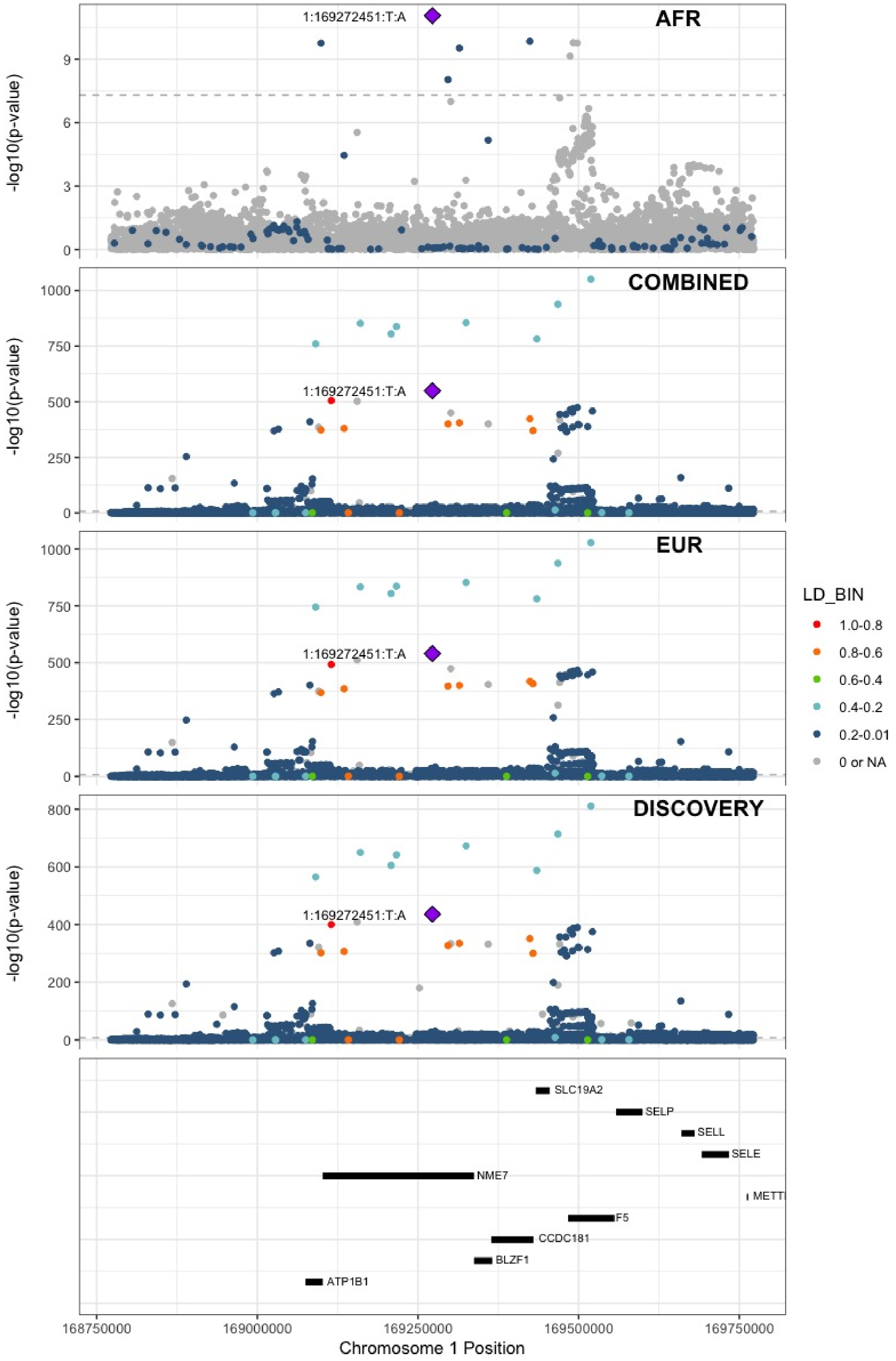

**Supplementary Figure 8.2** – rs182293241 (chr2:128029746) in locus 18 (ERCC3)

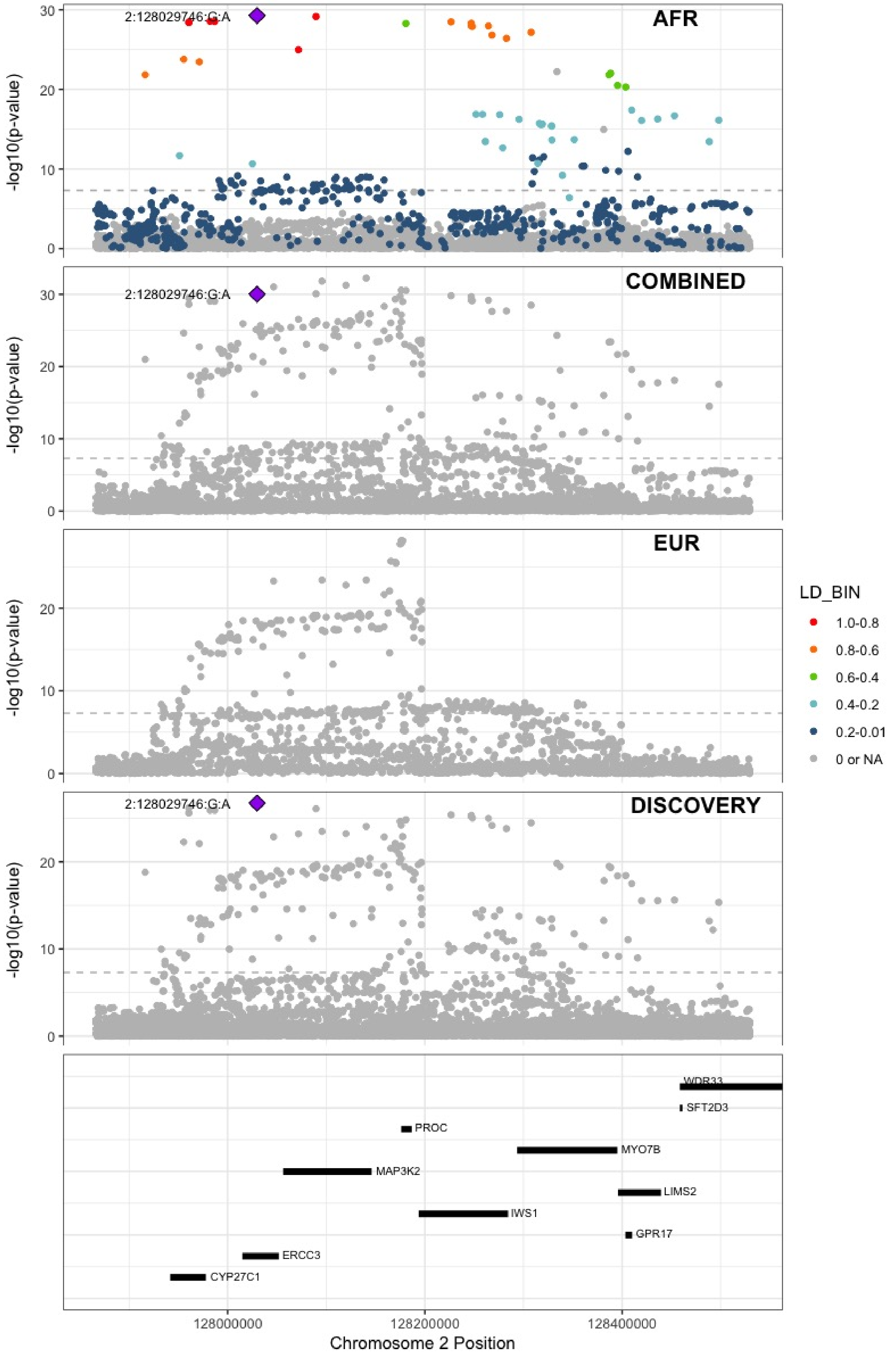

**Supplementary Figure 8.3** – rs529326506 (chr3:90503764) in locus 26 (EPHA3 ;NONE)

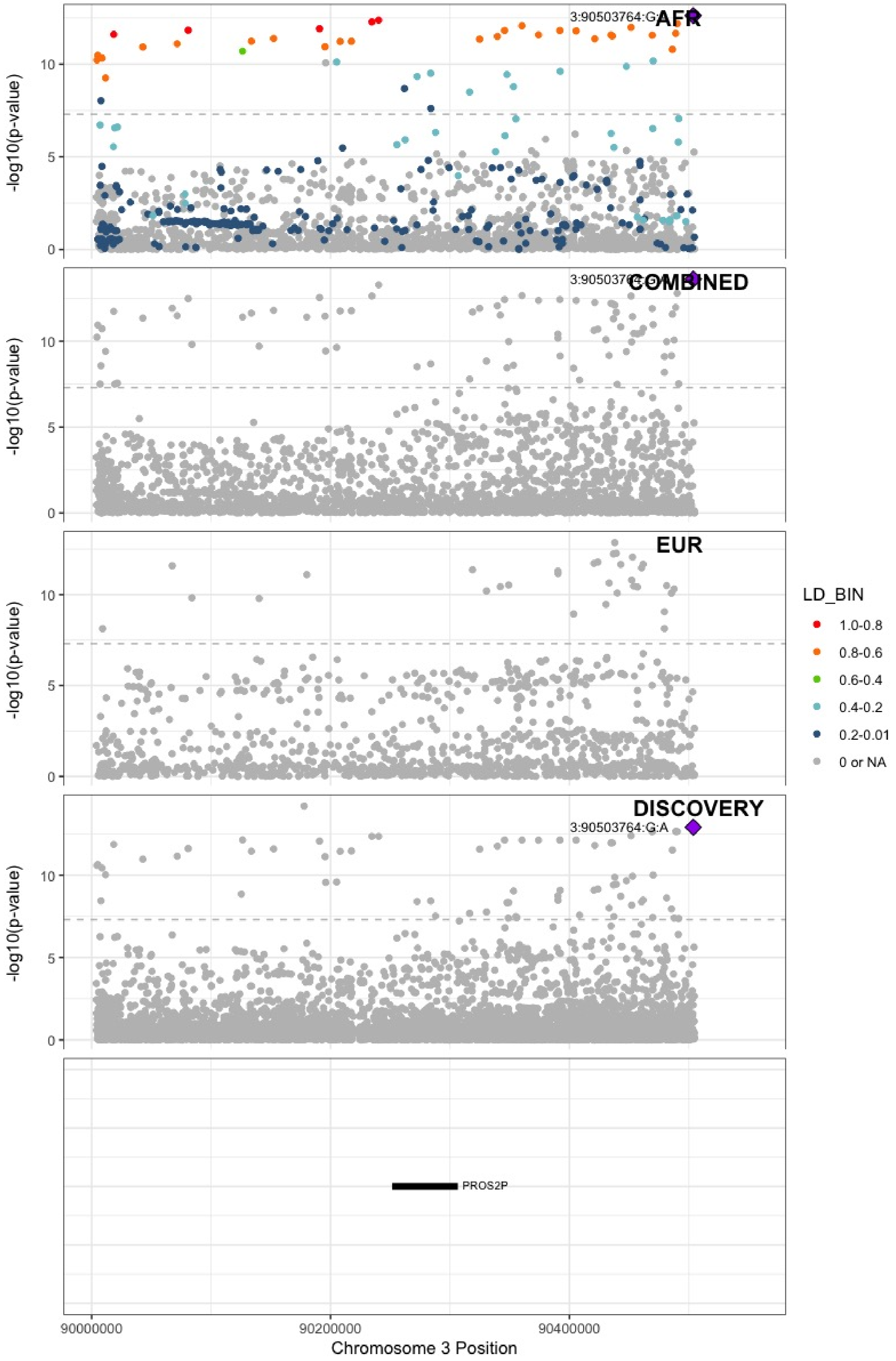

**Supplementary Figure 8.4** – rs80133433 (chr3:93791551) in locus 27 (NSUN3)

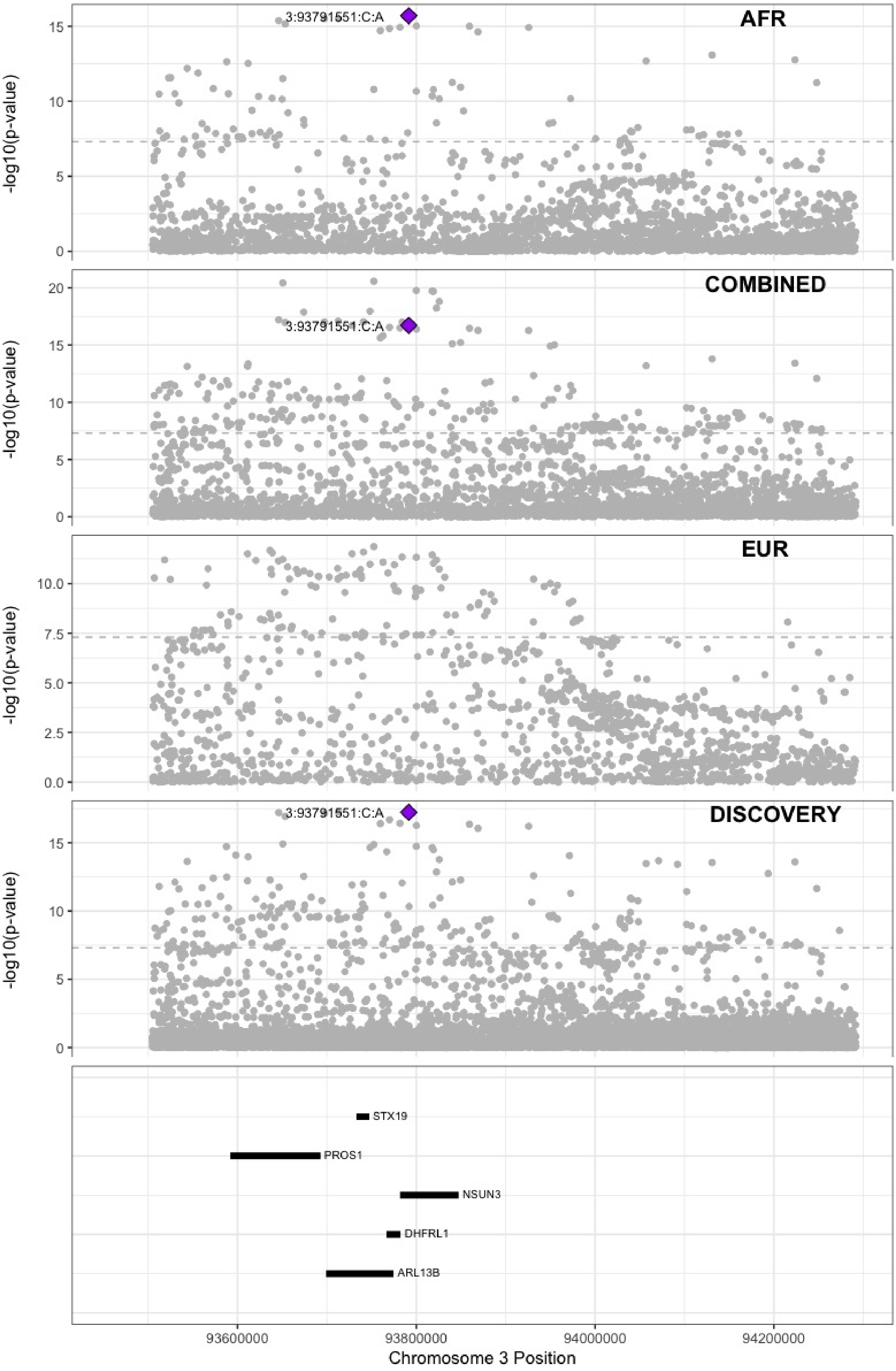

**Supplementary Figure 8.5** – rs2066864 (chr4:155525695) in locus 39 (FGG)

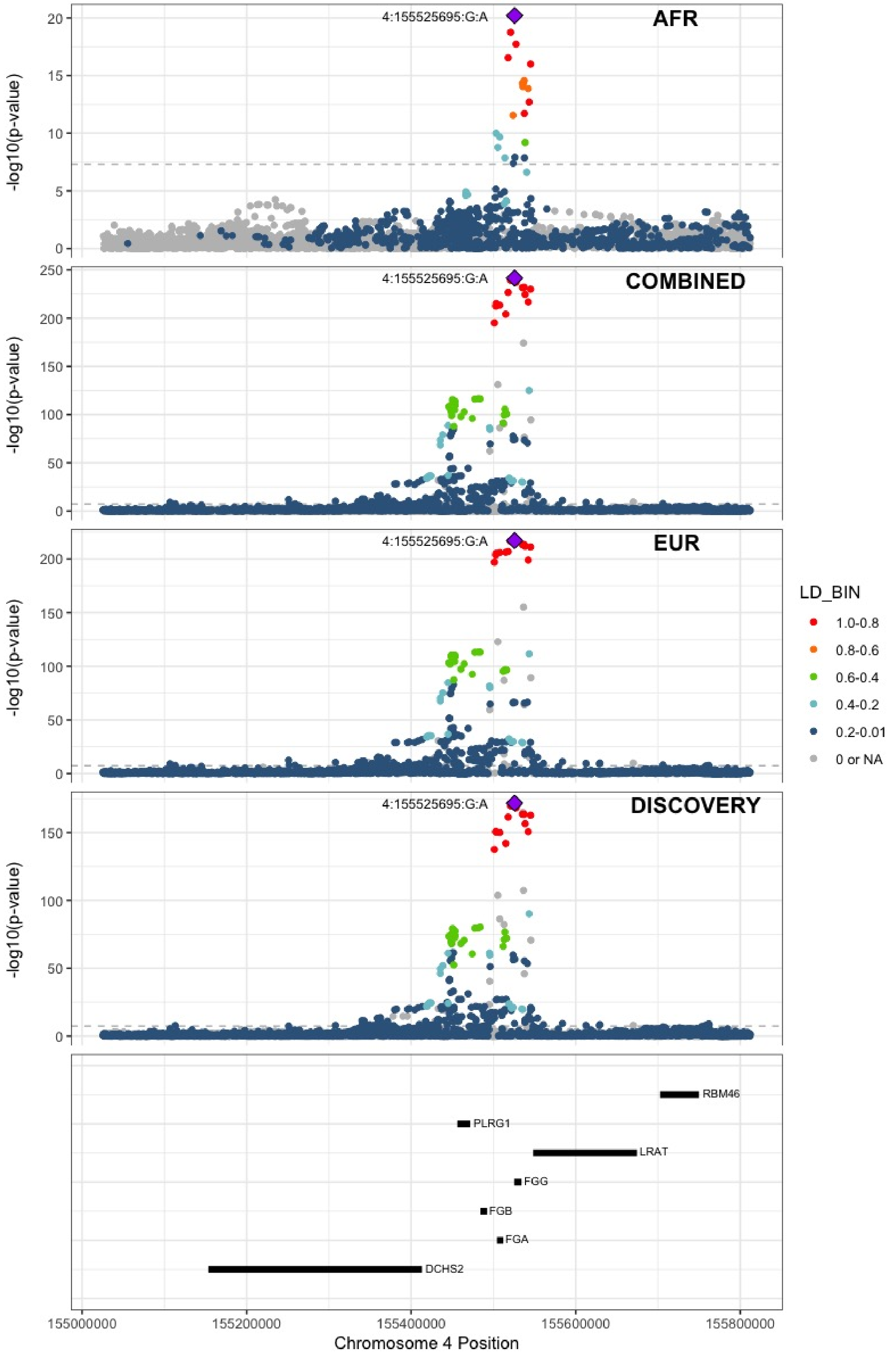

**Supplementary Figure 8.6** – rs56810541 (chr4:187200550) in locus 40 (F11)

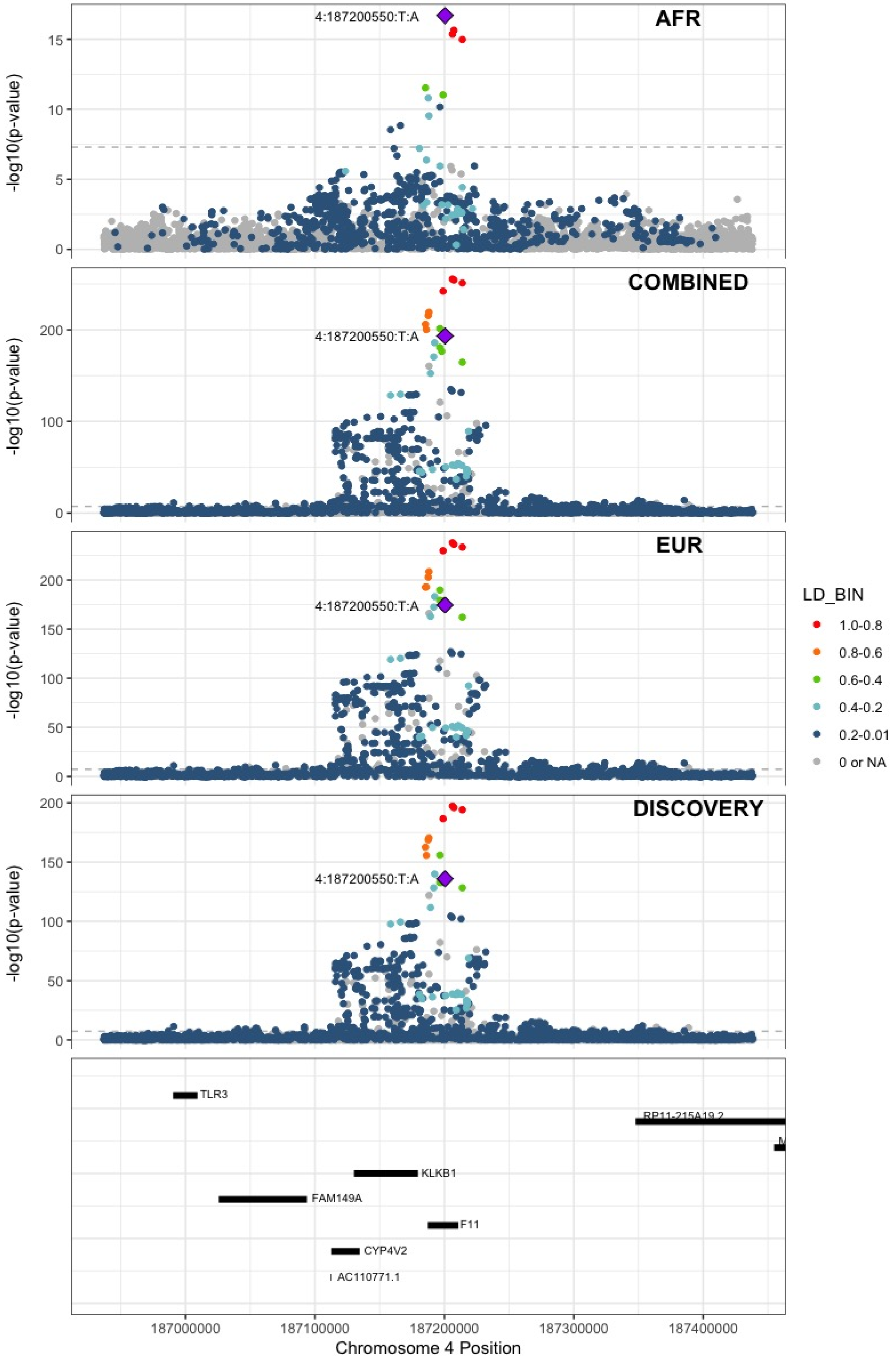

**Supplementary Figure 8.7** – rs8176719 (chr9:136132908) in locus 66 (ABO)

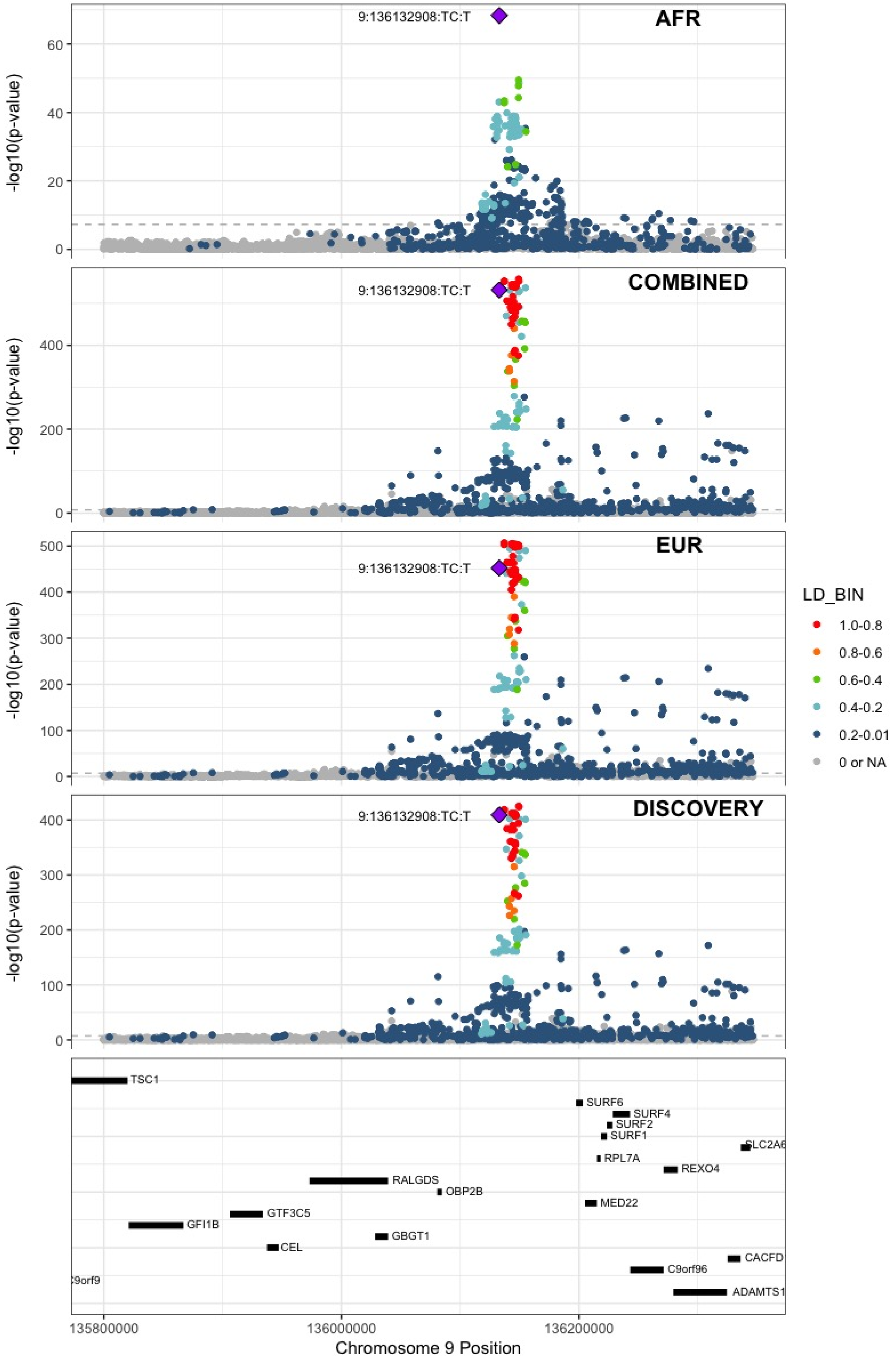

**Supplementary Figure 8.8** – rs57950734 (chr12:6145649) in locus 87 (VWF)

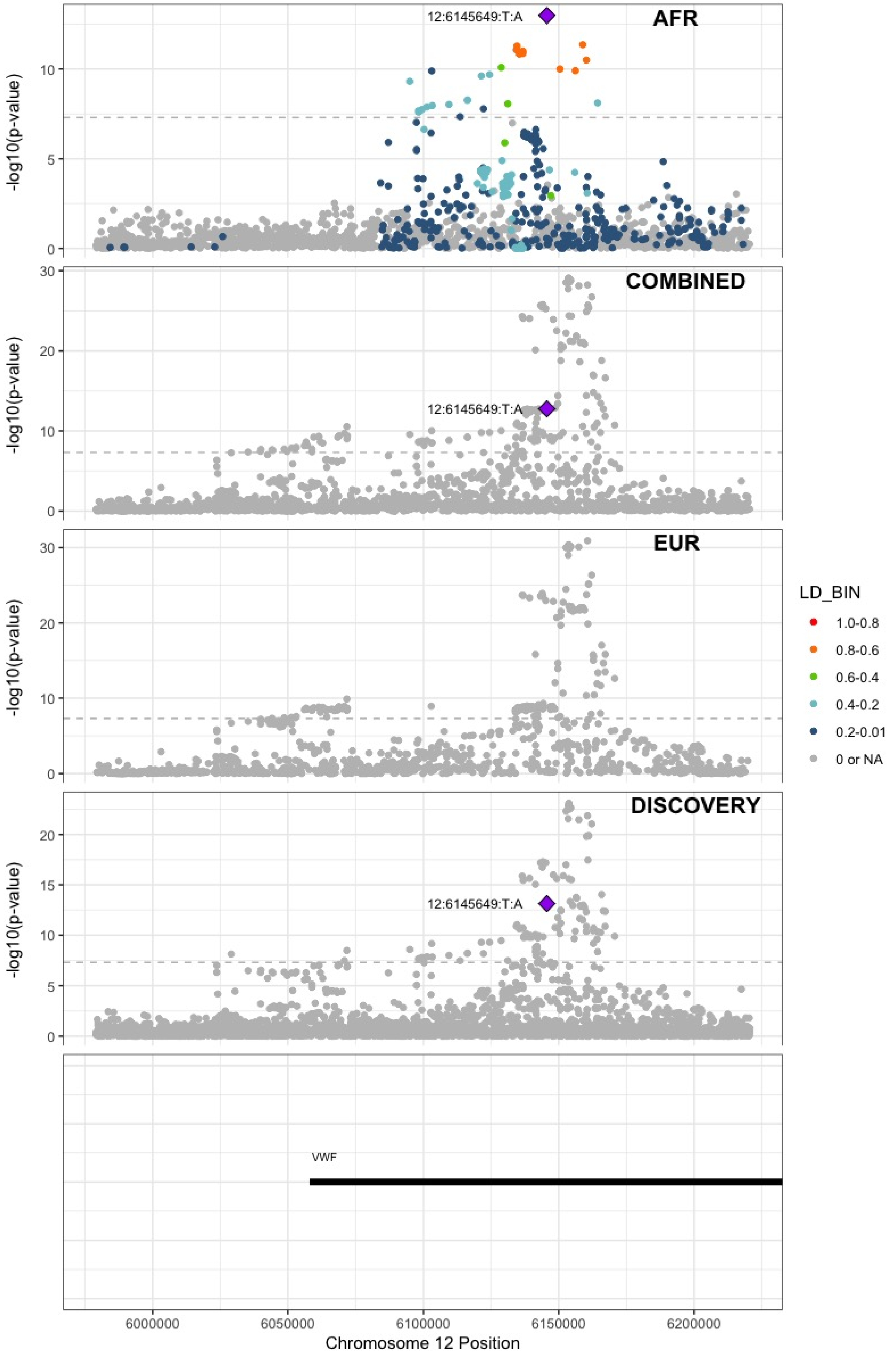

**Supplementary Figure 8.9** – rs76668186 (chr16:6686083) in locus 104 (RBFOX1)

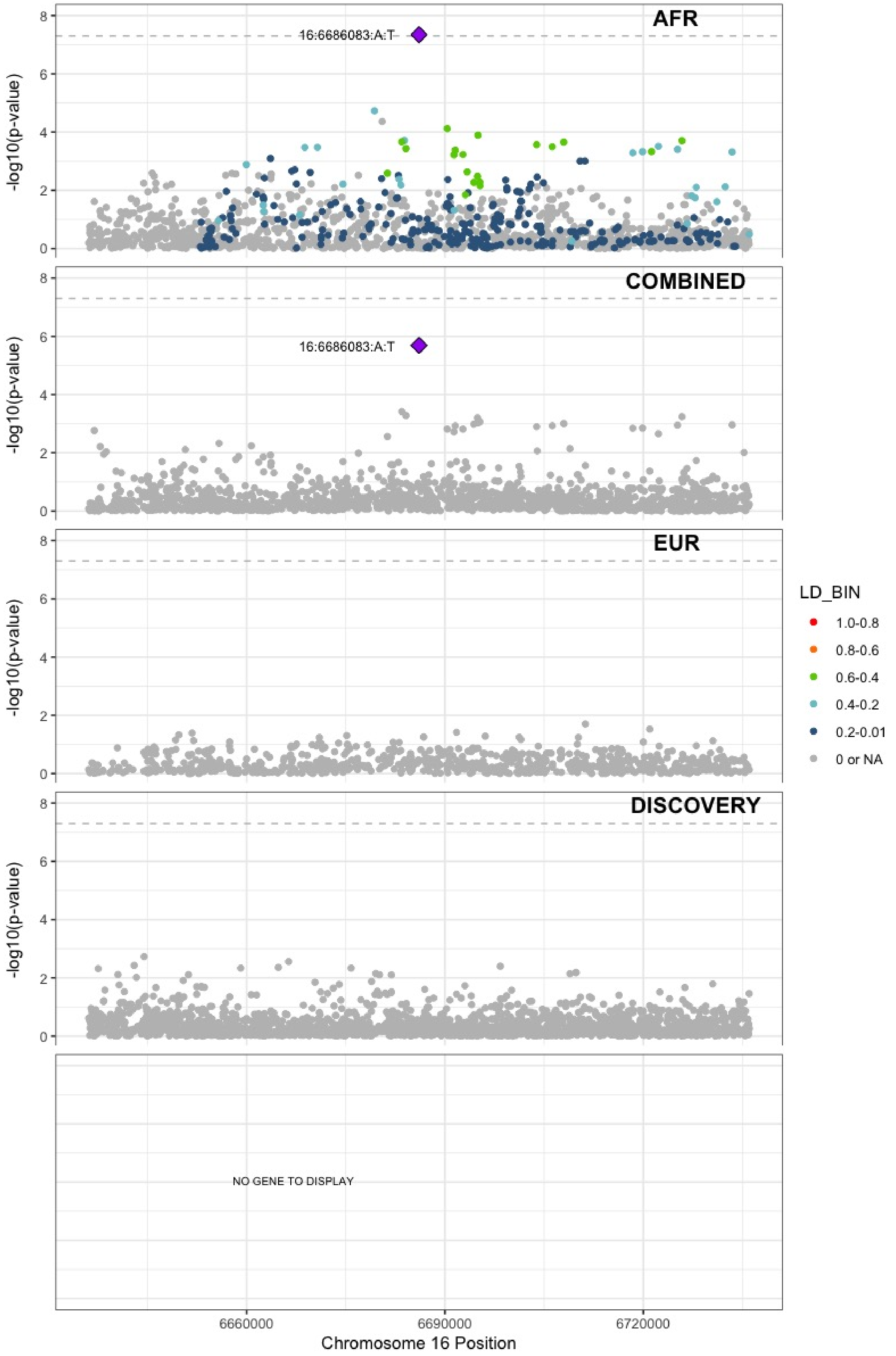

**Supplementary Figure 8.10** – rs13306849 (chr20:23030199) in locus 126 (THBD)

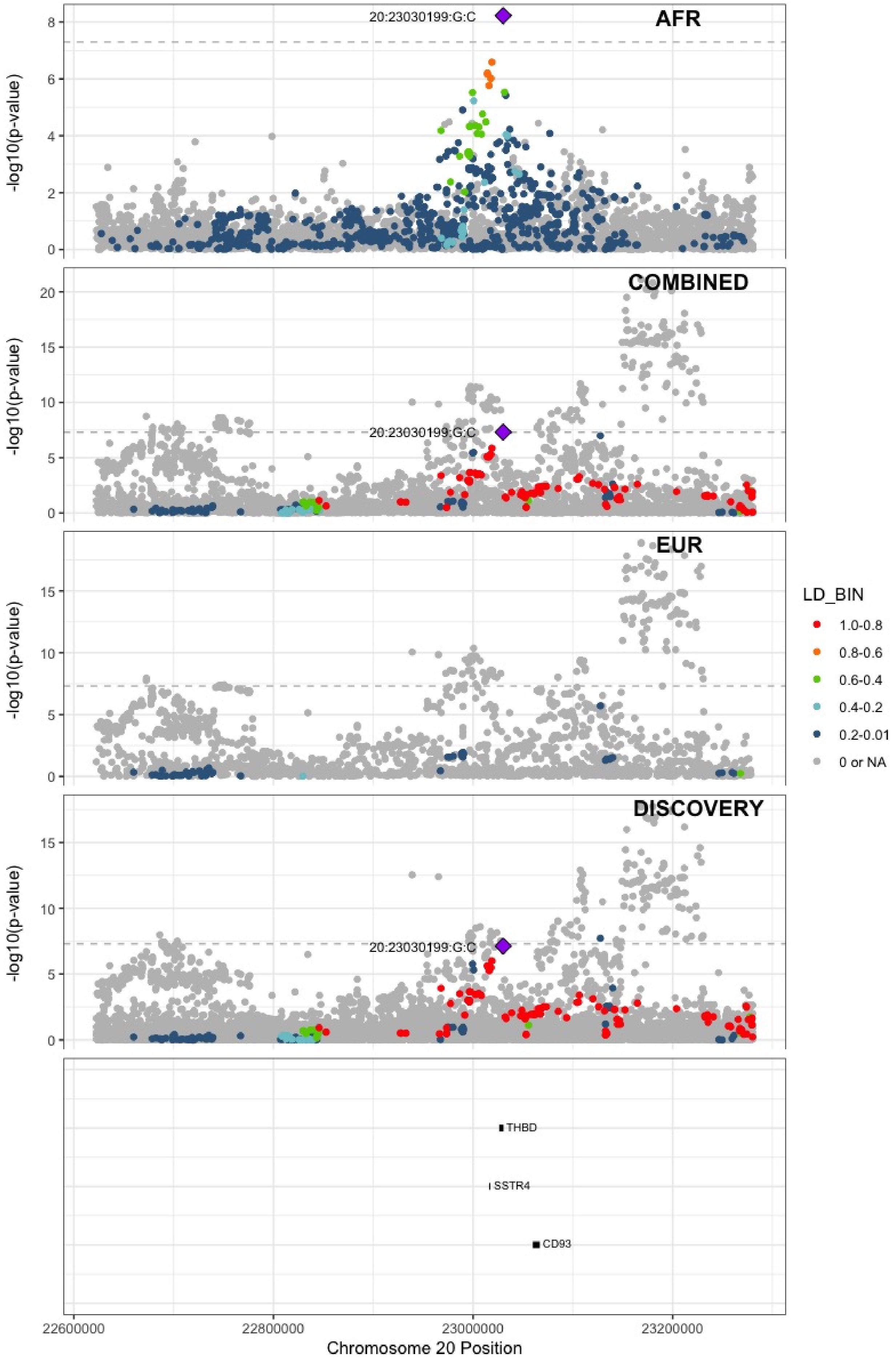

**Supplementary Figure 8.11** – rs1376223093 (chr20:33756333) in locus 127 (MMP24-AS1-EDEM2)

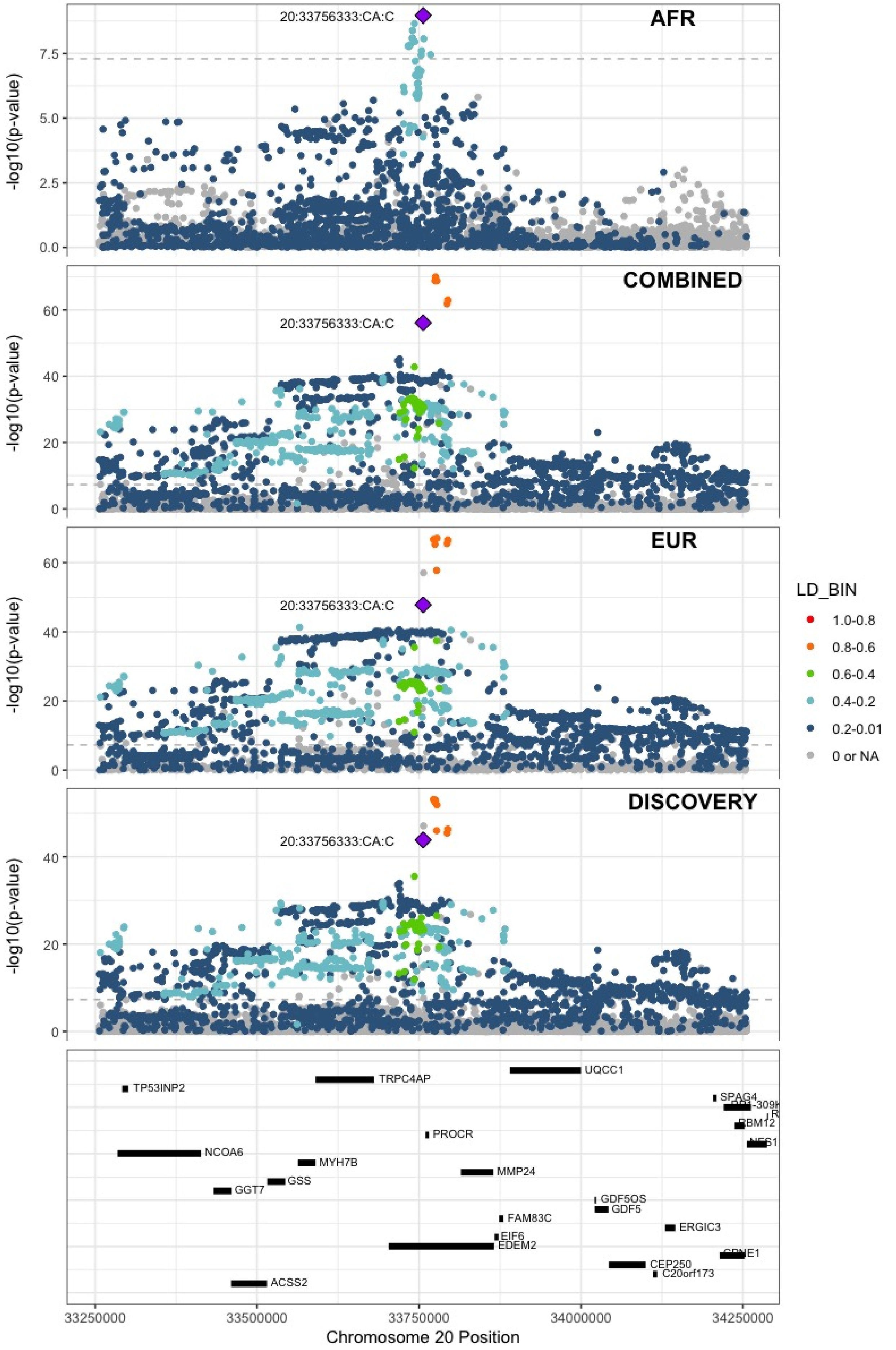

**Supplementary Figure 8.12** – rs114102448 (chr21:47523605) in locus 128 (COL6A2)

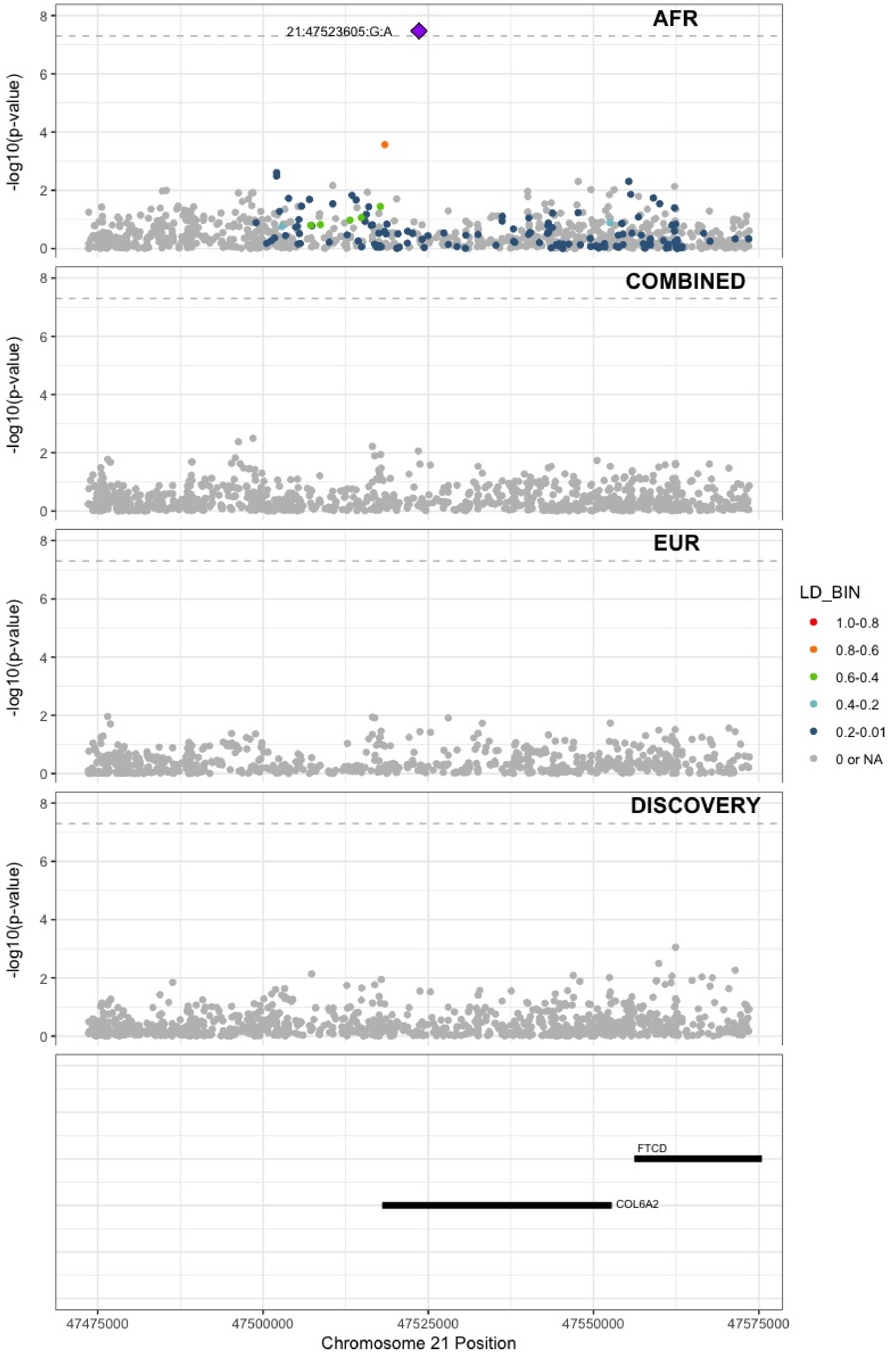

**Supplementary Figure 8.13** – rs73174952 (chr22:43111733) in locus 130 (A4GALT)

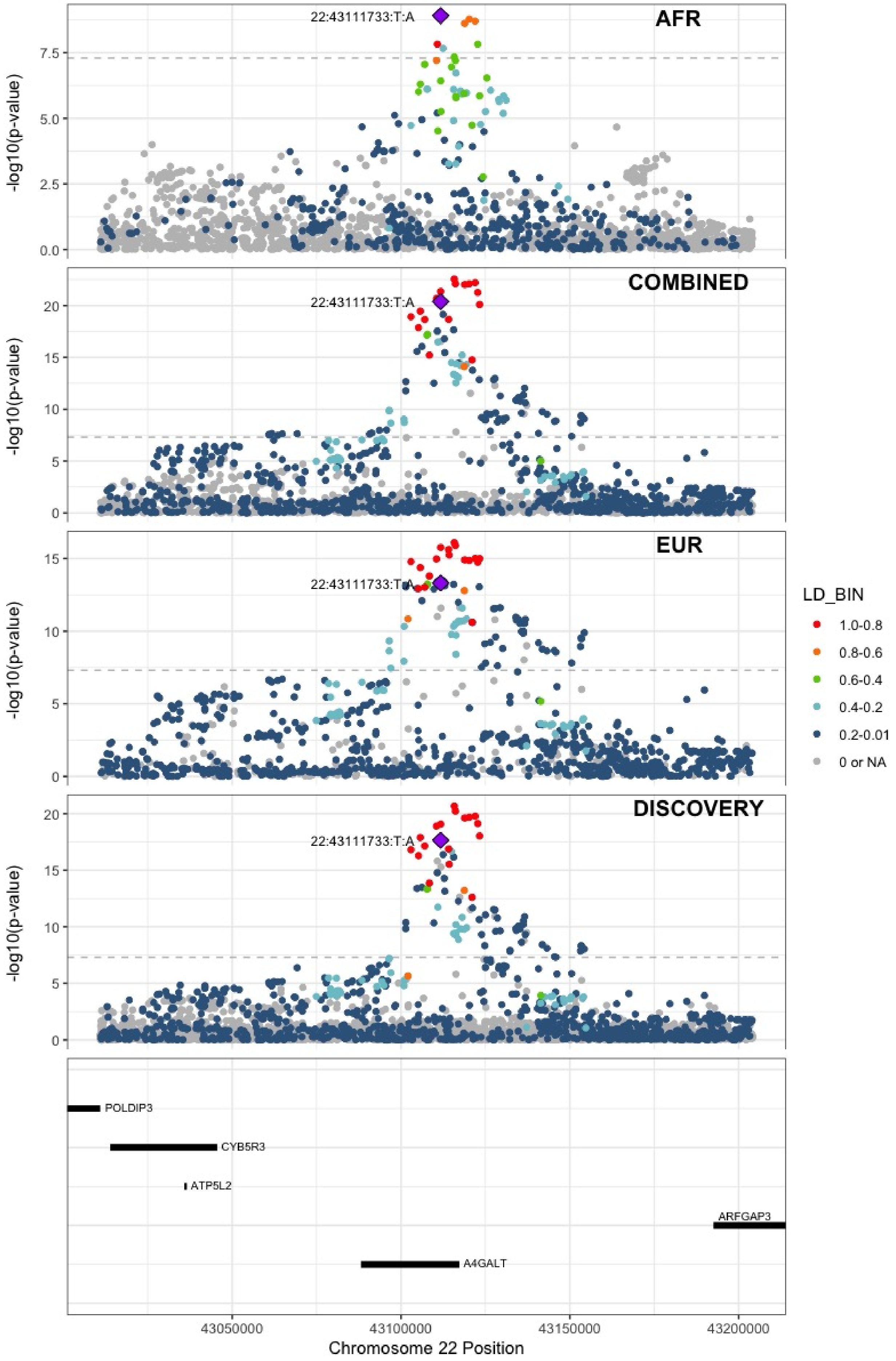
