## Supplemental Methods for "Cross-Ancestry Investigation of Venous Thromboembolism Genomic Predictors"

### Study Descriptions

The **23andMe, Inc. cohort** is a population based cohort. Participants provided informed consent to participate in research, under a protocol approved by the external AAHRPP-accredited IRB, Ethical & Independent Review Services (E&I Review). Cases were defined as those individuals that reported having had DVT through the survey questions "*What types of blood clot or stroke were you diagnosed with? Answer: A blood clot in your arms or legs (deep vein thrombosis or DVT)*" and "*Have you ever been diagnosed with deep vein thrombosis? Answer: Yes*". Controls were those individuals who reported not having had DVT. For the analysis, a set of unrelated individuals was chosen using a segmental identity-by-descent (IBD) estimation algorithm (https://www.23andme.com/ancestry-composition-guide/). Individuals were defined as related if they shared more than 700 cM IBD. The selection process was done by preferentially retaining cases over controls to maximize statistical power. A total of 59,143 DVT cases and 2,835,159 controls were included in this study.
The variant-level data for the 23andMe replication dataset are fully disclosed in the manuscript. Individual-level data are not publicly available due to participant confidentiality, and in accordance with the IRB-approved protocol under which the study was conducted. Participants provided informed consent and participated in the research online, under a protocol approved by the external AAHRPP-accredited IRB, Ethical & Independent Review Services (E&I Review). Participants were included in the analysis on the basis of consent status as checked at the time data analyses were initiated.

The **Atherosclerosis Risk in Communities (ARIC)** study has been described in detail previously.[1] Men and women aged 45-64 years at baseline were recruited from four communities: Forsyth County, North Carolina; Jackson, Mississippi; Minneapolis, Minnesota; and Washington County, Maryland. A total of 15,792 individuals, predominantly White and African American, participated in the baseline examination in 1987-1989, with 6 reexamination visits conducted from 1990-2019.

The **BioBank Japan (BBJ)** is a hospital-based Japanese national biobank project including data from approximately 200,000 patients enrolled between 2003 and 2007.[2,3] Participants were recruited at 12 medical institutes throughout Japan (Osaka Medical Center for Cancer and Cardiovascular Diseases, Cancer Institute Hospital of Japanese Foundation for Cancer Research, Juntendo University, Tokyo Metropolitan Geriatric Hospital, Nippon Medical School, Nihon University School of Medicine, Iwate Medical University, Tokushukai Hospitals, Shiga University of Medical Science, Fukujuji Hospital, National Hospital Organization Osaka National Hospital and Iizuka Hospital).

**BioME**: Mount Sinai's BioMe Biobank is an electronic health record (EHR)-linked clinical care cohort ('Biobank'), consisting of ~60,000 participants from diverse ancestries characterized by a broad spectrum of (longitudinal) biomedical traits. Enrolled participants consent to be followed throughout their clinical care (past, present, and future) at Mount Sinai in real-time, integrating their genomic information with their electronic health records for discovery research. Recontacting of participants is permitted by consent, enabling a broad range of additional studies including in-depth clinical and OMICS phenotyping, mobile Health applications, recruitment for prospective studies/trials, and return-of-results clinical care implementation projects. The BioMe Cohort, launched in September 2007, is an ongoing, consented EHR-linked bio- and data repository that enrolls participants non-selectively from the Mount Sinai Health System patient population. Starting December of 2014, we began in addition enrollment of participants at the Institute of Family Health, a network of 19 full-time Federally-Qualified Community Health Centers (FQHC) throughout New York City with an unified implementation of electronic health records (Epic systems) under a single administration. The Institute of Family Health facilities has over 20 years of successful involvement in community-based research to address health disparities. With IFH participation, BioMe further affords unique opportunities to extend comprehensive studies of genome sequence variation underlying rare and common diseases in underserved, urban communities. As of Oct 2018, 45,479 adult participants were enrolled. On average 1000 new participants are consented each month. BioMe participants represent a broad ancestral, ethnic and socioeconomic diversity with a distinct and population-specific disease burden, characteristic of Northern Manhattan communities served by Mount Sinai. BioMe participants are predominantly of African (35%) or Hispanic/Latino (36%) ancestry. Participants who self-identify as Hispanic/Latino further report to be of Puerto Rican (39%), Dominican (23%), Central/South American (17%), Mexican (5%) or other Hispanic (16%) ancestry.

The **Cardiovascular Health Study (CHS)** is a population-based cohort study of risk factors for coronary heart disease and stroke in adults ≥65 years conducted across four field centers.[4,5] The original predominantly European -ancestry cohort of 5,201 persons was recruited in 1989-1990 from random samples of the Medicare eligibility lists; subsequently, an additional predominantly African-American cohort of 687 persons were enrolled for a total sample of 5,888. Blood samples were drawn from all participants at their baseline examination and DNA was subsequently extracted from available samples. CHS was approved by institutional review committees at each field center. Participants included in the present analyses had available DNA and gave informed consent including consent to use of genetic information for the study of cardiovascular disease.

The **Early Onset Venous Thrombosis Study (EOVT):** In the EOVT study, 453 patients of European origin with early age of onset of VT (<50 years) recruited in 4 different French centers (Grenoble, Marseille, Montpellier and Paris) were compared to 1,327 healthy French subjects from the Suvimax study.[6] VT patients had a first DVT and/or PE event documented by venography, Doppler ultrasound, angiography, and/or ventilation/perfusion lung scan. They were free of any acquired risk factors at the time of VTE (including surgery, hospitalization, pregnancy, puerperium, oral contraception, cancer, and autoimmune disease); and strong known genetic risk factors, including anti-thrombin, protein C or protein S deficiency, homozygosity for FV Leiden or FII 20210A, and lupus anticoagulant. Criteria for inclusion of the healthy controls were European origin, no chronic conditions, and no regular medicines.

The **eMERGE (electronic MEdical Records GEnomics) Network** is a national initiative to combine biobanks with electronic medical records.[7] As part of eMERGE, more than 83,000 individuals across 13 sites have been genotyped. Recruitment criteria varied between sites.[7] For this study, four sites contributed data to the European ancestry analysis (Geisinger Health System, Group Health, Marshfield Clinic and Vanderbilt University). In total, 1,558 cases and 10,027 controls of European ancestry were included. For the African-American studies, two eMERGE sites contributed to the analyses (Vanderbilt and Mount Sinai). In total, 436 cases and 14,353 controls of African-American ancestry were included. VTE was ascertained through a NPL-based Electronic Health Records-driven algorithm that leveraged structured data (ICD codes) and unstructured data (clinical notes).[8]

The **Estonian Biobank** is a population-based cohort of the Estonian Genome Center at the University of Tartu (EGCUT), Estonia.[9] The current cohort size is ca 200000, from 18 years of age and up and reflects closely the age distribution in the adult Estonian population. This project is conducted according to the Estonian Gene Research Act and all participants have signed the broad informed consent. Upon recruitment, the biobank participants filled out a thorough questionnaire, covering lifestyle, diet and clinical diagnoses defined according to the ICD10 coding. Illumina GSA arrays were used for genotyping and imputation was performed by using the Estonian-specific reference panel.[10]

The **FARIVE Study** is a multicenter case-control study of 607 patients with a first episode of proximal deep VT and/or pulmonary embolism. Patients younger than 18 years, with previous VT event, that had a diagnosis of active cancer or a history of malignancy less than 5 years previously, or have a short life expectancy because of other causes, were excluded. The control group consists of age- and sex-matched individuals free of venous and arterial thrombotic disease. Potential control subjects with cancer, liver or kidney failure, or a history of venous and/or arterial thrombotic disease are ineligible.[11]

The **FinnGen** study is a public–private partnership project. Six regional and three country-wide Finnish biobanks participate in FinnGen. Additionally, data from previously established population and disease-based cohorts are utilized. Participants' health outcomes are followed up by linking to the national health registries (since 1969), which collect information from birth to death. Summary statistics for VTE (from release 5, code I9_VTE) were publicly available and retrieved from https://www.finngen.fi/en/access_results.

The **Framingham Heart Study (FHS)** was started in 1948 with 5,209 randomly ascertained participants from Framingham, Massachusetts, US, who had undergone biannual examinations to investigate cardiovascular disease and its risk factors.[12] In 1971, the Offspring cohort (comprising 5,124 children of the original cohort and the children's spouses) and in 2002, the Third Generation (consisting of 4,095 children of the Offspring cohort) were recruited. FHS participants in this study are of European ancestry. The methods of recruitment and data collection for the Offspring and Third Generation cohorts have been described elsewhere.[13]

The **GAIT (Genetic Analysis of Idiopathic Thrombophilia**) project is a family based study where 935 subjects in 35 extended pedigrees were collected.[14,15] To be included in the study, a family was required to have at least 10 living individuals in 3 or more generations. Families were selected through a proband with idiopathic thrombophilia, which was defined as recurrent thrombotic events (at least one of which was spontaneous), a single spontaneous thrombotic episode plus a first-degree relative also affected, or onset of thrombosis before age 45. Thrombosis in these probands was considered idiopathic when biological causes as antithrombin deficiency, protein S and C deficiencies, activated protein C resistance, plasminogen deficiency, heparin cofactor II deficiency, Factor V Leiden, dysfibrogenemia, lupus anticoagulant and antiphospholipid antibodies, were excluded. Subjects were interviewed by a physician to determine their health and reproductive history, current medications, alcohol consumption, use of sex hormones (oral contraceptives or hormonal replacement therapy) and their smoking history. The study was performed according to the Declaration of Helsinki. All procedures of the study were reviewed by the Institutional Review Board of the Hospital de la Santa Creu i Sant Pau, Barcelona, Spain. Adult subjects gave informed consent for themselves and for their minor children.

The **Global Biobank Meta-analysis Initiative** (GBMI) is a collaborative network of 19 biobanks from 4 continents representing more than 2.1 million consented individuals with genetic data linked to electronic health records. Standard ICD based definitions of VTE were used for identifying VTE cases and excluding related diseases from population based controls. Contributing cohorts for a meta-analysis of VTE for a non-overlapping cohort replication of genome-wide significant findings included the China Kadoorie Biobank, East London Genes and Health, UCLA ATLAS Community Health Initiative, and the Michigan Genomics Initiative.

The **Heart and Vascular Health (HVH) VTE Study** is a case-control study of risk factors for cardiovascular outcomes set at Group Health (GH), an integrated health care delivery system in western Washington State. Cases include venous thromboembolism (VTE), myocardial infarction (MI), stroke, and atrial fibrillation, with a shared common control group frequency matched to MI cases on age (within decade) sex, treated hypertension, and calendar year of identification. Study approval was granted by the human subjects committee at GH, and written informed consent was provided by all study participants. Methods for the study have been described previously.[16,17] VTE cases and controls with no prior VTE were utilized for these analyses. All study participants were GH members, with women aged 18-89 years old and men aged 30-89 years old. Deep venous thrombosis (DVT) and pulmonary embolism (PE) events were identified using hospital discharge diagnosis codes and urgent care diagnosis codes. Additionally, subjects who received a prescription for low molecular weight heparin were screened as potential cases of DVT. Eligibility and risk factor information were collected by trained medical record abstractors from a review of the GH medical record using only data available prior to the index date. A venous blood sample was collected from all consenting subjects, and DNA was extracted from white blood cells using standard procedures.

The **HUNT (Nord-Trøndelag Health) Study** is a large population-based health study started in 1984 for the inhabitants of Nord-Trøndelag county in central Norway. A comprehensive description of the study population has been previously reported.[18] In brief, approximately every 10 years the entire adult population of Nord-Trøndelag (~90,000 adults in 1995) is invited to attend a health survey which includes comprehensive questionnaires, an interview, clinical examination, and detailed phenotypic measurements (HUNT1 [1984 to 1986]; HUNT2 [1995 to 1997] and HUNT3 [2006 to 2008]). Approximately 90% of participants from HUNT2 and HUNT3 were genotyped in 2015 (n = 69,422). All VTE cases are validated based on thorough review of the electronic health record.

The **JUPITER study** (Justification for the Use of statins in Prevention: an Intervention Trial Evaluating Rosuvastatin) was an international, randomized, placebo-controlled trial of rosuvastatin (20mg/day) in the primary prevention of cardiovascular disease conducted among 17,802 apparently healthy men and women with LDL-C < 130 mg/dL and hsCRP > 2 mg/L conducted between 2003 and 2008, with median and maximum follow-up times of 1.9 and 5.0 years, respectively.[19–21] Approximately 71.2% of JUPITER participants had European ancestry among whom 71.4% provided DNA and consent for genetic analysis. Ascertainment of incident venous thromboembolism (VTE) was pre-specified in the trial protocol and included all cases of diagnosed pulmonary embolism or deep-vein thrombosis as well as corroborating evidence from confirmatory diagnostic tests, the initiation of anticoagulation therapy, or death that was considered likely to have been due to a pulmonary embolism. The genetic sub-sample for the current analysis included 8,749 unrelated individuals with self-reported European ancestry confirmed by genetic analysis.

The **MARseille THrombosis Association (MARTHA)** project has already been described extensively.[22–24] It is composed of unrelated subjects of European origin, with the majority being of French ancestry, consecutively recruited at the Thrombophilia center of La Timone hospital (Marseille, France) between January 1994 and October 2012. All patients had a documented history of VT and were free of well characterized genetic risk factors including AT, PC, or PS deficiency, homozygosity for FV Leiden or FII 20210A, and lupus anticoagulant. They were interviewed by a physician on their medical history, which emphasized manifestations of deep vein thrombosis and pulmonary embolism using a standardized questionnaire. The thrombotic events were confirmed by venography, Doppler ultrasound, spiral computed tomographic scanning angiography, and/or ventilation/perfusion lung scan. Controls were healthy individuals randomly selected from the 3C study, a population-based study carried out in 3 French cities composed of 8707 non institutionalized individuals aged over 65 randomly selected from the electoral rolls and free of any chronic diseases and for which biological (DNA, plasma) samples could have been obtained.[25]

The **MARseille THrombosis Association study of 2010-2012 (MARTHA12)** is composed of an independent sample of 1,245 VT patients. Patients have been recruited between 2010 and 2012 according to the same criteria as the MARTHA patients.[24]

The **Mass General Brigham Biobank (MGBB)** is a hospital-based research cohort containing genotypic and clinical data from >105,000 individuals enrolled across 7 regional hospitals with median 3 years of follow-up.[8] Genotyping was performed for ~36,000 MGBB participants. Venous thromboembolism case status was ascertained from EHR query of relevant ICD-9 codes (451.11, 451.19, 453.2, 453.4, 415.1) and ICD-10 codes (I80.1, I80.2, I82.22, I82.4, I82.5, I26.0, I26.9) when a minimum of two hospital (inpatient or outpatient) encounters had occurred.[26] Control status was defined among the genotyped population as lacking the above ICD-9/10 codes.

The **Mayo VTE Study** recruited consecutive Mayo Clinic outpatients who resided in the upper midwest United States and who were referred to the Mayo Clinic Special Coagulation Laboratory or Thrombophilia Center.[27] We prospectively selected clinic-based controls from persons undergoing outpatient general medical examinations in 2004 - 2009 within the Mayo Clinic Divisions of General Internal Medicine and Primary Care Internal Medicine, Department of Internal Medicine, and general internal medicine practices that care for patients (> 10 000 per year) from the upper Midwest United States. Additional controls were recruited from the Department of Family Medicine and the Mayo Clinic Sports Medicine Center. Controls were frequency matched on the age group, sex, state of residence and myocardial infarction(MI)/stroke status distribution of the cases, and had no previous diagnosis of VT or superficial vein thrombosis.[27]

The **Multiple Environmental and Genetic Assessment of risk factors for venous thrombosis (MEGA)** study is a large population-based case-control study.[28] Data collection and ascertainment of venous thrombotic events have been previously described in detail. In short, patients with a first deep vein thrombosis or pulmonary embolism were recruited at six anticoagulation clinics in the Netherlands between 1999 and 2004. The diagnosis of a deep vein thrombosis was based on compression ultrasonography, whereas a pulmonary embolism was confirmed by perfusion and ventilation scintigraphy, helical computed tomography or pulmonary angiography. Blood samples were taken at least 3 months after discontinuation of vitamin K antagonist treatment, unless patients were still receiving anticoagulant therapy one year after their VT event.

The **Multi-Ethnic Study of Atherosclerosis (MESA)** is a study of the characteristics of subclinical cardiovascular disease (disease detected non-invasively before it has produced clinical signs and symptoms) and the risk factors that predict progression to clinically overt cardiovascular disease or progression of the subclinical disease. MESA researchers study a diverse, population-based sample of 6,814 asymptomatic men and women aged 45-84 from six field centers across the United States. Approximately 38 percent of the recruited participants are white, 28 percent African-American, 22 percent Hispanic, and 12 percent Asian, predominantly of Chinese descent. Five follow-up exams have been completed since 2000.[29] Blood was collected by venipuncture at baseline and at each follow-up visit. Participants provided informed consent for the use of DNA from the blood sample. Participants in the MESA cohort who consented to genetic analyses and data sharing (dbGaP) were genotyped using the Affymetrix Human SNP Array 6.0 (GWAS array) as part of the NHLBI SHARe (SNP Health Association Resource) project.

The **Million Veteran Program (MVP)** is a Department of Veterans Affair cohort study. We conducted a discovery genetic association analysis using DNA samples and phenotypic data from the Million Veteran Program (MVP). In MVP, individuals aged 18 to over 100 years have been recruited from 63 Veterans Affairs (VA) Medical Centers across the United States.[26,30]

The **Nurse's Health Studies (NHS and NHS-II)** These studies have been described previously and additional information is available at http://www.nurseshealthstudy.org.[31,32] NHS and NHS-II are longitudinal cohort studies of female nurses. In 1976, baseline questionnaires were sent to registered nurses from 11 populous US states, establishing a cohort of 121,700 women aged 30-55. There were no exclusions by race, but the majority (96%) were of European ancestry; corresponding to the demographics of nurses in 1976. NHS participants are mailed a questionnaire every two years that assesses risk factor status and interval disease events. Physician-diagnosed PE has been asked on every biennial NHS questionnaire since 1982. In NHS, the question reads: "Since [year], have you had any of these physician-diagnosed illnesses? ... Pulmonary Embolus." NHS questionnaires also ask whether the nurse had "Any other major diagnosis: ____." In NHS, DVT is captured when a nurse answers that she has had phlebitis or thrombophlebitis (ICD-9=453.x). Questionnaire-reported VTE diagnoses have proven to be highly accurate, with >95% validation of VTE events. A physician reviews medical records for all reported PEs, validating diagnoses when medical records include: a positive pulmonary angiogram, a high-probability ventilation/perfusion scan, or a positive CT pulmonary angiogram.

The **Penn Medicine BioBank (PMBB)** recruits patients from throughout the University of Pennsylvania Health System for genomic and precision medicine research. Participants actively consent to allow the linkage of biospecimens to their longitudinal EHR. Currently, >60 000 participants are enrolled in the PMBB. A subset of ~45000 individuals who have undergone whole exome sequencing and genotyping, performed through a collaboration with the Regeneron Genetics Center. A further subset of ~12000 subjects with imputed genotype data was used in this analysis.

The **Riesgo de Enfermedad TROmboembólica VEnosa study (RETROVE)** is a prospective case–control study that includes 400 consecutive patients with VTE (cancer associated thrombosis was excluded) and 400 healthy control volunteers. All individuals were ≥ 18 years. The diagnosis was confirmed with Doppler ultrasonography, tomography, magnetic resonance, arteriography, phlebography or pulmonary gammagraphy. Blood samples from the patients were taken at least 6 months after thrombosis to minimize the influence of the acute phase. None of the participants was using oral anticoagulants, heparin, or antiplatelet therapy at the time of blood collection. Controls were selected according to the age and sex distribution of the Spanish population (2001 census). A total of 5 ml of blood was obtained in a Vacutainer tube (BD Vacutainer Becton Dickinson and Company, New Jersey, USA) containing EDTA as anticoagulant. All individuals were genotyped using Infinium Global Screening Array-24 v3.0 kit from Illumina and imputed using the Haplotype Reference Consortium panel. Written informed consent was obtained for all participants and all procedures were approved by the Institutional Review Board of the Hospital de la Santa Creu i Sant Pau (Barcelona).

The **Tromsø Study** is a single-center, population-based cohort study of the inhabitants of Tromsø, Norway.[33] 27,158 individuals participated in the fourth survey of the Tromsø Study between 1994-1995; baseline characteristics were collected using self-reported questionnaires, physical examinations, and blood samples. Non-fasting blood was drawn from an antecubital vein to gather plasma and whole blood. Whole blood was used to prepare archive quality DNA, and was stored at the HUNT Biobank in Levanger, Norway. All 27,158 participants were followed from the date of enrollment through December 31, 2011. All cohort members that experienced an incident venous thromboembolism (VTE) during the study period were identified by searching the hospital discharge diagnosis registry, the autopsy registry, and the radiology procedure registry at the University Hospital of North Norway, the sole hospital in the Tromsø municipality.[34] The VTE events were thoroughly validated by review of medical records as previously described in detail.[34] For each case, a paired control, matched on age and sex, was randomly sampled from the cohort.

The **UK Biobank** (https://www.ukbiobank.ac.uk/) is a large population-based prospective cohort study of ~500,00 participants residing in the United Kingdom (UK).[35] All participants, aged 40-71 at recruitment in 2006-2010, attended a baseline exam at a local study assessment center and gave informed consent. DNA was extracted from the blood samples drawn at the baseline exam and genotyped at the Affymetrix Research Services Laboratory in Santa Clara, California, USA.

The **Women's Genome Health Study (WGHS):** WGHS is a prospective cohort of female North American health care professionals representing participants in the Women's Health Study (WHS) trial who provided a blood sample at baseline and consent for blood-based analyses.[10] Participants in the WHS were 45 years or older at enrollment and free of cardiovascular disease, cancer or other major chronic illness. The current data are derived from 23,294 WGHS participants for whom whole genome genotype information was available at the time of analysis and for whom self-reported European ancestry could be confirmed by multidimensional scaling analysis of 1,443 ancestry informative markers in PLINK v. 1.06. At baseline, BP and lifestyle habits related to smoking, consumption of alcohol, and physical activity as well as other general clinical information were ascertained by a self-reported questionnaire, an approach which has been validated in the WGHS demographic, namely female health care professionals.

In the WHS (the WGHS parent cohort), venous thromboembolism events are collected by self-report questionnaire and then confirmed by review of medical records. A diagnosis of deep-vein thrombosis is confirmed by a positive venous ultrasonography or venography report, whereas the diagnosis of pulmonary embolism is confirmed by a positive angiogram or computed tomography scan of the chest, or a ventilation-perfusion scan with 2 or more mismatched defects. Deaths due to pulmonary embolism are confirmed when autopsy reports, symptoms, circumstances, and medical history are consistent with this diagnosis.

The Women's Genome Health Study is supported by the National Heart, Lung, and Blood Institute (HL043851 and HL080467) and the National Cancer Institute (CA047988 and UM1CA182913), with funding for genotyping provided by Amgen.

The **Women's Health Initiative (WHI)** is a long-term national health study that focuses on strategies for preventing common diseases such as heart disease, cancer and fracture in postmenopausal women. A total of 161,838 women aged 50–79 years old were recruited from 40 clinical centers in the US between 1993 and 1998. WHI consists of an observational study, two clinical trials of postmenopausal hormone therapy (HT, estrogen alone or estrogen plus progestin), a calcium and vitamin D supplement trial, and a dietary modification trial.[36,37]

### Ethical Oversight

| Study | IRB / Ethics Committee | Status |
|---|---|---|
| FARIVE | Paris Broussais–Hopital Europeen Georges Pompidou ethics committee in Paris (ethical permit: 2002-034) | Approved |
| GAIT2 | Institutional Review Board of the Hospital de la Santa Creu i Sant Pau, Barcelona, Spain. | Approved |
| MARTHA12 | Health Department of the General Directorate for the French Ministry of Research and Innovation (Projects DC: 2008-880 and 09.576) | Approved |
| Mass General Brigham Biobank | Mass General Brigham Ethics Committee | Approved |
| MESA | The University of Washington IRB Committee D | Approved |
| Million Veteran Program | VA Central Institutional Review Board | Approved |
| Penn Medicine | IRB protocol# 813913 | Approved |
| RETROVE | Institutional Review Board of the Hospital de la Santa Creu i Sant Pau, Barcelona, Spain. | Approved |
| 23andMe | external AAHRPP accredited IRB, Ethical & Independent Review Services | Approved |
| INVENT-2019 (includes ARIC, CHS, EOVT, eMERGE, FHS, HVH, HUNT, JUPITER, MARTHA, MAYO, MEGA, NHS/NHSII/HPFS, Tromso, WGHS, WHI) | *N/A (Meta-analysis published [PMID:31420334] and available through dbGaP)* | |
| Biobank Japan | *N/A (biobank)* | |
| BioME Biobank | *N/A (biobank)* | |
| Estonian Biobank | *N/A (biobank)* | |
| FinnGen | *N/A (biobank)* | |
| GBMI | *N/A (biobank meta-analysis currently detailed in the following medrxiv preprint: doi.org/10.1101/2021.11.19.21266436)* | |
| UK Biobank | *N/A (biobank)* | |

**Funding Acknowledgement:**

**23andMe:** The variant-level data for the replication dataset are fully disclosed in the manuscript. Individual-level data are not publicly available due participant confidentiality, and in accordance with the IRB-approved protocol under which the study was conducted. We would like to thank the research participants and employees of 23andMe for making this work possible. The following members of the 23andMe Research Team contributed to this study: Stella Aslibekyan, Adam Auton, Elizabeth Babalola, Robert K. Bell, Jessica Bielenberg, Katarzyna Bryc, Emily Bullis, Daniella Coker, Devika Dhamija, Sayantan Das, Sarah L. Elson, Teresa Filshtein, Kipper Fletez-Brant, Pierre Fontanillas, Will Freyman, Pooja M. Gandhi, Karl Heilbron, Barry Hicks, David A. Hinds, Ethan M. Jewett, Yunxuan Jiang, Katelyn Kukar, Keng-Han Lin, Maya Lowe, Jey C. McCreight, Matthew H. McIntyre, Steven J. Micheletti, Meghan E. Moreno, Joanna L. Mountain, Priyanka Nandakumar, Elizabeth S. Noblin, Jared O'Connell, Aaron A. Petrakovitz, G. David Poznik, Morgan Schumacher, Anjali J. Shastri, Janie F. Shelton, Jingchunzi Shi, Suyash Shringarpure, Vinh Tran, Joyce Y. Tung, Wei Wang, Catherine H. Weldon, Peter Wilton, Alejandro Hernandez, Corinna Wong, Christophe Toukam Tchakouté, Gabriel Cuellar Partida, Xin Wang and 23andMe Research Team are employed by and hold stock or stock options in 23andMe, Inc.

**Atherosclerosis Risk in Communities (ARIC):** The Atherosclerosis Risk in Communities study has been funded in whole or in part with Federal funds from the National Heart, Lung, and Blood Institute, National Institutes of Health, Department of Health and Human Services, under Contract nos. (HHSN268201700001I, HHSN268201700002I, HHSN268201700003I, HHSN268201700004I, HHSN268201700005I). The authors thank the staff and participants of the ARIC study for their important contributions. Funding was also supported by R01HL087641, R01HL059367 and R01HL086694; National Human Genome Research Institute contract U01HG004402; and National Institutes of Health contract HHSN268200625226C. Infrastructure was partly supported by Grant Number UL1RR025005, a component of the National Institutes of Health and NIH Roadmap for Medical Research. Jack W. Pattee was supported by NIH T32GM108557.

**BioBank Japan (BBJ):** The BioBank was supported by the Tailor-Made Medical Treatment Program of the Ministry of Education, Culture, Sports, Science, and Technology and Japan Agency for Medical Research (AMED) under grant numbers JP17km0305002, JP17km0305001, JP20km0405209 and JP20ek0109487. S.K, K.I and I.K were funded by AMED under Grant Numbers JP20km0405209 and JP20ek0109487.

**BioME:** The BioMe Biobank is supported by the Andrea and Charles Bronfman Philanthropies.

**Cardiovascular Health Study:** This CHS research was supported by NHLBI contracts HHSN268201200036C, HHSN268200800007C, HHSN268201800001C, HHSN268200960009C, N01HC55222, N01HC85079, N01HC85080, N01HC85081, N01HC85082, N01HC85083, N01HC85086, 75N92021D00006; and NHLBI grants U01HL080295, R01HL085251, R01HL087652, R01HL105756, R01HL103612, R01HL120393, and U01HL130114 with additional contribution from the National Institute of Neurological Disorders and Stroke (NINDS). Additional support was provided through R01AG023629 from the National Institute on Aging (NIA). A full list of principal CHS investigators and institutions can be found at CHS-NHLBI.org. The provision of genotyping data was supported in part by the National Center for Advancing Translational Sciences, CTSI grant UL1TR001881, and the National Institute of Diabetes and Digestive and Kidney Disease Diabetes Research Center (DRC) grant DK063491 to the Southern California Diabetes Endocrinology Research Center.

The content is solely the responsibility of the authors and does not necessarily represent the official views of the National Institutes of Health.

**Early Onset Venous Thrombosis Study (EOVT):** No grants to acknowledge.
**eMERGE Consortium:** The eMERGE Network was initiated and funded by NHGRI through the following grants: U01HG006828 (Cincinnati Children's Hospital Medical Center/Boston Children's Hospital); U01HG006830 (Children's Hospital of Philadelphia); U01HG006389 (Essentia Institute of Rural Health, Marshfield Clinic Research Foundation and Pennsylvania State University); U01HG006382 (Geisinger Clinic); U01HG006375 (Group Health Cooperative); U01HG006379 (Mayo Clinic); U01HG006380 (Icahn School of Medicine at Mount Sinai); U01HG006388 (Northwestern University); U01HG006378 (Vanderbilt University Medical Center); and U01HG006385 (Vanderbilt University Medical Center serving as the Coordinating Center). *Group Health Cooperative/University of Washington:* Funding support for Alzheimer's Disease Patient Registry (ADPR) and Adult Changes in Thought (ACT) study was provided by a U01 from the National Institute on Aging (Eric B. Larson, PI, U01AG006781). A gift from the 3M Corporation was used to expand the ACT cohort. DNA aliquots sufficient for GWAS from ADPR Probable AD cases, who had been enrolled in Genetic Differences in Alzheimer's Cases and Controls (Walter Kukull, PI, R01 AG007584) and obtained under that grant, were made available to eMERGE without charge. Funding support for genotyping, which was performed at Johns Hopkins University, was provided by the NIH (U01HG004438). Genome-wide association analyses were supported through a Cooperative Agreement from the National Human Genome Research Institute, U01HG004610 (Eric B. Larson, PI). Assistance with phenotype harmonization and genotype data cleaning was provided by the eMERGE Administrative Coordinating Center (U01HG004603) and the National Center for Biotechnology Information (NCBI). *Marshfield Clinic Research Foundation:* Funding support for the Personalized Medicine Research Project (PMRP) was provided through a cooperative agreement (U01HG004608) with the National Human Genome Research Institute (NHGRI), with additional funding from the National Institute for General Medical Sciences (NIGMS) The samples used for PMRP analyses were obtained with funding from Marshfield Clinic, Health Resources Service Administration Office of Rural Health Policy grant number D1A RH00025, and Wisconsin Department of Commerce Technology Development Fund contract number TDF FYO10718. Funding support for genotyping, which was performed at Johns Hopkins University, was provided by the NIH (U01HG004438). Assistance with phenotype harmonization and genotype data cleaning was provided by the eMERGE Administrative Coordinating Center (U01HG004603) and the National Center for Biotechnology Information (NCBI). *Vanderbilt University:* Funding support for the Vanderbilt Genome-Electronic Records (VGER) project was provided through a cooperative agreement (U01HG004603) with the National Human Genome Research Institute (NHGRI) with additional funding from the National Institute of General Medical Sciences (NIGMS). The dataset and samples used for the VGER analyses were obtained from Vanderbilt University Medical Center's BioVU, which is supported by institutional funding and by the Vanderbilt CTSA grant UL1RR024975 from NCRR/NIH. Funding support for genotyping, which was performed at The Broad Institute, was provided by the NIH (U01HG004424). Assistance with phenotype harmonization and genotype data cleaning was provided by the eMERGE Administrative Coordinating Center (U01HG004603) and the National Center for Biotechnology Information (NCBI). *Geisinger Health System:* Samples and data in this obesity study were provided by the non-alcoholic steatohepatitis (NASH) project. Funding for the NASH project was provided by a grant from the Clinic Research Fund of Geisinger Clinic. Funding support for the genotyping of the NASH cohort was provided by a Geisinger Clinic operating funds and an award from the Clinic Research Fund. Samples and data in this study were provided by the abdominal aortic aneurysm (AAA) project. Funding for the AAA project was provided by a grant from the Clinic Research Fund of Geisinger Clinic. Funding support for the genotyping of the AAA cohort was

provided by a Geisinger Clinic operating funds and an award from the Clinic Research Fund. Samples and data in this study were provided by the Geisinger MyCode Project. Funding for the MyCode project was provided by a grant from Commonwealth of Pennsylvania and the Clinic Research Fund of Geisinger Clinic. Funding support for the genotyping of the MyCode cohort was provided by Geisinger Clinic operating funds and an award from the Clinic Research Fund. *Mount Sinai School of Medicine:* Samples and data used in this study were provided by the Mount Sinai School of Medicine (MSSM) Biobank Project funded by The Charles R. Bronfman Institute for Personalized Medicine (IPM) at Mount Sinai School of Medicine. The Coronary Artery Disease study (IPM BioBank GWAS) is a genome-wide association study funded by the Charles R. Bronfman Institute for Personalized Medicine. The datasets used for the analyses described in this manuscript were obtained from dbGaP at http://www.ncbi.nlm.nih.gov/gap through dbGaP accession number phs000888.v1.p1.

**Estonian Biobank**: This study was funded by EU H2020 grant 692145, Estonian Research Council Grant IUT20-60, IUT24-6, and European Union through the European Regional Development Fund Project No. 2014-2020.4.01.15-0012 GENTRANSMED and 2014-2020.4.01.16-0125. Data analyses were carried out in part in the High-Performance Computing Center of University of Tartu.

**FARIVE:** The FARIVE study was supported by Fondation pour la Recherche Médicale, Programme Hospitalier de Recherche Clinique (PHRC), Fondation de France, and the Leducq Foundation (LINAT Leducq International Network Against Thrombosis). FARIVE genetics research program was supported and funded by the GenMed LABEX (ANR-10-LBX-0013) and the French INvestigation Network on Venous Thrombo-Embolism (INNOVTE).

**FinnGen**: The FinnGen project is funded by two grants from Business Finland (HUS 4685/31/2016 and UH 4386/31/2016) and the following industry partners: AbbVie Inc., AstraZeneca UK Ltd, Biogen MA Inc., Bristol Myers Squibb (and Celgene Corporation & Celgene International II Sàrl), Genentech Inc., Merck Sharp & Dohme Corp, Pfizer Inc., GlaxoSmithKline Intellectual Property Development Ltd., Sanofi US Services Inc., Maze Therapeutics Inc., Janssen Biotech Inc, Novartis AG, and Boehringer Ingelheim. Following biobanks are acknowledged for delivering biobank samples to FinnGen: Auria Biobank, THL Biobank, Helsinki Biobank, Biobank Borealis of Northern Finland, Finnish Clinical Biobank Tampere, Biobank of Eastern Finland, Central Finland Biobank, Finnish Red Cross Blood Service Biobank and Terveystalo Biobank. All Finnish Biobanks are members of BBMRI.fi infrastructure. Finnish Biobank Cooperative -FINBB (https://finbb.fi/) is the coordinator of BBMRI-ERIC operations in Finland. The Finnish biobank data can be accessed through the Fingenious® services (https://site.fingenious.fi/en/) managed by FINBB.

**Framingham Heart Study (FHS):** Framingham Heart Study (FHS) was partially supported by the National Heart, Lung, and Blood Institute's (NHLBI's) Framingham Heart Study (Contract No. N01-HC-25195) and its contract with Affymetrix, Inc. for genotyping services (Contract No. N02-HL-6-4278). A portion of this research utilized the Linux Cluster for Genetic Analysis (LinGA-II) funded by the Robert Dawson Evans Endowment of the Department of Medicine at Boston University School of Medicine and Boston Medical Center. This project has been funded in whole or in part with Federal funds from the National Heart, Lung, and Blood Institute, National Institutes of Health, Department of Health and Human Services, under Contract No. 75N92019D00031. The analyses reflect intellectual input and resource development from the Framingham Heart Study investigators participating in the SNP Health Association Resource (SHARe) project. National Heart, Lung and Blood Institute Division of Intramural Research funds supported personnel involved in this work. The views expressed in this manuscript are those of the authors and do not necessarily represent the views of the National Heart, Lung and Blood Institute; the National Institutes of Health; or the U.S. Department of Health and Human Services.

**Genetic Analysis for Idiopathic Thrombophilia (GAIT)** project was supported partially by grants PI-11/0184, PI-14/0582 and Red Investigación Cardiovascular RD12/0042/0032 from the Spanish Health Institute (ISCIII), and acknowledges current support as a Consolidated Research Group from the Government of Catalonia and CERCA Program (2017 SGR 1736), and from the non-profit organization ”Associació Activa´TT per la Salut”. Maria Sabater-Lleal is supported by a Miguel Servet contract from the ISCIII Spanish Health Institute (CP17/00142) and co-financed by the European Social Fund.

**GBMI**: See supplement of GBMI flagship paper for contributing cohort funding acknowledgements (https://doi.org/10.1101/2021.11.19.21266436). W.Z. was supported by the National Human Genome Research Institute of the National Institutes of Health under award number T32HG010464. BNW and CJW were supported by the National Heart Lung Blood under award number R35HL135824.

**Heart and Vascular Health (HVH) VTE Study:** The HVH Study was supported by National Heart, Lung, and Blood Institute grants HL43201, HL60739, HL68986, HL73410, HL74745, HL85251, HL95080, and HL121414.

**HUNT:** The Nord-Trøndelag Health Study (The HUNT Study) is a collaboration between HUNT Research Center (Faculty of Medicine and Health Sciences, NTNU, Norwegian University of Science and Technology), Nord-Trøndelag County Council, Central Norway Regional Health Authority, and the Norwegian Institute of Public Health. Drs. Ben Brumpton and Kristian Hveem work in a research unit funded by Stiftelsen Kristian Gerhard Jebsen; Faculty of Medicine and Health Sciences, NTNU; The Liaison Committee for education, research and innovation in Central Norway; and the Joint Research Committee between St. Olavs Hospital and the Faculty of Medicine and Health Sciences, NTNU. The genotyping in HUNT was financed by the National Institute of Health (NIH); University of Michigan; The Research Council of Norway; The Liaison Committee for education, research and innovation in Central Norway; and the Joint Research Committee between St. Olavs Hospital and the Faculty of Medicine and Health Sciences, NTNU. The K.G. Jebsen Center for Genetic Epidemiology is financed by Stiftelsen Kristian Gerhard Jebsen, Faculty of Medicine and Health Sciences Norwegian University of Science and Technology (NTNU) and the Liaison Committee for education, research and innovation in Central Norway. Dr. Brumpton is financed by the Medical Research Council Integrative Epidemiology Unit at the University of Bristol which is supported by the Medical Research Council and the University of Bristol [MC_UU_12013/1].

**JUPITER**: The JUPITER trial and its genetic substudy were funded by AstraZeneca.

**MARTHA & MARTHA12:** The MARTHA and MARTHA12 projects were supported by grants from the Program Hospitalier de Recherche Clinique. MARTHA related genetics research programs were supported and funded by the GenMed LABEX (ANR-10-LBX-0013) and the French INvestigation Network on Venous Thrombo-Embolism (INNOVTE). David-Alexandre Trégouët was financially supported by the “EPIDEMIOM-VTE” Senior Chair from the Initiative of Excellence of the University of Bordeaux.

**Mass General Brigham Biobank (MGBB):** Pradeep Natarajan is supported by grants from the National Heart Lung and Blood Institute (R01HL142711, R01HL148050, R01HL151283,

R01HL127564, R01HL148565, R01HL135242, R01HL151152), National Institute of Diabetes and Digestive and Kidney Diseases (R01DK125782), Fondation Leducq (TNE-18CVD04), and Massachusetts General Hospital (Paul and Phyllis Fireman Endowed Chair in Vascular Medicine). Thomas Gilliland is supported by T32 grant 5T32HL125232 of the National Heart, Lung, and Blood Institute.

**Mayo VTE Study:** The research was funded, in part, by grants from the National Institutes of Health, National Heart, Lung, and Blood Institute (HL66216 and HL83141) and the National Human Genome Research Institute (HG04735, HG06379-07, and HG06379-08) and Mayo Foundation.

**Million Veteran Program:** The MVP is funded by the Department of Veterans Affairs Office of Research and Development, Million Veteran Program Grant #MVP000. This publication does not represent the views of the Department of Veterans Affairs or the United States Government. MVP was also supported by three additional Department of Veterans Affairs awards (I01-01BX03340, I01-BX003362, and I01-CX001025). Scott M. Damrauer is supported by the Veterans Administration (IK2-CX001780). Pradeep Natarajan is supported by the NIH/NHLBI K08HL140203 R01HL142711 and a Hassenfeld Award from the Massachusetts General Hospital.

**Multiple Environmental and Genetic Assessment of risk factors for VT study (MEGA):** The MEGA study was supported by Netherlands Heart Foundation (NHS 98.113), the Dutch Cancer Foundation (RUL 99/1992), the Netherlands Organisation for Scientific Research (912–03–033| 2003), and partially by the Laboratory of Excellence in Medical Genomics (GenMed LABEX ANR-10-LABX-0013). We would like to thank all colleagues from the French Centre National de Génotypage for the genotyping contribution

**Multi-Ethnic Study of Atherosclerosis (MESA):** The Multi-Ethnic Study of Atherosclerosis (MESA) and the MESA SHARe project are conducted and supported by the National Heart, Lung, and Blood Institute in collaboration with MESA investigators. Support for MESA is provided by contracts HHSN268201500003I, N01-HC-95159, N01-HC-95160, N01-HC-95161, N01-HC-95162, N01-HC-95163, N01-HC-95164, N01-HC-95165, N01-HC-95166, N01-HC-95167, N01-HC-95168, N01-HC-95169, UL1-TR-000040, UL1-TR-001079, UL1-TR-001420, UL1-TR-001881, and DK063491. Funding for SHARe genotyping was provided by National Heart, Lung, and Blood Institute contract N02-HL-64278. Genotyping was performed at Affymetrix (Santa Clara, CA) and the Broad Institute of Harvard and the Massachusetts Institute of Technology (Boston, MA) using the Affymetrix Genome-Wide Human SNP Array 6.0. Pamela Lutsey is supported by K24 grant HL159246 of the National Heart, Lung, and Blood Institute.

**Nurse's Health Study (NHS and NHS-II) and Health Professionals Follow Up Study:** The work for the current study was funded by grants from the NIH: P01CA87969, R01CA49449, R01HL034594, R01HL088521, R01CA50385, R01CA67262, P01CA055075, R01HL35464, R01HL116854.

**Penn Medicine Biobank (PMBB):** We thank the patient-participants of Penn Medicine who consented to participate in this research program. We would also like to thank the Penn Medicine BioBank team and Regeneron Genetics Center for providing genetic variant data for analysis. The PMBB is approved under IRB protocol# 813913 and supported by Perelman School of Medicine at University of Pennsylvania, a gift from the Smilow family, and the National Center for Advancing Translational Sciences of the National Institutes of Health under CTSA award number UL1TR001878.

**RETROVE:** The RETROVE study acknowledges support from the Government of Catalonia and CERCA Program (2017 SGR 1736), from the Spanish Health Institute (ISCIII grants PI12/000612, PI15/00269 and PI18/00434), and from the non-profit organization ”Associació Activa´TT per la Salut”. Maria Sabater-Lleal is supported by a Miguel Servet contract from the ISCIII Spanish Health Institute (CP17/00142) and co-financed by the European Social Fund. The genotyping service was carried out at CEGEN-PRB3-ISCIII; and it is supported by grant PT17/0019, of the PE I+D+i 2013-2016, funded by ISCIII and ERDF.

**Tromsø Study:** The Tromsø Study was supported by an independent grant from Stiftelsen Kristian Gerhard Jebsen in Norway (J.B.H.).

**UK Biobank:** UK Biobank has received funding from the UK Medical Research Council, Wellcome Trust, UK Department of Health, British Heart Foundation, Cancer Research UK, Diabetes UK, Northwest Regional Development Agency, Scottish Government, and Welsh Assembly Government. This research has been conducted using the UK Biobank Resource under Application number 40713.

**Women's Genome Health Study (WGHS):** The WGHS is supported by the National Heart, Lung, and Blood Institute (HL043851 and HL080467) and the National Cancer Institute (CA047988 and UM1CA182913), with funding for genotyping provided by Amgen.

**Women's Health Initiative (WHI):** The WHI program is funded by the National Heart, Lung, and Blood Institute, National Institutes of Health, U.S. Department of Health and Human Services through contracts 75N92021D00001, 75N92021D00002, 75N92021D00003, 75N92021D00004, and 75N92021D00005.
